# Macrophage-CD8⁺ T Cell Spatial Coupling Defines an Innate-Adaptive Injury Niche in Human Checkpoint Inhibitor Hepatotoxicity

**DOI:** 10.64898/2026.08.18.26360744

**Authors:** Jacob M. Bogdanov, Neil Zhao, Helia Alavifard, David E. Kleiner, Robert J. Fontana, Andrew A. Stolz, Akil Merchant, Jonathan Z. Sexton, Lily Dara

## Abstract

**Background & Aims:** Immune-mediated liver injury from immune checkpoint inhibitors (ILICI) is a major immune-related adverse event that limits cancer immunotherapy, yet its tissue-level immunobiology is poorly defined and its management is largely extrapolated from autoimmune hepatitis (AIH). We previously identified a tri-cellular CD8+ T cell-macrophage-hepatocyte injury niche in a murine model of ILICI; here, we tested whether this niche is recapitulated in human disease.

**Methods:** We applied imaging mass cytometry with a 32-marker panel to liver biopsies from patients with ILICI (n = 12), AIH as a disease comparator (n = 14), and healthy controls (n = 2), profiling approximately 297,000 single cells across 144 regions of interest with spatially resolved detection of apoptosis (cleaved caspase-3, cC3) and pyroptosis (cleaved gasdermin D, cGSDMD).

**Results:** We detected histiocyte-rich granulomas in ILICI consisting of macrophages and CD8+ T cells, including activated memory-effector subsets. Permutation-based spatial analysis identified CD8+ T cell-macrophage co-localization as the most frequent significant interaction in ILICI, organizing into integrated innate-adaptive cellular neighborhoods that concentrated cC3-and cGSDMD-positive cells. Descriptively, this contrasted with AIH, in which immune cells and stroma were more spatially compartmentalized. CD8+ T-cell and macrophage densities correlated with Ishak necroinflammation scores, jaundice, and granuloma formation.

**Conclusions:** These findings provide a single-cell spatial proteomic characterization of human ILICI in situ; they recapitulate the tri-cellular CD8-macrophage-hepatocyte niche we previously defined in a murine model and characterize ILICI as a spatially organized innate-adaptive inflammatory process, nominating myeloid signaling and CD8-macrophage interactions as candidate liver-directed targets to uncouple hepatotoxicity from anti-tumor immunity.

**Impact and implications:** This study provides a spatially resolved single-cell proteomic characterization of immune-mediated liver injury from checkpoint inhibitors (ILICI) in human tissue, revealing that macrophages and CD8⁺ T cells are not merely co-present but reproducibly co-localized into integrated inflammatory neighborhoods where apoptotic and pyroptotic cell death is concentrated. These findings are important for hepatologists and oncologists because they reframe ILICI as a spatially organized innate-adaptive process, distinct from autoimmune hepatitis, suggesting that current treatment strategies extrapolated from AIH may not optimally target the underlying pathobiology. Our results nominate myeloid signaling, inflammatory cell death pathways, and macrophage-CD8⁺ T cell interactions as candidate liver-directed therapeutic targets, offering a rationale for mechanism-based interventions that could uncouple hepatotoxicity from anti-tumor immunity and allow patients to remain on effective cancer immunotherapy.

## Introduction

The clinical efficacy of immune checkpoint inhibitor (ICI)-based cancer therapy has been one of the most significant therapeutic advances in modern oncology. By 2023 an estimated 56% of US patients with solid or hematologic malignancies were potentially eligible for some form of ICI treatment ^[1]^. The backbone of immunotherapy targets the cytotoxic T-lymphocyte-associated protein-4 (CTLA-4) and programmed cell death protein-1 (PD-1) pathways ^[2, 3]^, which tumors exploit to evade immune detection: programmed death-ligand 1 (PD-L1) is upregulated on tumor cells and within the tumor microenvironment, while CTLA-4 signaling restrains T-cell priming, co-opting endogenous pathways that evolved to limit self-reactivity and excessive inflammation in peripheral tissues ^[4, 5]^. Administered as monotherapy or in combination, anti-CTLA-4, anti-PD-1, and anti-PD-L1 agents exert their anti-tumor effect through global immune reprogramming, increasing T-lymphocyte clonal expansion, trafficking, and cytotoxicity ^[4–8]^.

The liver depends on these same inhibitory pathways to maintain immunotolerance ^[4–6]^, and ICI therapy has accordingly been associated with immune-related adverse events (irAEs) across nearly all organ systems, including immune-mediated liver injury from ICI use (ILICI) ^[7, 9–16]^. Aminotransferase elevations occur in over 30% of patients receiving combination ICI therapy, with clinically significant (grade 3-4) hepatotoxicity in a smaller subset, and treatment options remain limited to systemic immunosuppression and ICI discontinuation, to the detriment of patients’ oncologic care ^[10, 11]^. Uncoupling the tumoricidal effect of ICIs from ILICI is therefore of substantial clinical interest, yet this is a complex challenge in the liver, where tolerance is maintained by a diverse range of cells under intricate regulatory signaling ^[3–5, 17]^. Under physiologic conditions, hepatic T lymphocytes are exposed to constitutive co-inhibitory signals, including CTLA-4 on resident regulatory T cells and PD-L1 expression on dendritic cells, Kupffer cells, and liver sinusoidal endothelial cells ^[8, 10, 14, 18, 19]^. This constitutive tolerance is disrupted by ICIs, predisposing patients to aberrant inflammatory responses capable of causing severe liver injury ^[5, 9]^. An increase in the effector memory T-lymphocyte phenotype prior to treatment has been shown to predict the development of ILICI ^[11, 14, 20, 21]^, suggesting a predisposition in patients with chronic inflammation. Notably, although several host immune-response genes have been associated with ILICI ^[15]^, the human leukocyte antigen (HLA) alleles linked to autoimmune hepatitis are not over-represented, nor has an association with pre-existing autoantibodies been established^[15, 22, 23]^.

Aside from steroids and other systemic immunosuppressants^[22, 23]^, only tumor necrosis factor (TNF) blockade is currently under clinical investigation, and in a murine model of ILICI, it mitigated liver injury ^[24–28]^; however, anti-TNF use in liver disease is controversial ^[3]^. Few liver-specific therapeutic targets otherwise exist, underscoring the need for a liver-centric solution to what is fundamentally a global immune-manipulation problem. Treatment for ILICI is extrapolated from autoimmune hepatitis (AIH). Despite shared features, such as histologically dense immune infiltrates ^[29]^, they represent distinct disease processes ^[16, 30]^. ILICI presents with a predominantly hepatocellular injury pattern, with less severe aminotransferase elevations than AIH, infrequent anti-nuclear antibody positivity, normal gamma globulin levels, and a faster time to resolution ^[16, 30]^.

Pathologically, ILICI is characterized by lobular necrosis, mild-to-moderate portal inflammation, and abundant T lymphocytes and histiocytes with few plasma cells ^[16, 18, 22, 30–32]^. A frequent and distinguishing feature is the presence of microgranulomas containing aggregates of CD8+ cytotoxic T lymphocytes and activated histiocytes ^[11, 15, 18, 22]^, implicating innate-adaptive immune crosstalk in disease progression ^[33]^. Despite these observations, the spatial cellular architecture of human ILICI in situ remains poorly defined, limiting the identification of liver-specific therapeutic targets. We hypothesized that the tri-cellular CD8+ T cell-macrophage-hepatocyte injury niche we previously identified in a murine model of ILICI is conserved in human disease and distinguishes ILICI from AIH. To test this, we profiled liver biopsies from patients with ILICI, alongside AIH and healthy liver controls, using imaging mass cytometry to define the immune cell composition and spatial organization of human ILICI.

## Materials/Patients and methods

See *Supplementary Materials & methods* for full detail.

## Results

### ILICI manifests as hepatitis with histiocyte-rich granulomas

AIH is typically a more chronic, progressive, and frequently more severe hepatitis than the acute presentation of ILICI. To provide a comparator that still represented a distinct immune-mediated process, we selected AIH biopsies without advanced fibrosis (F0-F1) or overwhelming necrosis. The cohorts differed in demographics: AIH patients were typically younger Hispanic females (mean age 44 y, 86% Hispanic), whereas ILICI patients were mostly older Caucasian males (mean age 57 y, 92% Caucasian) (Fig. 1A). AIH patients were more frequently jaundiced (71% vs 25%) and had more frequent serum autoantibodies (ANA 86% vs 0%; anti-smooth-muscle 57% vs 8%) (Fig. 1A). Both cohorts were nearly universally treated with glucocorticoids; all-cause mortality was higher in the ILICI cohort (50% vs 0%), though only one death was liver-related (Fig. 1A). AIH patients presented with higher aminotransferases and bilirubin, whereas ILICI patients had higher serum alkaline phosphatase (p < 0.05) and lower serum immunoglobulins (IgG p < 0.001) at onset (Fig. 1B).

**Fig. 1.**
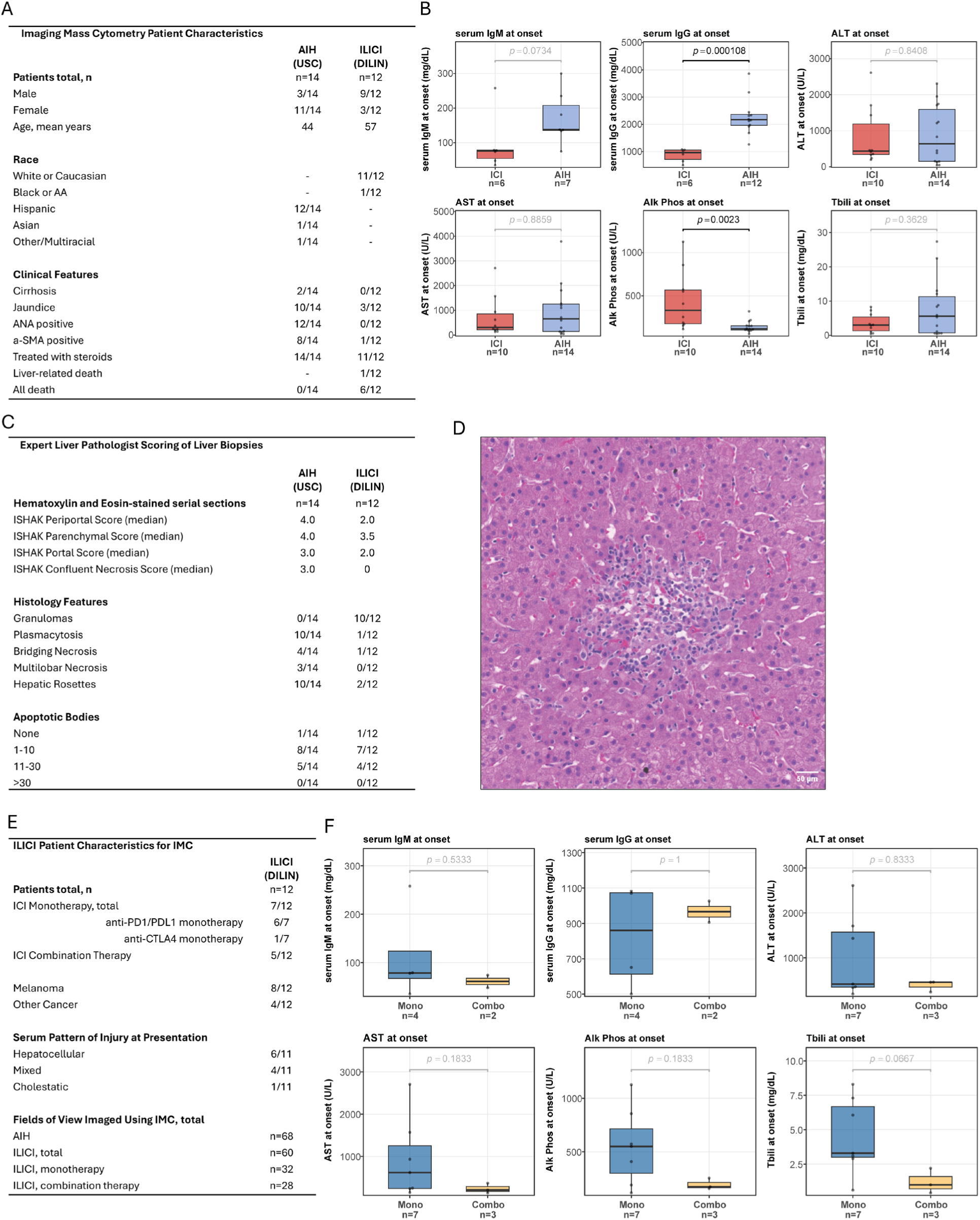
In situ characterization of human immune-mediated liver injury from immune checkpoint inhibitors (ILICI), with autoimmune hepatitis (AIH) as a disease comparator. (A) Summary of patient clinical characteristics. (B) Serum biochemistry profiles at injury onset for ILICI and AIH patients. (C) Summary of histological features in all included biopsies as evaluated by expert liver pathologist (DK). (D) Representative hematoxylin and eosin-stained (H&E) section of human liver biopsy showing focus of injury in ILICI. (E) ILICI patient characteristics for imaging mass cytometry (IMC) analysis including ICI therapy regimen breakdown and total fields of view (FOVs) imaged per group. (F) Serum laboratory values at injury onset stratified by ICI therapy subgroup (Mono = monotherapy; Combo = combination therapy). P-values in (B) and (F) by Wilcoxon rank-sum test. *Abbreviations: Alk Phos, alkaline phosphatase; ALT, alanine aminotransferase; AST, aspartate aminotransferase; Tbili, total bilirubin*.

H&E-stained sections from the biopsies (nILICI = 12; nAIH = 14) were graded by an expert liver pathologist (D.K.). AIH biopsies had higher median Ishak severity scores than ILICI biopsies (Fig. 1C). In ILICI, the highest necroinflammatory activity localized to spotty parenchymal (zone 2) foci rather than portal/periportal zones (Fig. 1C), and apoptotic-body counts were similar between cohorts (Fig. 1C). Plasmacytosis and hepatic rosettes, classical features of AIH, were common in AIH but rare in ILICI (Fig. 1C). Most notably, histiocyte-rich granulomas were present in 10 of 12 ILICI biopsies but none of the AIH biopsies (Fig. 1C,D), consistent with the histiocyte-rich granulomatous hepatitis described for ILICI.

Serial sections from these biopsies were imaged by IMC, yielding 144 regions of interest (ROIs; nILICI = 60, nAIH = 68, nCTRL = 16) (Fig. 1E). Within ILICI, we also compared ICI monotherapy and combination therapy (mono = 32 ROIs; combo = 28 ROIs); among monotherapy patients, six of seven received anti-PD-1/PD-L1 and one received anti-CTLA-4 (Fig. 1E). Monotherapy patients had non-significantly higher mean aminotransferases at onset than combination-therapy patients (Fig. 1F). Sections were stained with the 32-marker panel (Table S-2), and after acquisition and preprocessing (Fig. S-4), single-cell segmentation enabled phenotyping of every detected cell within each ROI (Fig. S-3). Single-cell features were then used for unsupervised cellular-neighborhood and spatial-interaction analyses (see supplemental methods).

### Single-cell proteomics maps CD8+ T cells and myeloid cells within intact liver architecture in ILICI

A maximum-intensity projection of all protein channels reconstructed the tissue image of a representative ILICI ROI, illustrating preservation of liver microarchitecture by IMC, with paired nuclear and cytoplasmic segmentation masks resolving parenchymal and non-parenchymal cells (Fig. 2A). Mean marker intensities across all 144 ROIs are shown per biopsy (Fig. 2B). Semi-supervised phenotyping identified 15 coarse cell classes, each with a distinct expression signature (Fig. 2C); finer classification, including T-lymphocyte subsets and cell-death markers (cC3, cGSDMD), resolved up to 23 subclasses (Table S-4). Spatial maps revealed discrete immune aggregates in ILICI composed of intermixed CD8+ T lymphocytes and macrophages (Fig. 2D; Fig. S-3). Per-ROI densities are shown across samples (Fig. 2E), with mean cell type densities and proportions summarized by disease group (Fig. 2F,G). In these ROI-level overviews, AIH differed from control in both density and proportion (p ≤ 0.001; Fig. 2F,G).

**Fig. 2.**
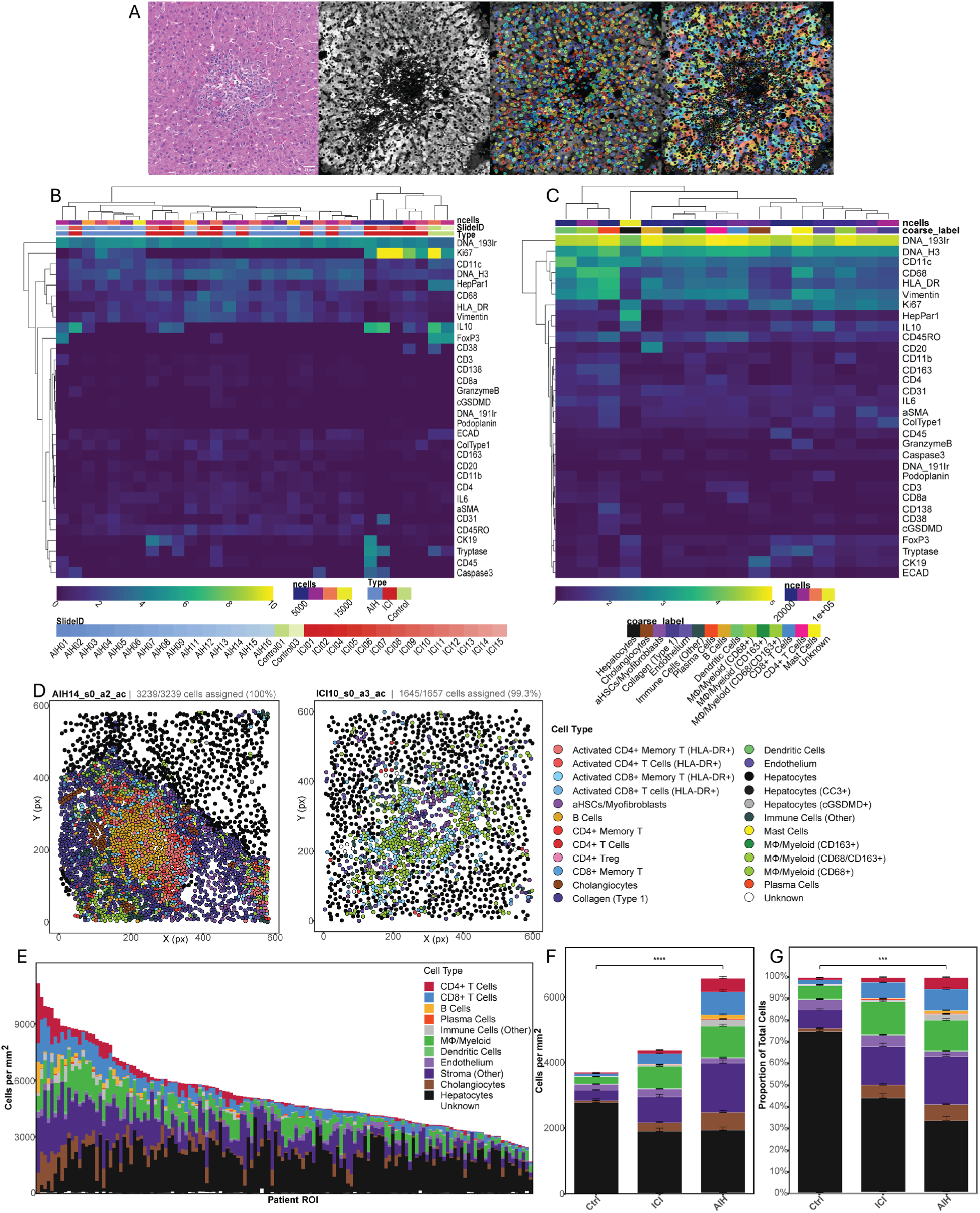
Single-cell proteomics characterization of regions-of-interest ROIs (*n_Total_ = 144*) within preserved tissue microarchitecture. (A) Representative IMC workflow. *From left to right:* H&E-stained adjacent serial section, pseudocolored multiplexed IMC channel composite, and overlaid nuclear and cell boundary segmentation masks. (B–C) Hierarchically clustered heatmaps depicting number of cells detected and mean signal intensity for all protein markers per slide (B) and per identified cell class (C). Annotation bars indicate cell count (ncells), slide identity and disease group (B) or cell class label (C). (D) Two-dimensional spatial plotting of representative (*left*) AIH and (*right*) ILICI ROIs demonstrating resolution of single-cell phenotyping. Pseudo-colored by granular cell class label. Cell class assignment percentages shown above each panel. (E) Stacked bar chart describing phenotypic composition of each ROI by cell density (cells/mm^2^), sorted by descending total density. (F) Mean cell class densities (cells/mm²) by disease group. (G) Mean cell class proportions by disease group. Omnibus significance in (F) and (G) by Kruskal–Wallis test on ROI-level totals. \*\*\*\**p < 0.0001*, \*\*\**p < 0.001*. Error bars indicate standard error (SE).

### ILICI is enriched in macrophages and CD8+ T cells, including memory-effector CD8+ subsets

Cell-class proportions across the three groups are shown in Fig. 3A (Fig. S-5). Hepatocyte proportion was most reduced in AIH, with at least one significant pairwise difference among the three groups (p ≤ 0.05; Fig. 3A). Stromal cells were higher in ILICI (∼15%) and AIH (∼22%) than control (∼8%), reaching significance only for AIH versus control (p < 0.05; Fig. 3A). AIH had significantly greater CD4+ T-cell proportions than ILICI (∼5-fold; p < 0.001) and control (p < 0.05), whereas ILICI and control were similar (∼1%; Fig. 3A). B cells were also enriched in AIH versus ILICI (p < 0.0001) and control (p < 0.05) and explained the largest share of between-group variance among all cell types (marginal R^2^ = 0.22; Fig. 3A). CD8+ T-cell proportions were ∼3-fold higher in ILICI than control (∼6% vs ∼2%; p < 0.05), and highest in AIH (∼10%; AIH vs control p < 0.01; Fig. 3A). Macrophage/myeloid proportion was numerically greatest in ILICI (∼16%) versus AIH (∼13%) and control (∼6%), but this difference was not statistically significant (χ^2^_Wald_(2) = 4.59, p = 0.101; Fig. 3A). Endothelial-cell proportions are also reported in Fig. 3A; however, because CD31 staining failed across all AIH ROIs, ILICI-versus-AIH endothelial comparisons are not interpreted here.

**Fig. 3.**
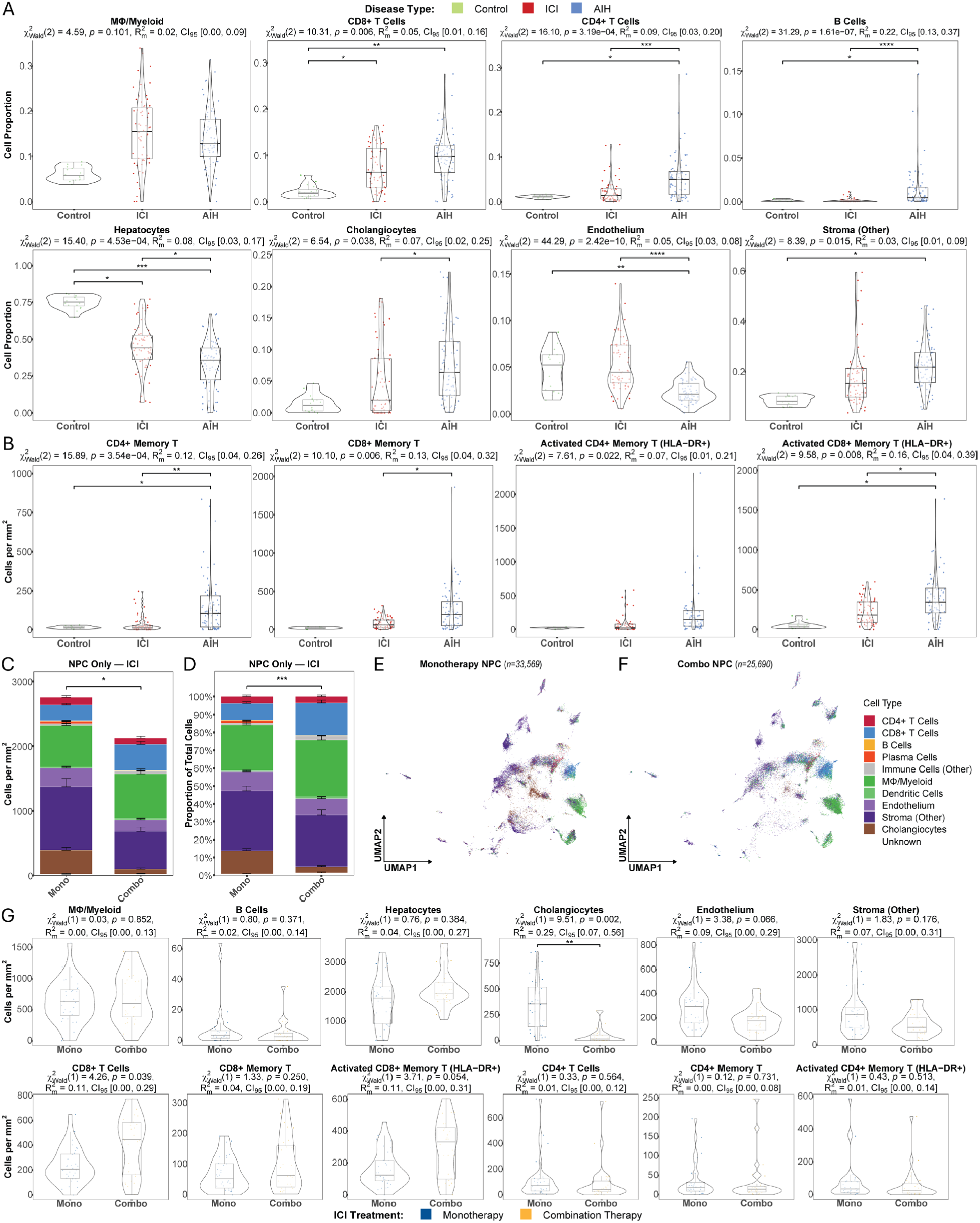
Single-cell phenotyping identifies dynamic cell class proportions across treatment conditions. (A) Combined box and violin plots quantifying designated cell class proportions by treatment condition. Each dot corresponds to a single ROI. (B) Cell densities (cells/mm²) for memory and activated T cell subclasses by treatment condition. (C-D) Stacked bar chart summarizing phenotypic composition in ICI monotherapy vs. combination therapy samples by cell density (cells/mm^2^) (C) and proportion (D). Pseudo-colored by cell type label, hepatocytes excluded. (E-F) Dimensionality reduction plotting using UMAP of all single-cell morphometrics quantified in (E) ICI monotherapy (*n_mono_* = 33,569 cells) and (F) combination therapy (*n_combo_* = 25,690 cells) samples. Pseudo-colored by cell class, hepatocytes excluded. (G) Combined box and violin plots quantifying designated cell class density (cells/mm^2^) in ICI monotherapy vs. combination therapy samples. Each panel in (A), (B), and (G) reports Wald χ² test statistic (with degrees of freedom), *p*-value, marginal R² (R²m), and 95% confidence interval from GLMMs with patient-level random intercepts. Pairwise comparisons by estimated marginal means with Benjamini–Hochberg correction. *Disease Type*: green = Control; red = ILICI; blue = AIH. *ICI Treatment*: blue = Monotherapy; gold = Combination Therapy. Omnibus significance in (C) and (D) by Kruskal–Wallis test on ROI-level totals. Error bars indicate SE. \*\*\*\**p < 0.0001*, \*\*\**p < 0.001*, \*\**p < 0.01*, \**p < 0.05*. *Abbreviations*: GLMM, generalized linear mixed-effects model; NPC, non-parenchymal cell; UMAP, Uniform Manifold Approximation and Projection.

We next quantified memory-effector T-lymphocyte (TEM) subsets previously implicated in ILICI ^[21]^ (Fig. 3B). Densities of CD4+ memory (CD45RO+) and activated CD4+ memory-effector (HLA-DR+) T cells were low and comparable between control and ILICI, while AIH showed ∼6-7-fold enrichment of both (CD4+ memory, p < 0.001; activated CD4+ memory, p < 0.05; Fig. 3B). CD8+ memory and activated CD8+ memory-effector T-cell densities were moderately higher in ILICI than control (CD8+ memory 62 vs 19 cells/mm^2^; activated CD8+ memory 183 vs 40 cells/mm^2^), though these pairwise differences were not significant after correction. AIH had the highest densities of both CD8+ memory subsets (198 and 346 cells/mm2) and differed significantly from ILICI and/or control (CD8+ memory, p < 0.01; activated CD8+ memory, p < 0.01; Fig. 3B).

Composition also differed between ICI monotherapy and combination therapy (Fig. 3C,D). UMAP embeddings of non-parenchymal cells showed qualitative differences between subgroups, including a larger CD8+ T-cell cluster and reduced cholangiocyte representation in combination therapy (Fig. 3E,F). Combination therapy had significantly higher CD8+ T-cell density than monotherapy (445 vs 208 cells/mm2, p = 0.039; Fig. 3G). Activated CD8+ TEM density was numerically about twice as high in combination therapy (332 vs 124 cells/mm^2^) but did not reach significance (Fig. 3G). Combination therapy was associated with significantly lower cholangiocyte density, which explained the largest between-group variance among cell types tested (R^2^ = 0.29; p = 0.002; Fig. 3G). Endothelial-cell density was ∼40% lower in combination therapy (p = 0.066; proportion p = 0.009; Fig. 3G); because both subgroups are ILICI and ran in the same batch, this within-ILICI endothelial comparison is not affected by the AIH CD31 failure. No further significant differences were detected between subgroups, including among CD4+ T cells and myeloid cells.

### Spatial analysis reveals distinct immune organization in ILICI and AIH

To interrogate spatial organization, permutation-based pairwise interaction analysis tested cell-cell attraction and avoidance among all cell classes in ILICI and AIH (Fig. 4A,B; Fig. S-6). The summed signed interaction score (Sigma-sigval) is the number of ROIs in which a pair reached significance within a disease group. In ILICI, the most frequently significant pairwise attraction was between CD8+ T cells and macrophages (Fig. 4A). ILICI also showed significant attraction among a broader set of classes, including myeloid cells, dendritic cells, and lymphocyte subsets, whereas AIH interactions were dominated by CD4+ T-cell and B-cell attraction, with fewer cross-compartment (innate-adaptive) interactions reaching significance (Fig. 4B).

**Fig. 4.**
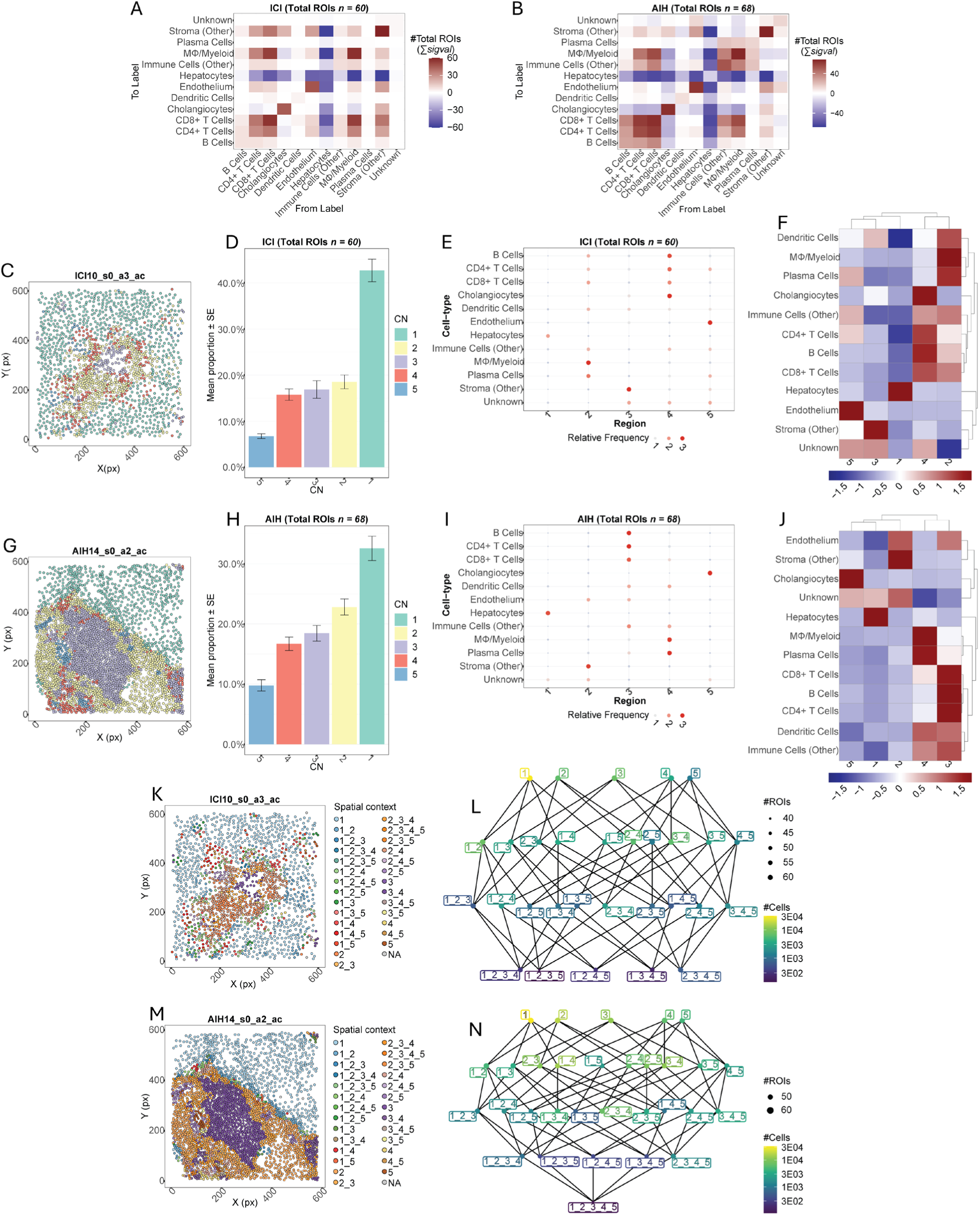
Computational spatial analysis reveals distinct spatial relationships and cellular neighborhoods in ILICI and AIH. (A-B) Pairwise cell-cell expansion interaction heatmaps for (A) ILICI (*n =* 60) and (B) AIH (*n =* 68) ROIs. Sigval denotes the number of ROIs in which each pairwise spatial relationship is statistically significant. Red indicates spatial attraction; blue indicates spatial avoidance. (C, G) Two-dimensional spatial plots of representative (C) ILICI and (G) AIH ROIs with all single cells pseudocolored by their assigned cellular neighborhood. (D, H) Mean proportion (± SE) of cells assigned to each CN for (D) ILICI and (H) AIH. (E, I) Region maps denoting cell class composition of five distinct cellular neighborhoods (CN) detected using unsupervised Delaunay triangulation (k=5) in (E) ILICI samples and (I) *AIH* samples independently. Dot size indicates relative frequency of cell class by CN. (F, J) Heatmaps quantifying cell class enrichment z-scores per CN for (F) ILICI and (J) AIH samples. (K, M) Representative (K) ILICI and (M) AIH ROIs with all single cells pseudocolored by assigned spatial context node (unique combination of CN memberships). (L, N) Network graphs of spatial context nodes summarizing inter-CN organization across (L) ILICI and (N) AIH ROIs. Node labels indicate CN combinations. Node size indicates number of ROIs containing that context. Node color indicates total cell count in that context (log-scale).

Cellular neighborhoods (CNs) were then identified by clustering cells on the cell-type composition of their spatial neighbors, with five CNs per disease group numbered by descending cell count (Fig. 4C-F, G-J; Fig. S-7). CNs were derived separately within ILICI and within AIH, so neighborhood identities are not directly equivalent across diseases and the comparison below is descriptive. In ILICI, the dominant immune neighborhood (CN2) was enriched for both adaptive and innate immune cells (CD8+ T cells, macrophages, and dendritic cells), indicating intermixing of these compartments across ILICI ROIs, while a less prevalent CN (CN4) contained lymphocytes and cholangiocytes consistent with portal-tract localization (Fig. 4C-F). In AIH, by contrast, immune neighborhoods were more segregated: one CN was enriched for adaptive immune cells (CD4+ T cells, B cells, plasma cells) and a separate CN for myeloid cells, while the largest non-hepatocyte CN was dominated by stromal cells including myofibroblasts (Fig. 4G-J), consistent with the greater fibrosis and chronicity seen on histology (Fig. 1C; Table S-1).

Spatial-context analysis, which labels each cell by the CN identities of its neighbors, showed that in ILICI the largest higher-order contexts contained cells simultaneously bordered by both innate and adaptive immune neighborhoods, whereas in AIH immune and stromal neighborhoods were more separated (Fig. 4K-N; Fig. S-7). Together, these descriptive analyses indicate that macrophages and CD8+ T cells are spatially intermixed at sites of injury in ILICI, in contrast to the more compartmentalized immune architecture of AIH.

### Spatial mapping of apoptotic and pyroptotic cell death in ILICI and AIH

The antibody panel included cleaved caspase-3 (cC3) and cleaved N-terminal gasdermin D (cGSDMD), the activated executioner proteases of apoptosis and pyroptosis. In representative images, cC3+ cells were prominent along the interface between intact parenchyma and dense innate-adaptive infiltrates in regions of hepatocyte dropout (Fig. 5A). Cells positive for cC3 and/or cGSDMD were quantified across all ROIs (Fig. 5A) and mapped by cell type, CN address, and death state in alluvial diagrams (Fig. 5B,C). Cell death marker-positive cells comprised 1.1% of detected cells in ILICI (1,172/104,412) and 0.7% in AIH (1,183/171,106).

**Fig. 5.**
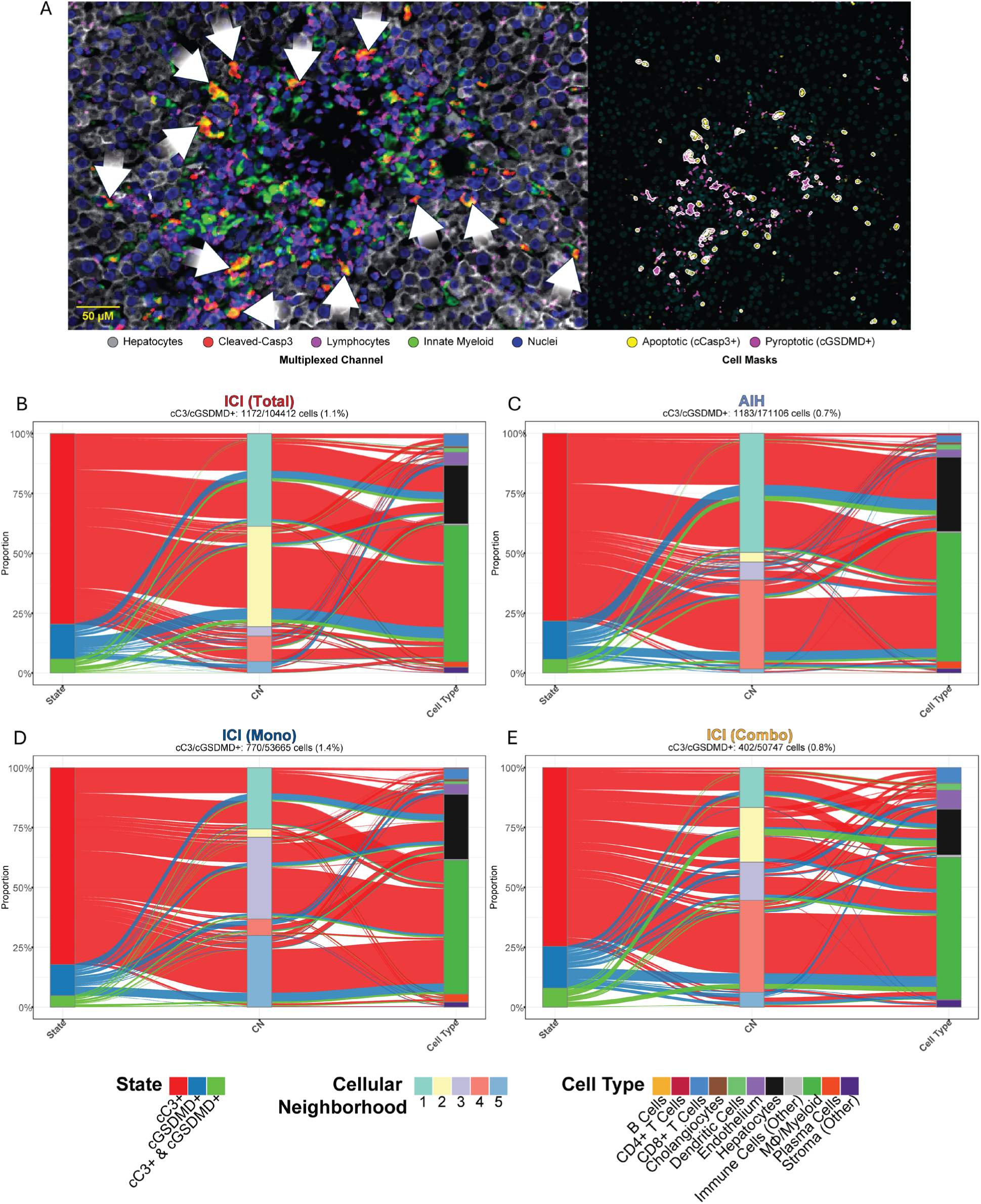
Spatial proteomics applied to map cC3-and cGSDMD-positive events in ILICI and AIH. (A) (*left*) Multiplexed image of a representative ILICI ROI pseudocolored to identify innate and adaptive immune activity and active apoptosis (cCasp3+) occurring in-situ. White arrows denote apoptotic cells on the front of hepatic injury. (*right*) Cell segmentation mask depicting all detected single cells dying by apoptosis (cCasp3+) and pyroptosis (N-terminal cleaved-GSDMD) in same representative ILICI ROI. *Scale bar* = 50 μm. (B-C) Alluvial diagrams characterizing the cell class identity, cellular neighborhood location, and mode of cell death for all detected cC3-and cGSDMD-positive cells in (B) ILICI total (*n =* 60) and (C) AIH (*n =* 68) ROIs. (D-E) Alluvial plots comparing single-cell apoptotic and pyroptotic events detected in (D) ICI monotherapy (*n =* 32) and (E) ICI combination therapy (*n =* 28) ROIs. *State*: red = cC3+; blue = cGSDMD+; green = cC3+ & cGSDMD+ co-positive. *CN:* colored 1–5*. Cell Type:* colored by aggregated cell type identity.

In both diseases, apoptosis (cC3+) predominated, with pyroptotic (cGSDMD+) events accounting for less than a quarter of the total (Fig. 5B,C), and macrophages/myeloid cells were the largest population of cC3/cGSDMD+ cells. A larger proportion of cell death marker-positive hepatocytes was seen in AIH and of CD8+ T cells in ILICI; endothelial events appeared more frequently in ILICI, but this is confounded by the AIH CD31 failure and is not interpreted across groups. Neither total cell-death events nor their compositional distribution differed significantly between ILICI and AIH when formally tested. Within ILICI, cGSDMD positivity was detected mainly among myeloid cells, hepatocytes, and endothelial cells, while CD8+ T cells were nearly exclusively cC3+ (Fig. 5B).

Mapping cC3+/cGSDMD-positive events onto neighborhoods, the integrated immune neighborhood (CN2) contained the largest share of cell-death events in ILICI despite having fewer than half the cells of the largest neighborhood (CN1); this may partly reflect that immune-rich neighborhoods favor foci detection (Fig. 4D; Fig. 5B). Pyroptotic events in CN2 were predominantly myeloid, while CN1 contained pyroptotic hepatocytes and CN5 pyroptotic endothelial cells (Fig. 5B). By therapy subgroup (Fig. 5D,E), combination therapy showed a greater proportion of cGSDMD+ events and cC3+ CD8+ T cells, and a lower proportion of hepatocyte death events than monotherapy. cC3+/cGSDMD+ co-positive cells were observed only in the combination-therapy group. However, combination therapy had fewer total marker-positive events and a lower overall event fraction (0.8%, 402/50,747) than monotherapy (1.4%, 770/53,665), and neither the total number nor the composition of events differed significantly between subgroups.

Across ILICI disease and therapy comparisons, cC3/cGSDMD-positive events shared several consistent features: apoptosis predominated, myeloid cells were the majority of marker-positive cells, and death events concentrated within the integrated innate-adaptive neighborhood where there was hepatocyte loss and CD8+ T cells and macrophages co-localize. Although the compositional differences between ILICI and AIH and between therapy subgroups were not statistically significant, the spatial concentration of cell death within immune-rich neighborhoods was consistent across ILICI samples.

### IMC-derived immune densities correlate with histological and clinical measures of injury in ILICI

We next related IMC-derived features to clinical and histopathological metadata (Fig. 6A; Fig. S-8). By DILIN severity grade, only cholangiocyte (χ^2^_Wald_(2) = 3.22, p = 0.073) density differed significantly, both highest in the pooled moderate/severe group; the immune populations emphasized here; CD8+ T cells, macrophages, and cell-death events, did not differ significantly by severity grade (Fig. 6A). At a descriptive level, mild cases had the highest CD8+ T-cell (including activated CD8+ memory-effector) density, while moderate/severe cases had the highest myeloid density and the highest densities of death marker-positive hepatocytes and total death events (Fig. 6A; Fig. S-8).

**Fig. 6.**
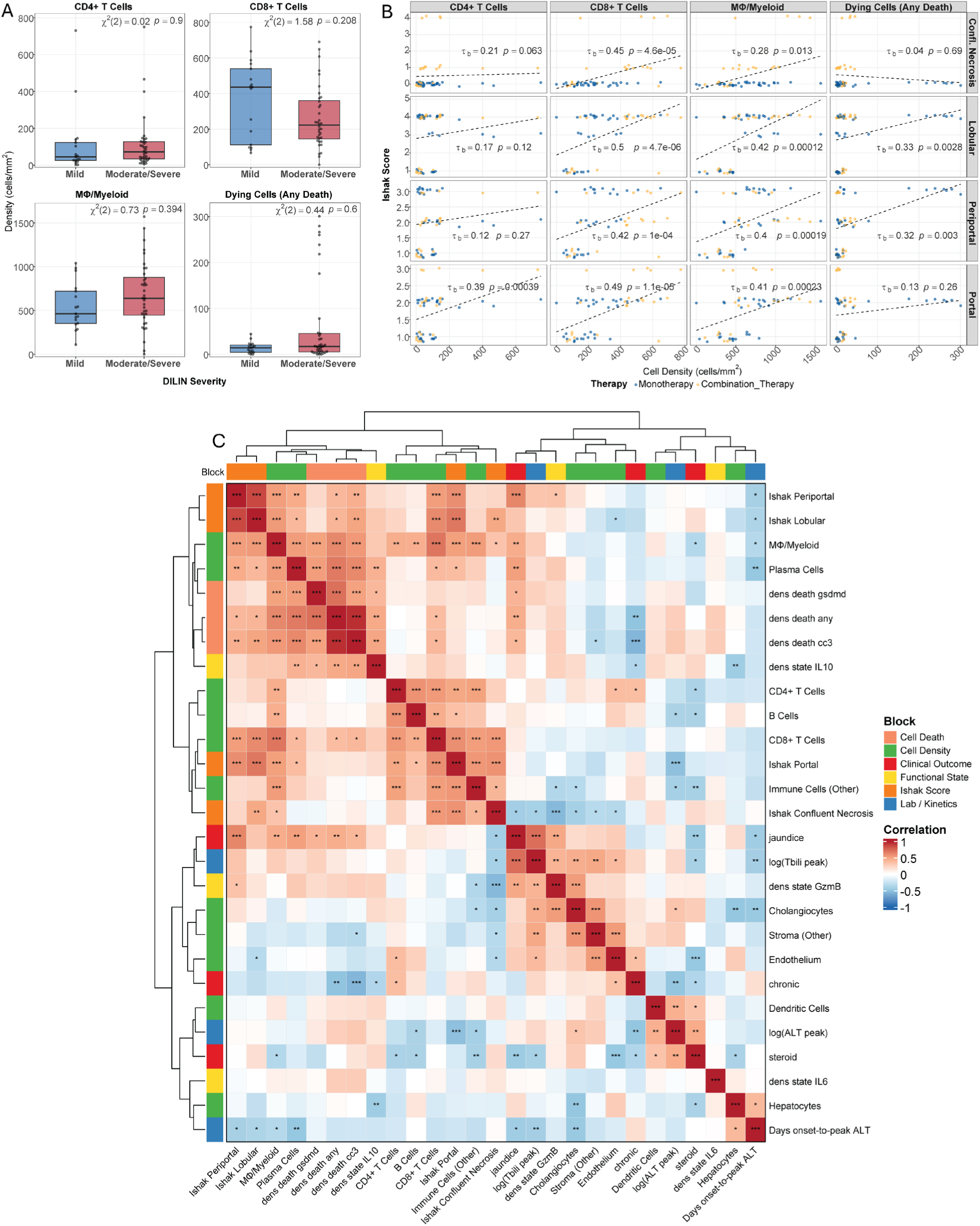
IMC immunophenotyping and spatial analysis correlates with clinical and histological features of ILICI. (A) Boxplots quantifying per-ROI cell densities (cells/mm²) for CD4+ T cells, CD8+ T cells, MΦ/Myeloid, and dying cells (any death marker) stratified by DILIN severity grade (Mild, Moderate, Severe). Wald χ² and *p*-values by LMM with patient-level random intercepts. (B) Scatter plot ranking correlations between IMC-detected densities of cell classes indicated in (A) with Ishak necroinflammation, or modified histology activity index (HAI) scoring by expert liver pathologist (DK). Each dot corresponds to single ILICI ROI, pseudo-colored by therapy subgroup. Correlation coefficients and *p*-values by Kendall’s *τ_b*. Dashed lines indicate linear trend. (C) Hierarchically clustered focused mixed-type correlation heatmap (Spearman, rank-biserial, phi) integrating IMC-derived, clinical, and histological variables in ILICI samples (*n_total_ =* 53 ROIs, 12 ILICI patients). Significance asterisks reflect Benjamini–Hochberg-adjusted *p*-values. Block annotations indicate variable category: Cell Death (*salmon*), Cell Density (*green*), Clinical Outcome (*pink*), Functional State (*light pink*), Ishak Score (*purple*), Lab/Kinetics (*yellow*). \*\*\**p* < 0.001, \*\**p* < 0.01, \**p* < 0.05. *Abbreviations*: DILIN, Drug-Induced Liver Injury Network; LMM, linear mixed-effects model.

By contrast, Ishak necroinflammation scores correlated consistently with IMC densities (Fig. 6B; Fig. S-9). CD8+ T-cell density correlated with all four Ishak scores (τ-b = 0.42-0.50, all p ≤ 0.001), as did macrophage/myeloid density (τ-b = 0.28-0.42, p ≤ 0.013). CD4+ T-cell density correlated selectively with portal inflammation (τ-b = 0.39, p < 0.001) but not significantly with lobular, periportal, or confluent necrosis scores. Total cell-death density correlated with lobular (τ-b = 0.33) and periportal (τ-b = 0.32) scores but not with confluent necrosis or portal scores (Fig. 6B; Fig. S-9).

An exploratory correlogram integrating 210 variables identified a large number of significant pairwise correlations (BH-adjusted FDR < 0.05; Table S-1; Fig. S-1). Among a focused set of ILICI ROI-level variables (Fig. 6C), macrophage/myeloid and CD8+ T-cell densities were positively correlated with jaundice, granulomas, higher Ishak scores, and cell-death density. Peak serum bilirubin correlated with densities of plasma cells, macrophages, cC3+ cells, and total death events, and with Ishak periportal score and ductular reaction (Fig. 6C). Clinical jaundice correlated with cC3+ and Granzyme-B+ cell densities and with histological apoptotic bodies and ductular reaction. Functional-state marker densities (IL-6, IL-10, Ki67) correlated with cell-death density, whereas stromal density was inversely correlated with it.

Collectively, these correlations indicate that IMC-derived immune densities and cell-death events track established measures of liver-injury severity in ILICI, with CD8+ T-cell and macrophage density co-associating with necroinflammatory activity, jaundice, and granuloma formation across clinical, histological, and proteomic readouts.

## Discussion

ILICI remains a major clinical barrier to effective cancer immunotherapy, yet its tissue-level immunobiology has remained incompletely defined. By integrating high-dimensional spatial proteomics in human liver biopsies with our prior mechanistic work in a murine model ^[8]^, we find that human ILICI is spatially organized around integrated innate-adaptive immune niches rather than a purely cytotoxic CD8+ T cell process. We compared ILICI to AIH in this manuscript. Although an imperfect comparator, AIH is the closest clinically relevant immune-mediated hepatitis for which liver biopsies are available and reflects current clinical management, where ILICI is largely treated by analogy to AIH.

A central feature of this architecture is the spatial coupling of macrophages and CD8+ T cells in ILICI. Our protein-level data in the DILIN patients confirm our findings in mice^[8]^, and indicate that CD8+ T cells reside within a microenvironment enriched for macrophages and Kupffer cells, and spatial modeling shows that these populations are not merely co-present but consistently co-localized across multiple patient biopsies, forming reproducible niches enriched for both adaptive and innate immune cells. Descriptively, this contrasts with AIH, in which immune populations appear more spatially compartmentalized, with CD4+ T cells and B cells predominating alongside stromal fractions.

Our findings converge with those of a concurrent, independent study. Uzun and colleagues recently reported a multimodal atlas of ILICI in 35 patients, combining multiplex immunofluorescence, bulk T-cell receptor sequencing, and spatial transcriptomics^[34]^. In that cohort, CD8+ T cells and myeloid/macrophage populations were the only cell types whose density and fraction increased significantly relative to uninflamed liver, and CD8+ cytotoxic T cells aggregated into patches in which they interacted preferentially with CD68+CD163Hi and CD68+CD163Lo macrophages. Using paired liver and tumour TCR sequencing, they further identified T-cell clones shared between the two sites in every patient examined, predominantly with a cytotoxic CD8+ phenotype, providing in situ evidence that tumour-reactive clones traffic to the liver in ILICI. That two independent cohorts, profiled on different platforms in different countries, converge on the same architectural feature substantially strengthens the inference that macrophage-CD8+ T cell coupling is a reproducible property of human ILICI rather than a cohort-specific observation; in our case that architecture was predicted a priori from our murine model^[8]^.

Our study addresses three questions that the concurrent work leaves open. First, that study did not include markers of regulated cell death, so neither the mode nor the location of hepatocyte and immune cell death was assessed; we localize apoptotic and pyroptotic pathway activation to the integrated innate-adaptive neighborhood. Second, its comparator was histologically uninflamed liver from untreated patients, which establishes that these features distinguish ILICI from normal liver but not from other immune-mediated hepatitides; our AIH comparator addresses that question directly, and the granulomatous phenotype we observe in 10 of 12 ILICI biopsies and none of 14 AIH biopsies is not accounted for by an antigen-restricted adaptive mechanism. Third, we relate spatial features to formal histopathological grading and to clinical severity. Notably, the tumor-shared T-cell clones that are the focus of that analysis accounted for 0.4-2.58% of the hepatic T-cell receptor repertoire, indicating that the hepatic infiltrate is predominantly polyclonal; the mechanisms that recruit and sustain the remainder of the infiltrate, and that execute hepatocyte death, remain undefined, and are the processes to which our data speak.

These observations are concordant with our prior murine work, in which hepatocyte apoptosis and NLRP3 inflammasome activation were linked through interactions between CD8+ T cells and myeloid cells ^[8]^. In that model, apoptotic hepatocytes were spatially associated with macrophages and NLRP3-high myeloid cells, and zones of three-way interaction between CD8+ T cells, macrophages, and dying hepatocytes were observed. The current human data recapitulate this architecture at single-cell resolution, consistent with a tri-cellular inflammatory niche that is conserved between the murine model and human disease. While traditional immunohistochemistry has identified the non-specific presence of CD8+ T cells, macrophages in patients with liver injury^[35]^, our spatially resolved, single-cell data detect an architecture invisible to compartment-level immunohistochemistry: in ILICI, macrophages and CD8+ T cells are not merely co-present but reproducibly co-localized into integrated neighborhoods that concentrate cell-death pathway markers.

These patterns suggest that CD8+ T-cell activation in ILICI may occur in a different immunologic context than in AIH. Recent peripheral immunophenotyping identified an exhausted circulating CD8+ effector memory signature specific to ILICI^[32]^. In AIH, the enrichment of CD4+ T cells and B cells is consistent with classical antigen-driven adaptive immunity, in which CD8+ T cells act downstream of CD4-mediated help in an antigen-restricted manner. In ILICI, by contrast, we observed minimal CD4+ and B-cell involvement but prominent macrophage and CD8+ effector (including memory) populations. We therefore hypothesize that, in ILICI, CD8+ T cells are activated in a myeloid-rich, cytokine-driven environment, consistent with checkpoint blockade lowering activation thresholds and enabling innate-driven licensing of cytotoxic responses. How these innate cells are activated, and what recruits CD8+ T cells to the liver, remain open questions.

Confirming our prior findings in mice^[8]^, our spatial analysis shows that innate and adaptive immune cells are not only co-localized but organized into mixed innate-adaptive neighborhoods that concentrate both cC3+ and cGSDMD+ cells; whether this co-localization reflects functional coupling, and whether it sustains a feed-forward inflammatory loop in which pyroptotic signaling amplifies injury and recruits additional immune cells, cannot be determined from static tissue and remains a hypothesis. This integrated-versus-compartmentalized distinction parallels spatial immunotypes described in hepatocellular carcinoma^[36]^, linking the liver-injury architecture we observe to the anti-tumor immune contexts in which ICIs act. Our panel focused on apoptotic and pyroptotic pathways because these emerged as the dominant mechanisms in our prior mechanistic murine studies^[8]^; additional regulated cell death pathways such as ferroptosis, warrant future investigation. Within ILICI, dual-checkpoint blockade trended toward a more immune-centric distribution of dying hepatocytes and a greater proportion of pyroptotic and cC3+/cGSDMD+ events, consistent with dose-dependent innate activation, though this subgroup comparison did not reach significance and awaits validation in larger cohorts.

These findings test the hypothesis, generated from our murine model, that a spatially coupled macrophage-CD8+ T-cell niche drives hepatocyte injury in human ILICI, and support targeting myeloid signaling, pyroptotic pathways, or specific CD8-macrophage interactions as more precise therapeutic strategies, building on prior work showing that modulating apoptosis and inflammasome pathways alters disease severity in our murine model^[8]^. Because all-cause mortality in ILICI is high but predominantly cancer-related, the goal of such liver-directed strategies would be to uncouple hepatotoxicity from anti-tumor immunity so that patients can remain on effective therapy. Overall, our findings characterize ILICI as a spatially organized innate-adaptive inflammatory process and provide a basis for mechanism-focused studies aimed at separating anti-tumor immunity from liver toxicity.

Our study has several limitations. First, although the panel was designed to interrogate inflammasome activity, antibody performance precluded reliable detection of NLRP3 in human tissue. Cleaved gasdermin D (cGSDMD) therefore served as a downstream proxy for pyroptotic pathway activation rather than direct evidence of inflammasome assembly. Together with our murine data^[8]^, these findings support inflammasome activity in human ILICI. Second, our sample size is moderate, reflecting the rarity of well-phenotyped, biopsy-confirmed ILICI. As an observational study, causal inferences cannot be established, and cC3 and cGSDMD positivity should be interpreted as localization of these pathway markers rather than definitive evidence of the terminal fate of individual immune cells. However, the concordance between the human findings and preclinical mechanistic data strengthens the biological plausibility of the observed associations. Third, technical factors may contribute to variability. The ILICI and AIH cohorts were acquired at different institutions, and imaging was performed sequentially over several days, introducing the potential for batch effects that cannot be fully disentangled from biological variation. To minimize these effects, we applied batch correction. ROI acquisitions also spanned multiple sessions with varying detector sensitivities, necessitating per-ROI threshold optimization for cell classification. These thresholds were empirically validated to achieve consistent signal-to-background performance across acquisitions. Fourth, the study design imposes analytical considerations. ROIs were intentionally selected from histologically active regions to maximize characterization of inflammatory microenvironments; consequently, portal, periportal, and uninvolved parenchyma are underrepresented, and compositional findings should not be extrapolated to the entire biopsy or liver. ROI area also varied across groups. While cell densities were normalized for area, neighborhood analyses rely on absolute cell counts and graph topology and therefore may be influenced by ROI size. Differences in age, sex, ethnicity, and underlying malignancy between cohorts reflect the clinical populations studied and could not be adjusted for at the present sample size. The monotherapy subgroup encompassed mechanistically distinct immune checkpoint inhibitors (anti-PD-1/PD-L1 and anti-CTLA-4), and subgroup as well as ROI-level clinical analyses should be interpreted as exploratory pending validation in larger cohorts. Finally, we did not perform T-cell receptor sequencing, and therefore cannot determine the antigen specificity or clonal structure of the CD8+ T cells within these niches; the inference that a substantial proportion of the infiltrate is not tumor-cross-reactive rests on the repertoire data reported by Uzun et al^[34]^.

Taken together, these data reframe ILICI not as a diffuse, purely cytotoxic infiltrate but as a spatially organized macrophage-CD8+ T cell circuit that recapitulates, in human tissue, the injury niche we defined in mice. This spatial atlas nominates myeloid signaling and CD8-macrophage interactions as liver-directed targets and provides a framework for future studies aimed at uncoupling hepatotoxicity from anti-tumor immunity, the central challenge in making ICI therapy safer without making it less effective.

## Abbreviations

AIH: autoimmune hepatitis
Alk Phos: alkaline phosphatase
ALT: alanine aminotransferase
ANA: anti-nuclear antibody
AST: aspartate aminotransferase
cC3: cleaved caspase-3
cGSDMD: cleaved N-terminal gasdermin D
CN: cellular neighborhood
CTL: cytotoxic T lymphocyte
CTLA-4: cytotoxic T-lymphocyte-associated protein-4
DILIN: Drug-Induced Liver Injury Network
FOV: field of view
GLMM: generalized linear mixed-effects model
HAI: histology activity index
HLA: human leukocyte antigen
ICI: immune checkpoint inhibitor
ILICI: immune-mediated liver injury from ICI use
IMC: imaging mass cytometry
irAE: immune-related adverse event
LMM: linear mixed model
NPC: non-parenchymal cell
PD-1: programmed cell death protein-1
PD-L1: programmed death-ligand 1
ROI: region of interest
TEM: effector memory T cell
Tbili: total bilirubin
TNF: tumor necrosis factor
UMAP: uniform manifold approximation and projection

## Acknowledgements

We thank Ying Li (Cedars-Sinai Medical Center) for technical support with imaging mass cytometry post-acquisition QC troubleshooting, and Benjamin Halligan for technical assistance with data preprocessing and implementation. We acknowledge the patients and families who participated in the Drug-Induced Liver Injury Network. The DILIN is structured as a cooperative agreement, and the NIDDK program scientists participated in the design and conduct of the study and reviewed the manuscript. We also thank the investigators and coordinators at the clinical sites participating in the DILIN network.

## Declaration of generative AI and AI-assisted technologies in the manuscript preparation process

During the preparation of this work, the author(s) used Claude (Anthropic) for drafting and implementing template software code for data preprocessing and visualization, and for proofreading, grammatical review, and editing of the manuscript text. The author(s) reviewed and edited all output as needed and take full responsibility for the content of the published article.

## Author contributions

**JBM**: Conceptualization, Methodology, Validation, Formal analysis, Investigation, Data curation, Writing - original draft, Writing - review & editing, Visualization, Supervision, Project administration. **NZ:** Conceptualization, Methodology, Software, Validation, Formal analysis, Investigation, Resources, Data curation, Writing - original draft, Writing - review & editing, Visualization, Supervision, Project administration. **HA:** Methodology, Investigation, Writing - review & editing. **DEK:** Investigation, Resources, Writing - review & editing. **RJF:** Resources, Writing - review & editing. **AAS:** Conceptualization, Resources, Writing - review & editing. **AM:** Conceptualization, Methodology, Investigation, Resources, Writing - review & editing. **JZS:** Conceptualization, Methodology, Software, Formal analysis, Investigation, Resources, Data Curation, Writing - original draft, Writing - review & editing, Supervision, Project administration. **LD:** Conceptualization, Writing - original draft, Writing - review & editing, Supervision, Project administration, Funding acquisition.

## Data availability statement

Raw IMC images, processed single-cell objects, and per-ROI metadata are available in BioImage Archive Submission - S-BIAD3614 (DOI: 10.6019/S-BIAD3614).

## Clinical trial number

Not applicable.

## References

[1] Haslam A, Olivier T, Prasad V. How many people in the US are eligible for and respond to checkpoint inhibitors: An empirical analysis. Int J Cancer. 2025;156:2352–2359.

[2] Ennin E, Mallepally N, Ali M, et al. High Grade Hepatotoxicity From Dual Checkpoint Inhibitors Is More Common in Hepatocellular Carcinoma Than Other Cancers. Liver international : official journal of the International Association for the Study of the Liver. 2025;45:e70255.

[3] Dara L, De Martin E. Immune-Mediated Liver Injury From Checkpoint Inhibitor: An Evolving Frontier With Emerging Challenges. Liver international : official journal of the International Association for the Study of the Liver. 2025;45:e16198.

[4] Wang T, Yeh MM, Avigan MI, et al. Deciphering the Dynamic Complexities of the Liver Microenvironment - Toward a Better Understanding of Immune-Mediated liver Injury Caused by Immune Checkpoint Inhibitors (ILICI). AAPS J. 2021;23:99.

[5] Shojaie L, Ali M, Iorga A, et al. Mechanisms of immune checkpoint inhibitor-mediated liver injury. Acta Pharm Sin B. 2021;11:3727–3739.

[6] Affolter T, Llewellyn HP, Bartlett DW, et al. Inhibition of immune checkpoints PD-1, CTLA-4, and IDO1 coordinately induces immune-mediated liver injury in mice. PLoS One. 2019;14:e0217276.

[7] Regev A, Avigan MI, Kiazand A, et al. Best practices for detection, assessment and management of suspected immune-mediated liver injury caused by immune checkpoint inhibitors during drug development. J Autoimmun. 2020;114:102514.

[8] Shojaie L, Bogdanov JM, Alavifard H, et al. Innate and adaptive immune cell interaction drives inflammasome activation and hepatocyte apoptosis in murine liver injury from immune checkpoint inhibitors. Cell Death Dis. 2024;15:140.

[9] Adam K, Iuga A, Tocheva AS, et al. A novel mouse model for checkpoint inhibitor-induced adverse events. PLoS One. 2021;16:e0246168.

[10] Gudd CLC, Au L, Triantafyllou E, et al. Activation and transcriptional profile of monocytes and CD8(+) T cells are altered in checkpoint inhibitor-related hepatitis. J Hepatol. 2021;75:177–189.

[11] Triantafyllou E, Gudd CLC, Possamai LA. Immune-mediated liver injury from checkpoint inhibitors: mechanisms, clinical characteristics and management. Nat Rev Gastroenterol Hepatol. 2025;22:112–126.

[12] Saarela M, Parviainen E, Lleo A, et al. Increased PD-1 expression in livers associated with PD-1-antibody-induced hepatotoxicity. BMC Immunol. 2025;26:4.

[13] Pesch G, Piseddu I, Gaertig J, et al. Deep immune phenotyping reveals distinct immunopathogenesis in checkpoint inhibitor-induced colitis compared to ulcerative colitis. Cancer Immunol Res. 2025.

[14] Ji C, Kumpf S, Qian J, et al. Transcriptomic and proteomic characterization of cell and protein biomarkers of checkpoint inhibitor-induced liver injury. Cancer Immunol Immunother. 2025;74:190.

[15] Fontana RJ, Li YJ, Chen V, et al. Genetic variants associated with immune-mediated liver injury from checkpoint inhibitors. Hepatol Commun. 2024;8.

[16] Sunago K, Abe M, Yoshida O, et al. Clinical and Pathological Features of Immune Checkpoint Inhibitor-induced Liver Injury in Comparison with Drug-induced Liver Injury and Autoimmune Hepatitis. J Gastrointestin Liver Dis. 2023;32:488–496.

[17] Wang SJ, Dougan SK, Dougan M. Immune mechanisms of toxicity from checkpoint inhibitors. Trends Cancer. 2023;9:543–553.

[18] Parlati L, Marcin K, Terris B, et al. Histological Characteristics and Management of Hepatitis on Immune Checkpoint Inhibitors: A Retrospective Descriptive Study. J Clin Med. 2023;12.

[19] Llewellyn HP, Arat S, Gao J, et al. T cells and monocyte-derived myeloid cells mediate immunotherapy-related hepatitis in a mouse model. J Hepatol. 2021;75:1083–1095.

[20] Hutchinson JA, Kronenberg K, Riquelme P, et al. Virus-specific memory T cell responses unmasked by immune checkpoint blockade cause hepatitis. Nat Commun. 2021;12:1439.

[21] Chuah S, Lee J, Song Y, et al. Uncoupling immune trajectories of response and adverse events from anti-PD-1 immunotherapy in hepatocellular carcinoma. J Hepatol. 2022;77:683–694.

[22] De Martin E, Michot JM, Papouin B, et al. Characterization of liver injury induced by cancer immunotherapy using immune checkpoint inhibitors. J Hepatol. 2018;68:1181–1190.

[23] Hountondji L, Ferreira De Matos C, Lebosse F, et al. Clinical pattern of checkpoint inhibitor-induced liver injury in a multicentre cohort. JHEP Rep. 2023;5:100719.

[24] Perez-Ruiz E, Minute L, Otano I, et al. Prophylactic TNF blockade uncouples efficacy and toxicity in dual CTLA-4 and PD-1 immunotherapy. Nature. 2019;569:428–432.

[25] Alvarez M, Otano I, Minute L, et al. Impact of prophylactic TNF blockade in the dual PD-1 and CTLA-4 immunotherapy efficacy and toxicity. Cell Stress. 2019;3:236–239.

[26] Corrigan M, Haydon G, Thompson F, et al. Infliximab for the treatment of refractory immune-related hepatitis secondary to checkpoint inhibitors: A case report. JHEP Rep. 2019;1:66–69.

[27] Parvathareddy V, Selamet U, Sen AA, et al. Infliximab for Treatment of Immune Adverse Events and Its Impact on Tumor Response. Cancers (Basel). 2023;15.

[28] Araujo DV, Muniz TP, Yang A, et al. Real World Outcomes and Hepatotoxicity of Infliximab in the Treatment of Steroid-Refractory Immune-Related Adverse Events. Curr Oncol. 2021;28:2173–2179.

[29] Bozward AG, Davies SP, Morris SM, et al. Cellular interactions in self-directed immune-mediated liver diseases. J Hepatol. 2025;82:1110–1124.

[30] Wang Y, Su Y, Guo T, et al. Immune-mediated liver injury caused by immune checkpoint inhibitors exhibits distinct clinical features that differ from autoimmune hepatitis. Expert Opin Drug Metab Toxicol. 2024:1–9.

[31] Yasuda T, Ito T, Ishikawa T, et al. Clinical features and pathological findings by liver biopsy in patients with immune-related sclerosing cholangitis induced by immune checkpoint inhibitors. Dig Liver Dis. 2025;57:877–884.

[32] Astbury S, Atallah E, Grove JI, et al. Circulating exhausted CD8+ effector memory cells differentiate immune checkpoint inhibitor-induced liver injury from other acute immune-mediated liver injuries. J Immunother Cancer. 2026;14.

[33] Huang LR, Wohlleber D, Reisinger F, et al. Intrahepatic myeloid-cell aggregates enable local proliferation of CD8(+) T cells and successful immunotherapy against chronic viral liver infection. Nat Immunol. 2013;14:574–583.

[34] Uzun S, Haefliger S, Zinner CP, et al. An integrated single-cell atlas of checkpoint inhibitor-induced liver injury links shared liver-tumour CD8+ T cell clones to cytotoxicity and macrophage crosstalks. medRxiv. 2026:2026.2004.2027.26351449.

[35] Foureau DM, Walling TL, Maddukuri V, et al. Comparative analysis of portal hepatic infiltrating leucocytes in acute drug-induced liver injury, idiopathic autoimmune and viral hepatitis. Clin Exp Immunol. 2015;180:40–51.

[36] Salie H, Wischer L, D’Alessio A, et al. Spatial single-cell profiling and neighbourhood analysis reveal the determinants of immune architecture connected to checkpoint inhibitor therapy outcome in hepatocellular carcinoma. Gut. 2025;74:451–466.

