## Supplementary material for "Macrophage-CD8⁺ T Cell Spatial Coupling Defines an Innate-Adaptive Injury Niche in Human Checkpoint Inhibitor Hepatotoxicity": Figure S-1

Global Clinical−Histopathological Correlogram (ICI, FOV−level, n=53)
210 variables | 4992 sig. pairs (FDR<0.05) | Spearman | Rank−biserial | Phi | Cramér's V | KW−e

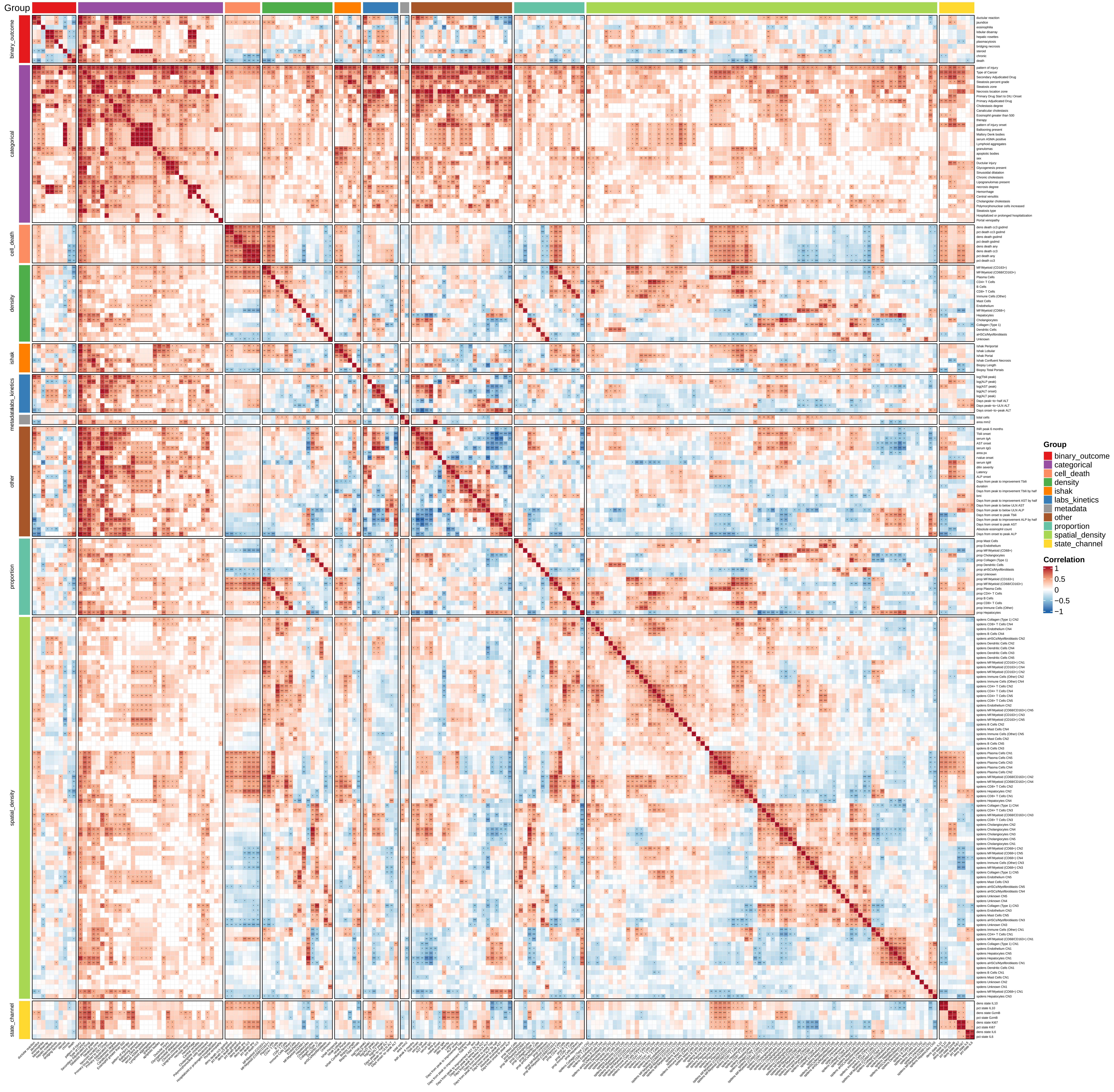
