## Supplementary figures and images for "Macrophage-CD8⁺ T Cell Spatial Coupling Defines an Innate-Adaptive Injury Niche in Human Checkpoint Inhibitor Hepatotoxicity"

### Figure S-2

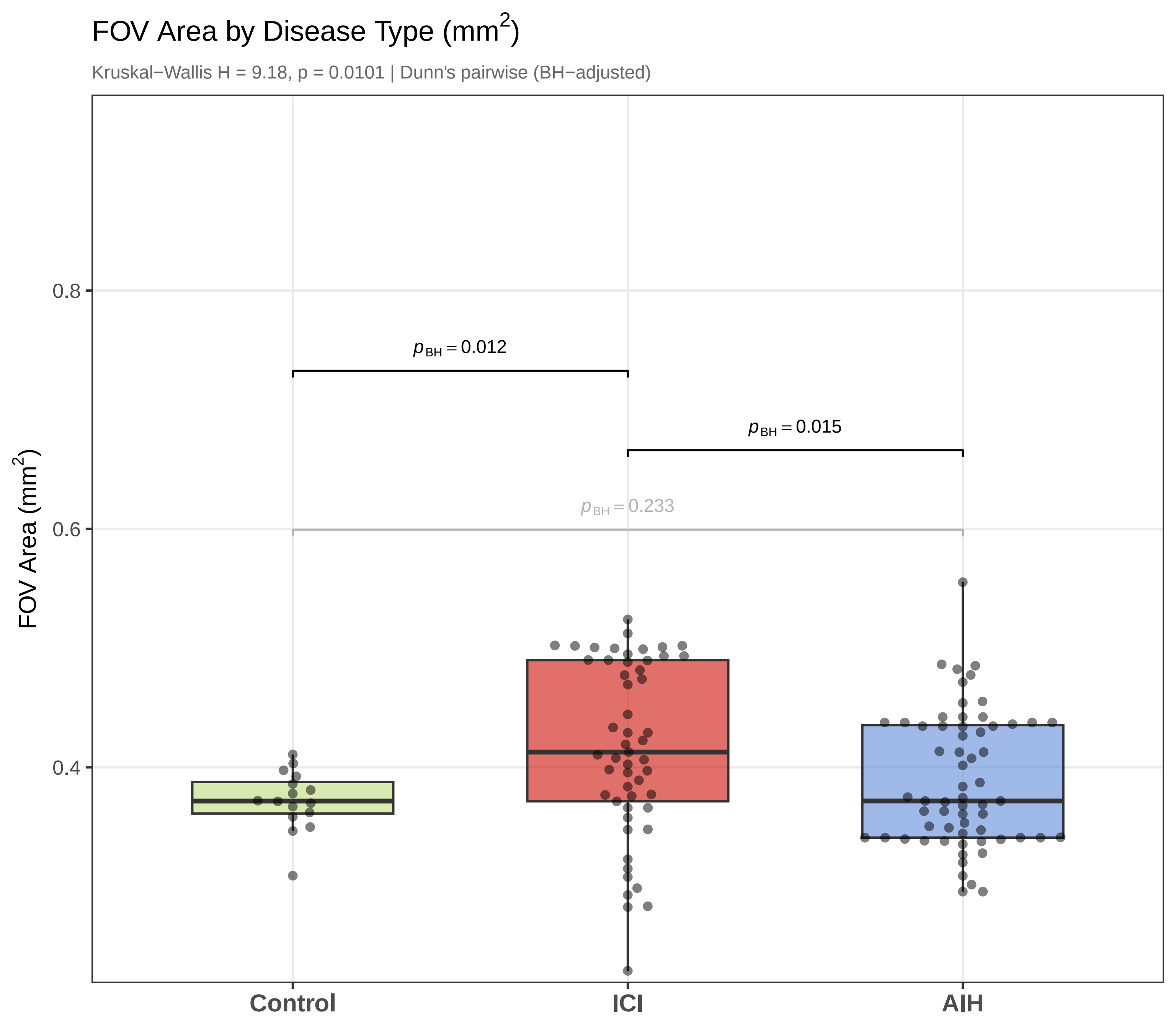

### Figure S-4

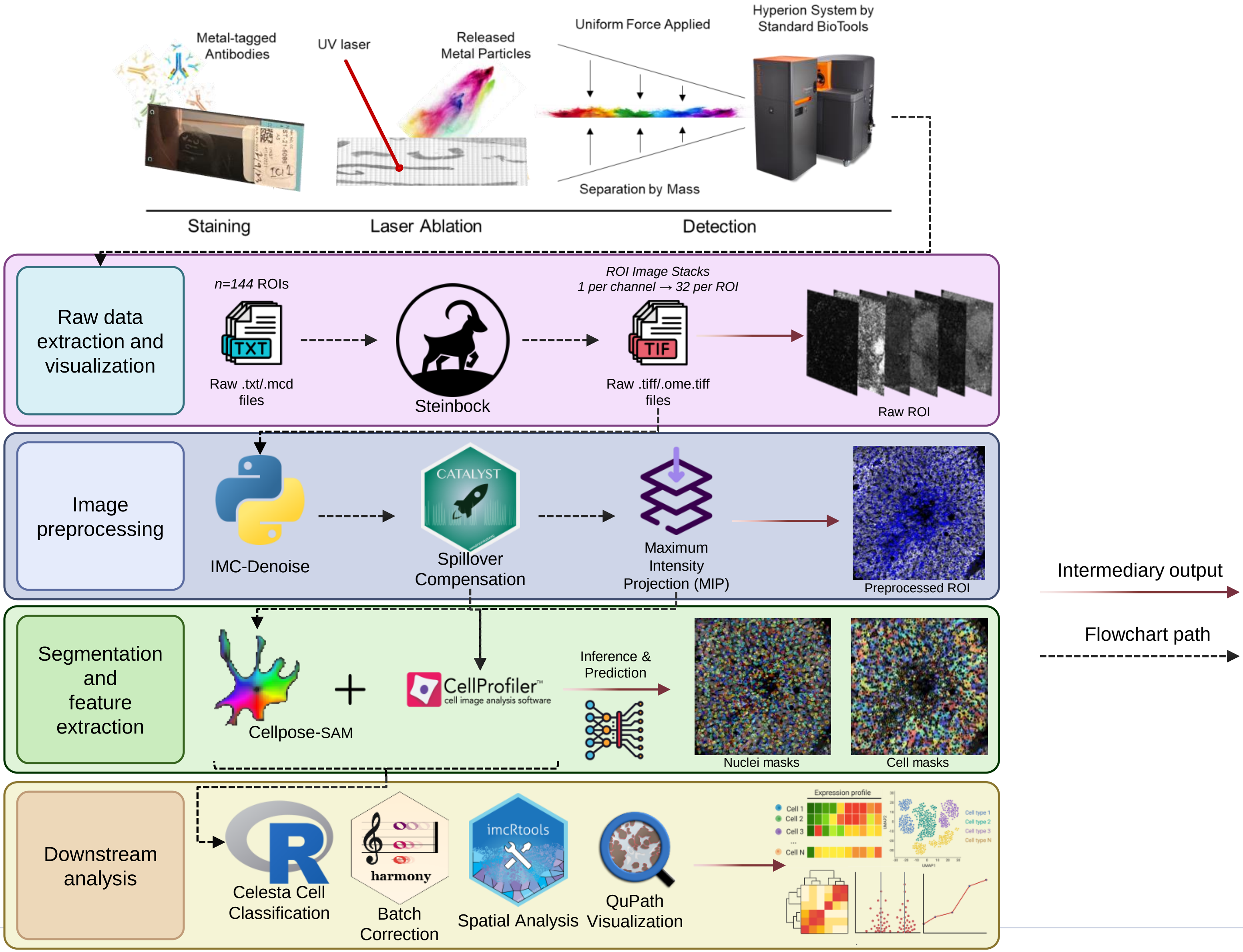

### Figure S-6

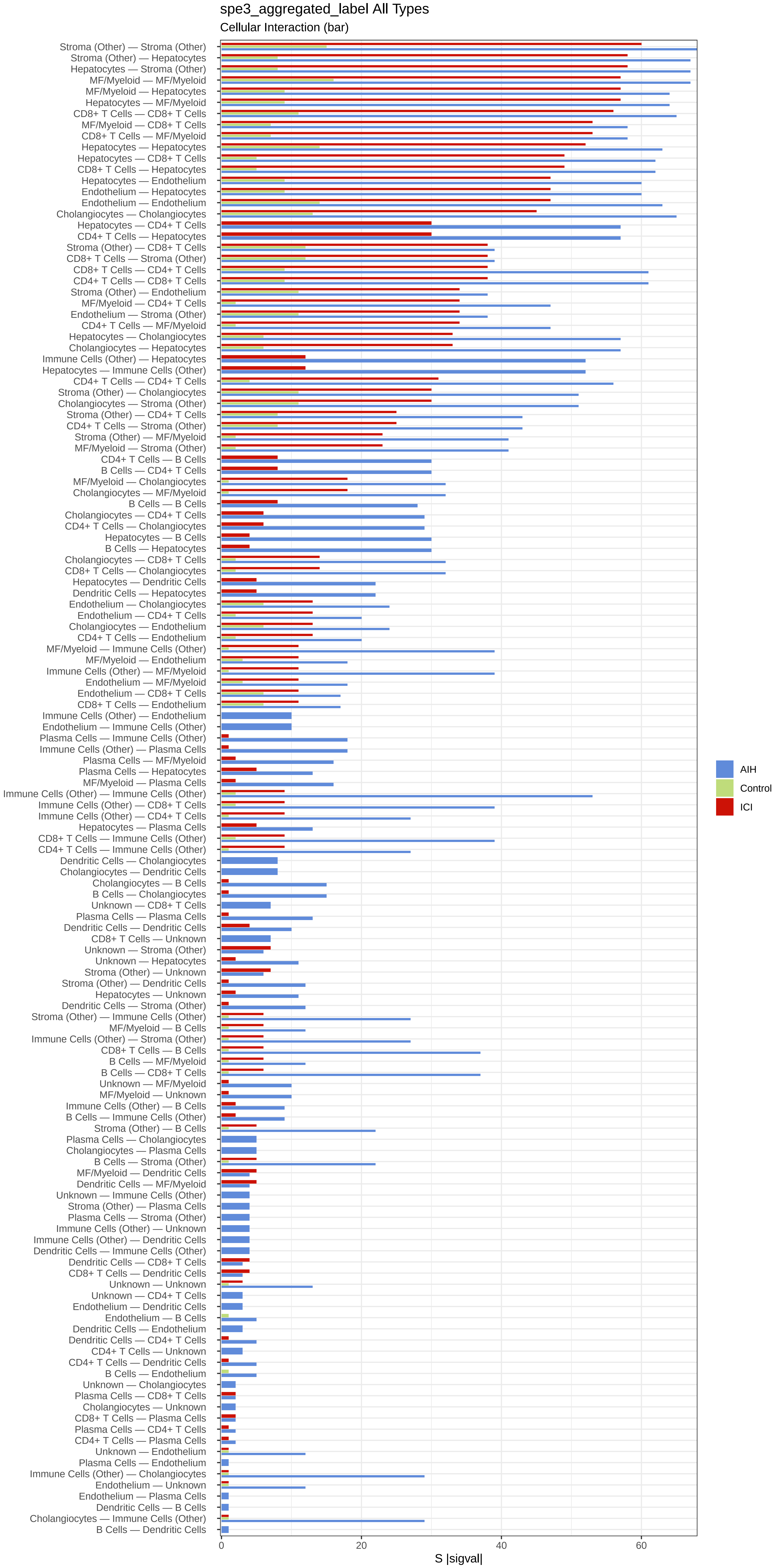
