## Supplementary material for "Macrophage-CD8⁺ T Cell Spatial Coupling Defines an Innate-Adaptive Injury Niche in Human Checkpoint Inhibitor Hepatotoxicity": Figure S-3

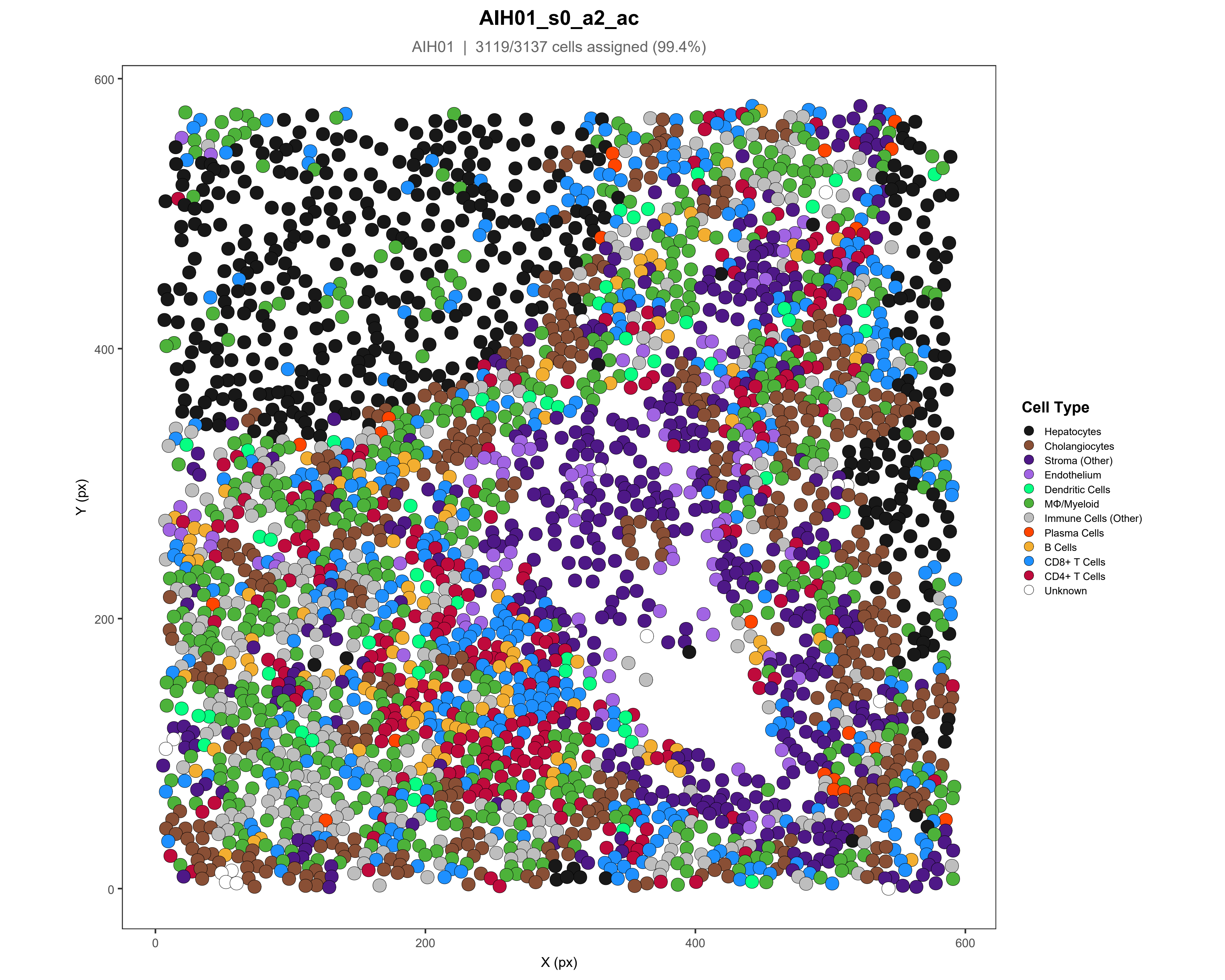

AIH01_s0_a2_ac
AIH01 | 3119/3137 cells assigned (99.4%)
Y (px)
X (px)
0
200
400
600
Cell Type
Hepatocytes
Cholangiocytes
Stroma (Other)
Endothelium
Dendritic Cells
MΦ/Myeloid
Immune Cells (Other)
Plasma Cells
B Cells
CD8+ T Cells
CD4+ T Cells
Unknown

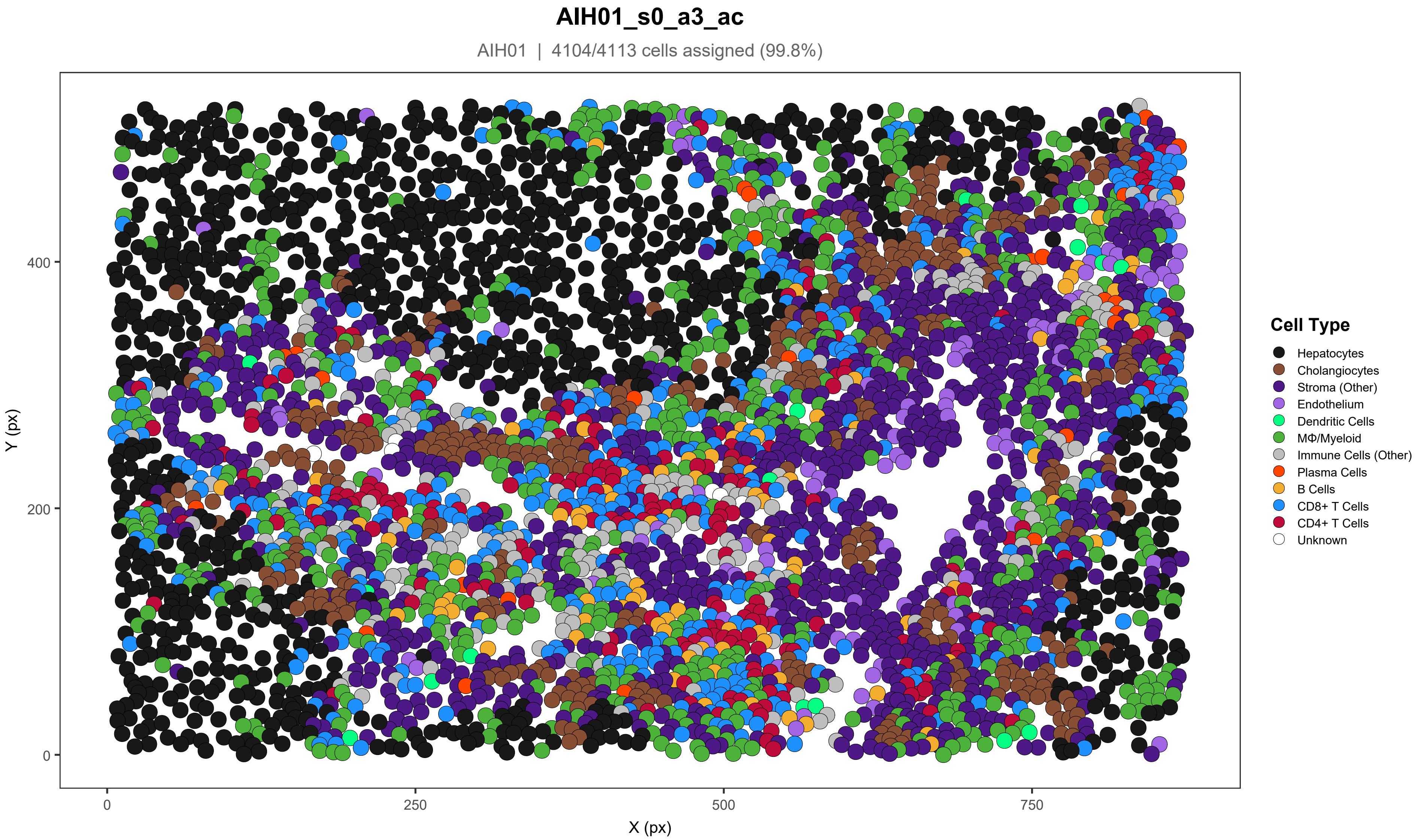
AIH01_s0_a3_ac
AIH01 | 4104/4113 cells assigned (99.8%)
Y (px)
X (px)
0
200
400
0
250
500
750
Cell Type
Hepatocytes
Cholangiocytes
Stroma (Other)
Endothelium
Dendritic Cells
MΦ/Myeloid
Immune Cells (Other)
Plasma Cells
B Cells
CD8+ T Cells
CD4+ T Cells
Unknown

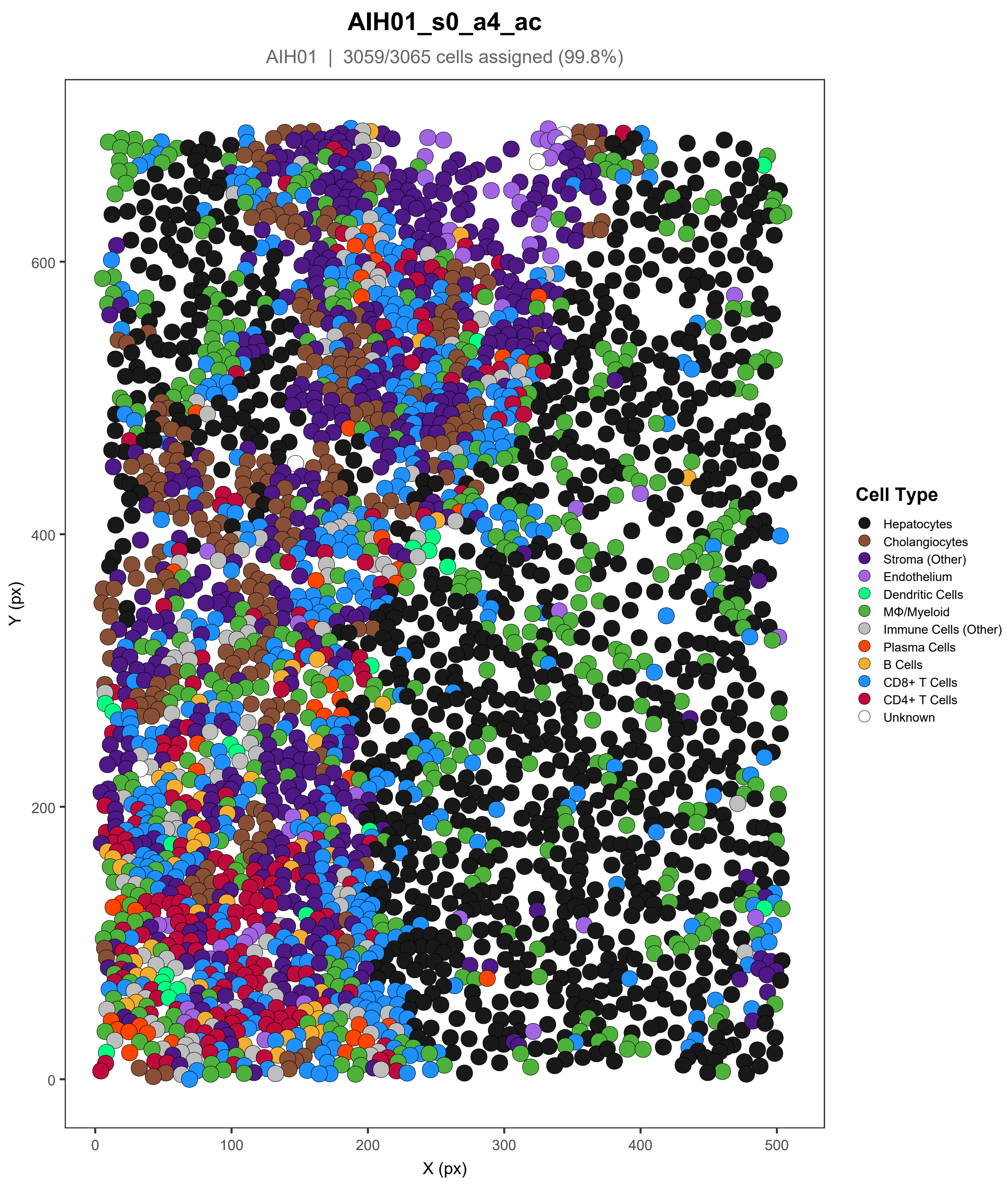

AIH01_s0_a4_ac
AIH01 | 3059/3065 cells assigned (99.8%)
Y (px)
X (px)
0
200
400
600
0
100
200
300
400
500
Cell Type
Hepatocytes
Cholangiocytes
Stroma (Other)
Endothelium
Dendritic Cells
MΦ/Myeloid
Immune Cells (Other)
Plasma Cells
B Cells
CD8+ T Cells
CD4+ T Cells
Unknown

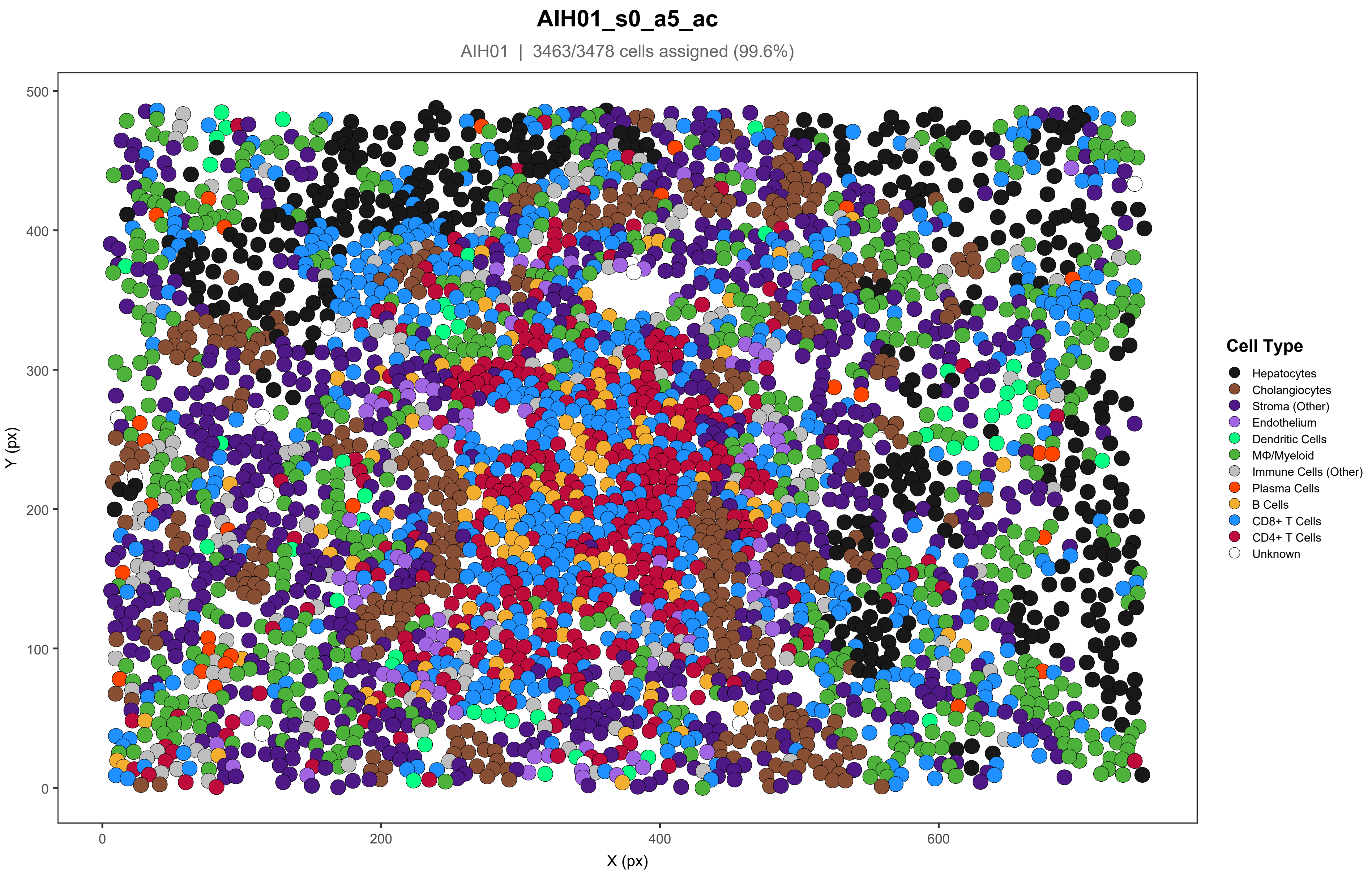
AIH01_s0_a5_ac
AIH01 | 3463/3478 cells assigned (99.6%)
500
400
300
200
100
0
Y (px)
0
200
400
600
X (px)
Cell Type
Hepatocytes
Cholangiocytes
Stroma (Other)
Endothelium
Dendritic Cells
MΦ/Myeloid
Immune Cells (Other)
Plasma Cells
B Cells
CD8+ T Cells
CD4+ T Cells
Unknown

### AIH01_s0_a6_ac

AIH01 | 2979/2989 cells assigned (99.7%)

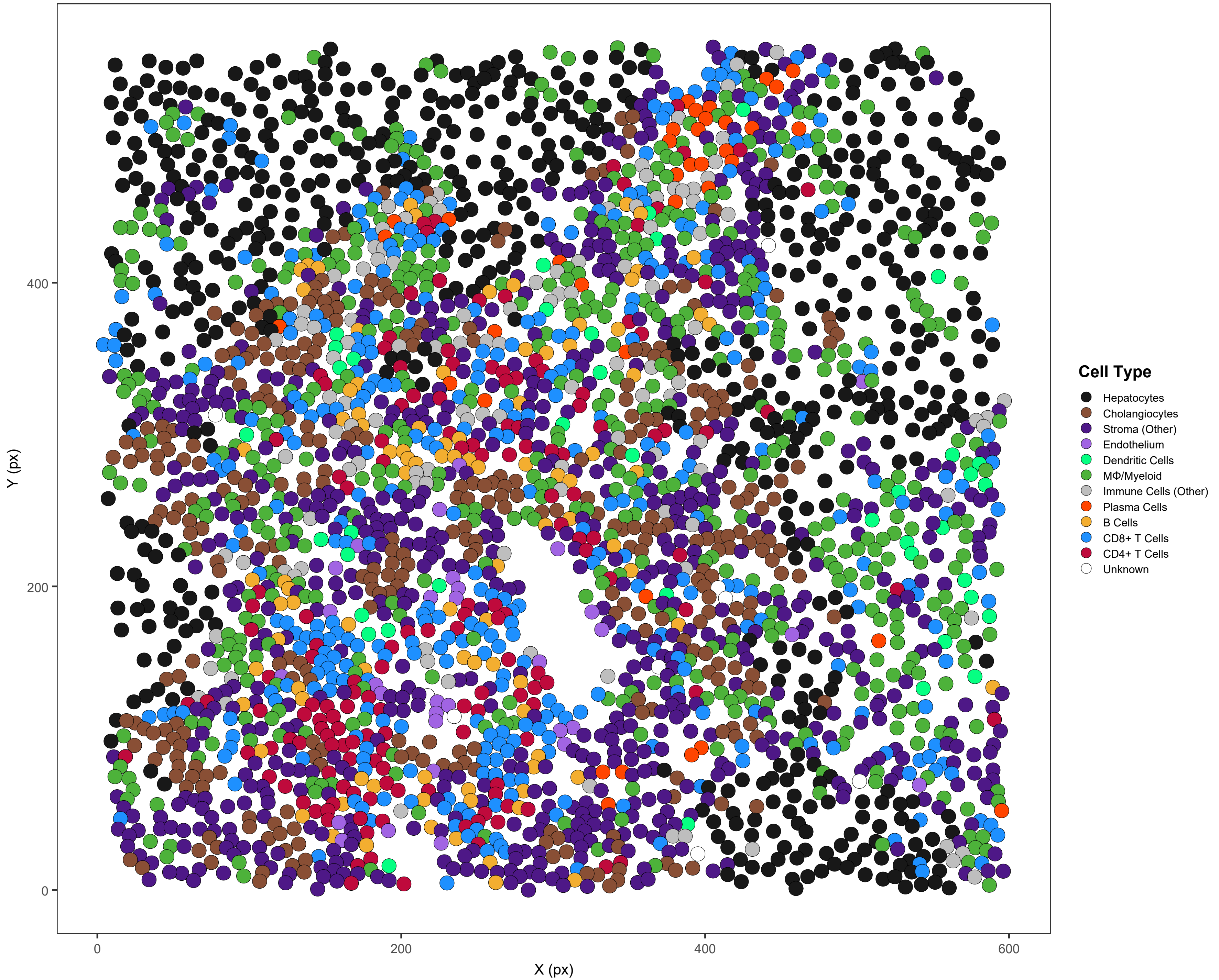

#### AIH02_s0_a1_ac

AIH02 | 3358/3358 cells assigned (100%)

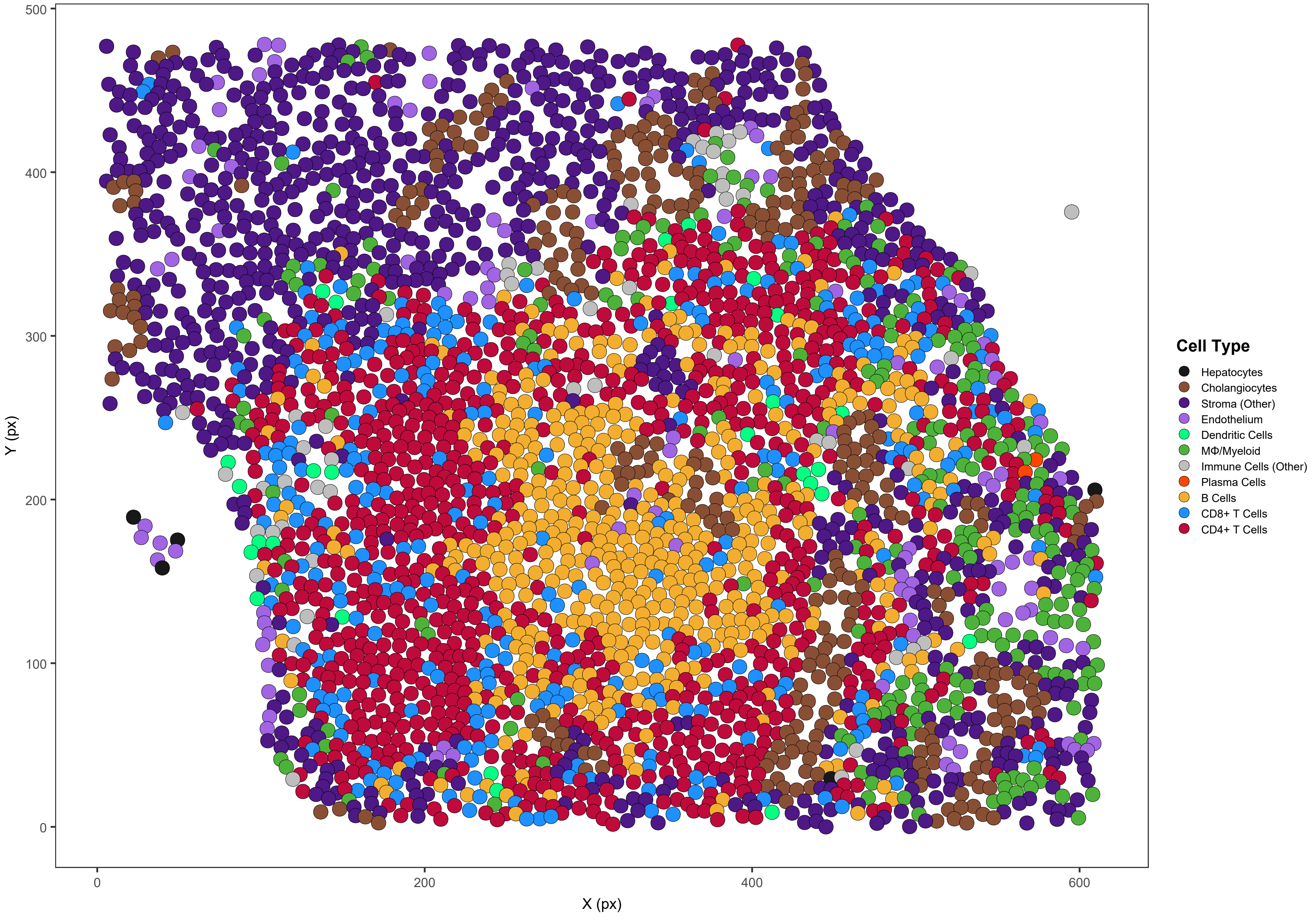

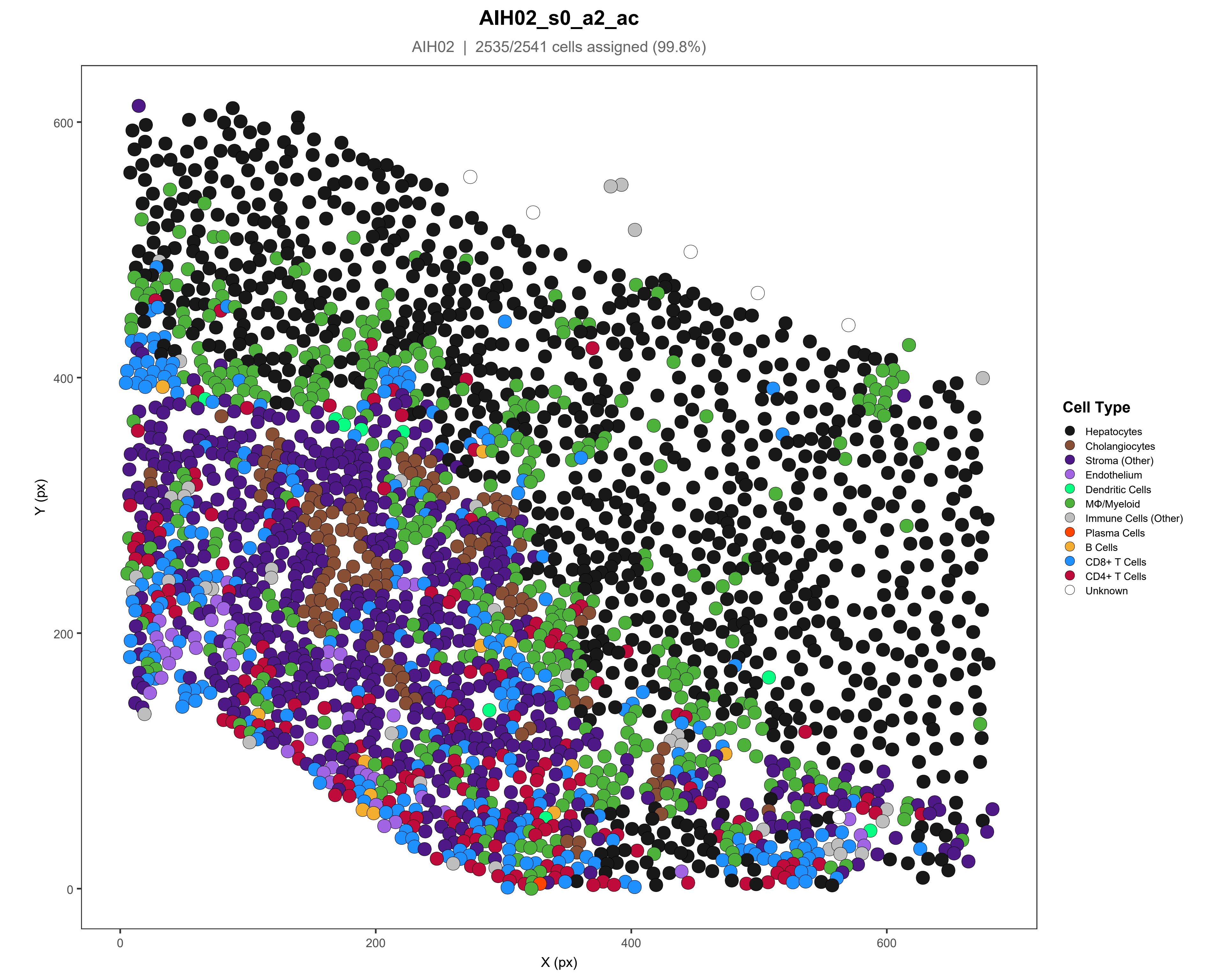
AIH02_s0_a2_ac
AIH02 | 2535/2541 cells assigned (99.8%)
Y (px)
X (px)
0
200
400
600
Cell Type
Hepatocytes
Cholangiocytes
Stroma (Other)
Endothelium
Dendritic Cells
MΦ/Myeloid
Immune Cells (Other)
Plasma Cells
B Cells
CD8+ T Cells
CD4+ T Cells
Unknown

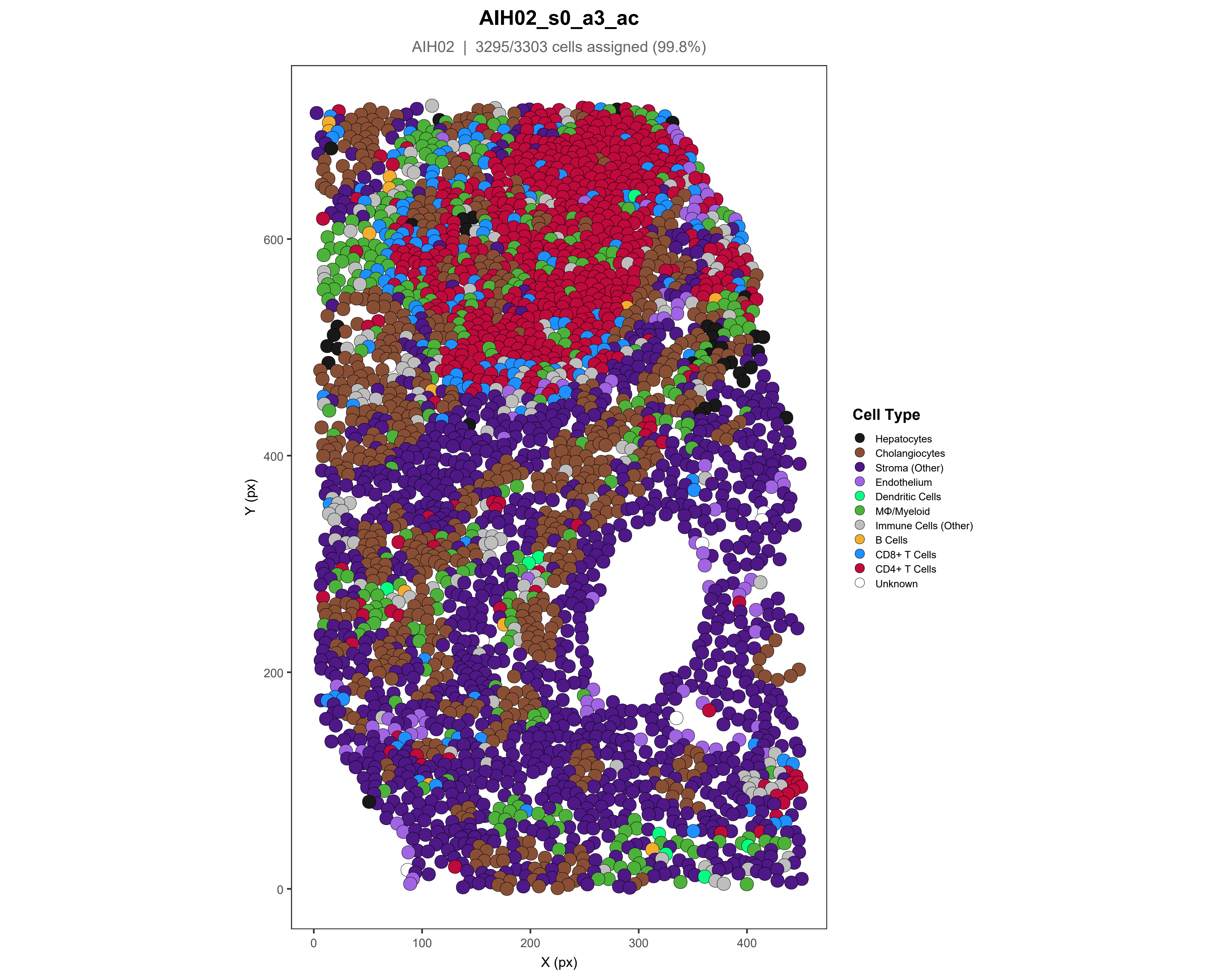
AIH02_s0_a3_ac
AIH02 | 3295/3303 cells assigned (99.8%)
Y (px)
X (px)
0
100
200
300
400
600
Cell Type
Hepatocytes
Cholangiocytes
Stroma (Other)
Endothelium
Dendritic Cells
MΦ/Myeloid
Immune Cells (Other)
B Cells
CD8+ T Cells
CD4+ T Cells
Unknown

### AIH02_s0_a4_ac

AIH02 | 1824/1831 cells assigned (99.6%)

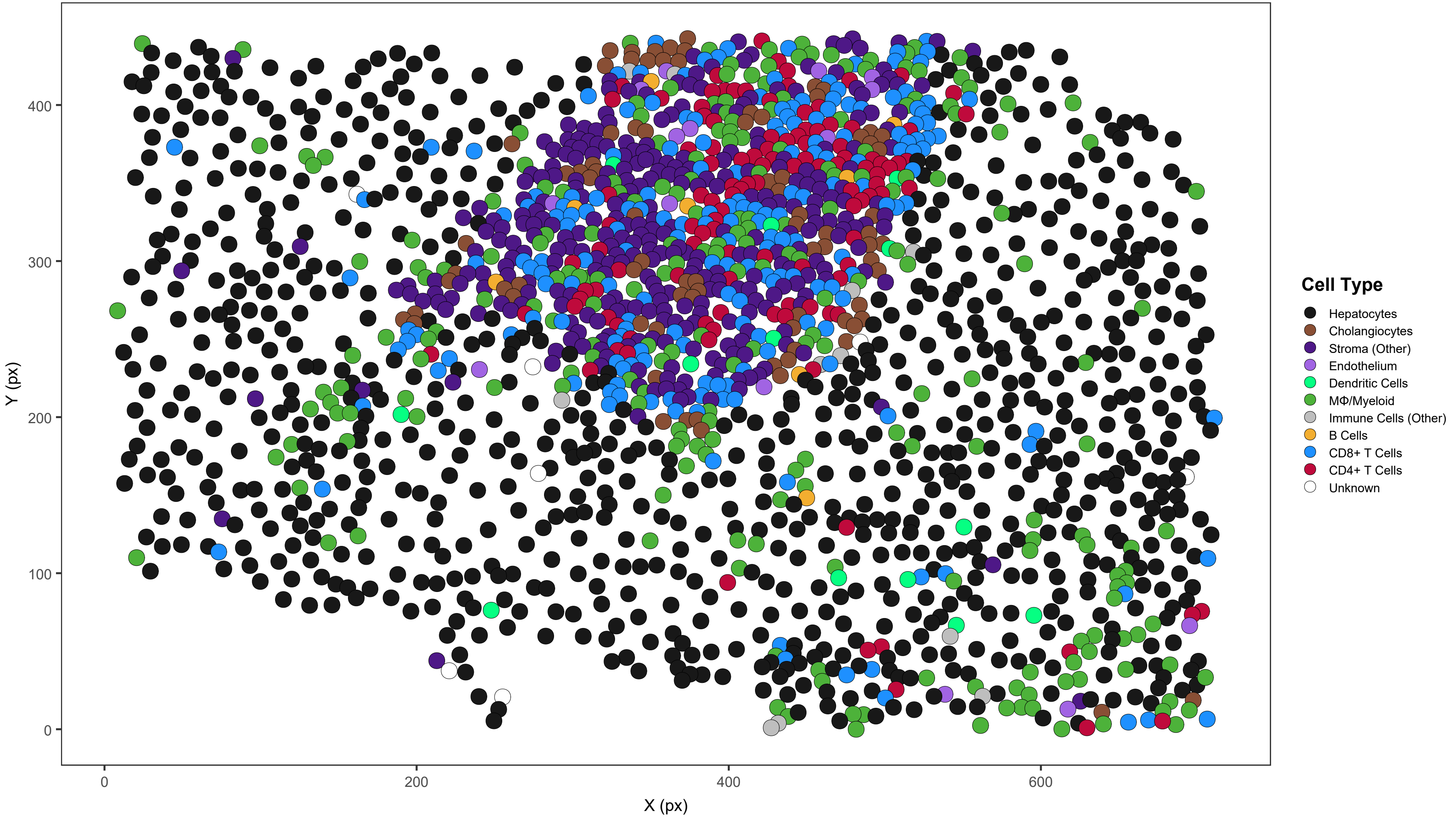

### AIH02_s0_a5_ac

AIH02 | 2240/2242 cells assigned (99.9%)

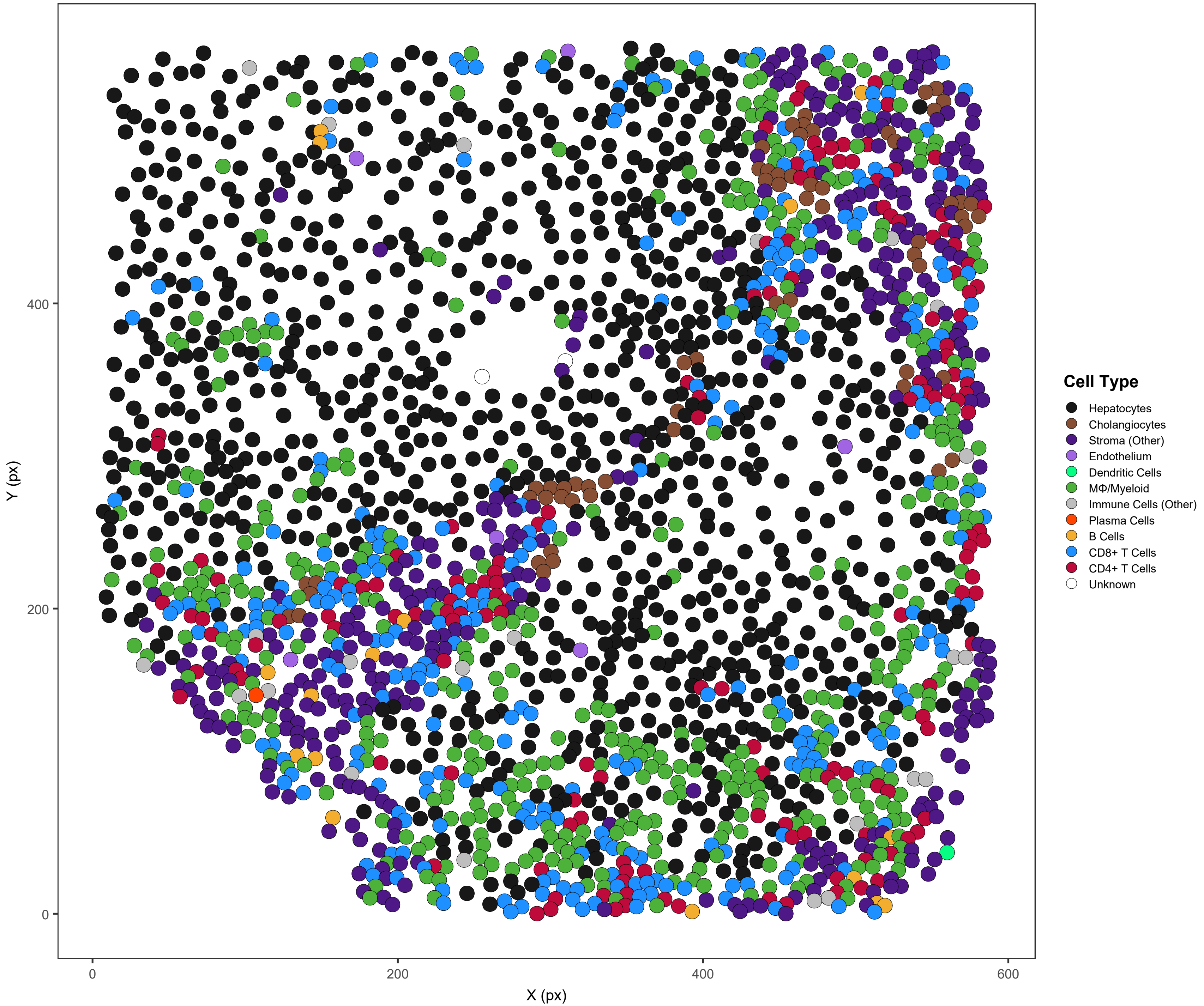

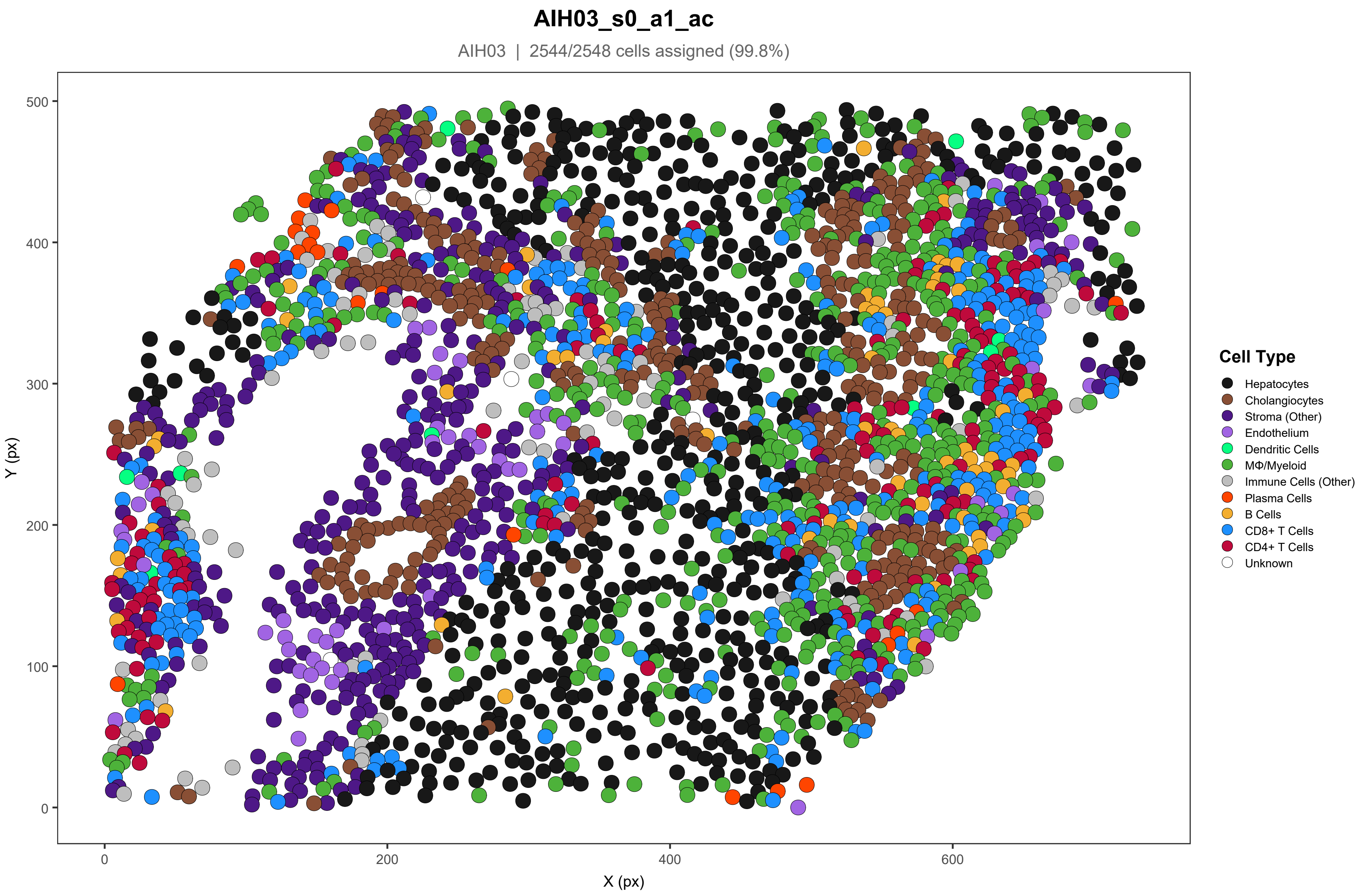
AIH03_s0_a1_ac
AIH03 | 2544/2548 cells assigned (99.8%)
Y (px)
X (px)
0
100
200
300
400
500
0
200
400
600
Cell Type
Hepatocytes
Cholangiocytes
Stroma (Other)
Endothelium
Dendritic Cells
MΦ/Myeloid
Immune Cells (Other)
Plasma Cells
B Cells
CD8+ T Cells
CD4+ T Cells
Unknown

### AIH03_s0_a2_ac

AIH03 | 2025/2033 cells assigned (99.6%)

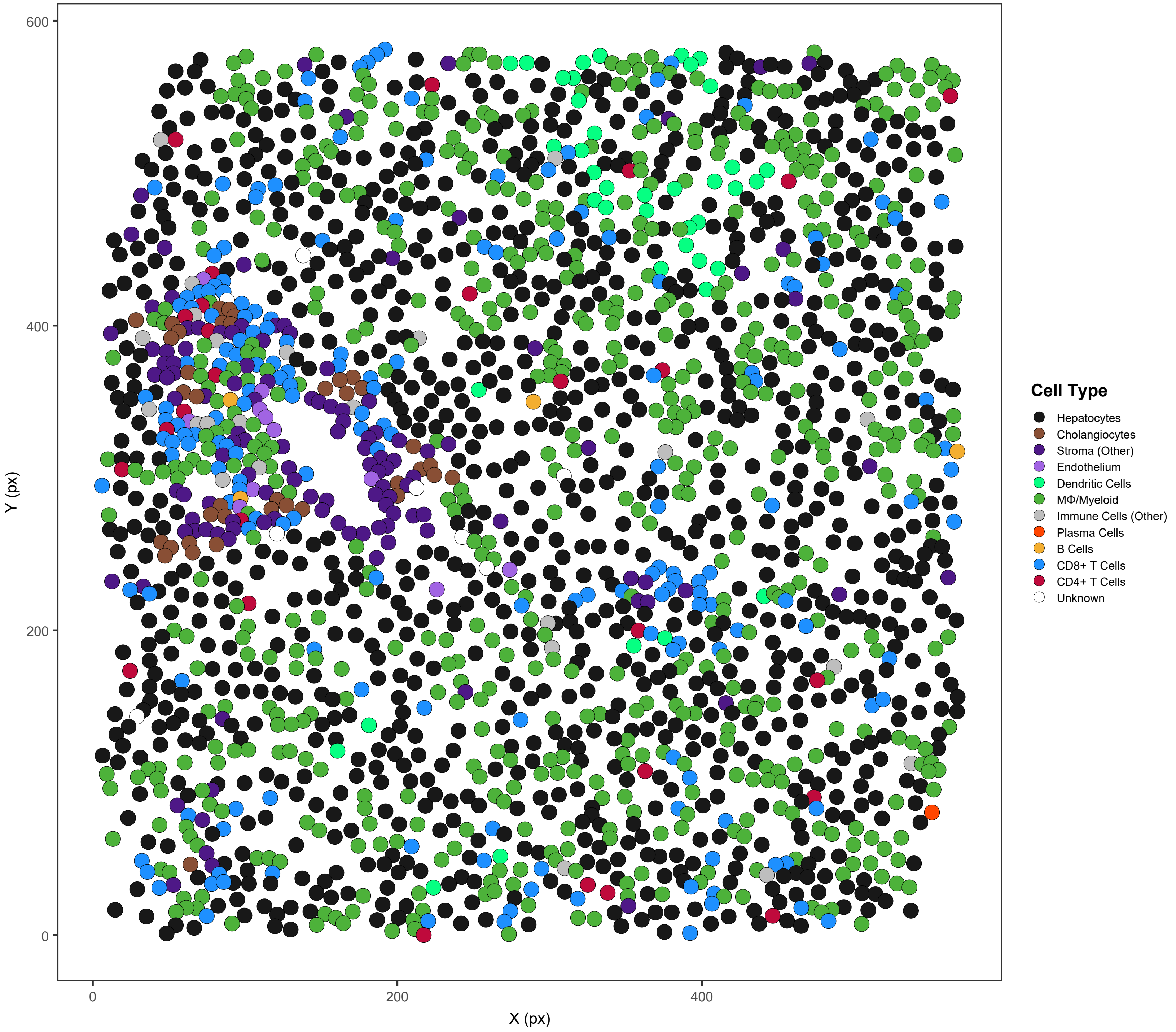

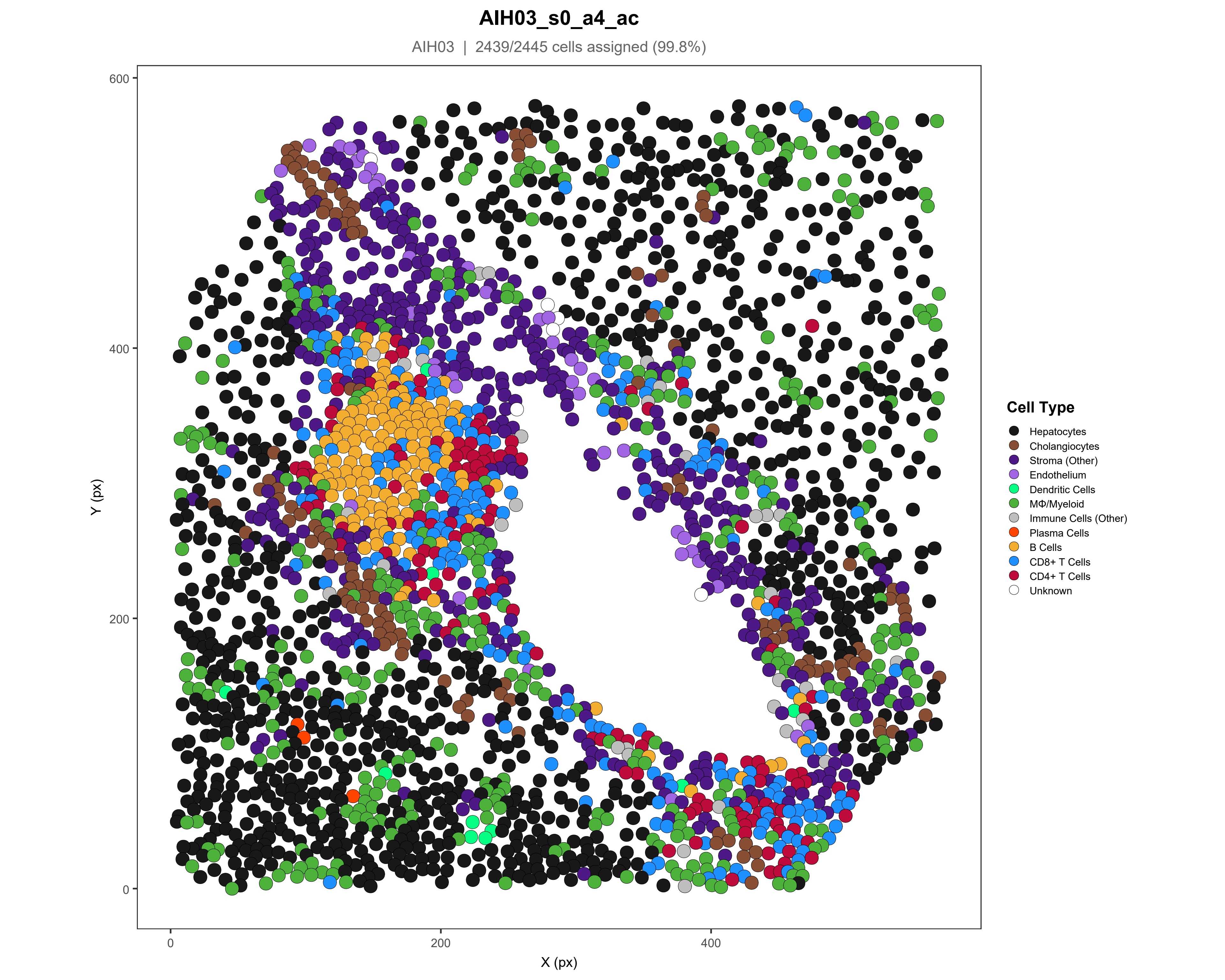

AIH03_s0_a4_ac
AIH03 | 2439/2445 cells assigned (99.8%)
Y (px)
X (px)
600
400
200
0
0
200
400
Cell Type
Hepatocytes
Cholangiocytes
Stroma (Other)
Endothelium
Dendritic Cells
MΦ/Myeloid
Immune Cells (Other)
Plasma Cells
B Cells
CD8+ T Cells
CD4+ T Cells
Unknown

### AIH03_s0_a5_ac

AIH03 | 2072/2077 cells assigned (99.8%)

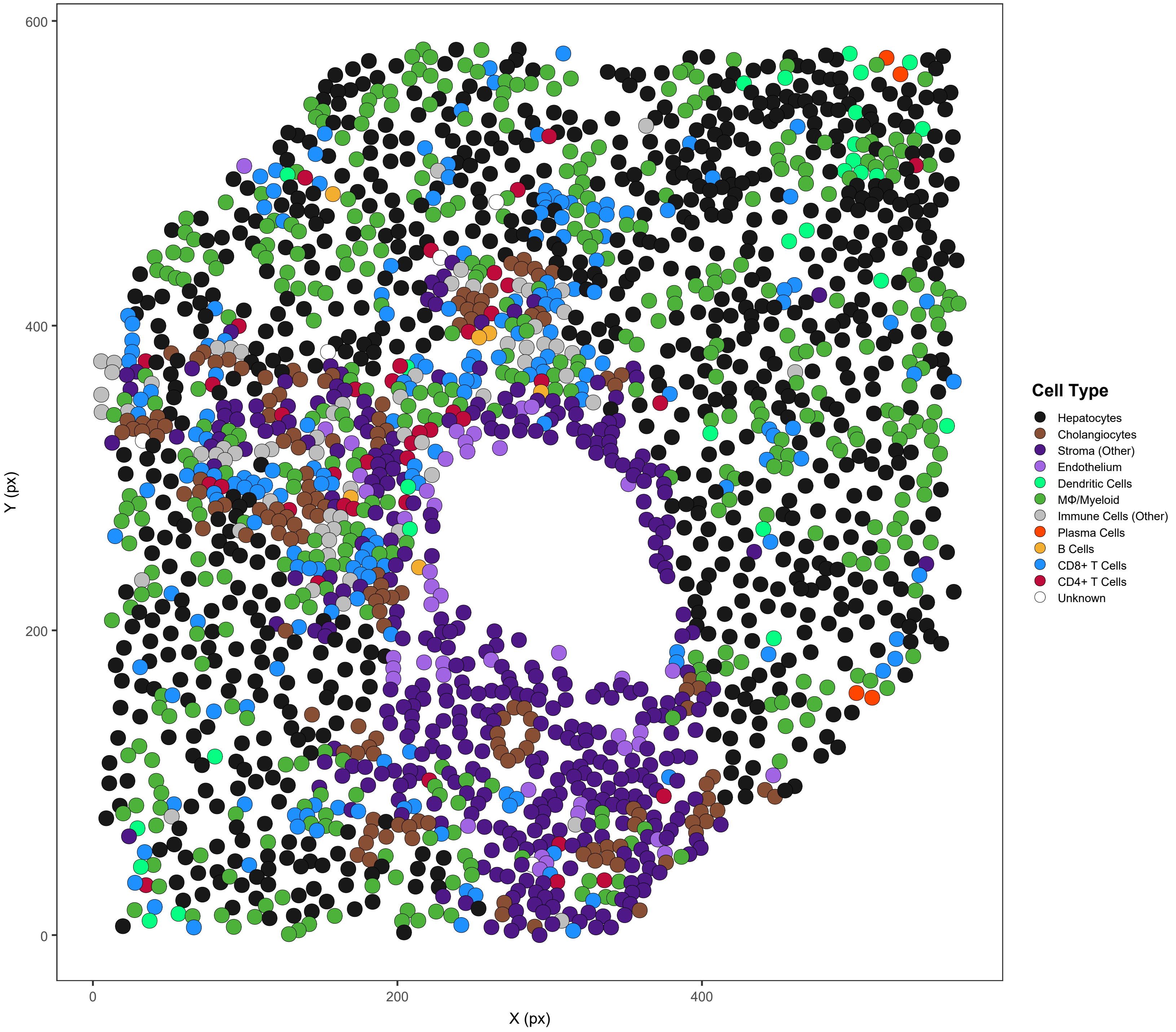

#### AIH04_s0_a1_ac

AIH04 | 2687/2699 cells assigned (99.6%)

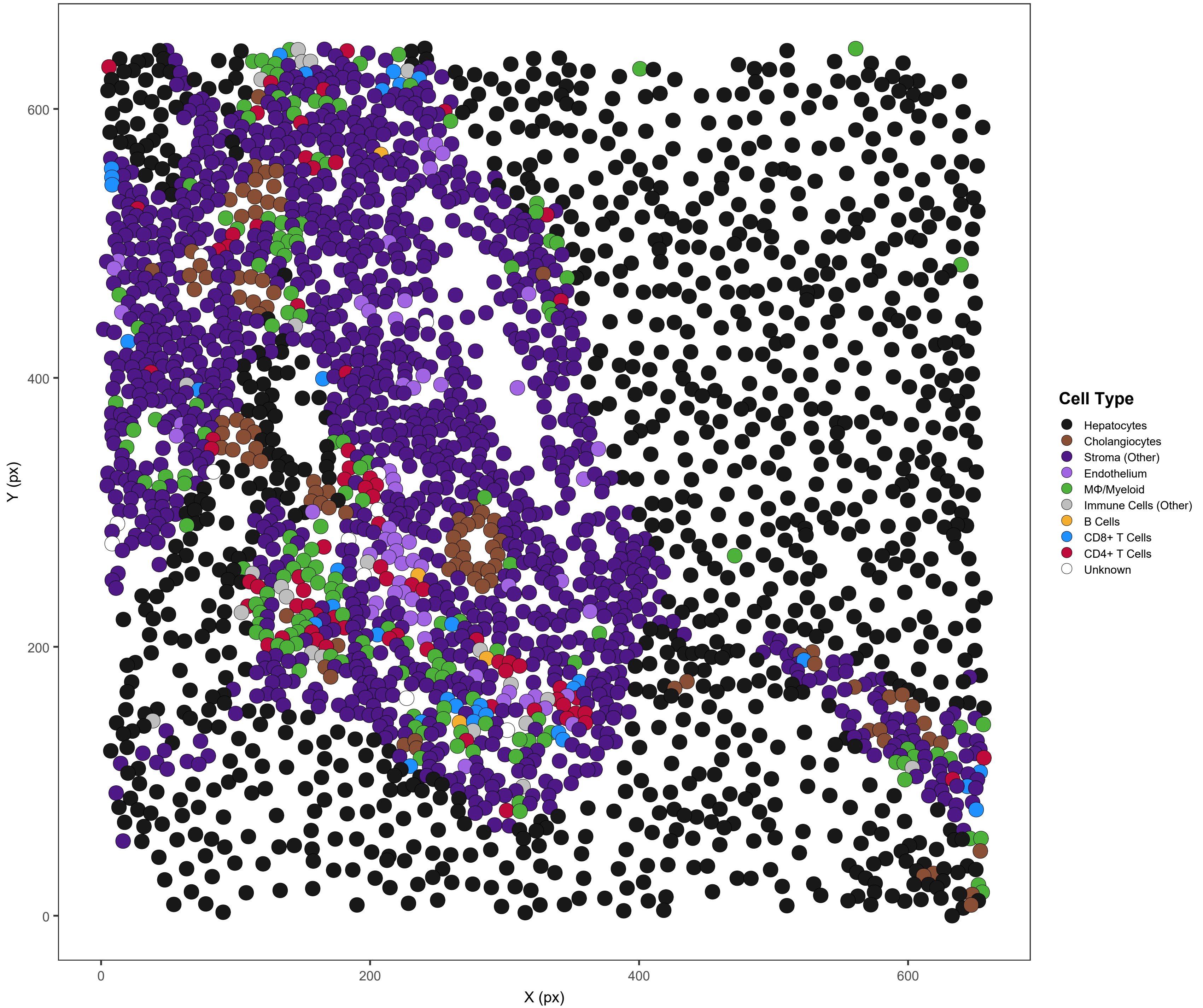

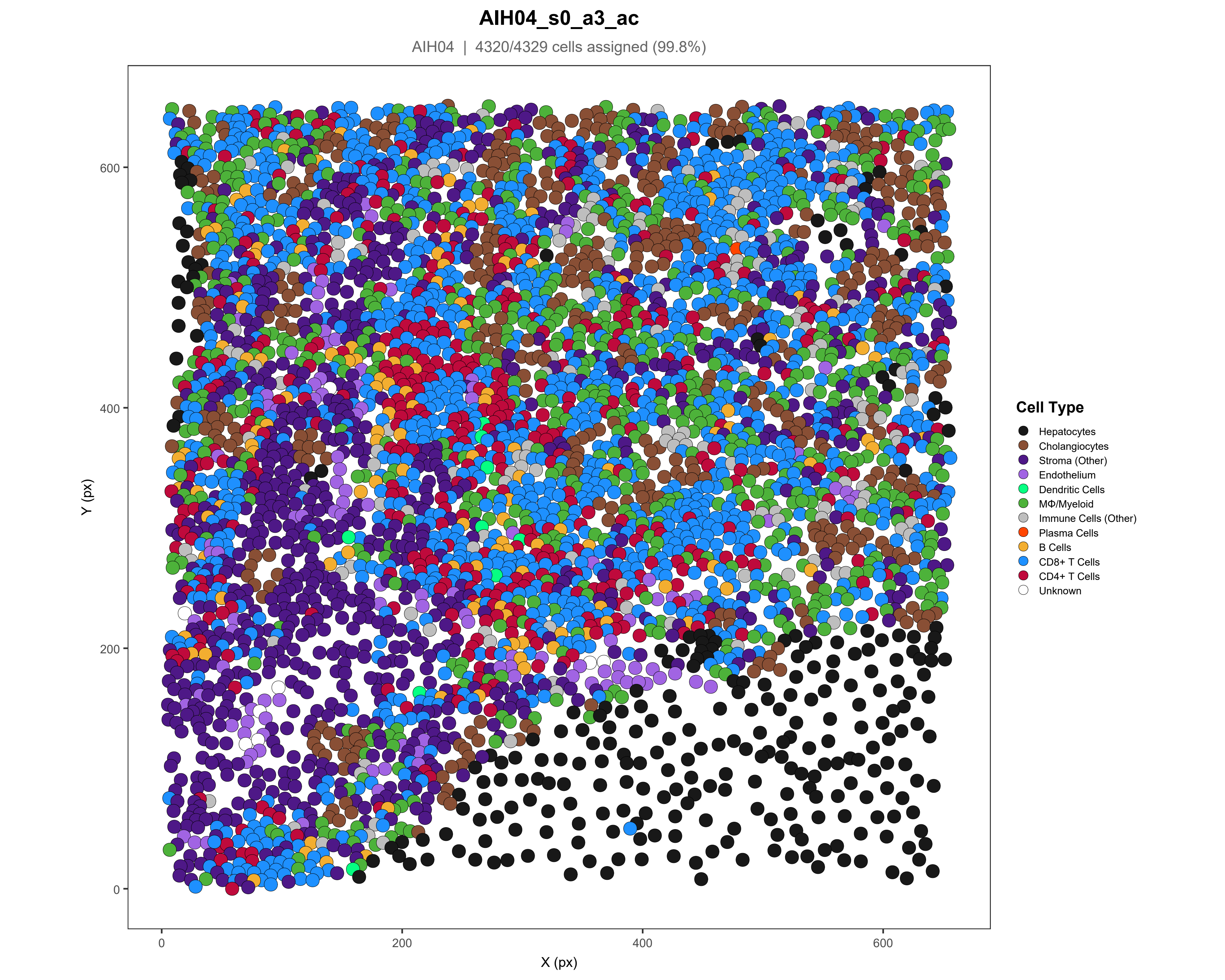

AIH04_s0_a3_ac
AIH04 | 4320/4329 cells assigned (99.8%)
Y (px)
X (px)
0
200
400
600
Cell Type
Hepatocytes
Cholangiocytes
Stroma (Other)
Endothelium
Dendritic Cells
MΦ/Myeloid
Immune Cells (Other)
Plasma Cells
B Cells
CD8+ T Cells
CD4+ T Cells
Unknown

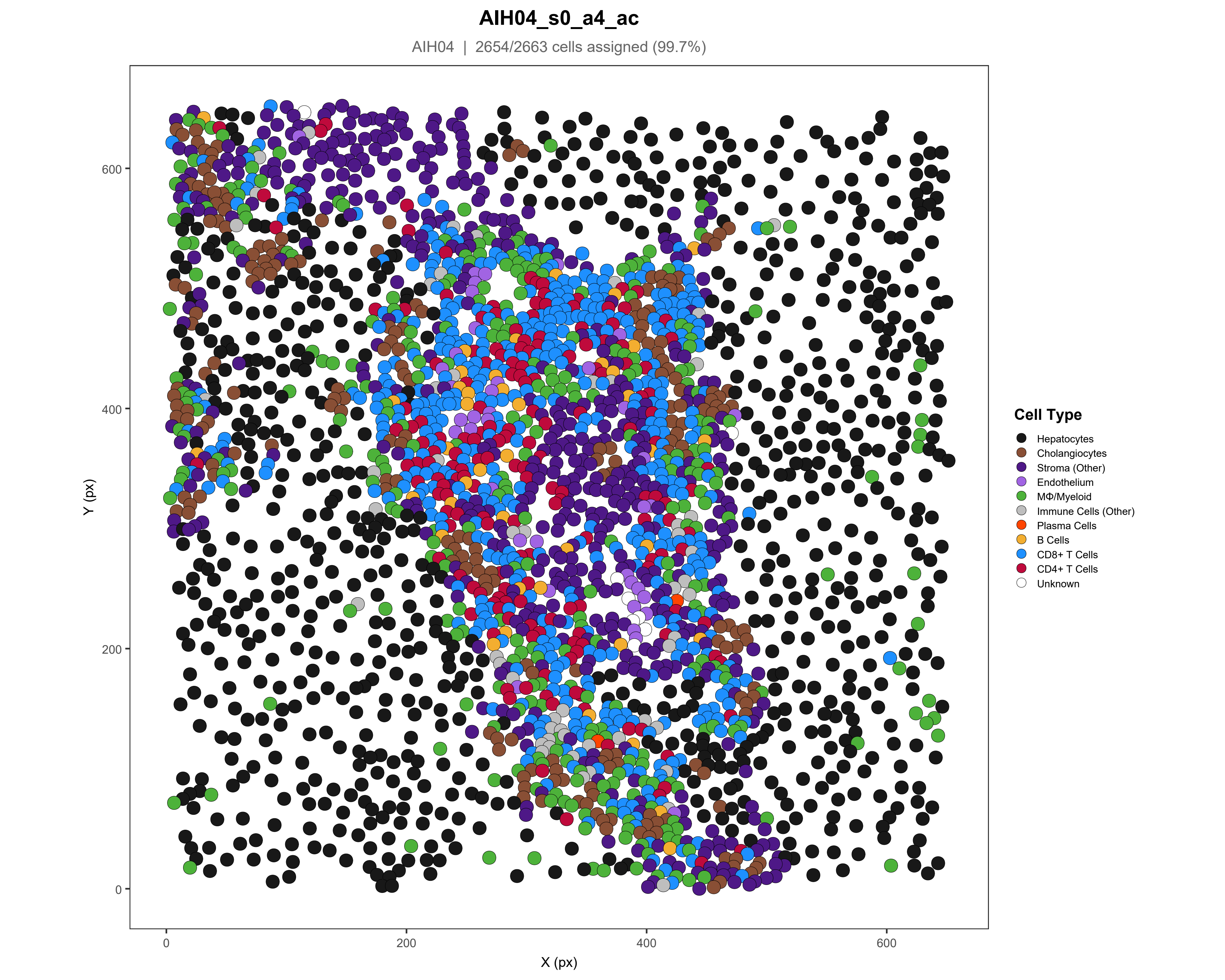
AIH04_s0_a4_ac
AIH04 | 2654/2663 cells assigned (99.7%)
Y (px)
X (px)
0
200
400
600
Cell Type
Hepatocytes
Cholangiocytes
Stroma (Other)
Endothelium
MΦ/Myeloid
Immune Cells (Other)
Plasma Cells
B Cells
CD8+ T Cells
CD4+ T Cells
Unknown

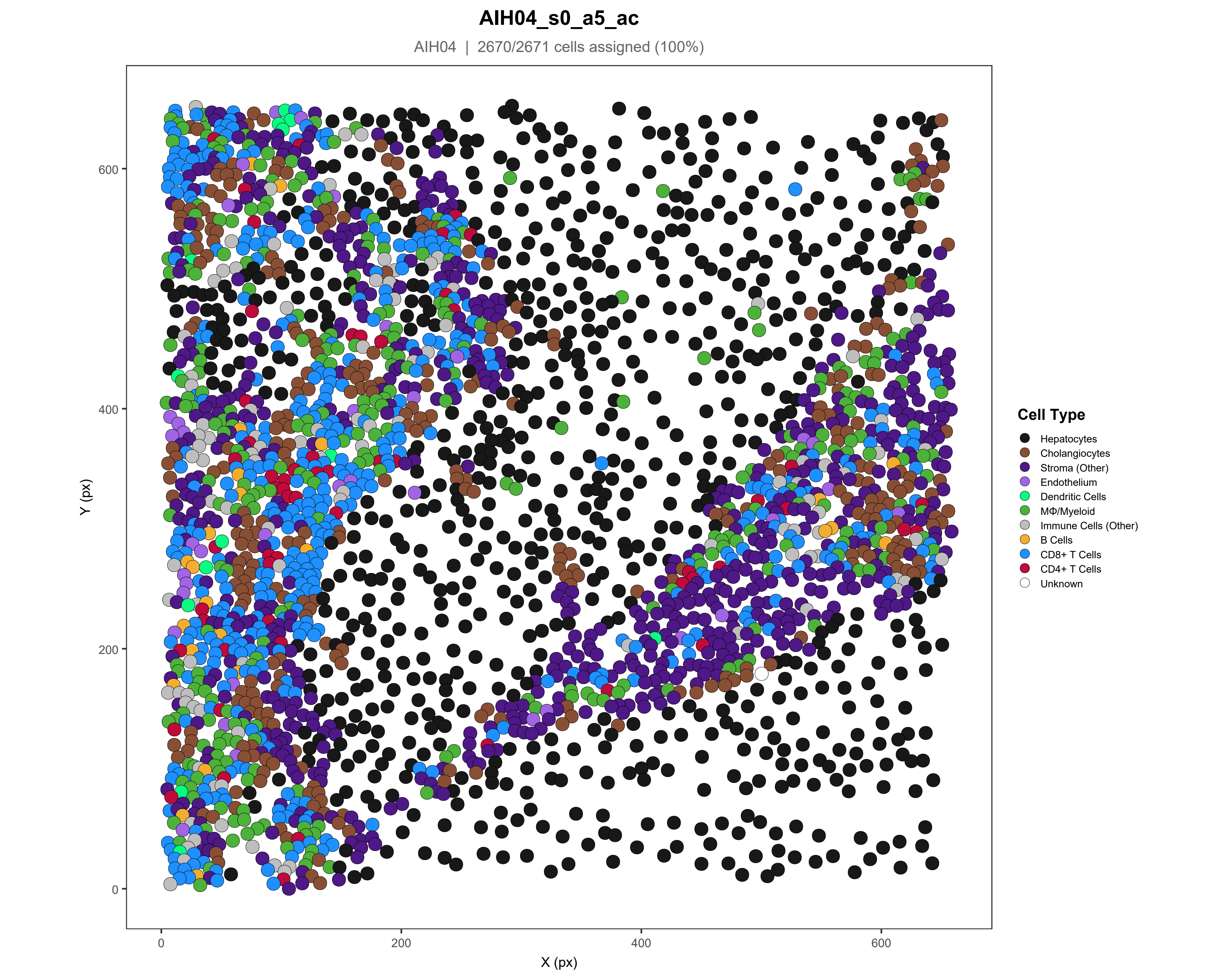
AIH04_s0_a5_ac
AIH04 | 2670/2671 cells assigned (100%)
Y (px)
X (px)
0
200
400
600
Cell Type
Hepatocytes
Cholangiocytes
Stroma (Other)
Endothelium
Dendritic Cells
MΦ/Myeloid
Immune Cells (Other)
B Cells
CD8+ T Cells
CD4+ T Cells
Unknown

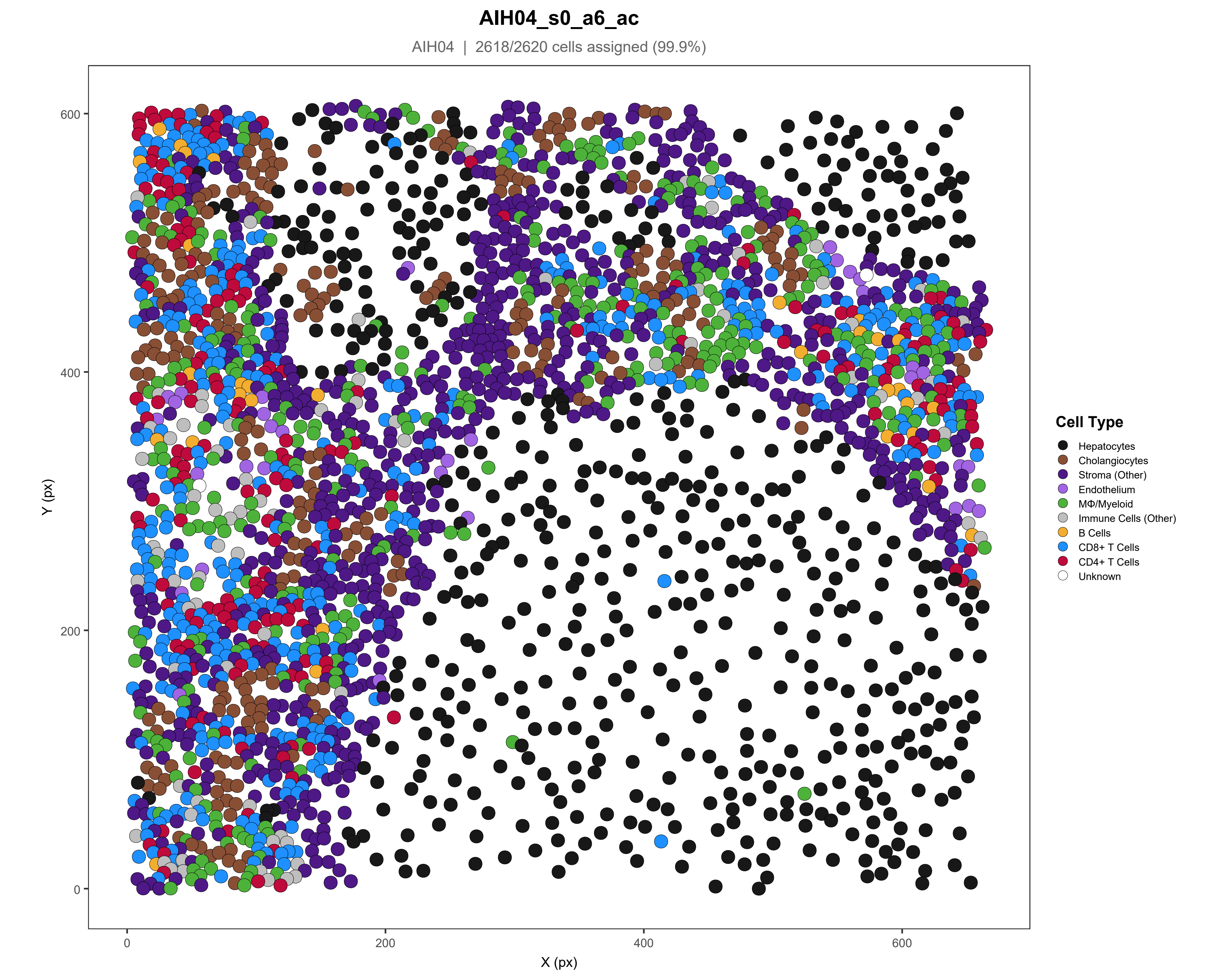

AIH04_s0_a6_ac
AIH04 | 2618/2620 cells assigned (99.9%)
Y (px)
X (px)
0
200
400
600
Cell Type
Hepatocytes
Cholangiocytes
Stroma (Other)
Endothelium
MΦ/Myeloid
Immune Cells (Other)
B Cells
CD8+ T Cells
CD4+ T Cells
Unknown

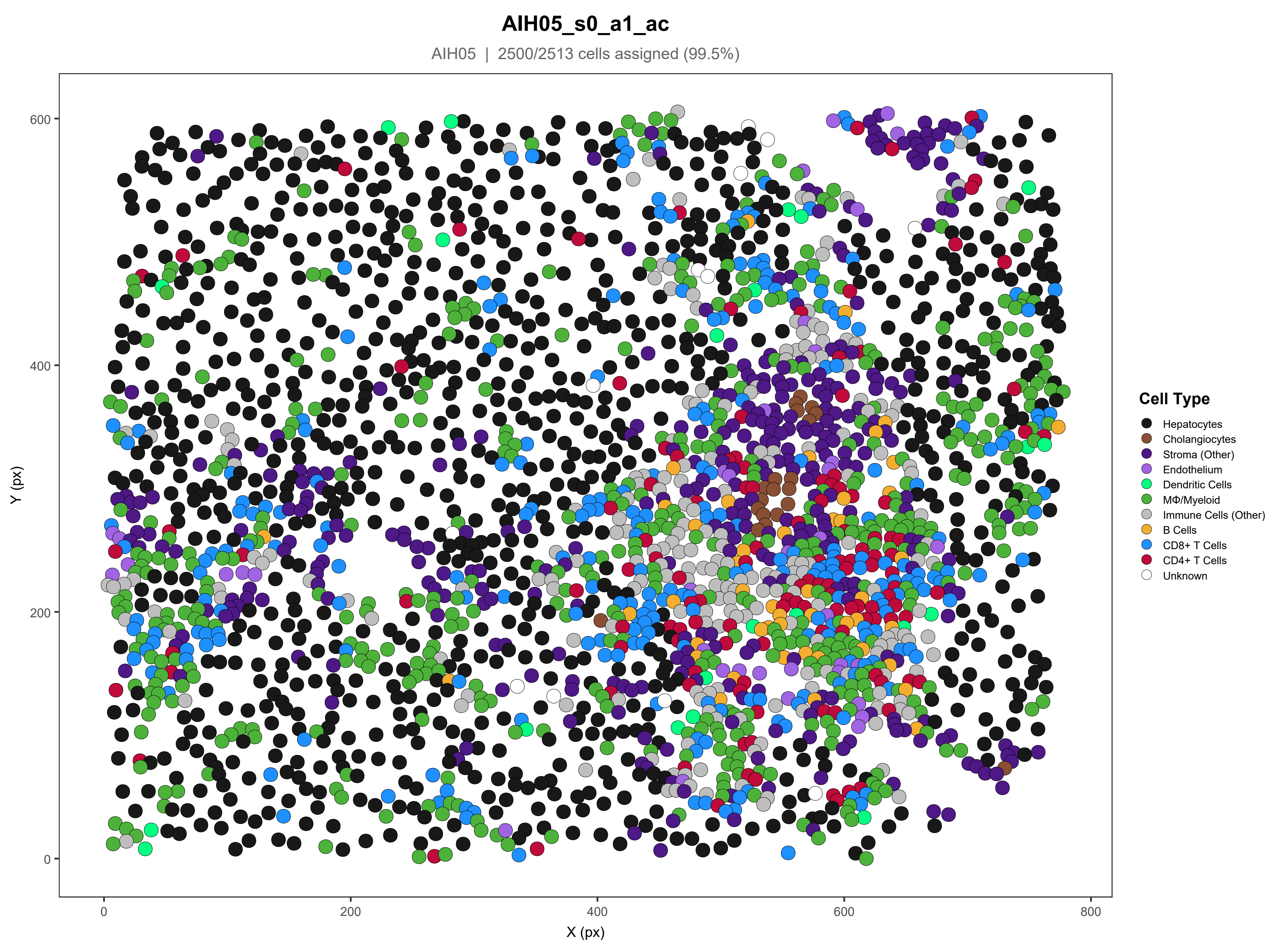

AIH05_s0_a1_ac
AIH05 | 2500/2513 cells assigned (99.5%)
Y (px)
X (px)
0
200
400
600
800
Cell Type
Hepatocytes
Cholangiocytes
Stroma (Other)
Endothelium
Dendritic Cells
MΦ/Myeloid
Immune Cells (Other)
B Cells
CD8+ T Cells
CD4+ T Cells
Unknown

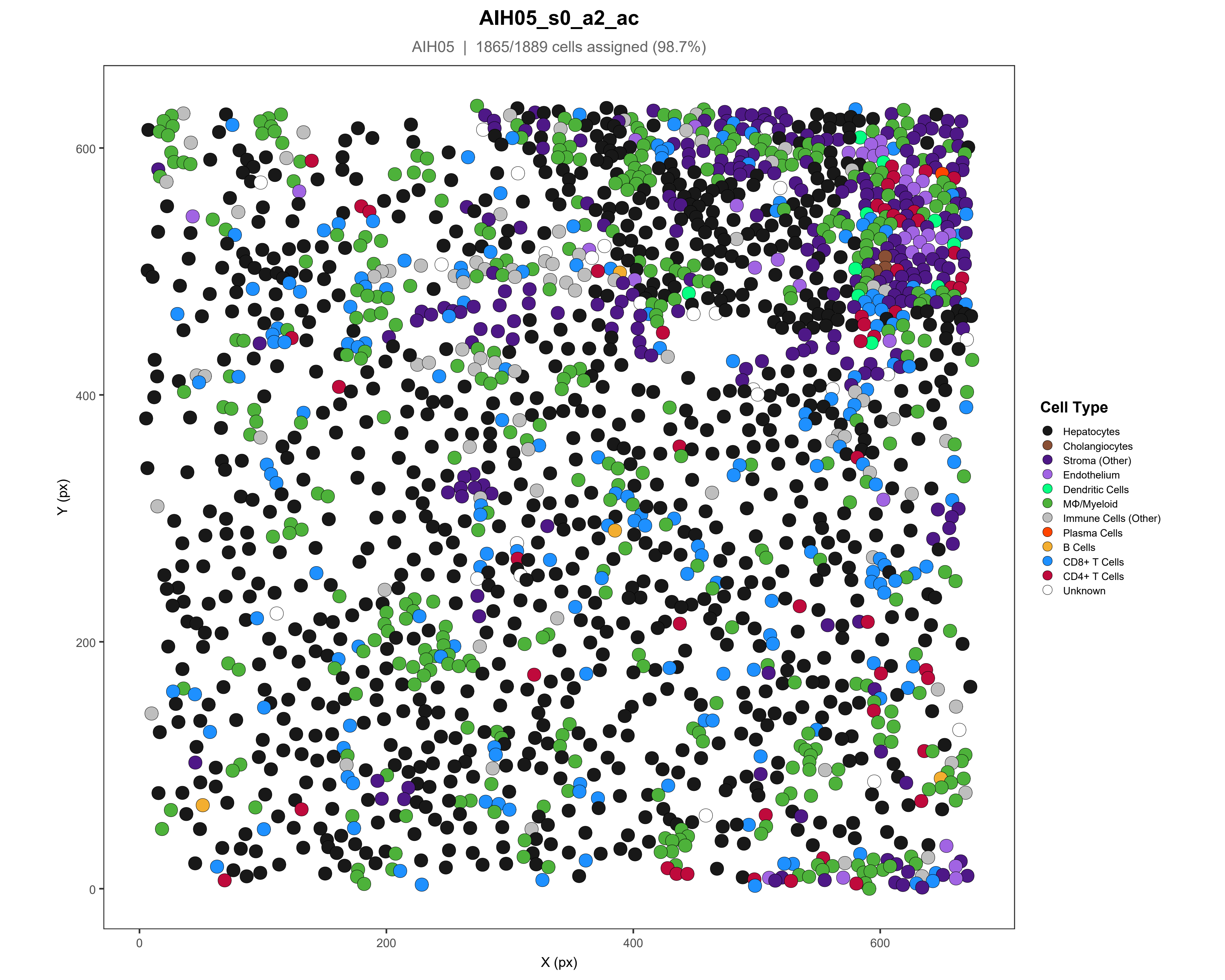
AIH05_s0_a2_ac
AIH05 | 1865/1889 cells assigned (98.7%)
Y (px)
X (px)
0
200
400
600
Cell Type
Hepatocytes
Cholangiocytes
Stroma (Other)
Endothelium
Dendritic Cells
MΦ/Myeloid
Immune Cells (Other)
Plasma Cells
B Cells
CD8+ T Cells
CD4+ T Cells
Unknown

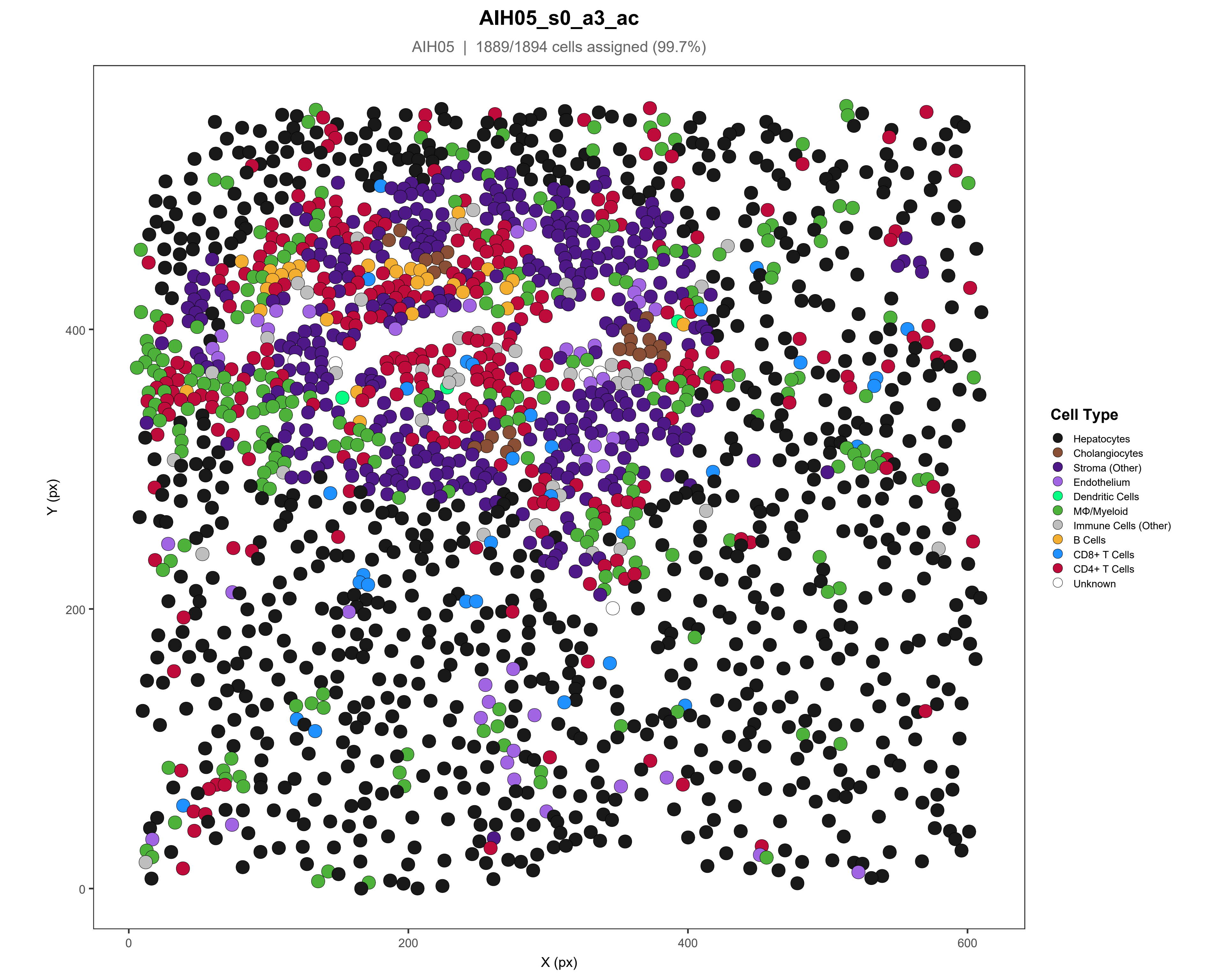
AIH05_s0_a3_ac
AIH05 | 1889/1894 cells assigned (99.7%)
Y (px)
X (px)
0
200
400
600
Cell Type
Hepatocytes
Cholangiocytes
Stroma (Other)
Endothelium
Dendritic Cells
MΦ/Myeloid
Immune Cells (Other)
B Cells
CD8+ T Cells
CD4+ T Cells
Unknown

### AIH05_s0_a5_ac

AIH05 | 1818/1834 cells assigned (99.1%)

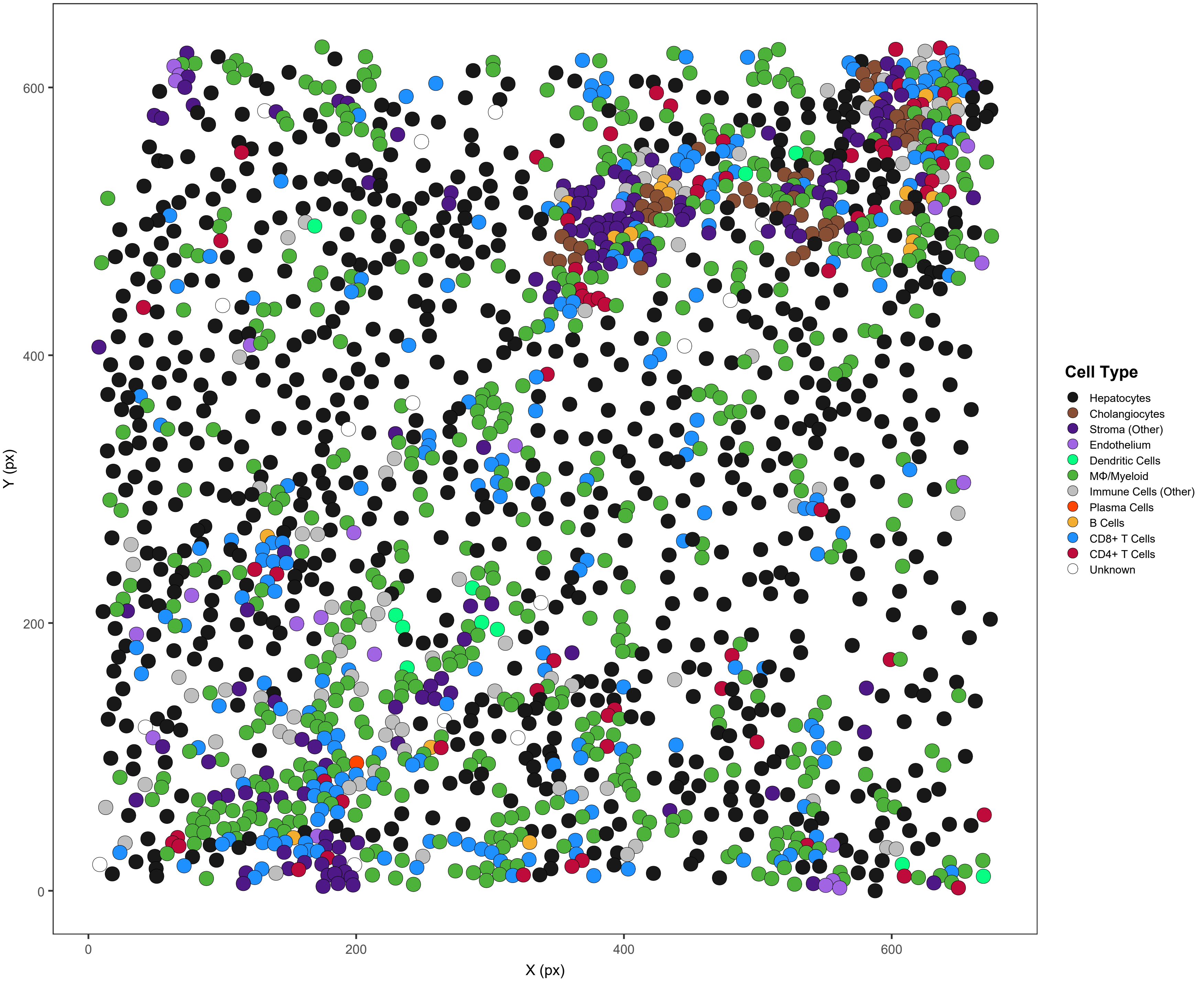

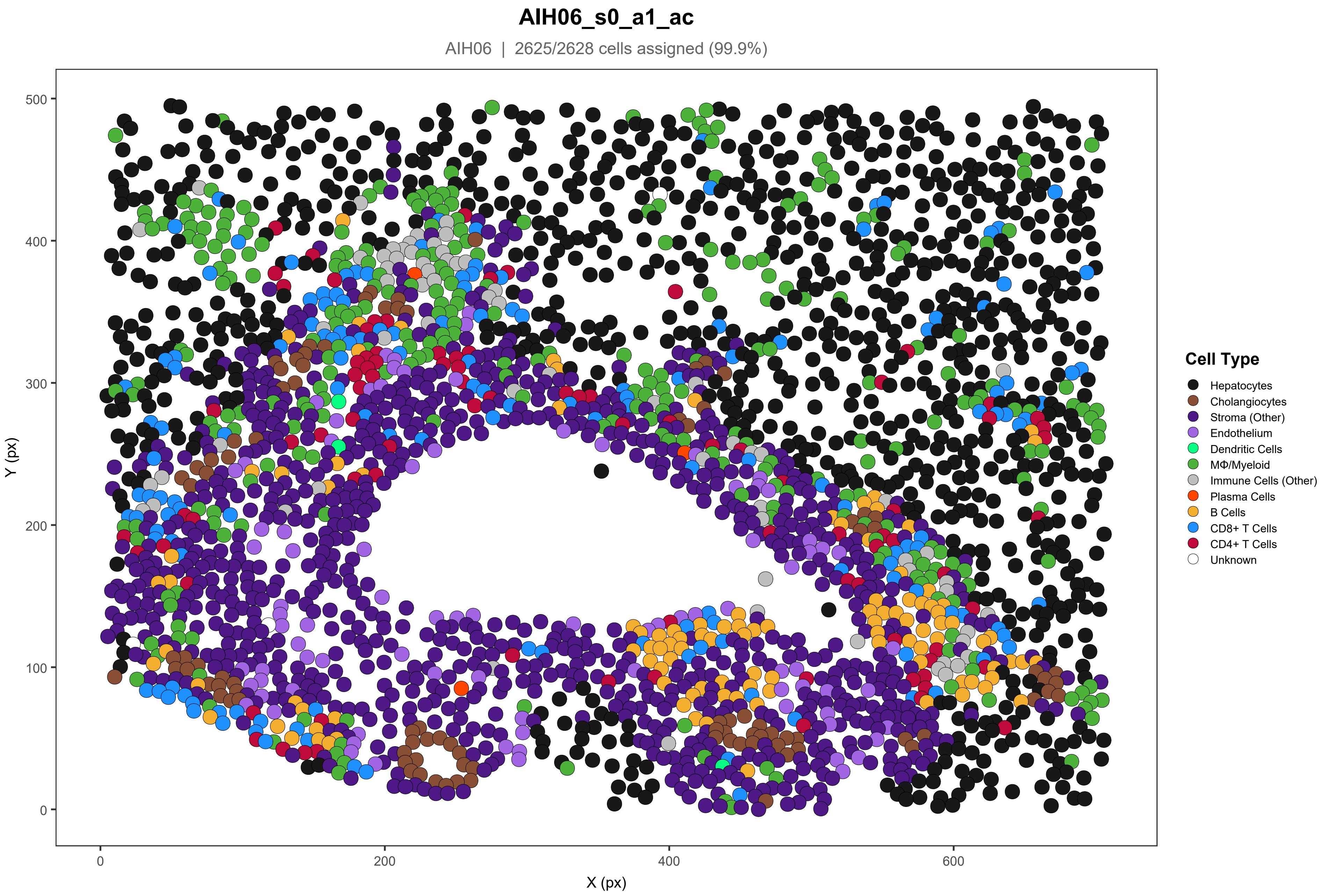

AIH06_s0_a1_ac
AIH06 | 2625/2628 cells assigned (99.9%)
Y (px)
X (px)
0
100
200
300
400
500
0
200
400
600
Cell Type
Hepatocytes
Cholangiocytes
Stroma (Other)
Endothelium
Dendritic Cells
MΦ/Myeloid
Immune Cells (Other)
Plasma Cells
B Cells
CD8+ T Cells
CD4+ T Cells
Unknown

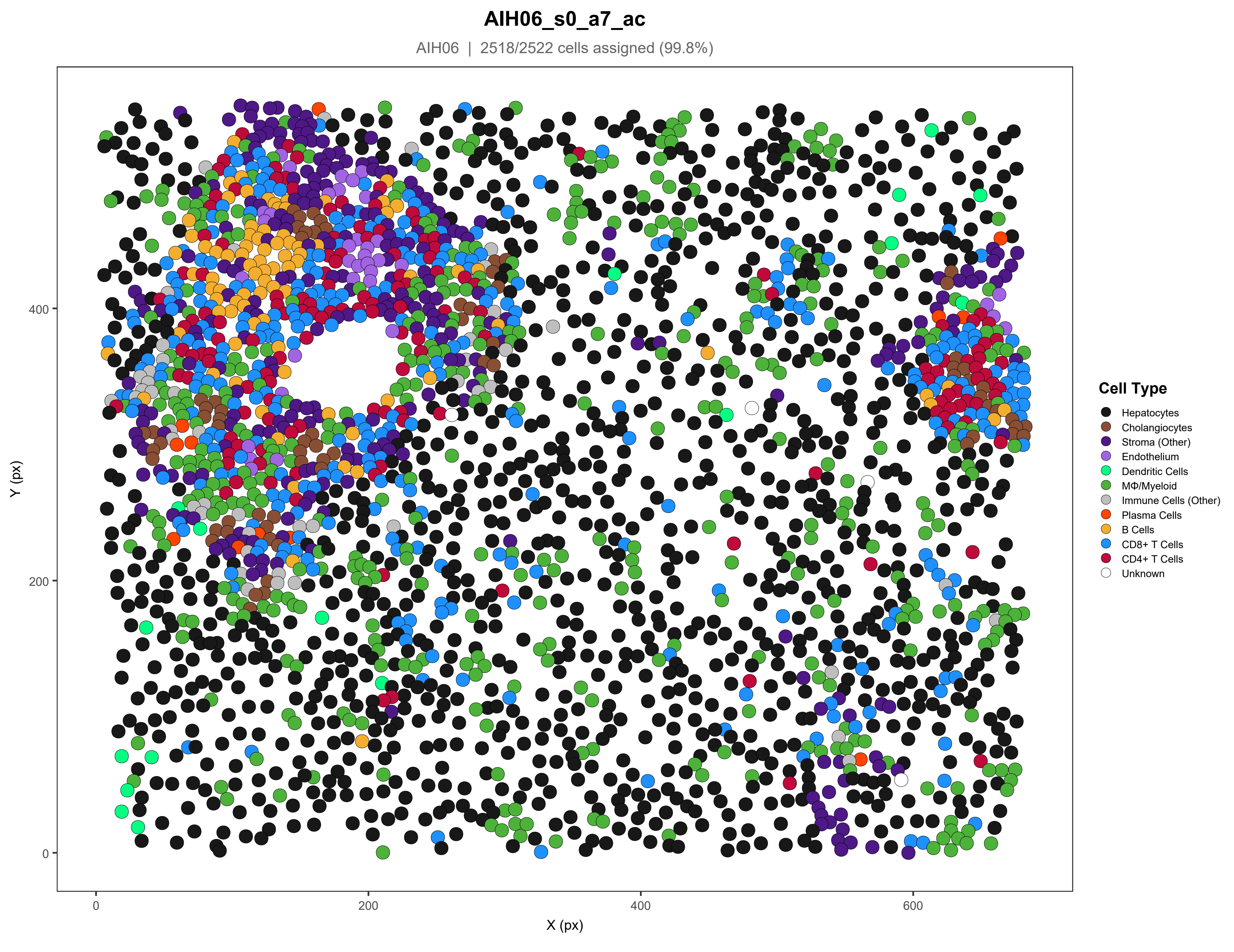
AIH06_s0_a7_ac
AIH06 | 2518/2522 cells assigned (99.8%)
Y (px)
X (px)
0
200
400
600
Cell Type
Hepatocytes
Cholangiocytes
Stroma (Other)
Endothelium
Dendritic Cells
MΦ/Myeloid
Immune Cells (Other)
Plasma Cells
B Cells
CD8+ T Cells
CD4+ T Cells
Unknown

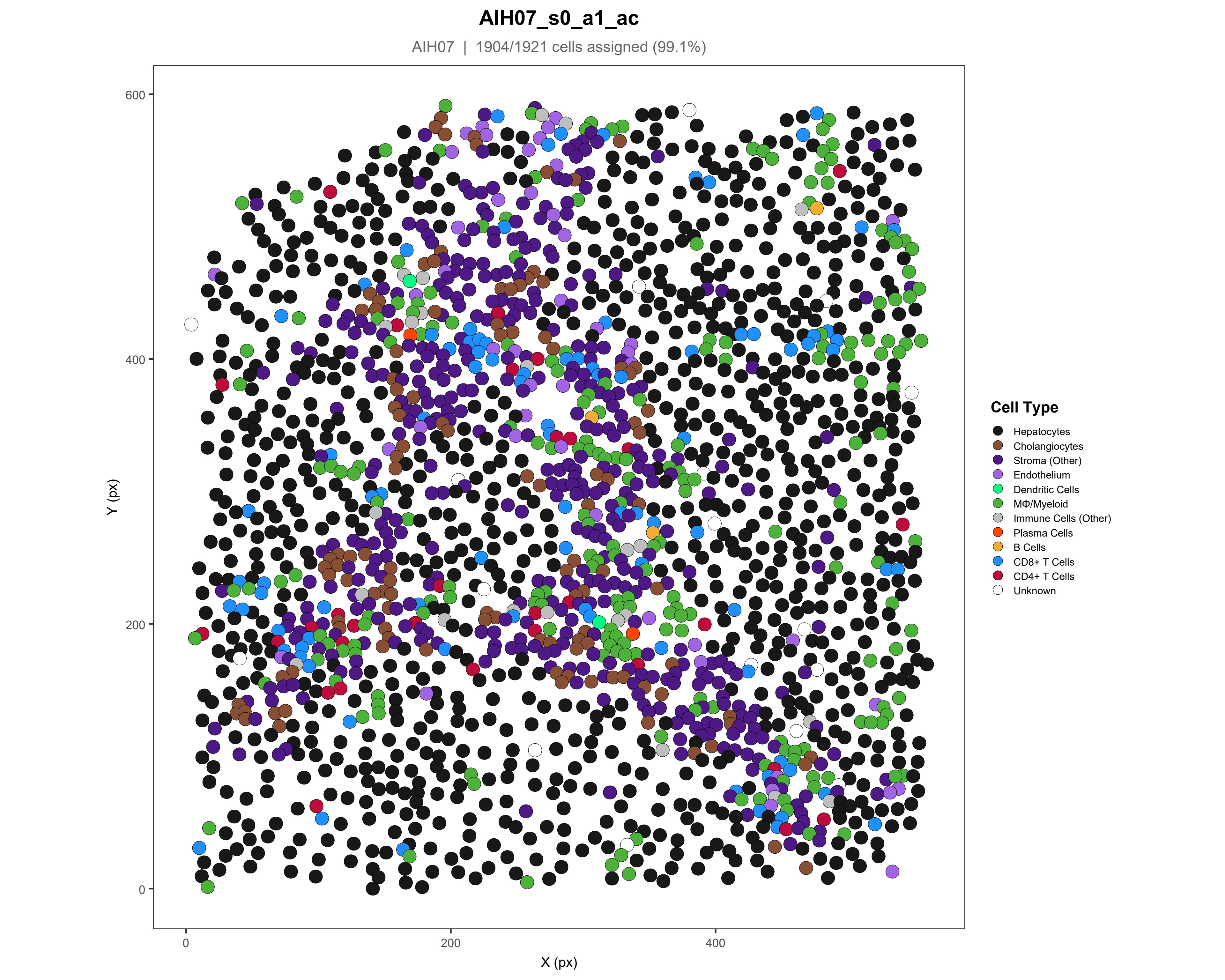
AIH07_s0_a1_ac
AIH07 | 1904/1921 cells assigned (99.1%)
Y (px)
X (px)
0
200
400
600
Cell Type
Hepatocytes
Cholangiocytes
Stroma (Other)
Endothelium
Dendritic Cells
MΦ/Myeloid
Immune Cells (Other)
Plasma Cells
B Cells
CD8+ T Cells
CD4+ T Cells
Unknown

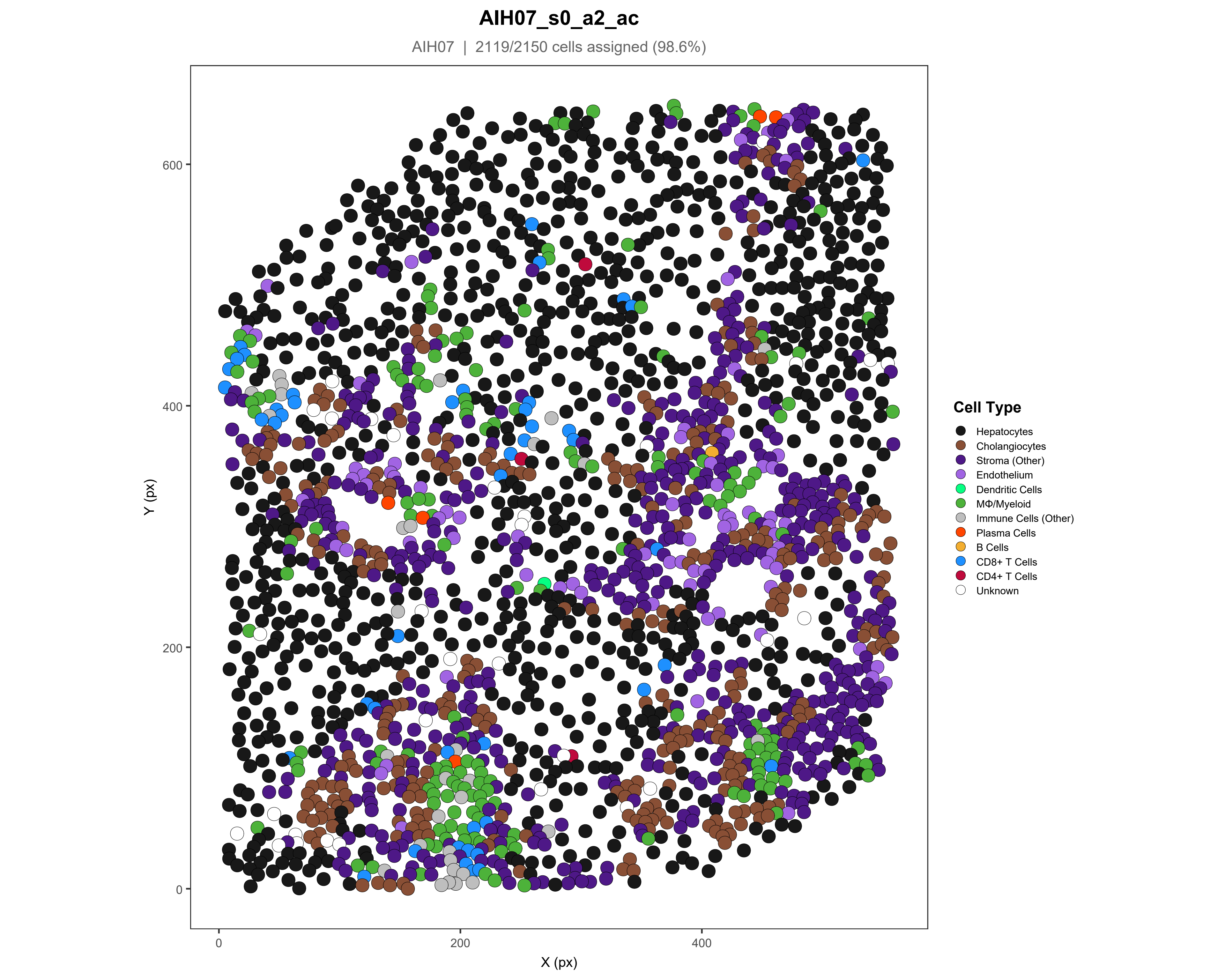
AIH07_s0_a2_ac
AIH07 | 2119/2150 cells assigned (98.6%)
Y (px)
X (px)
0
200
400
600
Cell Type
Hepatocytes
Cholangiocytes
Stroma (Other)
Endothelium
Dendritic Cells
MΦ/Myeloid
Immune Cells (Other)
Plasma Cells
B Cells
CD8+ T Cells
CD4+ T Cells
Unknown

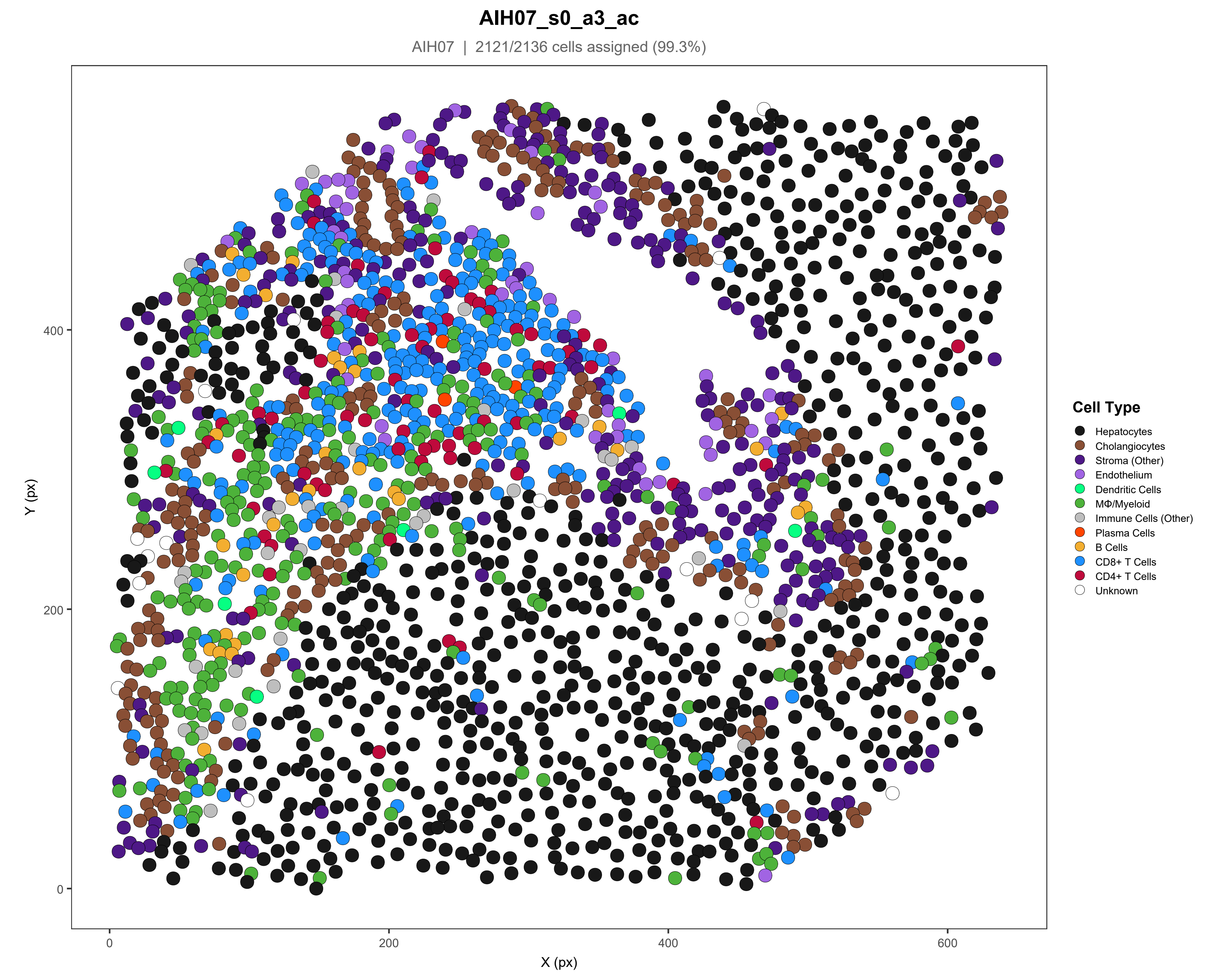
AIH07_s0_a3_ac
AIH07 | 2121/2136 cells assigned (99.3%)
Cell Type
Hepatocytes
Cholangiocytes
Stroma (Other)
Endothelium
Dendritic Cells
MΦ/Myeloid
Immune Cells (Other)
Plasma Cells
B Cells
CD8+ T Cells
CD4+ T Cells
Unknown
Y (px)
X (px)
0
200
400
600

### AIH07_s0_a4_ac

AIH07 | 2105/2135 cells assigned (98.6%)

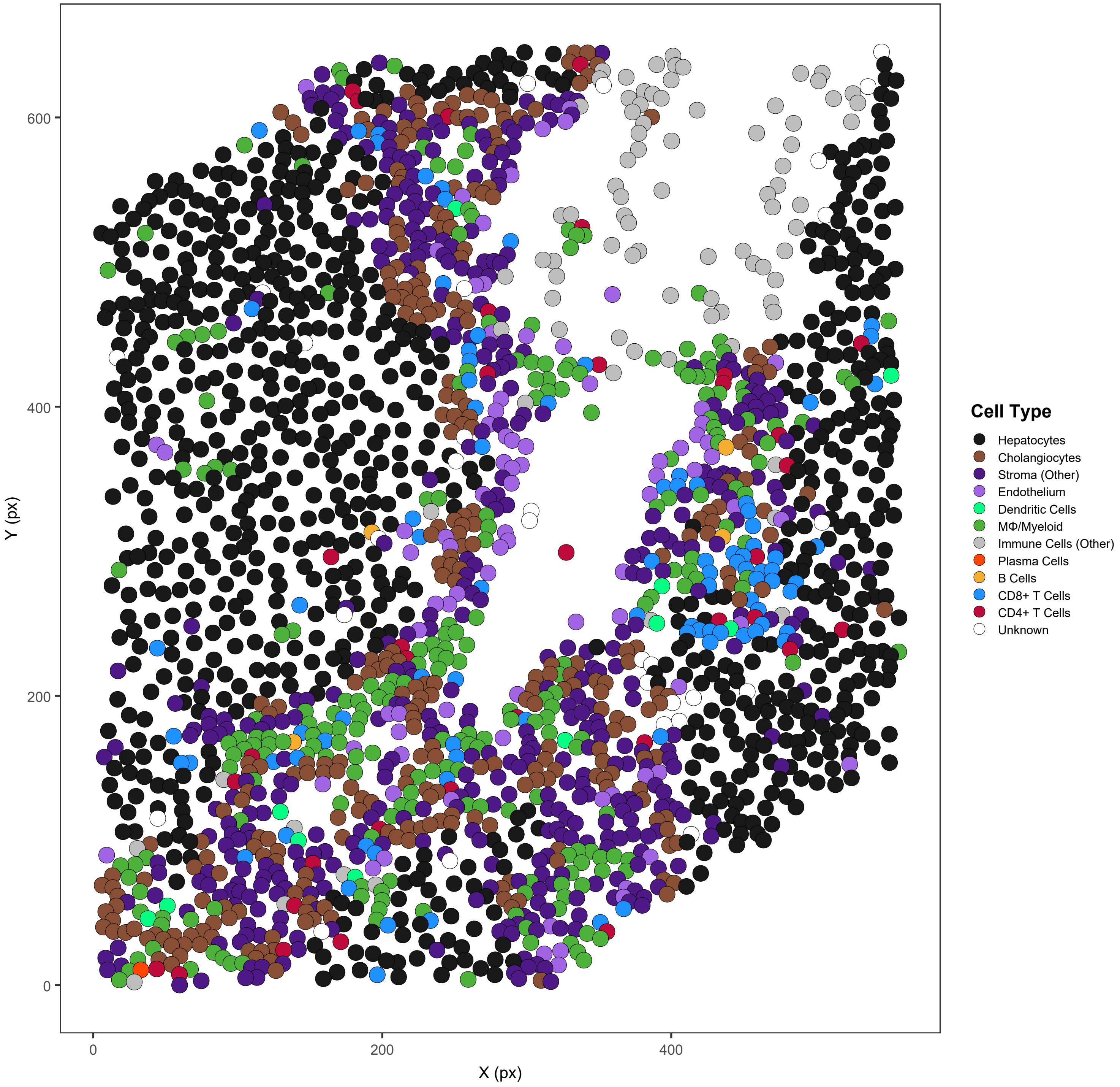

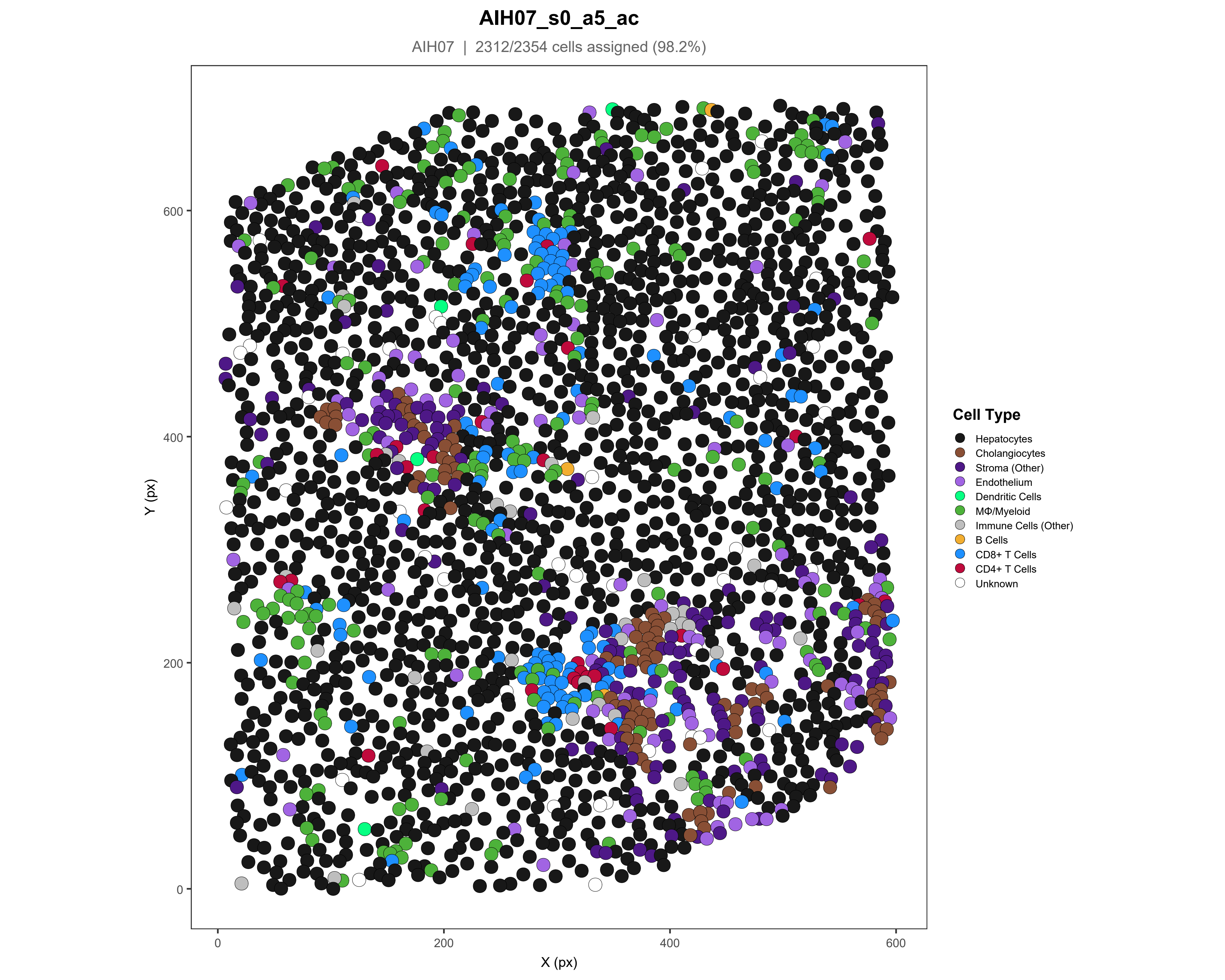

AIH07_s0_a5_ac
AIH07 | 2312/2354 cells assigned (98.2%)
Y (px)
X (px)
0
200
400
600
Cell Type
Hepatocytes
Cholangiocytes
Stroma (Other)
Endothelium
Dendritic Cells
MΦ/Myeloid
Immune Cells (Other)
B Cells
CD8+ T Cells
CD4+ T Cells
Unknown

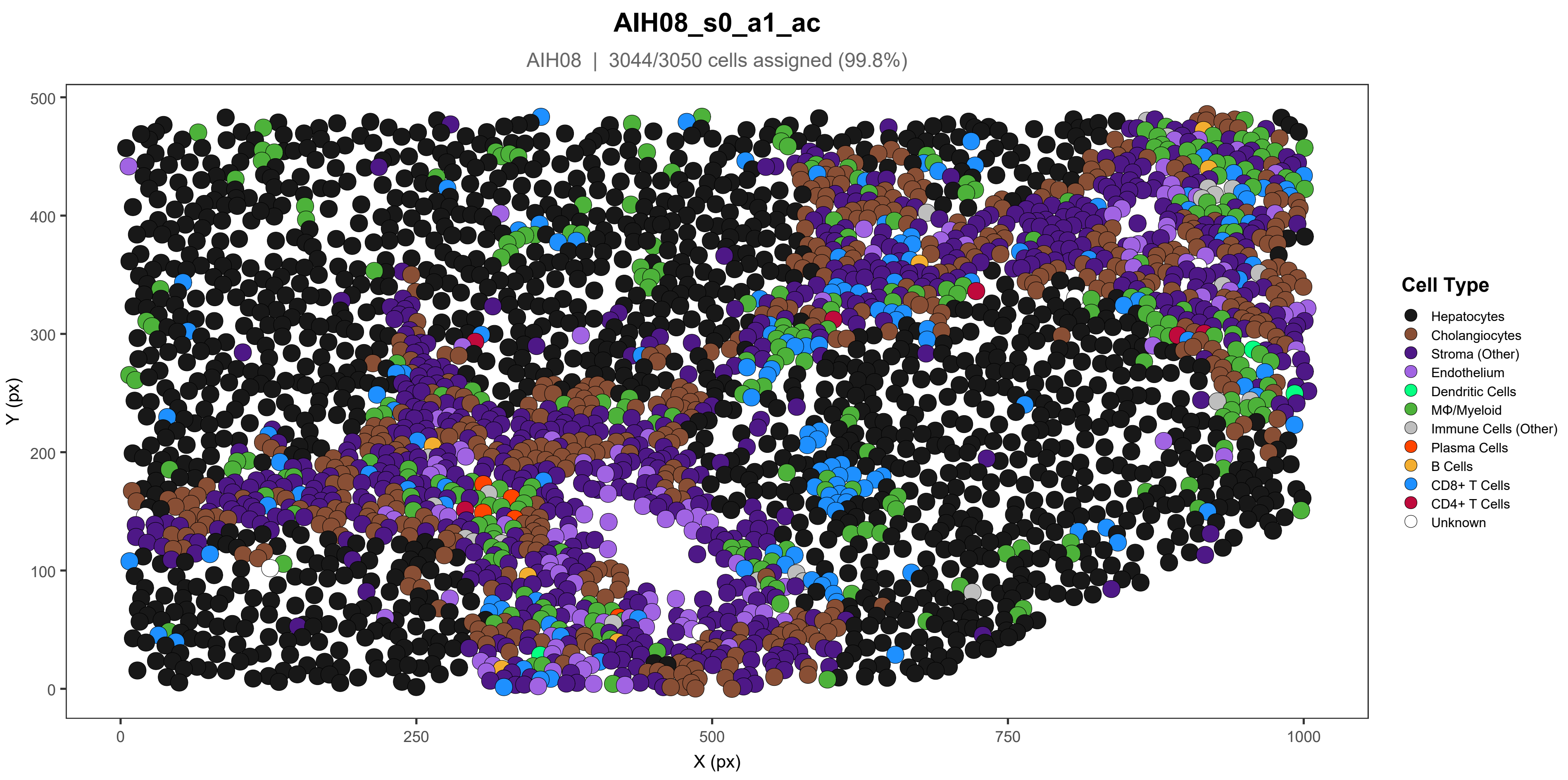
AIH08_s0_a1_ac
AIH08 | 3044/3050 cells assigned (99.8%)
Y (px)
X (px)
0
100
200
300
400
500
0
250
500
750
1000
Cell Type
Hepatocytes
Cholangiocytes
Stroma (Other)
Endothelium
Dendritic Cells
MΦ/Myeloid
Immune Cells (Other)
Plasma Cells
B Cells
CD8+ T Cells
CD4+ T Cells
Unknown

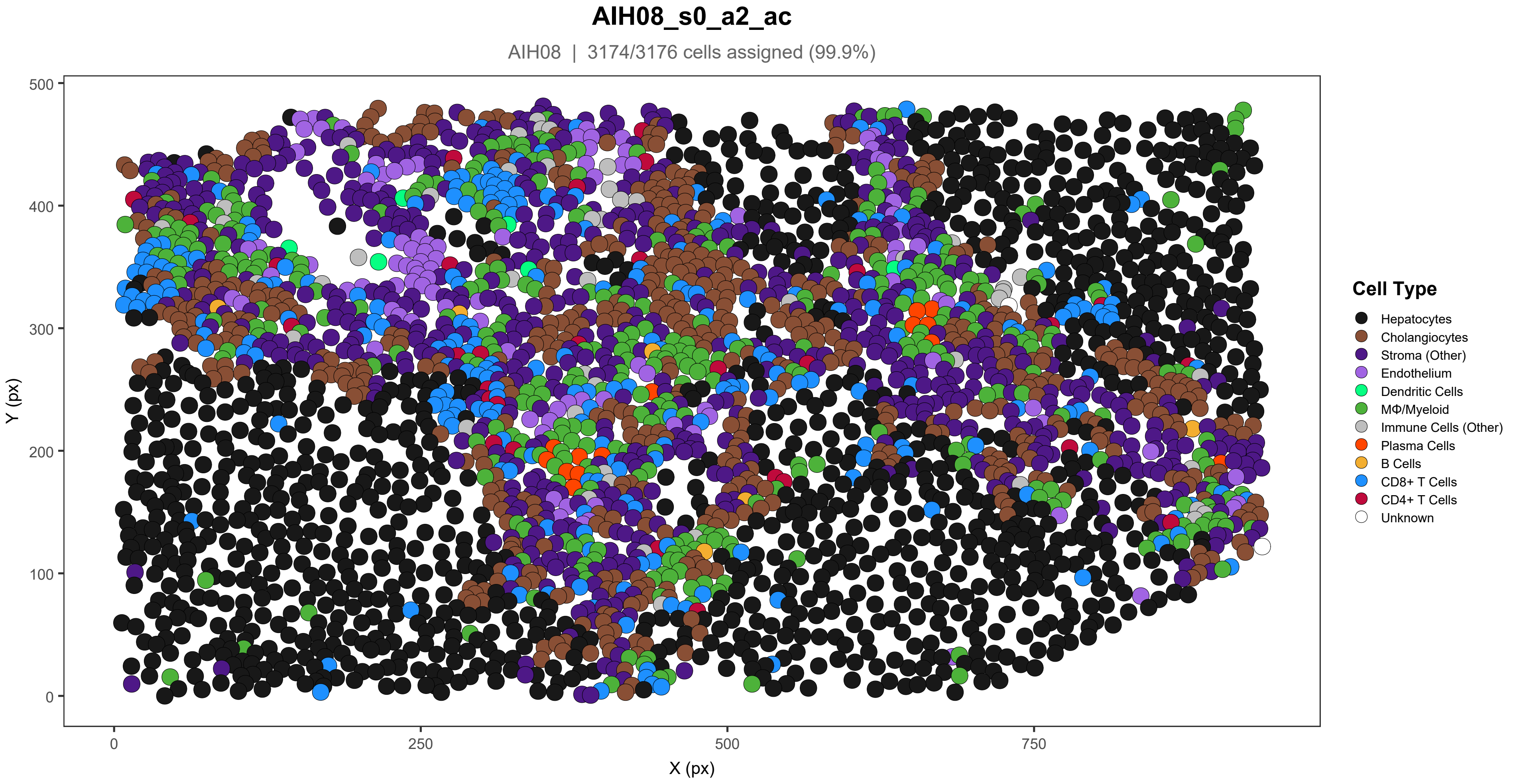

AIH08_s0_a2_ac
AIH08 | 3174/3176 cells assigned (99.9%)
Y (px)
X (px)
0
100
200
300
400
500
0
250
500
750
Cell Type
Hepatocytes
Cholangiocytes
Stroma (Other)
Endothelium
Dendritic Cells
MΦ/Myeloid
Immune Cells (Other)
Plasma Cells
B Cells
CD8+ T Cells
CD4+ T Cells
Unknown

#### AIH08_s0_a3_ac

AIH08 | 2208/2231 cells assigned (99%)

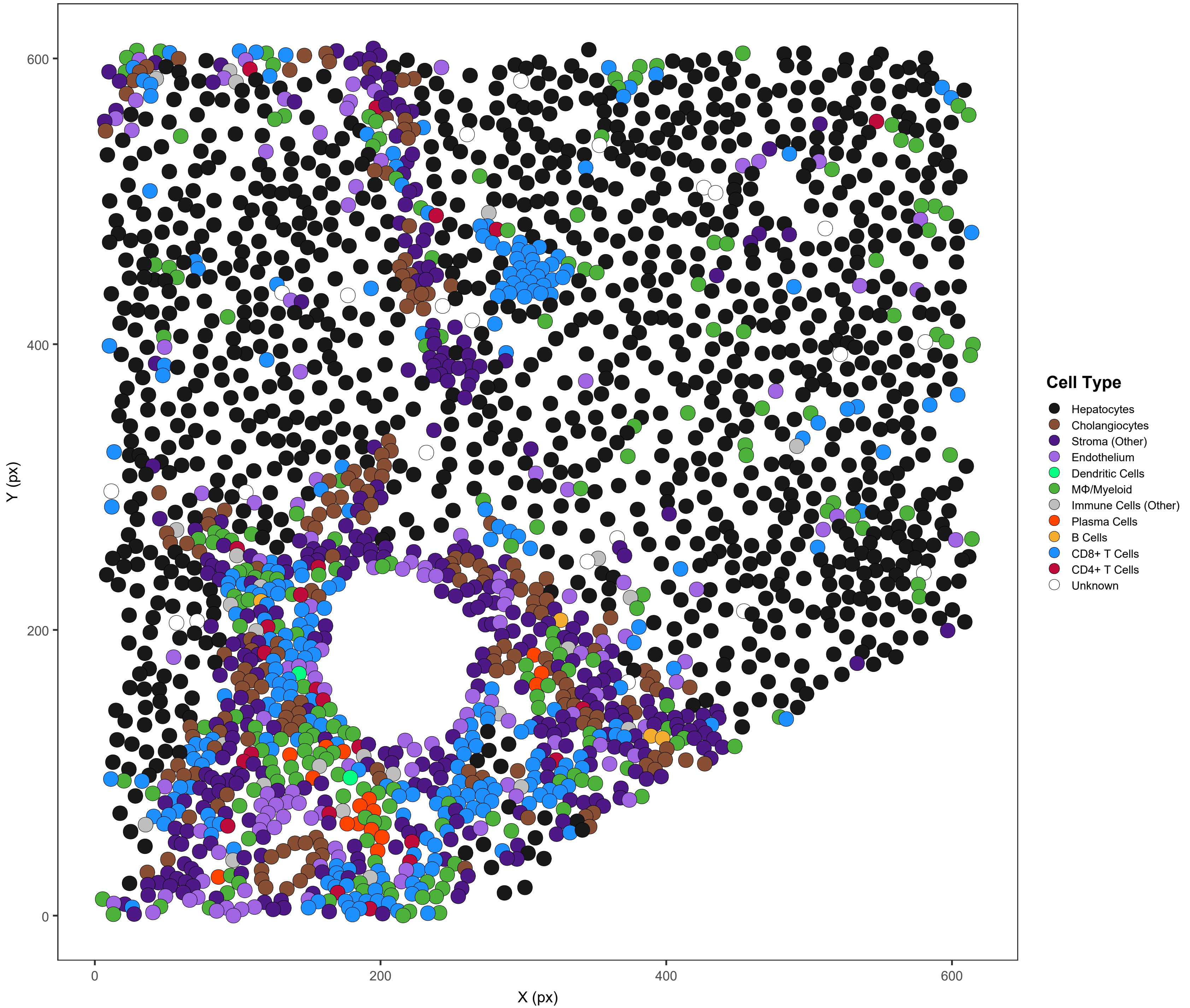

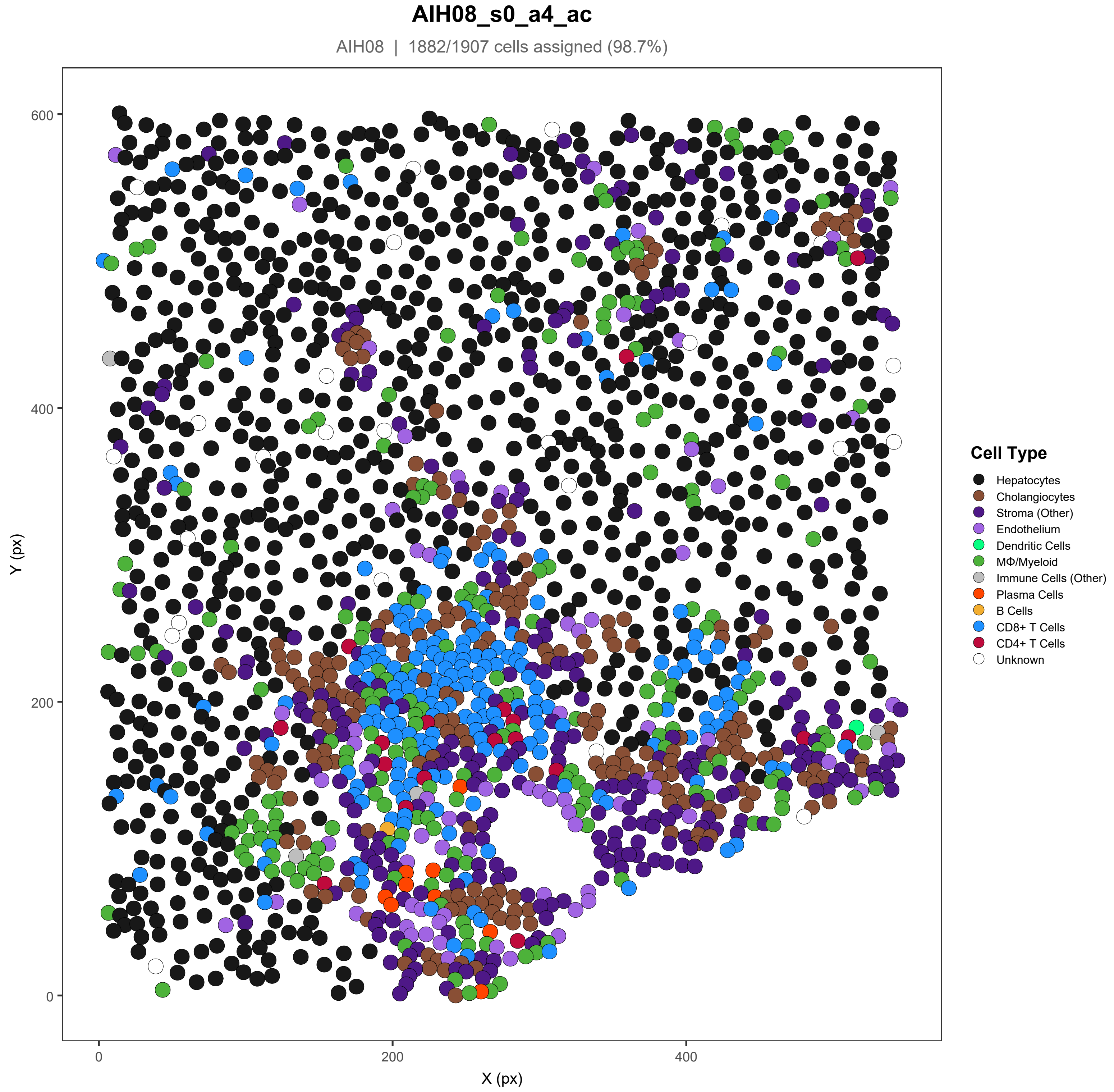
AIH08_s0_a4_ac
AIH08 | 1882/1907 cells assigned (98.7%)
Y (px)
X (px)
0
200
400
600
Cell Type
Hepatocytes
Cholangiocytes
Stroma (Other)
Endothelium
Dendritic Cells
MΦ/Myeloid
Immune Cells (Other)
Plasma Cells
B Cells
CD8+ T Cells
CD4+ T Cells
Unknown

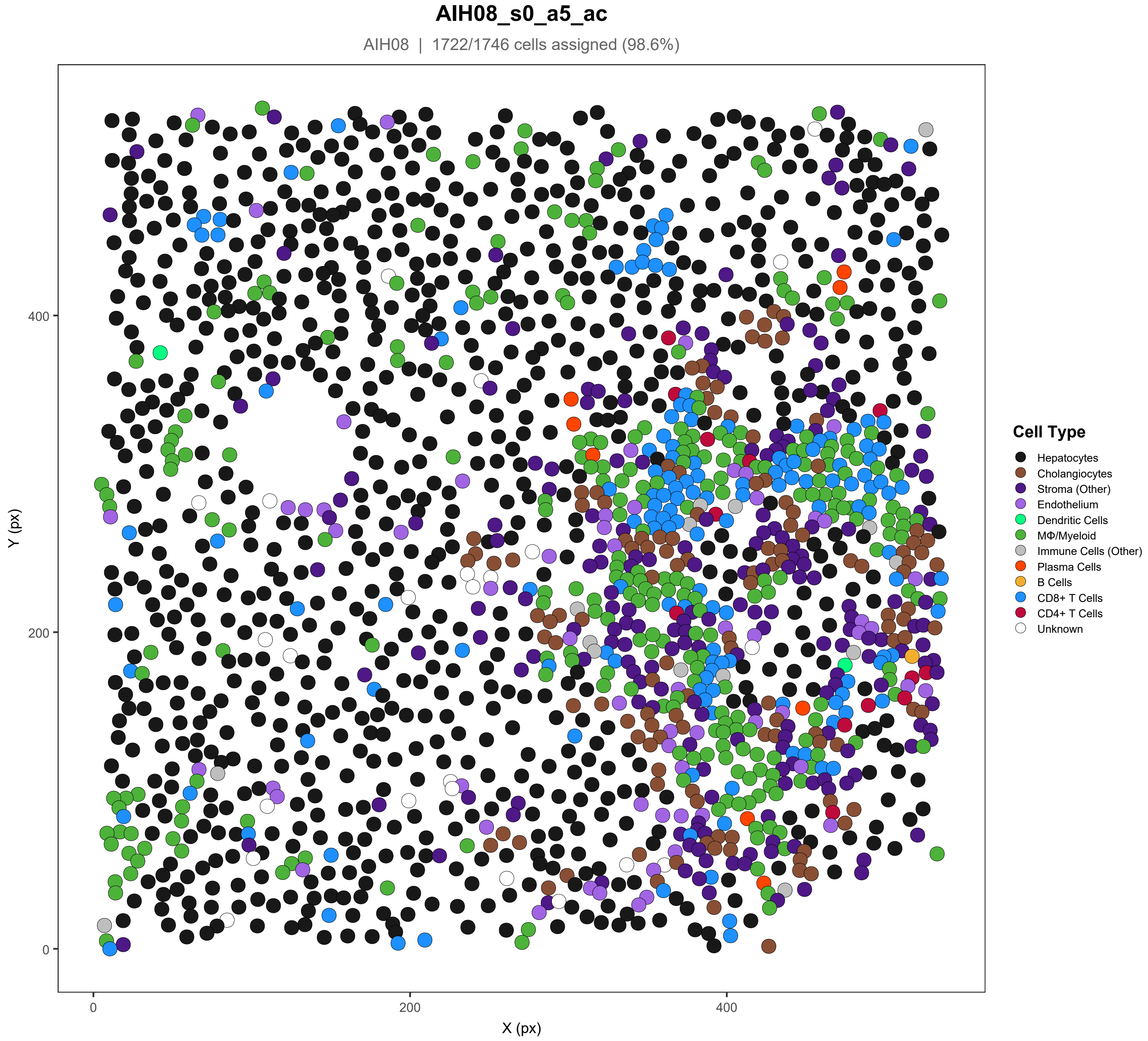

AIH08_s0_a5_ac
AIH08 | 1722/1746 cells assigned (98.6%)
Y (px)
X (px)
0
200
400
Cell Type
Hepatocytes
Cholangiocytes
Stroma (Other)
Endothelium
Dendritic Cells
MΦ/Myeloid
Immune Cells (Other)
Plasma Cells
B Cells
CD8+ T Cells
CD4+ T Cells
Unknown

#### AIH09_s0_a1_ac

AIH09 | 4345/4370 cells assigned (99.4%)

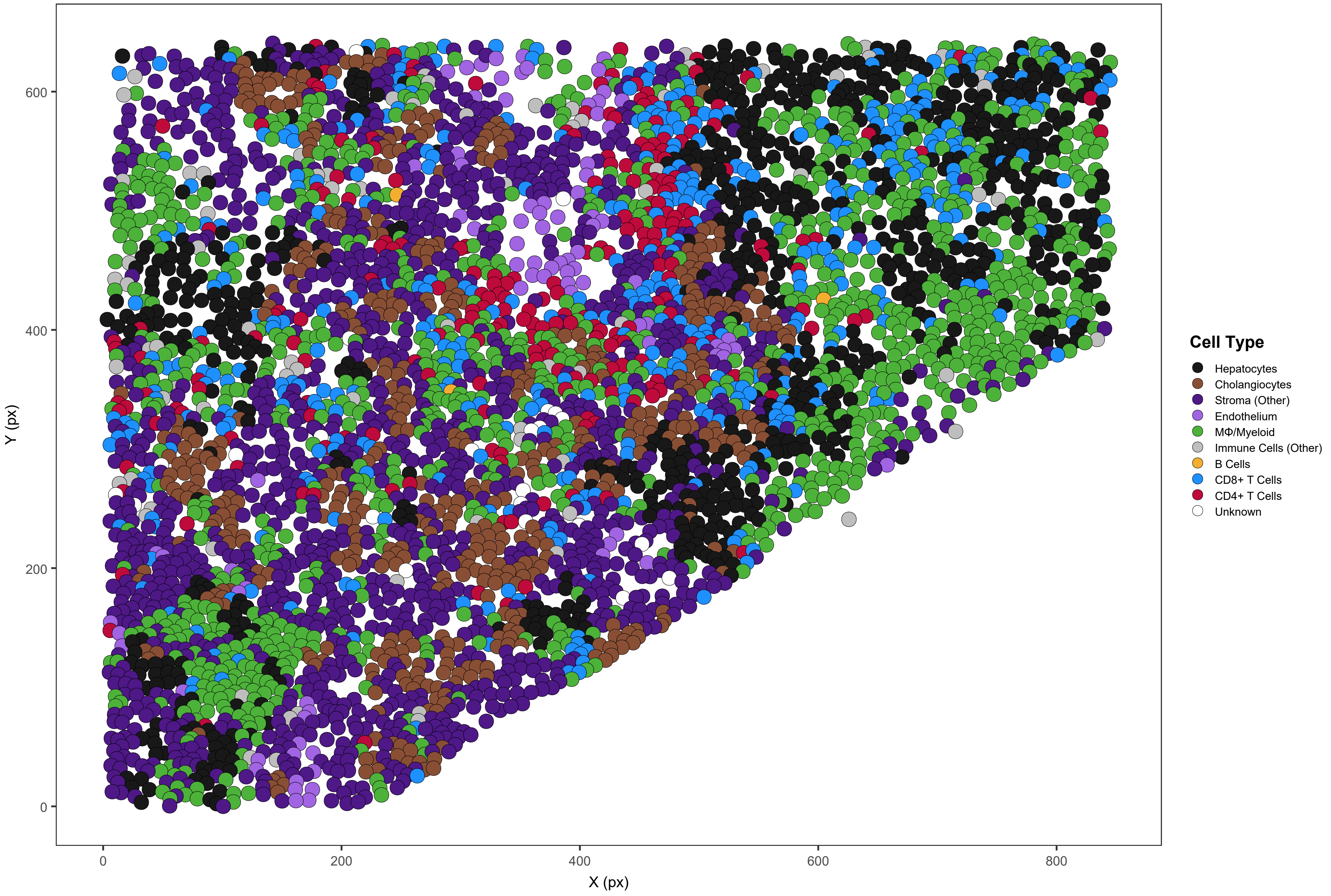

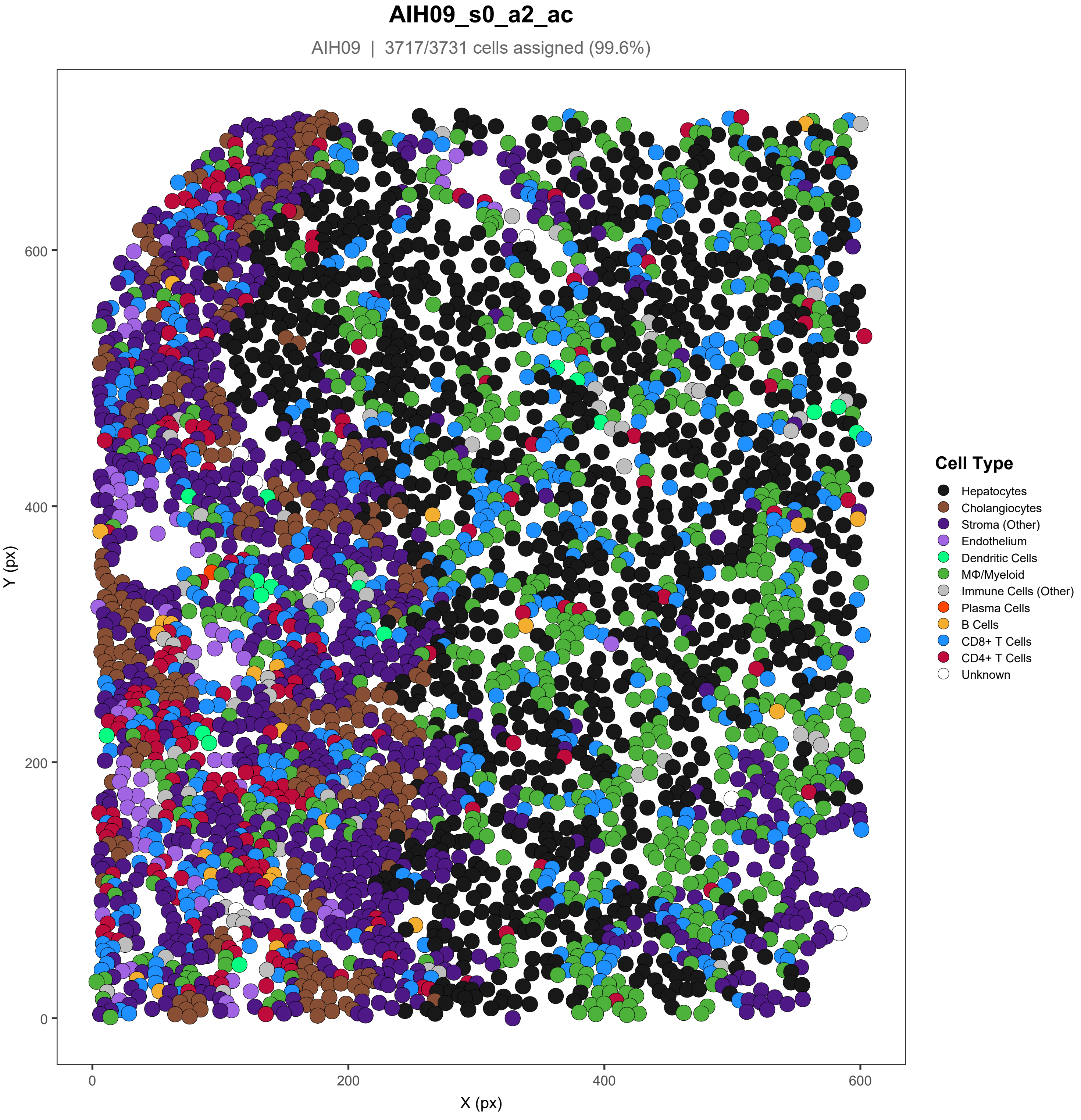
AIH09_s0_a2_ac
AIH09 | 3717/3731 cells assigned (99.6%)
Y (px)
X (px)
0
200
400
600
Cell Type
Hepatocytes
Cholangiocytes
Stroma (Other)
Endothelium
Dendritic Cells
MΦ/Myeloid
Immune Cells (Other)
Plasma Cells
B Cells
CD8+ T Cells
CD4+ T Cells
Unknown

### AIH09_s0_a3_ac

AIH09 | 3408/3413 cells assigned (99.9%)

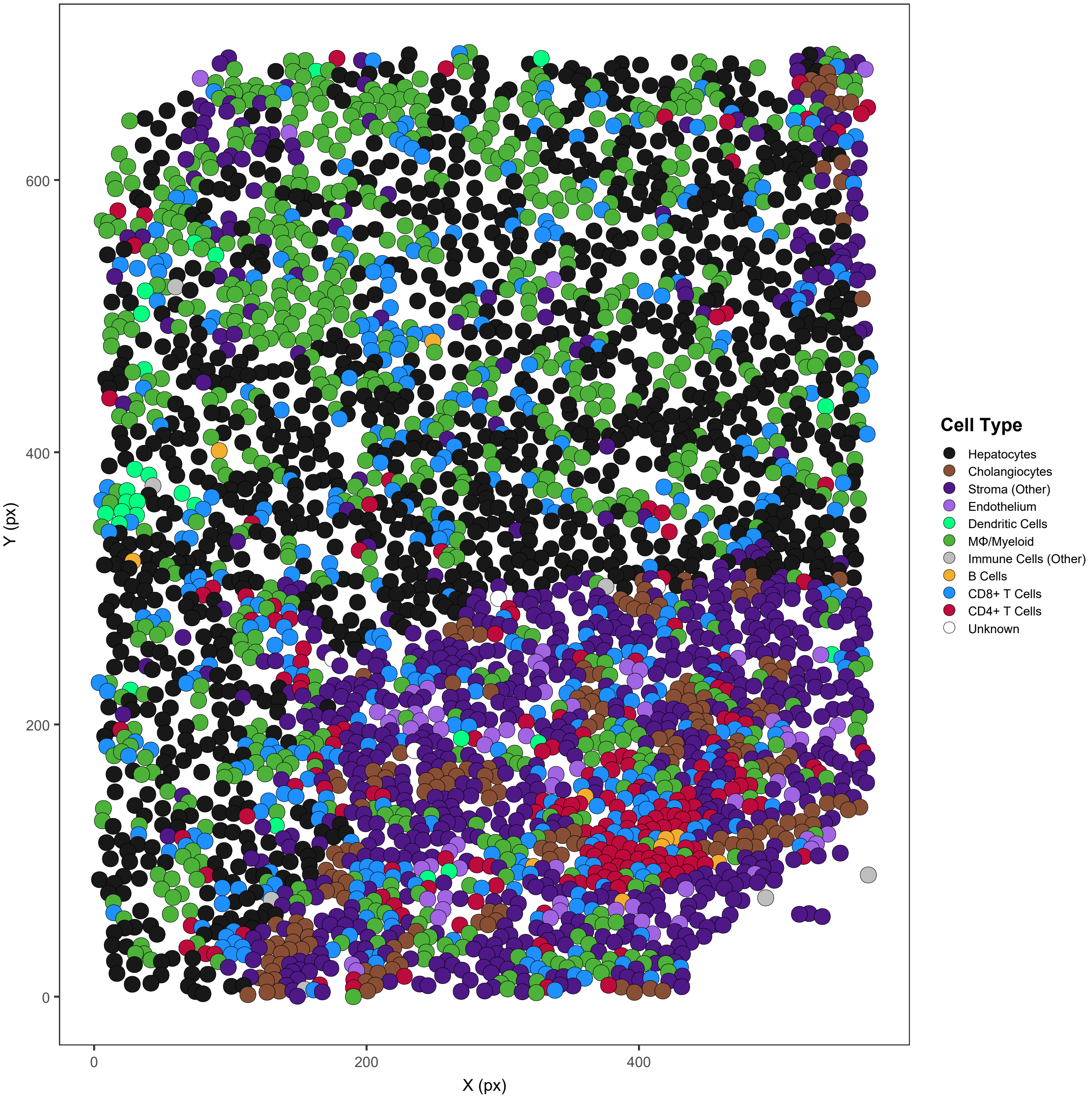

### AIH09_s0_a4_ac

AIH09 | 3654/3686 cells assigned (99.1%)

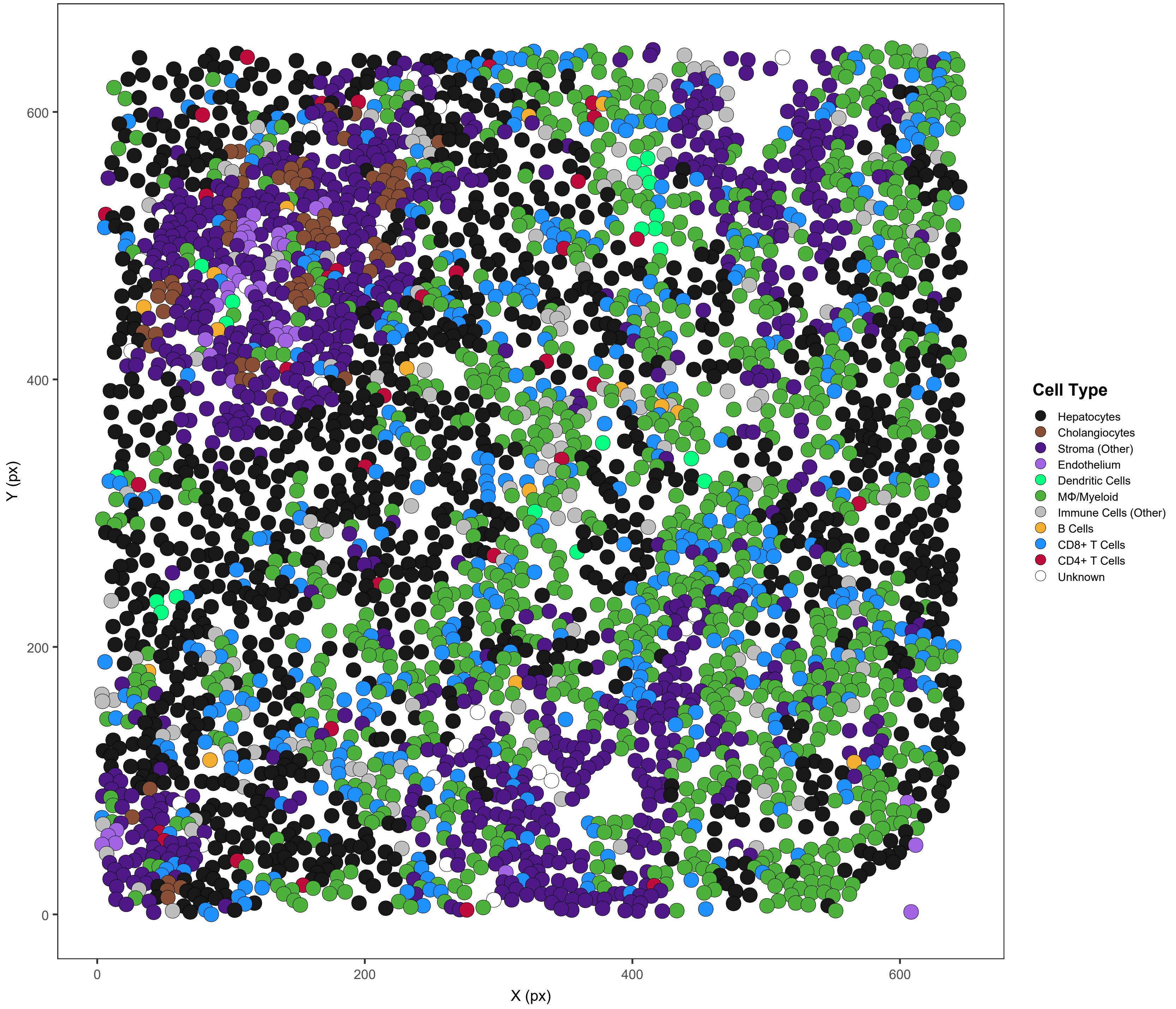

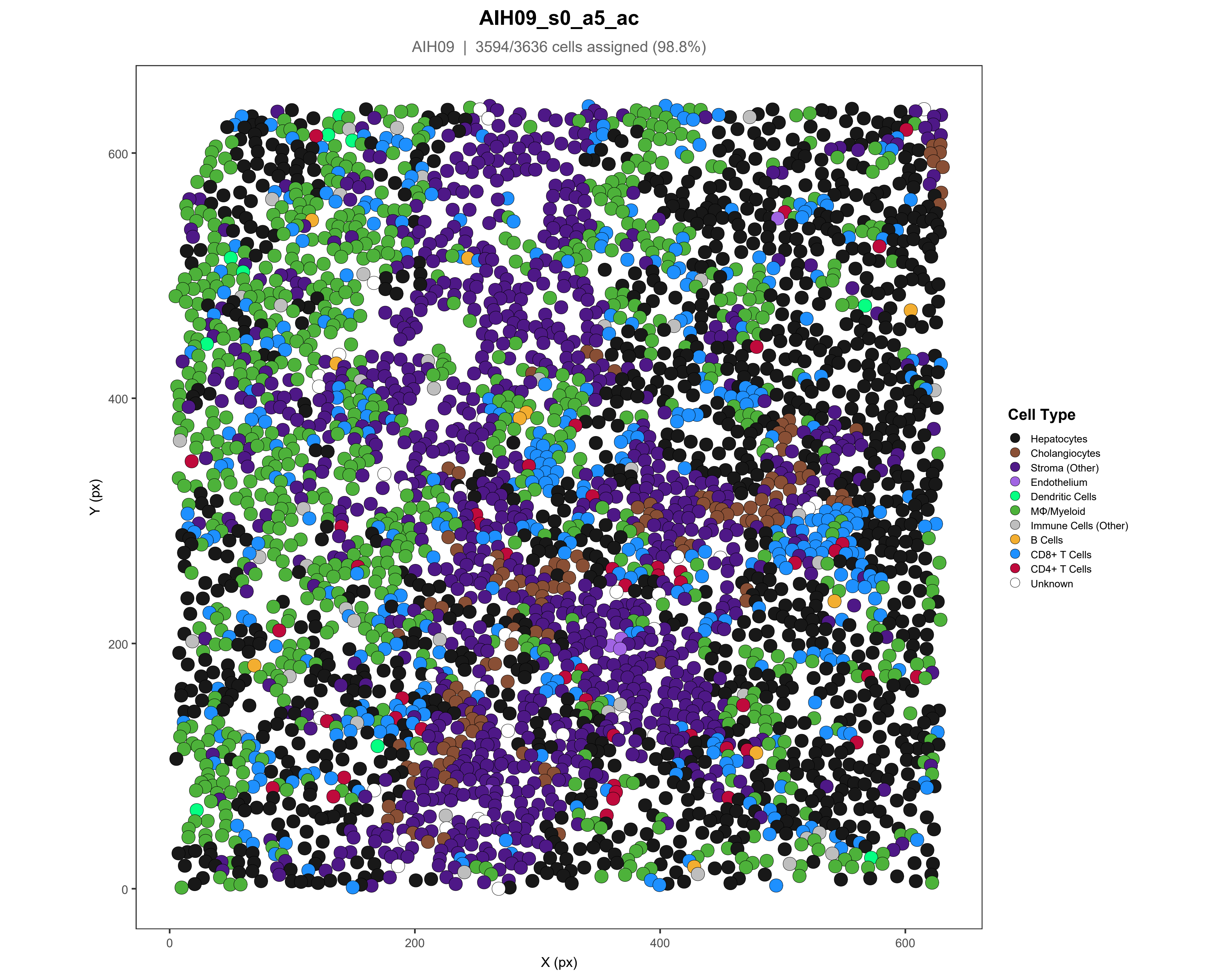
AIH09_s0_a5_ac
AIH09 | 3594/3636 cells assigned (98.8%)
Y (px)
X (px)
0
200
400
600
Cell Type
Hepatocytes
Cholangiocytes
Stroma (Other)
Endothelium
Dendritic Cells
MΦ/Myeloid
Immune Cells (Other)
B Cells
CD8+ T Cells
CD4+ T Cells
Unknown

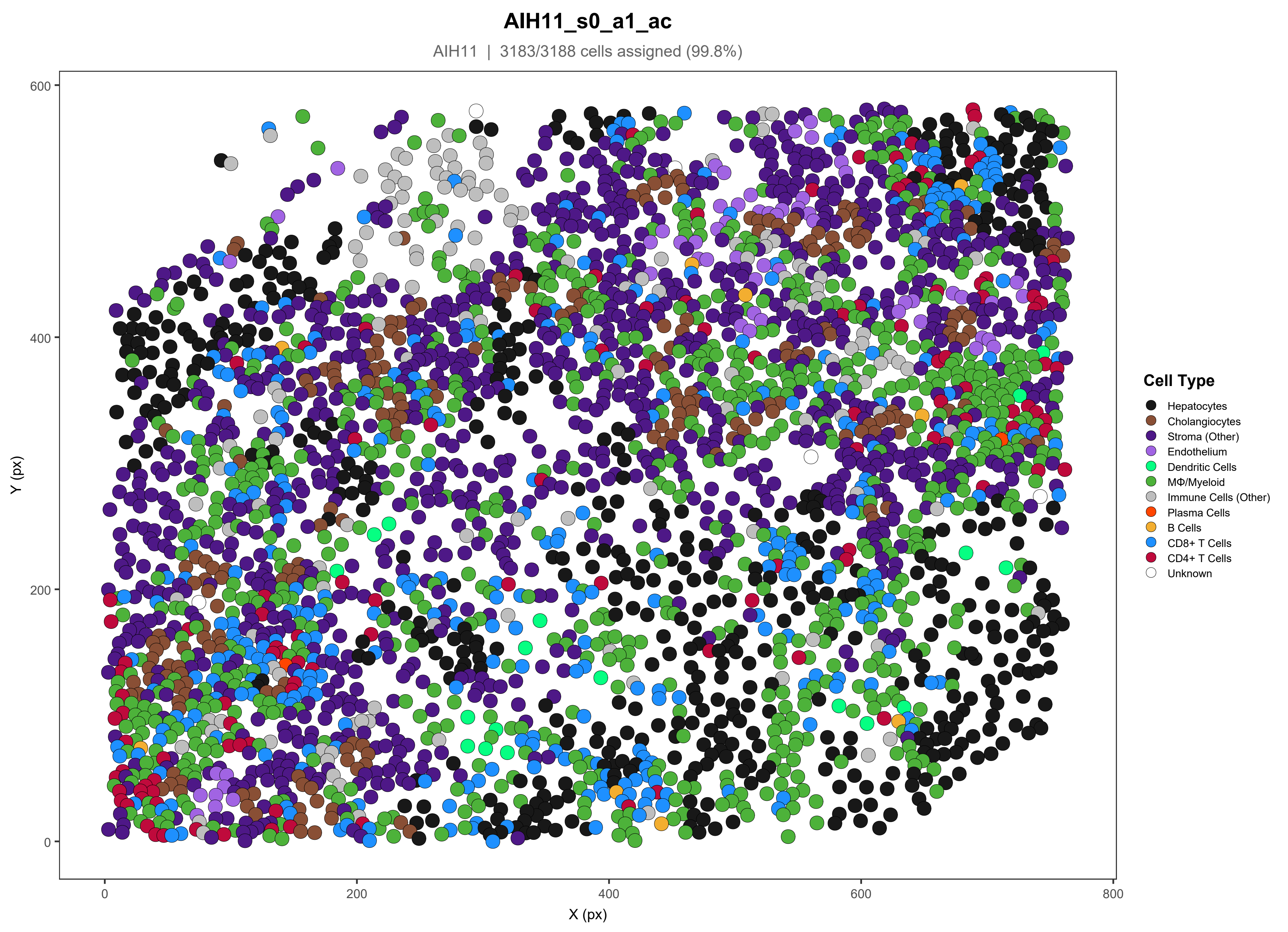
AIH11_s0_a1_ac
AIH11 | 3183/3188 cells assigned (99.8%)
Y (px)
X (px)
0
200
400
600
800
Cell Type
Hepatocytes
Cholangiocytes
Stroma (Other)
Endothelium
Dendritic Cells
MΦ/Myeloid
Immune Cells (Other)
Plasma Cells
B Cells
CD8+ T Cells
CD4+ T Cells
Unknown

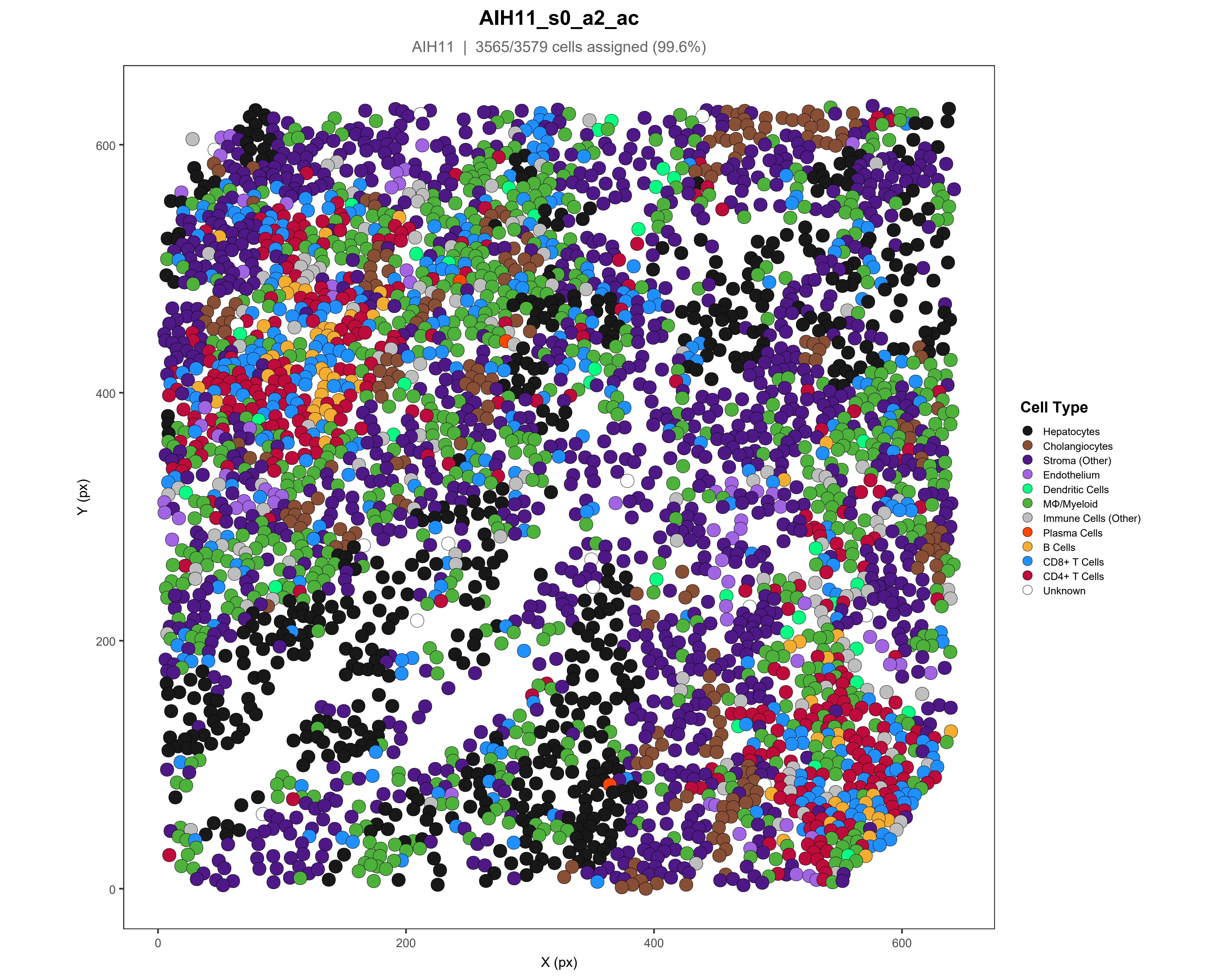
AIH11_s0_a2_ac
AIH11 | 3565/3579 cells assigned (99.6%)
Y (px)
X (px)
0
200
400
600
Cell Type
Hepatocytes
Cholangiocytes
Stroma (Other)
Endothelium
Dendritic Cells
MΦ/Myeloid
Immune Cells (Other)
Plasma Cells
B Cells
CD8+ T Cells
CD4+ T Cells
Unknown

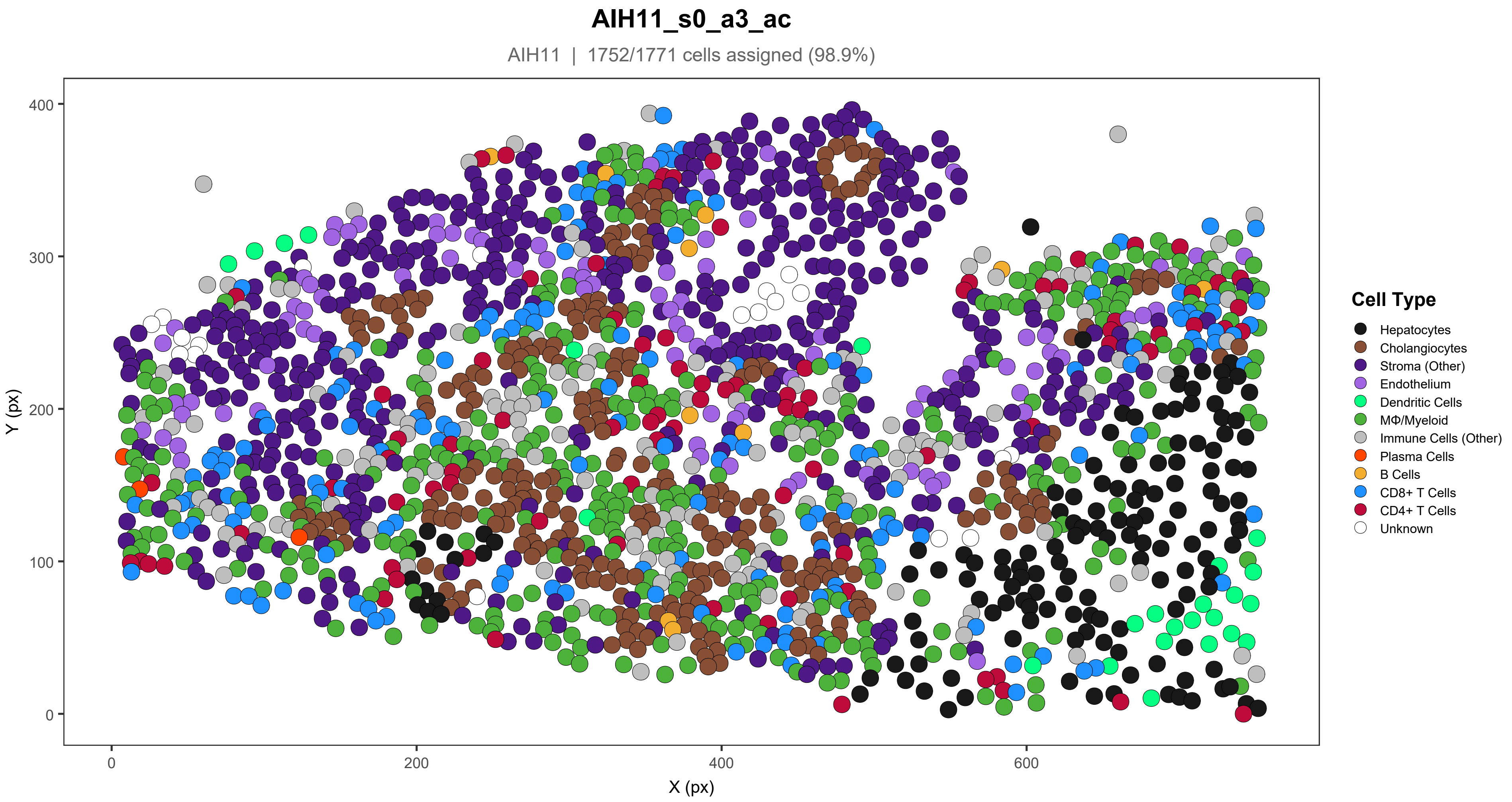

AIH11_s0_a3_ac
AIH11 | 1752/1771 cells assigned (98.9%)
Y (px)
X (px)
0
100
200
300
400
0
200
400
600
Cell Type
Hepatocytes
Cholangiocytes
Stroma (Other)
Endothelium
Dendritic Cells
MΦ/Myeloid
Immune Cells (Other)
Plasma Cells
B Cells
CD8+ T Cells
CD4+ T Cells
Unknown

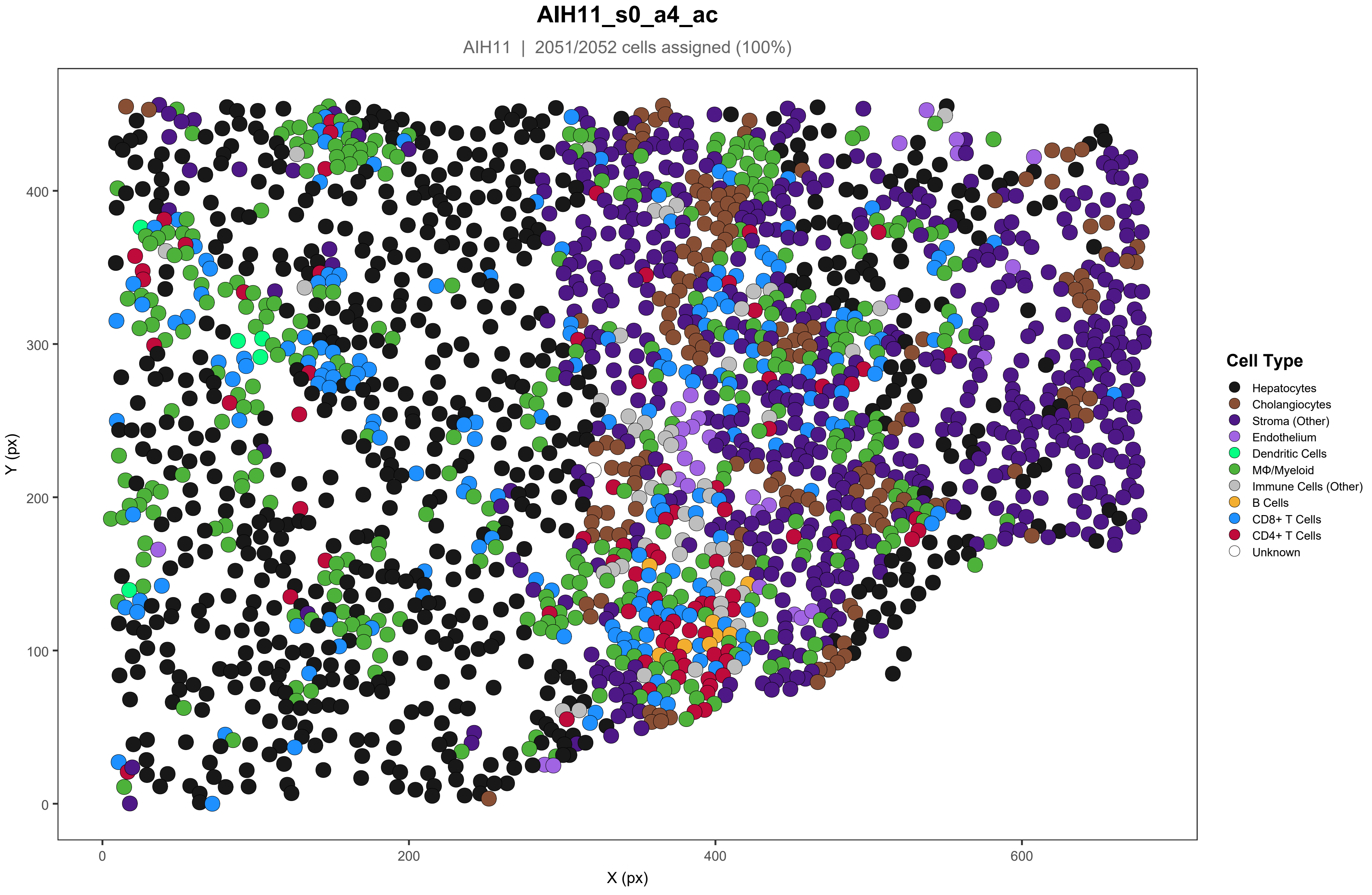
AIH11_s0_a4_ac
AIH11 | 2051/2052 cells assigned (100%)
Y (px)
X (px)
0
100
200
300
400
0
200
400
600
Cell Type
Hepatocytes
Cholangiocytes
Stroma (Other)
Endothelium
Dendritic Cells
MΦ/Myeloid
Immune Cells (Other)
B Cells
CD8+ T Cells
CD4+ T Cells
Unknown

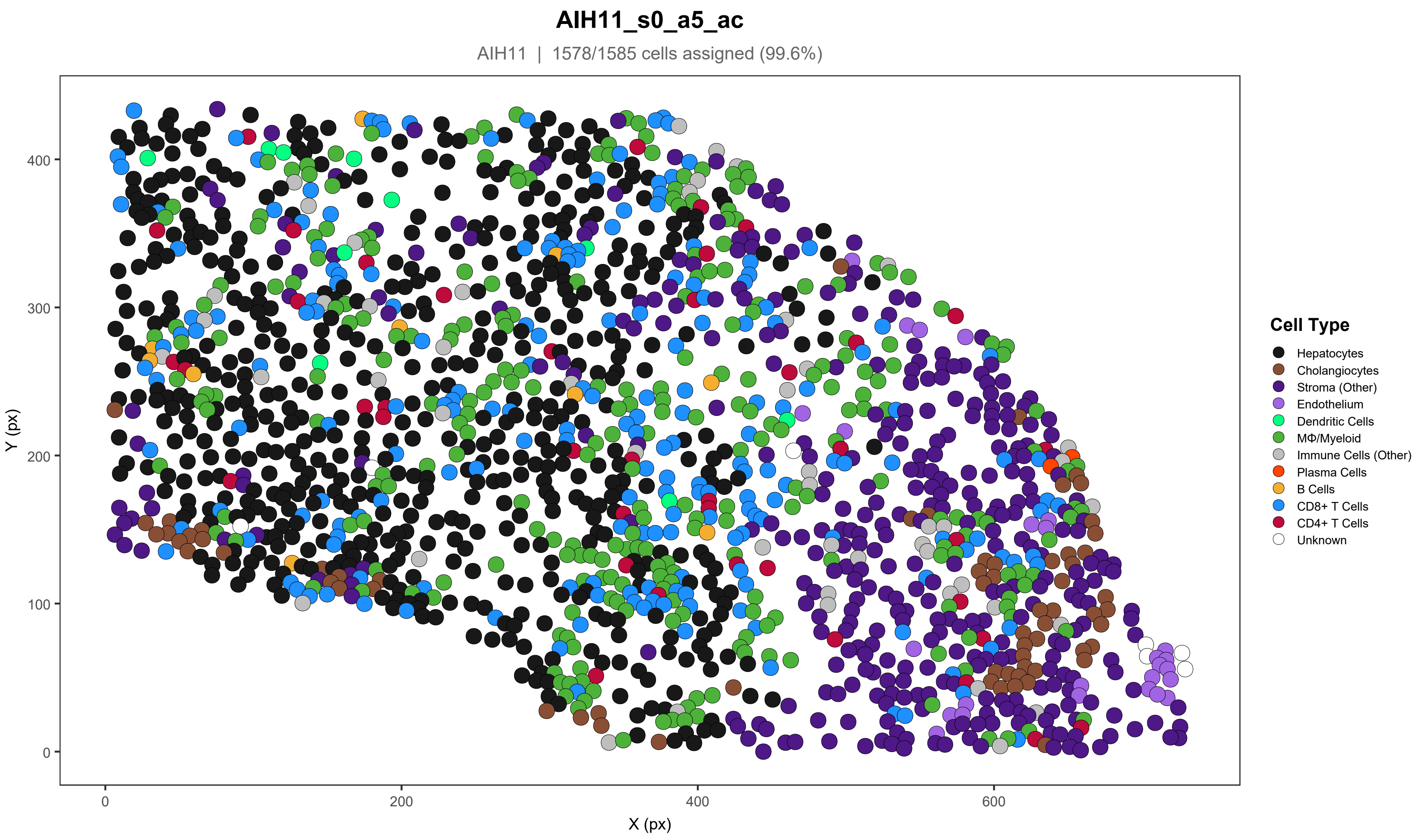

AIH11_s0_a5_ac
AIH11 | 1578/1585 cells assigned (99.6%)
Y (px)
X (px)
0
100
200
300
400
600
Cell Type
Hepatocytes
Cholangiocytes
Stroma (Other)
Endothelium
Dendritic Cells
MΦ/Myeloid
Immune Cells (Other)
Plasma Cells
B Cells
CD8+ T Cells
CD4+ T Cells
Unknown

### AIH12_s0_a1_ac

AIH12 | 2319/2332 cells assigned (99.4%)

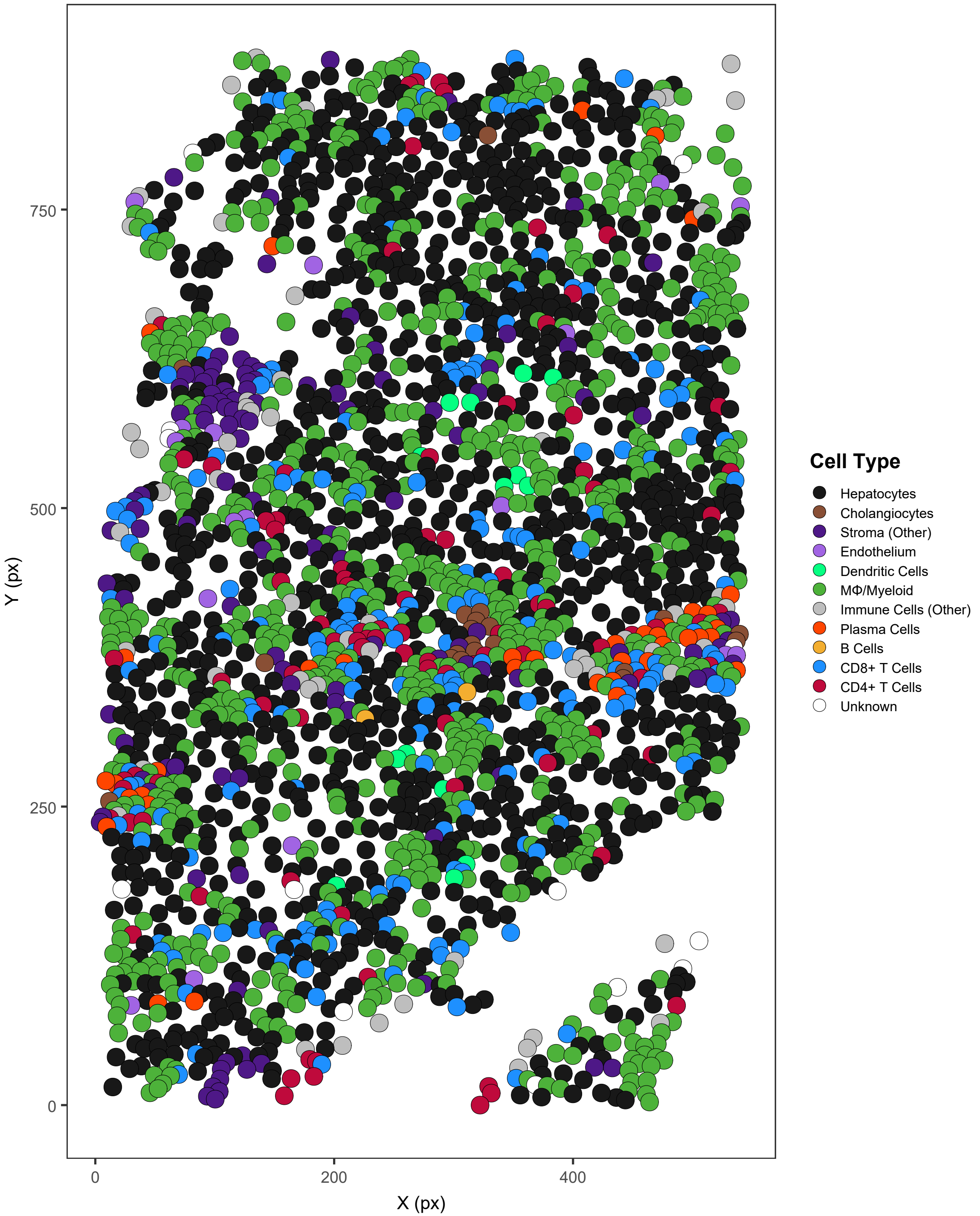

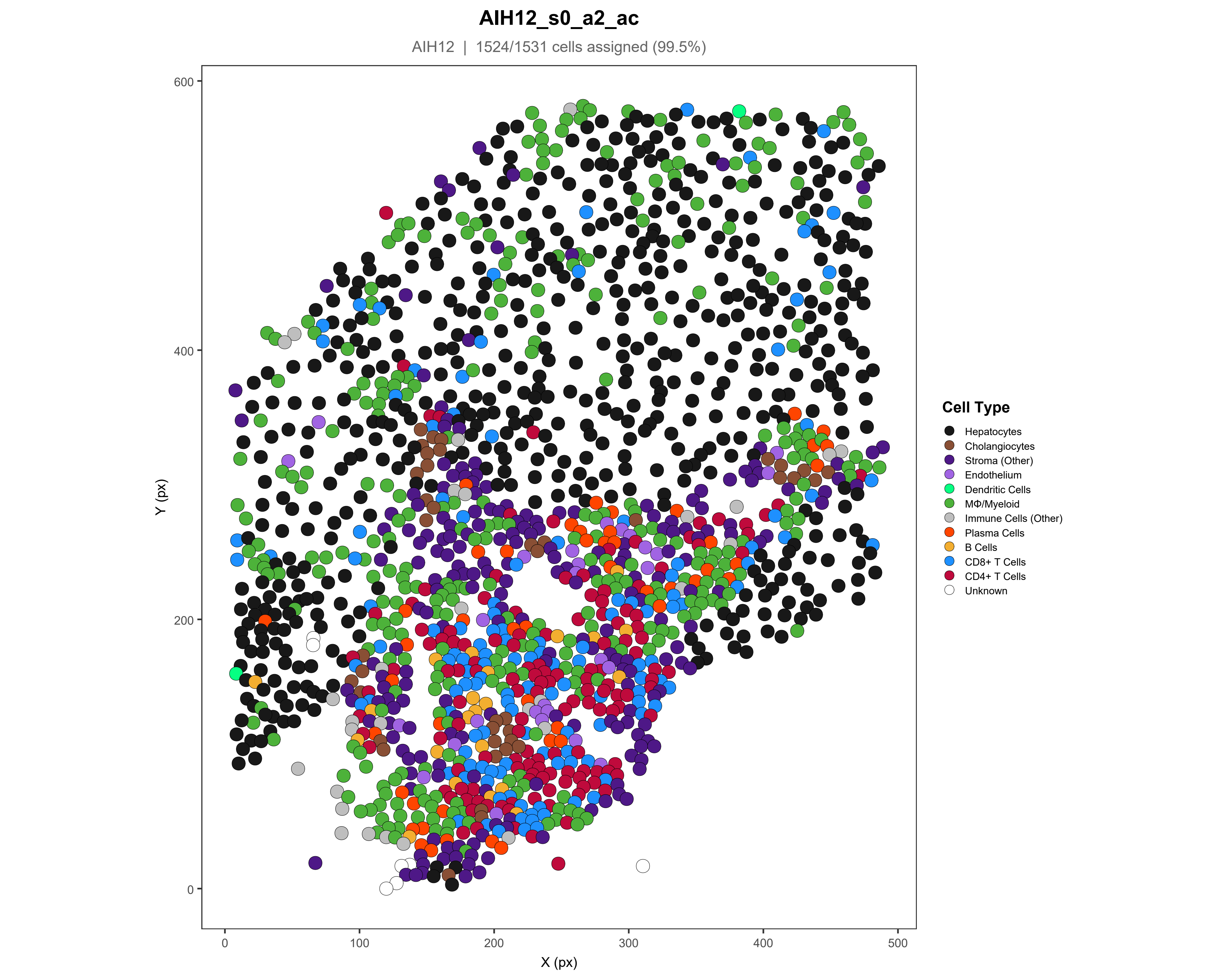

AIH12_s0_a2_ac
AIH12 | 1524/1531 cells assigned (99.5%)
Y (px)
X (px)
0
100
200
300
400
500
600
Cell Type
Hepatocytes
Cholangiocytes
Stroma (Other)
Endothelium
Dendritic Cells
MΦ/Myeloid
Immune Cells (Other)
Plasma Cells
B Cells
CD8+ T Cells
CD4+ T Cells
Unknown

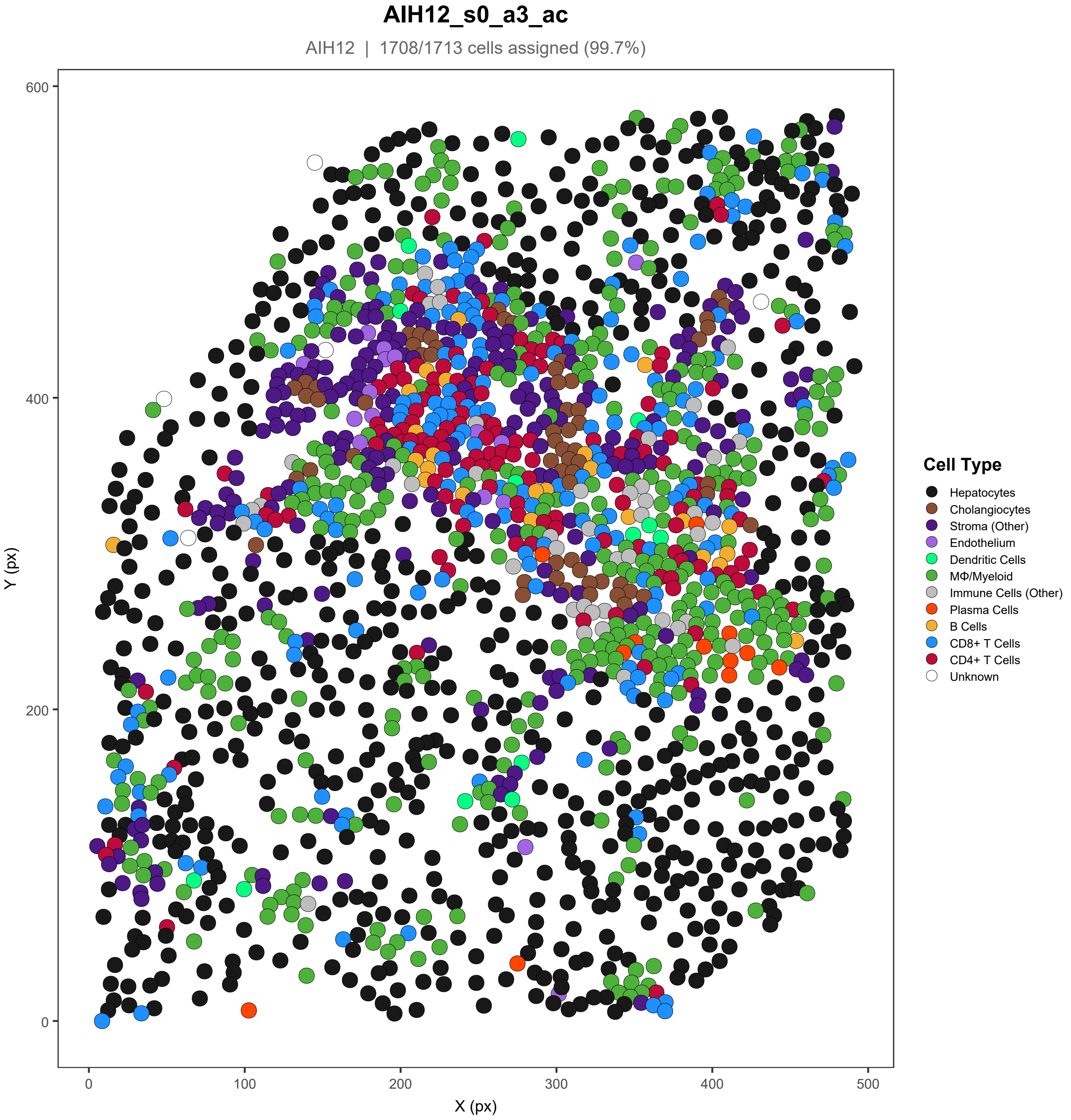
AIH12_s0_a3_ac
AIH12 | 1708/1713 cells assigned (99.7%)
Y (px)
X (px)
0
100
200
300
400
500
600
Cell Type
Hepatocytes
Cholangiocytes
Stroma (Other)
Endothelium
Dendritic Cells
MΦ/Myeloid
Immune Cells (Other)
Plasma Cells
B Cells
CD8+ T Cells
CD4+ T Cells
Unknown

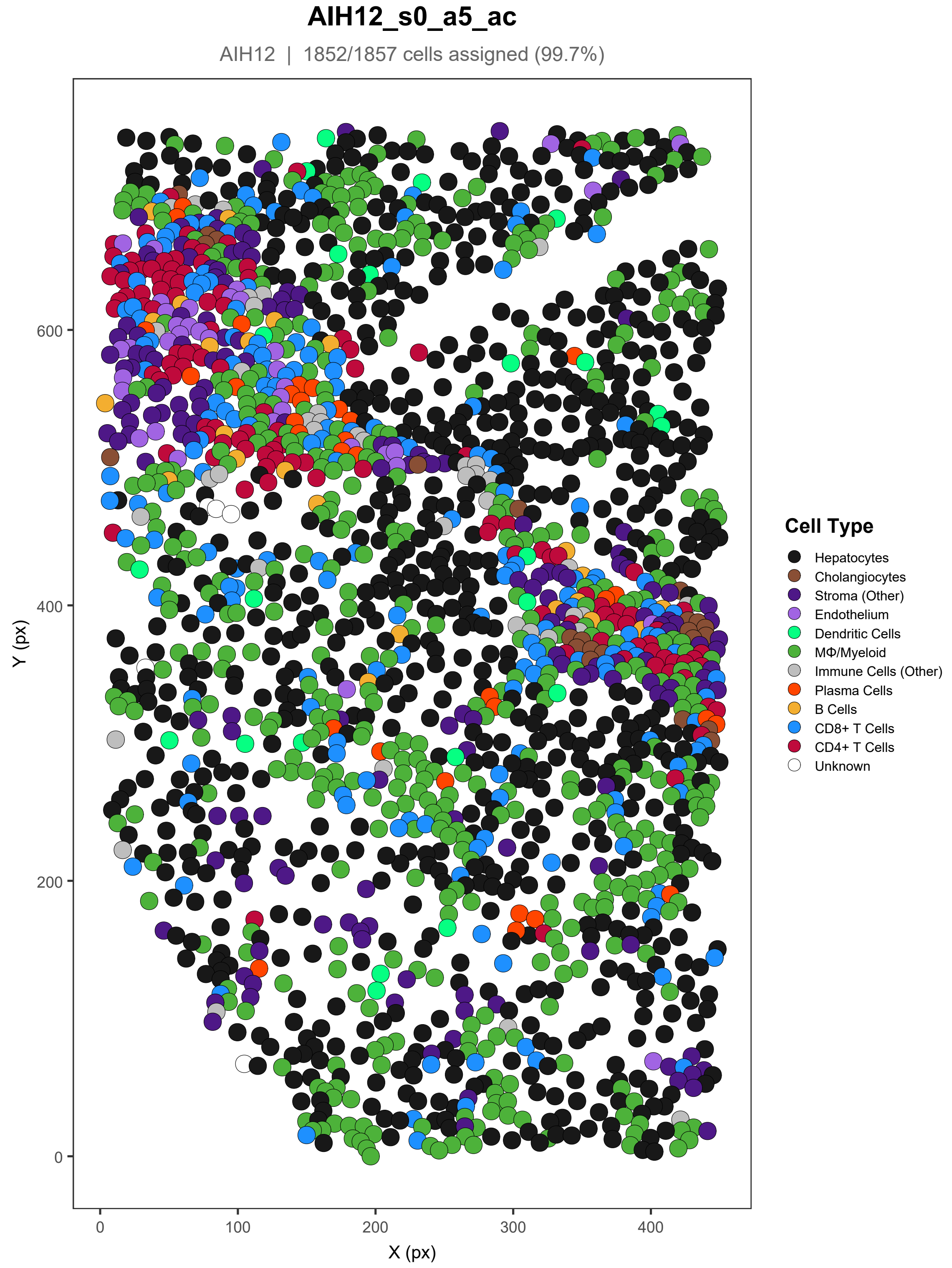

AIH12_s0_a5_ac
AIH12 | 1852/1857 cells assigned (99.7%)
Y (px)
X (px)
0
100
200
300
400
600
Cell Type
Hepatocytes
Cholangiocytes
Stroma (Other)
Endothelium
Dendritic Cells
MΦ/Myeloid
Immune Cells (Other)
Plasma Cells
B Cells
CD8+ T Cells
CD4+ T Cells
Unknown

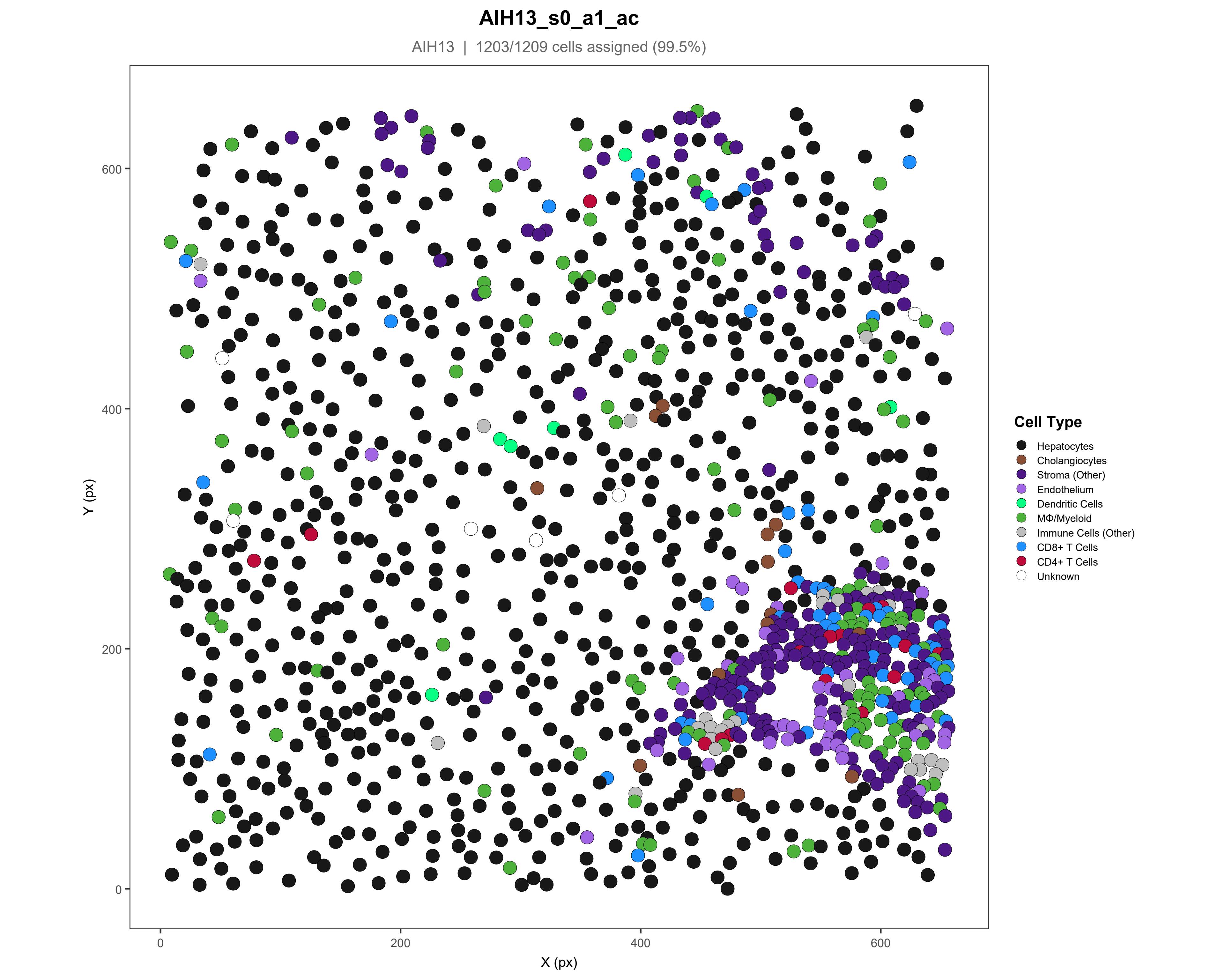

AIH13_s0_a1_ac
AIH13 | 1203/1209 cells assigned (99.5%)
Y (px)
X (px)
0
200
400
600
Cell Type
Hepatocytes
Cholangiocytes
Stroma (Other)
Endothelium
Dendritic Cells
MΦ/Myeloid
Immune Cells (Other)
CD8+ T Cells
CD4+ T Cells
Unknown

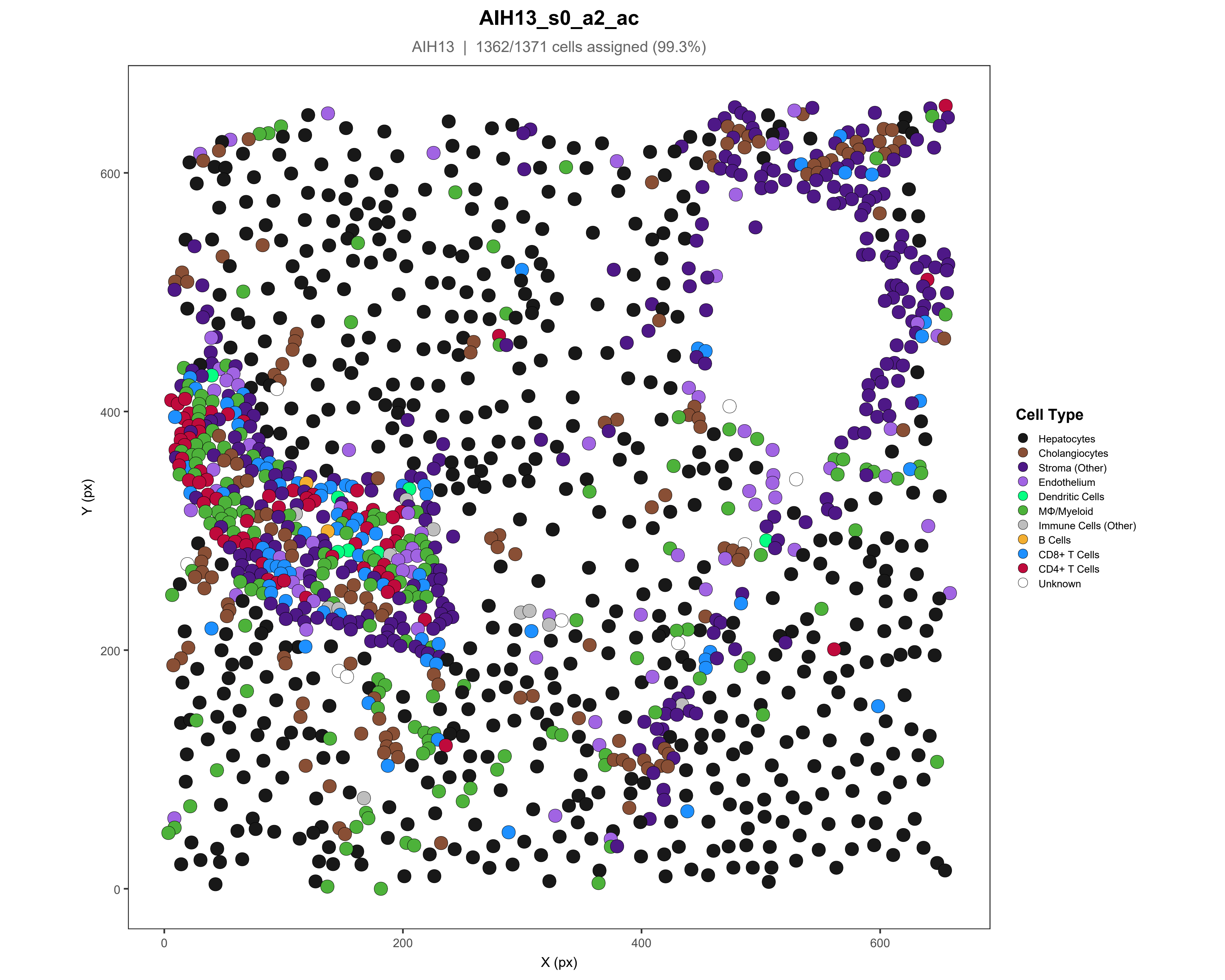
AIH13_s0_a2_ac
AIH13 | 1362/1371 cells assigned (99.3%)
Y (px)
X (px)
0
200
400
600
Cell Type
Hepatocytes
Cholangiocytes
Stroma (Other)
Endothelium
Dendritic Cells
MΦ/Myeloid
Immune Cells (Other)
B Cells
CD8+ T Cells
CD4+ T Cells
Unknown

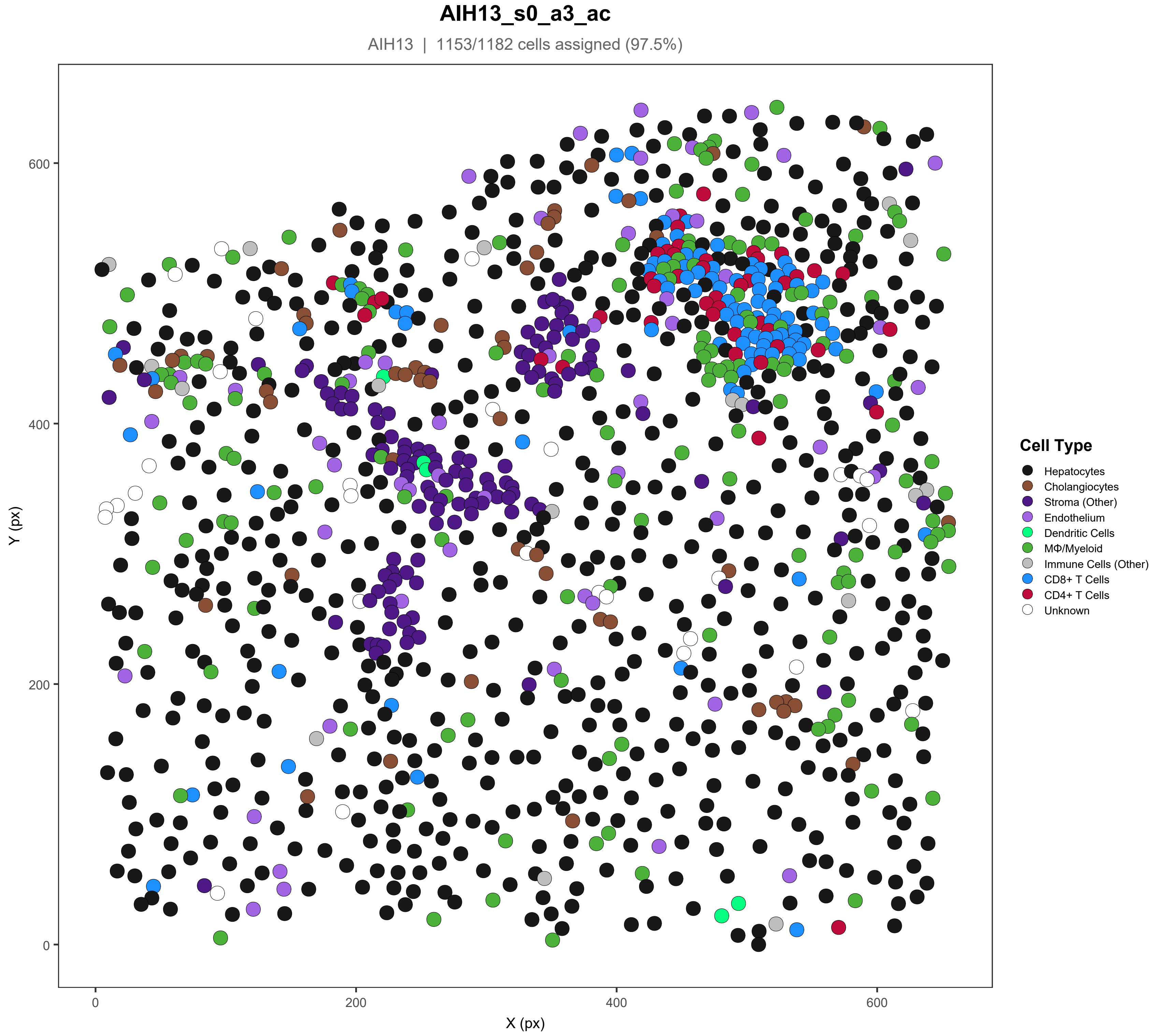
AIH13_s0_a3_ac
AIH13 | 1153/1182 cells assigned (97.5%)
Y (px)
X (px)
0
200
400
600
Cell Type
Hepatocytes
Cholangiocytes
Stroma (Other)
Endothelium
Dendritic Cells
MΦ/Myeloid
Immune Cells (Other)
CD8+ T Cells
CD4+ T Cells
Unknown

### AIH13_s0_a4_ac

AIH13 | 1636/1641 cells assigned (99.7%)

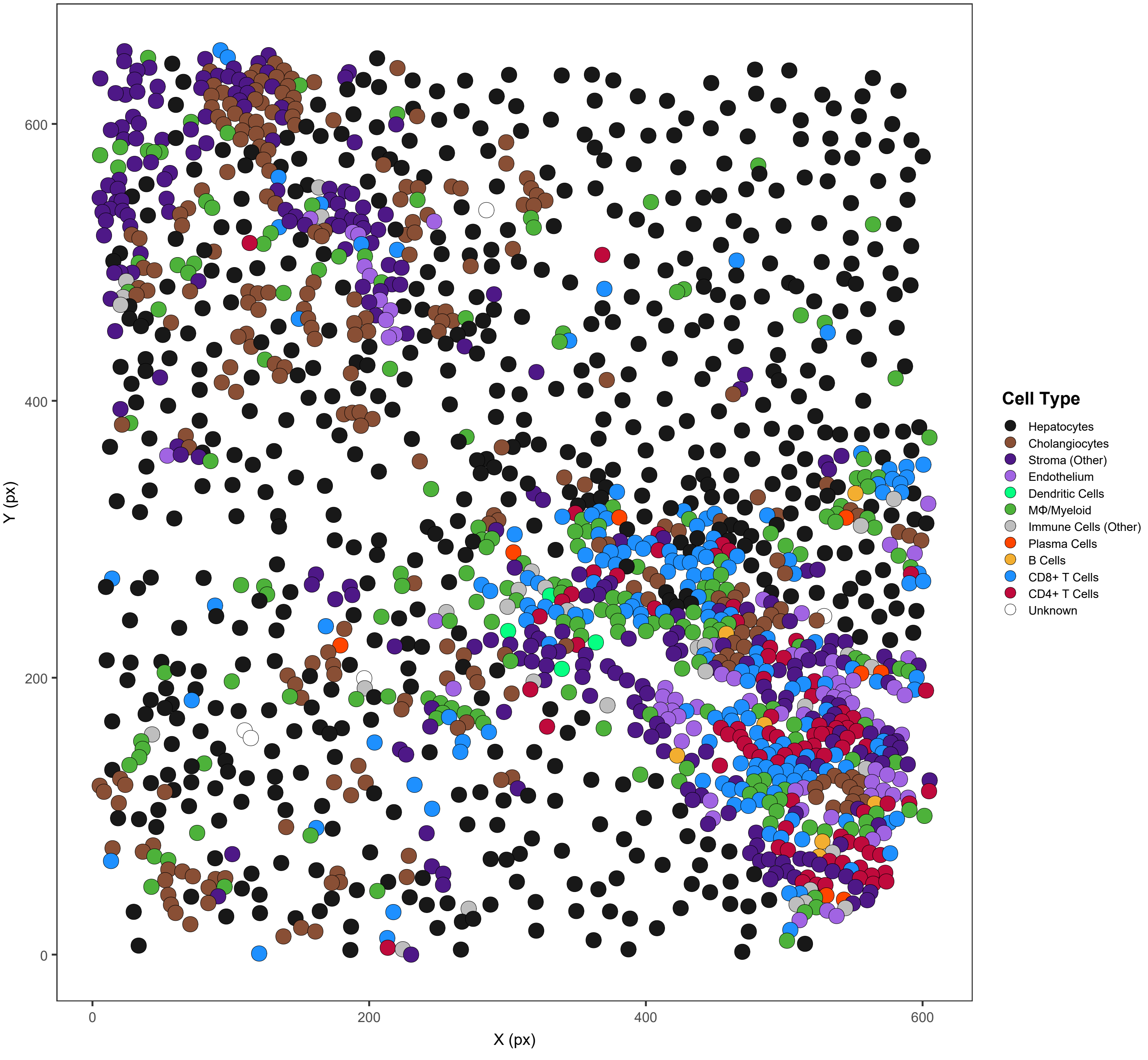

#### AIH13_s0_a5_ac

AIH13 | 1138/1141 cells assigned (99.7%)

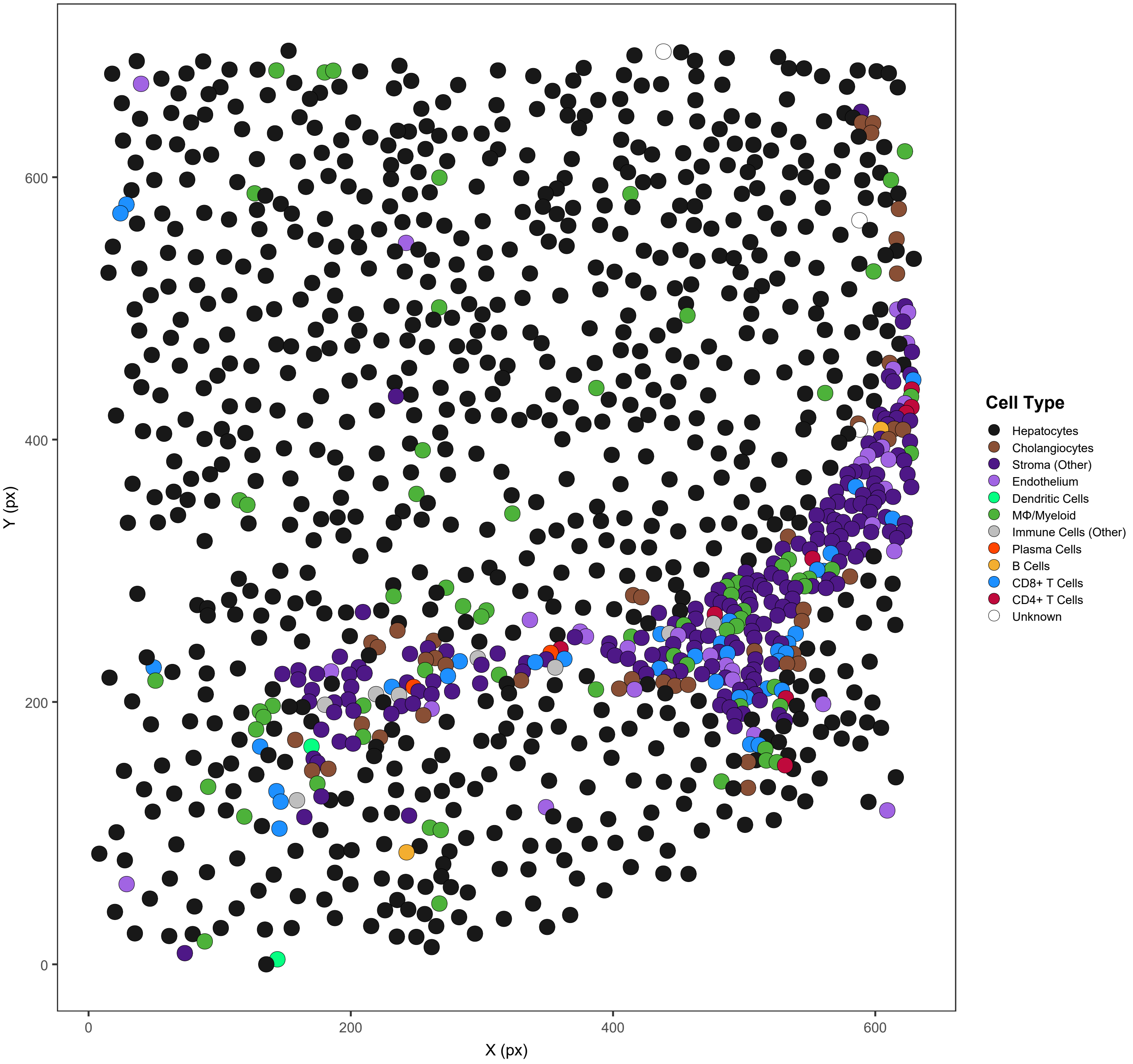

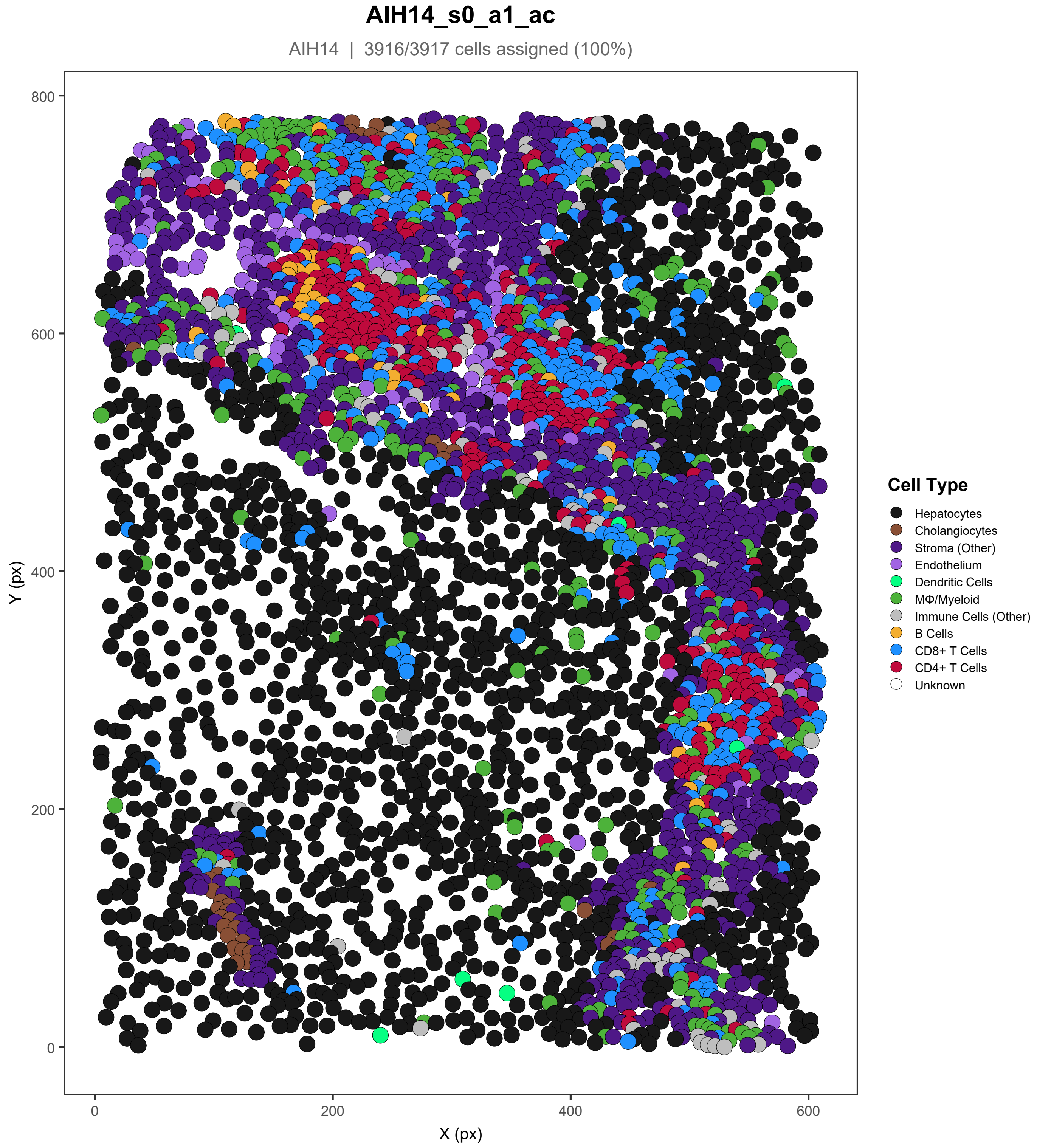
AIH14_s0_a1_ac
AIH14 | 3916/3917 cells assigned (100%)
Y (px)
X (px)
800
600
400
200
0
0
200
400
600
Cell Type
Hepatocytes
Cholangiocytes
Stroma (Other)
Endothelium
Dendritic Cells
MΦ/Myeloid
Immune Cells (Other)
B Cells
CD8+ T Cells
CD4+ T Cells
Unknown

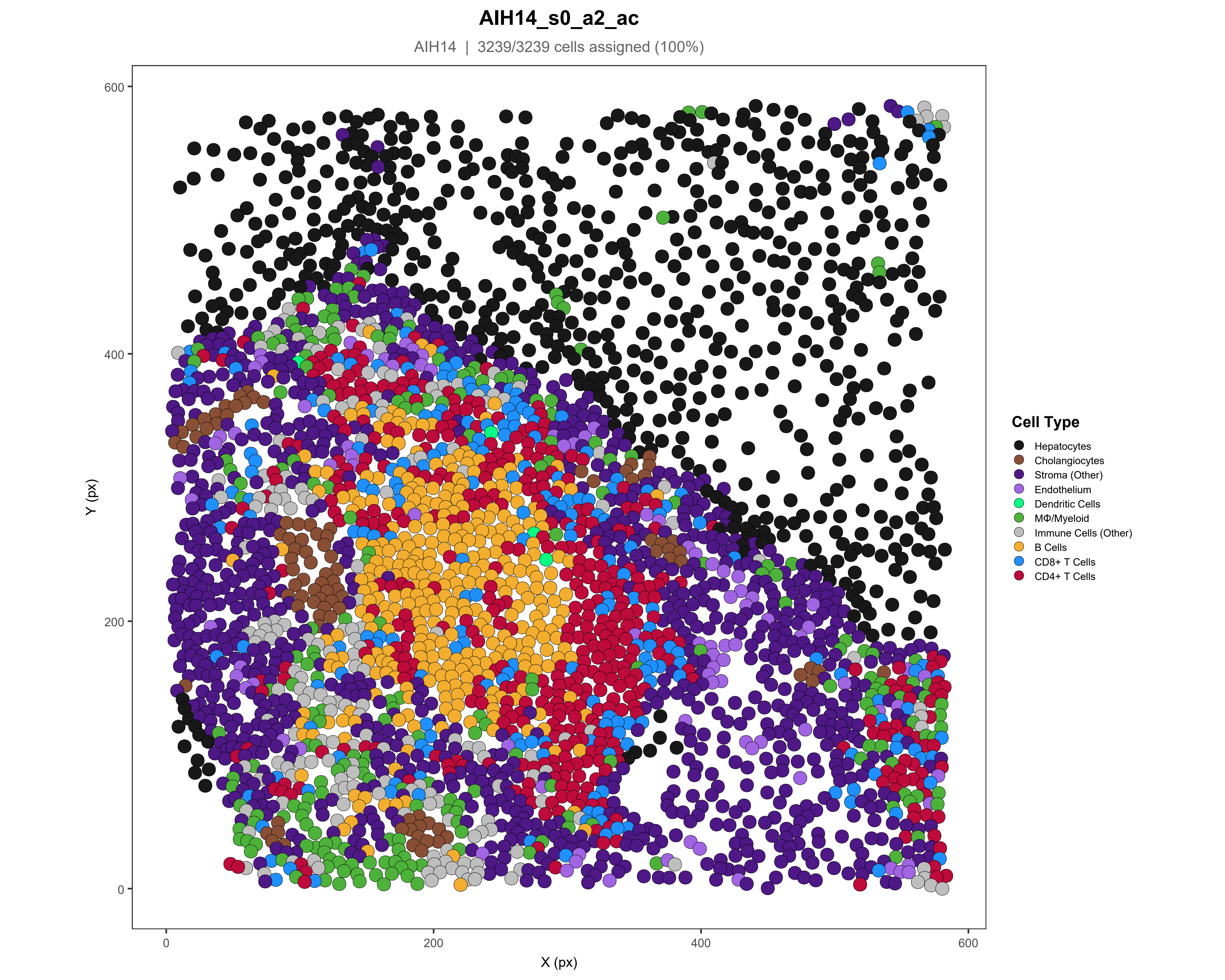
AIH14_s0_a2_ac
AIH14 | 3239/3239 cells assigned (100%)
Y (px)
X (px)
0
200
400
600
Cell Type
Hepatocytes
Cholangiocytes
Stroma (Other)
Endothelium
Dendritic Cells
MΦ/Myeloid
Immune Cells (Other)
B Cells
CD8+ T Cells
CD4+ T Cells

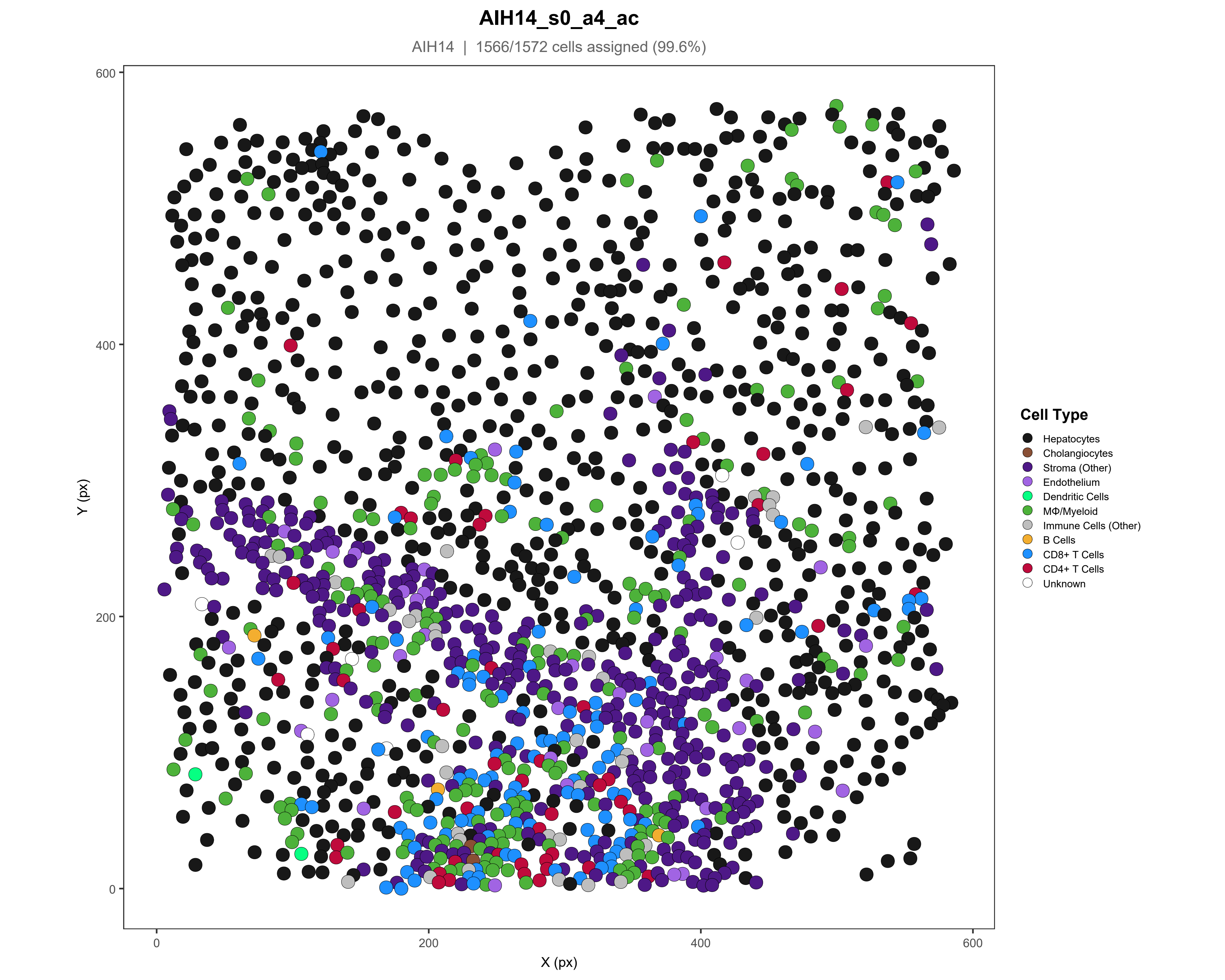
AIH14_s0_a4_ac
AIH14 | 1566/1572 cells assigned (99.6%)
Y (px)
X (px)
0
200
400
600
Cell Type
Hepatocytes
Cholangiocytes
Stroma (Other)
Endothelium
Dendritic Cells
MΦ/Myeloid
Immune Cells (Other)
B Cells
CD8+ T Cells
CD4+ T Cells
Unknown

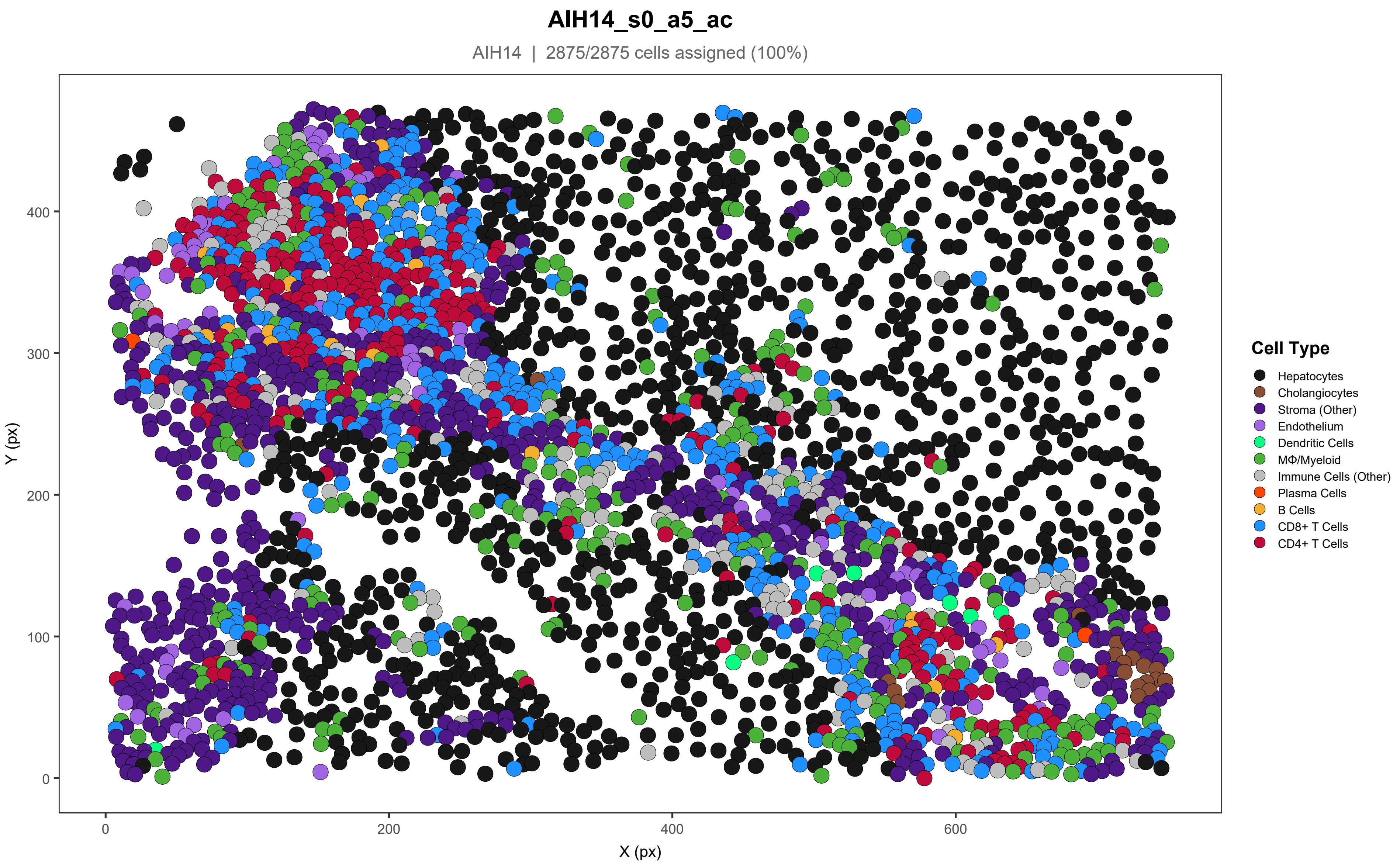

AIH14_s0_a5_ac
AIH14 | 2875/2875 cells assigned (100%)
Y (px)
X (px)
0
100
200
300
400
0
200
400
600
Cell Type
Hepatocytes
Cholangiocytes
Stroma (Other)
Endothelium
Dendritic Cells
MΦ/Myeloid
Immune Cells (Other)
Plasma Cells
B Cells
CD8+ T Cells
CD4+ T Cells

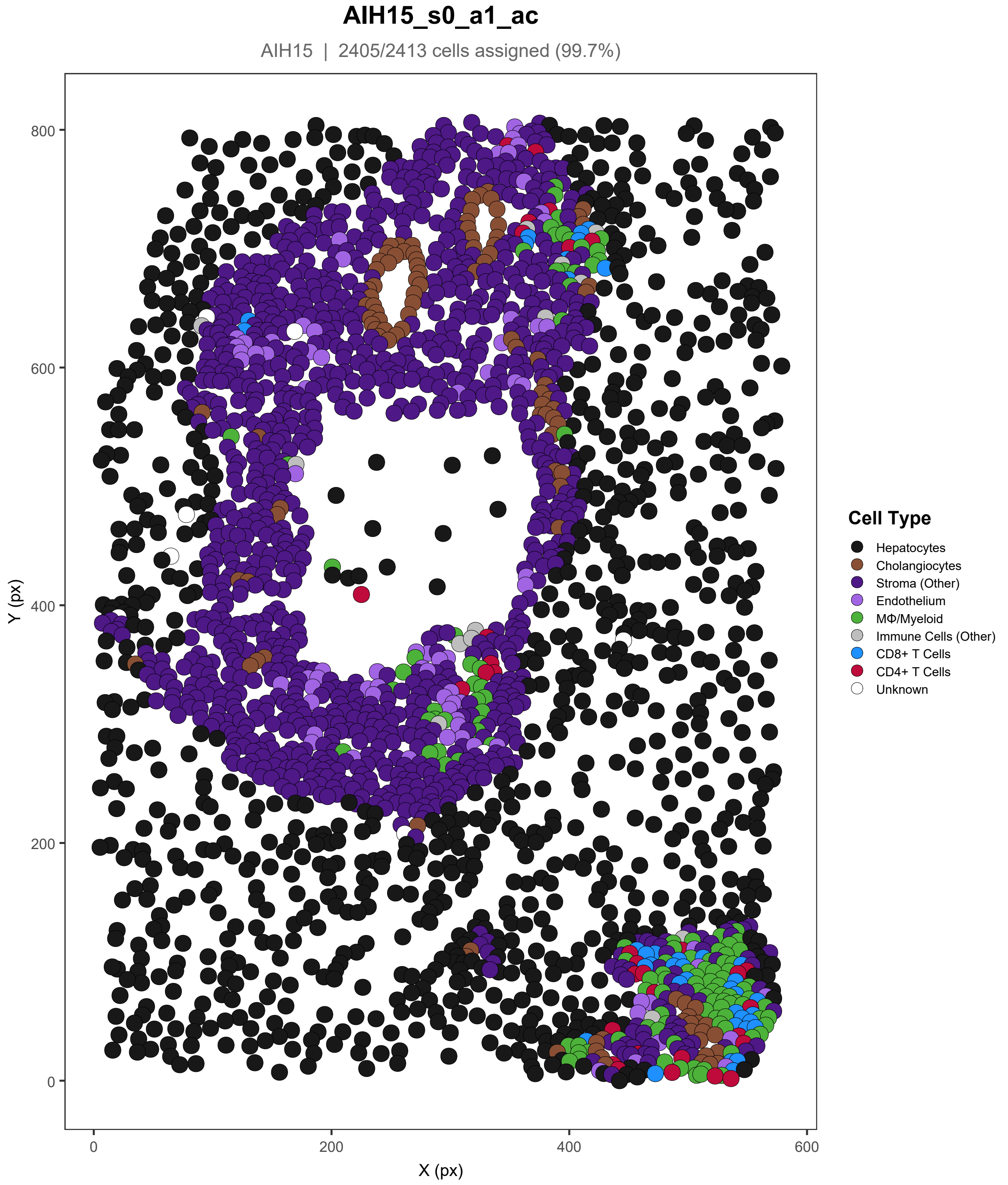

AIH15_s0_a1_ac
AIH15 | 2405/2413 cells assigned (99.7%)
Y (px)
X (px)
800
600
400
200
0
0
200
400
600
Cell Type
Hepatocytes
Cholangiocytes
Stroma (Other)
Endothelium
MΦ/Myeloid
Immune Cells (Other)
CD8+ T Cells
CD4+ T Cells
Unknown

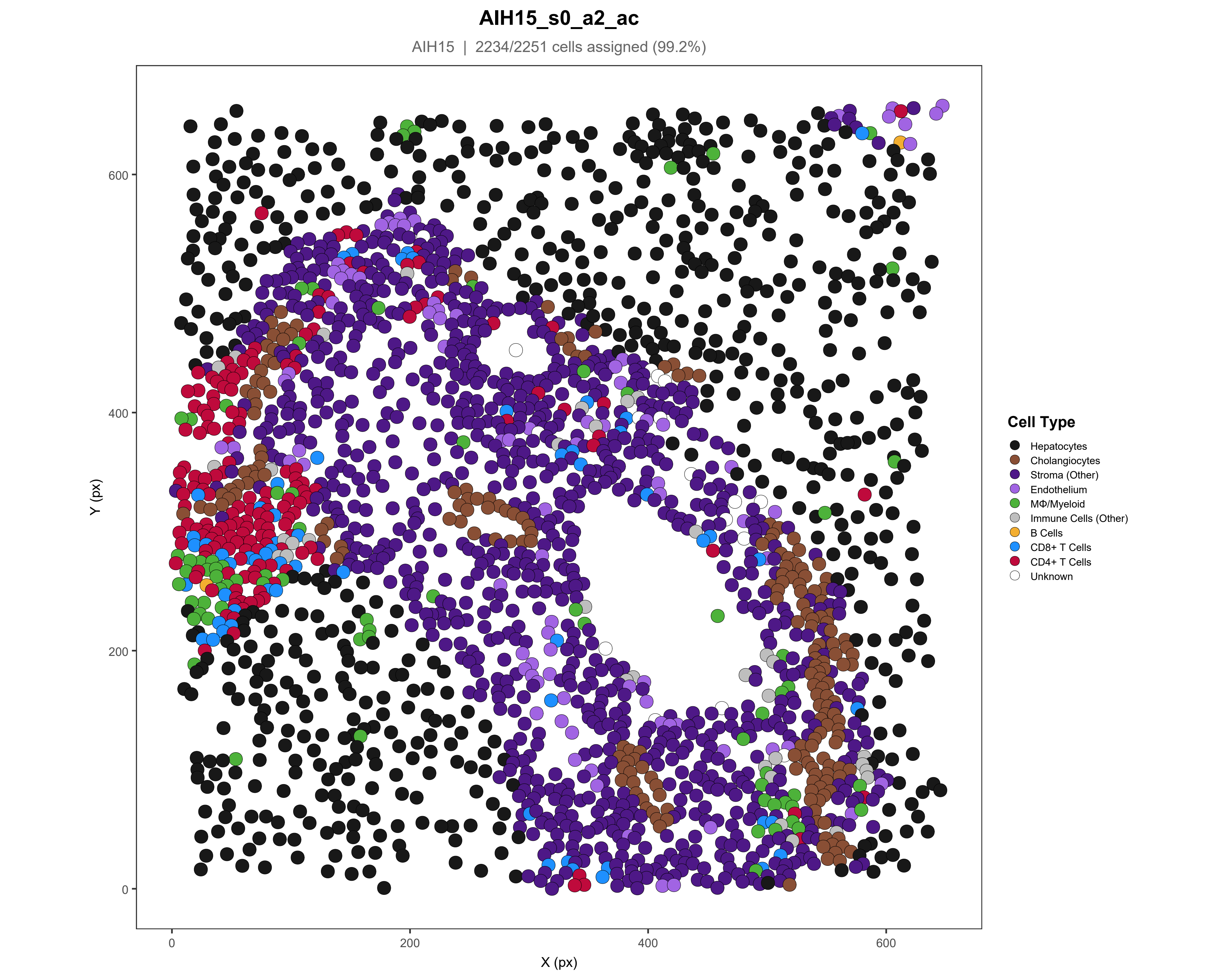
AIH15_s0_a2_ac
AIH15 | 2234/2251 cells assigned (99.2%)
Y (px)
X (px)
0
200
400
600
Cell Type
Hepatocytes
Cholangiocytes
Stroma (Other)
Endothelium
MΦ/Myeloid
Immune Cells (Other)
B Cells
CD8+ T Cells
CD4+ T Cells
Unknown

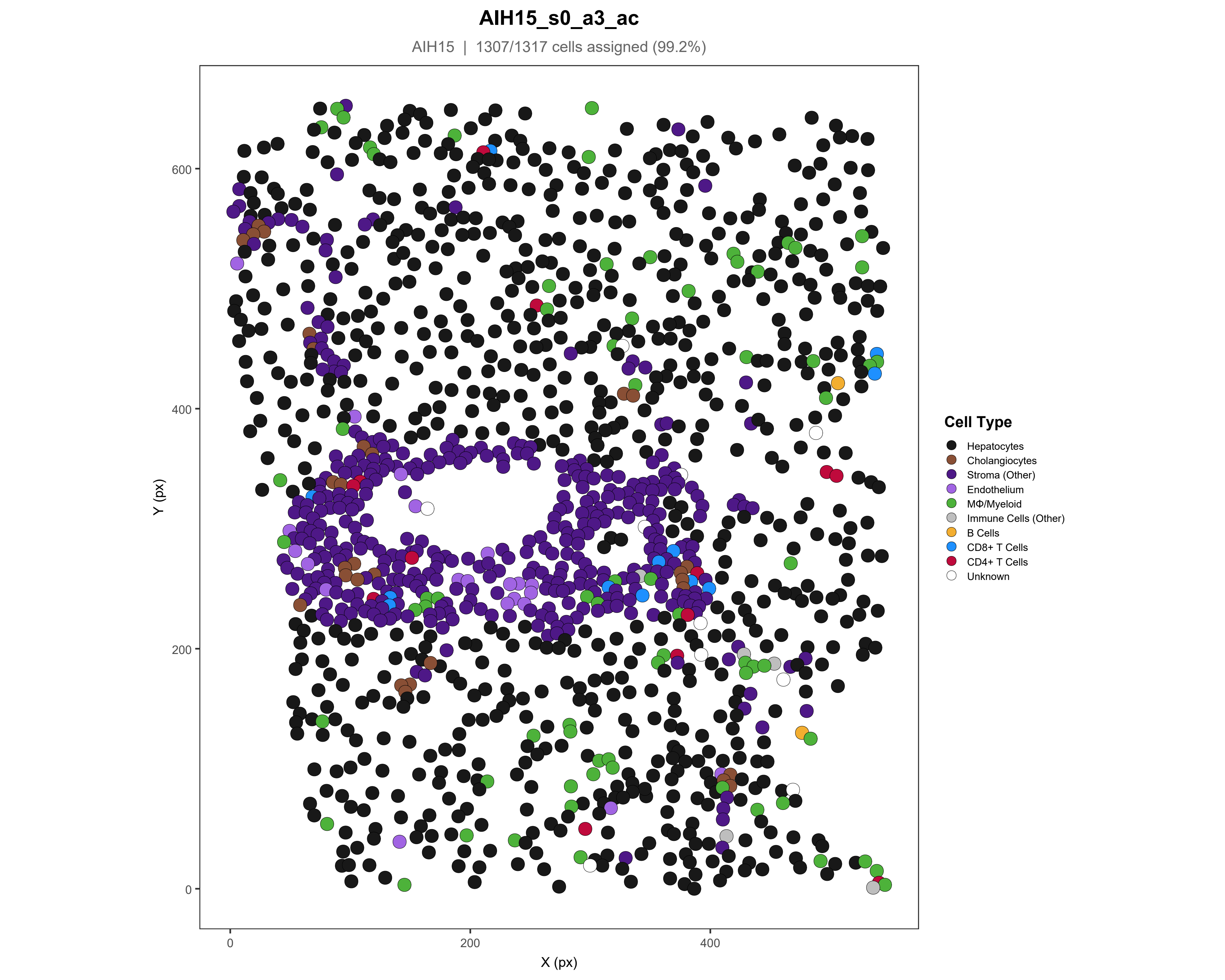

AIH15_s0_a3_ac
AIH15 | 1307/1317 cells assigned (99.2%)
Y (px)
X (px)
0
200
400
600
Cell Type
Hepatocytes
Cholangiocytes
Stroma (Other)
Endothelium
MΦ/Myeloid
Immune Cells (Other)
B Cells
CD8+ T Cells
CD4+ T Cells
Unknown

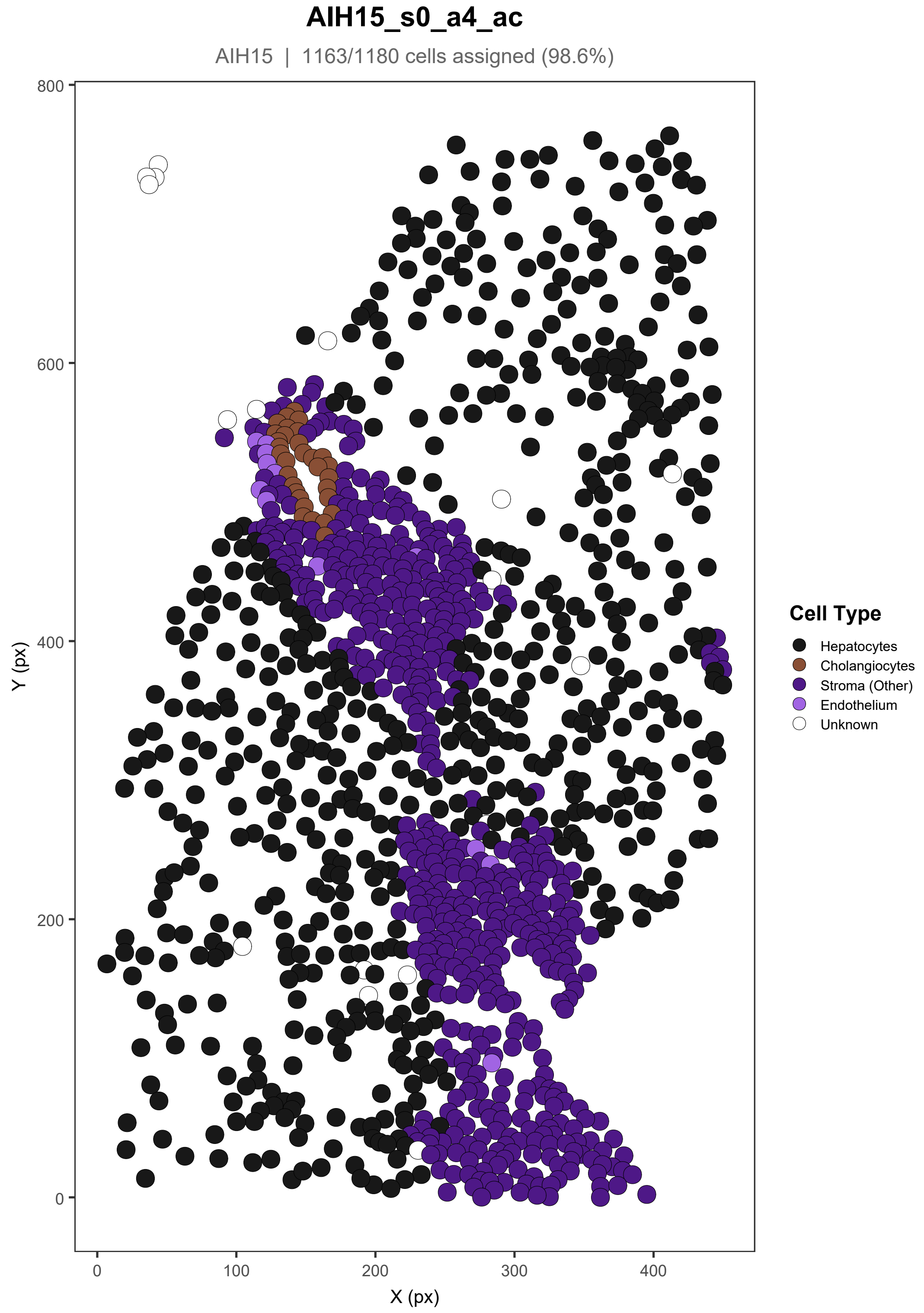
AIH15_s0_a4_ac
AIH15 | 1163/1180 cells assigned (98.6%)
Y (px)
X (px)
800
600
400
200
0
0
100
200
300
400
Cell Type
Hepatocytes
Cholangiocytes
Stroma (Other)
Endothelium
Unknown

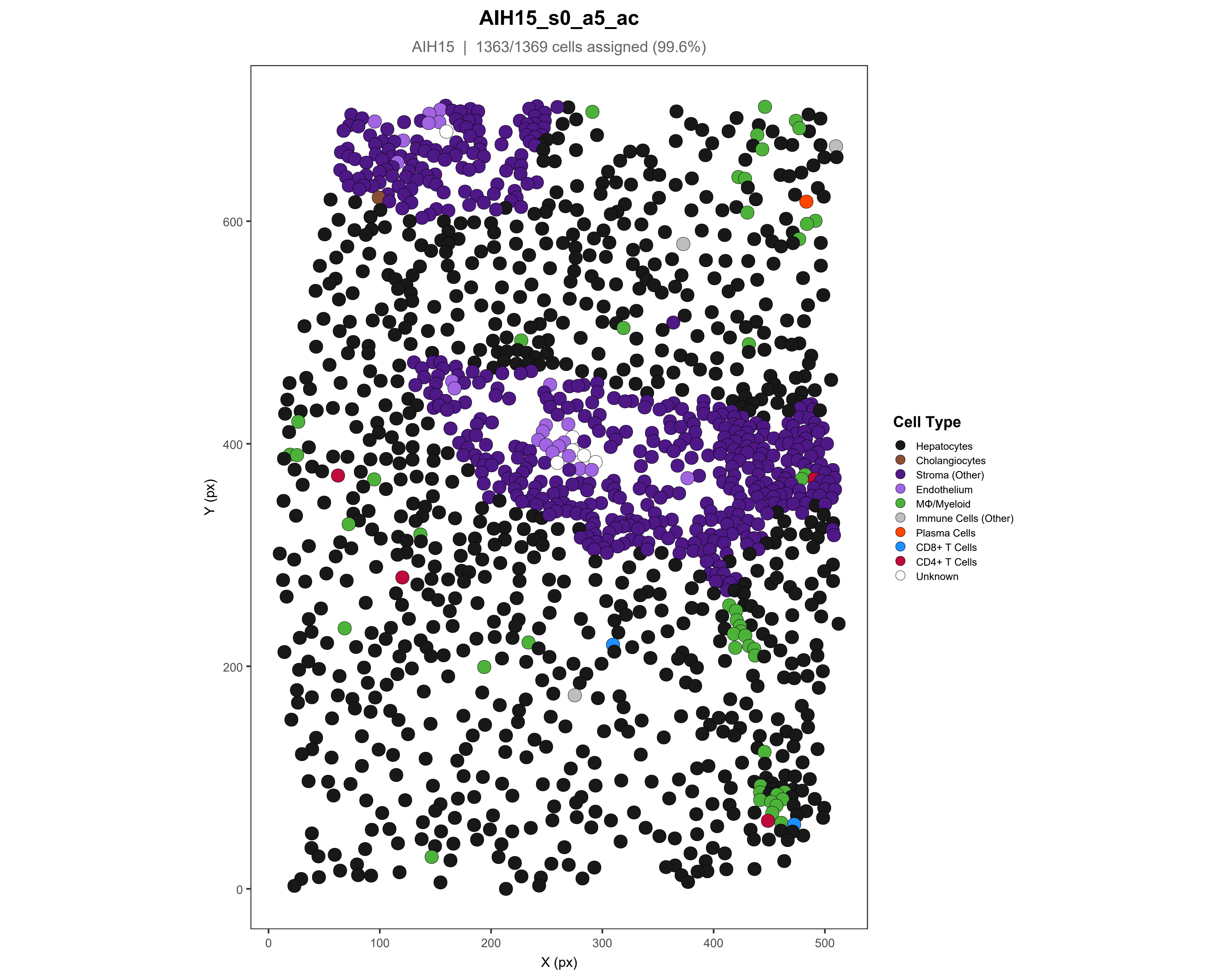

AIH15_s0_a5_ac
AIH15 | 1363/1369 cells assigned (99.6%)
Y (px)
X (px)
0
200
400
600
0
100
200
300
400
500
Cell Type
Hepatocytes
Cholangiocytes
Stroma (Other)
Endothelium
MΦ/Myeloid
Immune Cells (Other)
Plasma Cells
CD8+ T Cells
CD4+ T Cells
Unknown

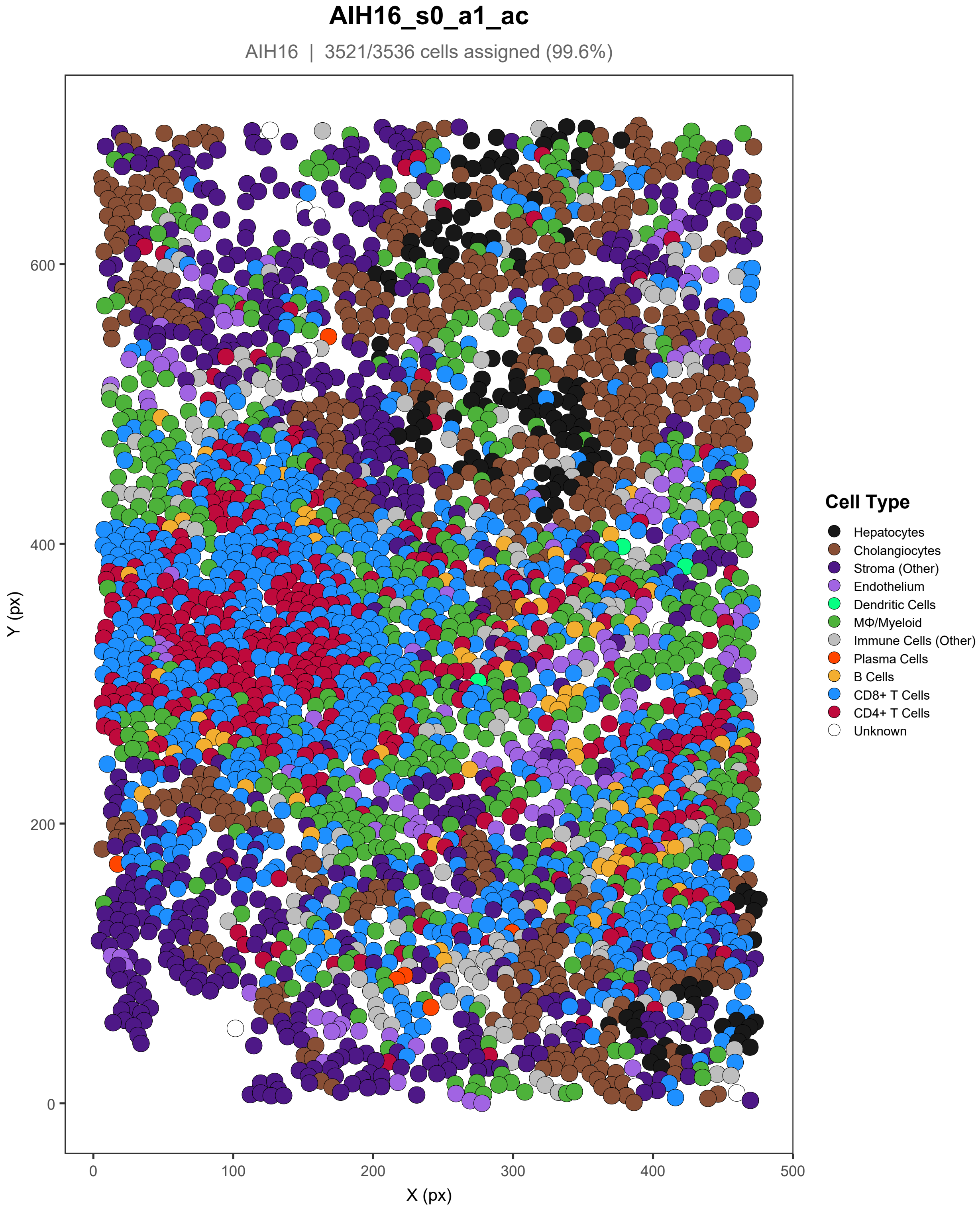

AIH16_s0_a1_ac
AIH16 | 3521/3536 cells assigned (99.6%)
Y (px)
600
400
200
0
0
100
200
300
400
500
X (px)
Cell Type
Hepatocytes
Cholangiocytes
Stroma (Other)
Endothelium
Dendritic Cells
MΦ/Myeloid
Immune Cells (Other)
Plasma Cells
B Cells
CD8+ T Cells
CD4+ T Cells
Unknown

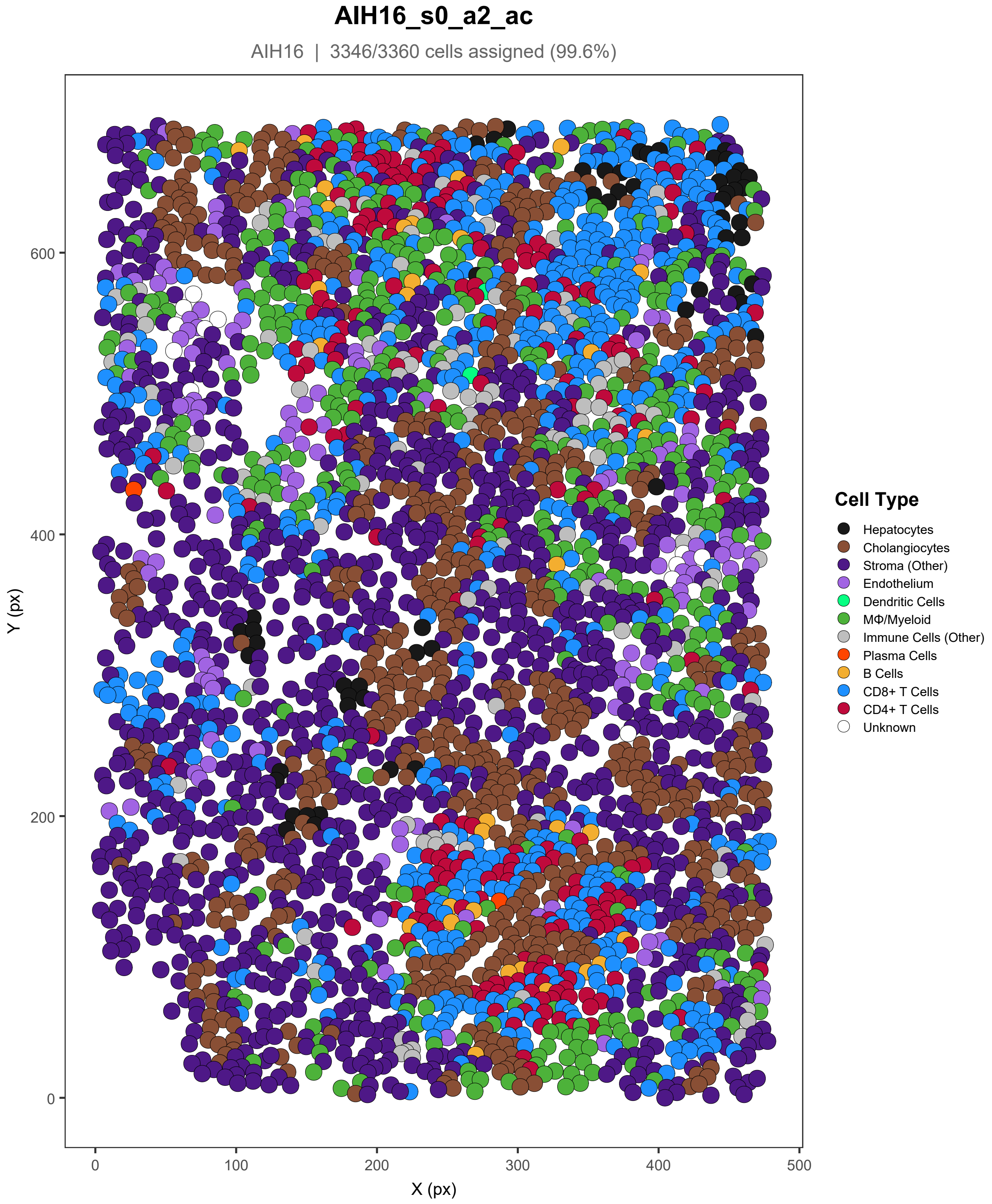

AIH16_s0_a2_ac
AIH16 | 3346/3360 cells assigned (99.6%)
Y (px)
X (px)
0
200
400
600
0
100
200
300
400
500
Cell Type
Hepatocytes
Cholangiocytes
Stroma (Other)
Endothelium
Dendritic Cells
MΦ/Myeloid
Immune Cells (Other)
Plasma Cells
B Cells
CD8+ T Cells
CD4+ T Cells
Unknown

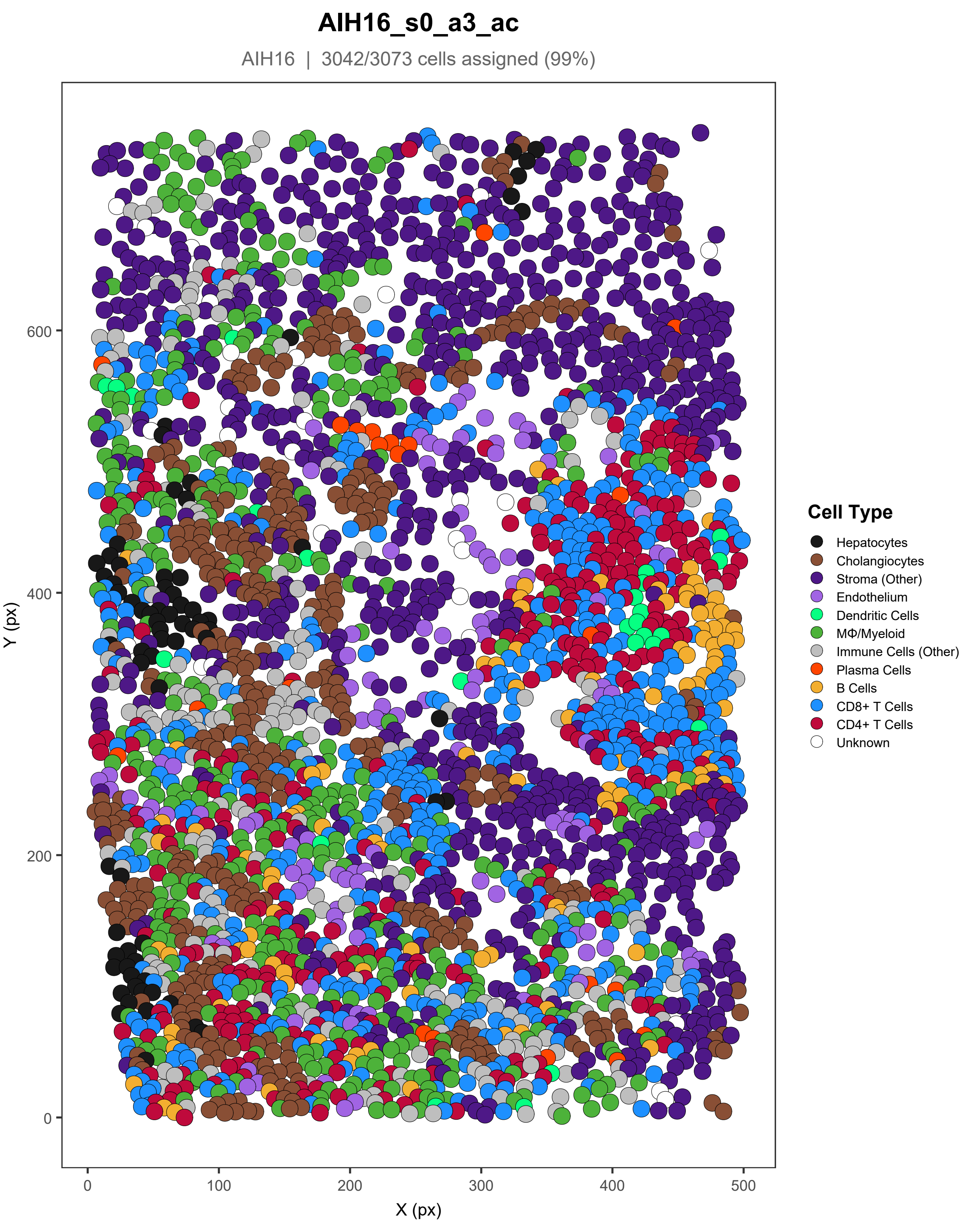
AIH16_s0_a3_ac
AIH16 | 3042/3073 cells assigned (99%)
Y (px)
X (px)
0
100
200
300
400
500
600
Cell Type
Hepatocytes
Cholangiocytes
Stroma (Other)
Endothelium
Dendritic Cells
MΦ/Myeloid
Immune Cells (Other)
Plasma Cells
B Cells
CD8+ T Cells
CD4+ T Cells
Unknown

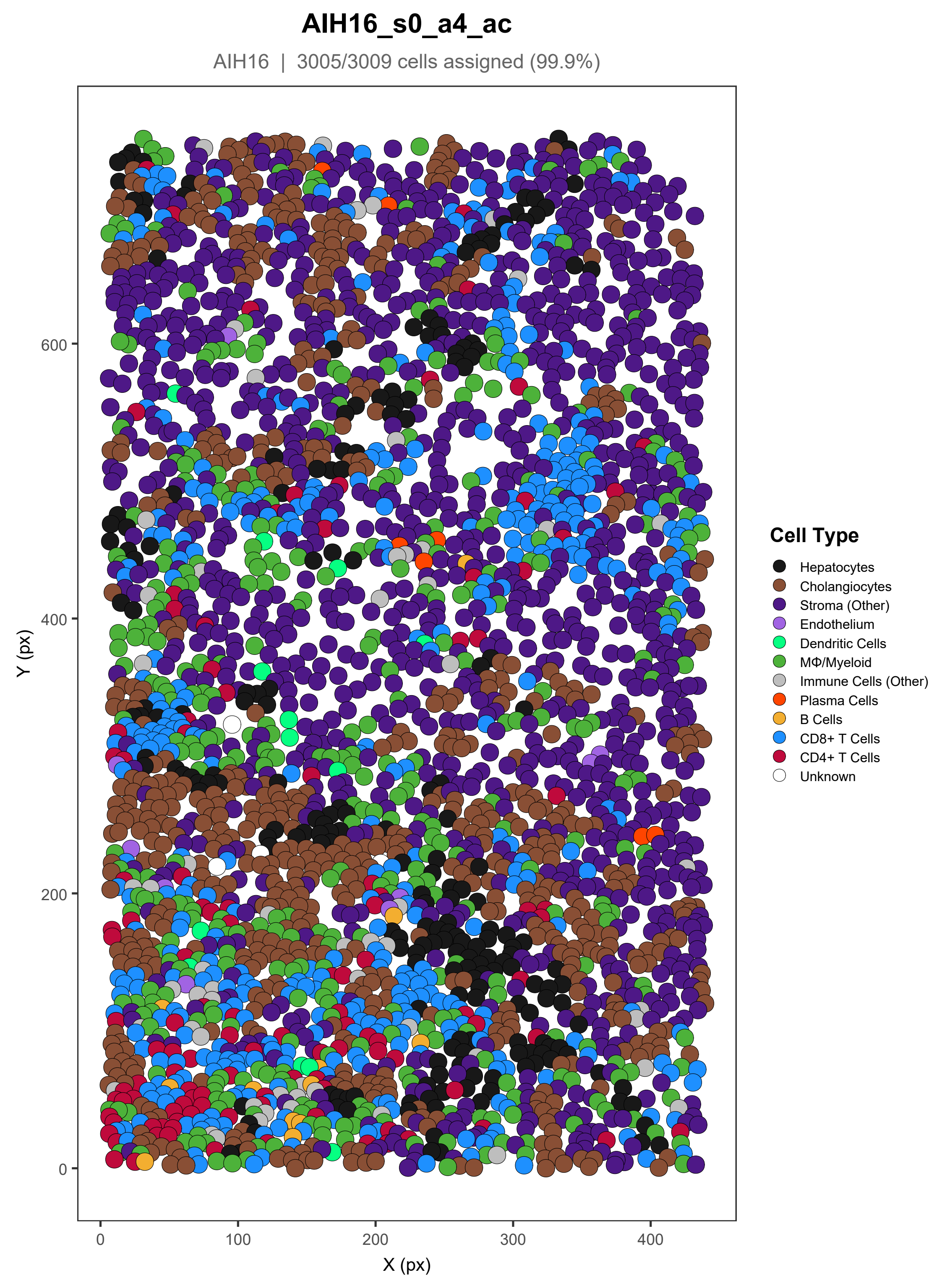
AIH16_s0_a4_ac
AIH16 | 3005/3009 cells assigned (99.9%)
Y (px)
X (px)
0
100
200
300
400
600
Cell Type
Hepatocytes
Cholangiocytes
Stroma (Other)
Endothelium
Dendritic Cells
MΦ/Myeloid
Immune Cells (Other)
Plasma Cells
B Cells
CD8+ T Cells
CD4+ T Cells
Unknown

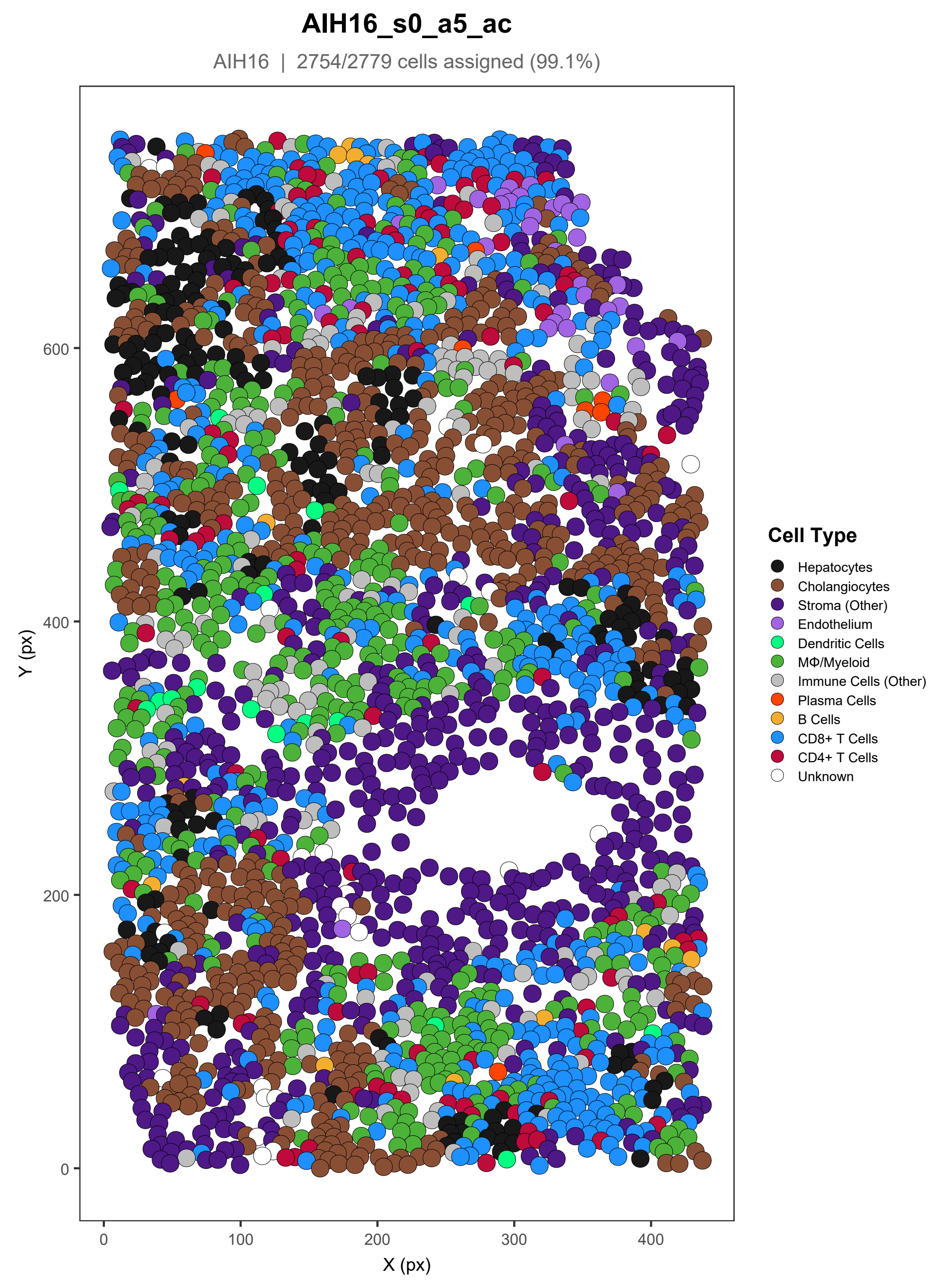
AIH16_s0_a5_ac
AIH16 | 2754/2779 cells assigned (99.1%)
Y (px)
X (px)
0
200
400
600
0
100
200
300
400
Cell Type
Hepatocytes
Cholangiocytes
Stroma (Other)
Endothelium
Dendritic Cells
MΦ/Myeloid
Immune Cells (Other)
Plasma Cells
B Cells
CD8+ T Cells
CD4+ T Cells
Unknown

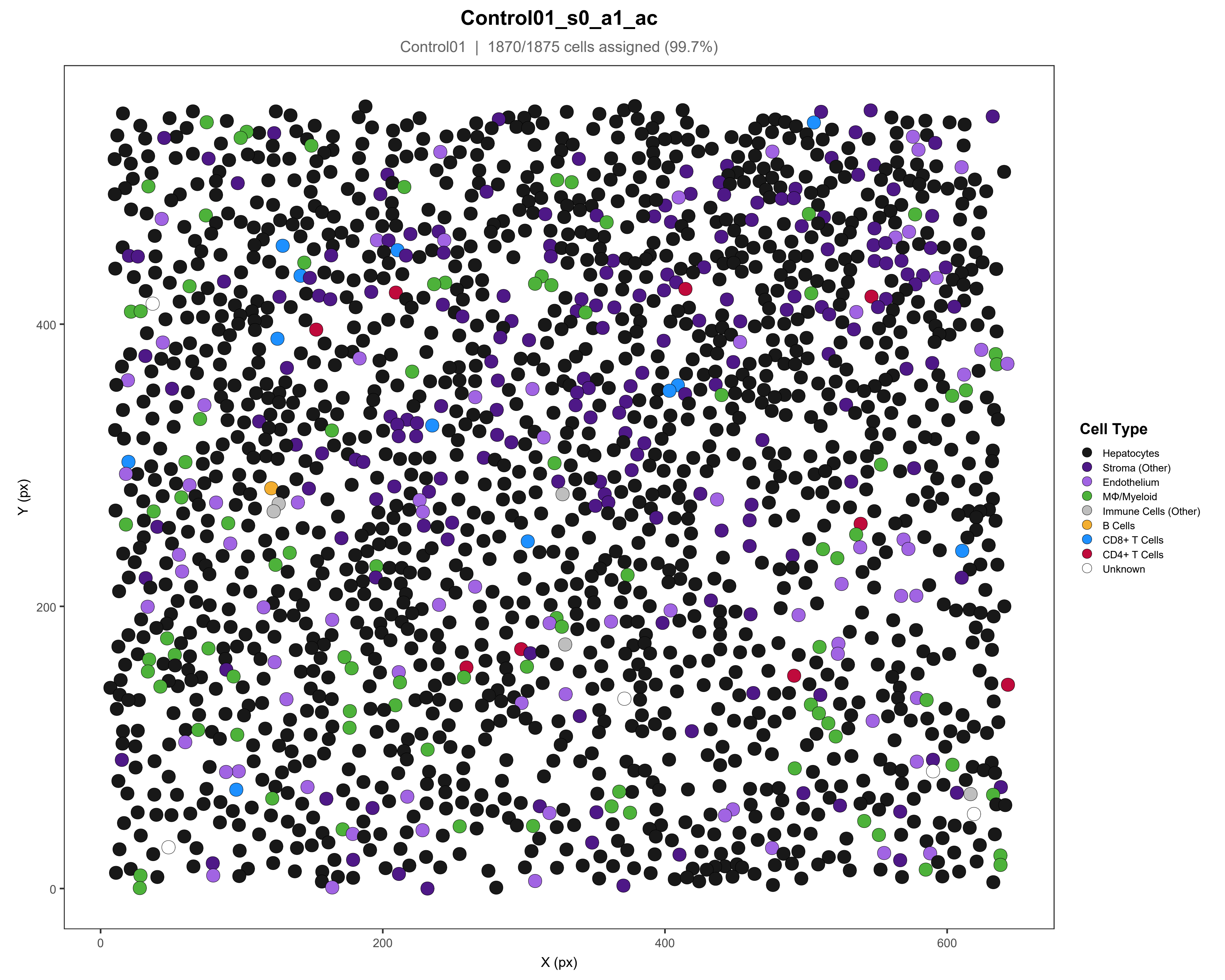

Control01_s0_a1_ac
Control01 | 1870/1875 cells assigned (99.7%)
Y (px)
X (px)
0
200
400
600
Cell Type
Hepatocytes
Stroma (Other)
Endothelium
MΦ/Myeloid
Immune Cells (Other)
B Cells
CD8+ T Cells
CD4+ T Cells
Unknown

### Control01_s0_a2_ac

Control01 | 1450/1451 cells assigned (99.9%)

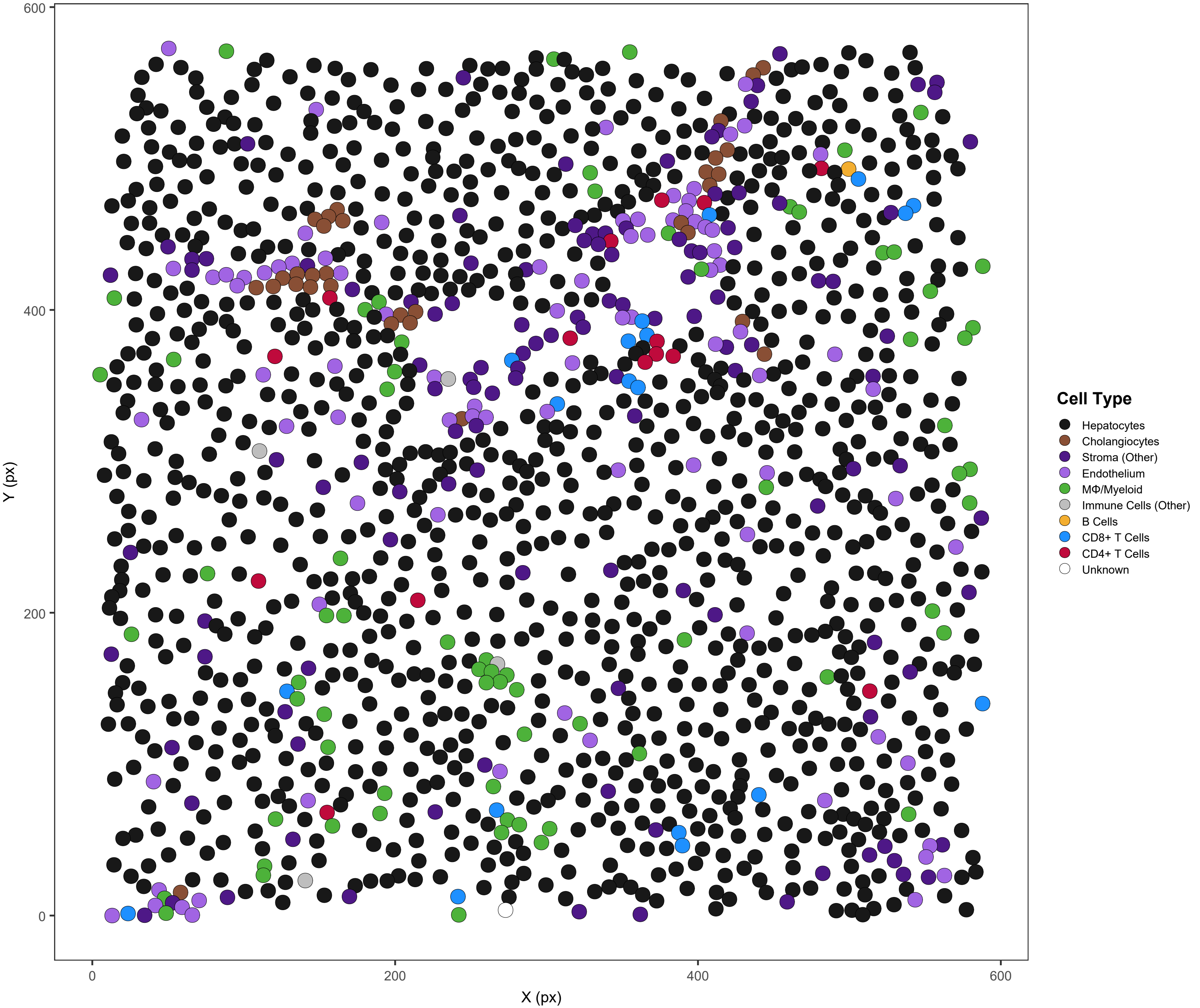

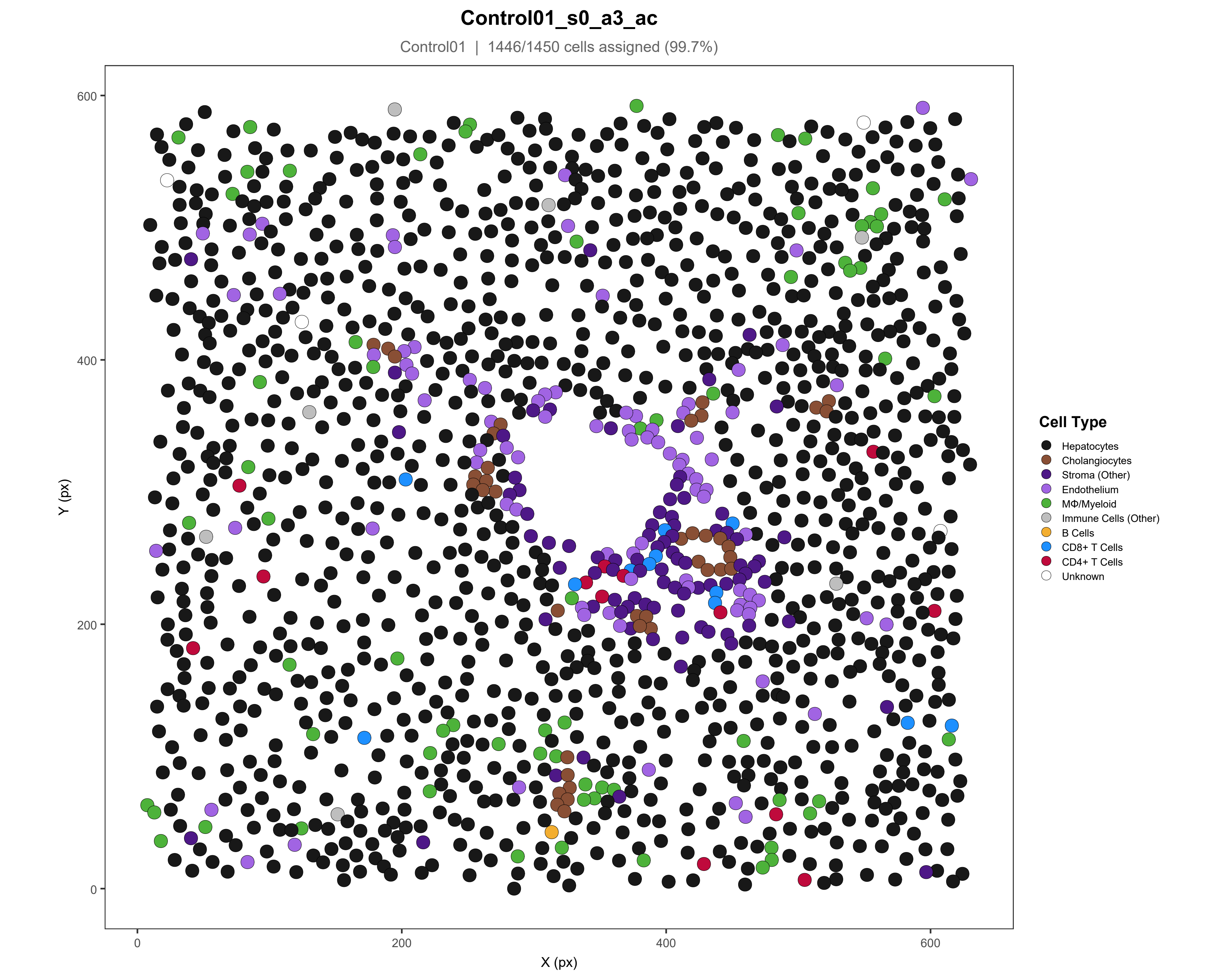

Control01_s0_a3_ac
Control01 | 1446/1450 cells assigned (99.7%)
Y (px)
X (px)
0
200
400
600
Cell Type
Hepatocytes
Cholangiocytes
Stroma (Other)
Endothelium
MΦ/Myeloid
Immune Cells (Other)
B Cells
CD8+ T Cells
CD4+ T Cells
Unknown

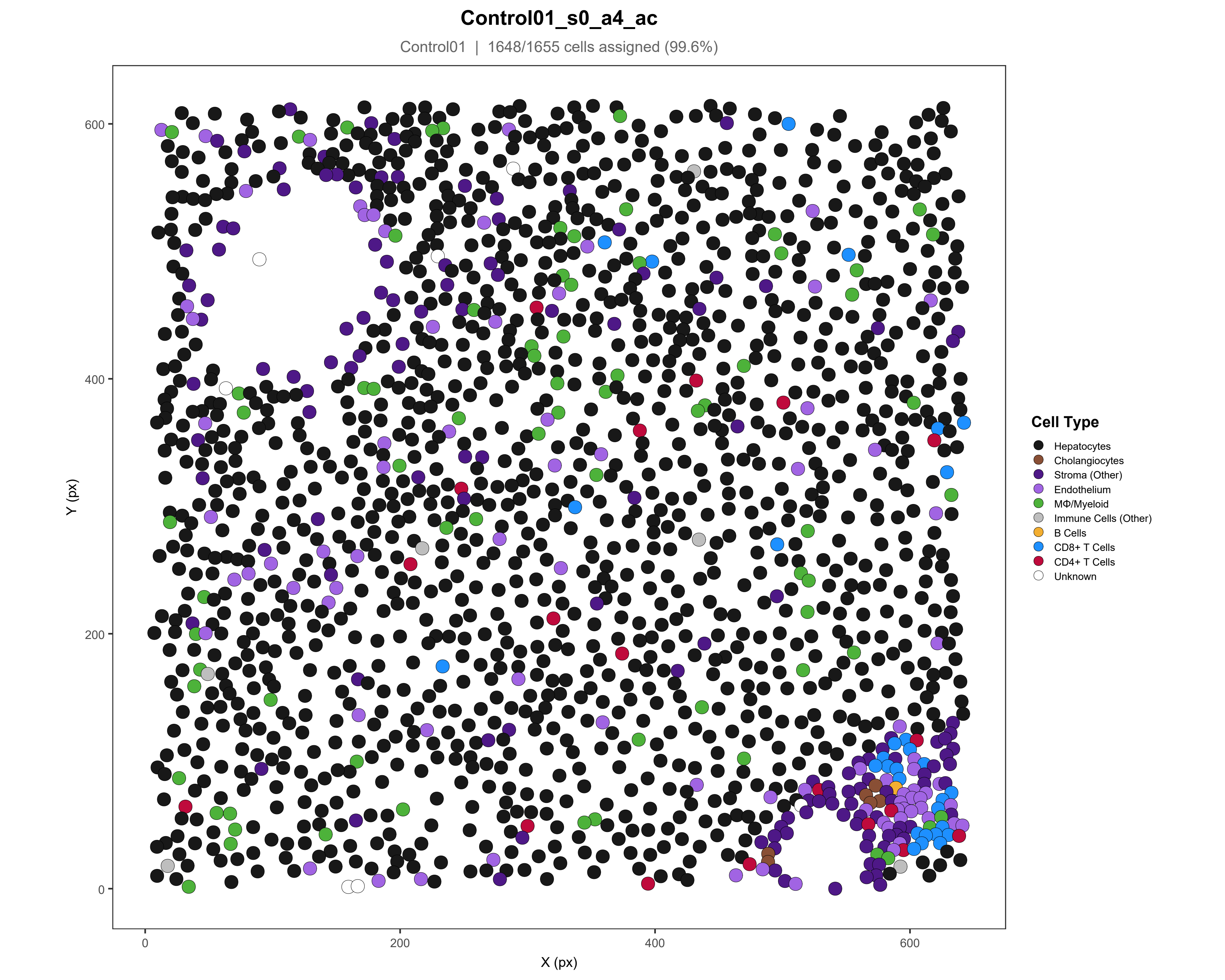

Control01_s0_a4_ac
Control01 | 1648/1655 cells assigned (99.6%)
Y (px)
X (px)
0
200
400
600
Cell Type
Hepatocytes
Cholangiocytes
Stroma (Other)
Endothelium
MΦ/Myeloid
Immune Cells (Other)
B Cells
CD8+ T Cells
CD4+ T Cells
Unknown

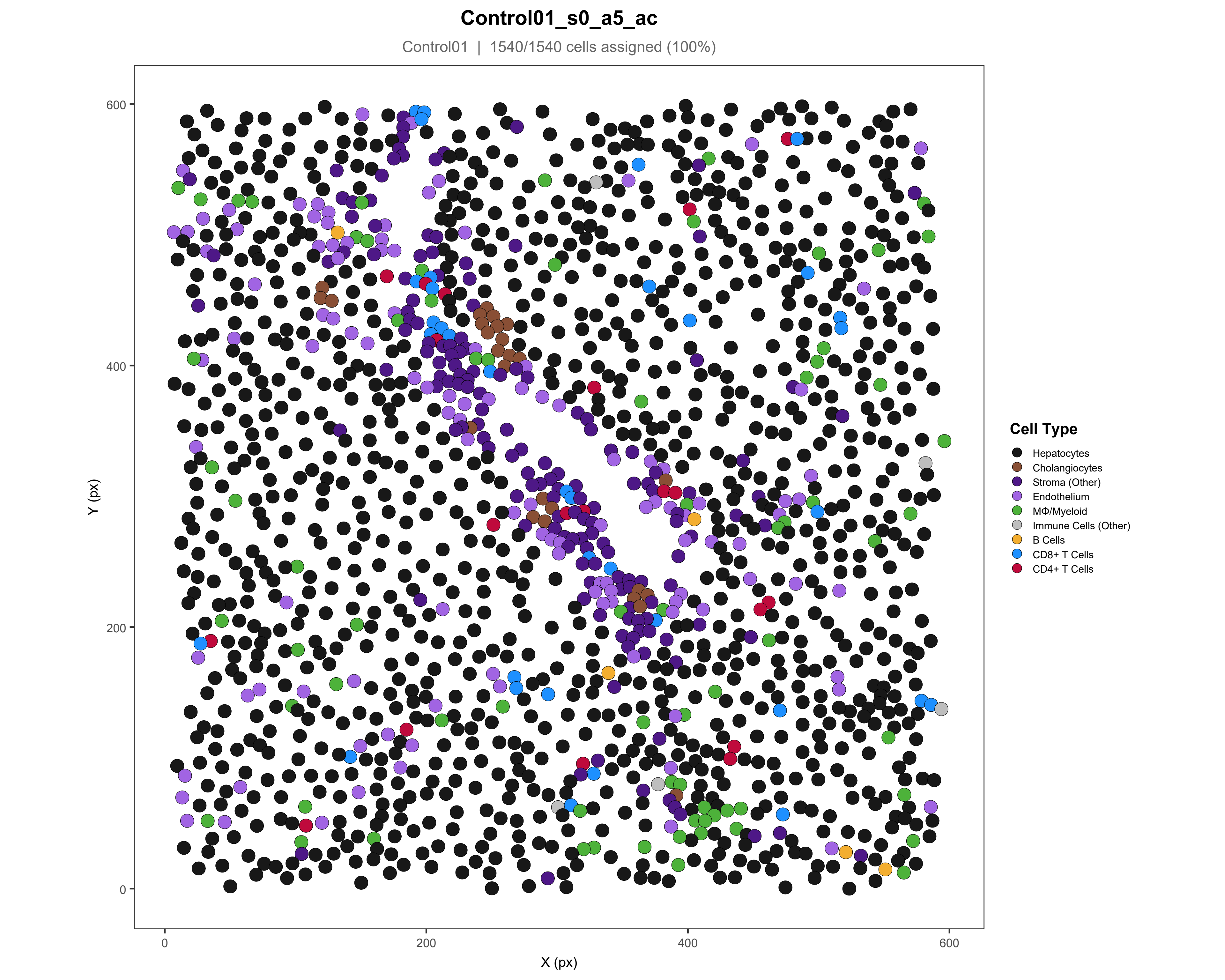
Control01_s0_a5_ac
Control01 | 1540/1540 cells assigned (100%)
Y (px)
X (px)
0
200
400
600
Cell Type
Hepatocytes
Cholangiocytes
Stroma (Other)
Endothelium
MΦ/Myeloid
Immune Cells (Other)
B Cells
CD8+ T Cells
CD4+ T Cells

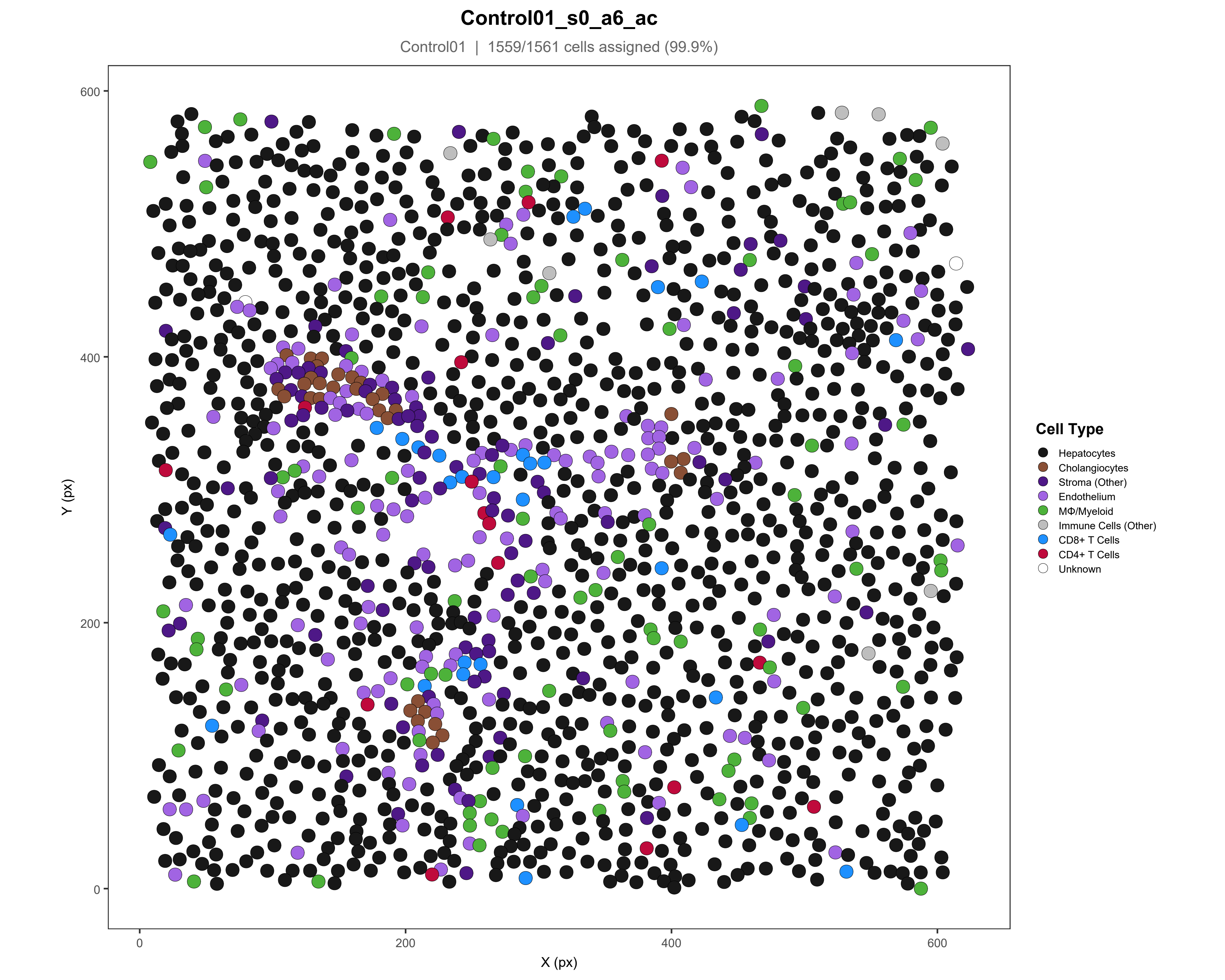
Control01_s0_a6_ac
Control01 | 1559/1561 cells assigned (99.9%)
Y (px)
X (px)
0
200
400
600
Cell Type
Hepatocytes
Cholangiocytes
Stroma (Other)
Endothelium
MΦ/Myeloid
Immune Cells (Other)
CD8+ T Cells
CD4+ T Cells
Unknown

### Control01_s0_a7_ac

Control01 | 1500/1504 cells assigned (99.7%)

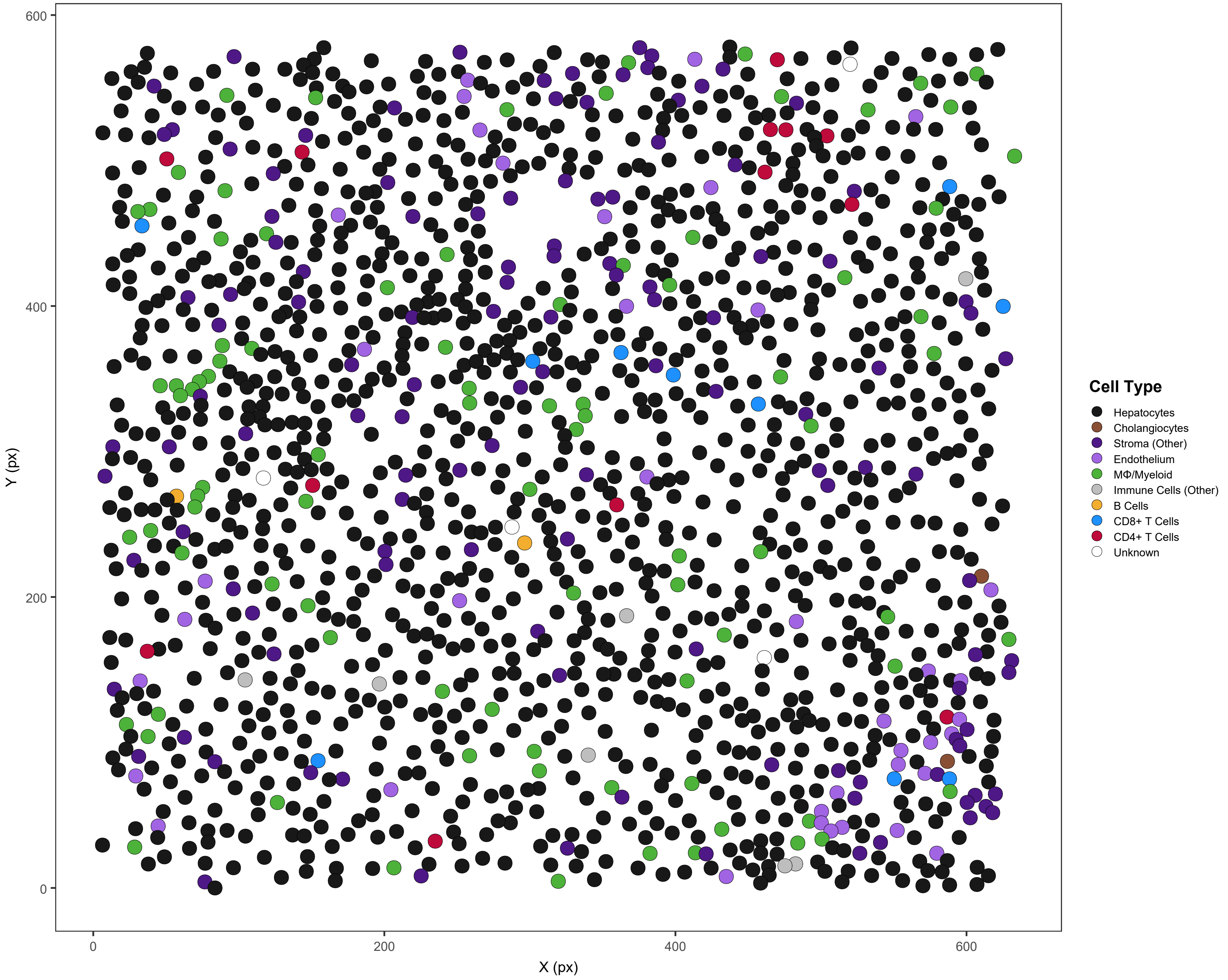

### Control01_s0_a8_ac

Control01 | 1659/1660 cells assigned (99.9%)

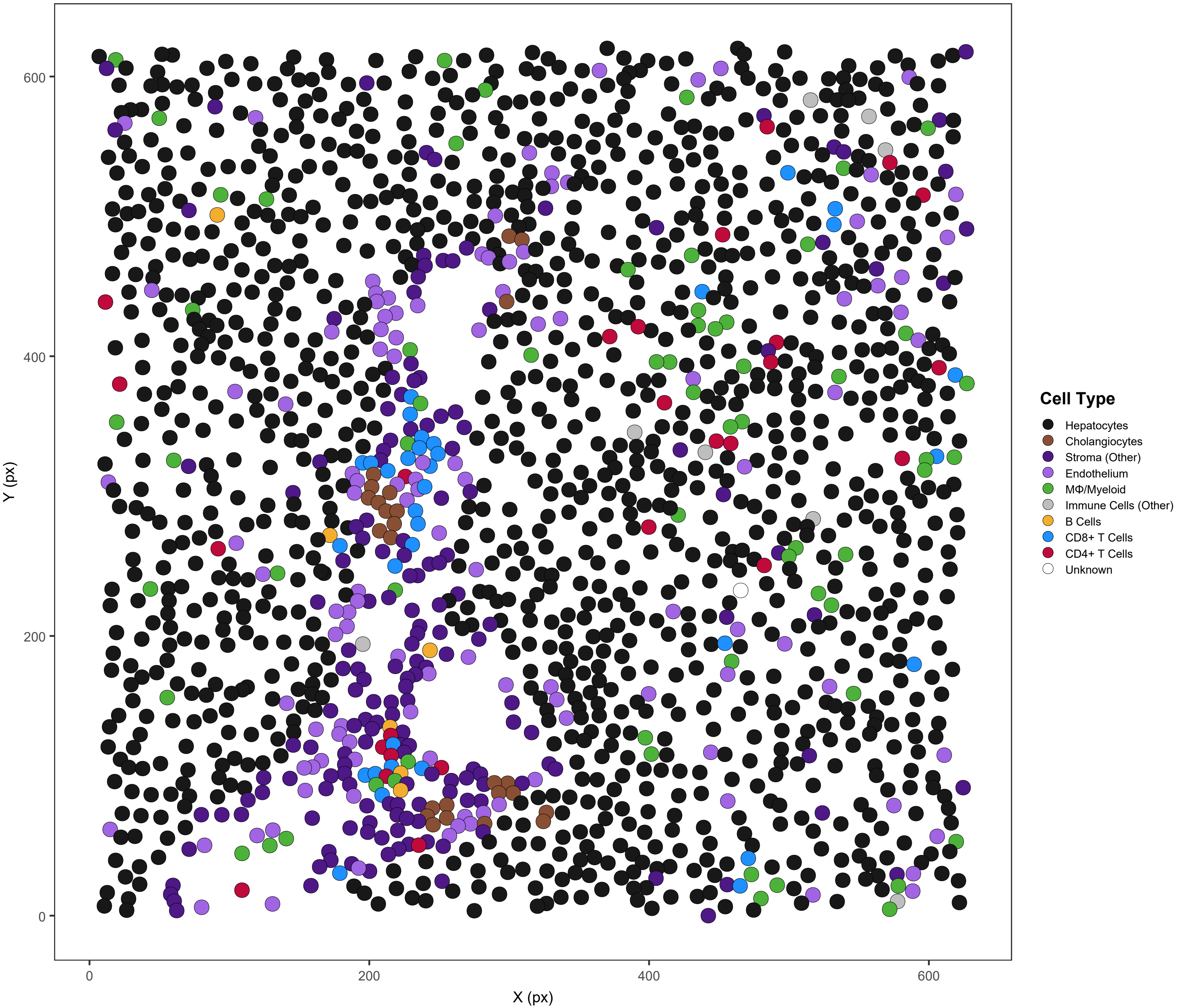

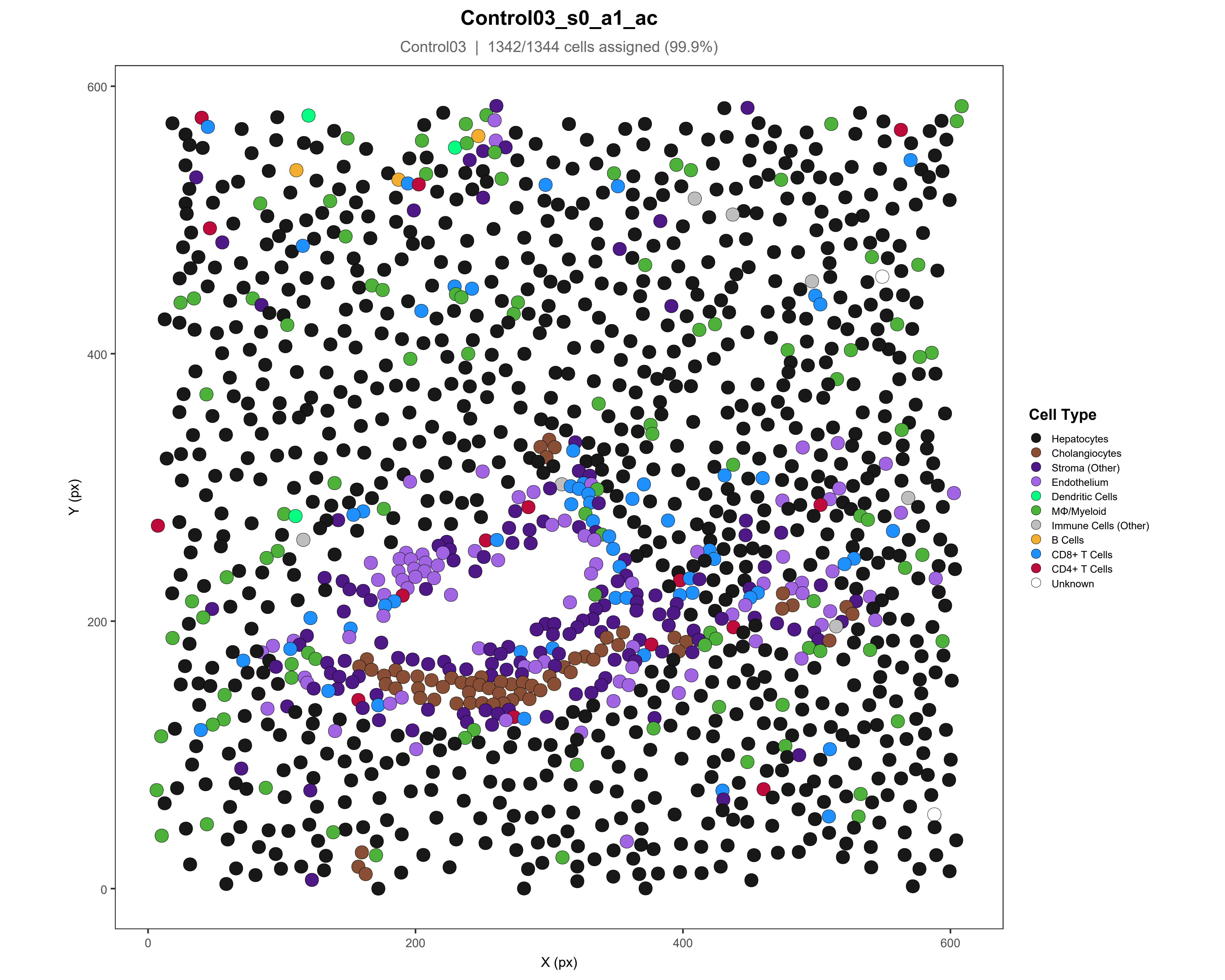
Control03_s0_a1_ac
Control03 | 1342/1344 cells assigned (99.9%)
Y (px)
X (px)
0
200
400
600
Cell Type
Hepatocytes
Cholangiocytes
Stroma (Other)
Endothelium
Dendritic Cells
MΦ/Myeloid
Immune Cells (Other)
B Cells
CD8+ T Cells
CD4+ T Cells
Unknown

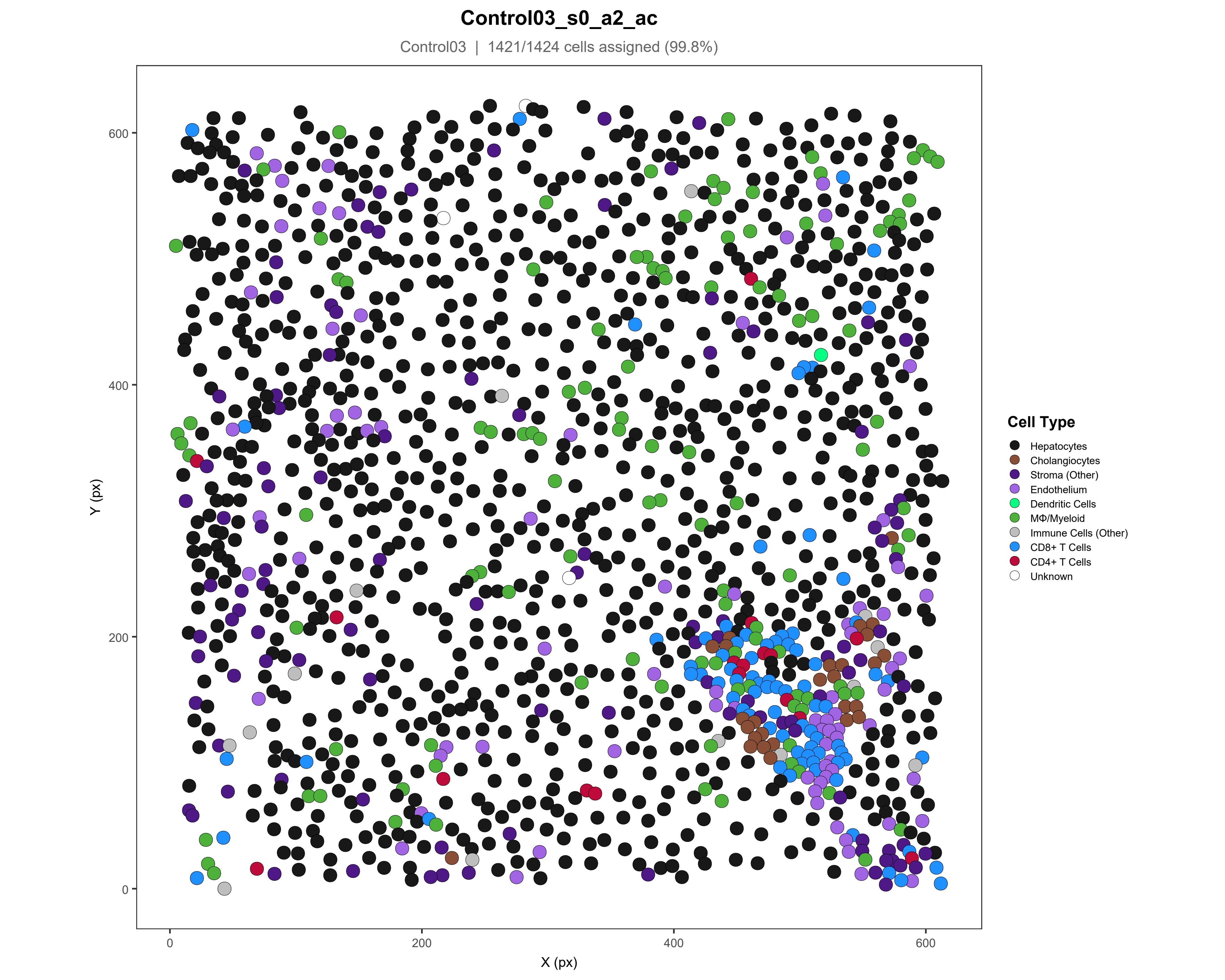

Control03_s0_a2_ac
Control03 | 1421/1424 cells assigned (99.8%)
Y (px)
X (px)
0
200
400
600
Cell Type
Hepatocytes
Cholangiocytes
Stroma (Other)
Endothelium
Dendritic Cells
MΦ/Myeloid
Immune Cells (Other)
CD8+ T Cells
CD4+ T Cells
Unknown

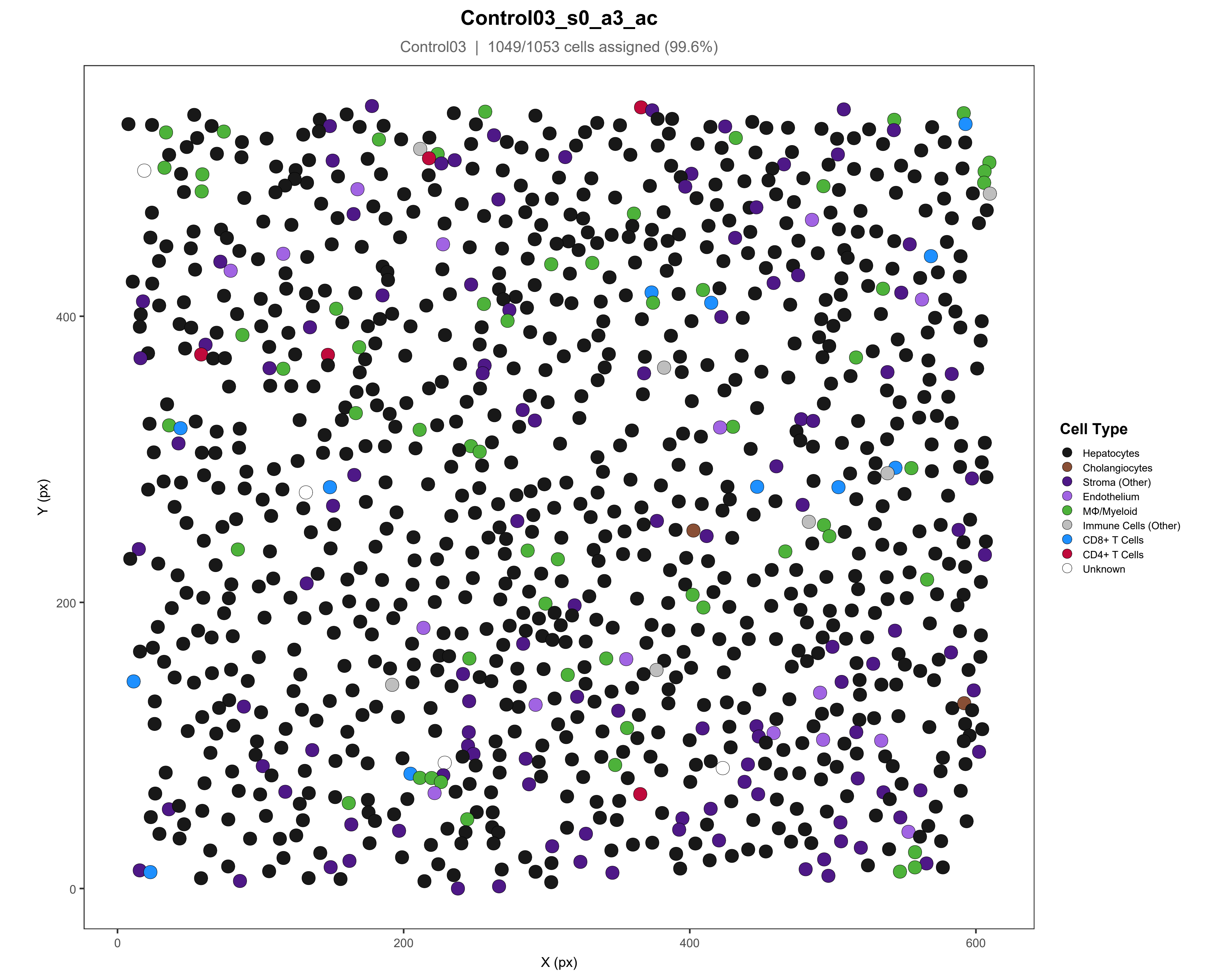

Control03_s0_a3_ac
Control03 | 1049/1053 cells assigned (99.6%)
Y (px)
X (px)
0
200
400
600
Cell Type
Hepatocytes
Cholangiocytes
Stroma (Other)
Endothelium
MΦ/Myeloid
Immune Cells (Other)
CD8+ T Cells
CD4+ T Cells
Unknown

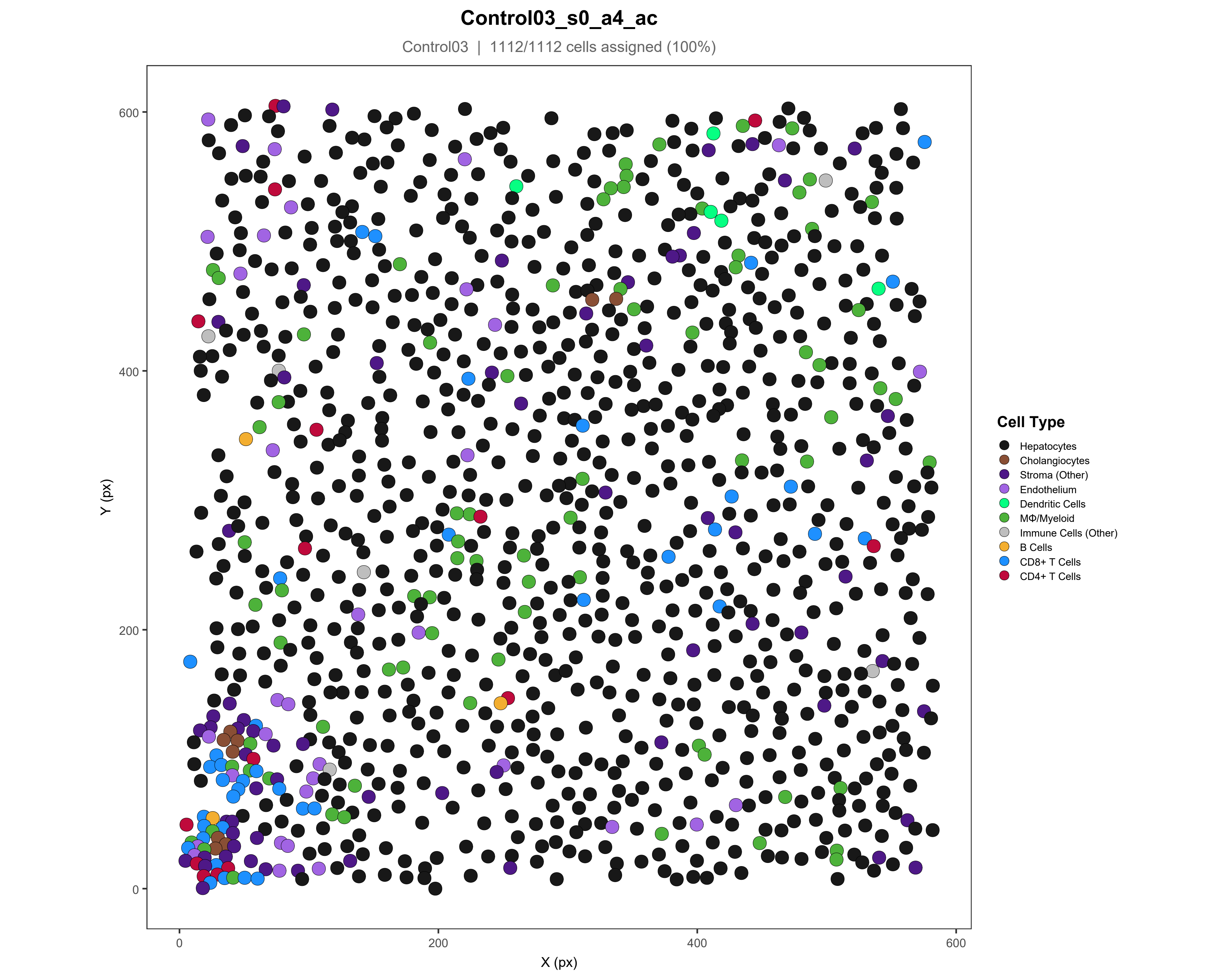

Control03_s0_a4_ac
Control03 | 1112/1112 cells assigned (100%)
Y (px)
X (px)
0
200
400
600
Cell Type
Hepatocytes
Cholangiocytes
Stroma (Other)
Endothelium
Dendritic Cells
MΦ/Myeloid
Immune Cells (Other)
B Cells
CD8+ T Cells
CD4+ T Cells

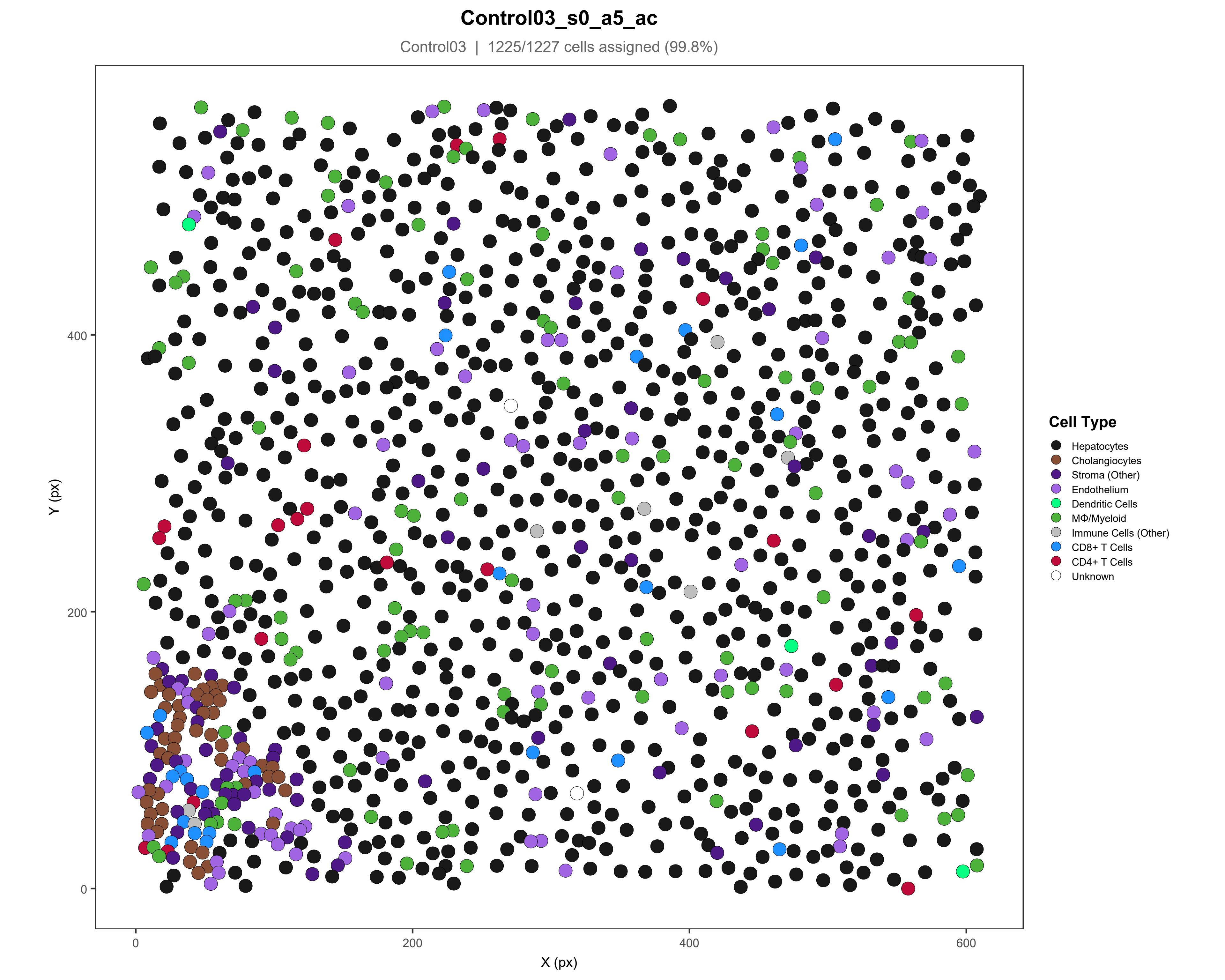
Control03_s0_a5_ac
Control03 | 1225/1227 cells assigned (99.8%)
Y (px)
X (px)
0
200
400
600
Cell Type
Hepatocytes
Cholangiocytes
Stroma (Other)
Endothelium
Dendritic Cells
MΦ/Myeloid
Immune Cells (Other)
CD8+ T Cells
CD4+ T Cells
Unknown

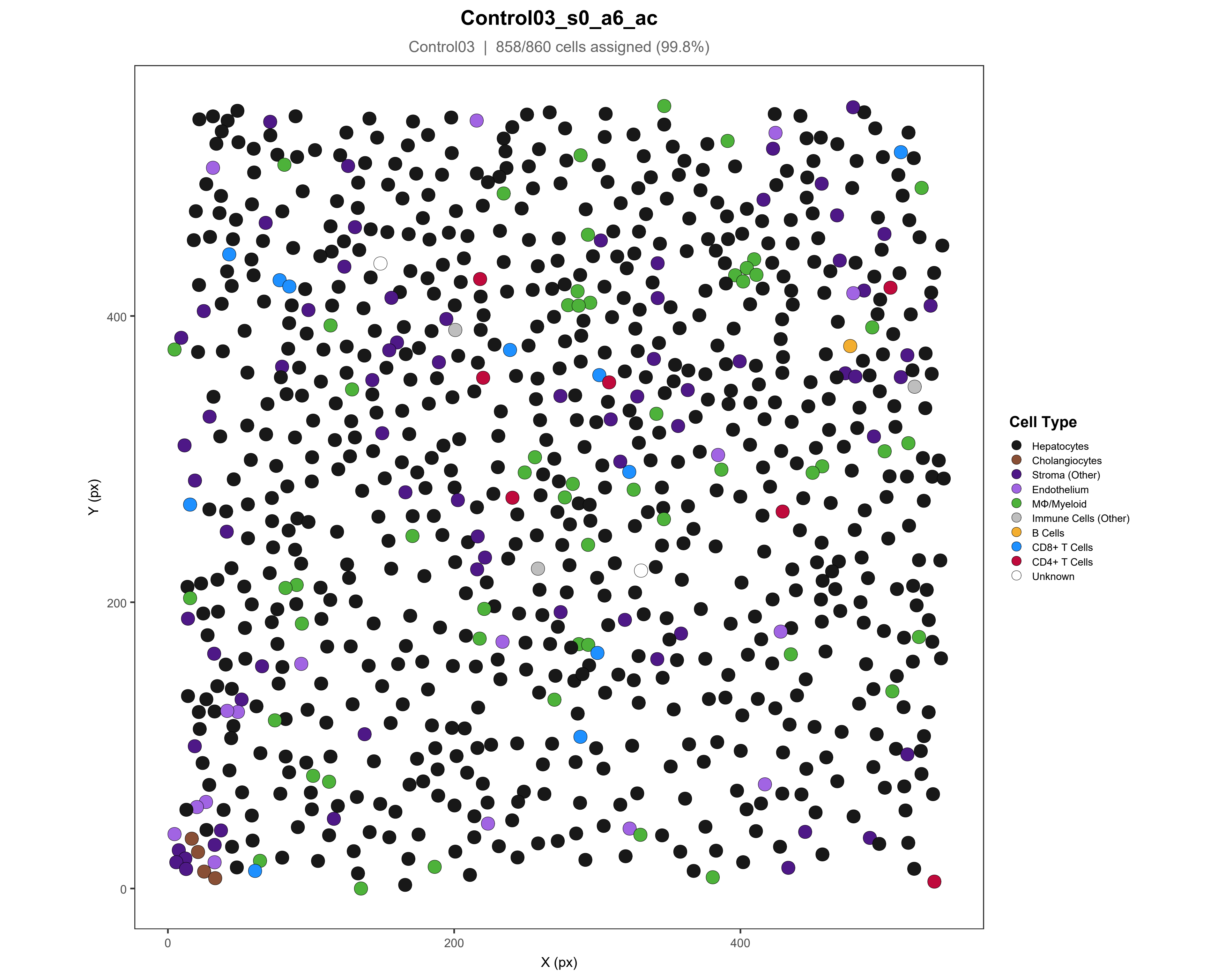

Control03_s0_a6_ac
Control03 | 858/860 cells assigned (99.8%)
Y (px)
X (px)
0
200
400
Cell Type
Hepatocytes
Cholangiocytes
Stroma (Other)
Endothelium
MΦ/Myeloid
Immune Cells (Other)
B Cells
CD8+ T Cells
CD4+ T Cells
Unknown

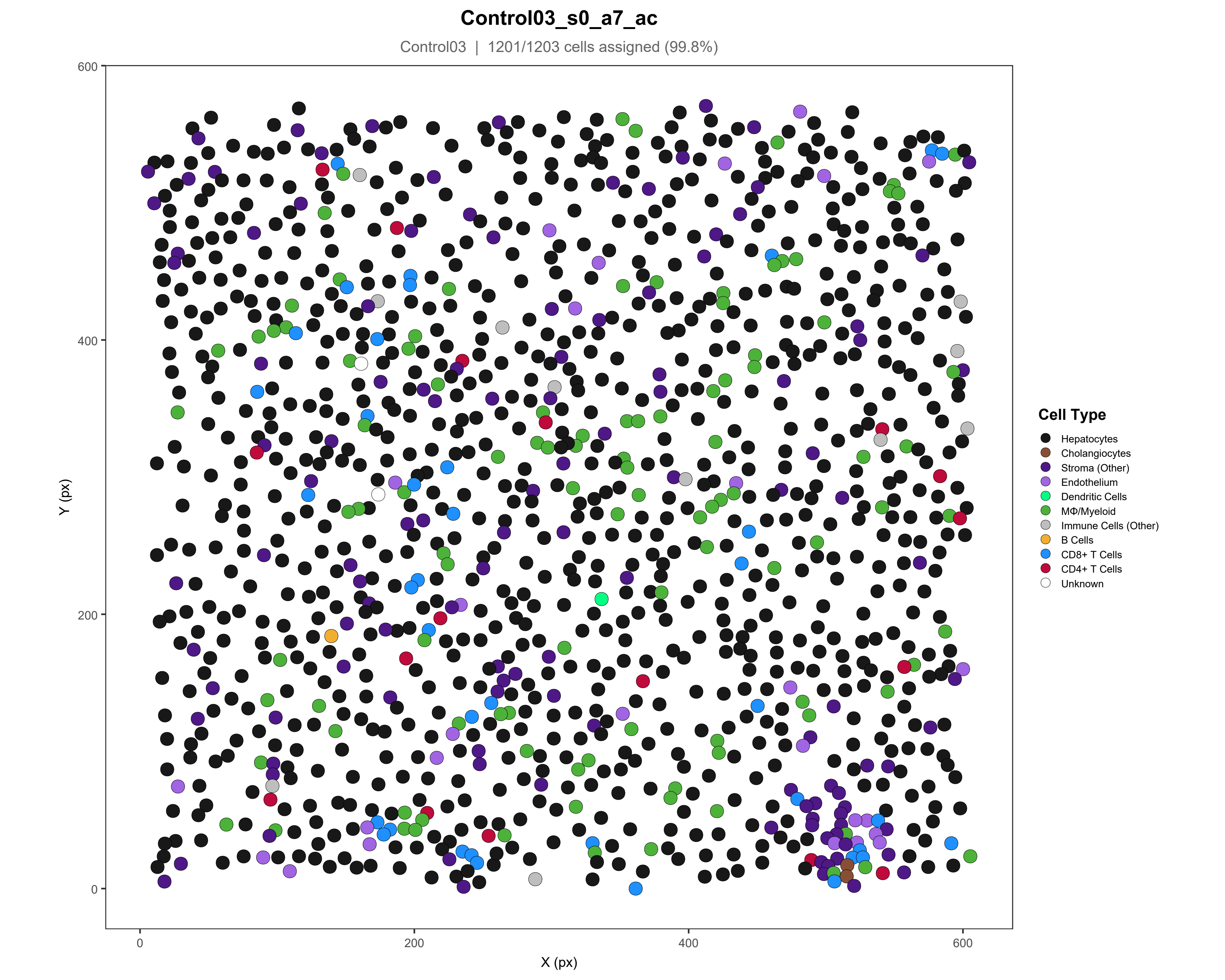
Control03_s0_a7_ac
Control03 | 1201/1203 cells assigned (99.8%)
Y (px)
X (px)
0
200
400
600
Cell Type
Hepatocytes
Cholangiocytes
Stroma (Other)
Endothelium
Dendritic Cells
MΦ/Myeloid
Immune Cells (Other)
B Cells
CD8+ T Cells
CD4+ T Cells
Unknown

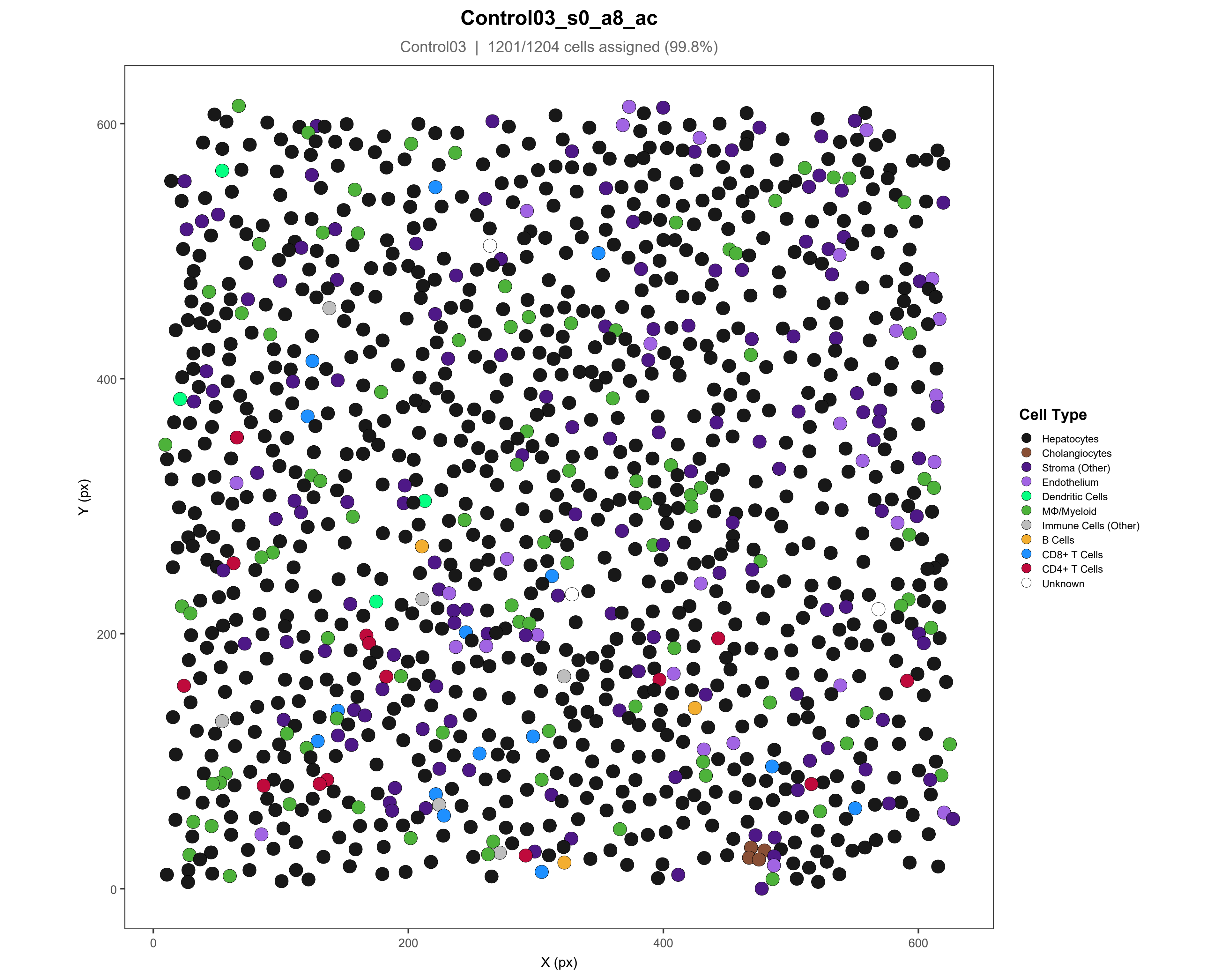
Control03_s0_a8_ac
Control03 | 1201/1204 cells assigned (99.8%)
Y (px)
X (px)
0
200
400
600
Cell Type
Hepatocytes
Cholangiocytes
Stroma (Other)
Endothelium
Dendritic Cells
MΦ/Myeloid
Immune Cells (Other)
B Cells
CD8+ T Cells
CD4+ T Cells
Unknown

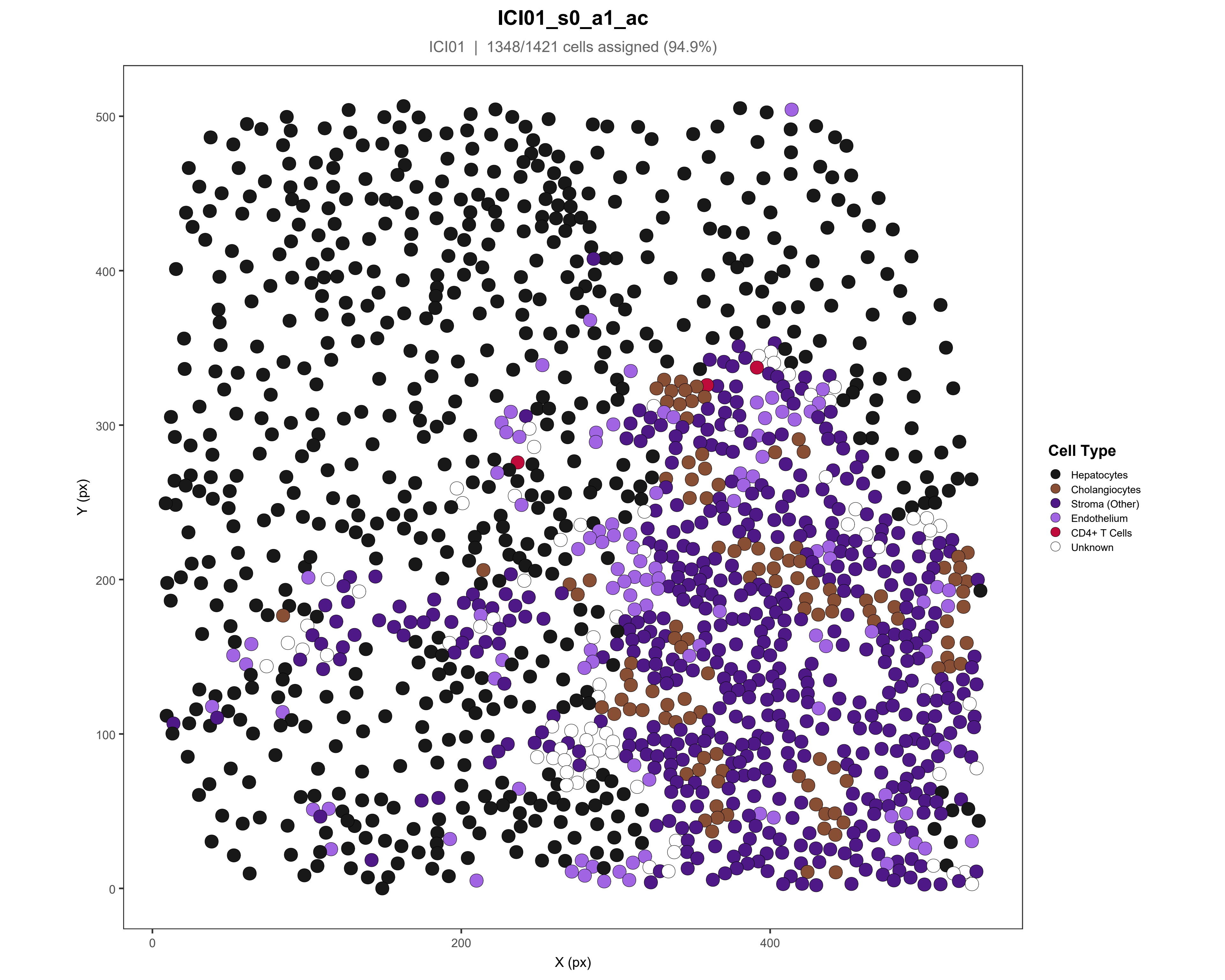
ICI01_s0_a1_ac
ICI01 | 1348/1421 cells assigned (94.9%)
Y (px)
X (px)
0
200
400
100
300
500
Cell Type
Hepatocytes
Cholangiocytes
Stroma (Other)
Endothelium
CD4+ T Cells
Unknown

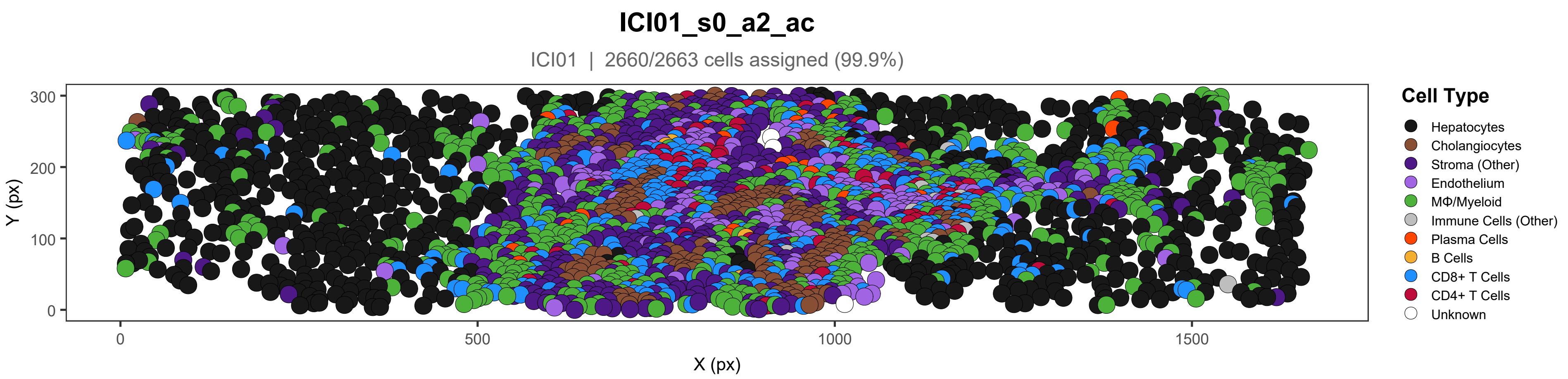

ICI01_s0_a2_ac
ICI01 | 2660/2663 cells assigned (99.9%)
300
200
100
0
Y (px)
0
500
1000
1500
X (px)
Cell Type
Hepatocytes
Cholangiocytes
Stroma (Other)
Endothelium
MΦ/Myeloid
Immune Cells (Other)
Plasma Cells
B Cells
CD8+ T Cells
CD4+ T Cells
Unknown

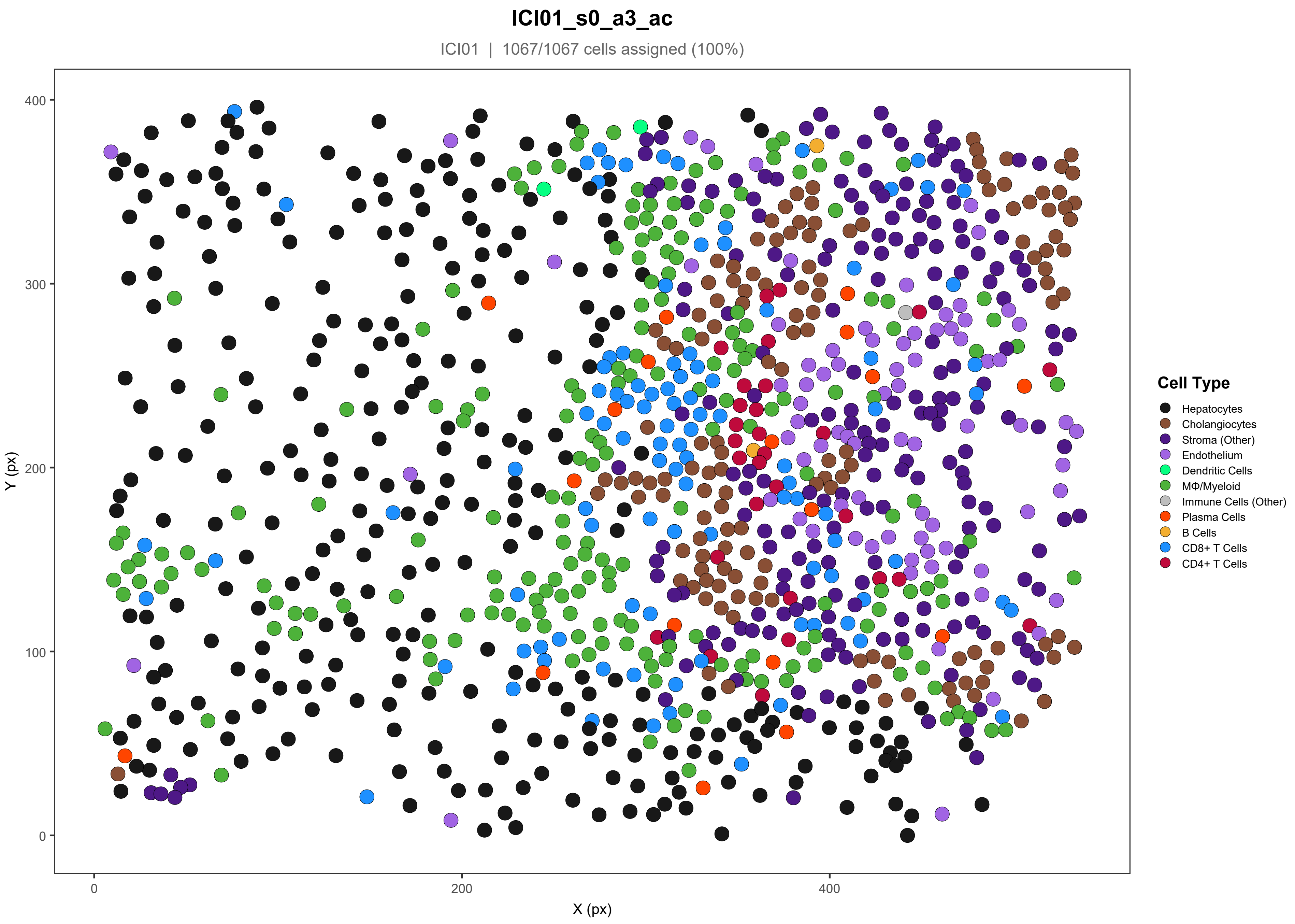
ICI01_s0_a3_ac
ICI01 | 1067/1067 cells assigned (100%)
Y (px)
X (px)
0
100
200
300
400
Cell Type
Hepatocytes
Cholangiocytes
Stroma (Other)
Endothelium
Dendritic Cells
MΦ/Myeloid
Immune Cells (Other)
Plasma Cells
B Cells
CD8+ T Cells
CD4+ T Cells

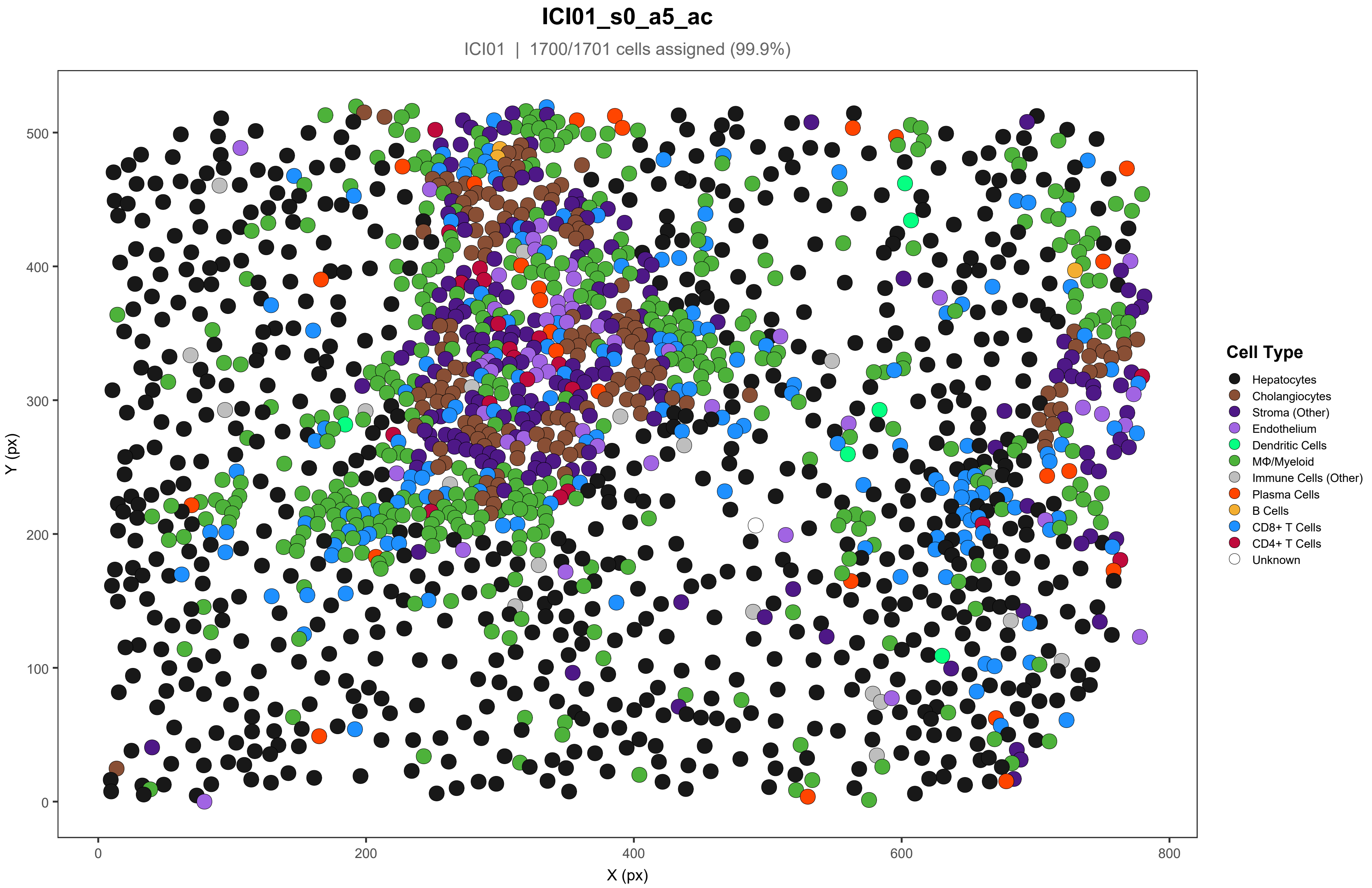
ICI01_s0_a5_ac
ICI01 | 1700/1701 cells assigned (99.9%)
Y (px)
X (px)
0
100
200
300
400
500
0
200
400
600
800
Cell Type
Hepatocytes
Cholangiocytes
Stroma (Other)
Endothelium
Dendritic Cells
MΦ/Myeloid
Immune Cells (Other)
Plasma Cells
B Cells
CD8+ T Cells
CD4+ T Cells
Unknown

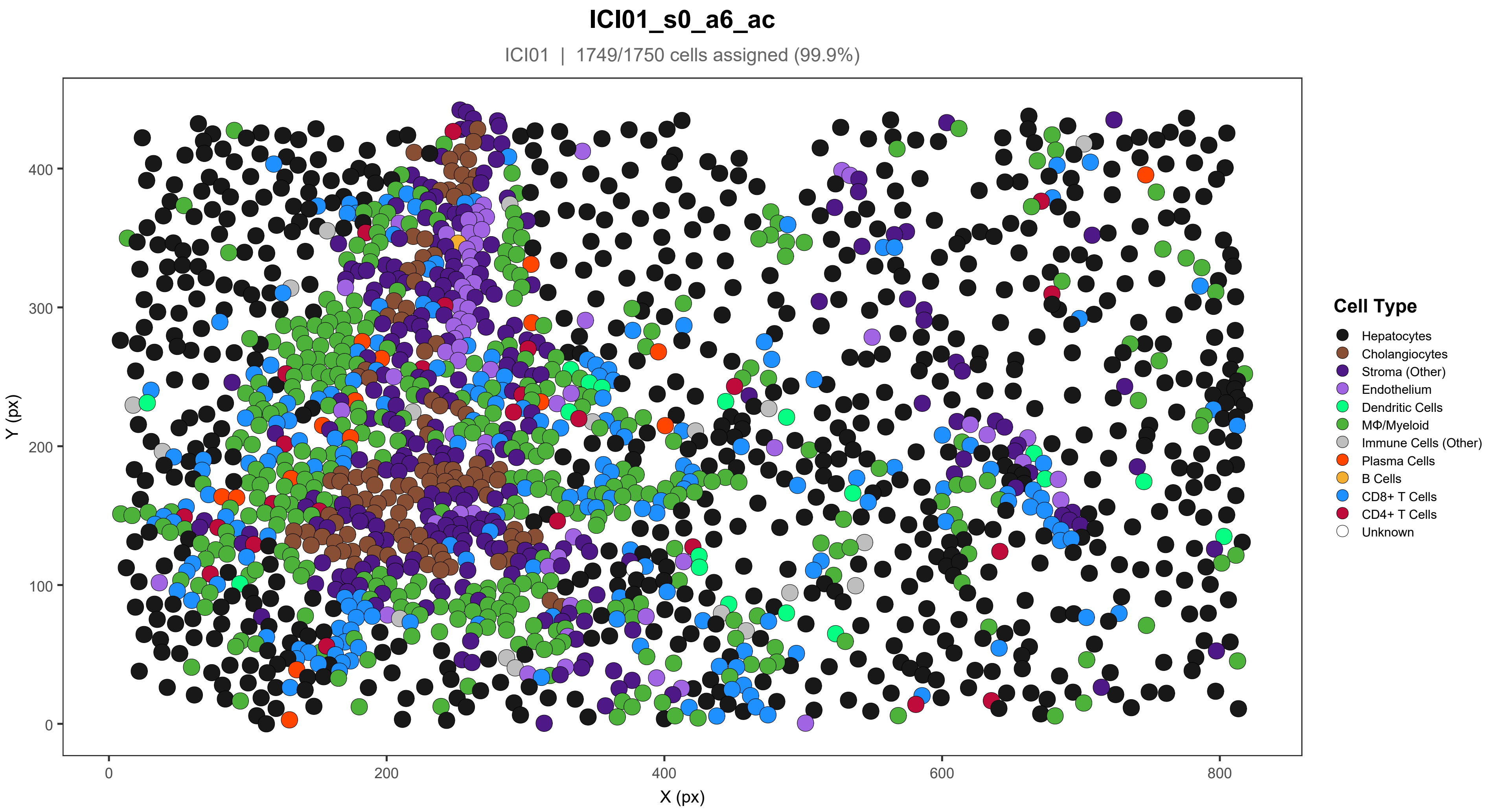

ICI01_s0_a6_ac
ICI01 | 1749/1750 cells assigned (99.9%)
Y (px)
X (px)
0
100
200
300
400
0
200
400
600
800
Cell Type
Hepatocytes
Cholangiocytes
Stroma (Other)
Endothelium
Dendritic Cells
MΦ/Myeloid
Immune Cells (Other)
Plasma Cells
B Cells
CD8+ T Cells
CD4+ T Cells
Unknown

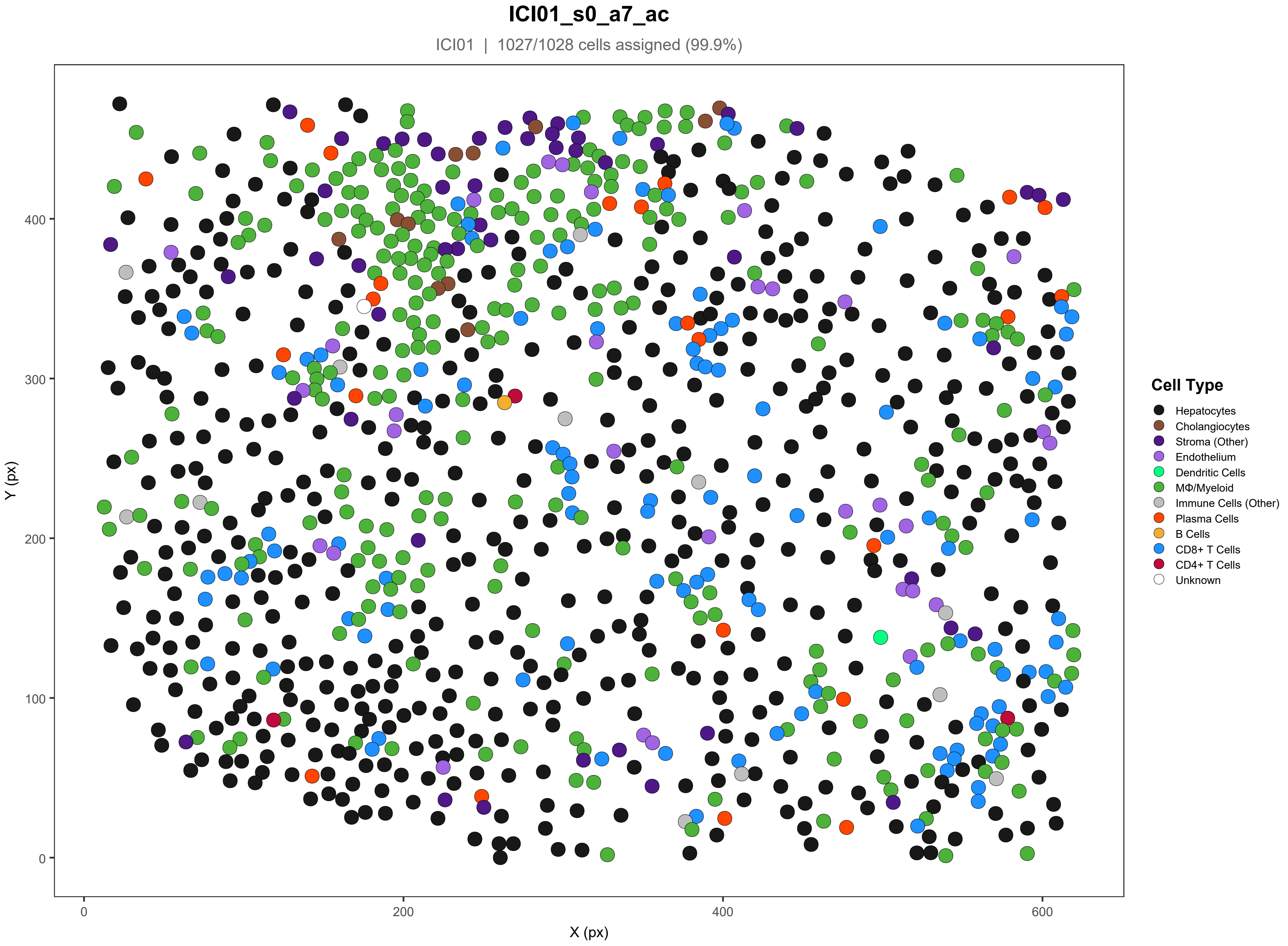
ICI01_s0_a7_ac
ICI01 | 1027/1028 cells assigned (99.9%)
Y (px)
X (px)
0
100
200
300
400
0
200
400
600
Cell Type
Hepatocytes
Cholangiocytes
Stroma (Other)
Endothelium
Dendritic Cells
MΦ/Myeloid
Immune Cells (Other)
Plasma Cells
B Cells
CD8+ T Cells
CD4+ T Cells
Unknown

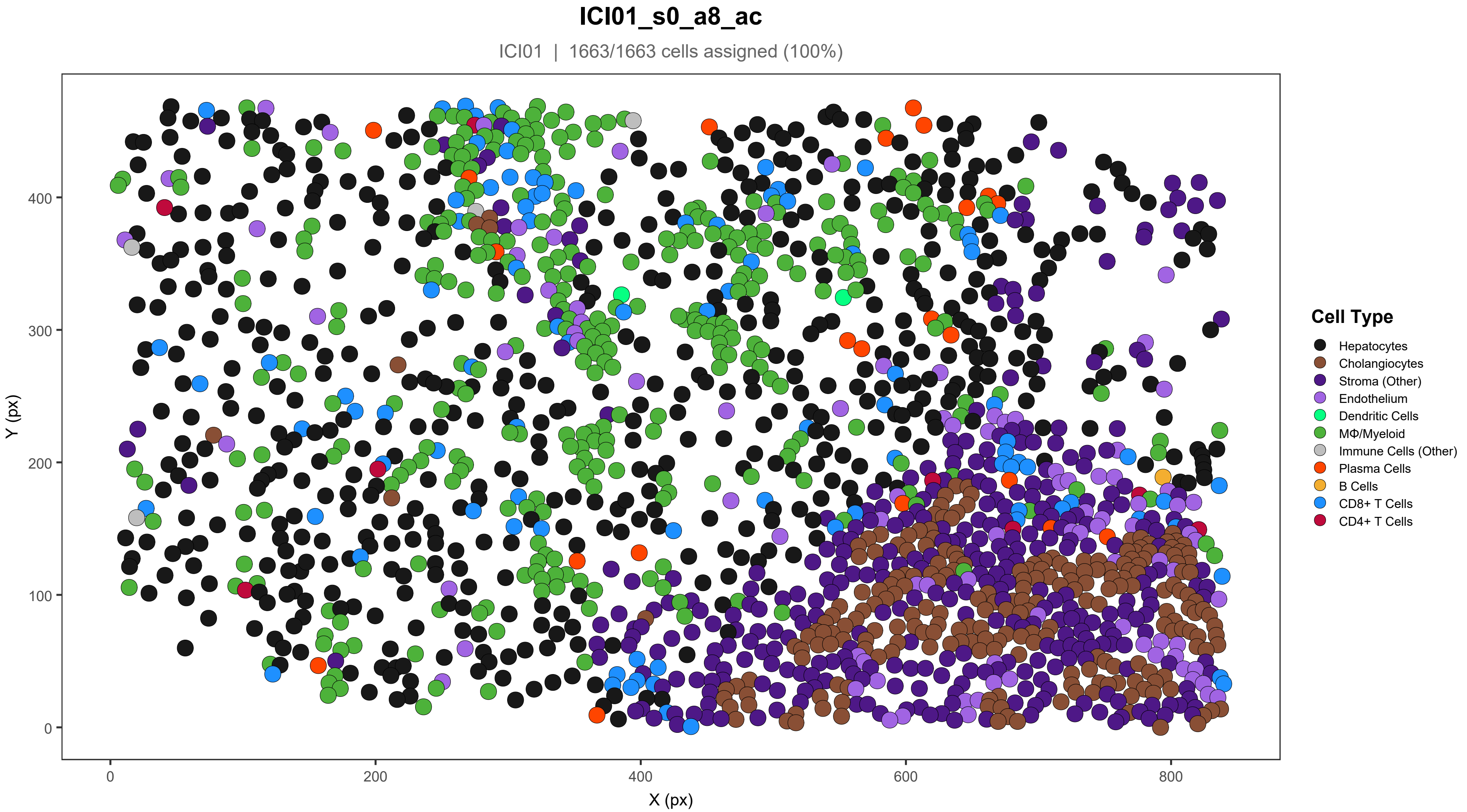
ICI01_s0_a8_ac
ICI01 | 1663/1663 cells assigned (100%)
Y (px)
X (px)
0
100
200
300
400
0
200
400
600
800
Cell Type
Hepatocytes
Cholangiocytes
Stroma (Other)
Endothelium
Dendritic Cells
MΦ/Myeloid
Immune Cells (Other)
Plasma Cells
B Cells
CD8+ T Cells
CD4+ T Cells

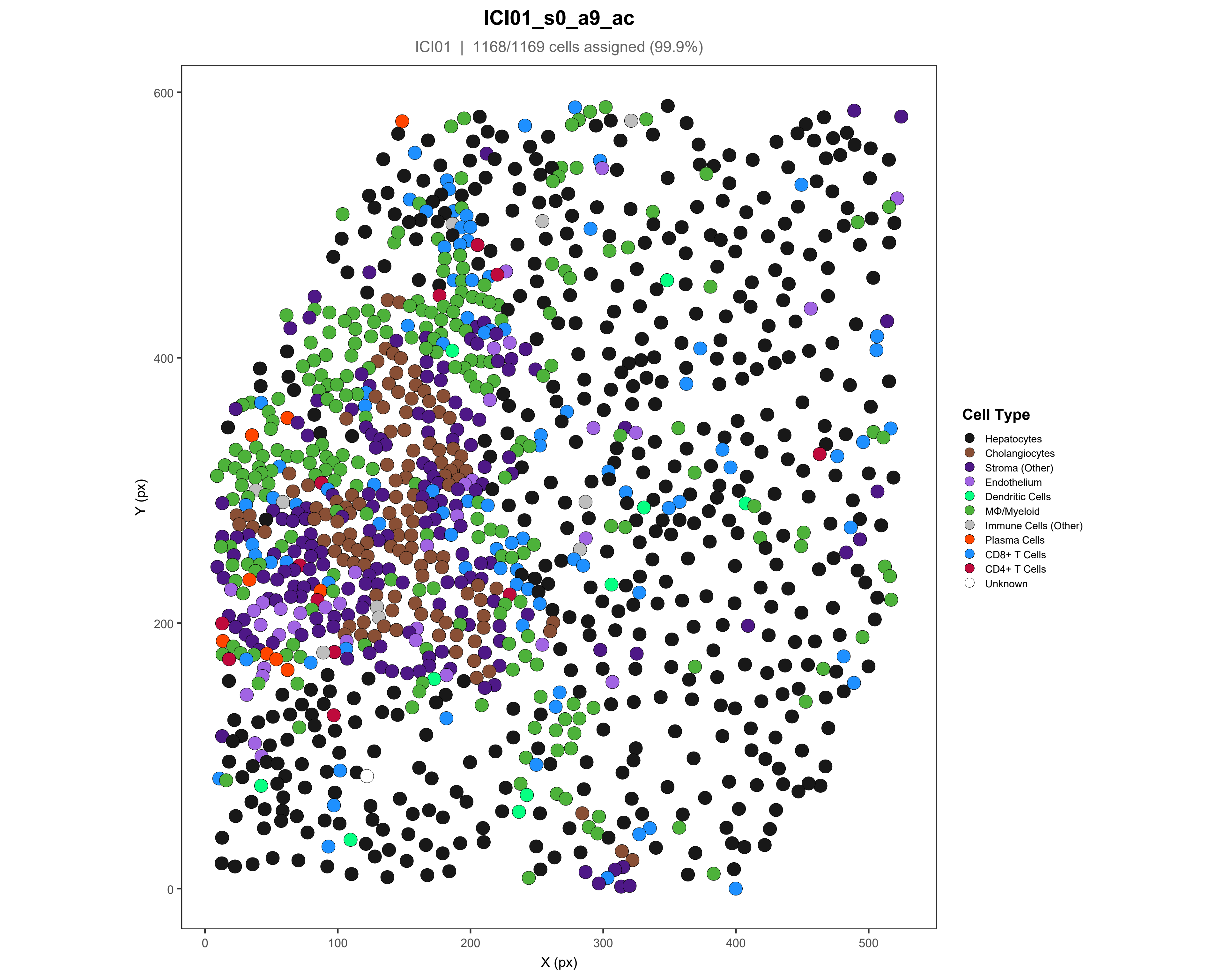

ICI01_s0_a9_ac
ICI01 | 1168/1169 cells assigned (99.9%)
Y (px)
X (px)
0
200
400
600
0
100
200
300
400
500
Cell Type
Hepatocytes
Cholangiocytes
Stroma (Other)
Endothelium
Dendritic Cells
MΦ/Myeloid
Immune Cells (Other)
Plasma Cells
CD8+ T Cells
CD4+ T Cells
Unknown

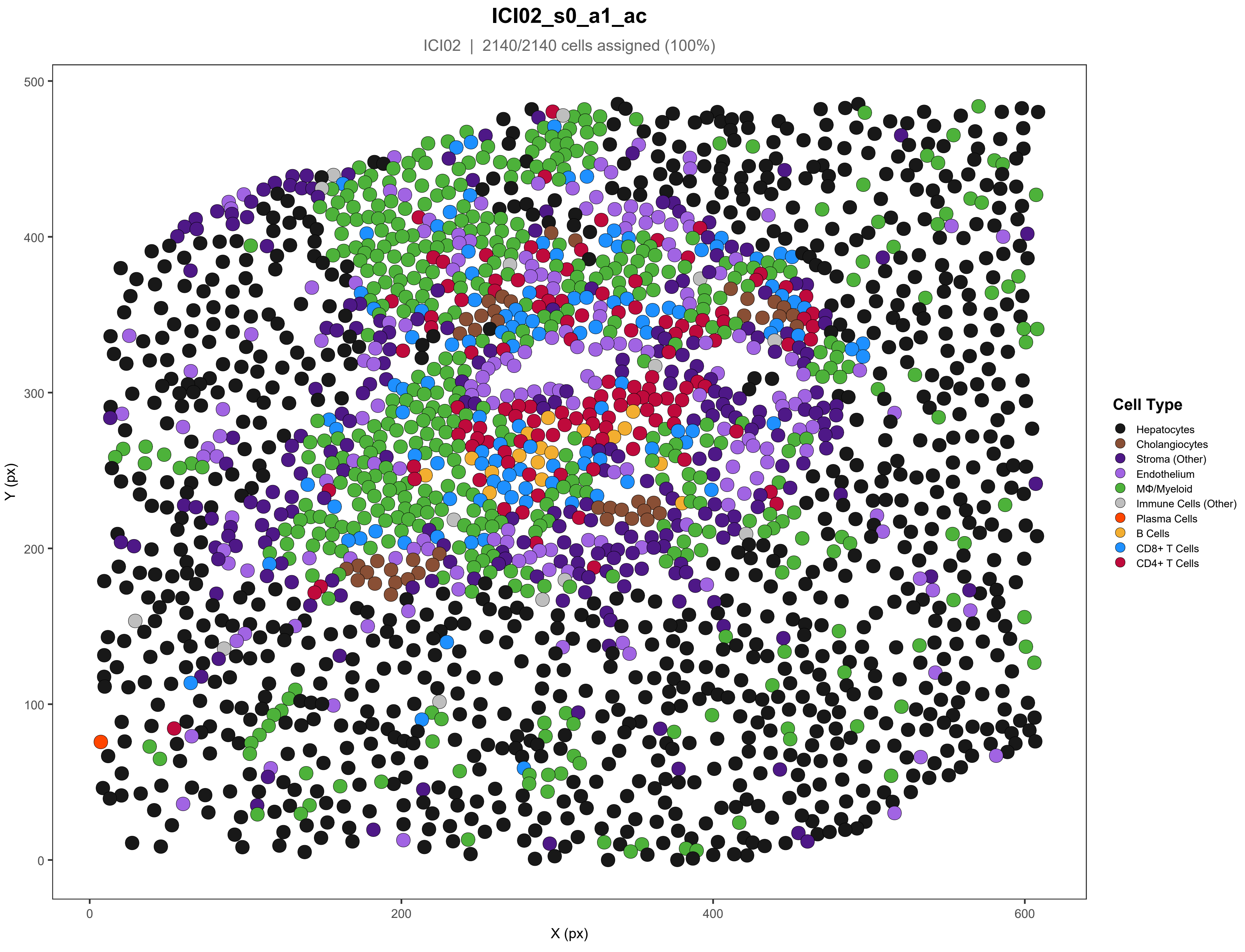

ICI02_s0_a1_ac
ICI02 | 2140/2140 cells assigned (100%)
Y (px)
X (px)
0
100
200
300
400
500
0
200
400
600
Cell Type
Hepatocytes
Cholangiocytes
Stroma (Other)
Endothelium
MΦ/Myeloid
Immune Cells (Other)
Plasma Cells
B Cells
CD8+ T Cells
CD4+ T Cells

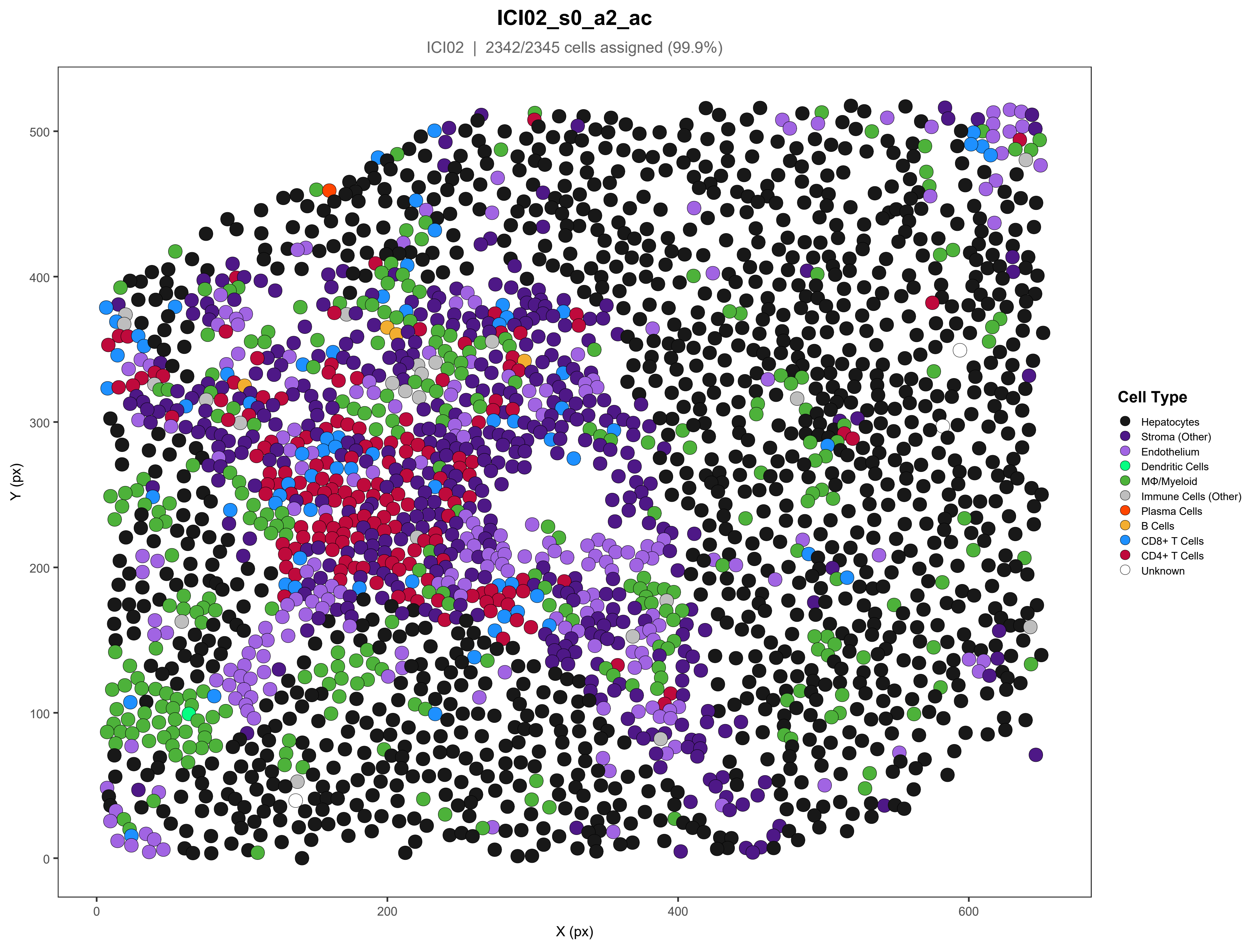
ICI02_s0_a2_ac
ICI02 | 2342/2345 cells assigned (99.9%)
Y (px)
X (px)
0
100
200
300
400
500
0
200
400
600
Cell Type
Hepatocytes
Stroma (Other)
Endothelium
Dendritic Cells
MΦ/Myeloid
Immune Cells (Other)
Plasma Cells
B Cells
CD8+ T Cells
CD4+ T Cells
Unknown

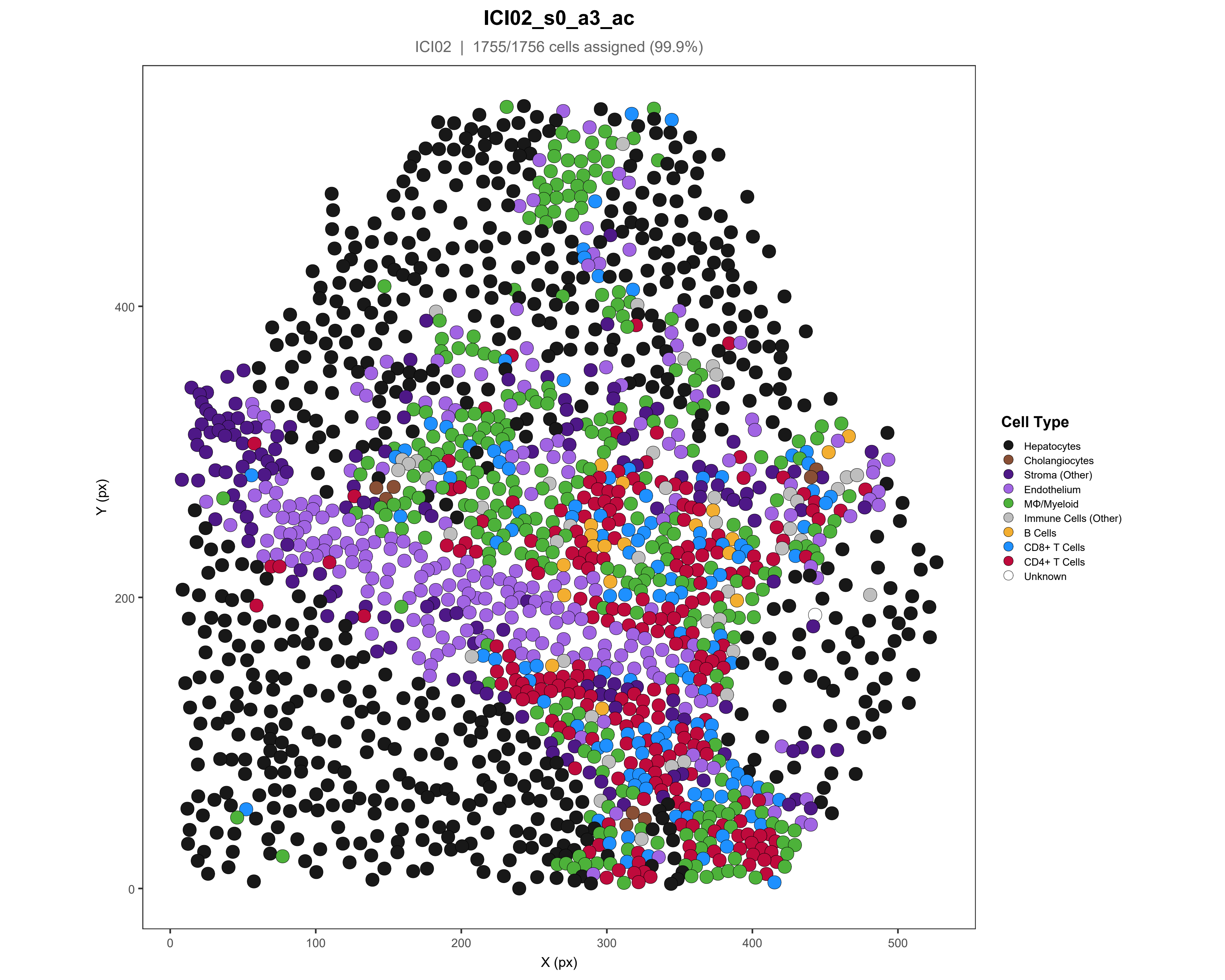
ICI02_s0_a3_ac
ICI02 | 1755/1756 cells assigned (99.9%)
Y (px)
X (px)
0
100
200
300
400
500
0
200
400
Cell Type
Hepatocytes
Cholangiocytes
Stroma (Other)
Endothelium
MΦ/Myeloid
Immune Cells (Other)
B Cells
CD8+ T Cells
CD4+ T Cells
Unknown

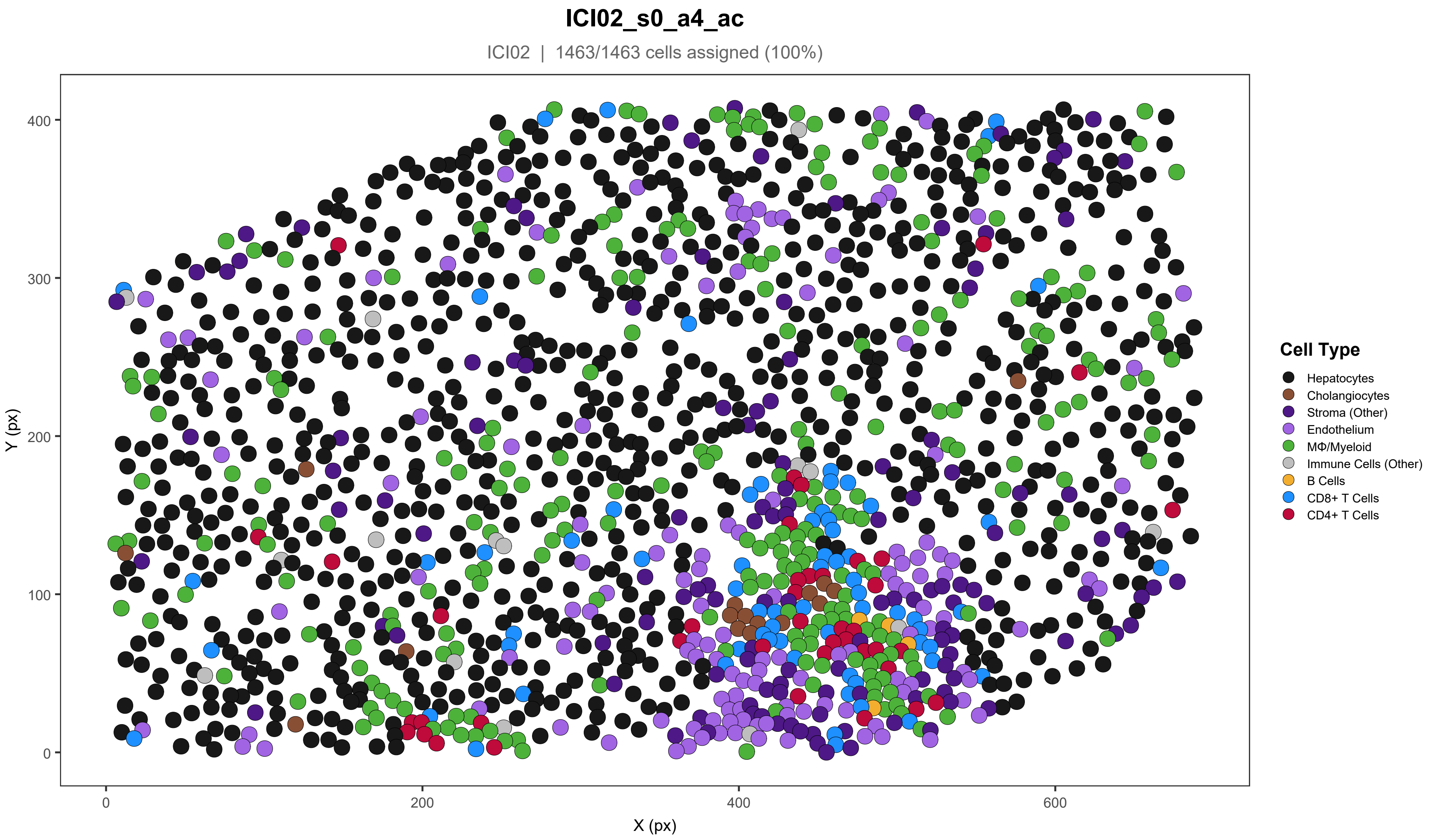

ICI02_s0_a4_ac
ICI02 | 1463/1463 cells assigned (100%)
Y (px)
X (px)
0
100
200
300
400
0
200
400
600
Cell Type
Hepatocytes
Cholangiocytes
Stroma (Other)
Endothelium
MΦ/Myeloid
Immune Cells (Other)
B Cells
CD8+ T Cells
CD4+ T Cells

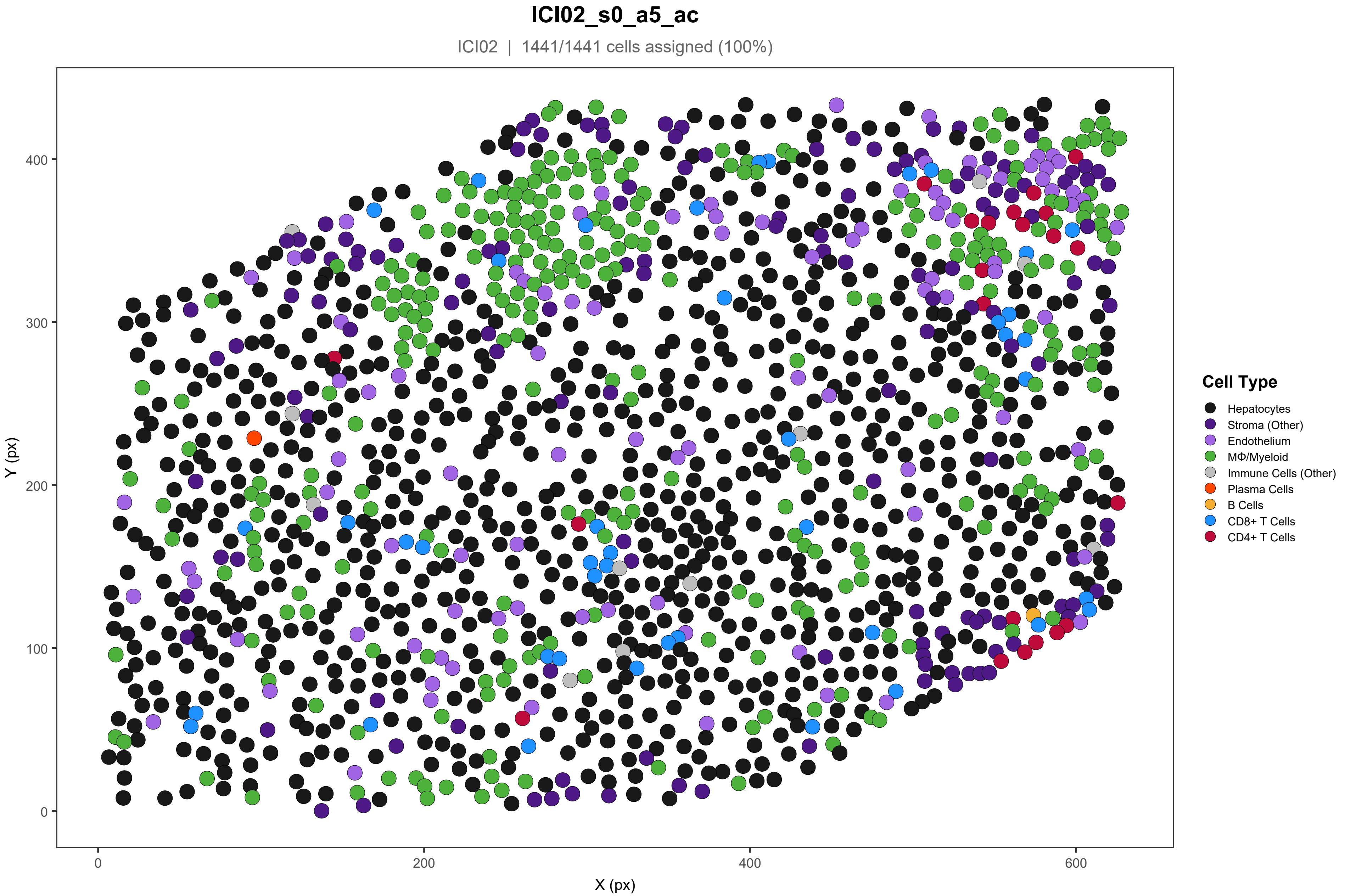
ICI02_s0_a5_ac
ICI02 | 1441/1441 cells assigned (100%)
Y (px)
X (px)
0
100
200
300
400
0
200
400
600
Cell Type
Hepatocytes
Stroma (Other)
Endothelium
MΦ/Myeloid
Immune Cells (Other)
Plasma Cells
B Cells
CD8+ T Cells
CD4+ T Cells

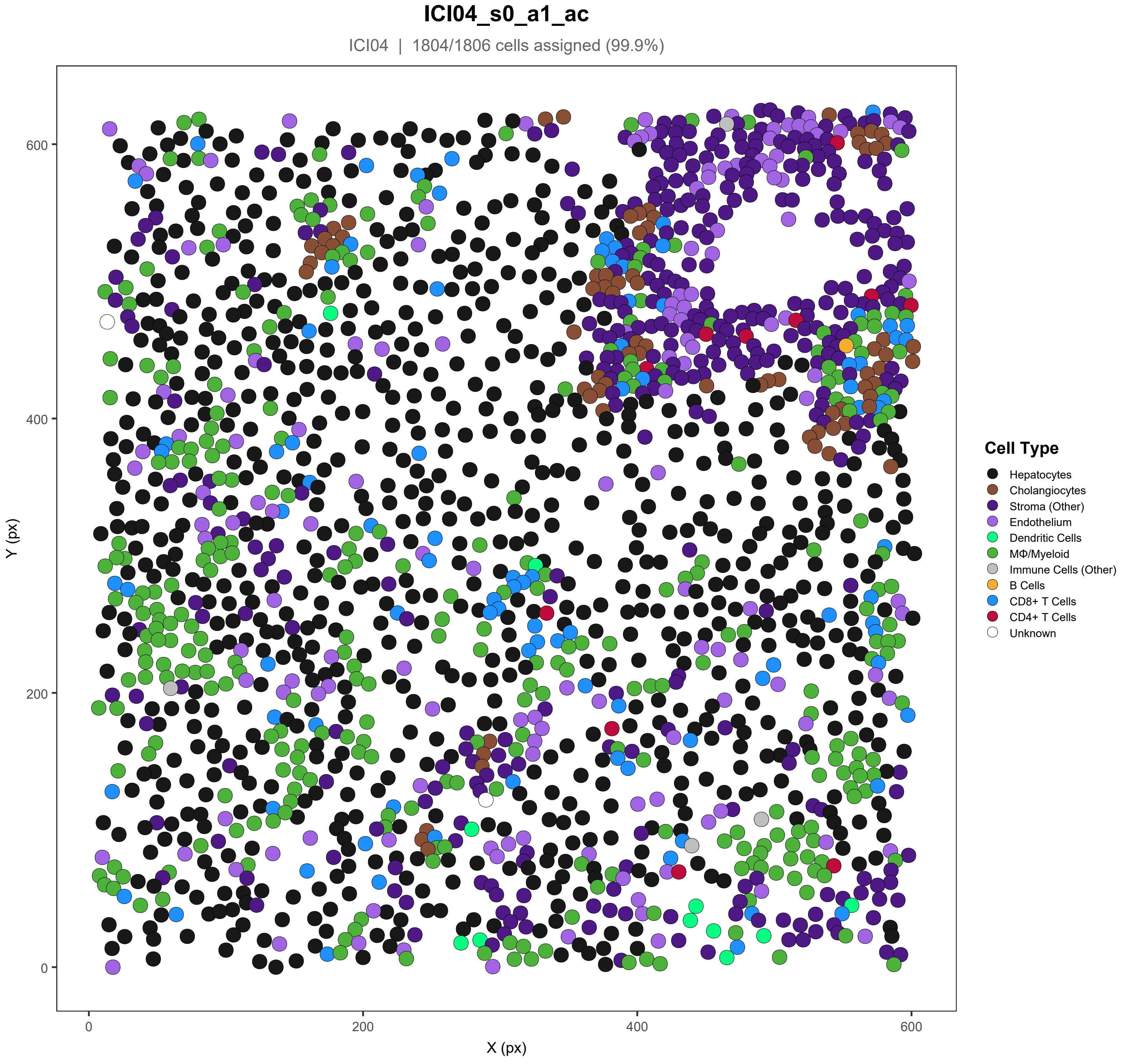

ICI04_s0_a1_ac
ICI04 | 1804/1806 cells assigned (99.9%)
Y (px)
X (px)
0
200
400
600
Cell Type
Hepatocytes
Cholangiocytes
Stroma (Other)
Endothelium
Dendritic Cells
MΦ/Myeloid
Immune Cells (Other)
B Cells
CD8+ T Cells
CD4+ T Cells
Unknown

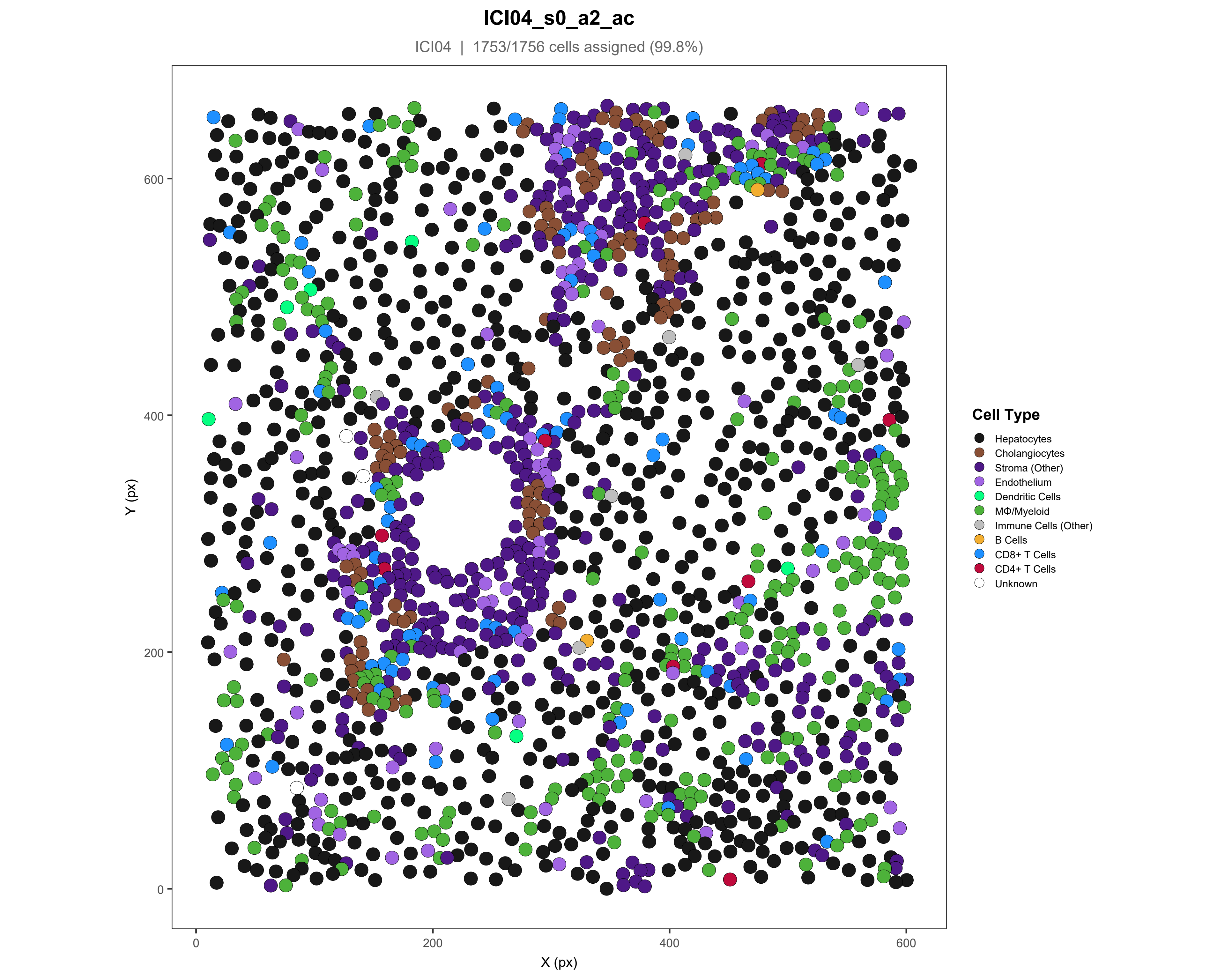
ICI04_s0_a2_ac
ICI04 | 1753/1756 cells assigned (99.8%)
Y (px)
X (px)
0
200
400
600
Cell Type
Hepatocytes
Cholangiocytes
Stroma (Other)
Endothelium
Dendritic Cells
MΦ/Myeloid
Immune Cells (Other)
B Cells
CD8+ T Cells
CD4+ T Cells
Unknown

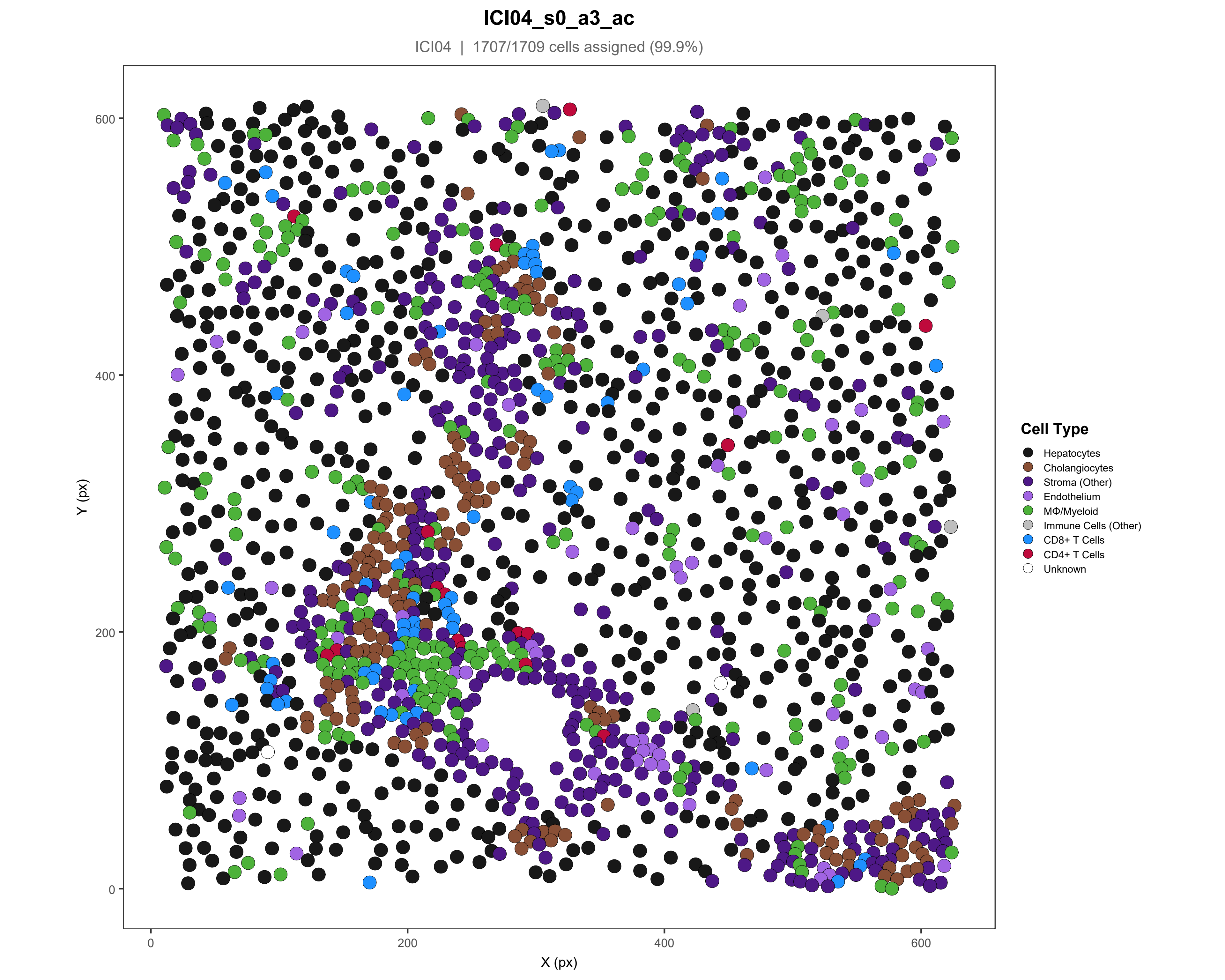

ICI04_s0_a3_ac
ICI04 | 1707/1709 cells assigned (99.9%)
Y (px)
X (px)
0
200
400
600
Cell Type
Hepatocytes
Cholangiocytes
Stroma (Other)
Endothelium
MΦ/Myeloid
Immune Cells (Other)
CD8+ T Cells
CD4+ T Cells
Unknown

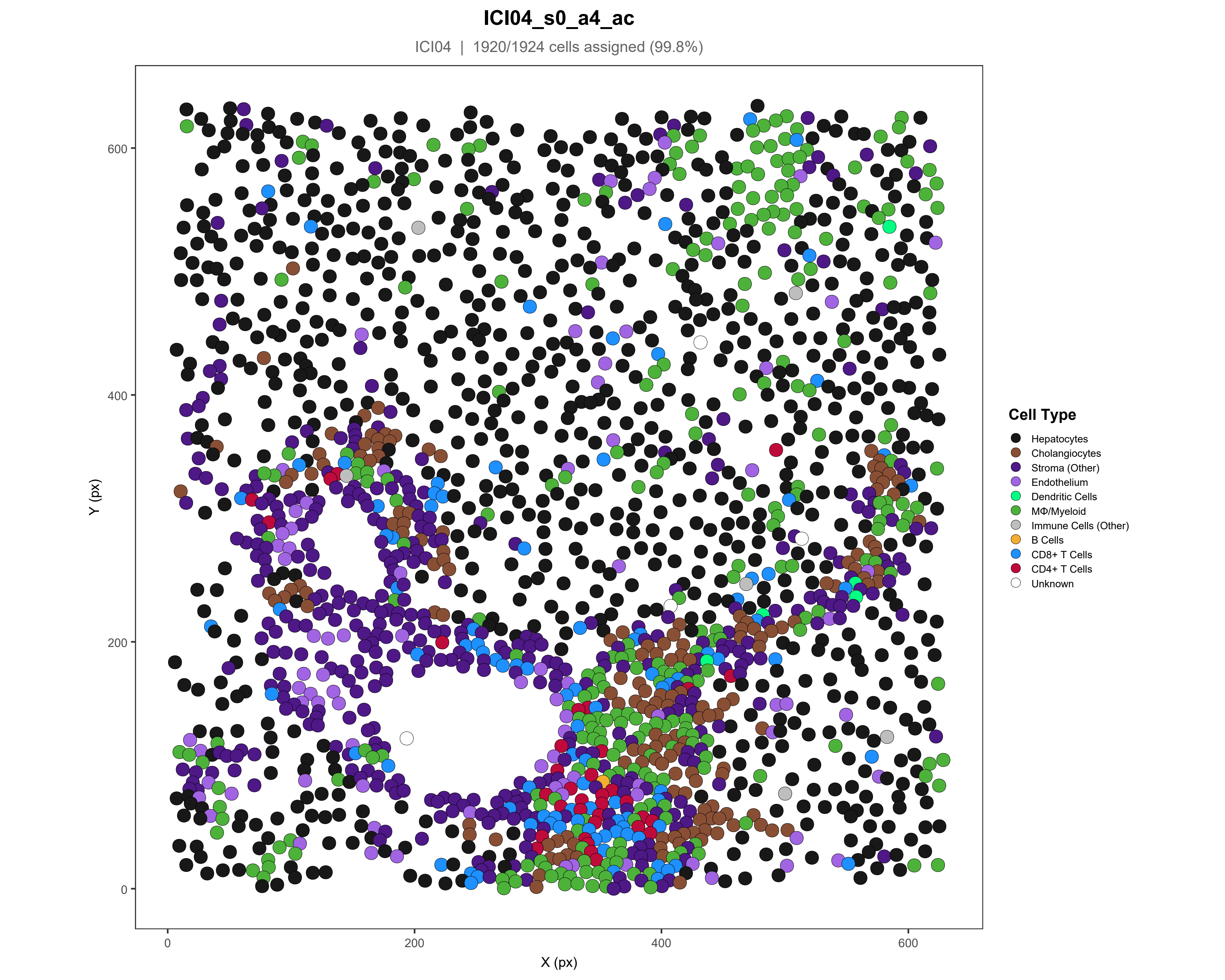
ICI04_s0_a4_ac
ICI04 | 1920/1924 cells assigned (99.8%)
Y (px)
X (px)
0
200
400
600
Cell Type
Hepatocytes
Cholangiocytes
Stroma (Other)
Endothelium
Dendritic Cells
MΦ/Myeloid
Immune Cells (Other)
B Cells
CD8+ T Cells
CD4+ T Cells
Unknown

### ICI04_s0_a5_ac

ICI04 | 1920/1920 cells assigned (100%)

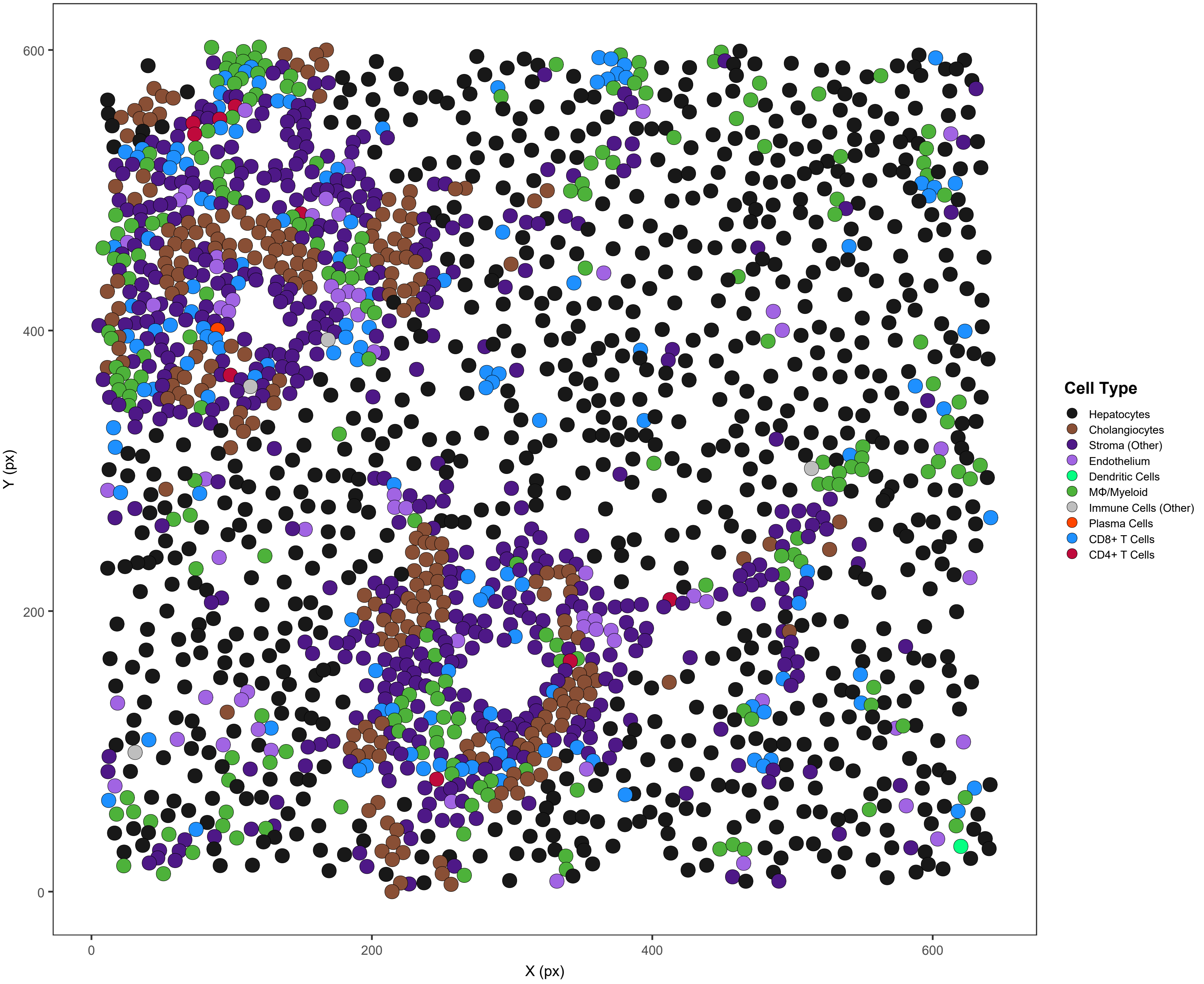

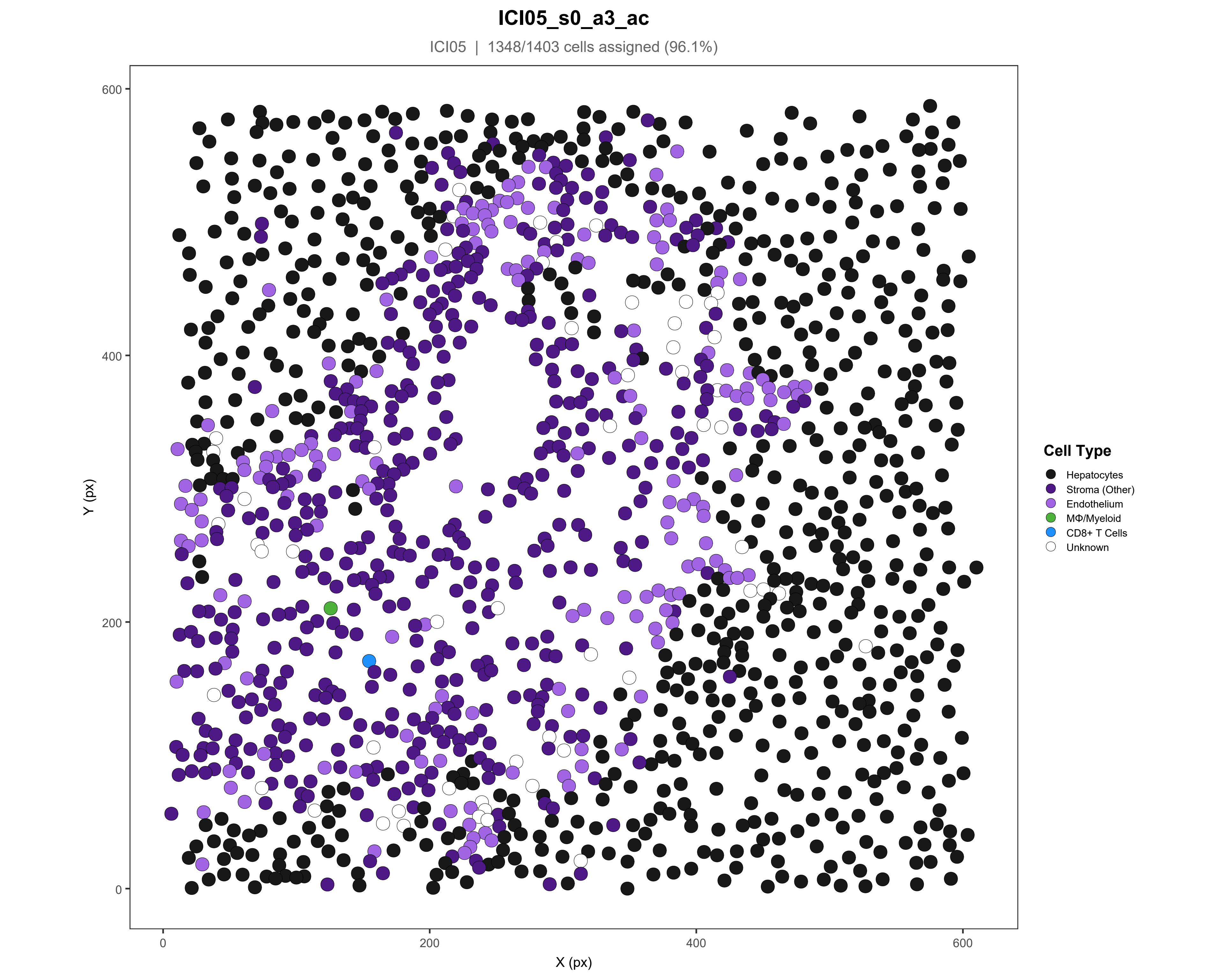
ICI05_s0_a3_ac
ICI05 | 1348/1403 cells assigned (96.1%)
Y (px)
X (px)
0
200
400
600
Cell Type
Hepatocytes
Stroma (Other)
Endothelium
MΦ/Myeloid
CD8+ T Cells
Unknown

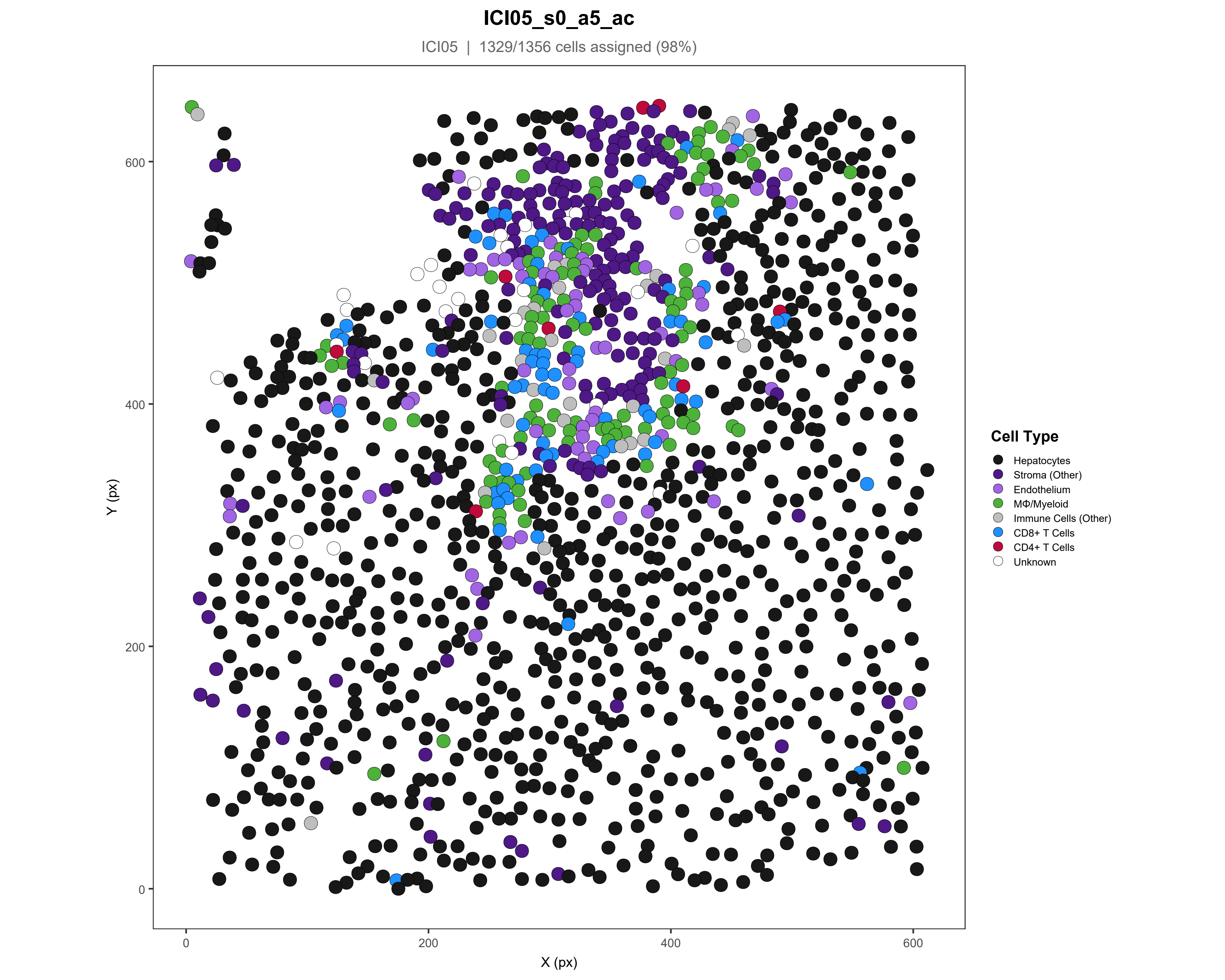
ICI05_s0_a5_ac
ICI05 | 1329/1356 cells assigned (98%)
Y (px)
600
400
200
0
X (px)
0
200
400
600
Cell Type
Hepatocytes
Stroma (Other)
Endothelium
MΦ/Myeloid
Immune Cells (Other)
CD8+ T Cells
CD4+ T Cells
Unknown

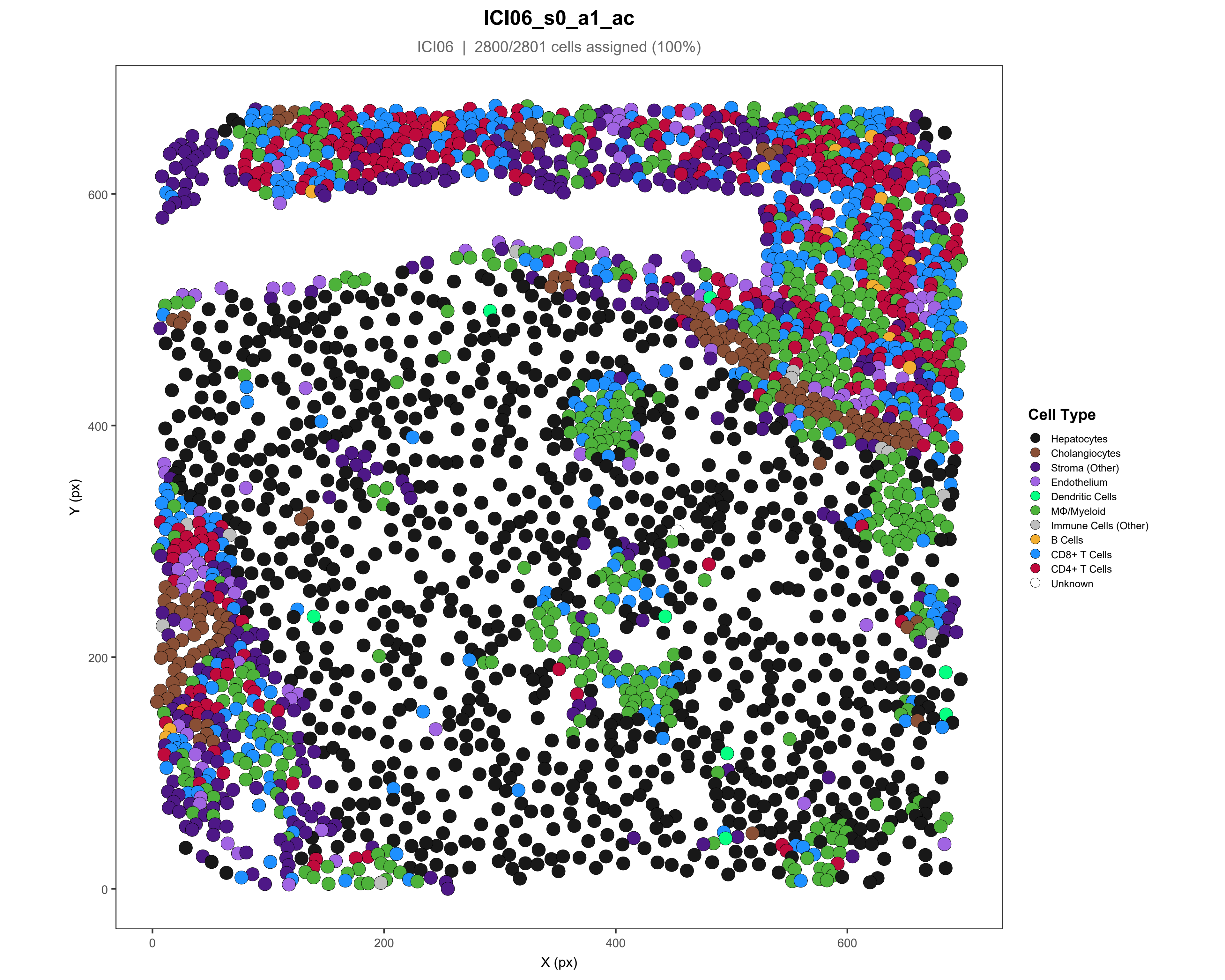
ICI06_s0_a1_ac
ICI06 | 2800/2801 cells assigned (100%)
Y (px)
X (px)
0
200
400
600
Cell Type
Hepatocytes
Cholangiocytes
Stroma (Other)
Endothelium
Dendritic Cells
MΦ/Myeloid
Immune Cells (Other)
B Cells
CD8+ T Cells
CD4+ T Cells
Unknown

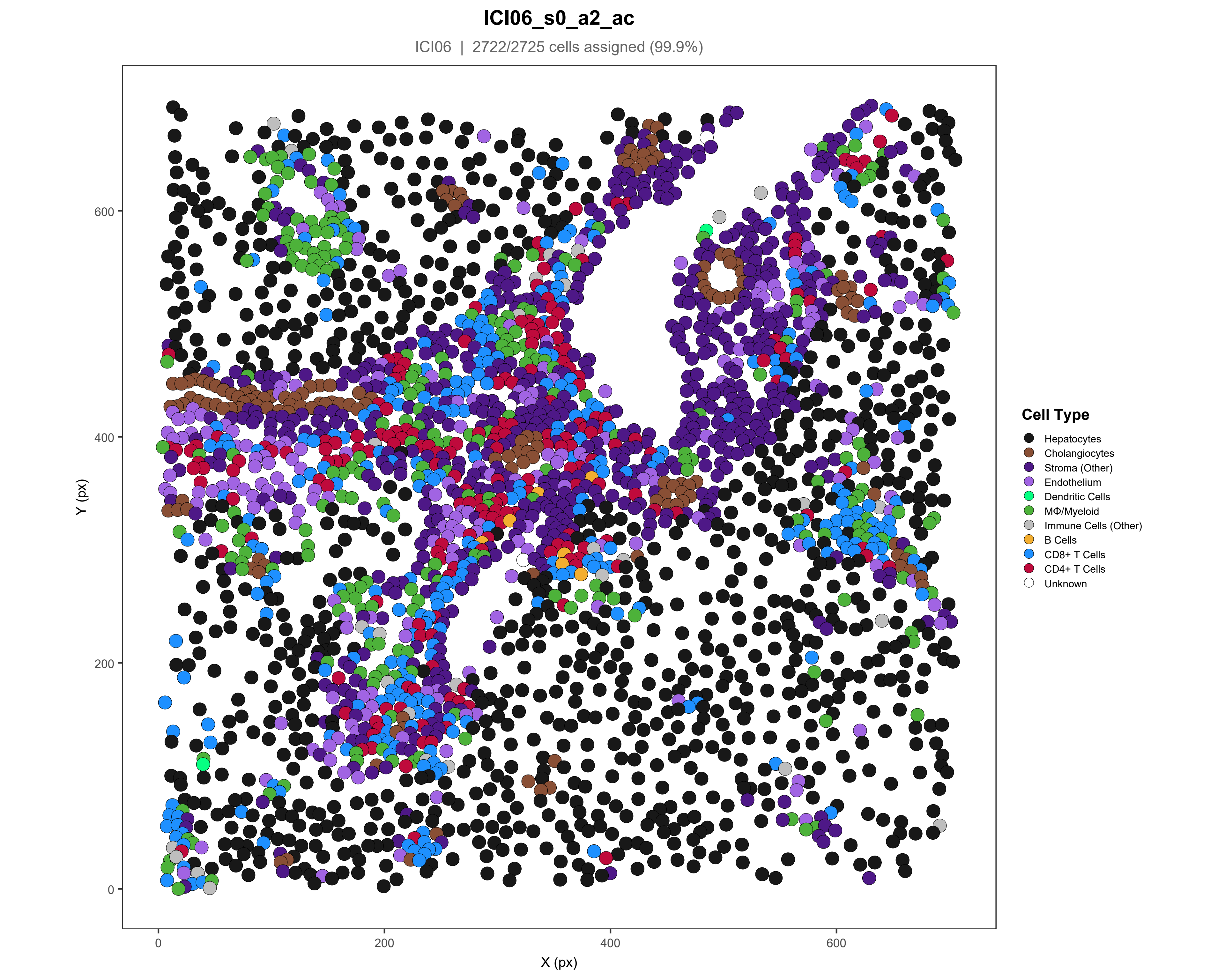

ICI06_s0_a2_ac
ICI06 | 2722/2725 cells assigned (99.9%)
Y (px)
X (px)
0
200
400
600
Cell Type
Hepatocytes
Cholangiocytes
Stroma (Other)
Endothelium
Dendritic Cells
MΦ/Myeloid
Immune Cells (Other)
B Cells
CD8+ T Cells
CD4+ T Cells
Unknown

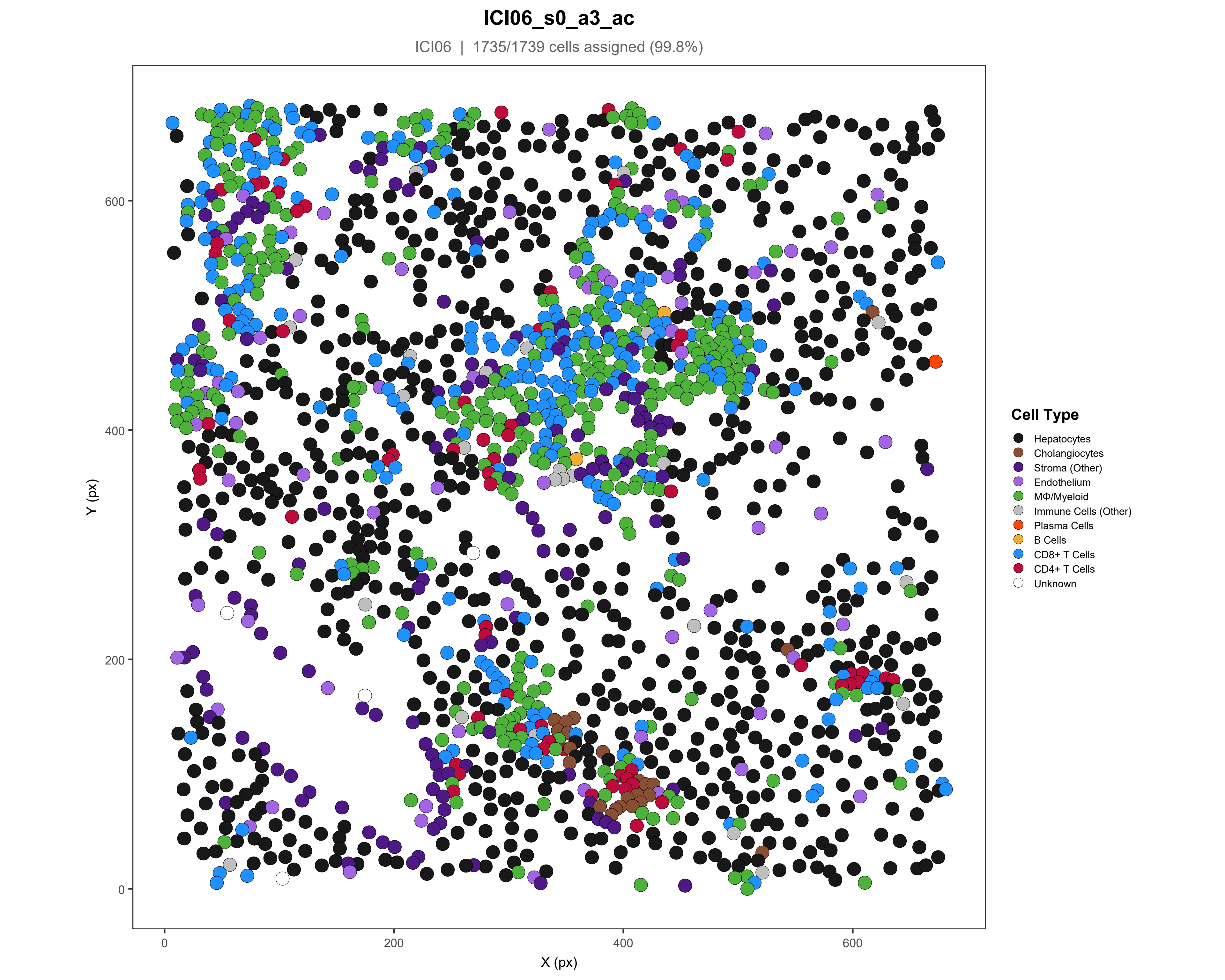

ICI06_s0_a3_ac
ICI06 | 1735/1739 cells assigned (99.8%)
Cell Type
Hepatocytes
Cholangiocytes
Stroma (Other)
Endothelium
MΦ/Myeloid
Immune Cells (Other)
Plasma Cells
B Cells
CD8+ T Cells
CD4+ T Cells
Unknown
Y (px)
X (px)
0
200
400
600

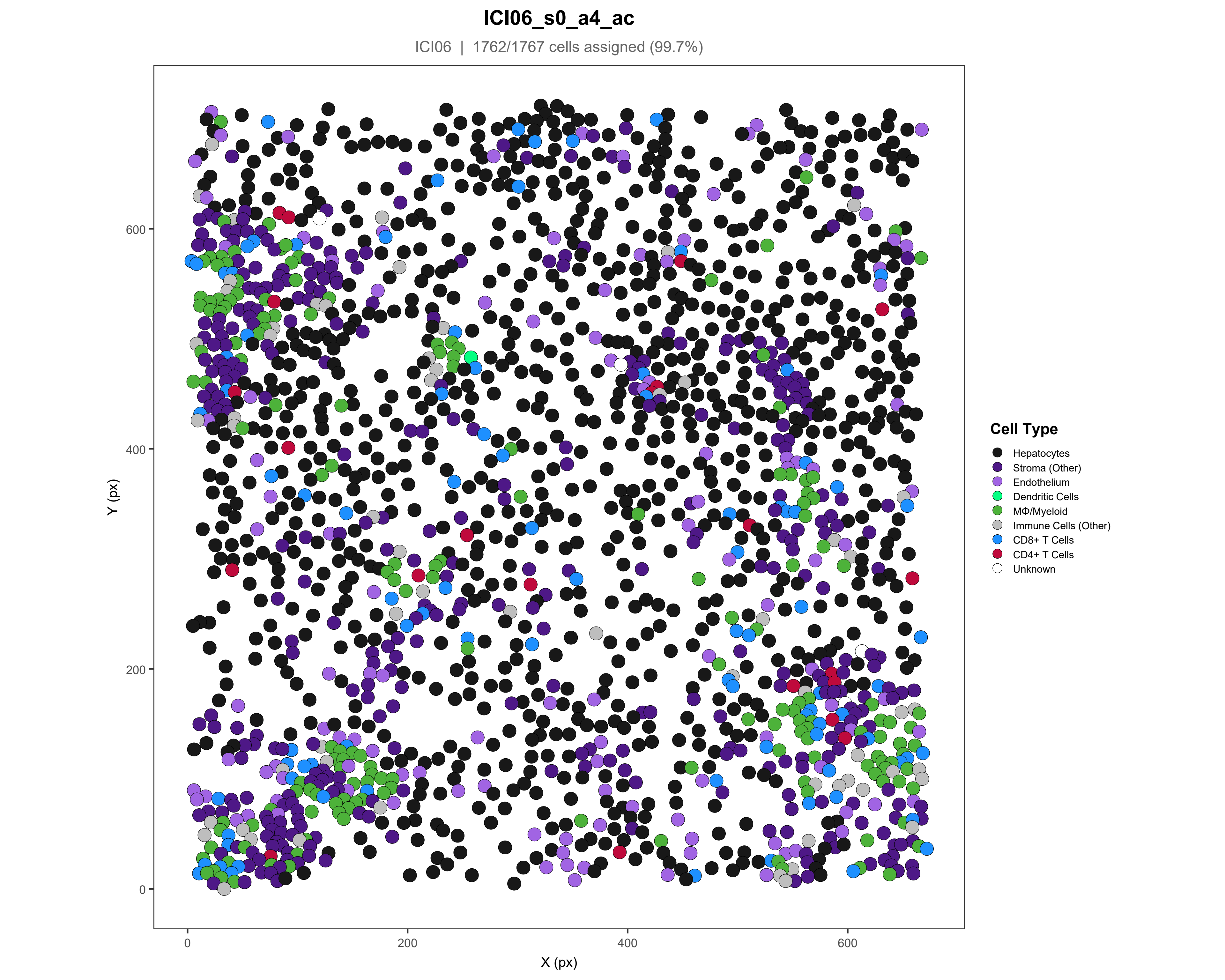
ICI06_s0_a4_ac
ICI06 | 1762/1767 cells assigned (99.7%)
Y (px)
X (px)
0
200
400
600
Cell Type
Hepatocytes
Stroma (Other)
Endothelium
Dendritic Cells
MΦ/Myeloid
Immune Cells (Other)
CD8+ T Cells
CD4+ T Cells
Unknown

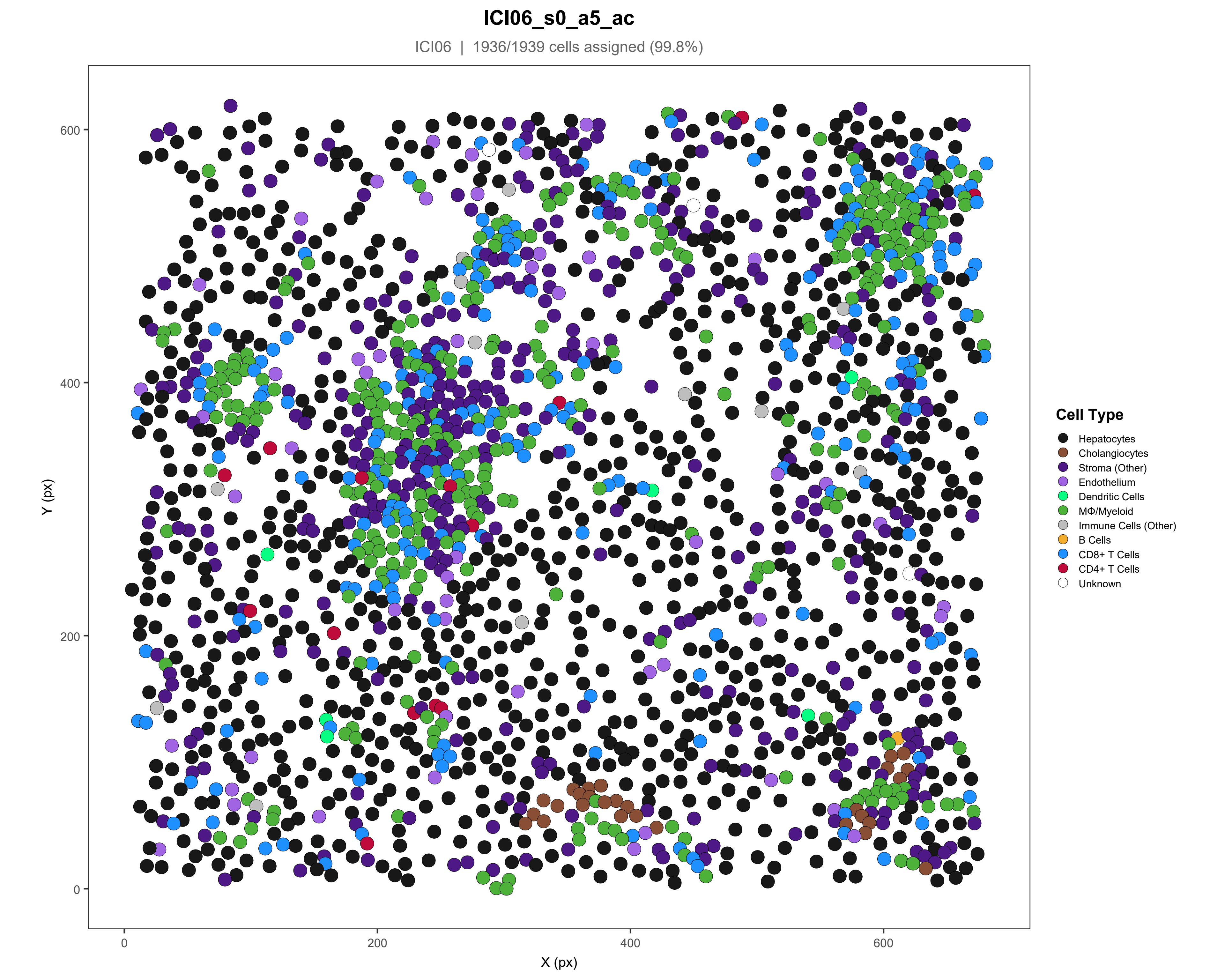
ICI06_s0_a5_ac
ICI06 | 1936/1939 cells assigned (99.8%)
Y (px)
X (px)
0
200
400
600
Cell Type
Hepatocytes
Cholangiocytes
Stroma (Other)
Endothelium
Dendritic Cells
MΦ/Myeloid
Immune Cells (Other)
B Cells
CD8+ T Cells
CD4+ T Cells
Unknown

### ICI07_s0_a2_ac

ICI07 | 1871/1876 cells assigned (99.7%)

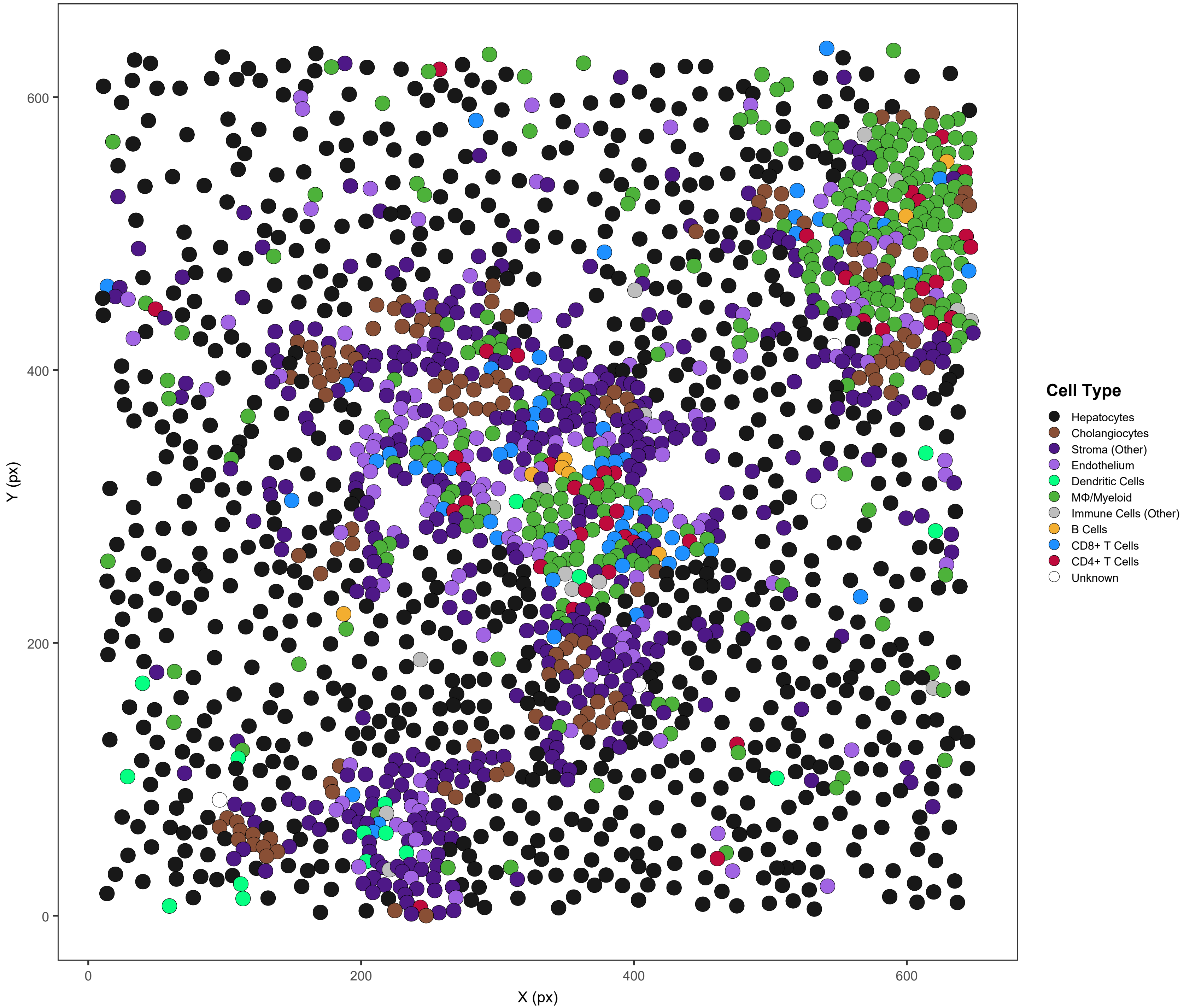

### ICI08_s0_a1_ac

ICI08 | 1190/1190 cells assigned (100%)

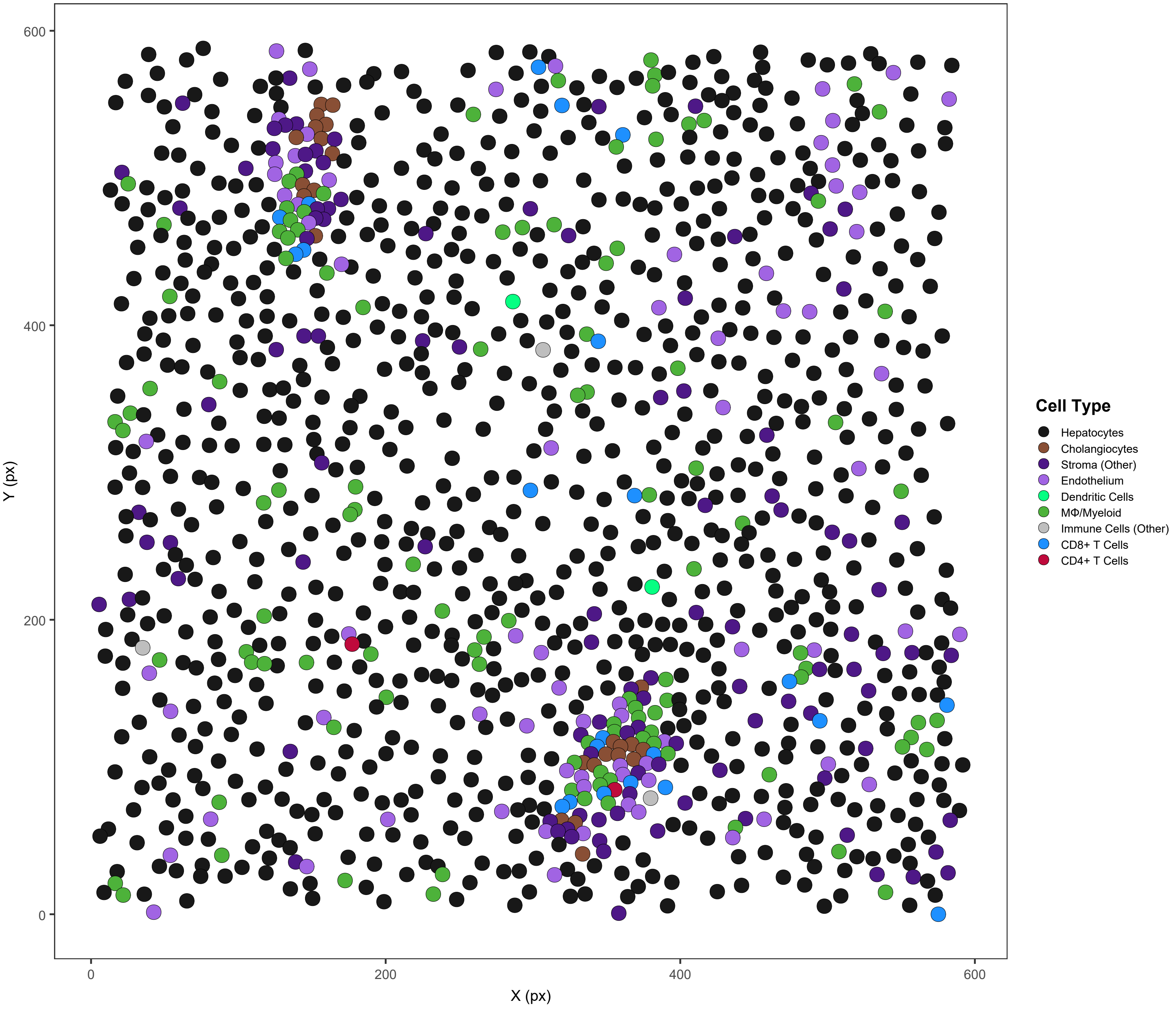

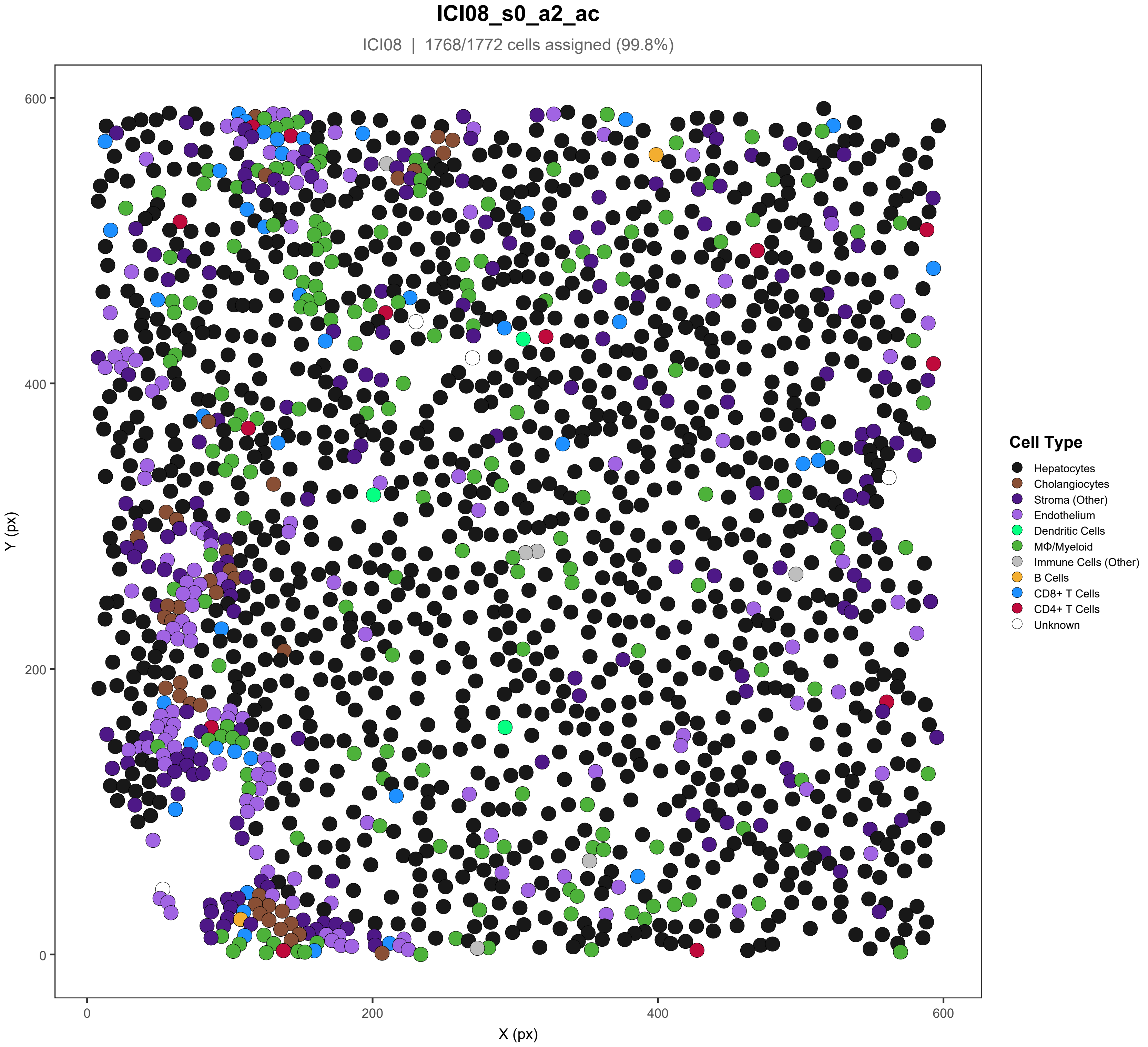
ICI08_s0_a2_ac
ICI08 | 1768/1772 cells assigned (99.8%)
Y (px)
X (px)
0
200
400
600
Cell Type
Hepatocytes
Cholangiocytes
Stroma (Other)
Endothelium
Dendritic Cells
MΦ/Myeloid
Immune Cells (Other)
B Cells
CD8+ T Cells
CD4+ T Cells
Unknown

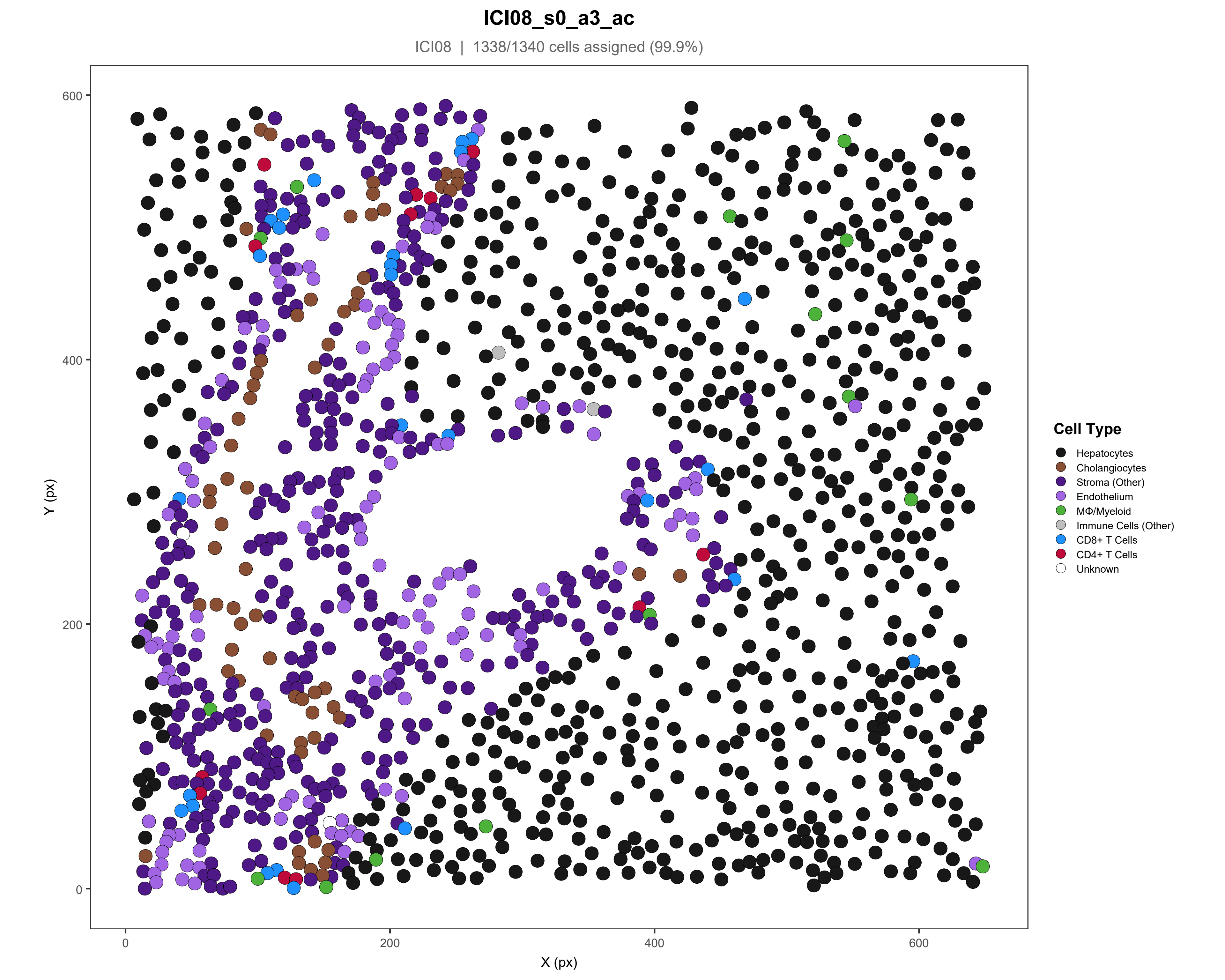

ICI08_s0_a3_ac
ICI08 | 1338/1340 cells assigned (99.9%)
Y (px)
X (px)
0
200
400
600
Cell Type
Hepatocytes
Cholangiocytes
Stroma (Other)
Endothelium
MΦ/Myeloid
Immune Cells (Other)
CD8+ T Cells
CD4+ T Cells
Unknown

### ICI08_s0_a5_ac

ICI08 | 1211/1215 cells assigned (99.7%)

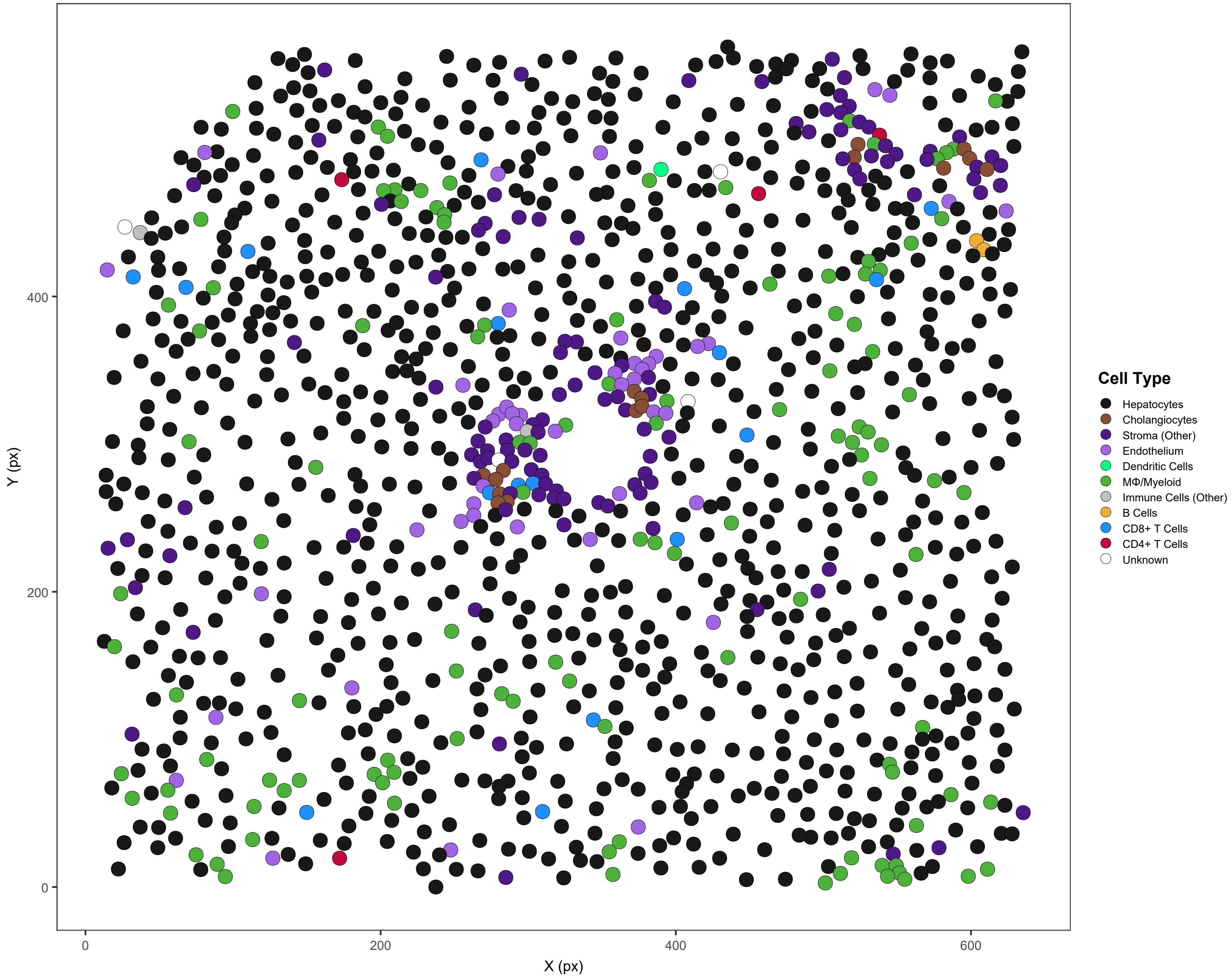

#### ICI09_s0_a1_ac

ICI09 | 1921/1921 cells assigned (100%)

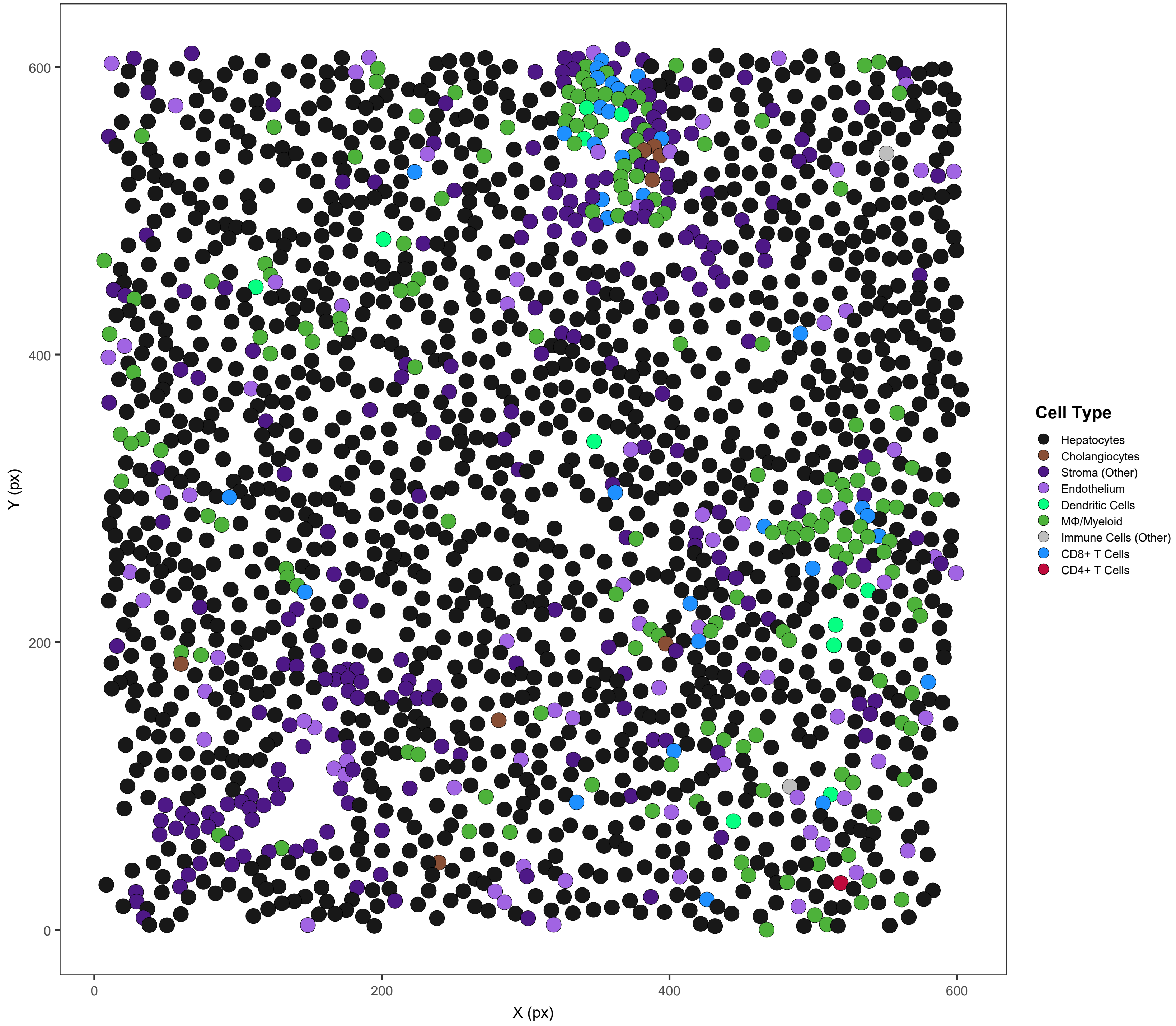

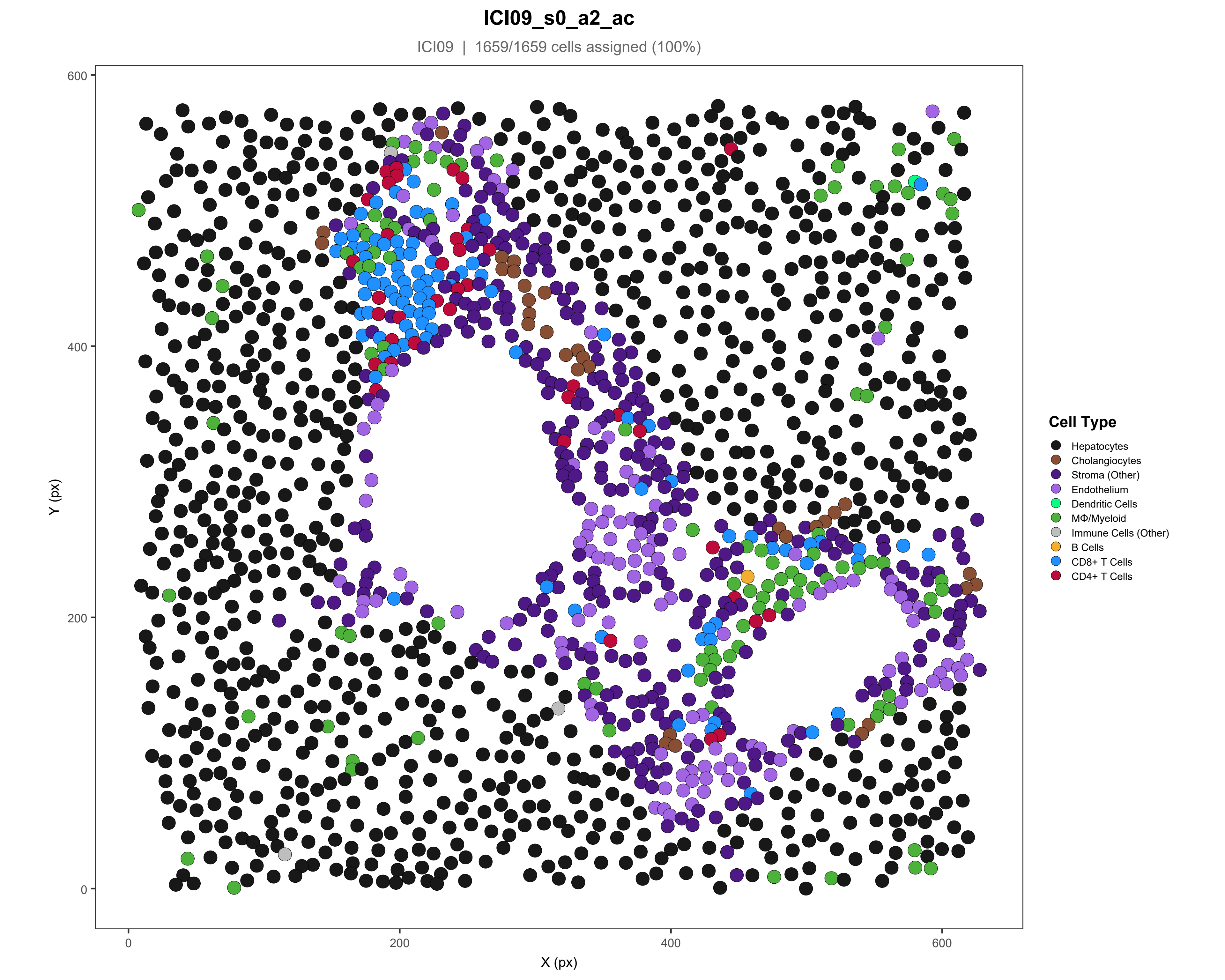
ICI09_s0_a2_ac
ICI09 | 1659/1659 cells assigned (100%)
Y (px)
X (px)
0
200
400
600
Cell Type
Hepatocytes
Cholangiocytes
Stroma (Other)
Endothelium
Dendritic Cells
MΦ/Myeloid
Immune Cells (Other)
B Cells
CD8+ T Cells
CD4+ T Cells

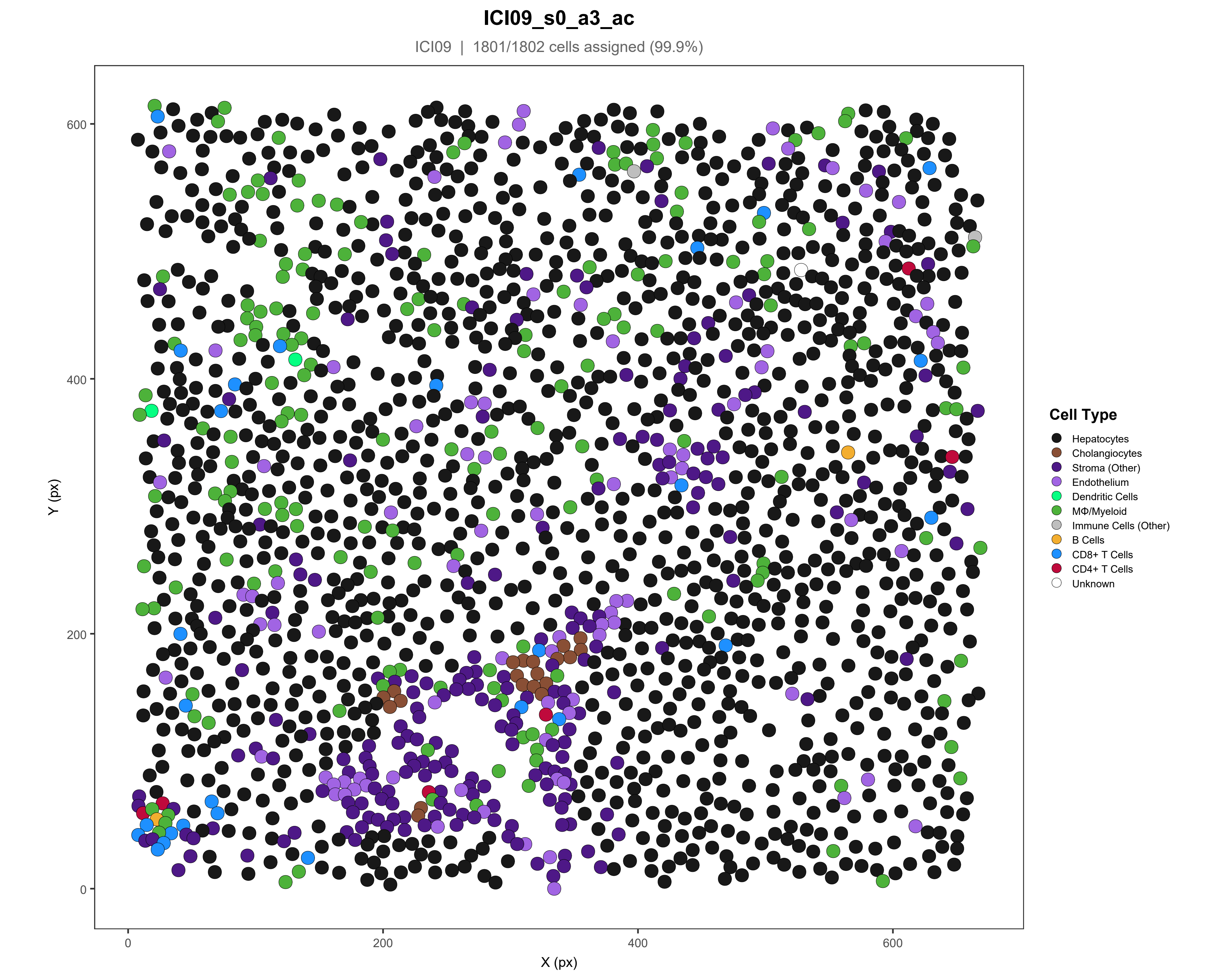
ICI09_s0_a3_ac
ICI09 | 1801/1802 cells assigned (99.9%)
Y (px)
X (px)
0
200
400
600
Cell Type
Hepatocytes
Cholangiocytes
Stroma (Other)
Endothelium
Dendritic Cells
MΦ/Myeloid
Immune Cells (Other)
B Cells
CD8+ T Cells
CD4+ T Cells
Unknown

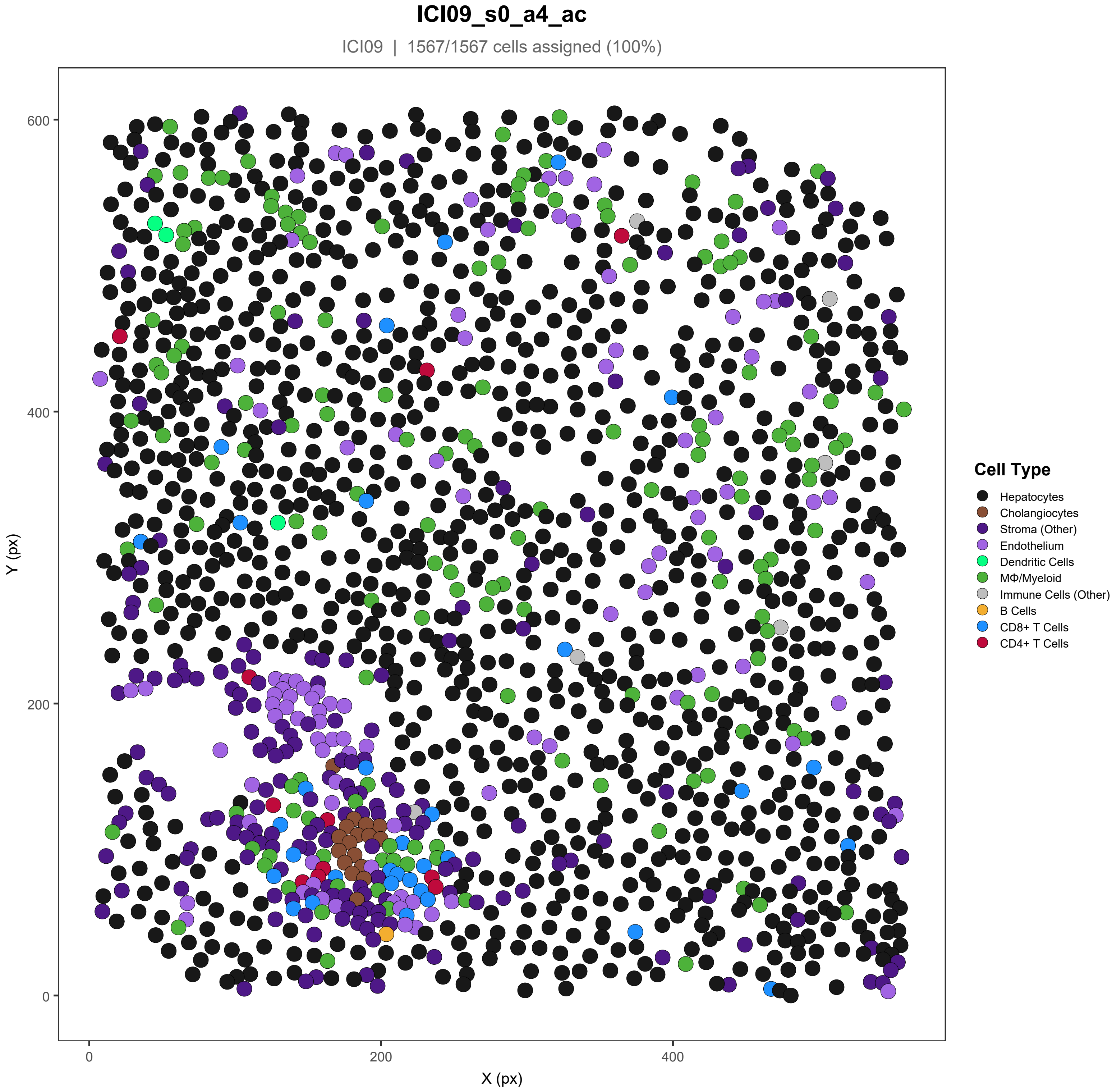
ICI09_s0_a4_ac
ICI09 | 1567/1567 cells assigned (100%)
Y (px)
X (px)
0
200
400
600
Cell Type
Hepatocytes
Cholangiocytes
Stroma (Other)
Endothelium
Dendritic Cells
MΦ/Myeloid
Immune Cells (Other)
B Cells
CD8+ T Cells
CD4+ T Cells

### ICI09_s0_a5_ac

ICI09 | 1680/1681 cells assigned (99.9%)

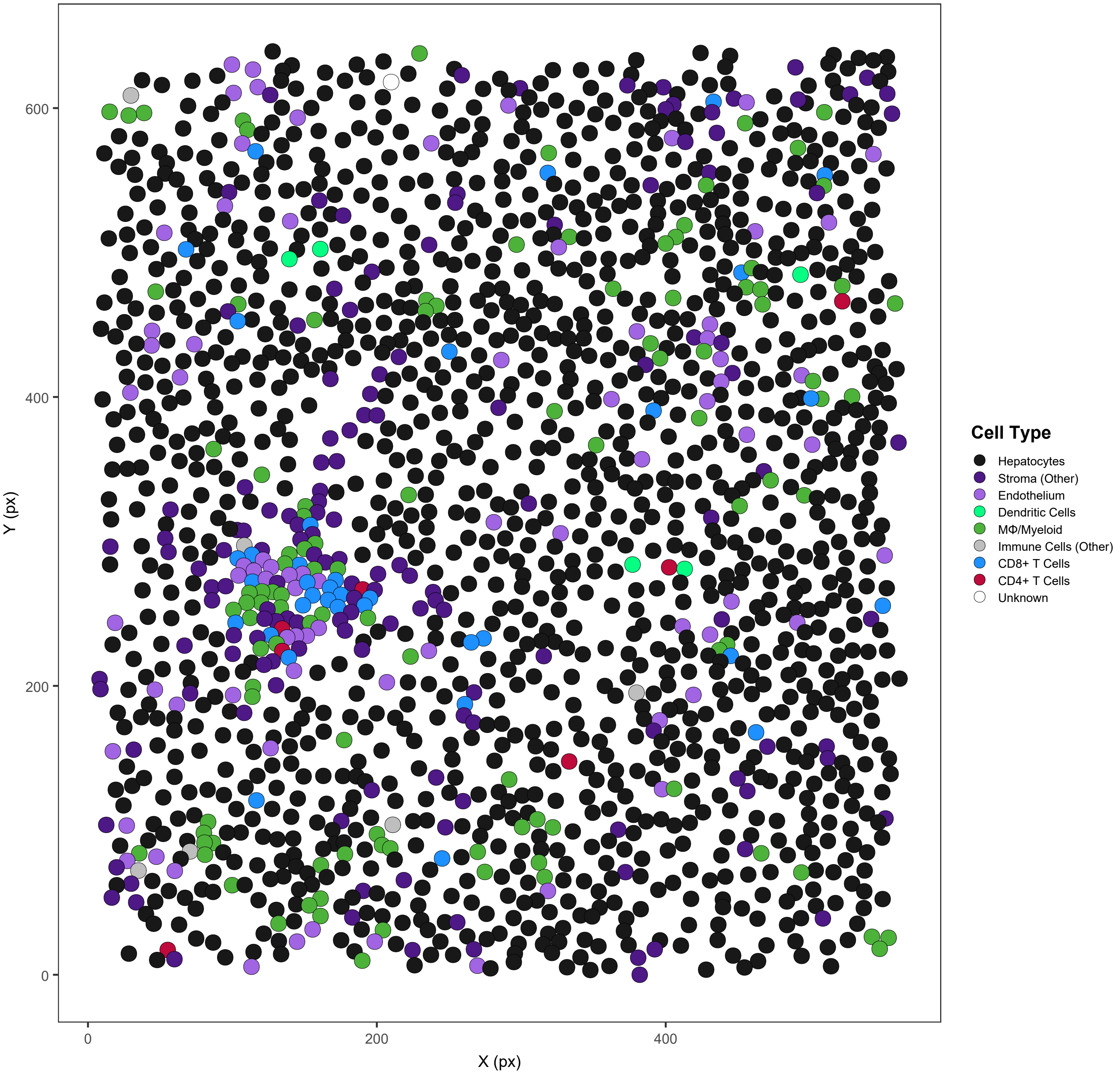

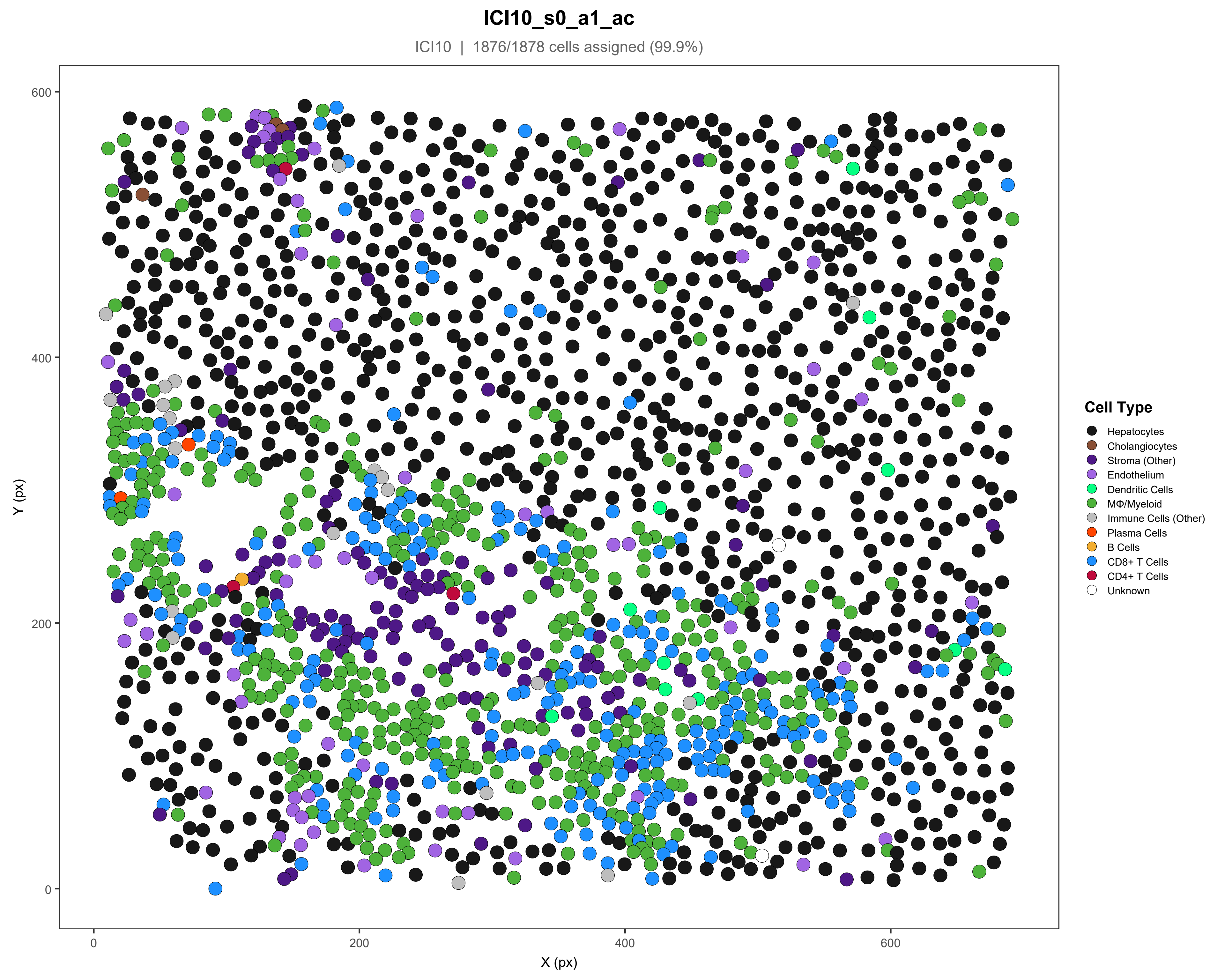
ICI10_s0_a1_ac
ICI10 | 1876/1878 cells assigned (99.9%)
Y (px)
X (px)
0
200
400
600
Cell Type
Hepatocytes
Cholangiocytes
Stroma (Other)
Endothelium
Dendritic Cells
MΦ/Myeloid
Immune Cells (Other)
Plasma Cells
B Cells
CD8+ T Cells
CD4+ T Cells
Unknown

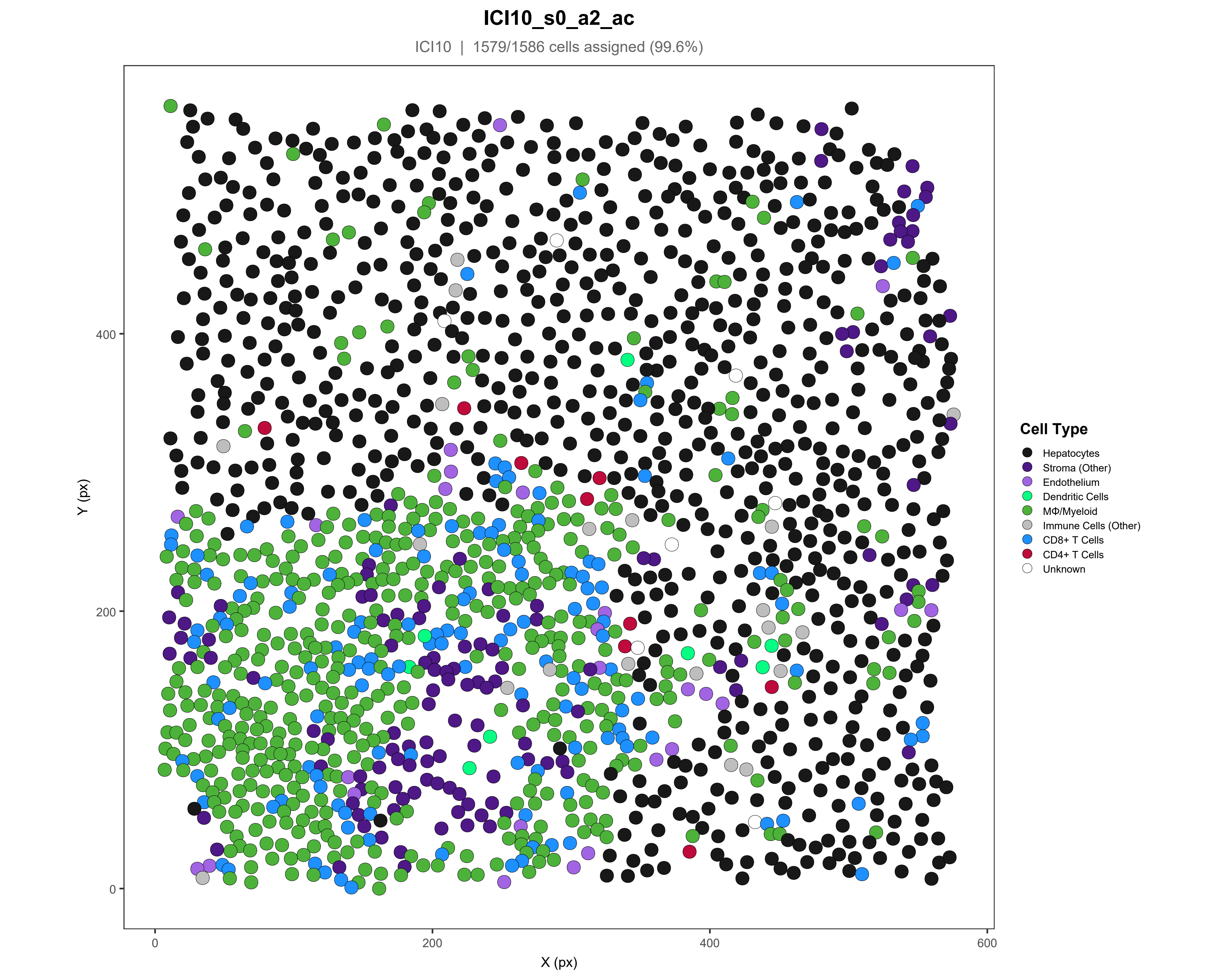
ICI10_s0_a2_ac
ICI10 | 1579/1586 cells assigned (99.6%)
Y (px)
400
200
0
X (px)
0
200
400
600
Cell Type
Hepatocytes
Stroma (Other)
Endothelium
Dendritic Cells
MΦ/Myeloid
Immune Cells (Other)
CD8+ T Cells
CD4+ T Cells
Unknown

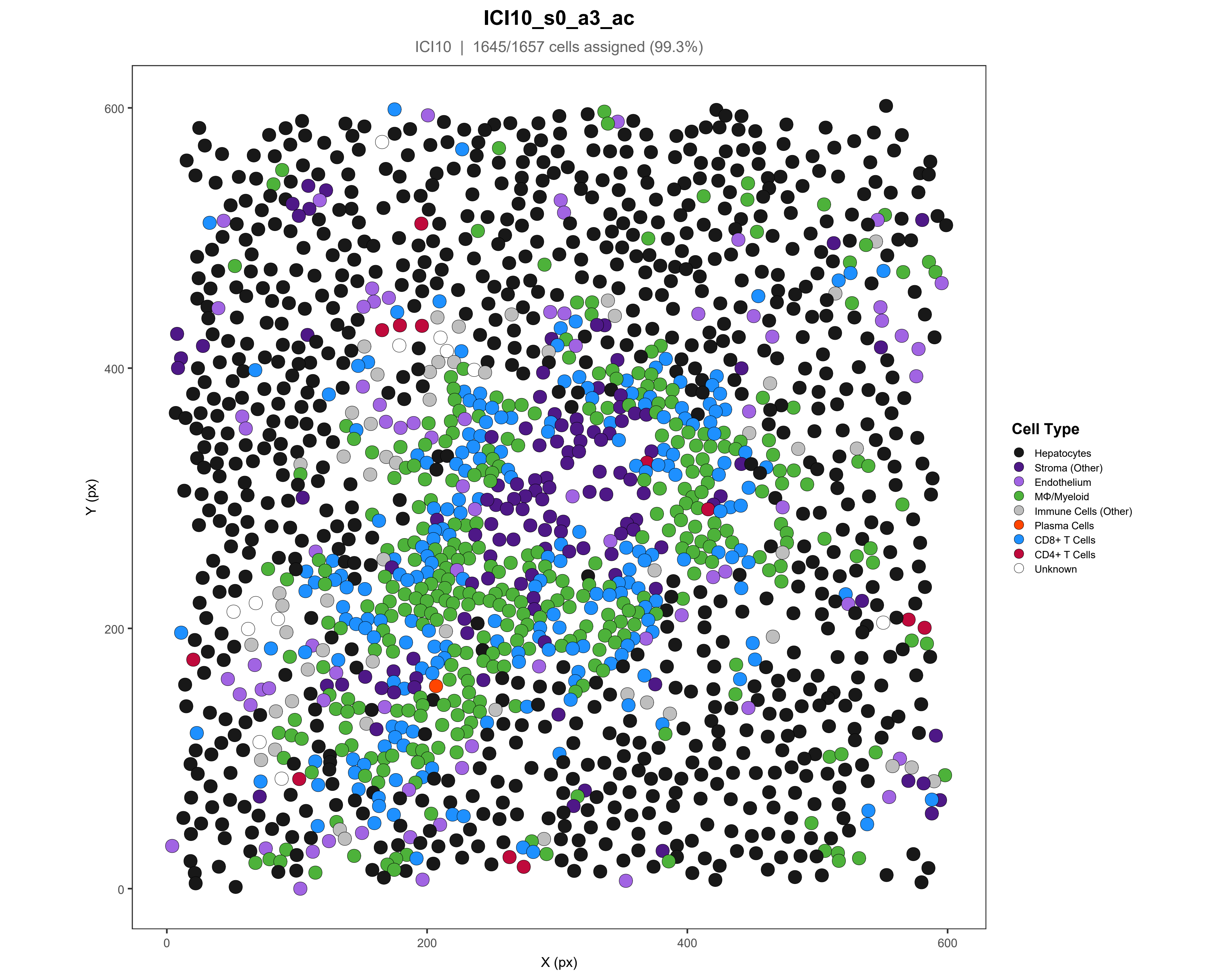
ICI10_s0_a3_ac
ICI10 | 1645/1657 cells assigned (99.3%)
Y (px)
X (px)
0
200
400
600
Cell Type
Hepatocytes
Stroma (Other)
Endothelium
MΦ/Myeloid
Immune Cells (Other)
Plasma Cells
CD8+ T Cells
CD4+ T Cells
Unknown

### ICI10_s0_a4_ac

ICI10 | 2348/2349 cells assigned (100%)

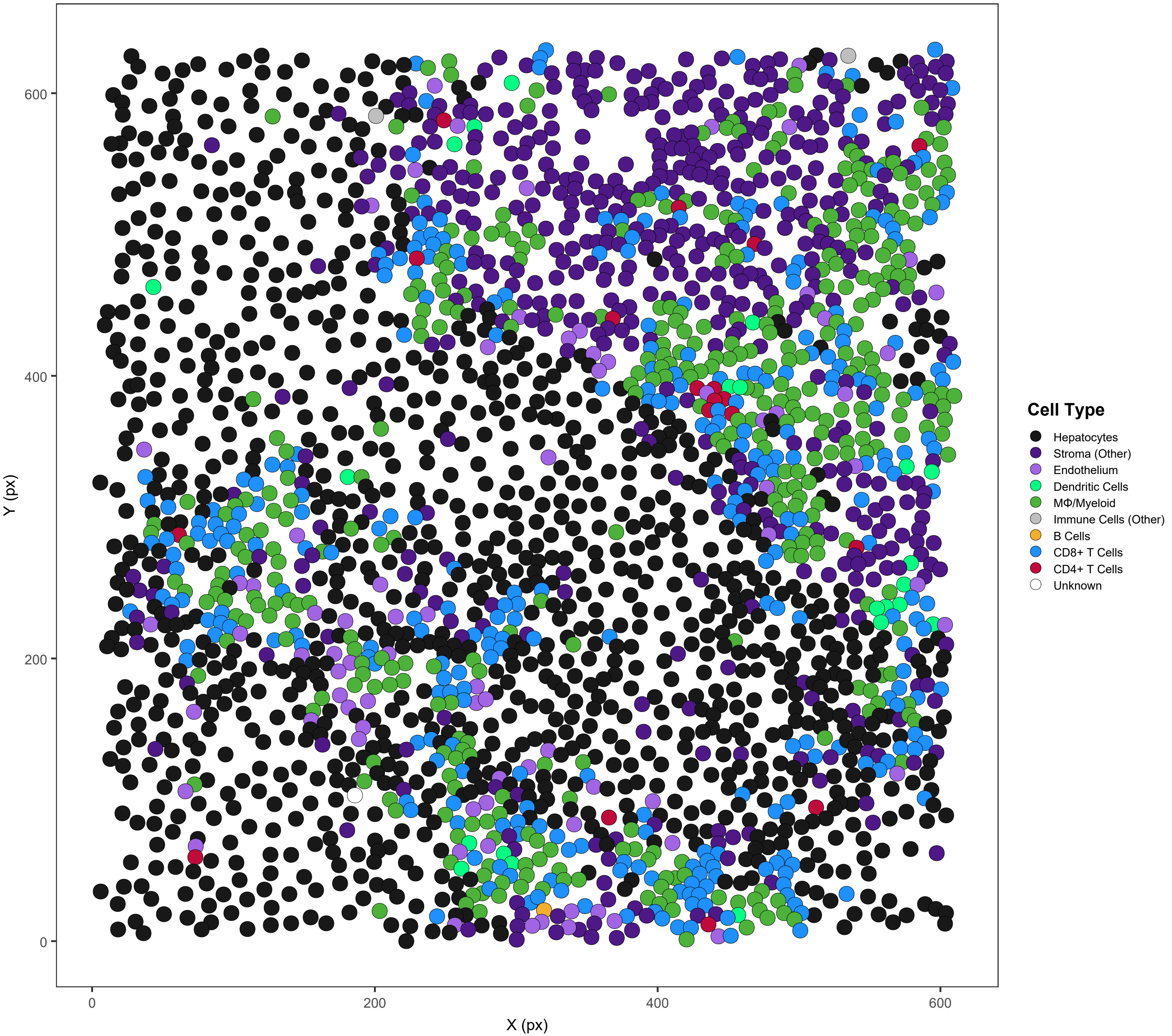

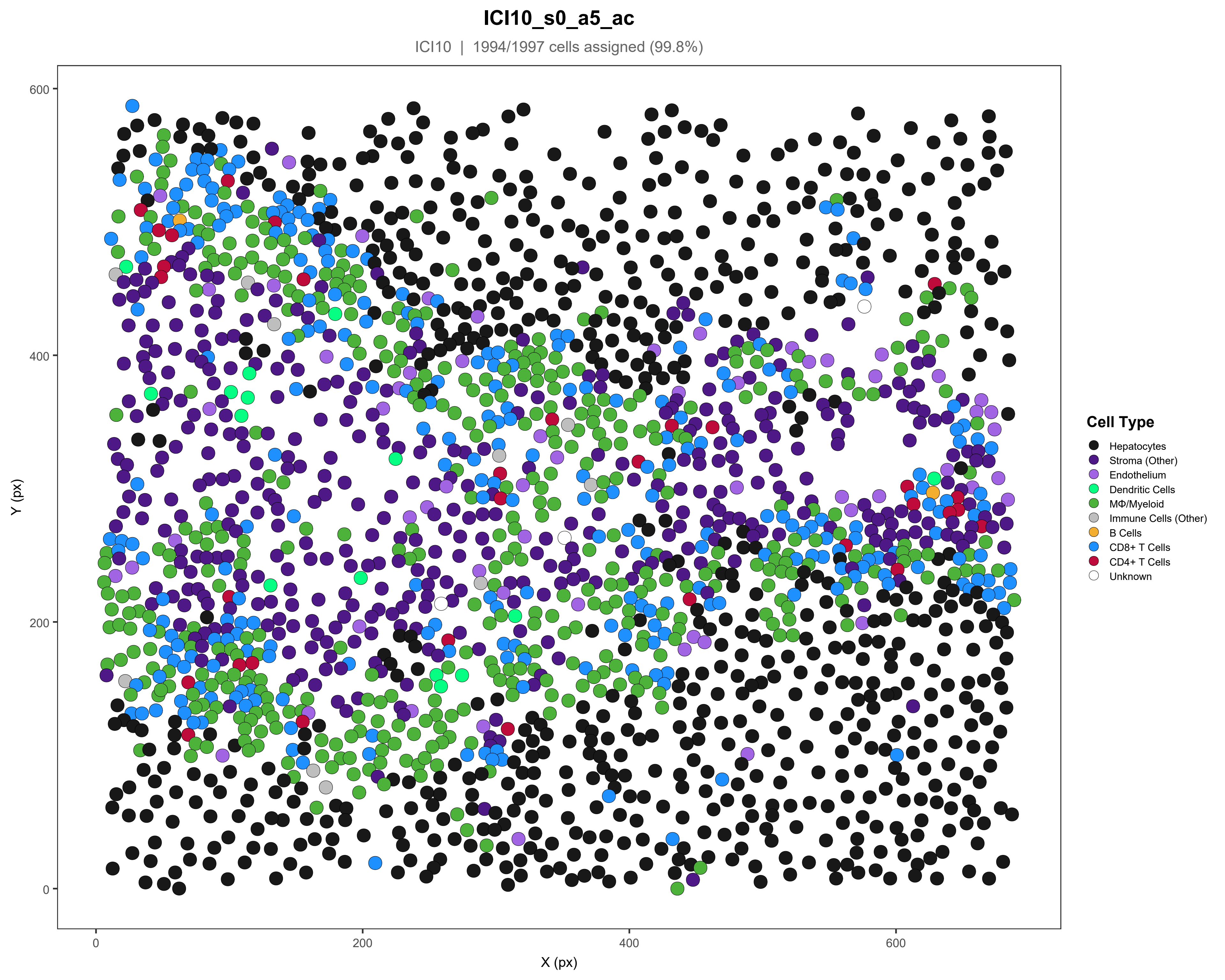
ICI10_s0_a5_ac
ICI10 | 1994/1997 cells assigned (99.8%)
Y (px)
X (px)
0
200
400
600
Cell Type
Hepatocytes
Stroma (Other)
Endothelium
Dendritic Cells
MΦ/Myeloid
Immune Cells (Other)
B Cells
CD8+ T Cells
CD4+ T Cells
Unknown

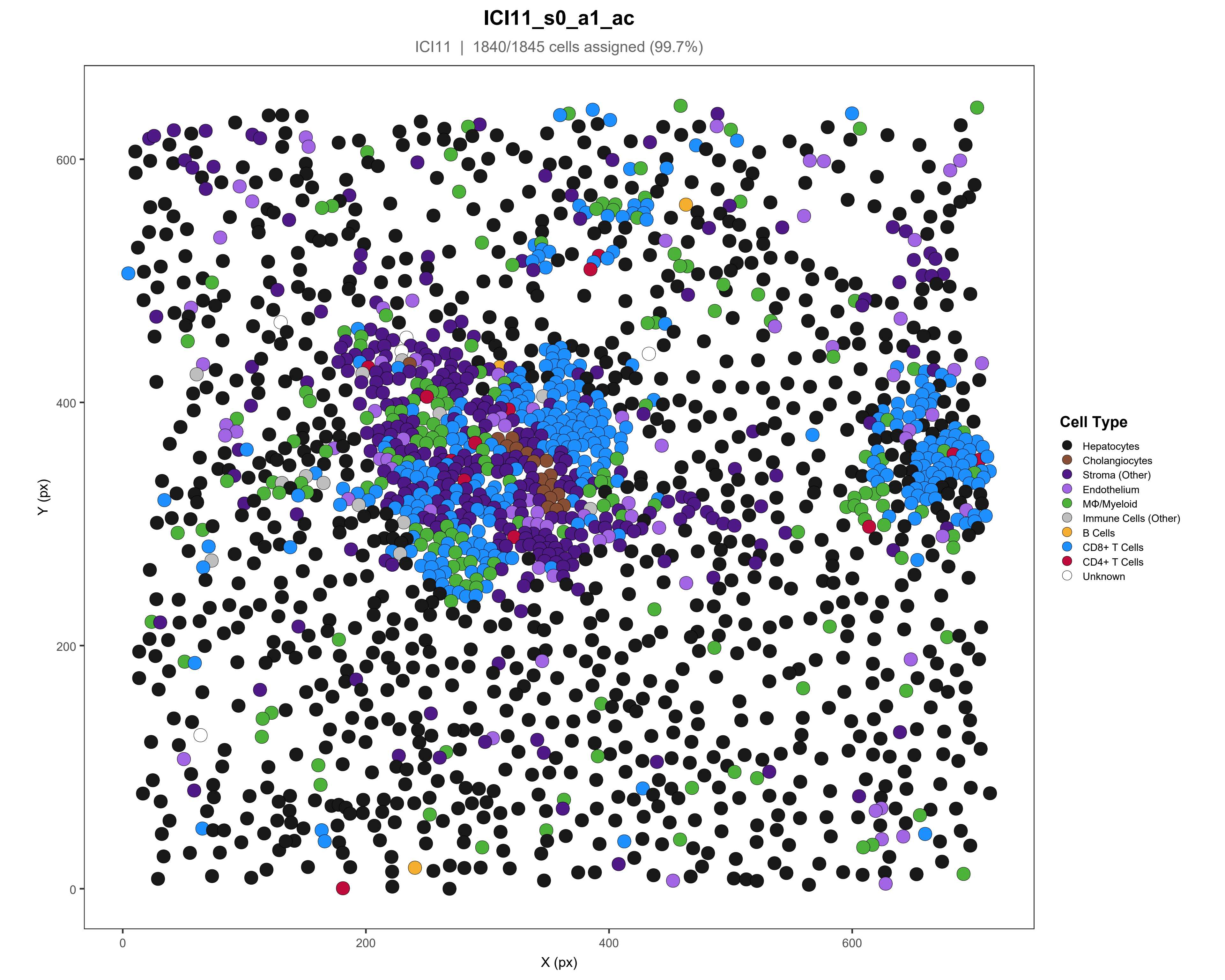

ICI11_s0_a1_ac
ICI11 | 1840/1845 cells assigned (99.7%)
Y (px)
X (px)
0
200
400
600
Cell Type
Hepatocytes
Cholangiocytes
Stroma (Other)
Endothelium
MΦ/Myeloid
Immune Cells (Other)
B Cells
CD8+ T Cells
CD4+ T Cells
Unknown

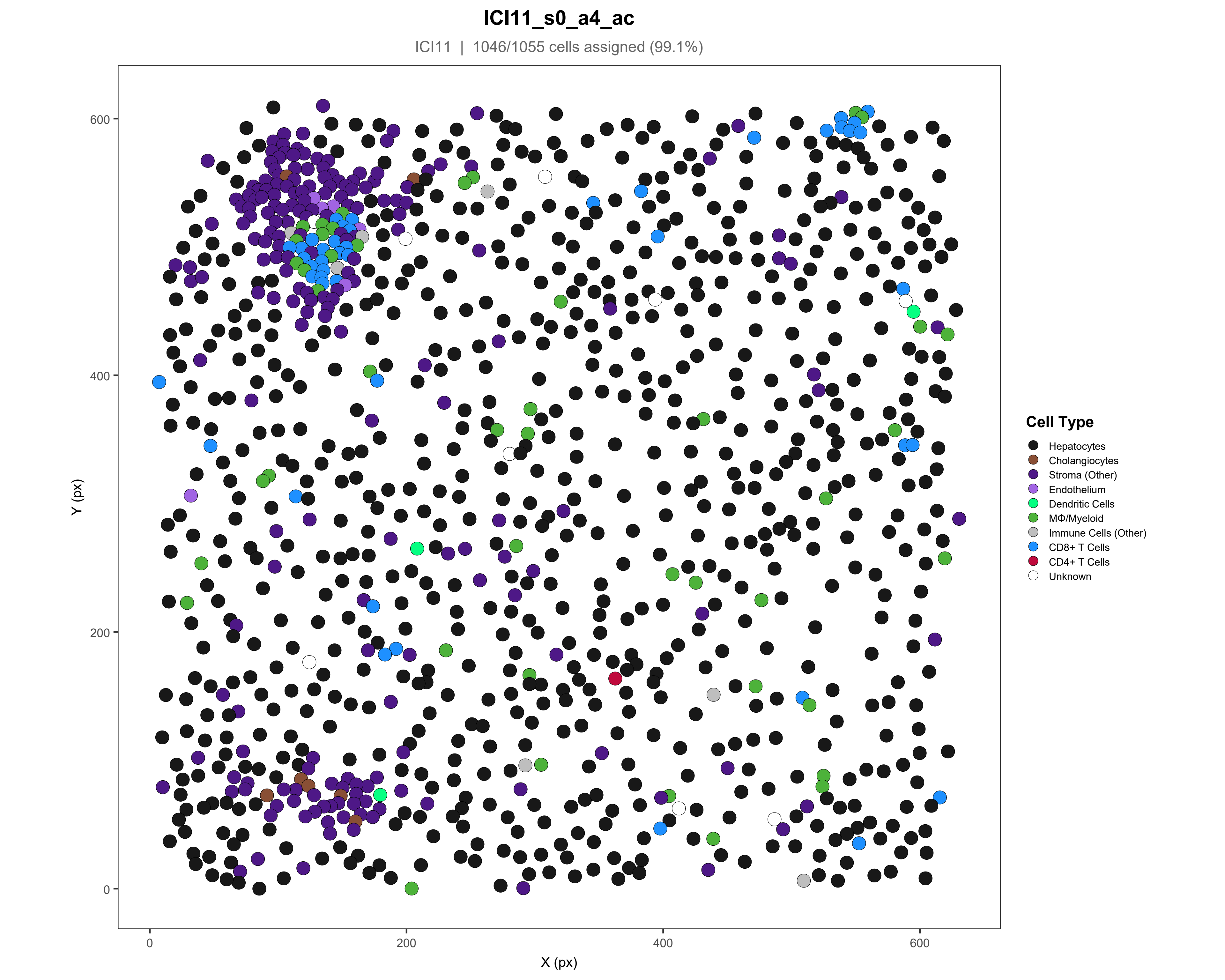

ICI11_s0_a4_ac
ICI11 | 1046/1055 cells assigned (99.1%)
Y (px)
X (px)
0
200
400
600
Cell Type
Hepatocytes
Cholangiocytes
Stroma (Other)
Endothelium
Dendritic Cells
MΦ/Myeloid
Immune Cells (Other)
CD8+ T Cells
CD4+ T Cells
Unknown

### ICI12_s0_a1_ac

ICI12 | 1177/1229 cells assigned (95.8%)

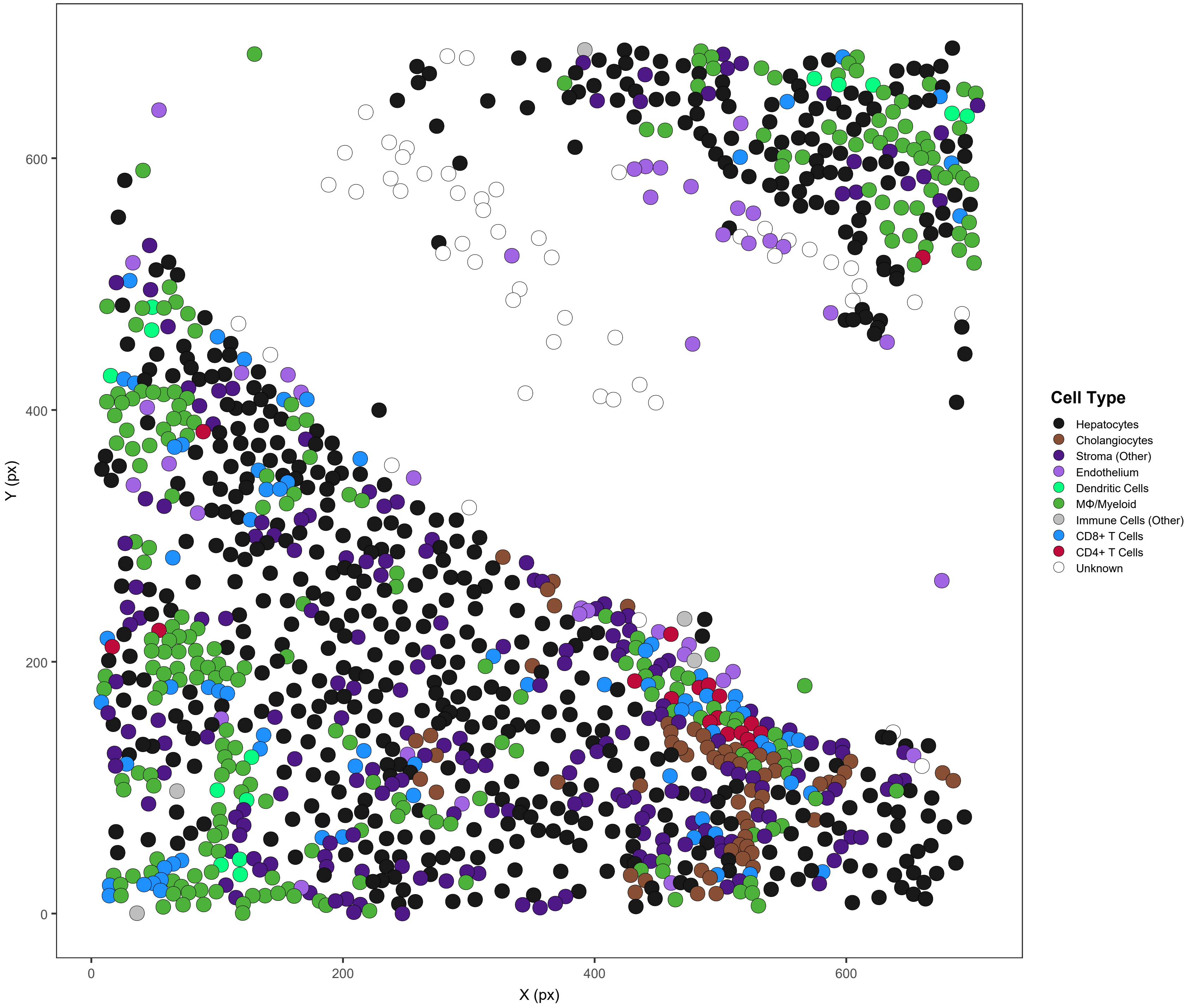

### ICI12_s0_a2_ac

ICI12 | 2154/2154 cells assigned (100%)

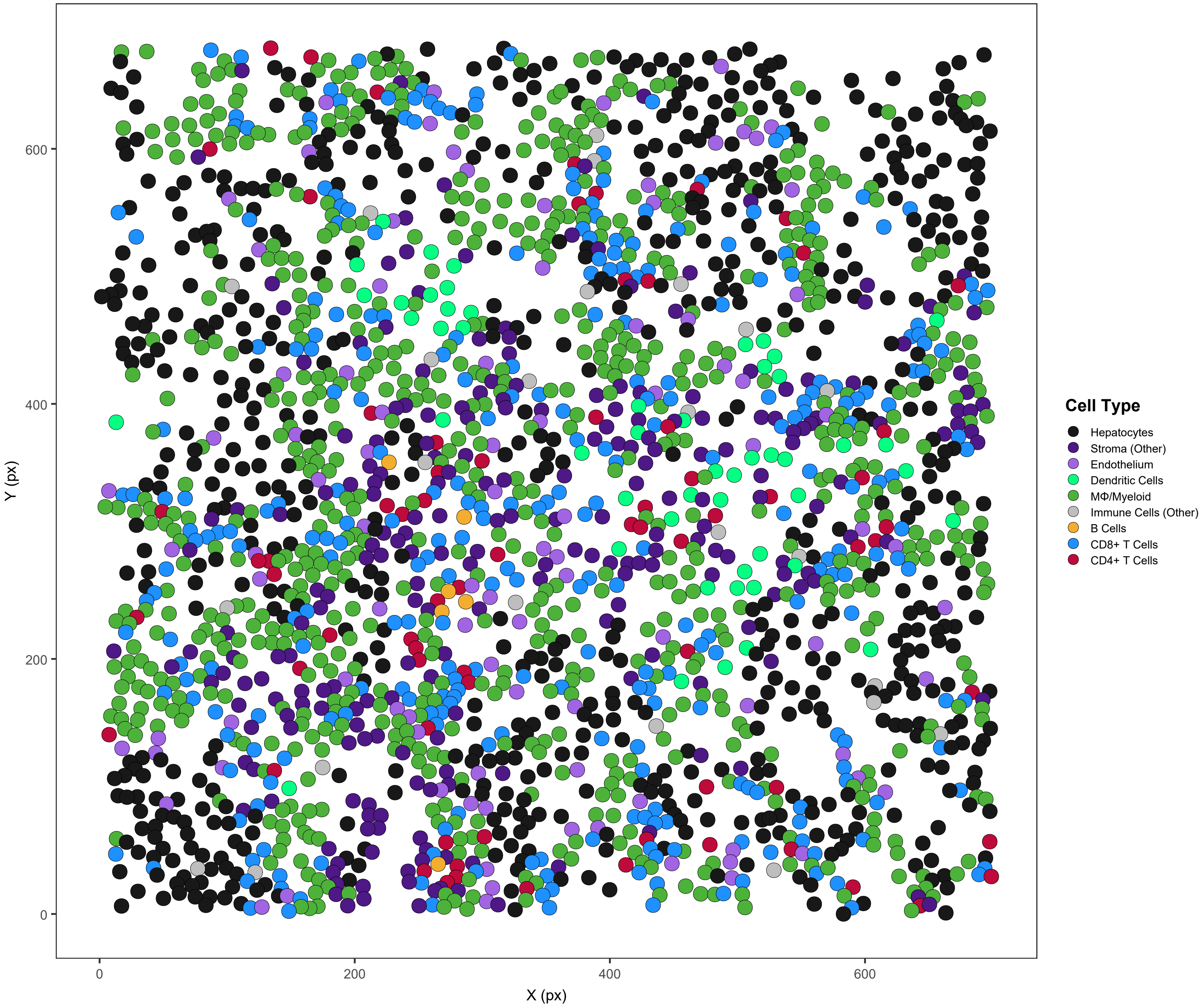

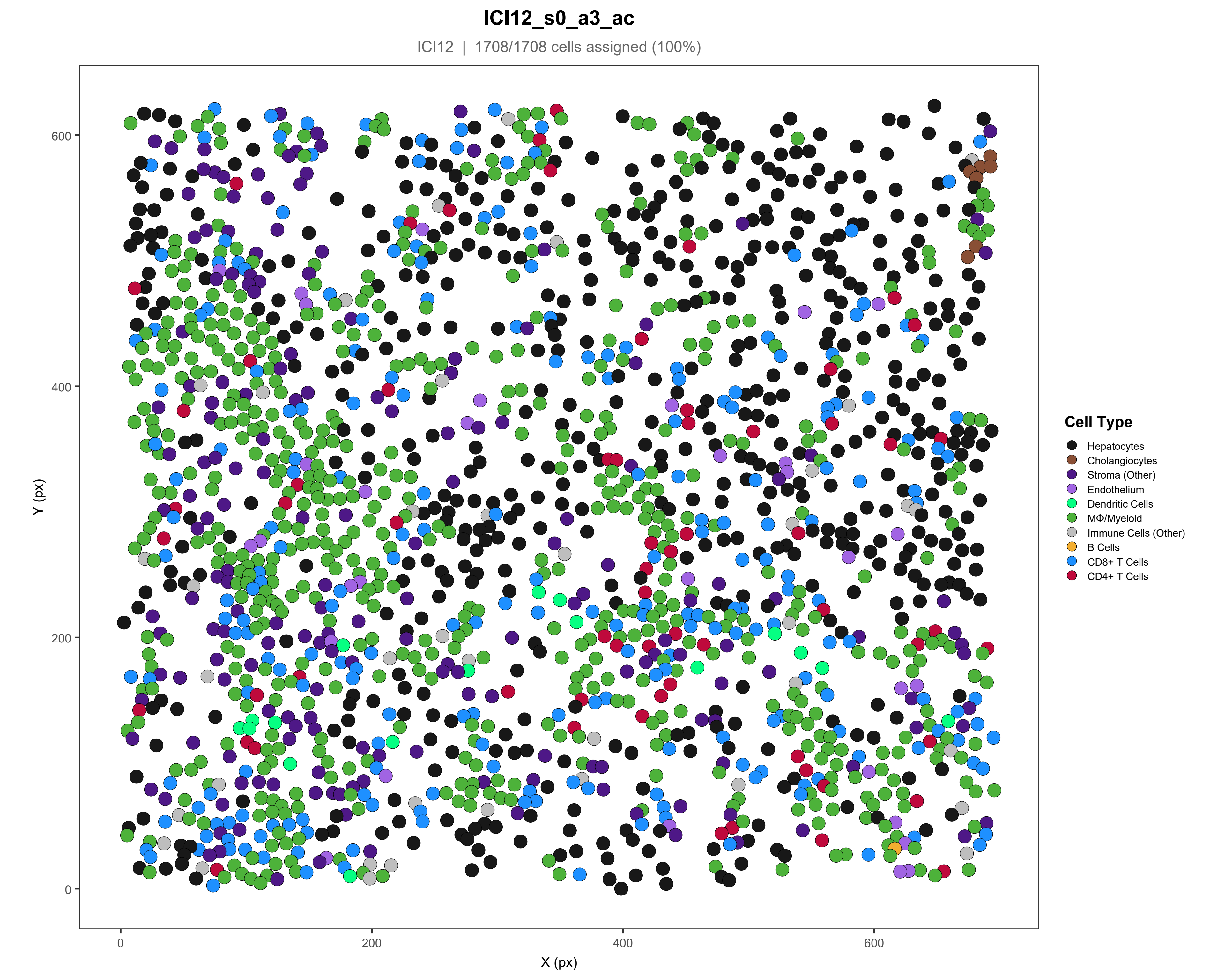

ICI12_s0_a3_ac
ICI12 | 1708/1708 cells assigned (100%)
Y (px)
X (px)
0
200
400
600
Cell Type
Hepatocytes
Cholangiocytes
Stroma (Other)
Endothelium
Dendritic Cells
MΦ/Myeloid
Immune Cells (Other)
B Cells
CD8+ T Cells
CD4+ T Cells

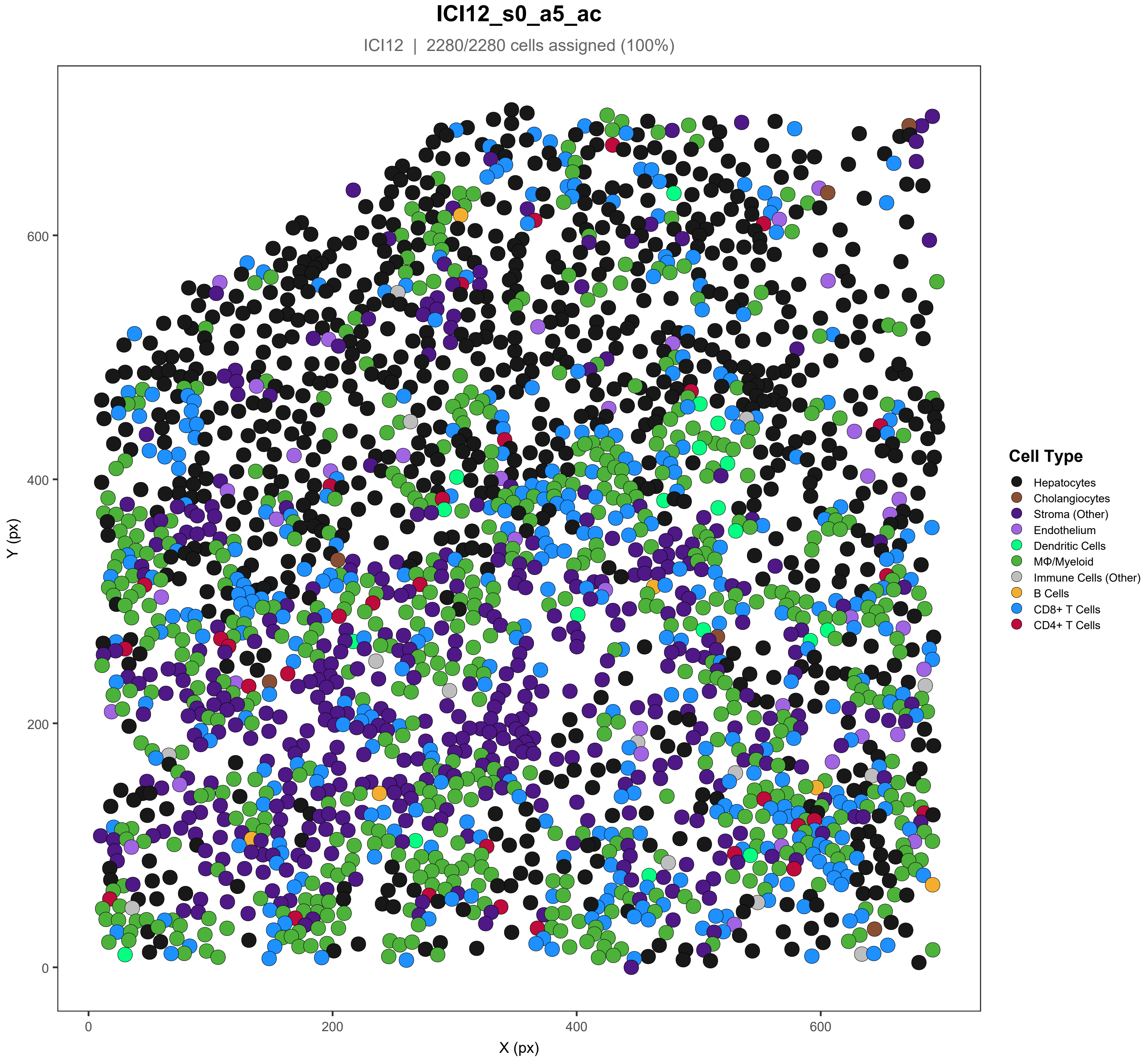
ICI12_s0_a5_ac
ICI12 | 2280/2280 cells assigned (100%)
Y (px)
X (px)
0
200
400
600
Cell Type
Hepatocytes
Cholangiocytes
Stroma (Other)
Endothelium
Dendritic Cells
MΦ/Myeloid
Immune Cells (Other)
B Cells
CD8+ T Cells
CD4+ T Cells

### ICI13_s0_a1_ac

ICI13 | 1955/1956 cells assigned (99.9%)

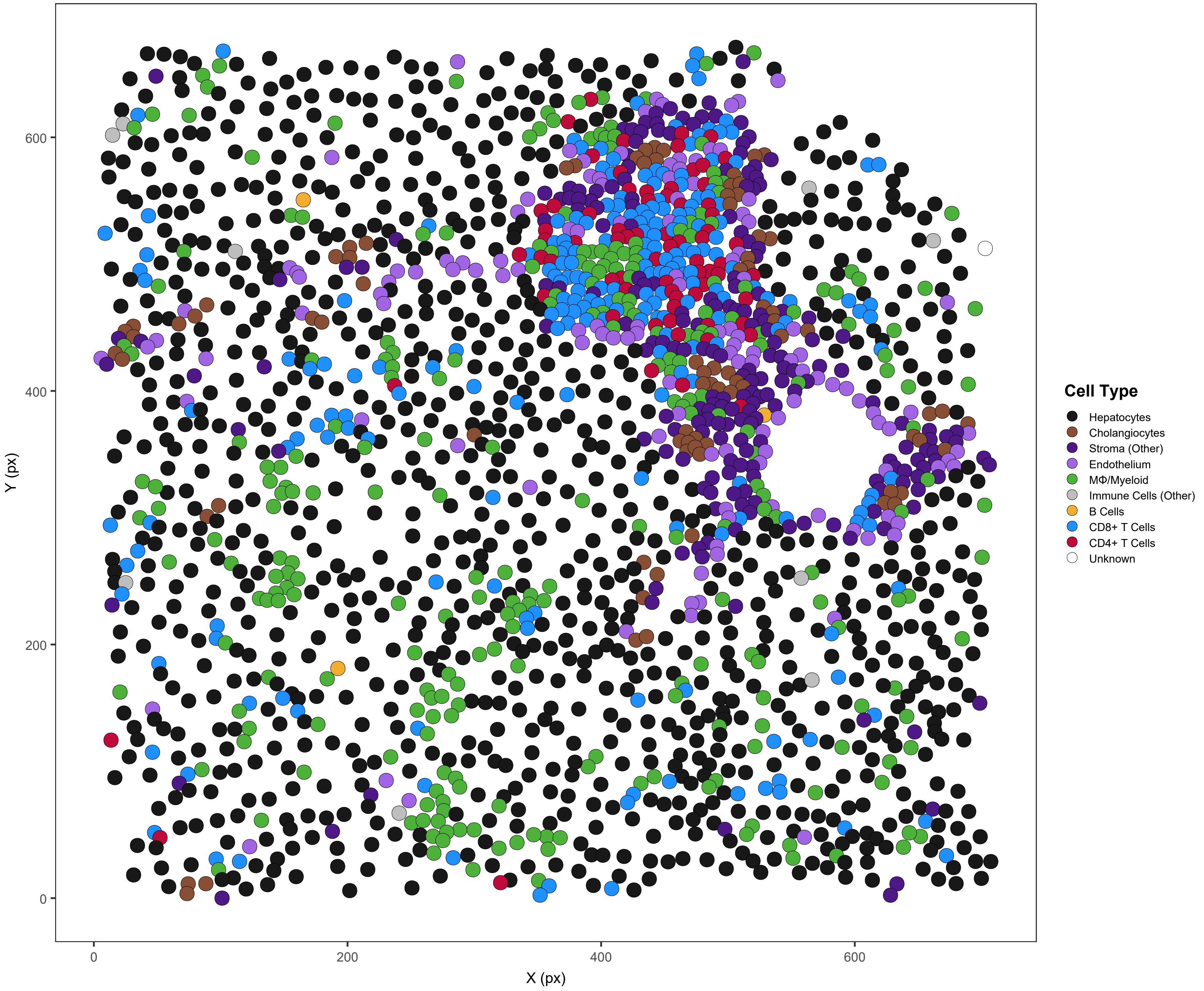

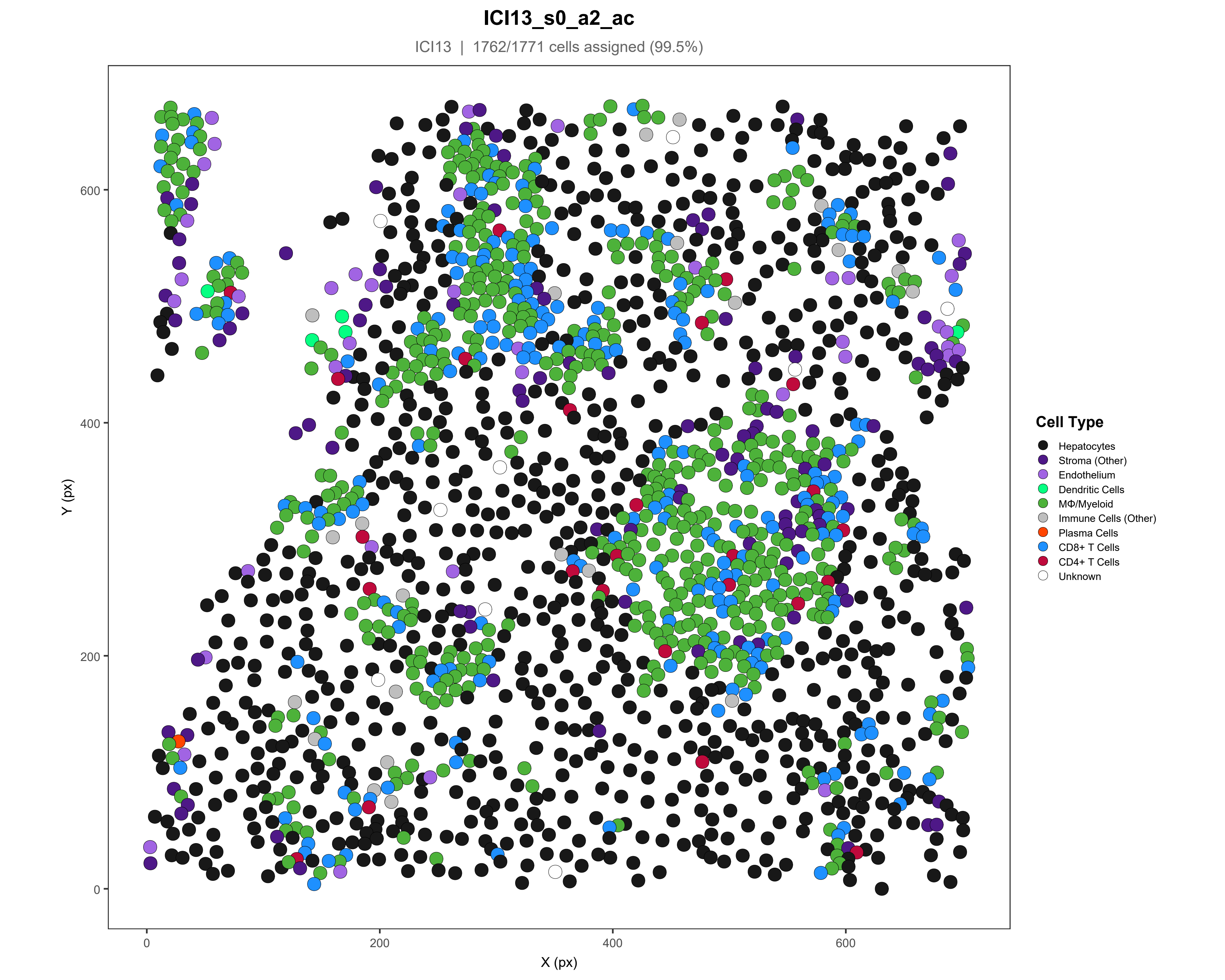
ICI13_s0_a2_ac
ICI13 | 1762/1771 cells assigned (99.5%)
Y (px)
X (px)
0
200
400
600
Cell Type
Hepatocytes
Stroma (Other)
Endothelium
Dendritic Cells
MΦ/Myeloid
Immune Cells (Other)
Plasma Cells
CD8+ T Cells
CD4+ T Cells
Unknown

**ICI13_s0_a3_ac**

ICI13 | 1879/1880 cells assigned (99.9%)

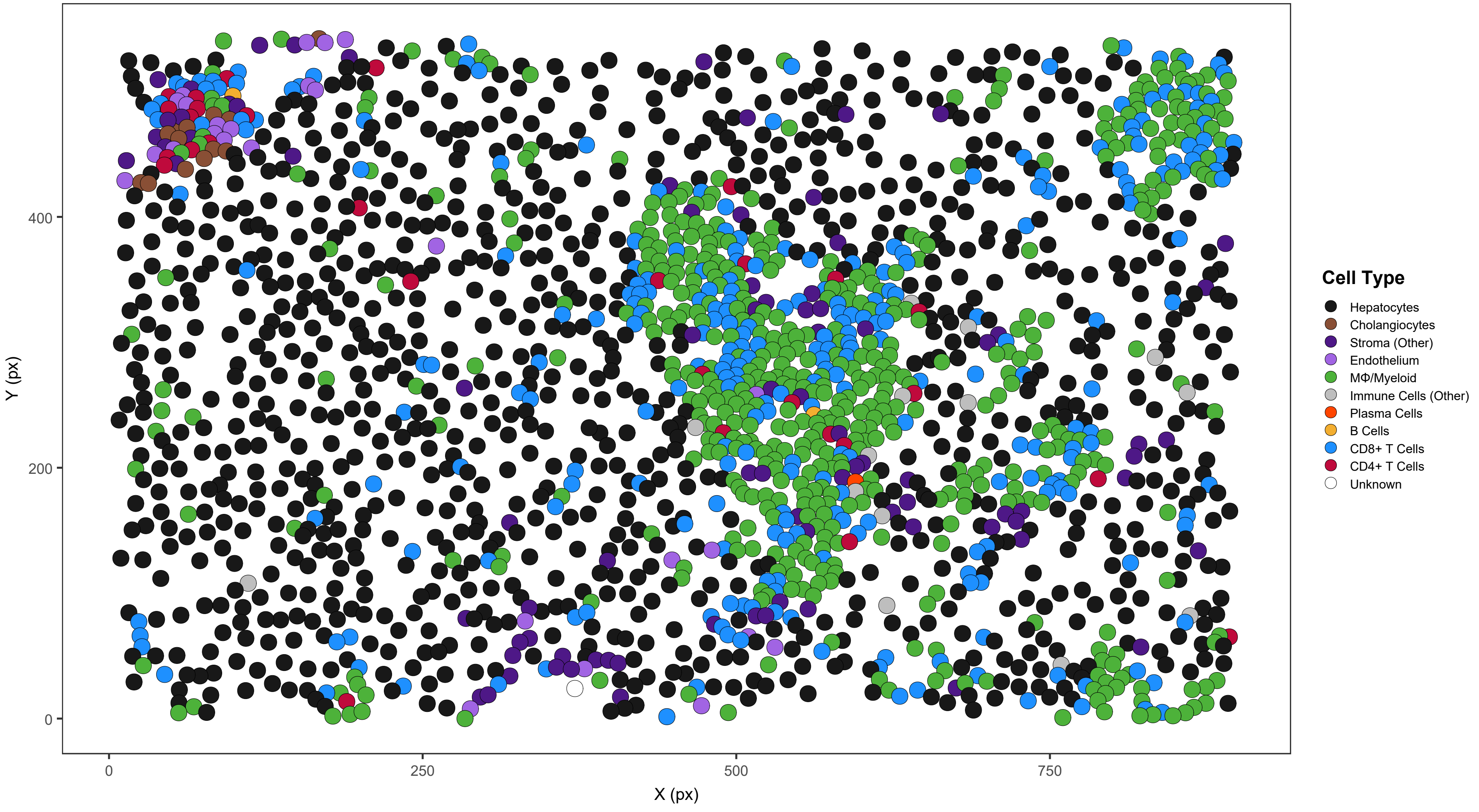

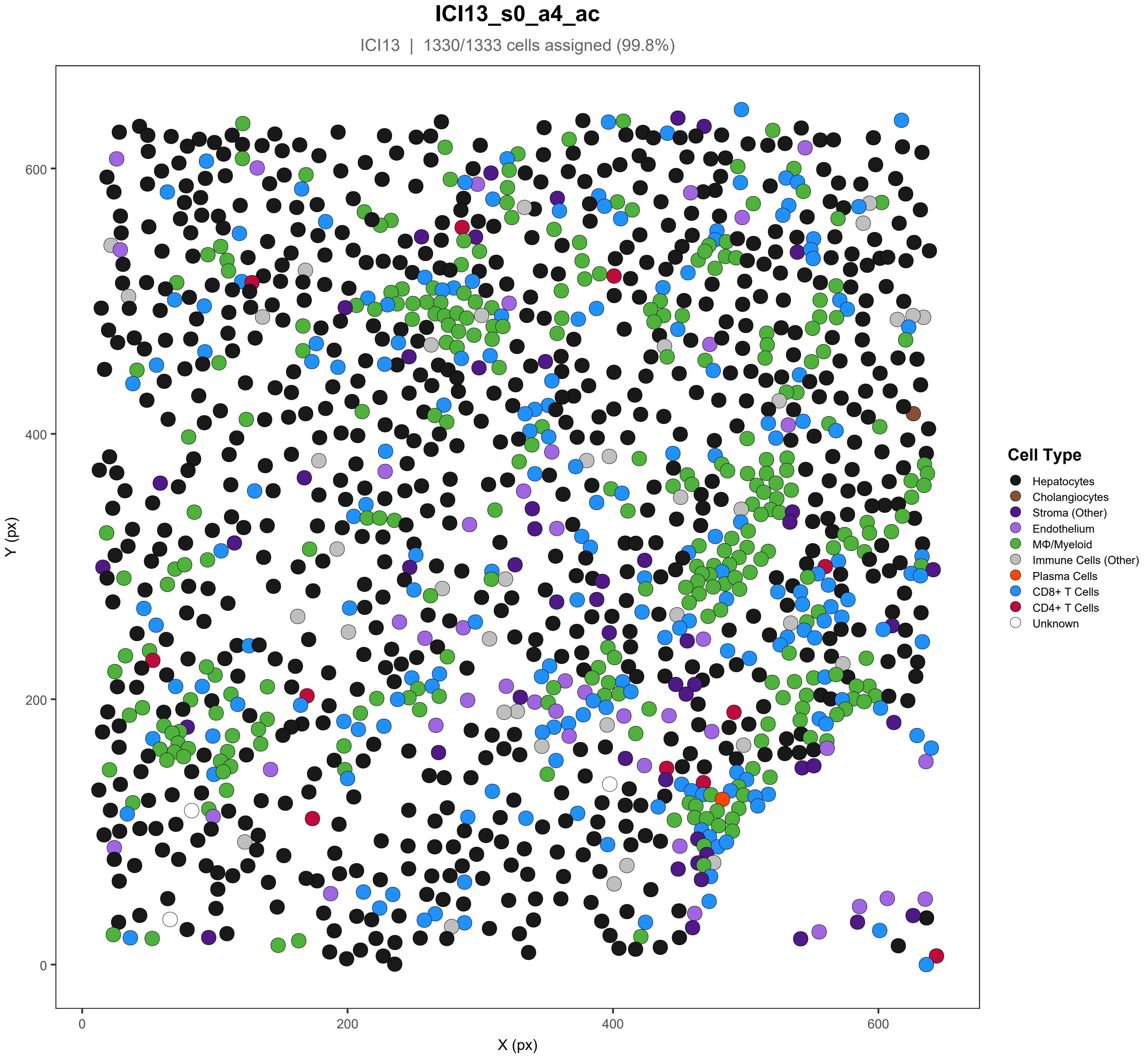
ICI13_s0_a4_ac
ICI13 | 1330/1333 cells assigned (99.8%)
Y (px)
X (px)
0
200
400
600
Cell Type
Hepatocytes
Cholangiocytes
Stroma (Other)
Endothelium
MΦ/Myeloid
Immune Cells (Other)
Plasma Cells
CD8+ T Cells
CD4+ T Cells
Unknown

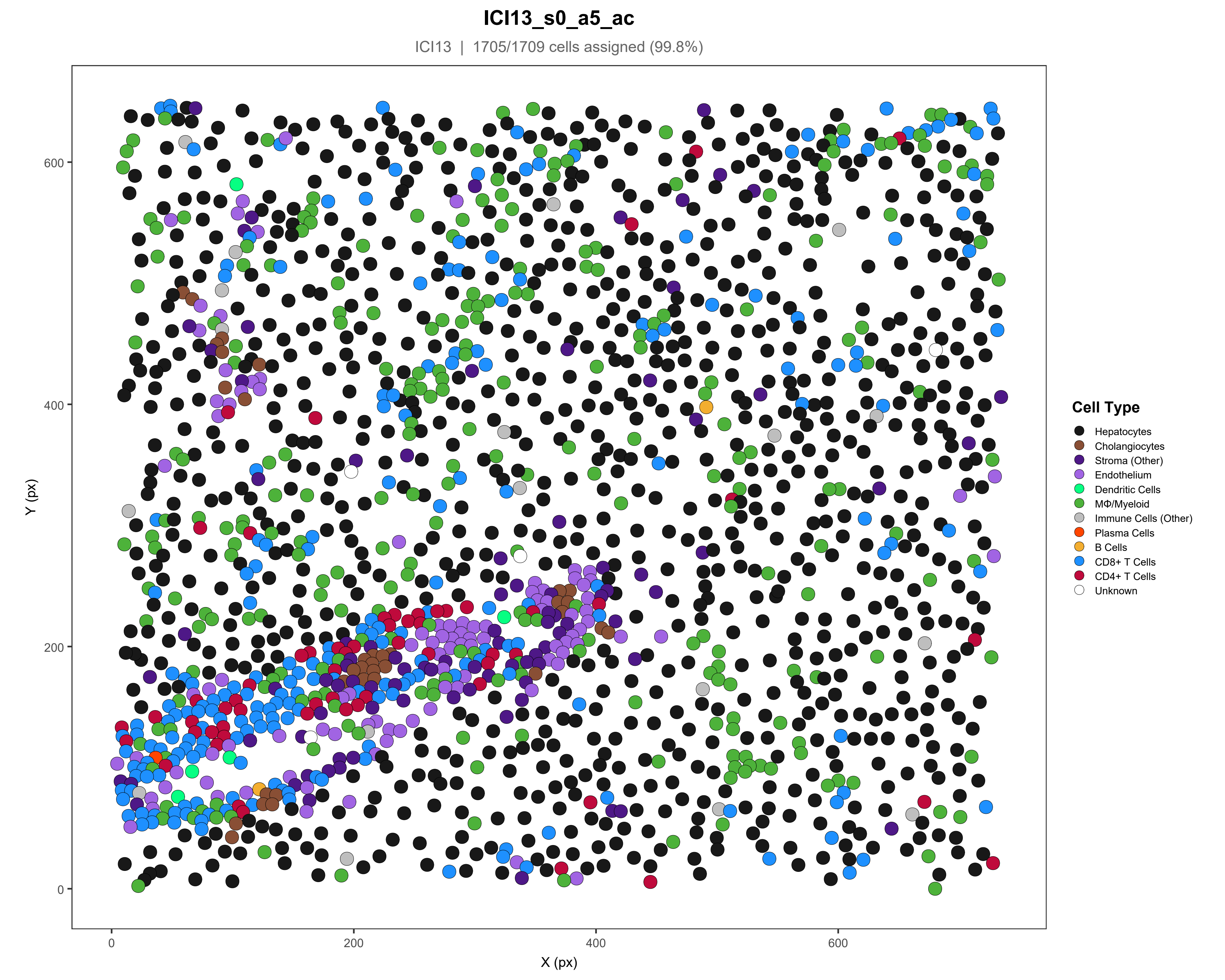

ICI13_s0_a5_ac
ICI13 | 1705/1709 cells assigned (99.8%)
Y (px)
X (px)
0
200
400
600
Cell Type
Hepatocytes
Cholangiocytes
Stroma (Other)
Endothelium
Dendritic Cells
MΦ/Myeloid
Immune Cells (Other)
Plasma Cells
B Cells
CD8+ T Cells
CD4+ T Cells
Unknown

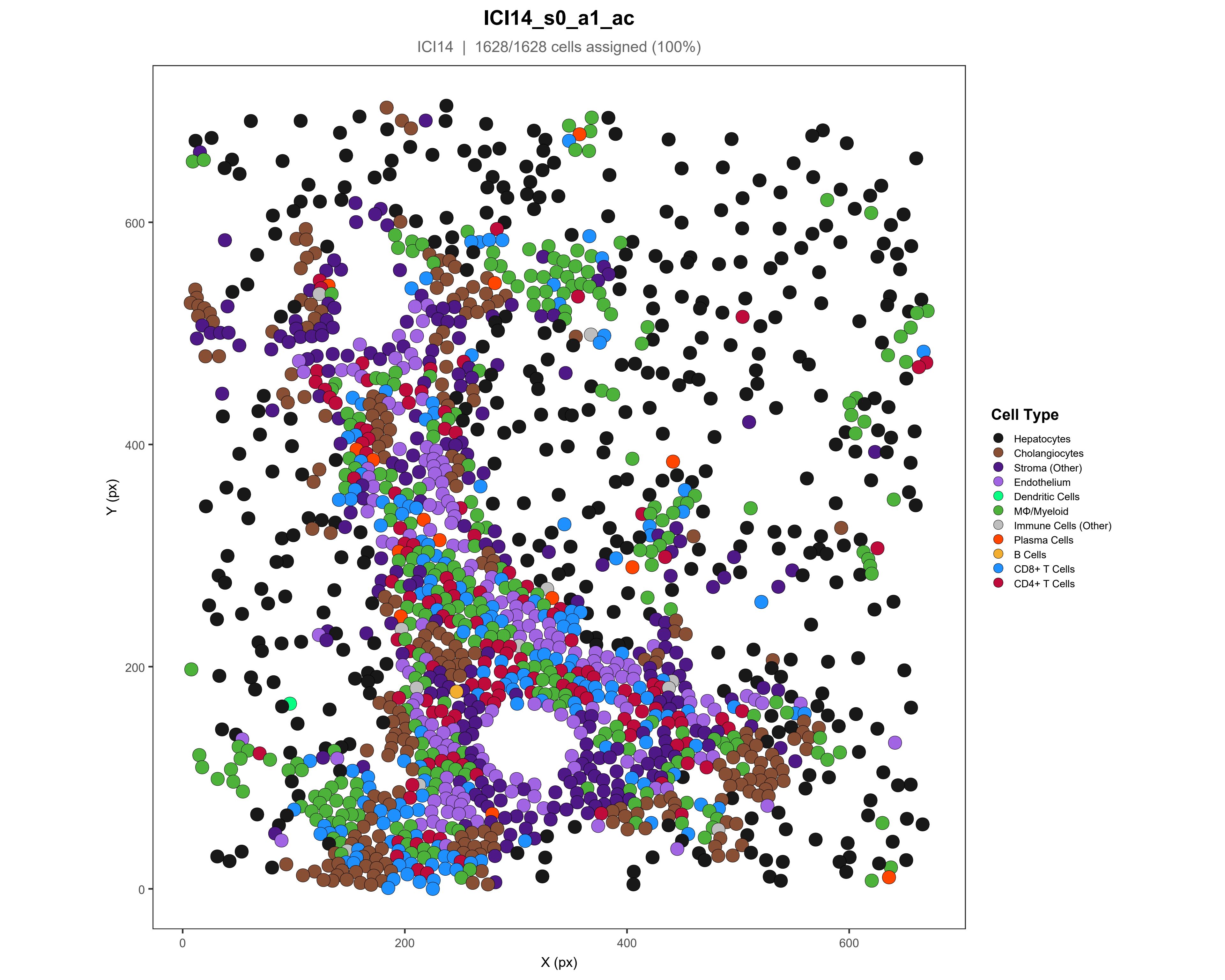

ICI14_s0_a1_ac
ICI14 | 1628/1628 cells assigned (100%)
Y (px)
X (px)
0
200
400
600
Cell Type
Hepatocytes
Cholangiocytes
Stroma (Other)
Endothelium
Dendritic Cells
MΦ/Myeloid
Immune Cells (Other)
Plasma Cells
B Cells
CD8+ T Cells
CD4+ T Cells

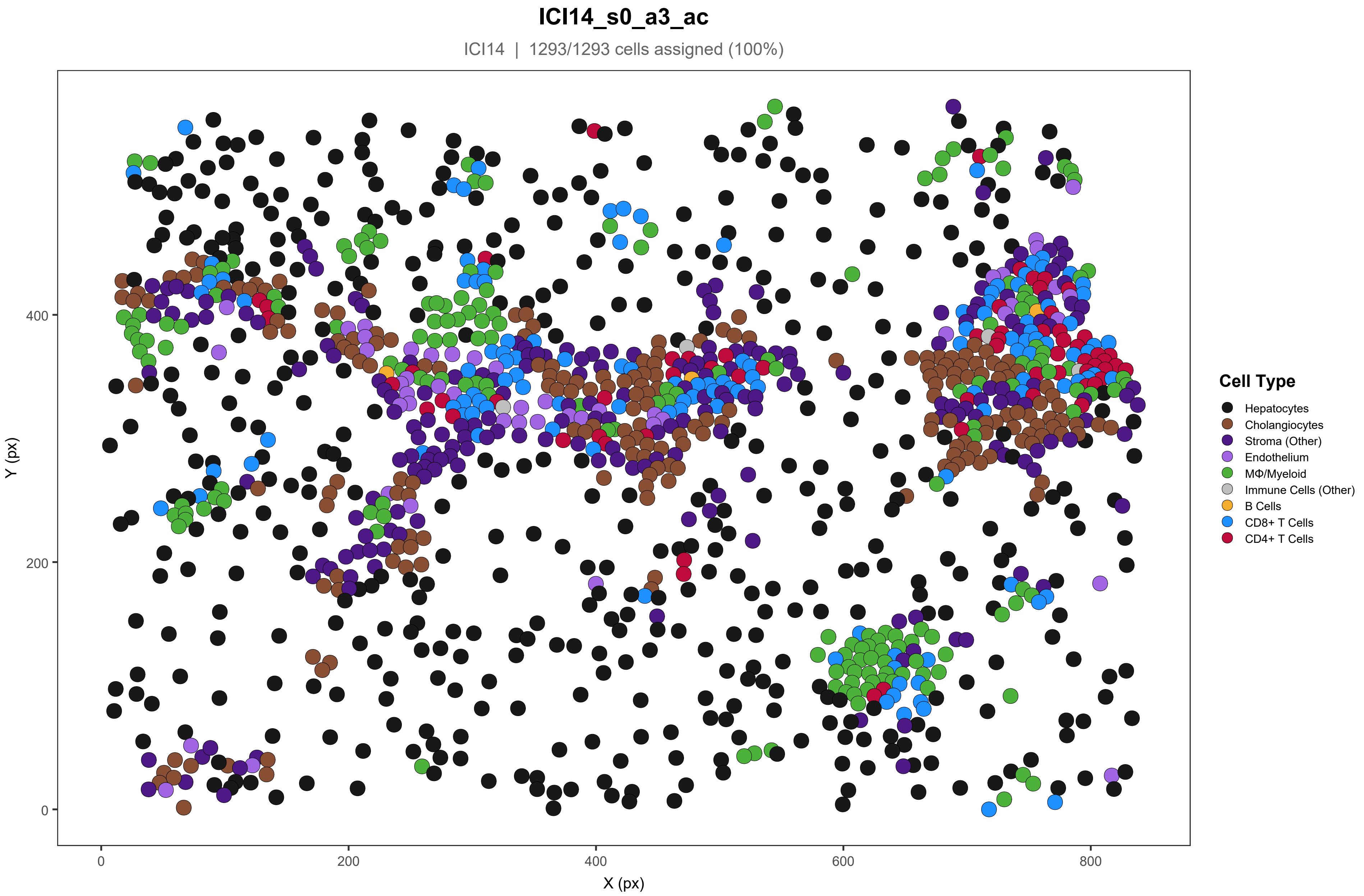
ICI14_s0_a3_ac
ICI14 | 1293/1293 cells assigned (100%)
Y (px)
X (px)
0
200
400
600
800
Cell Type
Hepatocytes
Cholangiocytes
Stroma (Other)
Endothelium
MΦ/Myeloid
Immune Cells (Other)
B Cells
CD8+ T Cells
CD4+ T Cells

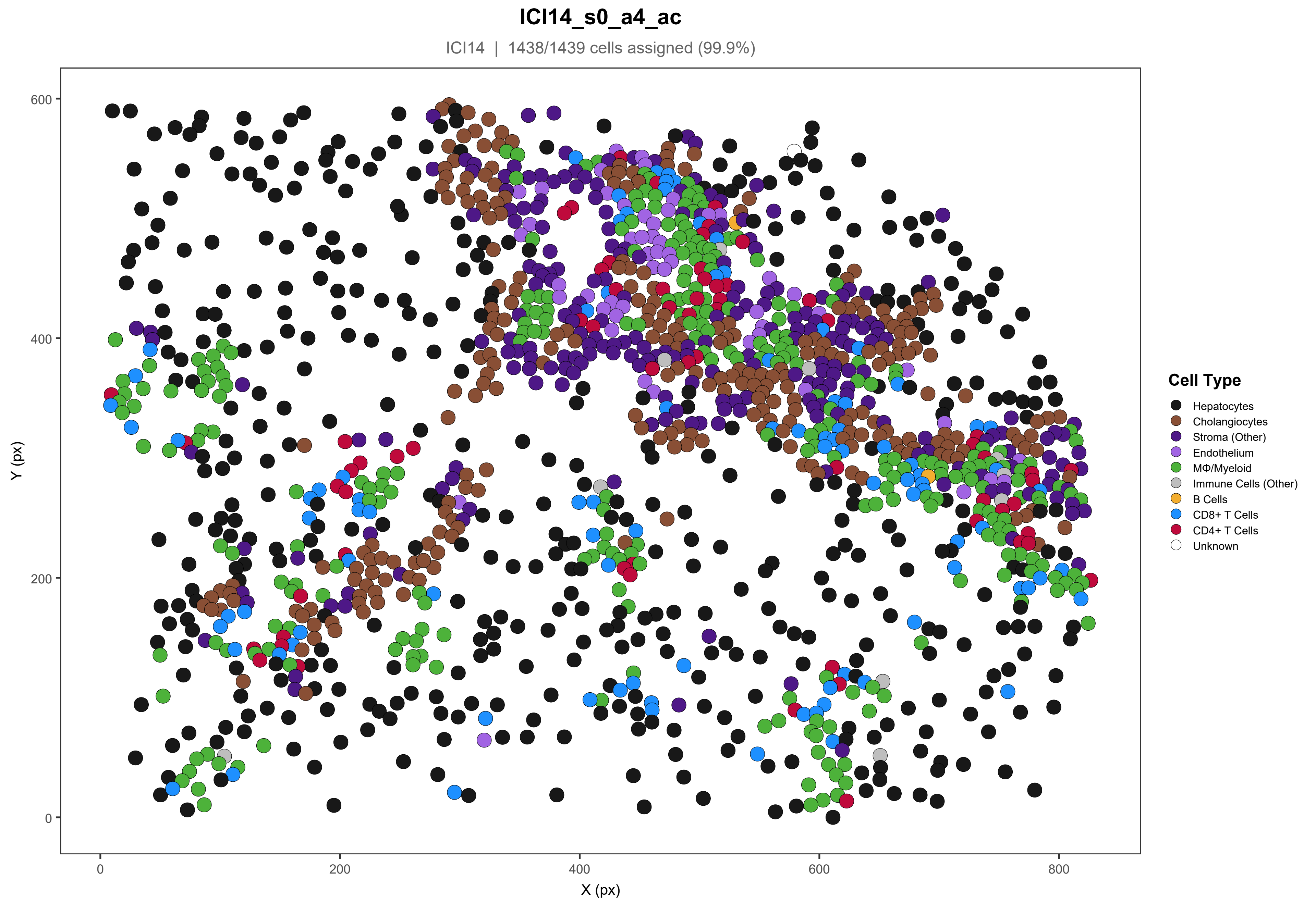

ICI14_s0_a4_ac
ICI14 | 1438/1439 cells assigned (99.9%)
Y (px)
X (px)
0
200
400
600
800
Cell Type
Hepatocytes
Cholangiocytes
Stroma (Other)
Endothelium
MΦ/Myeloid
Immune Cells (Other)
B Cells
CD8+ T Cells
CD4+ T Cells
Unknown

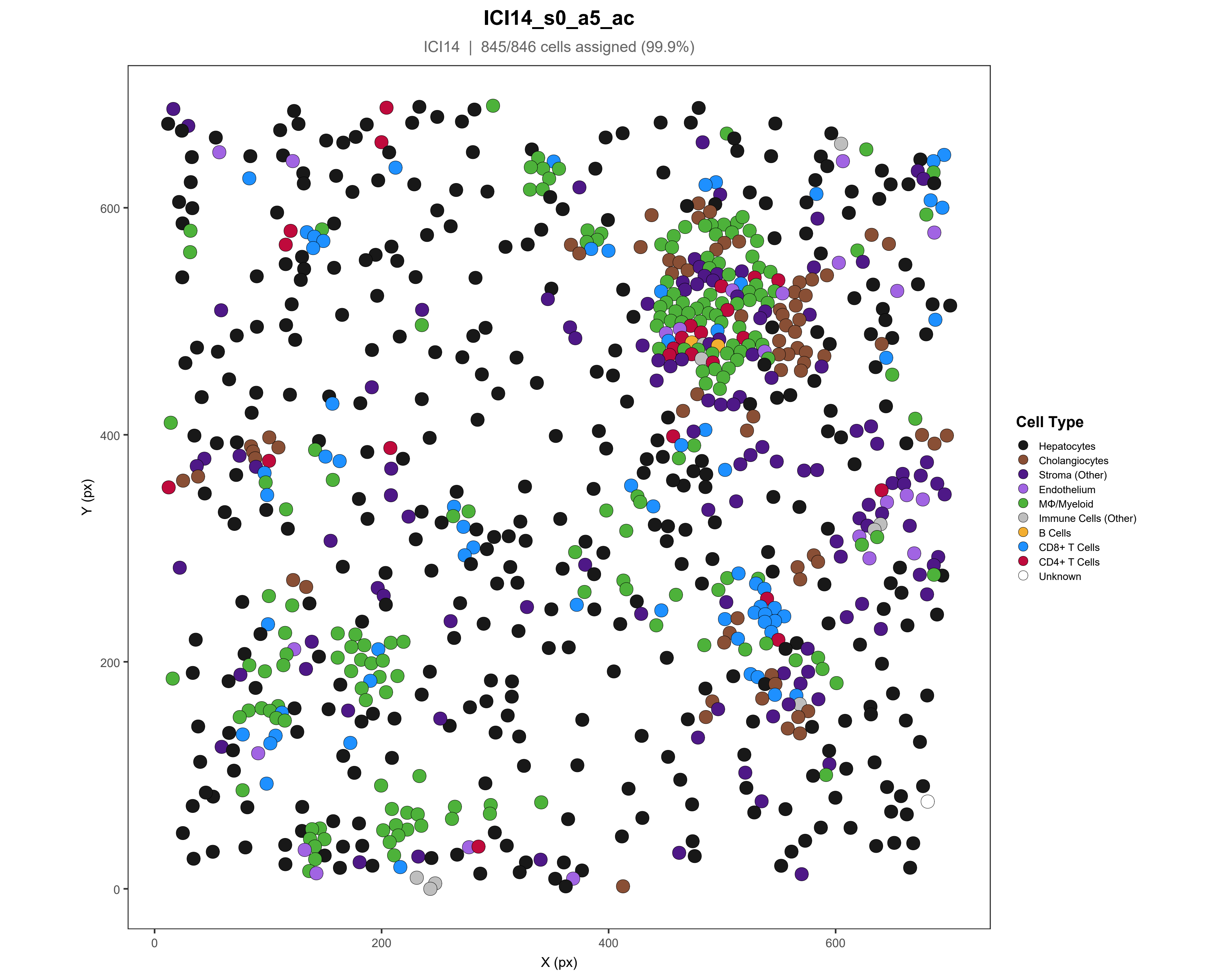

ICI14_s0_a5_ac
ICI14 | 845/846 cells assigned (99.9%)
Y (px)
X (px)
0
200
400
600
Cell Type
Hepatocytes
Cholangiocytes
Stroma (Other)
Endothelium
MΦ/Myeloid
Immune Cells (Other)
B Cells
CD8+ T Cells
CD4+ T Cells
Unknown

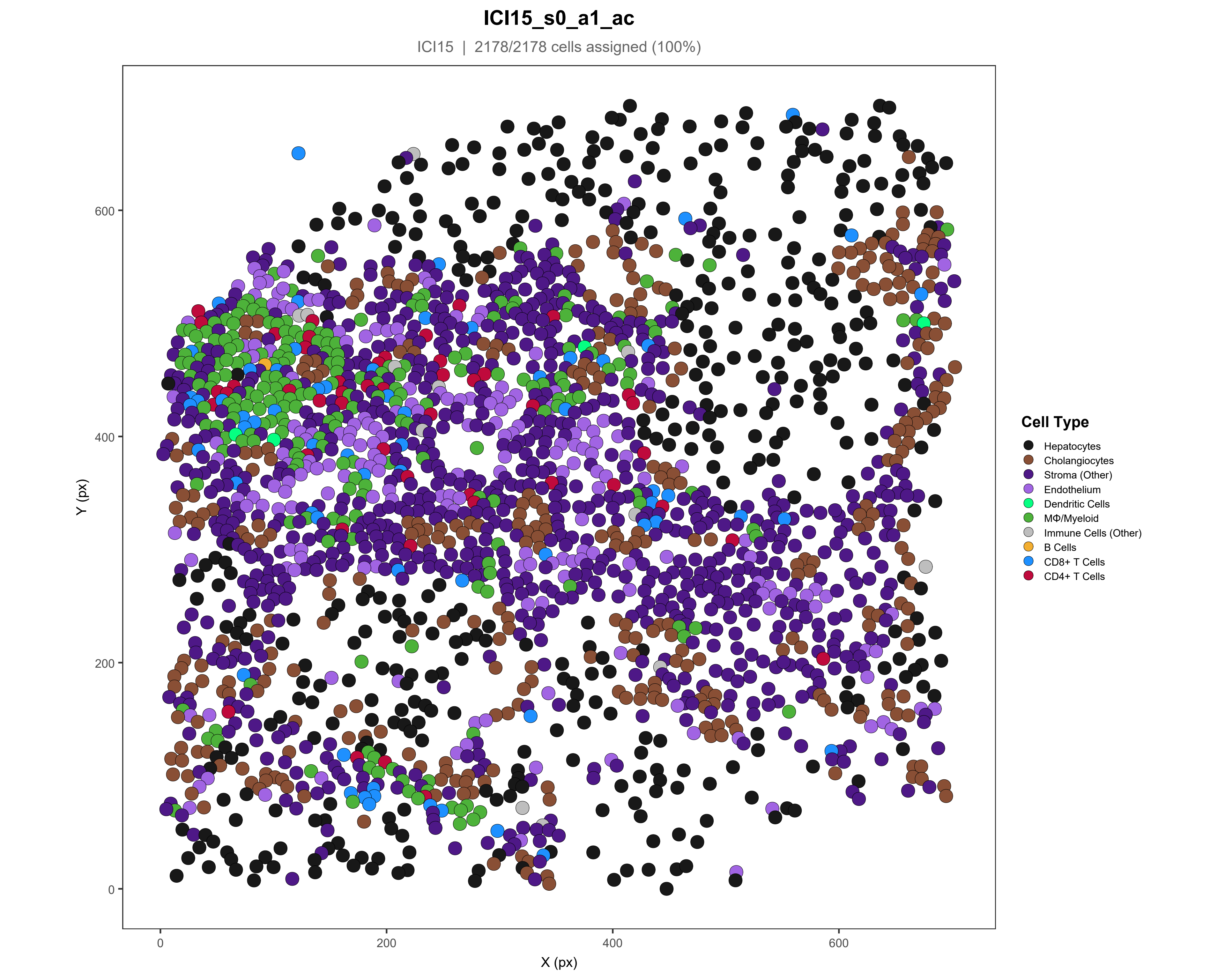

ICI15_s0_a1_ac
ICI15 | 2178/2178 cells assigned (100%)
Y (px)
X (px)
0
200
400
600
Cell Type
Hepatocytes
Cholangiocytes
Stroma (Other)
Endothelium
Dendritic Cells
MΦ/Myeloid
Immune Cells (Other)
B Cells
CD8+ T Cells
CD4+ T Cells

#### ICI15_s0_a2_ac

ICI15 | 2448/2473 cells assigned (99%)

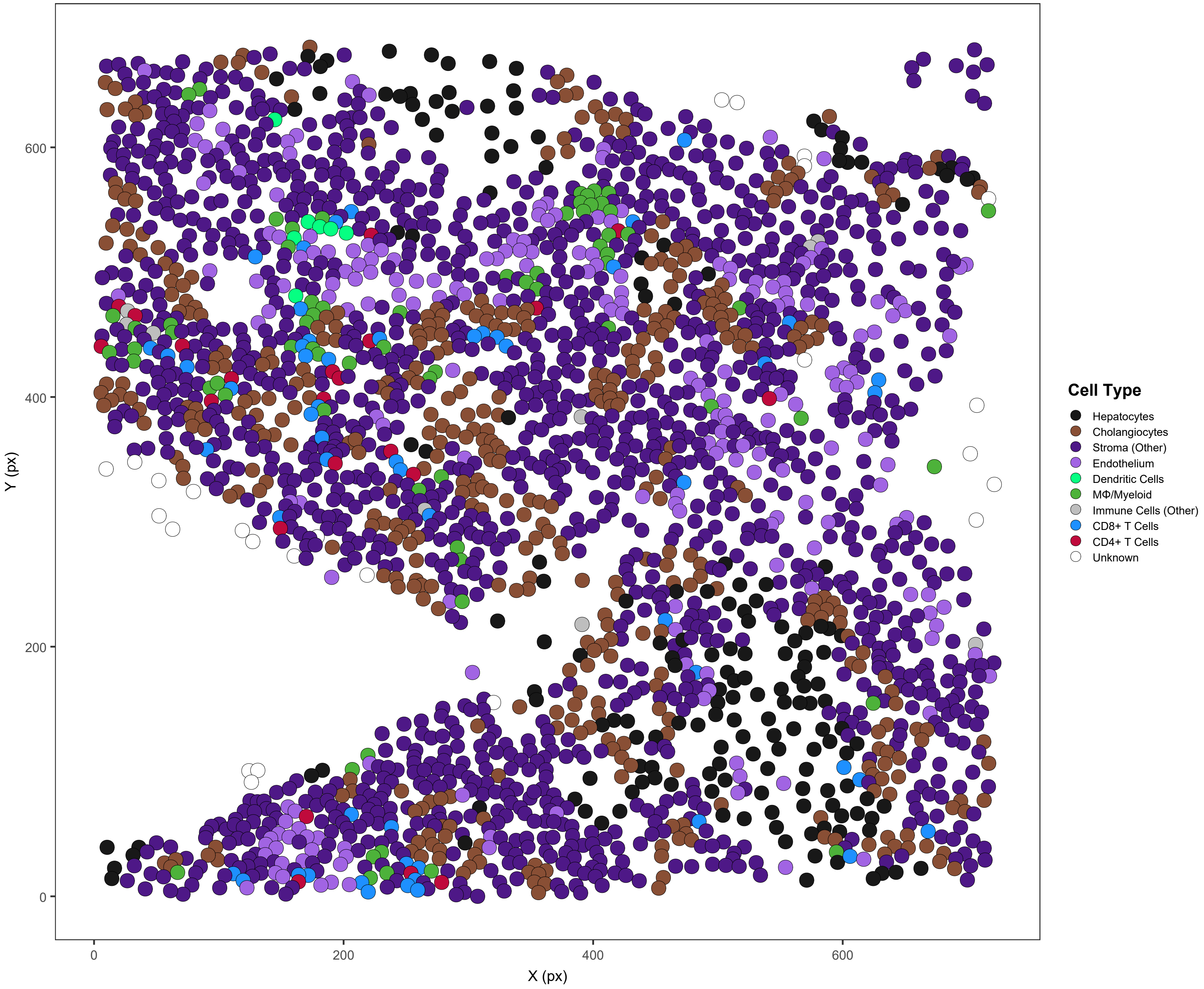

### ICI15_s0_a3_ac

ICI15 | 2080/2086 cells assigned (99.7%)

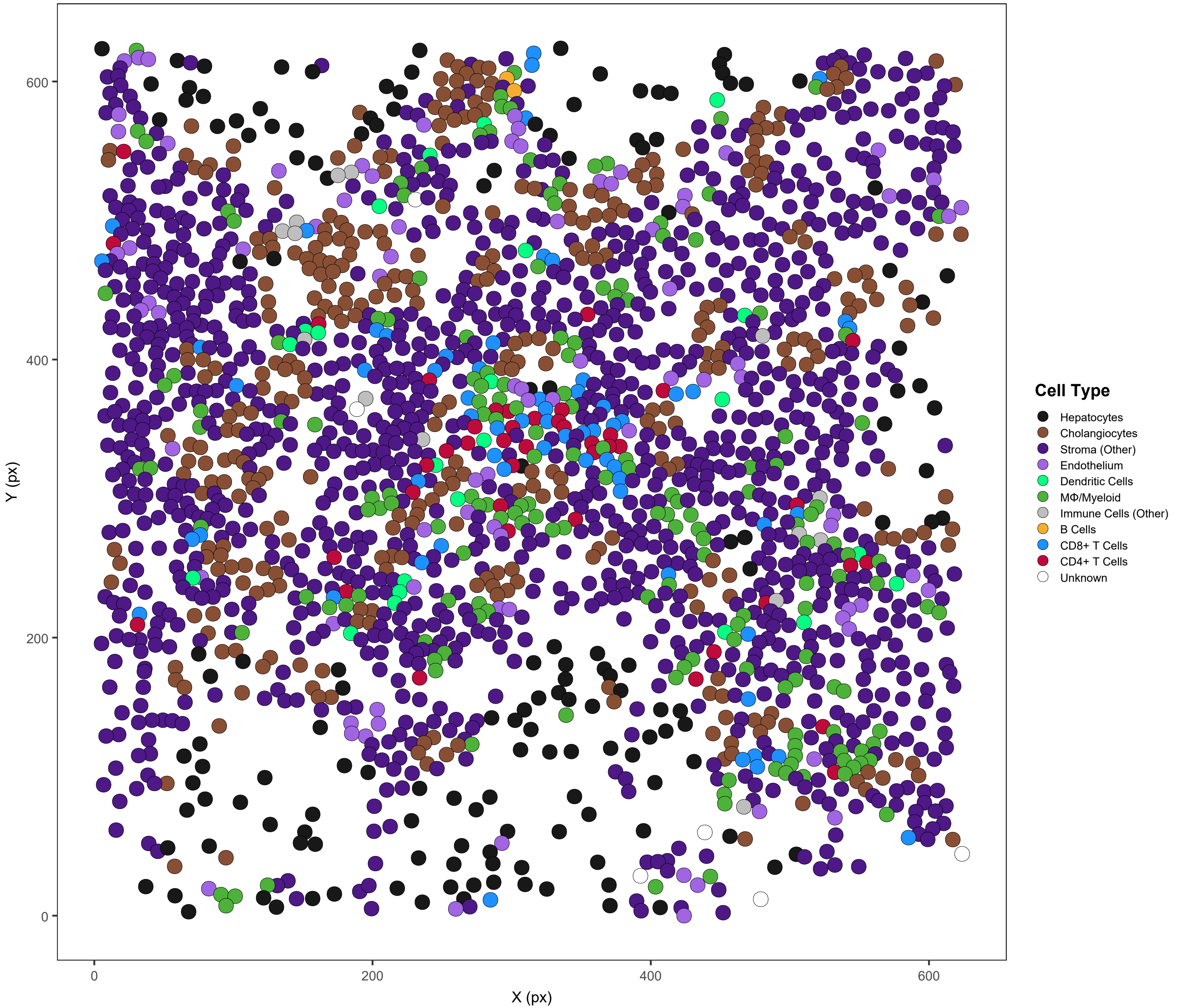

### ICI15_s0_a4_ac

ICI15 | 1559/1560 cells assigned (99.9%)

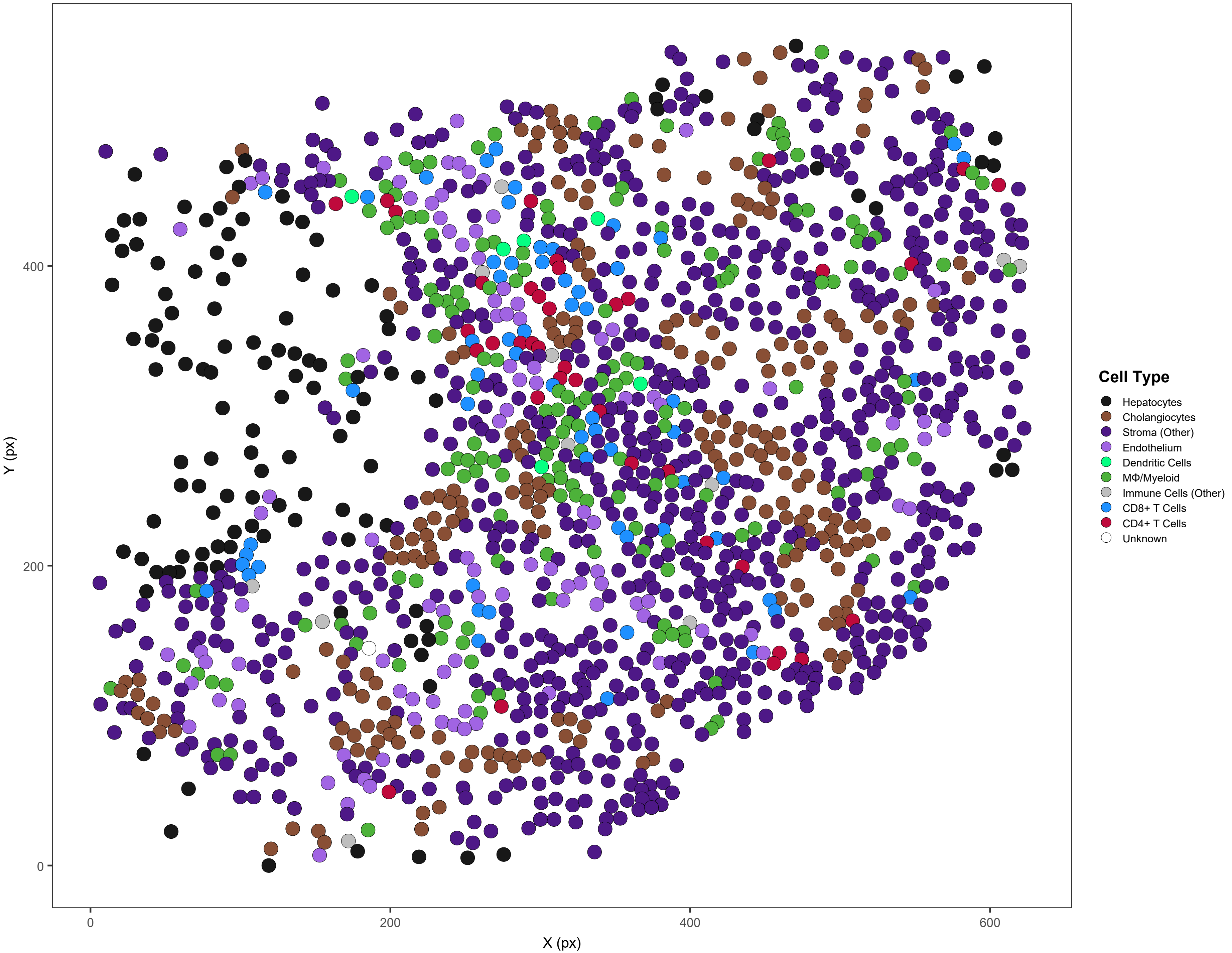

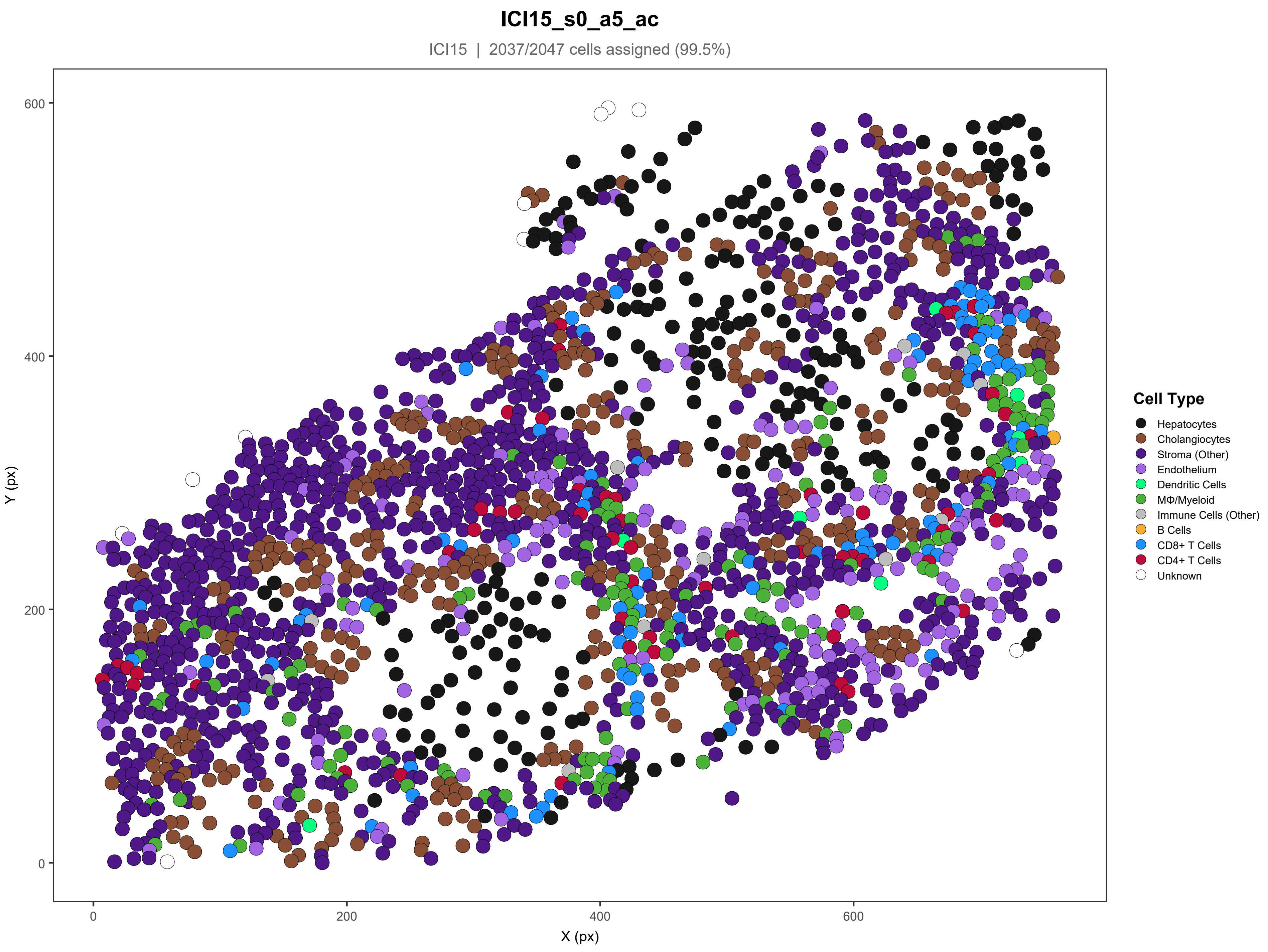

ICI15_s0_a5_ac
ICI15 | 2037/2047 cells assigned (99.5%)
Y (px)
X (px)
0
200
400
600
Cell Type
Hepatocytes
Cholangiocytes
Stroma (Other)
Endothelium
Dendritic Cells
MΦ/Myeloid
Immune Cells (Other)
B Cells
CD8+ T Cells
CD4+ T Cells
Unknown

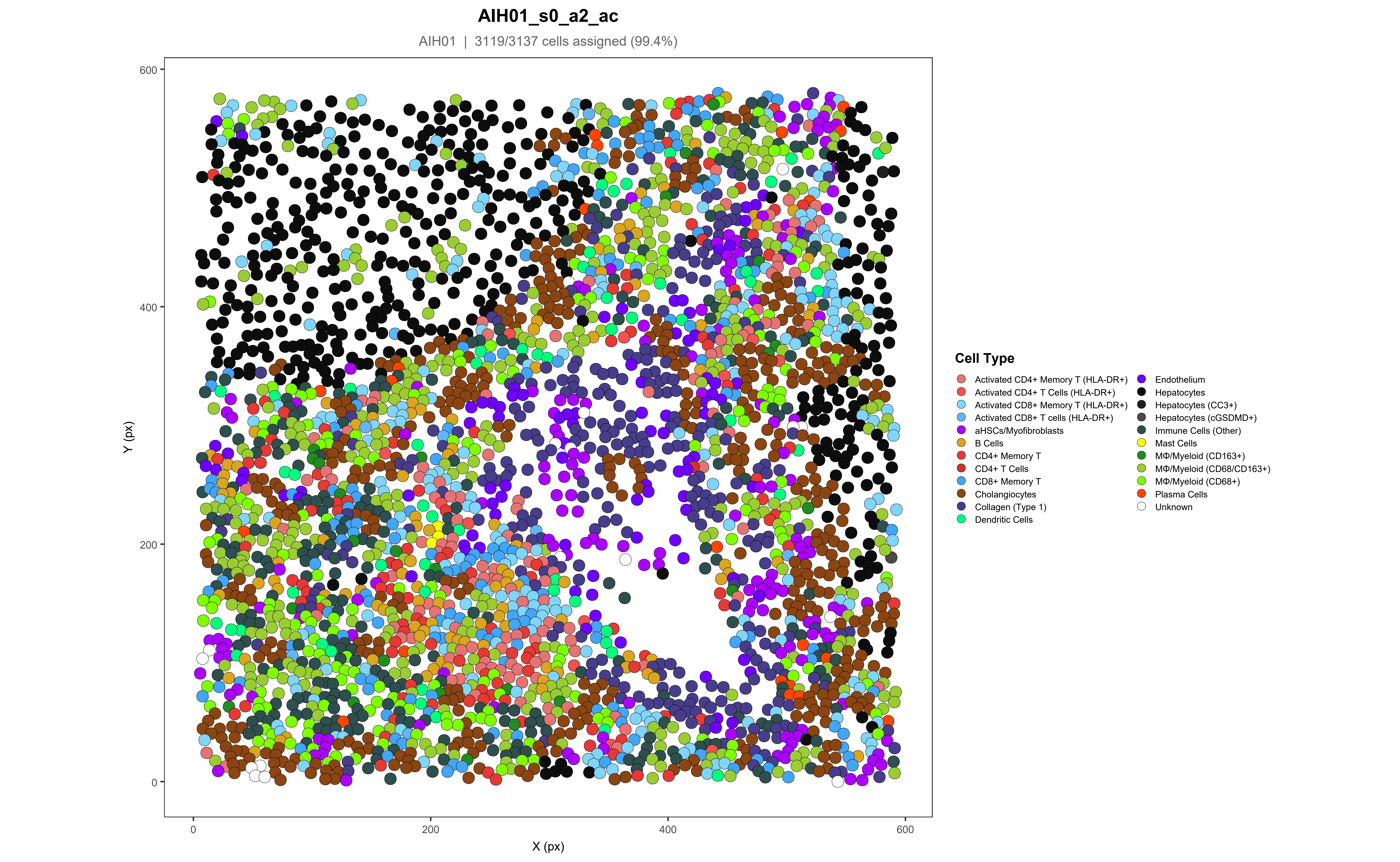

AIH01_s0_a2_ac
AIH01 | 3119/3137 cells assigned (99.4%)
Y (px)
X (px)
0
200
400
600
Cell Type
Activated CD4+ Memory T (HLA-DR+)
Activated CD4+ T Cells (HLA-DR+)
Activated CD8+ Memory T (HLA-DR+)
Activated CD8+ T cells (HLA-DR+)
aHSCs/Myofibroblasts
B Cells
CD4+ Memory T
CD4+ T Cells
CD8+ Memory T
Cholangiocytes
Collagen (Type 1)
Dendritic Cells
Endothelium
Hepatocytes
Hepatocytes (CC3+)
Hepatocytes (cGSDMD+)
Immune Cells (Other)
Mast Cells
MΦ/Myeloid (CD163+)
MΦ/Myeloid (CD68/CD163+)
MΦ/Myeloid (CD68+)
Plasma Cells
Unknown

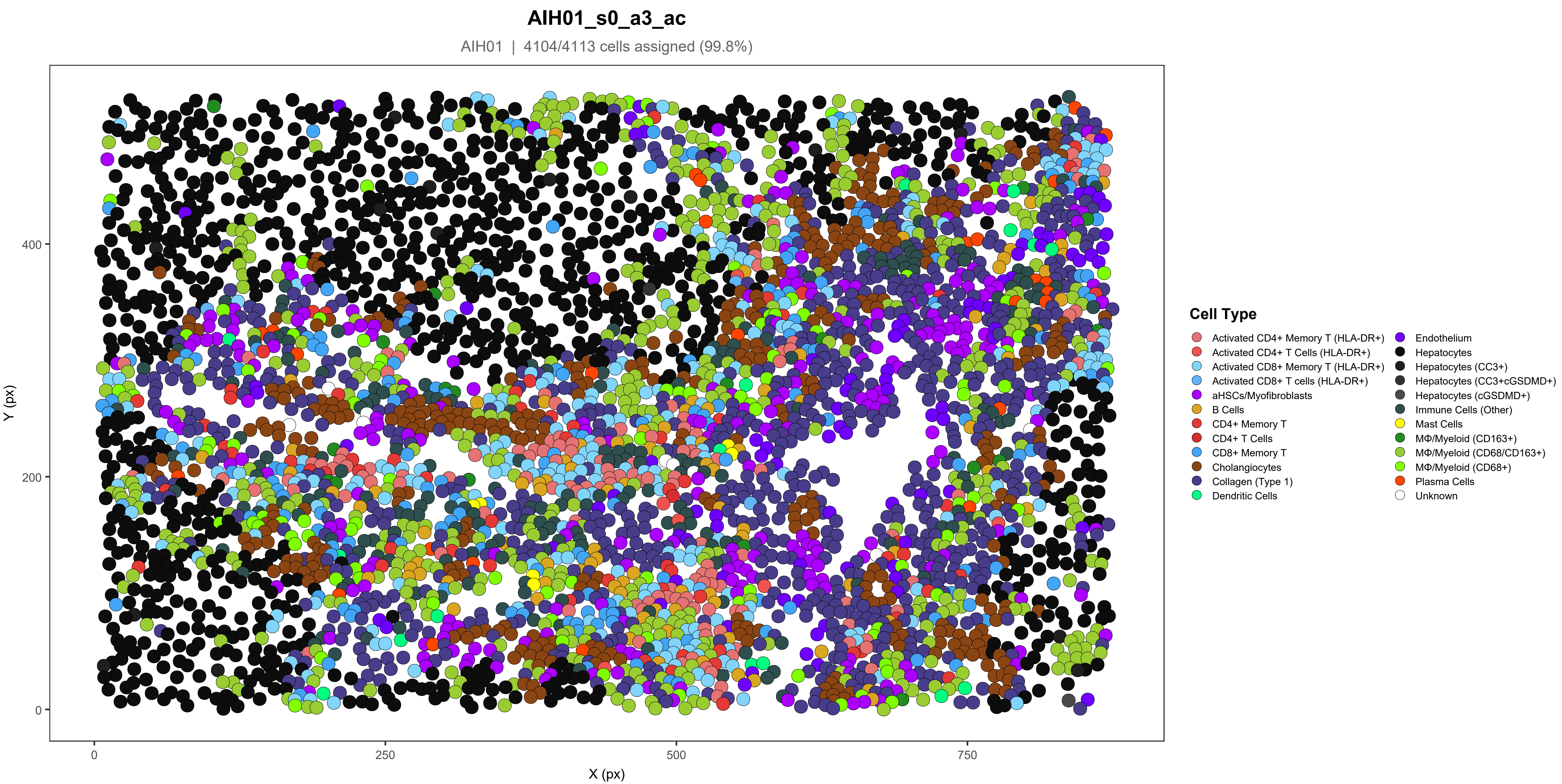

AIH01_s0_a3_ac
AIH01 | 4104/4113 cells assigned (99.8%)
Y (px)
X (px)
0
200
400
0
250
500
750
Cell Type
Activated CD4+ Memory T (HLA-DR+)
Activated CD4+ T Cells (HLA-DR+)
Activated CD8+ Memory T (HLA-DR+)
Activated CD8+ T cells (HLA-DR+)
aHSCs/Myofibroblasts
B Cells
CD4+ Memory T
CD4+ T Cells
CD8+ Memory T
Cholangiocytes
Collagen (Type 1)
Dendritic Cells
Endothelium
Hepatocytes
Hepatocytes (CC3+)
Hepatocytes (CC3+cGSDMD+)
Hepatocytes (cGSDMD+)
Immune Cells (Other)
Mast Cells
MΦ/Myeloid (CD163+)
MΦ/Myeloid (CD68/CD163+)
MΦ/Myeloid (CD68+)
Plasma Cells
Unknown

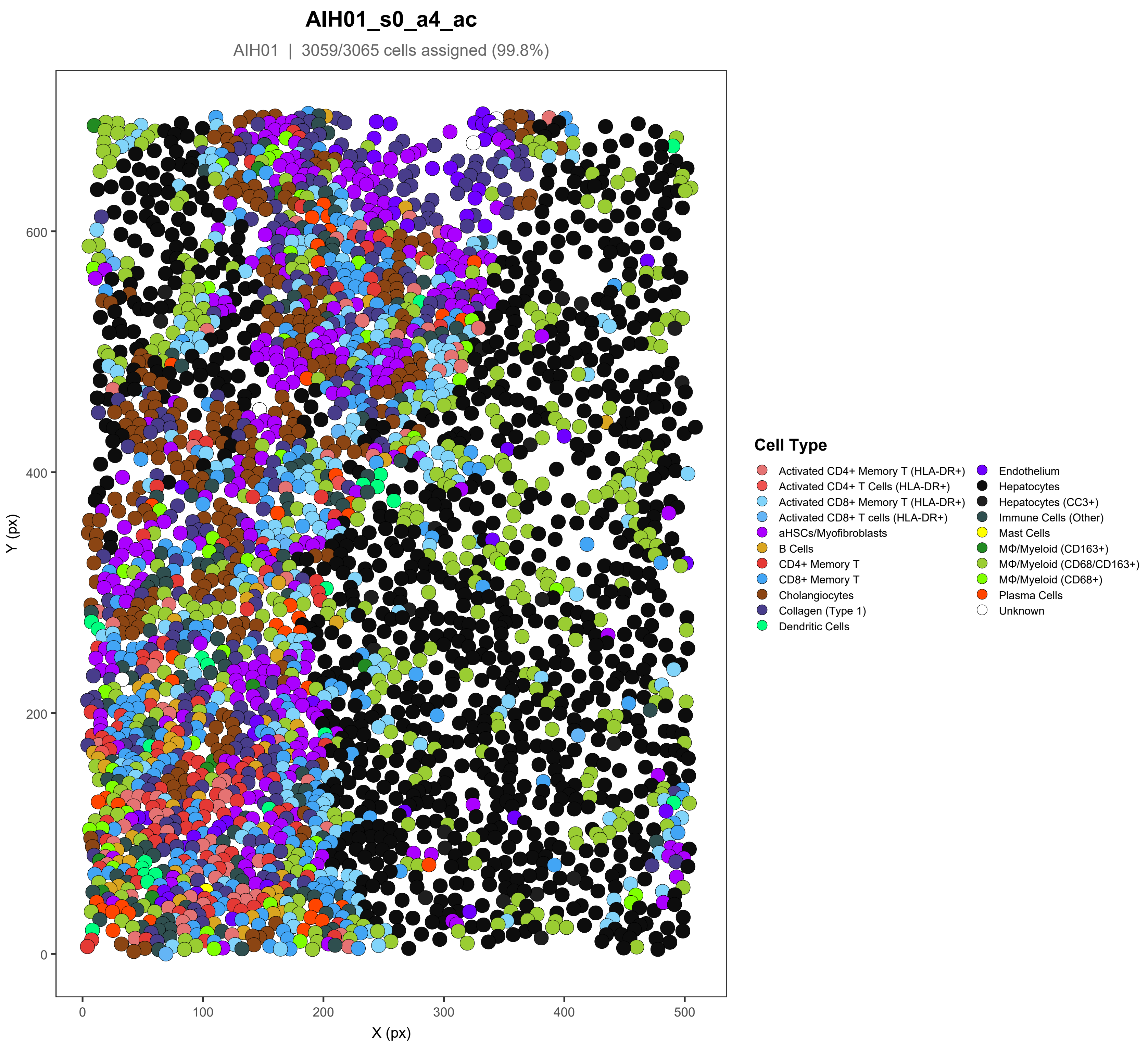
AIH01_s0_a4_ac
AIH01 | 3059/3065 cells assigned (99.8%)
Y (px)
X (px)
0
100
200
300
400
500
600
Cell Type
Activated CD4+ Memory T (HLA-DR+)
Activated CD4+ T Cells (HLA-DR+)
Activated CD8+ Memory T (HLA-DR+)
Activated CD8+ T cells (HLA-DR+)
aHSCs/Myofibroblasts
B Cells
CD4+ Memory T
CD8+ Memory T
Cholangiocytes
Collagen (Type 1)
Dendritic Cells
Endothelium
Hepatocytes
Hepatocytes (CC3+)
Immune Cells (Other)
Mast Cells
MΦ/Myeloid (CD163+)
MΦ/Myeloid (CD68/CD163+)
MΦ/Myeloid (CD68+)
Plasma Cells
Unknown

**AIH01_s0_a5_ac**

AIH01 | 3463/3478 cells assigned (99.6%)

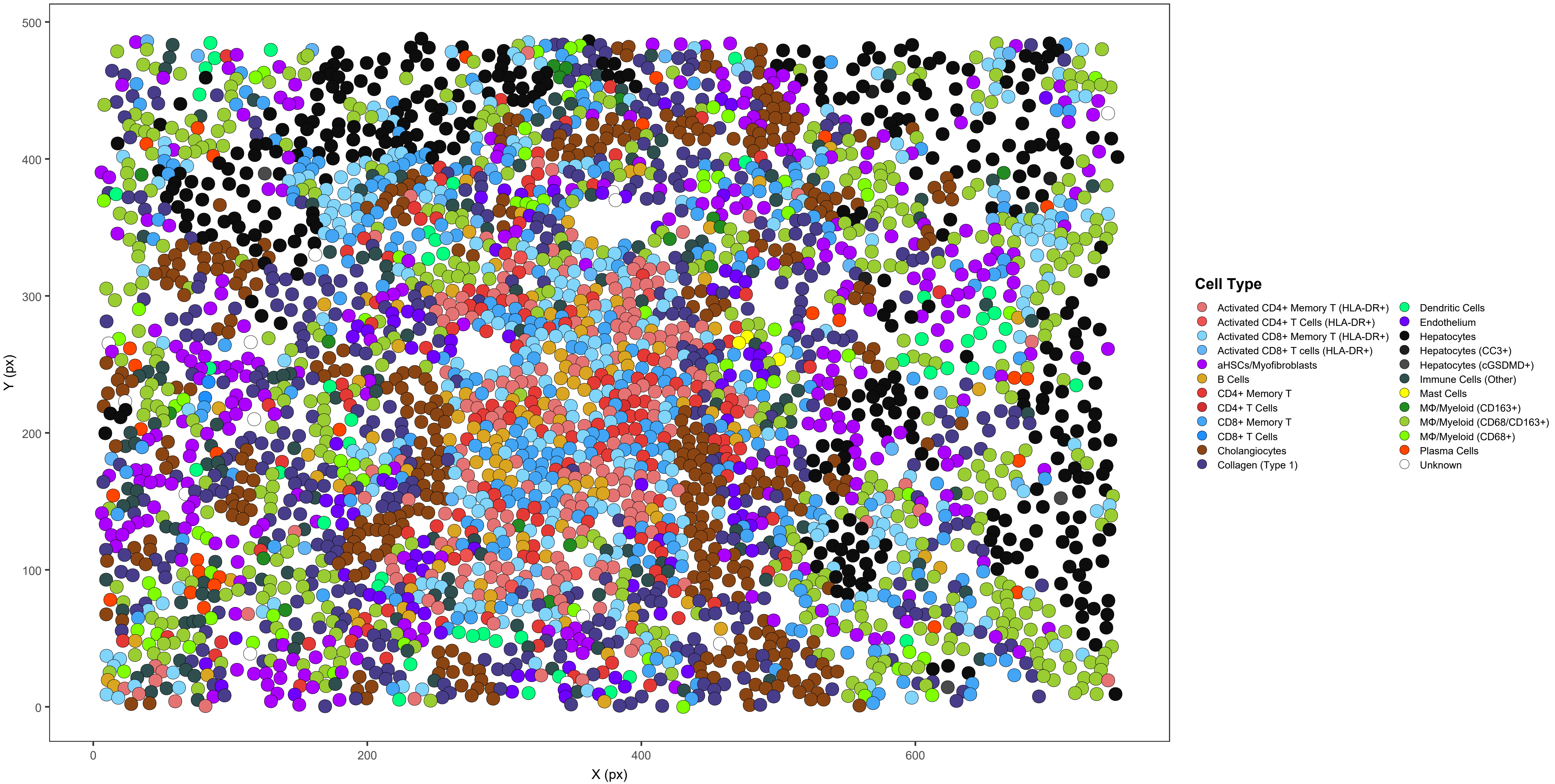

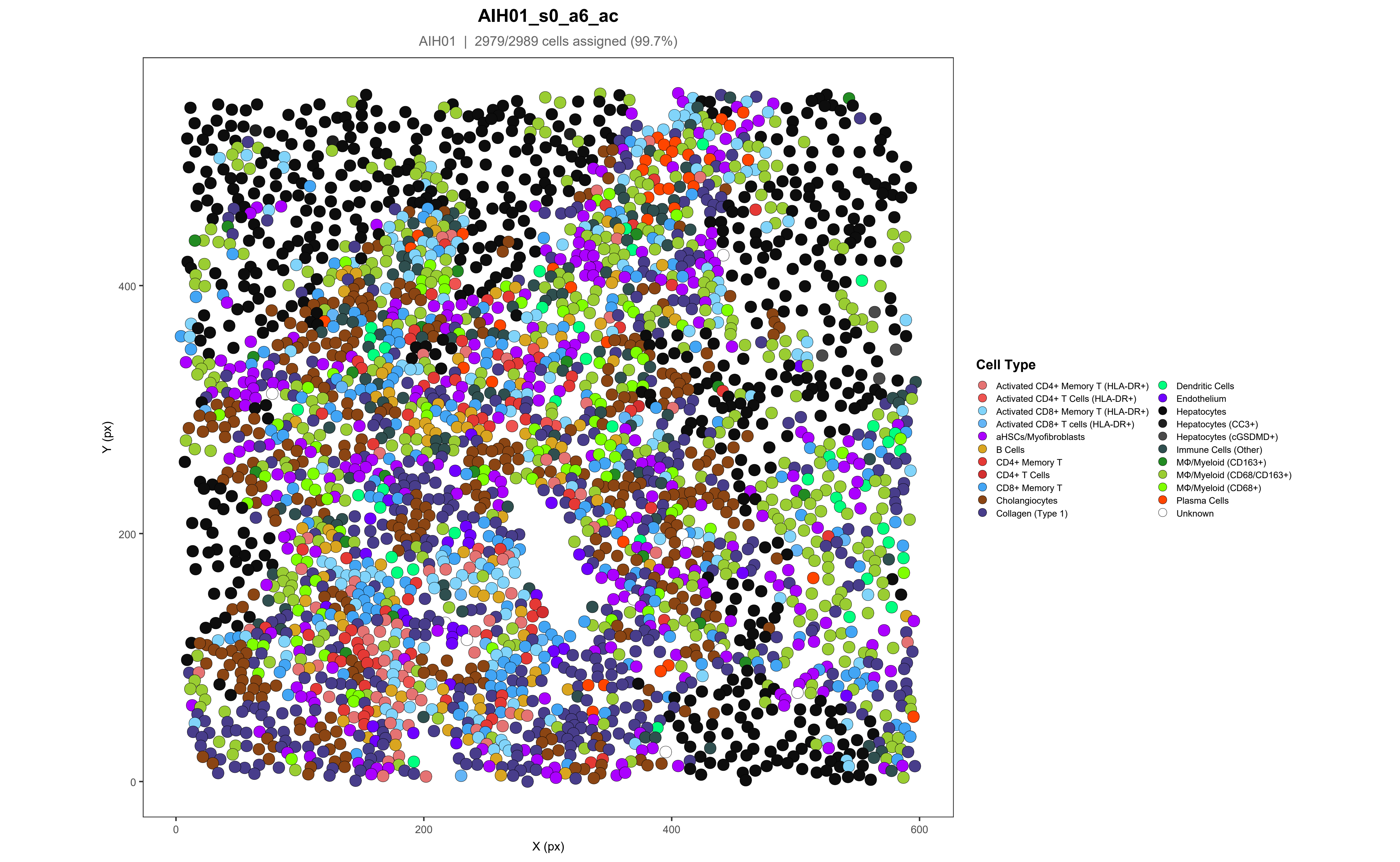

AIH01_s0_a6_ac
AIH01 | 2979/2989 cells assigned (99.7%)
Y (px)
X (px)
0
200
400
600
Cell Type
Activated CD4+ Memory T (HLA-DR+)
Activated CD4+ T Cells (HLA-DR+)
Activated CD8+ Memory T (HLA-DR+)
Activated CD8+ T cells (HLA-DR+)
aHSCs/Myofibroblasts
B Cells
CD4+ Memory T
CD4+ T Cells
CD8+ Memory T
Cholangiocytes
Collagen (Type 1)
Dendritic Cells
Endothelium
Hepatocytes
Hepatocytes (CC3+)
Hepatocytes (cGSDMD+)
Immune Cells (Other)
MΦ/Myeloid (CD163+)
MΦ/Myeloid (CD68/CD163+)
MΦ/Myeloid (CD68+)
Plasma Cells
Unknown

**AIH02_s0_a1_ac**

AIH02 | 3358/3358 cells assigned (100%)

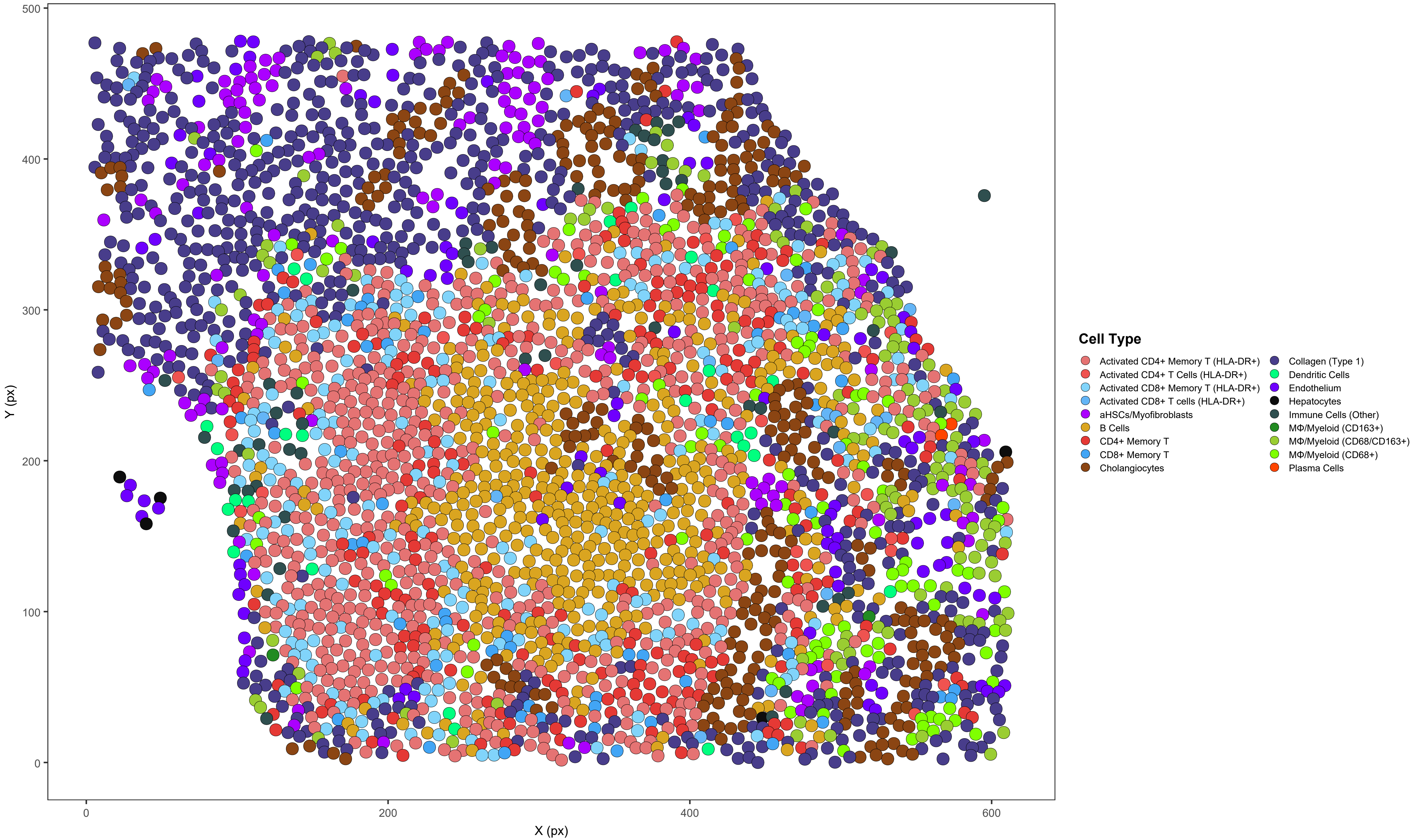

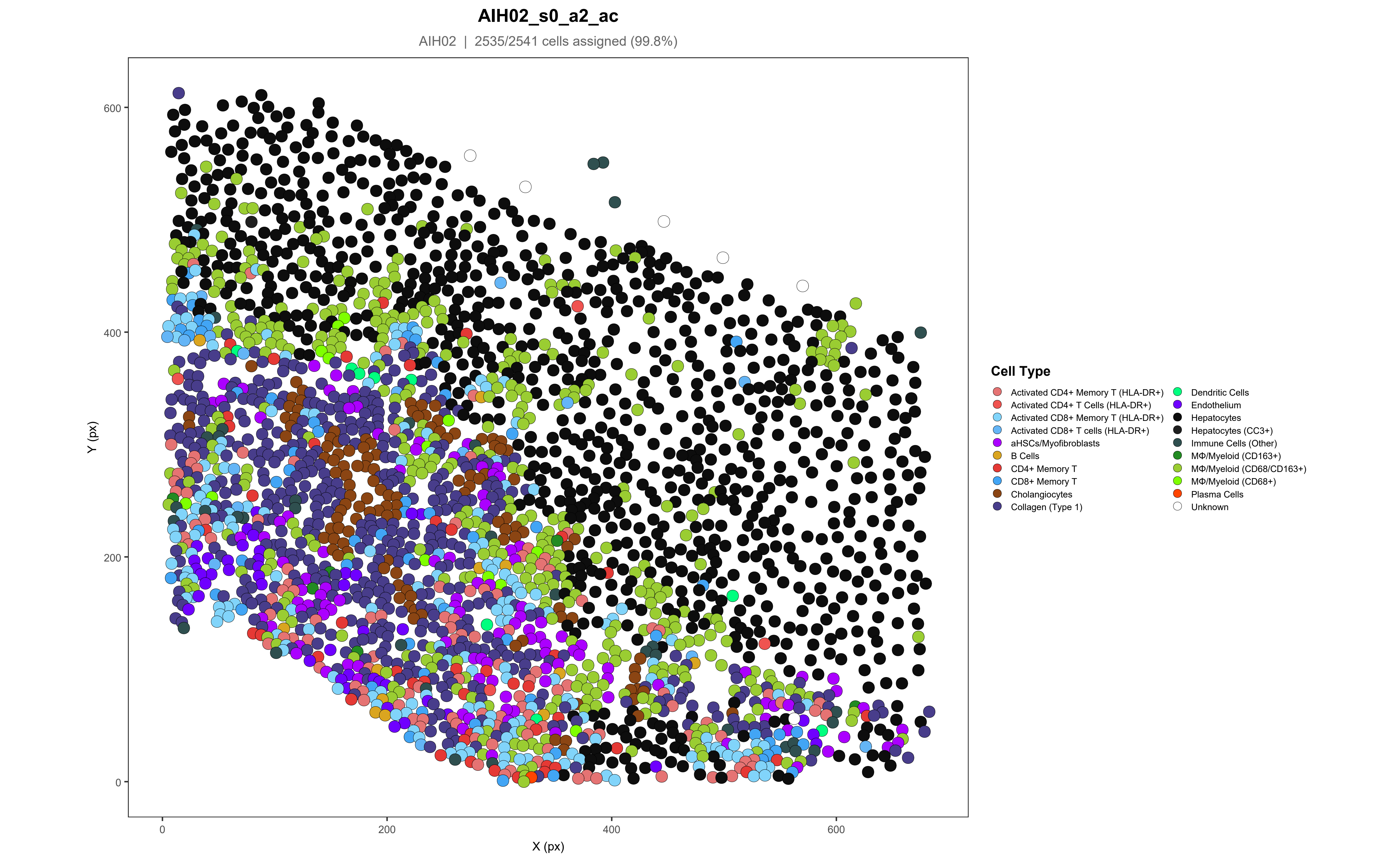

AIH02_s0_a2_ac
AIH02 | 2535/2541 cells assigned (99.8%)
Y (px)
X (px)
0
200
400
600
Cell Type
Activated CD4+ Memory T (HLA-DR+)
Activated CD4+ T Cells (HLA-DR+)
Activated CD8+ Memory T (HLA-DR+)
Activated CD8+ T cells (HLA-DR+)
aHSCs/Myofibroblasts
B Cells
CD4+ Memory T
CD8+ Memory T
Cholangiocytes
Collagen (Type 1)
Dendritic Cells
Endothelium
Hepatocytes
Hepatocytes (CC3+)
Immune Cells (Other)
MΦ/Myeloid (CD163+)
MΦ/Myeloid (CD68/CD163+)
MΦ/Myeloid (CD68+)
Plasma Cells
Unknown

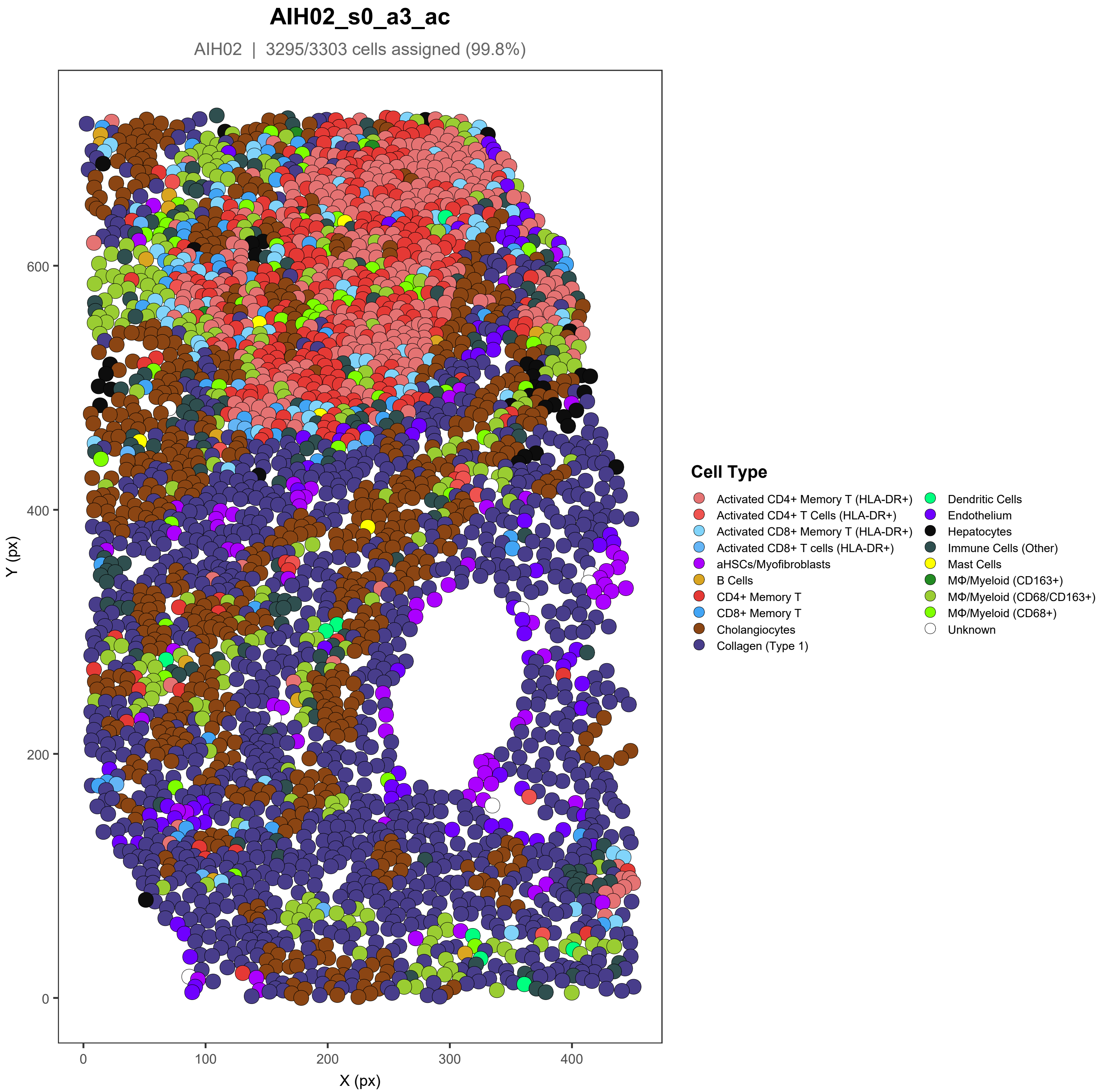

AIH02_s0_a3_ac
AIH02 | 3295/3303 cells assigned (99.8%)
Y (px)
X (px)
0
200
400
600
0
100
200
300
400
Cell Type
Activated CD4+ Memory T (HLA-DR+)
Activated CD4+ T Cells (HLA-DR+)
Activated CD8+ Memory T (HLA-DR+)
Activated CD8+ T cells (HLA-DR+)
aHSCs/Myofibroblasts
B Cells
CD4+ Memory T
CD8+ Memory T
Cholangiocytes
Collagen (Type 1)
Dendritic Cells
Endothelium
Hepatocytes
Immune Cells (Other)
Mast Cells
MΦ/Myeloid (CD163+)
MΦ/Myeloid (CD68/CD163+)
MΦ/Myeloid (CD68+)
Unknown

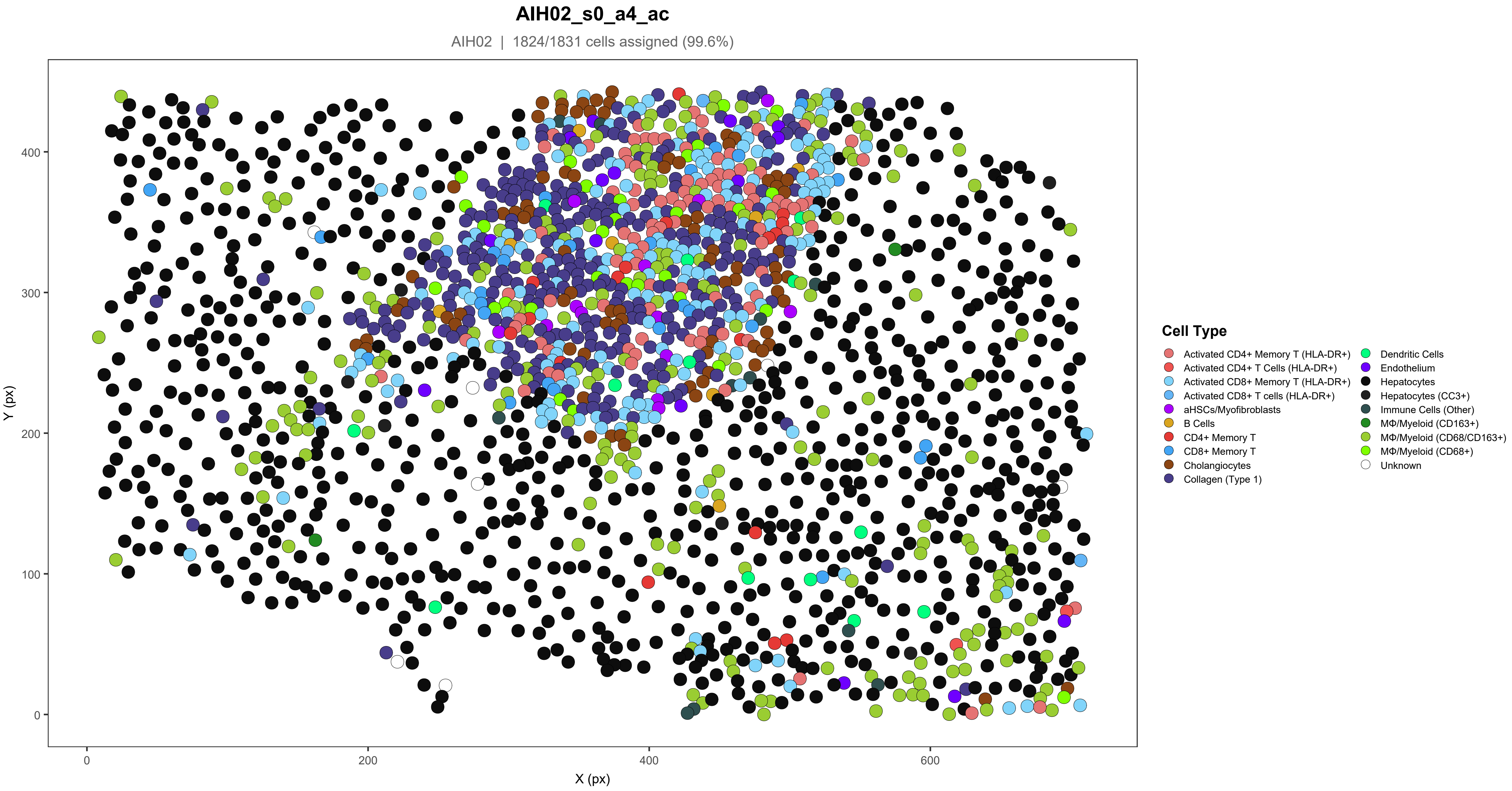
AIH02_s0_a4_ac
AIH02 | 1824/1831 cells assigned (99.6%)
Y (px)
X (px)
0
100
200
300
400
600
Cell Type
Activated CD4+ Memory T (HLA-DR+)
Activated CD4+ T Cells (HLA-DR+)
Activated CD8+ Memory T (HLA-DR+)
Activated CD8+ T cells (HLA-DR+)
aHSCs/Myofibroblasts
B Cells
CD4+ Memory T
CD8+ Memory T
Cholangiocytes
Collagen (Type 1)
Dendritic Cells
Endothelium
Hepatocytes
Hepatocytes (CC3+)
Immune Cells (Other)
MΦ/Myeloid (CD163+)
MΦ/Myeloid (CD68/CD163+)
MΦ/Myeloid (CD68+)
Unknown

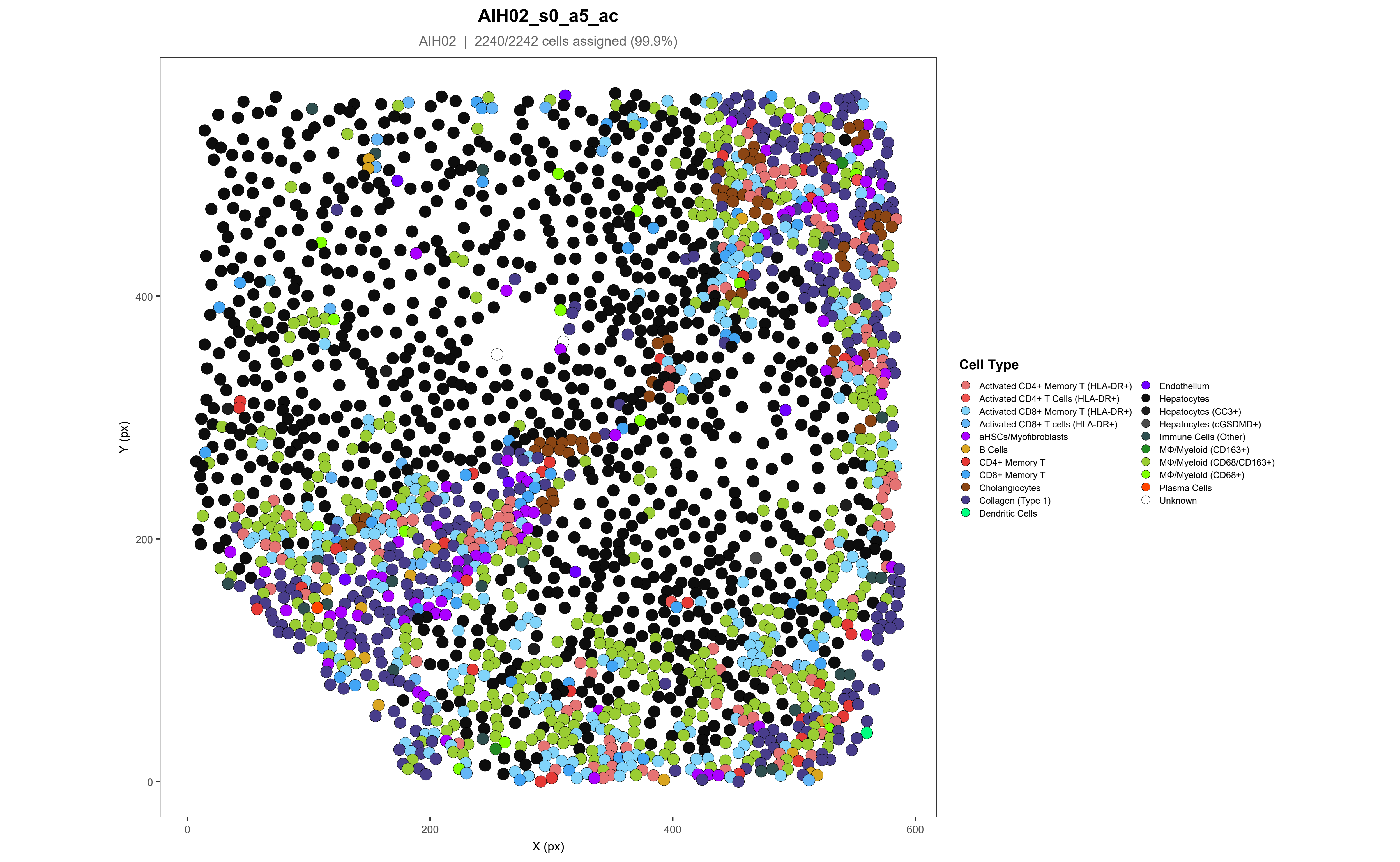
AIH02_s0_a5_ac
AIH02 | 2240/2242 cells assigned (99.9%)
Y (px)
X (px)
0
200
400
600
Cell Type
Activated CD4+ Memory T (HLA-DR+)
Activated CD4+ T Cells (HLA-DR+)
Activated CD8+ Memory T (HLA-DR+)
Activated CD8+ T cells (HLA-DR+)
aHSCs/Myofibroblasts
B Cells
CD4+ Memory T
CD8+ Memory T
Cholangiocytes
Collagen (Type 1)
Dendritic Cells
Endothelium
Hepatocytes
Hepatocytes (CC3+)
Hepatocytes (cGSDMD+)
Immune Cells (Other)
MΦ/Myeloid (CD163+)
MΦ/Myeloid (CD68/CD163+)
MΦ/Myeloid (CD68+)
Plasma Cells
Unknown

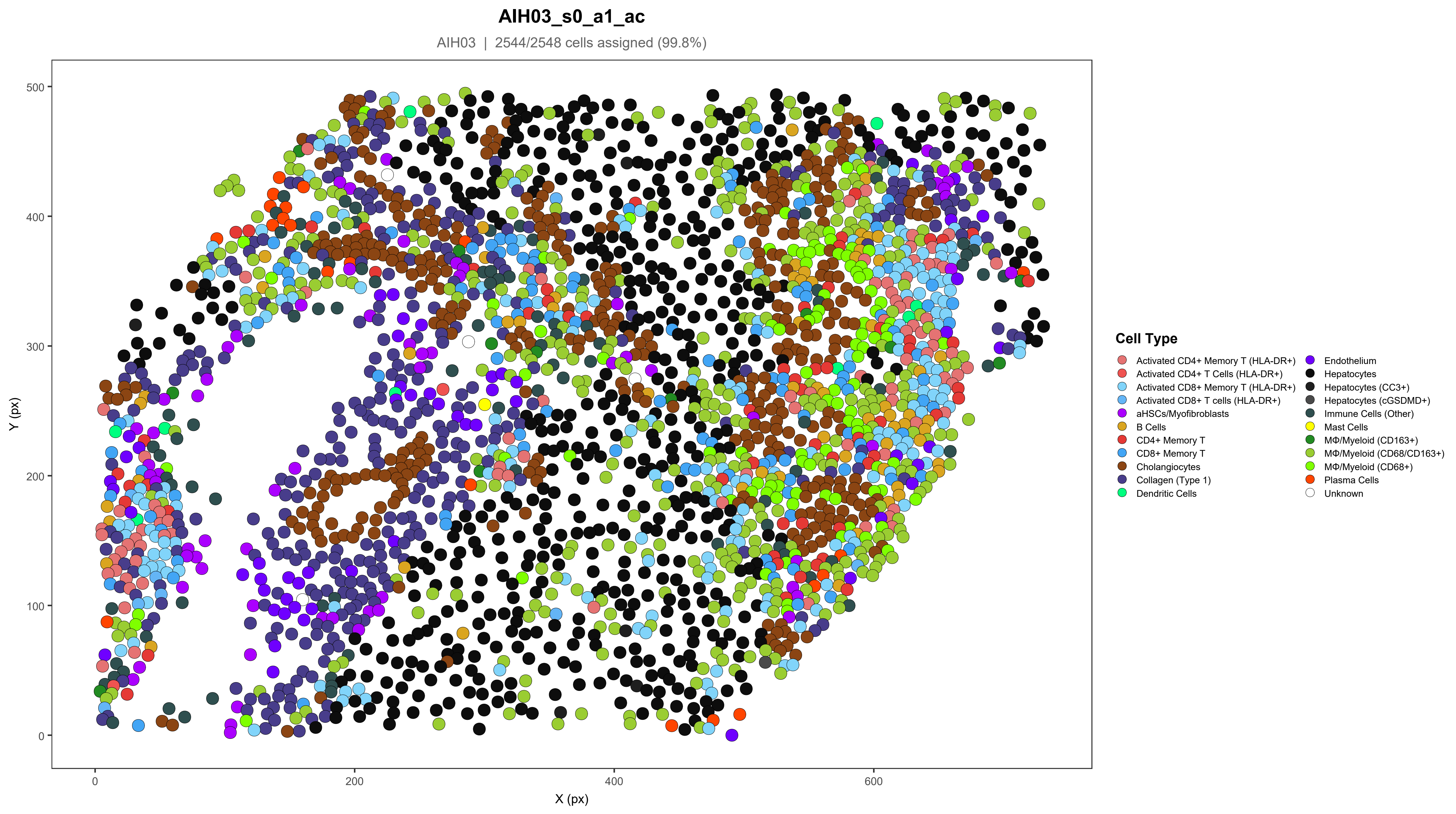
AIH03_s0_a1_ac
AIH03 | 2544/2548 cells assigned (99.8%)
Y (px)
X (px)
0
100
200
300
400
500
0
200
400
600
Cell Type
Activated CD4+ Memory T (HLA-DR+)
Activated CD4+ T Cells (HLA-DR+)
Activated CD8+ Memory T (HLA-DR+)
Activated CD8+ T cells (HLA-DR+)
aHSCs/Myofibroblasts
B Cells
CD4+ Memory T
CD8+ Memory T
Cholangiocytes
Collagen (Type 1)
Dendritic Cells
Endothelium
Hepatocytes
Hepatocytes (CC3+)
Hepatocytes (cGSDMD+)
Immune Cells (Other)
Mast Cells
MΦ/Myeloid (CD163+)
MΦ/Myeloid (CD68/CD163+)
MΦ/Myeloid (CD68+)
Plasma Cells
Unknown

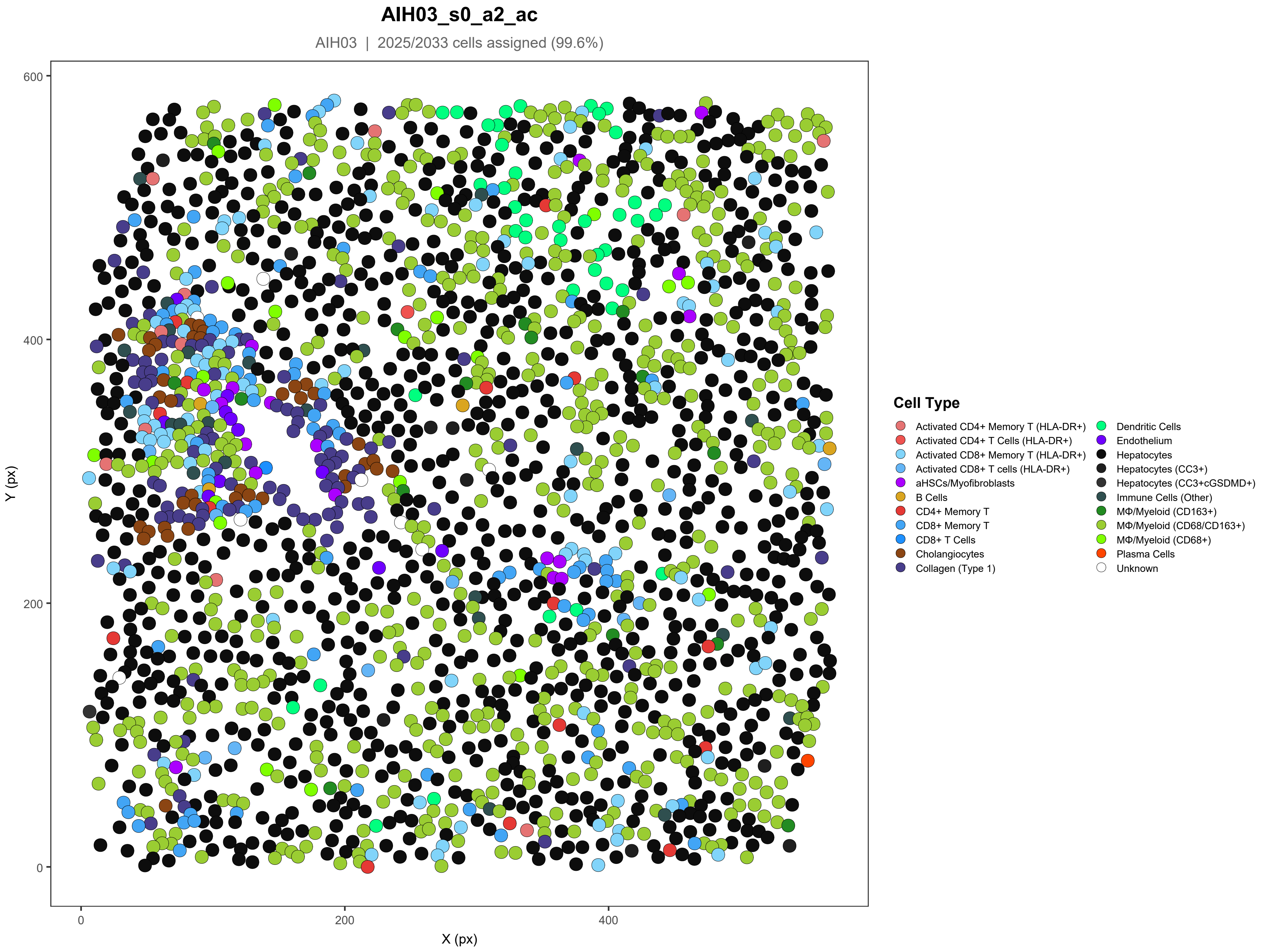
AIH03_s0_a2_ac
AIH03 | 2025/2033 cells assigned (99.6%)
Y (px)
X (px)
0
200
400
600
Cell Type
Activated CD4+ Memory T (HLA-DR+)
Activated CD4+ T Cells (HLA-DR+)
Activated CD8+ Memory T (HLA-DR+)
Activated CD8+ T cells (HLA-DR+)
aHSCs/Myofibroblasts
B Cells
CD4+ Memory T
CD8+ Memory T
CD8+ T Cells
Cholangiocytes
Collagen (Type 1)
Dendritic Cells
Endothelium
Hepatocytes
Hepatocytes (CC3+)
Hepatocytes (CC3+cGSDMD+)
Immune Cells (Other)
MΦ/Myeloid (CD163+)
MΦ/Myeloid (CD68/CD163+)
MΦ/Myeloid (CD68+)
Plasma Cells
Unknown

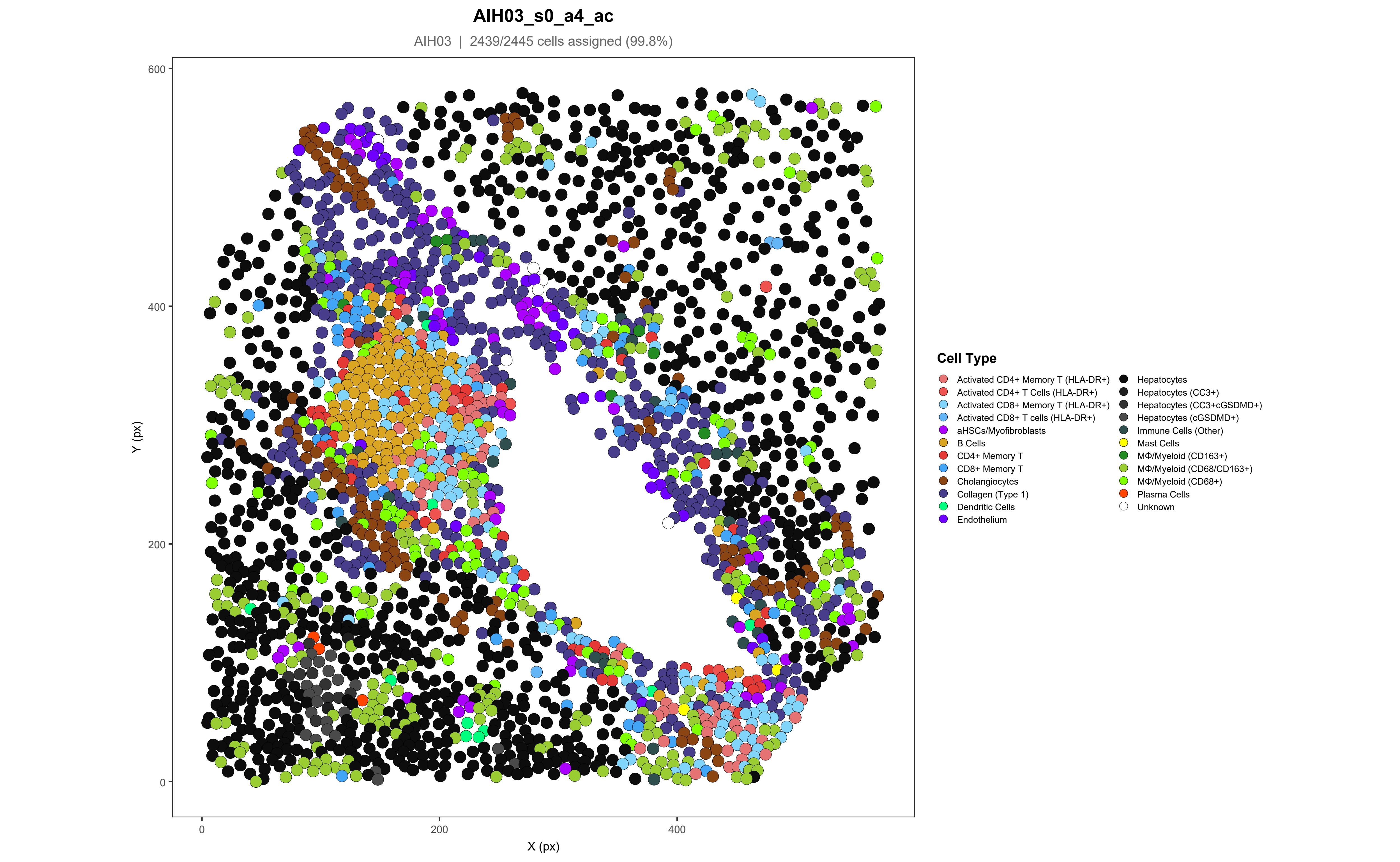
AIH03_s0_a4_ac
AIH03 | 2439/2445 cells assigned (99.8%)
600
400
200
0
Y (px)
0
200
400
X (px)
Cell Type
Activated CD4+ Memory T (HLA-DR+)
Activated CD4+ T Cells (HLA-DR+)
Activated CD8+ Memory T (HLA-DR+)
Activated CD8+ T cells (HLA-DR+)
aHSCs/Myofibroblasts
B Cells
CD4+ Memory T
CD8+ Memory T
Cholangiocytes
Collagen (Type 1)
Dendritic Cells
Endothelium
Hepatocytes
Hepatocytes (CC3+)
Hepatocytes (CC3+cGSDMD+)
Hepatocytes (cGSDMD+)
Immune Cells (Other)
Mast Cells
MΦ/Myeloid (CD163+)
MΦ/Myeloid (CD68/CD163+)
MΦ/Myeloid (CD68+)
Plasma Cells
Unknown

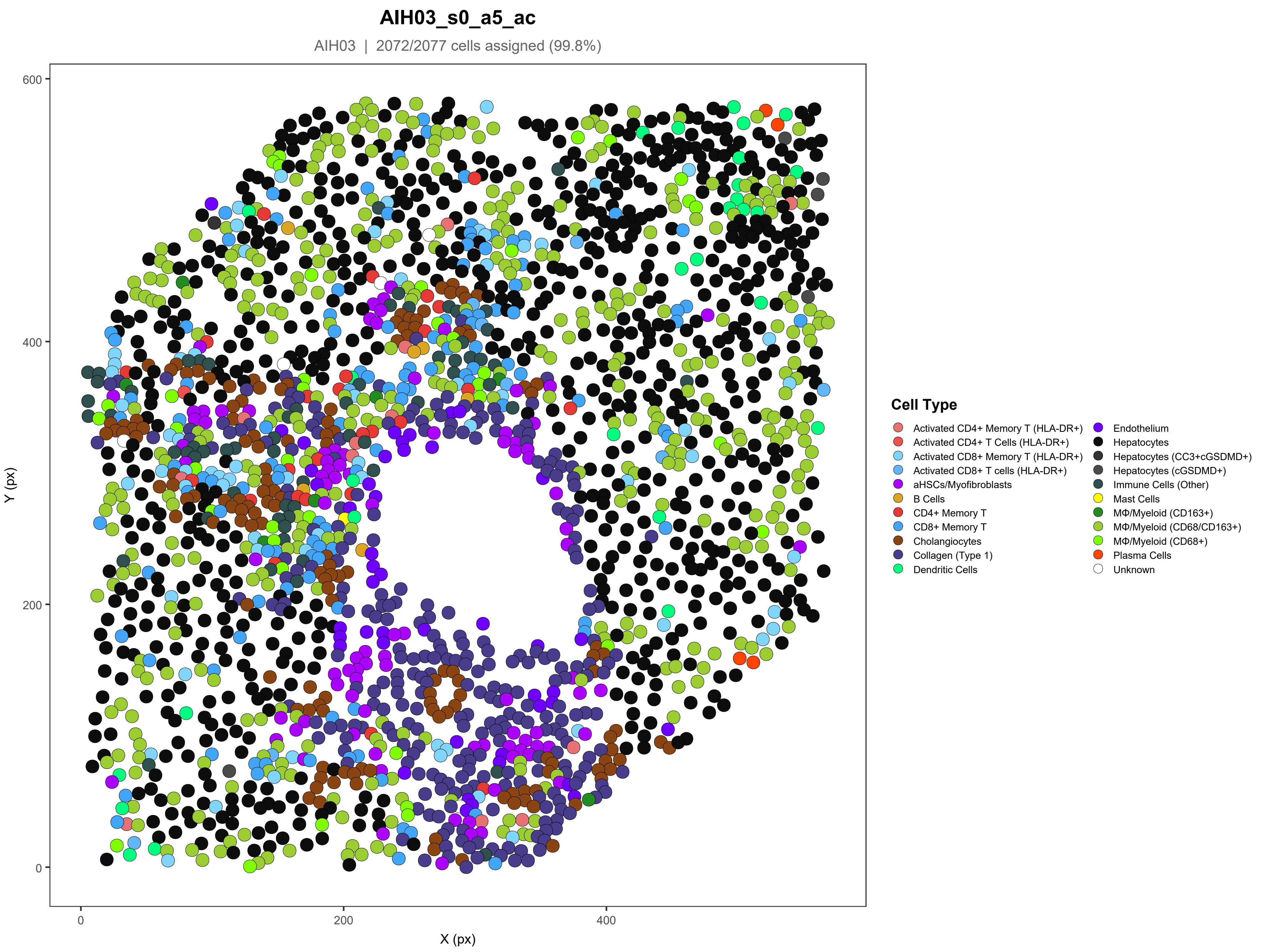

AIH03_s0_a5_ac
AIH03 | 2072/2077 cells assigned (99.8%)
Y (px)
X (px)
0
200
400
600
Cell Type
Activated CD4+ Memory T (HLA-DR+)
Activated CD4+ T Cells (HLA-DR+)
Activated CD8+ Memory T (HLA-DR+)
Activated CD8+ T cells (HLA-DR+)
aHSCs/Myofibroblasts
B Cells
CD4+ Memory T
CD8+ Memory T
Cholangiocytes
Collagen (Type 1)
Dendritic Cells
Endothelium
Hepatocytes
Hepatocytes (CC3+cGSDMD+)
Hepatocytes (cGSDMD+)
Immune Cells (Other)
Mast Cells
MΦ/Myeloid (CD163+)
MΦ/Myeloid (CD68/CD163+)
MΦ/Myeloid (CD68+)
Plasma Cells
Unknown

#### AIH04_s0_a1_ac

AIH04 | 2687/2699 cells assigned (99.6%)

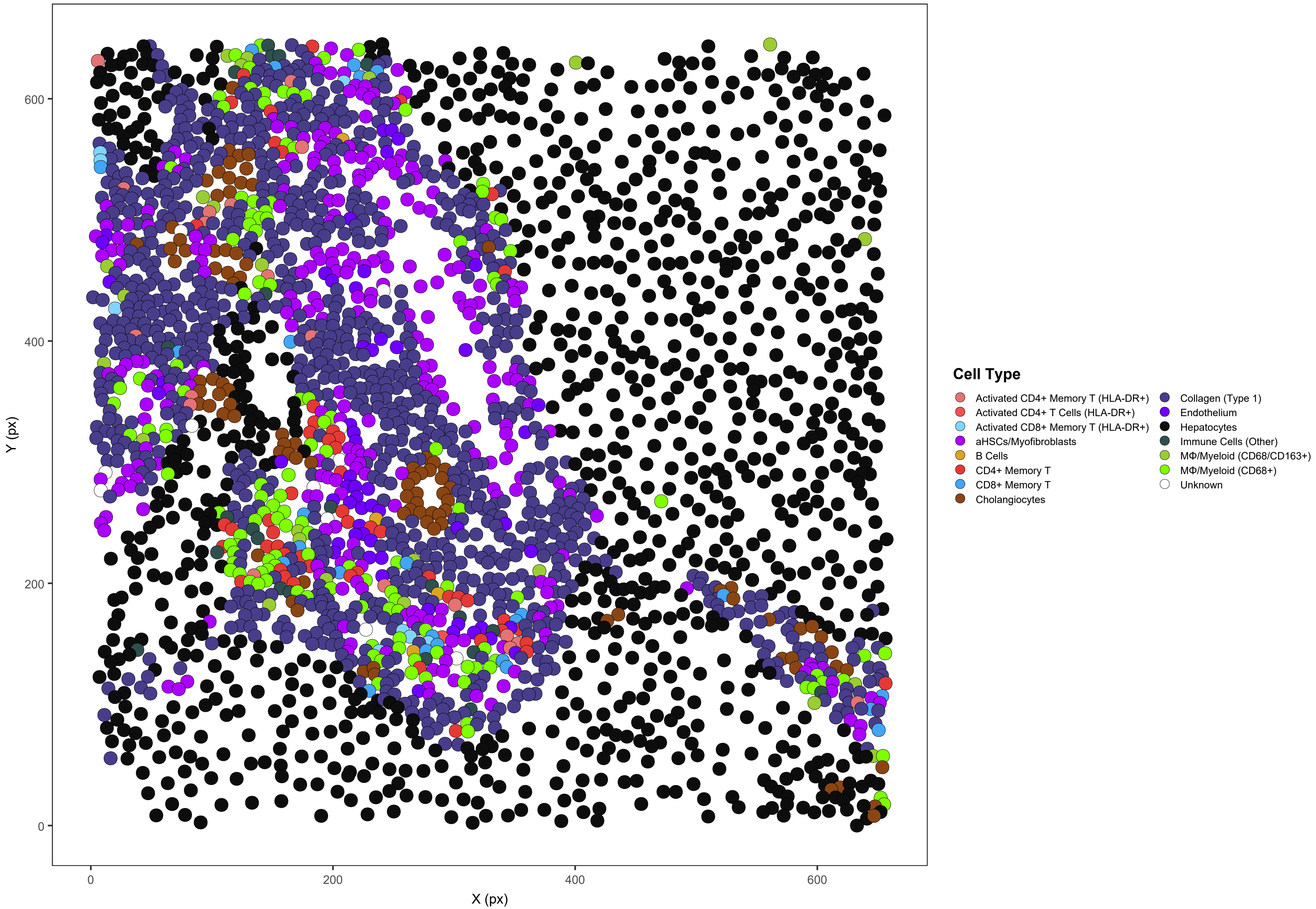

### AIH04_s0_a3_ac

AIH04 | 4320/4329 cells assigned (99.8%)

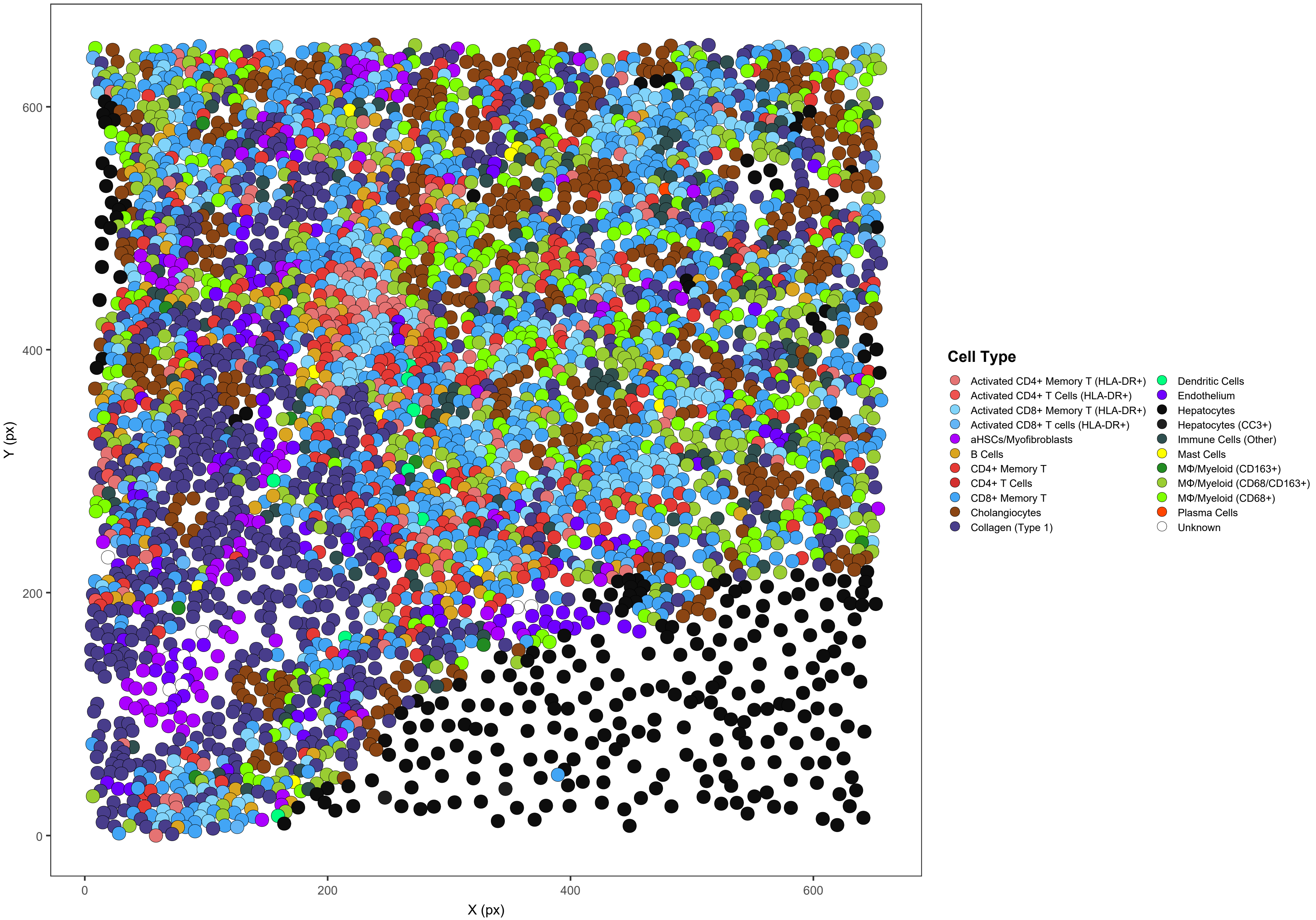

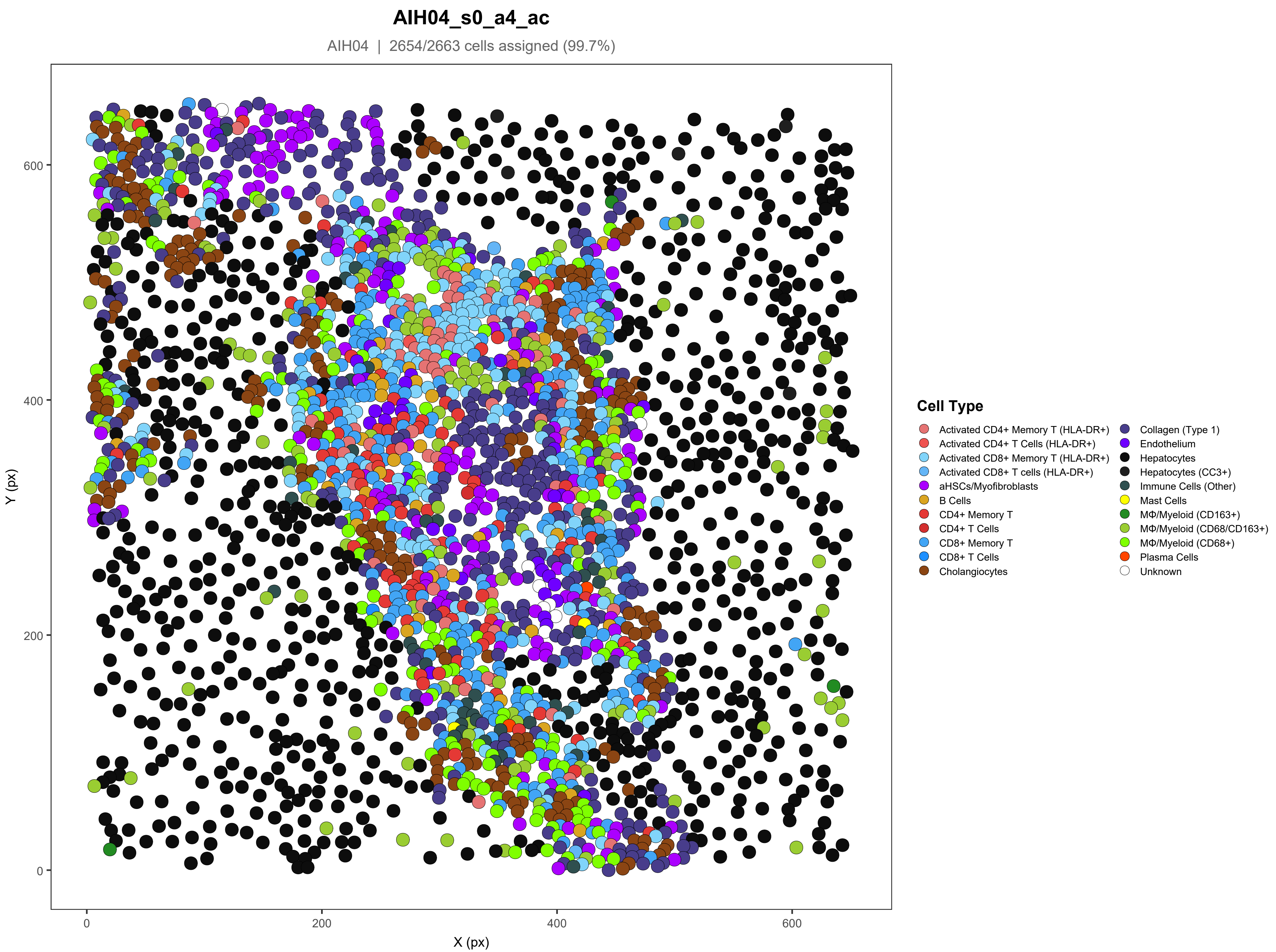

AIH04_s0_a4_ac
AIH04 | 2654/2663 cells assigned (99.7%)
Y (px)
X (px)
0
200
400
600
Cell Type
Activated CD4+ Memory T (HLA-DR+)
Activated CD4+ T Cells (HLA-DR+)
Activated CD8+ Memory T (HLA-DR+)
Activated CD8+ T cells (HLA-DR+)
aHSCs/Myofibroblasts
B Cells
CD4+ Memory T
CD4+ T Cells
CD8+ Memory T
CD8+ T Cells
Cholangiocytes
Collagen (Type 1)
Endothelium
Hepatocytes
Hepatocytes (CC3+)
Immune Cells (Other)
Mast Cells
MΦ/Myeloid (CD163+)
MΦ/Myeloid (CD68/CD163+)
MΦ/Myeloid (CD68+)
Plasma Cells
Unknown

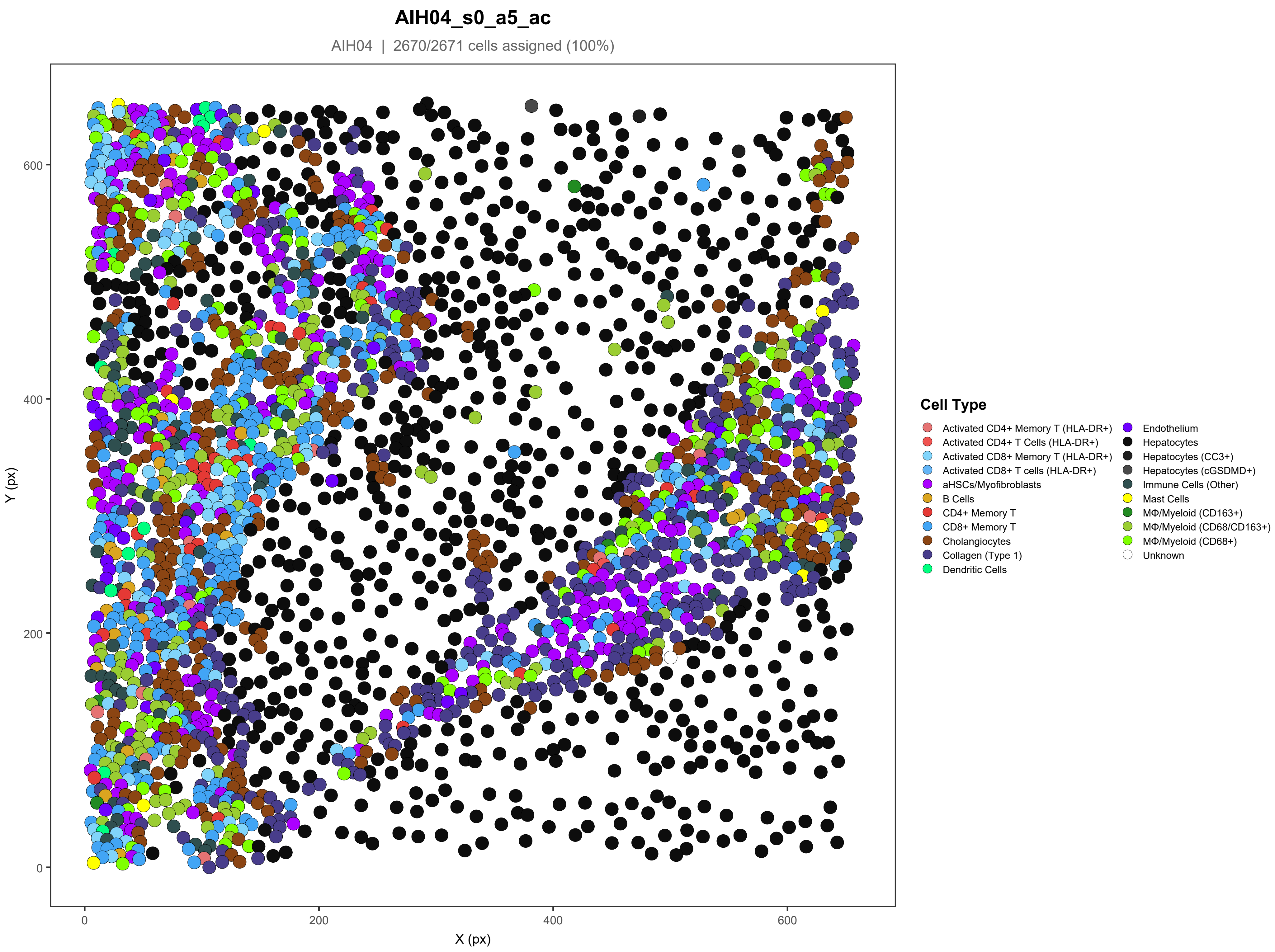
AIH04_s0_a5_ac
AIH04 | 2670/2671 cells assigned (100%)
Y (px)
X (px)
0
200
400
600
Cell Type
Activated CD4+ Memory T (HLA-DR+)
Activated CD4+ T Cells (HLA-DR+)
Activated CD8+ Memory T (HLA-DR+)
Activated CD8+ T cells (HLA-DR+)
aHSCs/Myofibroblasts
B Cells
CD4+ Memory T
CD8+ Memory T
Cholangiocytes
Collagen (Type 1)
Dendritic Cells
Endothelium
Hepatocytes
Hepatocytes (CC3+)
Hepatocytes (cGSDMD+)
Immune Cells (Other)
Mast Cells
MΦ/Myeloid (CD163+)
MΦ/Myeloid (CD68/CD163+)
MΦ/Myeloid (CD68+)
Unknown

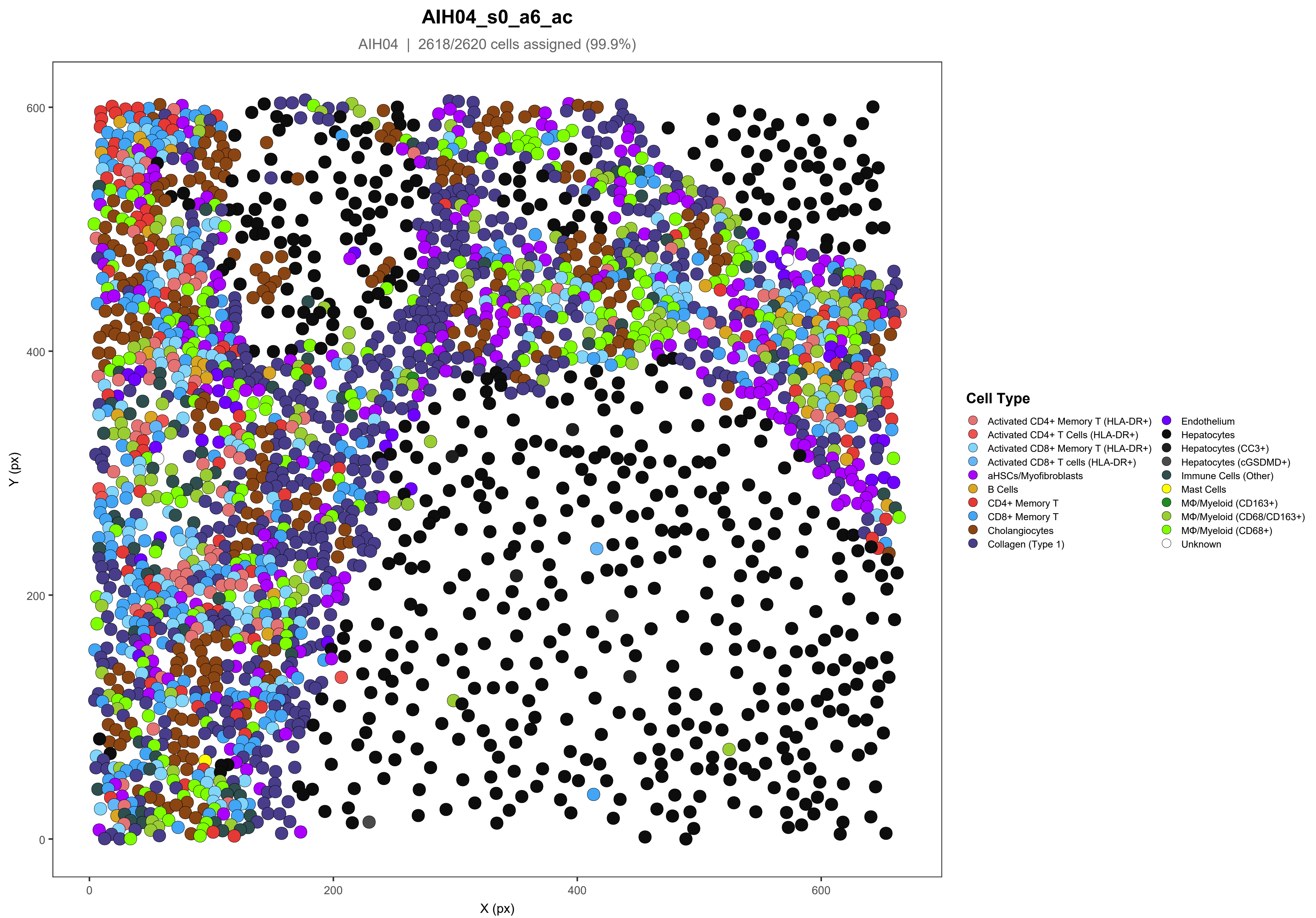

AIH04_s0_a6_ac
AIH04 | 2618/2620 cells assigned (99.9%)
Y (px)
X (px)
0
200
400
600
Cell Type
Activated CD4+ Memory T (HLA-DR+)
Activated CD4+ T Cells (HLA-DR+)
Activated CD8+ Memory T (HLA-DR+)
Activated CD8+ T cells (HLA-DR+)
aHSCs/Myofibroblasts
B Cells
CD4+ Memory T
CD8+ Memory T
Cholangiocytes
Collagen (Type 1)
Endothelium
Hepatocytes
Hepatocytes (CC3+)
Hepatocytes (cGSDMD+)
Immune Cells (Other)
Mast Cells
MΦ/Myeloid (CD163+)
MΦ/Myeloid (CD68/CD163+)
MΦ/Myeloid (CD68+)
Unknown

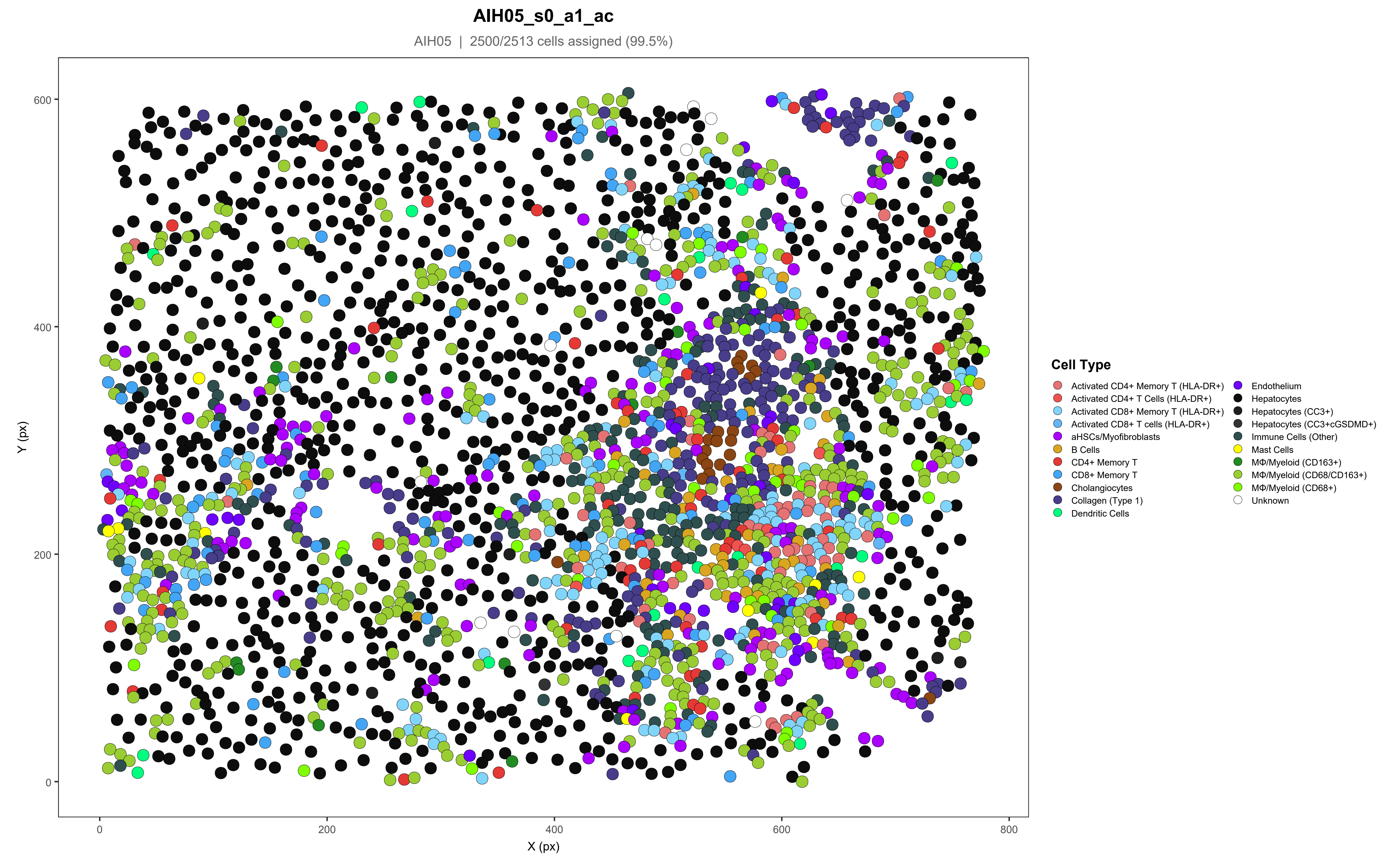
AIH05_s0_a1_ac
AIH05 | 2500/2513 cells assigned (99.5%)
Y (px)
X (px)
0
200
400
600
800
Cell Type
Activated CD4+ Memory T (HLA-DR+)
Activated CD4+ T Cells (HLA-DR+)
Activated CD8+ Memory T (HLA-DR+)
Activated CD8+ T cells (HLA-DR+)
aHSCs/Myofibroblasts
B Cells
CD4+ Memory T
CD8+ Memory T
Cholangiocytes
Collagen (Type 1)
Dendritic Cells
Endothelium
Hepatocytes
Hepatocytes (CC3+)
Hepatocytes (CC3+cGSDMD+)
Immune Cells (Other)
Mast Cells
MΦ/Myeloid (CD163+)
MΦ/Myeloid (CD68/CD163+)
MΦ/Myeloid (CD68+)
Unknown

### AIH05_s0_a2_ac

AIH05 | 1865/1889 cells assigned (98.7%)

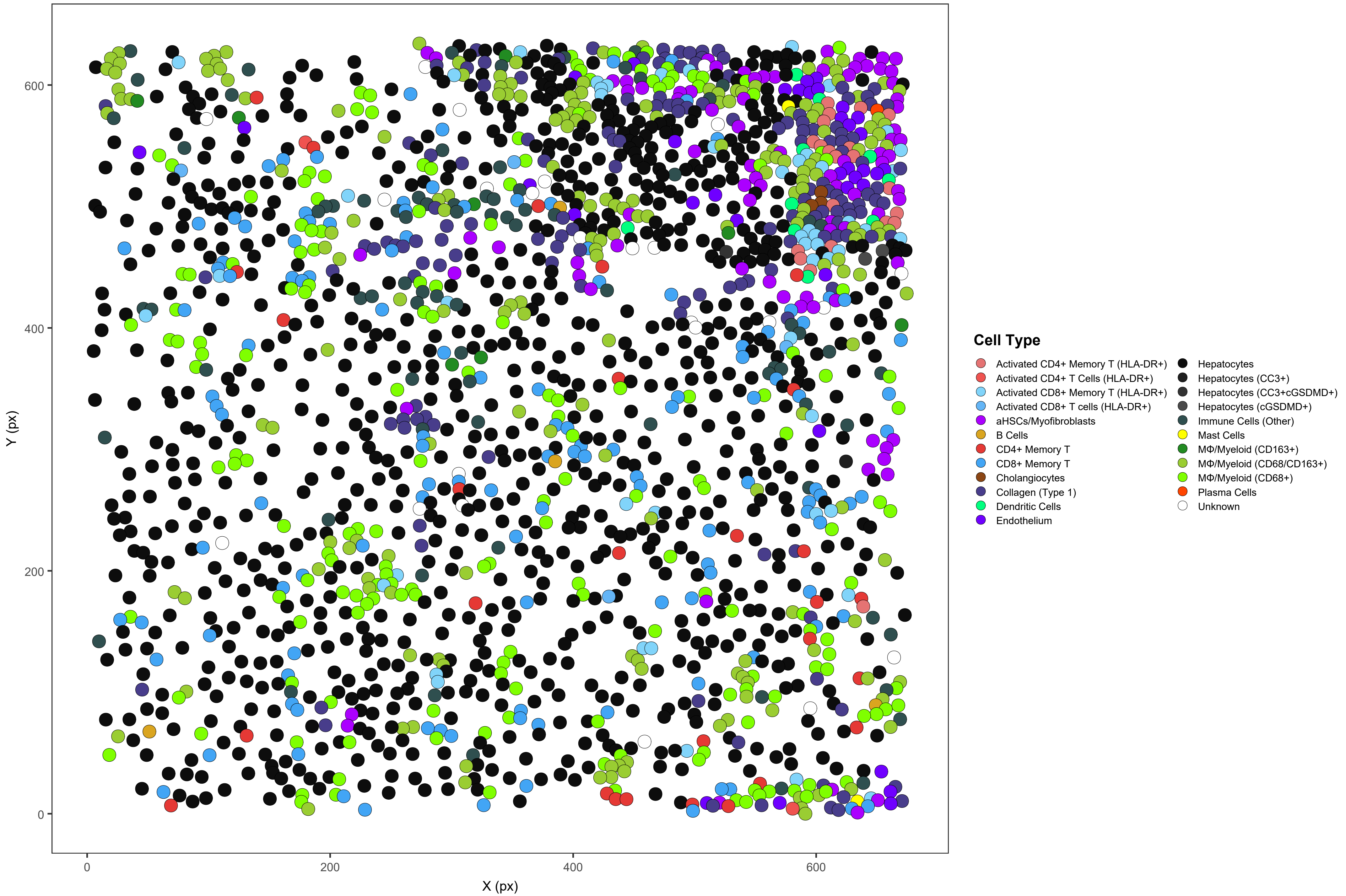

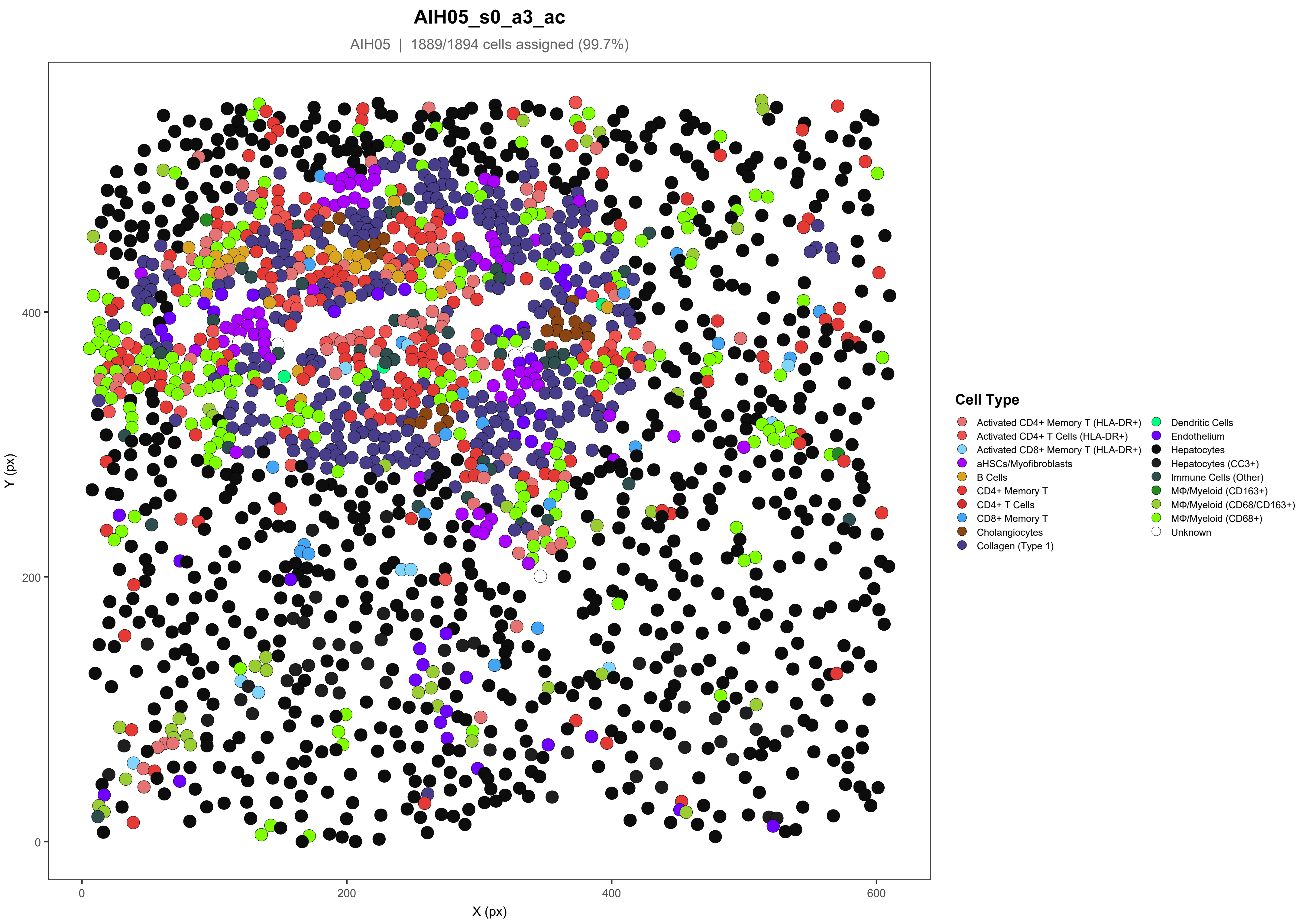
AIH05_s0_a3_ac
AIH05 | 1889/1894 cells assigned (99.7%)
Y (px)
X (px)
0
200
400
600
Cell Type
Activated CD4+ Memory T (HLA-DR+)
Activated CD4+ T Cells (HLA-DR+)
Activated CD8+ Memory T (HLA-DR+)
aHSCs/Myofibroblasts
B Cells
CD4+ Memory T
CD4+ T Cells
CD8+ Memory T
Cholangiocytes
Collagen (Type 1)
Dendritic Cells
Endothelium
Hepatocytes
Hepatocytes (CC3+)
Immune Cells (Other)
MΦ/Myeloid (CD163+)
MΦ/Myeloid (CD68/CD163+)
MΦ/Myeloid (CD68+)
Unknown

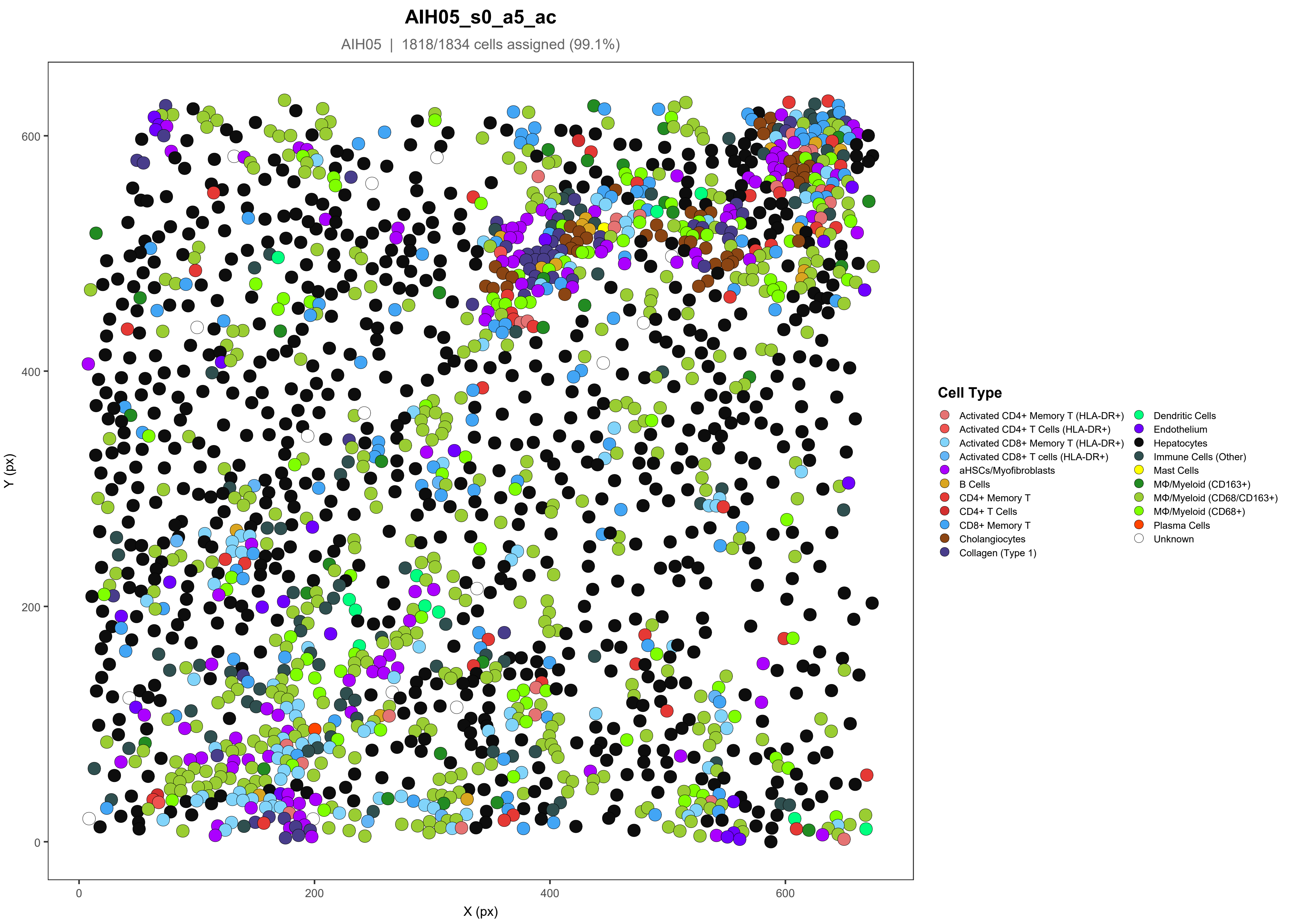
AIH05_s0_a5_ac
AIH05 | 1818/1834 cells assigned (99.1%)
Y (px)
X (px)
0
200
400
600
Cell Type
Activated CD4+ Memory T (HLA-DR+)
Activated CD4+ T Cells (HLA-DR+)
Activated CD8+ Memory T (HLA-DR+)
Activated CD8+ T cells (HLA-DR+)
aHSCs/Myofibroblasts
B Cells
CD4+ Memory T
CD4+ T Cells
CD8+ Memory T
Cholangiocytes
Collagen (Type 1)
Dendritic Cells
Endothelium
Hepatocytes
Immune Cells (Other)
Mast Cells
MΦ/Myeloid (CD163+)
MΦ/Myeloid (CD68/CD163+)
MΦ/Myeloid (CD68+)
Plasma Cells
Unknown

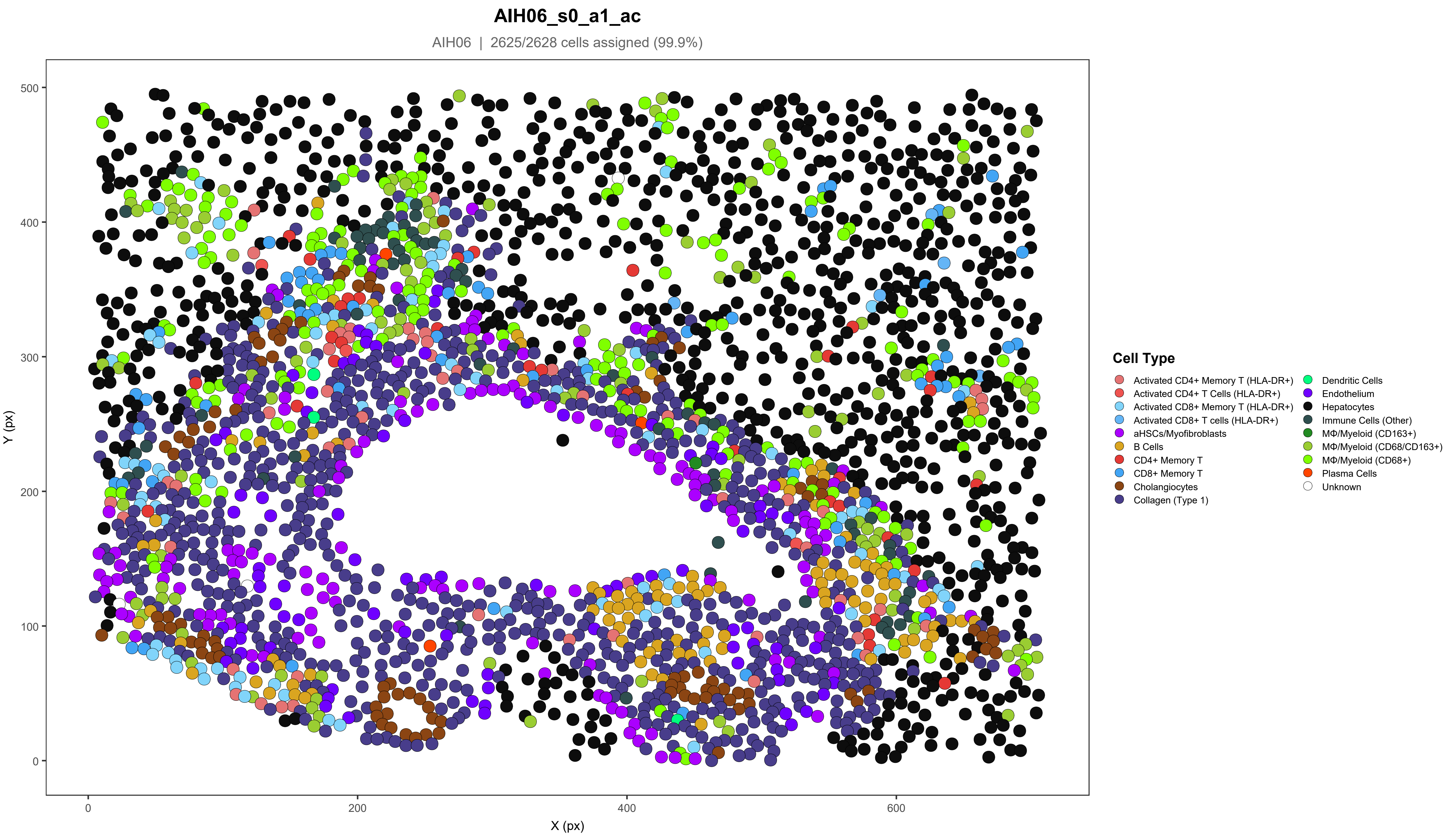
AIH06_s0_a1_ac
AIH06 | 2625/2628 cells assigned (99.9%)
Y (px)
X (px)
0
100
200
300
400
500
0
200
400
600
Cell Type
Activated CD4+ Memory T (HLA-DR+)
Activated CD4+ T Cells (HLA-DR+)
Activated CD8+ Memory T (HLA-DR+)
Activated CD8+ T cells (HLA-DR+)
aHSCs/Myofibroblasts
B Cells
CD4+ Memory T
CD8+ Memory T
Cholangiocytes
Collagen (Type 1)
Dendritic Cells
Endothelium
Hepatocytes
Immune Cells (Other)
MΦ/Myeloid (CD163+)
MΦ/Myeloid (CD68/CD163+)
MΦ/Myeloid (CD68+)
Plasma Cells
Unknown

### AIH06_s0_a7_ac

AIH06 | 2518/2522 cells assigned (99.8%)

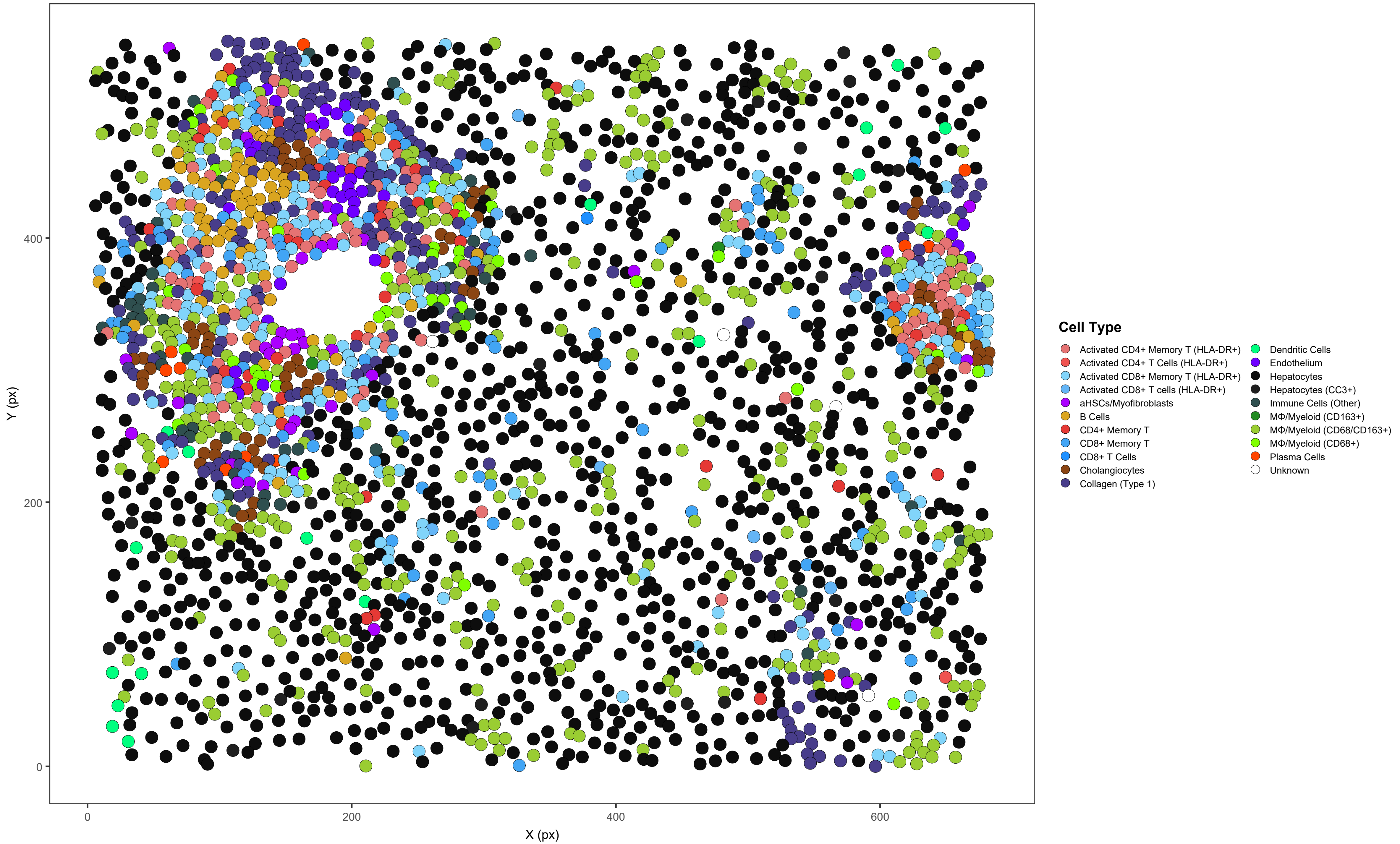

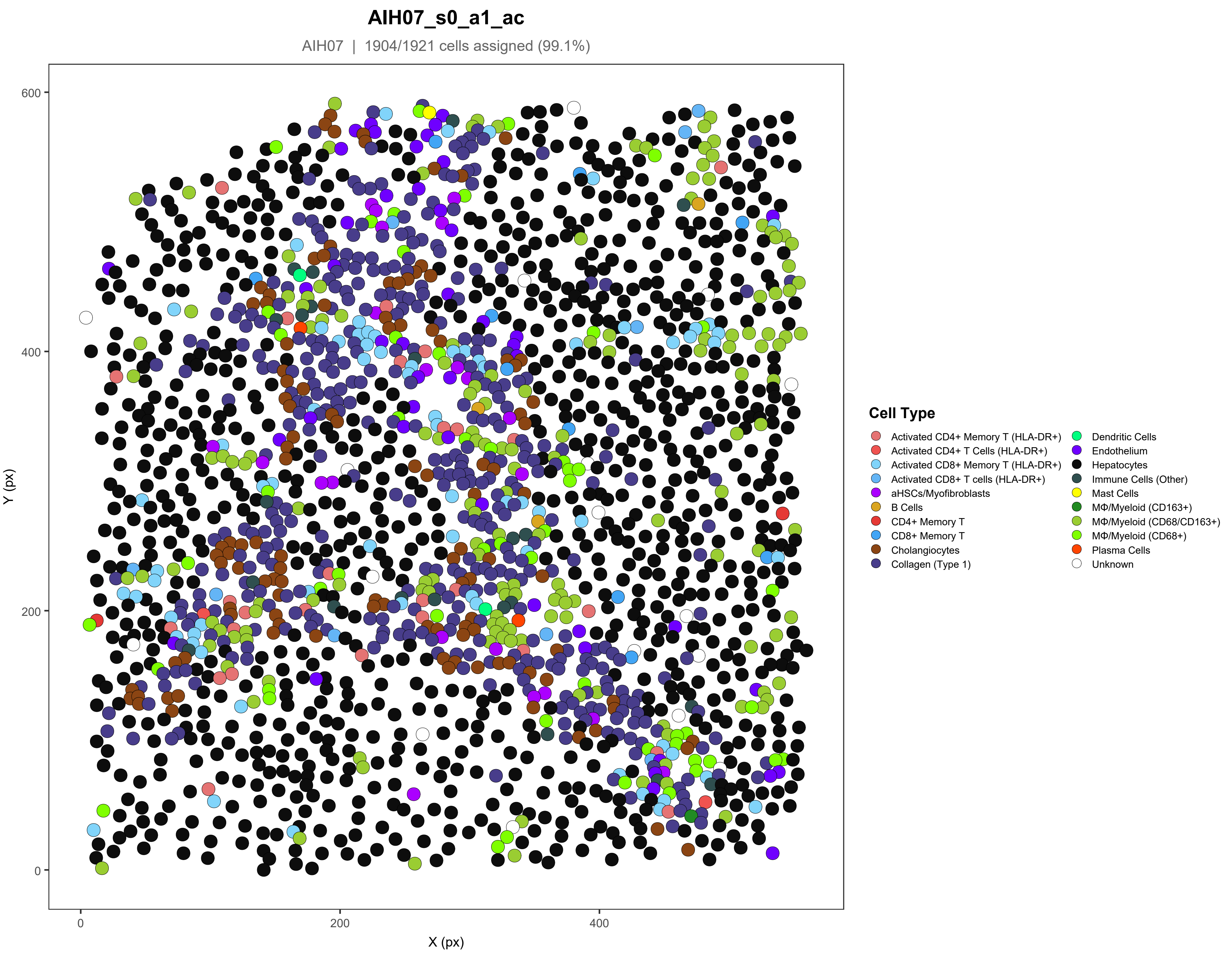
AIH07_s0_a1_ac
AIH07 | 1904/1921 cells assigned (99.1%)
Y (px)
X (px)
0
200
400
600
Cell Type
Activated CD4+ Memory T (HLA-DR+)
Activated CD4+ T Cells (HLA-DR+)
Activated CD8+ Memory T (HLA-DR+)
Activated CD8+ T cells (HLA-DR+)
aHSCs/Myofibroblasts
B Cells
CD4+ Memory T
CD8+ Memory T
Cholangiocytes
Collagen (Type 1)
Dendritic Cells
Endothelium
Hepatocytes
Immune Cells (Other)
Mast Cells
MΦ/Myeloid (CD163+)
MΦ/Myeloid (CD68/CD163+)
MΦ/Myeloid (CD68+)
Plasma Cells
Unknown

#### AIH07_s0_a2_ac

AIH07 | 2119/2150 cells assigned (98.6%)

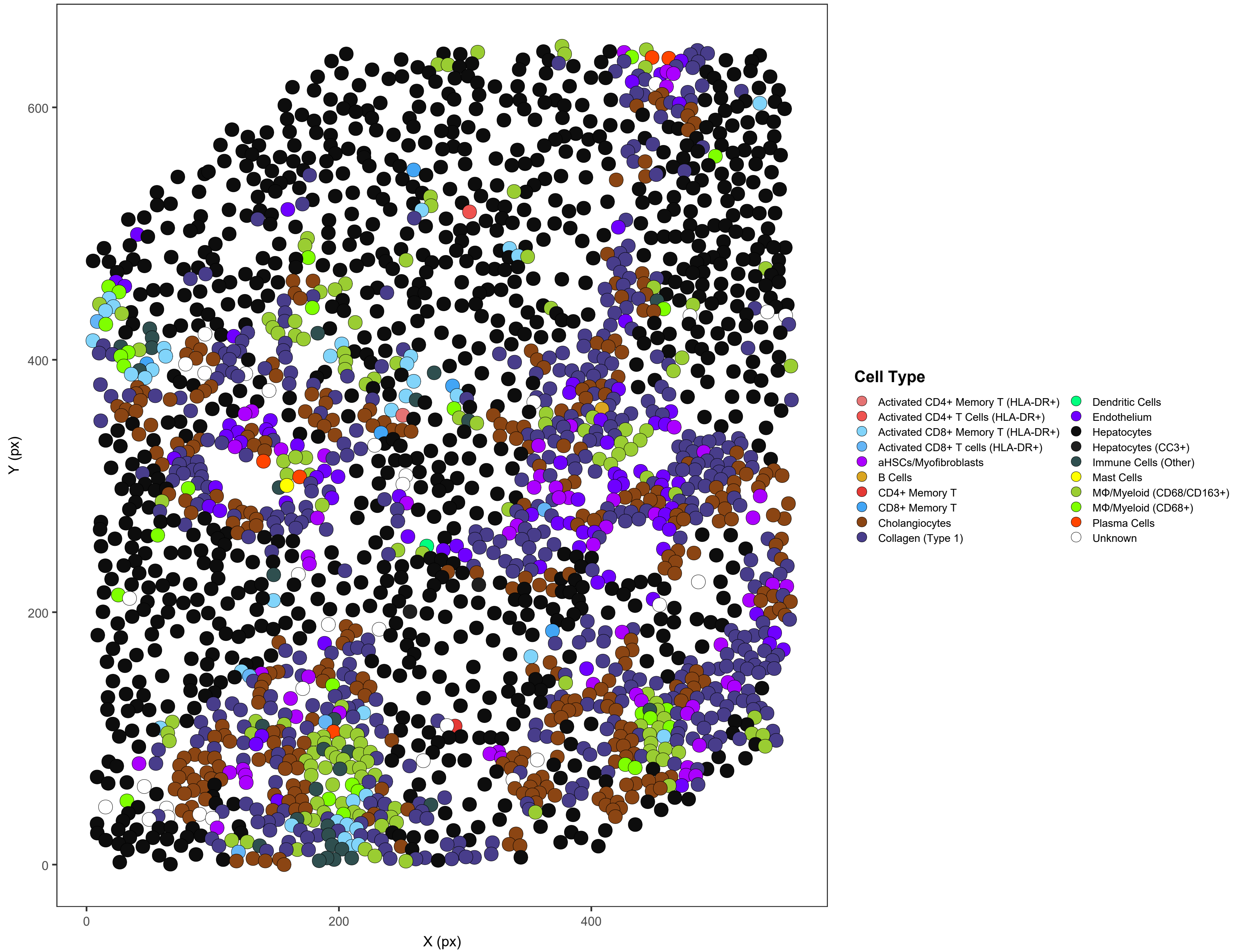

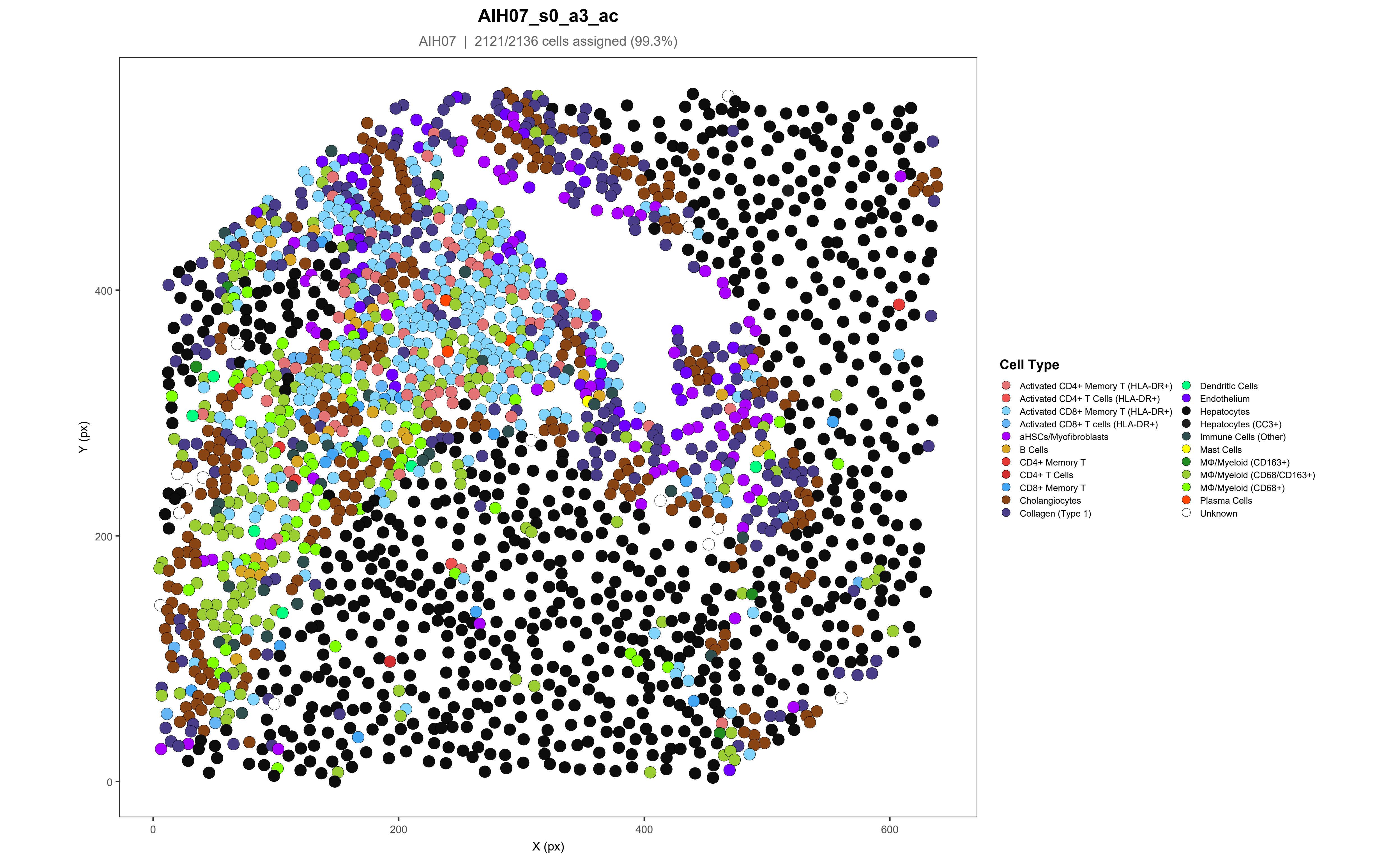
AIH07_s0_a3_ac
AIH07 | 2121/2136 cells assigned (99.3%)
Y (px)
X (px)
0
200
400
600
Cell Type
Activated CD4+ Memory T (HLA-DR+)
Activated CD4+ T Cells (HLA-DR+)
Activated CD8+ Memory T (HLA-DR+)
Activated CD8+ T cells (HLA-DR+)
aHSCs/Myofibroblasts
B Cells
CD4+ Memory T
CD4+ T Cells
CD8+ Memory T
Cholangiocytes
Collagen (Type 1)
Dendritic Cells
Endothelium
Hepatocytes
Hepatocytes (CC3+)
Immune Cells (Other)
Mast Cells
MΦ/Myeloid (CD163+)
MΦ/Myeloid (CD68/CD163+)
MΦ/Myeloid (CD68+)
Plasma Cells
Unknown

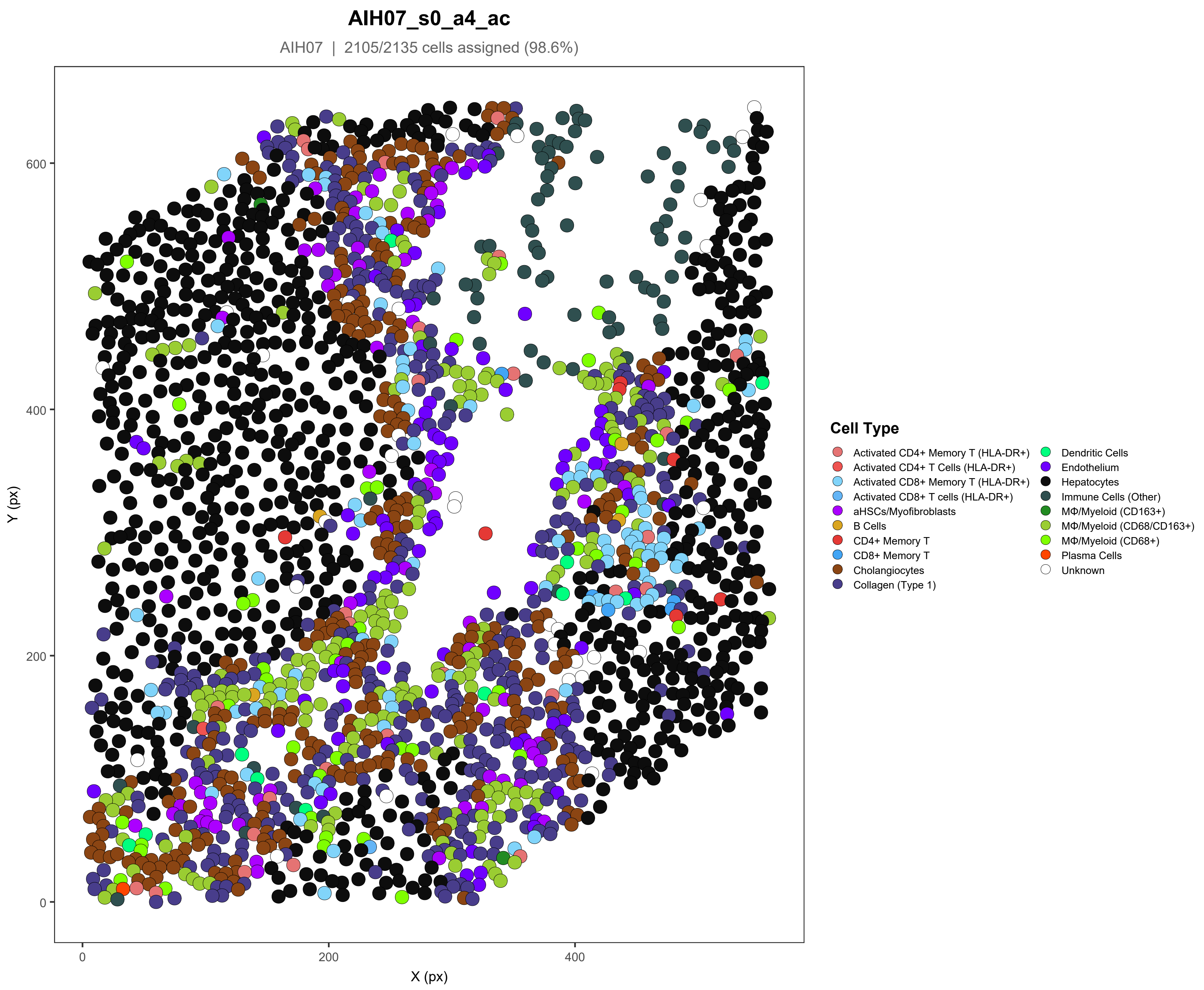
AIH07_s0_a4_ac
AIH07 | 2105/2135 cells assigned (98.6%)
Y (px)
X (px)
0
200
400
600
0
200
400
Cell Type
Activated CD4+ Memory T (HLA-DR+)
Activated CD4+ T Cells (HLA-DR+)
Activated CD8+ Memory T (HLA-DR+)
Activated CD8+ T cells (HLA-DR+)
aHSCs/Myofibroblasts
B Cells
CD4+ Memory T
CD8+ Memory T
Cholangiocytes
Collagen (Type 1)
Dendritic Cells
Endothelium
Hepatocytes
Immune Cells (Other)
MΦ/Myeloid (CD163+)
MΦ/Myeloid (CD68/CD163+)
MΦ/Myeloid (CD68+)
Plasma Cells
Unknown

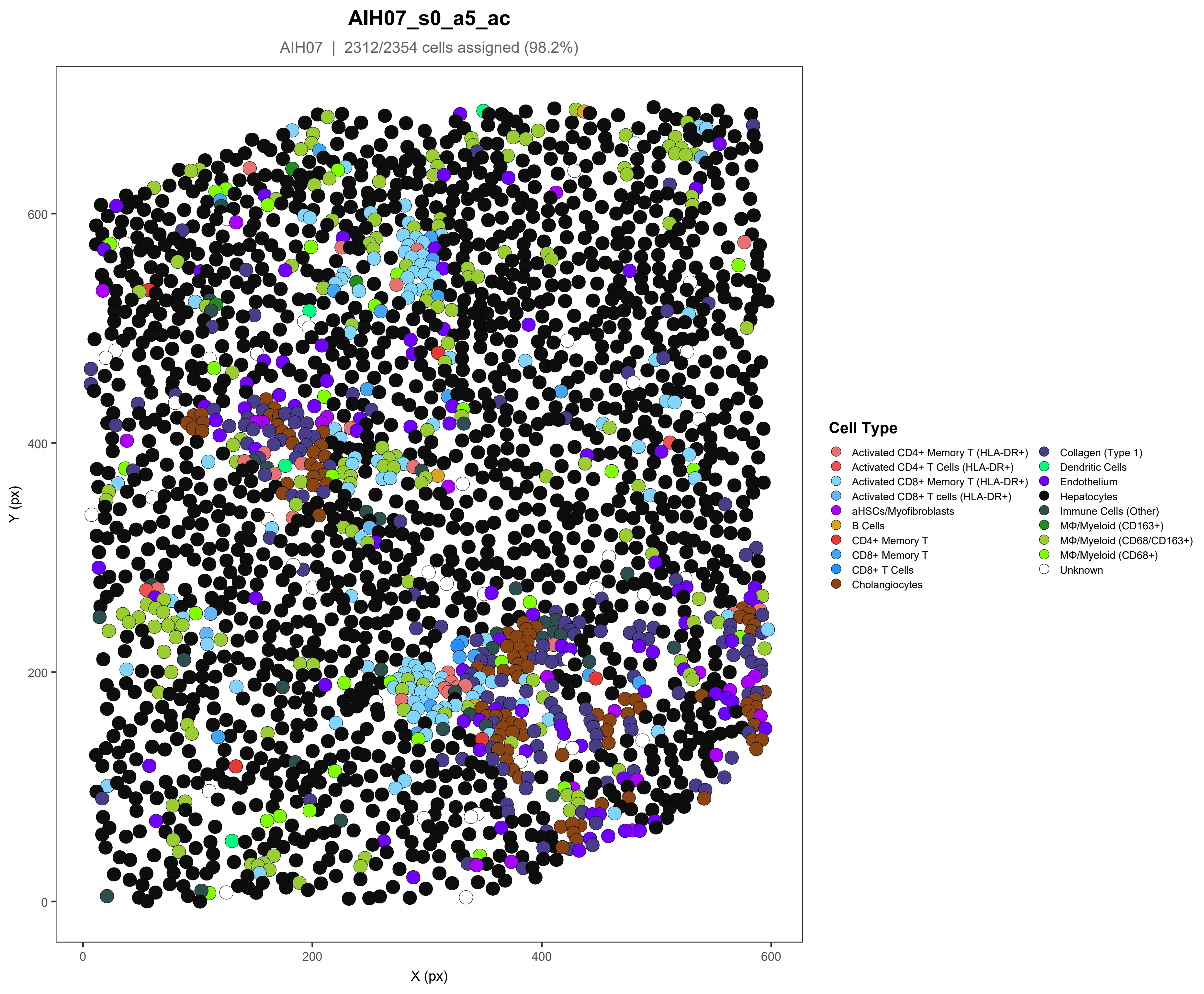
AIH07_s0_a5_ac
AIH07 | 2312/2354 cells assigned (98.2%)
Y (px)
X (px)
0
200
400
600
Cell Type
Activated CD4+ Memory T (HLA-DR+)
Activated CD4+ T Cells (HLA-DR+)
Activated CD8+ Memory T (HLA-DR+)
Activated CD8+ T cells (HLA-DR+)
aHSCs/Myofibroblasts
B Cells
CD4+ Memory T
CD8+ Memory T
CD8+ T Cells
Cholangiocytes
Collagen (Type 1)
Dendritic Cells
Endothelium
Hepatocytes
Immune Cells (Other)
MΦ/Myeloid (CD163+)
MΦ/Myeloid (CD68/CD163+)
MΦ/Myeloid (CD68+)
Unknown

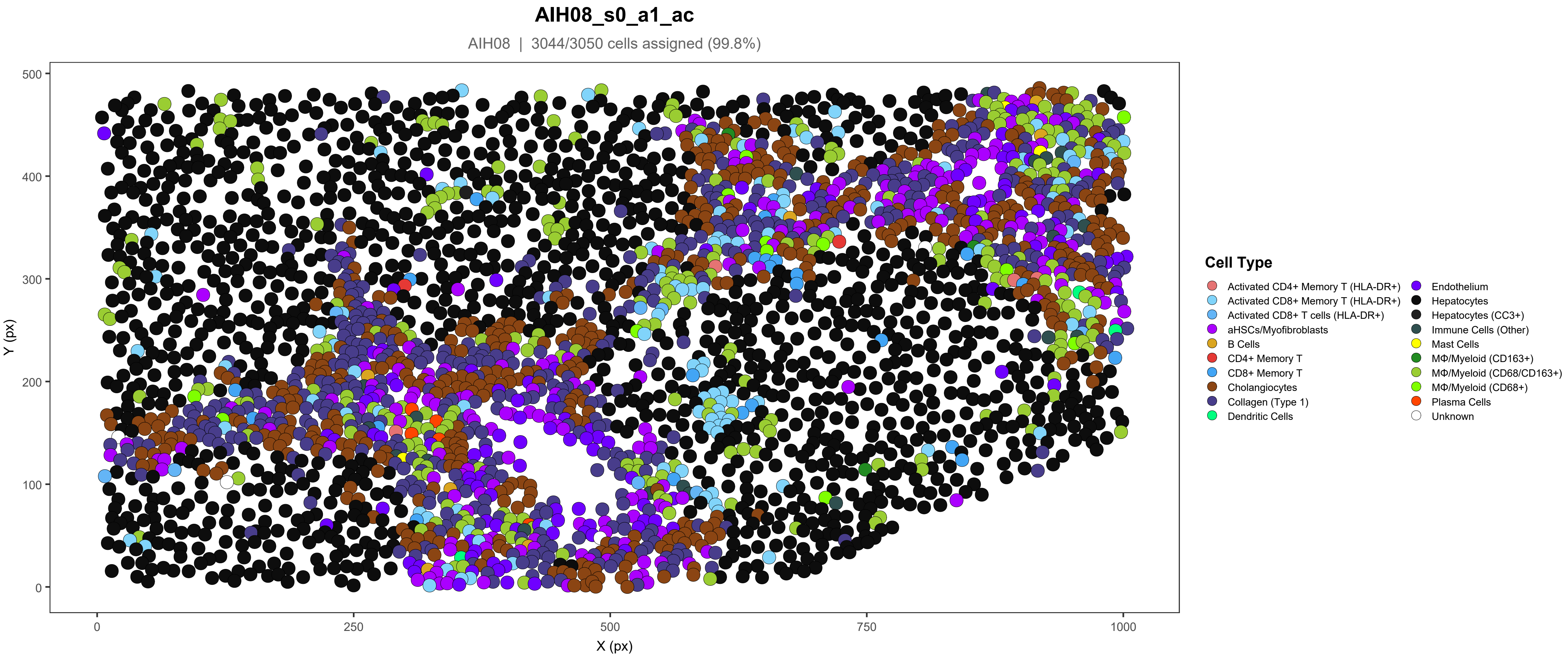
AIH08_s0_a1_ac
AIH08 | 3044/3050 cells assigned (99.8%)
Y (px)
X (px)
0
100
200
300
400
500
0
250
500
750
1000
Cell Type
Activated CD4+ Memory T (HLA-DR+)
Activated CD8+ Memory T (HLA-DR+)
Activated CD8+ T cells (HLA-DR+)
aHSCs/Myofibroblasts
B Cells
CD4+ Memory T
CD8+ Memory T
Cholangiocytes
Collagen (Type 1)
Dendritic Cells
Endothelium
Hepatocytes
Hepatocytes (CC3+)
Immune Cells (Other)
Mast Cells
MΦ/Myeloid (CD163+)
MΦ/Myeloid (CD68/CD163+)
MΦ/Myeloid (CD68+)
Plasma Cells
Unknown

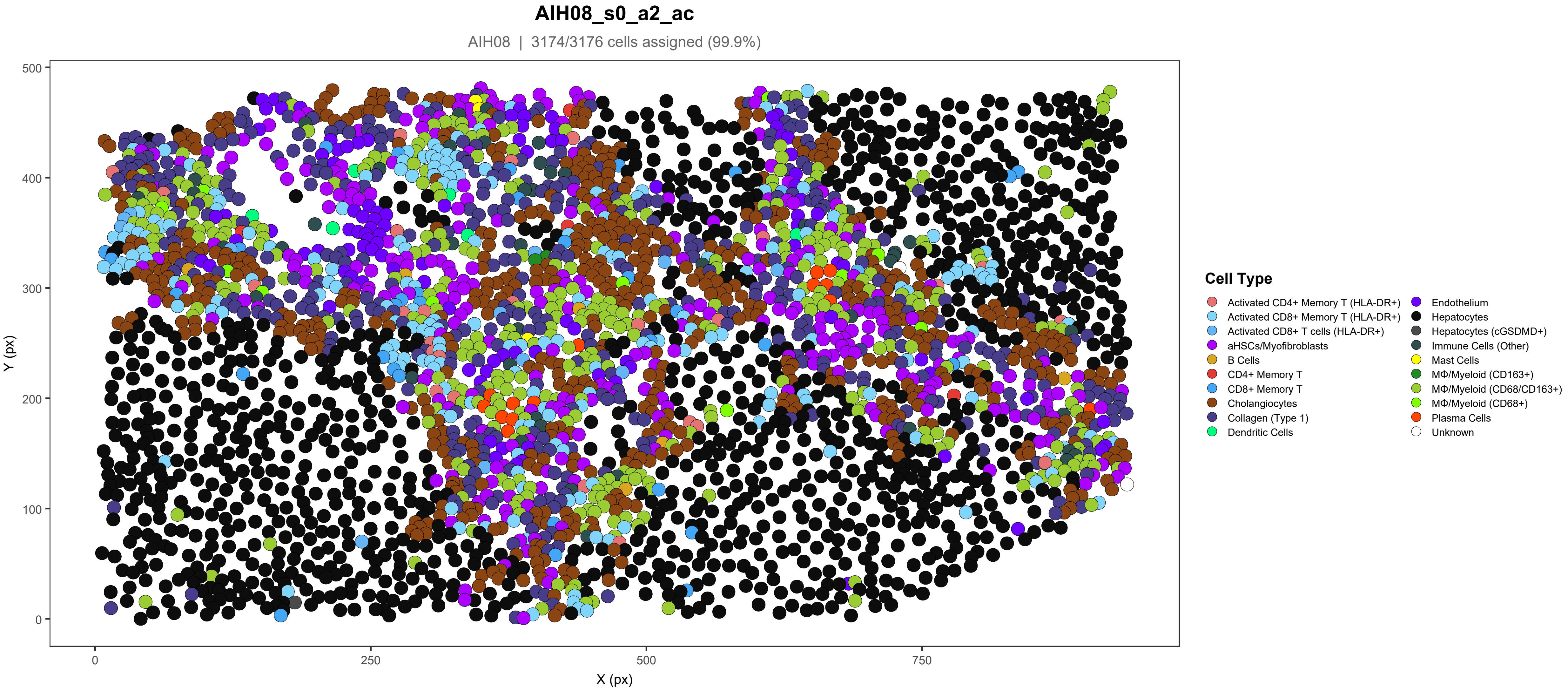
AIH08_s0_a2_ac
AIH08 | 3174/3176 cells assigned (99.9%)
Y (px)
X (px)
0
100
200
300
400
500
0
250
500
750
Cell Type
Activated CD4+ Memory T (HLA-DR+)
Activated CD8+ Memory T (HLA-DR+)
Activated CD8+ T cells (HLA-DR+)
aHSCs/Myofibroblasts
B Cells
CD4+ Memory T
CD8+ Memory T
Cholangiocytes
Collagen (Type 1)
Dendritic Cells
Endothelium
Hepatocytes
Hepatocytes (cGSDMD+)
Immune Cells (Other)
Mast Cells
MΦ/Myeloid (CD163+)
MΦ/Myeloid (CD68/CD163+)
MΦ/Myeloid (CD68+)
Plasma Cells
Unknown

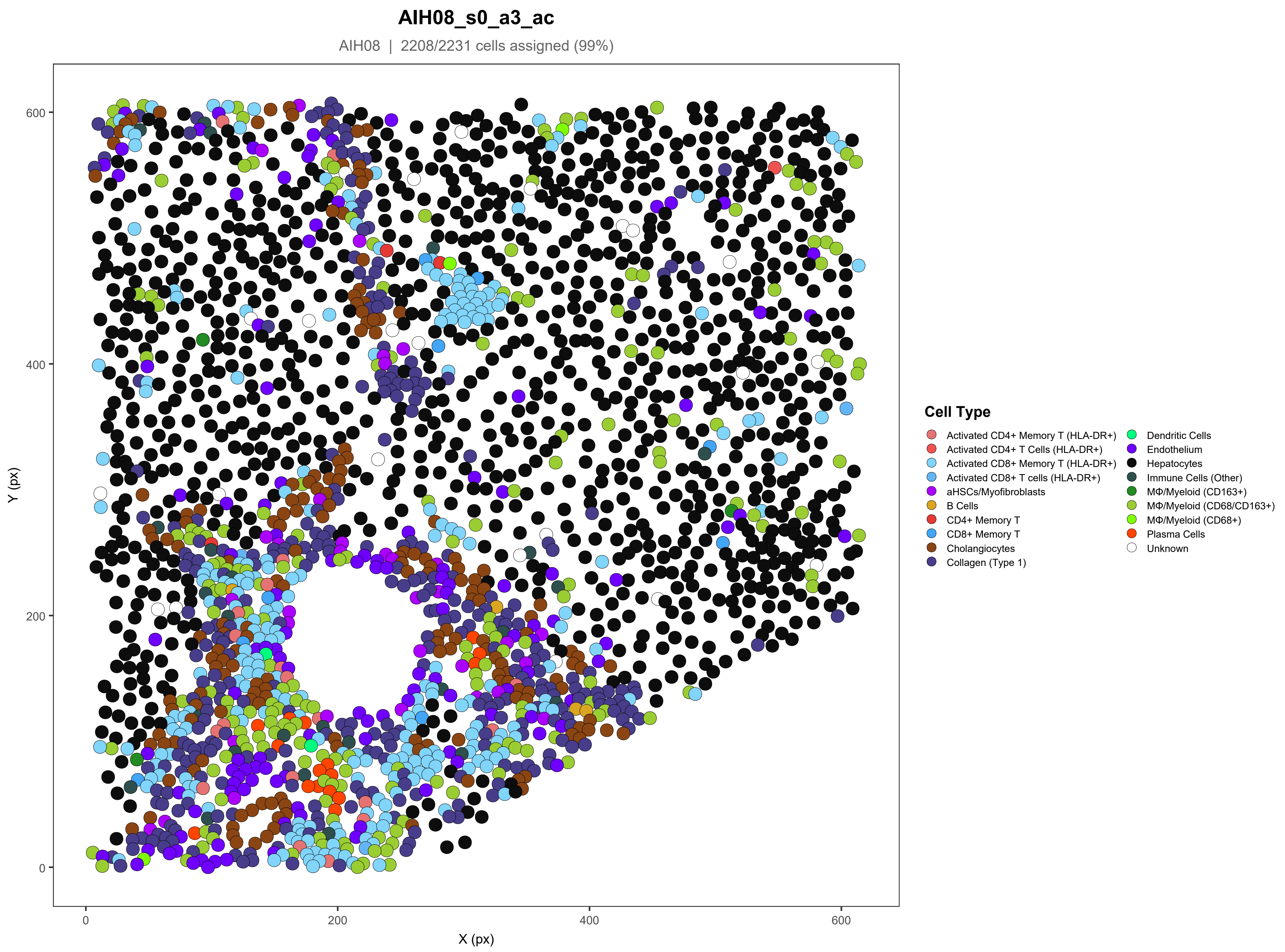
AIH08_s0_a3_ac
AIH08 | 2208/2231 cells assigned (99%)
Y (px)
X (px)
0
200
400
600
Cell Type
Activated CD4+ Memory T (HLA-DR+)
Activated CD4+ T Cells (HLA-DR+)
Activated CD8+ Memory T (HLA-DR+)
Activated CD8+ T cells (HLA-DR+)
aHSCs/Myofibroblasts
B Cells
CD4+ Memory T
CD8+ Memory T
Cholangiocytes
Collagen (Type 1)
Dendritic Cells
Endothelium
Hepatocytes
Immune Cells (Other)
MΦ/Myeloid (CD163+)
MΦ/Myeloid (CD68/CD163+)
MΦ/Myeloid (CD68+)
Plasma Cells
Unknown

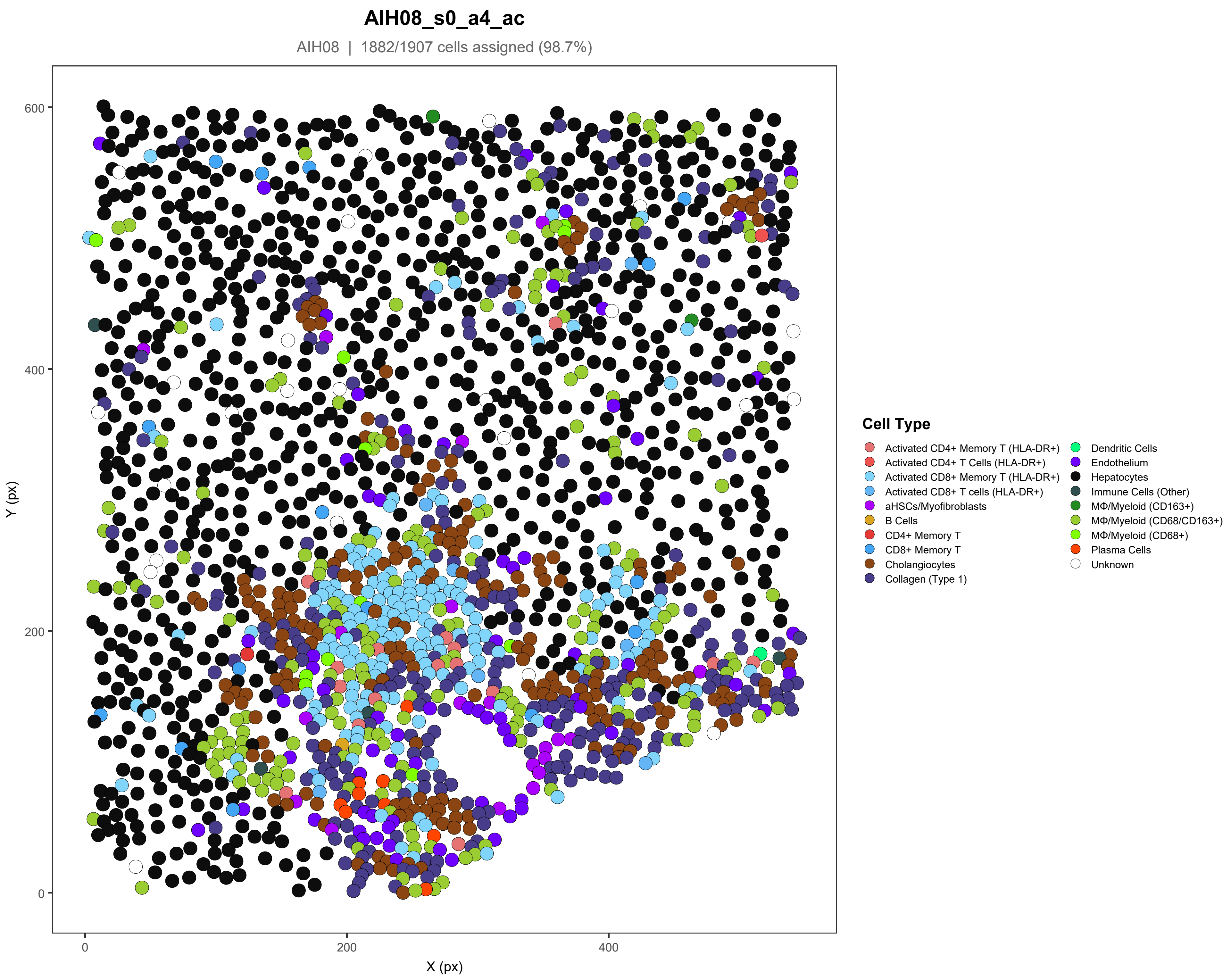
AIH08_s0_a4_ac
AIH08 | 1882/1907 cells assigned (98.7%)
Y (px)
X (px)
0
200
400
600
Cell Type
Activated CD4+ Memory T (HLA-DR+)
Activated CD4+ T Cells (HLA-DR+)
Activated CD8+ Memory T (HLA-DR+)
Activated CD8+ T cells (HLA-DR+)
aHSCs/Myofibroblasts
B Cells
CD4+ Memory T
CD8+ Memory T
Cholangiocytes
Collagen (Type 1)
Dendritic Cells
Endothelium
Hepatocytes
Immune Cells (Other)
MΦ/Myeloid (CD163+)
MΦ/Myeloid (CD68/CD163+)
MΦ/Myeloid (CD68+)
Plasma Cells
Unknown

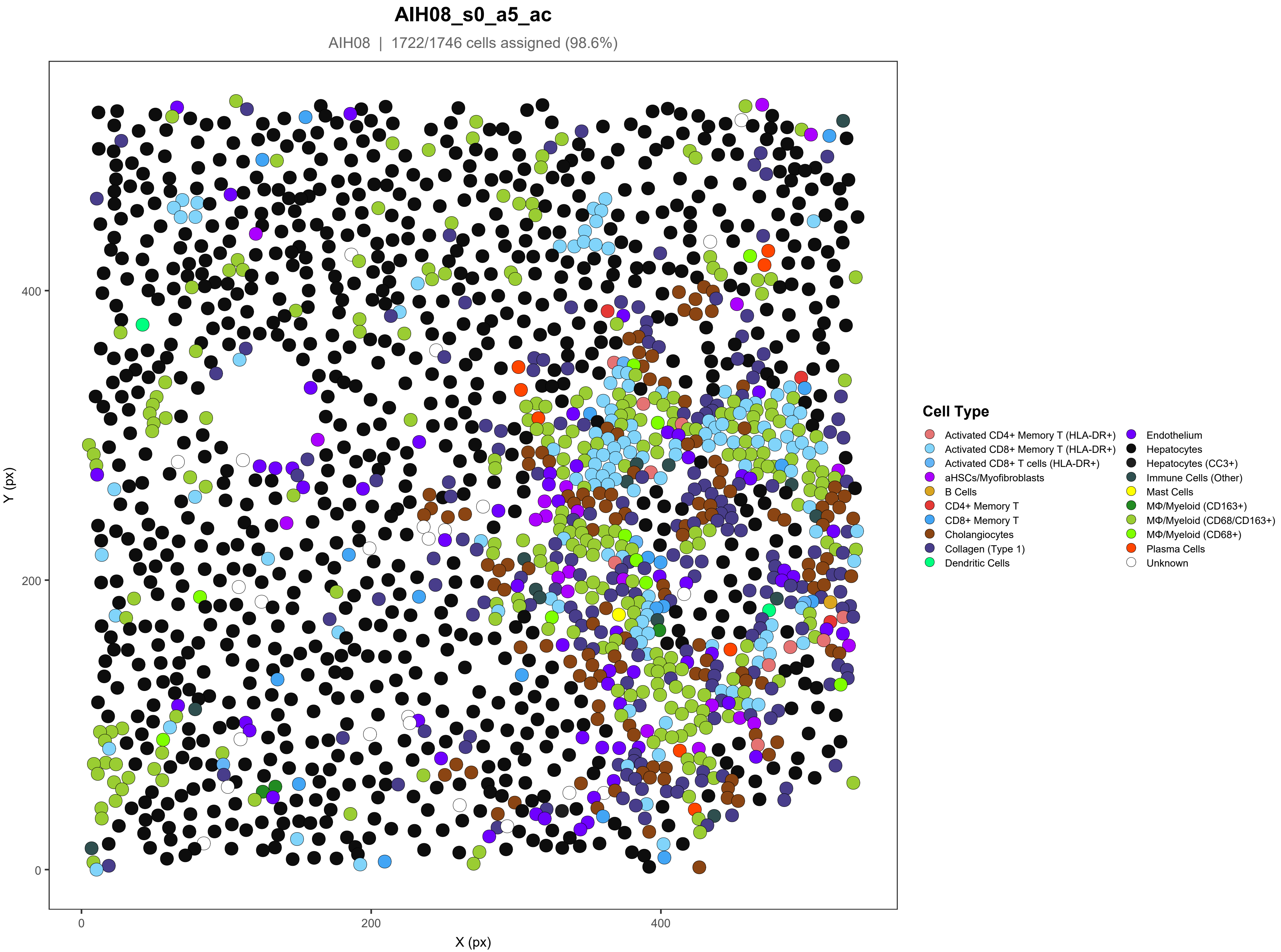

AIH08_s0_a5_ac
AIH08 | 1722/1746 cells assigned (98.6%)
Y (px)
X (px)
0
200
400
Cell Type
Activated CD4+ Memory T (HLA-DR+)
Activated CD8+ Memory T (HLA-DR+)
Activated CD8+ T cells (HLA-DR+)
aHSCs/Myofibroblasts
B Cells
CD4+ Memory T
CD8+ Memory T
Cholangiocytes
Collagen (Type 1)
Dendritic Cells
Endothelium
Hepatocytes
Hepatocytes (CC3+)
Immune Cells (Other)
Mast Cells
MΦ/Myeloid (CD163+)
MΦ/Myeloid (CD68/CD163+)
MΦ/Myeloid (CD68+)
Plasma Cells
Unknown

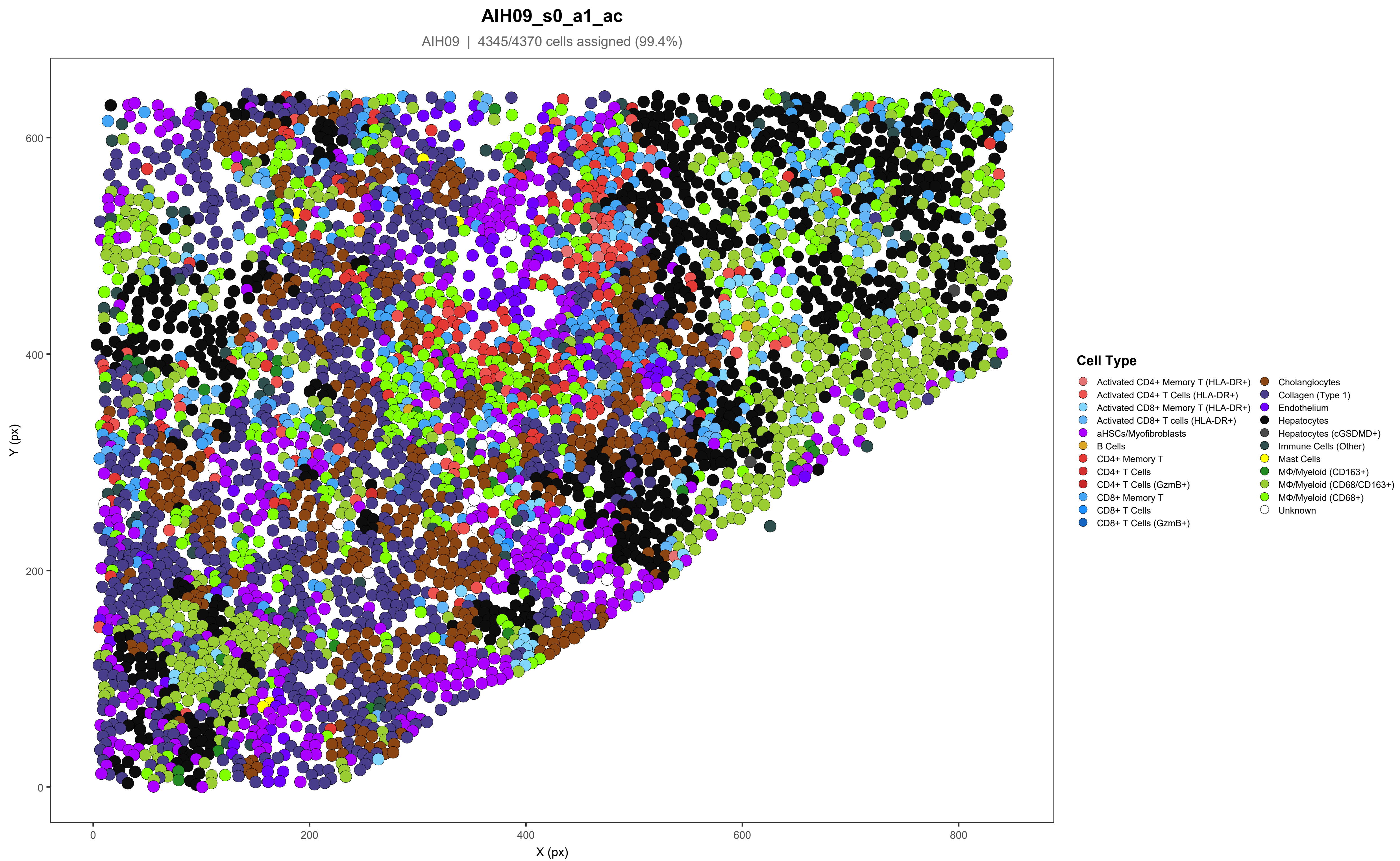
AIH09_s0_a1_ac
AIH09 | 4345/4370 cells assigned (99.4%)
Y (px)
X (px)
0
200
400
600
800
Cell Type
Activated CD4+ Memory T (HLA-DR+)
Activated CD4+ T Cells (HLA-DR+)
Activated CD8+ Memory T (HLA-DR+)
Activated CD8+ T cells (HLA-DR+)
aHSCs/Myofibroblasts
B Cells
CD4+ Memory T
CD4+ T Cells
CD4+ T Cells (GzmB+)
CD8+ Memory T
CD8+ T Cells
CD8+ T Cells (GzmB+)
Cholangiocytes
Collagen (Type 1)
Endothelium
Hepatocytes
Hepatocytes (cGSDMD+)
Immune Cells (Other)
Mast Cells
MΦ/Myeloid (CD163+)
MΦ/Myeloid (CD68/CD163+)
MΦ/Myeloid (CD68+)
Unknown

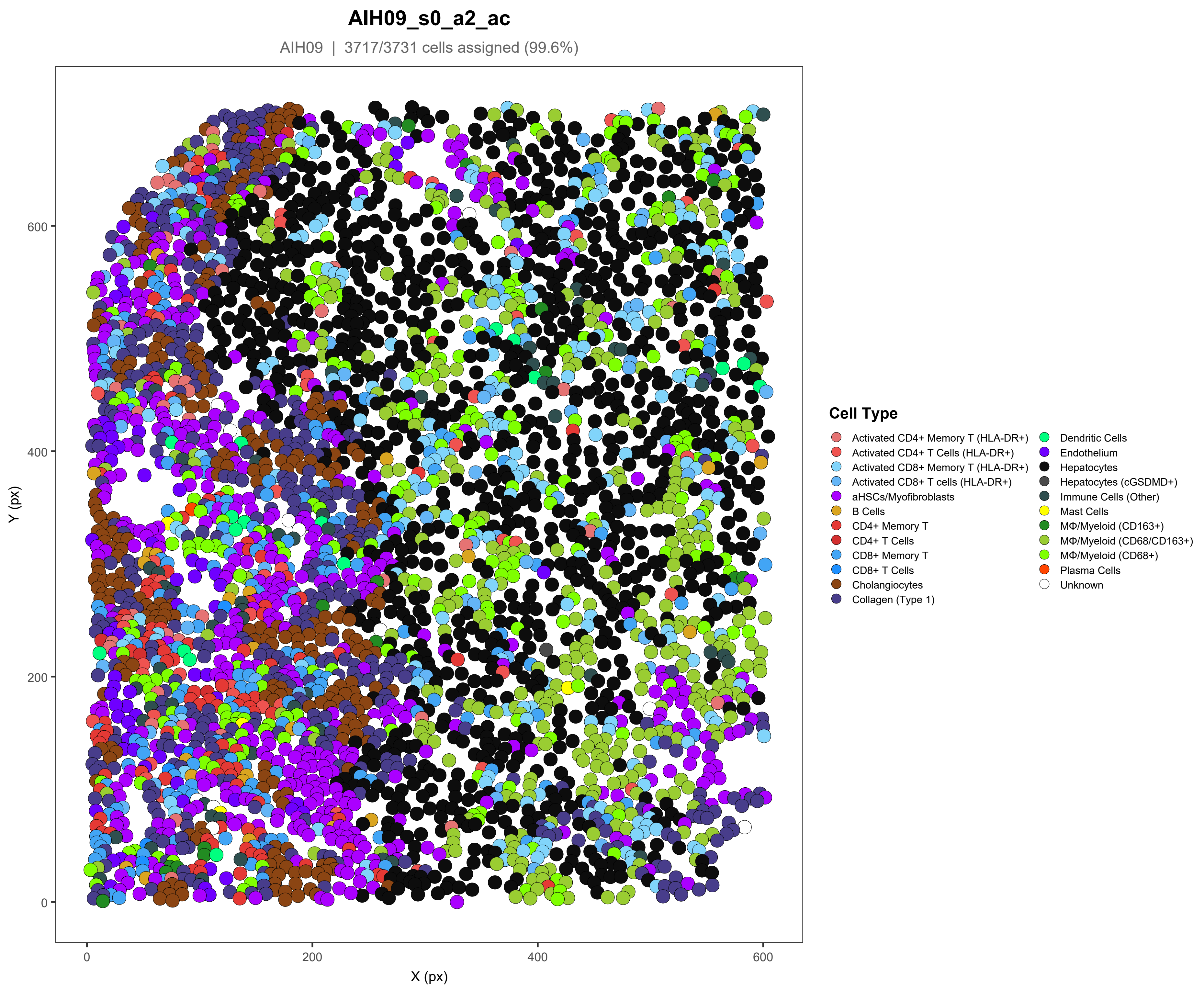
AIH09_s0_a2_ac
AIH09 | 3717/3731 cells assigned (99.6%)
Y (px)
X (px)
0
200
400
600
Cell Type
Activated CD4+ Memory T (HLA-DR+)
Activated CD4+ T Cells (HLA-DR+)
Activated CD8+ Memory T (HLA-DR+)
Activated CD8+ T cells (HLA-DR+)
aHSCs/Myofibroblasts
B Cells
CD4+ Memory T
CD4+ T Cells
CD8+ Memory T
CD8+ T Cells
Cholangiocytes
Collagen (Type 1)
Dendritic Cells
Endothelium
Hepatocytes
Hepatocytes (cGSDMD+)
Immune Cells (Other)
Mast Cells
MΦ/Myeloid (CD163+)
MΦ/Myeloid (CD68/CD163+)
MΦ/Myeloid (CD68+)
Plasma Cells
Unknown

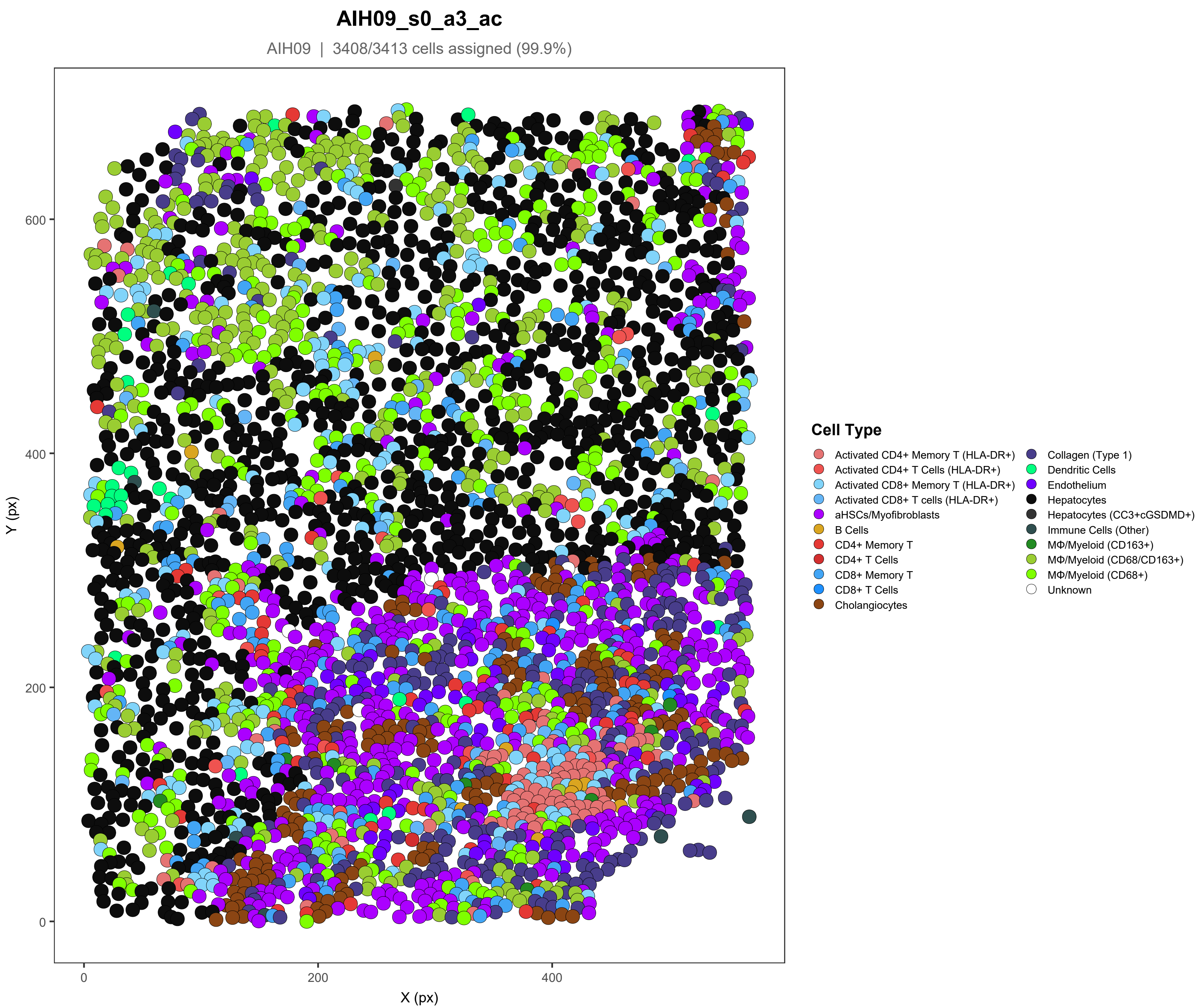
AIH09_s0_a3_ac
AIH09 | 3408/3413 cells assigned (99.9%)
Y (px)
X (px)
0
200
400
600
Cell Type
Activated CD4+ Memory T (HLA-DR+)
Activated CD4+ T Cells (HLA-DR+)
Activated CD8+ Memory T (HLA-DR+)
Activated CD8+ T cells (HLA-DR+)
aHSCs/Myofibroblasts
B Cells
CD4+ Memory T
CD4+ T Cells
CD8+ Memory T
CD8+ T Cells
Cholangiocytes
Collagen (Type 1)
Dendritic Cells
Endothelium
Hepatocytes
Hepatocytes (CC3+cGSDMD+)
Immune Cells (Other)
MΦ/Myeloid (CD163+)
MΦ/Myeloid (CD68/CD163+)
MΦ/Myeloid (CD68+)
Unknown

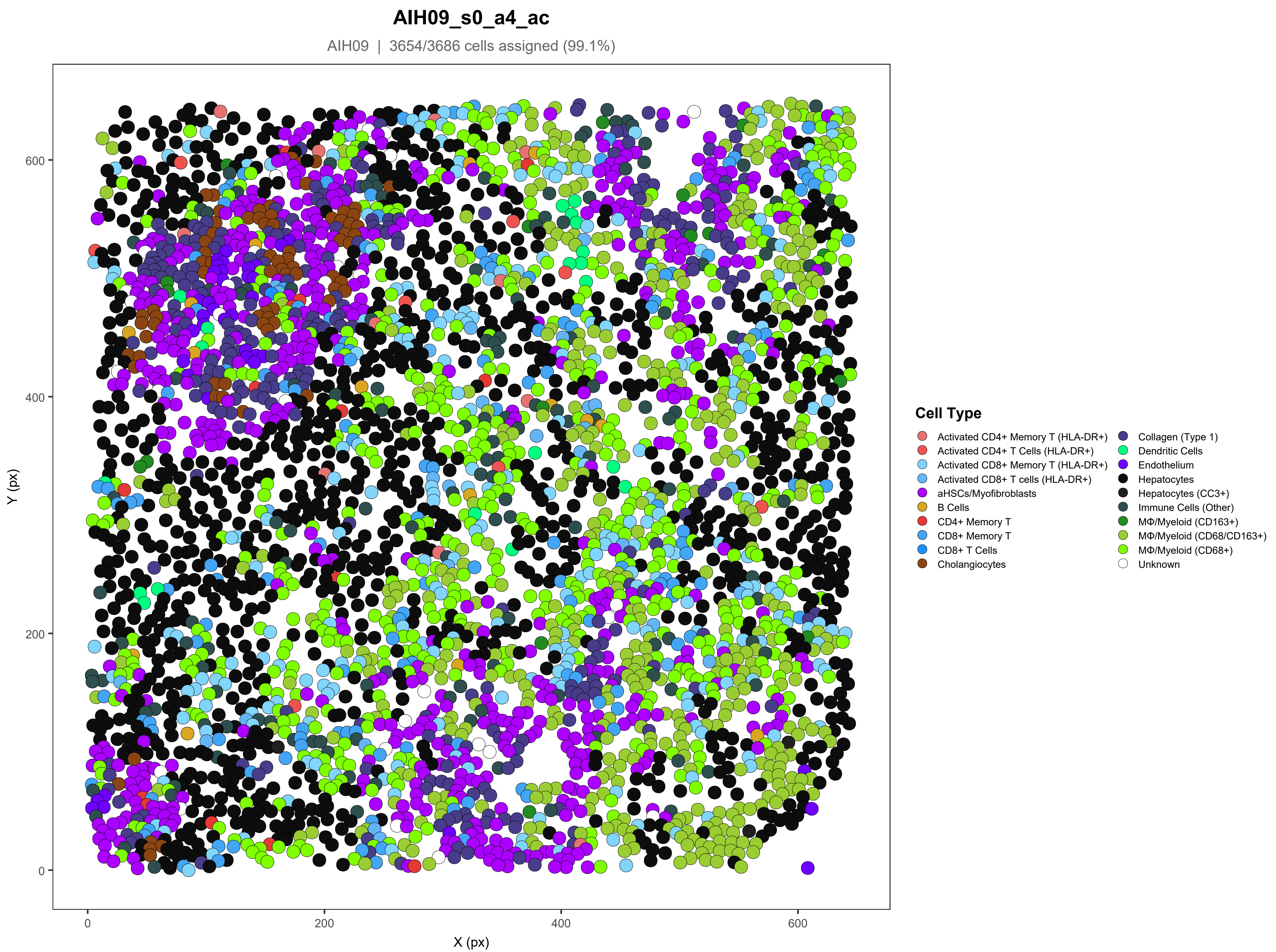
AIH09_s0_a4_ac
AIH09 | 3654/3686 cells assigned (99.1%)
Y (px)
X (px)
0
200
400
600
Cell Type
Activated CD4+ Memory T (HLA-DR+)
Activated CD4+ T Cells (HLA-DR+)
Activated CD8+ Memory T (HLA-DR+)
Activated CD8+ T cells (HLA-DR+)
aHSCs/Myofibroblasts
B Cells
CD4+ Memory T
CD8+ Memory T
CD8+ T Cells
Cholangiocytes
Collagen (Type 1)
Dendritic Cells
Endothelium
Hepatocytes
Hepatocytes (CC3+)
Immune Cells (Other)
MΦ/Myeloid (CD163+)
MΦ/Myeloid (CD68/CD163+)
MΦ/Myeloid (CD68+)
Unknown

### AIH09_s0_a5_ac

AIH09 | 3594/3636 cells assigned (98.8%)

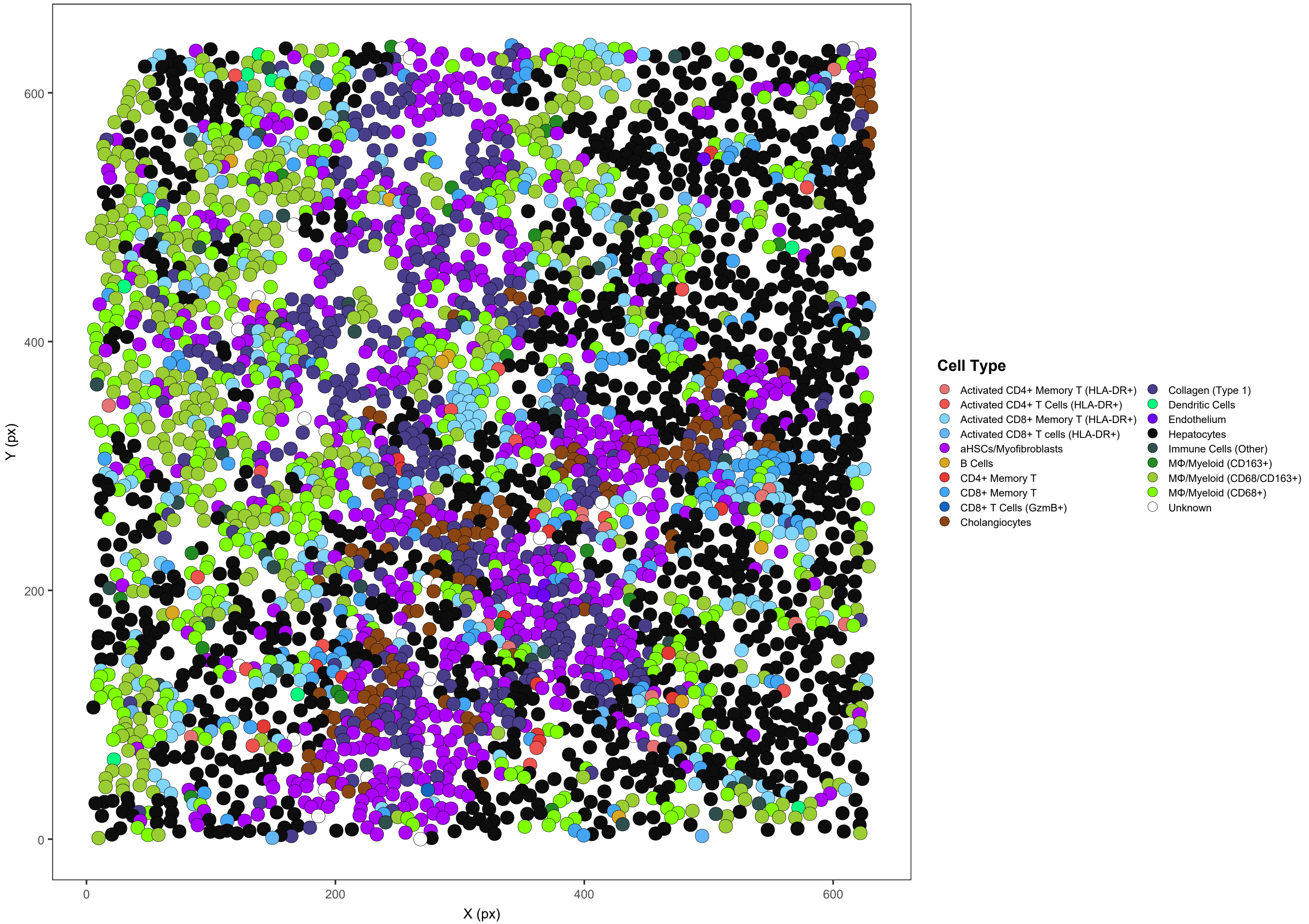

### AIH11_s0_a1_ac

AIH11 | 3183/3188 cells assigned (99.8%)

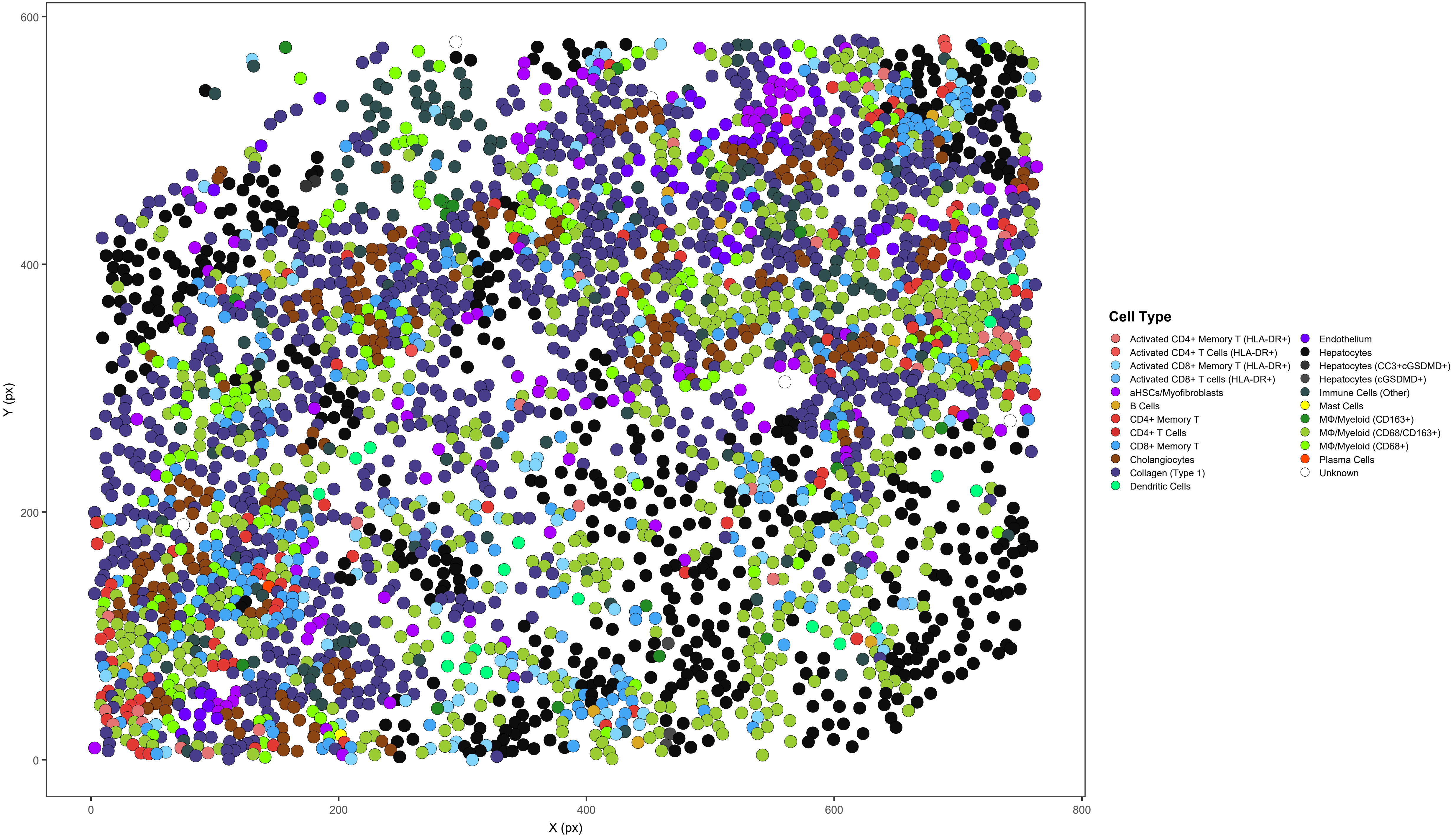

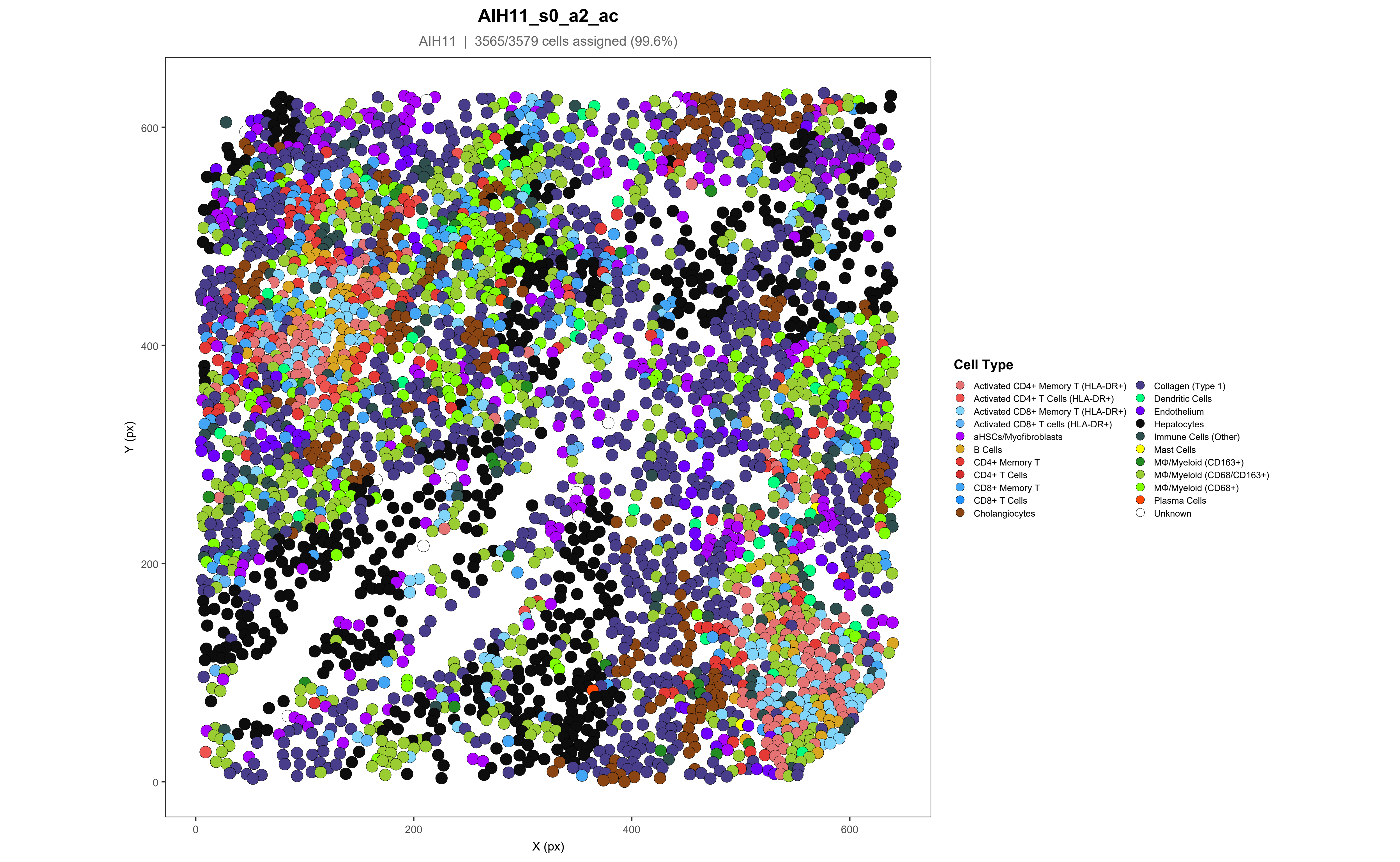

AIH11_s0_a2_ac
AIH11 | 3565/3579 cells assigned (99.6%)
Y (px)
X (px)
0
200
400
600
Cell Type
Activated CD4+ Memory T (HLA-DR+)
Activated CD4+ T Cells (HLA-DR+)
Activated CD8+ Memory T (HLA-DR+)
Activated CD8+ T cells (HLA-DR+)
aHSCs/Myofibroblasts
B Cells
CD4+ Memory T
CD4+ T Cells
CD8+ Memory T
CD8+ T Cells
Cholangiocytes
Collagen (Type 1)
Dendritic Cells
Endothelium
Hepatocytes
Immune Cells (Other)
Mast Cells
MΦ/Myeloid (CD163+)
MΦ/Myeloid (CD68/CD163+)
MΦ/Myeloid (CD68+)
Plasma Cells
Unknown

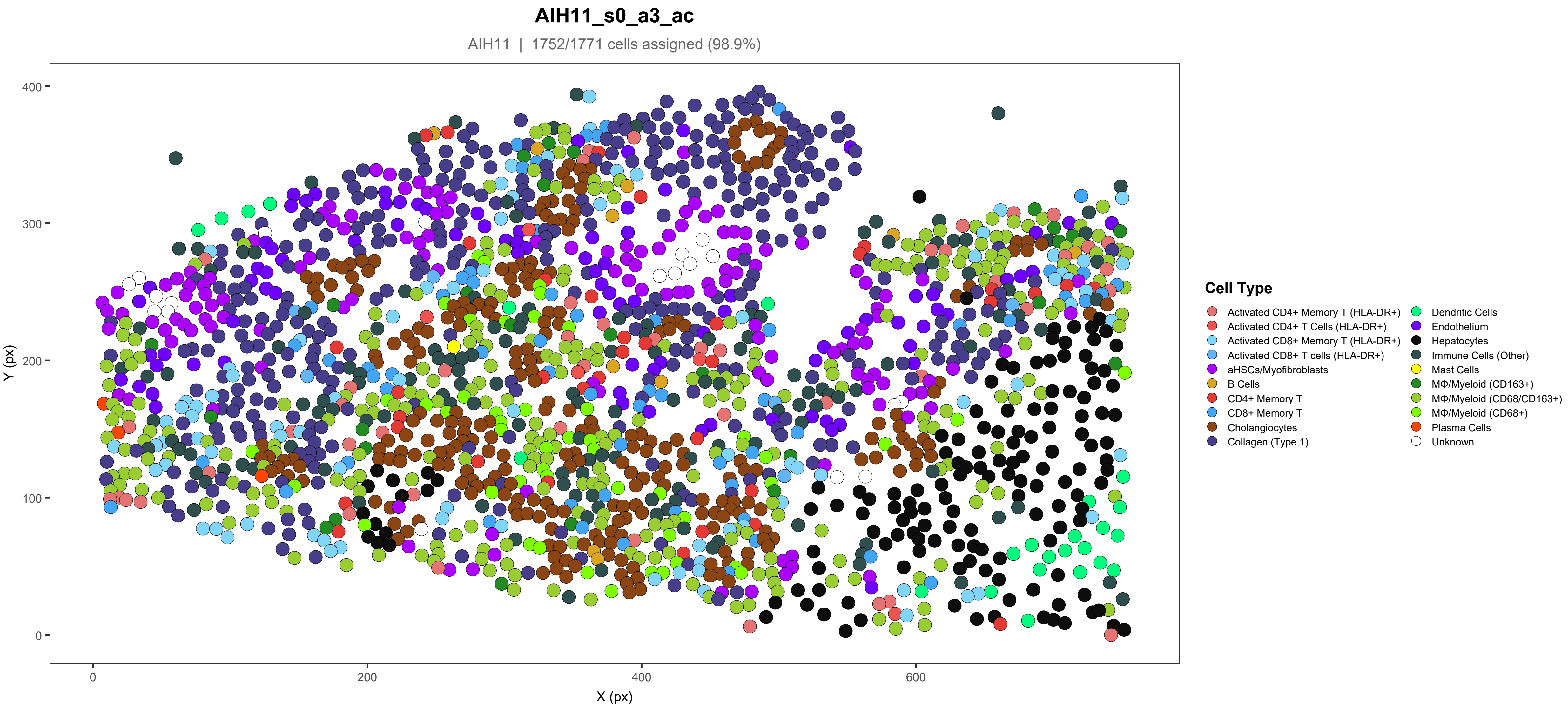
AIH11_s0_a3_ac
AIH11 | 1752/1771 cells assigned (98.9%)
Y (px)
X (px)
0
200
400
600
100
300
Cell Type
Activated CD4+ Memory T (HLA-DR+)
Activated CD4+ T Cells (HLA-DR+)
Activated CD8+ Memory T (HLA-DR+)
Activated CD8+ T cells (HLA-DR+)
aHSCs/Myofibroblasts
B Cells
CD4+ Memory T
CD8+ Memory T
Cholangiocytes
Collagen (Type 1)
Dendritic Cells
Endothelium
Hepatocytes
Immune Cells (Other)
Mast Cells
MΦ/Myeloid (CD163+)
MΦ/Myeloid (CD68/CD163+)
MΦ/Myeloid (CD68+)
Plasma Cells
Unknown

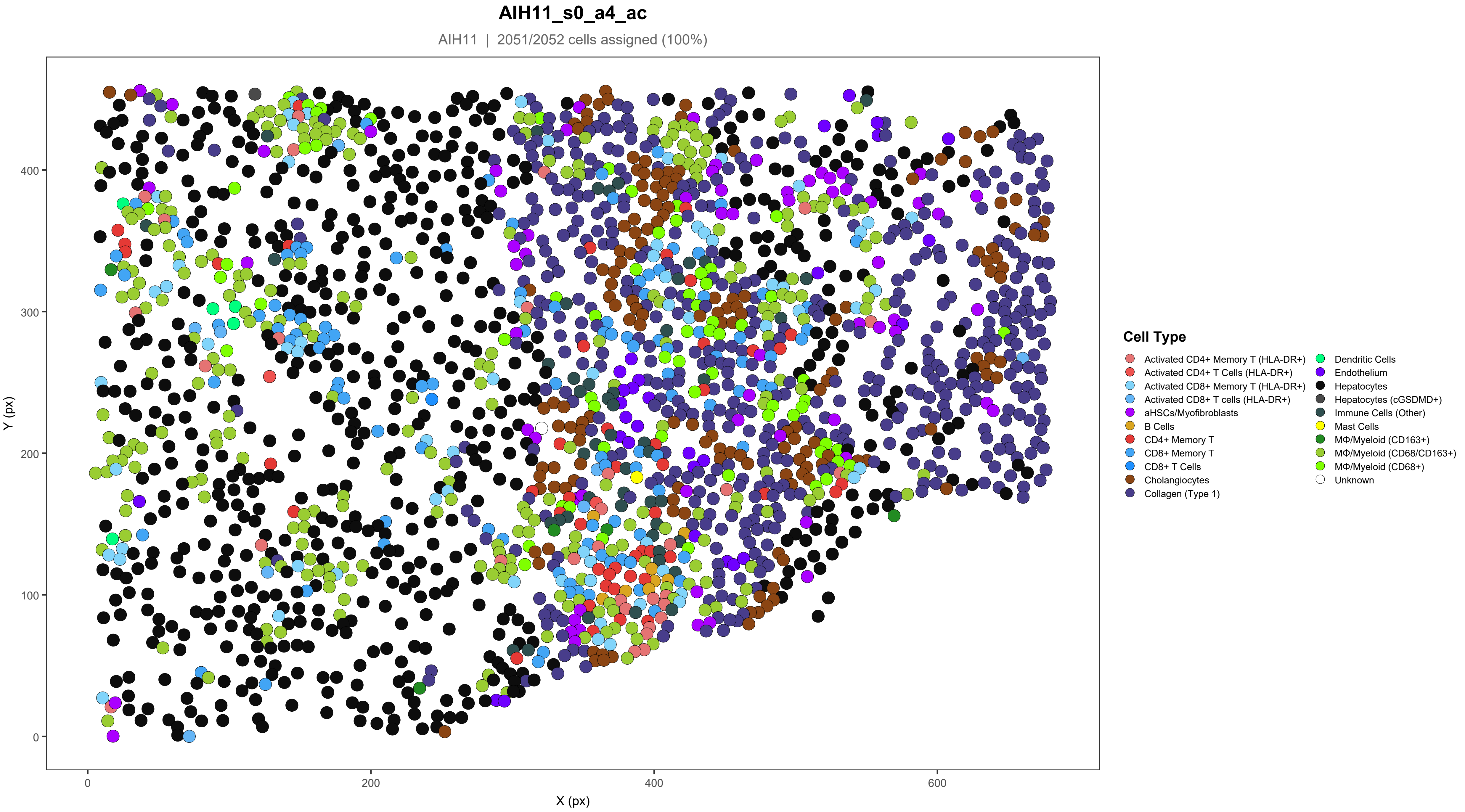

AIH11_s0_a4_ac
AIH11 | 2051/2052 cells assigned (100%)
Y (px)
X (px)
0
100
200
300
400
0
200
400
600
Cell Type
Activated CD4+ Memory T (HLA-DR+)
Activated CD4+ T Cells (HLA-DR+)
Activated CD8+ Memory T (HLA-DR+)
Activated CD8+ T cells (HLA-DR+)
aHSCs/Myofibroblasts
B Cells
CD4+ Memory T
CD8+ Memory T
CD8+ T Cells
Cholangiocytes
Collagen (Type 1)
Dendritic Cells
Endothelium
Hepatocytes
Hepatocytes (cGSDMD+)
Immune Cells (Other)
Mast Cells
MΦ/Myeloid (CD163+)
MΦ/Myeloid (CD68/CD163+)
MΦ/Myeloid (CD68+)
Unknown

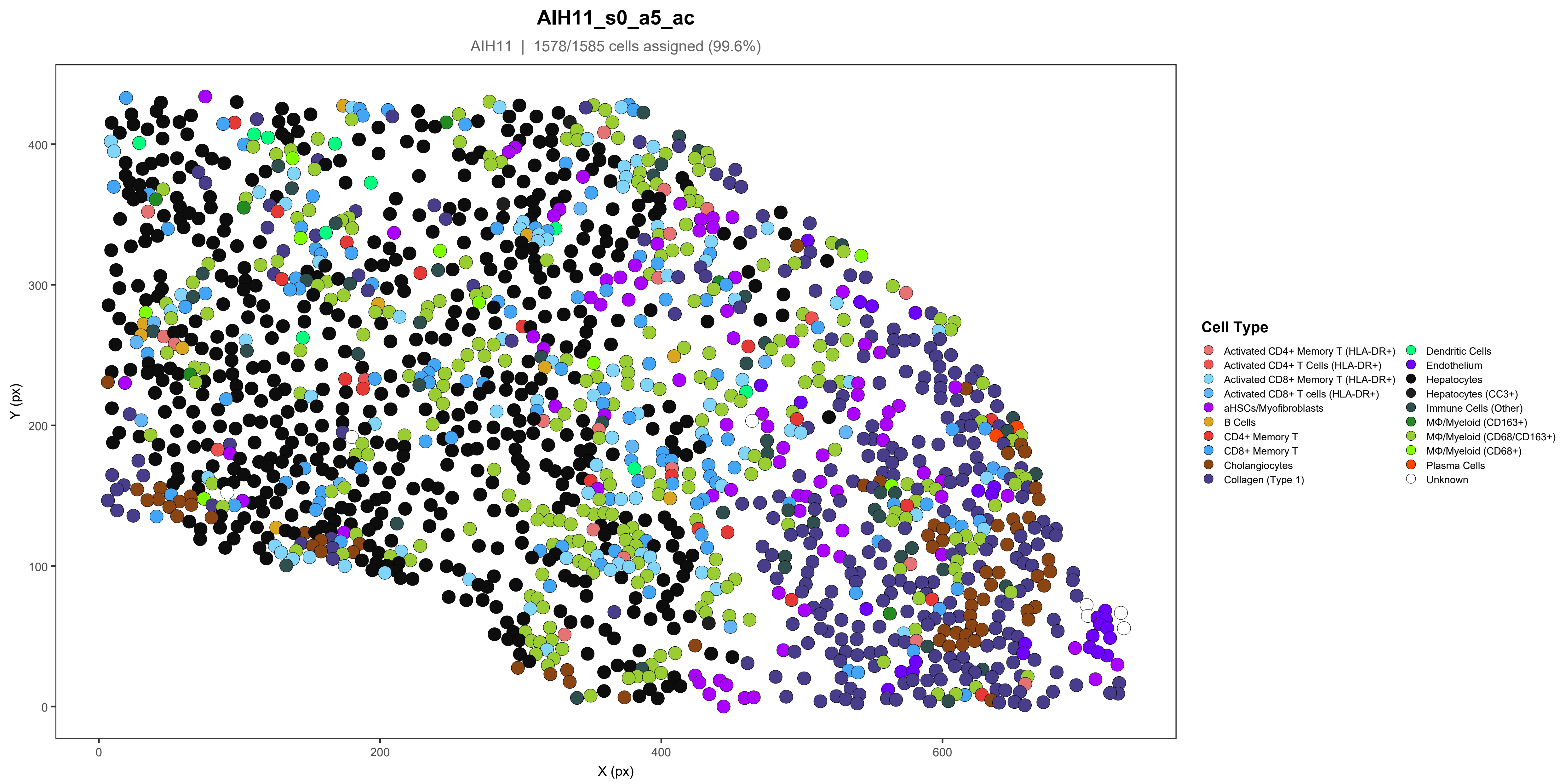
AIH11_s0_a5_ac
AIH11 | 1578/1585 cells assigned (99.6%)
Y (px)
X (px)
0
100
200
300
400
0
200
400
600
Cell Type
Activated CD4+ Memory T (HLA-DR+)
Activated CD4+ T Cells (HLA-DR+)
Activated CD8+ Memory T (HLA-DR+)
Activated CD8+ T cells (HLA-DR+)
aHSCs/Myofibroblasts
B Cells
CD4+ Memory T
CD8+ Memory T
Cholangiocytes
Collagen (Type 1)
Dendritic Cells
Endothelium
Hepatocytes
Hepatocytes (CC3+)
Immune Cells (Other)
MΦ/Myeloid (CD163+)
MΦ/Myeloid (CD68/CD163+)
MΦ/Myeloid (CD68+)
Plasma Cells
Unknown

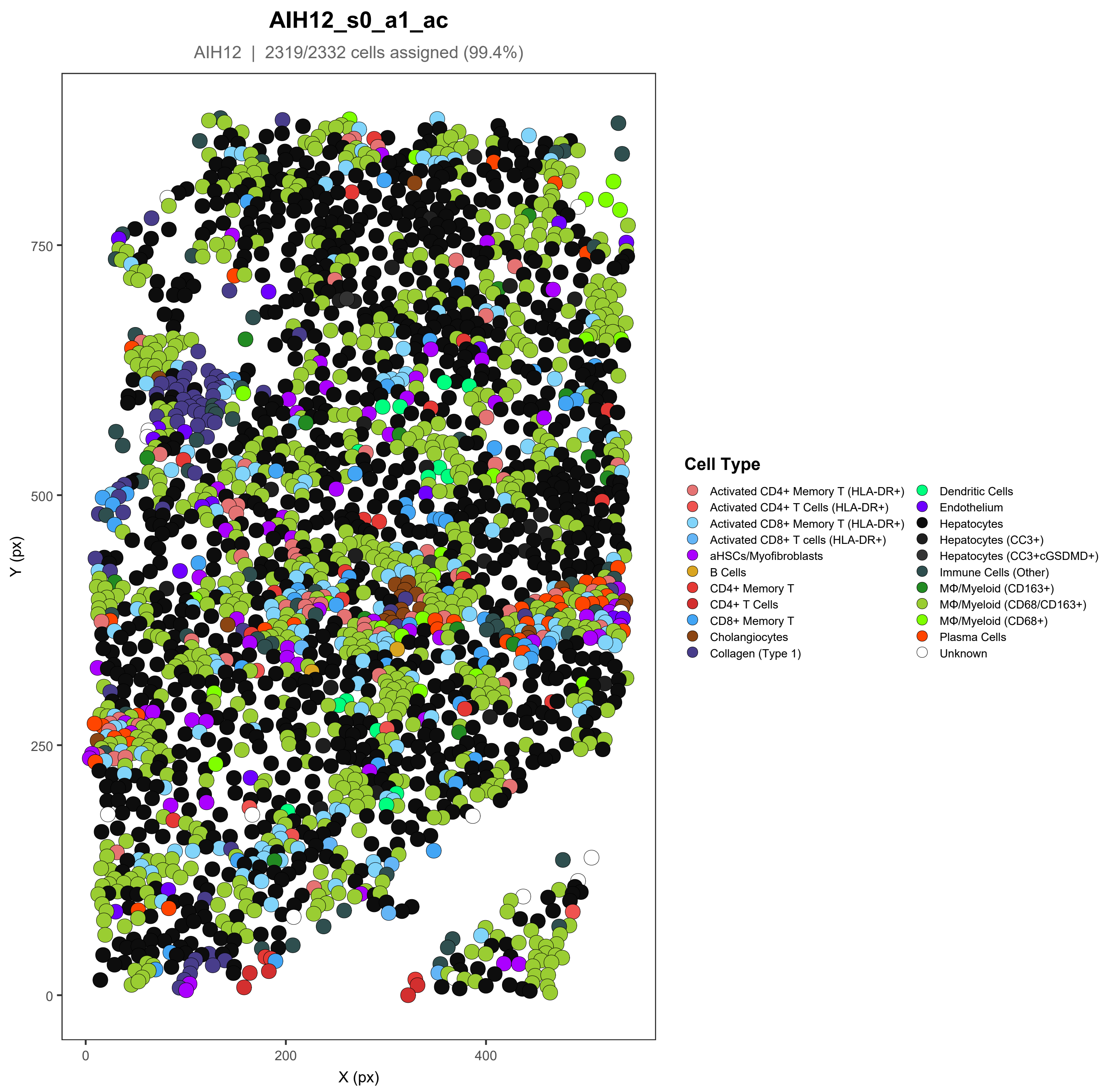

AIH12_s0_a1_ac
AIH12 | 2319/2332 cells assigned (99.4%)
Y (px)
X (px)
0
250
500
750
0
200
400
Cell Type
Activated CD4+ Memory T (HLA-DR+)
Activated CD4+ T Cells (HLA-DR+)
Activated CD8+ Memory T (HLA-DR+)
Activated CD8+ T cells (HLA-DR+)
aHSCs/Myofibroblasts
B Cells
CD4+ Memory T
CD4+ T Cells
CD8+ Memory T
Cholangiocytes
Collagen (Type 1)
Dendritic Cells
Endothelium
Hepatocytes
Hepatocytes (CC3+)
Hepatocytes (CC3+cGSDMD+)
Immune Cells (Other)
MΦ/Myeloid (CD163+)
MΦ/Myeloid (CD68/CD163+)
MΦ/Myeloid (CD68+)
Plasma Cells
Unknown

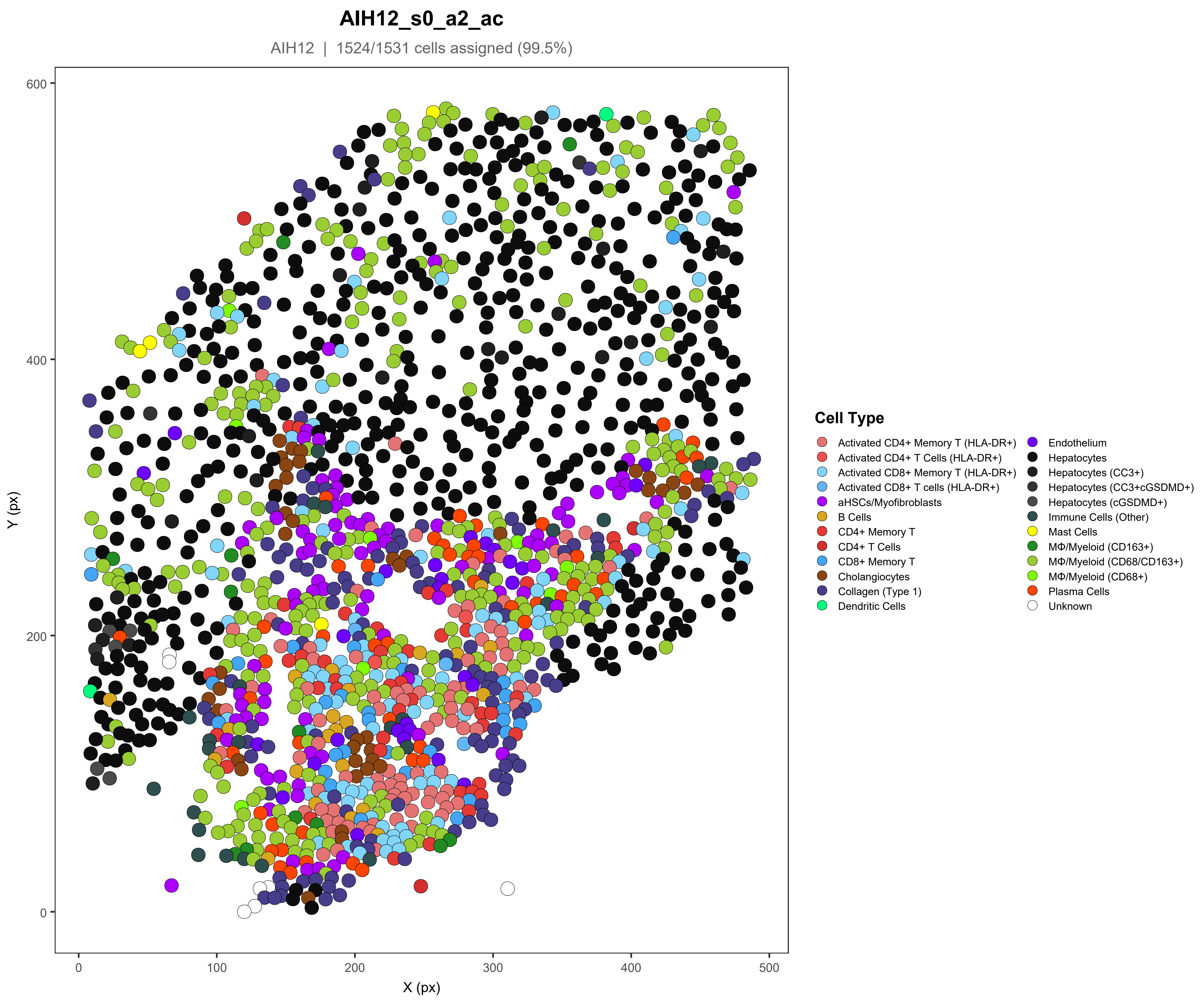

AIH12_s0_a2_ac
AIH12 | 1524/1531 cells assigned (99.5%)
Y (px)
X (px)
0
100
200
300
400
500
600
Cell Type
Activated CD4+ Memory T (HLA-DR+)
Activated CD4+ T Cells (HLA-DR+)
Activated CD8+ Memory T (HLA-DR+)
Activated CD8+ T cells (HLA-DR+)
aHSCs/Myofibroblasts
B Cells
CD4+ Memory T
CD4+ T Cells
CD8+ Memory T
Cholangiocytes
Collagen (Type 1)
Dendritic Cells
Endothelium
Hepatocytes
Hepatocytes (CC3+)
Hepatocytes (CC3+cGSDMD+)
Hepatocytes (cGSDMD+)
Immune Cells (Other)
Mast Cells
MΦ/Myeloid (CD163+)
MΦ/Myeloid (CD68/CD163+)
MΦ/Myeloid (CD68+)
Plasma Cells
Unknown

### AIH12_s0_a3_ac

AIH12 | 1708/1713 cells assigned (99.7%)

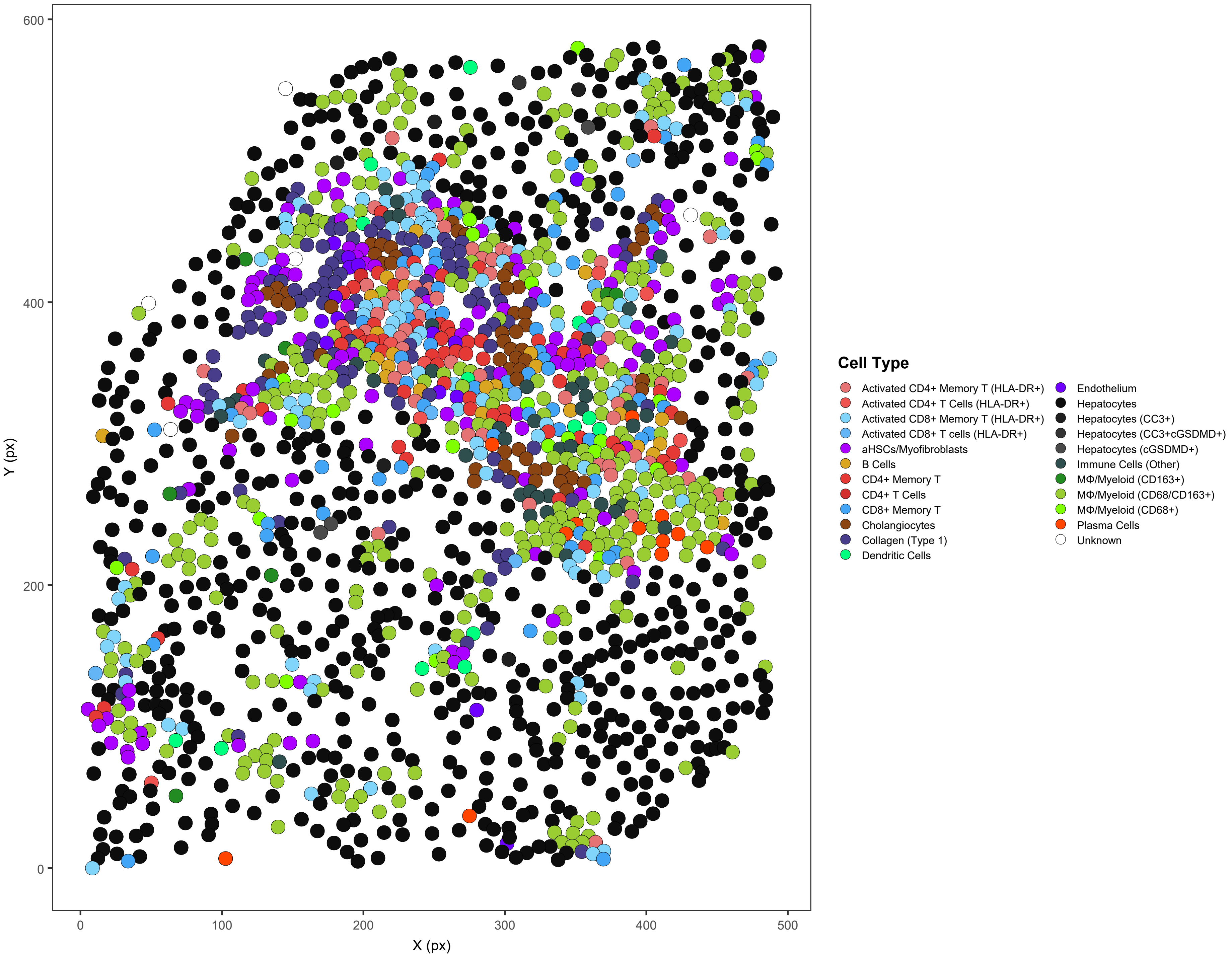

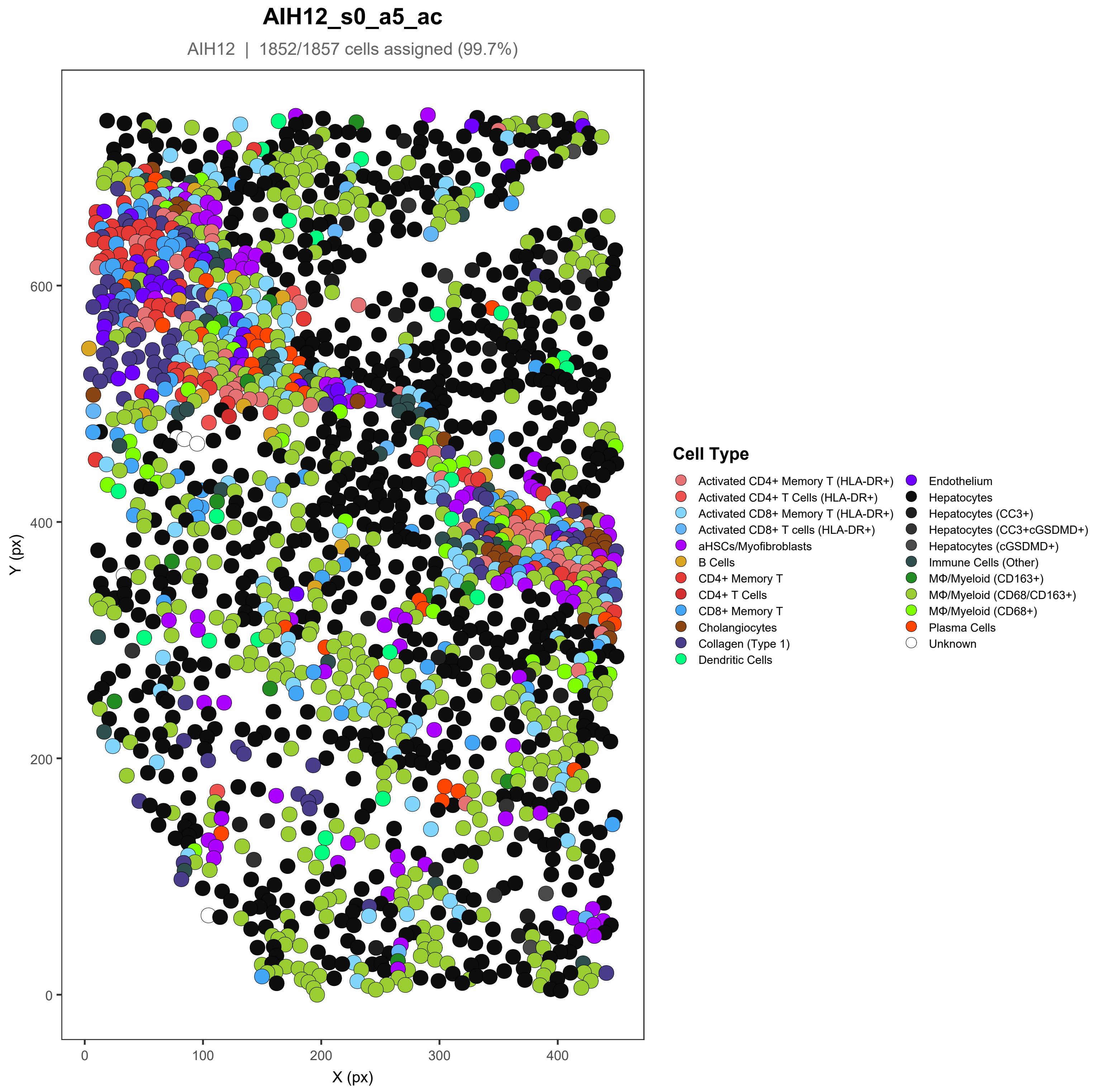

AIH12_s0_a5_ac
AIH12 | 1852/1857 cells assigned (99.7%)
Y (px)
X (px)
0
200
400
600
0
100
200
300
400
Cell Type
Activated CD4+ Memory T (HLA-DR+)
Activated CD4+ T Cells (HLA-DR+)
Activated CD8+ Memory T (HLA-DR+)
Activated CD8+ T cells (HLA-DR+)
aHSCs/Myofibroblasts
B Cells
CD4+ Memory T
CD4+ T Cells
CD8+ Memory T
Cholangiocytes
Collagen (Type 1)
Dendritic Cells
Endothelium
Hepatocytes
Hepatocytes (CC3+)
Hepatocytes (CC3+cGSDMD+)
Hepatocytes (cGSDMD+)
Immune Cells (Other)
MΦ/Myeloid (CD163+)
MΦ/Myeloid (CD68/CD163+)
MΦ/Myeloid (CD68+)
Plasma Cells
Unknown

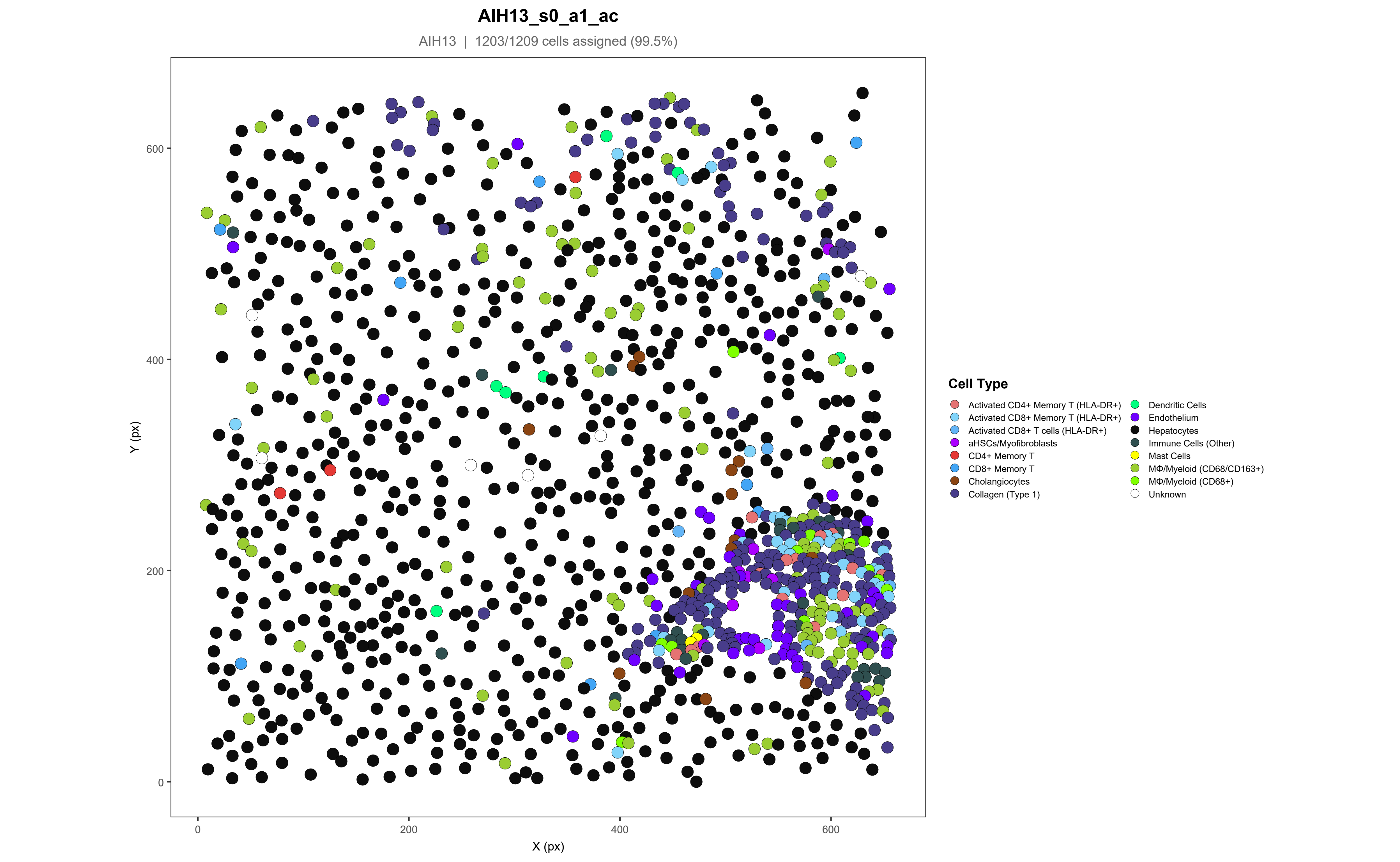
AIH13_s0_a1_ac
AIH13 | 1203/1209 cells assigned (99.5%)
Y (px)
X (px)
0
200
400
600
Cell Type
Activated CD4+ Memory T (HLA-DR+)
Activated CD8+ Memory T (HLA-DR+)
Activated CD8+ T cells (HLA-DR+)
aHSCs/Myofibroblasts
CD4+ Memory T
CD8+ Memory T
Cholangiocytes
Collagen (Type 1)
Dendritic Cells
Endothelium
Hepatocytes
Immune Cells (Other)
Mast Cells
MΦ/Myeloid (CD68/CD163+)
MΦ/Myeloid (CD68+)
Unknown

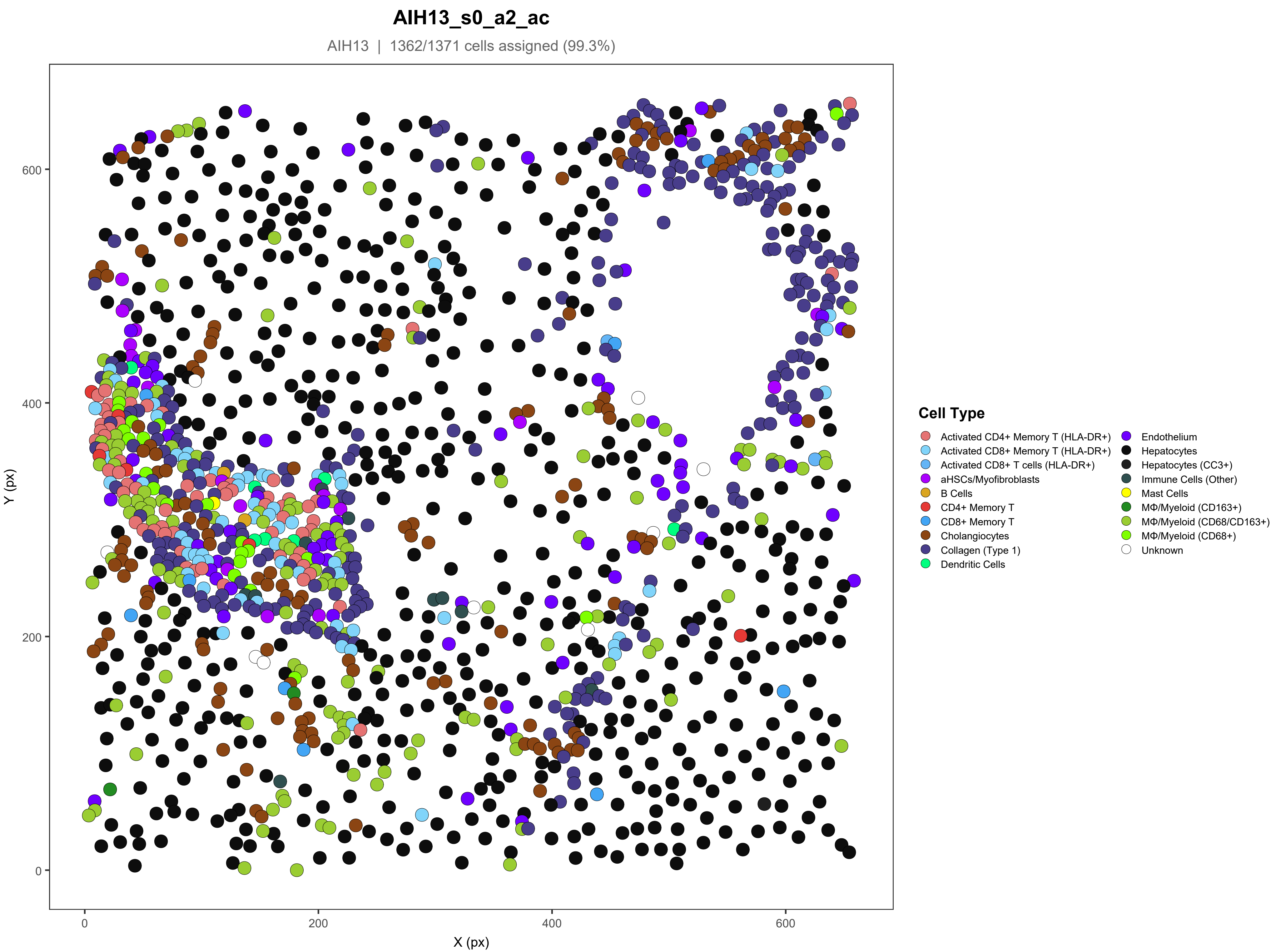

AIH13_s0_a2_ac
AIH13 | 1362/1371 cells assigned (99.3%)
Y (px)
X (px)
0
200
400
600
Cell Type
Activated CD4+ Memory T (HLA-DR+)
Activated CD8+ Memory T (HLA-DR+)
Activated CD8+ T cells (HLA-DR+)
aHSCs/Myofibroblasts
B Cells
CD4+ Memory T
CD8+ Memory T
Cholangiocytes
Collagen (Type 1)
Dendritic Cells
Endothelium
Hepatocytes
Hepatocytes (CC3+)
Immune Cells (Other)
Mast Cells
MΦ/Myeloid (CD163+)
MΦ/Myeloid (CD68/CD163+)
MΦ/Myeloid (CD68+)
Unknown

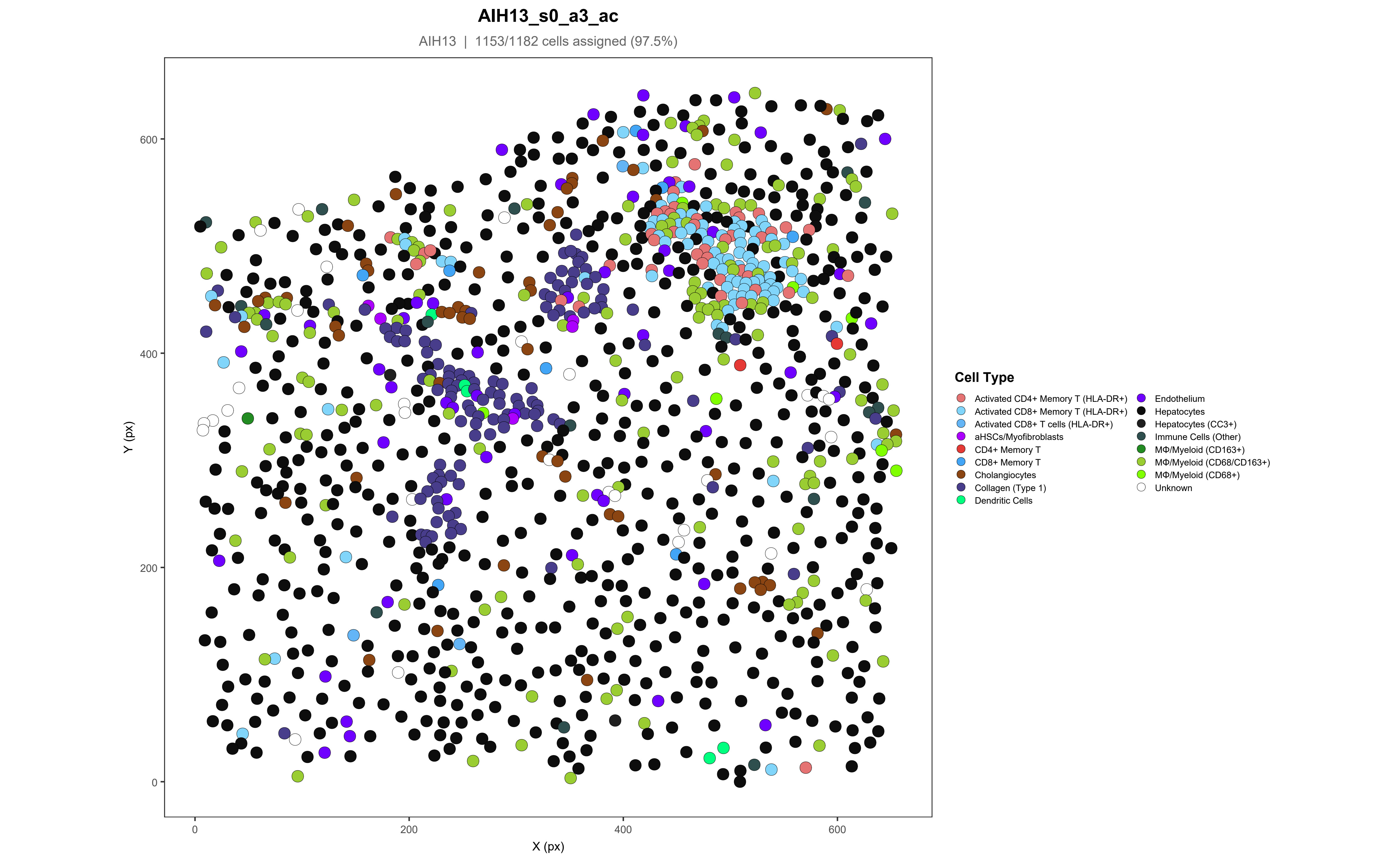

AIH13_s0_a3_ac
AIH13 | 1153/1182 cells assigned (97.5%)
Y (px)
X (px)
0
200
400
600
Cell Type
Activated CD4+ Memory T (HLA-DR+)
Activated CD8+ Memory T (HLA-DR+)
Activated CD8+ T cells (HLA-DR+)
aHSCs/Myofibroblasts
CD4+ Memory T
CD8+ Memory T
Cholangiocytes
Collagen (Type 1)
Dendritic Cells
Endothelium
Hepatocytes
Hepatocytes (CC3+)
Immune Cells (Other)
MΦ/Myeloid (CD163+)
MΦ/Myeloid (CD68/CD163+)
MΦ/Myeloid (CD68+)
Unknown

### AIH13_s0_a4_ac

AIH13 | 1636/1641 cells assigned (99.7%)

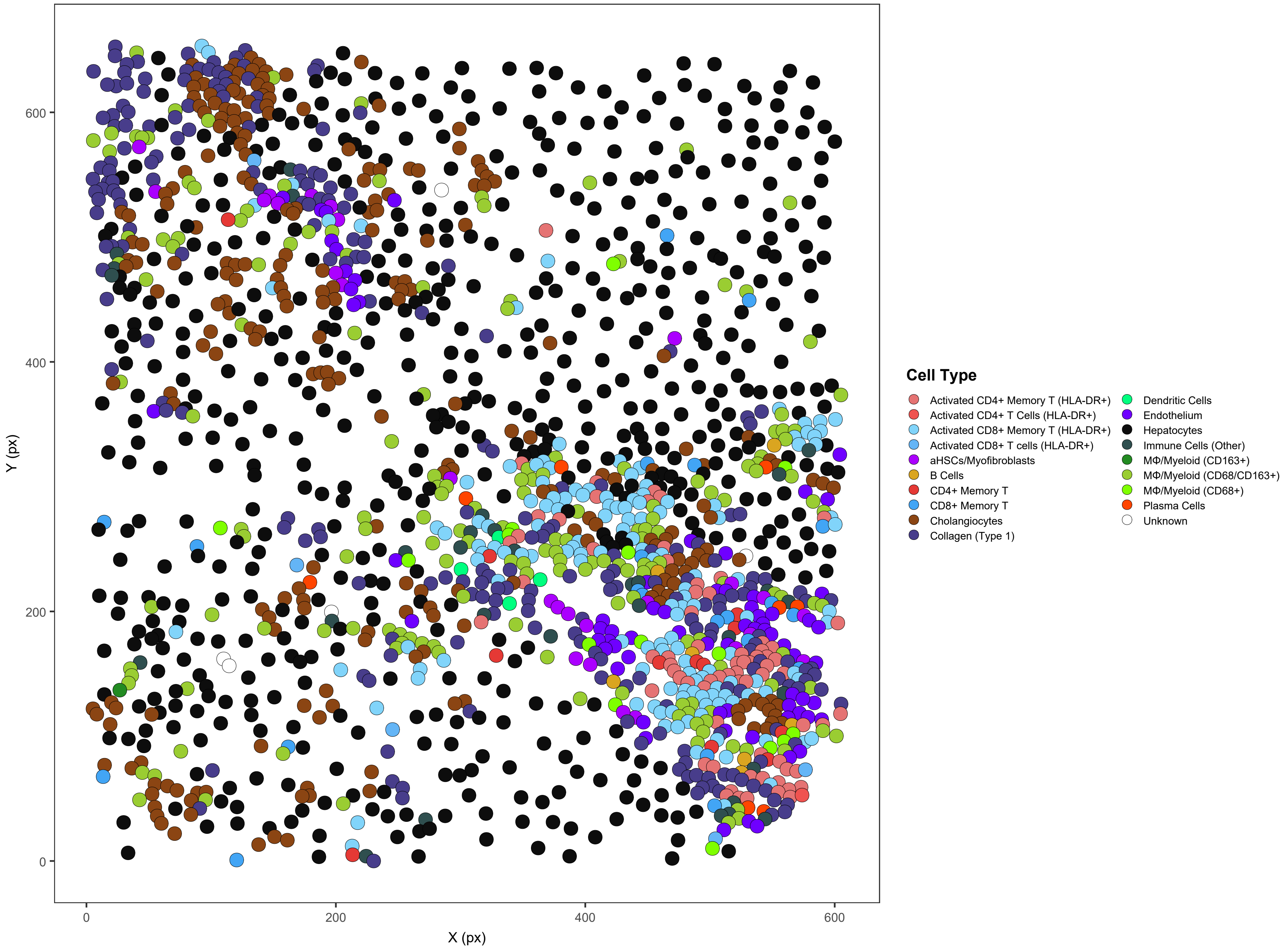

#### AIH13_s0_a5_ac

AIH13 | 1138/1141 cells assigned (99.7%)

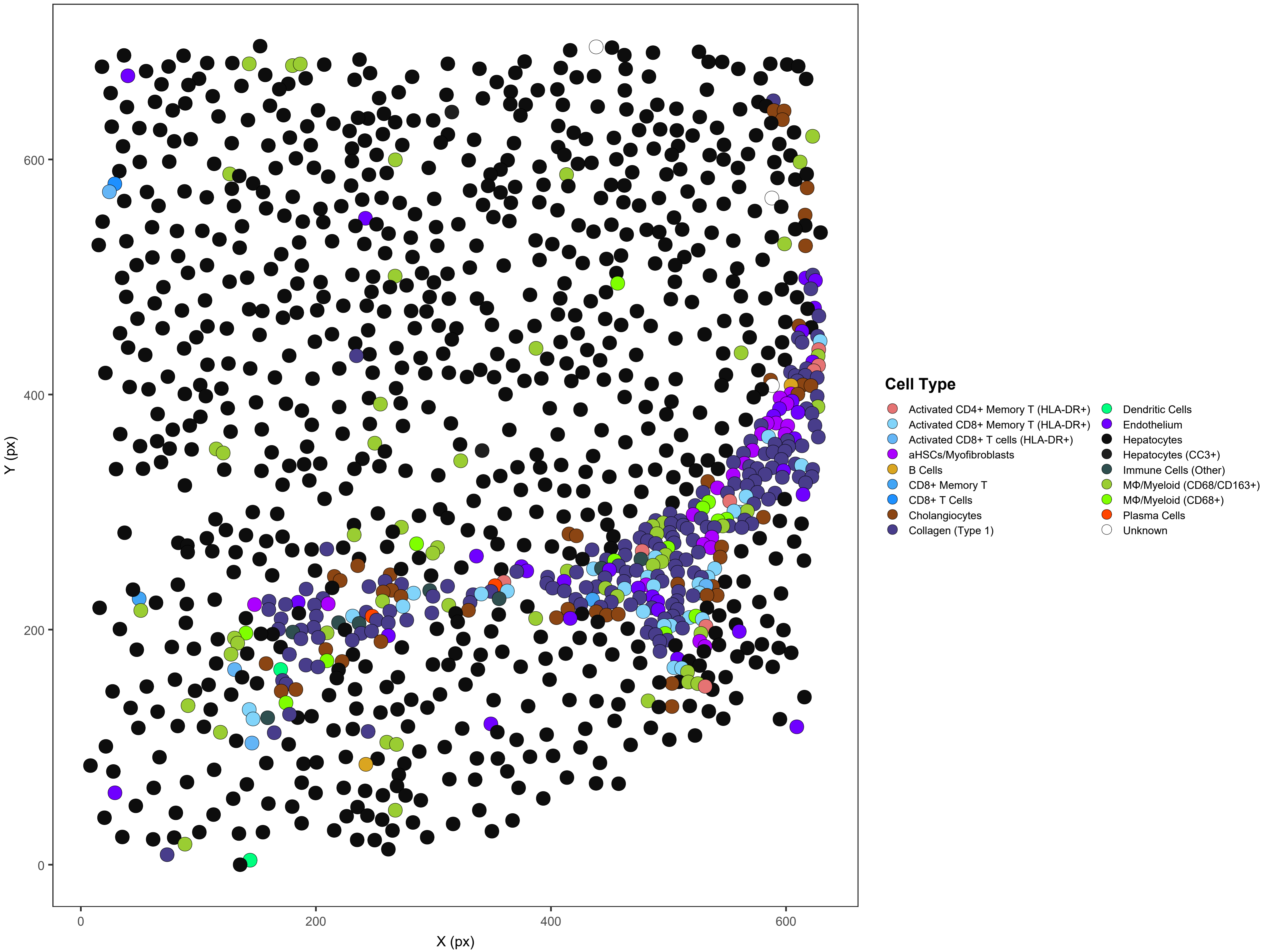

### AIH14_s0_a1_ac

AIH14 | 3916/3917 cells assigned (100%)

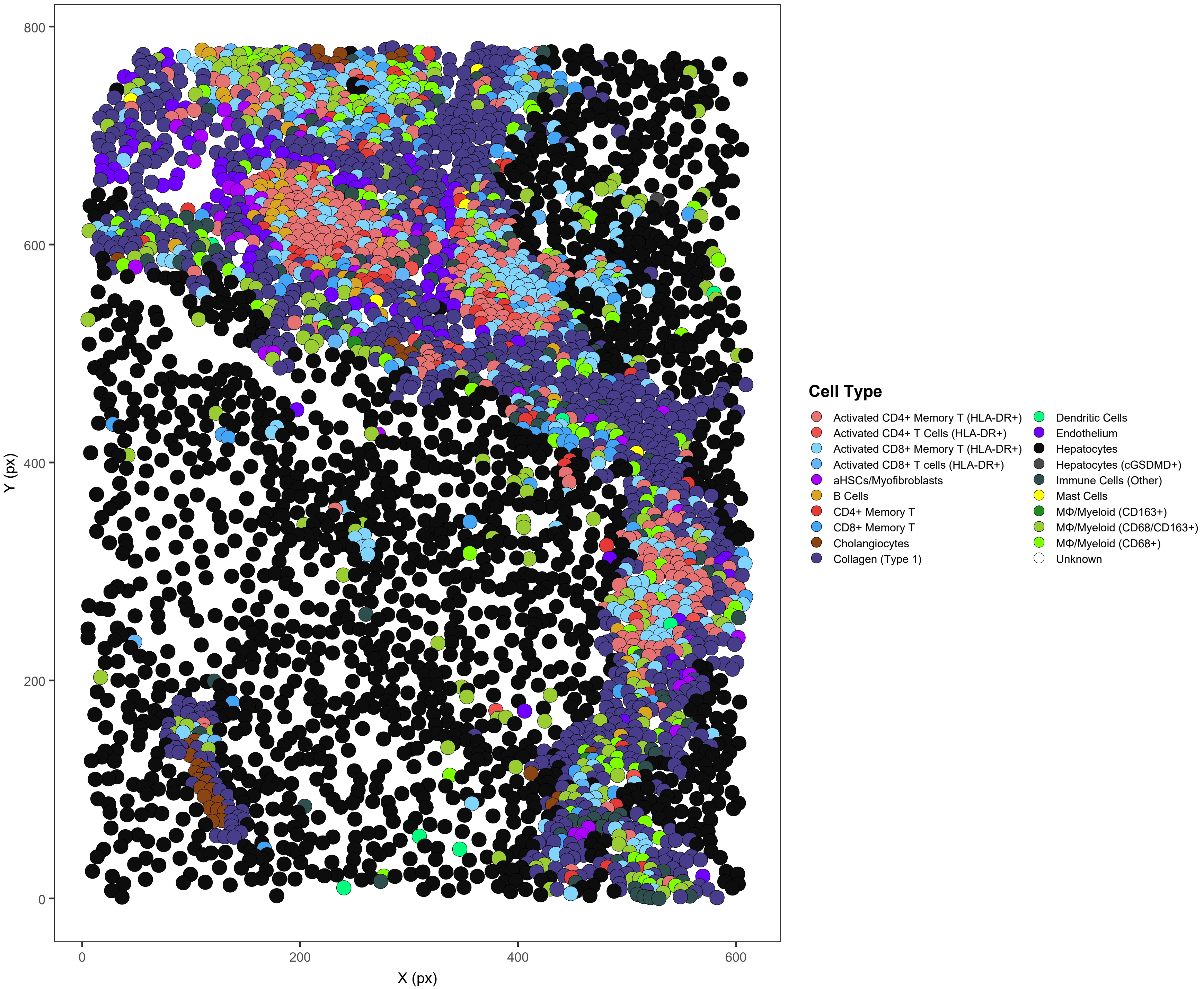

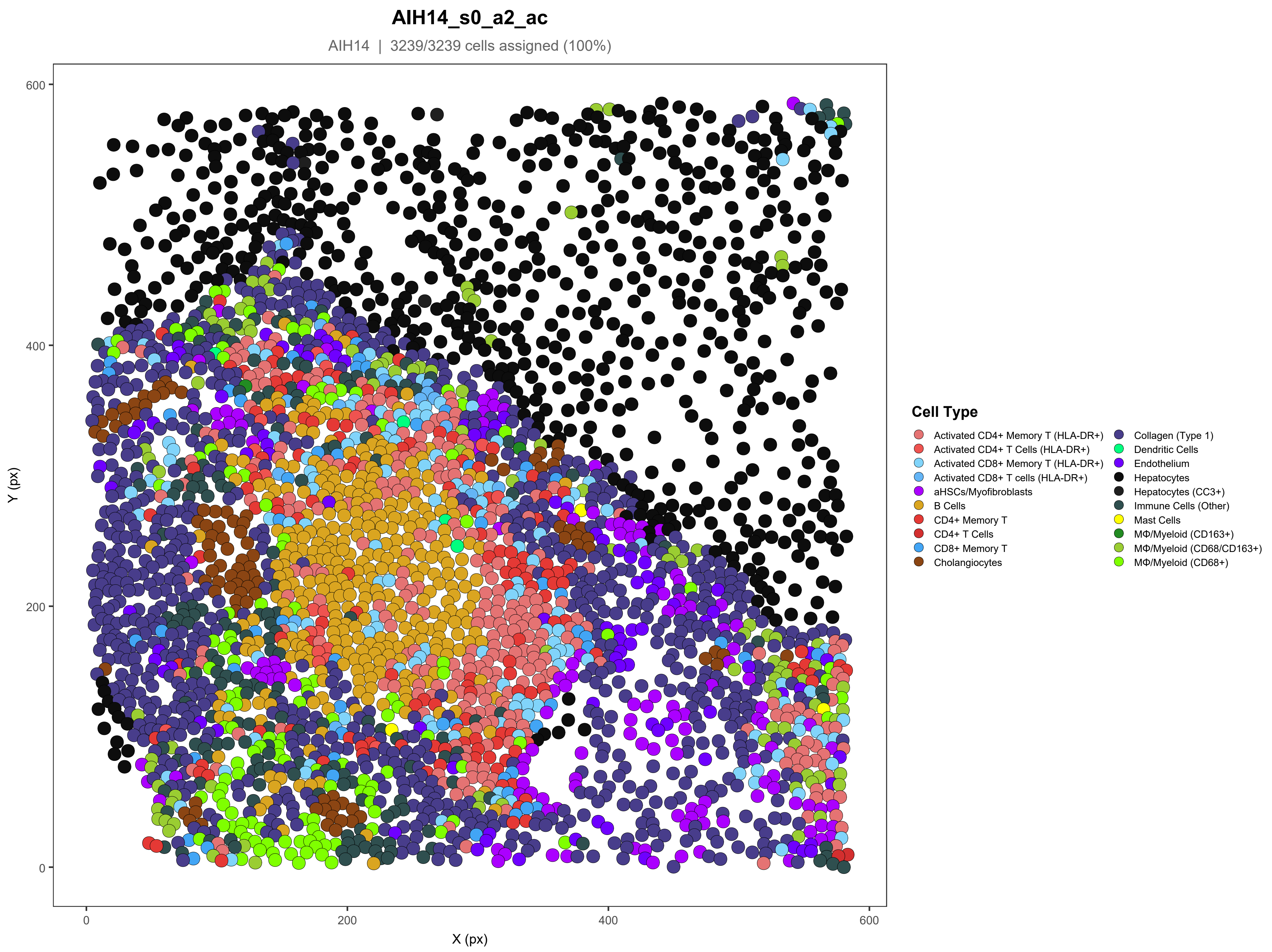
AIH14_s0_a2_ac
AIH14 | 3239/3239 cells assigned (100%)
Y (px)
X (px)
0
200
400
600
Cell Type
Activated CD4+ Memory T (HLA-DR+)
Activated CD4+ T Cells (HLA-DR+)
Activated CD8+ Memory T (HLA-DR+)
Activated CD8+ T cells (HLA-DR+)
aHSCs/Myofibroblasts
B Cells
CD4+ Memory T
CD4+ T Cells
CD8+ Memory T
Cholangiocytes
Collagen (Type 1)
Dendritic Cells
Endothelium
Hepatocytes
Hepatocytes (CC3+)
Immune Cells (Other)
Mast Cells
MΦ/Myeloid (CD163+)
MΦ/Myeloid (CD68/CD163+)
MΦ/Myeloid (CD68+)

#### AIH14_s0_a4_ac

AIH14 | 1566/1572 cells assigned (99.6%)

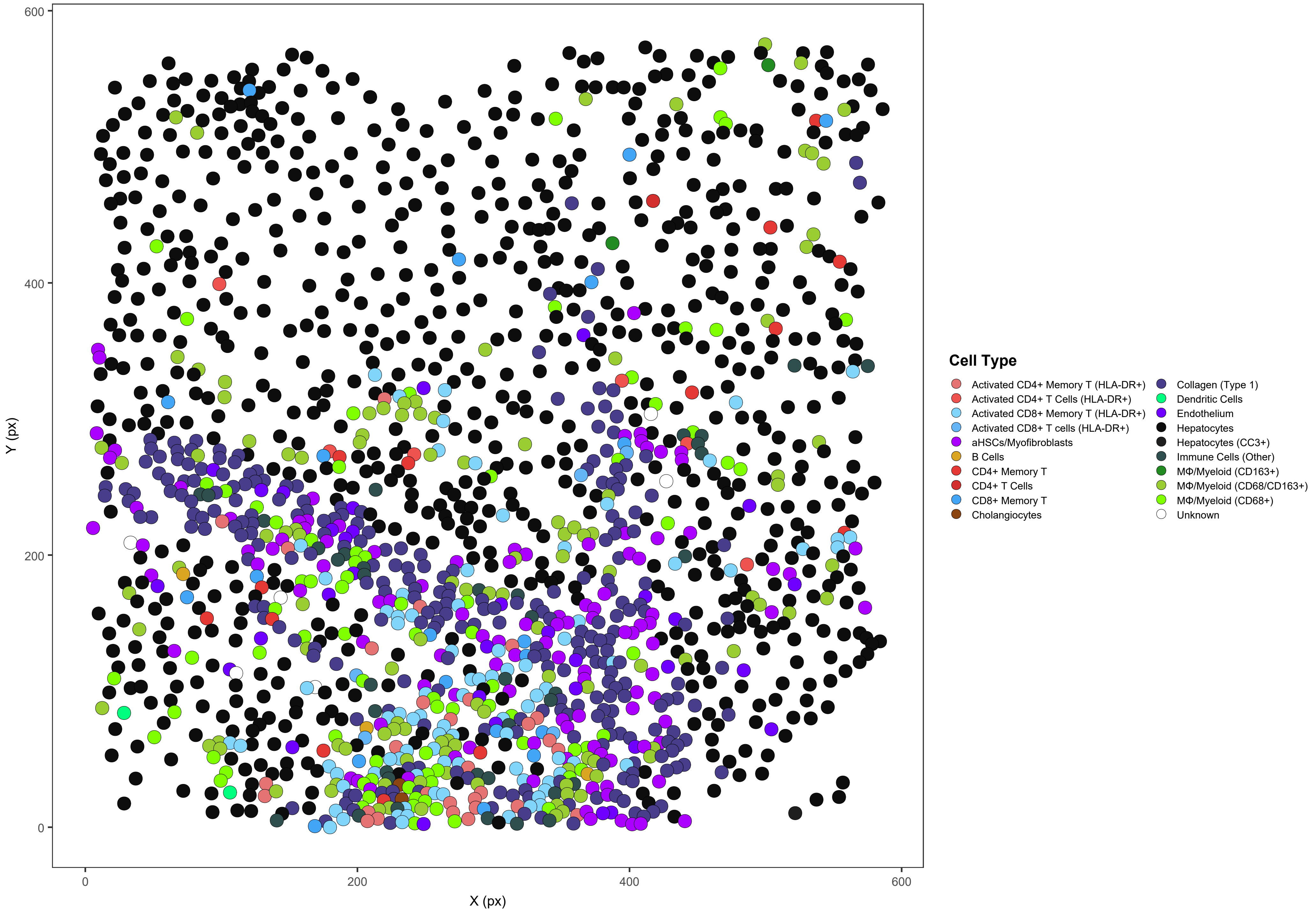

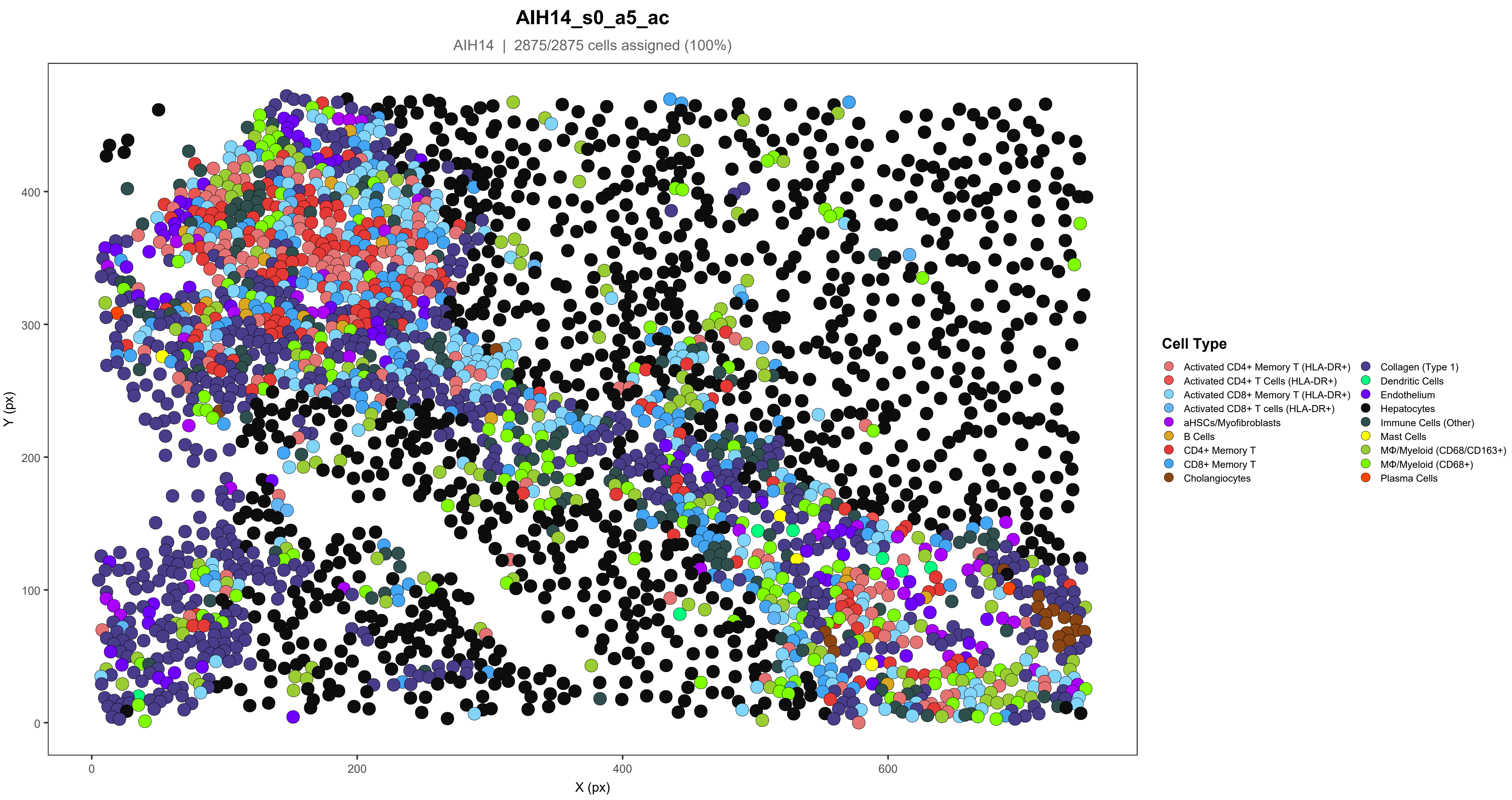
AIH14_s0_a5_ac
AIH14 | 2875/2875 cells assigned (100%)
Y (px)
X (px)
0
100
200
300
400
600
Cell Type
Activated CD4+ Memory T (HLA-DR+)
Activated CD4+ T Cells (HLA-DR+)
Activated CD8+ Memory T (HLA-DR+)
Activated CD8+ T cells (HLA-DR+)
aHSCs/Myofibroblasts
B Cells
CD4+ Memory T
CD8+ Memory T
Cholangiocytes
Collagen (Type 1)
Dendritic Cells
Endothelium
Hepatocytes
Immune Cells (Other)
Mast Cells
MΦ/Myeloid (CD68/CD163+)
MΦ/Myeloid (CD68+)
Plasma Cells

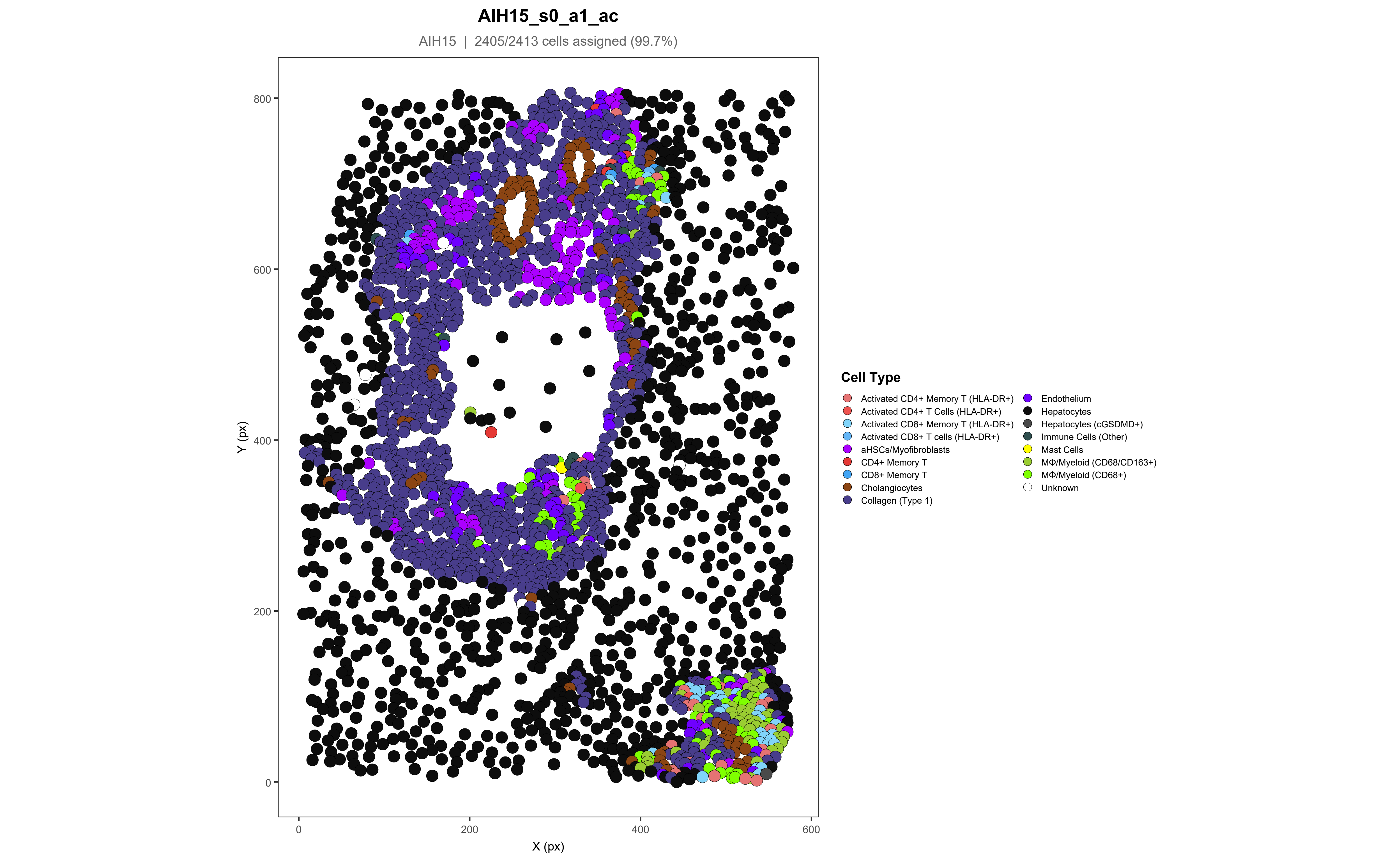
AIH15_s0_a1_ac
AIH15 | 2405/2413 cells assigned (99.7%)
Y (px)
X (px)
0
200
400
600
800
Cell Type
Activated CD4+ Memory T (HLA-DR+)
Activated CD4+ T Cells (HLA-DR+)
Activated CD8+ Memory T (HLA-DR+)
Activated CD8+ T cells (HLA-DR+)
aHSCs/Myofibroblasts
CD4+ Memory T
CD8+ Memory T
Cholangiocytes
Collagen (Type 1)
Endothelium
Hepatocytes
Hepatocytes (cGSDMD+)
Immune Cells (Other)
Mast Cells
MΦ/Myeloid (CD68/CD163+)
MΦ/Myeloid (CD68+)
Unknown

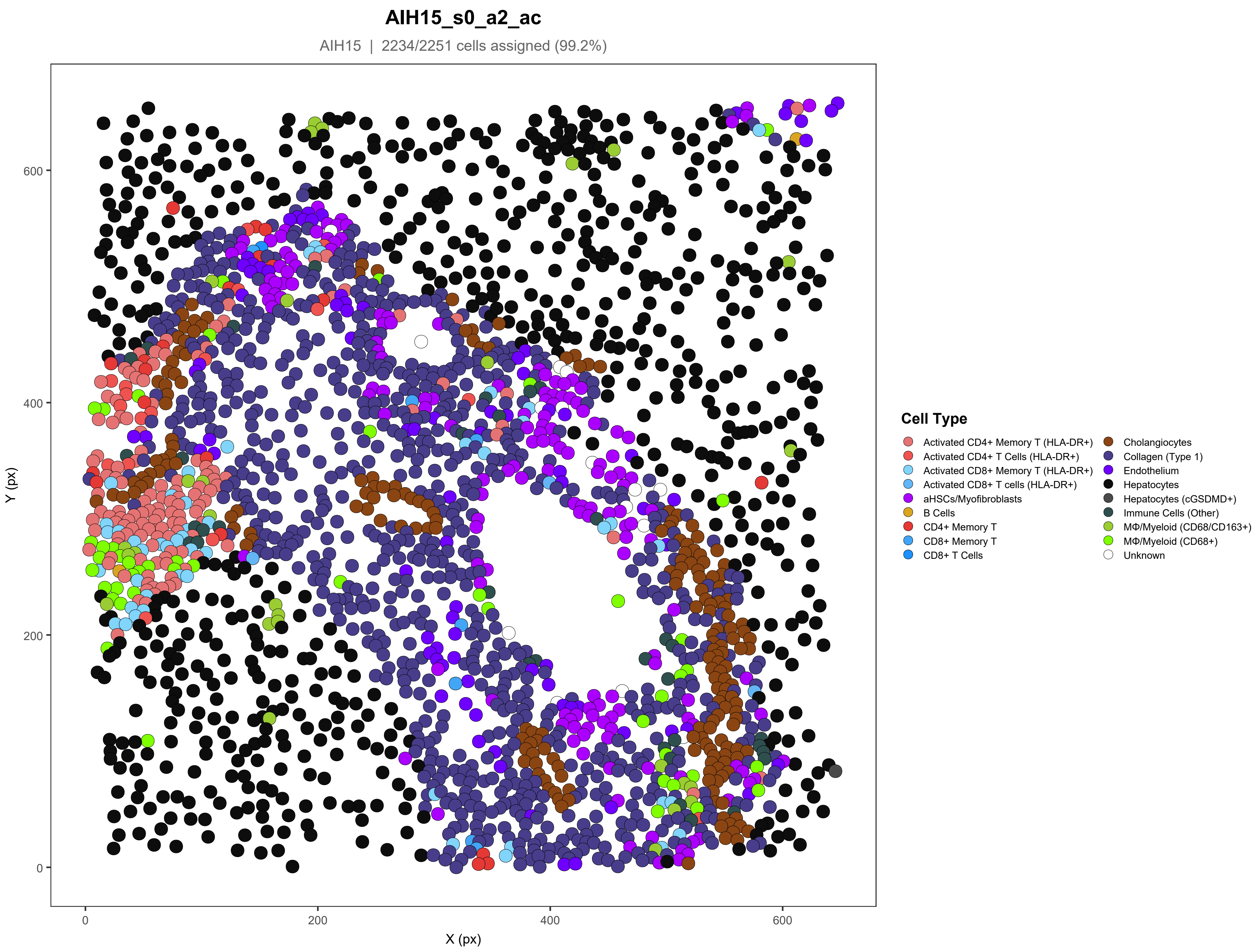
AIH15_s0_a2_ac
AIH15 | 2234/2251 cells assigned (99.2%)
Y (px)
X (px)
0
200
400
600
Cell Type
Activated CD4+ Memory T (HLA-DR+)
Activated CD4+ T Cells (HLA-DR+)
Activated CD8+ Memory T (HLA-DR+)
Activated CD8+ T cells (HLA-DR+)
aHSCs/Myofibroblasts
B Cells
CD4+ Memory T
CD8+ Memory T
CD8+ T Cells
Cholangiocytes
Collagen (Type 1)
Endothelium
Hepatocytes
Hepatocytes (cGSDMD+)
Immune Cells (Other)
MΦ/Myeloid (CD68/CD163+)
MΦ/Myeloid (CD68+)
Unknown

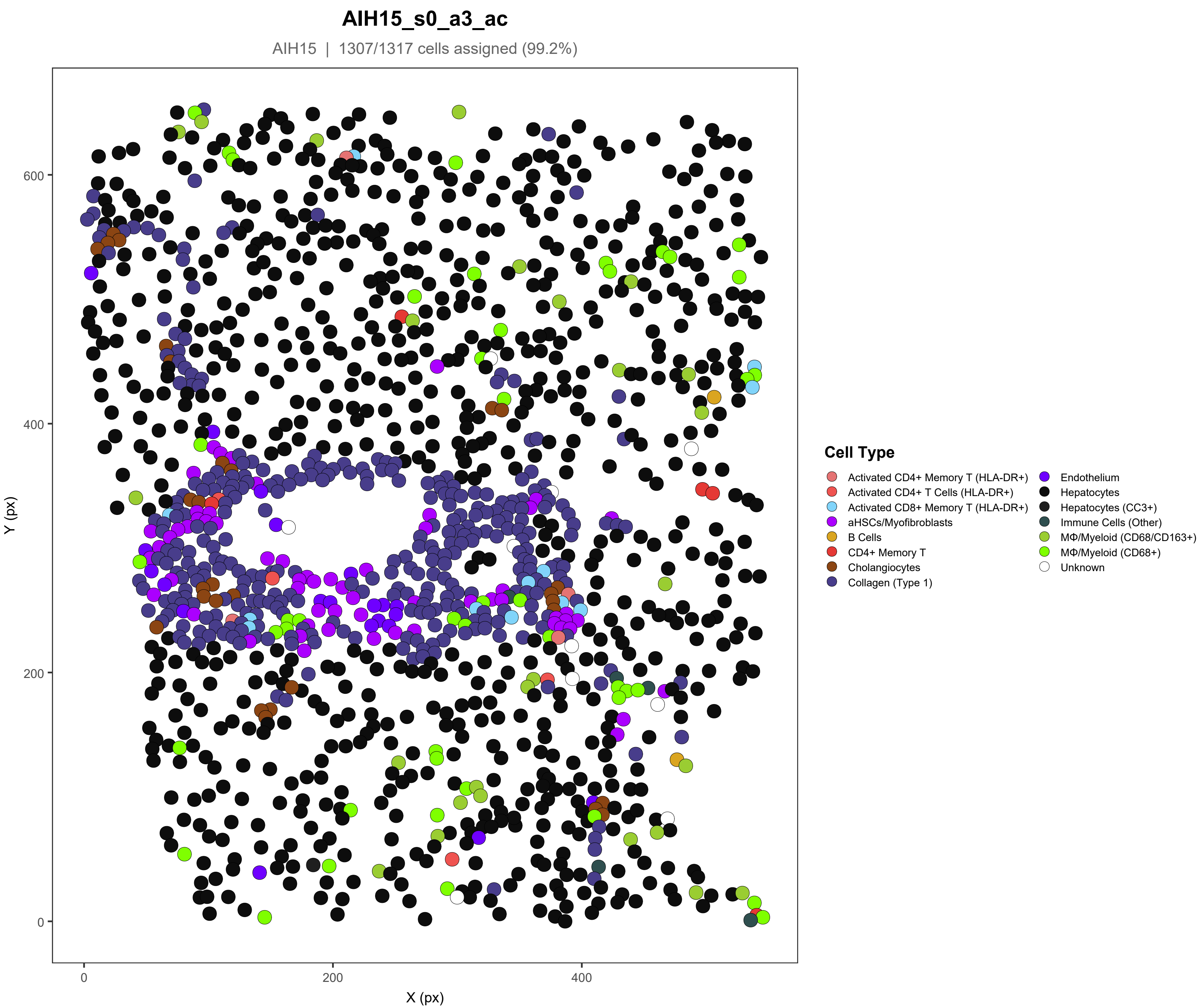
AIH15_s0_a3_ac
AIH15 | 1307/1317 cells assigned (99.2%)
Y (px)
X (px)
0
200
400
600
Cell Type
Activated CD4+ Memory T (HLA-DR+)
Activated CD4+ T Cells (HLA-DR+)
Activated CD8+ Memory T (HLA-DR+)
aHSCs/Myofibroblasts
B Cells
CD4+ Memory T
Cholangiocytes
Collagen (Type 1)
Endothelium
Hepatocytes
Hepatocytes (CC3+)
Immune Cells (Other)
MΦ/Myeloid (CD68/CD163+)
MΦ/Myeloid (CD68+)
Unknown

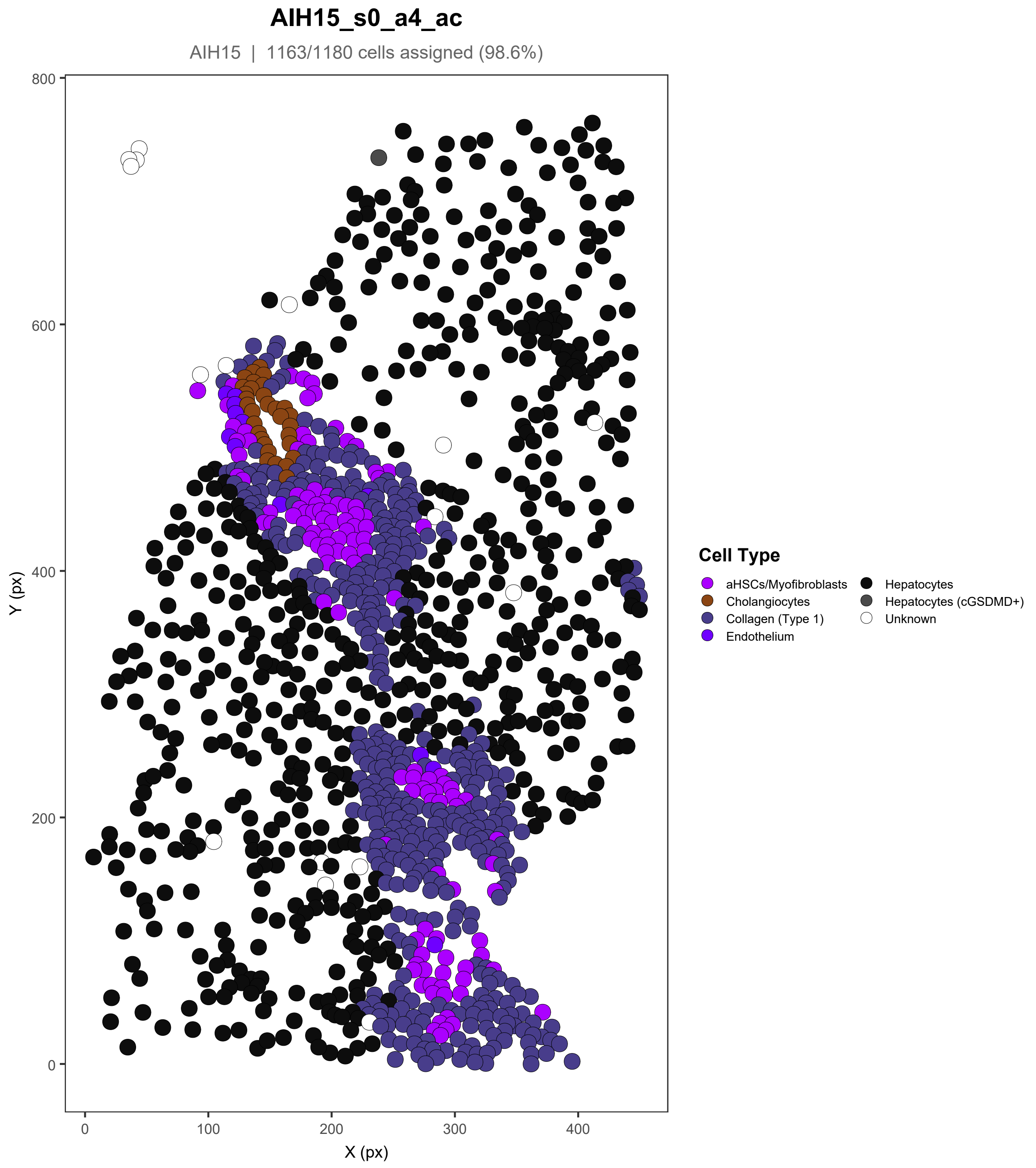

AIH15_s0_a4_ac
AIH15 | 1163/1180 cells assigned (98.6%)
Y (px)
X (px)
800
600
400
200
0
0
100
200
300
400
Cell Type
aHSCs/Myofibroblasts
Cholangiocytes
Collagen (Type 1)
Endothelium
Hepatocytes
Hepatocytes (cGSDMD+)
Unknown

#### AIH15_s0_a5_ac

AIH15 | 1363/1369 cells assigned (99.6%)

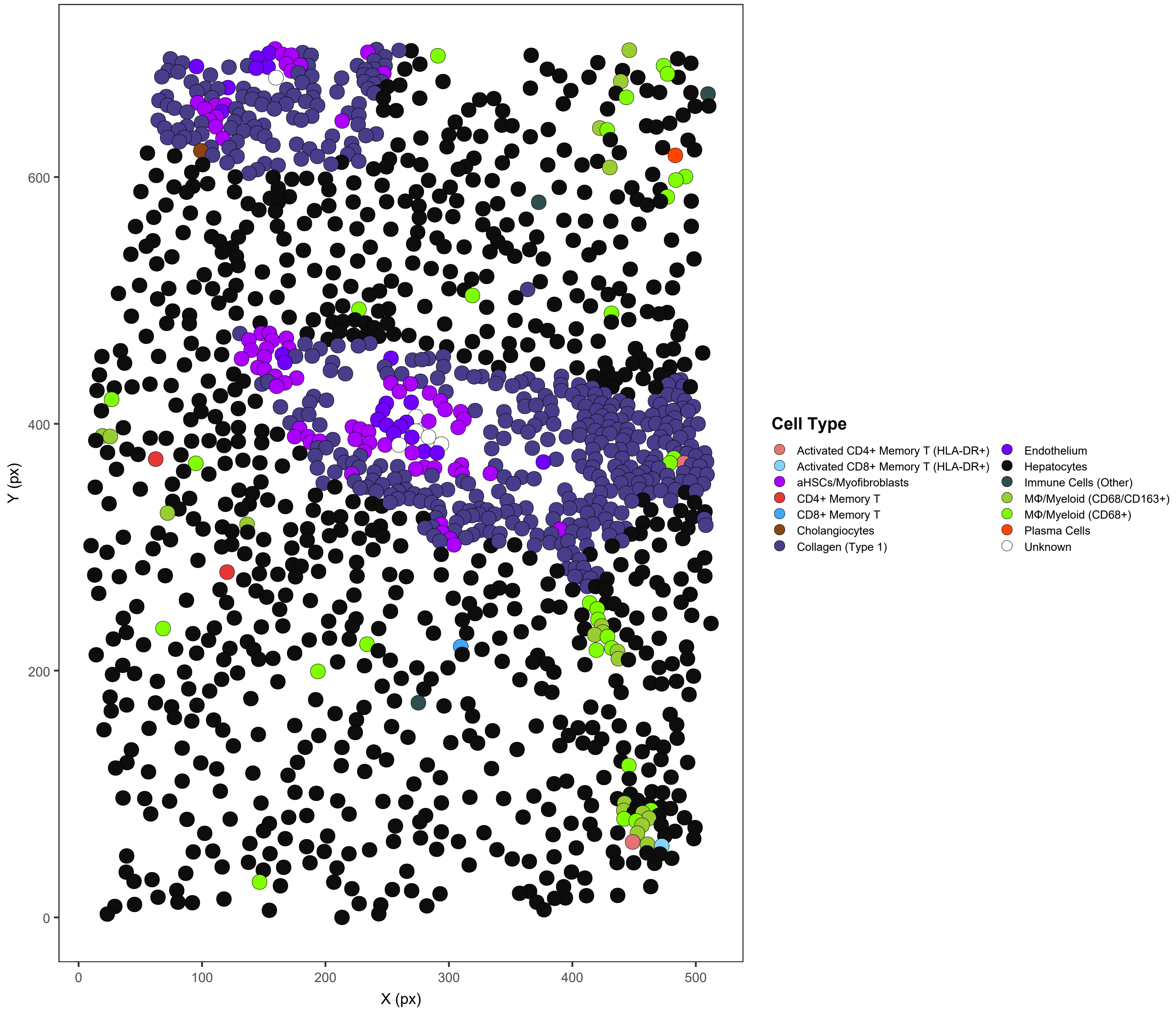

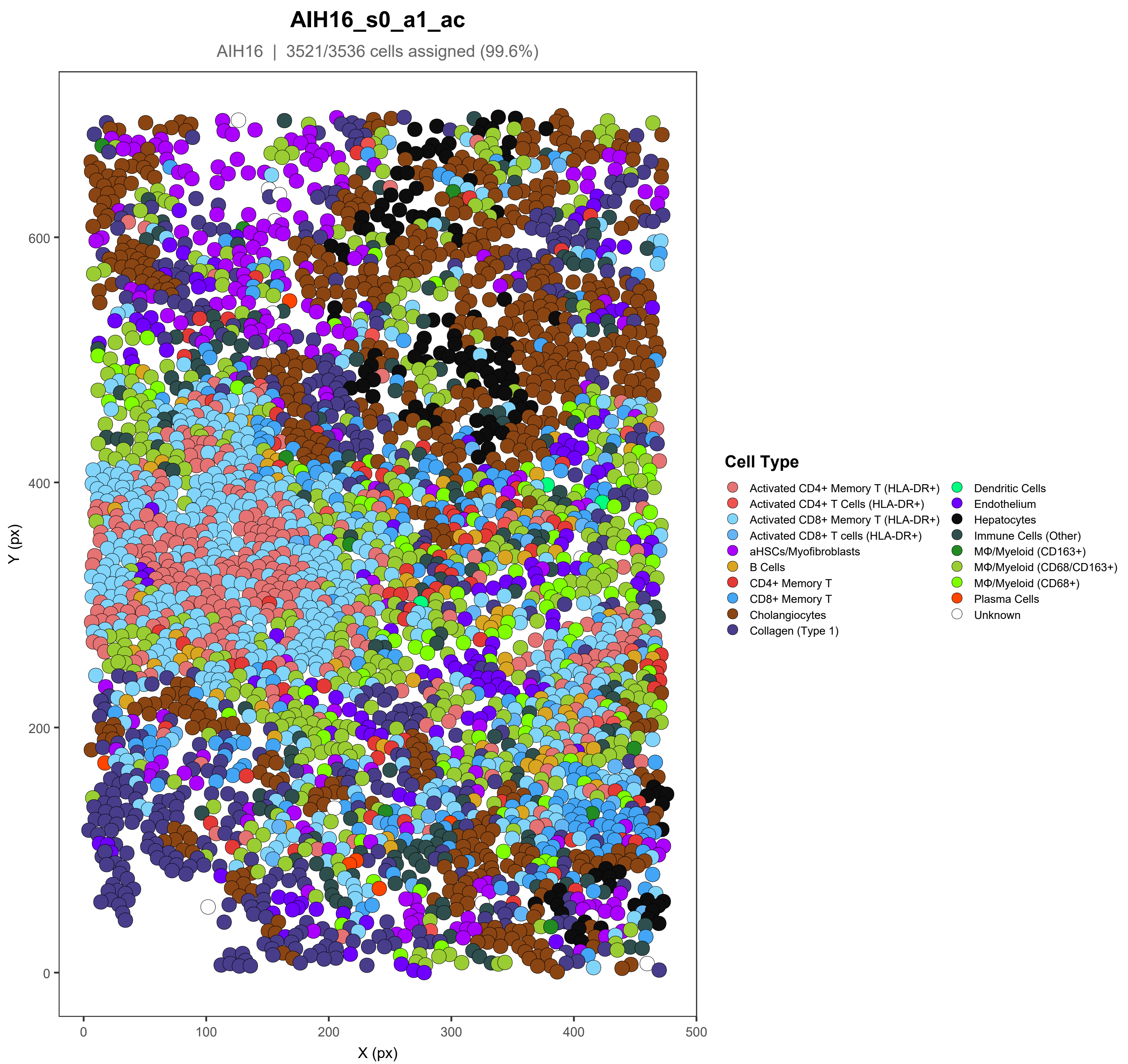
AIH16_s0_a1_ac
AIH16 | 3521/3536 cells assigned (99.6%)
Y (px)
X (px)
0
100
200
300
400
500
600
Cell Type
Activated CD4+ Memory T (HLA-DR+)
Activated CD4+ T Cells (HLA-DR+)
Activated CD8+ Memory T (HLA-DR+)
Activated CD8+ T cells (HLA-DR+)
aHSCs/Myofibroblasts
B Cells
CD4+ Memory T
CD8+ Memory T
Cholangiocytes
Collagen (Type 1)
Dendritic Cells
Endothelium
Hepatocytes
Immune Cells (Other)
MΦ/Myeloid (CD163+)
MΦ/Myeloid (CD68/CD163+)
MΦ/Myeloid (CD68+)
Plasma Cells
Unknown

#### AIH16_s0_a2_ac

AIH16 | 3346/3360 cells assigned (99.6%)

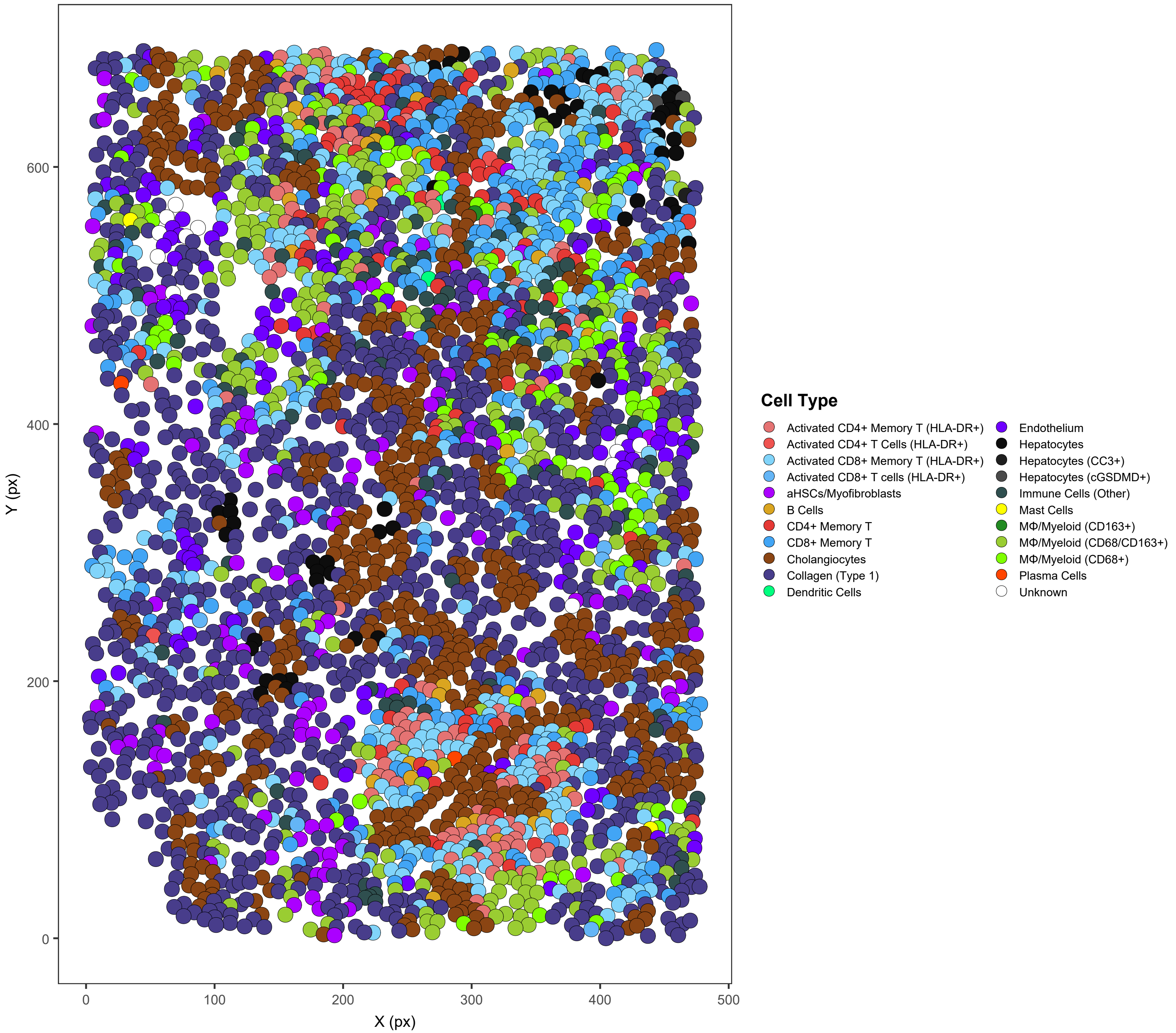

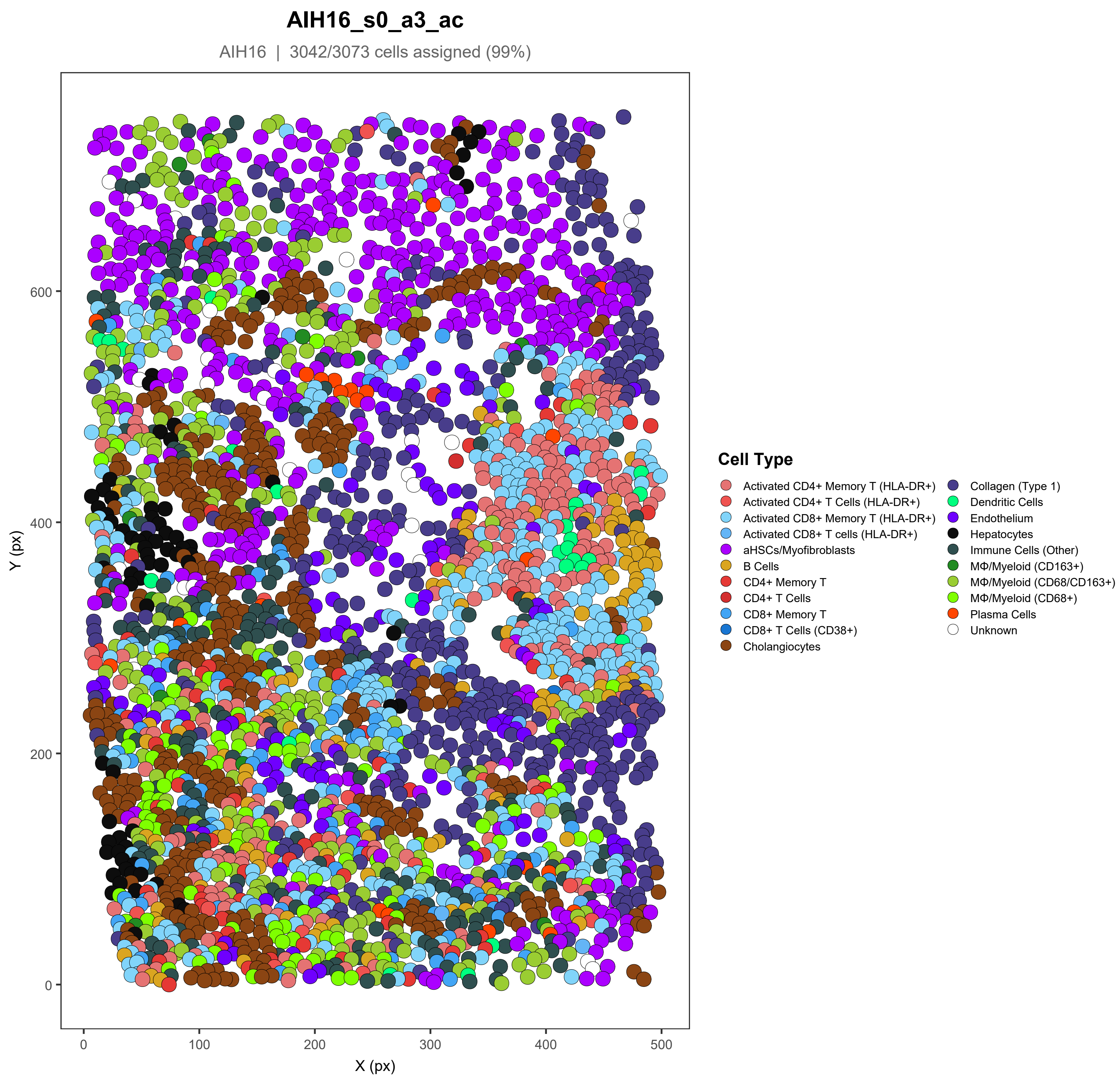

AIH16_s0_a3_ac
AIH16 | 3042/3073 cells assigned (99%)
Y (px)
X (px)
0
100
200
300
400
500
600
Cell Type
Activated CD4+ Memory T (HLA-DR+)
Activated CD4+ T Cells (HLA-DR+)
Activated CD8+ Memory T (HLA-DR+)
Activated CD8+ T cells (HLA-DR+)
aHSCs/Myofibroblasts
B Cells
CD4+ Memory T
CD4+ T Cells
CD8+ Memory T
CD8+ T Cells (CD38+)
Cholangiocytes
Collagen (Type 1)
Dendritic Cells
Endothelium
Hepatocytes
Immune Cells (Other)
MΦ/Myeloid (CD163+)
MΦ/Myeloid (CD68/CD163+)
MΦ/Myeloid (CD68+)
Plasma Cells
Unknown

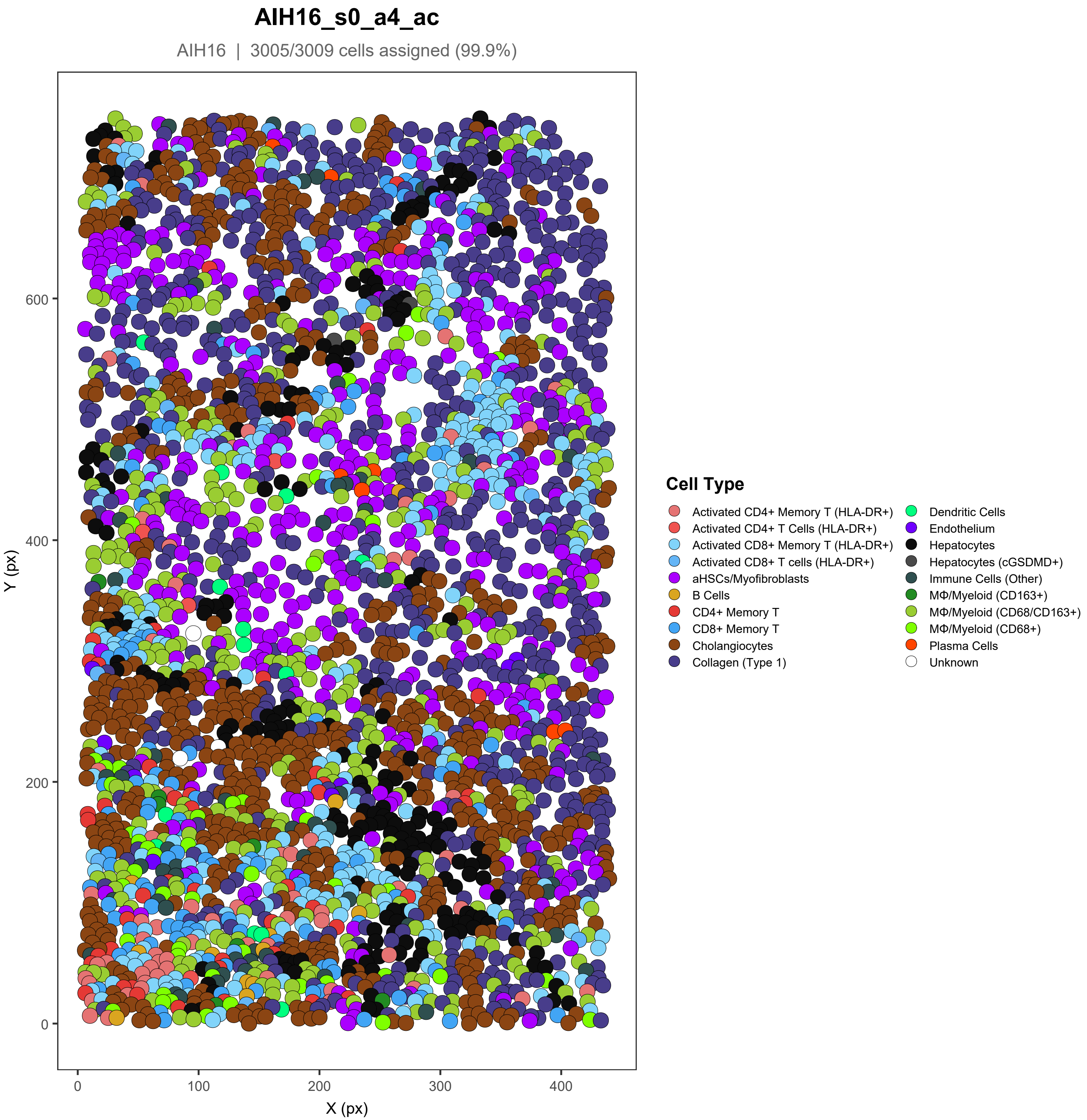

AIH16_s0_a4_ac
AIH16 | 3005/3009 cells assigned (99.9%)
Y (px)
X (px)
0
200
400
600
0
100
200
300
400
Cell Type
Activated CD4+ Memory T (HLA-DR+)
Activated CD4+ T Cells (HLA-DR+)
Activated CD8+ Memory T (HLA-DR+)
Activated CD8+ T cells (HLA-DR+)
aHSCs/Myofibroblasts
B Cells
CD4+ Memory T
CD8+ Memory T
Cholangiocytes
Collagen (Type 1)
Dendritic Cells
Endothelium
Hepatocytes
Hepatocytes (cGSDMD+)
Immune Cells (Other)
MΦ/Myeloid (CD163+)
MΦ/Myeloid (CD68/CD163+)
MΦ/Myeloid (CD68+)
Plasma Cells
Unknown

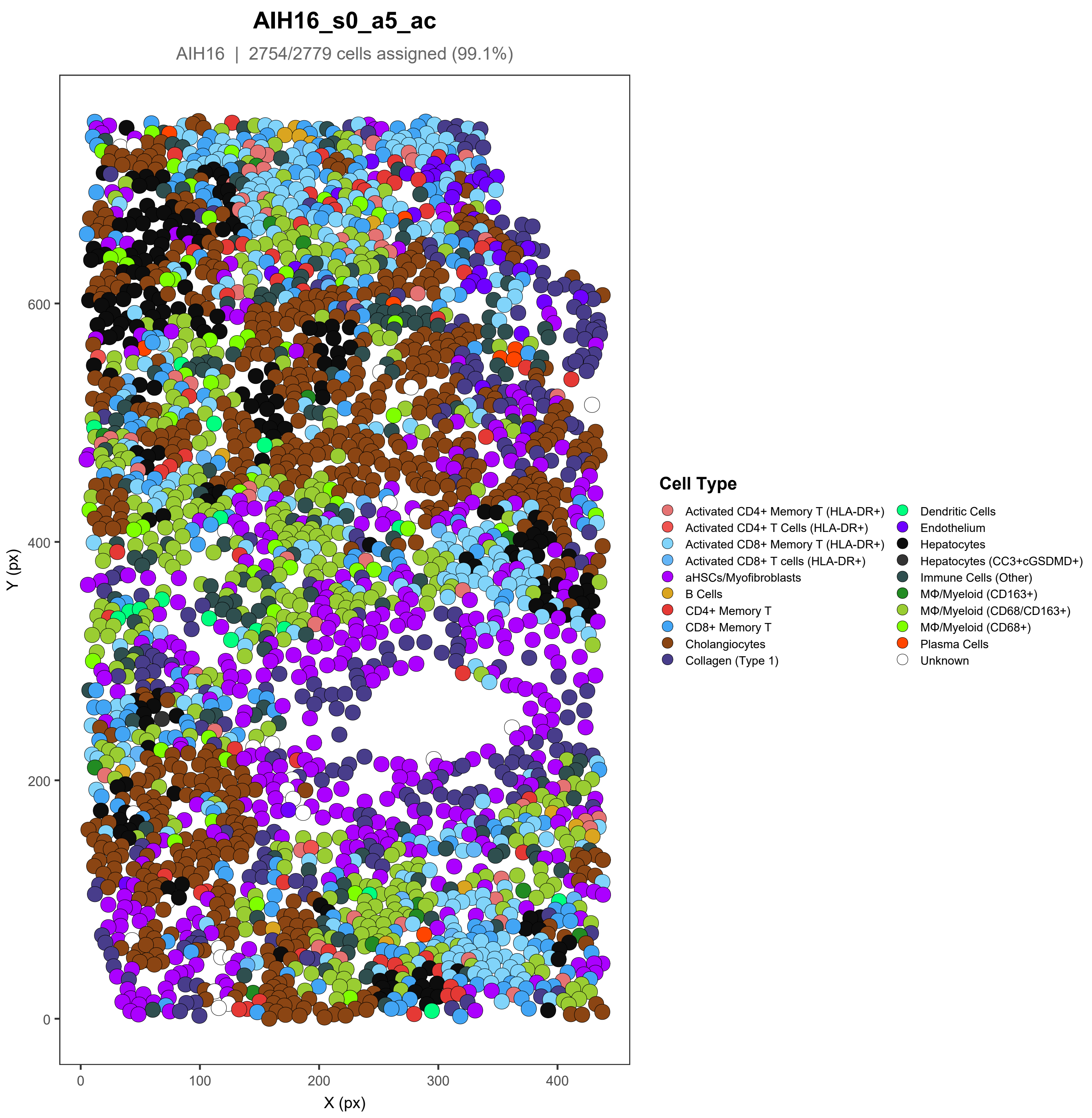
AIH16_s0_a5_ac
AIH16 | 2754/2779 cells assigned (99.1%)
Y (px)
X (px)
0
100
200
300
400
0
200
400
600
Cell Type
Activated CD4+ Memory T (HLA-DR+)
Activated CD4+ T Cells (HLA-DR+)
Activated CD8+ Memory T (HLA-DR+)
Activated CD8+ T cells (HLA-DR+)
aHSCs/Myofibroblasts
B Cells
CD4+ Memory T
CD8+ Memory T
Cholangiocytes
Collagen (Type 1)
Dendritic Cells
Endothelium
Hepatocytes
Hepatocytes (CC3+cGSDMD+)
Immune Cells (Other)
MΦ/Myeloid (CD163+)
MΦ/Myeloid (CD68/CD163+)
MΦ/Myeloid (CD68+)
Plasma Cells
Unknown

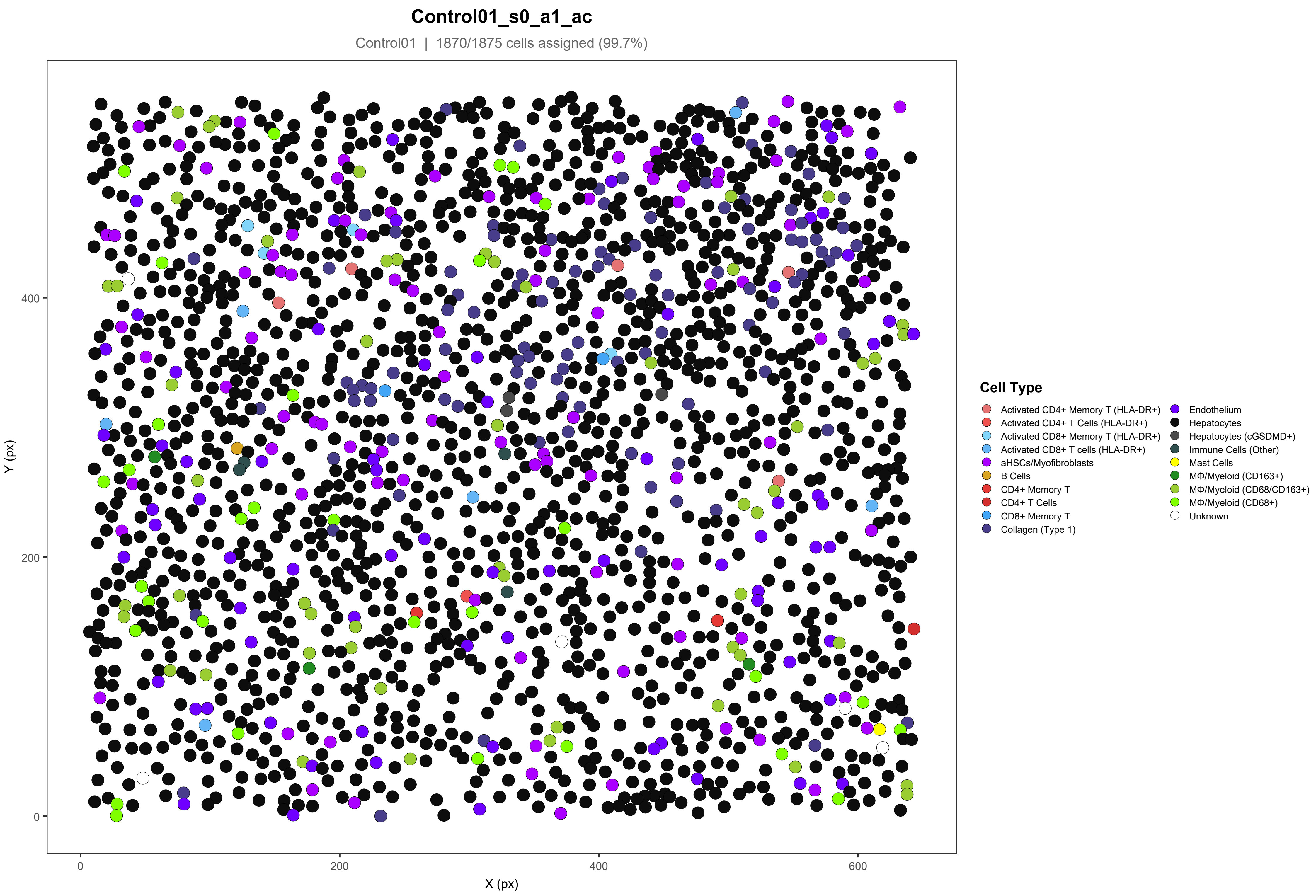

Control01_s0_a1_ac
Control01 | 1870/1875 cells assigned (99.7%)
Y (px)
X (px)
0
200
400
600
Cell Type
Activated CD4+ Memory T (HLA-DR+)
Activated CD4+ T Cells (HLA-DR+)
Activated CD8+ Memory T (HLA-DR+)
Activated CD8+ T cells (HLA-DR+)
aHSCs/Myofibroblasts
B Cells
CD4+ Memory T
CD4+ T Cells
CD8+ Memory T
Collagen (Type 1)
Endothelium
Hepatocytes
Hepatocytes (cGSDMD+)
Immune Cells (Other)
Mast Cells
MΦ/Myeloid (CD163+)
MΦ/Myeloid (CD68/CD163+)
MΦ/Myeloid (CD68+)
Unknown

#### Control01_s0_a2_ac

Control01 | 1450/1451 cells assigned (99.9%)

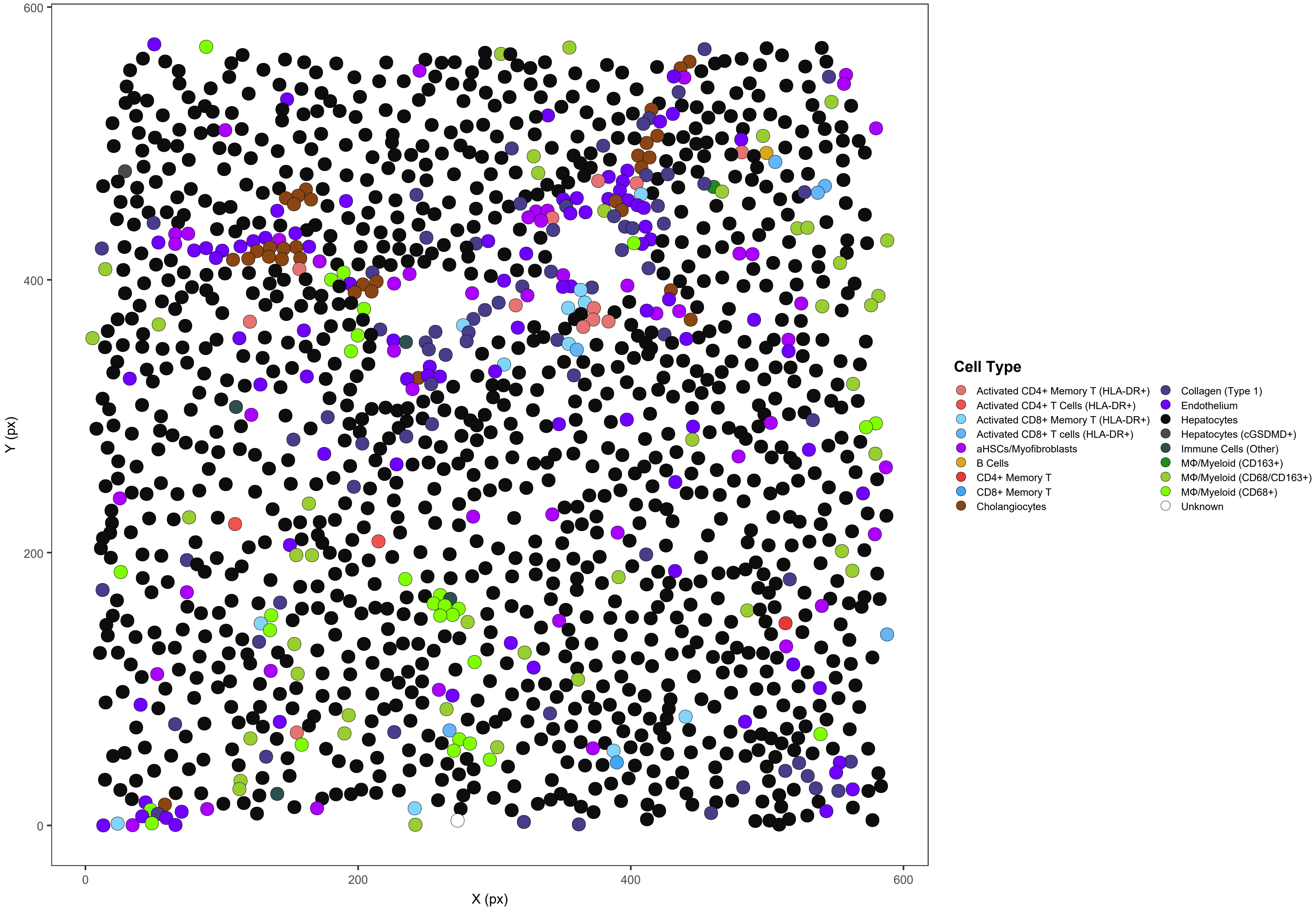

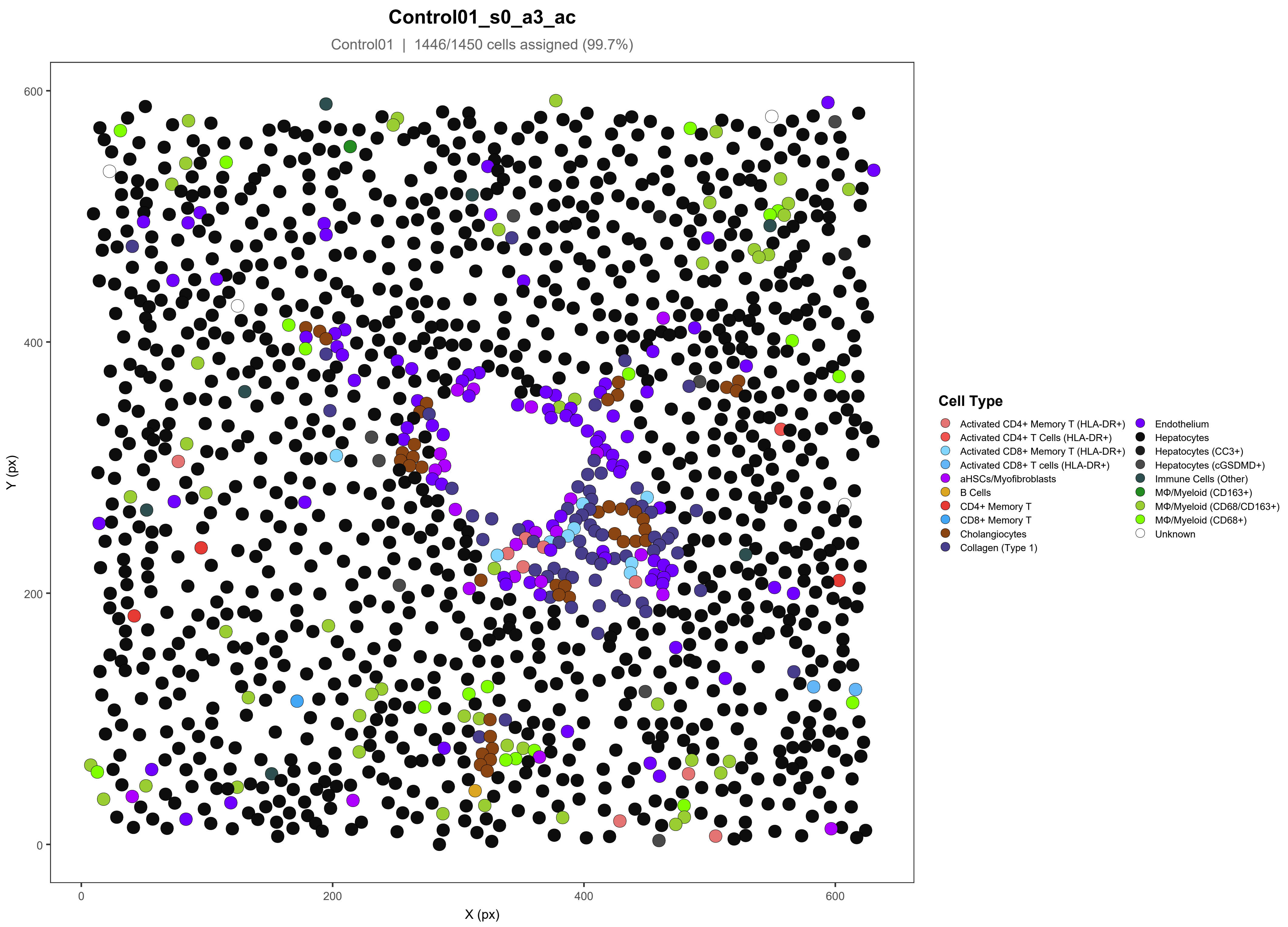
Control01_s0_a3_ac
Control01 | 1446/1450 cells assigned (99.7%)
Y (px)
X (px)
0
200
400
600
Cell Type
Activated CD4+ Memory T (HLA-DR+)
Activated CD4+ T Cells (HLA-DR+)
Activated CD8+ Memory T (HLA-DR+)
Activated CD8+ T cells (HLA-DR+)
aHSCs/Myofibroblasts
B Cells
CD4+ Memory T
CD8+ Memory T
Cholangiocytes
Collagen (Type 1)
Endothelium
Hepatocytes
Hepatocytes (CC3+)
Hepatocytes (cGSDMD+)
Immune Cells (Other)
MΦ/Myeloid (CD163+)
MΦ/Myeloid (CD68/CD163+)
MΦ/Myeloid (CD68+)
Unknown

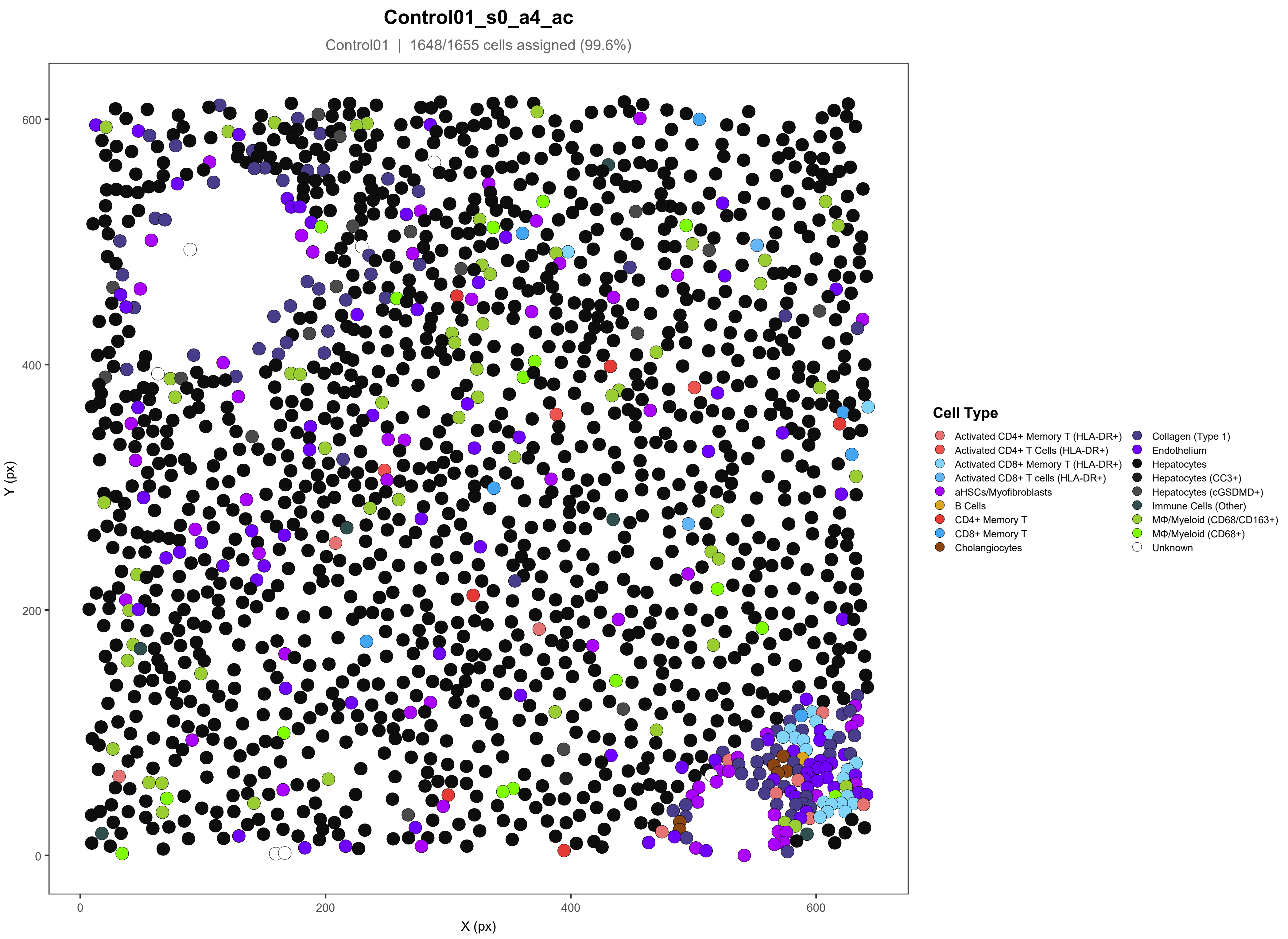

Control01_s0_a4_ac
Control01 | 1648/1655 cells assigned (99.6%)
Y (px)
X (px)
0
200
400
600
Cell Type
Activated CD4+ Memory T (HLA-DR+)
Activated CD4+ T Cells (HLA-DR+)
Activated CD8+ Memory T (HLA-DR+)
Activated CD8+ T cells (HLA-DR+)
aHSCs/Myofibroblasts
B Cells
CD4+ Memory T
CD8+ Memory T
Cholangiocytes
Collagen (Type 1)
Endothelium
Hepatocytes
Hepatocytes (CC3+)
Hepatocytes (cGSDMD+)
Immune Cells (Other)
MΦ/Myeloid (CD68/CD163+)
MΦ/Myeloid (CD68+)
Unknown

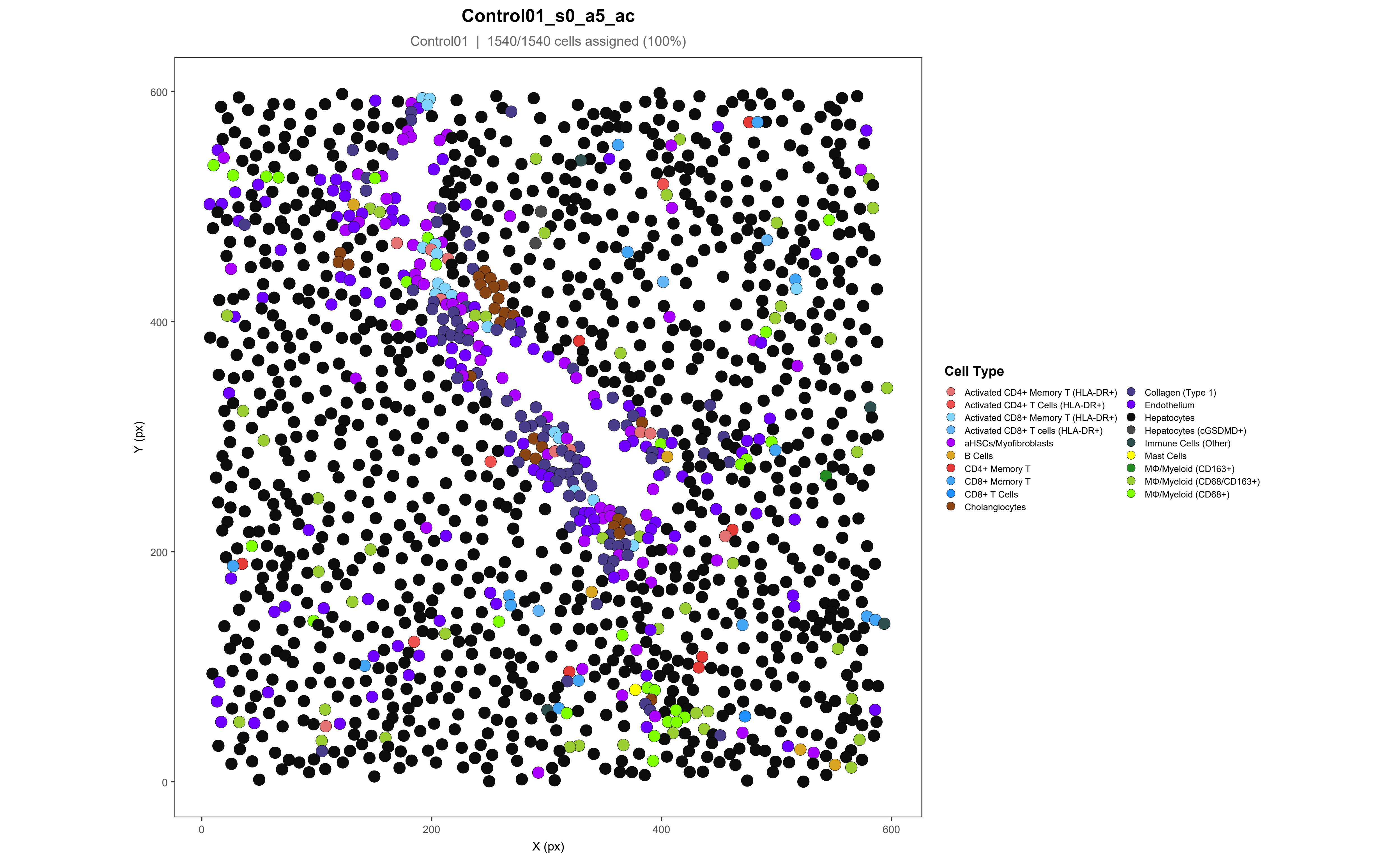
Control01_s0_a5_ac
Control01 | 1540/1540 cells assigned (100%)
Y (px)
X (px)
0
200
400
600
Cell Type
Activated CD4+ Memory T (HLA-DR+)
Activated CD4+ T Cells (HLA-DR+)
Activated CD8+ Memory T (HLA-DR+)
Activated CD8+ T cells (HLA-DR+)
aHSCs/Myofibroblasts
B Cells
CD4+ Memory T
CD8+ Memory T
CD8+ T Cells
Cholangiocytes
Collagen (Type 1)
Endothelium
Hepatocytes
Hepatocytes (cGSDMD+)
Immune Cells (Other)
Mast Cells
MΦ/Myeloid (CD163+)
MΦ/Myeloid (CD68/CD163+)
MΦ/Myeloid (CD68+)

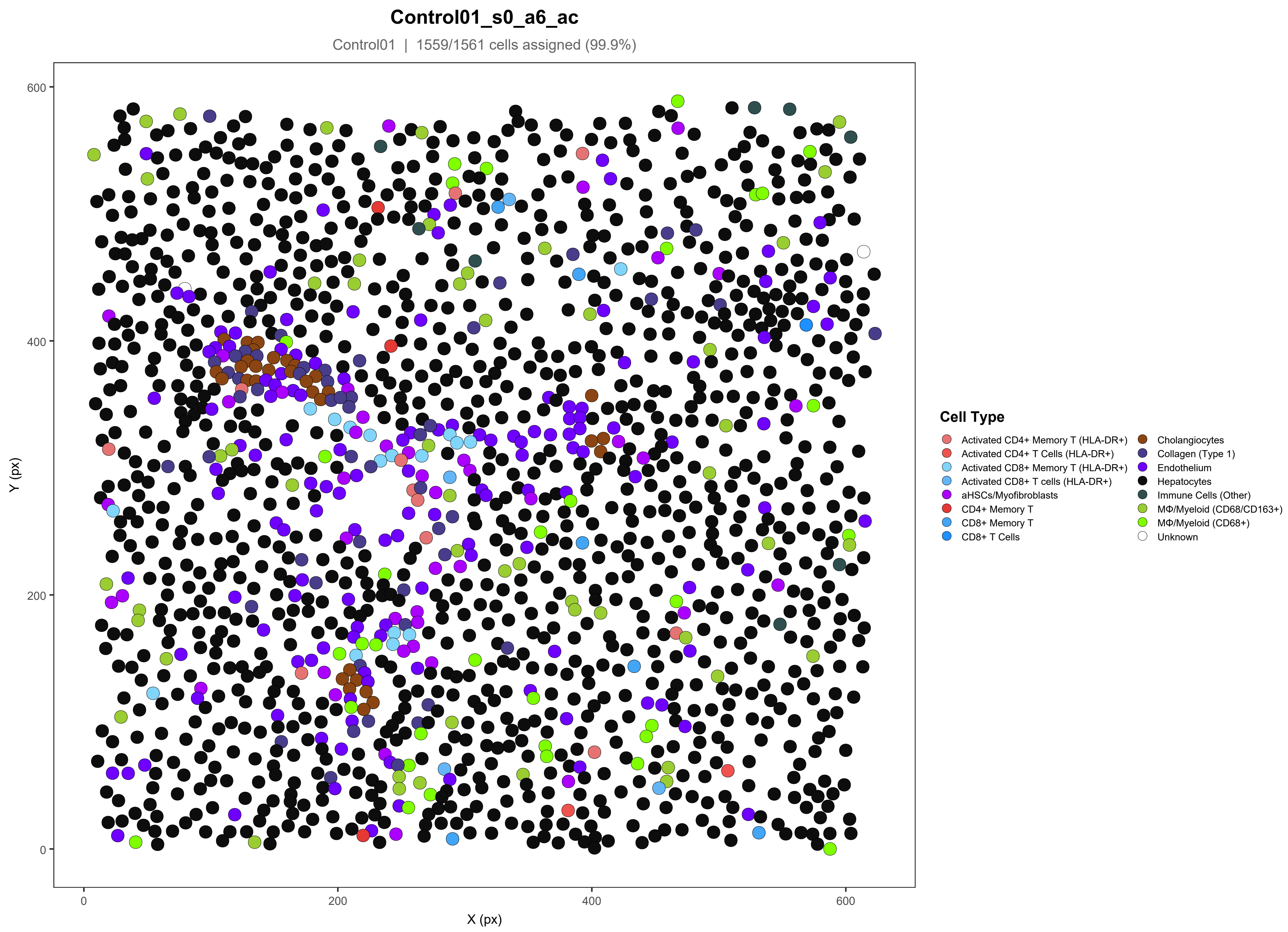
Control01_s0_a6_ac
Control01 | 1559/1561 cells assigned (99.9%)
Y (px)
X (px)
0
200
400
600
Cell Type
Activated CD4+ Memory T (HLA-DR+)
Activated CD4+ T Cells (HLA-DR+)
Activated CD8+ Memory T (HLA-DR+)
Activated CD8+ T cells (HLA-DR+)
aHSCs/Myofibroblasts
CD4+ Memory T
CD8+ Memory T
CD8+ T Cells
Cholangiocytes
Collagen (Type 1)
Endothelium
Hepatocytes
Immune Cells (Other)
MΦ/Myeloid (CD68/CD163+)
MΦ/Myeloid (CD68+)
Unknown

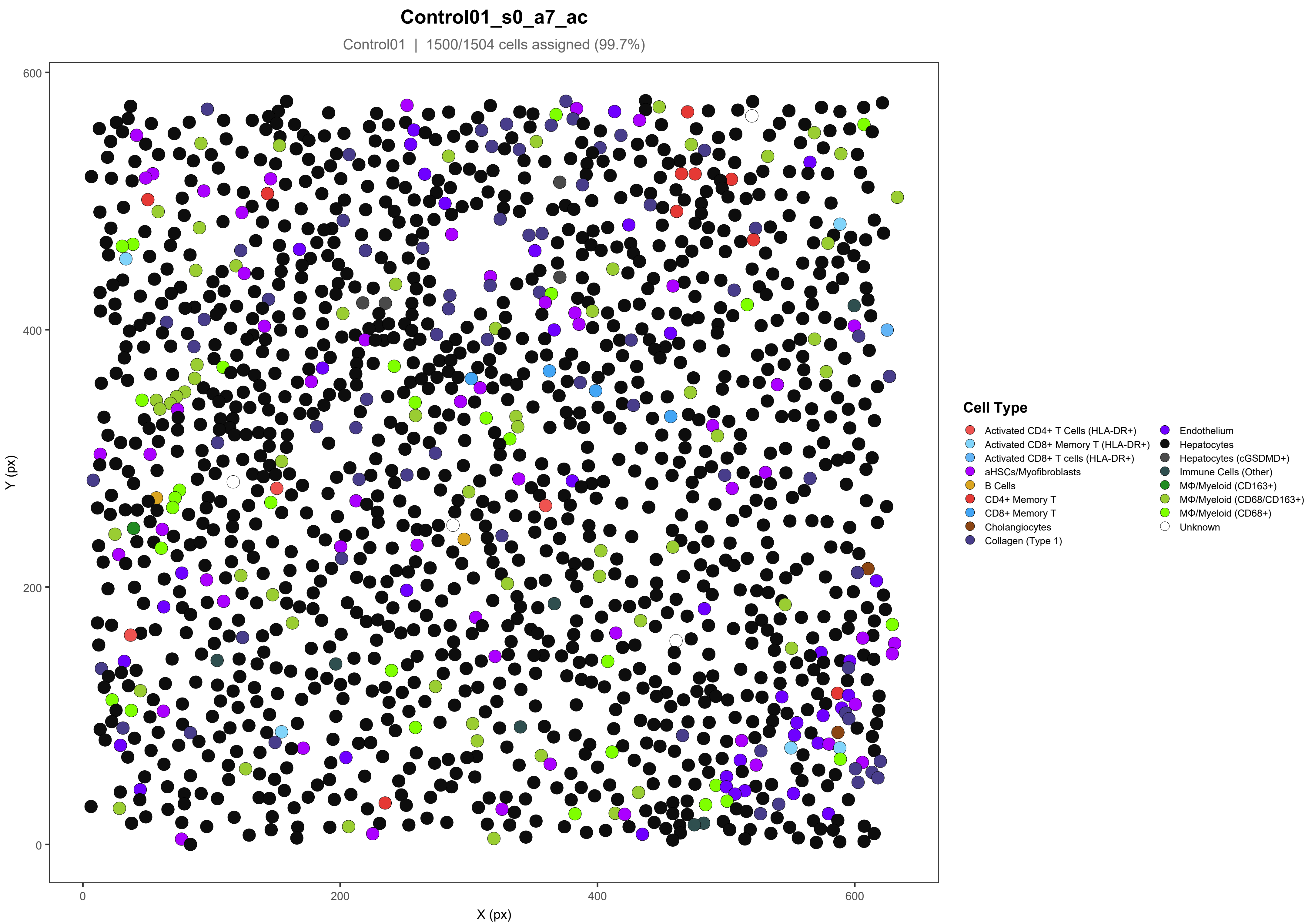
Control01_s0_a7_ac
Control01 | 1500/1504 cells assigned (99.7%)
Y (px)
X (px)
0
200
400
600
Cell Type
Activated CD4+ T Cells (HLA-DR+)
Activated CD8+ Memory T (HLA-DR+)
Activated CD8+ T cells (HLA-DR+)
aHSCs/Myofibroblasts
B Cells
CD4+ Memory T
CD8+ Memory T
Cholangiocytes
Collagen (Type 1)
Endothelium
Hepatocytes
Hepatocytes (cGSDMD+)
Immune Cells (Other)
MΦ/Myeloid (CD163+)
MΦ/Myeloid (CD68/CD163+)
MΦ/Myeloid (CD68+)
Unknown

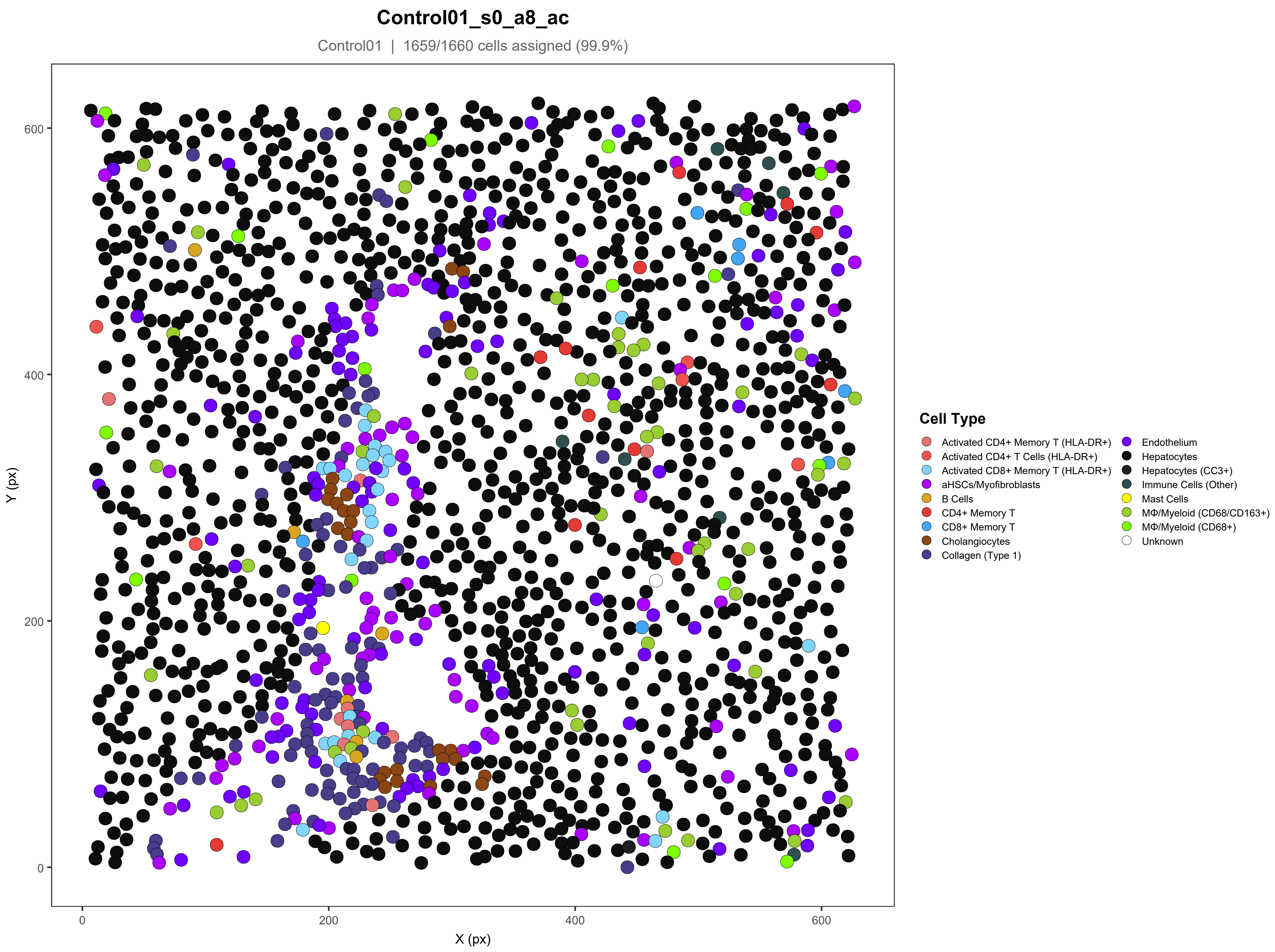
Control01_s0_a8_ac
Control01 | 1659/1660 cells assigned (99.9%)
Y (px)
X (px)
0
200
400
600
Cell Type
Activated CD4+ Memory T (HLA-DR+)
Activated CD4+ T Cells (HLA-DR+)
Activated CD8+ Memory T (HLA-DR+)
aHSCs/Myofibroblasts
B Cells
CD4+ Memory T
CD8+ Memory T
Cholangiocytes
Collagen (Type 1)
Endothelium
Hepatocytes
Hepatocytes (CC3+)
Immune Cells (Other)
Mast Cells
MΦ/Myeloid (CD68/CD163+)
MΦ/Myeloid (CD68+)
Unknown

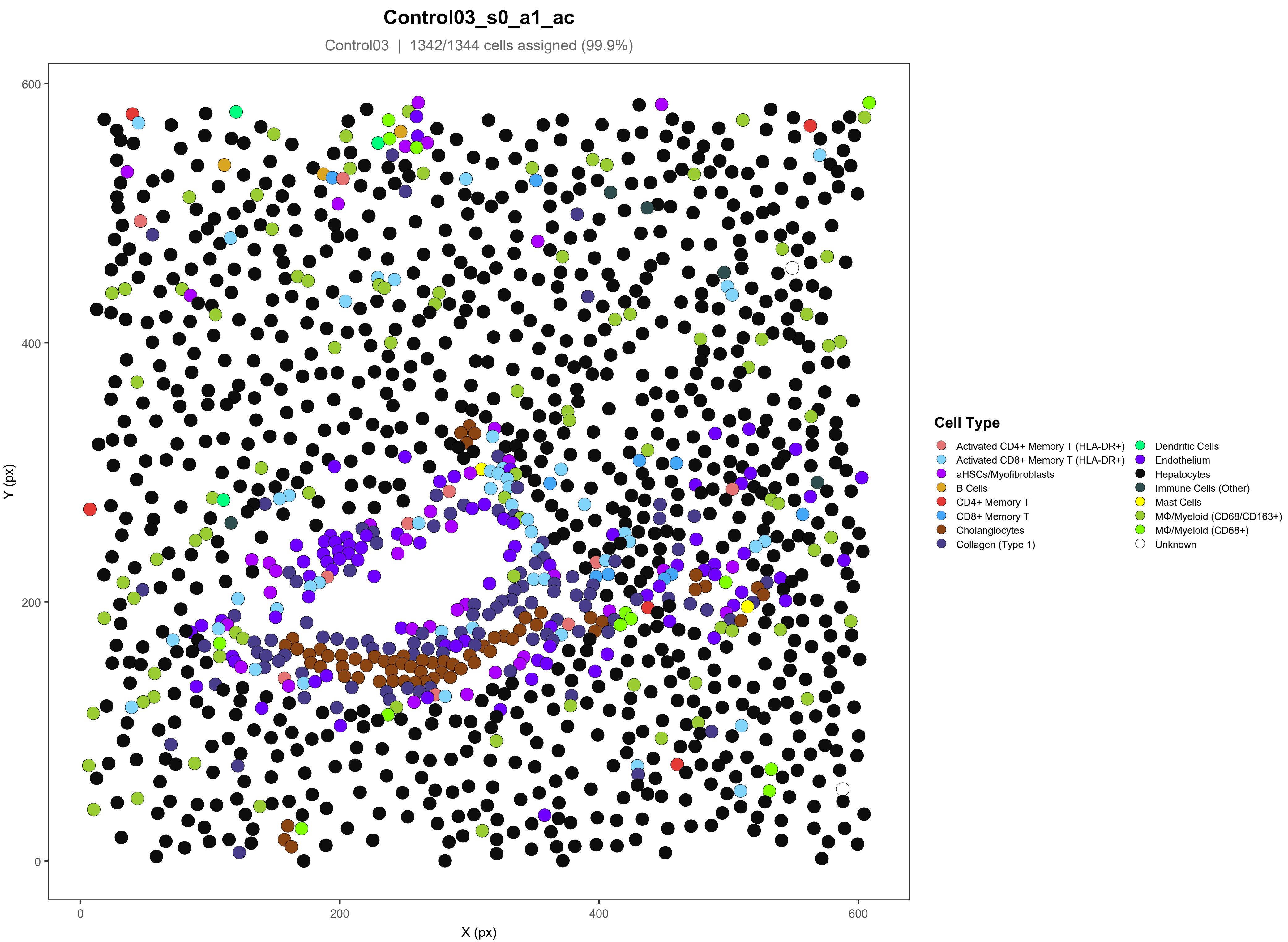

Control03_s0_a1_ac
Control03 | 1342/1344 cells assigned (99.9%)
Y (px)
X (px)
0
200
400
600
Cell Type
Activated CD4+ Memory T (HLA-DR+)
Activated CD8+ Memory T (HLA-DR+)
aHSCs/Myofibroblasts
B Cells
CD4+ Memory T
CD8+ Memory T
Cholangiocytes
Collagen (Type 1)
Dendritic Cells
Endothelium
Hepatocytes
Immune Cells (Other)
Mast Cells
MΦ/Myeloid (CD68/CD163+)
MΦ/Myeloid (CD68+)
Unknown

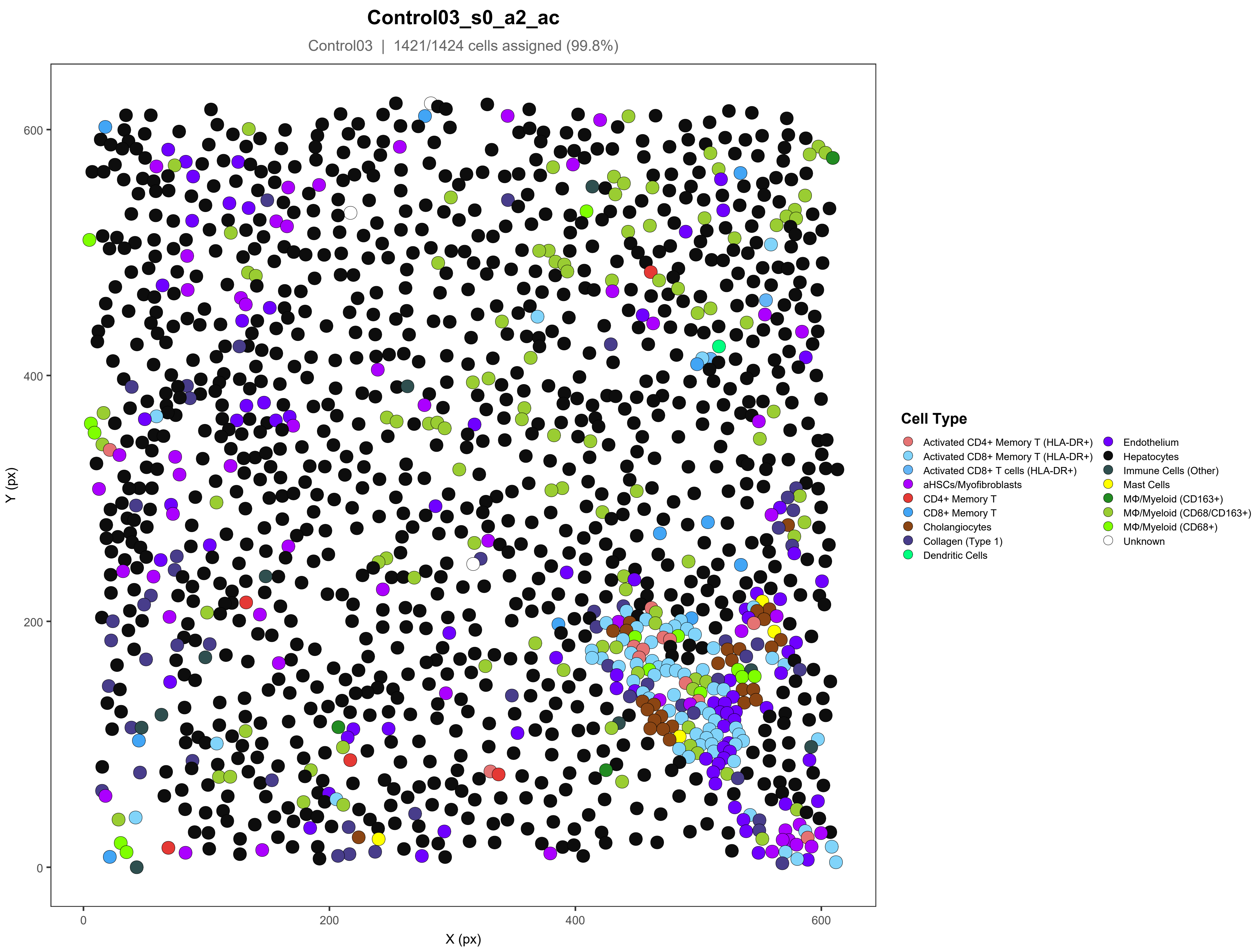
Control03_s0_a2_ac
Control03 | 1421/1424 cells assigned (99.8%)
Y (px)
X (px)
0
200
400
600
Cell Type
Activated CD4+ Memory T (HLA-DR+)
Activated CD8+ Memory T (HLA-DR+)
Activated CD8+ T cells (HLA-DR+)
aHSCs/Myofibroblasts
CD4+ Memory T
CD8+ Memory T
Cholangiocytes
Collagen (Type 1)
Dendritic Cells
Endothelium
Hepatocytes
Immune Cells (Other)
Mast Cells
MΦ/Myeloid (CD163+)
MΦ/Myeloid (CD68/CD163+)
MΦ/Myeloid (CD68+)
Unknown

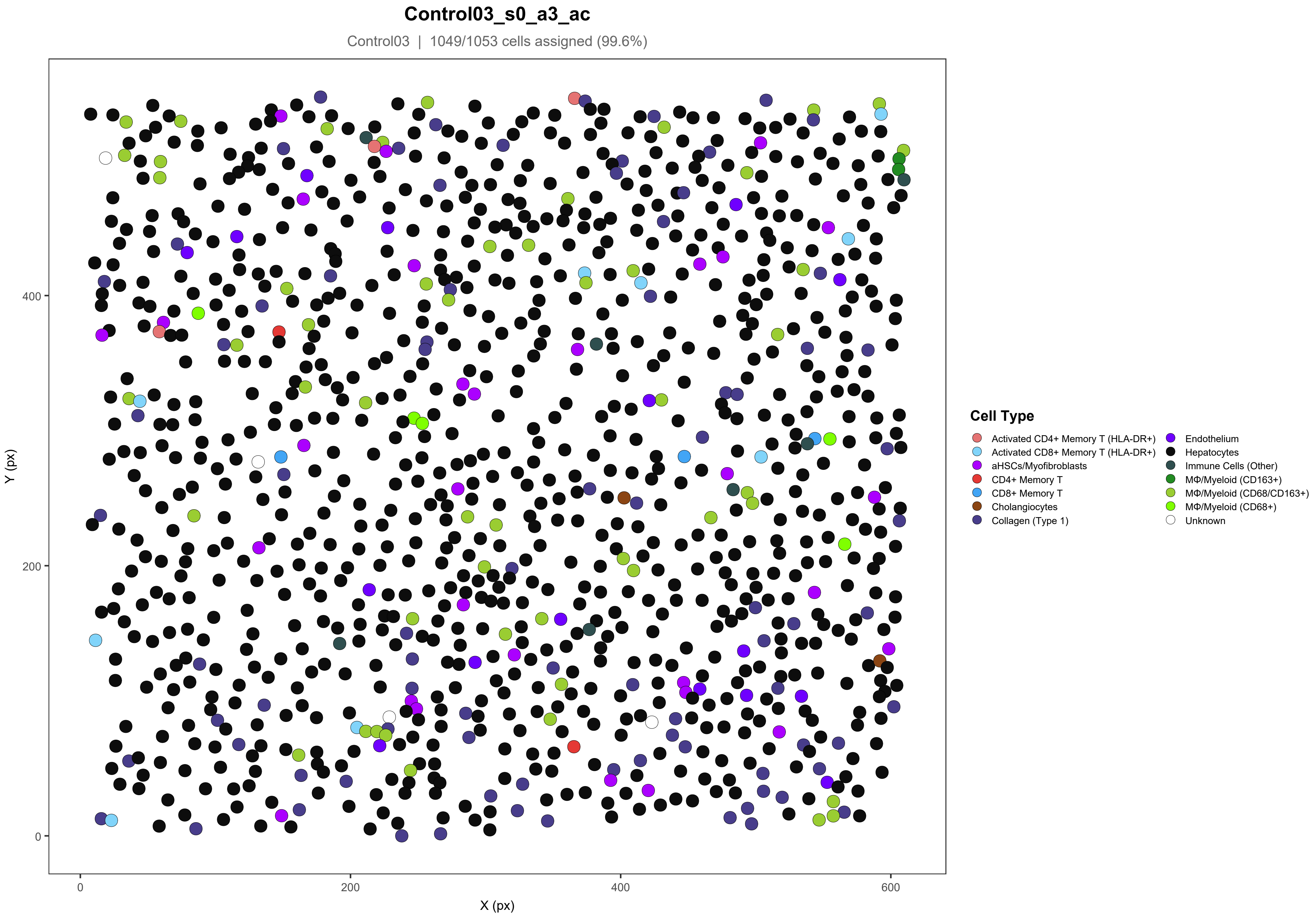

Control03_s0_a3_ac
Control03 | 1049/1053 cells assigned (99.6%)
Y (px)
X (px)
0
200
400
600
Cell Type
Activated CD4+ Memory T (HLA-DR+)
Activated CD8+ Memory T (HLA-DR+)
aHSCs/Myofibroblasts
CD4+ Memory T
CD8+ Memory T
Cholangiocytes
Collagen (Type 1)
Endothelium
Hepatocytes
Immune Cells (Other)
MΦ/Myeloid (CD163+)
MΦ/Myeloid (CD68/CD163+)
MΦ/Myeloid (CD68+)
Unknown

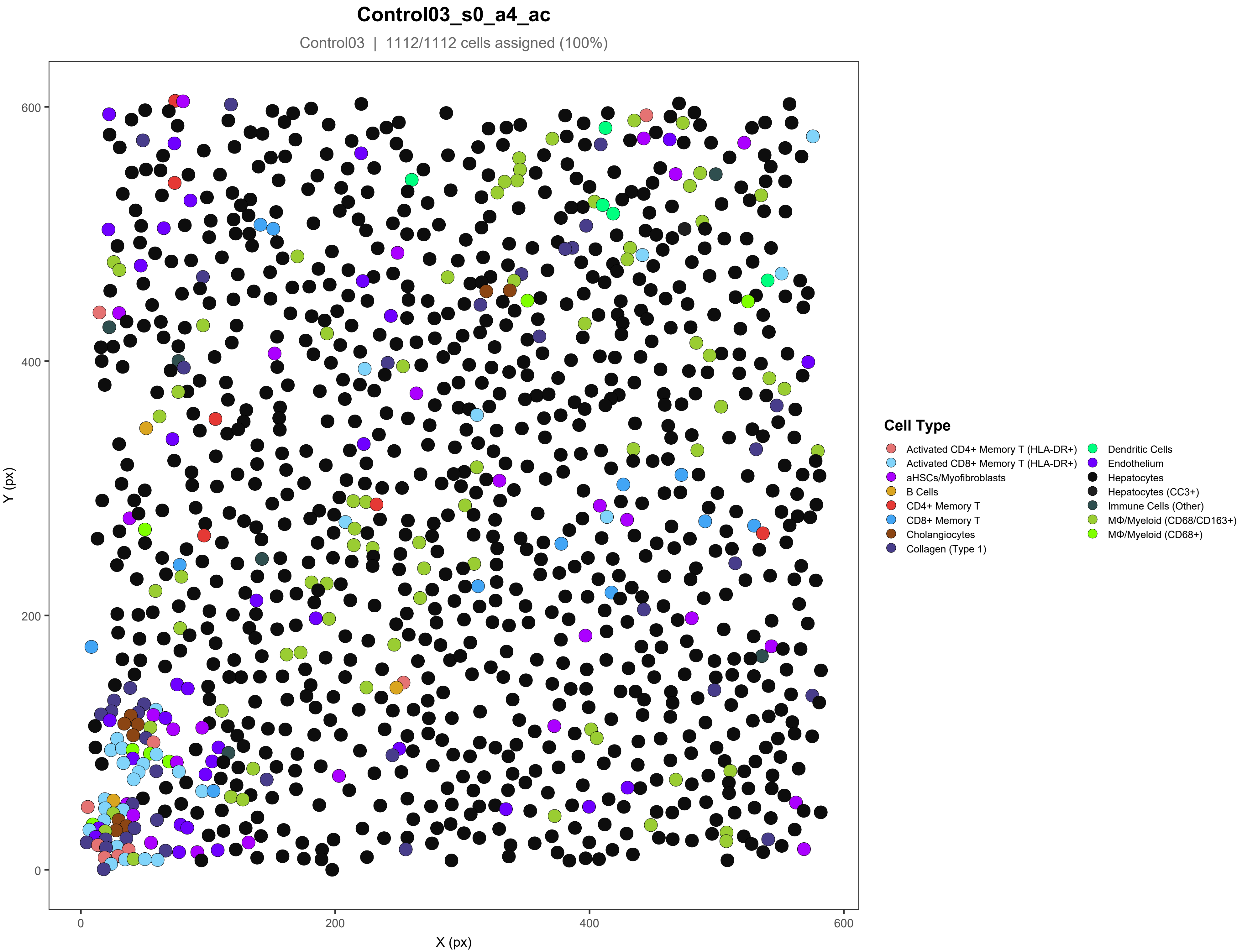
Control03_s0_a4_ac
Control03 | 1112/1112 cells assigned (100%)
Y (px)
X (px)
0
200
400
600
Cell Type
Activated CD4+ Memory T (HLA-DR+)
Activated CD8+ Memory T (HLA-DR+)
aHSCs/Myofibroblasts
B Cells
CD4+ Memory T
CD8+ Memory T
Cholangiocytes
Collagen (Type 1)
Dendritic Cells
Endothelium
Hepatocytes
Hepatocytes (CC3+)
Immune Cells (Other)
MΦ/Myeloid (CD68/CD163+)
MΦ/Myeloid (CD68+)

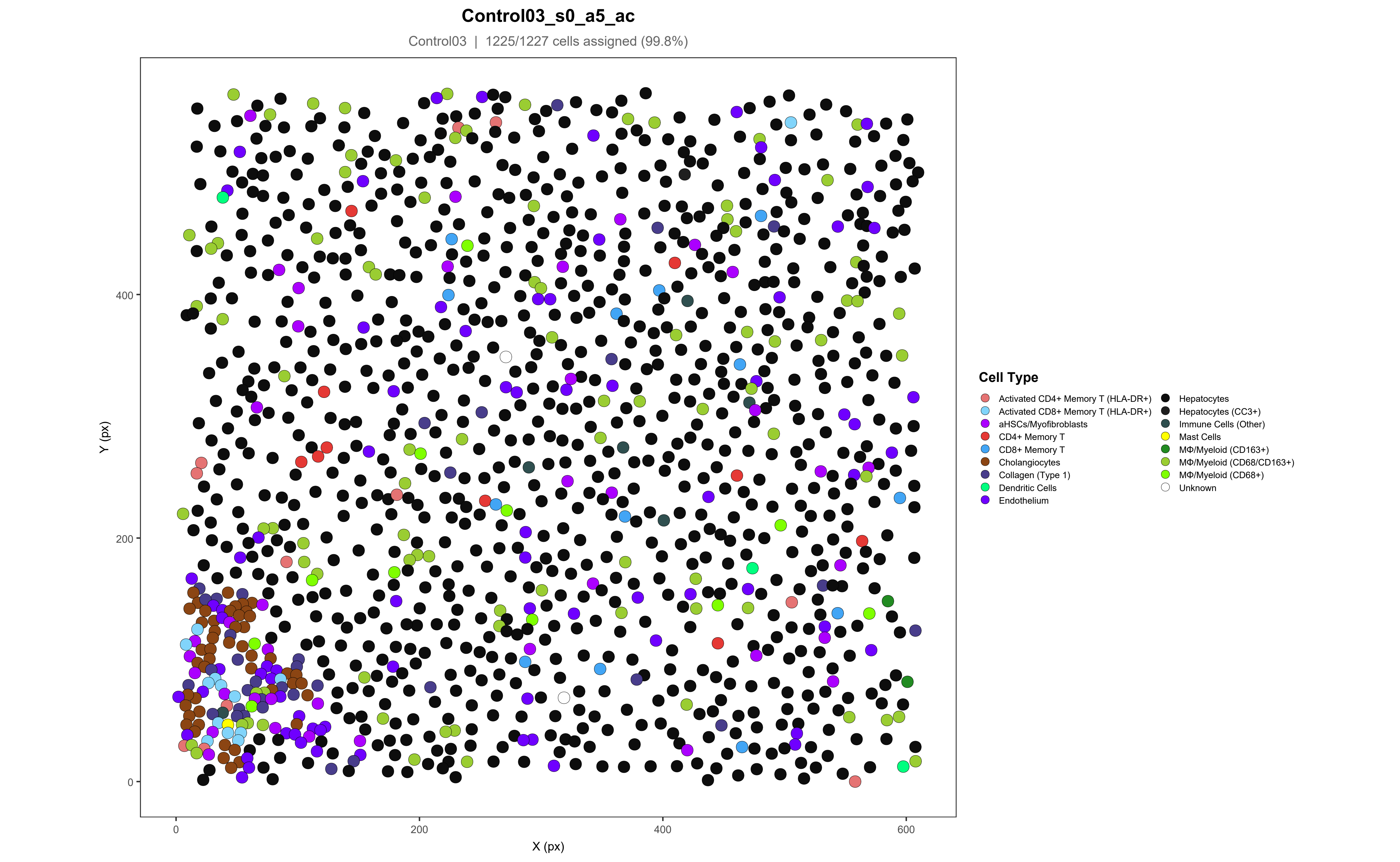
Control03_s0_a5_ac
Control03 | 1225/1227 cells assigned (99.8%)
Y (px)
X (px)
0
200
400
600
Cell Type
Activated CD4+ Memory T (HLA-DR+)
Activated CD8+ Memory T (HLA-DR+)
aHSCs/Myofibroblasts
CD4+ Memory T
CD8+ Memory T
Cholangiocytes
Collagen (Type 1)
Dendritic Cells
Endothelium
Hepatocytes
Hepatocytes (CC3+)
Immune Cells (Other)
Mast Cells
MΦ/Myeloid (CD163+)
MΦ/Myeloid (CD68/CD163+)
MΦ/Myeloid (CD68+)
Unknown

### Control03_s0_a6_ac

Control03 | 858/860 cells assigned (99.8%)

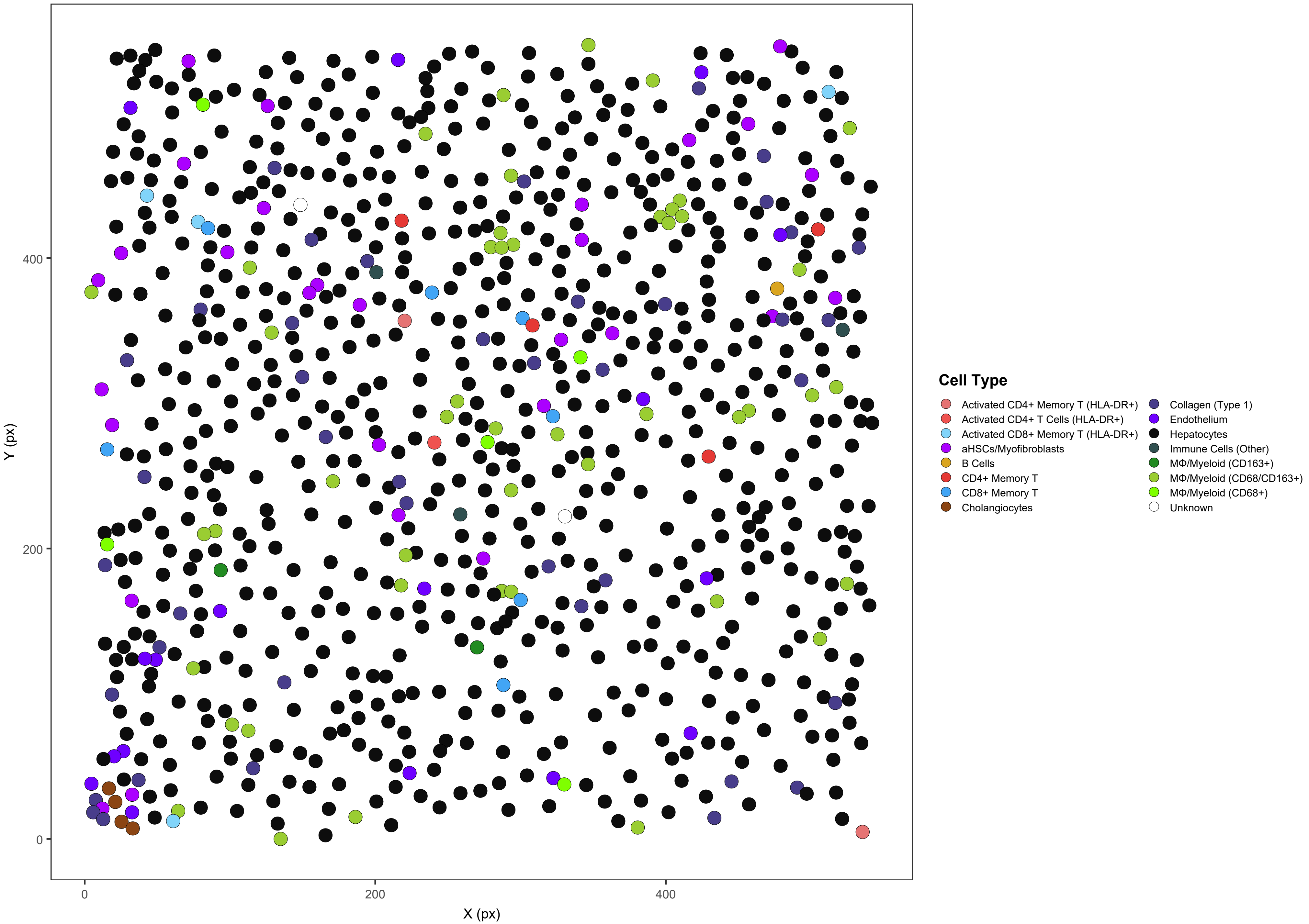

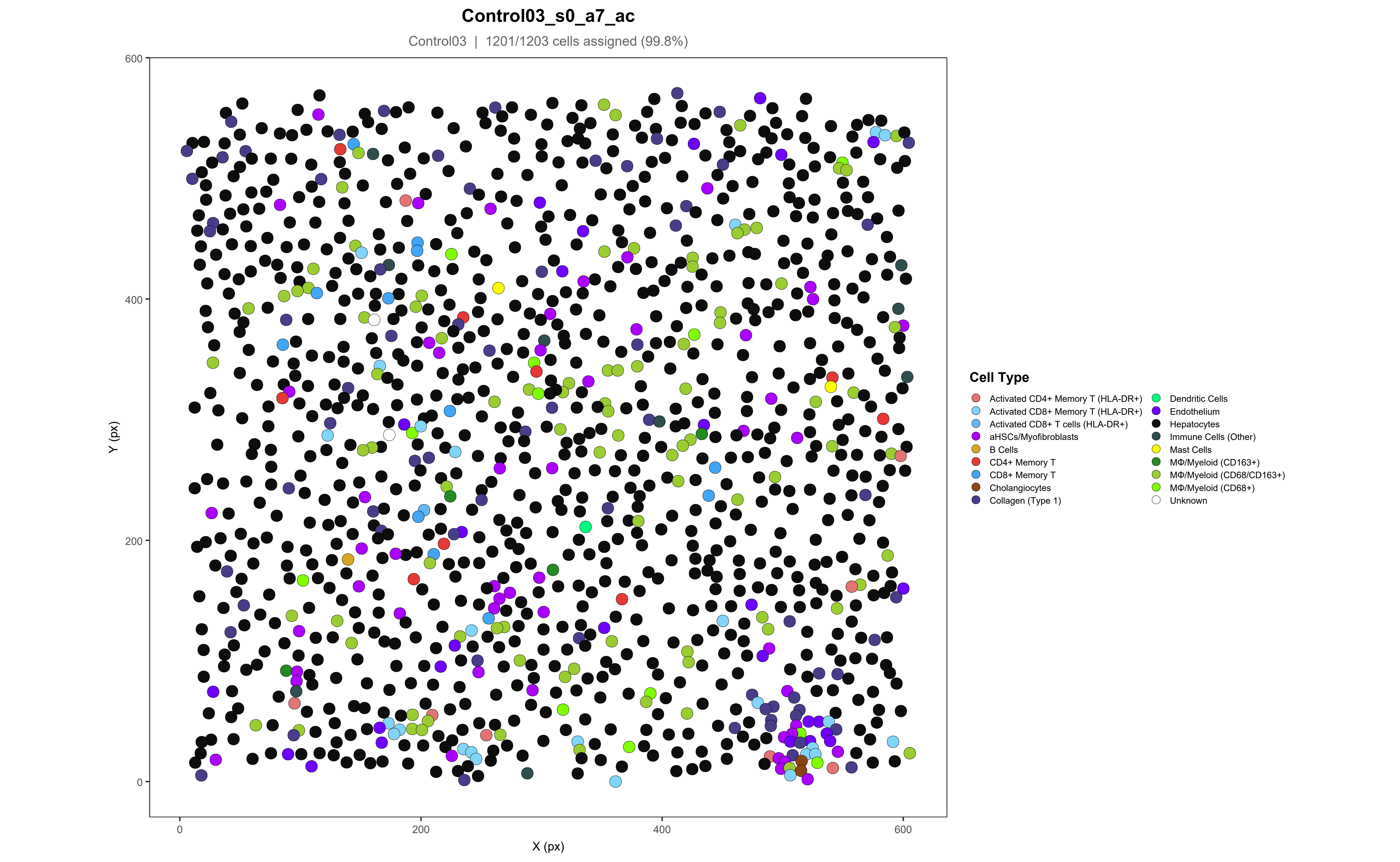

Control03_s0_a7_ac
Control03 | 1201/1203 cells assigned (99.8%)
Y (px)
X (px)
0
200
400
600
Cell Type
Activated CD4+ Memory T (HLA-DR+)
Activated CD8+ Memory T (HLA-DR+)
Activated CD8+ T cells (HLA-DR+)
aHSCs/Myofibroblasts
B Cells
CD4+ Memory T
CD8+ Memory T
Cholangiocytes
Collagen (Type 1)
Dendritic Cells
Endothelium
Hepatocytes
Immune Cells (Other)
Mast Cells
MΦ/Myeloid (CD163+)
MΦ/Myeloid (CD68/CD163+)
MΦ/Myeloid (CD68+)
Unknown

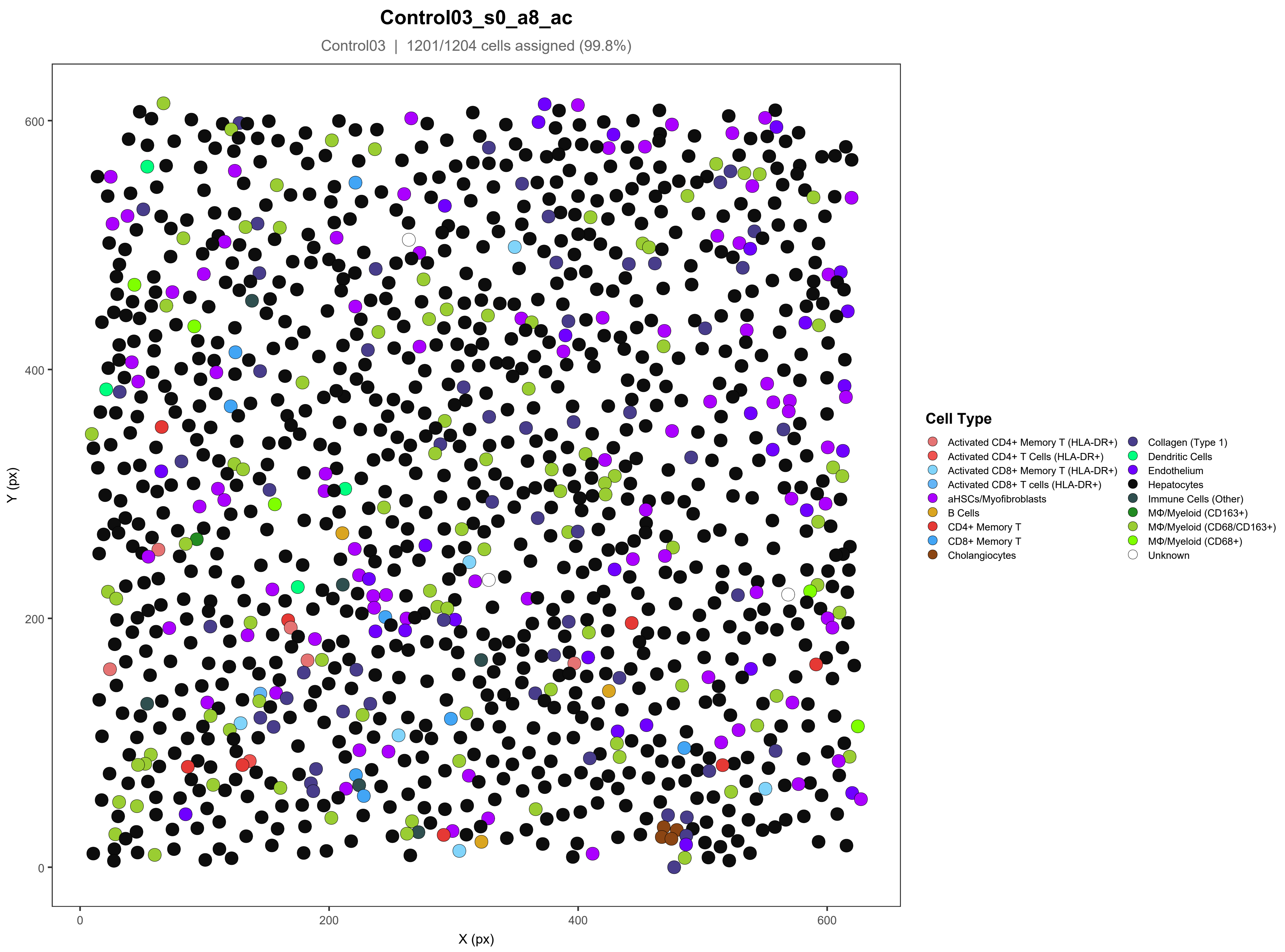

Control03_s0_a8_ac
Control03 | 1201/1204 cells assigned (99.8%)
Y (px)
X (px)
0
200
400
600
Cell Type
Activated CD4+ Memory T (HLA-DR+)
Activated CD4+ T Cells (HLA-DR+)
Activated CD8+ Memory T (HLA-DR+)
Activated CD8+ T cells (HLA-DR+)
aHSCs/Myofibroblasts
B Cells
CD4+ Memory T
CD8+ Memory T
Cholangiocytes
Collagen (Type 1)
Dendritic Cells
Endothelium
Hepatocytes
Immune Cells (Other)
MΦ/Myeloid (CD163+)
MΦ/Myeloid (CD68/CD163+)
MΦ/Myeloid (CD68+)
Unknown

#### ICI01_s0_a1_ac

ICI01 | 1348/1421 cells assigned (94.9%)

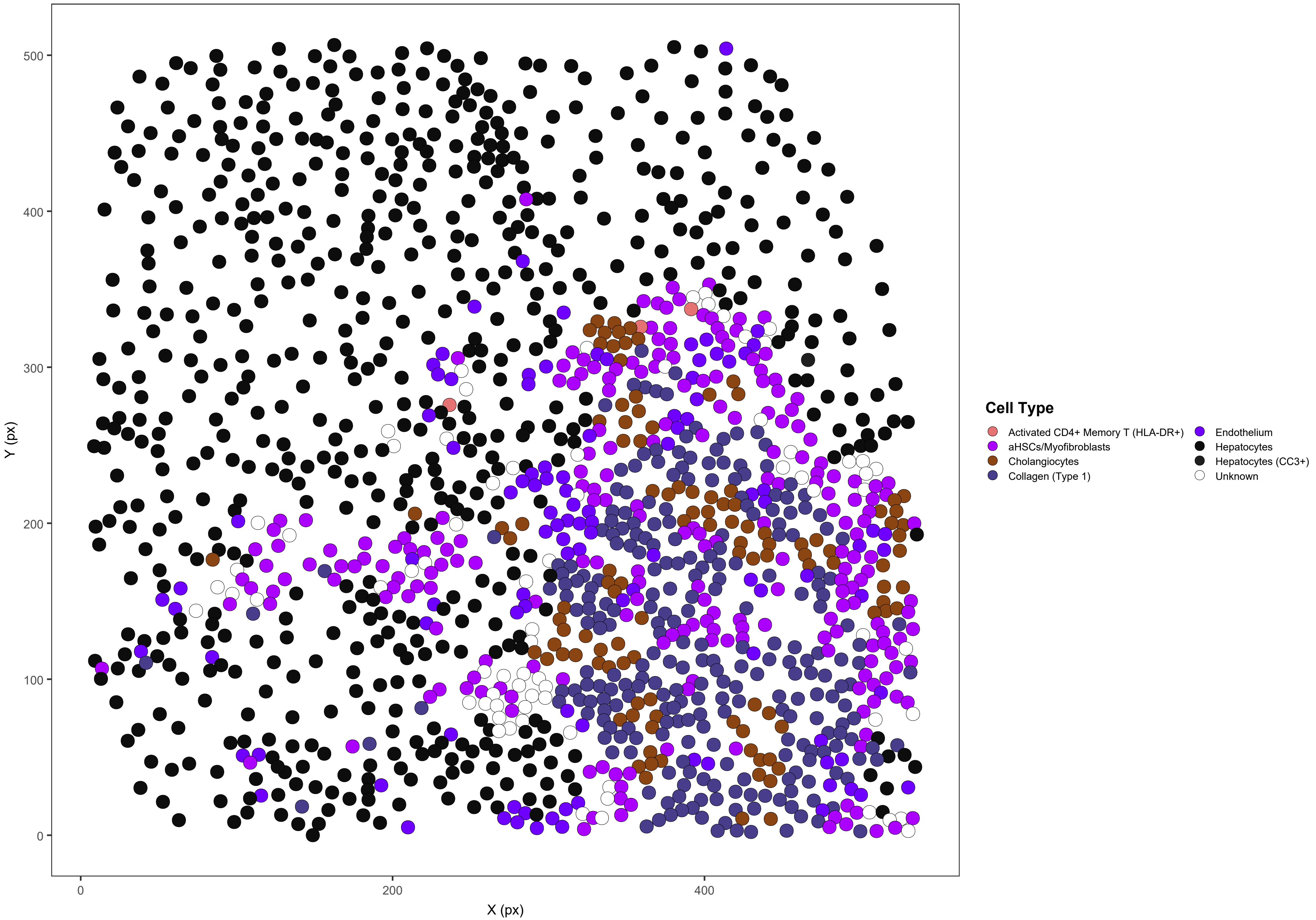

### ICI01_s0_a2_ac

ICI01 | 2660/2663 cells assigned (99.9%)

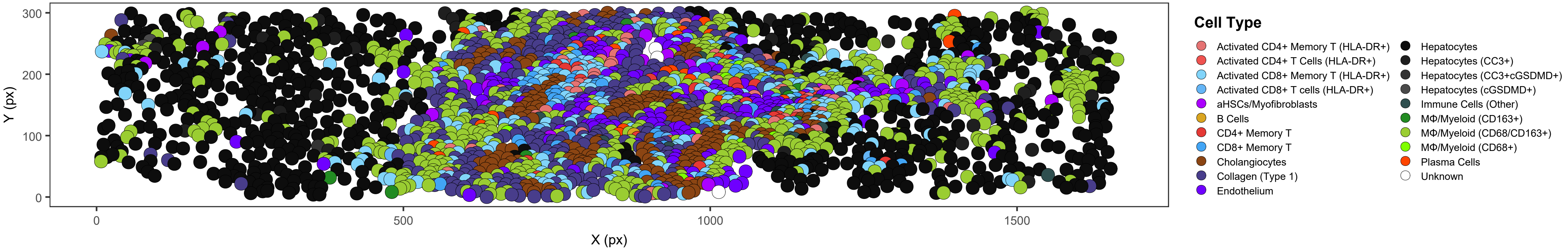

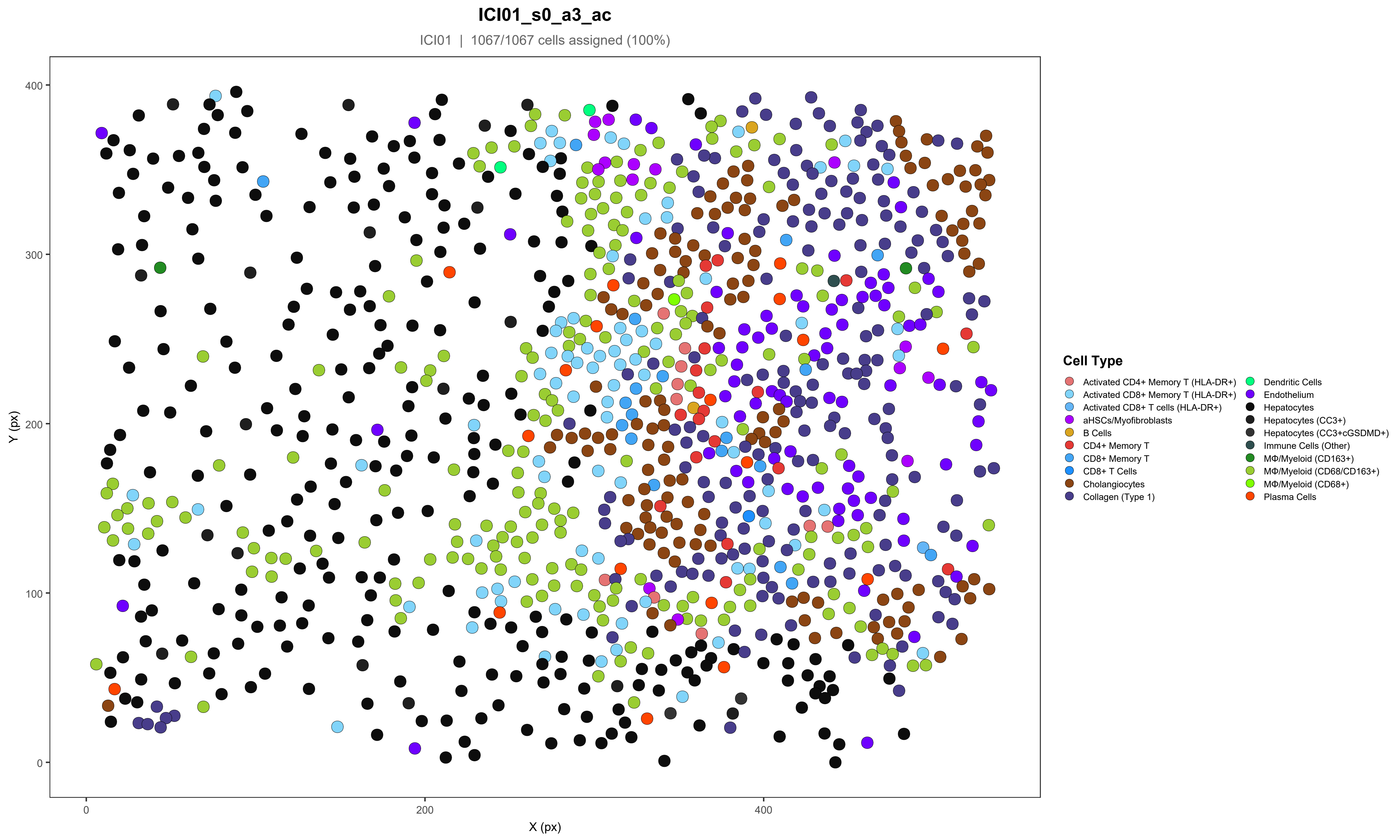

ICI01_s0_a3_ac
ICI01 | 1067/1067 cells assigned (100%)
Y (px)
X (px)
Cell Type
Activated CD4+ Memory T (HLA-DR+)
Activated CD8+ Memory T (HLA-DR+)
Activated CD8+ T cells (HLA-DR+)
aHSCs/Myofibroblasts
B Cells
CD4+ Memory T
CD8+ Memory T
CD8+ T Cells
Cholangiocytes
Collagen (Type 1)
Dendritic Cells
Endothelium
Hepatocytes
Hepatocytes (CC3+)
Hepatocytes (CC3+cGSDMD+)
Immune Cells (Other)
MΦ/Myeloid (CD163+)
MΦ/Myeloid (CD68/CD163+)
MΦ/Myeloid (CD68+)
Plasma Cells

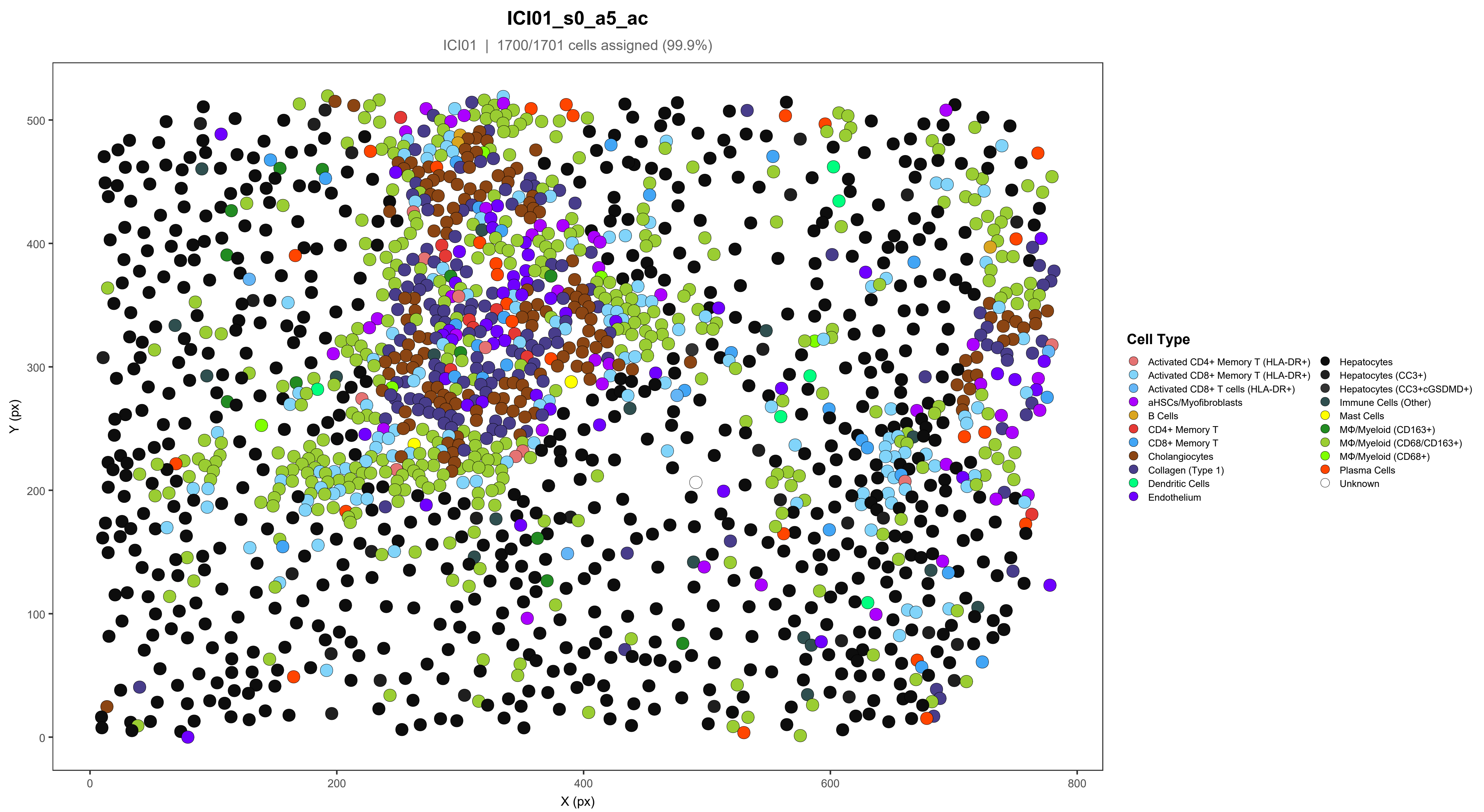

ICI01_s0_a5_ac
ICI01 | 1700/1701 cells assigned (99.9%)
Y (px)
X (px)
0
100
200
300
400
500
0
200
400
600
800
Cell Type
Activated CD4+ Memory T (HLA-DR+)
Activated CD8+ Memory T (HLA-DR+)
Activated CD8+ T cells (HLA-DR+)
aHSCs/Myofibroblasts
B Cells
CD4+ Memory T
CD8+ Memory T
Cholangiocytes
Collagen (Type 1)
Dendritic Cells
Endothelium
Hepatocytes
Hepatocytes (CC3+)
Hepatocytes (CC3+cGSDMD+)
Immune Cells (Other)
Mast Cells
MΦ/Myeloid (CD163+)
MΦ/Myeloid (CD68/CD163+)
MΦ/Myeloid (CD68+)
Plasma Cells
Unknown

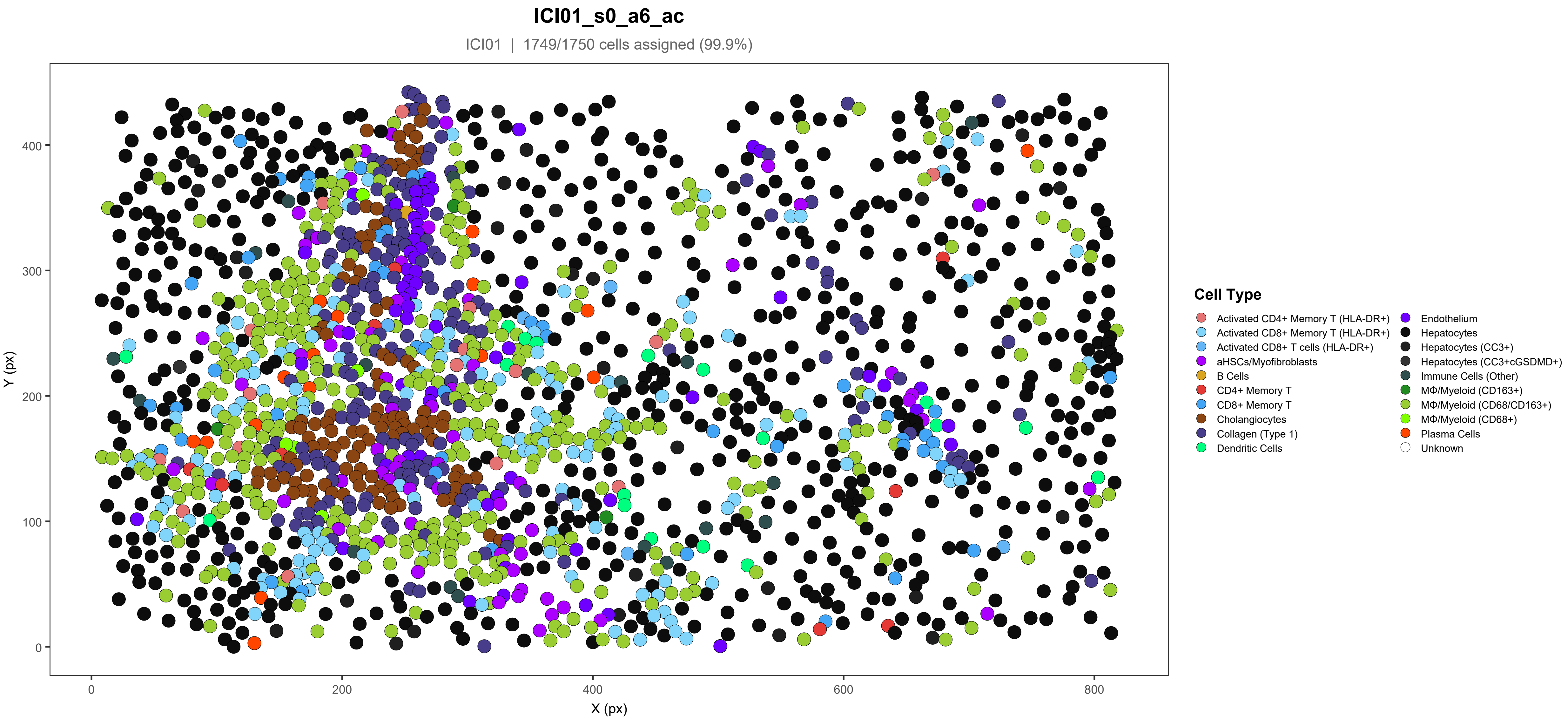

ICI01_s0_a6_ac
ICI01 | 1749/1750 cells assigned (99.9%)
Y (px)
X (px)
0
100
200
300
400
800
600
Cell Type
Activated CD4+ Memory T (HLA-DR+)
Activated CD8+ Memory T (HLA-DR+)
Activated CD8+ T cells (HLA-DR+)
aHSCs/Myofibroblasts
B Cells
CD4+ Memory T
CD8+ Memory T
Cholangiocytes
Collagen (Type 1)
Dendritic Cells
Endothelium
Hepatocytes
Hepatocytes (CC3+)
Hepatocytes (CC3+cGSDMD+)
Immune Cells (Other)
MΦ/Myeloid (CD163+)
MΦ/Myeloid (CD68/CD163+)
MΦ/Myeloid (CD68+)
Plasma Cells
Unknown

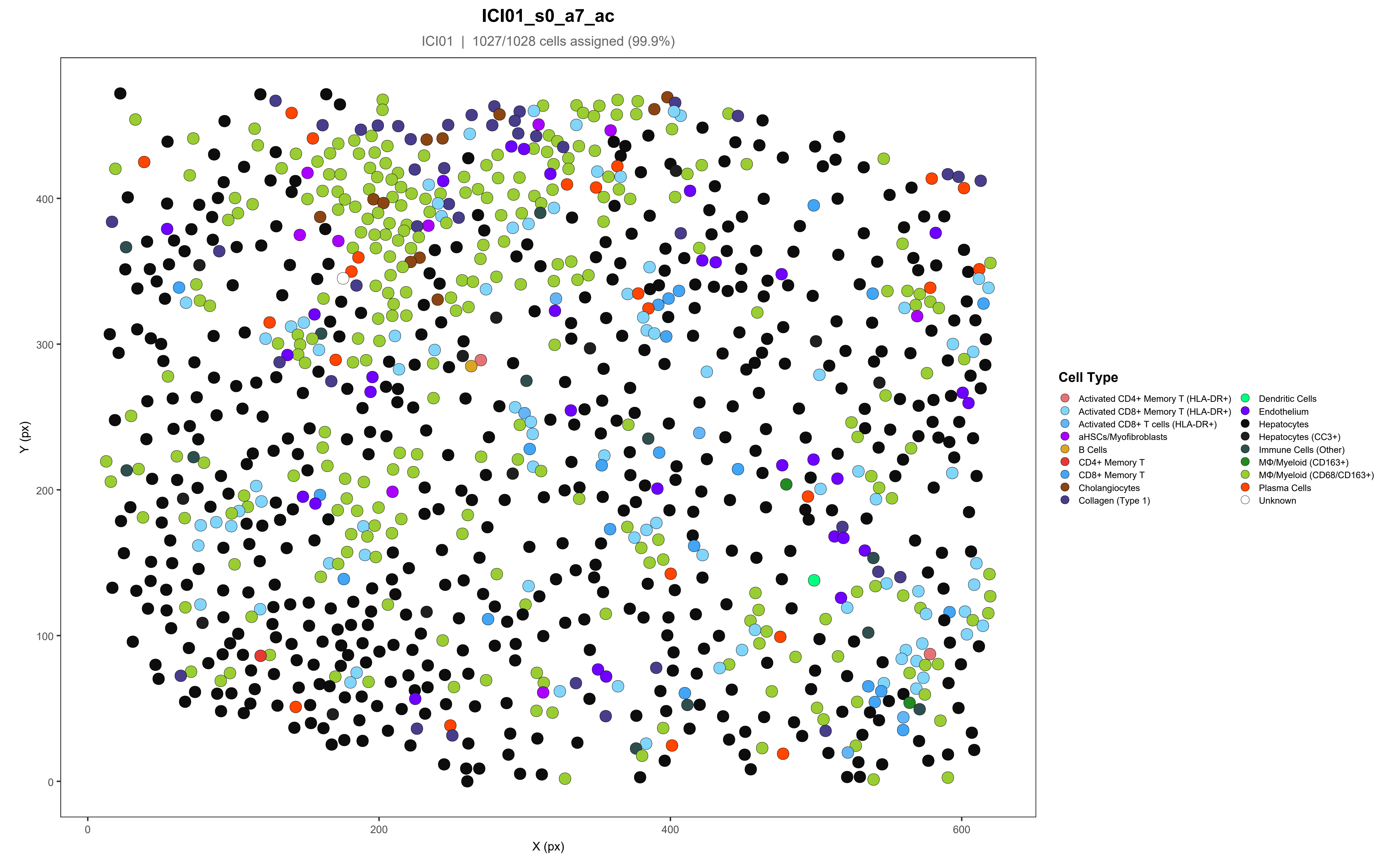
ICI01_s0_a7_ac
ICI01 | 1027/1028 cells assigned (99.9%)
Y (px)
X (px)
0
100
200
300
400
0
200
400
600
Cell Type
Activated CD4+ Memory T (HLA-DR+)
Activated CD8+ Memory T (HLA-DR+)
Activated CD8+ T cells (HLA-DR+)
aHSCs/Myofibroblasts
B Cells
CD4+ Memory T
CD8+ Memory T
Cholangiocytes
Collagen (Type 1)
Dendritic Cells
Endothelium
Hepatocytes
Hepatocytes (CC3+)
Immune Cells (Other)
MΦ/Myeloid (CD163+)
MΦ/Myeloid (CD68/CD163+)
Plasma Cells
Unknown

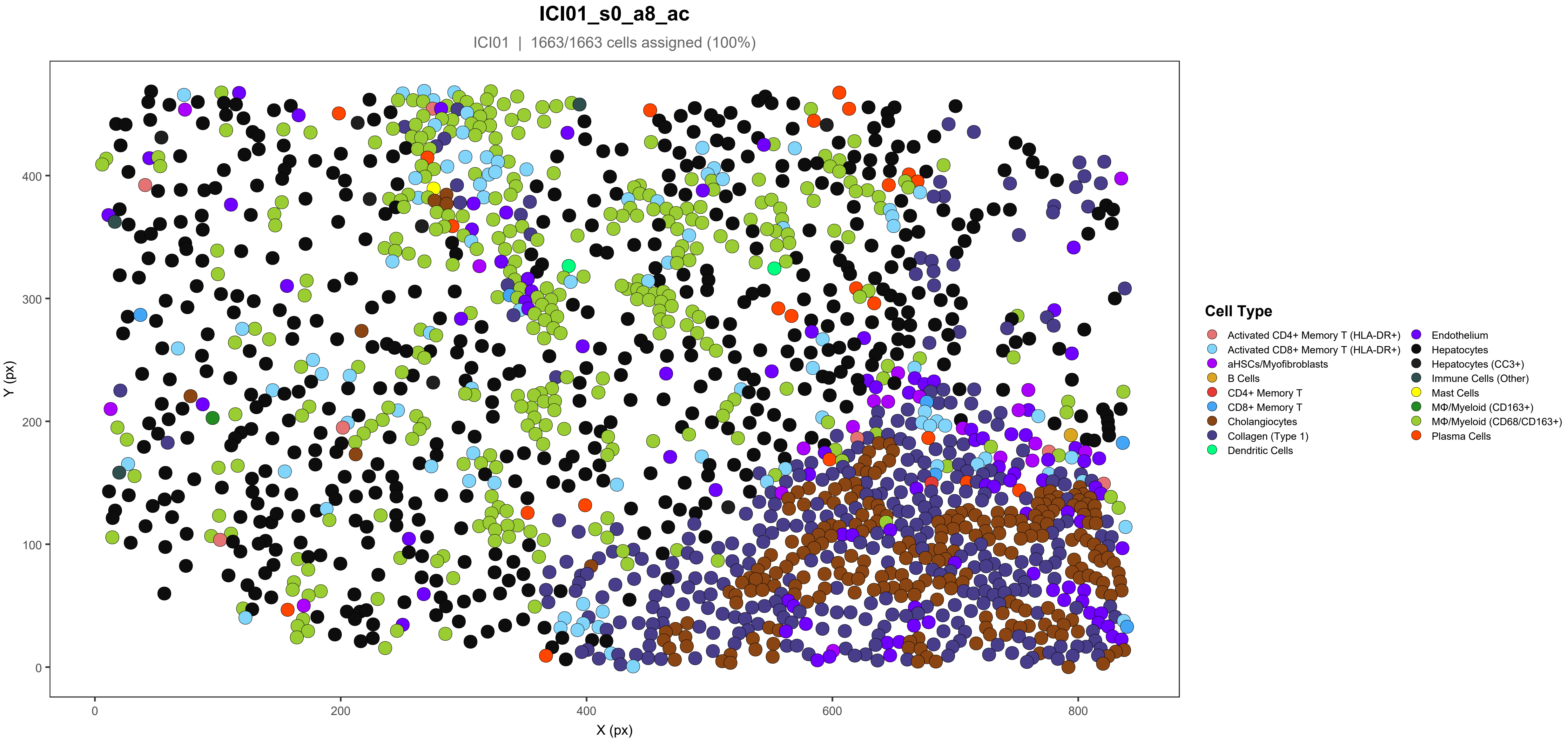
ICI01_s0_a8_ac
ICI01 | 1663/1663 cells assigned (100%)
Y (px)
X (px)
0
100
200
300
400
0
200
400
600
800
Cell Type
Activated CD4+ Memory T (HLA-DR+)
Activated CD8+ Memory T (HLA-DR+)
aHSCs/Myofibroblasts
B Cells
CD4+ Memory T
CD8+ Memory T
Cholangiocytes
Collagen (Type 1)
Dendritic Cells
Endothelium
Hepatocytes
Hepatocytes (CC3+)
Immune Cells (Other)
Mast Cells
MΦ/Myeloid (CD163+)
MΦ/Myeloid (CD68/CD163+)
Plasma Cells

#### ICI01_s0_a9_ac

ICI01 | 1168/1169 cells assigned (99.9%)

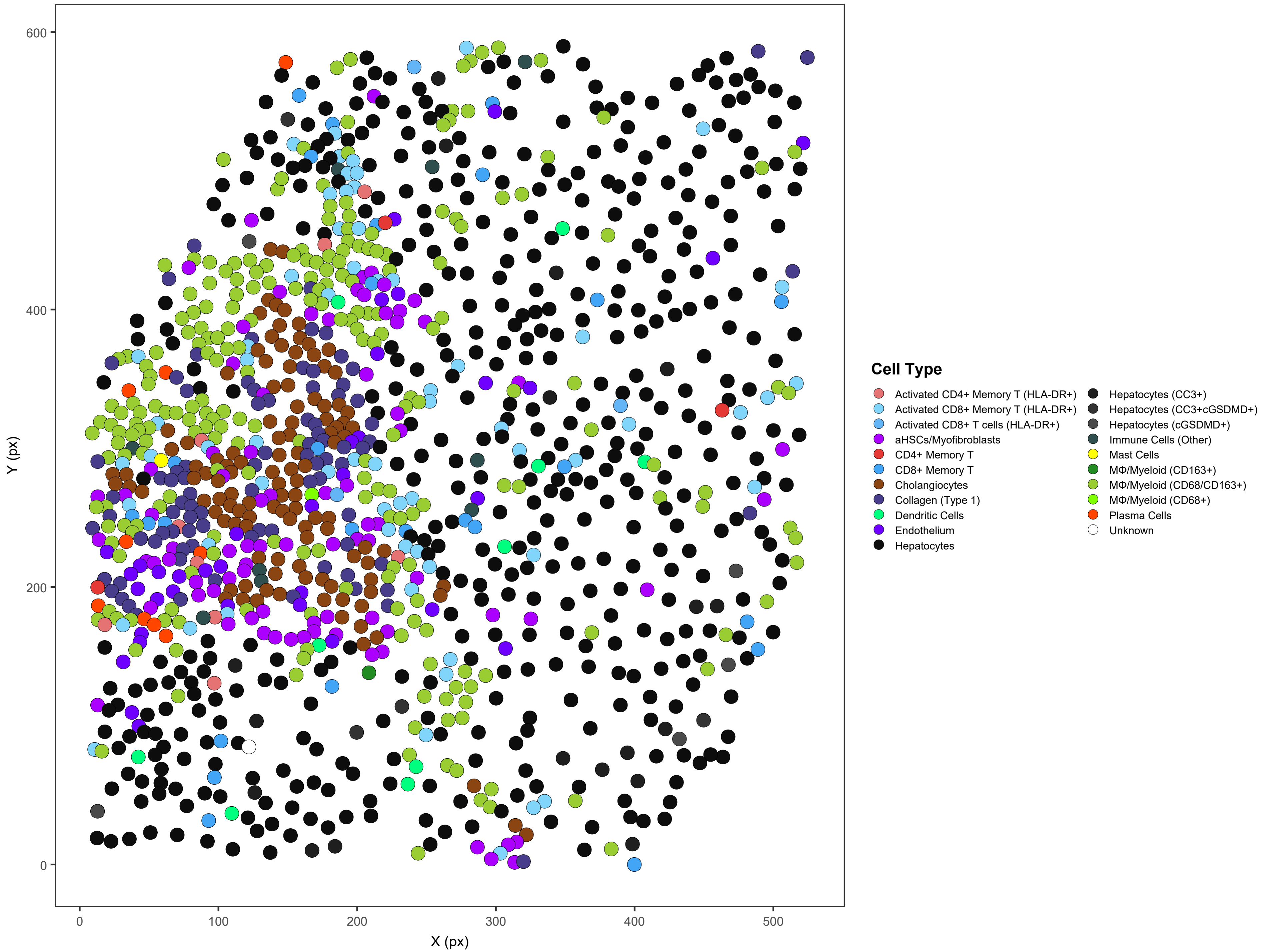

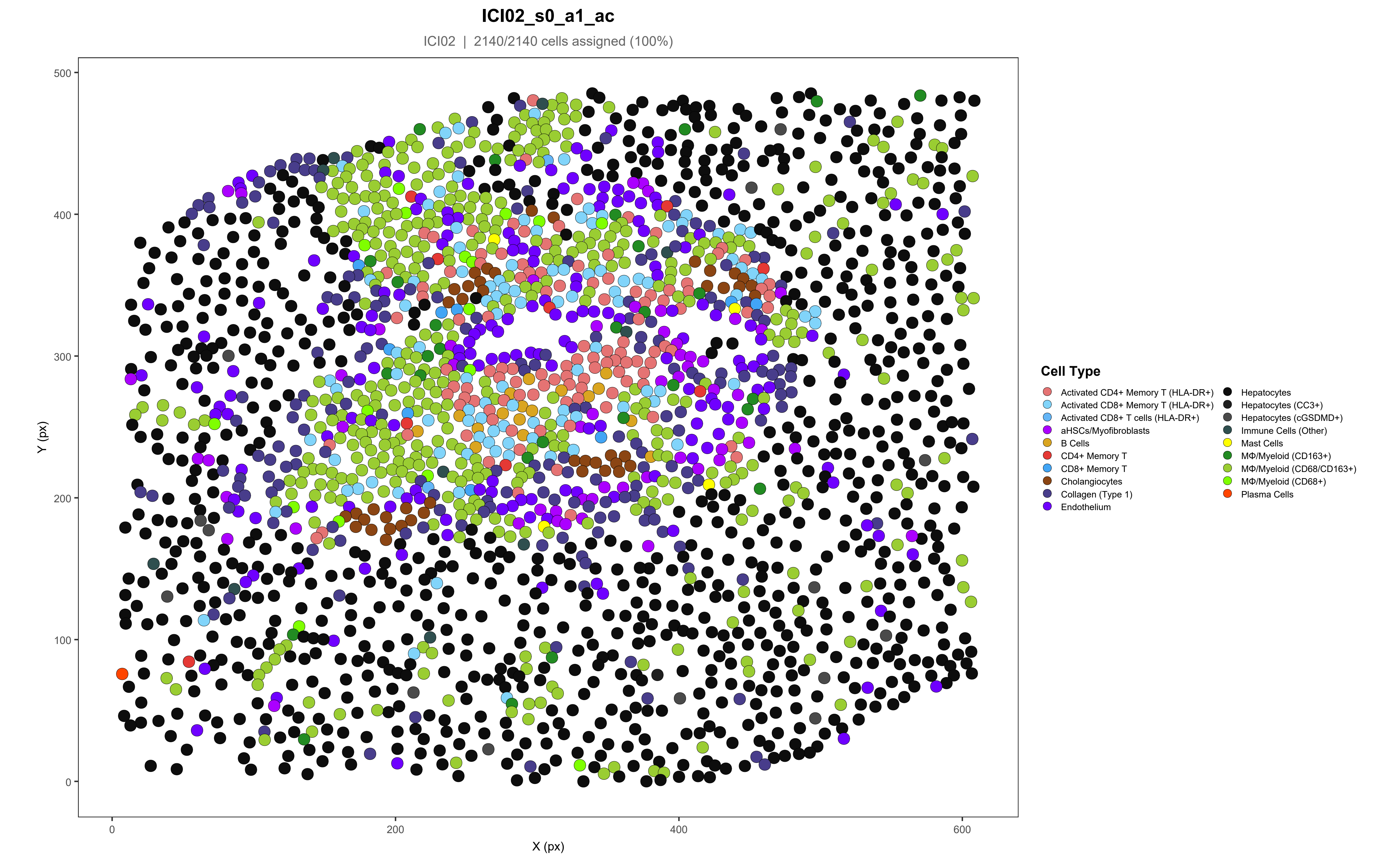
ICI02_s0_a1_ac
ICI02 | 2140/2140 cells assigned (100%)
Y (px)
X (px)
0
100
200
300
400
500
0
200
400
600
Cell Type
Activated CD4+ Memory T (HLA-DR+)
Activated CD8+ Memory T (HLA-DR+)
Activated CD8+ T cells (HLA-DR+)
aHSCs/Myofibroblasts
B Cells
CD4+ Memory T
CD8+ Memory T
Cholangiocytes
Collagen (Type 1)
Endothelium
Hepatocytes
Hepatocytes (CC3+)
Hepatocytes (cGSDMD+)
Immune Cells (Other)
Mast Cells
MΦ/Myeloid (CD163+)
MΦ/Myeloid (CD68/CD163+)
MΦ/Myeloid (CD68+)
Plasma Cells

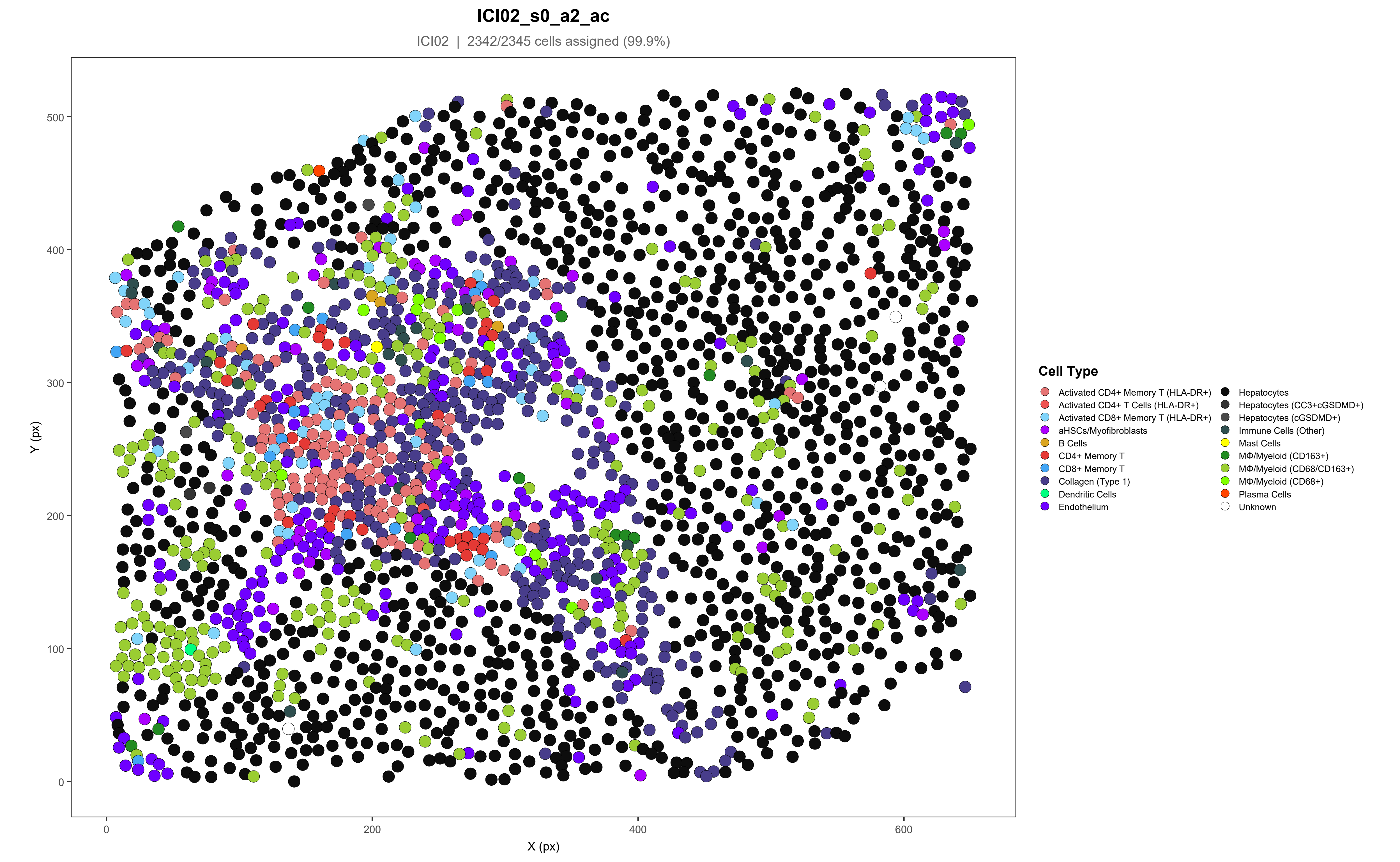
ICI02_s0_a2_ac
ICI02 | 2342/2345 cells assigned (99.9%)
Y (px)
X (px)
0
100
200
300
400
500
600
Cell Type
Activated CD4+ Memory T (HLA-DR+)
Activated CD4+ T Cells (HLA-DR+)
Activated CD8+ Memory T (HLA-DR+)
aHSCs/Myofibroblasts
B Cells
CD4+ Memory T
CD8+ Memory T
Collagen (Type 1)
Dendritic Cells
Endothelium
Hepatocytes
Hepatocytes (CC3+cGSDMD+)
Hepatocytes (cGSDMD+)
Immune Cells (Other)
Mast Cells
MΦ/Myeloid (CD163+)
MΦ/Myeloid (CD68/CD163+)
MΦ/Myeloid (CD68+)
Plasma Cells
Unknown

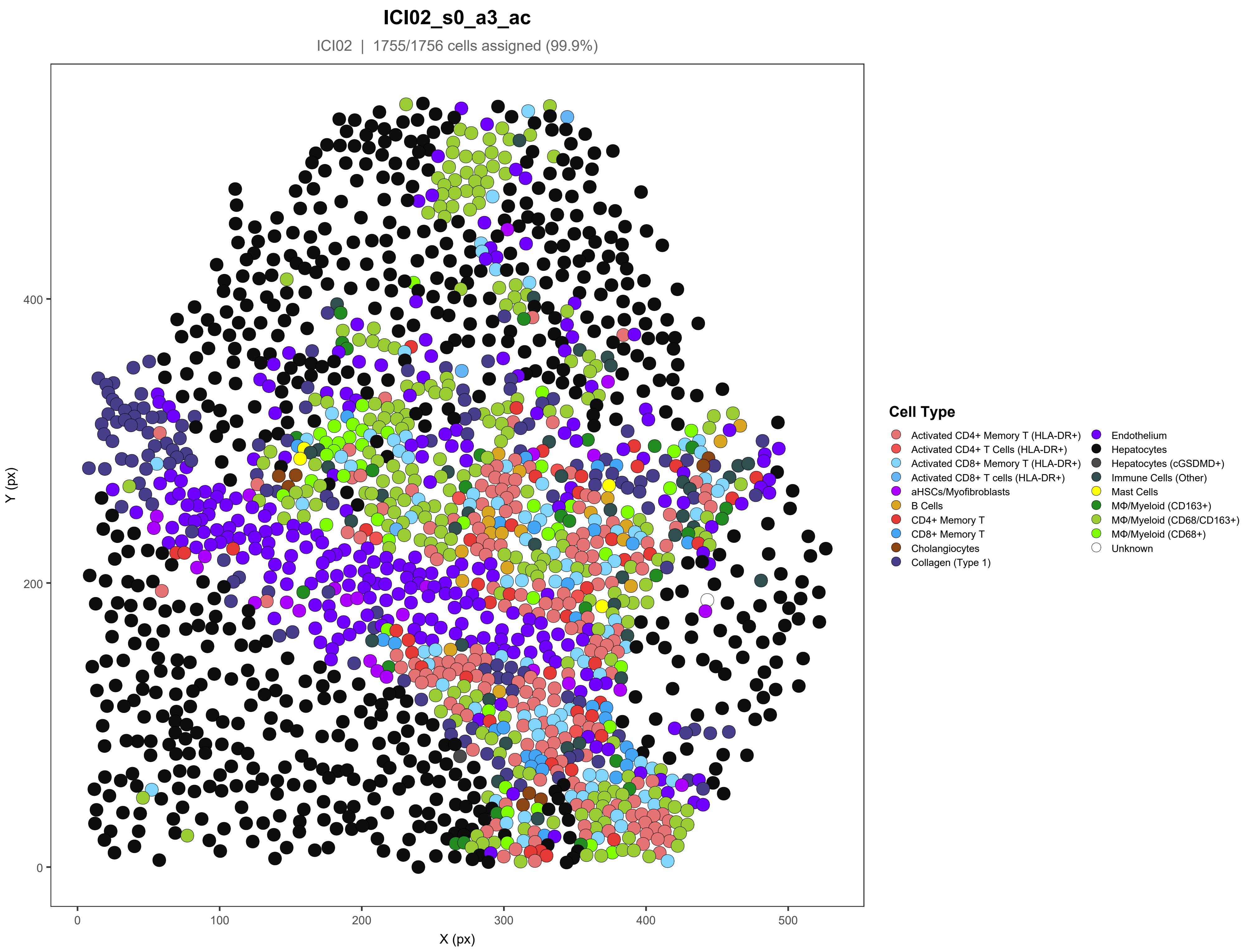
ICI02_s0_a3_ac
ICI02 | 1755/1756 cells assigned (99.9%)
Y (px)
X (px)
0
100
200
300
400
500
0
200
400
Cell Type
Activated CD4+ Memory T (HLA-DR+)
Activated CD4+ T Cells (HLA-DR+)
Activated CD8+ Memory T (HLA-DR+)
Activated CD8+ T cells (HLA-DR+)
aHSCs/Myofibroblasts
B Cells
CD4+ Memory T
CD8+ Memory T
Cholangiocytes
Collagen (Type 1)
Endothelium
Hepatocytes
Hepatocytes (cGSDMD+)
Immune Cells (Other)
Mast Cells
MΦ/Myeloid (CD163+)
MΦ/Myeloid (CD68/CD163+)
MΦ/Myeloid (CD68+)
Unknown

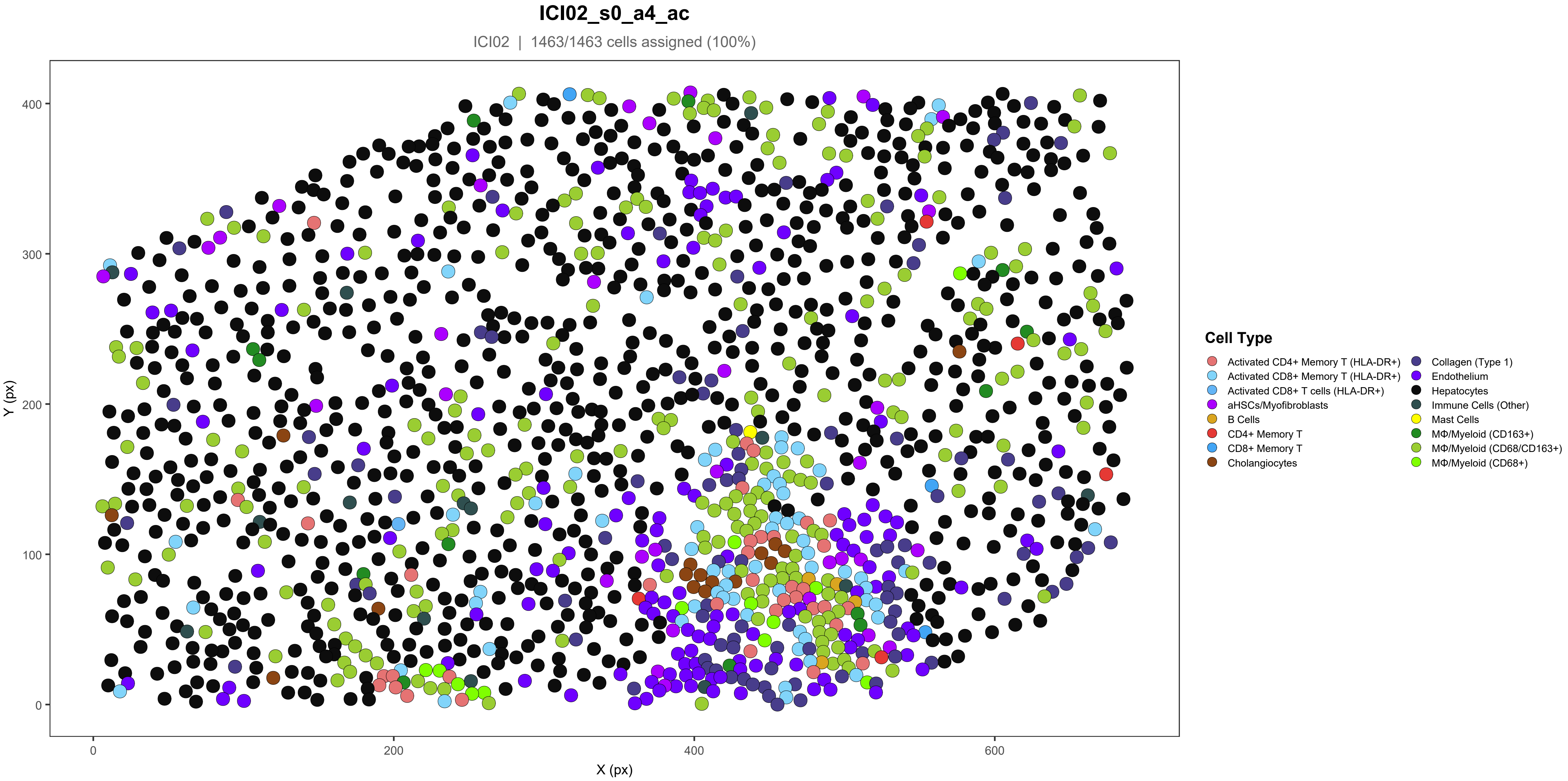
ICI02_s0_a4_ac
ICI02 | 1463/1463 cells assigned (100%)
Y (px)
X (px)
0
100
200
300
400
0
200
400
600
Cell Type
Activated CD4+ Memory T (HLA-DR+)
Activated CD8+ Memory T (HLA-DR+)
Activated CD8+ T cells (HLA-DR+)
aHSCs/Myofibroblasts
B Cells
CD4+ Memory T
CD8+ Memory T
Cholangiocytes
Collagen (Type 1)
Endothelium
Hepatocytes
Immune Cells (Other)
Mast Cells
MΦ/Myeloid (CD163+)
MΦ/Myeloid (CD68/CD163+)
MΦ/Myeloid (CD68+)

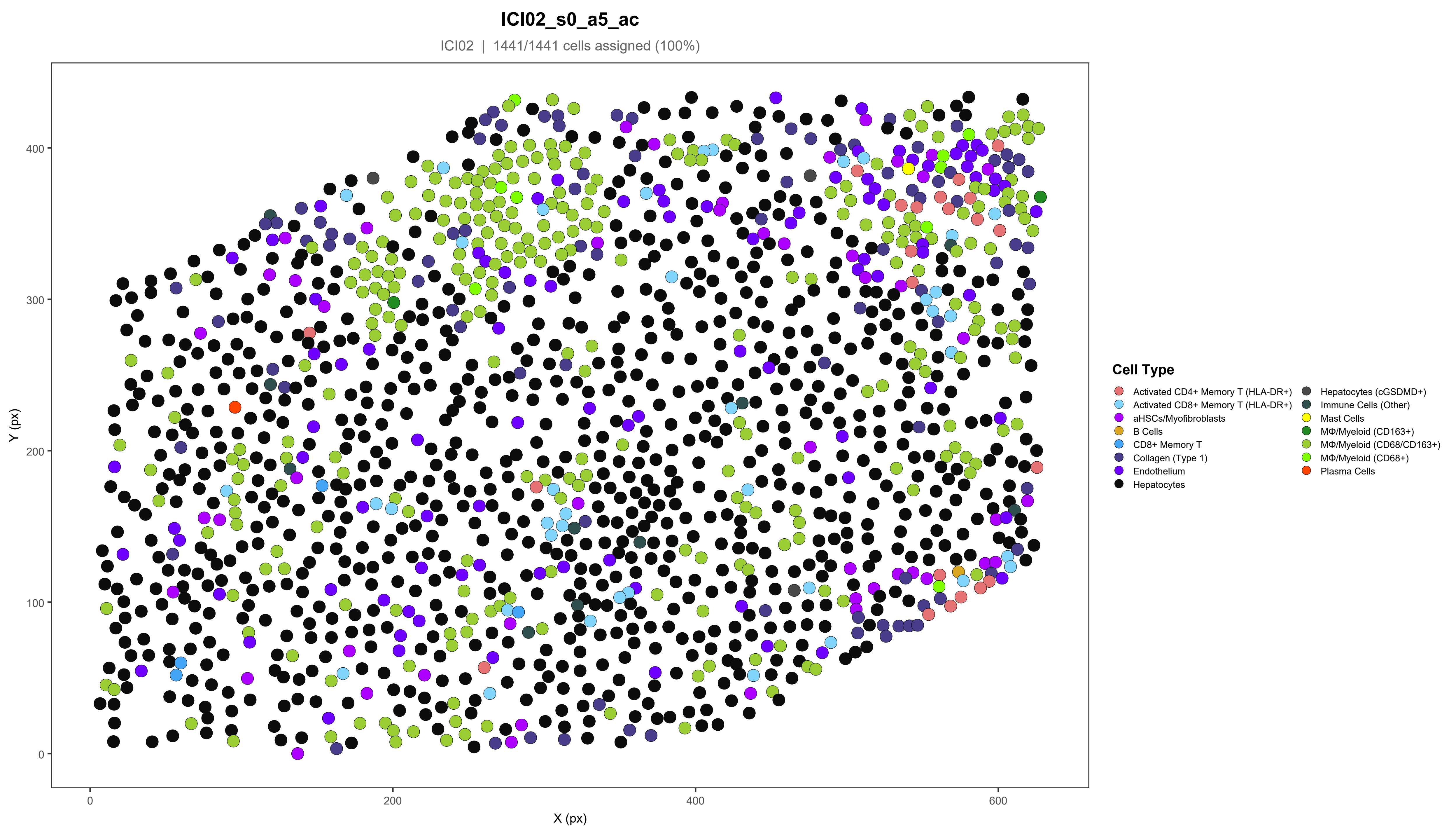
ICI02_s0_a5_ac
ICI02 | 1441/1441 cells assigned (100%)
Y (px)
X (px)
0
100
200
300
400
0
200
400
600
Cell Type
Activated CD4+ Memory T (HLA-DR+)
Activated CD8+ Memory T (HLA-DR+)
aHSCs/Myofibroblasts
B Cells
CD8+ Memory T
Collagen (Type 1)
Endothelium
Hepatocytes
Hepatocytes (cGSDMD+)
Immune Cells (Other)
Mast Cells
MΦ/Myeloid (CD163+)
MΦ/Myeloid (CD68/CD163+)
MΦ/Myeloid (CD68+)
Plasma Cells

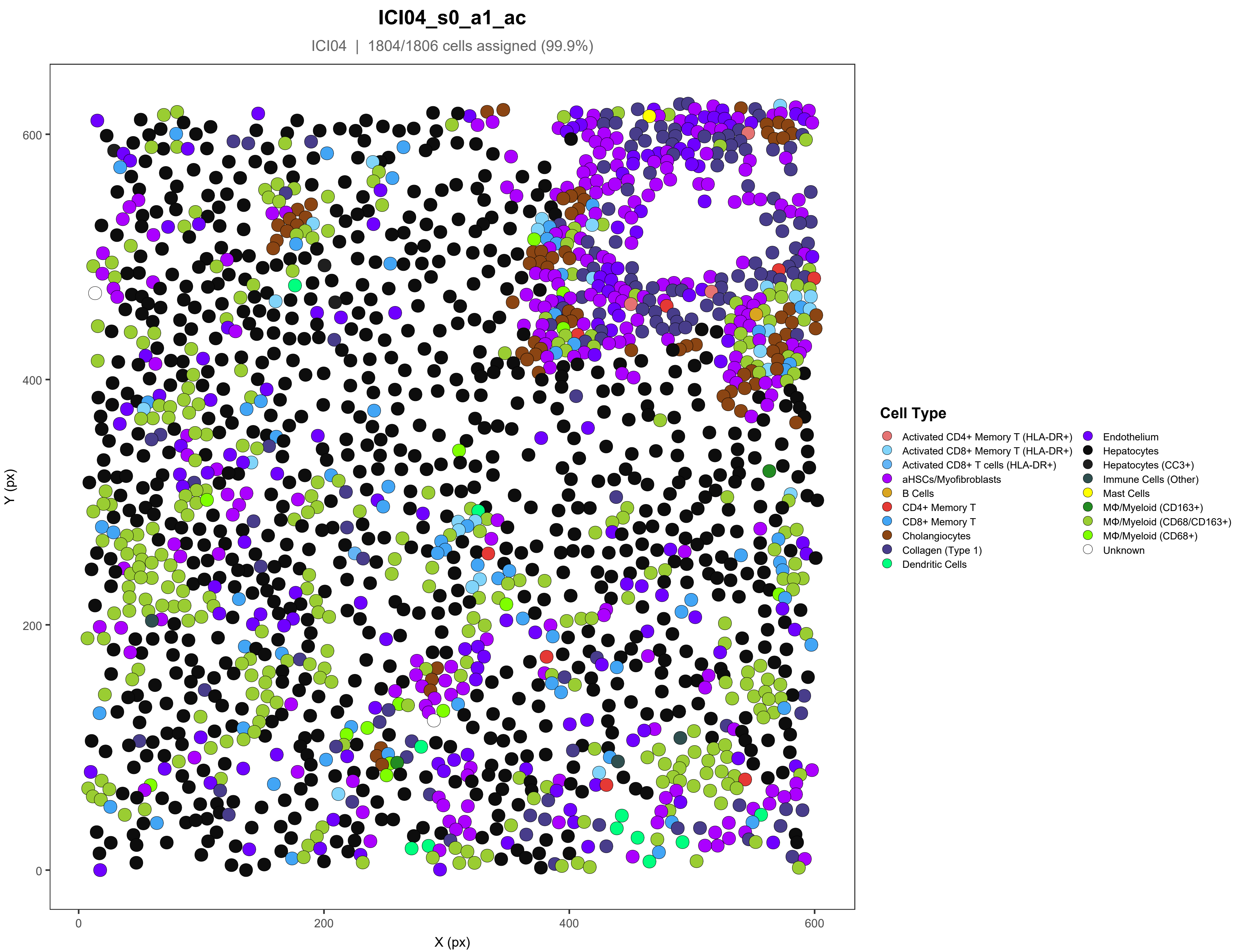
ICI04_s0_a1_ac
ICI04 | 1804/1806 cells assigned (99.9%)
Y (px)
X (px)
0
200
400
600
Cell Type
Activated CD4+ Memory T (HLA-DR+)
Activated CD8+ Memory T (HLA-DR+)
Activated CD8+ T cells (HLA-DR+)
aHSCs/Myofibroblasts
B Cells
CD4+ Memory T
CD8+ Memory T
Cholangiocytes
Collagen (Type 1)
Dendritic Cells
Endothelium
Hepatocytes
Hepatocytes (CC3+)
Immune Cells (Other)
Mast Cells
MΦ/Myeloid (CD163+)
MΦ/Myeloid (CD68/CD163+)
MΦ/Myeloid (CD68+)
Unknown

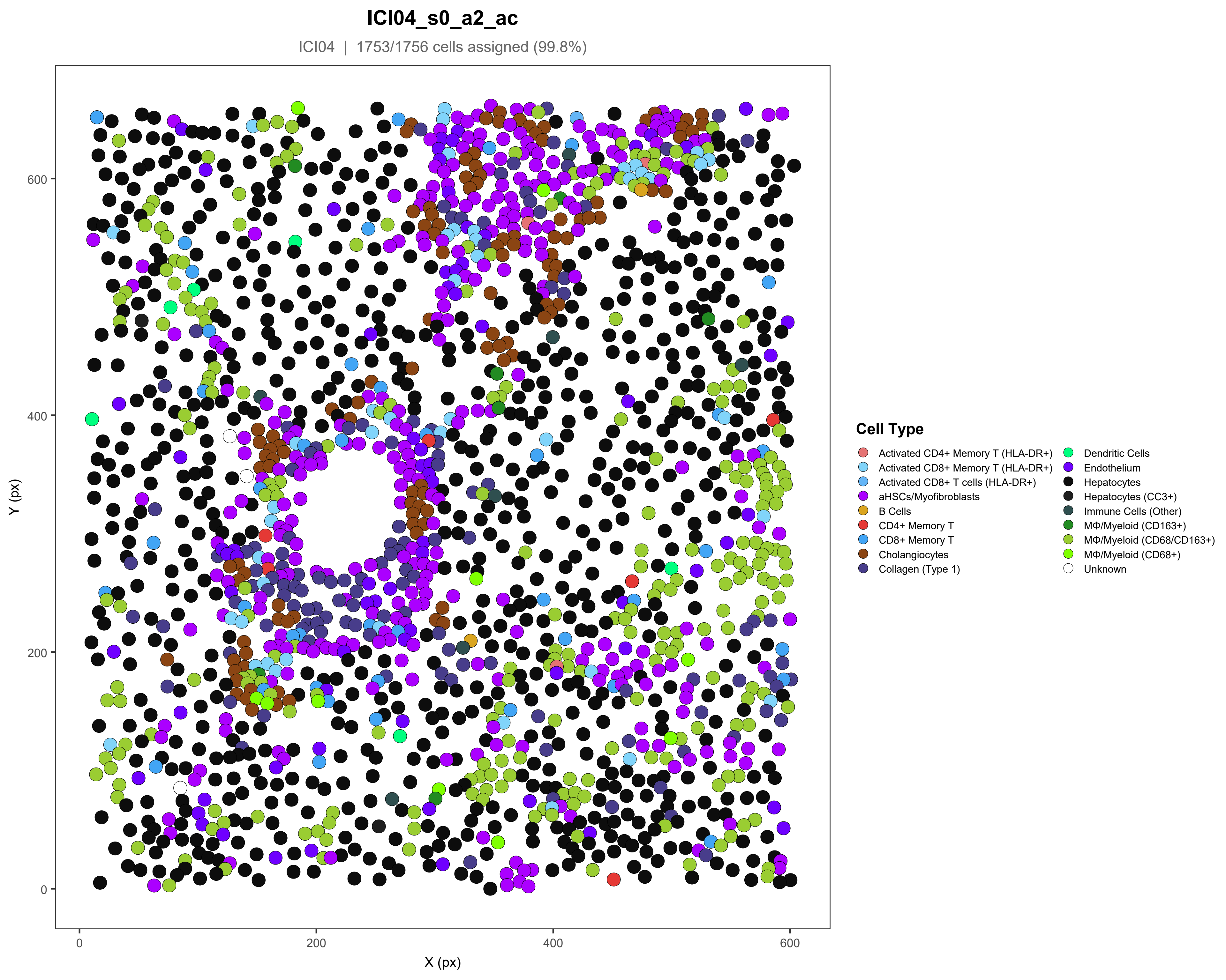

ICI04_s0_a2_ac
ICI04 | 1753/1756 cells assigned (99.8%)
Y (px)
X (px)
0
200
400
600
Cell Type
Activated CD4+ Memory T (HLA-DR+)
Activated CD8+ Memory T (HLA-DR+)
Activated CD8+ T cells (HLA-DR+)
aHSCs/Myofibroblasts
B Cells
CD4+ Memory T
CD8+ Memory T
Cholangiocytes
Collagen (Type 1)
Dendritic Cells
Endothelium
Hepatocytes
Hepatocytes (CC3+)
Immune Cells (Other)
MΦ/Myeloid (CD163+)
MΦ/Myeloid (CD68/CD163+)
MΦ/Myeloid (CD68+)
Unknown

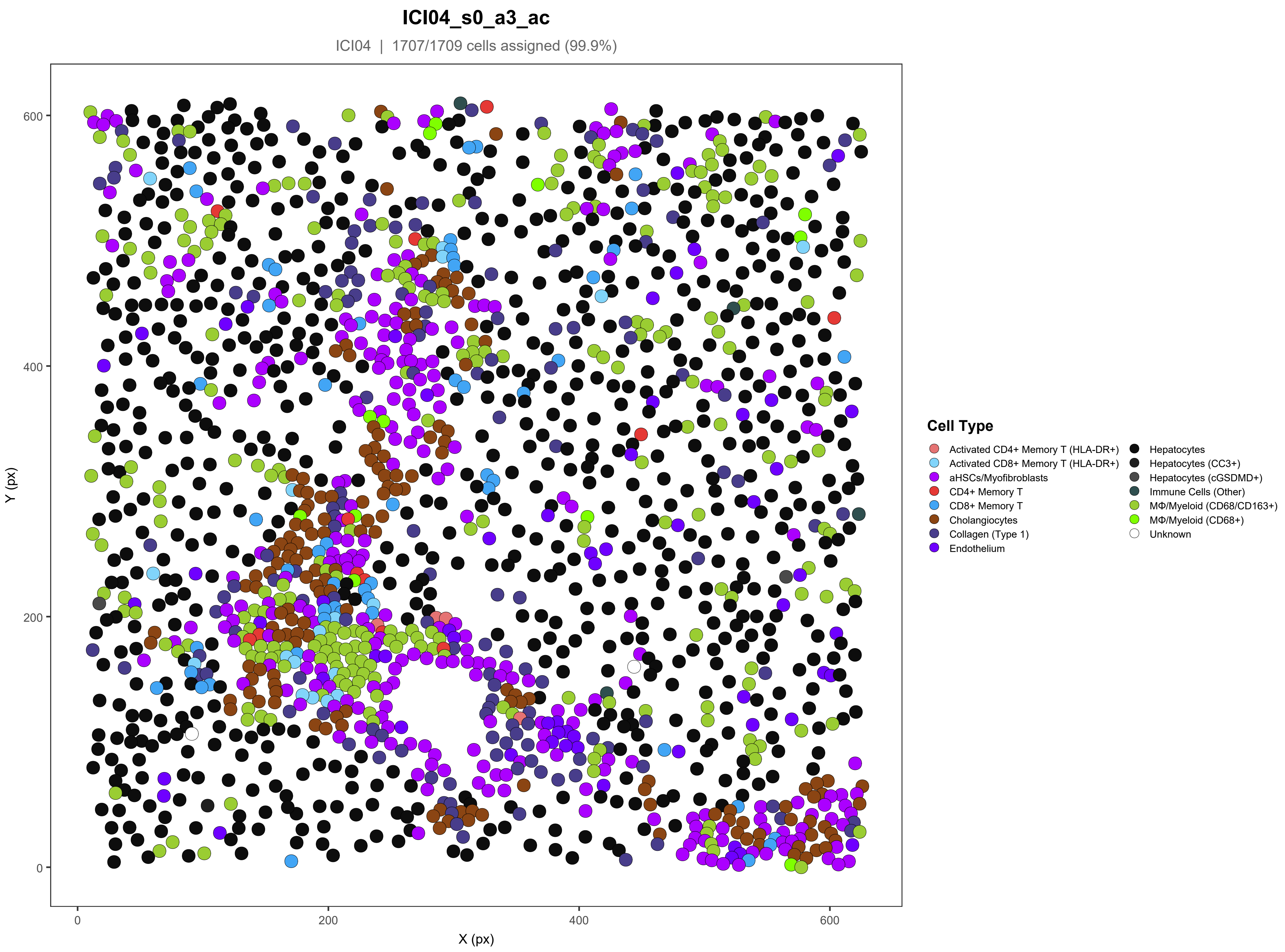

ICI04_s0_a3_ac
ICI04 | 1707/1709 cells assigned (99.9%)
Y (px)
X (px)
0
200
400
600
Cell Type
Activated CD4+ Memory T (HLA-DR+)
Activated CD8+ Memory T (HLA-DR+)
aHSCs/Myofibroblasts
CD4+ Memory T
CD8+ Memory T
Cholangiocytes
Collagen (Type 1)
Endothelium
Hepatocytes
Hepatocytes (CC3+)
Hepatocytes (cGSDMD+)
Immune Cells (Other)
MΦ/Myeloid (CD68/CD163+)
MΦ/Myeloid (CD68+)
Unknown

#### ICI04_s0_a4_ac

ICI04 | 1920/1924 cells assigned (99.8%)

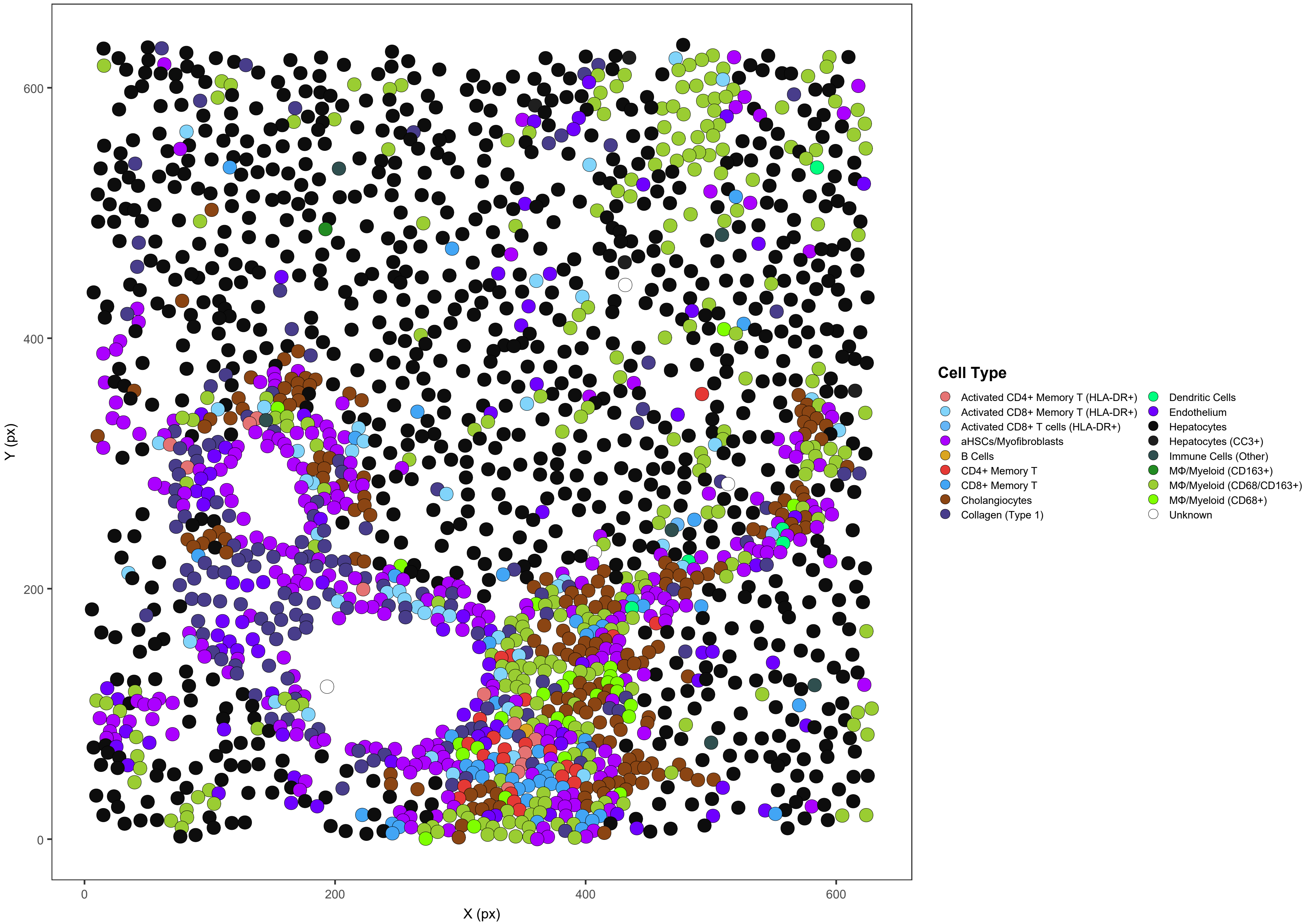

### ICI04_s0_a5_ac

ICI04 | 1920/1920 cells assigned (100%)

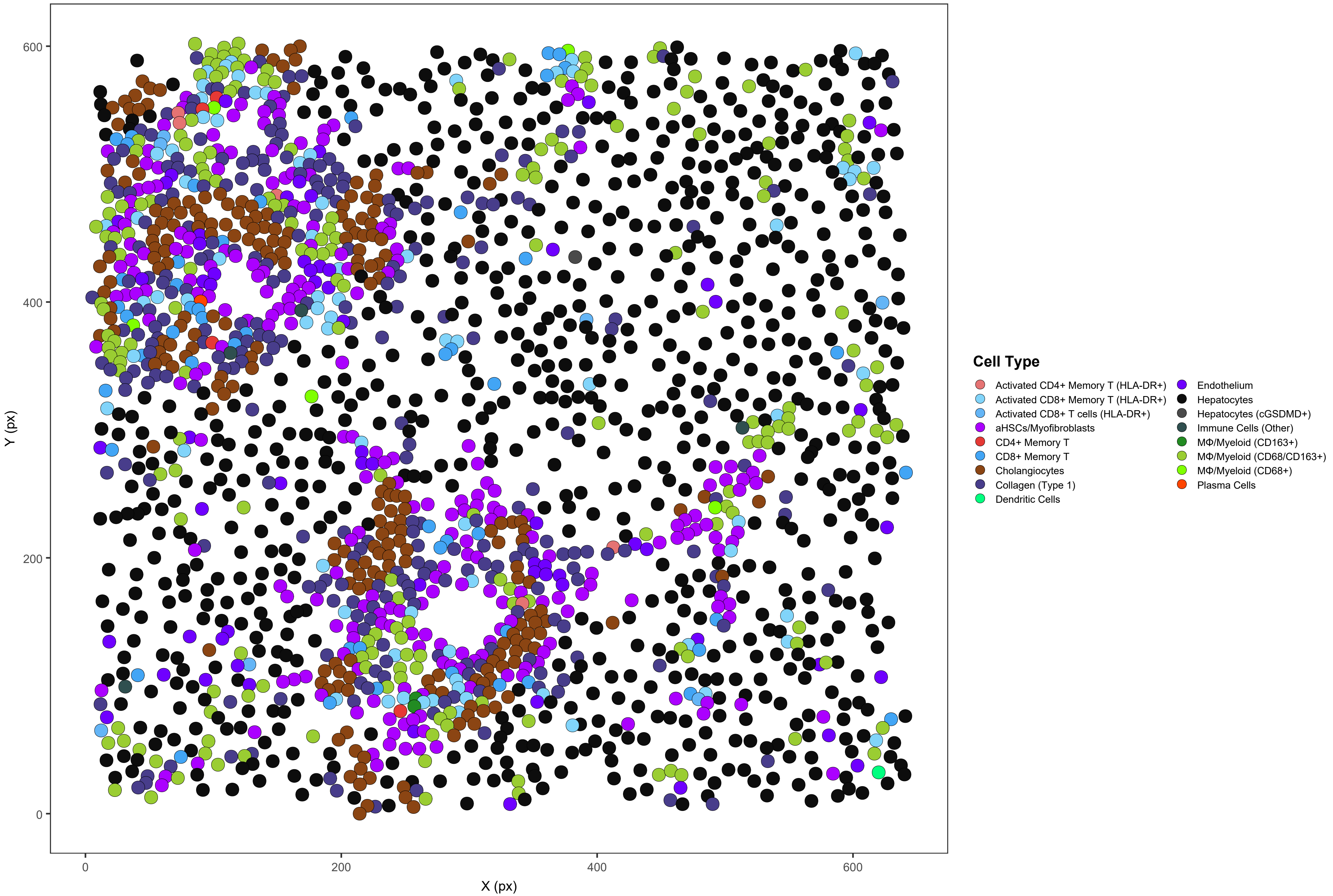

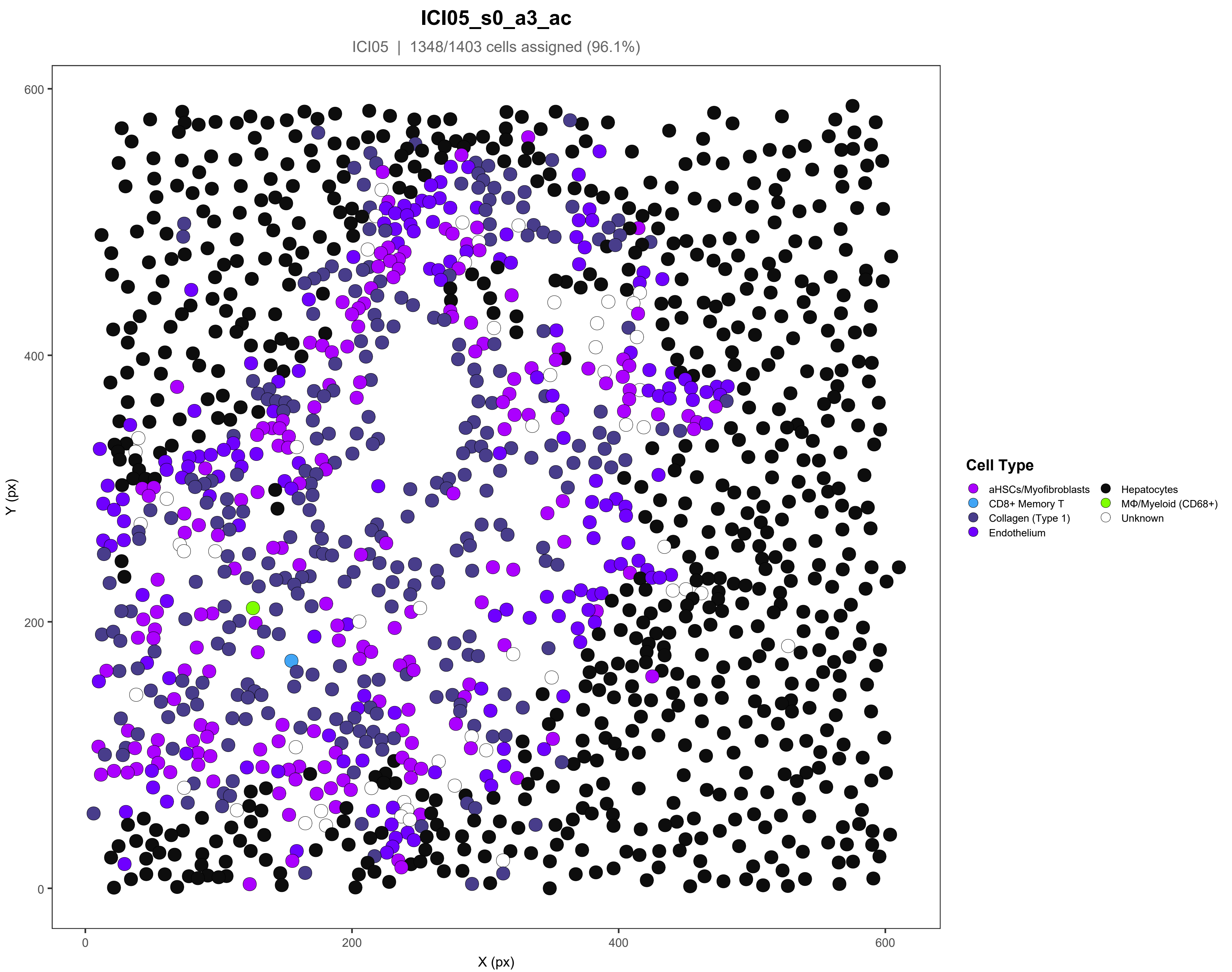
ICI05_s0_a3_ac
ICI05 | 1348/1403 cells assigned (96.1%)
Y (px)
X (px)
600
400
200
0
0
200
400
600
Cell Type
aHSCs/Myofibroblasts
CD8+ Memory T
Collagen (Type 1)
Endothelium
Hepatocytes
MΦ/Myeloid (CD68+)
Unknown

### ICI05_s0_a5_ac

ICI05 | 1329/1356 cells assigned (98%)

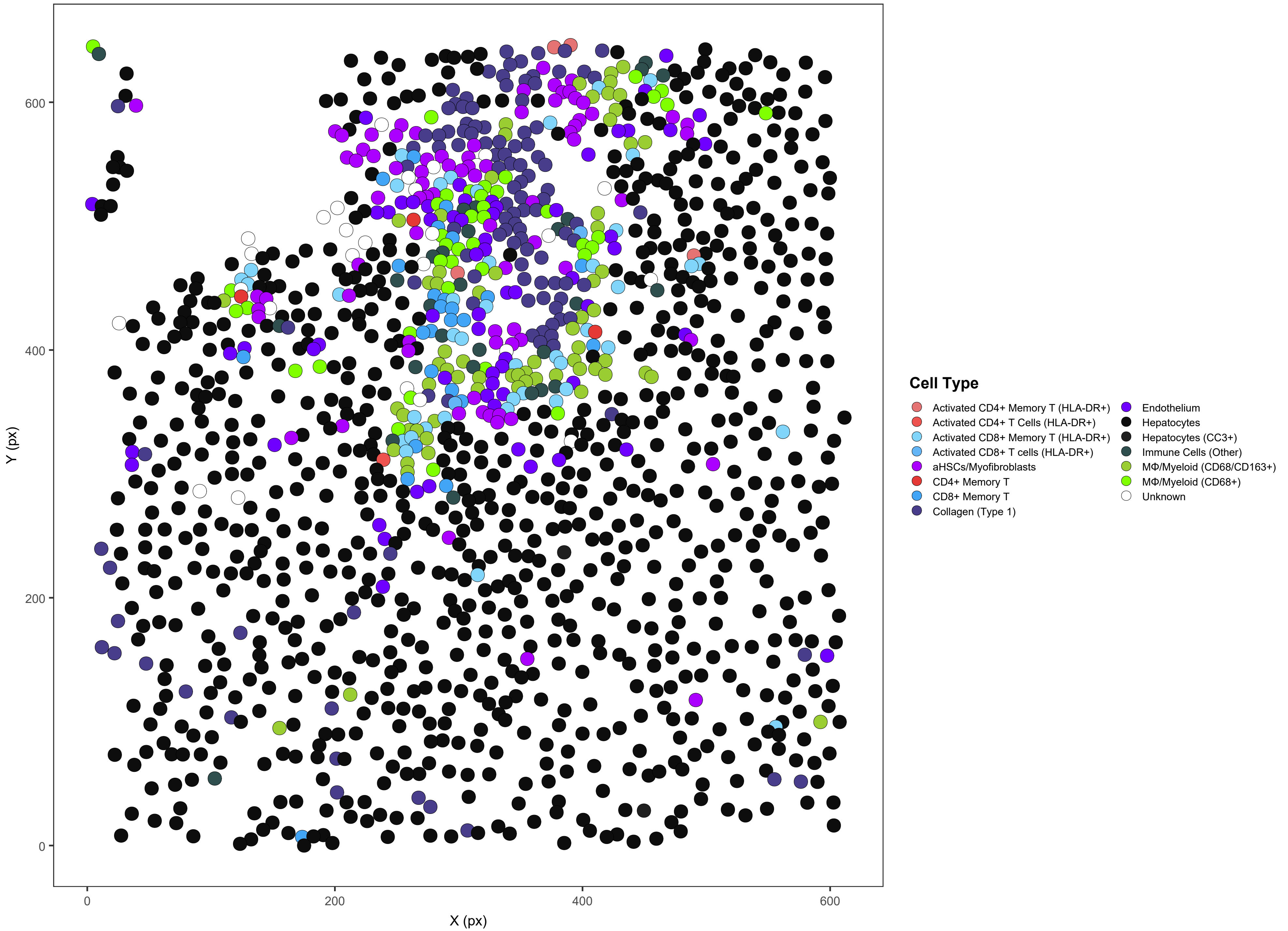

### ICI06_s0_a1_ac

ICI06 | 2800/2801 cells assigned (100%)

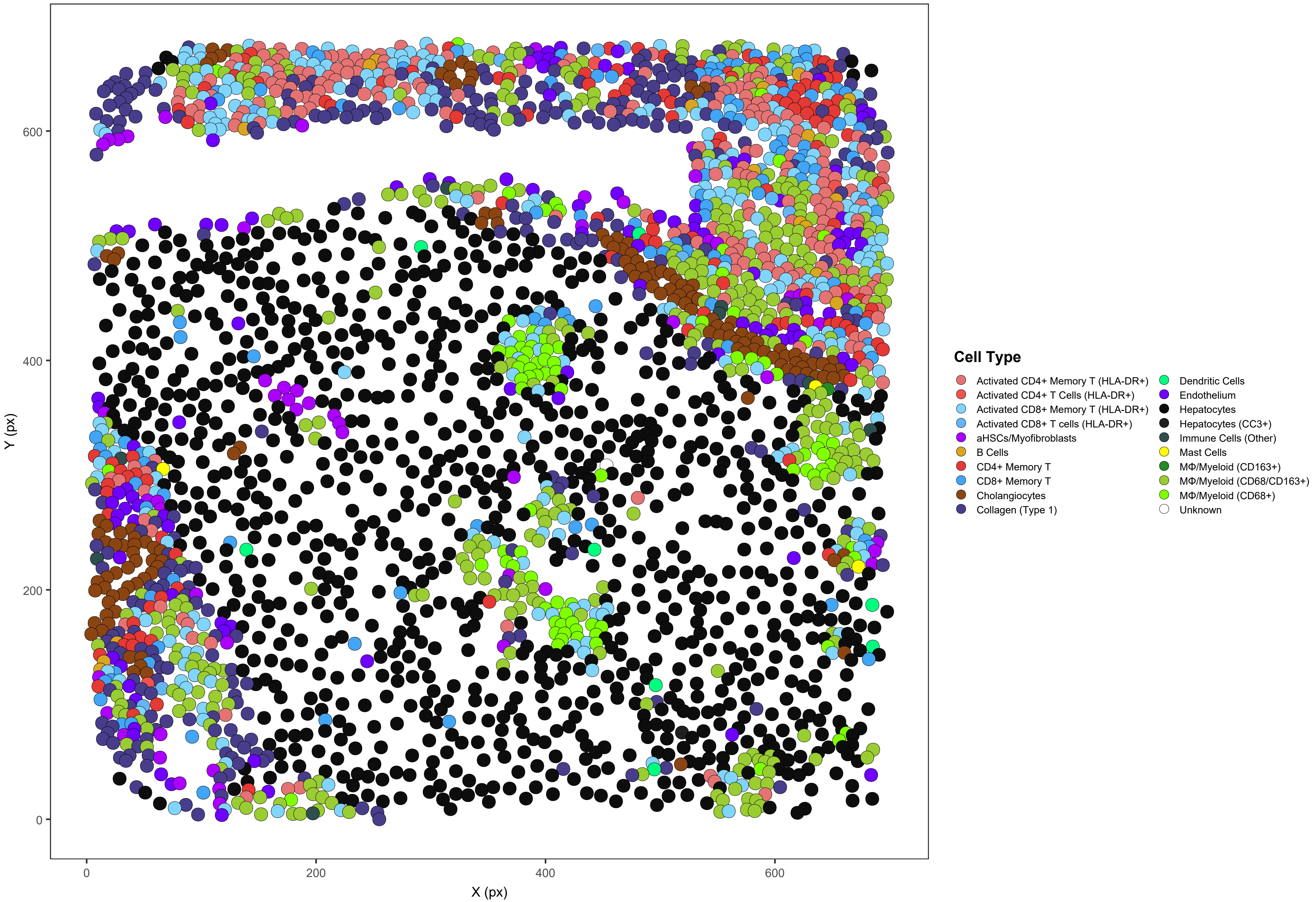

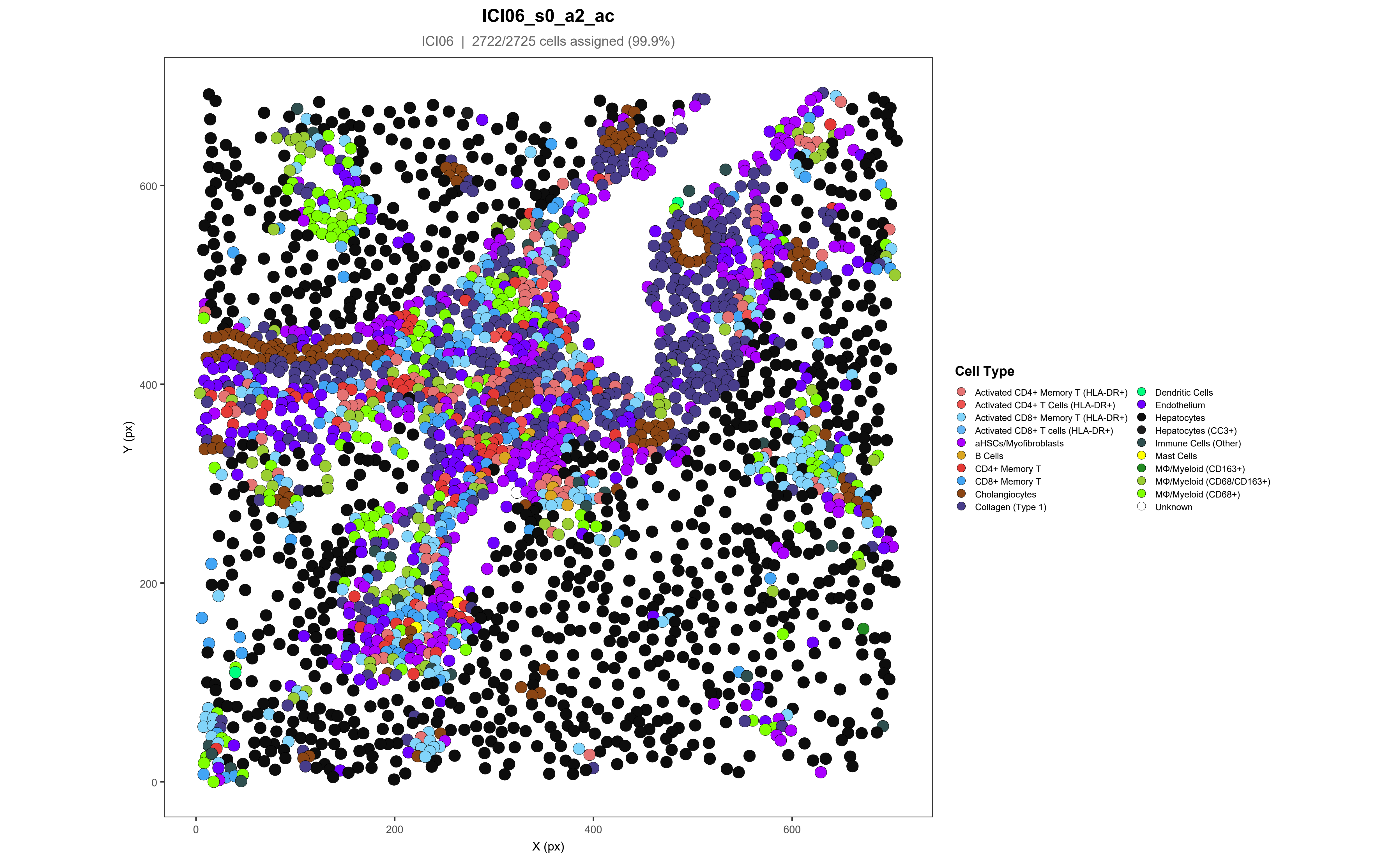
ICI06_s0_a2_ac
ICI06 | 2722/2725 cells assigned (99.9%)
Y (px)
X (px)
0
200
400
600
Cell Type
Activated CD4+ Memory T (HLA-DR+)
Activated CD4+ T Cells (HLA-DR+)
Activated CD8+ Memory T (HLA-DR+)
Activated CD8+ T cells (HLA-DR+)
aHSCs/Myofibroblasts
B Cells
CD4+ Memory T
CD8+ Memory T
Cholangiocytes
Collagen (Type 1)
Dendritic Cells
Endothelium
Hepatocytes
Hepatocytes (CC3+)
Immune Cells (Other)
Mast Cells
MΦ/Myeloid (CD163+)
MΦ/Myeloid (CD68/CD163+)
MΦ/Myeloid (CD68+)
Unknown

### ICI06_s0_a3_ac

ICI06 | 1735/1739 cells assigned (99.8%)

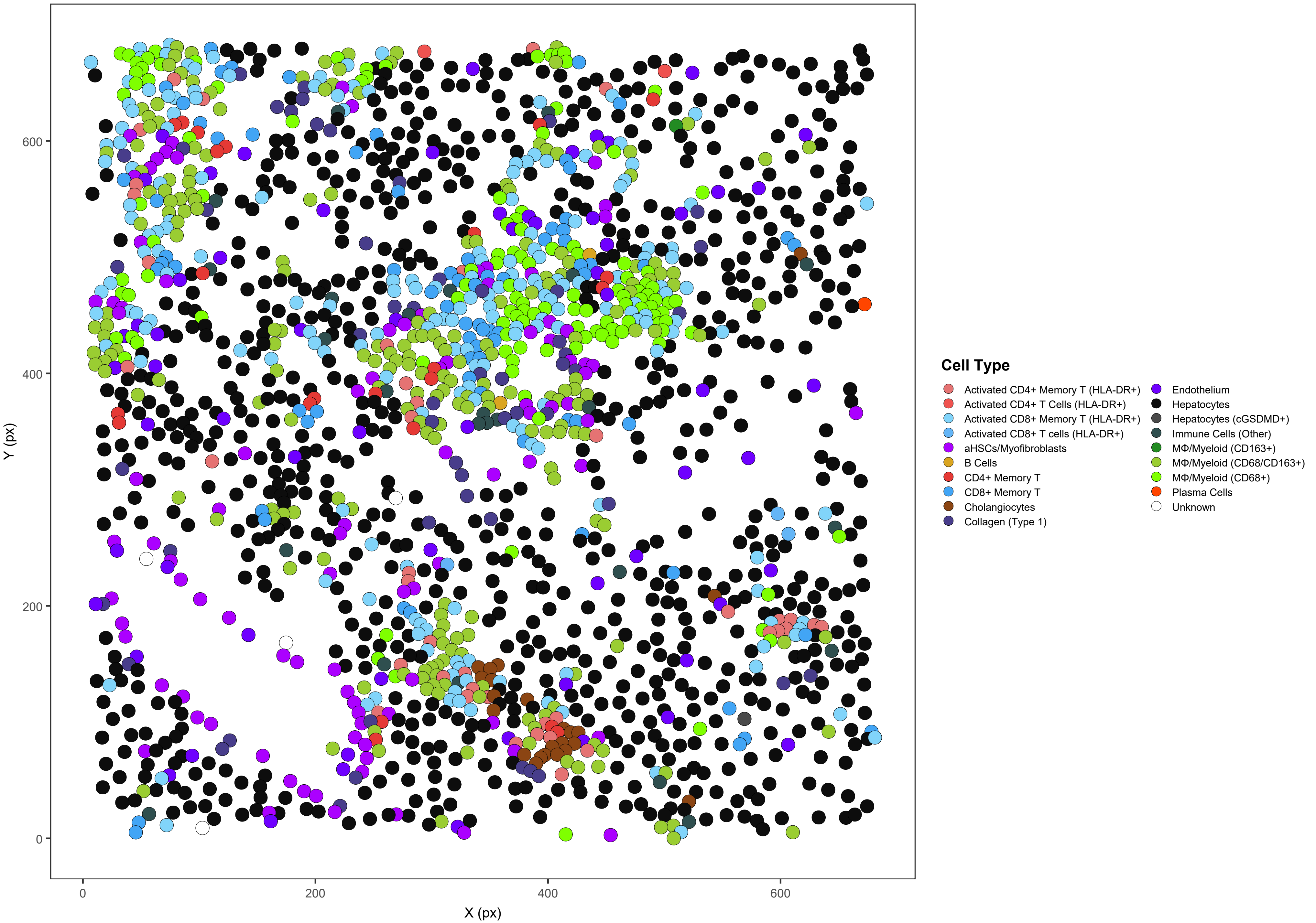

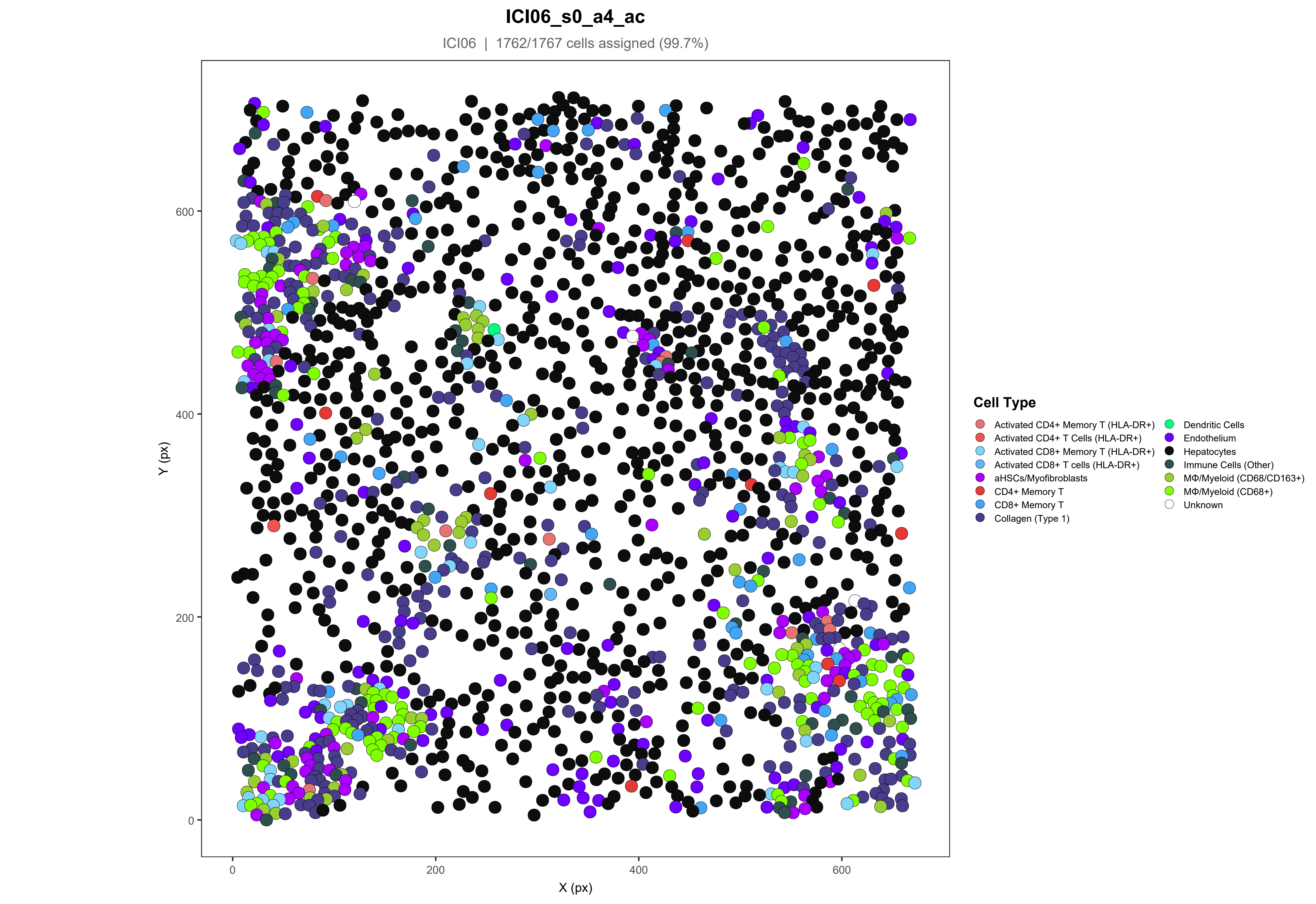
ICI06_s0_a4_ac
ICI06 | 1762/1767 cells assigned (99.7%)
Y (px)
X (px)
0
200
400
600
Cell Type
Activated CD4+ Memory T (HLA-DR+)
Activated CD4+ T Cells (HLA-DR+)
Activated CD8+ Memory T (HLA-DR+)
Activated CD8+ T cells (HLA-DR+)
aHSCs/Myofibroblasts
CD4+ Memory T
CD8+ Memory T
Collagen (Type 1)
Dendritic Cells
Endothelium
Hepatocytes
Immune Cells (Other)
MΦ/Myeloid (CD68/CD163+)
MΦ/Myeloid (CD68+)
Unknown

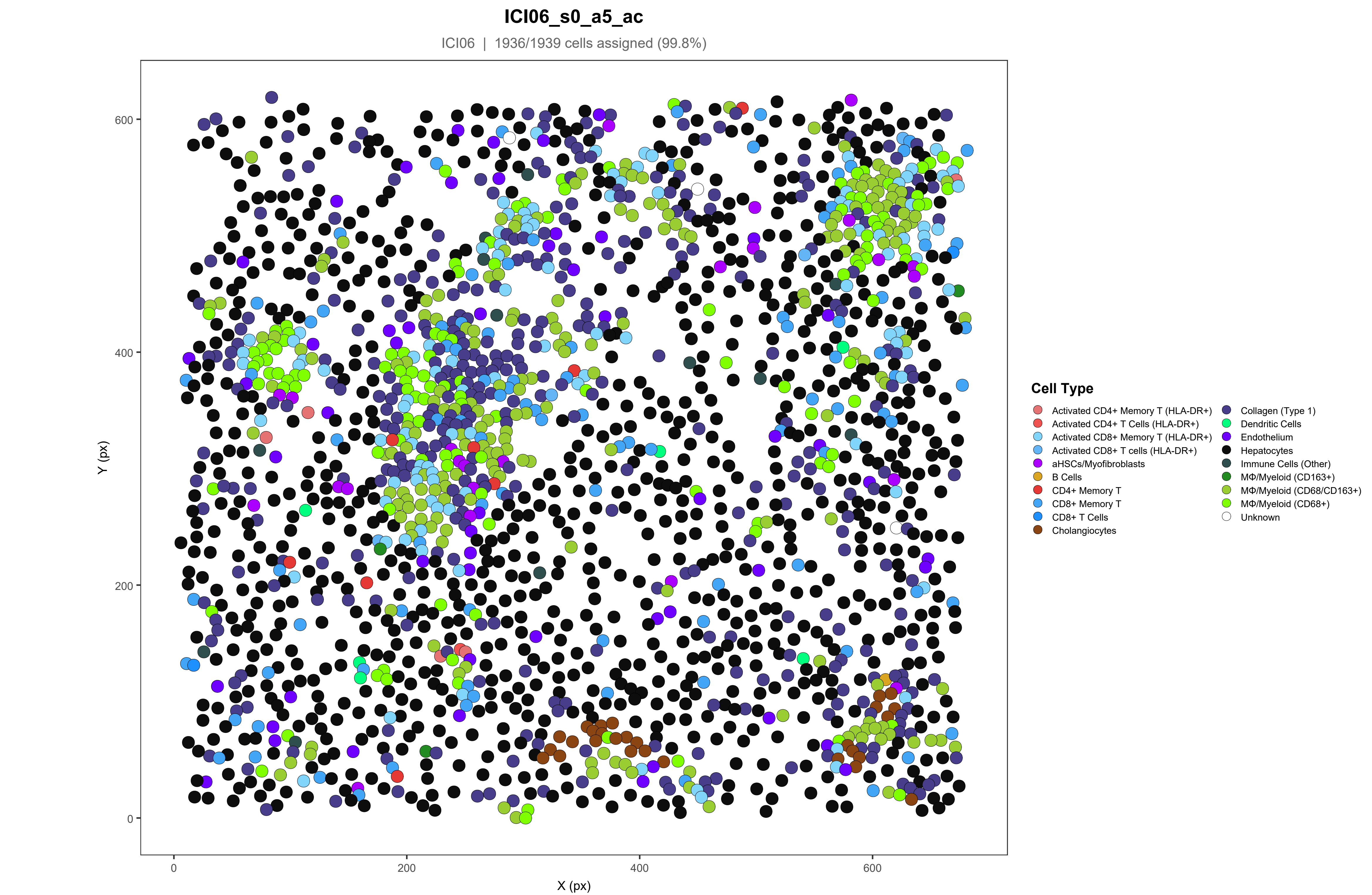
ICI06_s0_a5_ac
ICI06 | 1936/1939 cells assigned (99.8%)
Y (px)
X (px)
0
200
400
600
Cell Type
Activated CD4+ Memory T (HLA-DR+)
Activated CD4+ T Cells (HLA-DR+)
Activated CD8+ Memory T (HLA-DR+)
Activated CD8+ T cells (HLA-DR+)
aHSCs/Myofibroblasts
B Cells
CD4+ Memory T
CD8+ Memory T
CD8+ T Cells
Cholangiocytes
Collagen (Type 1)
Dendritic Cells
Endothelium
Hepatocytes
Immune Cells (Other)
MΦ/Myeloid (CD163+)
MΦ/Myeloid (CD68/CD163+)
MΦ/Myeloid (CD68+)
Unknown

#### ICI07_s0_a2_ac

ICI07 | 1871/1876 cells assigned (99.7%)

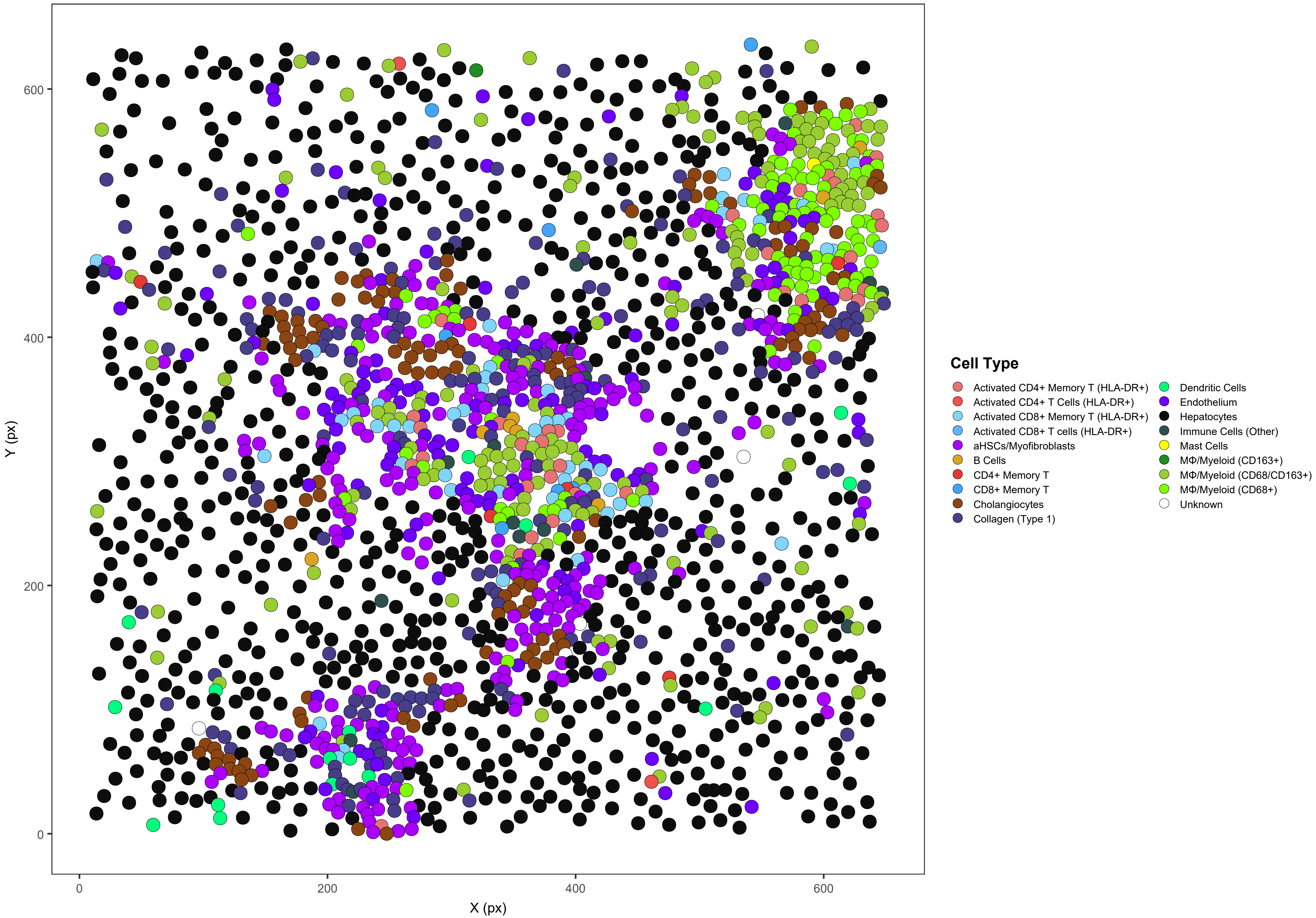

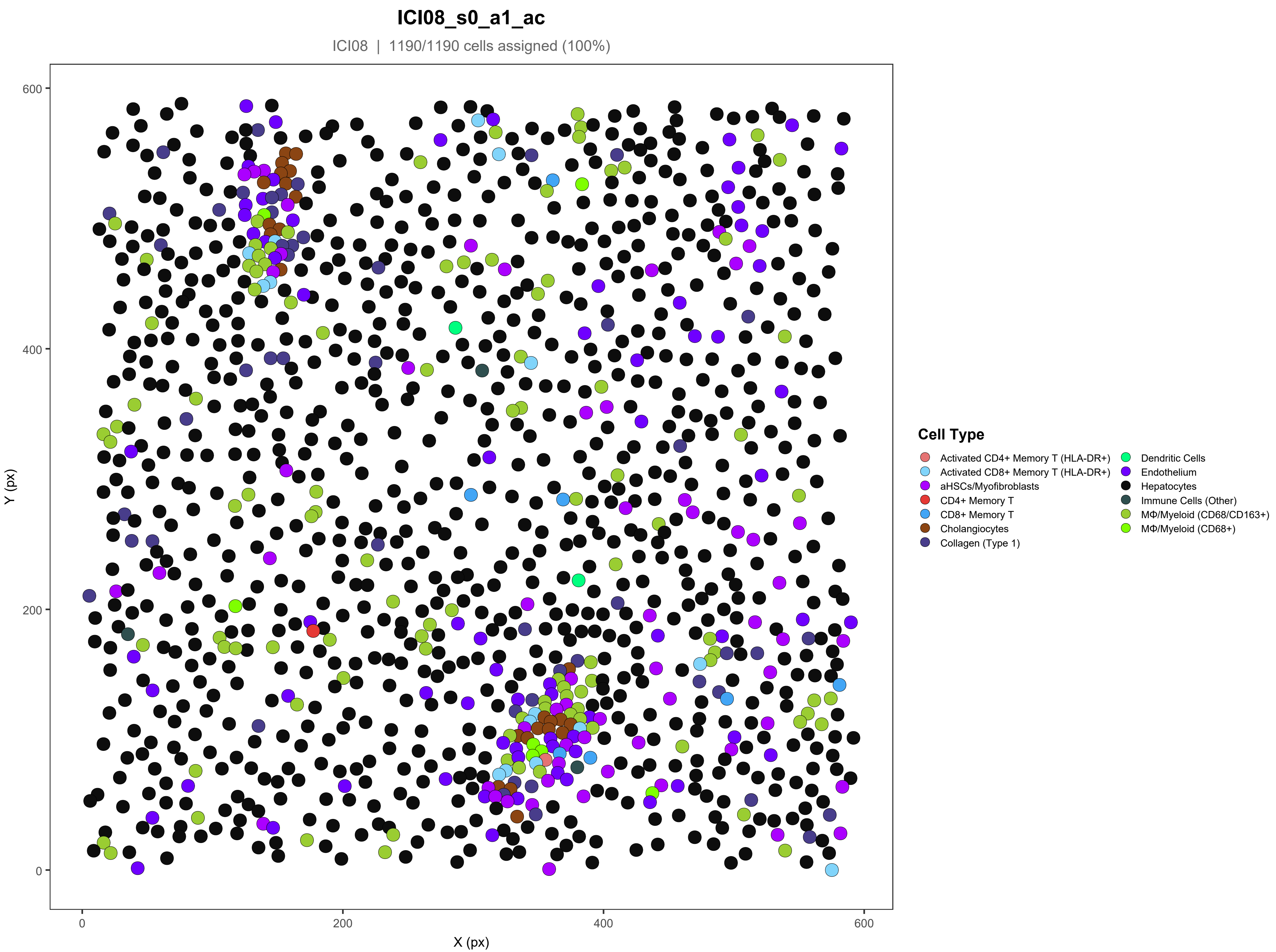
ICI08_s0_a1_ac
ICI08 | 1190/1190 cells assigned (100%)
Y (px)
X (px)
0
200
400
600
Cell Type
Activated CD4+ Memory T (HLA-DR+)
Activated CD8+ Memory T (HLA-DR+)
aHSCs/Myofibroblasts
CD4+ Memory T
CD8+ Memory T
Cholangiocytes
Collagen (Type 1)
Dendritic Cells
Endothelium
Hepatocytes
Immune Cells (Other)
MΦ/Myeloid (CD68/CD163+)
MΦ/Myeloid (CD68+)

### ICI08_s0_a2_ac

ICI08 | 1768/1772 cells assigned (99.8%)

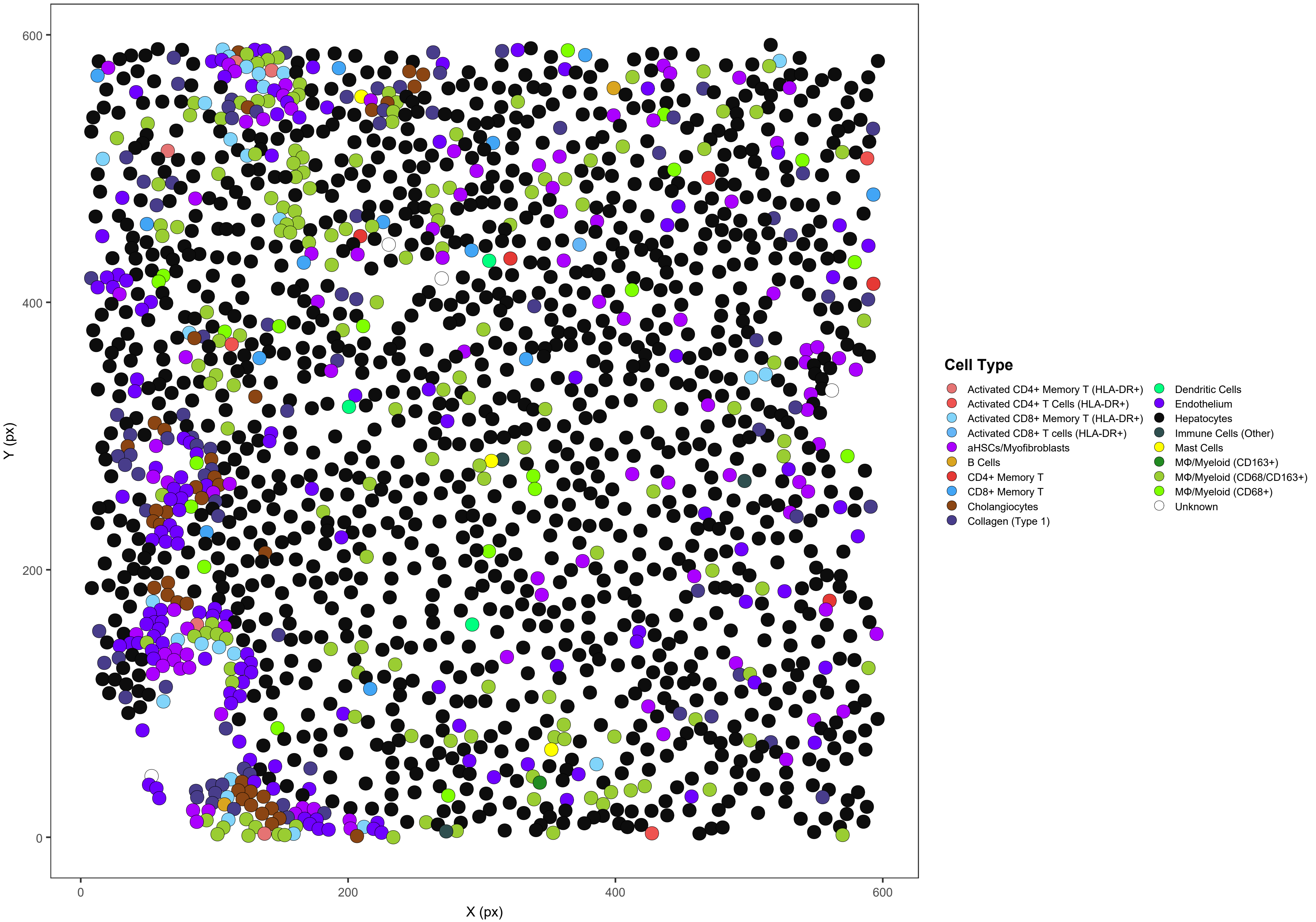

#### ICI08_s0_a3_ac

ICI08 | 1338/1340 cells assigned (99.9%)

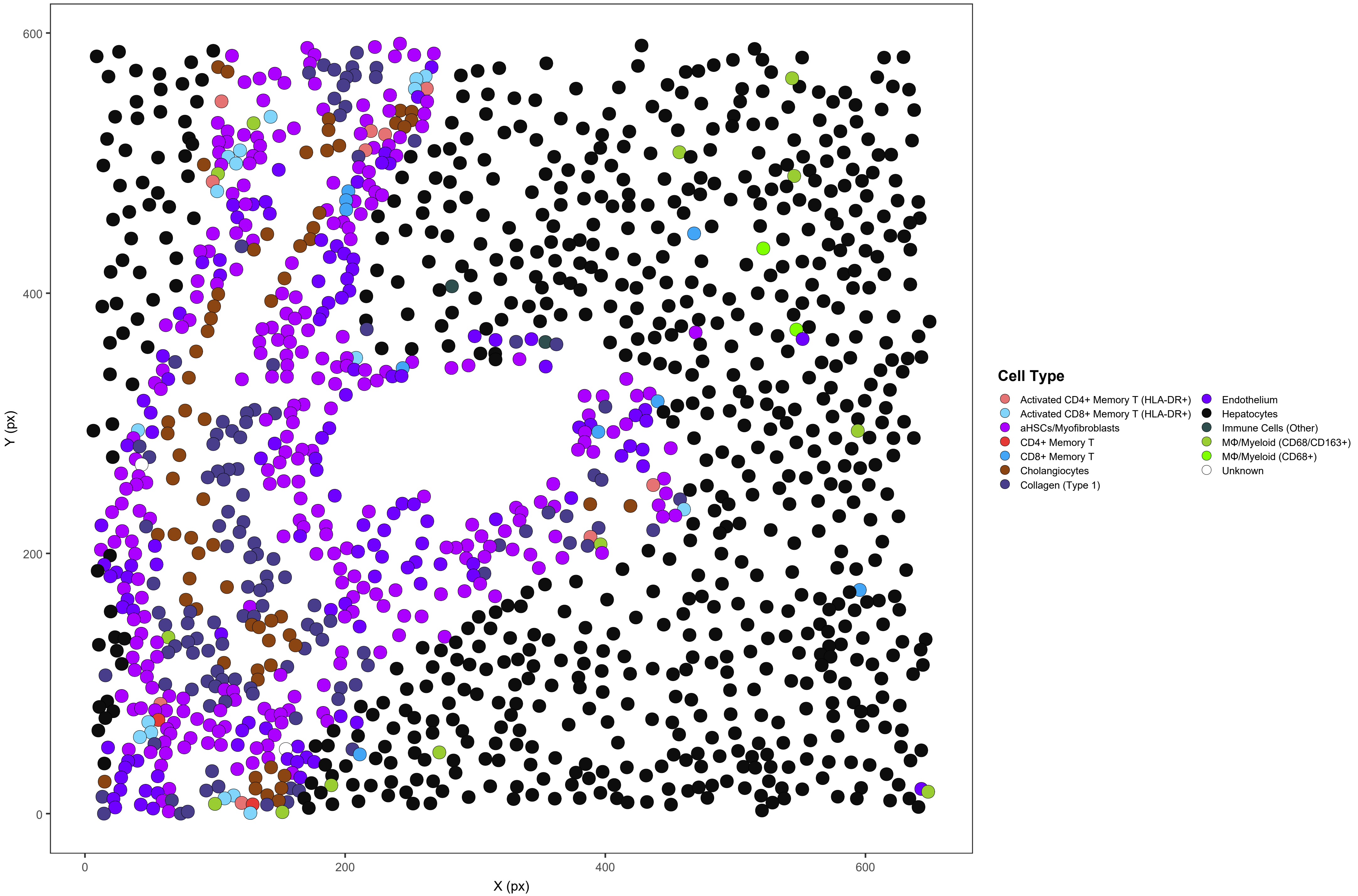

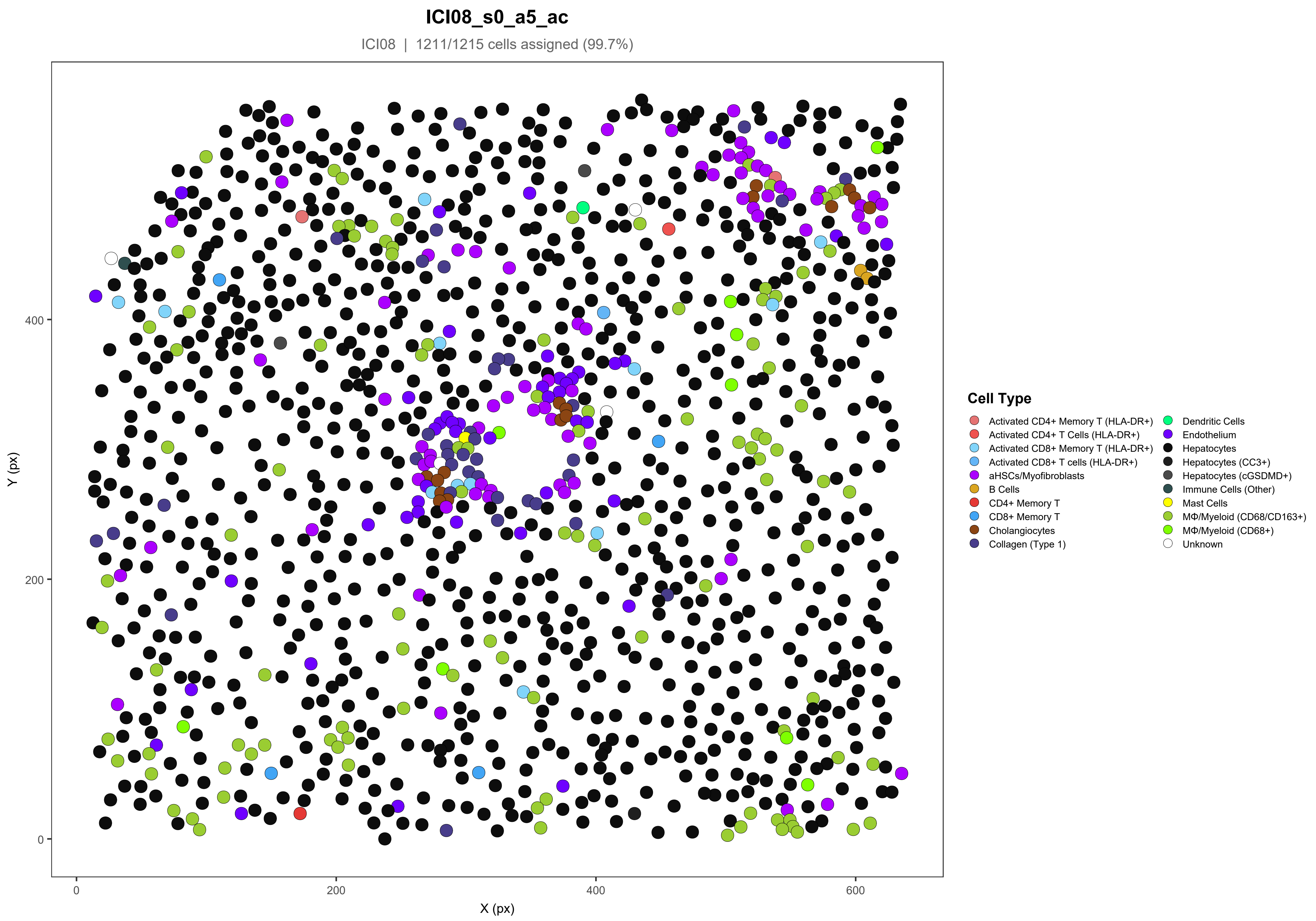
ICI08_s0_a5_ac
ICI08 | 1211/1215 cells assigned (99.7%)
Y (px)
X (px)
0
200
400
600
Cell Type
Activated CD4+ Memory T (HLA-DR+)
Activated CD4+ T Cells (HLA-DR+)
Activated CD8+ Memory T (HLA-DR+)
Activated CD8+ T cells (HLA-DR+)
aHSCs/Myofibroblasts
B Cells
CD4+ Memory T
CD8+ Memory T
Cholangiocytes
Collagen (Type 1)
Dendritic Cells
Endothelium
Hepatocytes
Hepatocytes (CC3+)
Hepatocytes (cGSDMD+)
Immune Cells (Other)
Mast Cells
MΦ/Myeloid (CD68/CD163+)
MΦ/Myeloid (CD68+)
Unknown

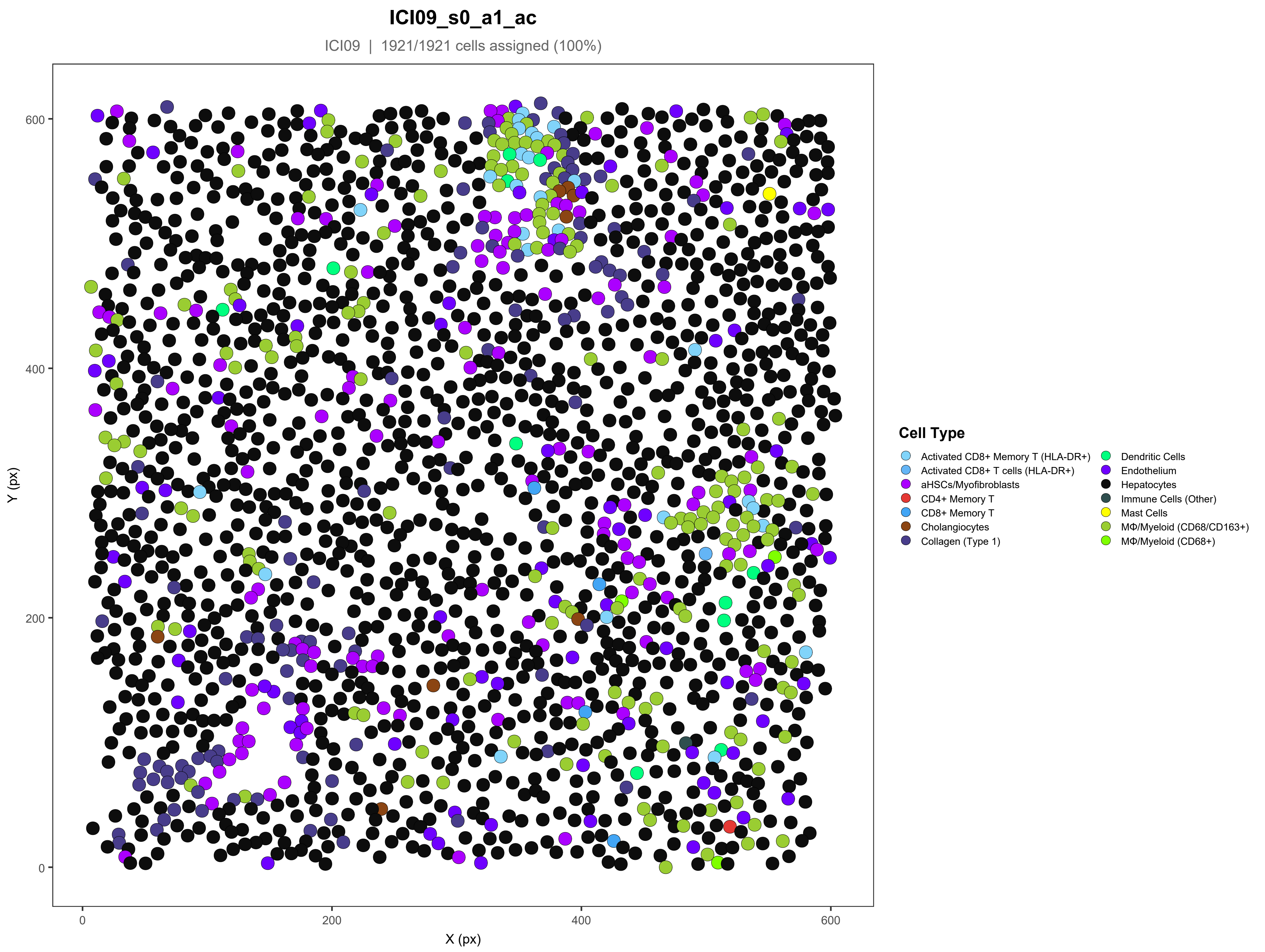
ICI09_s0_a1_ac
ICI09 | 1921/1921 cells assigned (100%)
Y (px)
X (px)
0
200
400
600
Cell Type
Activated CD8+ Memory T (HLA-DR+)
Activated CD8+ T cells (HLA-DR+)
aHSCs/Myofibroblasts
CD4+ Memory T
CD8+ Memory T
Cholangiocytes
Collagen (Type 1)
Dendritic Cells
Endothelium
Hepatocytes
Immune Cells (Other)
Mast Cells
MΦ/Myeloid (CD68/CD163+)
MΦ/Myeloid (CD68+)

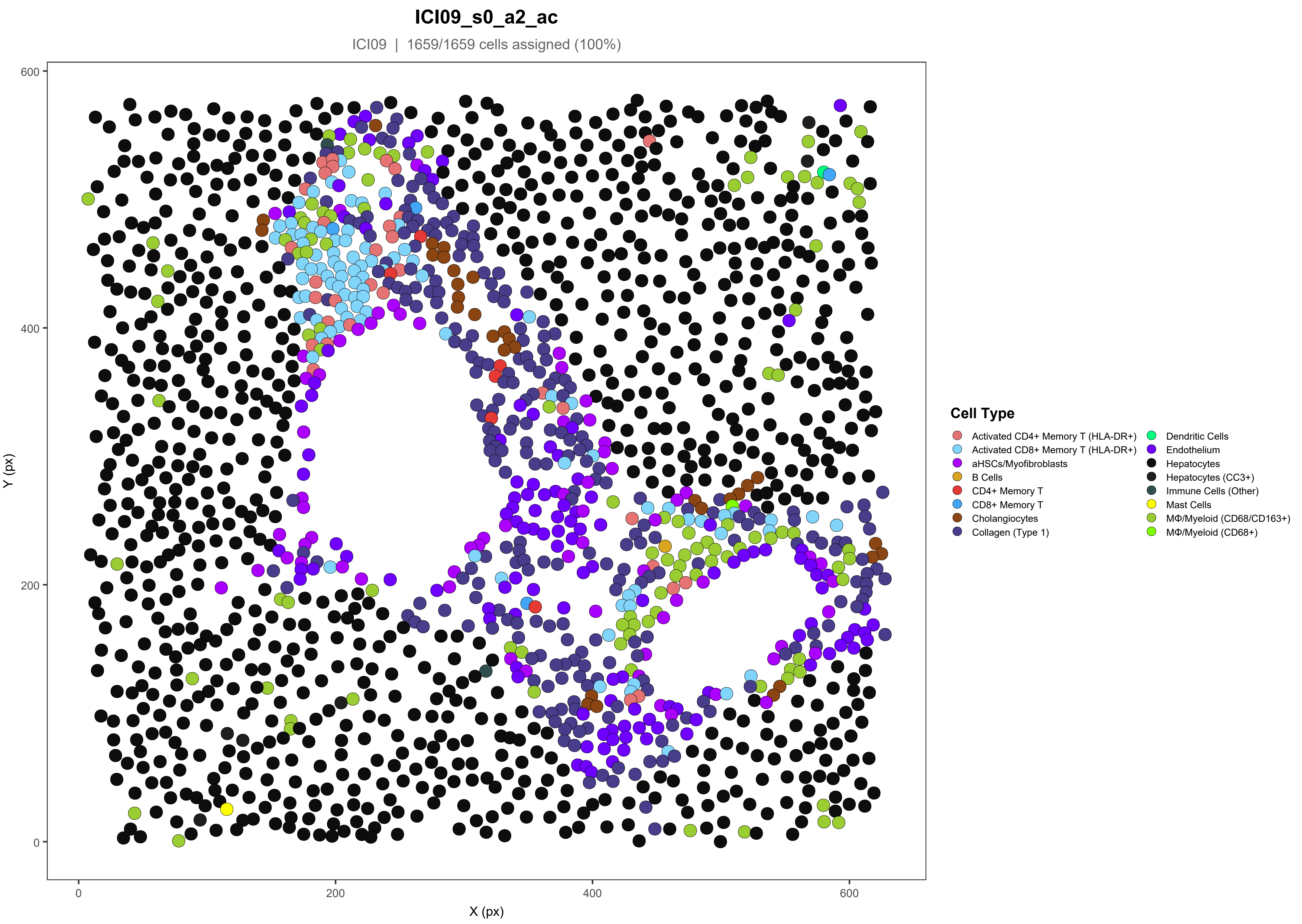

ICI09_s0_a2_ac
ICI09 | 1659/1659 cells assigned (100%)
Y (px)
X (px)
0
200
400
600
Cell Type
Activated CD4+ Memory T (HLA-DR+)
Activated CD8+ Memory T (HLA-DR+)
aHSCs/Myofibroblasts
B Cells
CD4+ Memory T
CD8+ Memory T
Cholangiocytes
Collagen (Type 1)
Dendritic Cells
Endothelium
Hepatocytes
Hepatocytes (CC3+)
Immune Cells (Other)
Mast Cells
MΦ/Myeloid (CD68/CD163+)
MΦ/Myeloid (CD68+)

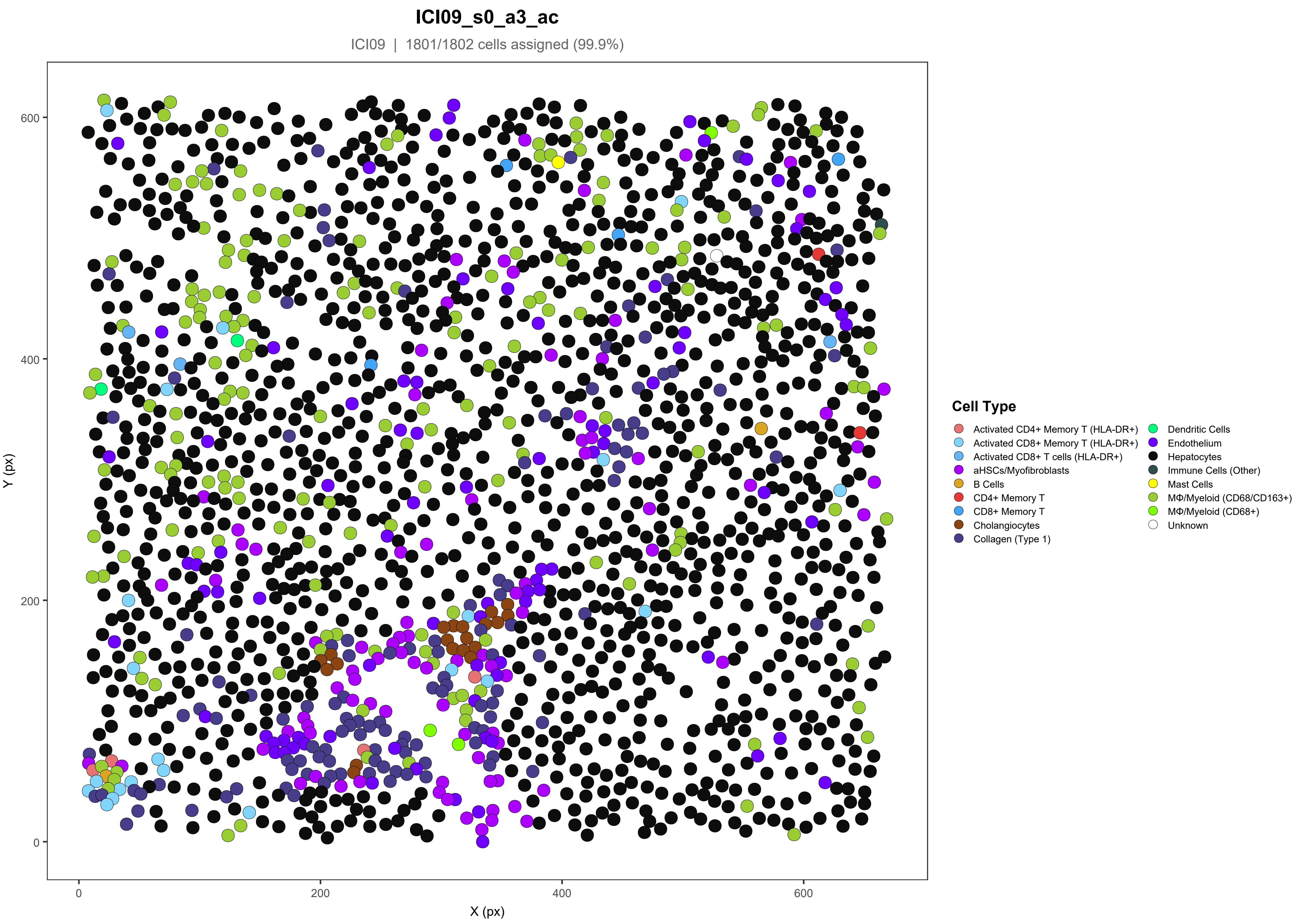

ICI09_s0_a3_ac
ICI09 | 1801/1802 cells assigned (99.9%)
Y (px)
X (px)
0
200
400
600
Cell Type
Activated CD4+ Memory T (HLA-DR+)
Activated CD8+ Memory T (HLA-DR+)
Activated CD8+ T cells (HLA-DR+)
aHSCs/Myofibroblasts
B Cells
CD4+ Memory T
CD8+ Memory T
Cholangiocytes
Collagen (Type 1)
Dendritic Cells
Endothelium
Hepatocytes
Immune Cells (Other)
Mast Cells
MΦ/Myeloid (CD68/CD163+)
MΦ/Myeloid (CD68+)
Unknown

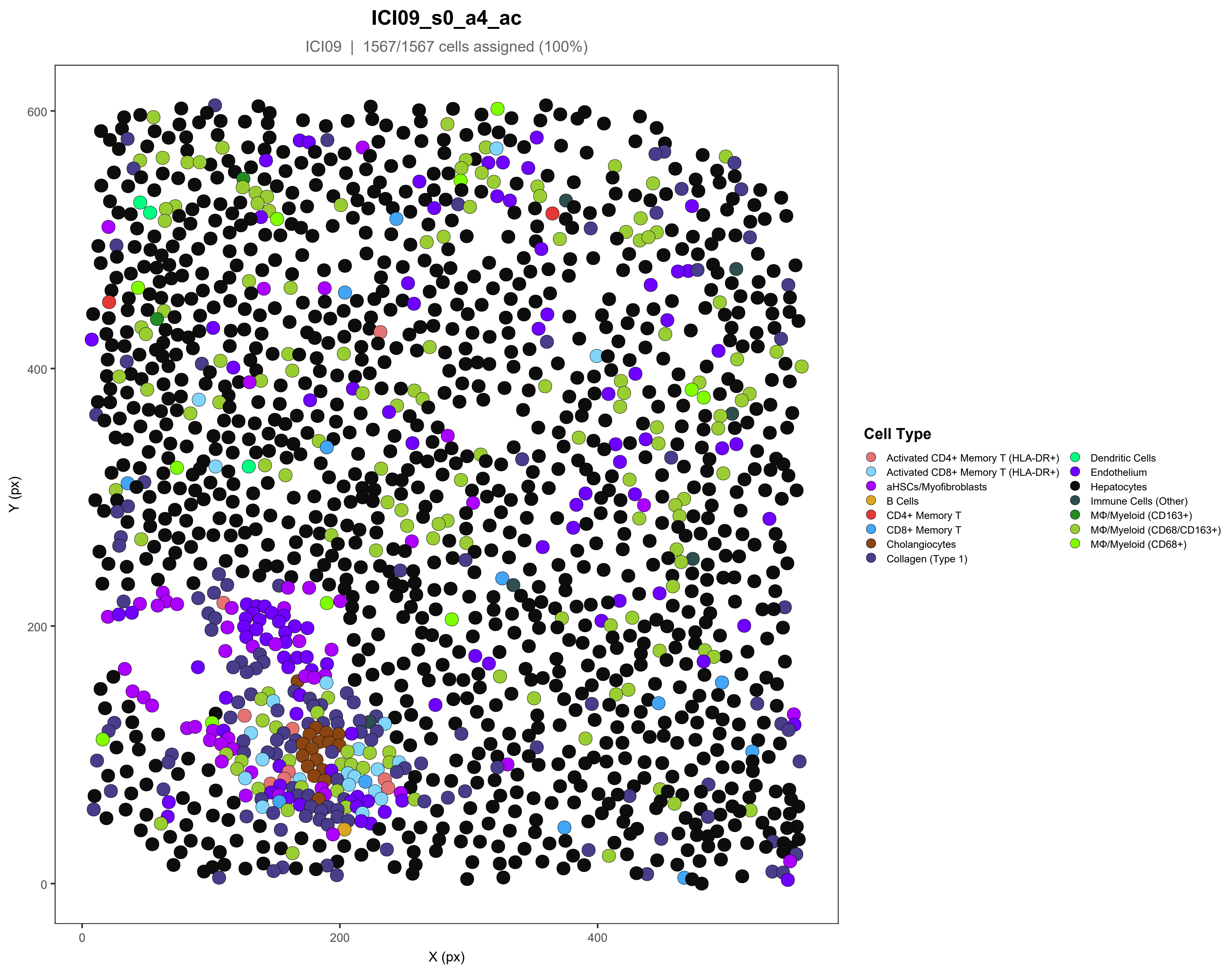

ICI09_s0_a4_ac
ICI09 | 1567/1567 cells assigned (100%)
Y (px)
X (px)
600
400
200
0
200
400
Cell Type
Activated CD4+ Memory T (HLA-DR+)
Activated CD8+ Memory T (HLA-DR+)
aHSCs/Myofibroblasts
B Cells
CD4+ Memory T
CD8+ Memory T
Cholangiocytes
Collagen (Type 1)
Dendritic Cells
Endothelium
Hepatocytes
Immune Cells (Other)
MΦ/Myeloid (CD163+)
MΦ/Myeloid (CD68/CD163+)
MΦ/Myeloid (CD68+)

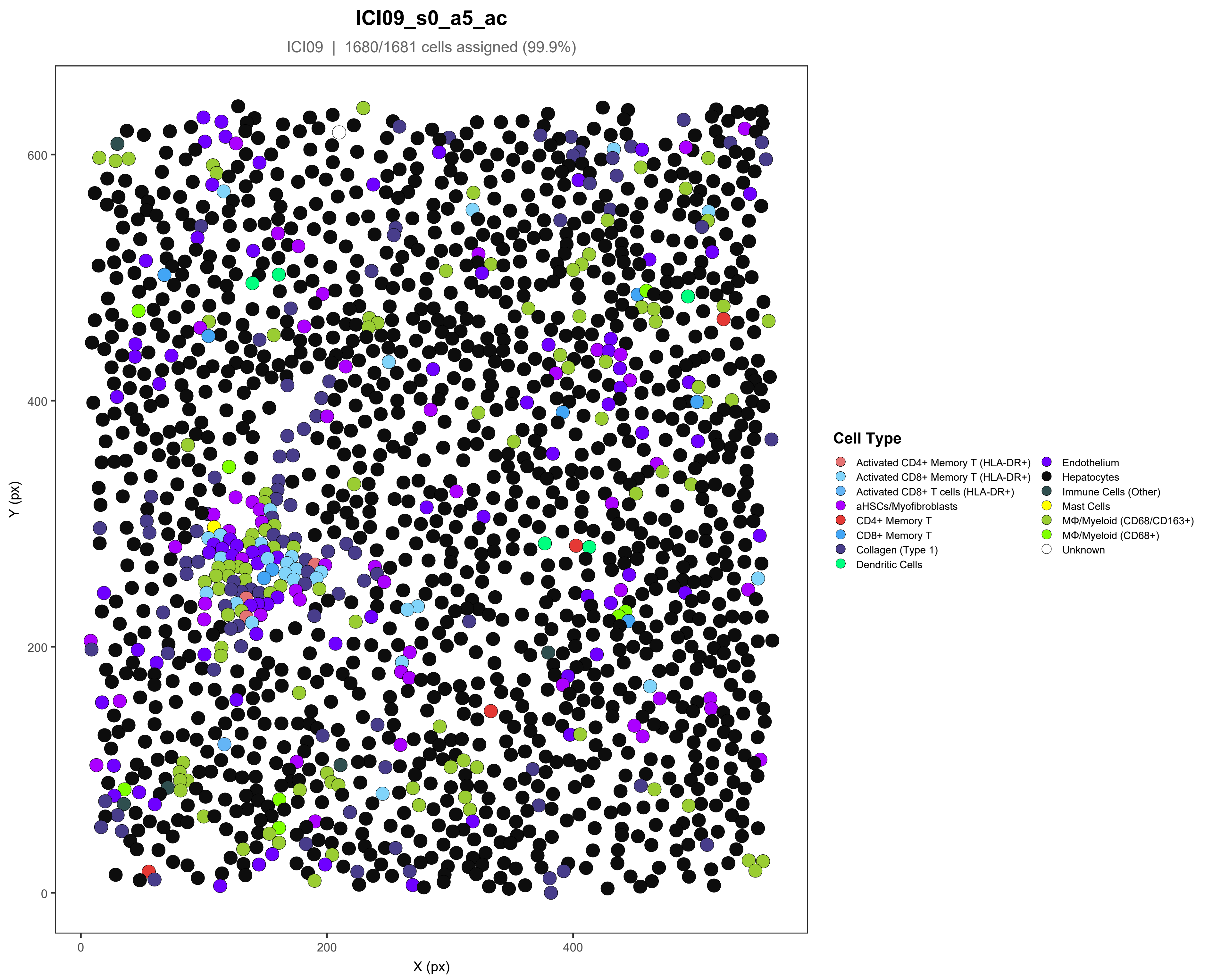

ICI09_s0_a5_ac
ICI09 | 1680/1681 cells assigned (99.9%)
Y (px)
X (px)
0
200
400
600
0
200
400
Cell Type
Activated CD4+ Memory T (HLA-DR+)
Activated CD8+ Memory T (HLA-DR+)
Activated CD8+ T cells (HLA-DR+)
aHSCs/Myofibroblasts
CD4+ Memory T
CD8+ Memory T
Collagen (Type 1)
Dendritic Cells
Endothelium
Hepatocytes
Immune Cells (Other)
Mast Cells
MΦ/Myeloid (CD68/CD163+)
MΦ/Myeloid (CD68+)
Unknown

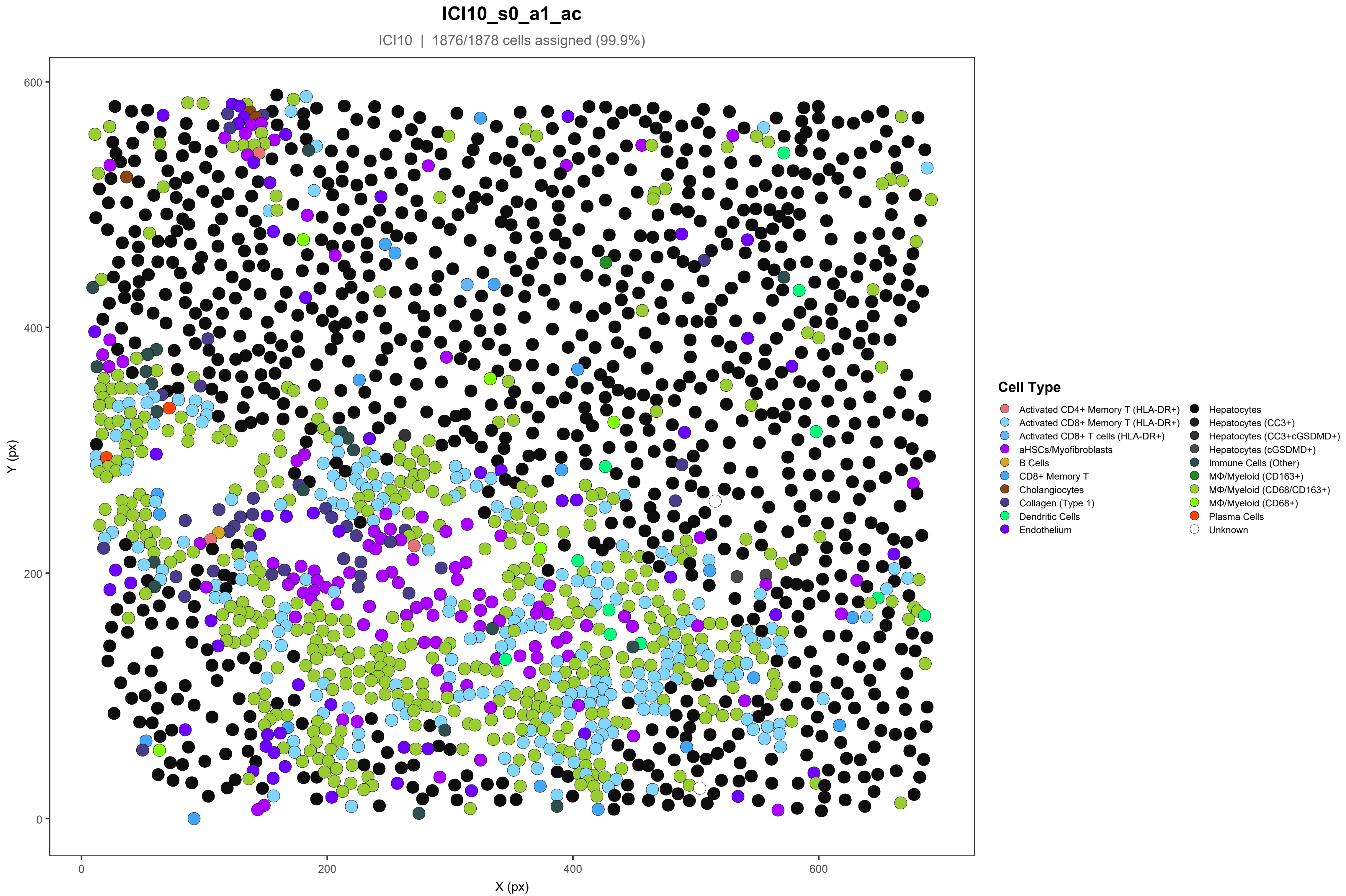
ICI10_s0_a1_ac
ICI10 | 1876/1878 cells assigned (99.9%)
Y (px)
X (px)
0
200
400
600
Cell Type
Activated CD4+ Memory T (HLA-DR+)
Activated CD8+ Memory T (HLA-DR+)
Activated CD8+ T cells (HLA-DR+)
aHSCs/Myofibroblasts
B Cells
CD8+ Memory T
Cholangiocytes
Collagen (Type 1)
Dendritic Cells
Endothelium
Hepatocytes
Hepatocytes (CC3+)
Hepatocytes (CC3+cGSDMD+)
Hepatocytes (cGSDMD+)
Immune Cells (Other)
MΦ/Myeloid (CD163+)
MΦ/Myeloid (CD68/CD163+)
MΦ/Myeloid (CD68+)
Plasma Cells
Unknown

### ICI10_s0_a2_ac

ICI10 | 1579/1586 cells assigned (99.6%)

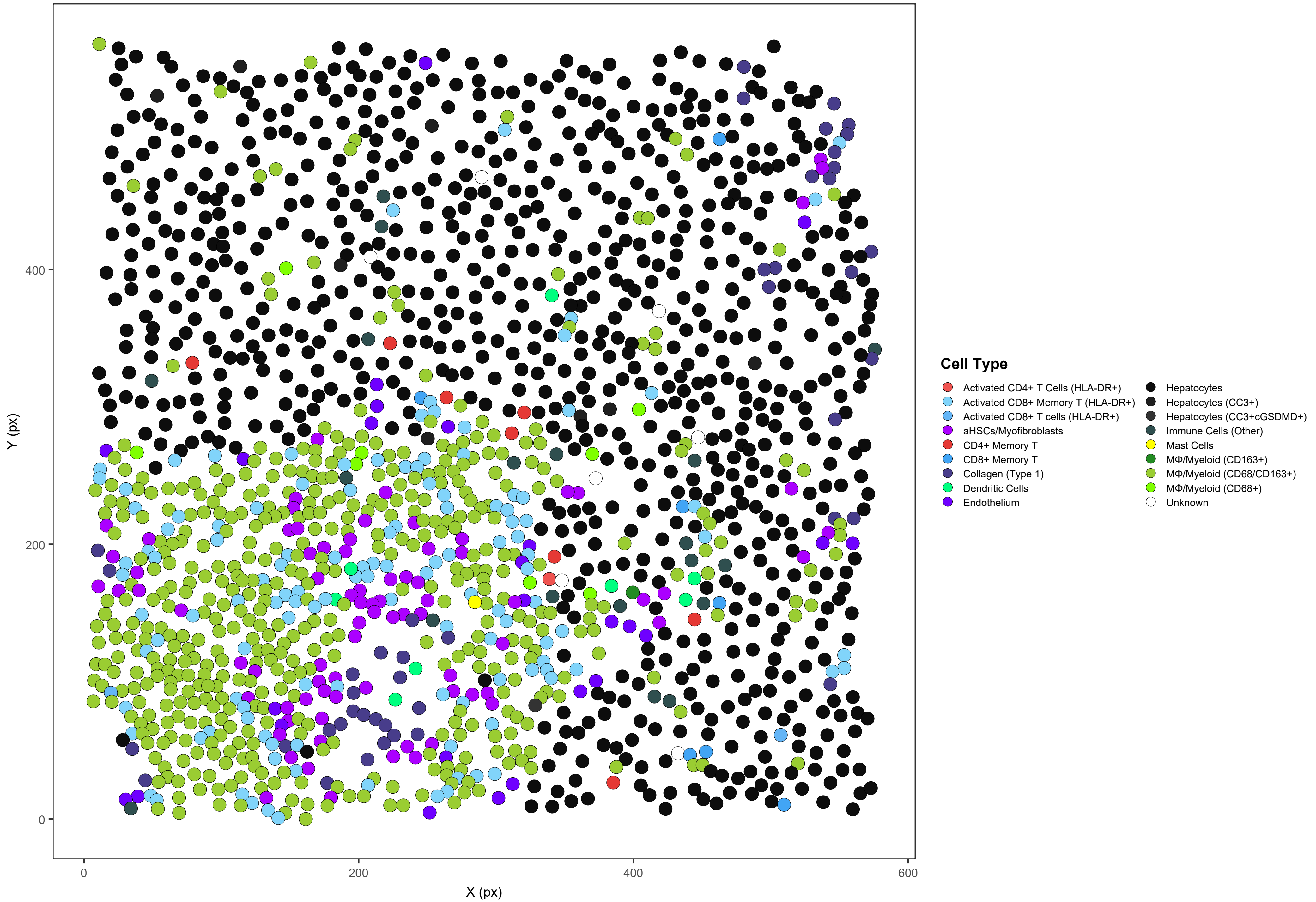

**ICI10_s0_a3_ac**

ICI10 | 1645/1657 cells assigned (99.3%)

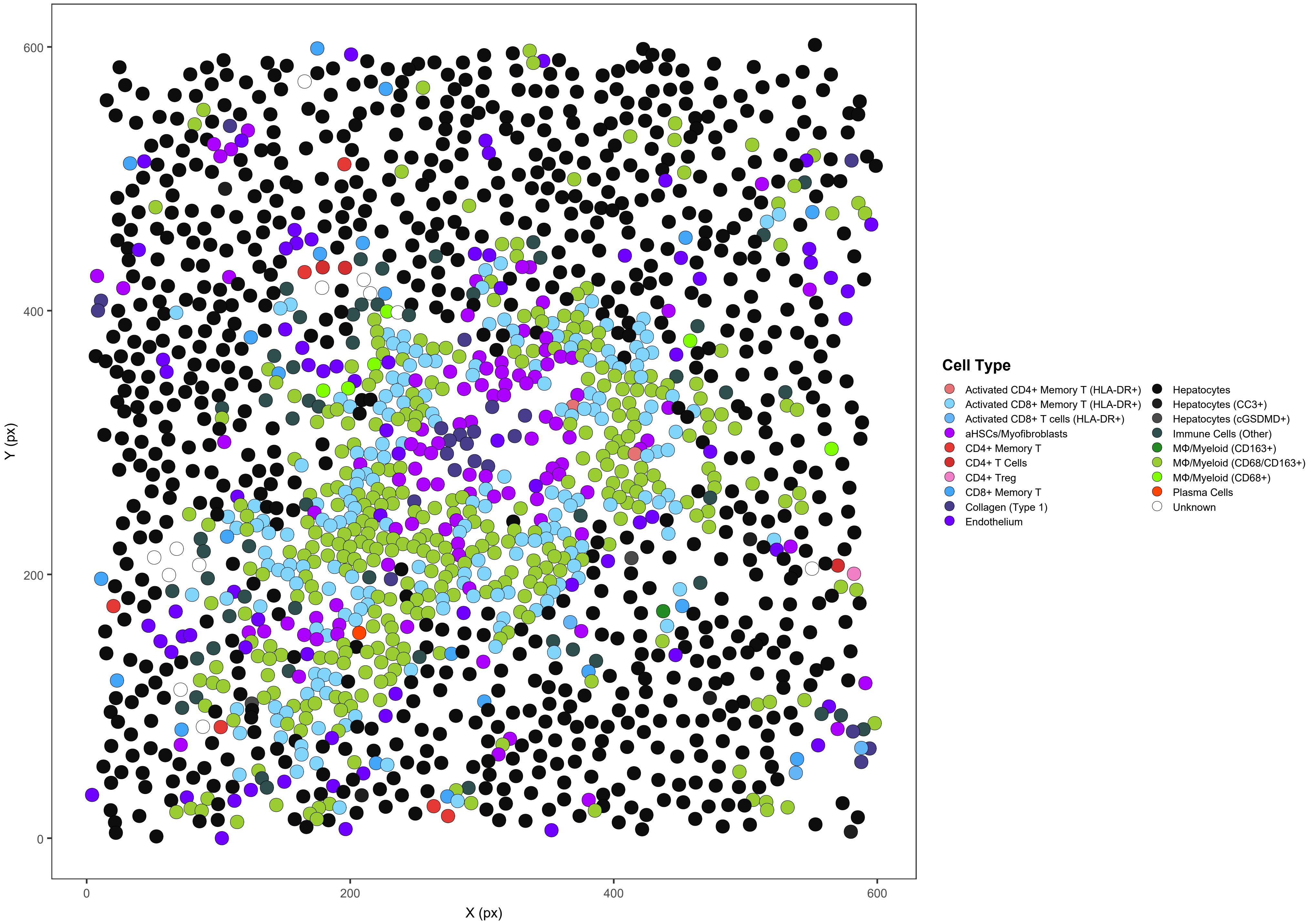

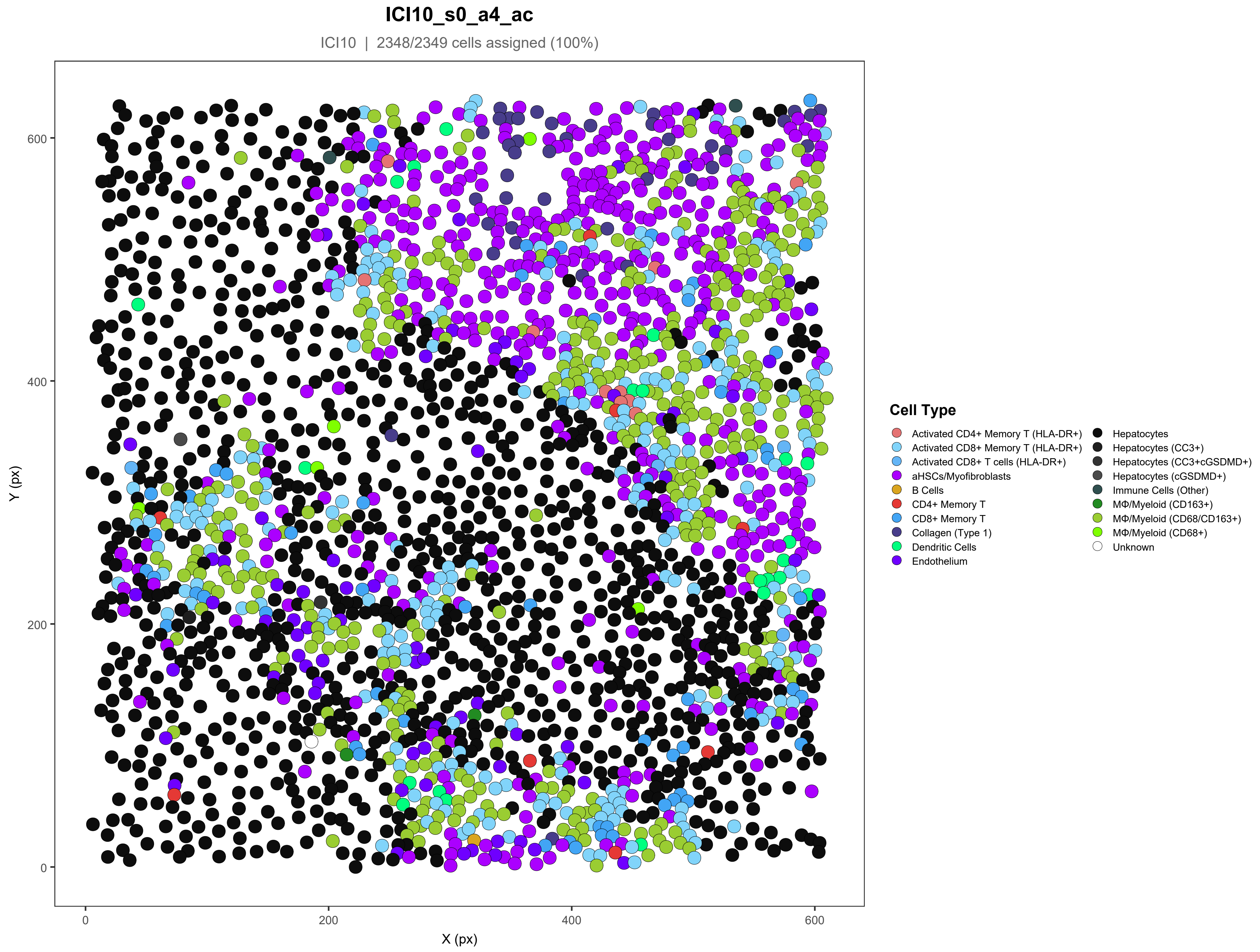
ICI10_s0_a4_ac
ICI10 | 2348/2349 cells assigned (100%)
Y (px)
X (px)
0
200
400
600
Cell Type
Activated CD4+ Memory T (HLA-DR+)
Activated CD8+ Memory T (HLA-DR+)
Activated CD8+ T cells (HLA-DR+)
aHSCs/Myofibroblasts
B Cells
CD4+ Memory T
CD8+ Memory T
Collagen (Type 1)
Dendritic Cells
Endothelium
Hepatocytes
Hepatocytes (CC3+)
Hepatocytes (CC3+cGSDMD+)
Hepatocytes (cGSDMD+)
Immune Cells (Other)
MΦ/Myeloid (CD163+)
MΦ/Myeloid (CD68/CD163+)
MΦ/Myeloid (CD68+)
Unknown

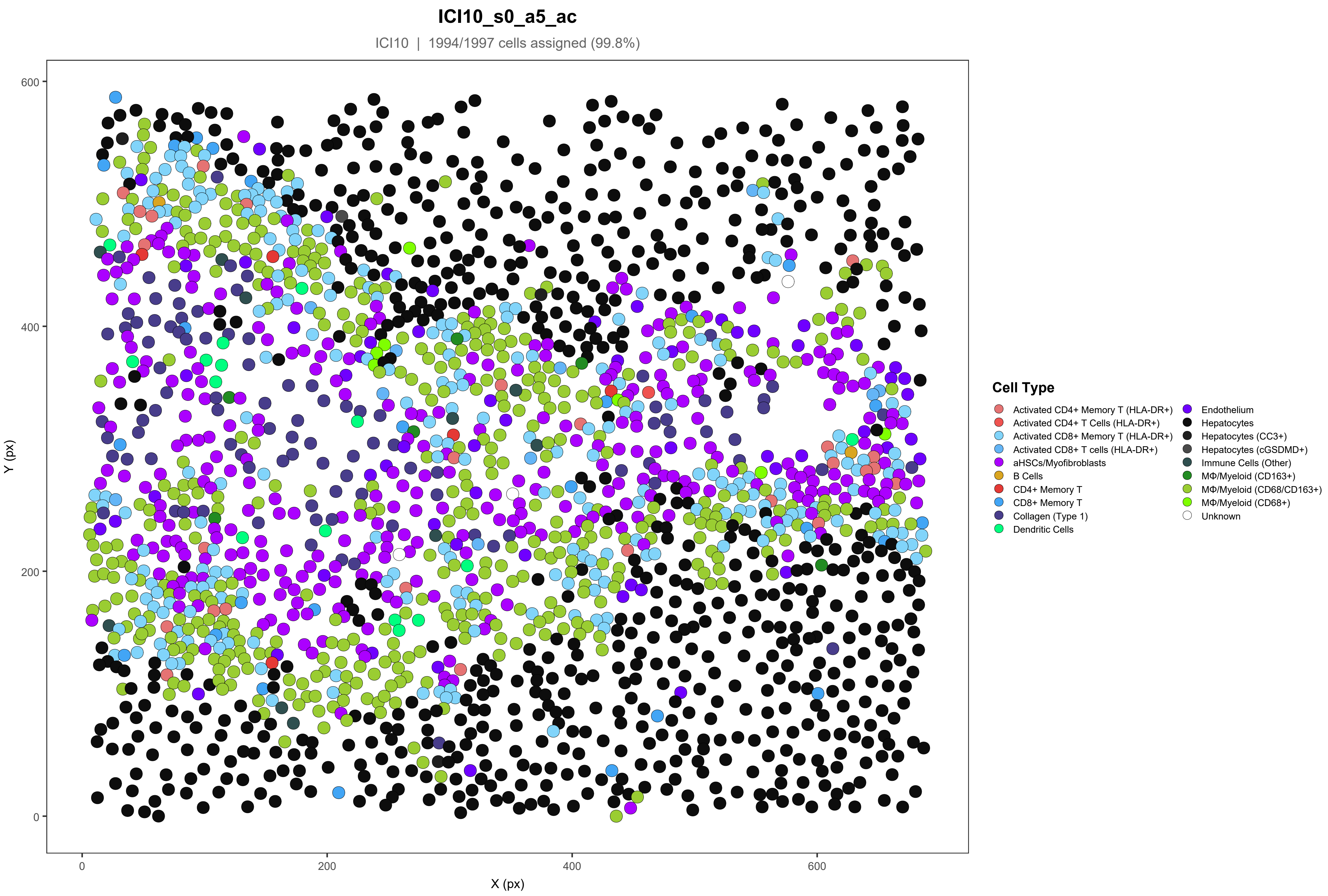
ICI10_s0_a5_ac
ICI10 | 1994/1997 cells assigned (99.8%)
Y (px)
X (px)
0
200
400
600
Cell Type
Activated CD4+ Memory T (HLA-DR+)
Activated CD4+ T Cells (HLA-DR+)
Activated CD8+ Memory T (HLA-DR+)
Activated CD8+ T cells (HLA-DR+)
aHSCs/Myofibroblasts
B Cells
CD4+ Memory T
CD8+ Memory T
Collagen (Type 1)
Dendritic Cells
Endothelium
Hepatocytes
Hepatocytes (CC3+)
Hepatocytes (cGSDMD+)
Immune Cells (Other)
MΦ/Myeloid (CD163+)
MΦ/Myeloid (CD68/CD163+)
MΦ/Myeloid (CD68+)
Unknown

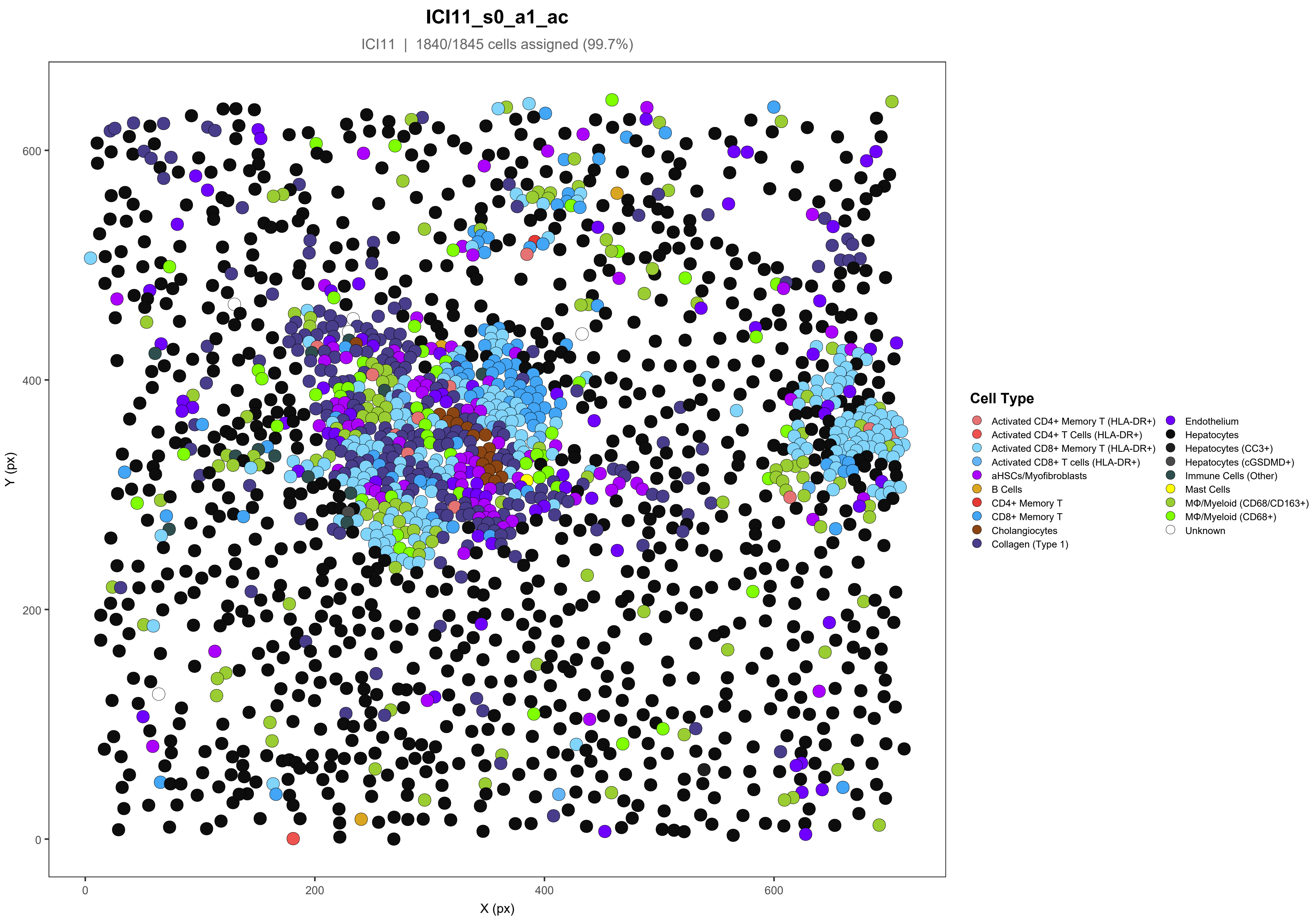
ICI11_s0_a1_ac
ICI11 | 1840/1845 cells assigned (99.7%)
Y (px)
X (px)
0
200
400
600
Cell Type
Activated CD4+ Memory T (HLA-DR+)
Activated CD4+ T Cells (HLA-DR+)
Activated CD8+ Memory T (HLA-DR+)
Activated CD8+ T cells (HLA-DR+)
aHSCs/Myofibroblasts
B Cells
CD4+ Memory T
CD8+ Memory T
Cholangiocytes
Collagen (Type 1)
Endothelium
Hepatocytes
Hepatocytes (CC3+)
Hepatocytes (cGSDMD+)
Immune Cells (Other)
Mast Cells
MΦ/Myeloid (CD68/CD163+)
MΦ/Myeloid (CD68+)
Unknown

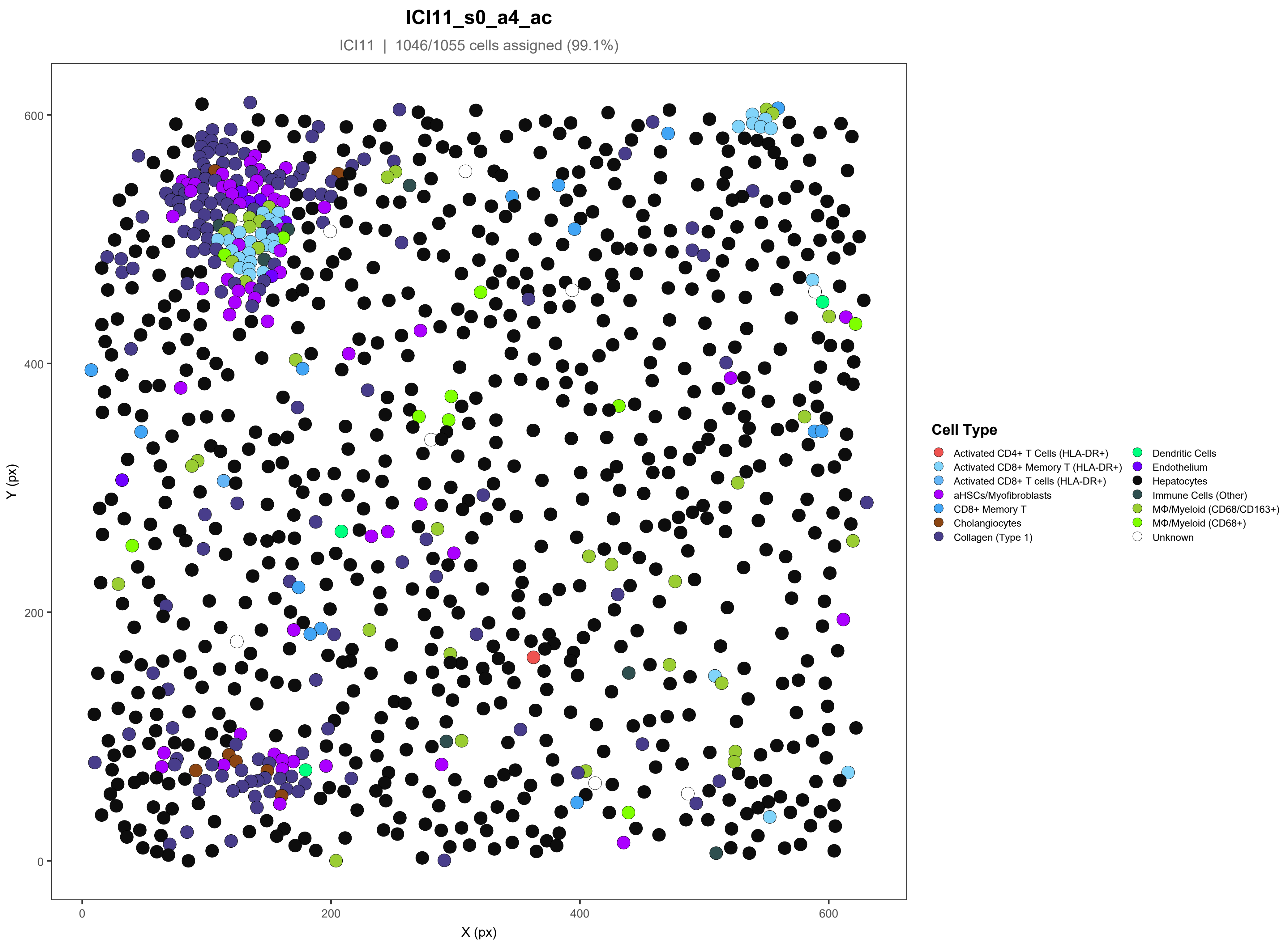
ICI11_s0_a4_ac
ICI11 | 1046/1055 cells assigned (99.1%)
Y (px)
X (px)
0
200
400
600
Cell Type
Activated CD4+ T Cells (HLA-DR+)
Activated CD8+ Memory T (HLA-DR+)
Activated CD8+ T cells (HLA-DR+)
aHSCs/Myofibroblasts
CD8+ Memory T
Cholangiocytes
Collagen (Type 1)
Dendritic Cells
Endothelium
Hepatocytes
Immune Cells (Other)
MΦ/Myeloid (CD68/CD163+)
MΦ/Myeloid (CD68+)
Unknown

#### ICI12_s0_a1_ac

ICI12 | 1177/1229 cells assigned (95.8%)

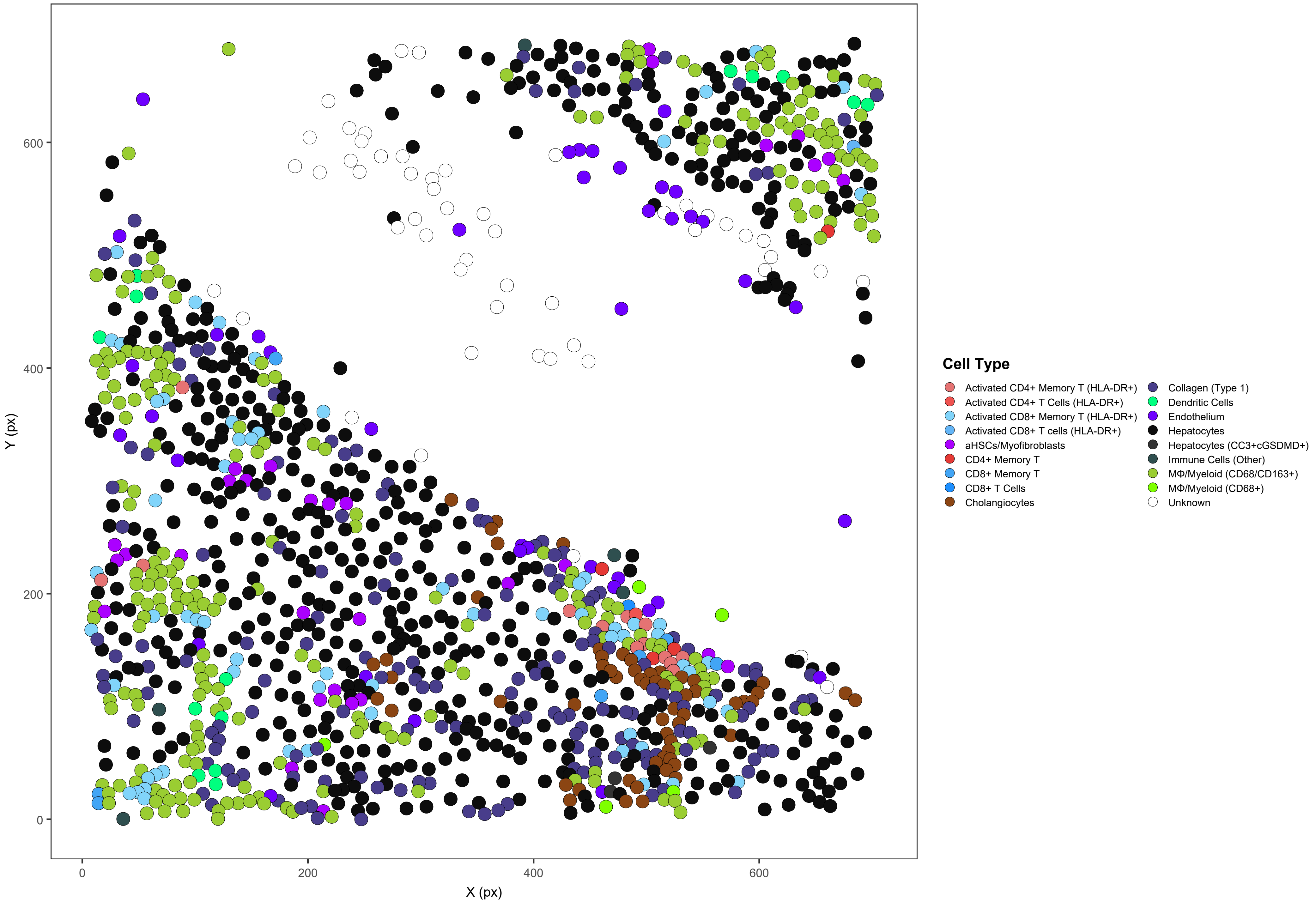

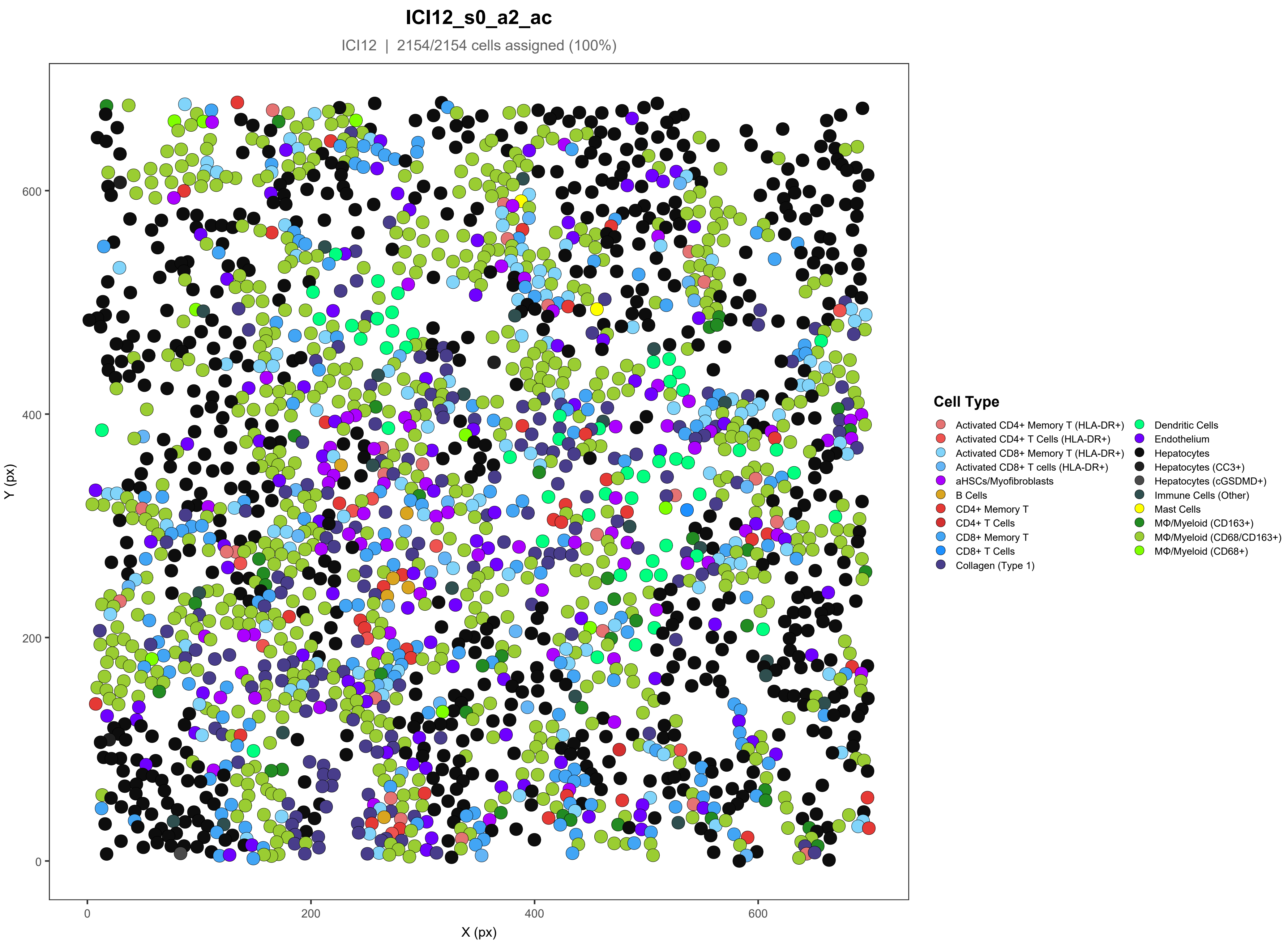
ICI12_s0_a2_ac
ICI12 | 2154/2154 cells assigned (100%)
Y (px)
X (px)
0
200
400
600
Cell Type
Activated CD4+ Memory T (HLA-DR+)
Activated CD4+ T Cells (HLA-DR+)
Activated CD8+ Memory T (HLA-DR+)
Activated CD8+ T cells (HLA-DR+)
aHSCs/Myofibroblasts
B Cells
CD4+ Memory T
CD4+ T Cells
CD8+ Memory T
CD8+ T Cells
Collagen (Type 1)
Dendritic Cells
Endothelium
Hepatocytes
Hepatocytes (CC3+)
Hepatocytes (cGSDMD+)
Immune Cells (Other)
Mast Cells
MΦ/Myeloid (CD163+)
MΦ/Myeloid (CD68/CD163+)
MΦ/Myeloid (CD68+)

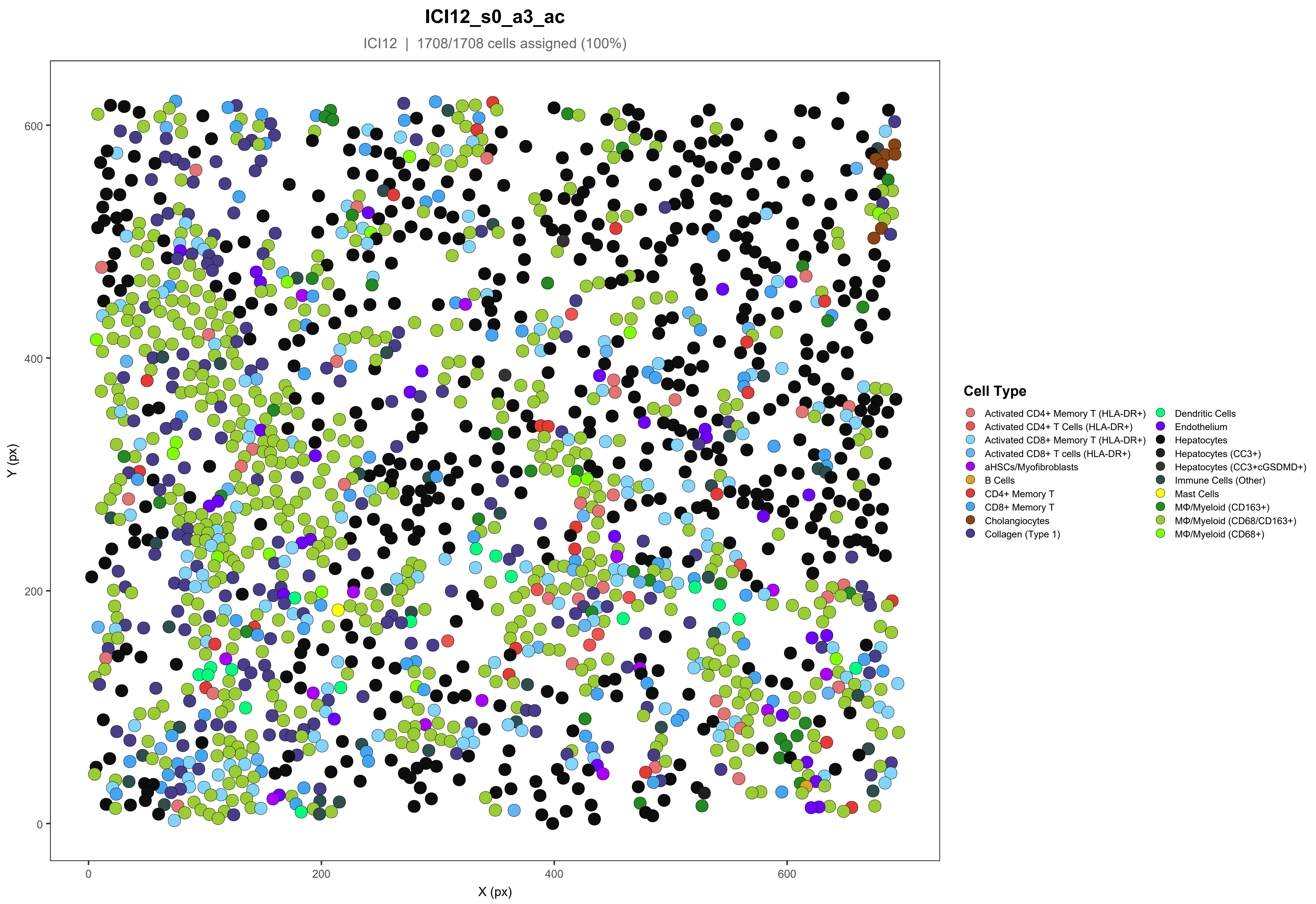
ICI12_s0_a3_ac
ICI12 | 1708/1708 cells assigned (100%)
600
400
200
0
Y (px)
0
200
400
600
X (px)
Cell Type
Activated CD4+ Memory T (HLA-DR+)
Activated CD4+ T Cells (HLA-DR+)
Activated CD8+ Memory T (HLA-DR+)
Activated CD8+ T cells (HLA-DR+)
aHSCs/Myofibroblasts
B Cells
CD4+ Memory T
CD8+ Memory T
Cholangiocytes
Collagen (Type 1)
Dendritic Cells
Endothelium
Hepatocytes
Hepatocytes (CC3+)
Hepatocytes (CC3+cGSDMD+)
Immune Cells (Other)
Mast Cells
MΦ/Myeloid (CD163+)
MΦ/Myeloid (CD68/CD163+)
MΦ/Myeloid (CD68+)

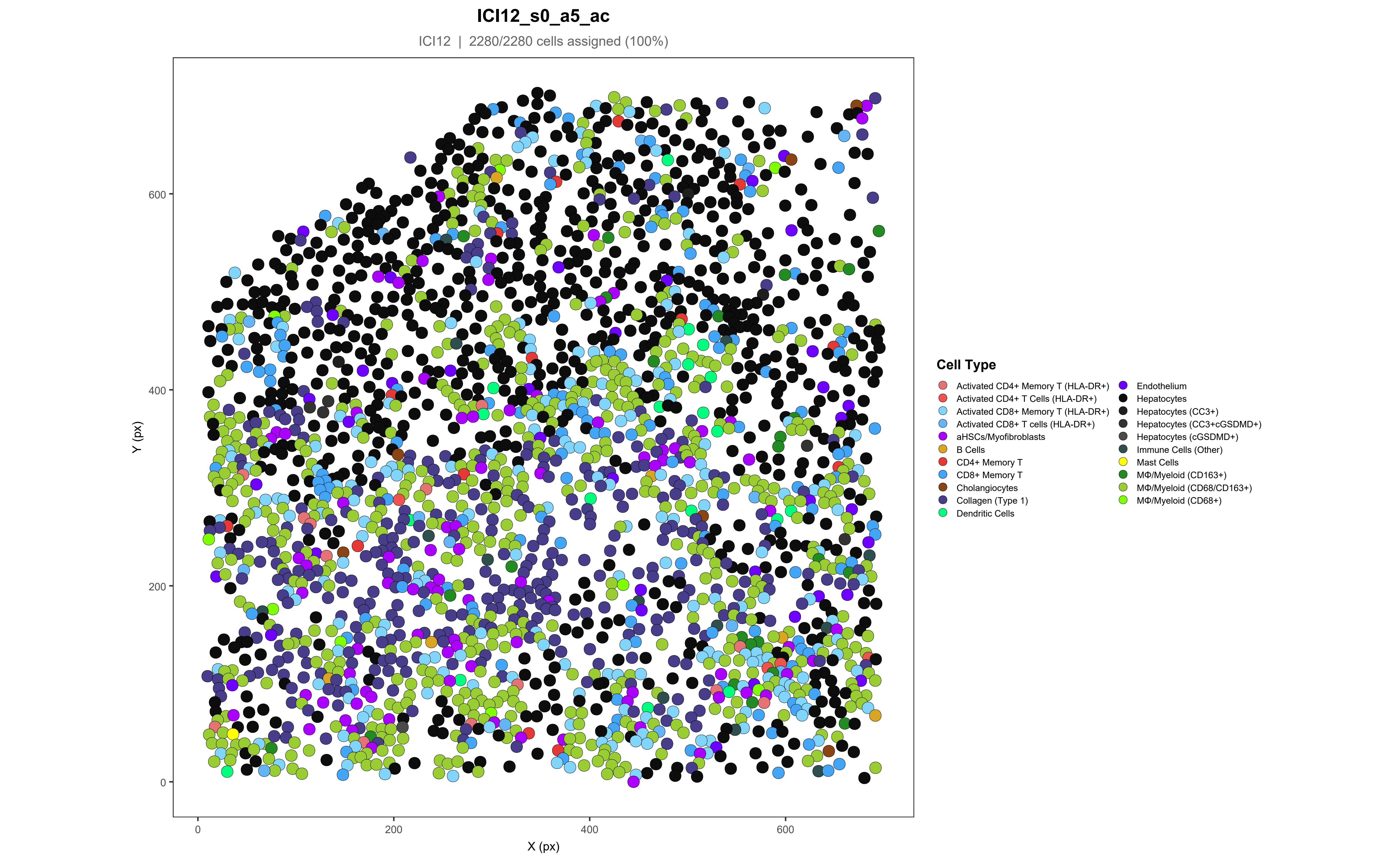

ICI12_s0_a5_ac
ICI12 | 2280/2280 cells assigned (100%)
Y (px)
X (px)
0
200
400
600
Cell Type
Activated CD4+ Memory T (HLA-DR+)
Activated CD4+ T Cells (HLA-DR+)
Activated CD8+ Memory T (HLA-DR+)
Activated CD8+ T cells (HLA-DR+)
aHSCs/Myofibroblasts
B Cells
CD4+ Memory T
CD8+ Memory T
Cholangiocytes
Collagen (Type 1)
Dendritic Cells
Endothelium
Hepatocytes
Hepatocytes (CC3+)
Hepatocytes (CC3+cGSDMD+)
Hepatocytes (cGSDMD+)
Immune Cells (Other)
Mast Cells
MΦ/Myeloid (CD163+)
MΦ/Myeloid (CD68/CD163+)
MΦ/Myeloid (CD68+)

### ICI13_s0_a1_ac

ICI13 | 1955/1956 cells assigned (99.9%)

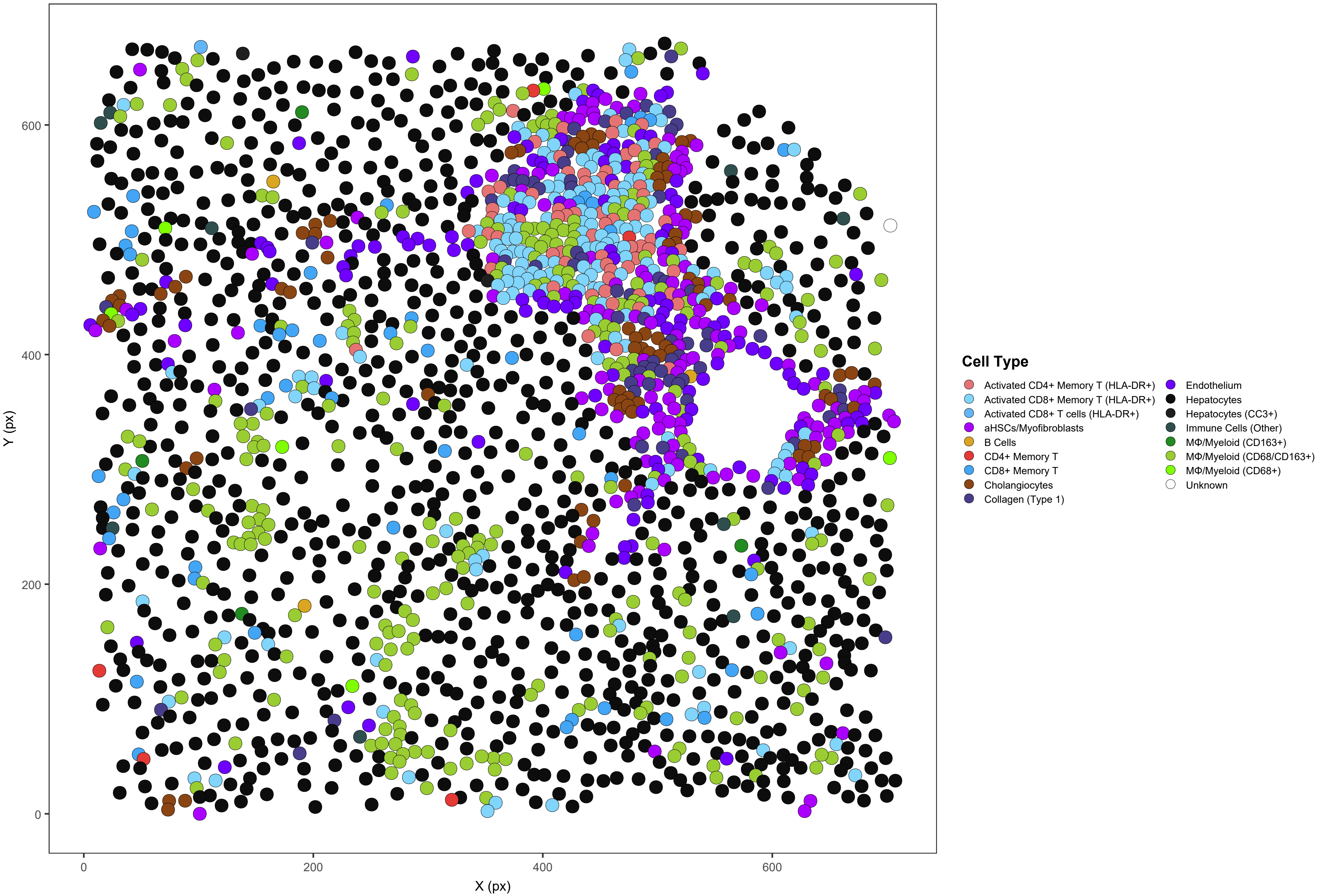

#### ICI13_s0_a2_ac

ICI13 | 1762/1771 cells assigned (99.5%)

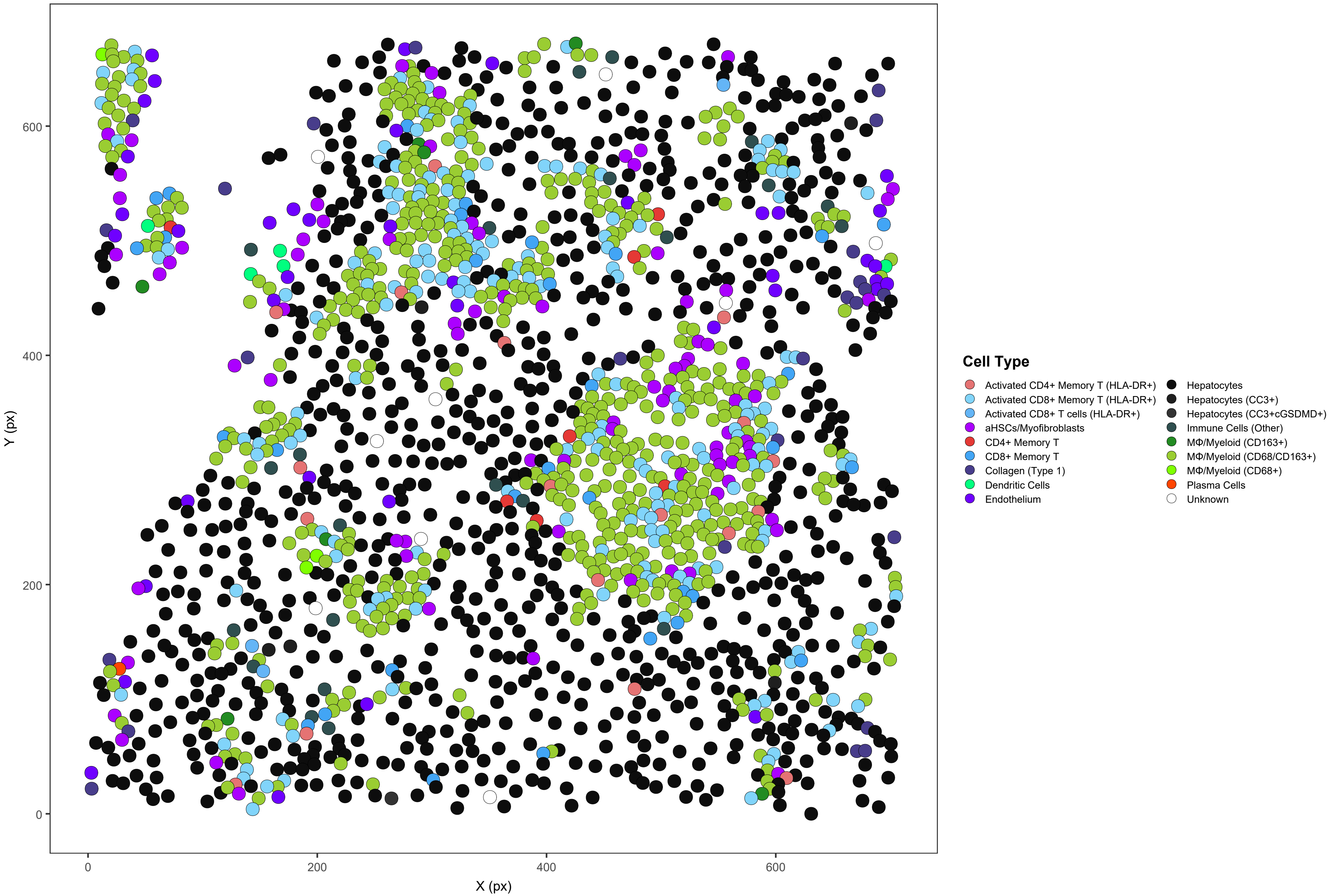

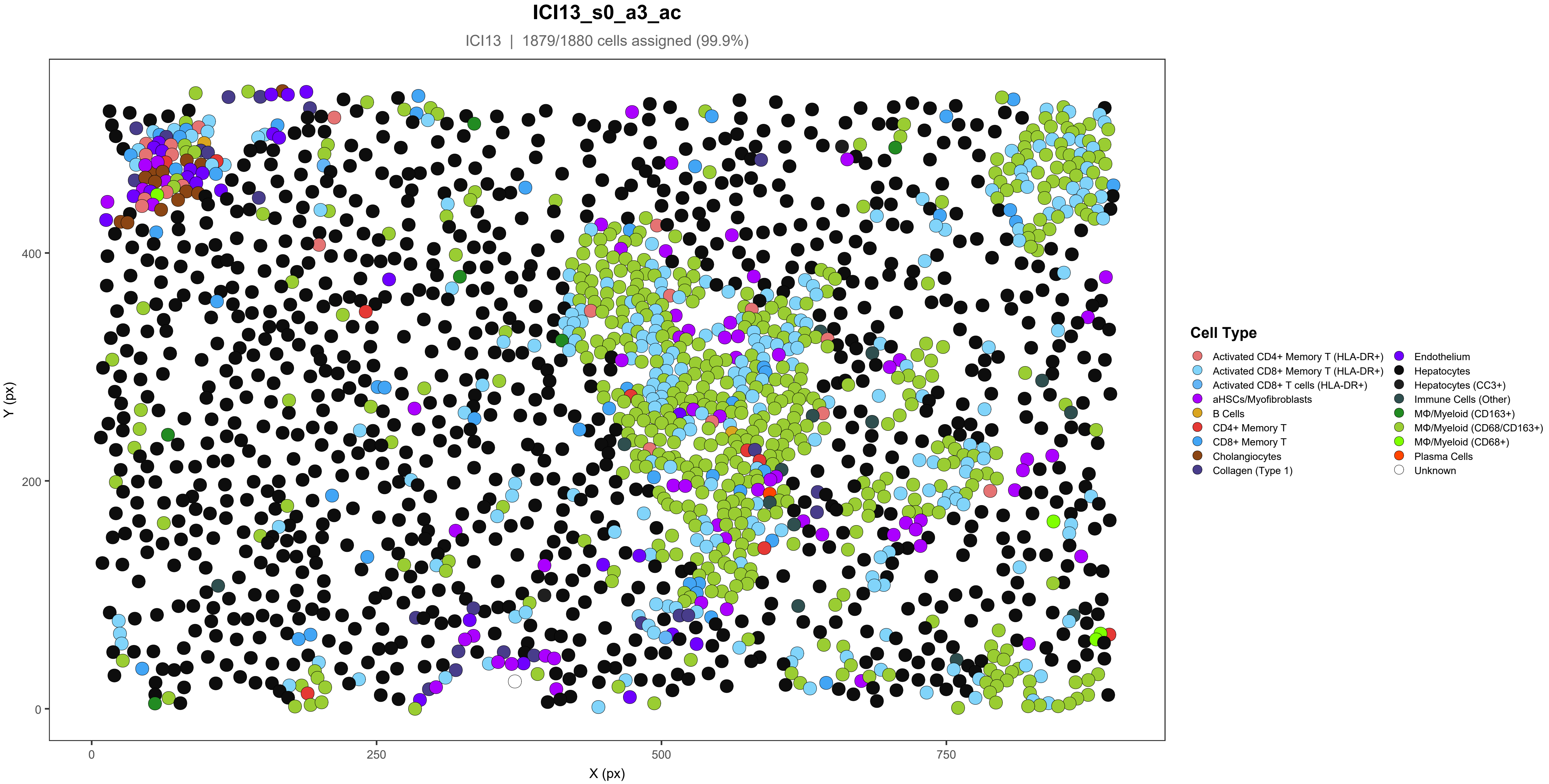
ICI13_s0_a3_ac
ICI13 | 1879/1880 cells assigned (99.9%)
Y (px)
X (px)
0
200
400
0
250
500
750
Cell Type
Activated CD4+ Memory T (HLA-DR+)
Activated CD8+ Memory T (HLA-DR+)
Activated CD8+ T cells (HLA-DR+)
aHSCs/Myofibroblasts
B Cells
CD4+ Memory T
CD8+ Memory T
Cholangiocytes
Collagen (Type 1)
Endothelium
Hepatocytes
Hepatocytes (CC3+)
Immune Cells (Other)
MΦ/Myeloid (CD163+)
MΦ/Myeloid (CD68/CD163+)
MΦ/Myeloid (CD68+)
Plasma Cells
Unknown

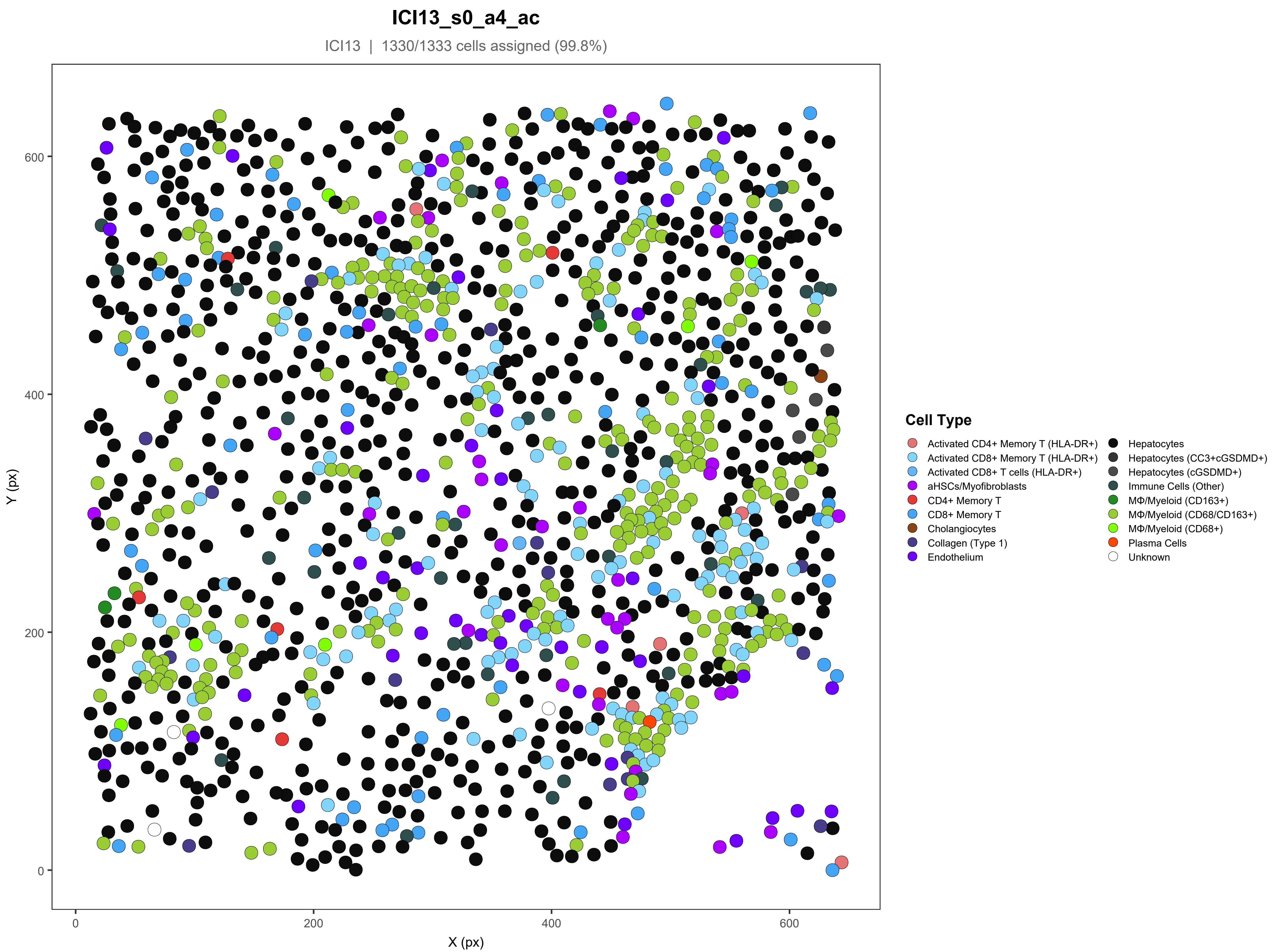

ICI13_s0_a4_ac
ICI13 | 1330/1333 cells assigned (99.8%)
Y (px)
X (px)
0
200
400
600
Cell Type
Activated CD4+ Memory T (HLA-DR+)
Activated CD8+ Memory T (HLA-DR+)
Activated CD8+ T cells (HLA-DR+)
aHSCs/Myofibroblasts
CD4+ Memory T
CD8+ Memory T
Cholangiocytes
Collagen (Type 1)
Endothelium
Hepatocytes
Hepatocytes (CC3+cGSDMD+)
Hepatocytes (cGSDMD+)
Immune Cells (Other)
MΦ/Myeloid (CD163+)
MΦ/Myeloid (CD68/CD163+)
MΦ/Myeloid (CD68+)
Plasma Cells
Unknown

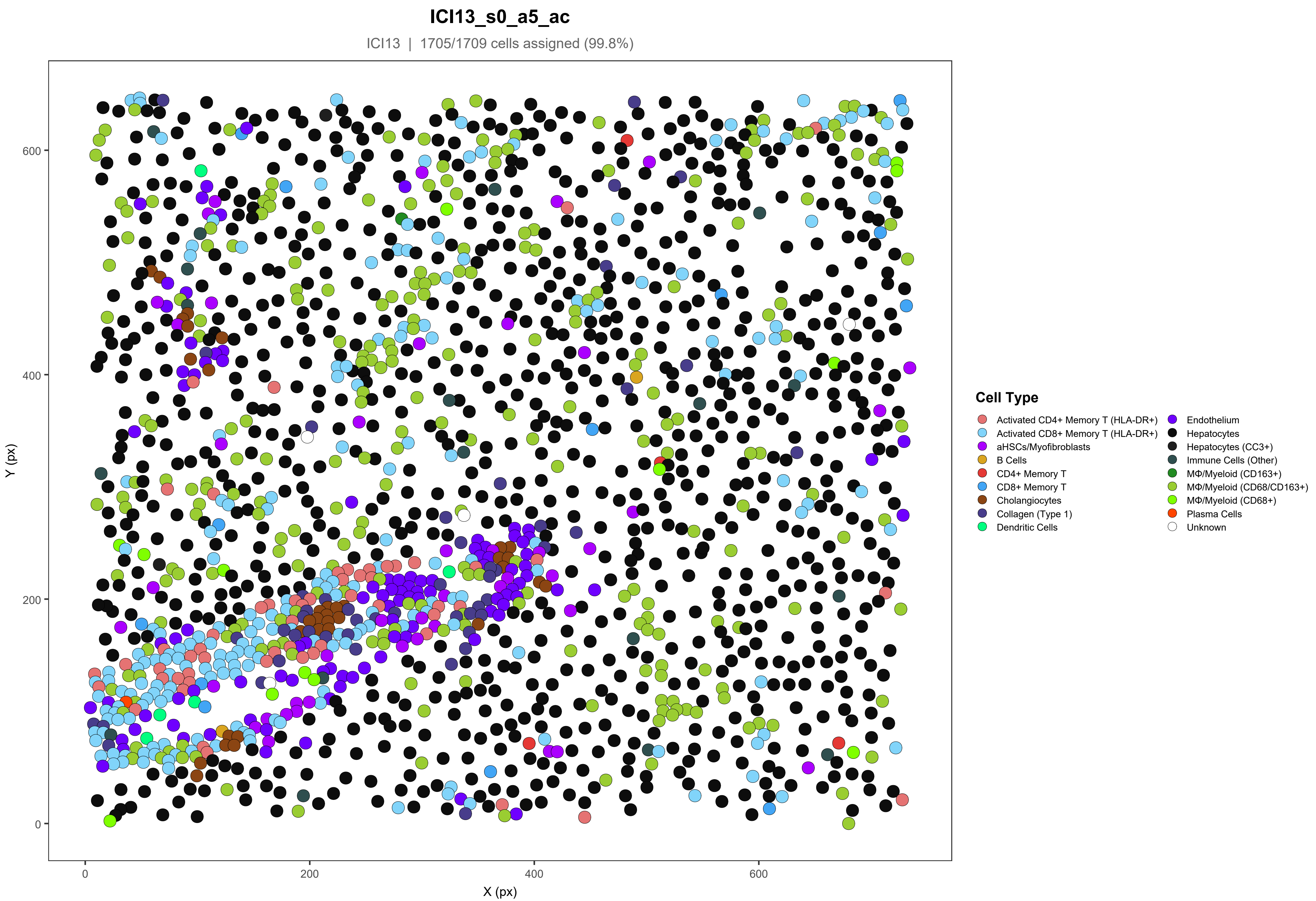
ICI13_s0_a5_ac
ICI13 | 1705/1709 cells assigned (99.8%)
Y (px)
X (px)
0
200
400
600
Cell Type
Activated CD4+ Memory T (HLA-DR+)
Activated CD8+ Memory T (HLA-DR+)
aHSCs/Myofibroblasts
B Cells
CD4+ Memory T
CD8+ Memory T
Cholangiocytes
Collagen (Type 1)
Dendritic Cells
Endothelium
Hepatocytes
Hepatocytes (CC3+)
Immune Cells (Other)
MΦ/Myeloid (CD163+)
MΦ/Myeloid (CD68/CD163+)
MΦ/Myeloid (CD68+)
Plasma Cells
Unknown

ICI14_s0_a1_ac

ICI14 | 1628/1628 cells assigned (100%)

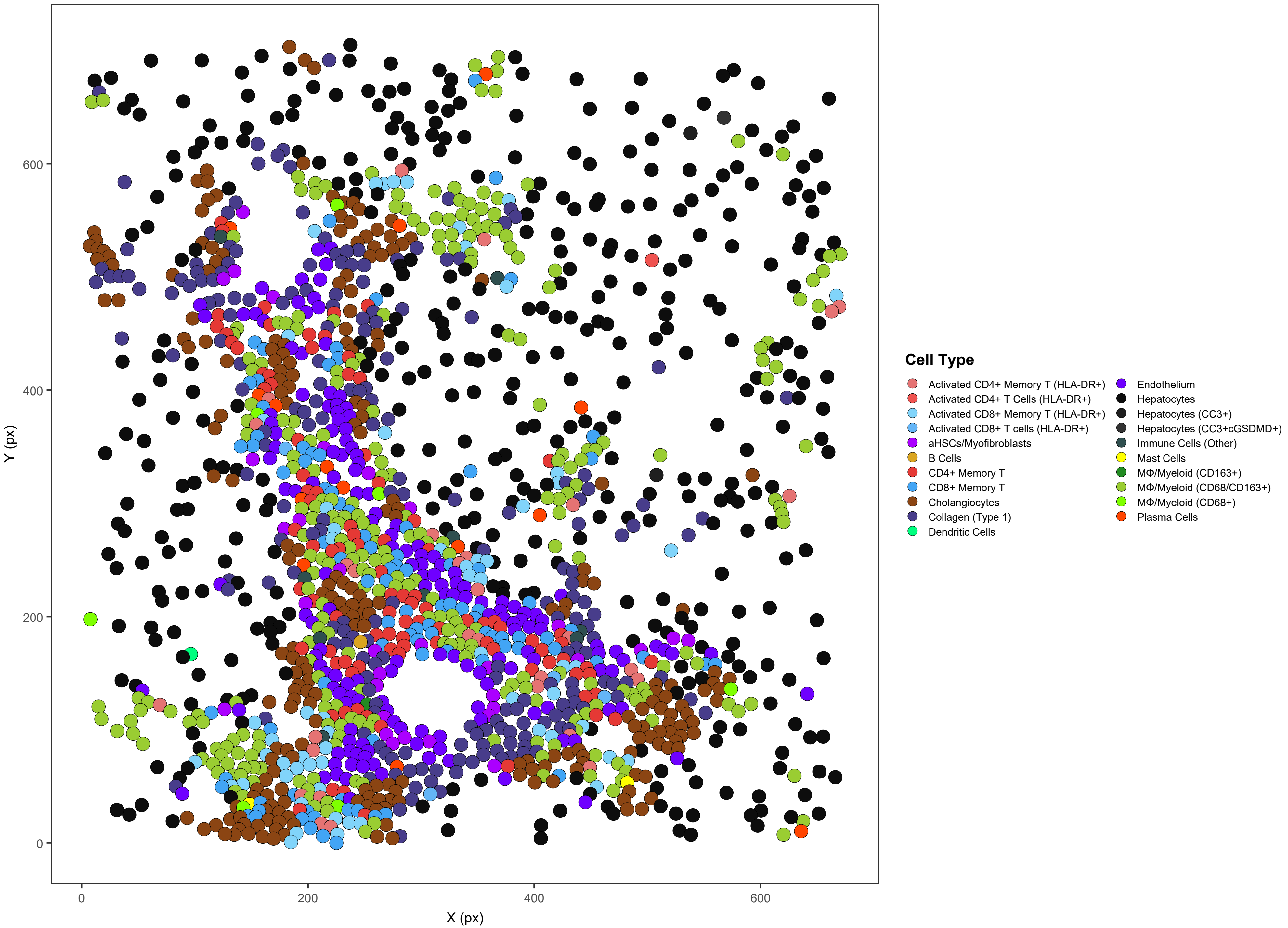

### ICI14_s0_a3_ac

ICI14 | 1293/1293 cells assigned (100%)

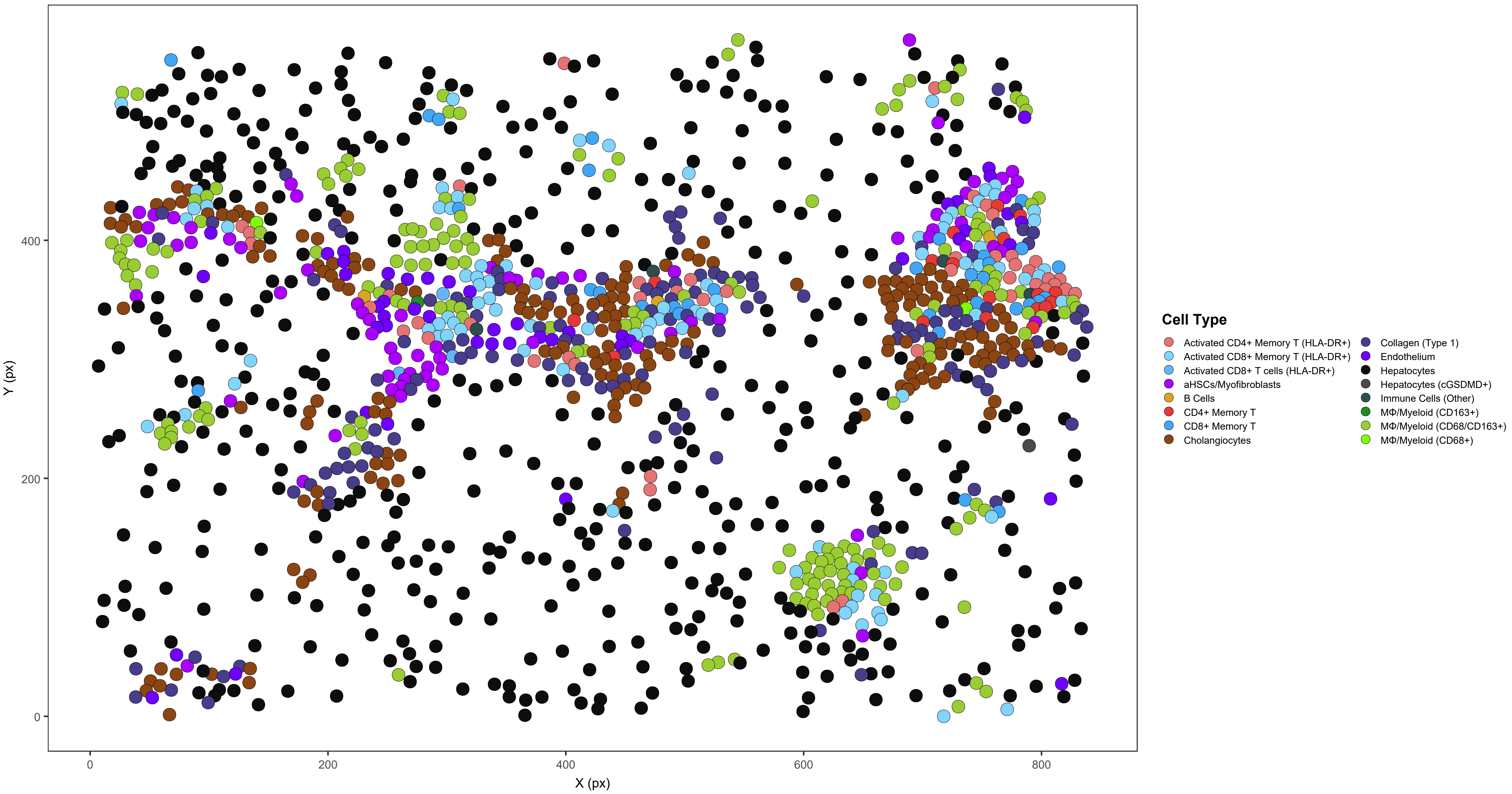

**ICI14_s0_a4_ac**

ICI14 | 1438/1439 cells assigned (99.9%)

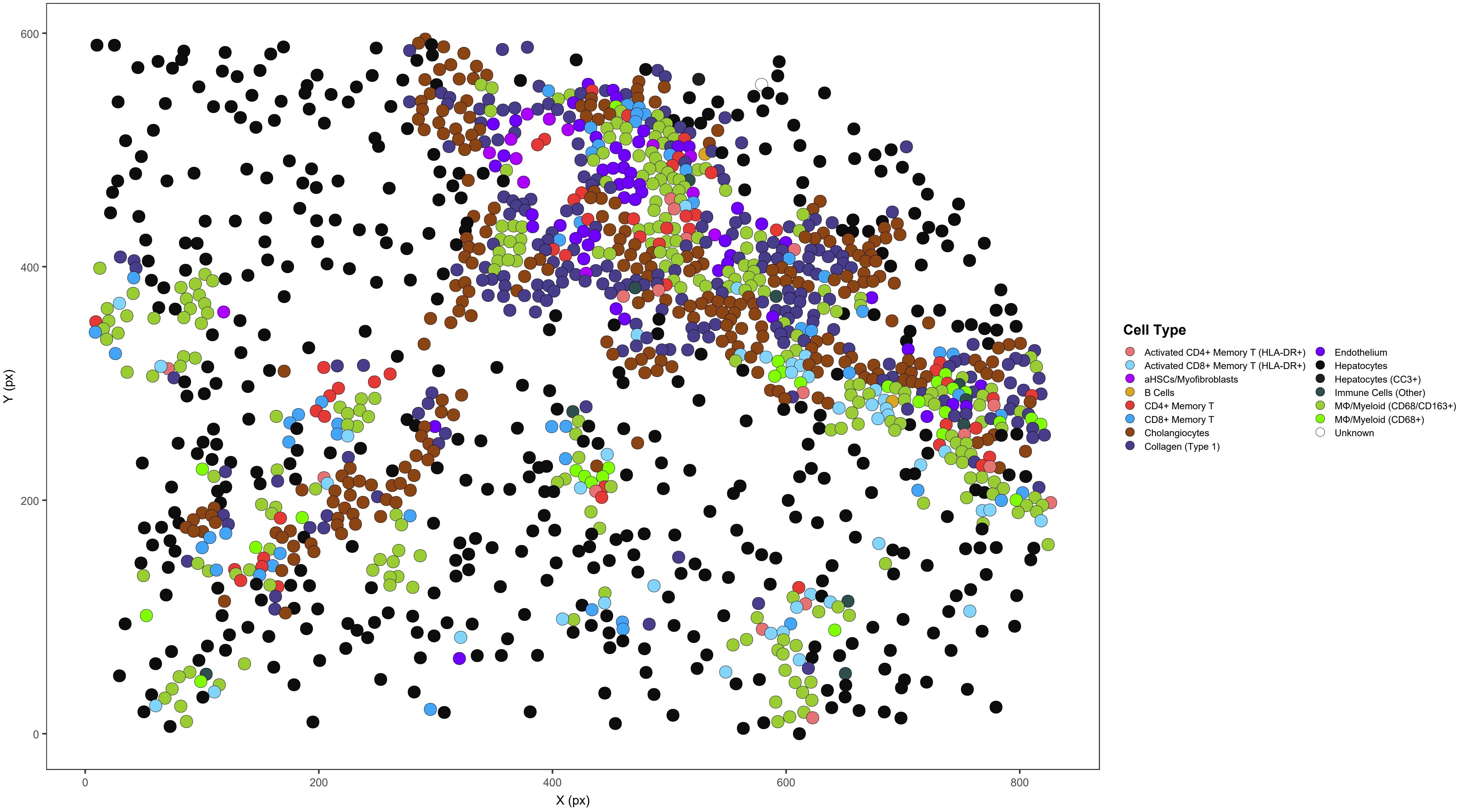

### ICI14_s0_a5_ac

ICI14 | 845/846 cells assigned (99.9%)

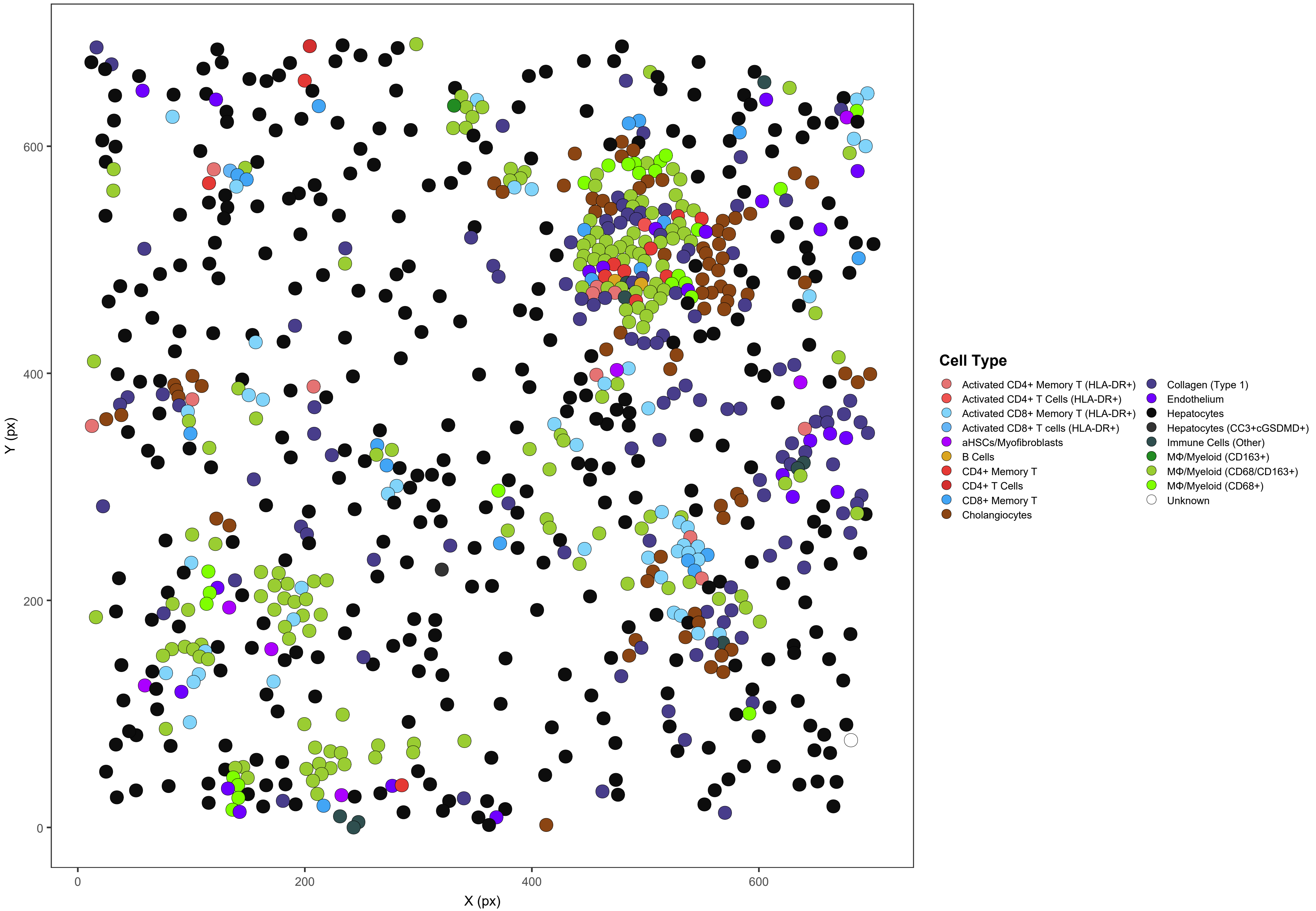

#### ICI15_s0_a1_ac

ICI15 | 2178/2178 cells assigned (100%)

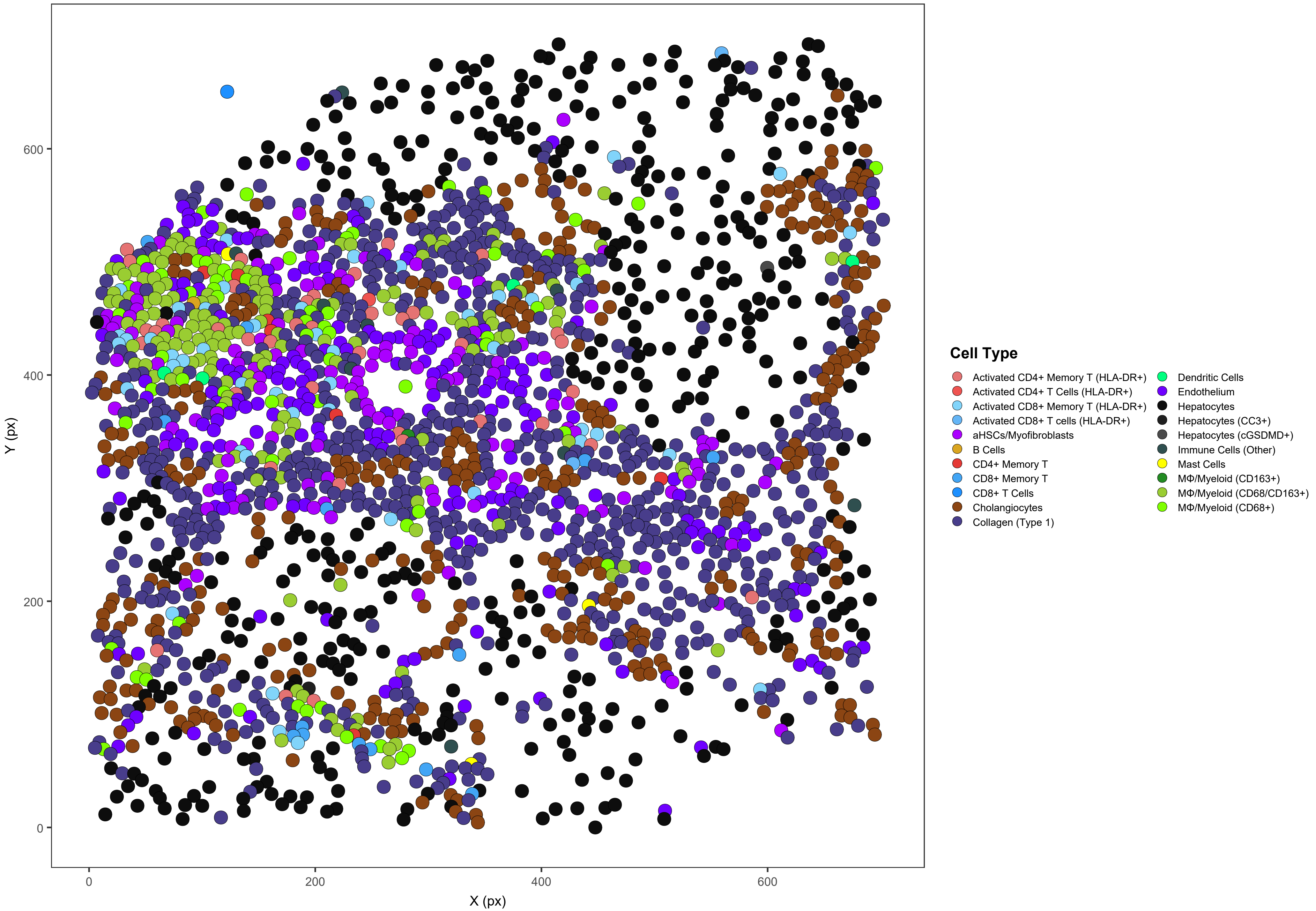

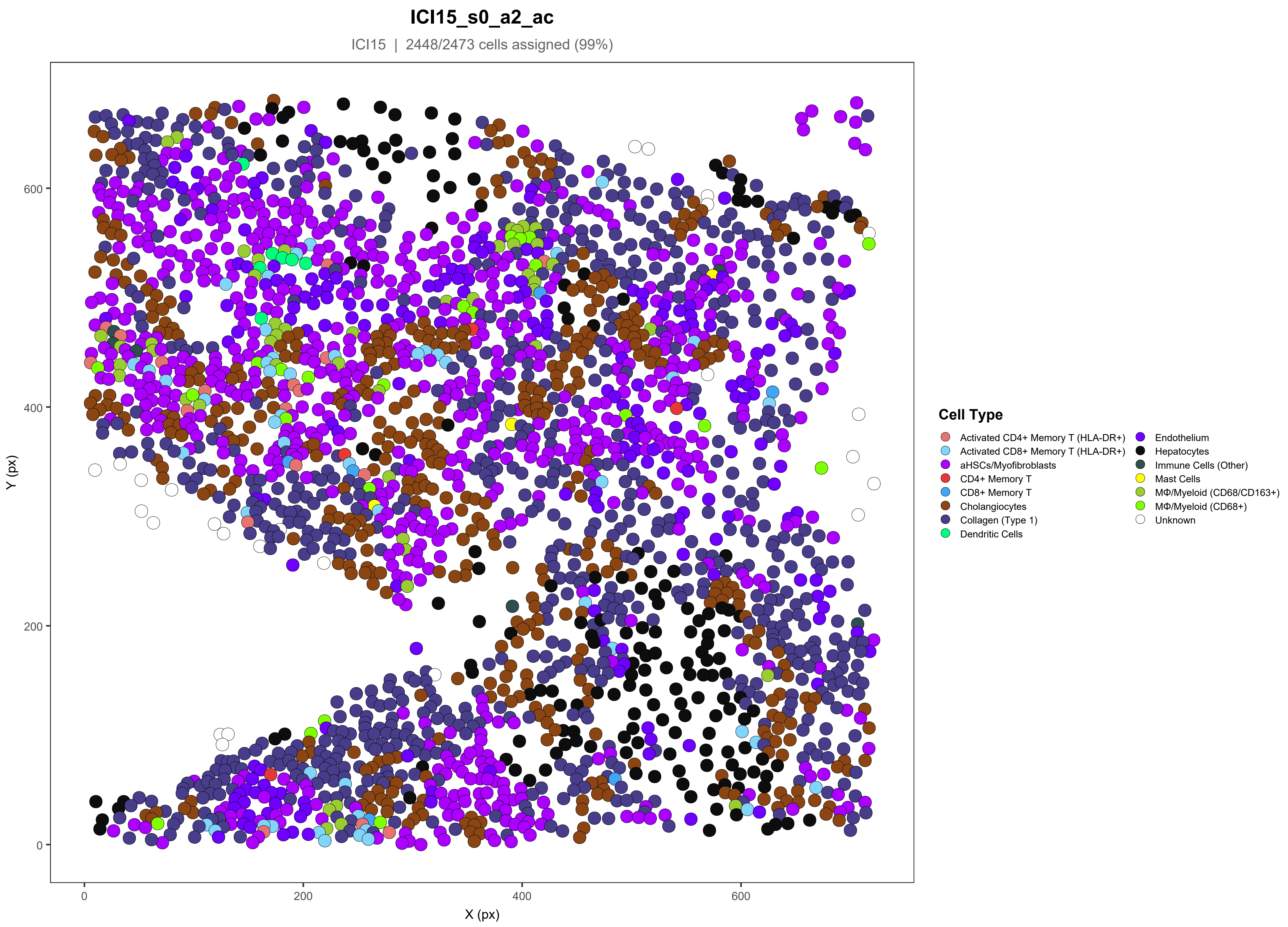
ICI15_s0_a2_ac
ICI15 | 2448/2473 cells assigned (99%)
Y (px)
X (px)
0
200
400
600
Cell Type
Activated CD4+ Memory T (HLA-DR+)
Activated CD8+ Memory T (HLA-DR+)
aHSCs/Myofibroblasts
CD4+ Memory T
CD8+ Memory T
Cholangiocytes
Collagen (Type 1)
Dendritic Cells
Endothelium
Hepatocytes
Immune Cells (Other)
Mast Cells
MΦ/Myeloid (CD68/CD163+)
MΦ/Myeloid (CD68+)
Unknown

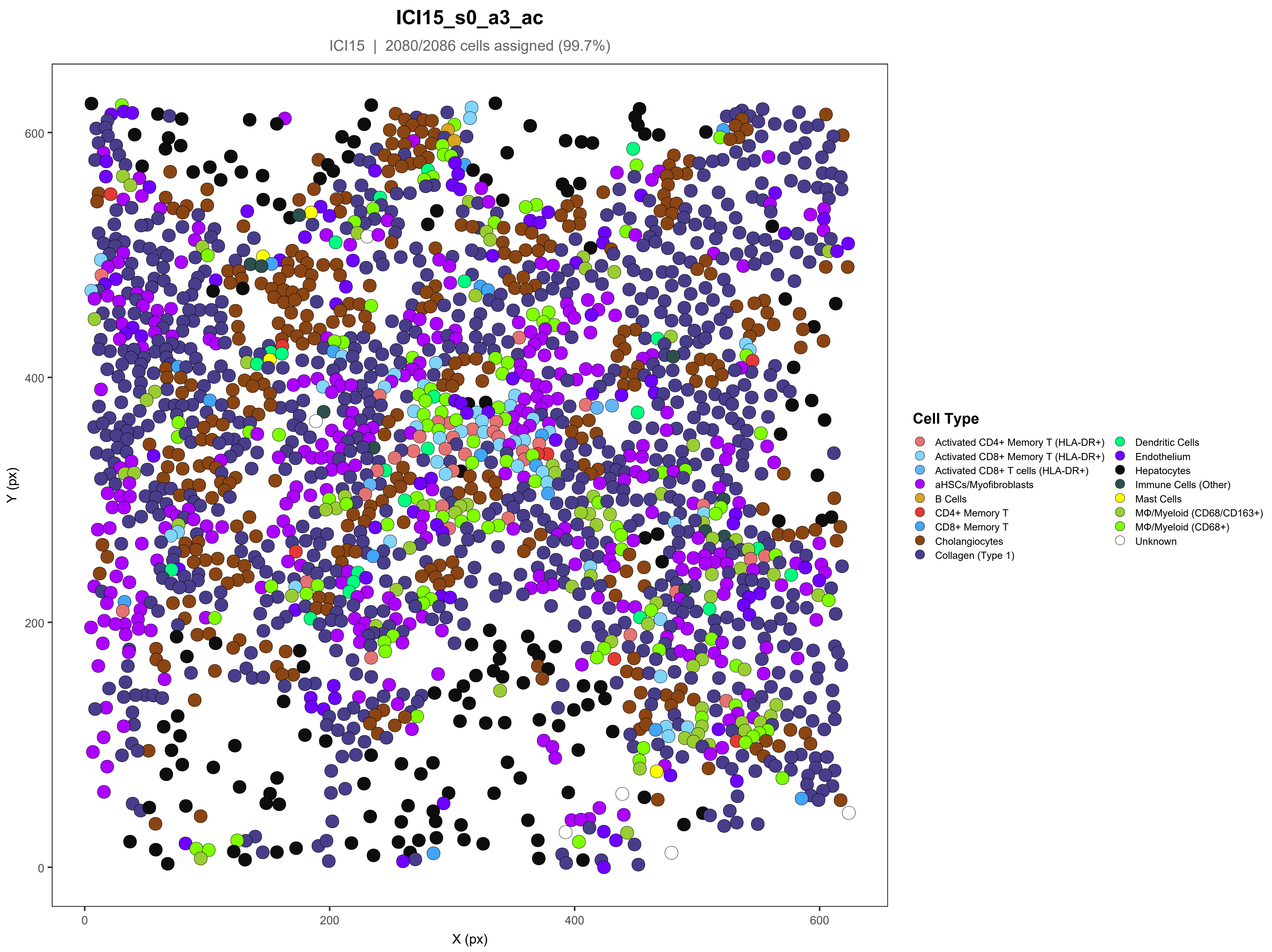
ICI15_s0_a3_ac
ICI15 | 2080/2086 cells assigned (99.7%)
Y (px)
X (px)
0
200
400
600
Cell Type
Activated CD4+ Memory T (HLA-DR+)
Activated CD8+ Memory T (HLA-DR+)
Activated CD8+ T cells (HLA-DR+)
aHSCs/Myofibroblasts
B Cells
CD4+ Memory T
CD8+ Memory T
Cholangiocytes
Collagen (Type 1)
Dendritic Cells
Endothelium
Hepatocytes
Immune Cells (Other)
Mast Cells
MΦ/Myeloid (CD68/CD163+)
MΦ/Myeloid (CD68+)
Unknown

### ICI15_s0_a4_ac

ICI15 | 1559/1560 cells assigned (99.9%)

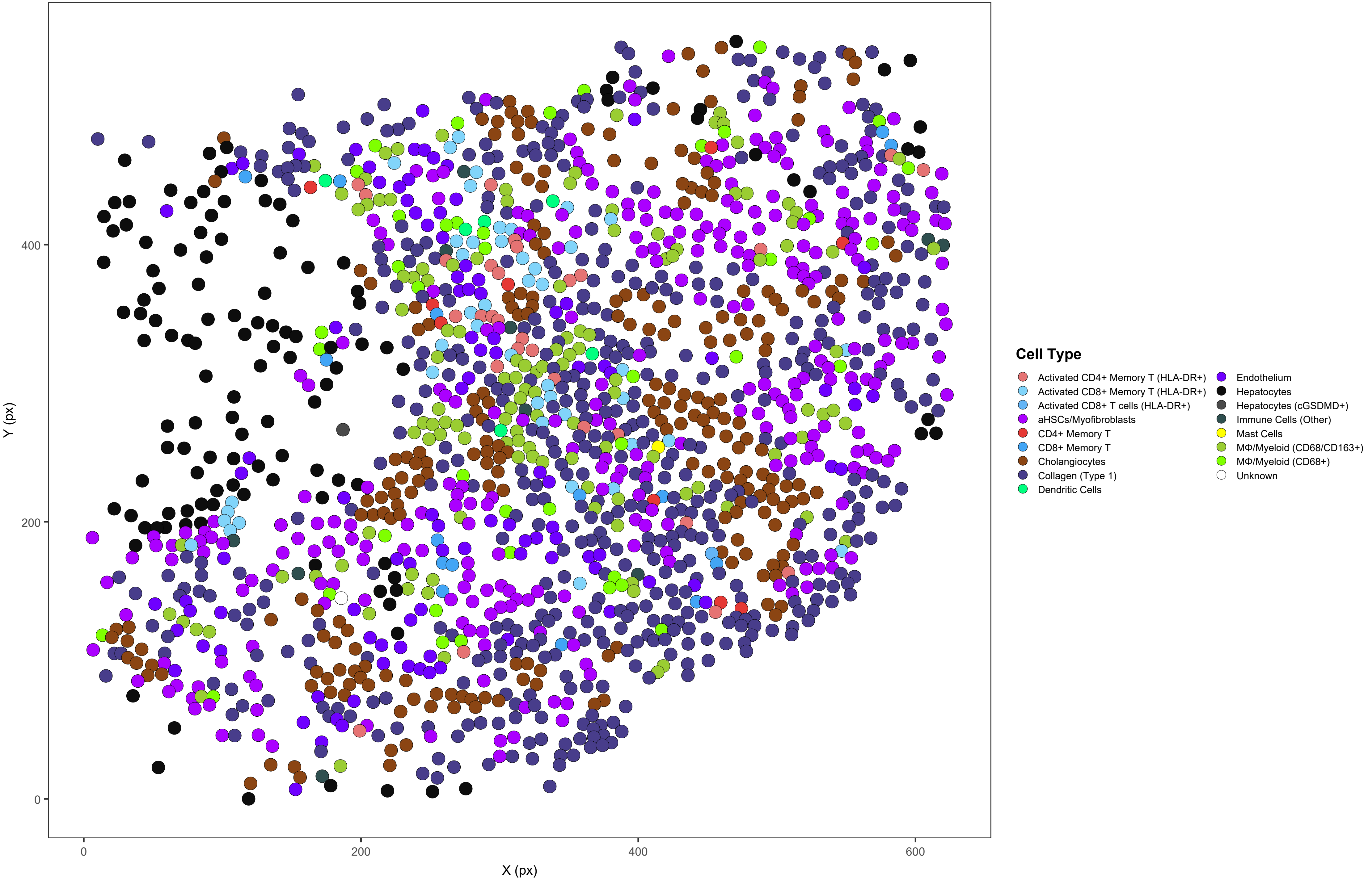

### ICI15_s0_a5_ac

ICI15 | 2037/2047 cells assigned (99.5%)

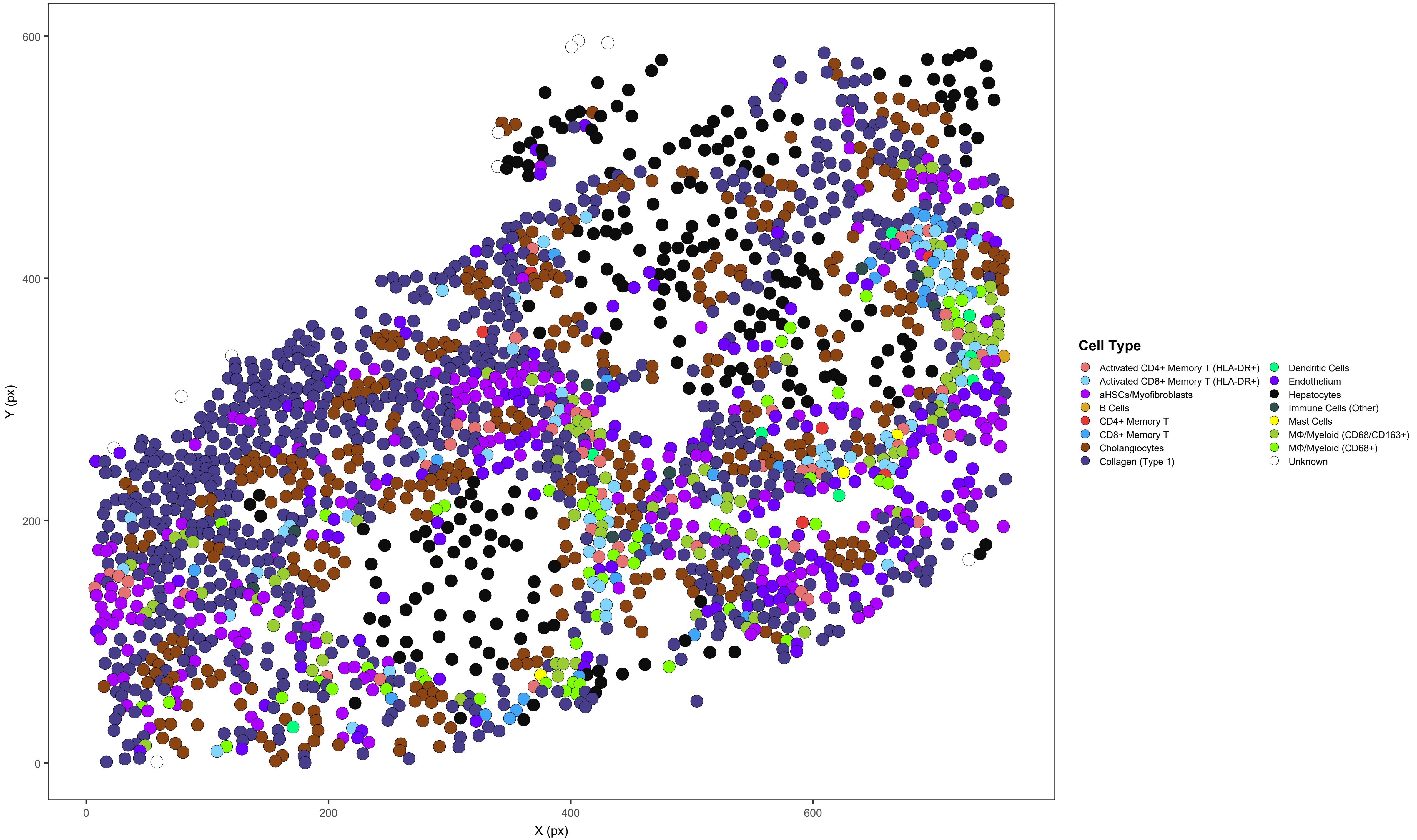
