## Supplementary material for "Macrophage-CD8⁺ T Cell Spatial Coupling Defines an Innate-Adaptive Injury Niche in Human Checkpoint Inhibitor Hepatotoxicity": Figure S-5

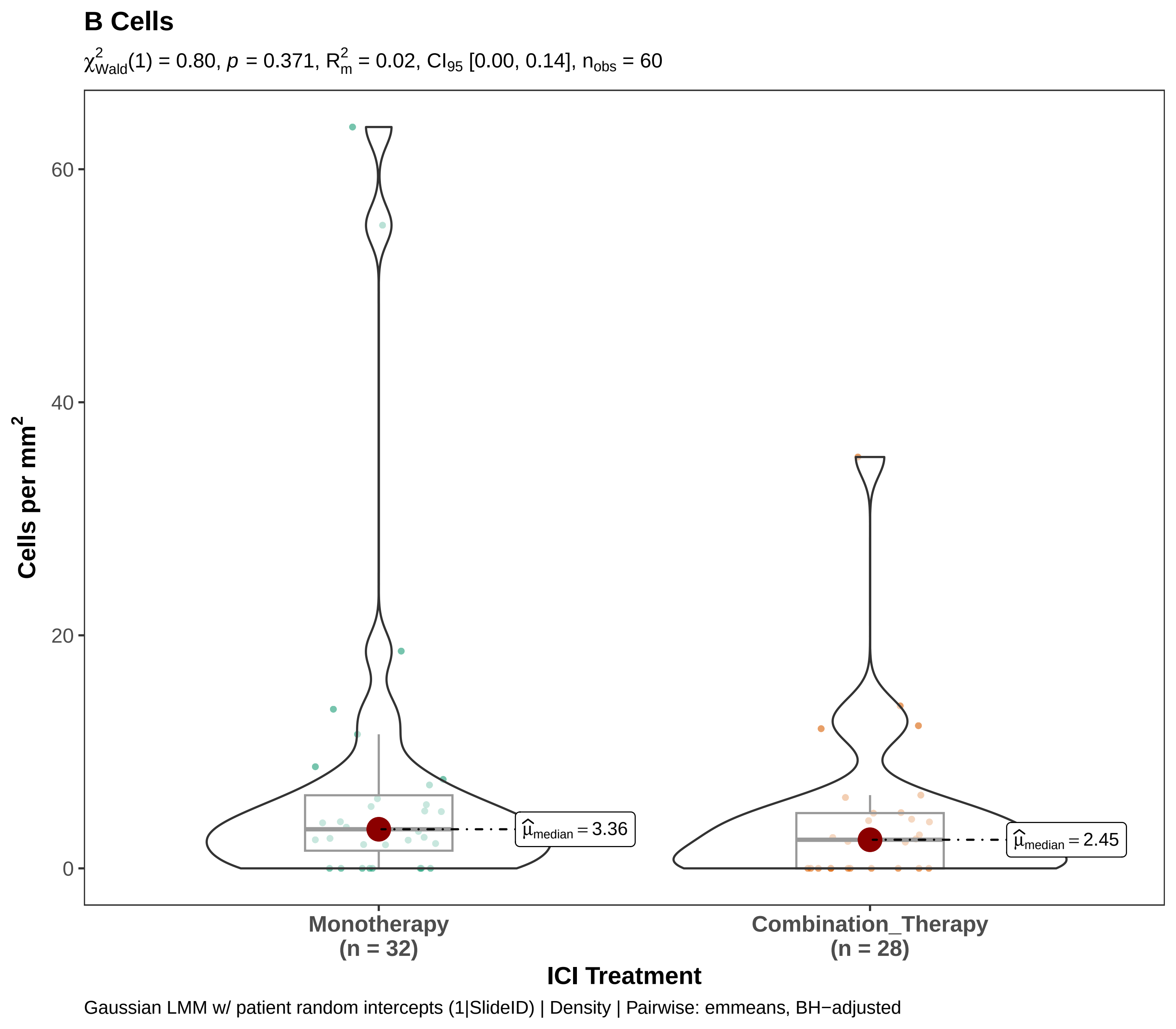
B Cells
$\chi^2_{Wald}(1) = 0.80$, $p = 0.371$, $R^2_m = 0.02$, $CI_{95}$ [0.00, 0.14], $n_{obs} = 60$
Cells per mm$^2$
60
40
20
0
$\hat{\mu}_{median} = 3.36$
$\hat{\mu}_{median} = 2.45$
Monotherapy
(n = 32)
Combination_Therapy
(n = 28)
ICI Treatment
Gaussian LMM w/ patient random intercepts (1|SlideID) | Density | Pairwise: emmeans, BH−adjusted

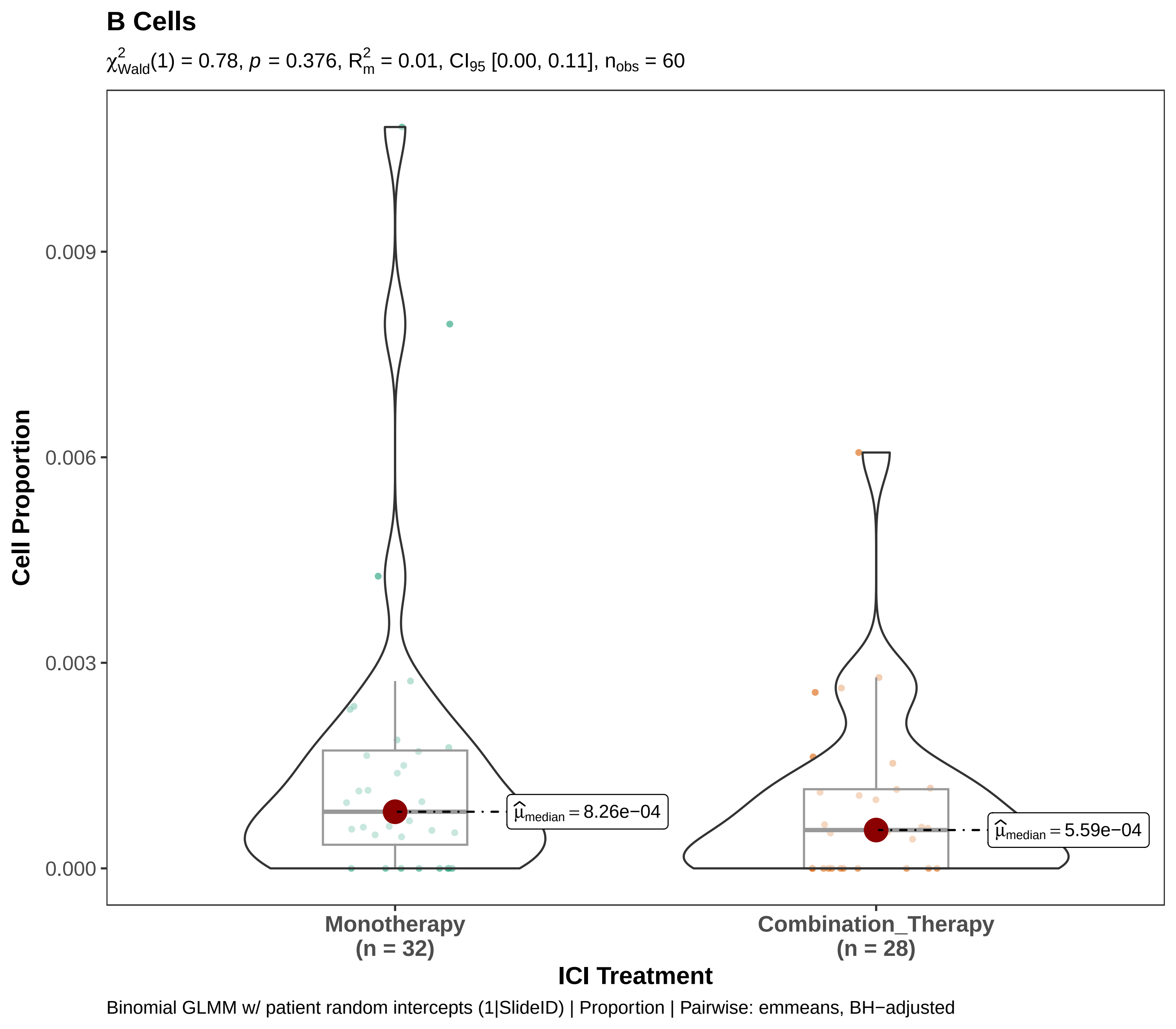
B Cells
$\chi^2_{Wald}(1) = 0.78$, $p = 0.376$, $R^2_m = 0.01$, $CI_{95}$ [0.00, 0.11], $n_{obs} = 60$
Cell Proportion
0.009
0.006
0.003
0.000
$\widehat{\mu}_{median} = 8.26e-04$
$\widehat{\mu}_{median} = 5.59e-04$
Monotherapy
(n = 32)
Combination_Therapy
(n = 28)
ICI Treatment
Binomial GLMM w/ patient random intercepts (1|SlideID) | Proportion | Pairwise: emmeans, BH−adjusted

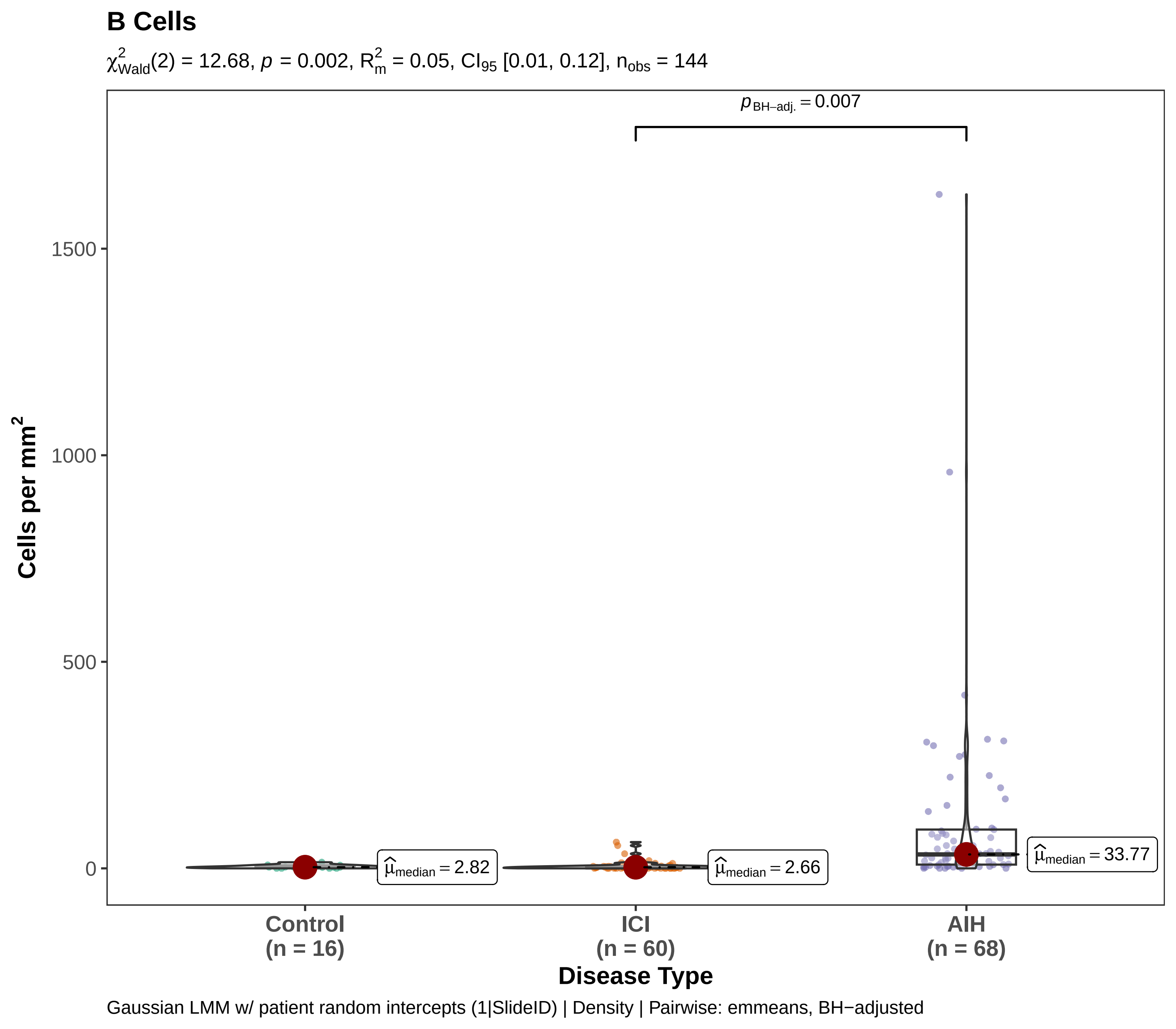

B Cells
$\chi^2_{Wald}(2) = 12.68$, $p = 0.002$, $R^2_m = 0.05$, $CI_{95}$ [0.01, 0.12], $n_{obs} = 144$
$p_{BH-adj.} = 0.007$
Cells per mm$^2$
1500
1000
500
0
$\hat{\mu}_{median} = 2.82$
$\hat{\mu}_{median} = 2.66$
$\hat{\mu}_{median} = 33.77$
Control (n = 16)
ICI (n = 60)
AIH (n = 68)
Disease Type
Gaussian LMM w/ patient random intercepts (1|SlideID) | Density | Pairwise: emmeans, BH−adjusted

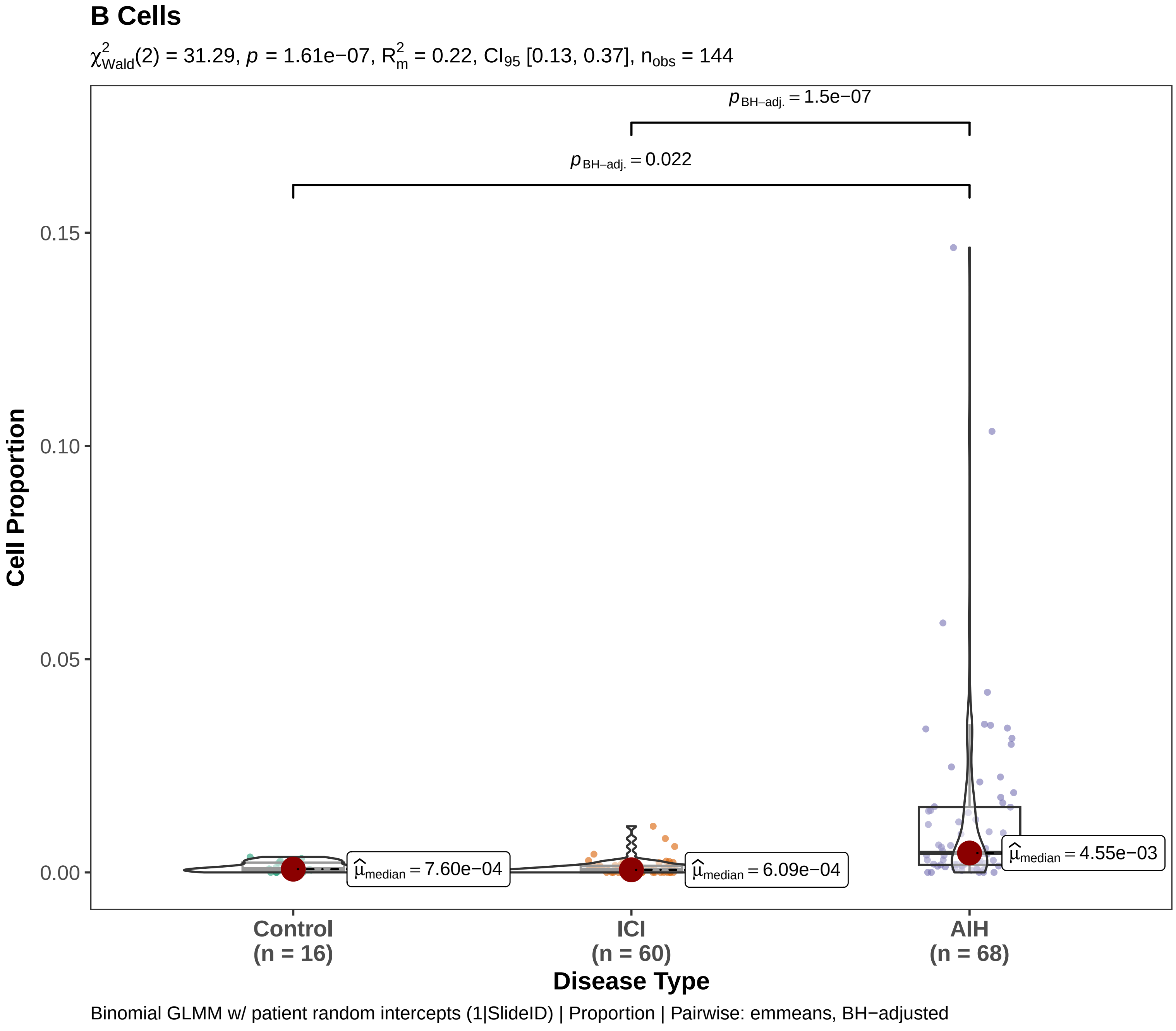
B Cells
$\chi^2_{Wald}(2) = 31.29$, $p = 1.61e-07$, $R^2_m = 0.22$, $CI_{95}$ [0.13, 0.37], $n_{obs} = 144$
$p_{BH-adj.} = 1.5e-07$
$p_{BH-adj.} = 0.022$
0.15
0.10
0.05
0.00
Cell Proportion
$\hat{\mu}_{median} = 7.60e-04$
$\hat{\mu}_{median} = 6.09e-04$
$\hat{\mu}_{median} = 4.55e-03$
Control
(n = 16)
ICI
(n = 60)
AIH
(n = 68)
Disease Type
Binomial GLMM w/ patient random intercepts (1|SlideID) | Proportion | Pairwise: emmeans, BH−adjusted

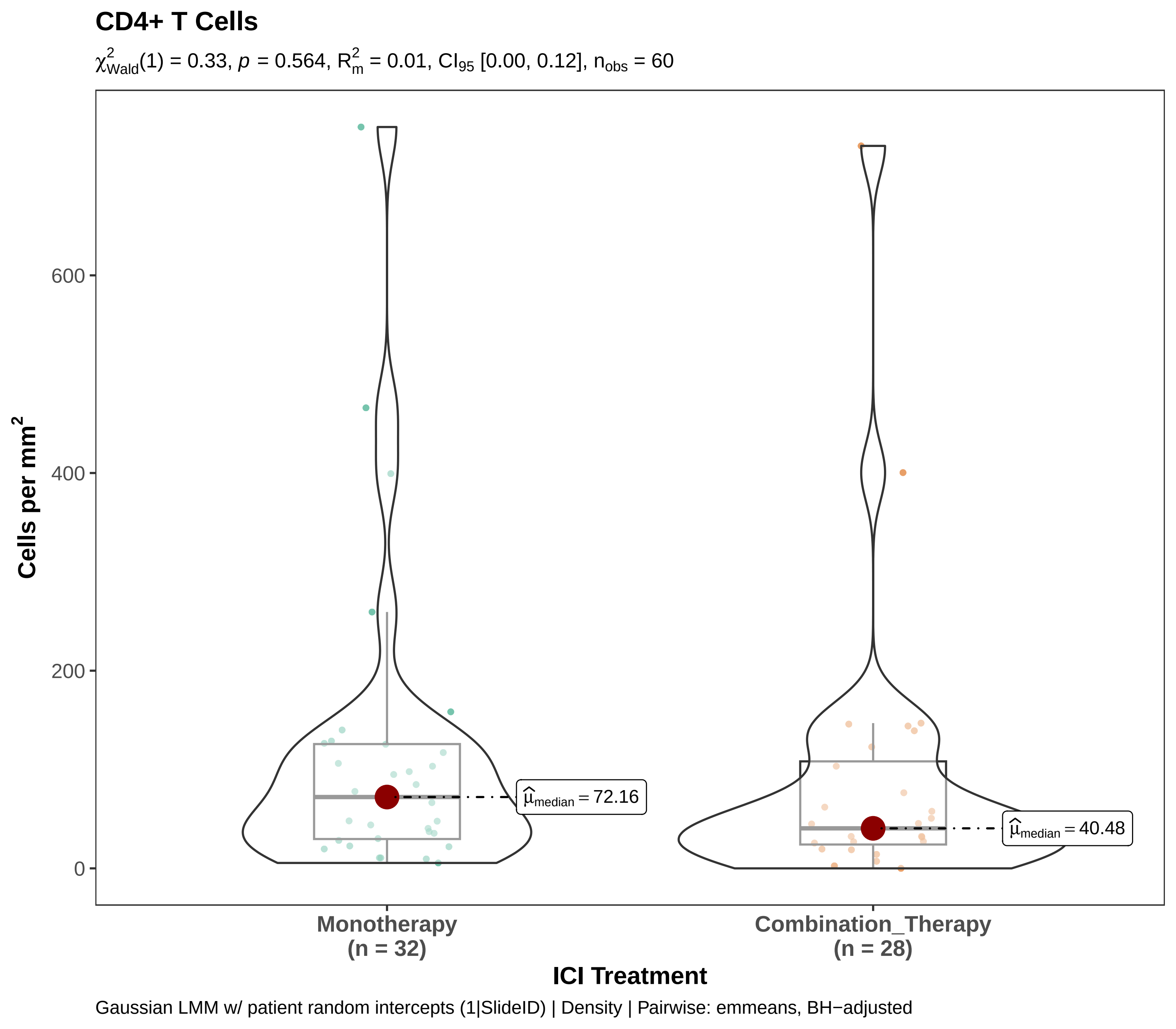
CD4+ T Cells
$\chi^2_{Wald}(1) = 0.33$, $p = 0.564$, $R^2_m = 0.01$, $CI_{95}$ [0.00, 0.12], $n_{obs} = 60$
Cells per mm$^2$
600
400
200
0
$\widehat{\mu}_{median} = 72.16$
$\widehat{\mu}_{median} = 40.48$
Monotherapy
(n = 32)
Combination_Therapy
(n = 28)
ICI Treatment
Gaussian LMM w/ patient random intercepts (1|SlideID) | Density | Pairwise: emmeans, BH−adjusted

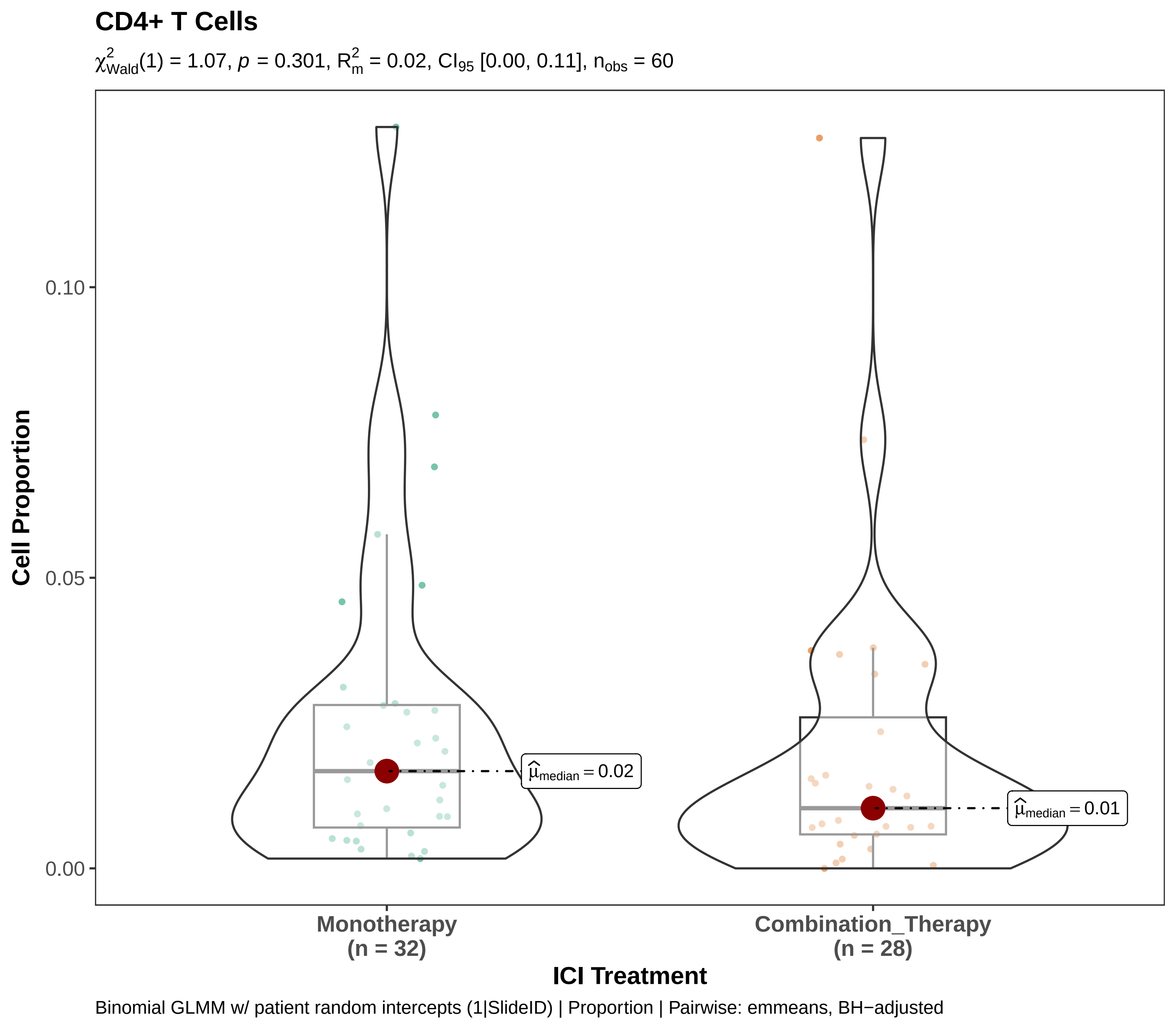
CD4+ T Cells
$\chi^2_{Wald}(1) = 1.07$, $p = 0.301$, $R^2_m = 0.02$, $CI_{95}$ [0.00, 0.11], $n_{obs} = 60$
Cell Proportion
0.10
0.05
0.00
$\hat{\mu}_{median} = 0.02$
$\hat{\mu}_{median} = 0.01$
Monotherapy
(n = 32)
Combination_Therapy
(n = 28)
ICI Treatment
Binomial GLMM w/ patient random intercepts (1|SlideID) | Proportion | Pairwise: emmeans, BH−adjusted

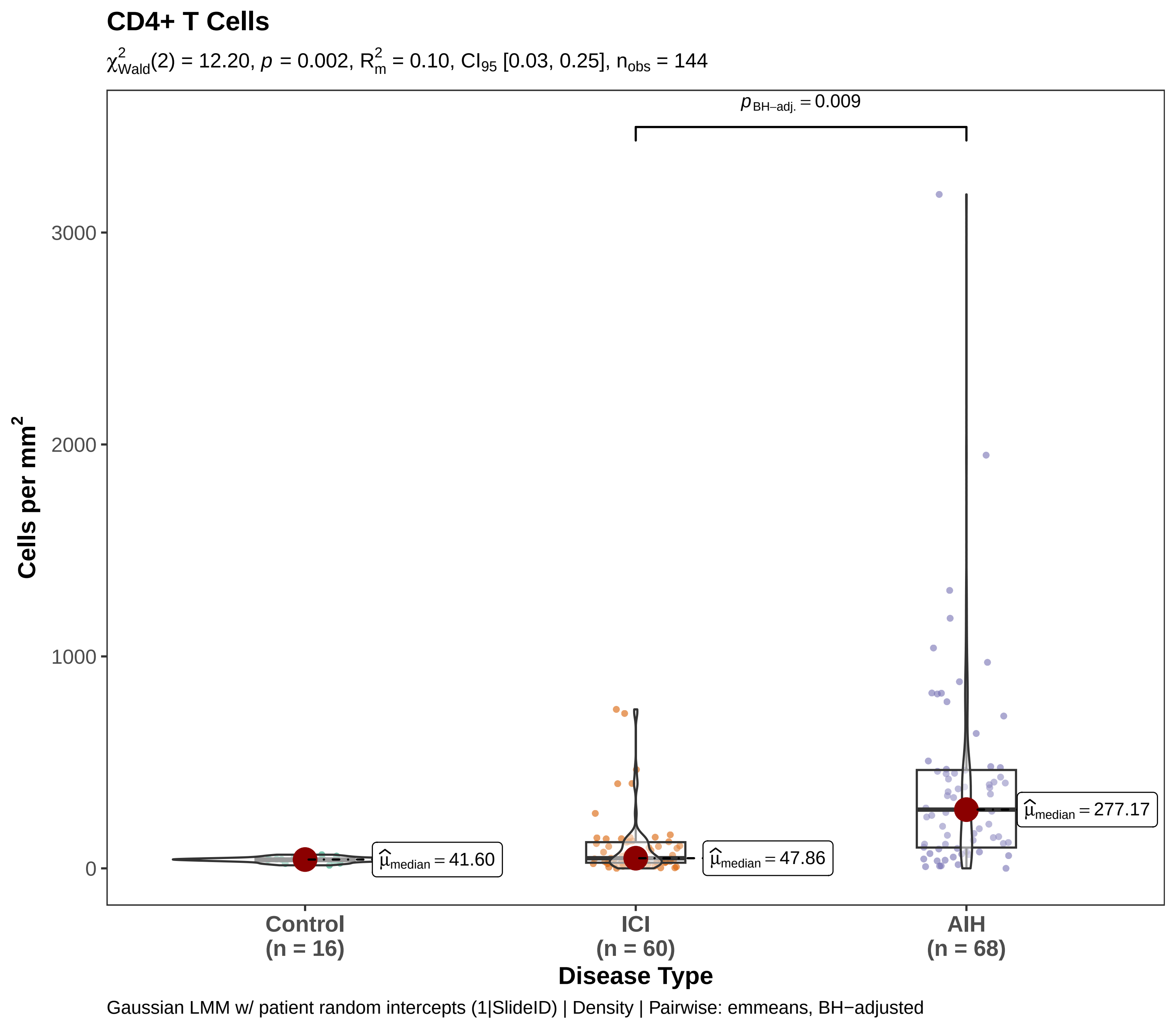
CD4+ T Cells
$\chi^2_{Wald}(2) = 12.20$, $p = 0.002$, $R^2_m = 0.10$, $CI_{95}$ [0.03, 0.25], $n_{obs} = 144$
$p_{BH-adj.} = 0.009$
Cells per mm$^2$
3000
2000
1000
0
$\widehat{\mu}_{median} = 41.60$
$\widehat{\mu}_{median} = 47.86$
$\widehat{\mu}_{median} = 277.17$
Control (n = 16)
ICI (n = 60)
AIH (n = 68)
Disease Type
Gaussian LMM w/ patient random intercepts (1|SlideID) | Density | Pairwise: emmeans, BH−adjusted

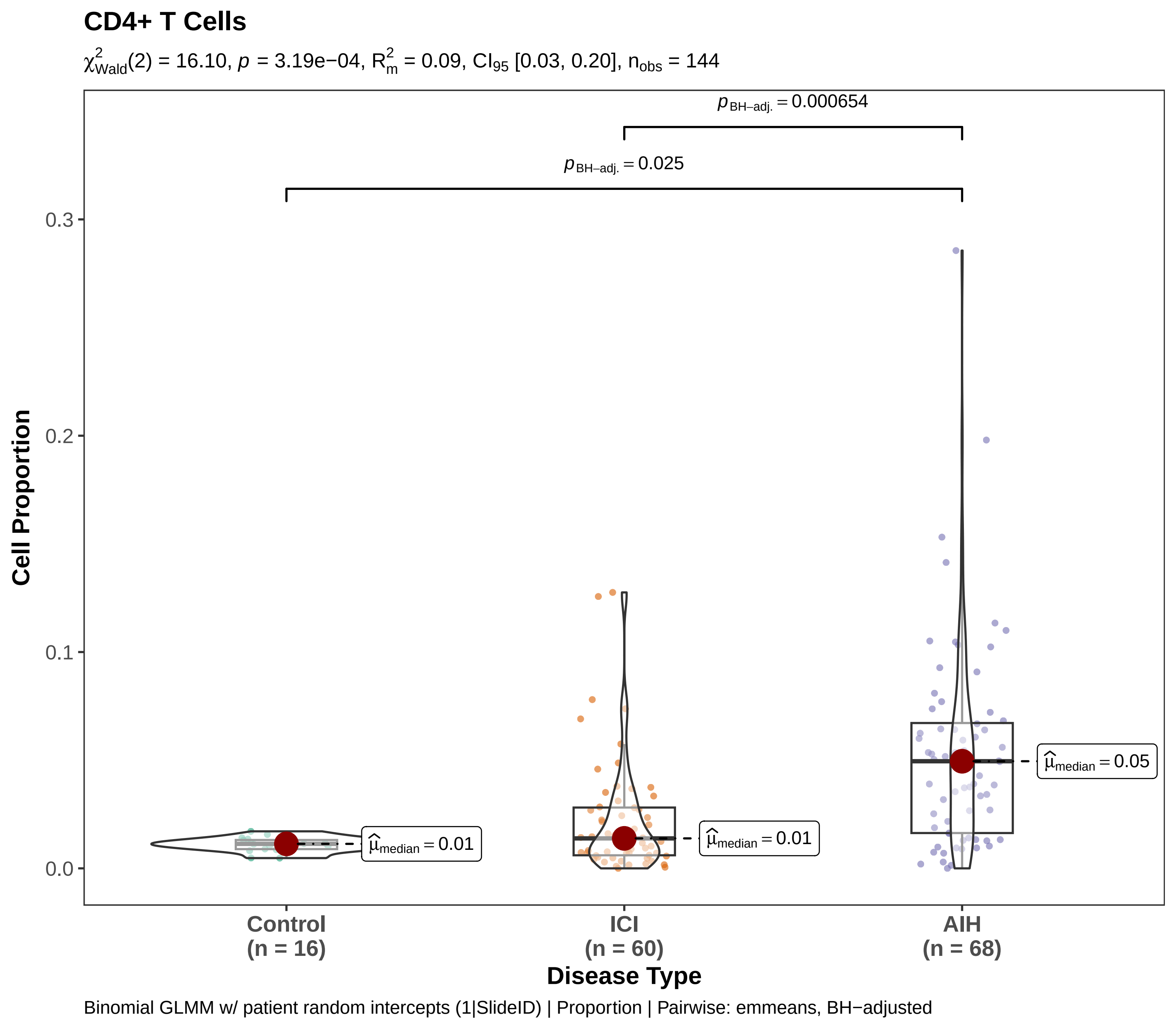

CD4+ T Cells
$\chi^2_{Wald}(2) = 16.10$, $p = 3.19e{-}04$, $R^2_m = 0.09$, $CI_{95}$ [0.03, 0.20], $n_{obs} = 144$
$p_{BH-adj.} = 0.000654$
$p_{BH-adj.} = 0.025$
Cell Proportion
0.3
0.2
0.1
0.0
$\hat{\mu}_{median} = 0.01$
$\hat{\mu}_{median} = 0.01$
$\hat{\mu}_{median} = 0.05$
Control
(n = 16)
ICI
(n = 60)
AIH
(n = 68)
Disease Type
Binomial GLMM w/ patient random intercepts (1|SlideID) | Proportion | Pairwise: emmeans, BH−adjusted

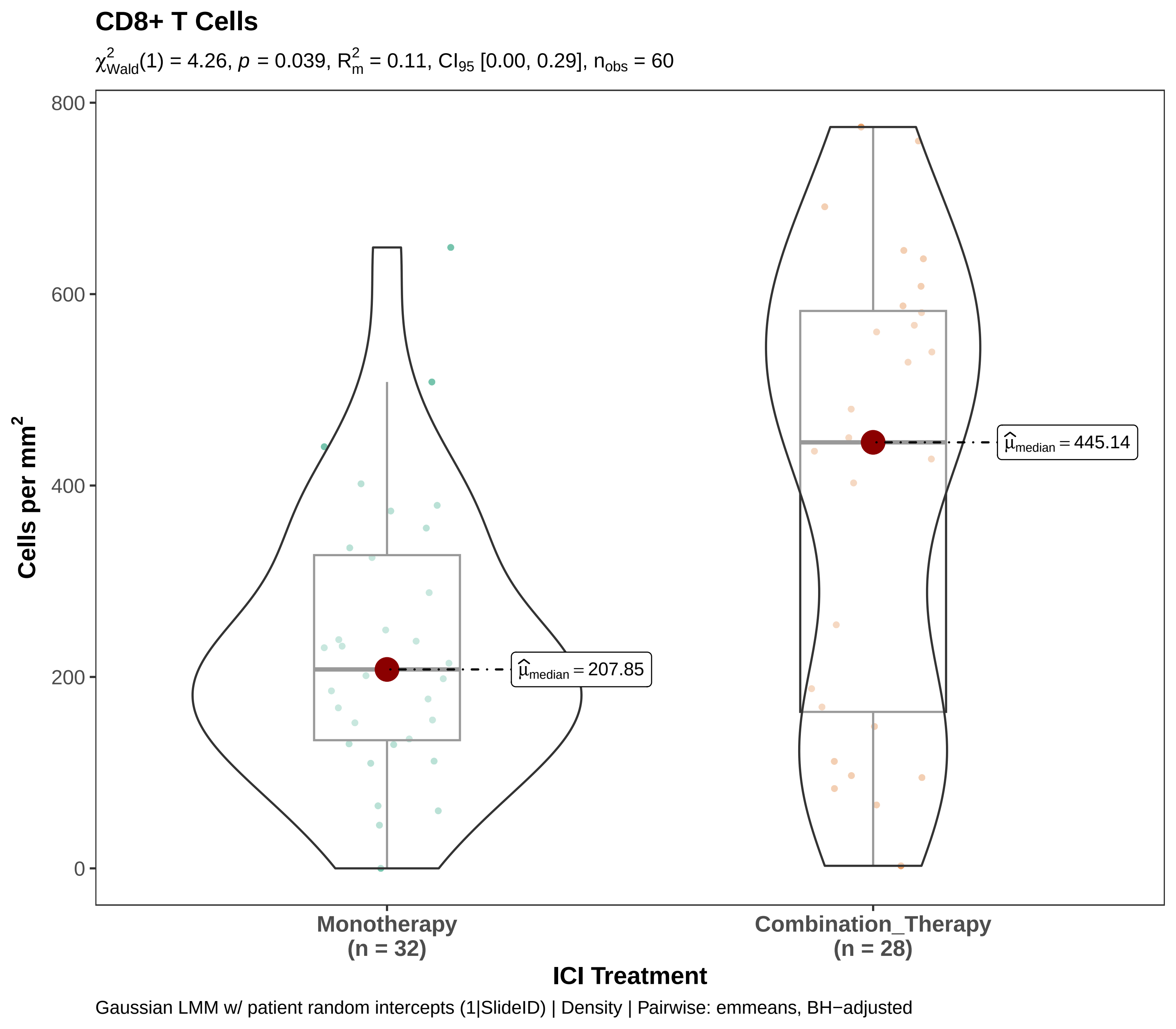

CD8+ T Cells
$\chi^2_{Wald}(1) = 4.26$, $p = 0.039$, $R^2_m = 0.11$, $CI_{95}$ [0.00, 0.29], $n_{obs} = 60$
Cells per mm²
800
600
400
200
0
$\hat{\mu}_{median} = 207.85$
$\hat{\mu}_{median} = 445.14$
Monotherapy
(n = 32)
Combination_Therapy
(n = 28)
ICI Treatment
Gaussian LMM w/ patient random intercepts (1|SlideID) | Density | Pairwise: emmeans, BH−adjusted

#### CD8+ T Cells

$\chi^2_{Wald}(1) = 3.12$, $p = 0.077$, $R^2_m = 0.03$, $CI_{95}$ [0.00, 0.09], $n_{obs} = 60$

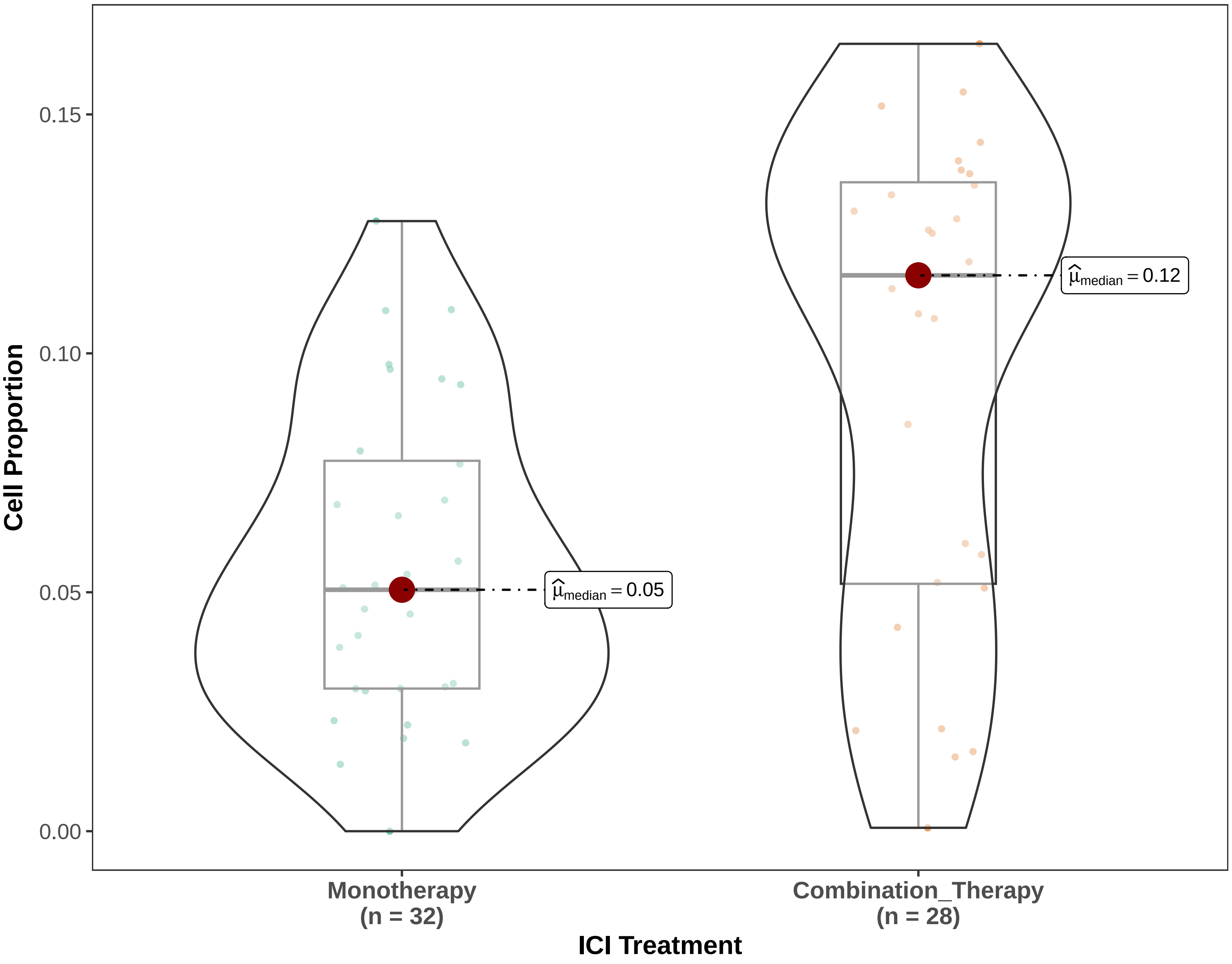


Binomial GLMM w/ patient random intercepts (1|SlideID) | Proportion | Pairwise: emmeans, BH−adjusted

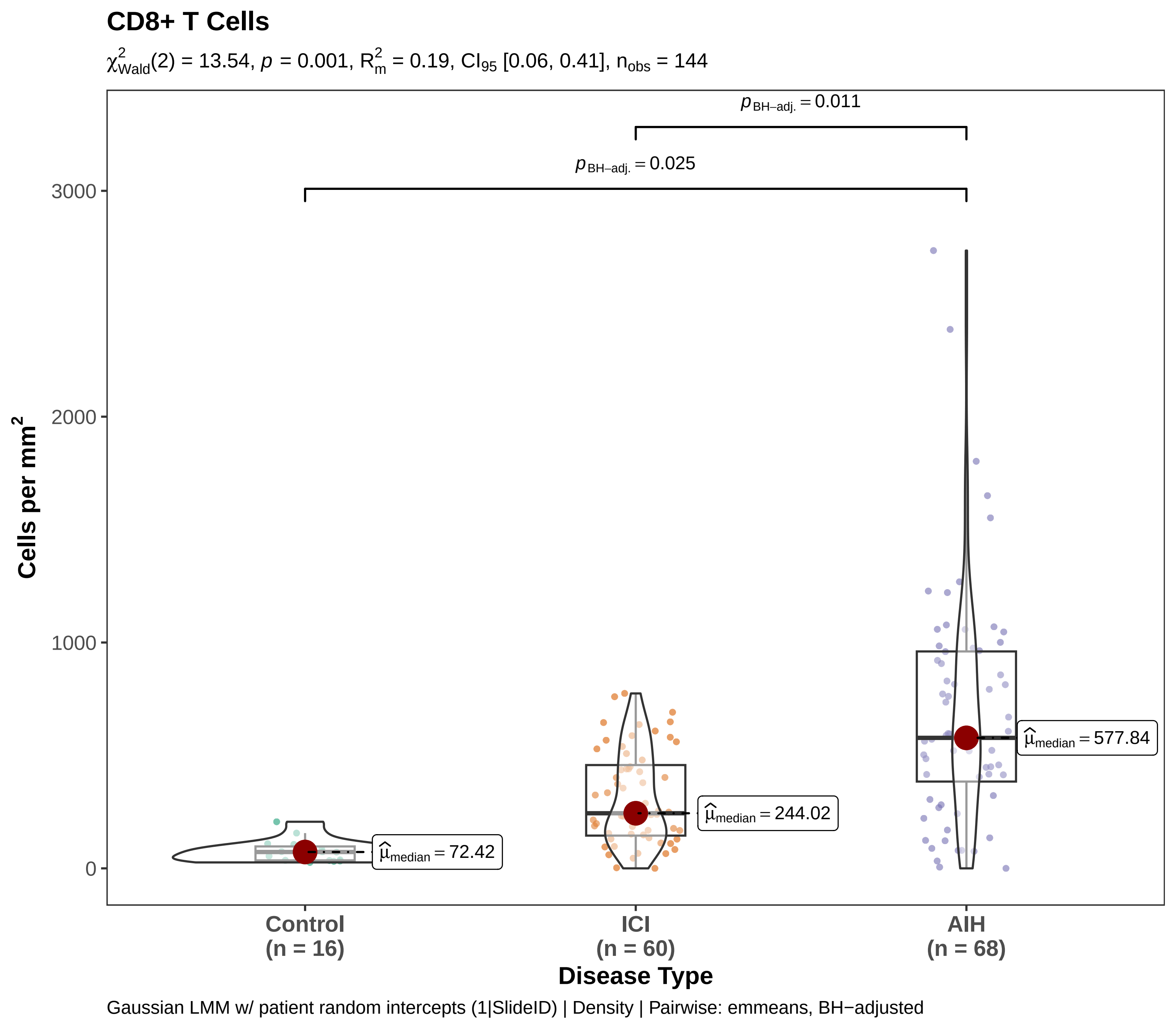

CD8+ T Cells
$\chi^2_{Wald}(2) = 13.54$, $p = 0.001$, $R^2_m = 0.19$, $CI_{95}$ [0.06, 0.41], $n_{obs} = 144$
$p_{BH-adj.} = 0.011$
$p_{BH-adj.} = 0.025$
3000
2000
1000
0
Cells per mm$^2$
$\widehat{\mu}_{median} = 72.42$
$\widehat{\mu}_{median} = 244.02$
$\widehat{\mu}_{median} = 577.84$
Control
(n = 16)
ICI
(n = 60)
AIH
(n = 68)
Disease Type
Gaussian LMM w/ patient random intercepts (1|SlideID) | Density | Pairwise: emmeans, BH−adjusted

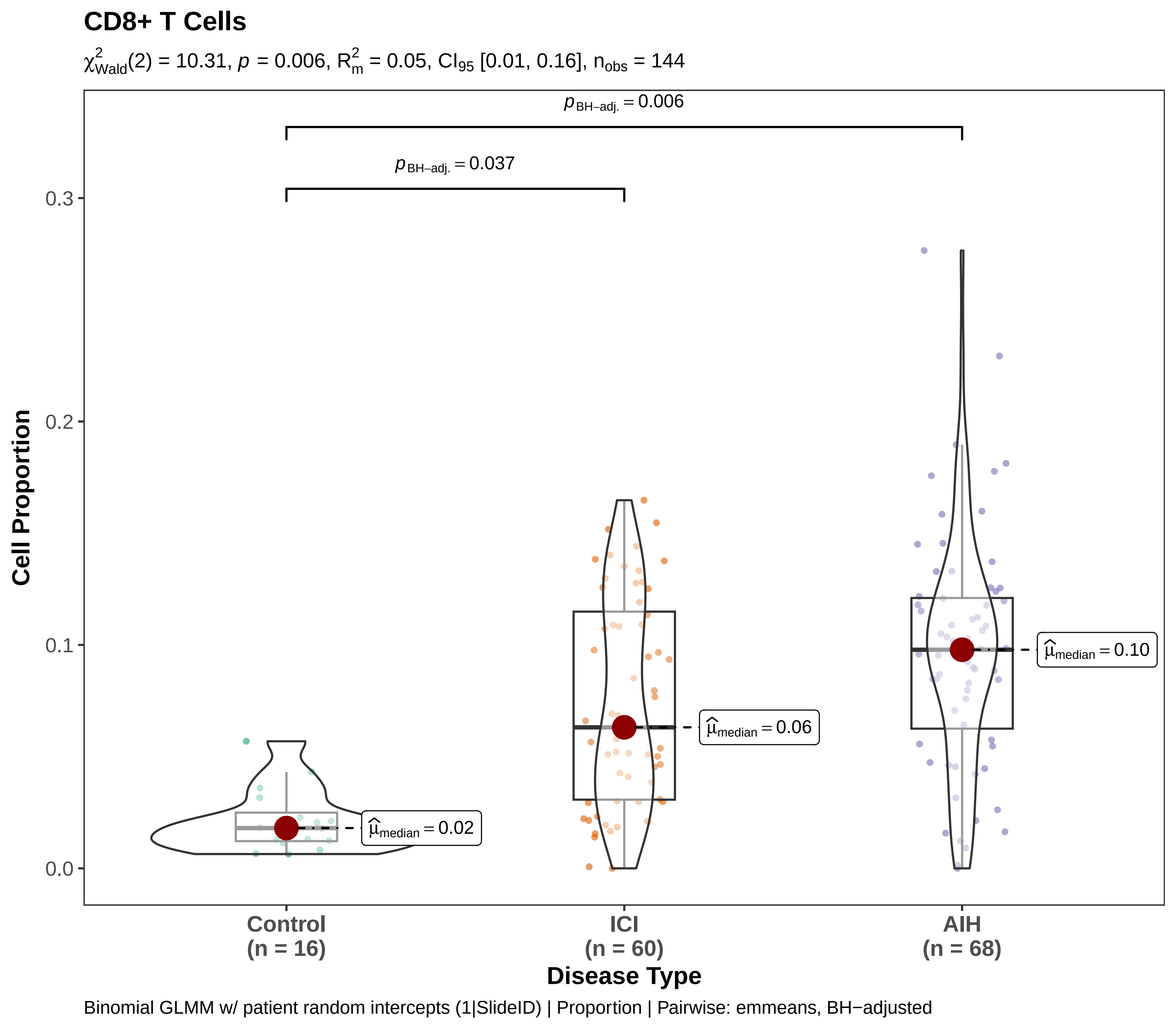
CD8+ T Cells
$\chi^2_{Wald}(2) = 10.31$, $p = 0.006$, $R^2_m = 0.05$, $CI_{95}$ [0.01, 0.16], $n_{obs} = 144$
$p_{BH-adj.} = 0.006$
$p_{BH-adj.} = 0.037$
0.3
0.2
0.1
0.0
Cell Proportion
$\widehat{\mu}_{median} = 0.02$
$\widehat{\mu}_{median} = 0.06$
$\widehat{\mu}_{median} = 0.10$
Control
(n = 16)
ICI
(n = 60)
AIH
(n = 68)
Disease Type
Binomial GLMM w/ patient random intercepts (1|SlideID) | Proportion | Pairwise: emmeans, BH−adjusted

**Cholangiocytes**

$\chi^2_{Wald}(1) = 9.51$, $p = 0.002$, $R^2_m = 0.29$, $CI_{95}$ [0.07, 0.56], $n_{obs} = 60$

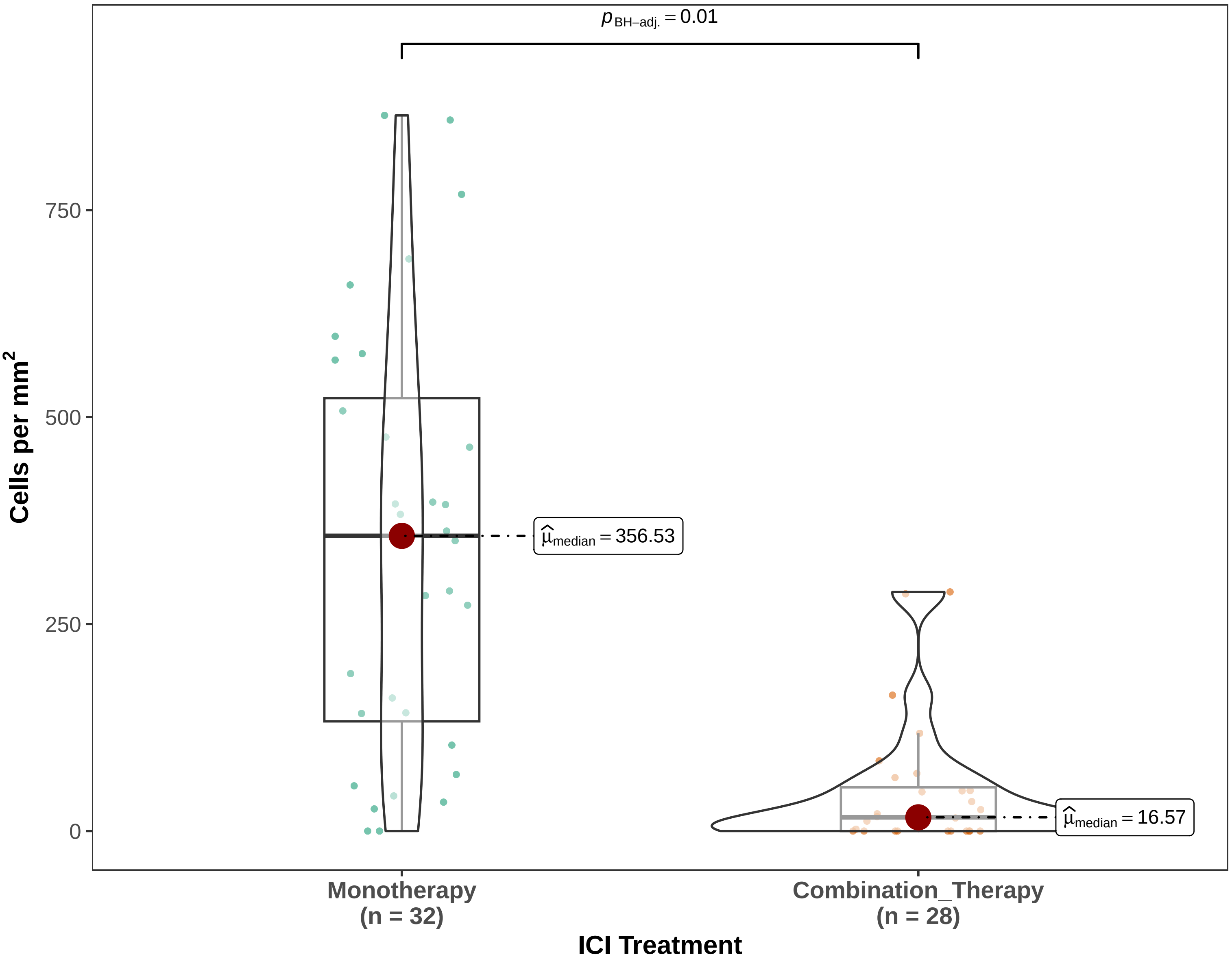


Gaussian LMM w/ patient random intercepts (1|SlideID) | Density | Pairwise: emmeans, BH−adjusted

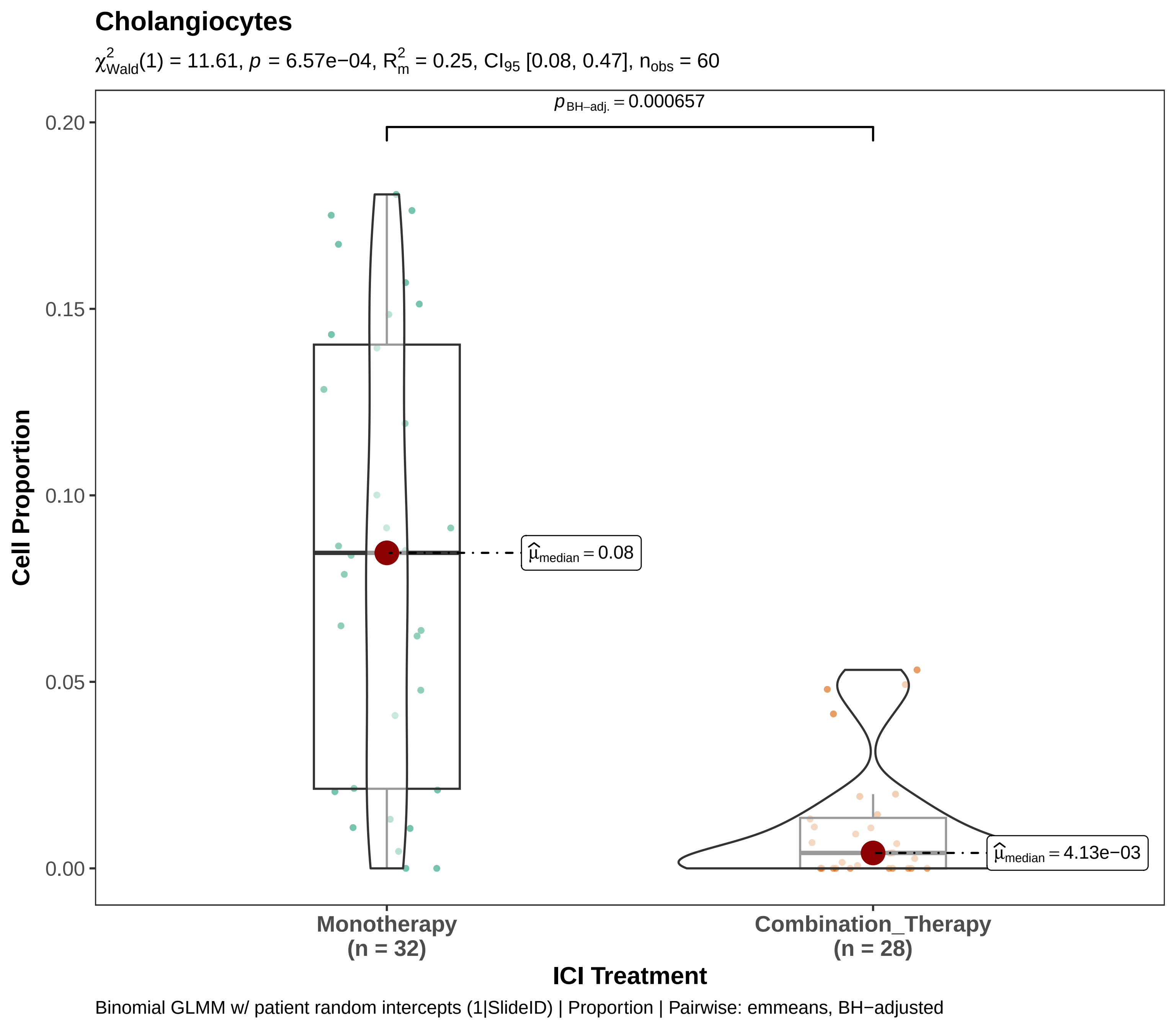

Cholangiocytes
$\chi^2_{Wald}(1) = 11.61$, $p = 6.57e{-}04$, $R^2_m = 0.25$, $CI_{95}$ [0.08, 0.47], $n_{obs} = 60$
$p_{BH-adj.} = 0.000657$
$\widehat{\mu}_{median} = 0.08$
$\widehat{\mu}_{median} = 4.13e{-}03$
0.20
0.15
0.10
0.05
0.00
Cell Proportion
Monotherapy
(n = 32)
Combination_Therapy
(n = 28)
ICI Treatment
Binomial GLMM w/ patient random intercepts (1|SlideID) | Proportion | Pairwise: emmeans, BH−adjusted

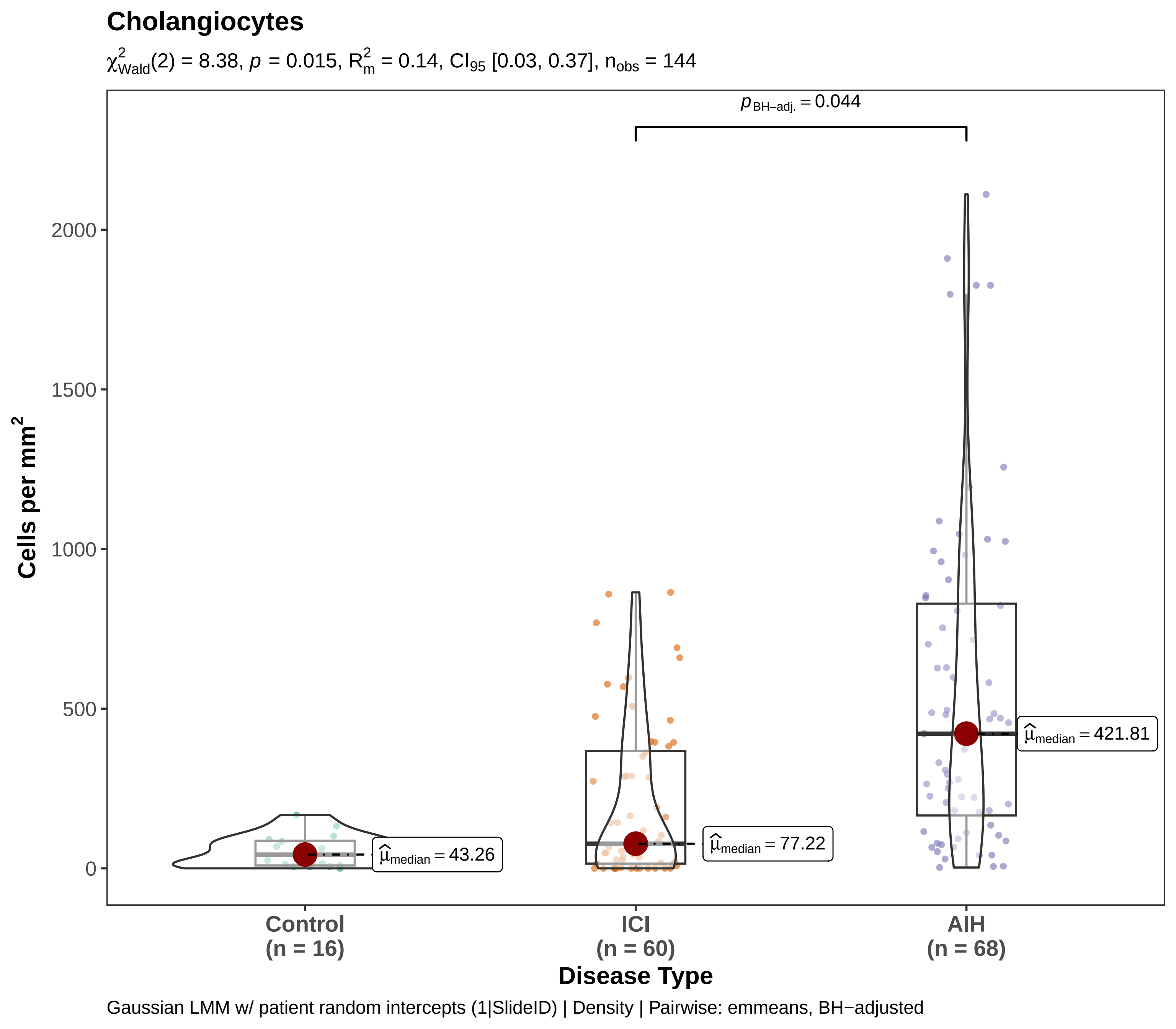

Cholangiocytes
$\chi^2_{Wald}(2) = 8.38$, $p = 0.015$, $R^2_m = 0.14$, $CI_{95}$ [0.03, 0.37], $n_{obs} = 144$
$p_{BH-adj.} = 0.044$
Cells per mm$^2$
2000
1500
1000
500
0
$\hat{\mu}_{median} = 43.26$
$\hat{\mu}_{median} = 77.22$
$\hat{\mu}_{median} = 421.81$
Control
(n = 16)
ICI
(n = 60)
AIH
(n = 68)
Disease Type
Gaussian LMM w/ patient random intercepts (1|SlideID) | Density | Pairwise: emmeans, BH−adjusted

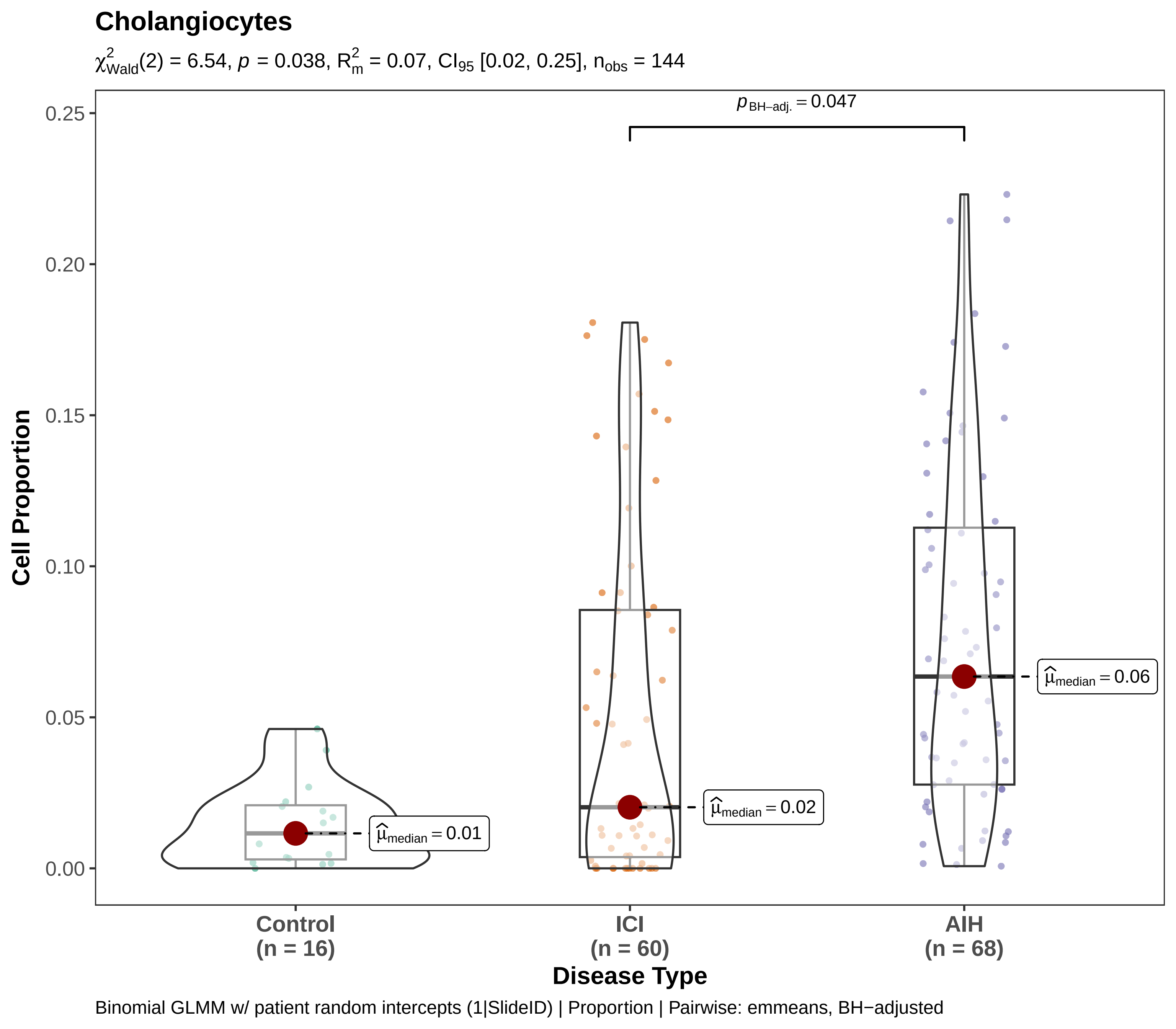
Cholangiocytes
$\chi^2_{Wald}(2) = 6.54$, $p = 0.038$, $R^2_m = 0.07$, $CI_{95}$ [0.02, 0.25], $n_{obs} = 144$
$p_{BH-adj.} = 0.047$
$\widehat{\mu}_{median} = 0.01$
$\widehat{\mu}_{median} = 0.02$
$\widehat{\mu}_{median} = 0.06$
0.25
0.20
0.15
0.10
0.05
0.00
Cell Proportion
Control
(n = 16)
ICI
(n = 60)
AIH
(n = 68)
Disease Type
Binomial GLMM w/ patient random intercepts (1|SlideID) | Proportion | Pairwise: emmeans, BH−adjusted

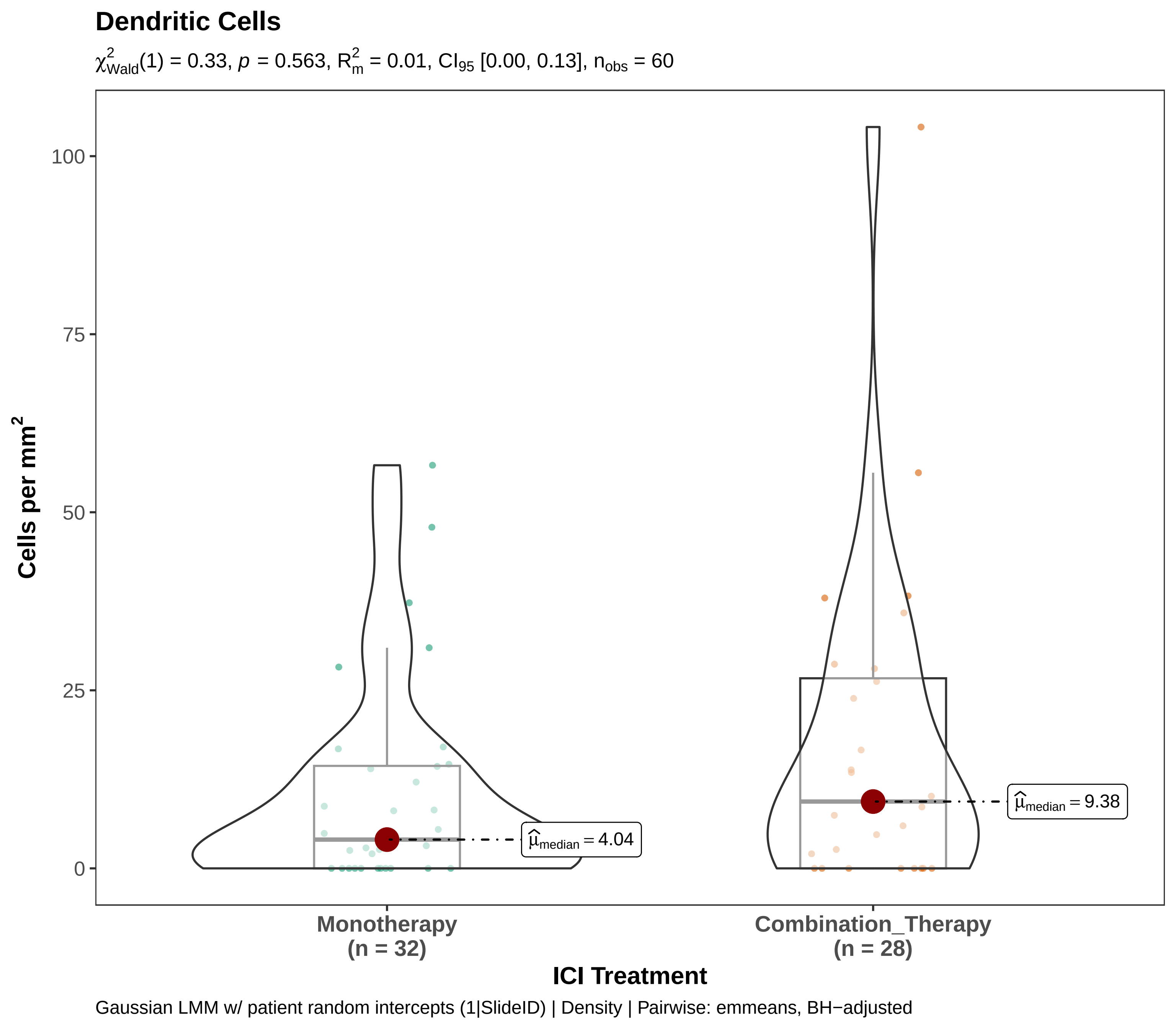
Dendritic Cells
$\chi^2_{Wald}(1) = 0.33$, $p = 0.563$, $R^2_m = 0.01$, $CI_{95}$ [0.00, 0.13], $n_{obs} = 60$
Cells per mm$^2$
100
75
50
25
0
$\hat{\mu}_{median} = 4.04$
$\hat{\mu}_{median} = 9.38$
Monotherapy
(n = 32)
Combination_Therapy
(n = 28)
ICI Treatment
Gaussian LMM w/ patient random intercepts (1|SlideID) | Density | Pairwise: emmeans, BH−adjusted

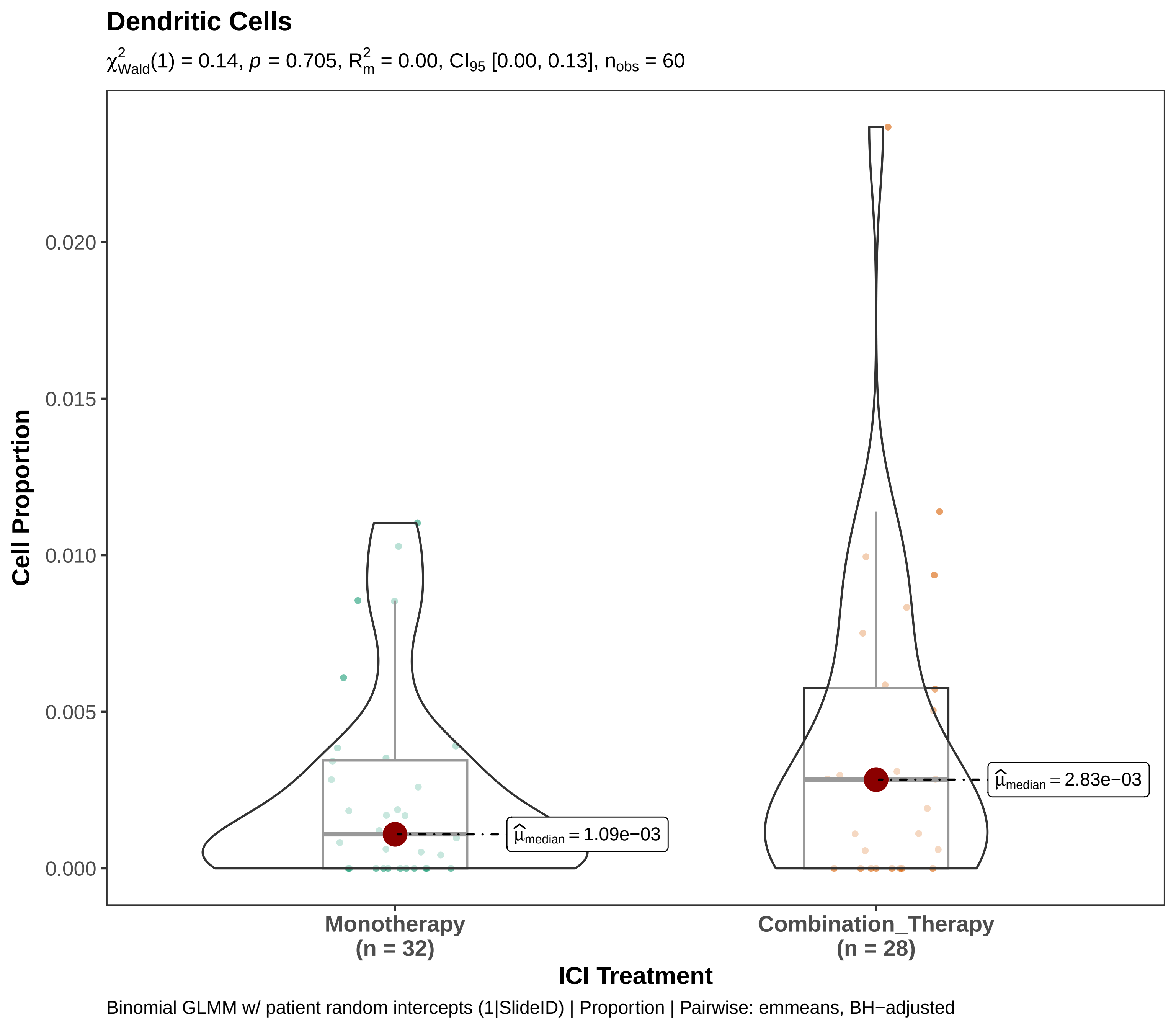

Dendritic Cells
$\chi^2_{Wald}(1) = 0.14$, $p = 0.705$, $R^2_m = 0.00$, $CI_{95}$ [0.00, 0.13], $n_{obs} = 60$
Cell Proportion
0.020
0.015
0.010
0.005
0.000
$\hat{\mu}_{median} = 1.09e{-}03$
$\hat{\mu}_{median} = 2.83e{-}03$
Monotherapy
(n = 32)
Combination_Therapy
(n = 28)
ICI Treatment
Binomial GLMM w/ patient random intercepts (1|SlideID) | Proportion | Pairwise: emmeans, BH−adjusted

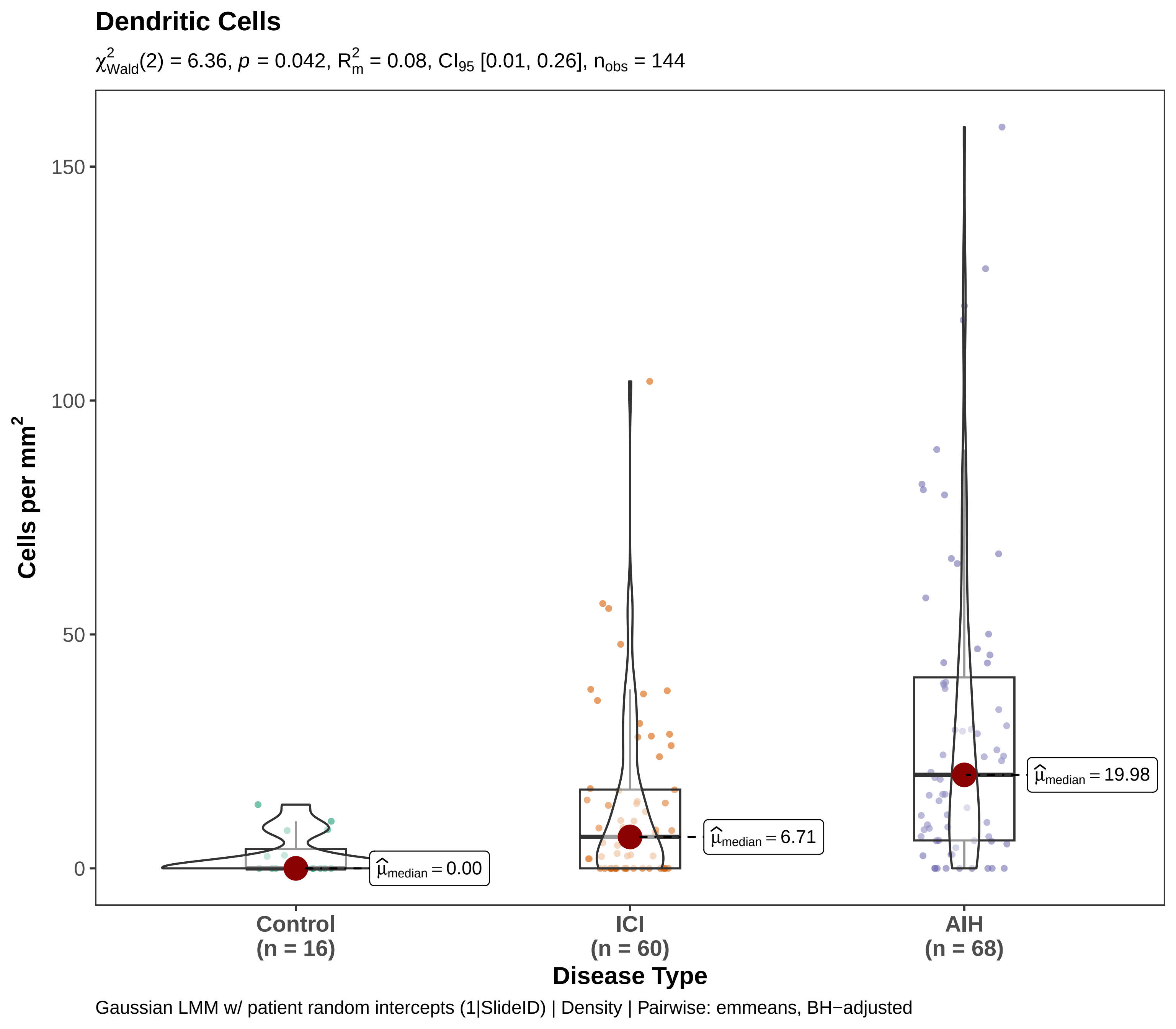
Dendritic Cells
$\chi^2_{Wald}(2) = 6.36$, $p = 0.042$, $R^2_m = 0.08$, $CI_{95}$ [0.01, 0.26], $n_{obs} = 144$
Cells per mm$^2$
150
100
50
0
$\hat{\mu}_{median} = 0.00$
$\hat{\mu}_{median} = 6.71$
$\hat{\mu}_{median} = 19.98$
Control
(n = 16)
ICI
(n = 60)
AIH
(n = 68)
Disease Type
Gaussian LMM w/ patient random intercepts (1|SlideID) | Density | Pairwise: emmeans, BH−adjusted

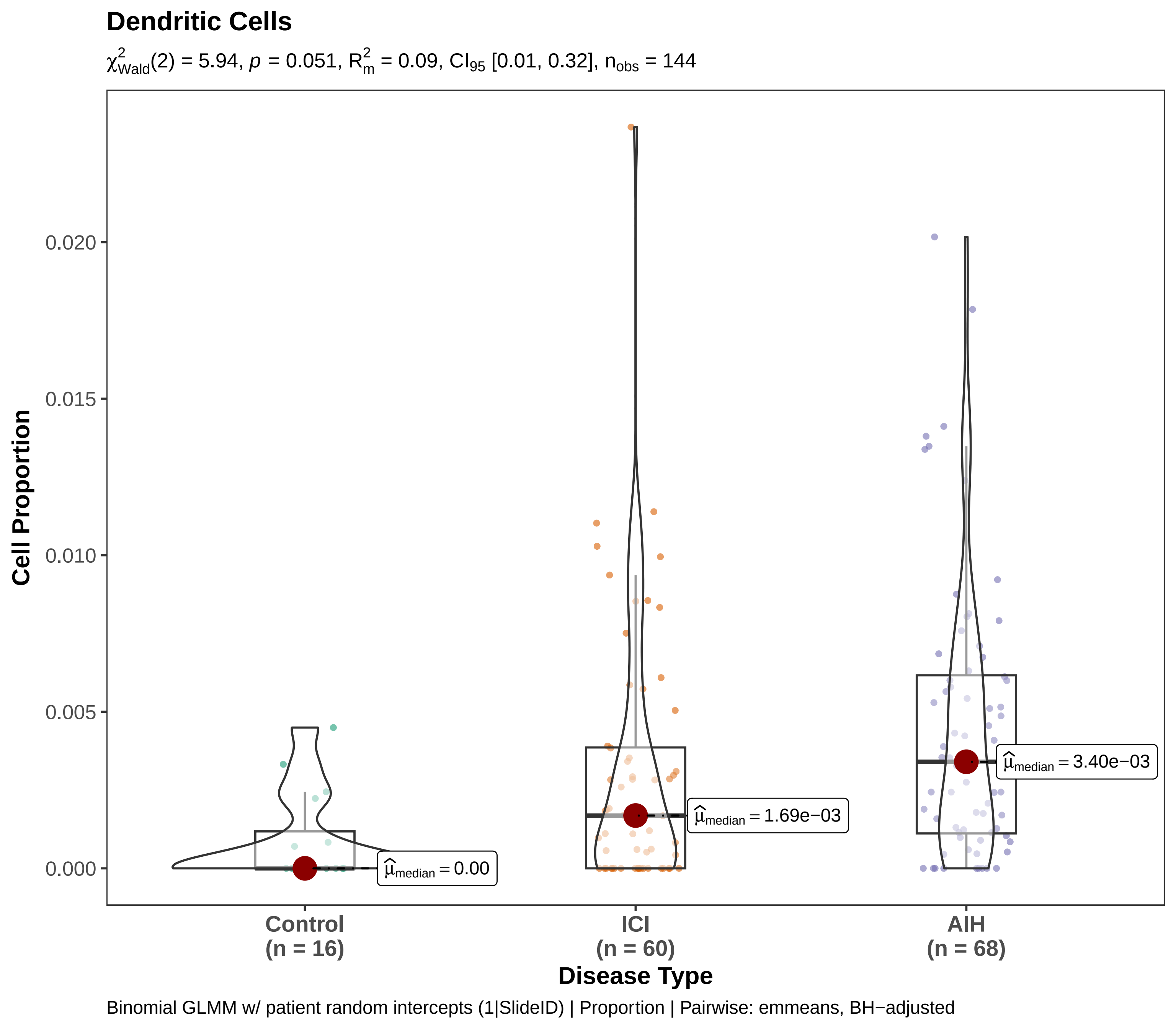
Dendritic Cells
$\chi^2_{Wald}(2) = 5.94$, $p = 0.051$, $R^2_m = 0.09$, $CI_{95}$ [0.01, 0.32], $n_{obs} = 144$
Cell Proportion
0.020
0.015
0.010
0.005
0.000
$\widehat{\mu}_{median} = 0.00$
$\widehat{\mu}_{median} = 1.69e{-}03$
$\widehat{\mu}_{median} = 3.40e{-}03$
Control
(n = 16)
ICI
(n = 60)
AIH
(n = 68)
Disease Type
Binomial GLMM w/ patient random intercepts (1|SlideID) | Proportion | Pairwise: emmeans, BH−adjusted

### Endothelium

$\chi^2_{Wald}(1) = 3.38$, $p = 0.066$, $R^2_m = 0.09$, $CI_{95}$ [0.00, 0.29], $n_{obs} = 60$

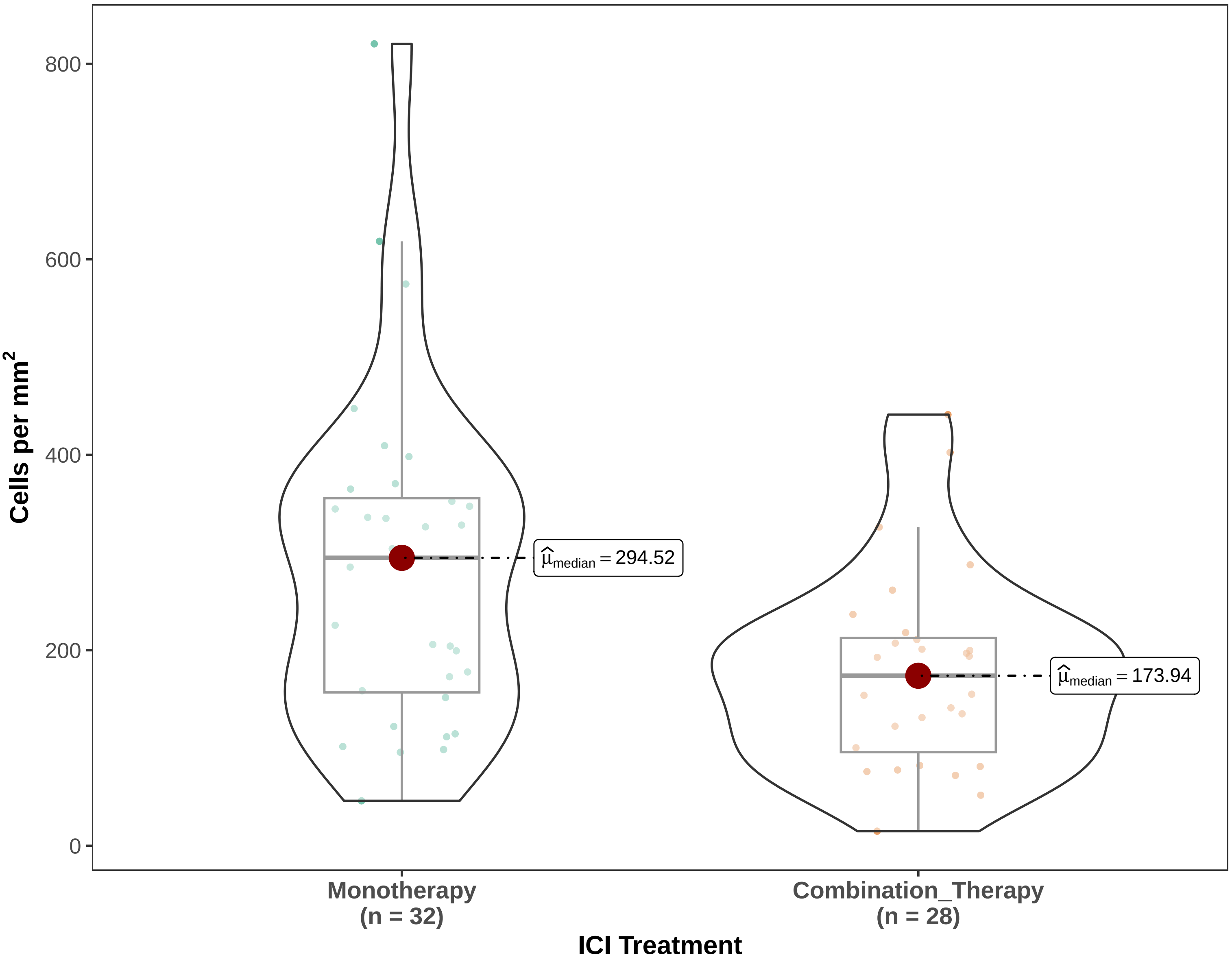


Gaussian LMM w/ patient random intercepts (1|SlideID) | Density | Pairwise: emmeans, BH−adjusted

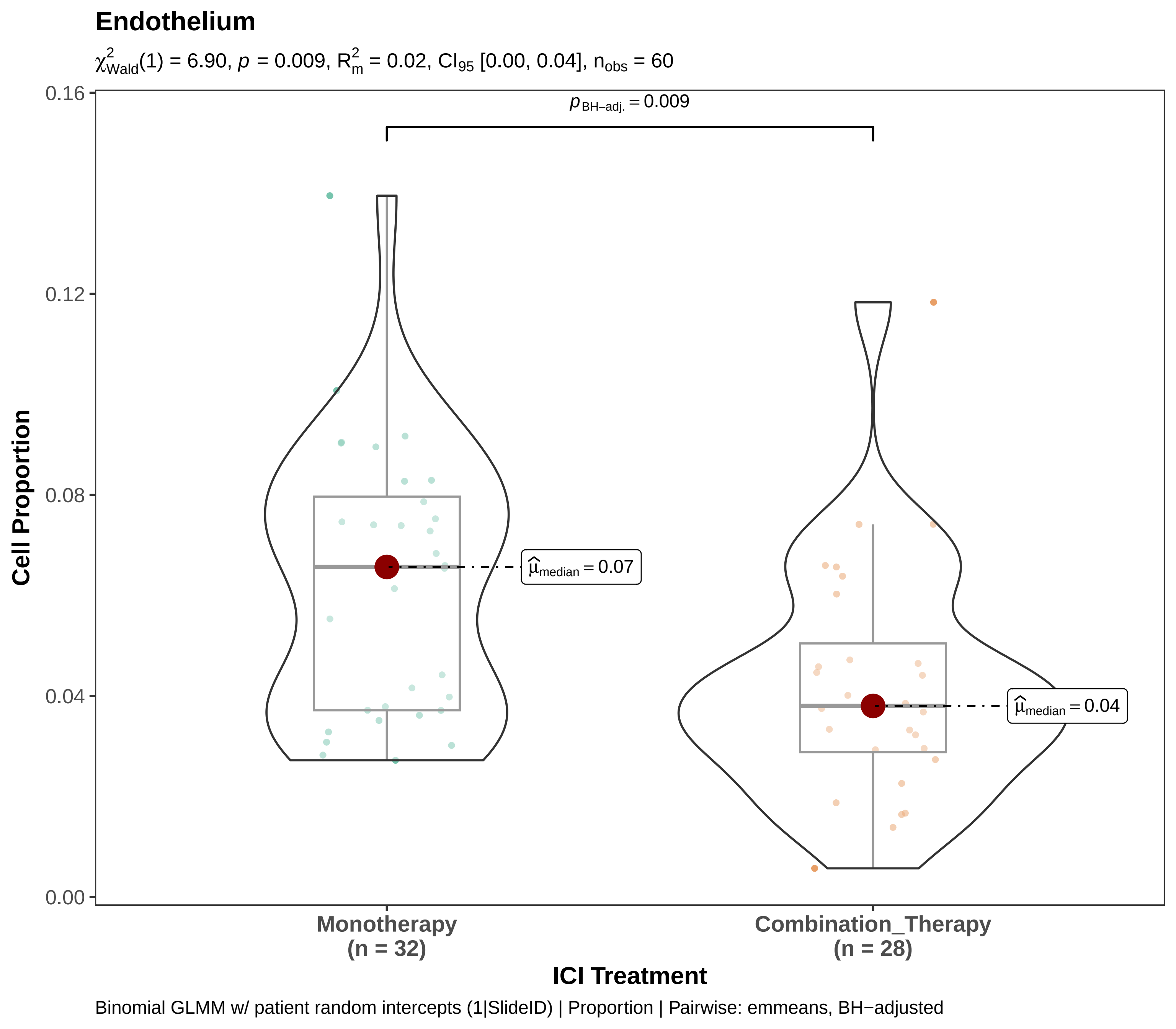

Endothelium
$\chi^2_{Wald}(1) = 6.90$, $p = 0.009$, $R^2_m = 0.02$, $CI_{95}$ [0.00, 0.04], $n_{obs} = 60$
$p_{BH-adj.} = 0.009$
$\widehat{\mu}_{median} = 0.07$
$\widehat{\mu}_{median} = 0.04$
0.16
0.12
0.08
0.04
0.00
Cell Proportion
Monotherapy
(n = 32)
Combination_Therapy
(n = 28)
ICI Treatment
Binomial GLMM w/ patient random intercepts (1|SlideID) | Proportion | Pairwise: emmeans, BH−adjusted

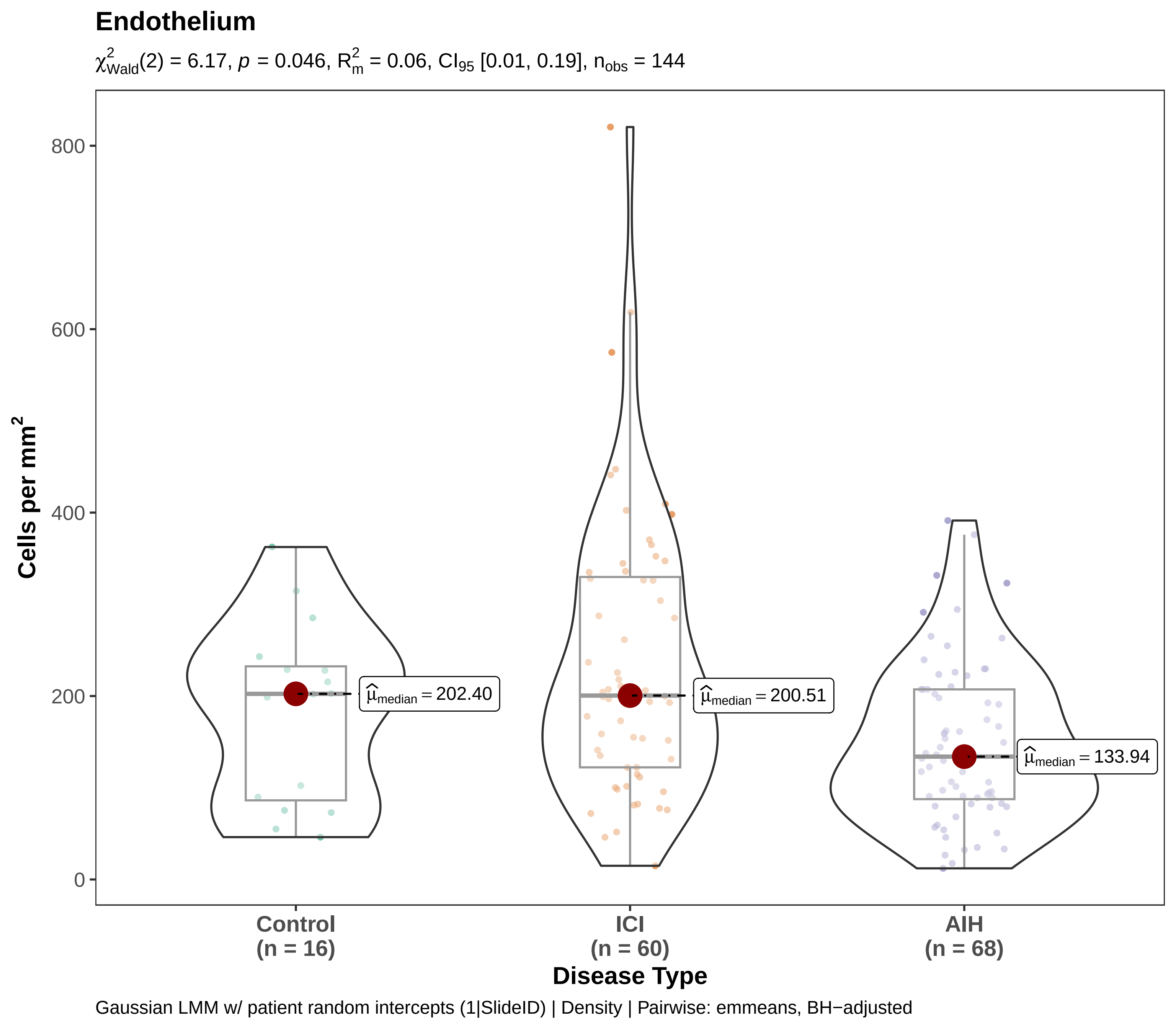

Endothelium
$\chi^2_{Wald}(2) = 6.17$, $p = 0.046$, $R^2_m = 0.06$, $CI_{95}$ [0.01, 0.19], $n_{obs} = 144$
Cells per mm²
800
600
400
200
0
$\widehat{\mu}_{median} = 202.40$
$\widehat{\mu}_{median} = 200.51$
$\widehat{\mu}_{median} = 133.94$
Control
(n = 16)
ICI
(n = 60)
AIH
(n = 68)
Disease Type
Gaussian LMM w/ patient random intercepts (1|SlideID) | Density | Pairwise: emmeans, BH−adjusted

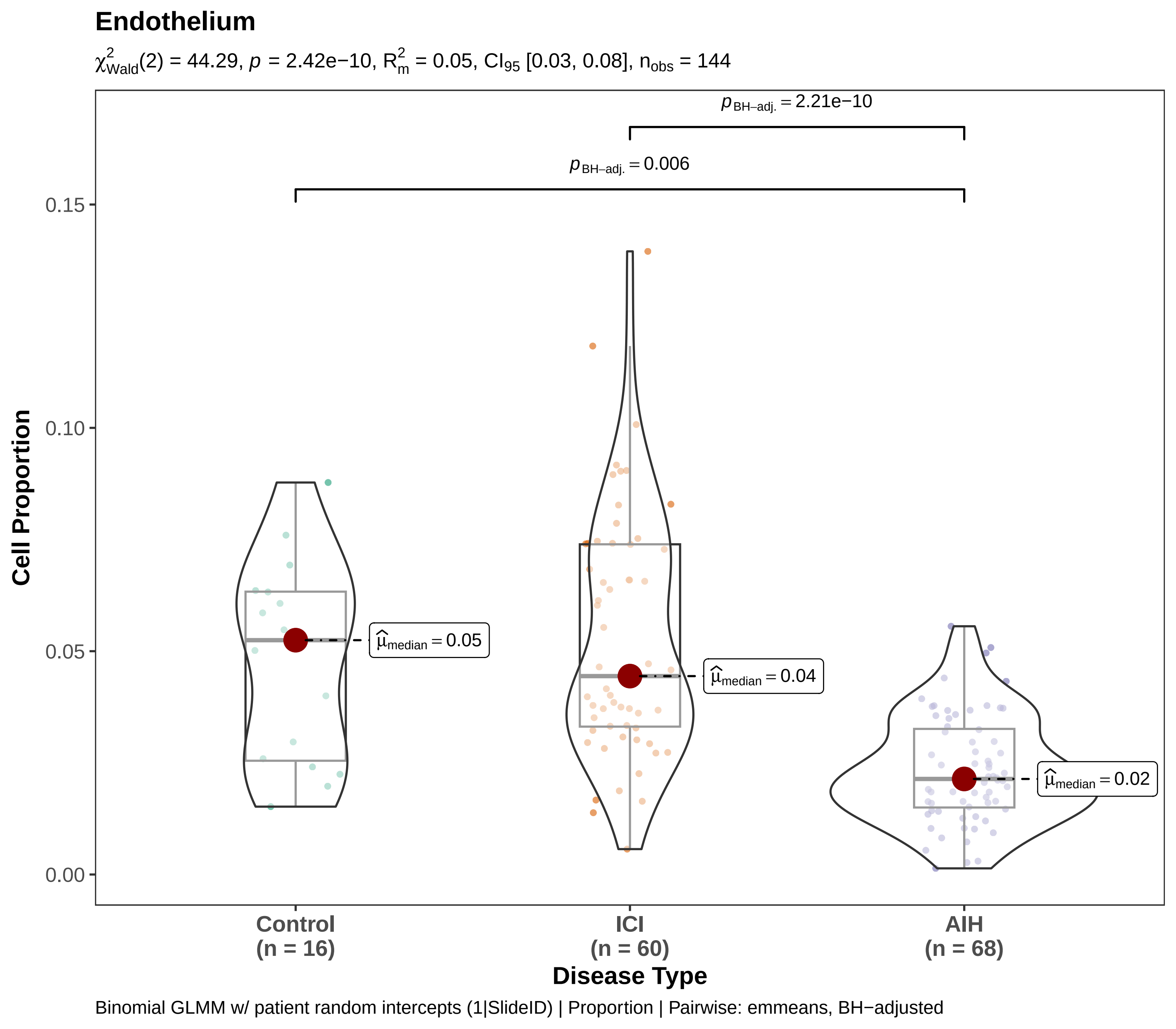
Endothelium
$\chi^2_{Wald}(2) = 44.29$, $p = 2.42e{-10}$, $R^2_m = 0.05$, $CI_{95}$ [0.03, 0.08], $n_{obs} = 144$
$p_{BH-adj.} = 2.21e{-10}$
$p_{BH-adj.} = 0.006$
$\hat{\mu}_{median} = 0.05$
$\hat{\mu}_{median} = 0.04$
$\hat{\mu}_{median} = 0.02$
Cell Proportion
0.15
0.10
0.05
0.00
Control
(n = 16)
ICI
(n = 60)
AIH
(n = 68)
Disease Type
Binomial GLMM w/ patient random intercepts (1|SlideID) | Proportion | Pairwise: emmeans, BH−adjusted

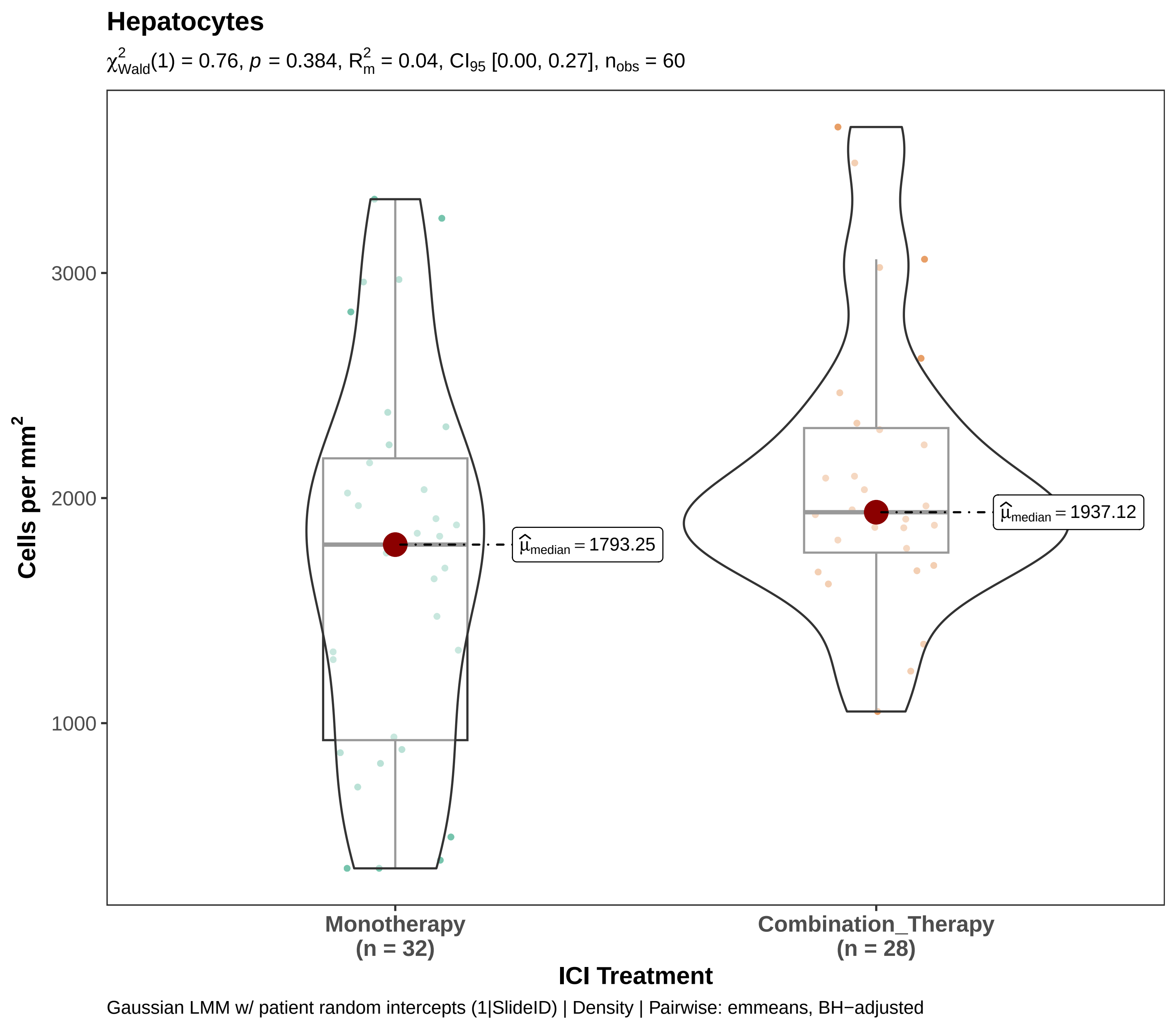
Hepatocytes
$\chi^2_{Wald}(1) = 0.76$, $p = 0.384$, $R^2_m = 0.04$, $CI_{95}$ [0.00, 0.27], $n_{obs} = 60$
Cells per mm²
3000
2000
1000
$\widehat{\mu}_{median} = 1793.25$
$\widehat{\mu}_{median} = 1937.12$
Monotherapy
(n = 32)
Combination_Therapy
(n = 28)
ICI Treatment
Gaussian LMM w/ patient random intercepts (1|SlideID) | Density | Pairwise: emmeans, BH−adjusted

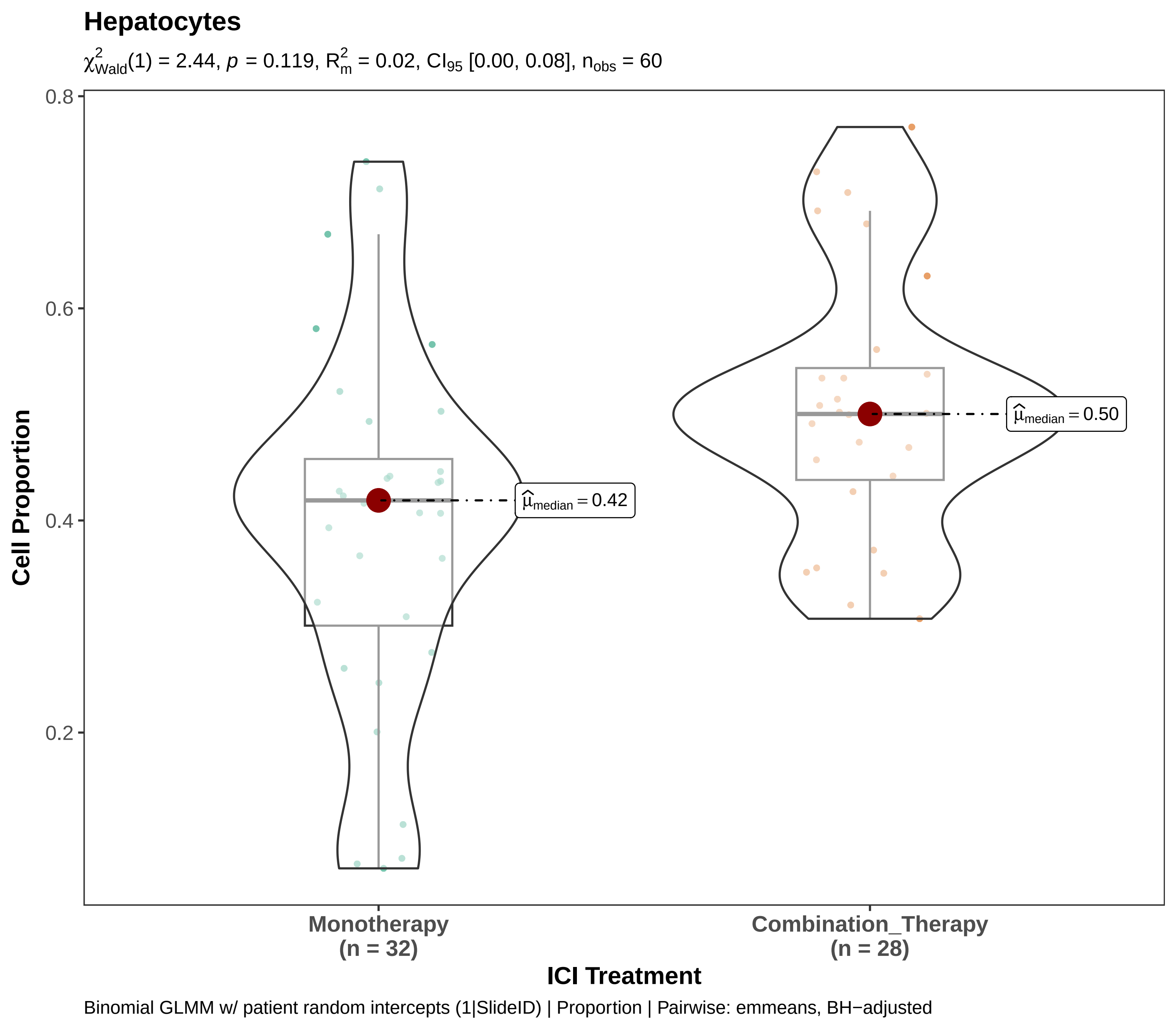
Hepatocytes
$\chi^2_{Wald}(1) = 2.44$, $p = 0.119$, $R^2_m = 0.02$, $CI_{95}$ [0.00, 0.08], $n_{obs} = 60$
Cell Proportion
0.8
0.6
0.4
0.2
$\widehat{\mu}_{median} = 0.42$
$\widehat{\mu}_{median} = 0.50$
Monotherapy
(n = 32)
Combination_Therapy
(n = 28)
ICI Treatment
Binomial GLMM w/ patient random intercepts (1|SlideID) | Proportion | Pairwise: emmeans, BH−adjusted

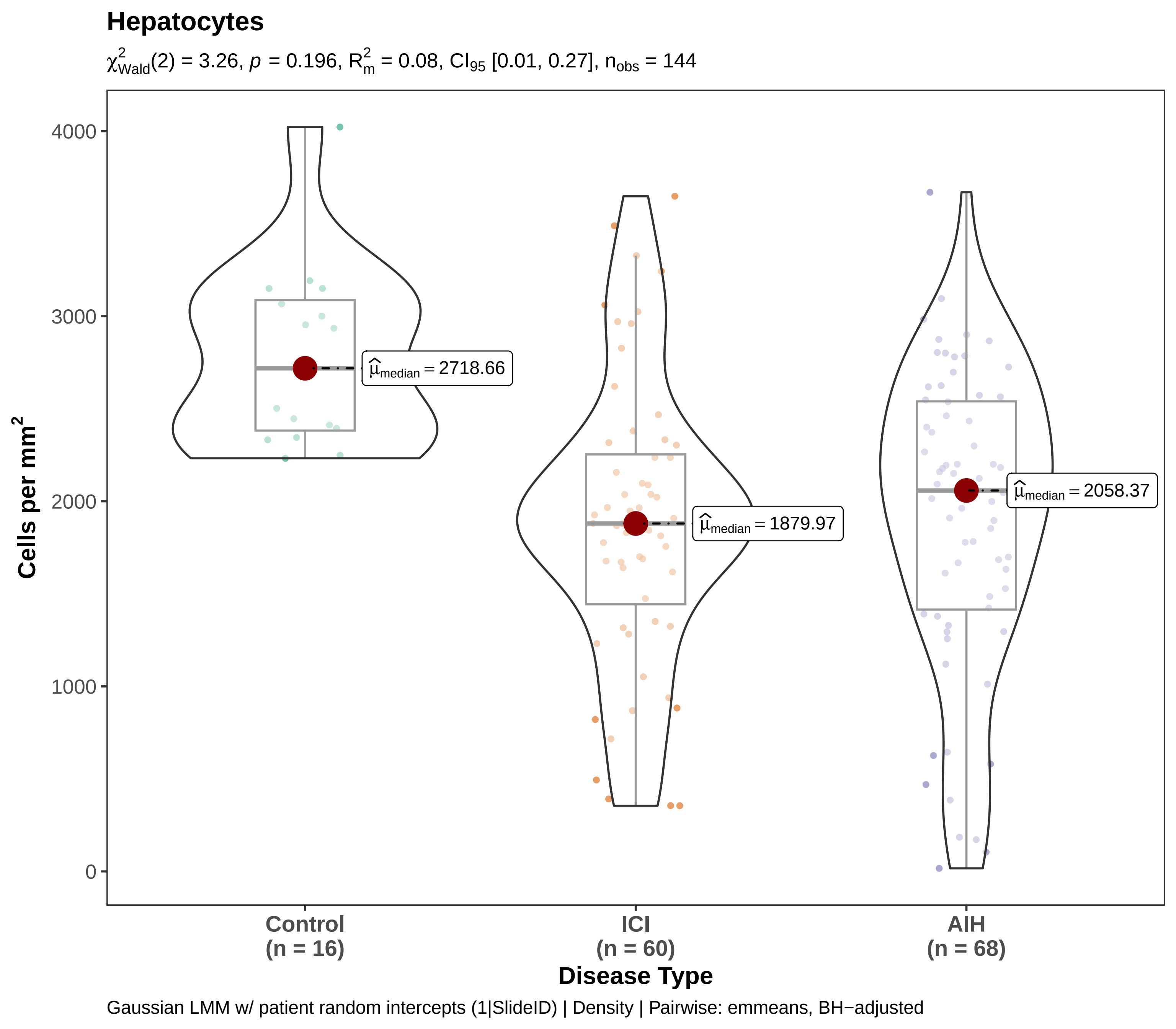

Hepatocytes
$\chi^2_{\text{Wald}}(2) = 3.26$, $p = 0.196$, $R^2_m = 0.08$, $CI_{95}$ [0.01, 0.27], $n_{obs} = 144$
Cells per mm$^2$
4000
3000
2000
1000
0
$\widehat{\mu}_{median} = 2718.66$
$\widehat{\mu}_{median} = 1879.97$
$\widehat{\mu}_{median} = 2058.37$
Control
(n = 16)
ICI
(n = 60)
AIH
(n = 68)
Disease Type
Gaussian LMM w/ patient random intercepts (1|SlideID) | Density | Pairwise: emmeans, BH−adjusted

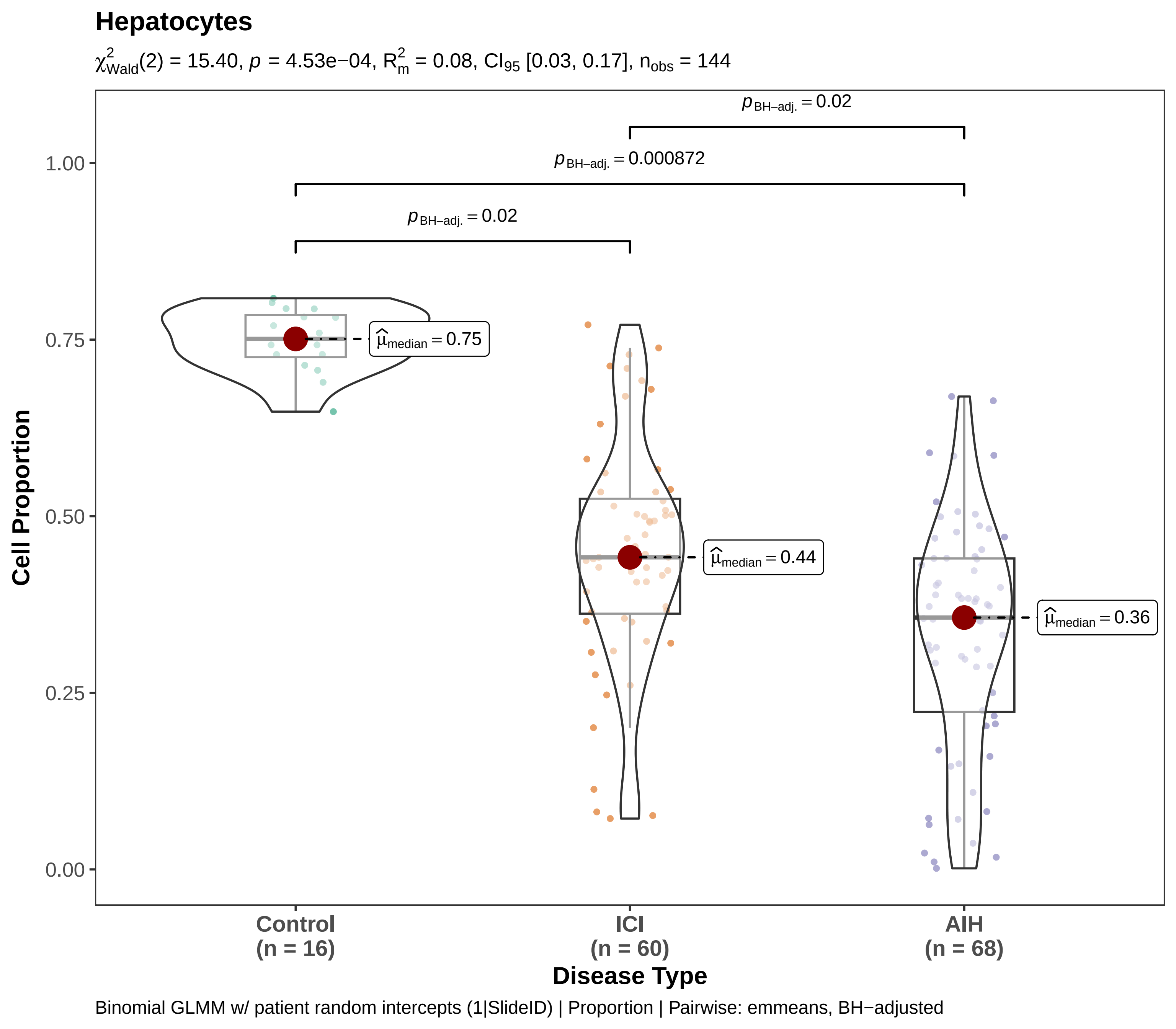
Hepatocytes
$\chi^2_{Wald}(2) = 15.40$, $p = 4.53e-04$, $R^2_m = 0.08$, $CI_{95}$ [0.03, 0.17], $n_{obs} = 144$
$p_{BH-adj.} = 0.02$
$p_{BH-adj.} = 0.000872$
$p_{BH-adj.} = 0.02$
$\hat{\mu}_{median} = 0.75$
$\hat{\mu}_{median} = 0.44$
$\hat{\mu}_{median} = 0.36$
1.00
0.75
0.50
0.25
0.00
Cell Proportion
Control (n = 16)
ICI (n = 60)
AIH (n = 68)
Disease Type
Binomial GLMM w/ patient random intercepts (1|SlideID) | Proportion | Pairwise: emmeans, BH−adjusted

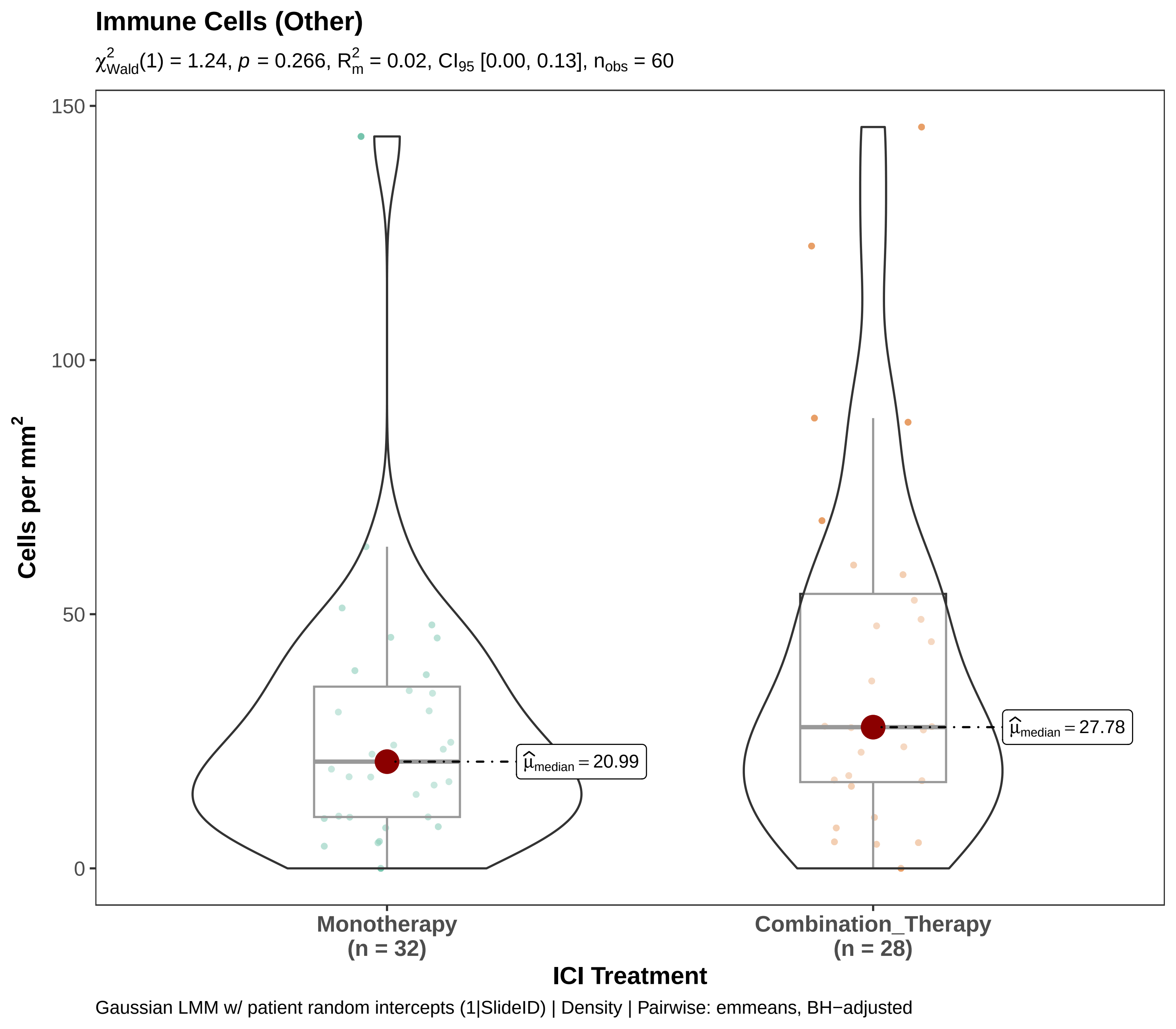
Immune Cells (Other)
$\chi^2_{Wald}(1) = 1.24$, $p = 0.266$, $R^2_m = 0.02$, $CI_{95}$ [0.00, 0.13], $n_{obs} = 60$
150
100
50
0
Cells per mm$^2$
$\widehat{\mu}_{median} = 20.99$
$\widehat{\mu}_{median} = 27.78$
Monotherapy
(n = 32)
Combination_Therapy
(n = 28)
ICI Treatment
Gaussian LMM w/ patient random intercepts (1|SlideID) | Density | Pairwise: emmeans, BH−adjusted

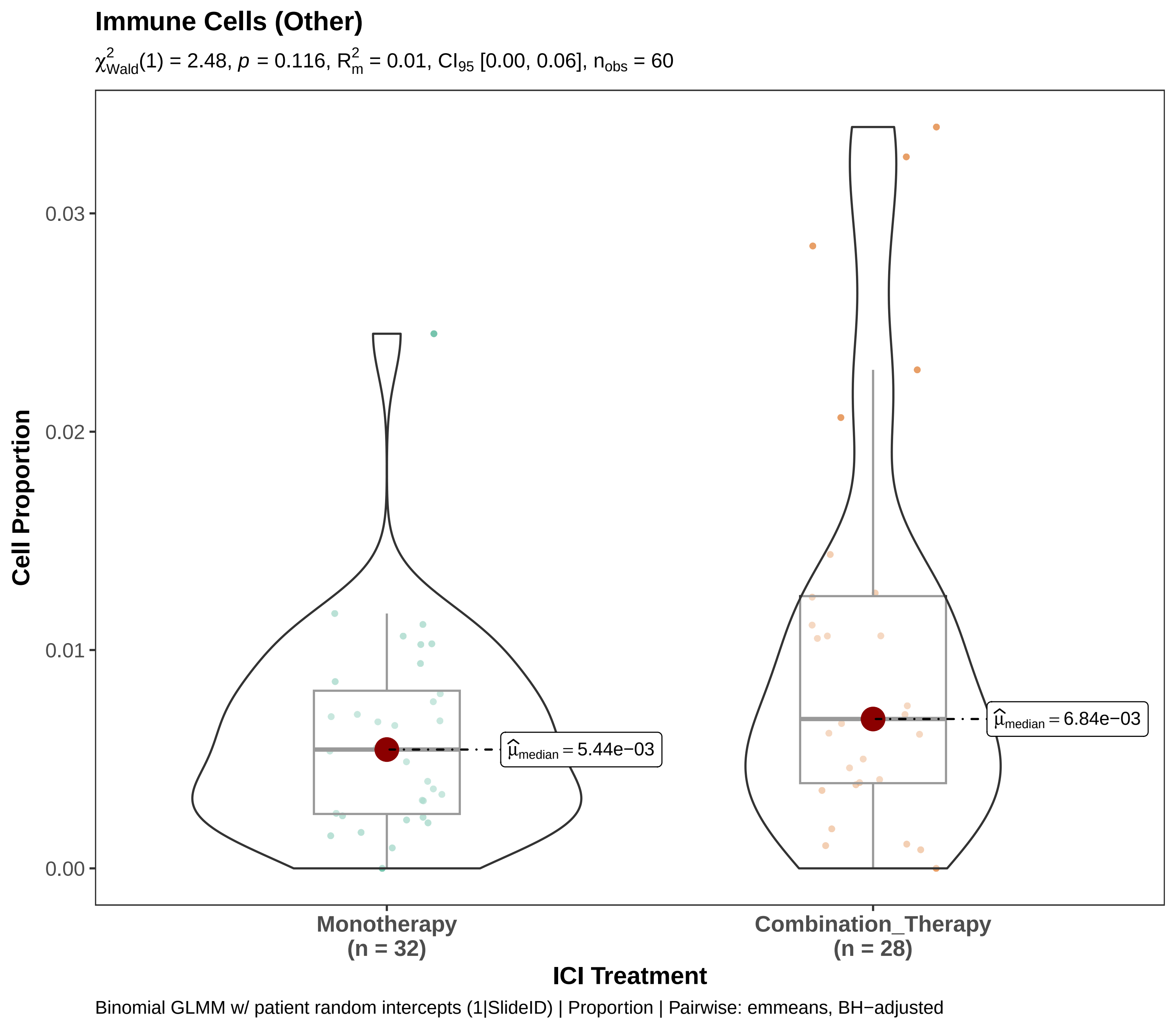
Immune Cells (Other)
$\chi^2_{Wald}(1) = 2.48$, $p = 0.116$, $R^2_m = 0.01$, $CI_{95}$ [0.00, 0.06], $n_{obs} = 60$
Cell Proportion
0.03
0.02
0.01
0.00
$\widehat{\mu}_{median} = 5.44e{-}03$
$\widehat{\mu}_{median} = 6.84e{-}03$
Monotherapy
(n = 32)
Combination_Therapy
(n = 28)
ICI Treatment
Binomial GLMM w/ patient random intercepts (1|SlideID) | Proportion | Pairwise: emmeans, BH−adjusted

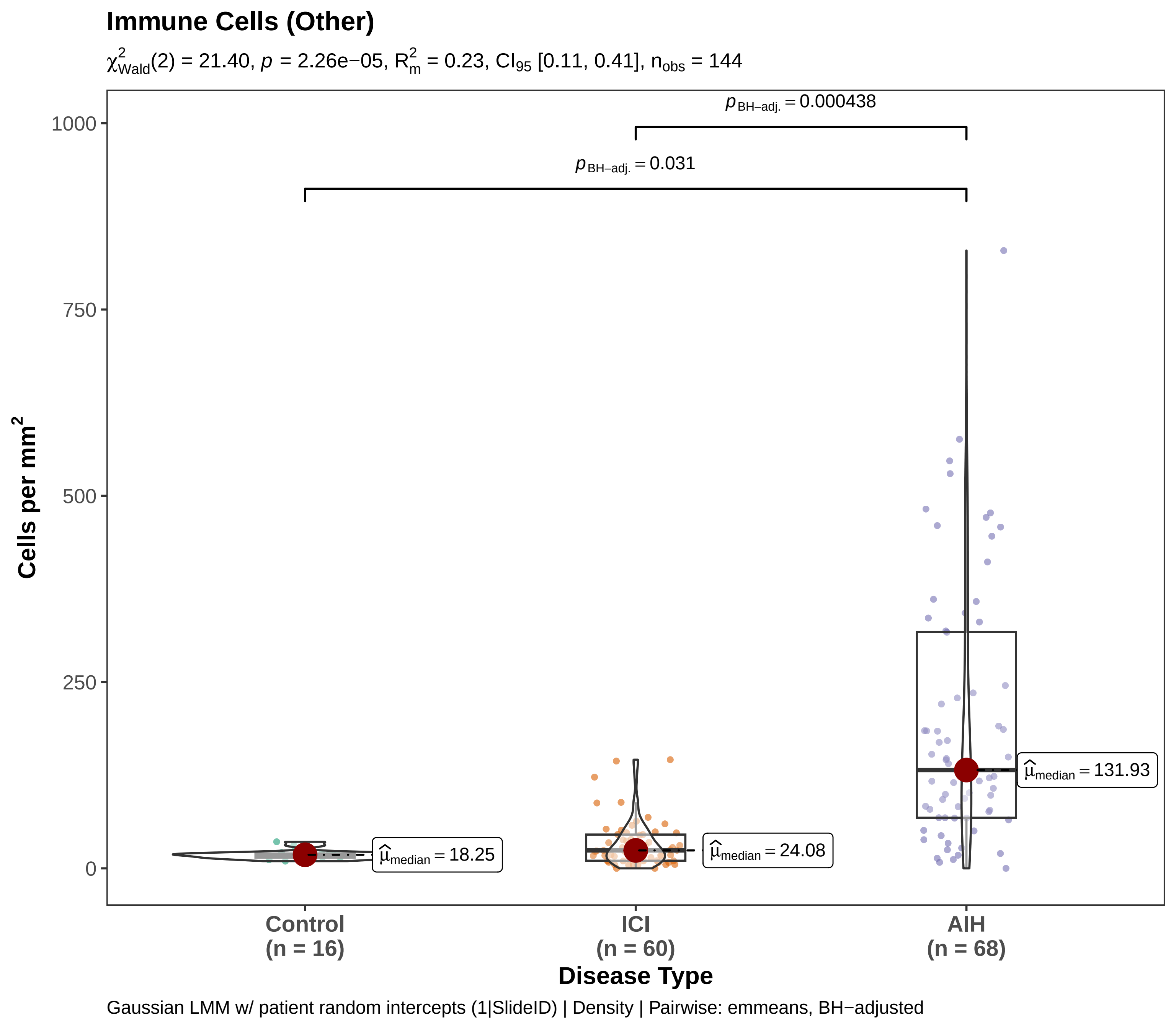

Immune Cells (Other)
$\chi^2_{Wald}(2) = 21.40$, $p = 2.26e{-}05$, $R^2_m = 0.23$, $CI_{95}$ [0.11, 0.41], $n_{obs} = 144$
$p_{BH-adj.} = 0.000438$
$p_{BH-adj.} = 0.031$
1000
750
500
250
0
Cells per mm$^2$
$\hat{\mu}_{median} = 18.25$
$\hat{\mu}_{median} = 24.08$
$\hat{\mu}_{median} = 131.93$
Control
(n = 16)
ICI
(n = 60)
AIH
(n = 68)
Disease Type
Gaussian LMM w/ patient random intercepts (1|SlideID) | Density | Pairwise: emmeans, BH−adjusted

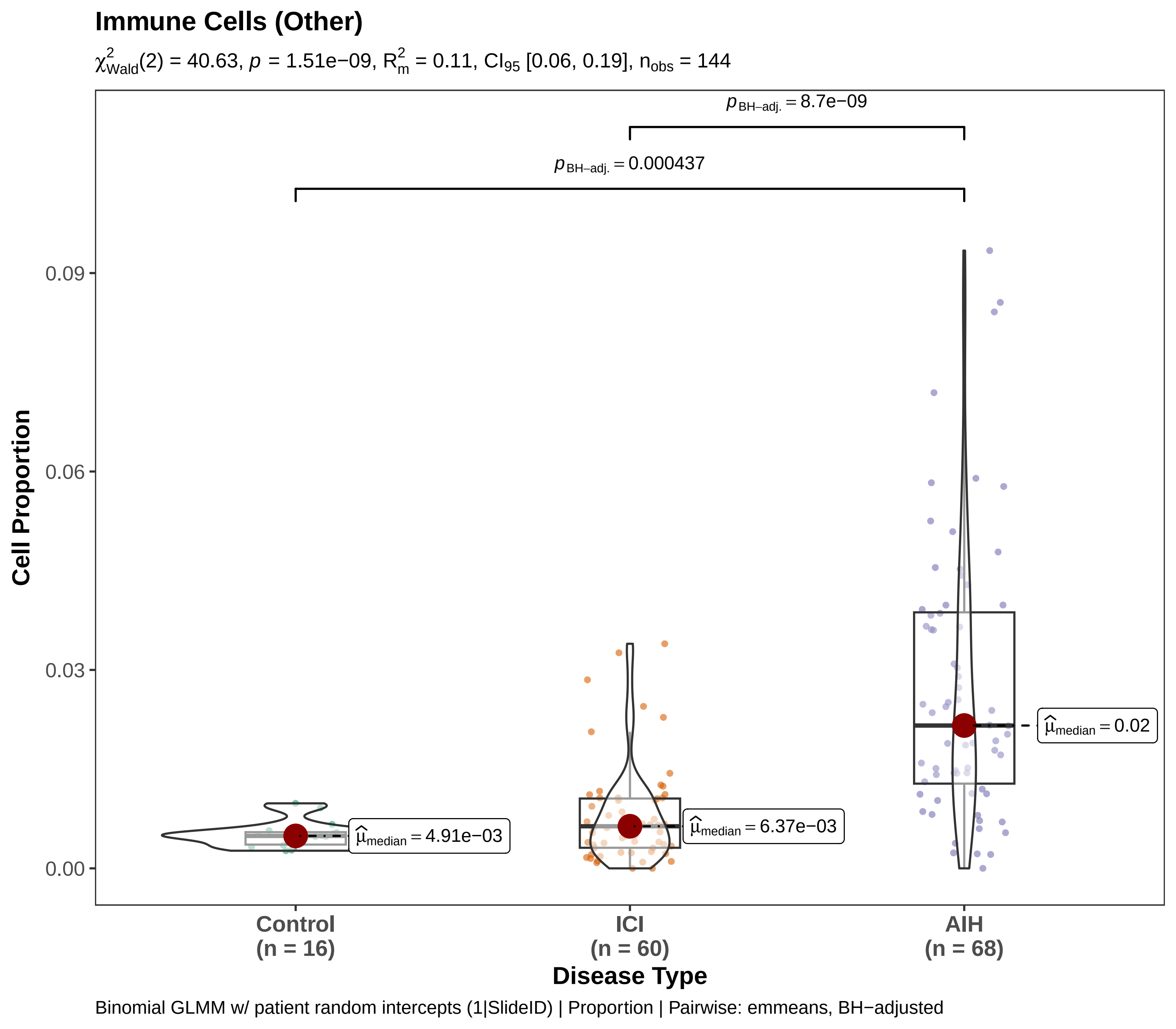

Immune Cells (Other)
$\chi^2_{Wald}(2)$ = 40.63, $p$ = 1.51e−09, $R^2_m$ = 0.11, $CI_{95}$ [0.06, 0.19], $n_{obs}$ = 144
$p_{BH-adj.} = 8.7e-09$
$p_{BH-adj.} = 0.000437$
0.09
0.06
0.03
0.00
Cell Proportion
$\hat{\mu}_{median} = 0.02$
$\hat{\mu}_{median} = 6.37e-03$
$\hat{\mu}_{median} = 4.91e-03$
Control
(n = 16)
ICI
(n = 60)
AIH
(n = 68)
Disease Type
Binomial GLMM w/ patient random intercepts (1|SlideID) | Proportion | Pairwise: emmeans, BH−adjusted

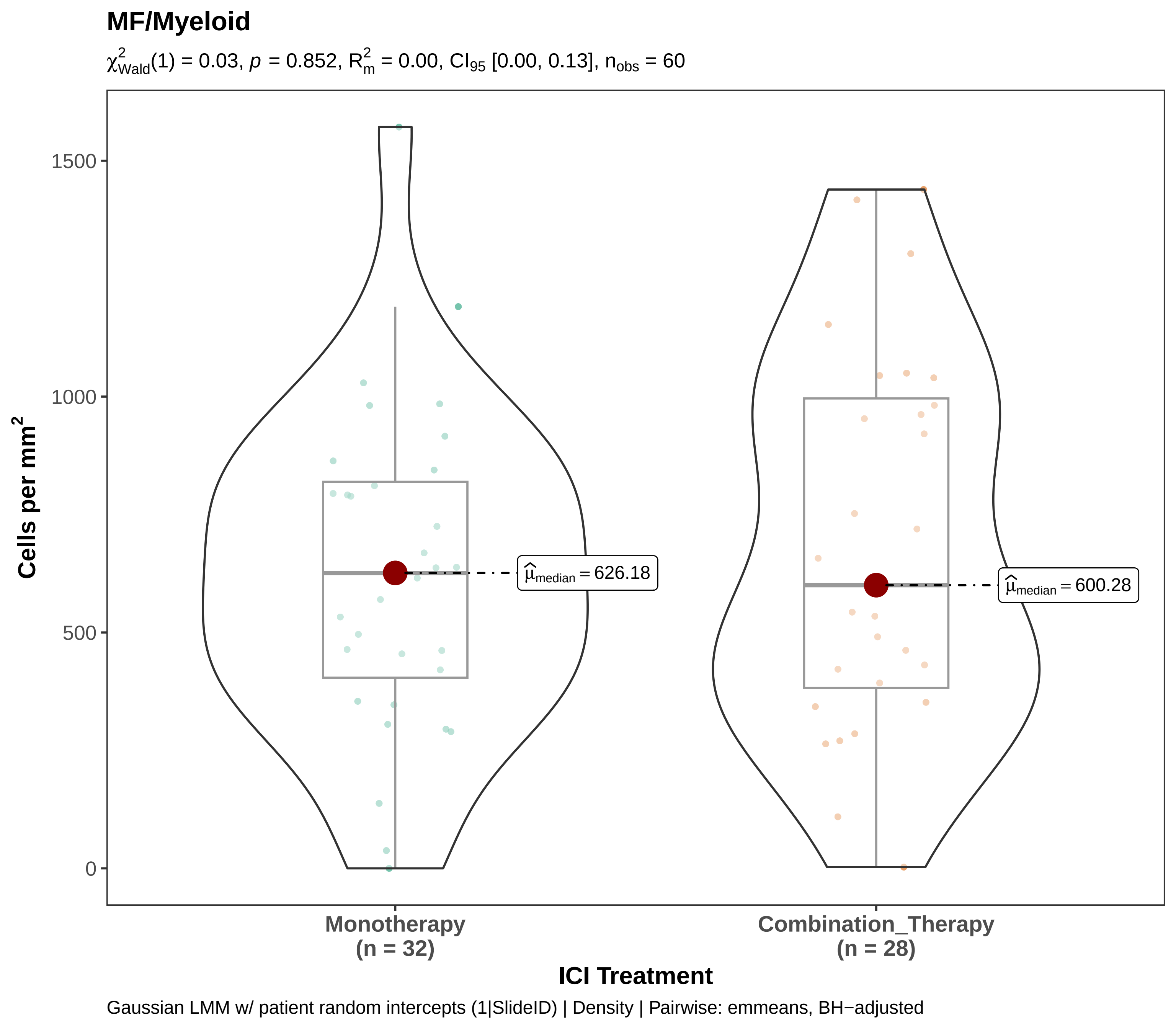
MF/Myeloid
$\chi^2_{Wald}(1) = 0.03$, $p = 0.852$, $R^2_m = 0.00$, $CI_{95}$ [0.00, 0.13], $n_{obs} = 60$
Cells per mm$^2$
1500
1000
500
0
$\widehat{\mu}_{median} = 626.18$
$\widehat{\mu}_{median} = 600.28$
Monotherapy
(n = 32)
Combination_Therapy
(n = 28)
ICI Treatment
Gaussian LMM w/ patient random intercepts (1|SlideID) | Density | Pairwise: emmeans, BH−adjusted

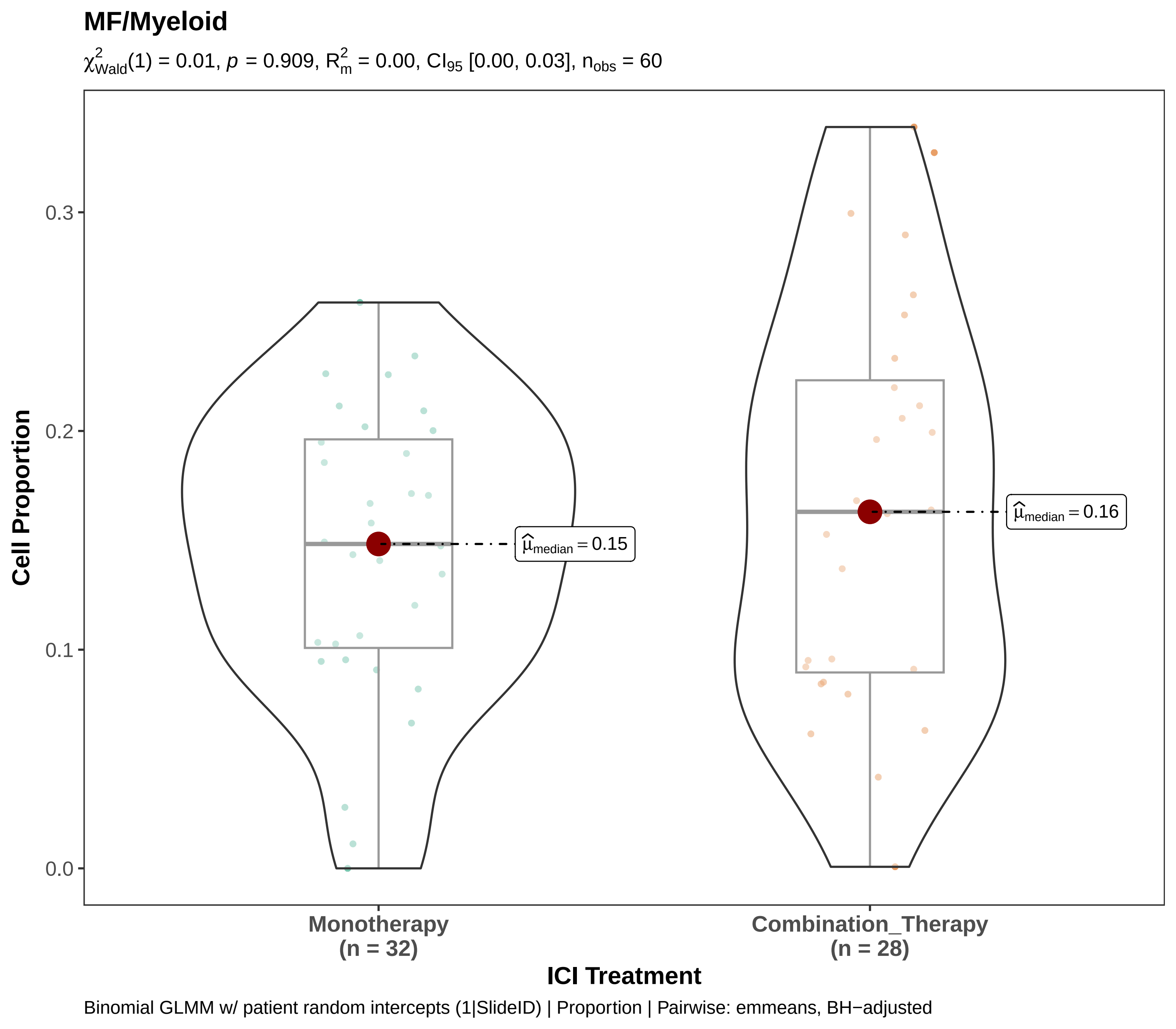
MF/Myeloid
$\chi^2_{Wald}(1) = 0.01$, $p = 0.909$, $R^2_m = 0.00$, $CI_{95}$ [0.00, 0.03], $n_{obs} = 60$
Cell Proportion
0.0
0.1
0.2
0.3
$\hat{\mu}_{median} = 0.15$
$\hat{\mu}_{median} = 0.16$
Monotherapy
(n = 32)
Combination_Therapy
(n = 28)
ICI Treatment
Binomial GLMM w/ patient random intercepts (1|SlideID) | Proportion | Pairwise: emmeans, BH−adjusted

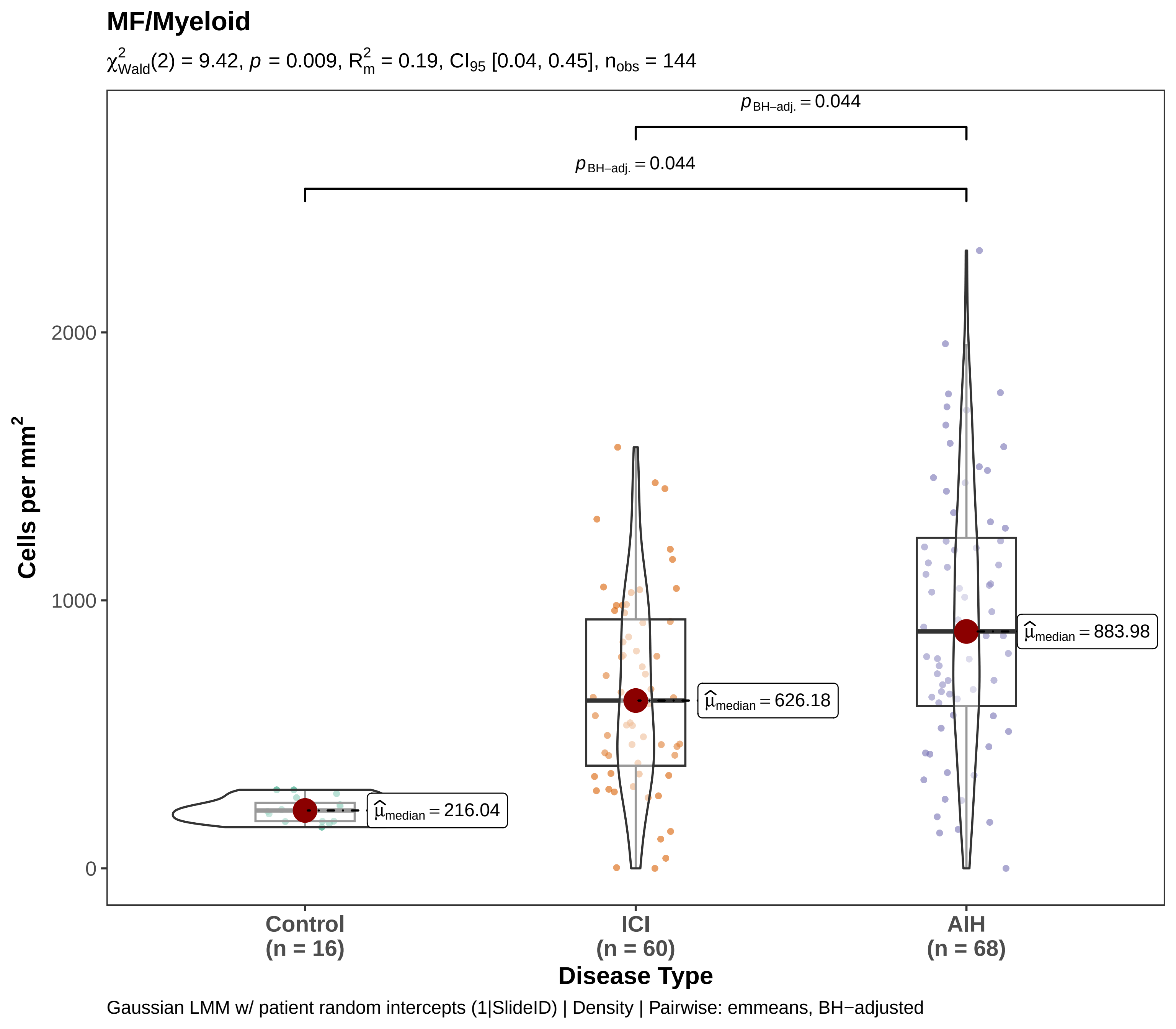

MF/Myeloid
$\chi^2_{Wald}(2) = 9.42$, $p = 0.009$, $R^2_m = 0.19$, $CI_{95}$ [0.04, 0.45], $n_{obs} = 144$
$p_{BH-adj.} = 0.044$
$p_{BH-adj.} = 0.044$
Cells per mm$^2$
2000
1000
0
$\hat{\mu}_{median} = 216.04$
$\hat{\mu}_{median} = 626.18$
$\hat{\mu}_{median} = 883.98$
Control (n = 16)
ICI (n = 60)
AIH (n = 68)
Disease Type
Gaussian LMM w/ patient random intercepts (1|SlideID) | Density | Pairwise: emmeans, BH−adjusted

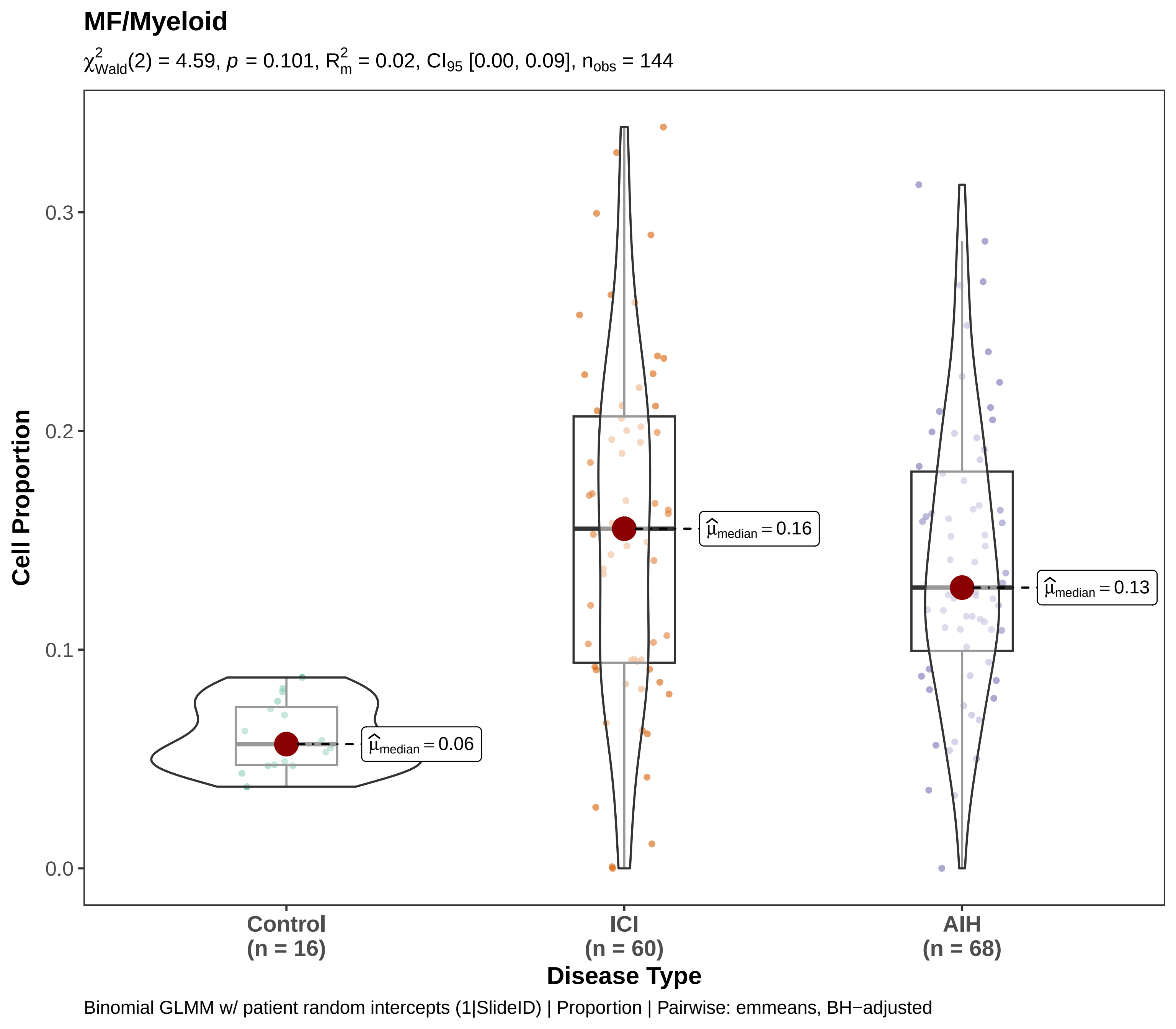
MF/Myeloid
$\chi^2_{Wald}(2) = 4.59$, $p = 0.101$, $R^2_m = 0.02$, $CI_{95}$ [0.00, 0.09], $n_{obs} = 144$
Cell Proportion
0.0
0.1
0.2
0.3
$\hat{\mu}_{median} = 0.06$
$\hat{\mu}_{median} = 0.16$
$\hat{\mu}_{median} = 0.13$
Control (n = 16)
ICI (n = 60)
AIH (n = 68)
Disease Type
Binomial GLMM w/ patient random intercepts (1|SlideID) | Proportion | Pairwise: emmeans, BH−adjusted

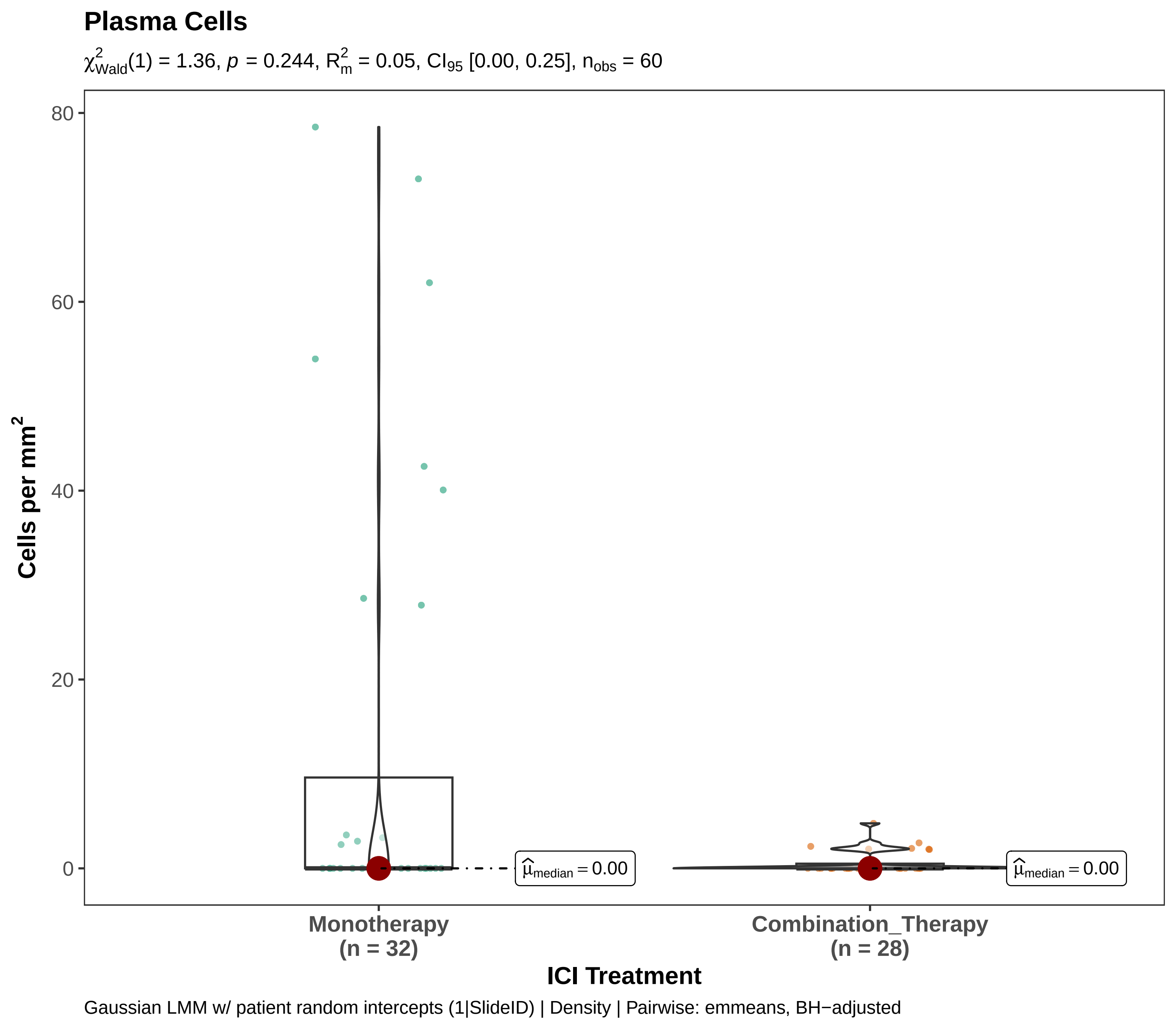
Plasma Cells
$\chi^2_{Wald}(1) = 1.36$, $p = 0.244$, $R^2_m = 0.05$, $CI_{95}$ [0.00, 0.25], $n_{obs} = 60$
Cells per mm$^2$
80
60
40
20
0
$\widehat{\mu}_{median} = 0.00$
$\widehat{\mu}_{median} = 0.00$
Monotherapy
(n = 32)
Combination_Therapy
(n = 28)
ICI Treatment
Gaussian LMM w/ patient random intercepts (1|SlideID) | Density | Pairwise: emmeans, BH−adjusted

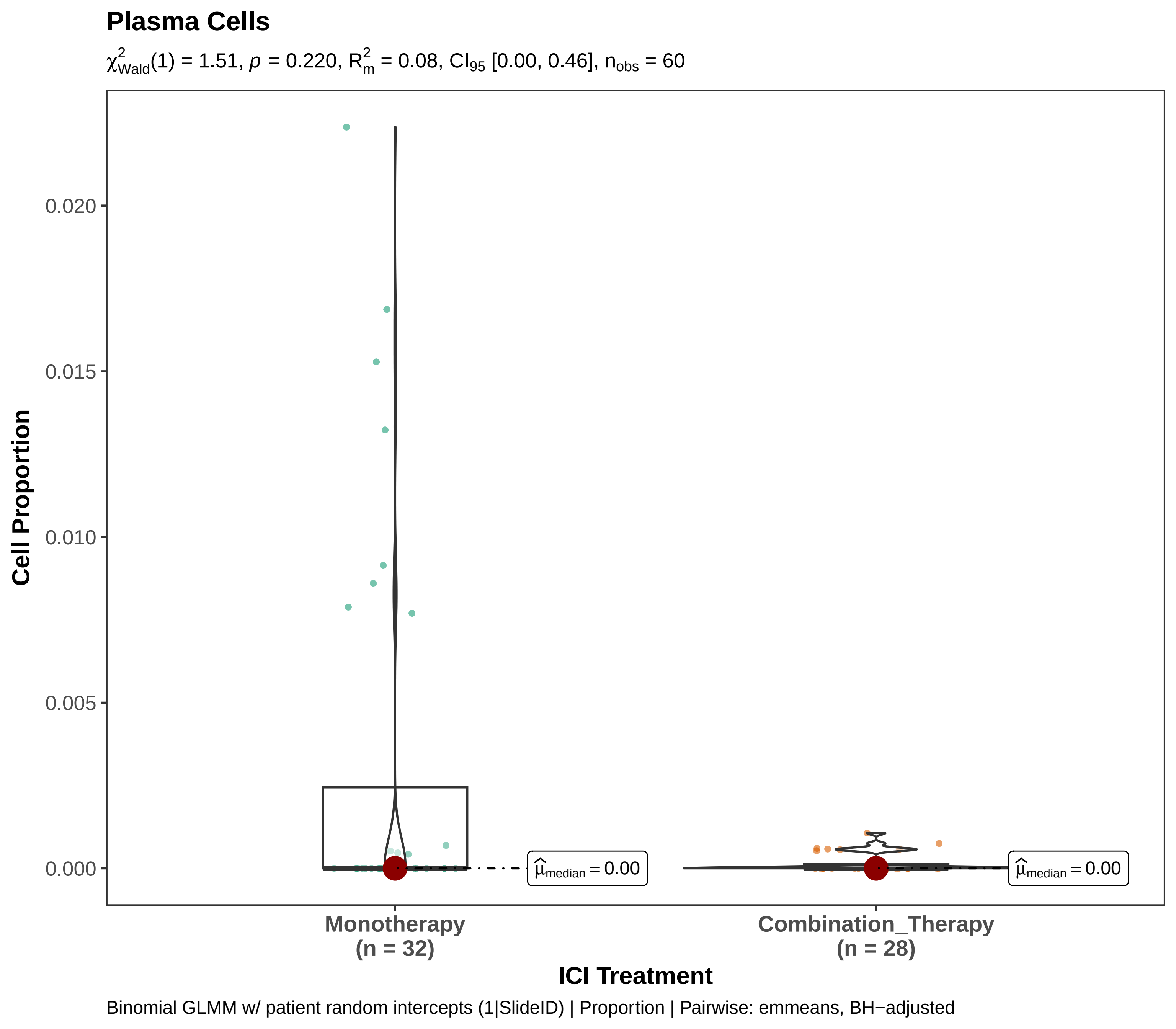
Plasma Cells
$\chi^2_{Wald}(1) = 1.51$, $p = 0.220$, $R^2_m = 0.08$, $CI_{95}$ [0.00, 0.46], $n_{obs} = 60$
Cell Proportion
0.020
0.015
0.010
0.005
0.000
$\hat{\mu}_{median} = 0.00$
$\hat{\mu}_{median} = 0.00$
Monotherapy
(n = 32)
Combination_Therapy
(n = 28)
ICI Treatment
Binomial GLMM w/ patient random intercepts (1|SlideID) | Proportion | Pairwise: emmeans, BH−adjusted

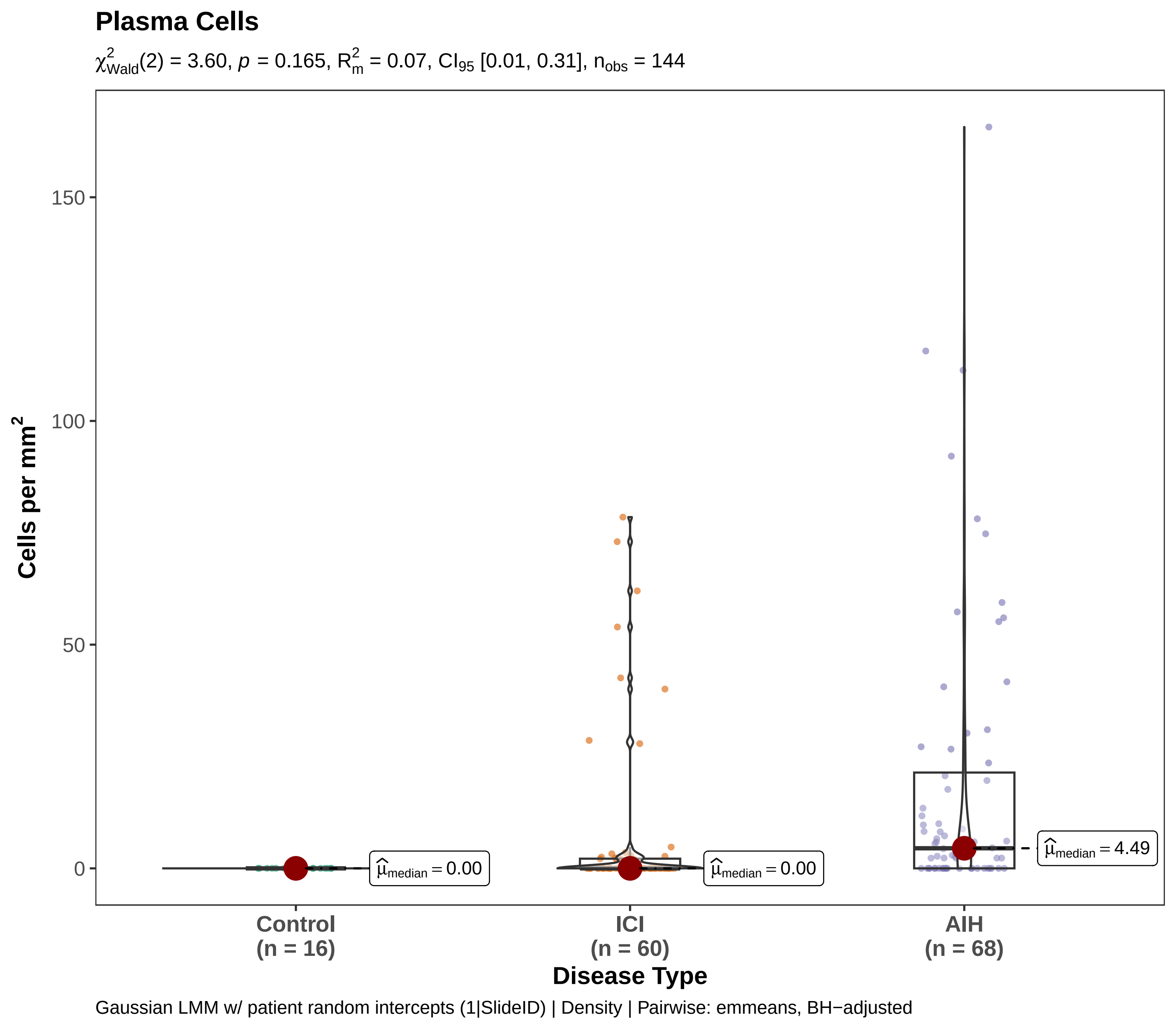

Plasma Cells
$\chi^2_{Wald}(2) = 3.60$, $p = 0.165$, $R^2_m = 0.07$, $CI_{95}$ [0.01, 0.31], $n_{obs} = 144$
Cells per mm$^2$
150
100
50
0
$\hat{\mu}_{median} = 0.00$
$\hat{\mu}_{median} = 0.00$
$\hat{\mu}_{median} = 4.49$
Control
(n = 16)
ICI
(n = 60)
AIH
(n = 68)
Disease Type
Gaussian LMM w/ patient random intercepts (1|SlideID) | Density | Pairwise: emmeans, BH−adjusted

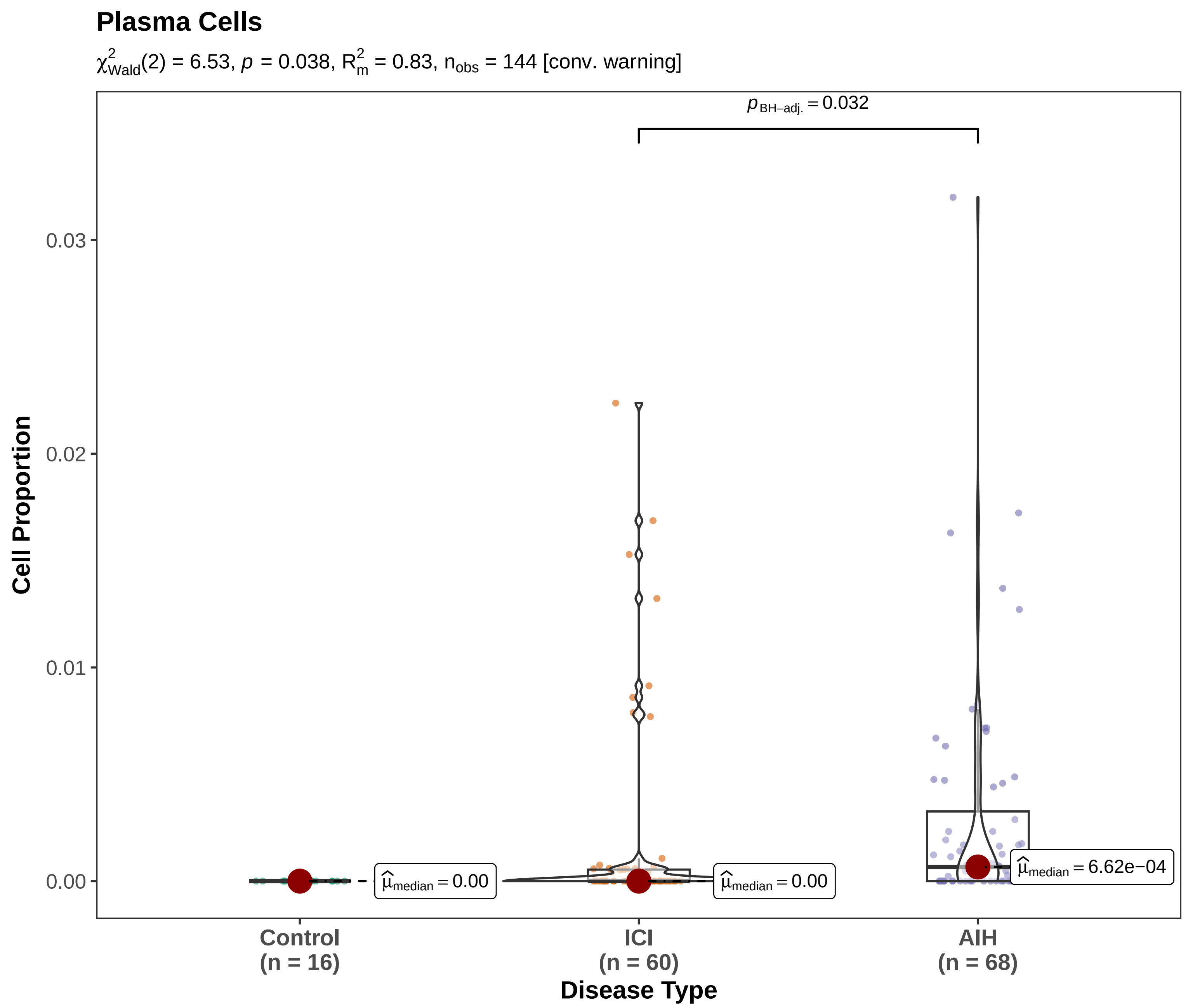

Binomial GLMM w/ patient random intercepts (1|SlideID) | Proportion | Pairwise: emmeans, BH−adjusted

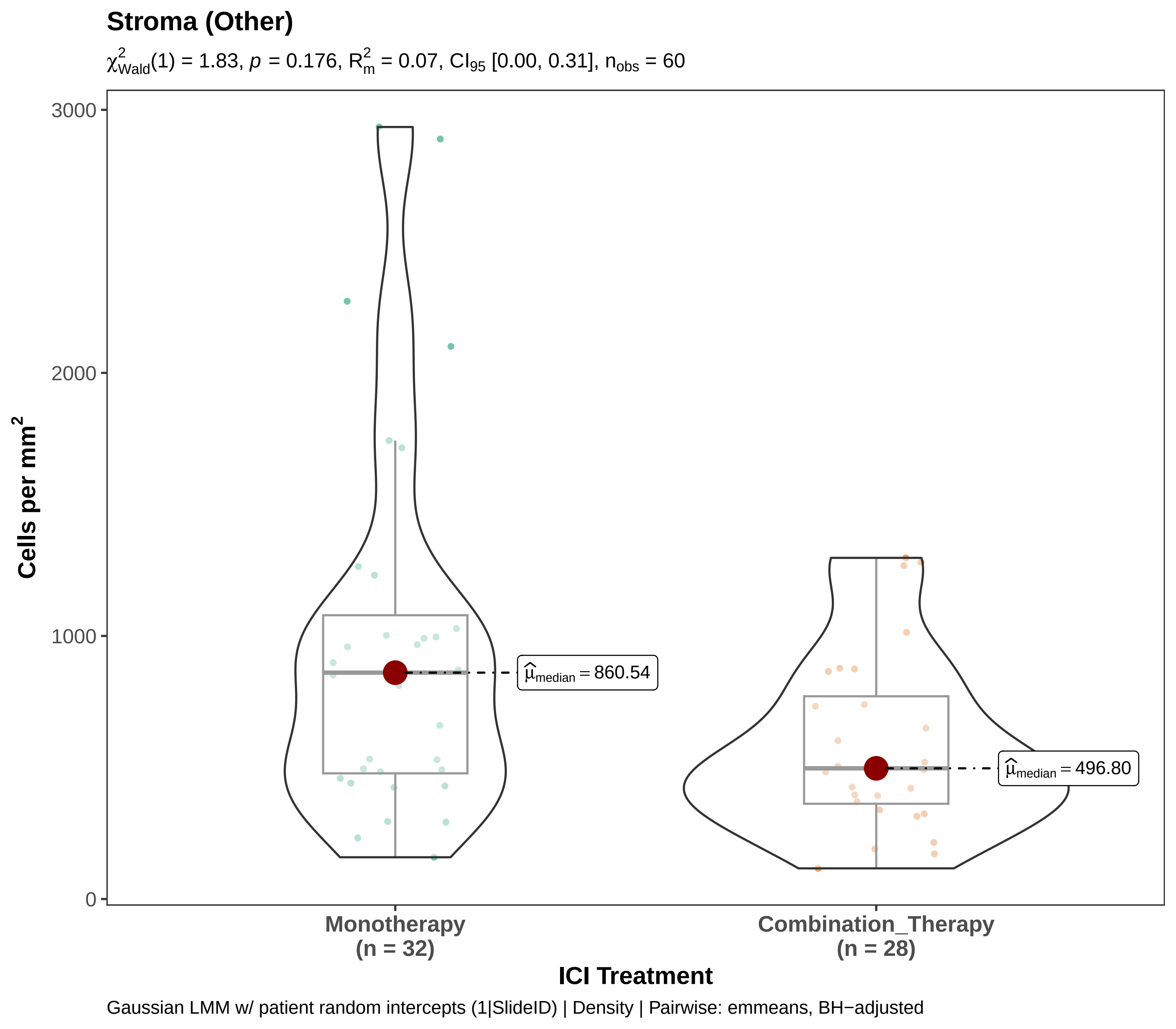
Stroma (Other)
$\chi^2_{Wald}(1) = 1.83$, $p = 0.176$, $R^2_m = 0.07$, $CI_{95}$ [0.00, 0.31], $n_{obs} = 60$
Cells per mm²
3000
2000
1000
0
$\hat{\mu}_{median} = 860.54$
$\hat{\mu}_{median} = 496.80$
Monotherapy
(n = 32)
Combination_Therapy
(n = 28)
ICI Treatment
Gaussian LMM w/ patient random intercepts (1|SlideID) | Density | Pairwise: emmeans, BH−adjusted

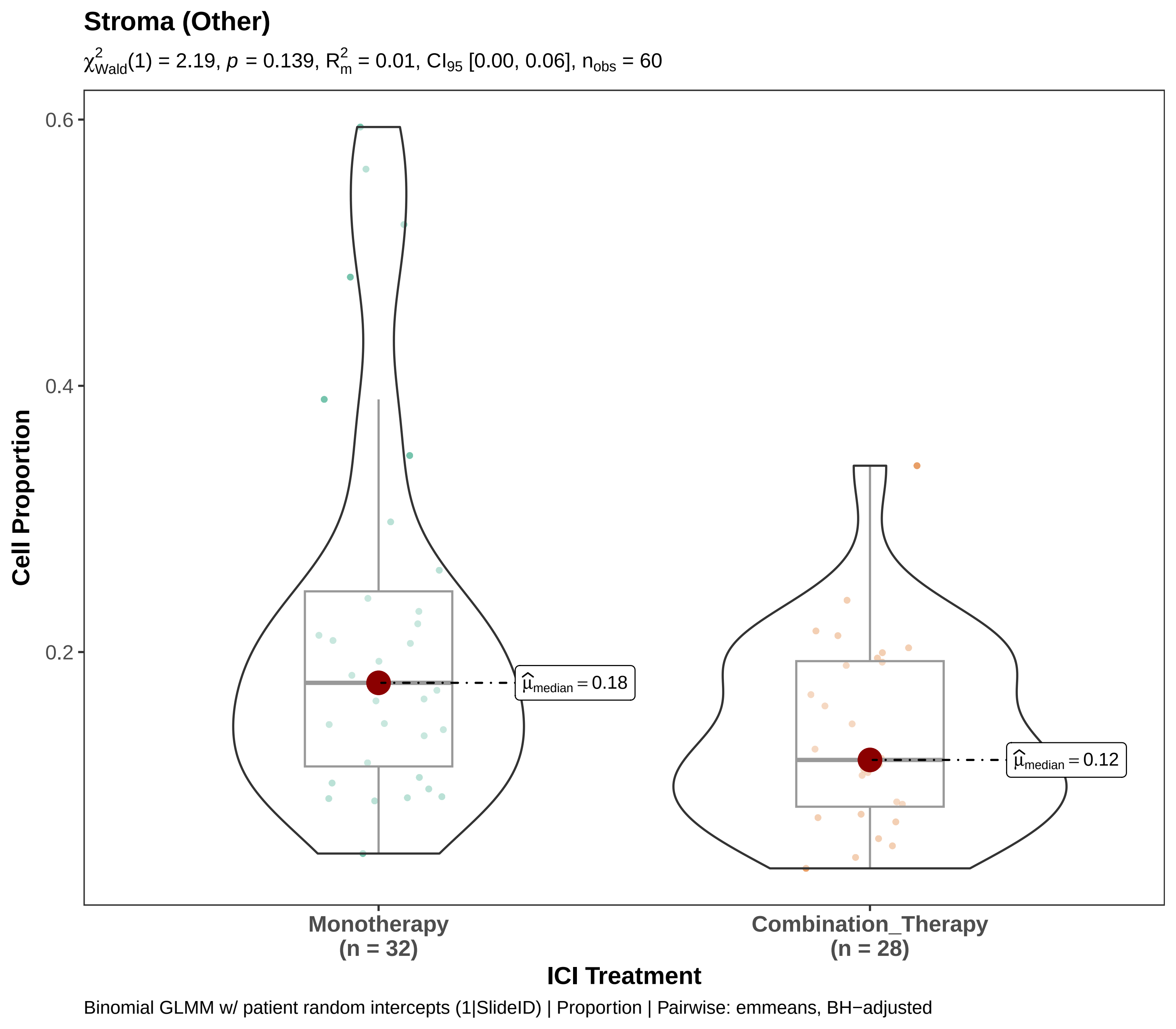

Stroma (Other)
$\chi^2_{Wald}(1) = 2.19$, $p = 0.139$, $R^2_m = 0.01$, $CI_{95}$ [0.00, 0.06], $n_{obs} = 60$
Cell Proportion
0.6
0.4
0.2
$\hat{\mu}_{median} = 0.18$
$\hat{\mu}_{median} = 0.12$
Monotherapy
(n = 32)
Combination_Therapy
(n = 28)
ICI Treatment
Binomial GLMM w/ patient random intercepts (1|SlideID) | Proportion | Pairwise: emmeans, BH−adjusted

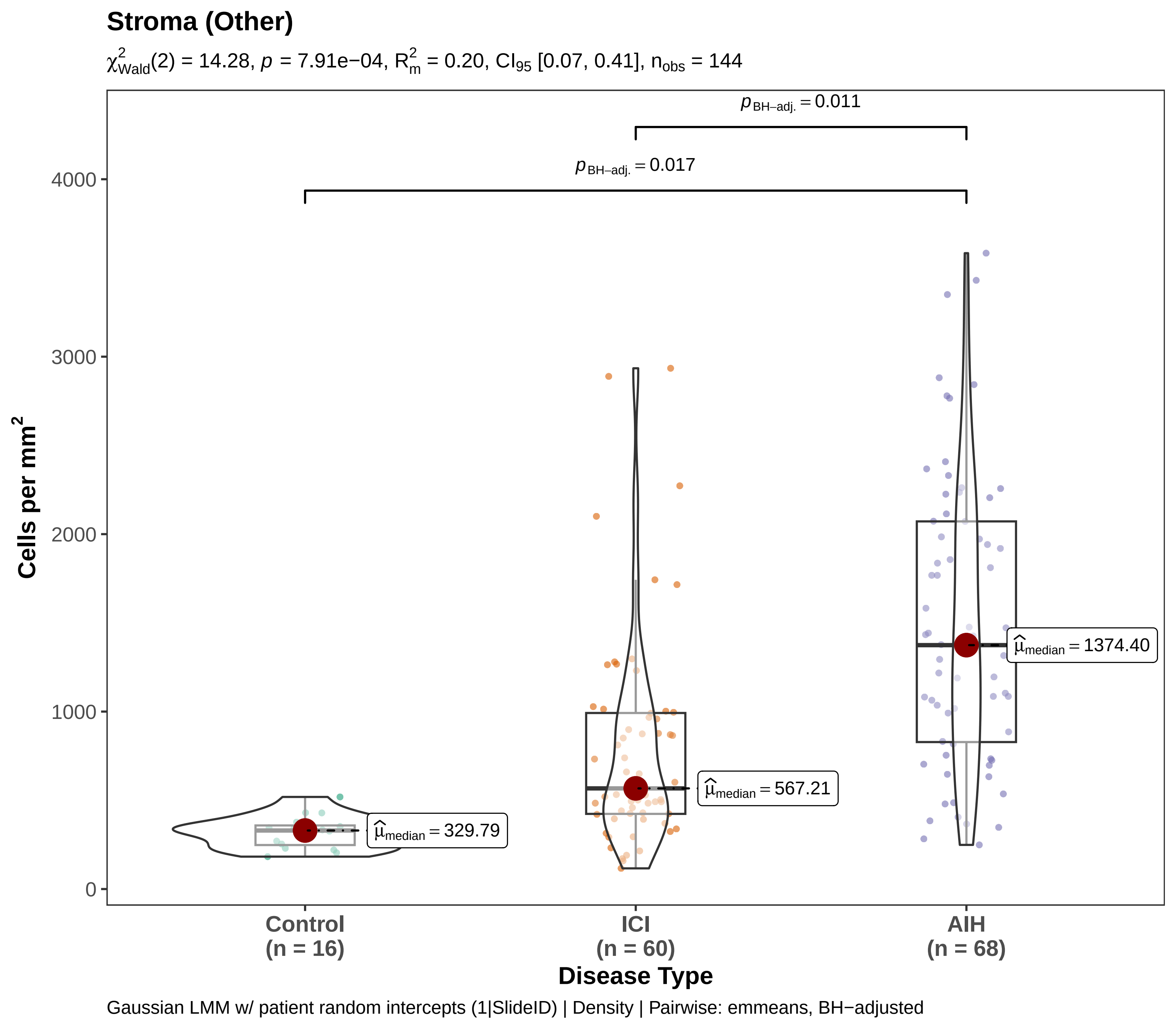

Stroma (Other)
$\chi^2_{Wald}(2) = 14.28$, $p = 7.91e{-}04$, $R^2_m = 0.20$, $CI_{95}$ [0.07, 0.41], $n_{obs} = 144$
$p_{BH-adj.} = 0.011$
$p_{BH-adj.} = 0.017$
4000
3000
2000
1000
0
Cells per mm$^2$
$\hat{\mu}_{median} = 329.79$
$\hat{\mu}_{median} = 567.21$
$\hat{\mu}_{median} = 1374.40$
Control
(n = 16)
ICI
(n = 60)
AIH
(n = 68)
Disease Type
Gaussian LMM w/ patient random intercepts (1|SlideID) | Density | Pairwise: emmeans, BH−adjusted

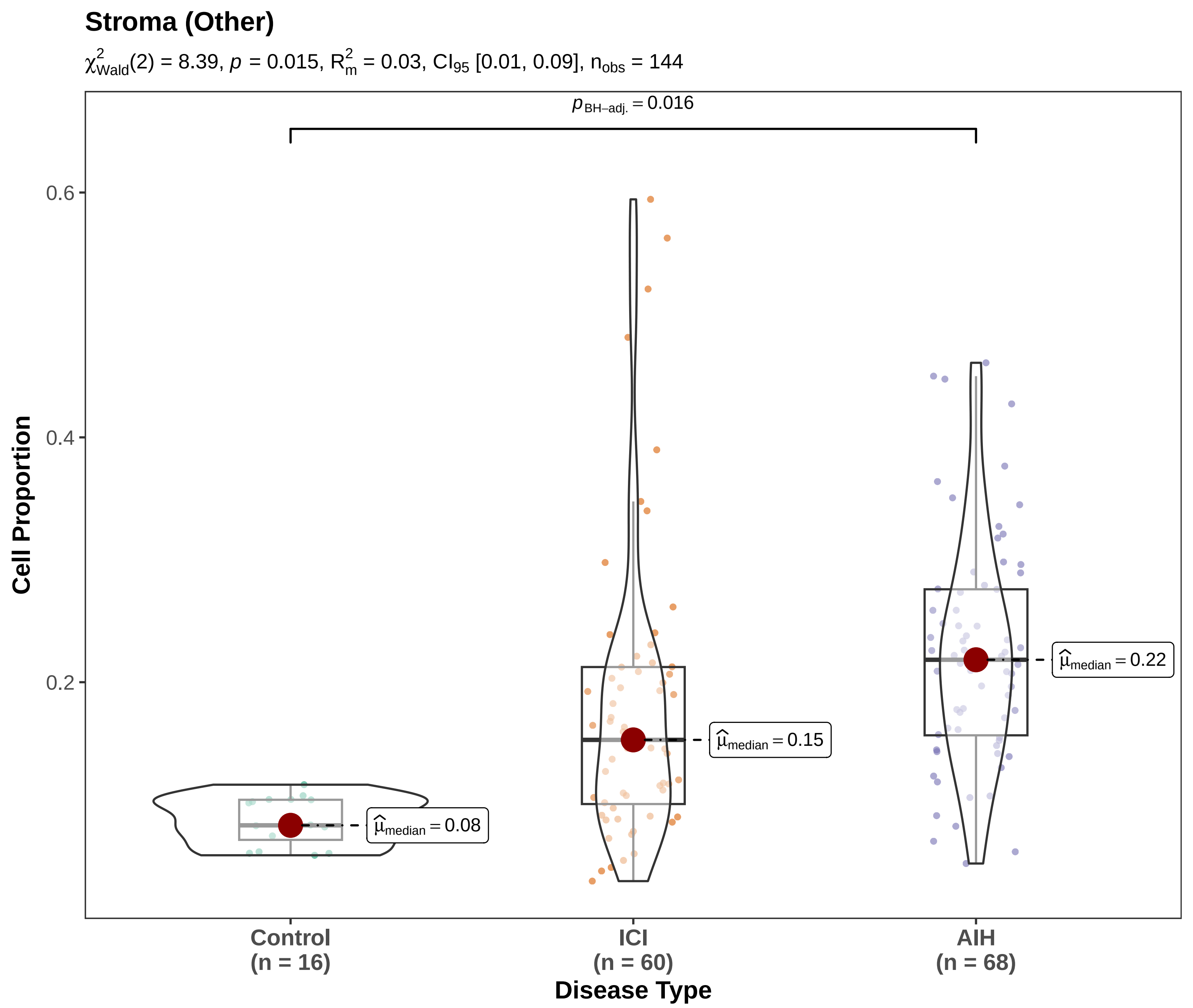

Binomial GLMM w/ patient random intercepts (1|SlideID) | Proportion | Pairwise: emmeans, BH−adjusted

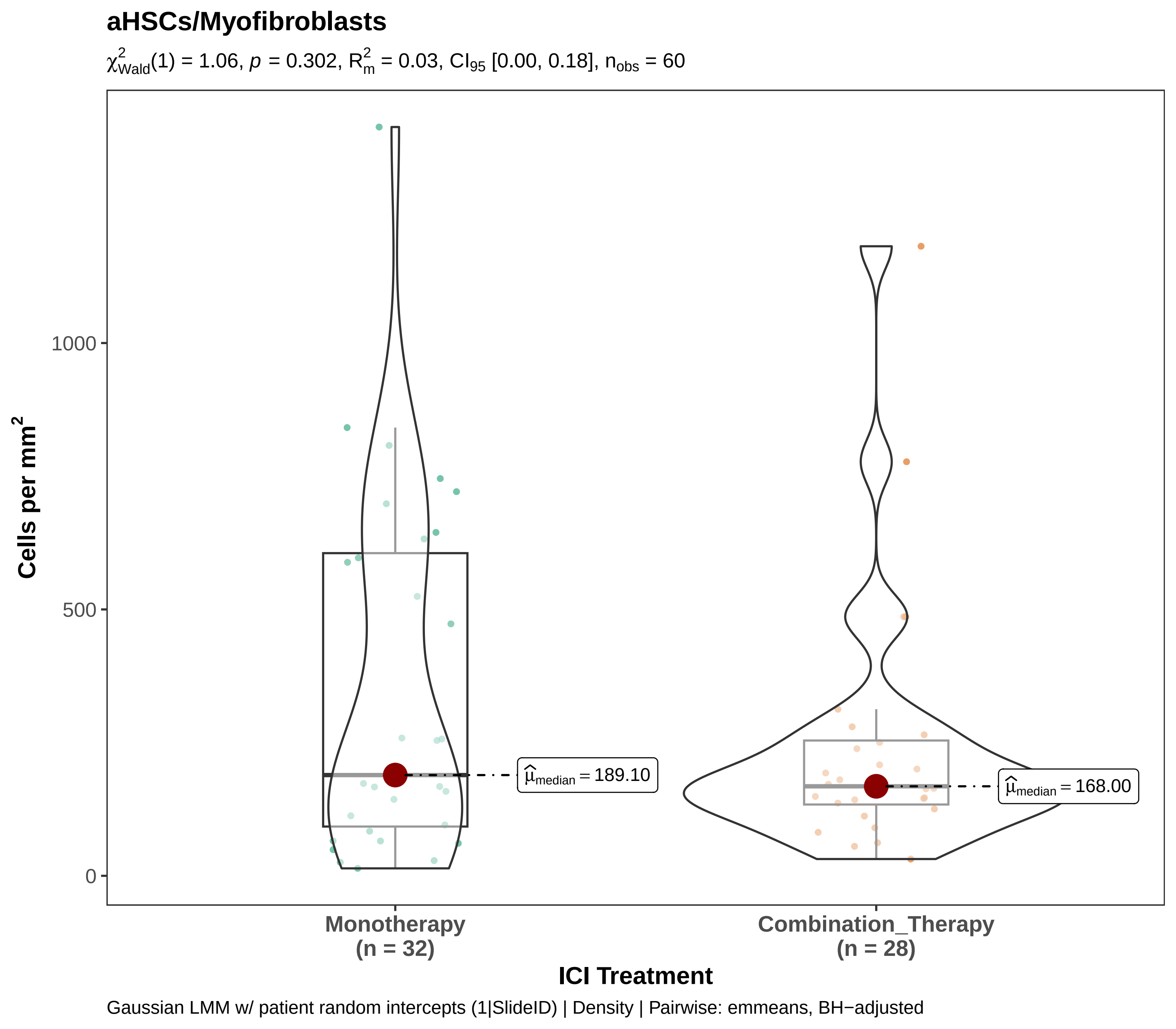
aHSCs/Myofibroblasts
$\chi^2_{Wald}(1) = 1.06$, $p = 0.302$, $R^2_m = 0.03$, $CI_{95}$ [0.00, 0.18], $n_{obs} = 60$
Cells per mm$^2$
1000
500
0
$\widehat{\mu}_{median} = 189.10$
$\widehat{\mu}_{median} = 168.00$
Monotherapy
(n = 32)
Combination_Therapy
(n = 28)
ICI Treatment
Gaussian LMM w/ patient random intercepts (1|SlideID) | Density | Pairwise: emmeans, BH−adjusted

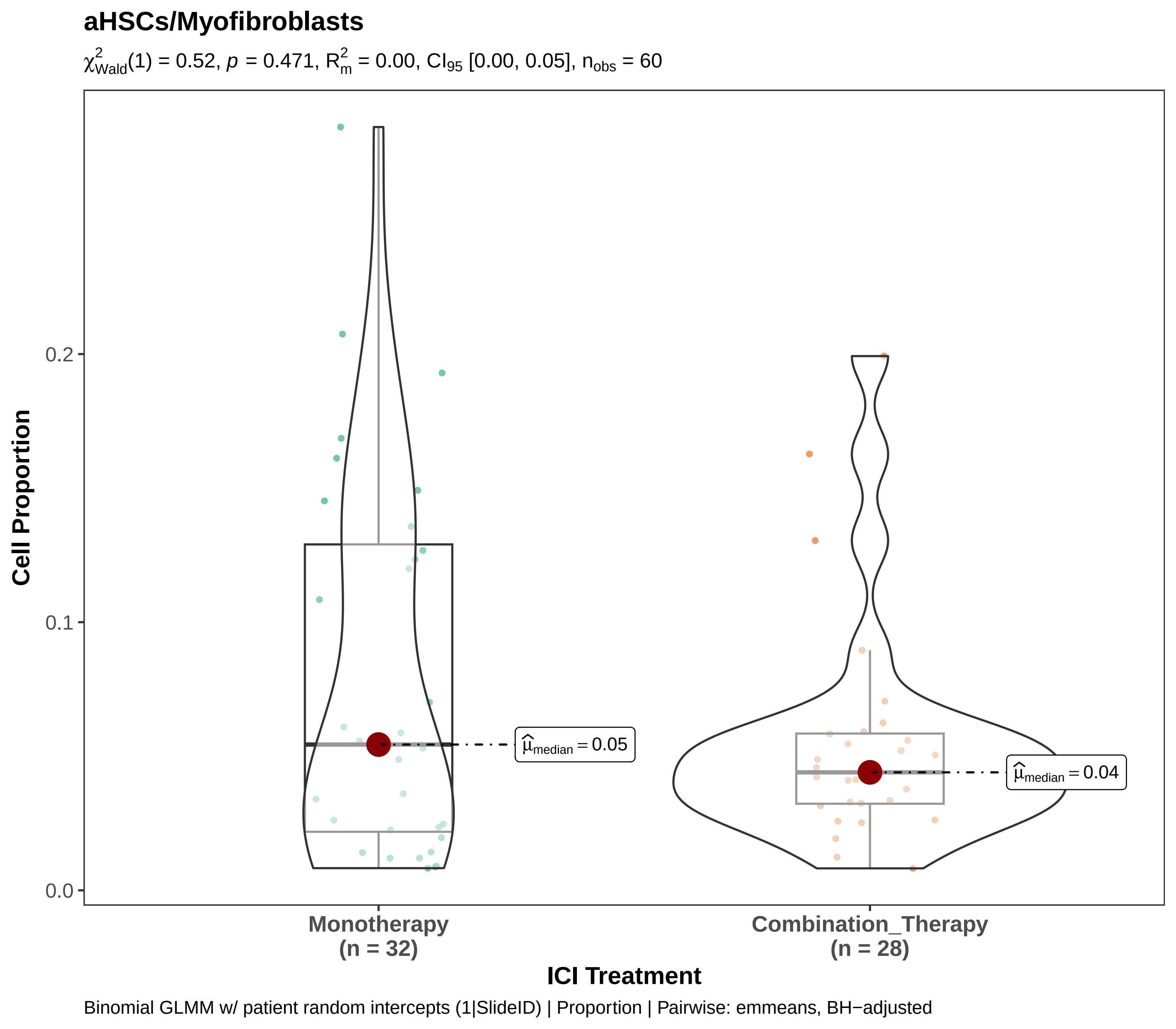

aHSCs/Myofibroblasts
$\chi^2_{Wald}(1) = 0.52$, $p = 0.471$, $R^2_m = 0.00$, $CI_{95}$ [0.00, 0.05], $n_{obs} = 60$
Cell Proportion
0.0
0.1
0.2
$\widehat{\mu}_{median} = 0.05$
$\widehat{\mu}_{median} = 0.04$
Monotherapy
(n = 32)
Combination_Therapy
(n = 28)
ICI Treatment
Binomial GLMM w/ patient random intercepts (1|SlideID) | Proportion | Pairwise: emmeans, BH−adjusted

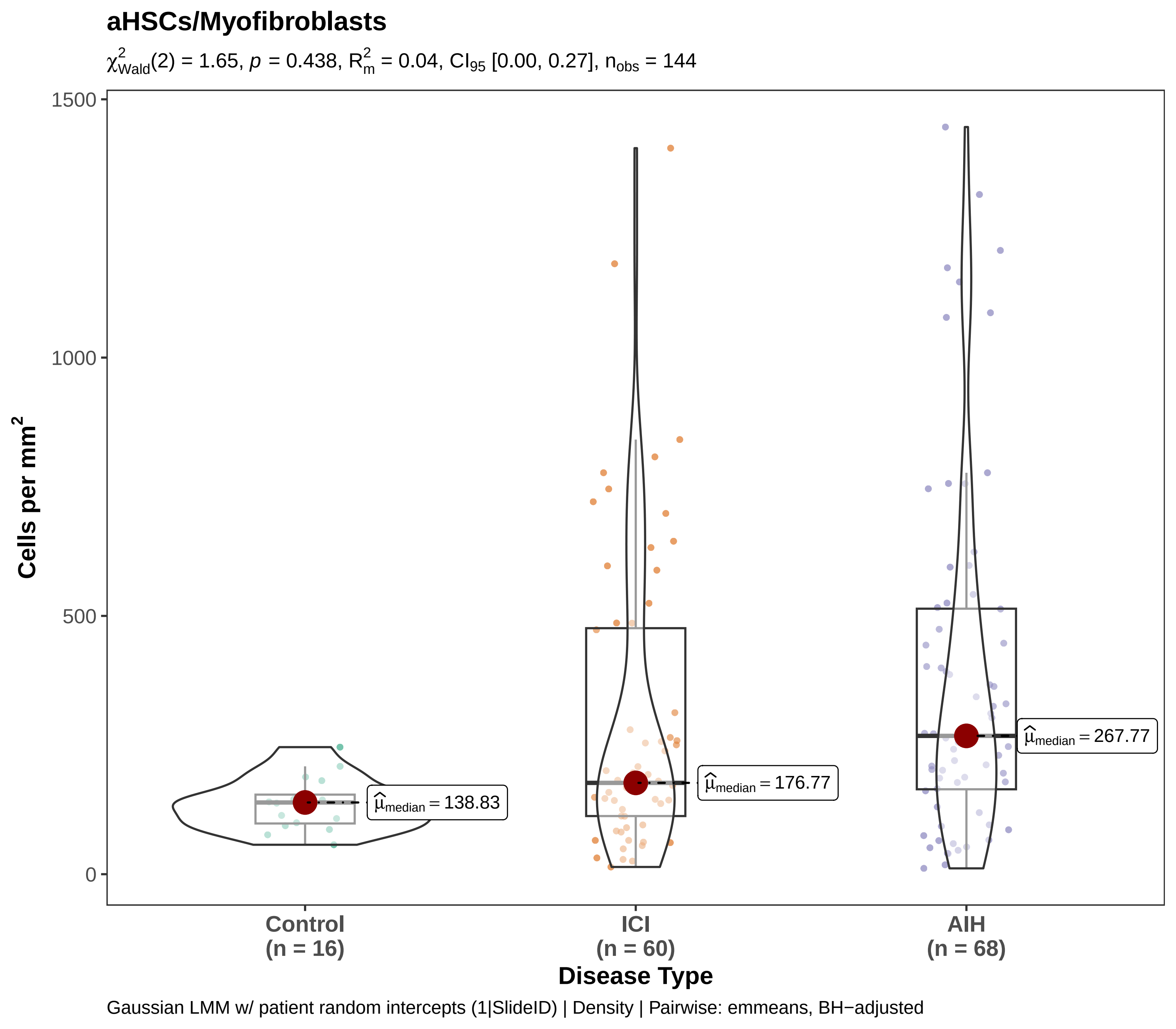

aHSCs/Myofibroblasts
$\chi^2_{Wald}(2) = 1.65$, $p = 0.438$, $R^2_m = 0.04$, $CI_{95}$ [0.00, 0.27], $n_{obs} = 144$
Cells per mm²
1500
1000
500
0
$\hat{\mu}_{median} = 138.83$
$\hat{\mu}_{median} = 176.77$
$\hat{\mu}_{median} = 267.77$
Control
(n = 16)
ICI
(n = 60)
AIH
(n = 68)
Disease Type
Gaussian LMM w/ patient random intercepts (1|SlideID) | Density | Pairwise: emmeans, BH−adjusted

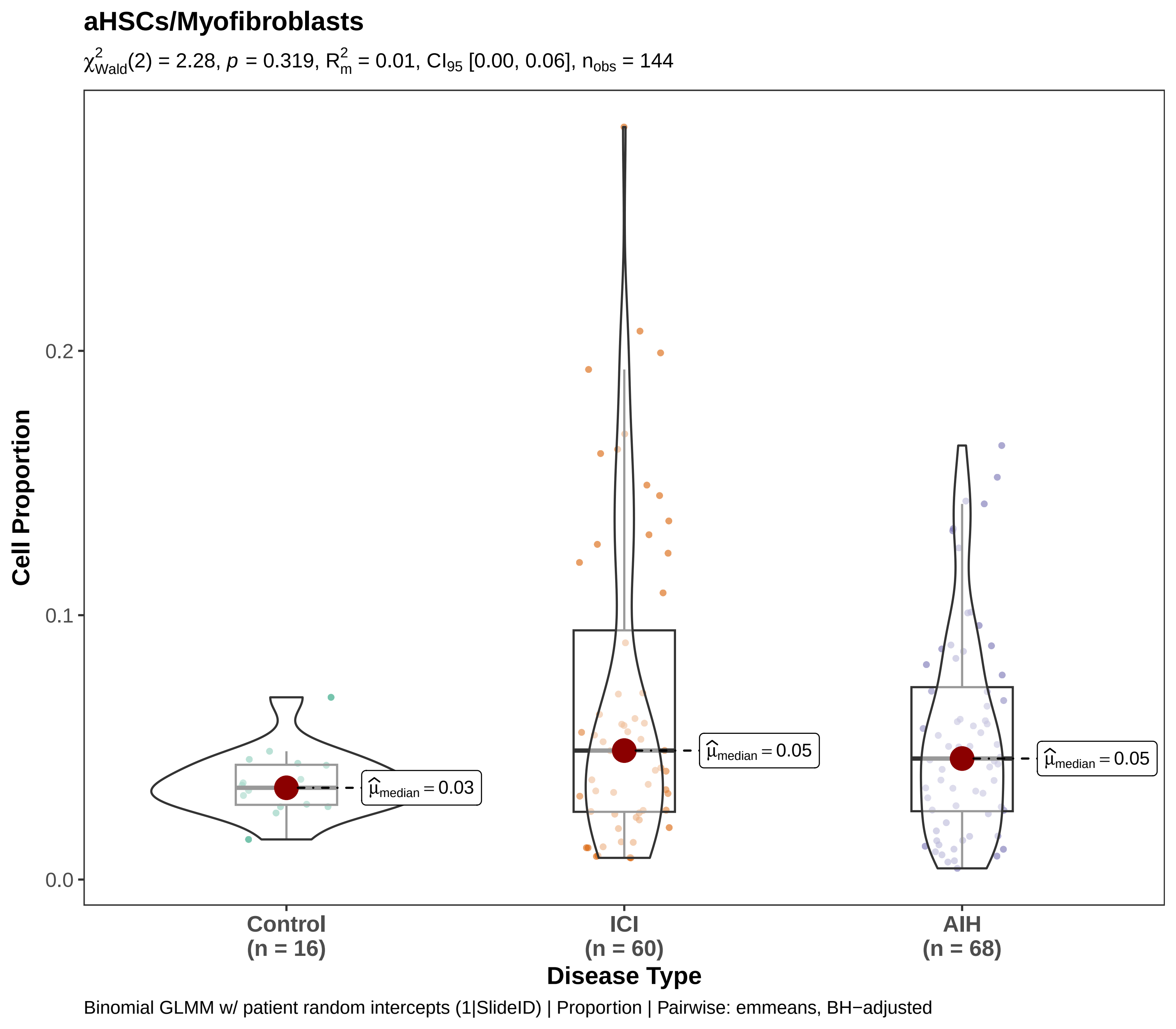

aHSCs/Myofibroblasts
$\chi^2_{Wald}(2) = 2.28$, $p = 0.319$, $R^2_m = 0.01$, $CI_{95}$ [0.00, 0.06], $n_{obs} = 144$
Cell Proportion
0.0
0.1
0.2
$\hat{\mu}_{median} = 0.03$
$\hat{\mu}_{median} = 0.05$
$\hat{\mu}_{median} = 0.05$
Control
(n = 16)
ICI
(n = 60)
AIH
(n = 68)
Disease Type
Binomial GLMM w/ patient random intercepts (1|SlideID) | Proportion | Pairwise: emmeans, BH−adjusted

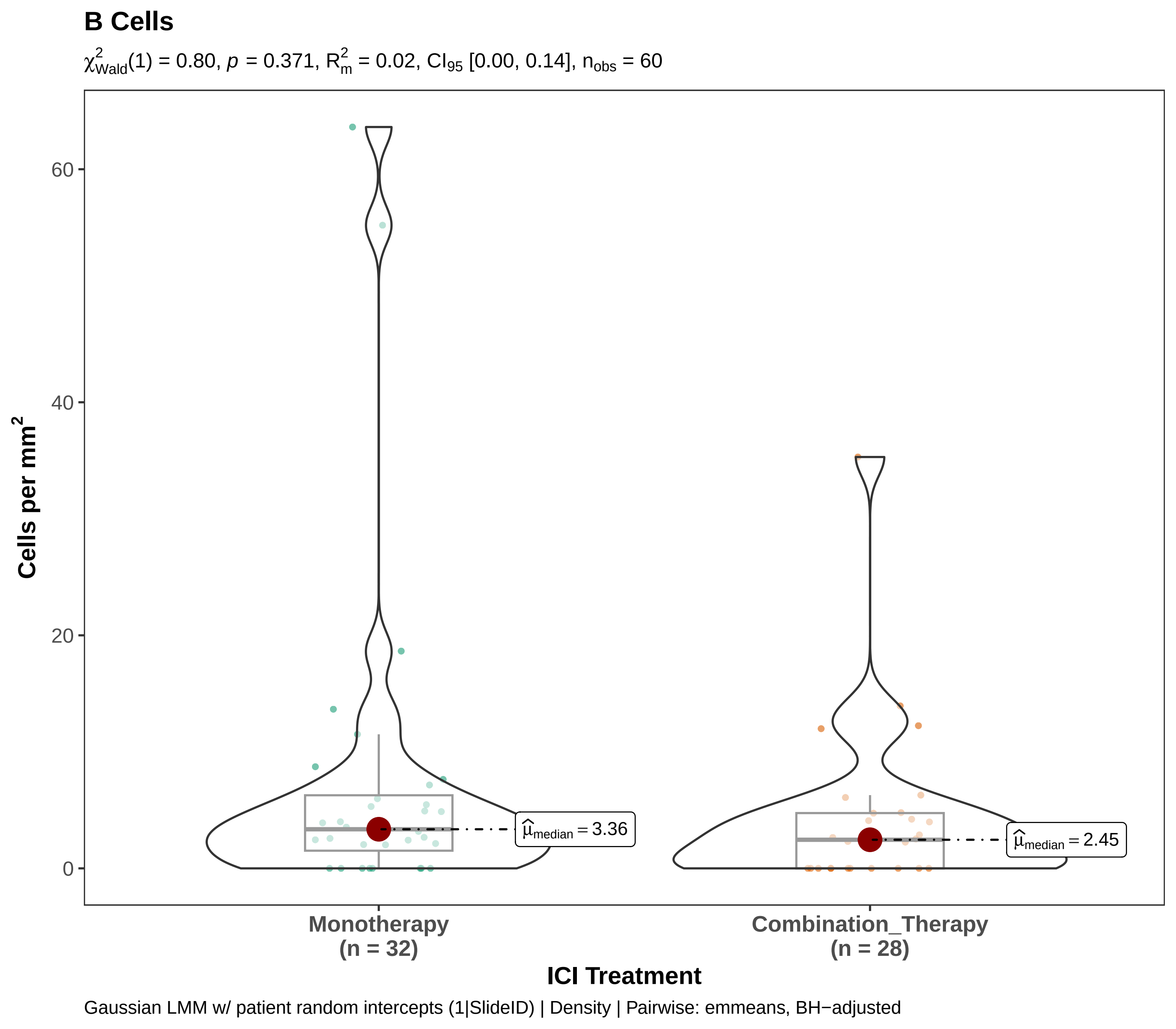
B Cells
$\chi^2_{Wald}(1) = 0.80$, $p = 0.371$, $R^2_m = 0.02$, $CI_{95}$ [0.00, 0.14], $n_{obs} = 60$
Cells per mm$^2$
60
40
20
0
$\hat{\mu}_{median} = 3.36$
$\hat{\mu}_{median} = 2.45$
Monotherapy
(n = 32)
Combination_Therapy
(n = 28)
ICI Treatment
Gaussian LMM w/ patient random intercepts (1|SlideID) | Density | Pairwise: emmeans, BH−adjusted

**B Cells**

$\chi^2_{Wald}(1) = 0.78$, $p = 0.376$, $R^2_m = 0.01$, $CI_{95}$ [0.00, 0.11], $n_{obs} = 60$

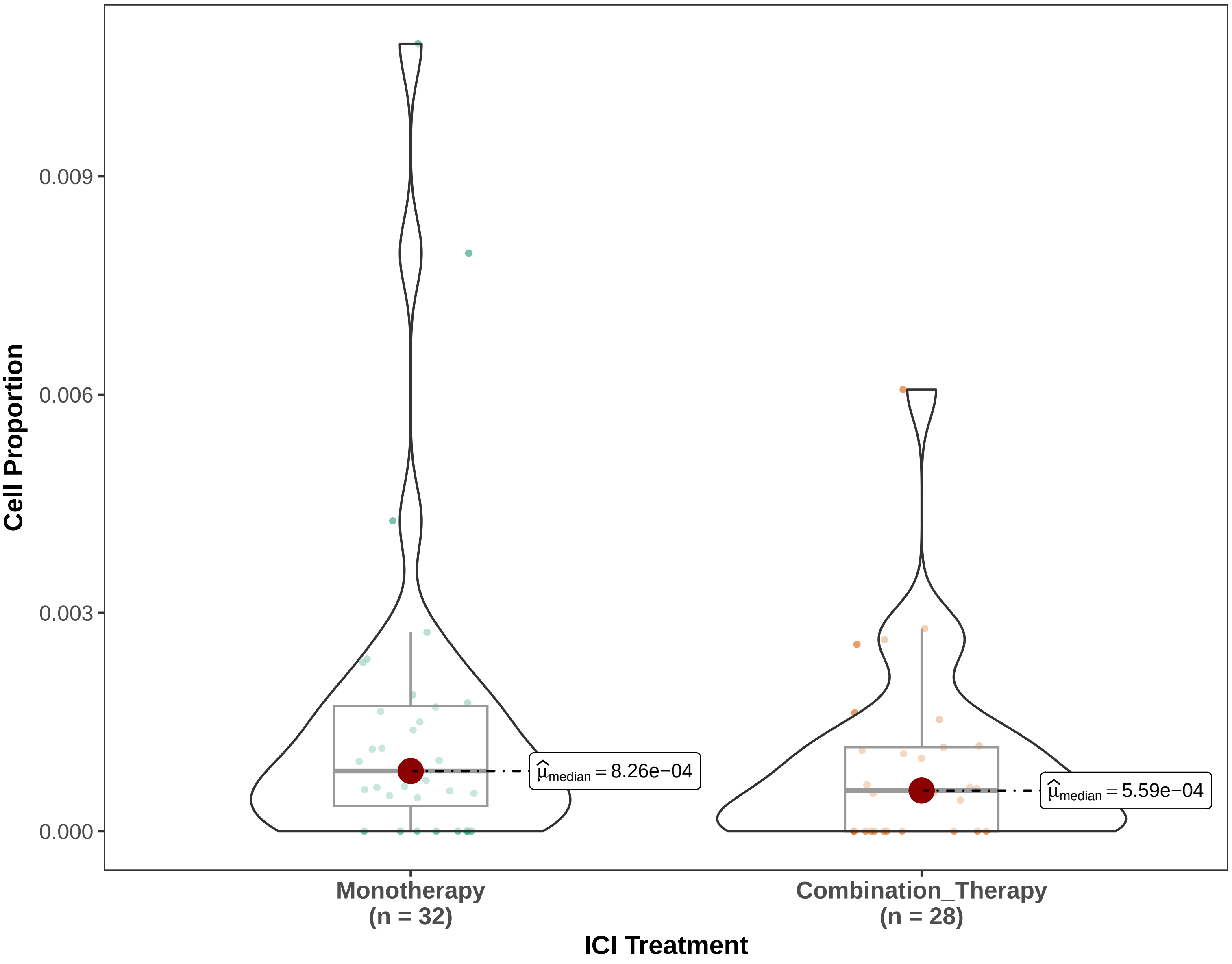


Binomial GLMM w/ patient random intercepts (1|SlideID) | Proportion | Pairwise: emmeans, BH−adjusted

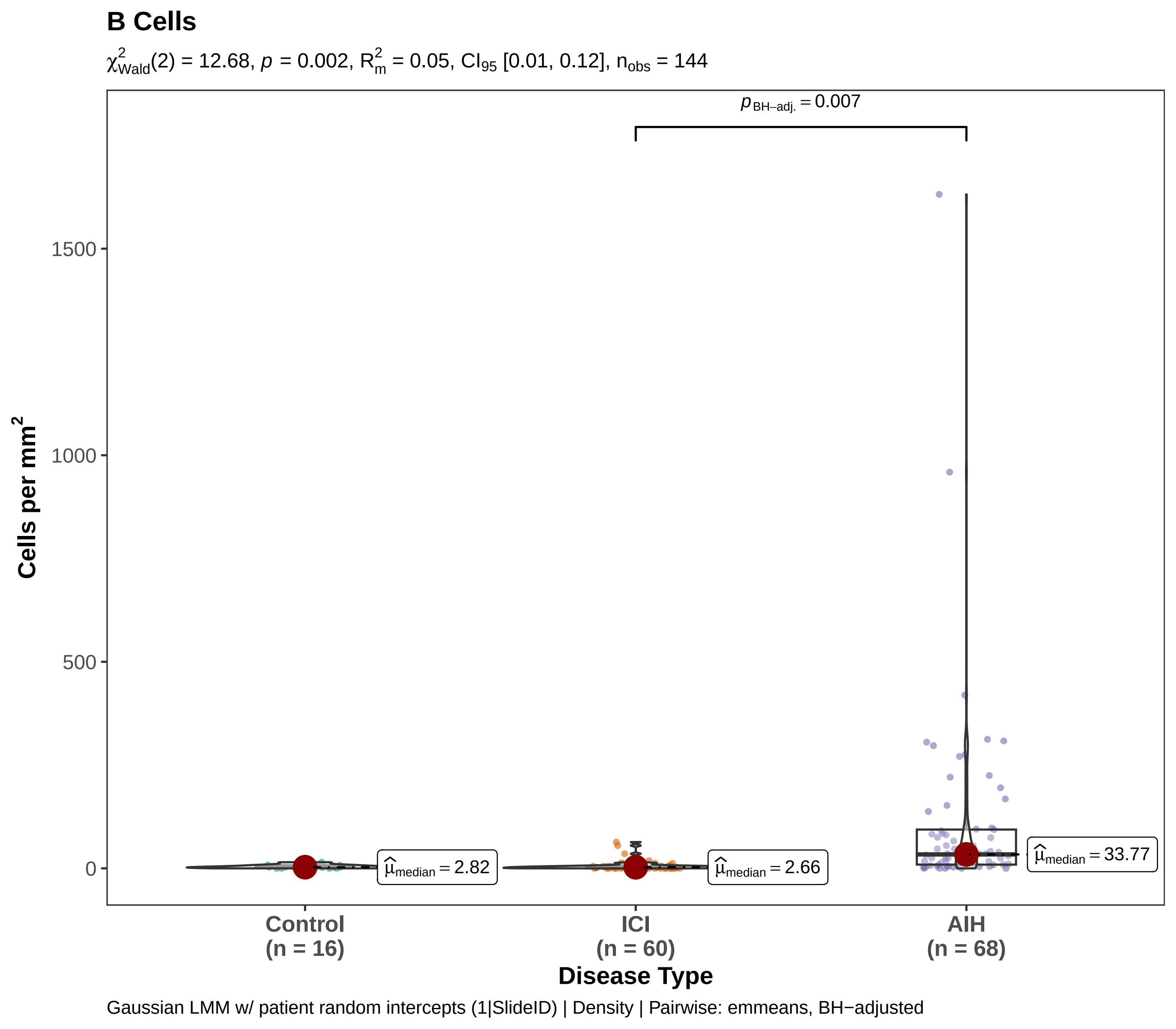
B Cells
$\chi^2_{Wald}(2) = 12.68$, $p = 0.002$, $R^2_m = 0.05$, $CI_{95}$ [0.01, 0.12], $n_{obs} = 144$
$p_{BH-adj.} = 0.007$
Cells per mm$^2$
1500
1000
500
0
$\hat{\mu}_{median} = 2.82$
$\hat{\mu}_{median} = 2.66$
$\hat{\mu}_{median} = 33.77$
Control
(n = 16)
ICI
(n = 60)
AIH
(n = 68)
Disease Type
Gaussian LMM w/ patient random intercepts (1|SlideID) | Density | Pairwise: emmeans, BH−adjusted

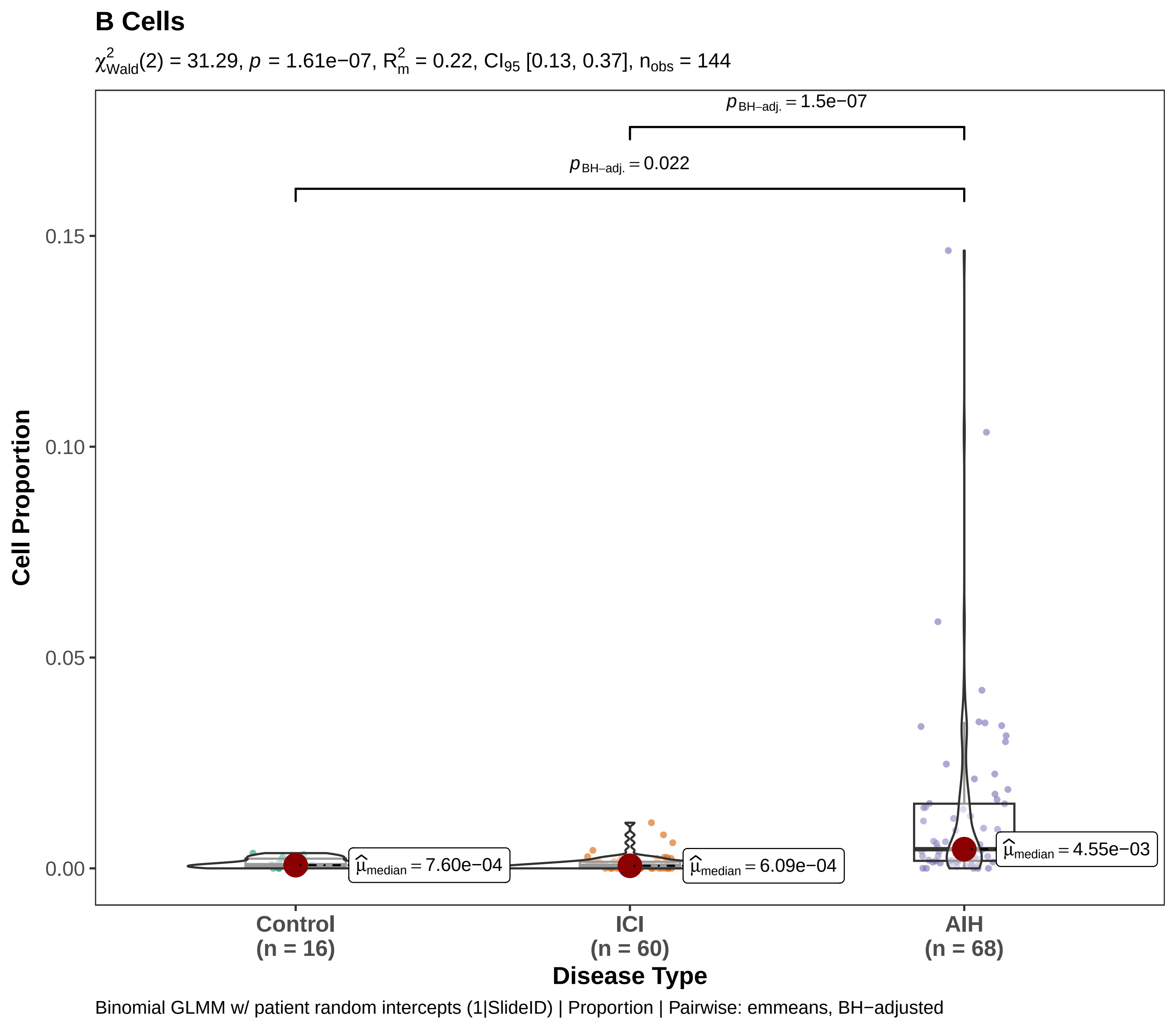

B Cells
$\chi^2_{Wald}(2) = 31.29$, $p = 1.61e{-}07$, $R^2_m = 0.22$, $CI_{95}$ [0.13, 0.37], $n_{obs} = 144$
$p_{BH-adj.} = 1.5e{-}07$
$p_{BH-adj.} = 0.022$
0.15
0.10
0.05
0.00
Cell Proportion
$\hat{\mu}_{median} = 7.60e{-}04$
$\hat{\mu}_{median} = 6.09e{-}04$
$\hat{\mu}_{median} = 4.55e{-}03$
Control
(n = 16)
ICI
(n = 60)
AIH
(n = 68)
Disease Type
Binomial GLMM w/ patient random intercepts (1|SlideID) | Proportion | Pairwise: emmeans, BH−adjusted

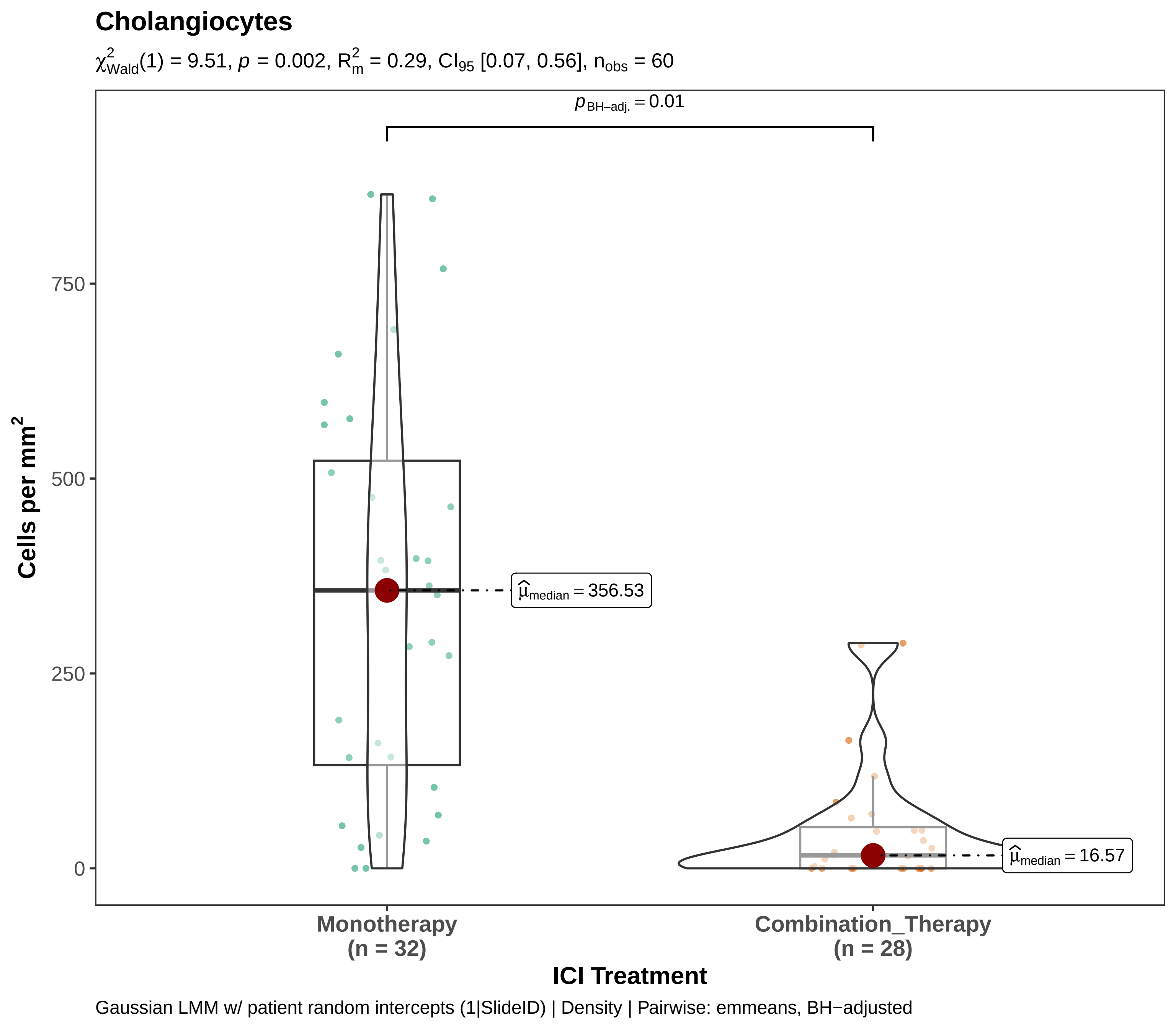

Cholangiocytes
$\chi^2_{Wald}(1) = 9.51$, $p = 0.002$, $R^2_m = 0.29$, $CI_{95}$ [0.07, 0.56], $n_{obs} = 60$
$p_{BH-adj.} = 0.01$
$\widehat{\mu}_{median} = 356.53$
$\widehat{\mu}_{median} = 16.57$
750
500
250
0
Cells per mm$^2$
Monotherapy
(n = 32)
Combination_Therapy
(n = 28)
ICI Treatment
Gaussian LMM w/ patient random intercepts (1|SlideID) | Density | Pairwise: emmeans, BH−adjusted

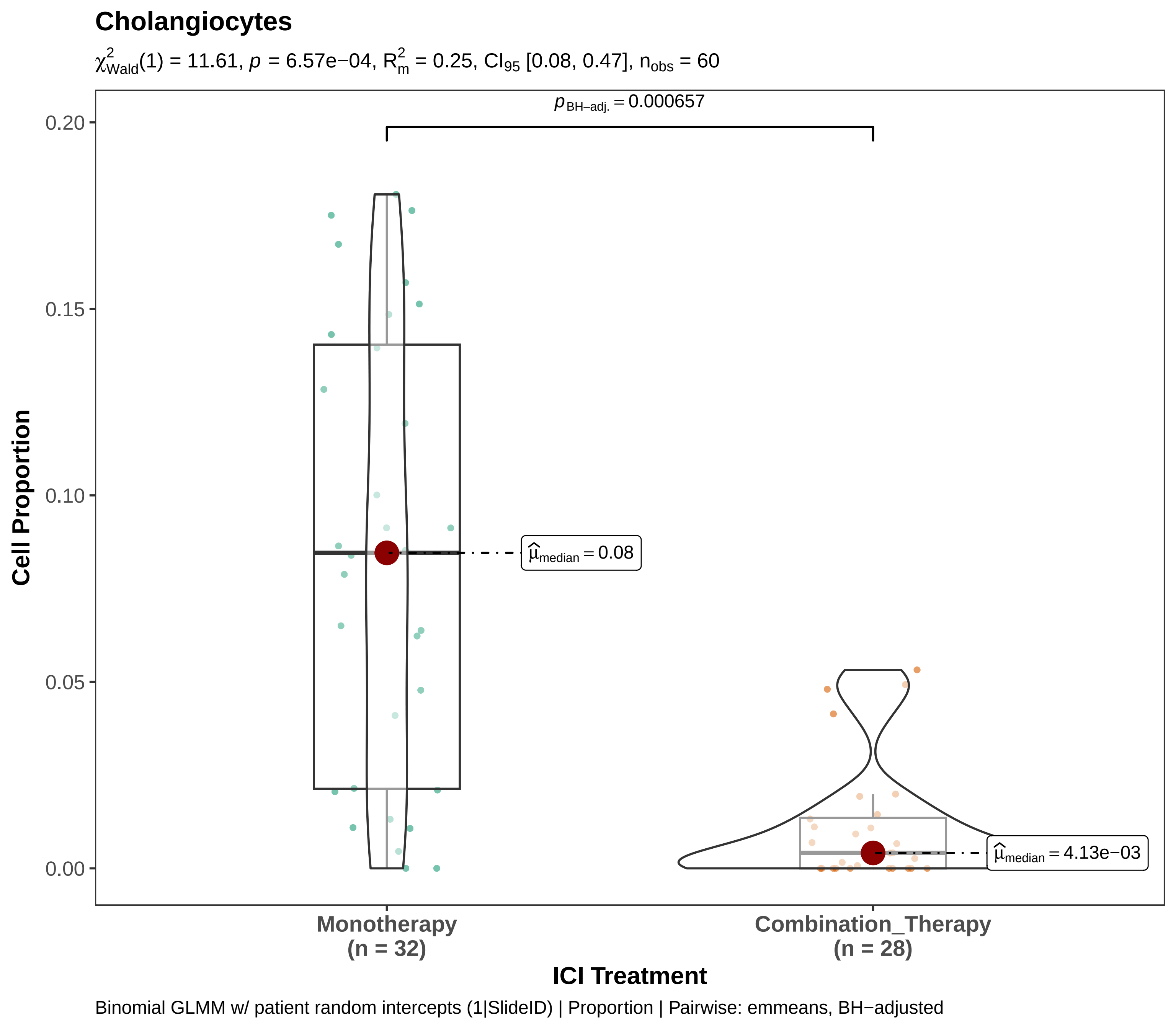
Cholangiocytes
χ²Wald(1) = 11.61, p = 6.57e−04, R²m = 0.25, CI95 [0.08, 0.47], nobs = 60
pBH−adj. = 0.000657
0.20
0.15
0.10
0.05
0.00
Cell Proportion
μ̂median = 0.08
μ̂median = 4.13e−03
Monotherapy
(n = 32)
Combination_Therapy
(n = 28)
ICI Treatment
Binomial GLMM w/ patient random intercepts (1|SlideID) | Proportion | Pairwise: emmeans, BH−adjusted

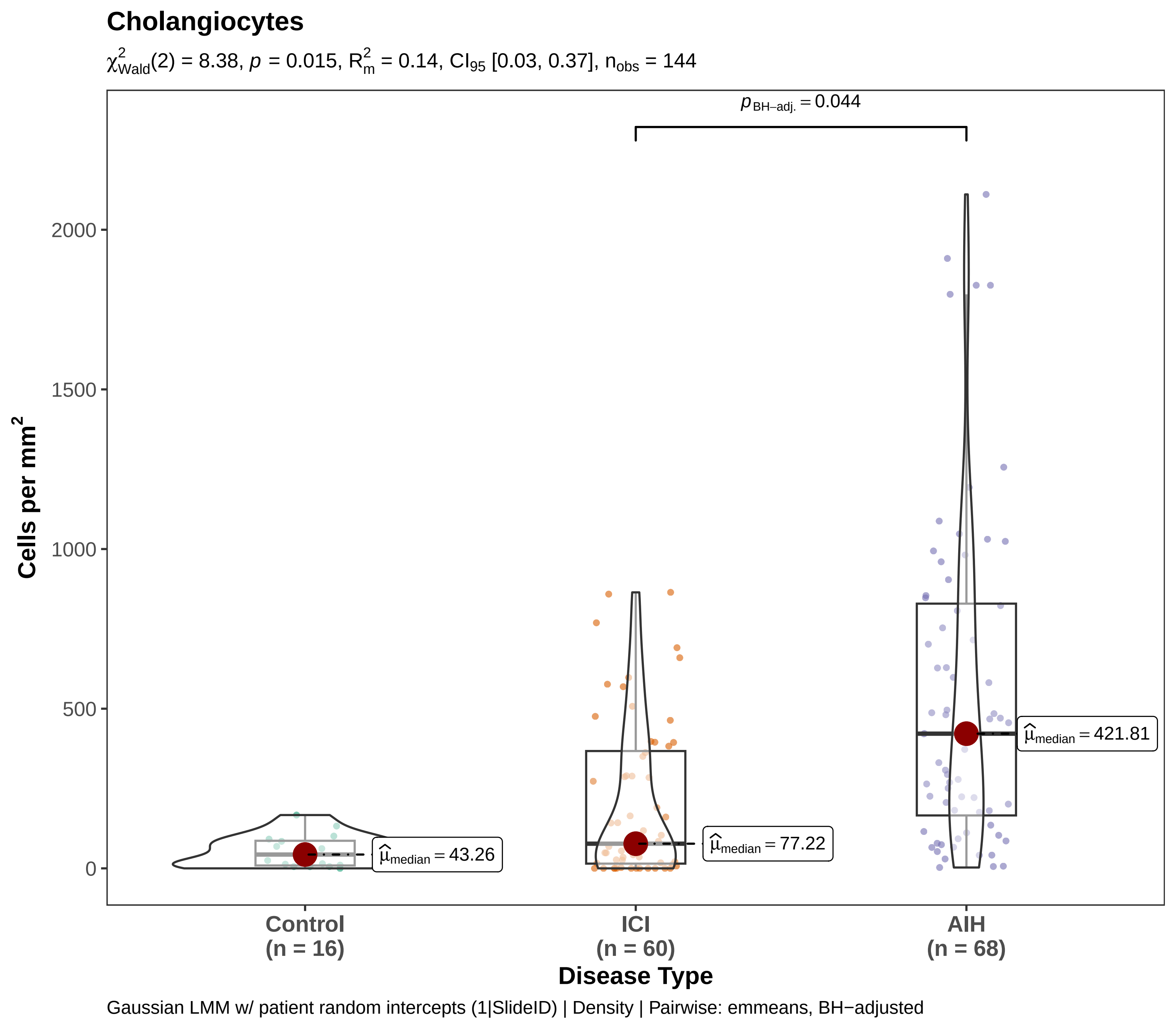
Cholangiocytes
$\chi^2_{Wald}(2) = 8.38$, $p = 0.015$, $R^2_m = 0.14$, $CI_{95}$ [0.03, 0.37], $n_{obs} = 144$
$p_{BH-adj.} = 0.044$
Cells per mm$^2$
2000
1500
1000
500
0
$\hat{\mu}_{median} = 43.26$
$\hat{\mu}_{median} = 77.22$
$\hat{\mu}_{median} = 421.81$
Control
(n = 16)
ICI
(n = 60)
AIH
(n = 68)
Disease Type
Gaussian LMM w/ patient random intercepts (1|SlideID) | Density | Pairwise: emmeans, BH−adjusted

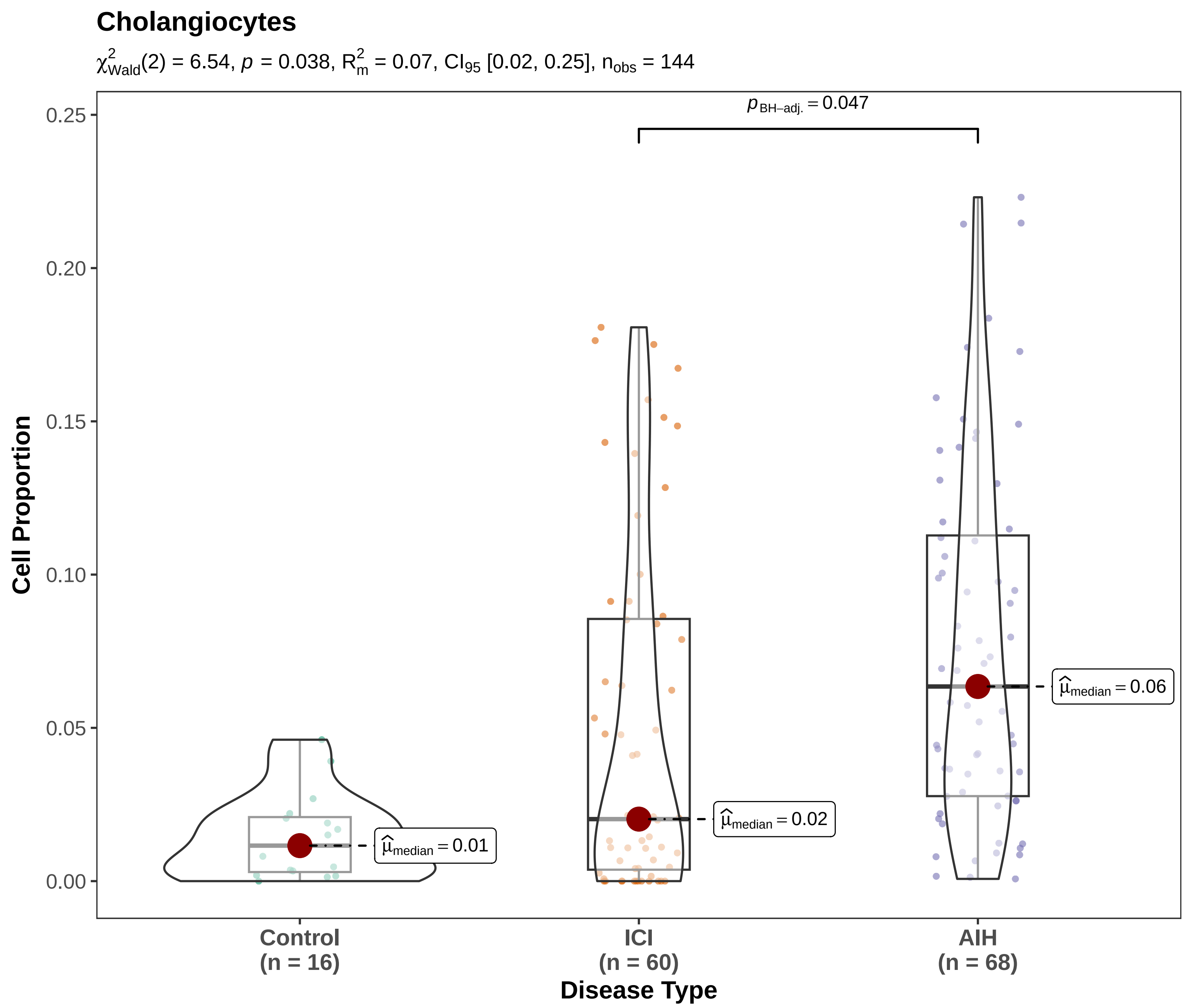


Binomial GLMM w/ patient random intercepts (1|SlideID) | Proportion | Pairwise: emmeans, BH−adjusted

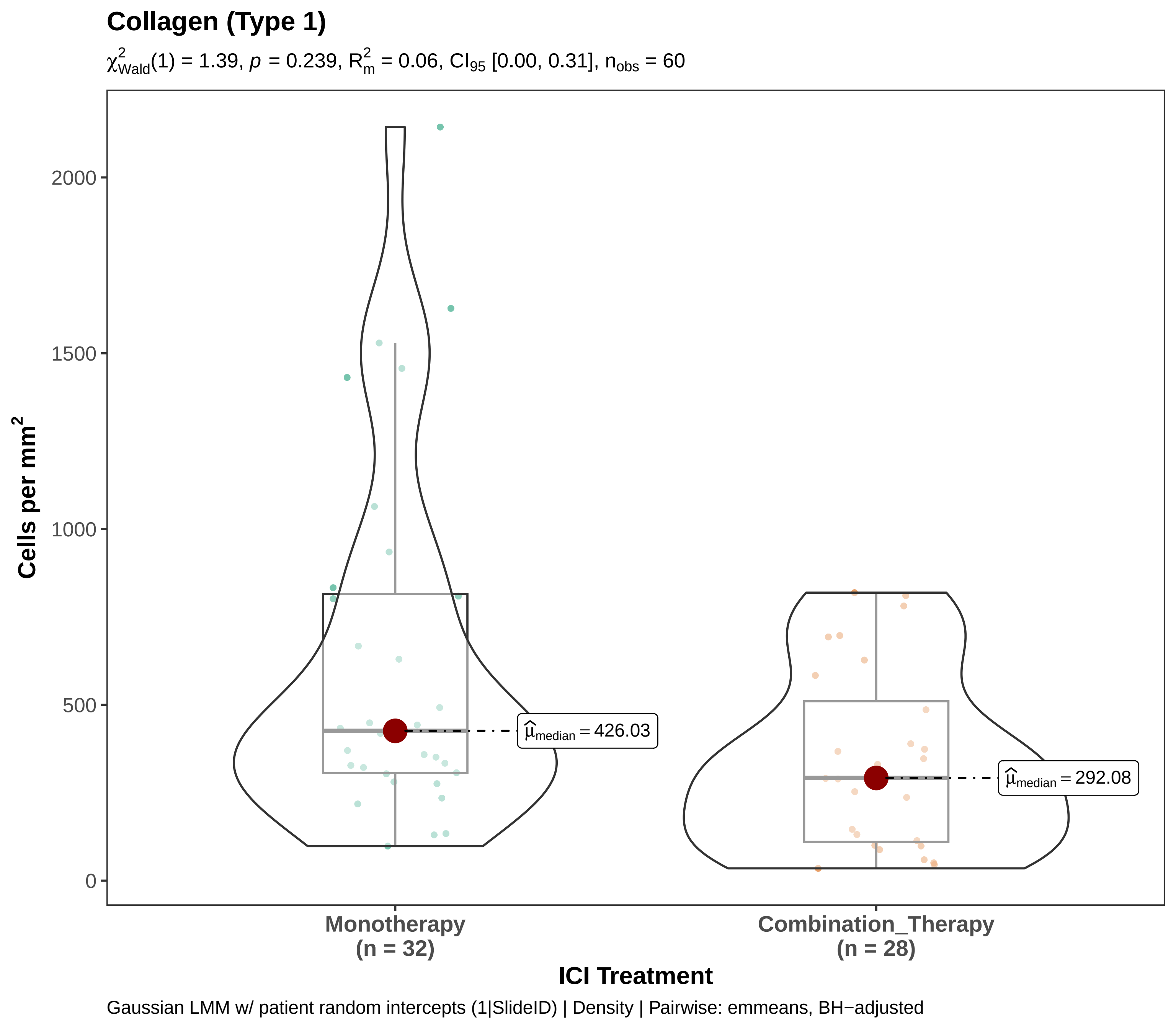
Collagen (Type 1)
$\chi^2_{Wald}(1) = 1.39$, $p = 0.239$, $R^2_m = 0.06$, $CI_{95}$ [0.00, 0.31], $n_{obs} = 60$
Cells per mm²
2000
1500
1000
500
0
$\hat{\mu}_{median} = 426.03$
$\hat{\mu}_{median} = 292.08$
Monotherapy
(n = 32)
Combination_Therapy
(n = 28)
ICI Treatment
Gaussian LMM w/ patient random intercepts (1|SlideID) | Density | Pairwise: emmeans, BH−adjusted

**Collagen (Type 1)**

$\chi^2_{Wald}(1) = 1.59$, $p = 0.208$, $R^2_m = 0.02$, $CI_{95}$ [0.00, 0.09], $n_{obs} = 60$

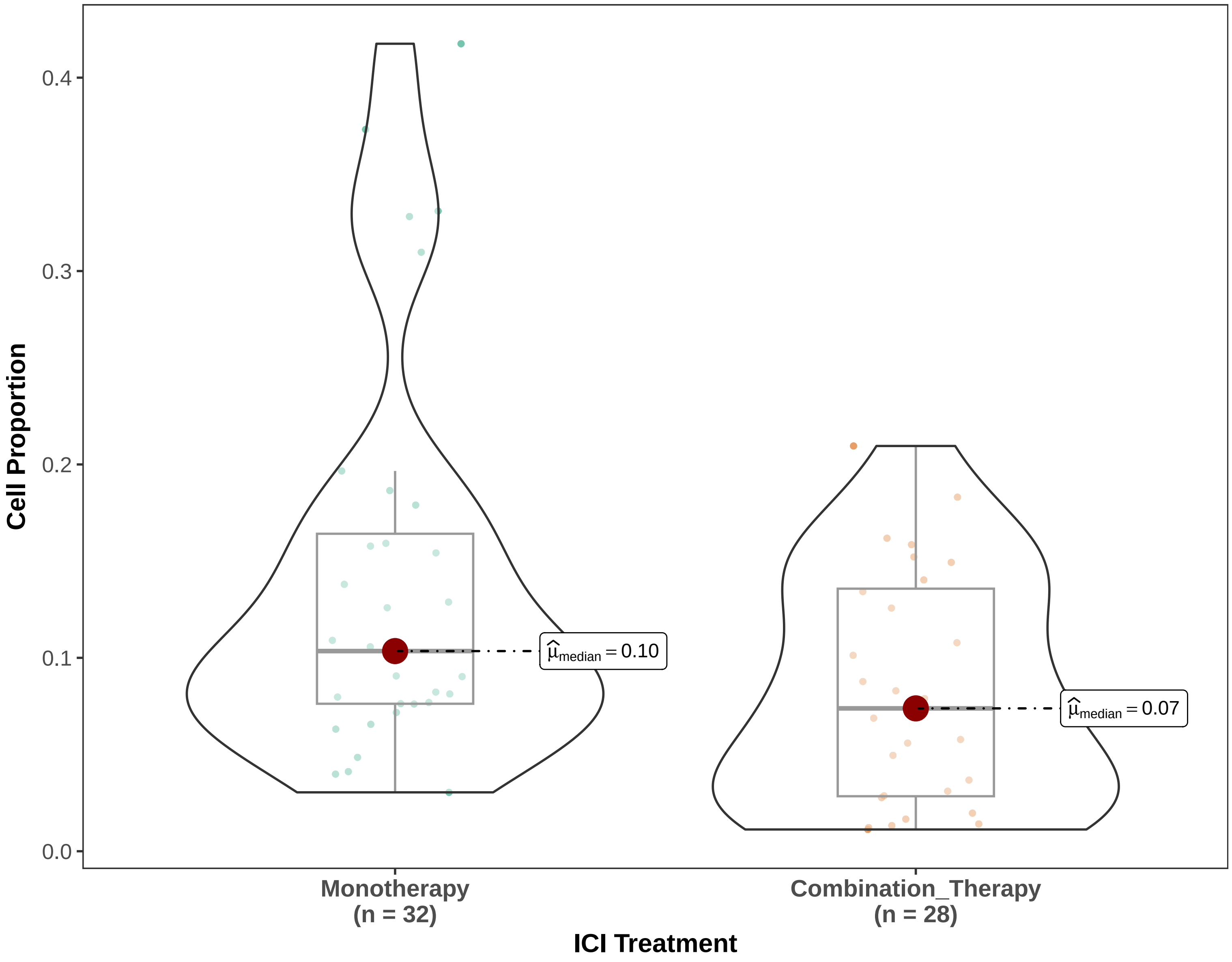


Binomial GLMM w/ patient random intercepts (1|SlideID) | Proportion | Pairwise: emmeans, BH−adjusted

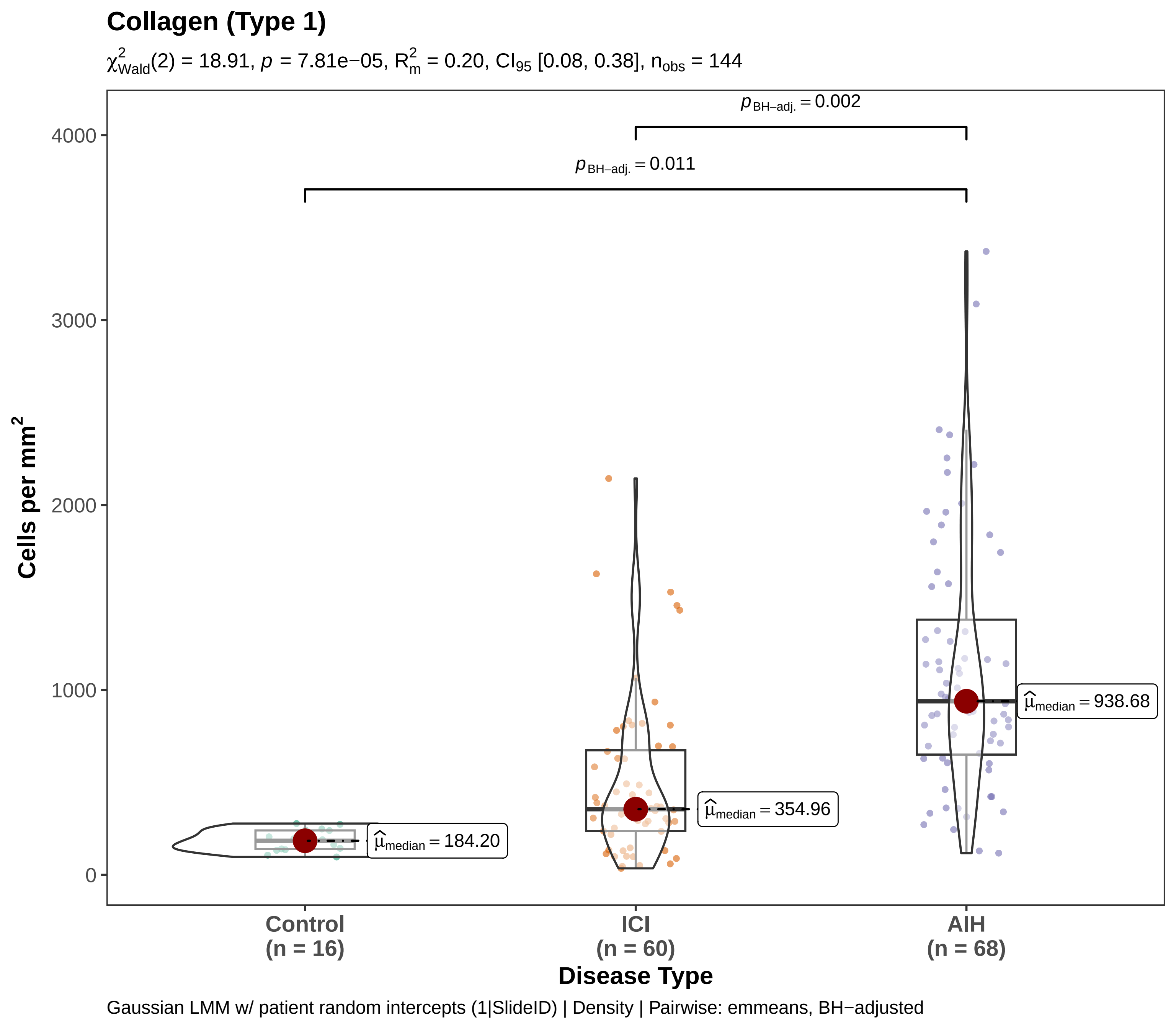

Collagen (Type 1)
$\chi^2_{Wald}(2) = 18.91$, $p = 7.81e{-}05$, $R^2_m = 0.20$, $CI_{95}$ [0.08, 0.38], $n_{obs} = 144$
$p_{BH-adj.} = 0.002$
$p_{BH-adj.} = 0.011$
4000
3000
2000
1000
0
Cells per mm$^2$
$\hat{\mu}_{median} = 184.20$
$\hat{\mu}_{median} = 354.96$
$\hat{\mu}_{median} = 938.68$
Control
(n = 16)
ICI
(n = 60)
AIH
(n = 68)
Disease Type
Gaussian LMM w/ patient random intercepts (1|SlideID) | Density | Pairwise: emmeans, BH−adjusted

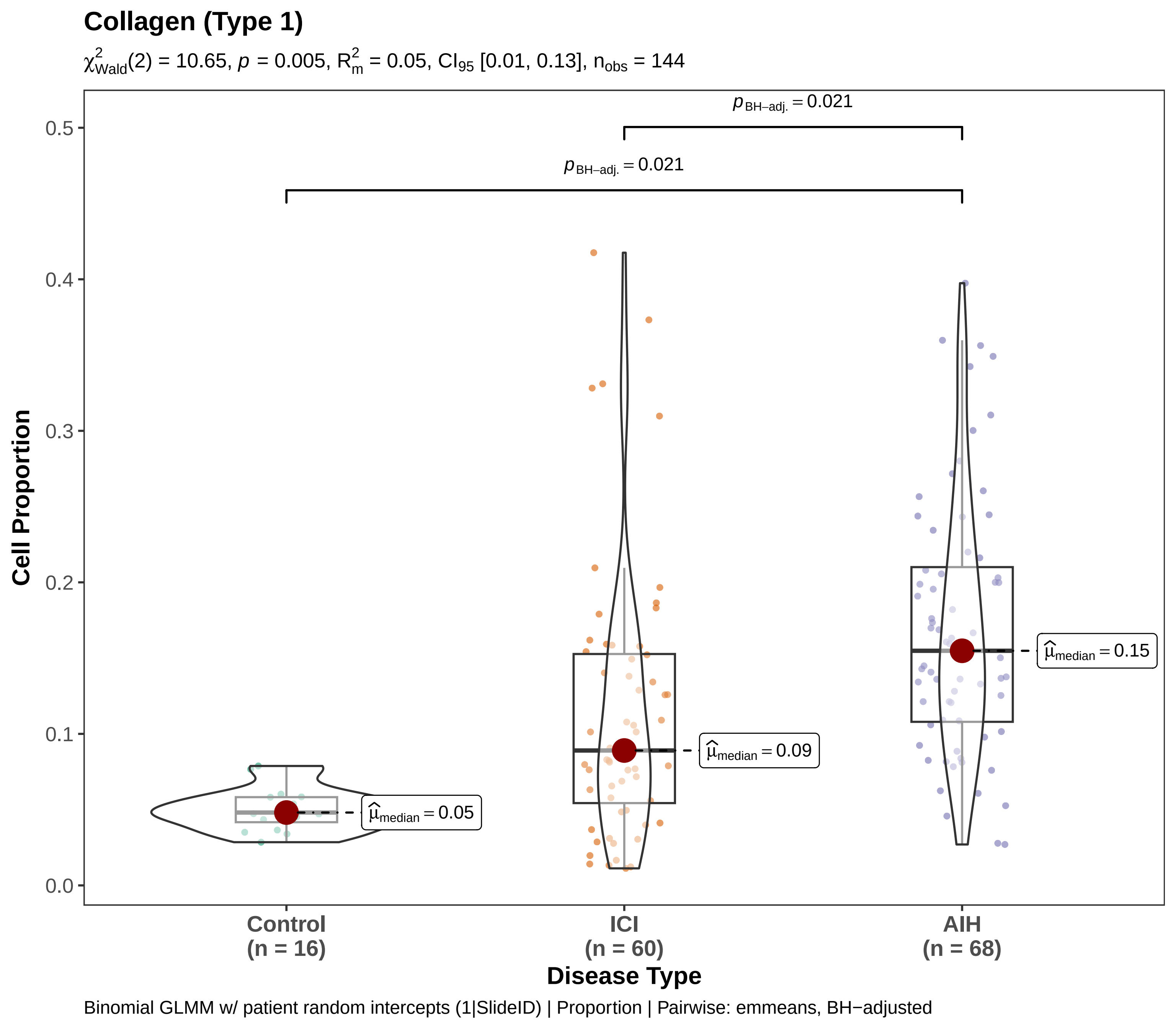

Collagen (Type 1)
$\chi^2_{\mathrm{Wald}}(2) = 10.65$, $p = 0.005$, $R^2_m = 0.05$, $CI_{95}$ [0.01, 0.13], $n_{obs} = 144$
$p_{\mathrm{BH-adj.}} = 0.021$
$p_{\mathrm{BH-adj.}} = 0.021$
$\widehat{\mu}_{\mathrm{median}} = 0.05$
$\widehat{\mu}_{\mathrm{median}} = 0.09$
$\widehat{\mu}_{\mathrm{median}} = 0.15$
Cell Proportion
0.0
0.1
0.2
0.3
0.4
0.5
Control (n = 16)
ICI (n = 60)
AIH (n = 68)
Disease Type
Binomial GLMM w/ patient random intercepts (1|SlideID) | Proportion | Pairwise: emmeans, BH−adjusted

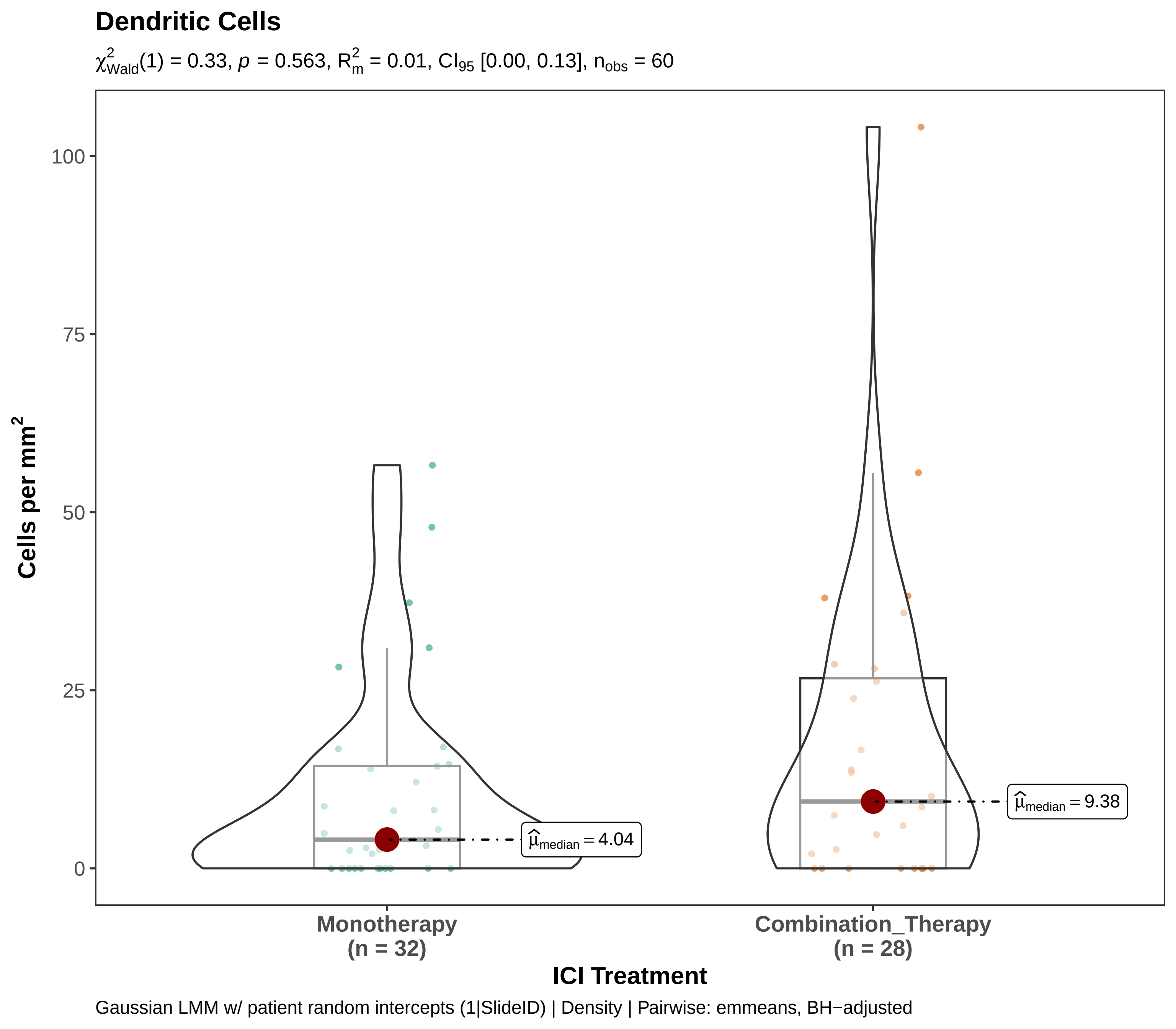

Dendritic Cells
$\chi^2_{Wald}(1) = 0.33$, $p = 0.563$, $R^2_m = 0.01$, $CI_{95}$ [0.00, 0.13], $n_{obs} = 60$
Cells per mm$^2$
100
75
50
25
0
$\widehat{\mu}_{median} = 4.04$
$\widehat{\mu}_{median} = 9.38$
Monotherapy
(n = 32)
Combination_Therapy
(n = 28)
ICI Treatment
Gaussian LMM w/ patient random intercepts (1|SlideID) | Density | Pairwise: emmeans, BH−adjusted

#### Dendritic Cells

$\chi^2_{Wald}(1) = 0.14$, $p = 0.705$, $R^2_m = 0.00$, $CI_{95}$ [0.00, 0.13], $n_{obs} = 60$

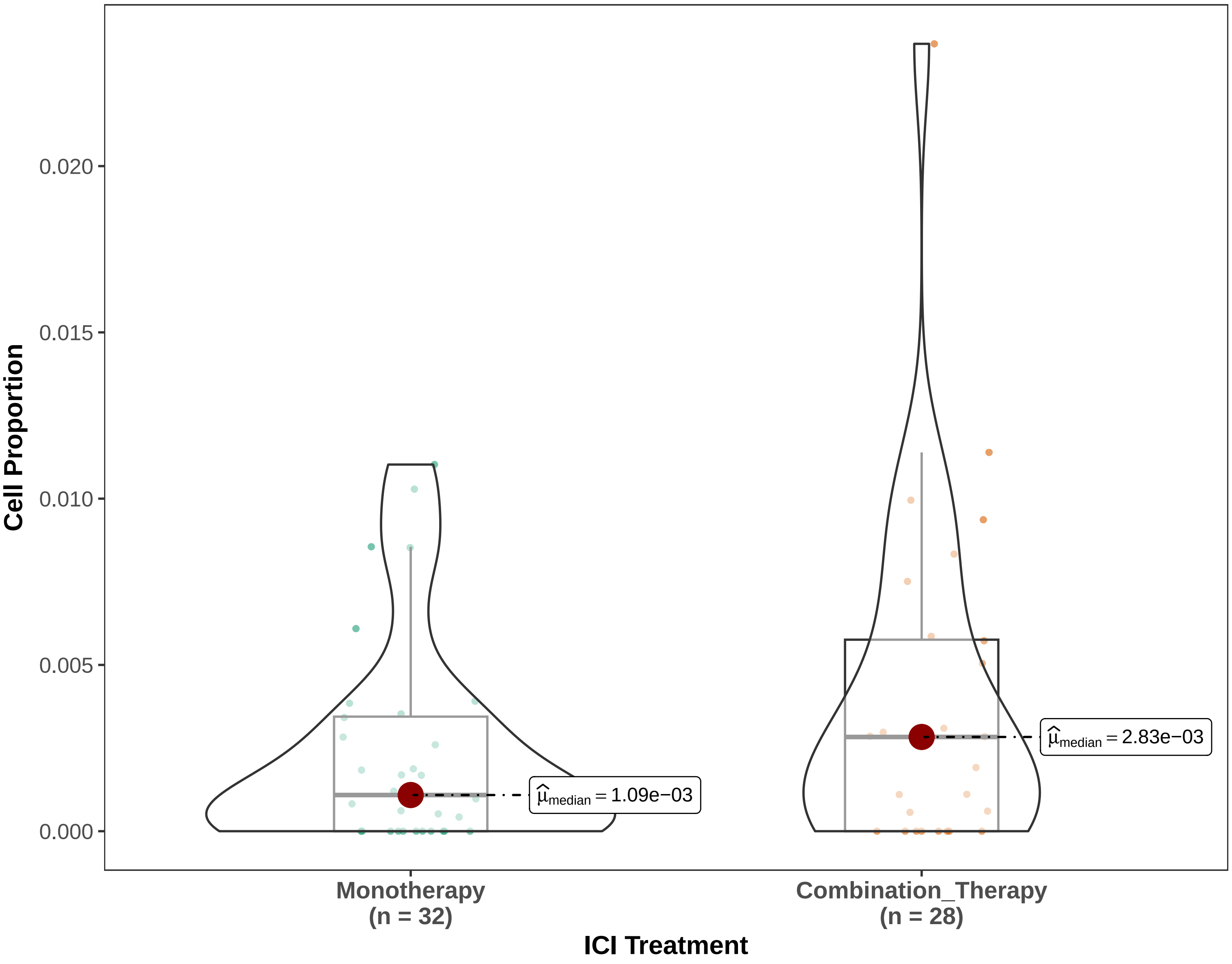


Binomial GLMM w/ patient random intercepts (1|SlideID) | Proportion | Pairwise: emmeans, BH−adjusted

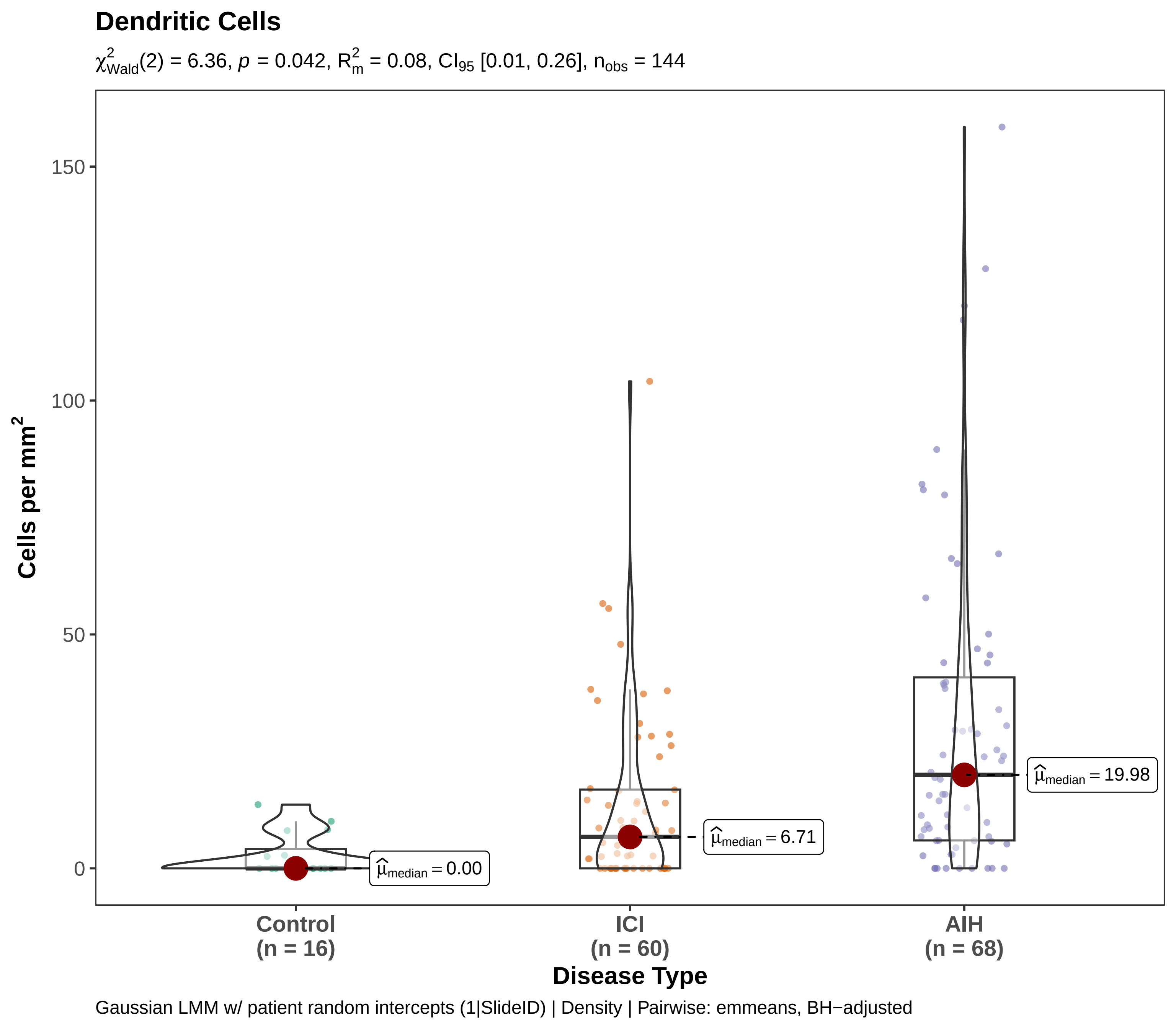
Dendritic Cells
$\chi^2_{Wald}(2) = 6.36$, $p = 0.042$, $R^2_m = 0.08$, $CI_{95}$ [0.01, 0.26], $n_{obs} = 144$
Cells per mm²
150
100
50
0
$\widehat{\mu}_{median} = 0.00$
$\widehat{\mu}_{median} = 6.71$
$\widehat{\mu}_{median} = 19.98$
Control
(n = 16)
ICI
(n = 60)
AIH
(n = 68)
Disease Type
Gaussian LMM w/ patient random intercepts (1|SlideID) | Density | Pairwise: emmeans, BH−adjusted

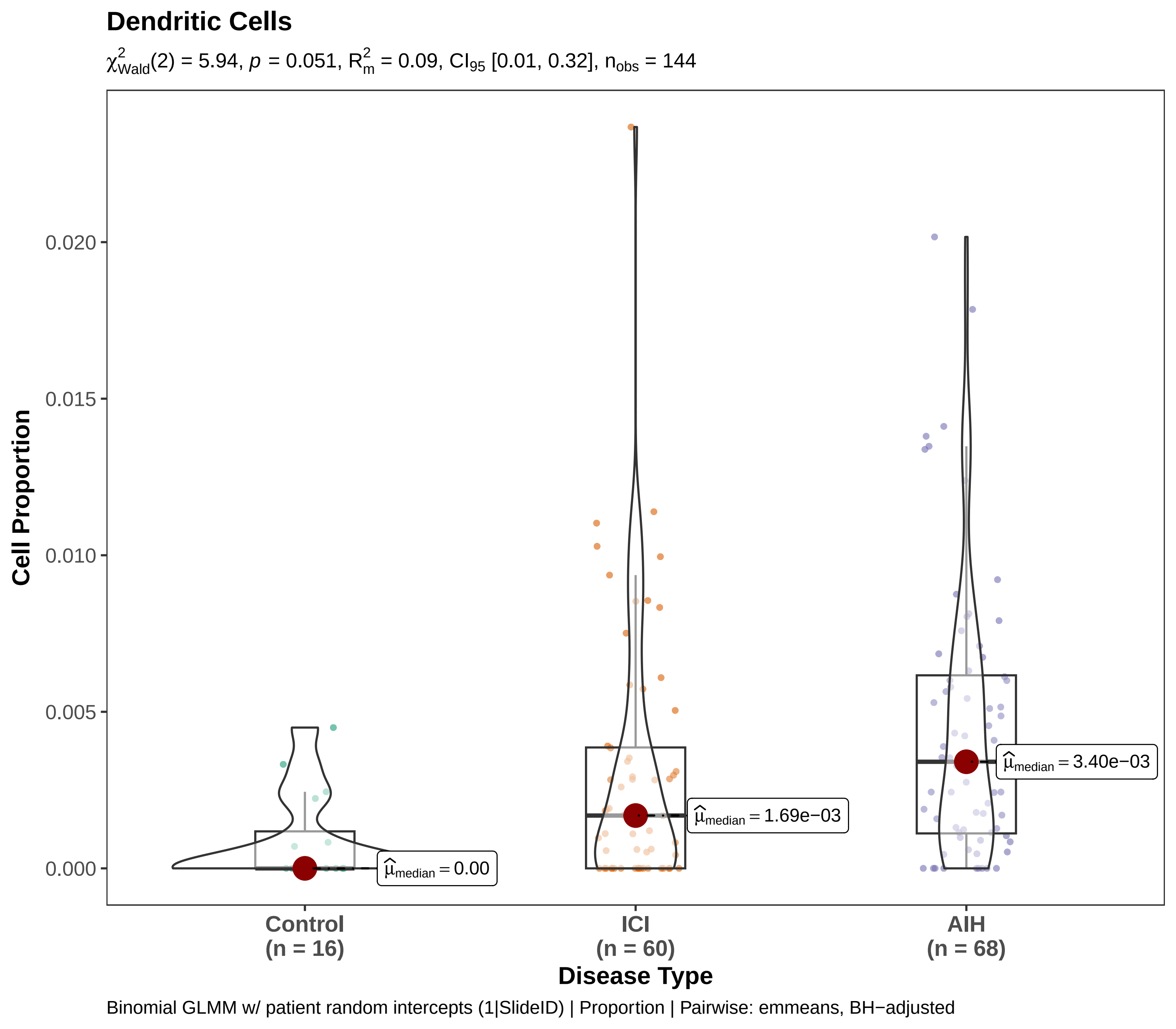

Dendritic Cells
$\chi^2_{Wald}(2) = 5.94$, $p = 0.051$, $R^2_m = 0.09$, $CI_{95}$ [0.01, 0.32], $n_{obs} = 144$
Cell Proportion
0.020
0.015
0.010
0.005
0.000
$\widehat{\mu}_{median} = 0.00$
$\widehat{\mu}_{median} = 1.69e-03$
$\widehat{\mu}_{median} = 3.40e-03$
Control
(n = 16)
ICI
(n = 60)
AIH
(n = 68)
Disease Type
Binomial GLMM w/ patient random intercepts (1|SlideID) | Proportion | Pairwise: emmeans, BH−adjusted

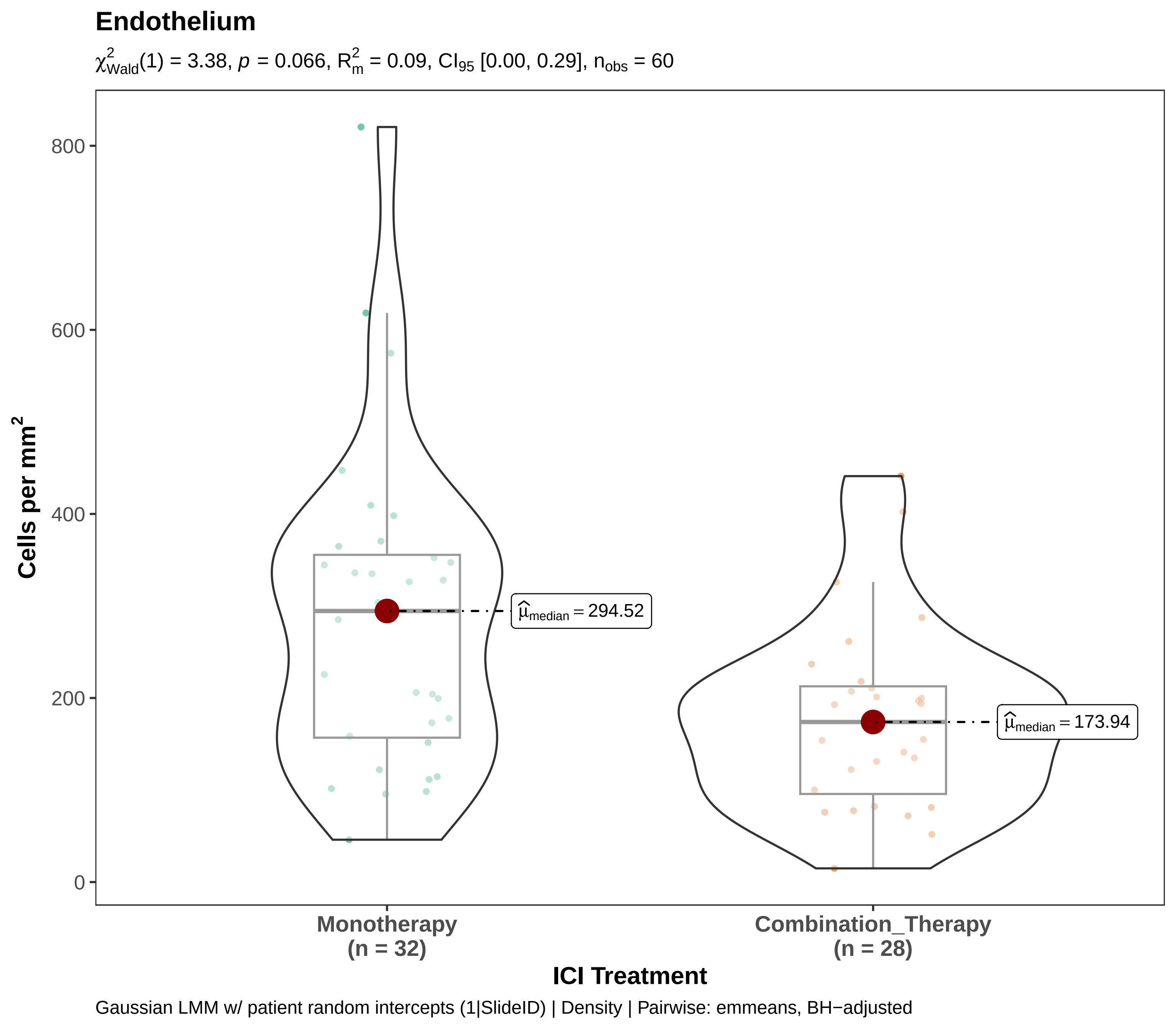

Endothelium
$\chi^2_{Wald}(1) = 3.38$, $p = 0.066$, $R^2_m = 0.09$, $CI_{95}$ [0.00, 0.29], $n_{obs} = 60$
Cells per mm$^2$
800
600
400
200
0
$\widehat{\mu}_{median} = 294.52$
$\widehat{\mu}_{median} = 173.94$
Monotherapy
(n = 32)
Combination_Therapy
(n = 28)
ICI Treatment
Gaussian LMM w/ patient random intercepts (1|SlideID) | Density | Pairwise: emmeans, BH−adjusted

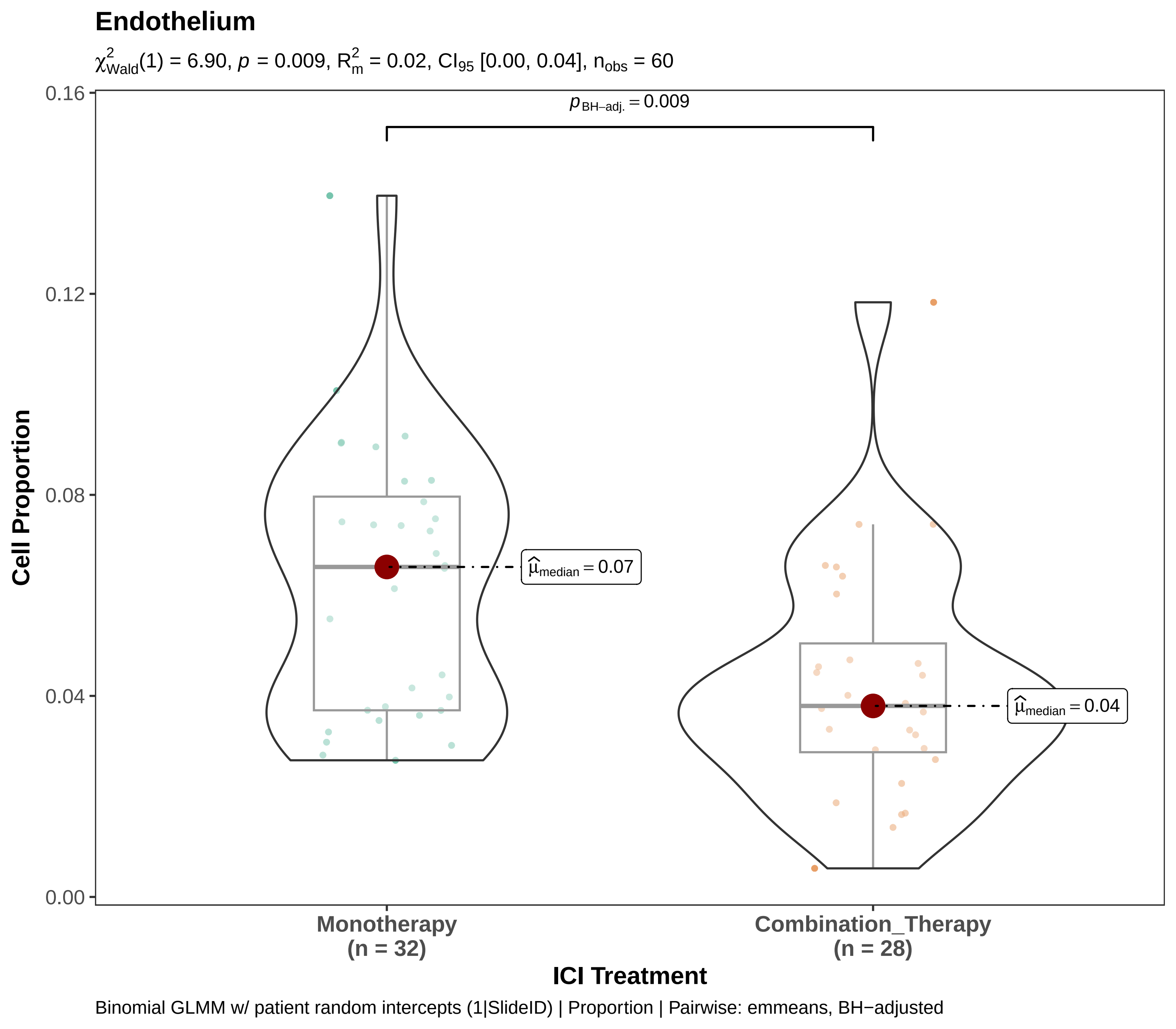
Endothelium
$\chi^2_{Wald}(1)$ = 6.90, $p$ = 0.009, $R^2_m$ = 0.02, $CI_{95}$ [0.00, 0.04], $n_{obs}$ = 60
$p_{BH-adj.} = 0.009$
$\hat{\mu}_{median} = 0.07$
$\hat{\mu}_{median} = 0.04$
0.16
0.12
0.08
0.04
0.00
Cell Proportion
Monotherapy
(n = 32)
Combination_Therapy
(n = 28)
ICI Treatment
Binomial GLMM w/ patient random intercepts (1|SlideID) | Proportion | Pairwise: emmeans, BH−adjusted

### Endothelium

$\chi^2_{Wald}(2) = 6.17$, $p = 0.046$, $R^2_m = 0.06$, $CI_{95}$ [0.01, 0.19], $n_{obs} = 144$

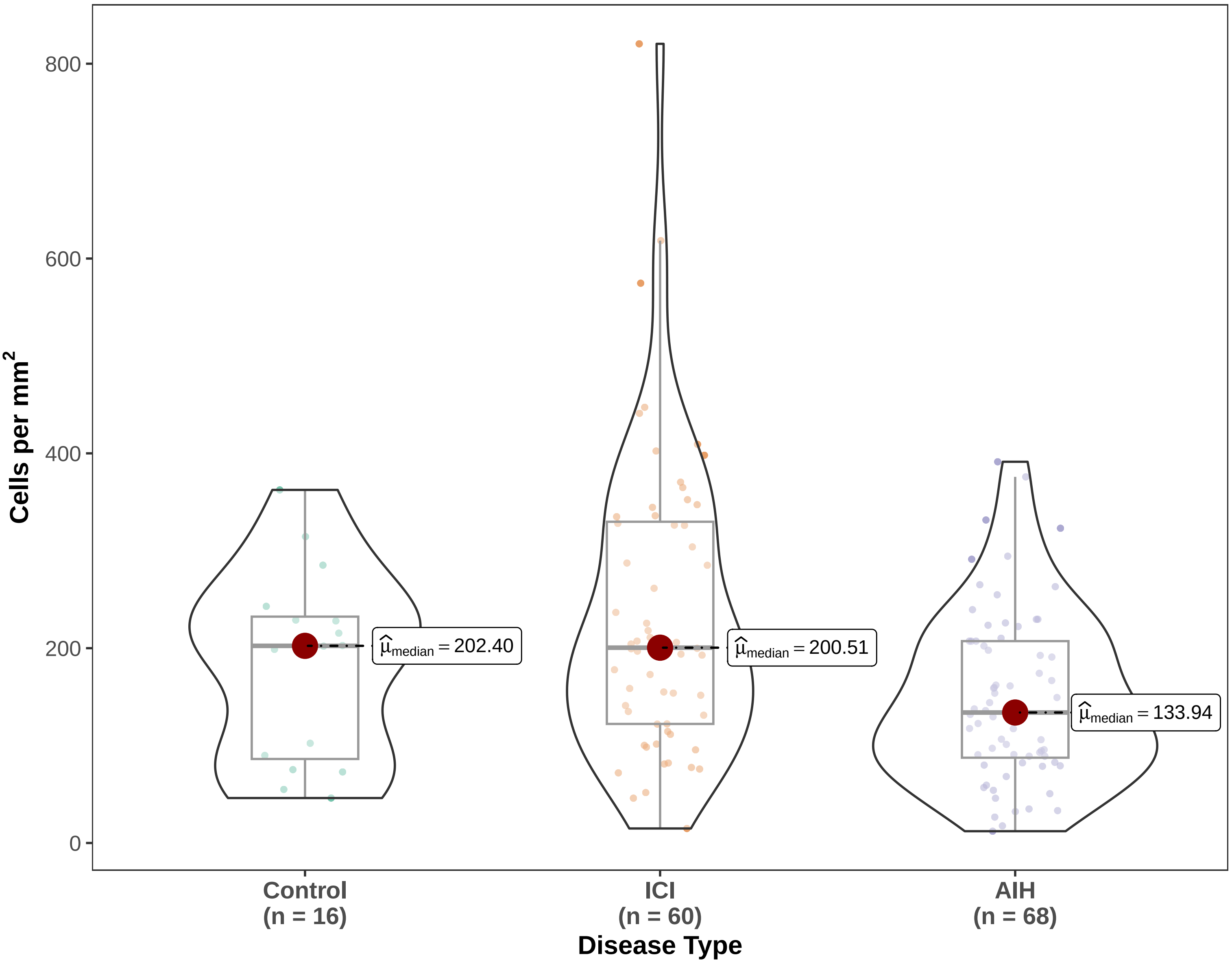


Gaussian LMM w/ patient random intercepts (1|SlideID) | Density | Pairwise: emmeans, BH−adjusted

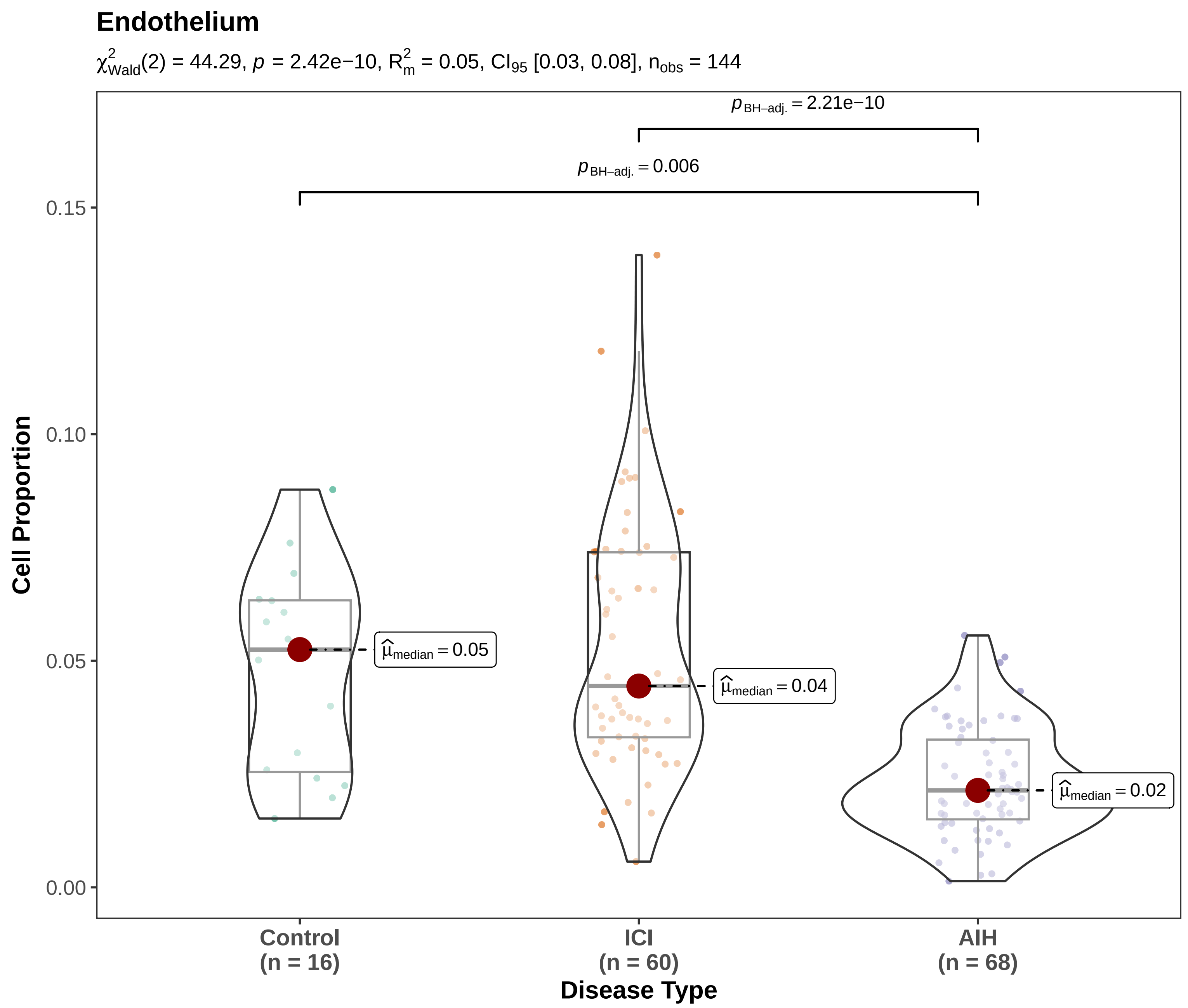

Binomial GLMM w/ patient random intercepts (1|SlideID) | Proportion | Pairwise: emmeans, BH−adjusted

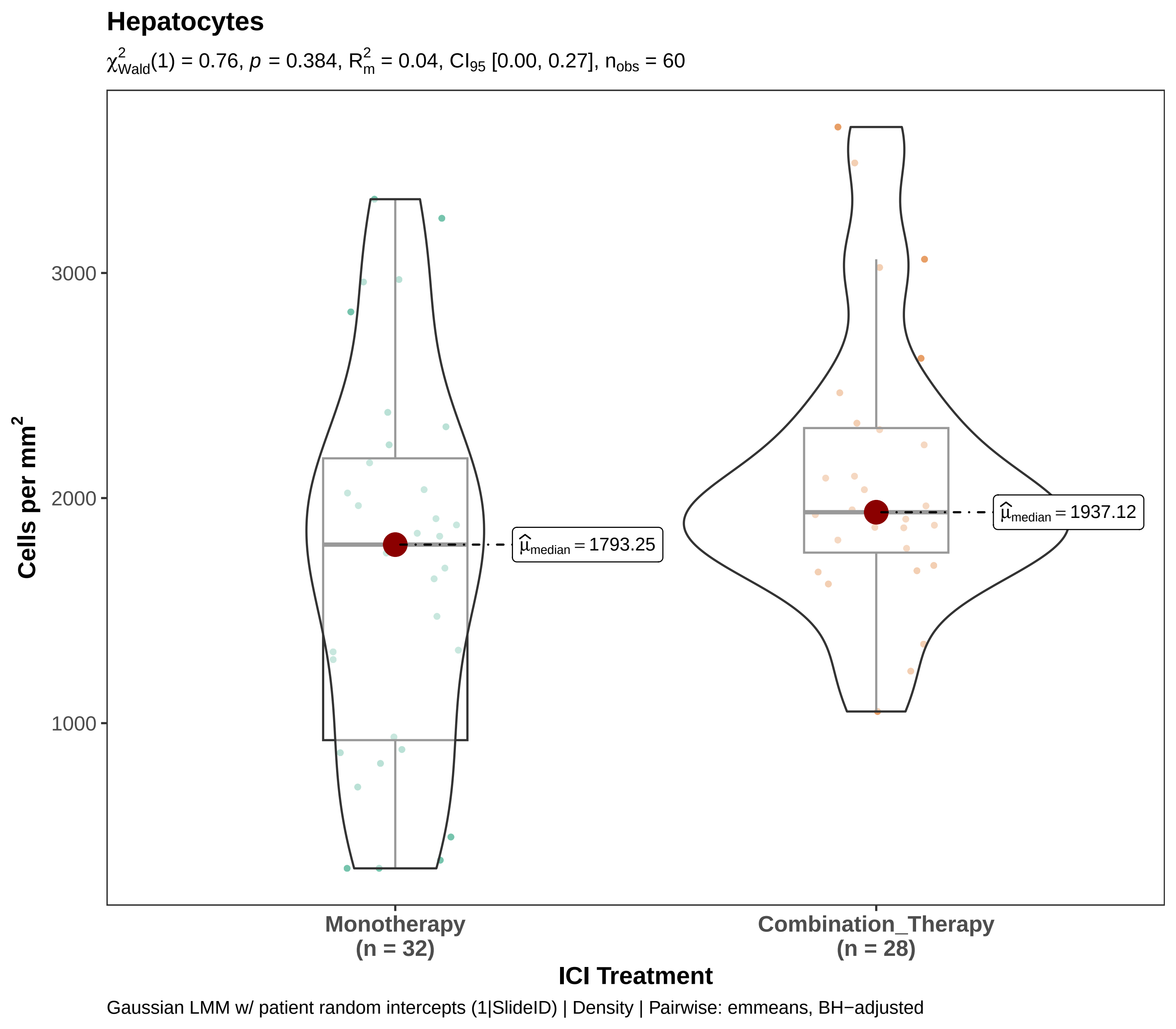

Hepatocytes
$\chi^2_{Wald}(1) = 0.76$, $p = 0.384$, $R^2_m = 0.04$, $CI_{95}$ [0.00, 0.27], $n_{obs} = 60$
Cells per mm²
3000
2000
1000
$\widehat{\mu}_{median} = 1793.25$
$\widehat{\mu}_{median} = 1937.12$
Monotherapy
(n = 32)
Combination_Therapy
(n = 28)
ICI Treatment
Gaussian LMM w/ patient random intercepts (1|SlideID) | Density | Pairwise: emmeans, BH−adjusted

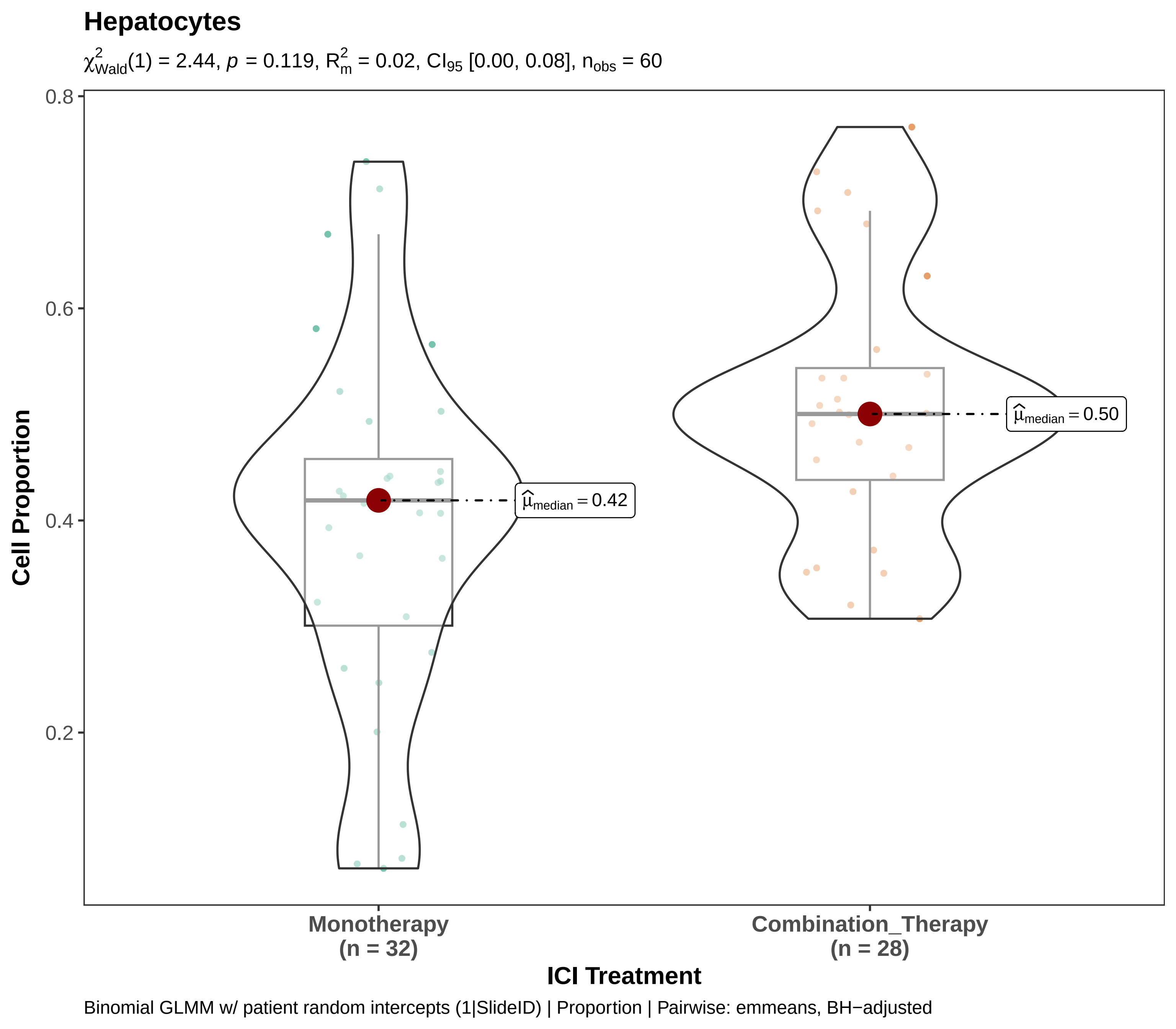

Hepatocytes
$\chi^2_{Wald}(1) = 2.44$, $p = 0.119$, $R^2_m = 0.02$, $CI_{95}$ [0.00, 0.08], $n_{obs} = 60$
0.8
0.6
0.4
0.2
Cell Proportion
$\widehat{\mu}_{median} = 0.42$
$\widehat{\mu}_{median} = 0.50$
Monotherapy
(n = 32)
Combination_Therapy
(n = 28)
ICI Treatment
Binomial GLMM w/ patient random intercepts (1|SlideID) | Proportion | Pairwise: emmeans, BH−adjusted

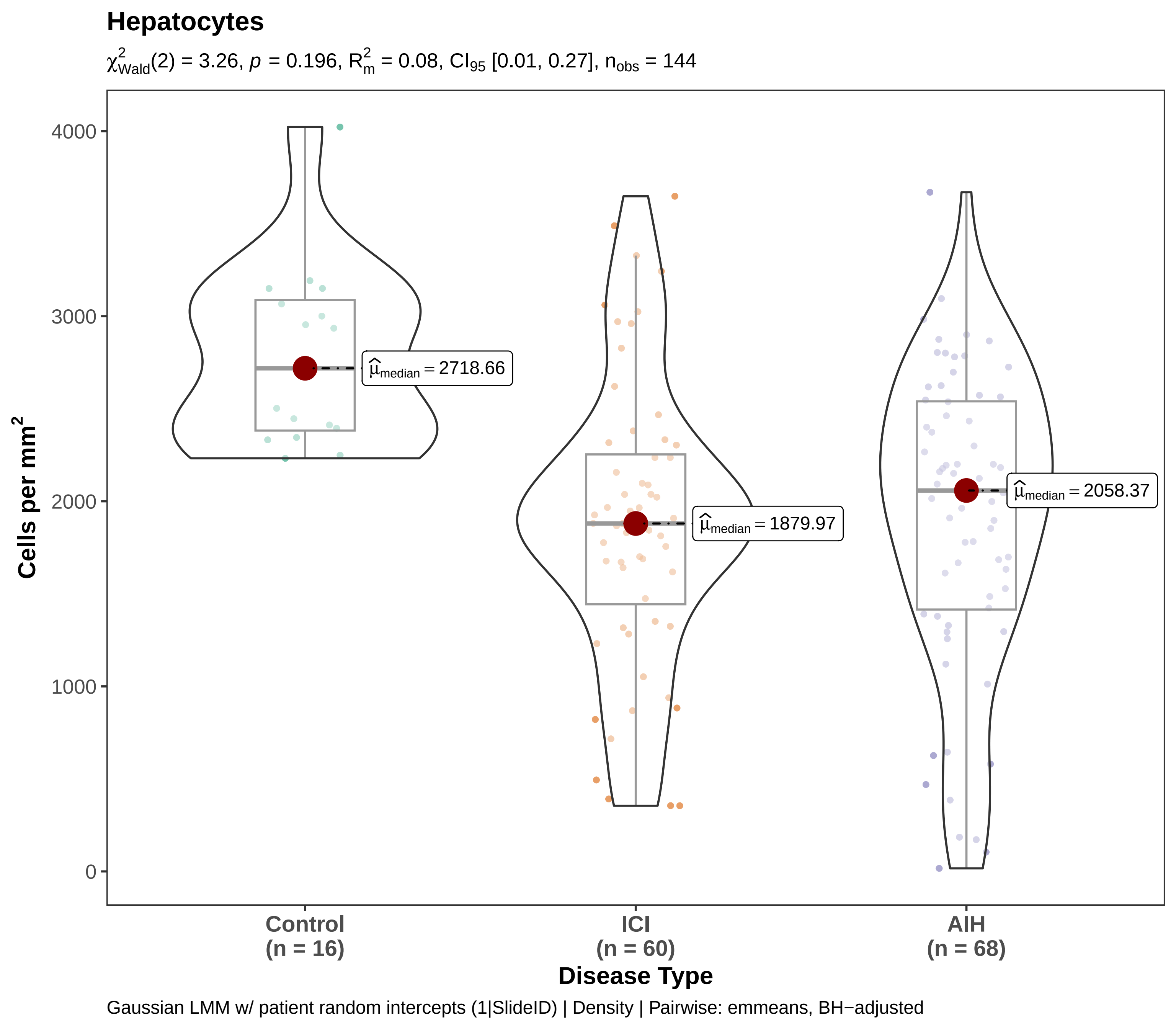

Hepatocytes
$\chi^2_{Wald}(2) = 3.26$, $p = 0.196$, $R^2_m = 0.08$, $CI_{95}$ [0.01, 0.27], $n_{obs} = 144$
4000
3000
2000
1000
0
Cells per mm$^2$
$\hat{\mu}_{median} = 2718.66$
$\hat{\mu}_{median} = 1879.97$
$\hat{\mu}_{median} = 2058.37$
Control
(n = 16)
ICI
(n = 60)
AIH
(n = 68)
Disease Type
Gaussian LMM w/ patient random intercepts (1|SlideID) | Density | Pairwise: emmeans, BH−adjusted

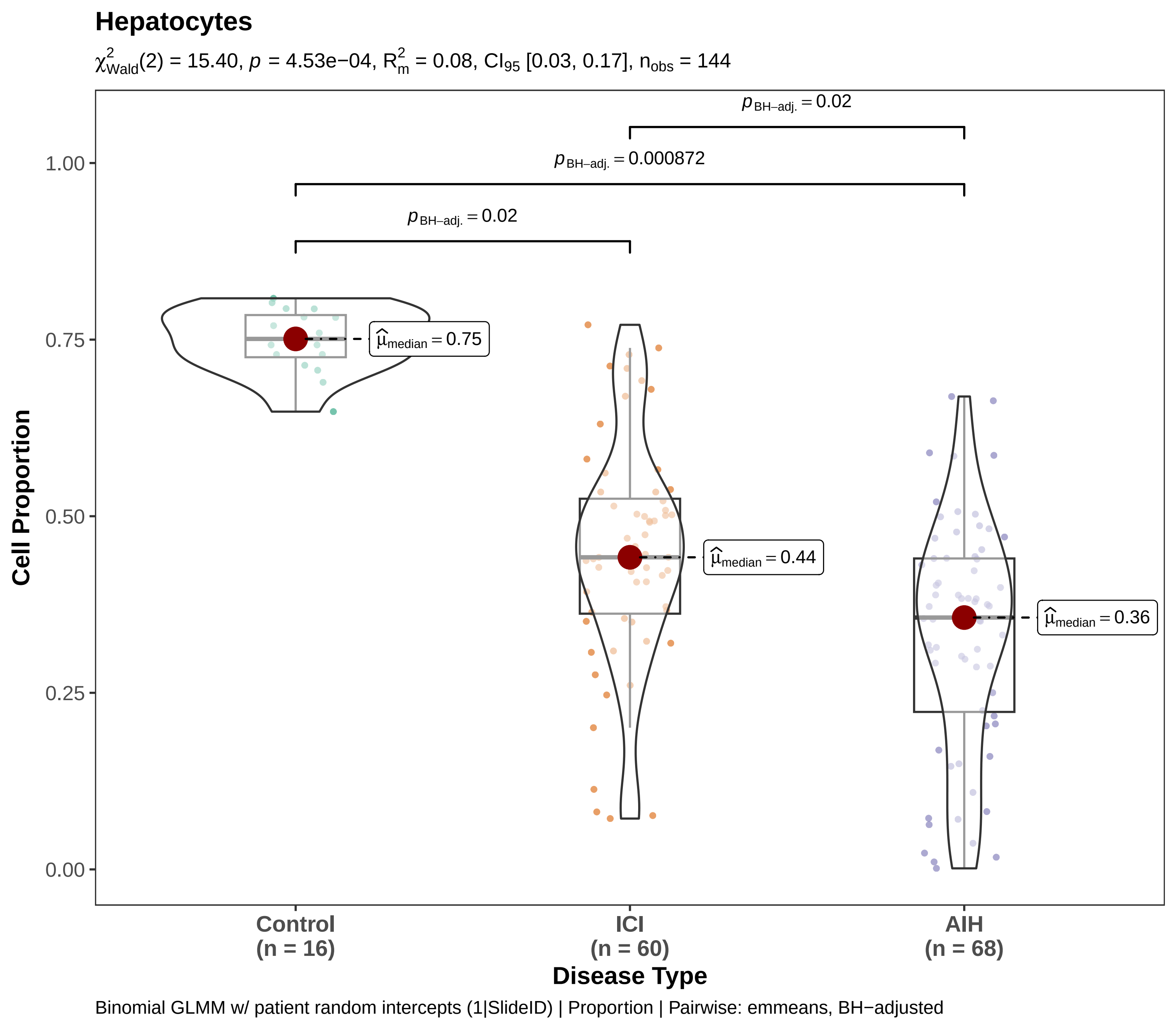
Hepatocytes
$\chi^2_{Wald}(2) = 15.40$, $p = 4.53e{-}04$, $R^2_m = 0.08$, $CI_{95}$ [0.03, 0.17], $n_{obs} = 144$
$p_{BH-adj.} = 0.02$
$p_{BH-adj.} = 0.000872$
$p_{BH-adj.} = 0.02$
$\hat{\mu}_{median} = 0.75$
$\hat{\mu}_{median} = 0.44$
$\hat{\mu}_{median} = 0.36$
1.00
0.75
0.50
0.25
0.00
Cell Proportion
Control (n = 16)
ICI (n = 60)
AIH (n = 68)
Disease Type
Binomial GLMM w/ patient random intercepts (1|SlideID) | Proportion | Pairwise: emmeans, BH−adjusted

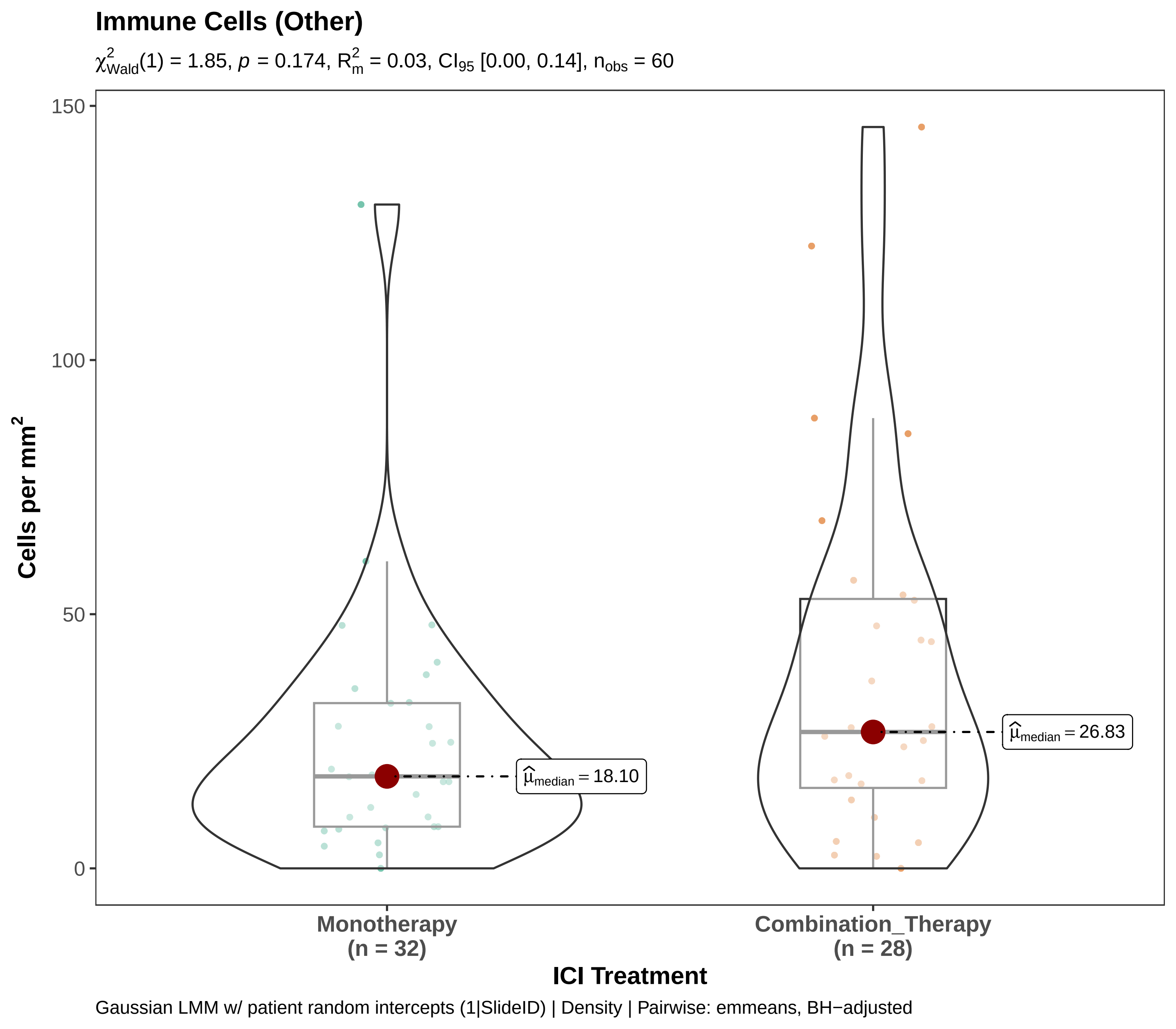

Immune Cells (Other)
$\chi^2_{Wald}(1) = 1.85$, $p = 0.174$, $R^2_m = 0.03$, $CI_{95}$ [0.00, 0.14], $n_{obs} = 60$
150
100
50
0
Cells per mm$^2$
$\widehat{\mu}_{median} = 18.10$
$\widehat{\mu}_{median} = 26.83$
Monotherapy
(n = 32)
Combination_Therapy
(n = 28)
ICI Treatment
Gaussian LMM w/ patient random intercepts (1|SlideID) | Density | Pairwise: emmeans, BH−adjusted

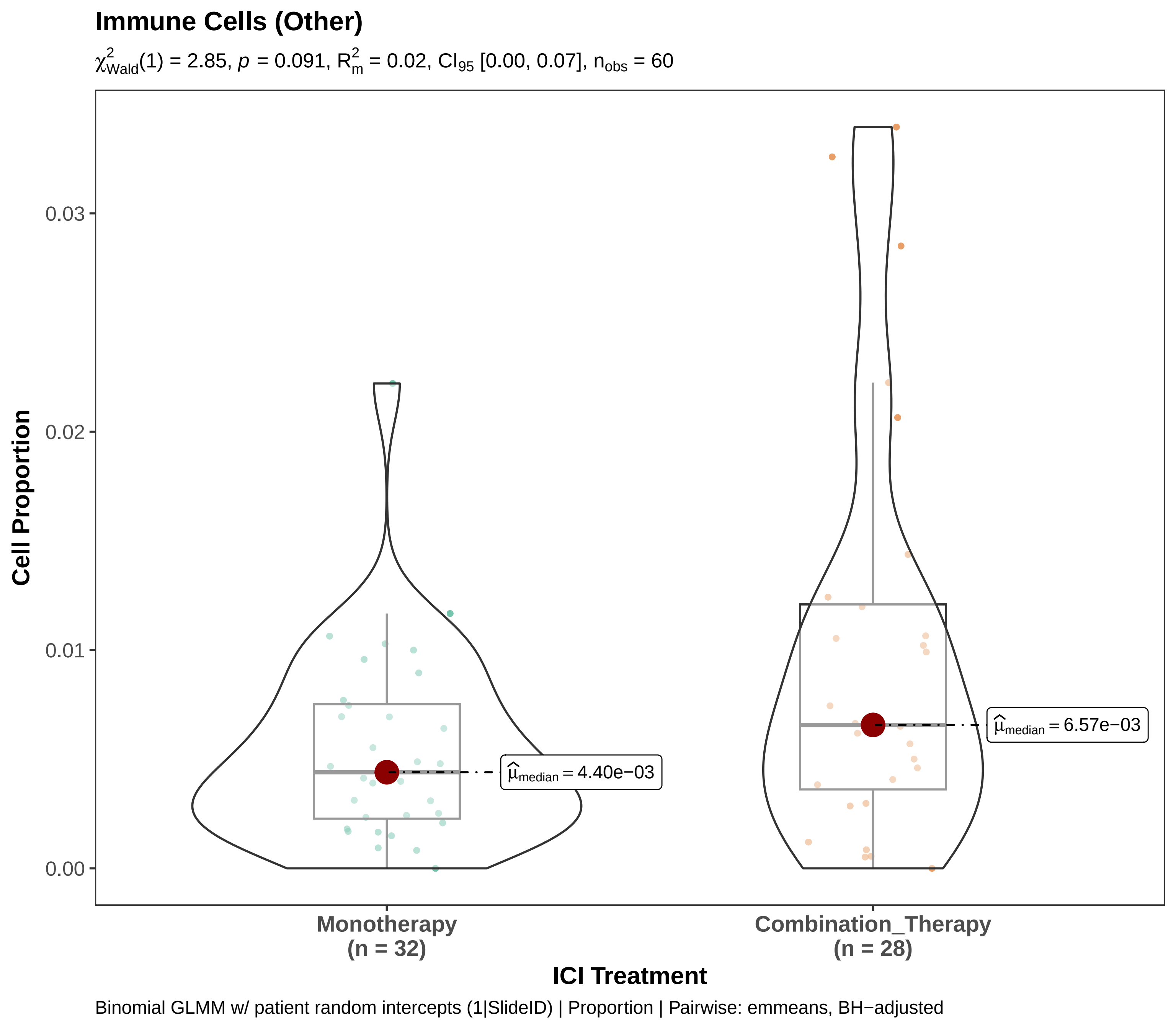
Immune Cells (Other)
$\chi^2_{Wald}(1) = 2.85$, $p = 0.091$, $R^2_m = 0.02$, $CI_{95}$ [0.00, 0.07], $n_{obs} = 60$
Cell Proportion
0.03
0.02
0.01
0.00
$\hat{\mu}_{median} = 4.40e-03$
$\hat{\mu}_{median} = 6.57e-03$
Monotherapy
(n = 32)
Combination_Therapy
(n = 28)
ICI Treatment
Binomial GLMM w/ patient random intercepts (1|SlideID) | Proportion | Pairwise: emmeans, BH−adjusted

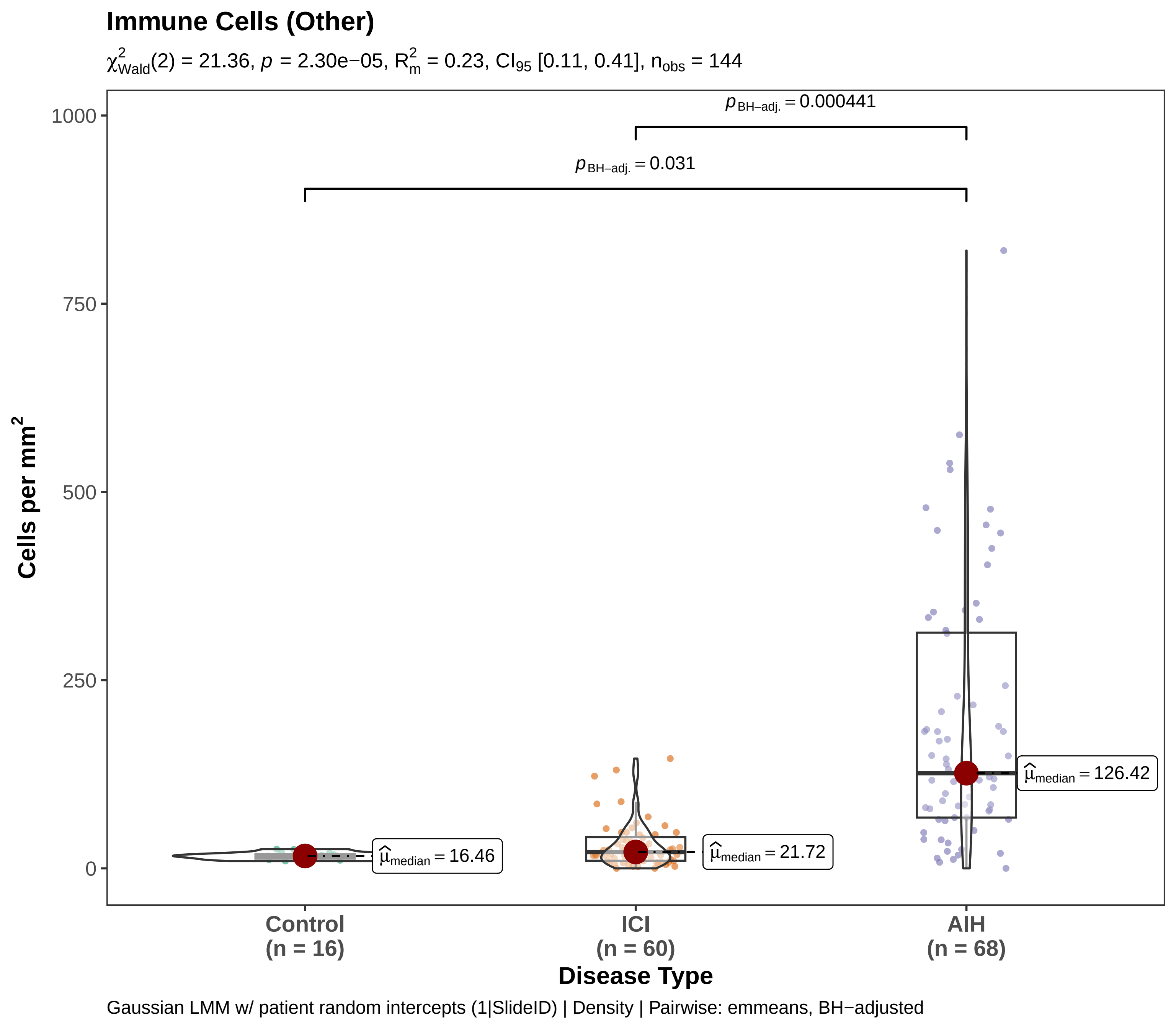

Immune Cells (Other)
$\chi^2_{Wald}(2) = 21.36$, $p = 2.30e-05$, $R^2_m = 0.23$, $CI_{95}$ [0.11, 0.41], $n_{obs} = 144$
$p_{BH-adj.} = 0.000441$
$p_{BH-adj.} = 0.031$
$\hat{\mu}_{median} = 16.46$
$\hat{\mu}_{median} = 21.72$
$\hat{\mu}_{median} = 126.42$
Cells per mm²
1000
750
500
250
0
Control (n = 16)
ICI (n = 60)
AIH (n = 68)
Disease Type
Gaussian LMM w/ patient random intercepts (1|SlideID) | Density | Pairwise: emmeans, BH−adjusted

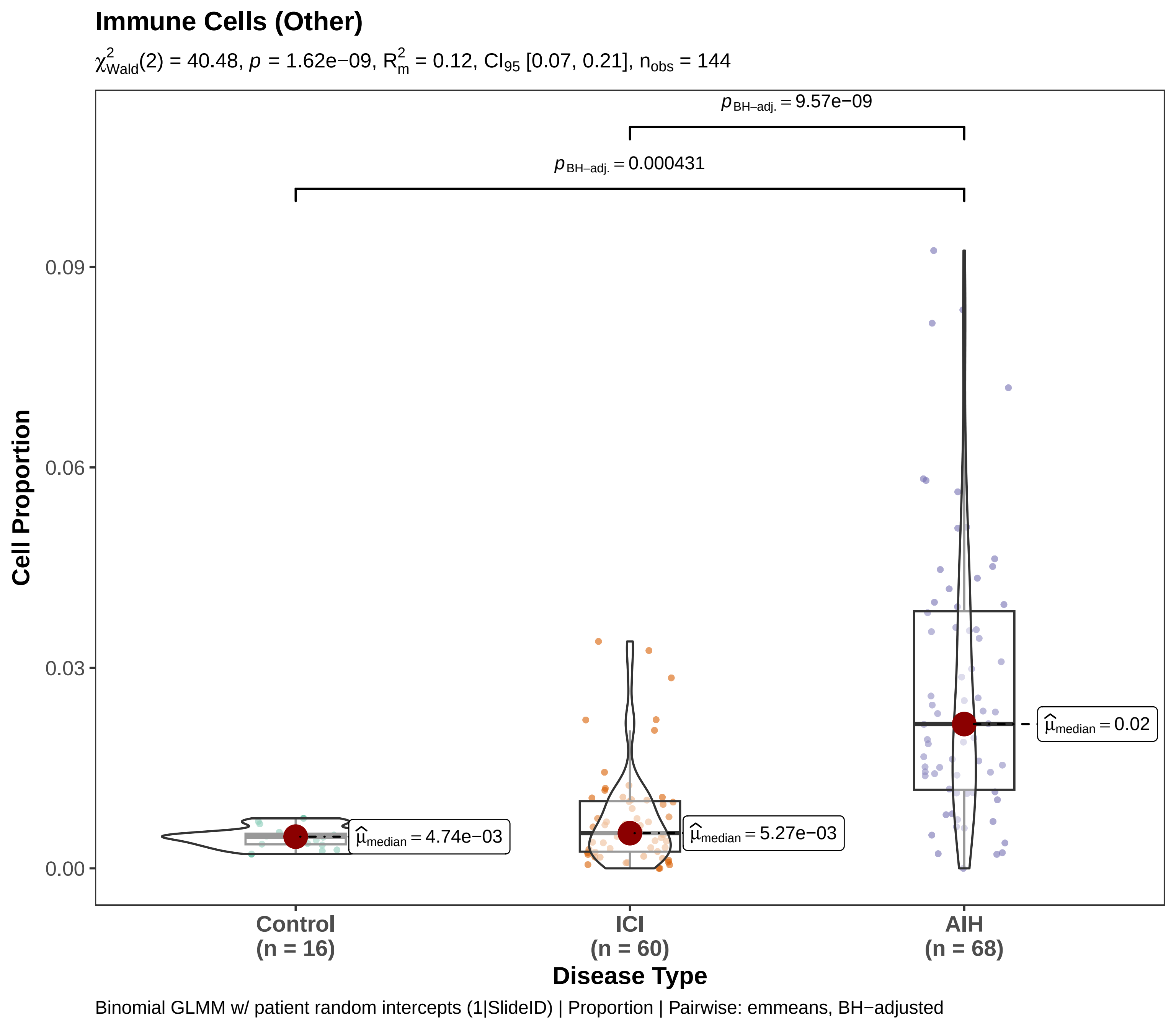

Immune Cells (Other)
$\chi^2_{Wald}(2) = 40.48$, $p = 1.62e-09$, $R^2_m = 0.12$, $CI_{95}$ [0.07, 0.21], $n_{obs} = 144$
$p_{BH-adj.} = 9.57e-09$
$p_{BH-adj.} = 0.000431$
0.09
0.06
0.03
0.00
Cell Proportion
$\widehat{\mu}_{median} = 4.74e-03$
$\widehat{\mu}_{median} = 5.27e-03$
$\widehat{\mu}_{median} = 0.02$
Control
(n = 16)
ICI
(n = 60)
AIH
(n = 68)
Disease Type
Binomial GLMM w/ patient random intercepts (1|SlideID) | Proportion | Pairwise: emmeans, BH−adjusted

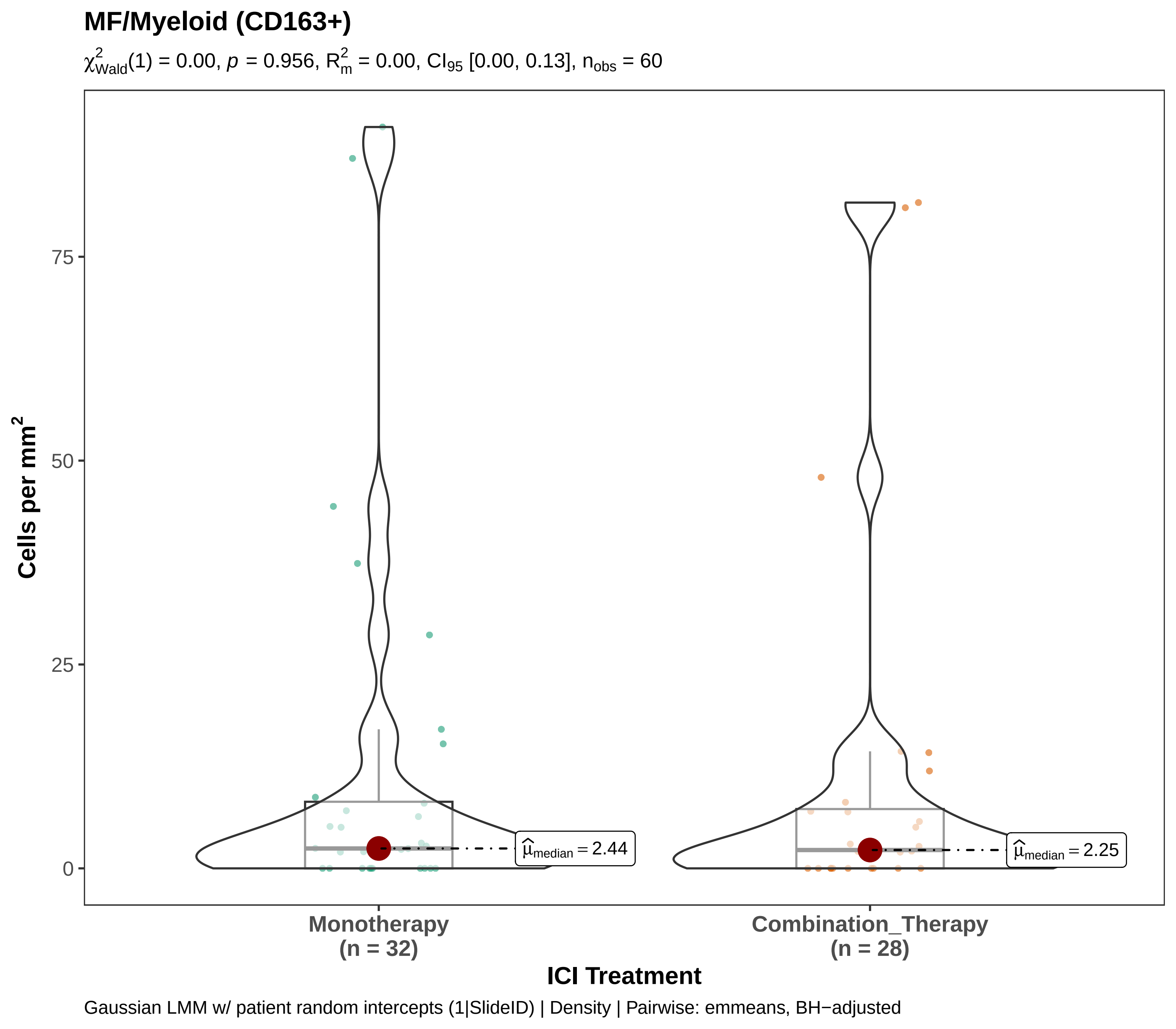

MF/Myeloid (CD163+)
$\chi^2_{Wald}(1) = 0.00$, $p = 0.956$, $R^2_m = 0.00$, $CI_{95}$ [0.00, 0.13], $n_{obs} = 60$
Cells per mm²
75
50
25
0
$\widehat{\mu}_{median} = 2.44$
$\widehat{\mu}_{median} = 2.25$
Monotherapy
(n = 32)
Combination_Therapy
(n = 28)
ICI Treatment
Gaussian LMM w/ patient random intercepts (1|SlideID) | Density | Pairwise: emmeans, BH−adjusted

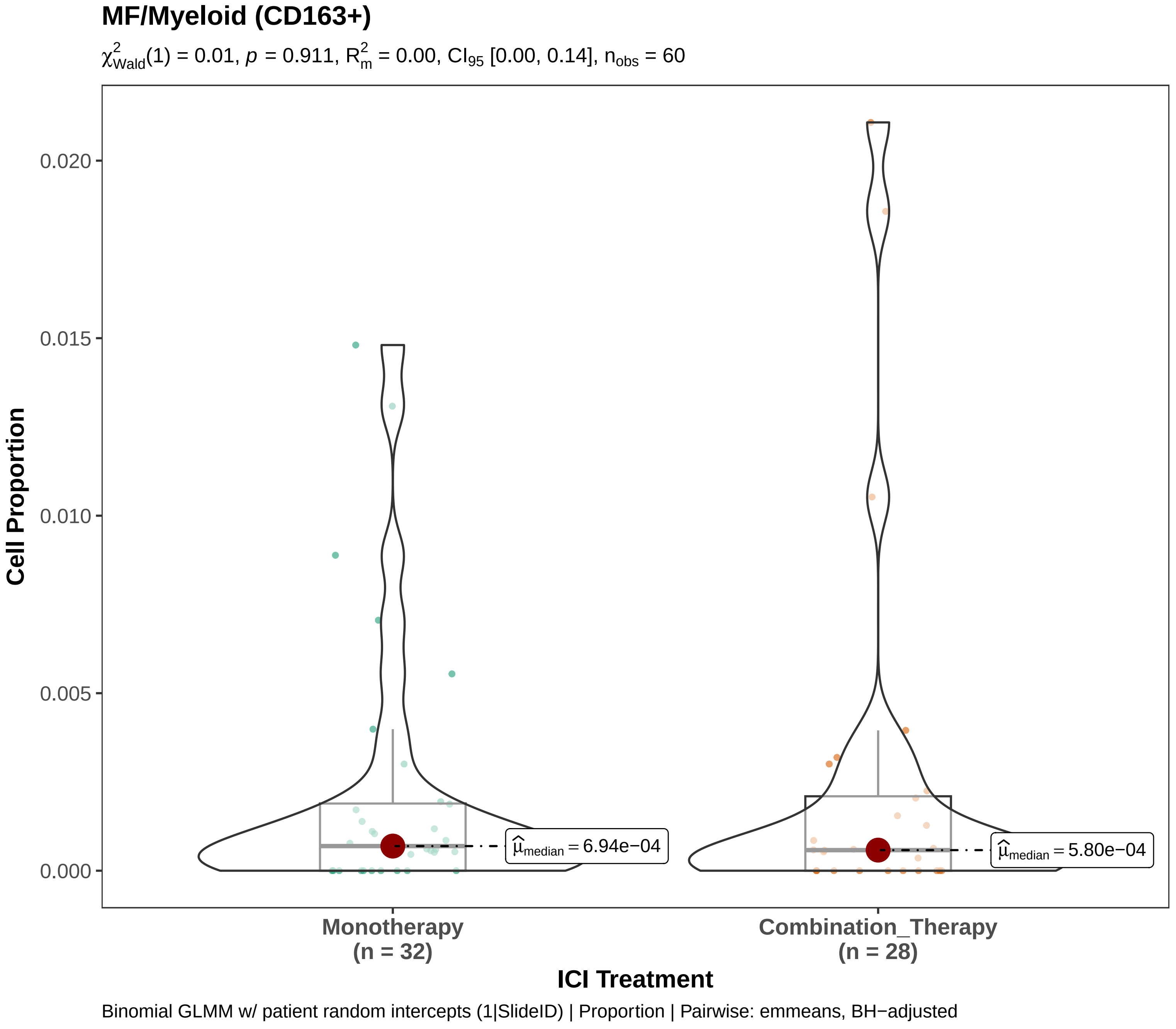
MF/Myeloid (CD163+)
$\chi^2_{Wald}(1) = 0.01$, $p = 0.911$, $R^2_m = 0.00$, $CI_{95}$ [0.00, 0.14], $n_{obs} = 60$
Cell Proportion
0.020
0.015
0.010
0.005
0.000
$\hat{\mu}_{median} = 6.94e-04$
$\hat{\mu}_{median} = 5.80e-04$
Monotherapy
(n = 32)
Combination_Therapy
(n = 28)
ICI Treatment
Binomial GLMM w/ patient random intercepts (1|SlideID) | Proportion | Pairwise: emmeans, BH−adjusted

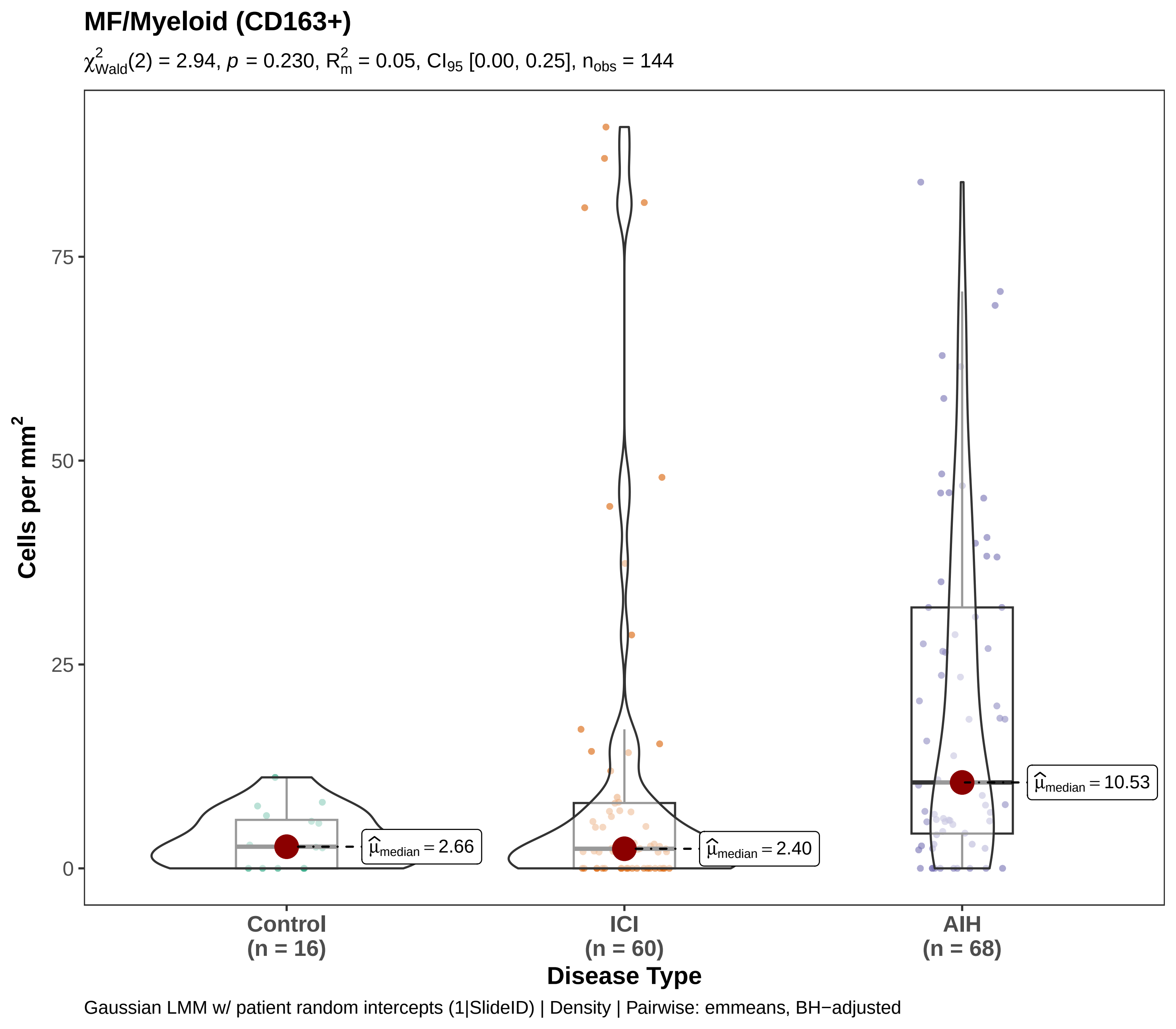
MF/Myeloid (CD163+)
$\chi^2_{Wald}(2) = 2.94$, $p = 0.230$, $R^2_m = 0.05$, $CI_{95}$ [0.00, 0.25], $n_{obs} = 144$
Cells per mm$^2$
75
50
25
0
$\widehat{\mu}_{median} = 2.66$
$\widehat{\mu}_{median} = 2.40$
$\widehat{\mu}_{median} = 10.53$
Control
(n = 16)
ICI
(n = 60)
AIH
(n = 68)
Disease Type
Gaussian LMM w/ patient random intercepts (1|SlideID) | Density | Pairwise: emmeans, BH−adjusted

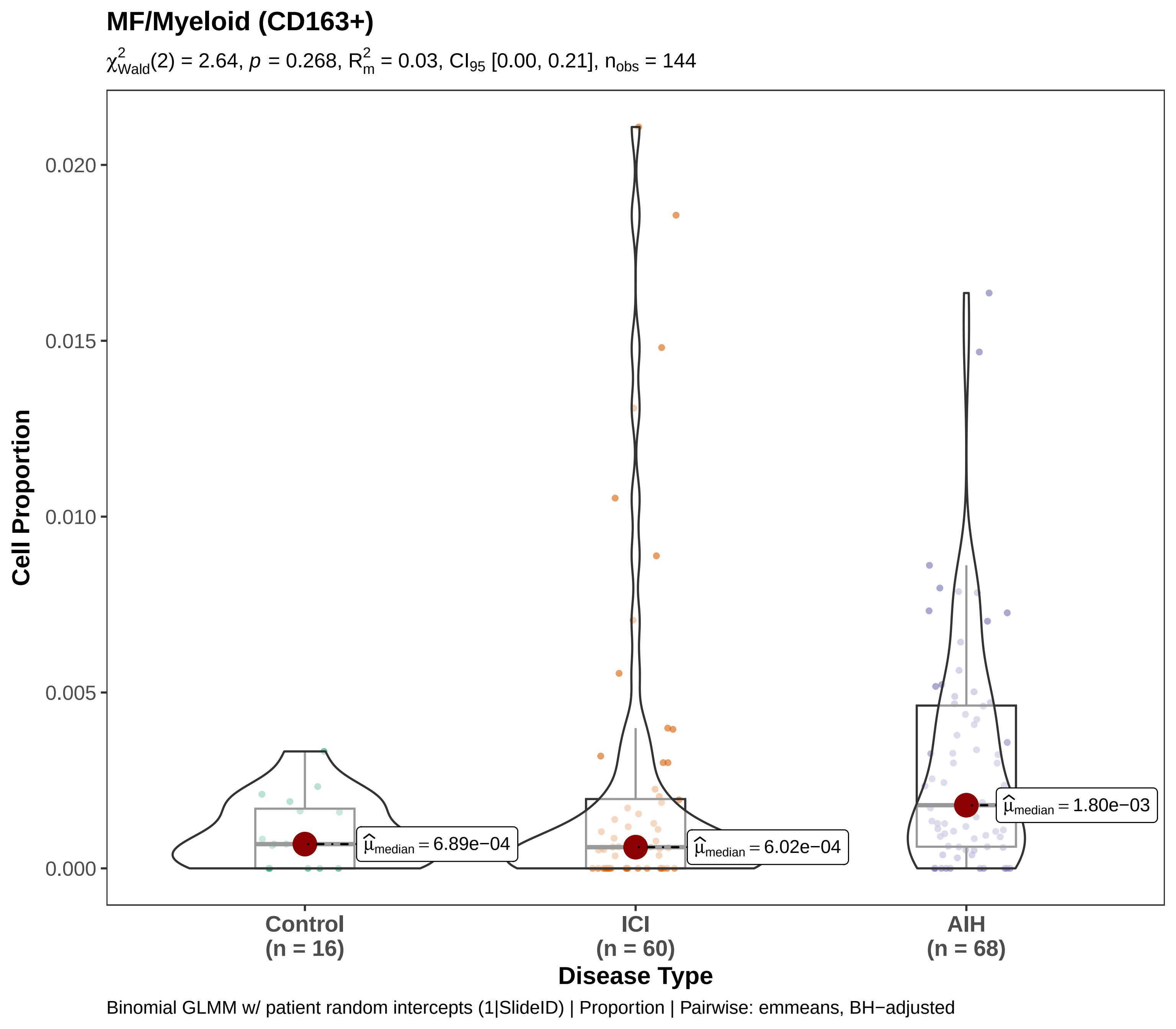
MF/Myeloid (CD163+)
$\chi^2_{Wald}(2) = 2.64$, $p = 0.268$, $R^2_m = 0.03$, $CI_{95}$ [0.00, 0.21], $n_{obs} = 144$
Cell Proportion
0.000
0.005
0.010
0.015
0.020
$\widehat{\mu}_{median} = 6.89e-04$
$\widehat{\mu}_{median} = 6.02e-04$
$\widehat{\mu}_{median} = 1.80e-03$
Control (n = 16)
ICI (n = 60)
AIH (n = 68)
Disease Type
Binomial GLMM w/ patient random intercepts (1|SlideID) | Proportion | Pairwise: emmeans, BH−adjusted

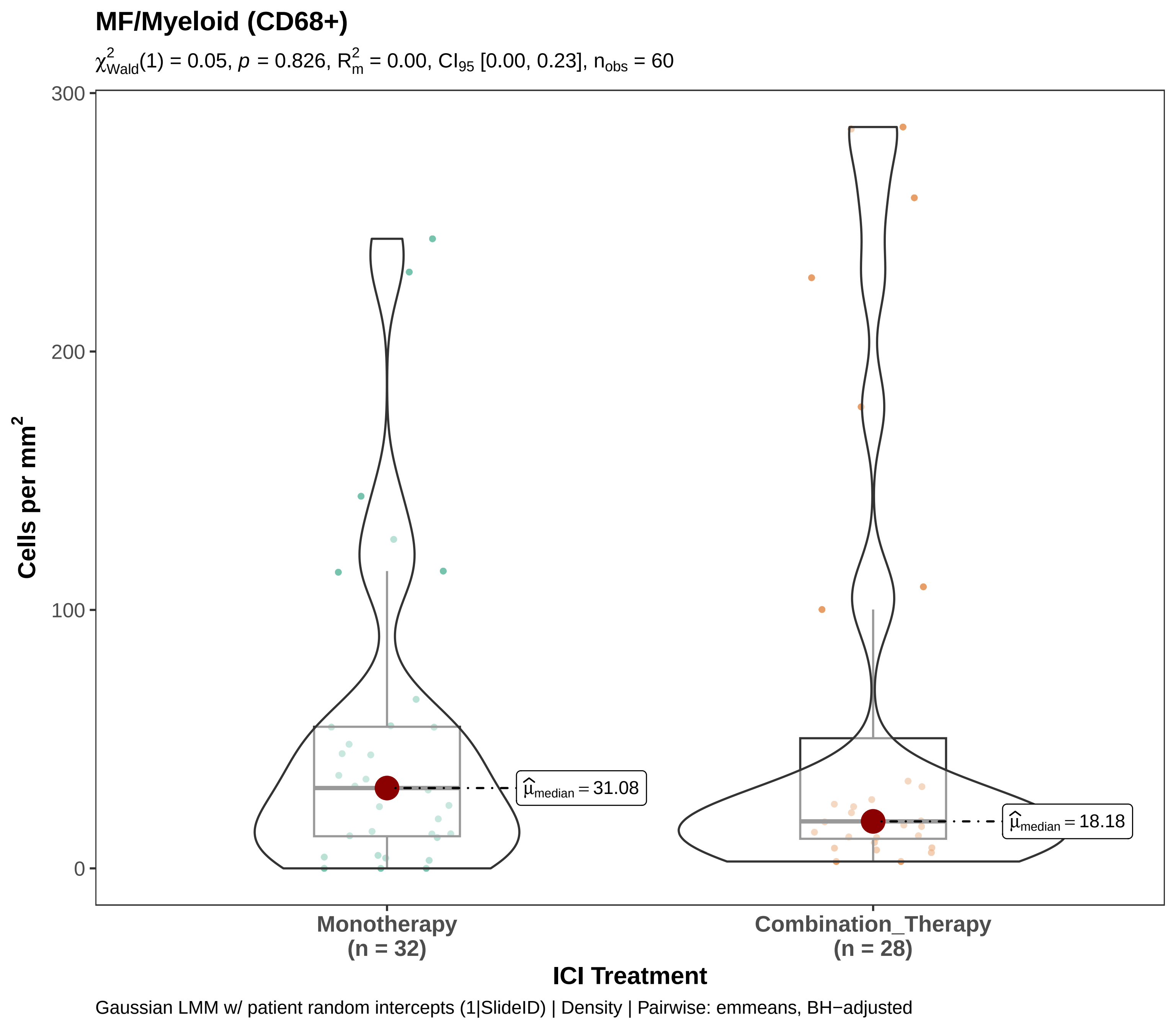

MF/Myeloid (CD68+)
$\chi^2_{Wald}(1) = 0.05$, $p = 0.826$, $R^2_m = 0.00$, $CI_{95}$ [0.00, 0.23], $n_{obs} = 60$
300
200
100
0
Cells per mm$^2$
$\hat{\mu}_{median} = 31.08$
$\hat{\mu}_{median} = 18.18$
Monotherapy
(n = 32)
Combination_Therapy
(n = 28)
ICI Treatment
Gaussian LMM w/ patient random intercepts (1|SlideID) | Density | Pairwise: emmeans, BH−adjusted

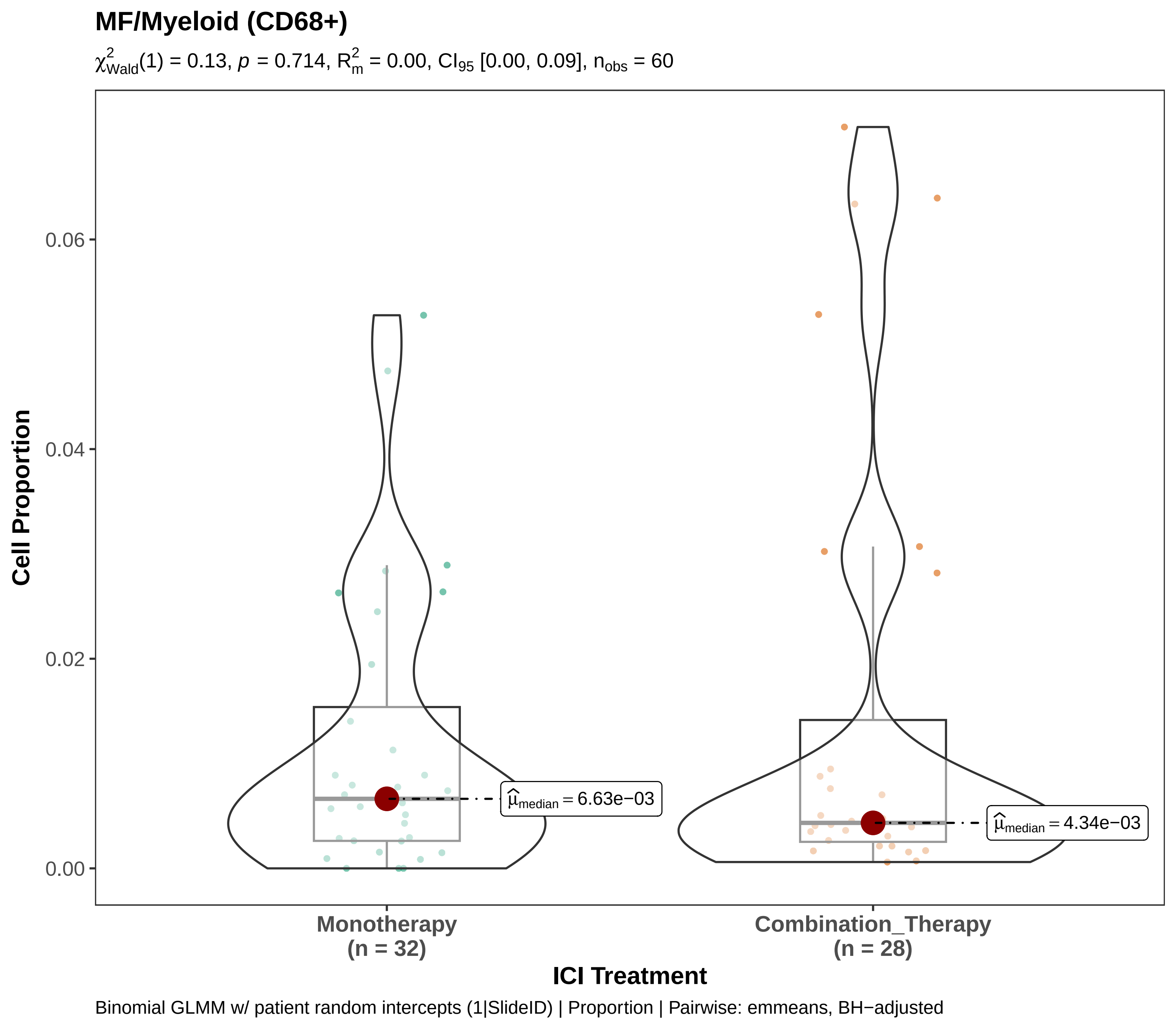
MF/Myeloid (CD68+)
$\chi^2_{Wald}(1) = 0.13$, $p = 0.714$, $R^2_m = 0.00$, $CI_{95}$ [0.00, 0.09], $n_{obs} = 60$
Cell Proportion
0.06
0.04
0.02
0.00
$\hat{\mu}_{median} = 6.63e{-}03$
$\hat{\mu}_{median} = 4.34e{-}03$
Monotherapy
(n = 32)
Combination_Therapy
(n = 28)
ICI Treatment
Binomial GLMM w/ patient random intercepts (1|SlideID) | Proportion | Pairwise: emmeans, BH−adjusted

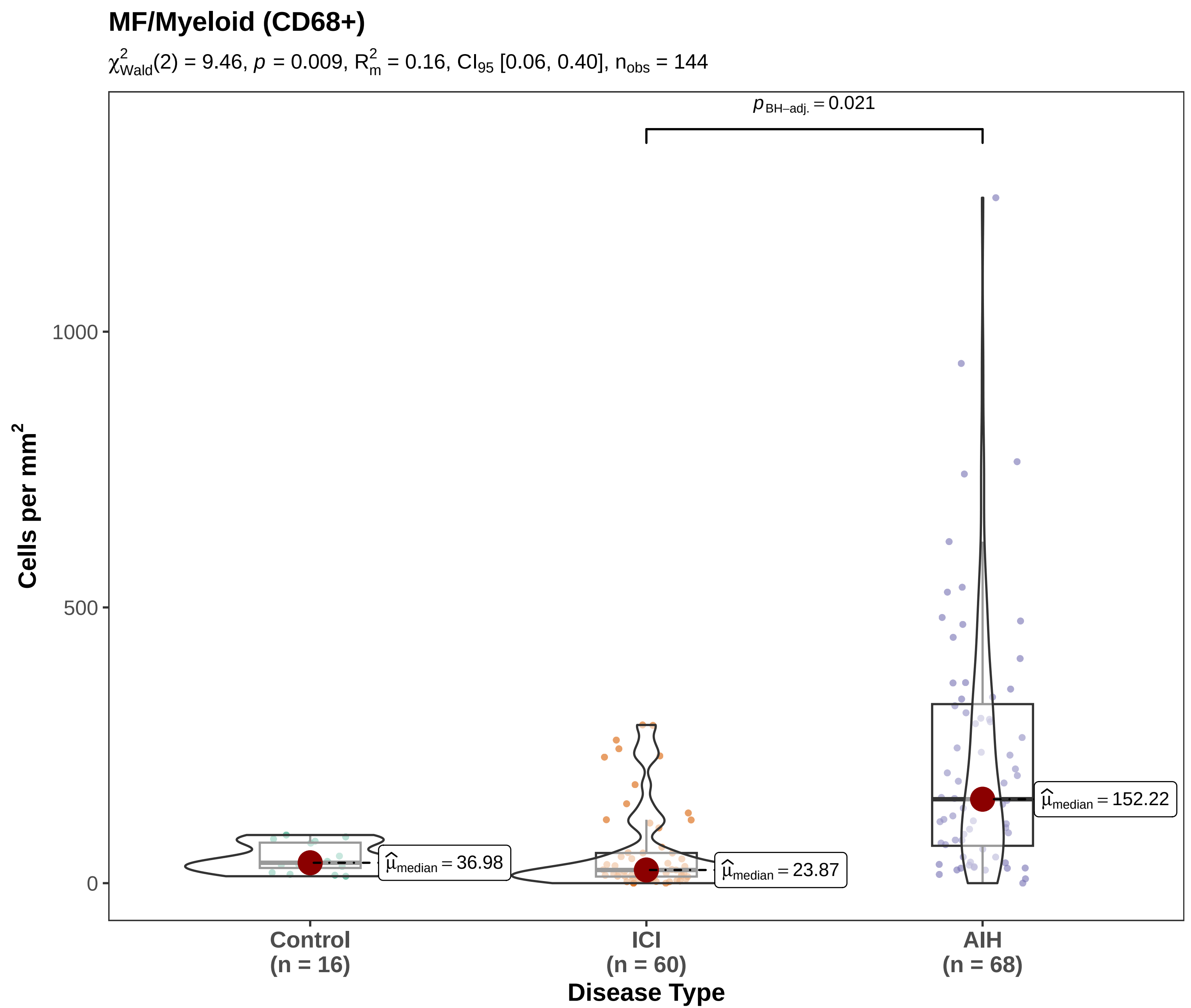

Gaussian LMM w/ patient random intercepts (1|SlideID) | Density | Pairwise: emmeans, BH−adjusted

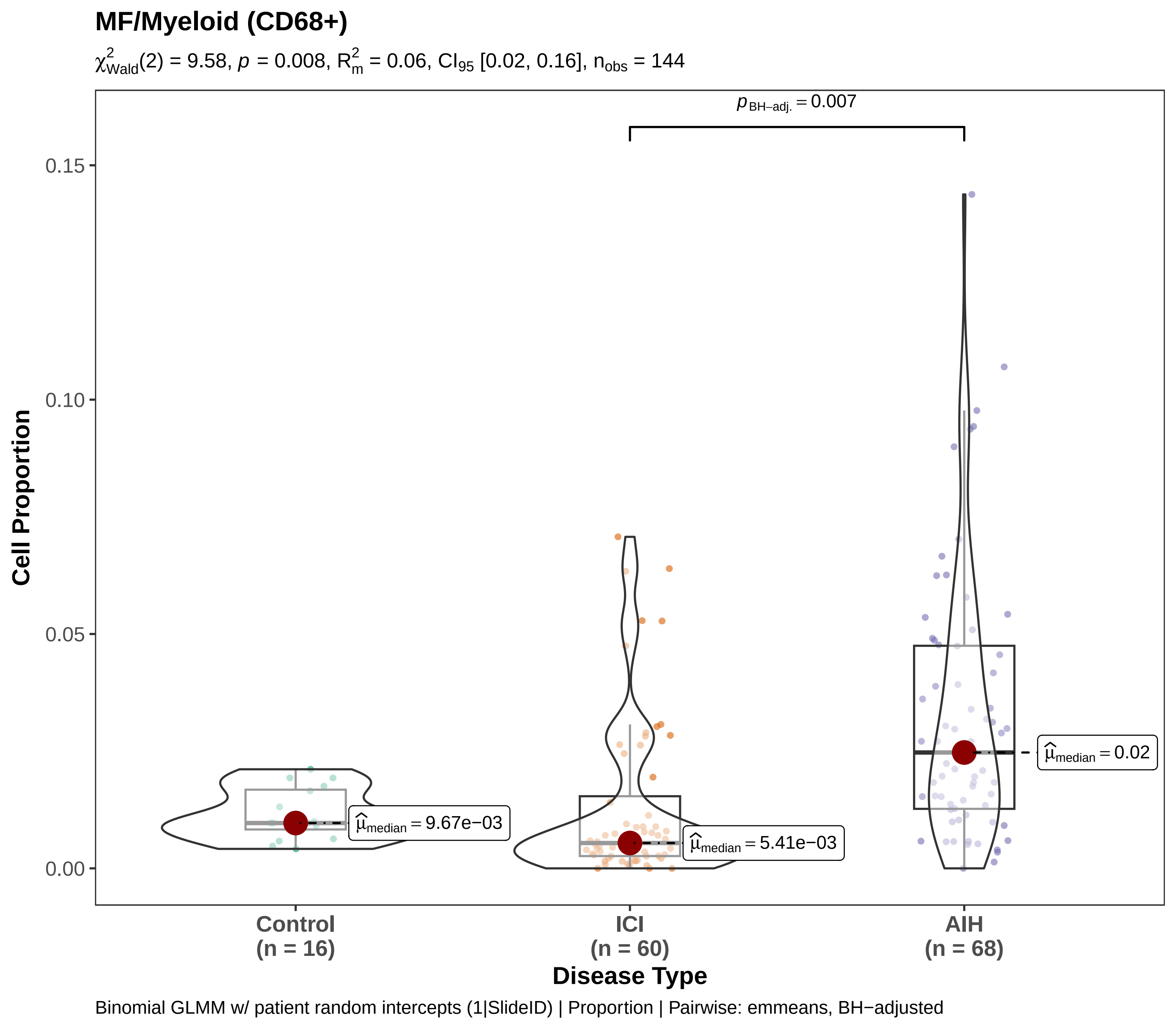

MF/Myeloid (CD68+)
$\chi^2_{Wald}(2) = 9.58$, $p = 0.008$, $R^2_m = 0.06$, $CI_{95}$ [0.02, 0.16], $n_{obs} = 144$
$p_{BH-adj.} = 0.007$
0.15
0.10
0.05
0.00
Cell Proportion
$\hat{\mu}_{median} = 9.67e-03$
$\hat{\mu}_{median} = 5.41e-03$
$\hat{\mu}_{median} = 0.02$
Control
(n = 16)
ICI
(n = 60)
AIH
(n = 68)
Disease Type
Binomial GLMM w/ patient random intercepts (1|SlideID) | Proportion | Pairwise: emmeans, BH−adjusted

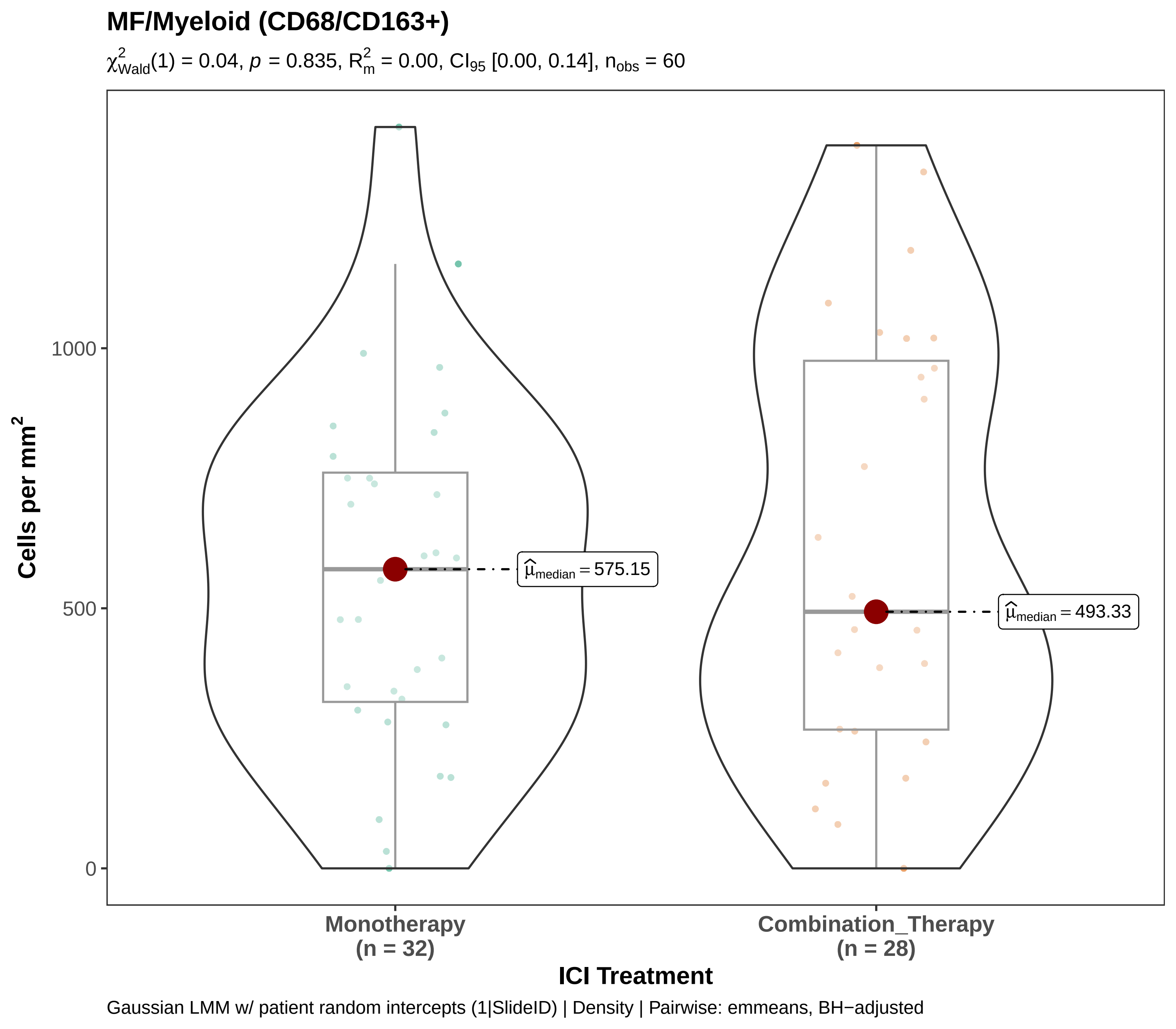

MF/Myeloid (CD68/CD163+)
χ²Wald(1) = 0.04, p = 0.835, R²m = 0.00, CI95 [0.00, 0.14], nobs = 60
Cells per mm²
1000
500
0
μ̂median = 575.15
μ̂median = 493.33
Monotherapy
(n = 32)
Combination_Therapy
(n = 28)
ICI Treatment
Gaussian LMM w/ patient random intercepts (1|SlideID) | Density | Pairwise: emmeans, BH−adjusted

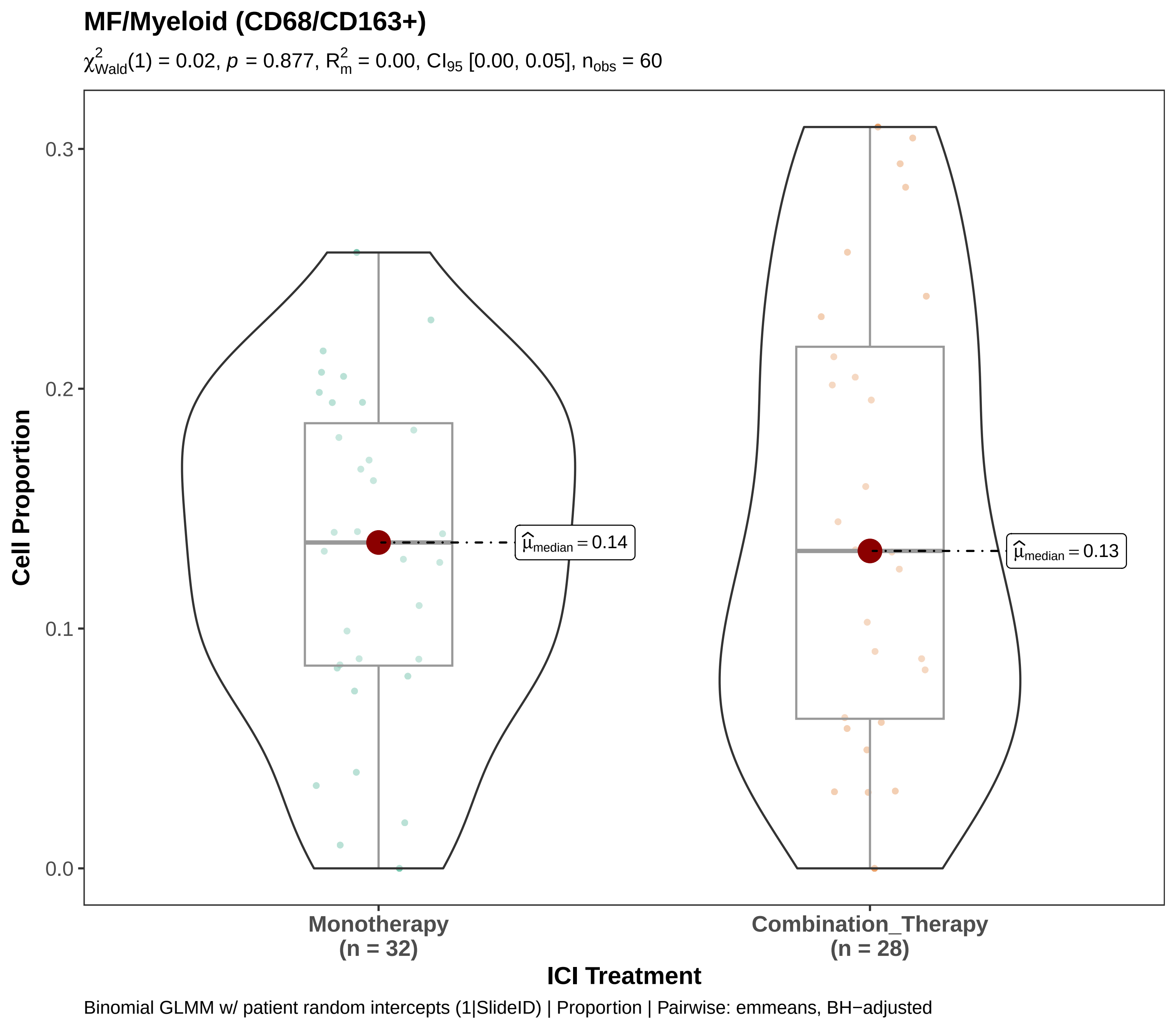
MF/Myeloid (CD68/CD163+)
$\chi^2_{Wald}(1) = 0.02$, $p = 0.877$, $R^2_m = 0.00$, $CI_{95}$ [0.00, 0.05], $n_{obs} = 60$
Cell Proportion
0.3
0.2
0.1
0.0
$\widehat{\mu}_{median} = 0.14$
$\widehat{\mu}_{median} = 0.13$
Monotherapy
(n = 32)
Combination_Therapy
(n = 28)
ICI Treatment
Binomial GLMM w/ patient random intercepts (1|SlideID) | Proportion | Pairwise: emmeans, BH−adjusted

### MF/Myeloid (CD68/CD163+)

$\chi^2_{Wald}(2) = 5.08$, $p = 0.079$, $R^2_m = 0.12$, $CI_{95}$ [0.01, 0.38], $n_{obs} = 144$

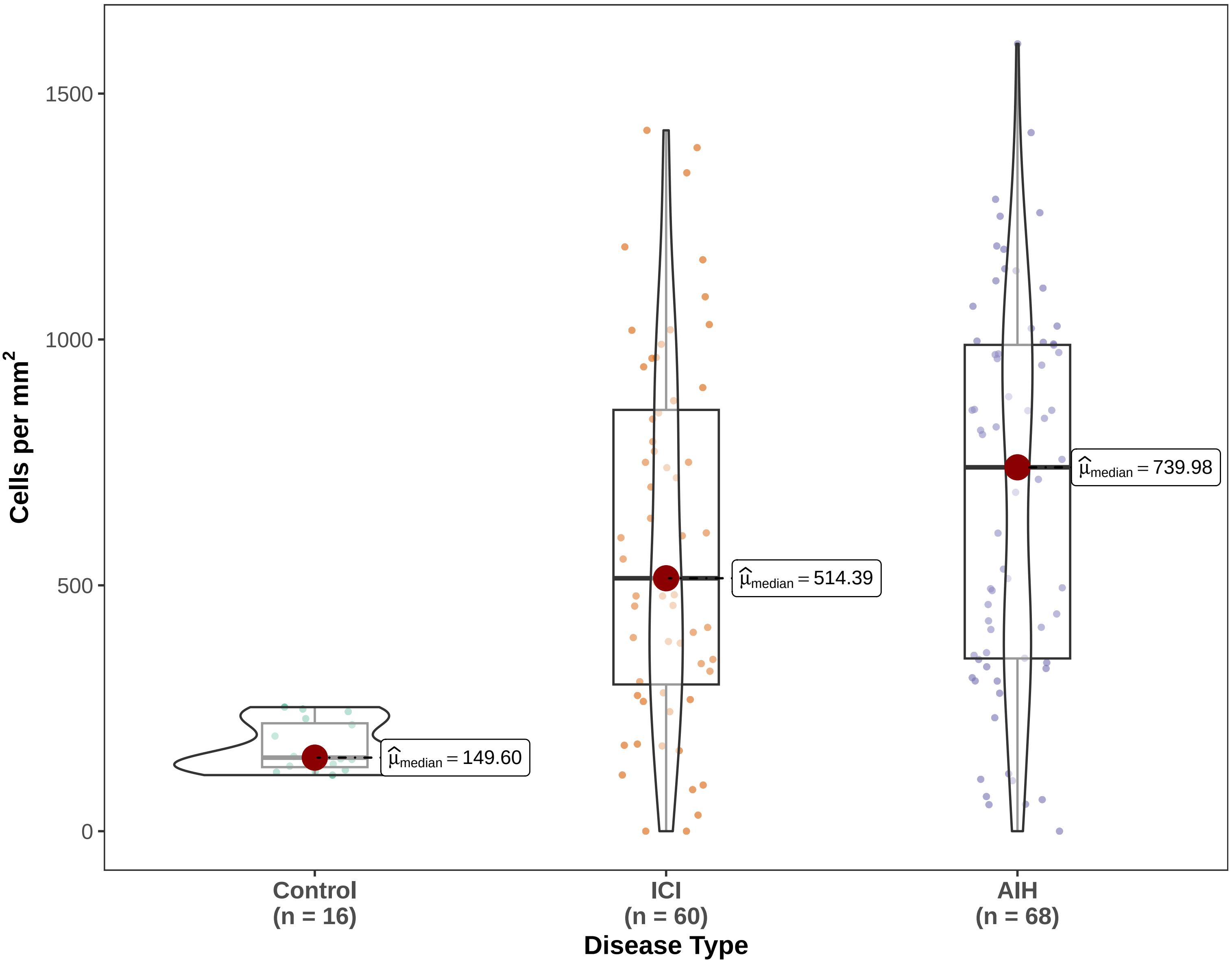


Gaussian LMM w/ patient random intercepts (1|SlideID) | Density | Pairwise: emmeans, BH−adjusted

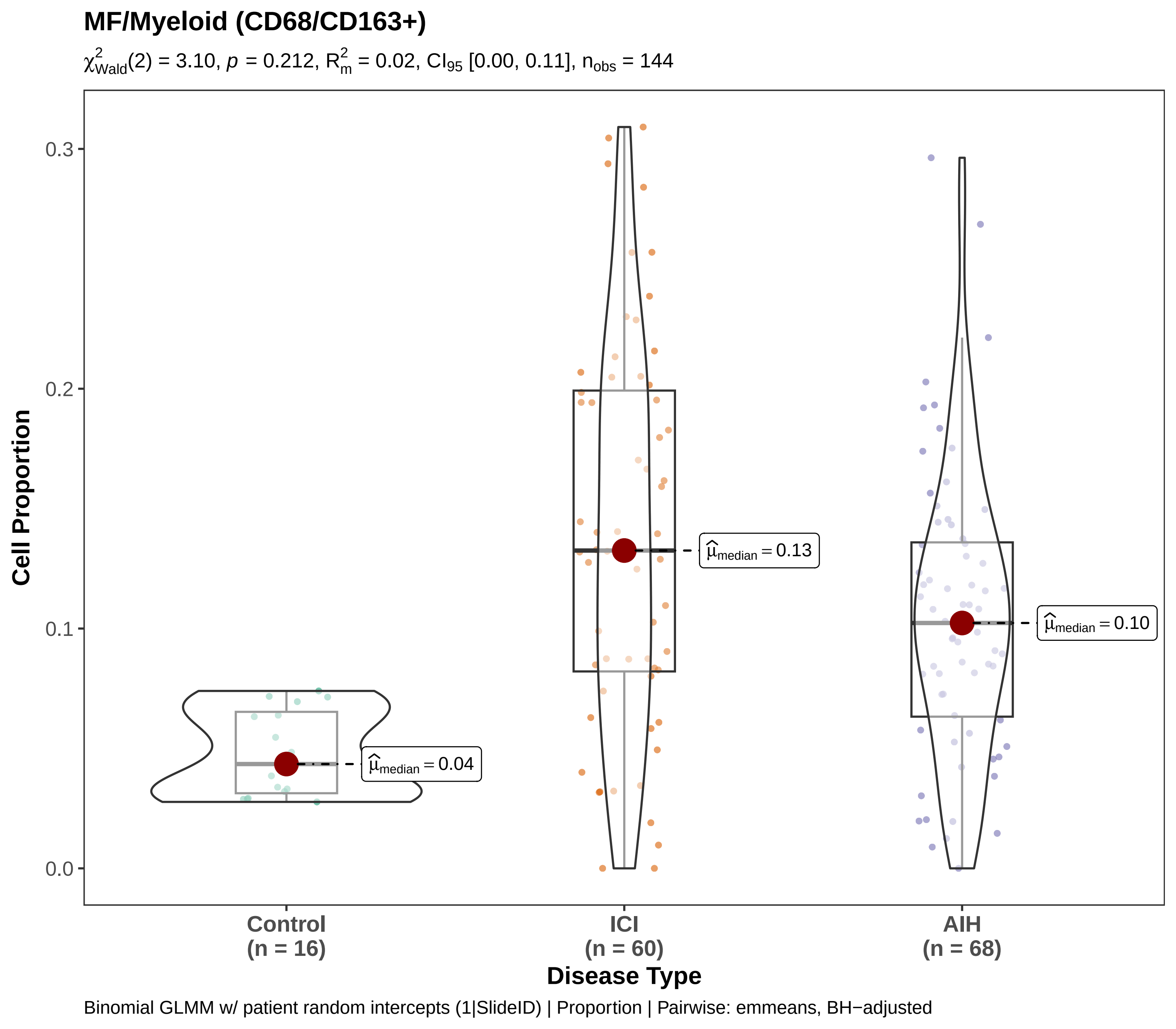

MF/Myeloid (CD68/CD163+)
$\chi^2_{Wald}(2) = 3.10$, $p = 0.212$, $R^2_m = 0.02$, $CI_{95}$ [0.00, 0.11], $n_{obs} = 144$
Cell Proportion
0.0
0.1
0.2
0.3
$\hat{\mu}_{median} = 0.04$
$\hat{\mu}_{median} = 0.13$
$\hat{\mu}_{median} = 0.10$
Control (n = 16)
ICI (n = 60)
AIH (n = 68)
Disease Type
Binomial GLMM w/ patient random intercepts (1|SlideID) | Proportion | Pairwise: emmeans, BH−adjusted

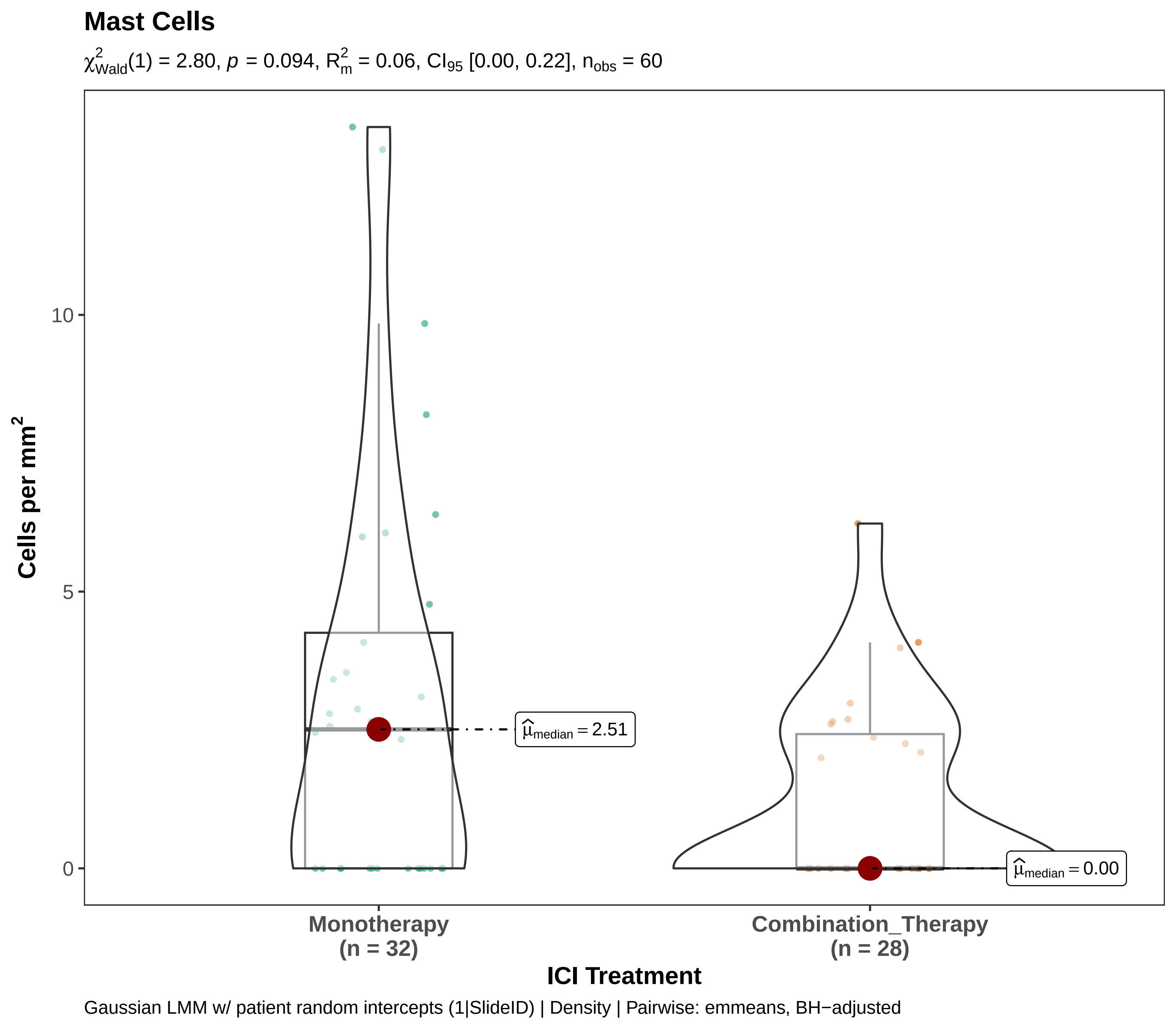

Mast Cells
$\chi^2_{Wald}(1) = 2.80$, $p = 0.094$, $R^2_m = 0.06$, $CI_{95}$ [0.00, 0.22], $n_{obs} = 60$
Cells per mm²
10
5
0
$\widehat{\mu}_{median} = 2.51$
$\widehat{\mu}_{median} = 0.00$
Monotherapy
(n = 32)
Combination_Therapy
(n = 28)
ICI Treatment
Gaussian LMM w/ patient random intercepts (1|SlideID) | Density | Pairwise: emmeans, BH−adjusted

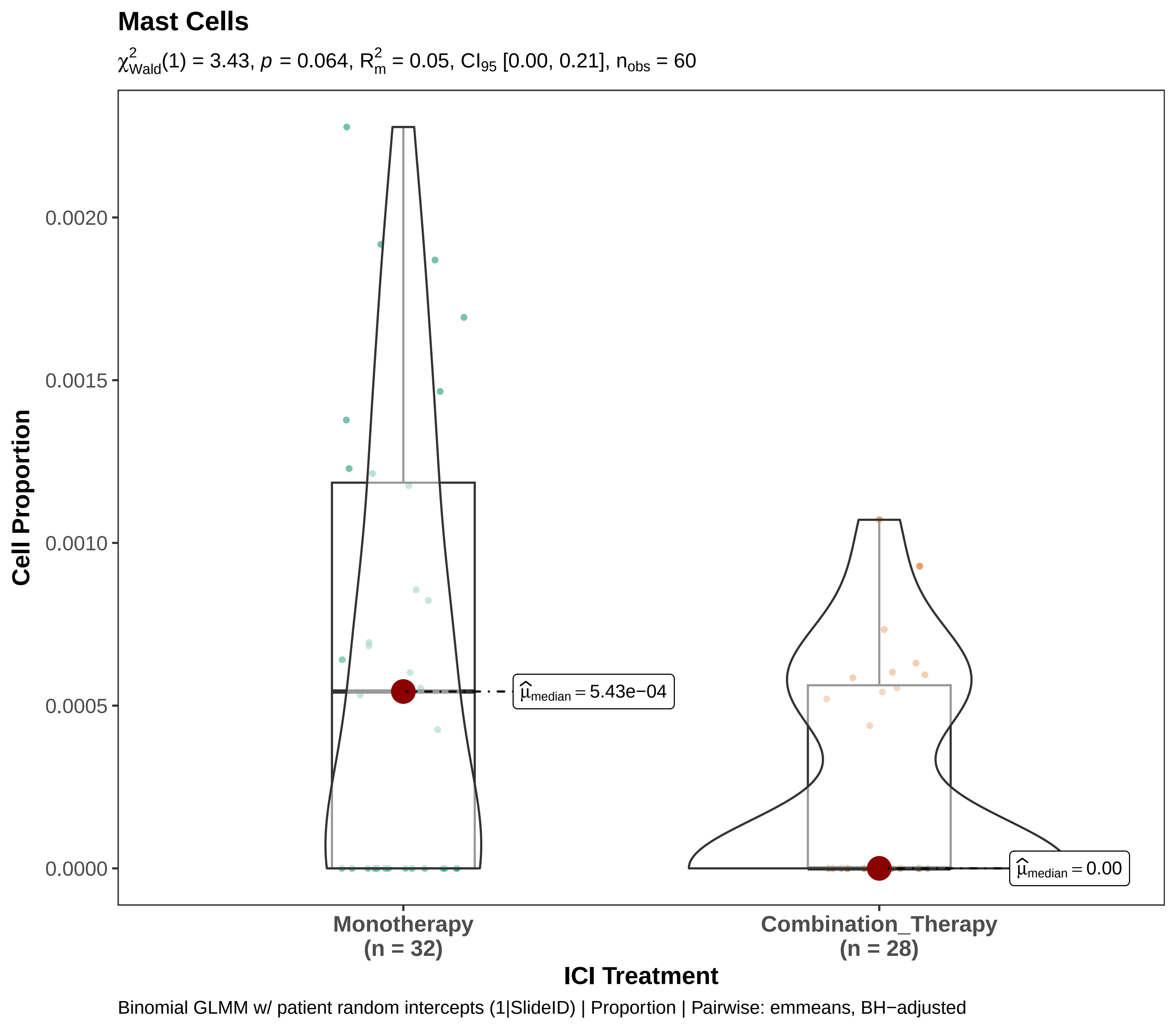

Mast Cells
$\chi^2_{Wald}(1) = 3.43$, $p = 0.064$, $R^2_m = 0.05$, $CI_{95}$ [0.00, 0.21], $n_{obs} = 60$
Cell Proportion
0.0020
0.0015
0.0010
0.0005
0.0000
$\hat{\mu}_{median} = 5.43e{-}04$
$\hat{\mu}_{median} = 0.00$
Monotherapy
(n = 32)
Combination_Therapy
(n = 28)
ICI Treatment
Binomial GLMM w/ patient random intercepts (1|SlideID) | Proportion | Pairwise: emmeans, BH−adjusted

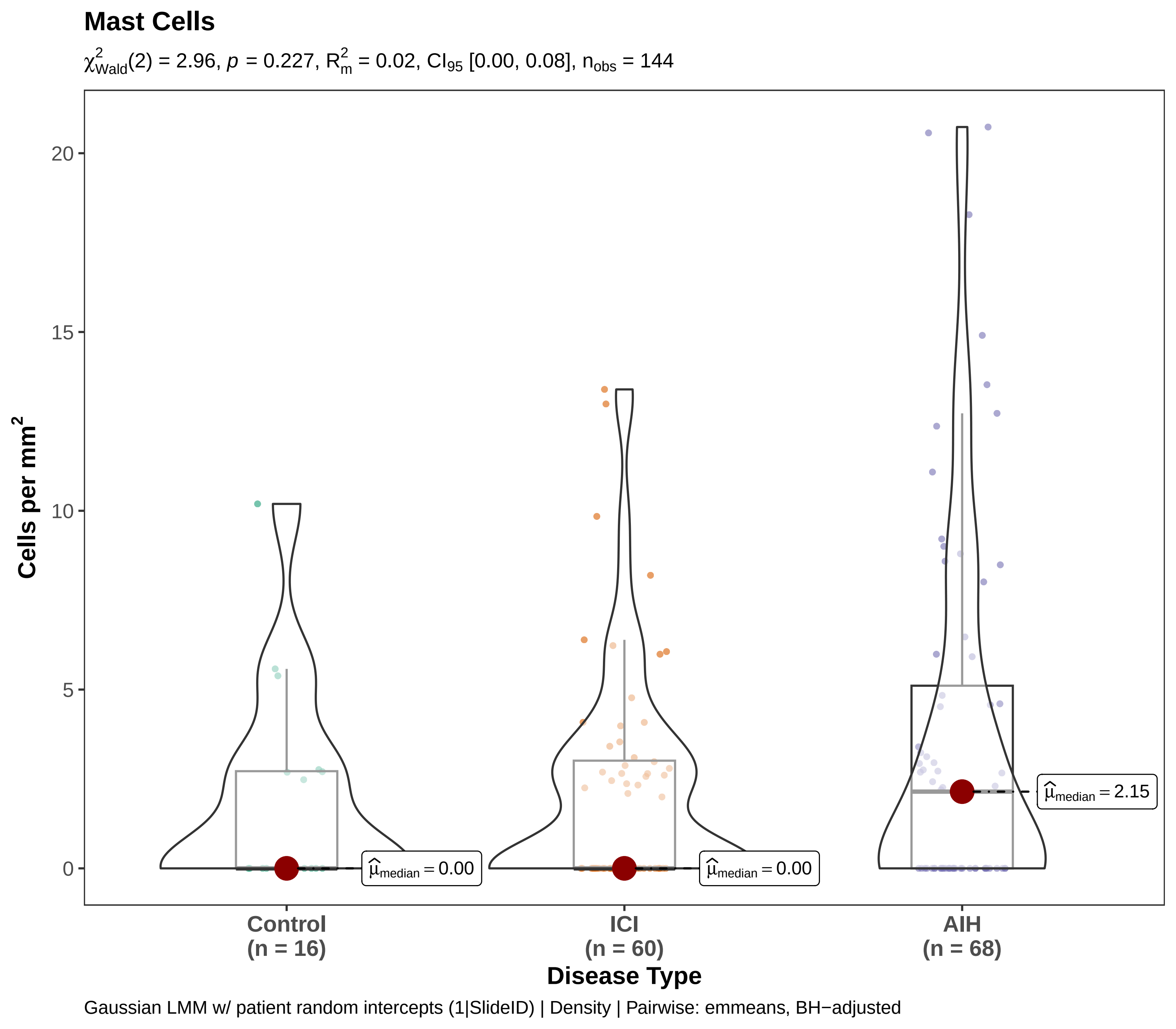

Mast Cells
χ²Wald(2) = 2.96, p = 0.227, R²m = 0.02, CI95 [0.00, 0.08], nobs = 144
Cells per mm²
Disease Type
Control
(n = 16)
ICI
(n = 60)
AIH
(n = 68)
μ̂median = 0.00
μ̂median = 0.00
μ̂median = 2.15
0
5
10
15
20
Gaussian LMM w/ patient random intercepts (1|SlideID) | Density | Pairwise: emmeans, BH−adjusted

#### Mast Cells

$\chi^2_{Wald}(2)$ = 12522.33, $p$ = 0.00e+00, $R^2_m$ = 0.00, $n_{obs}$ = 144 [conv. warning]

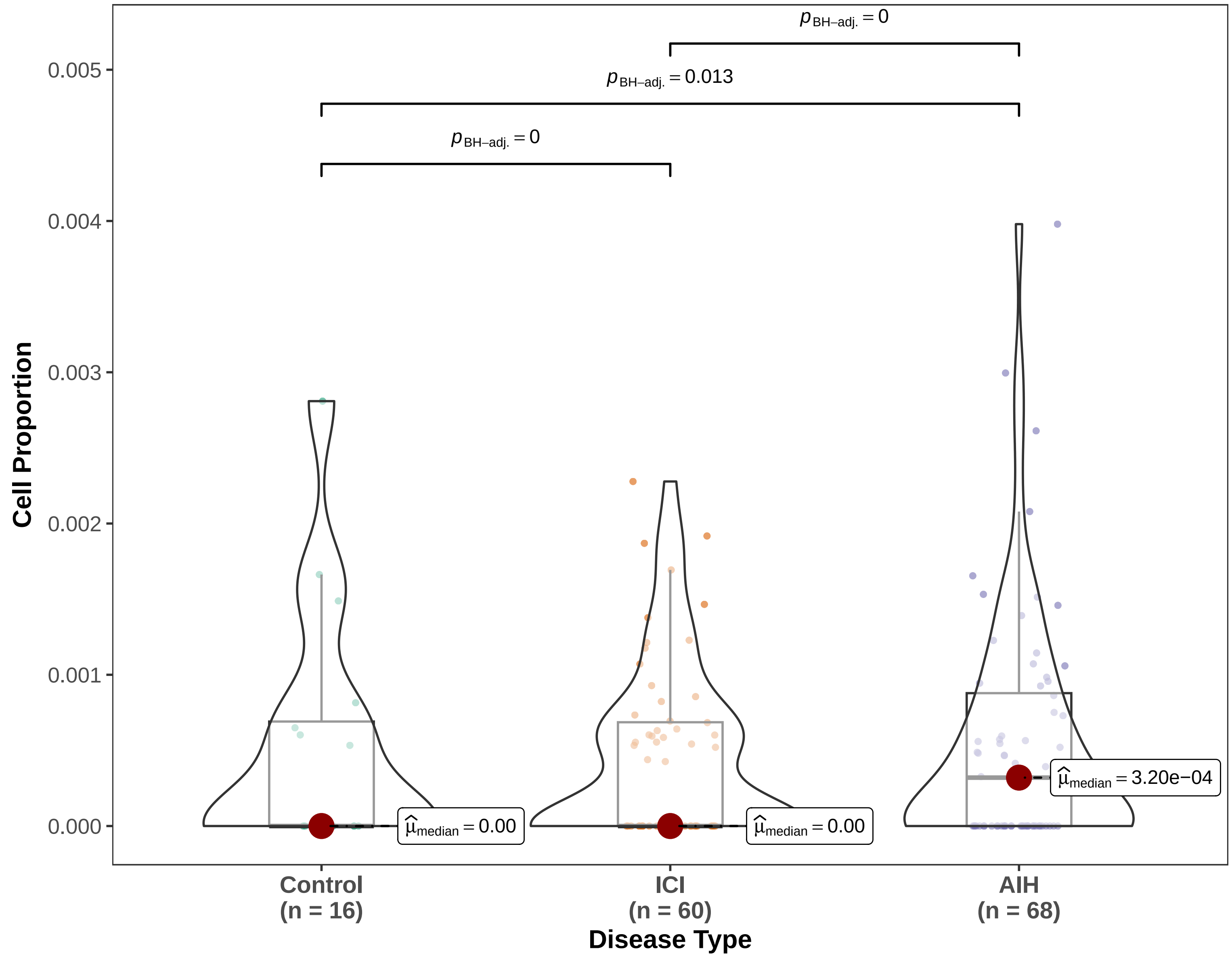


Binomial GLMM w/ patient random intercepts (1|SlideID) | Proportion | Pairwise: emmeans, BH−adjusted

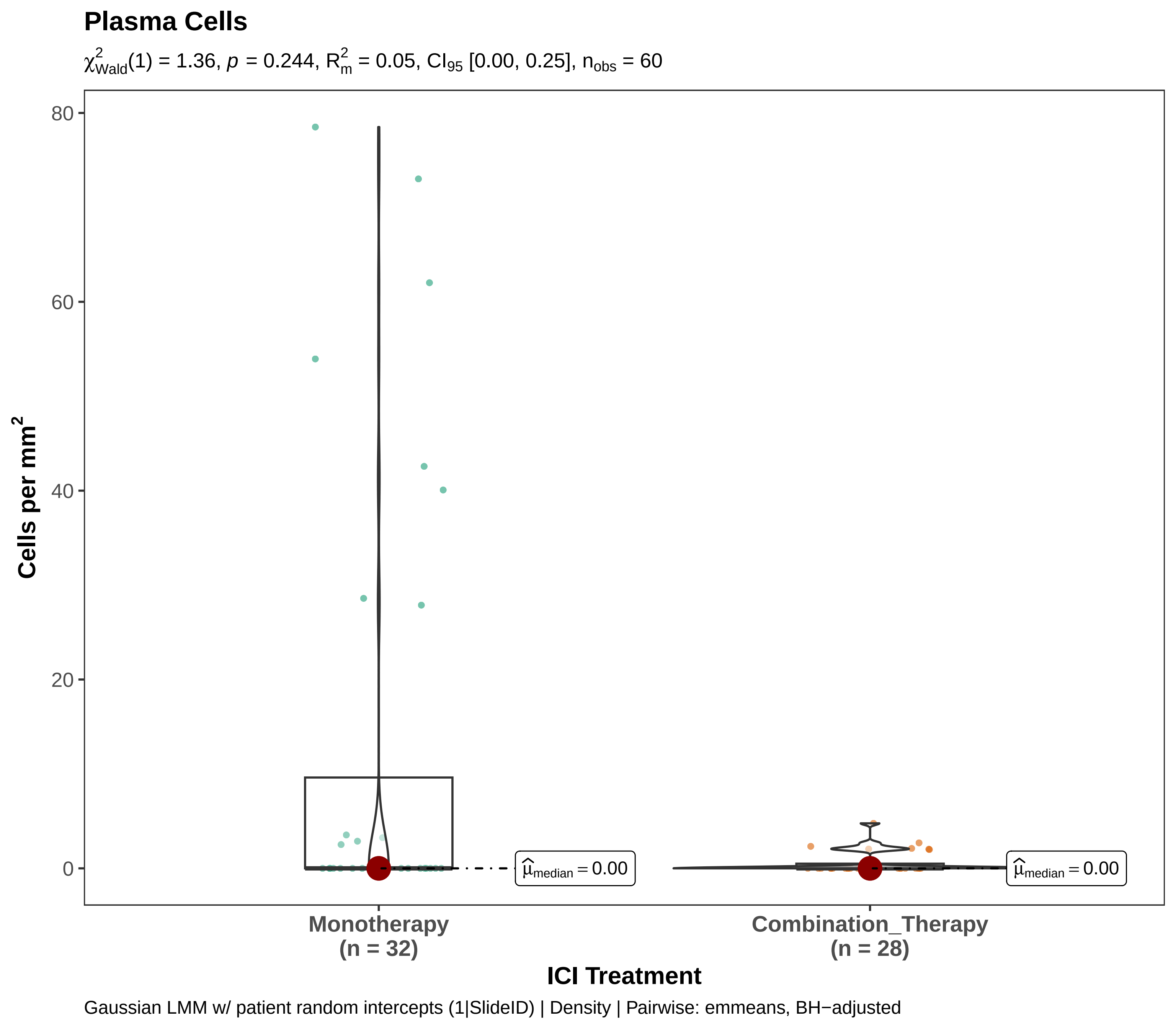
Plasma Cells
$\chi^2_{Wald}(1) = 1.36$, $p = 0.244$, $R^2_m = 0.05$, $CI_{95}$ [0.00, 0.25], $n_{obs} = 60$
80
60
40
20
0
Cells per mm²
$\hat{\mu}_{median} = 0.00$
$\hat{\mu}_{median} = 0.00$
Monotherapy
(n = 32)
Combination_Therapy
(n = 28)
ICI Treatment
Gaussian LMM w/ patient random intercepts (1|SlideID) | Density | Pairwise: emmeans, BH−adjusted

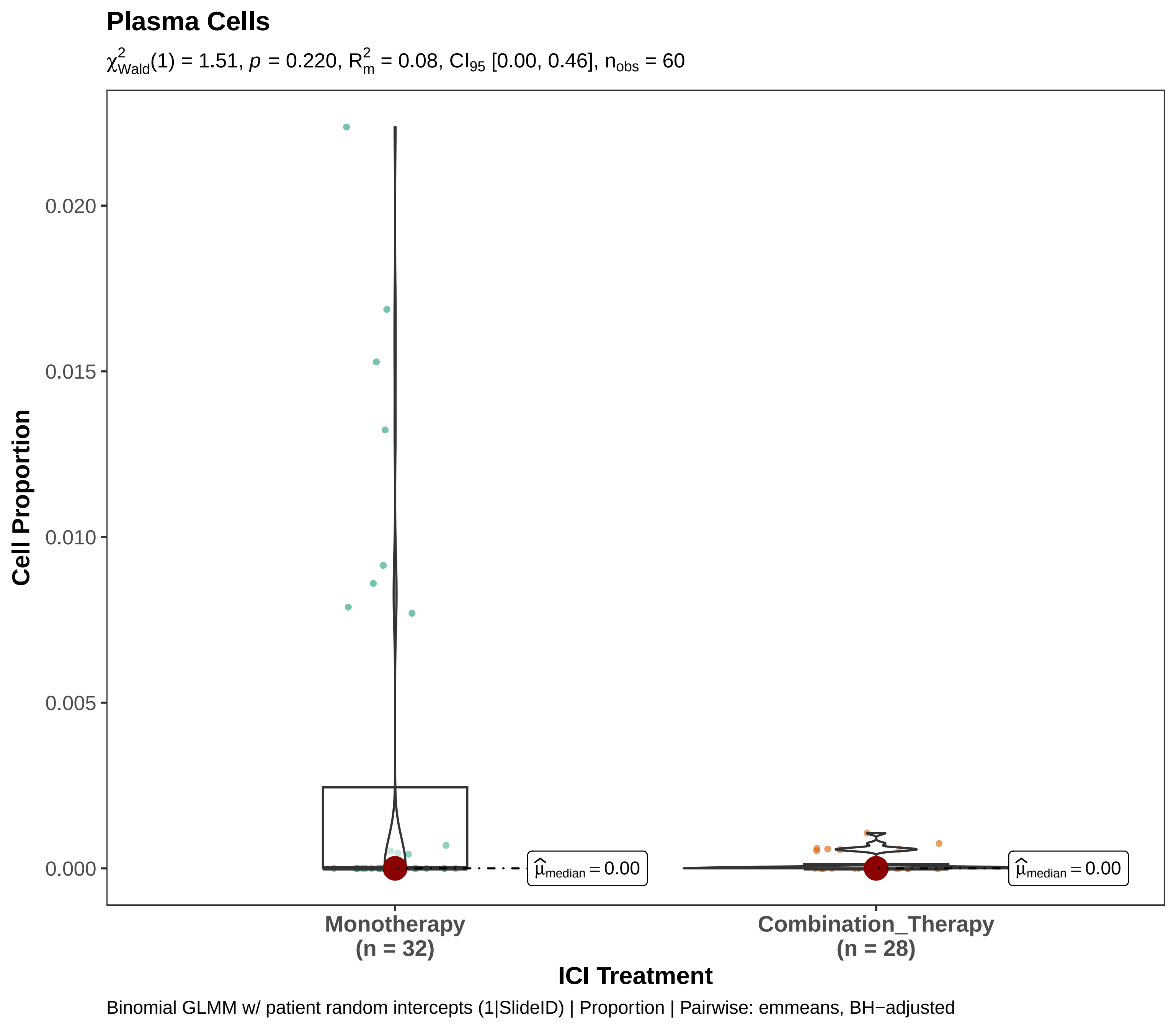
Plasma Cells
$\chi^2_{Wald}(1) = 1.51$, $p = 0.220$, $R^2_m = 0.08$, $CI_{95}$ [0.00, 0.46], $n_{obs} = 60$
Cell Proportion
0.020
0.015
0.010
0.005
0.000
$\hat{\mu}_{median} = 0.00$
$\hat{\mu}_{median} = 0.00$
Monotherapy
(n = 32)
Combination_Therapy
(n = 28)
ICI Treatment
Binomial GLMM w/ patient random intercepts (1|SlideID) | Proportion | Pairwise: emmeans, BH−adjusted

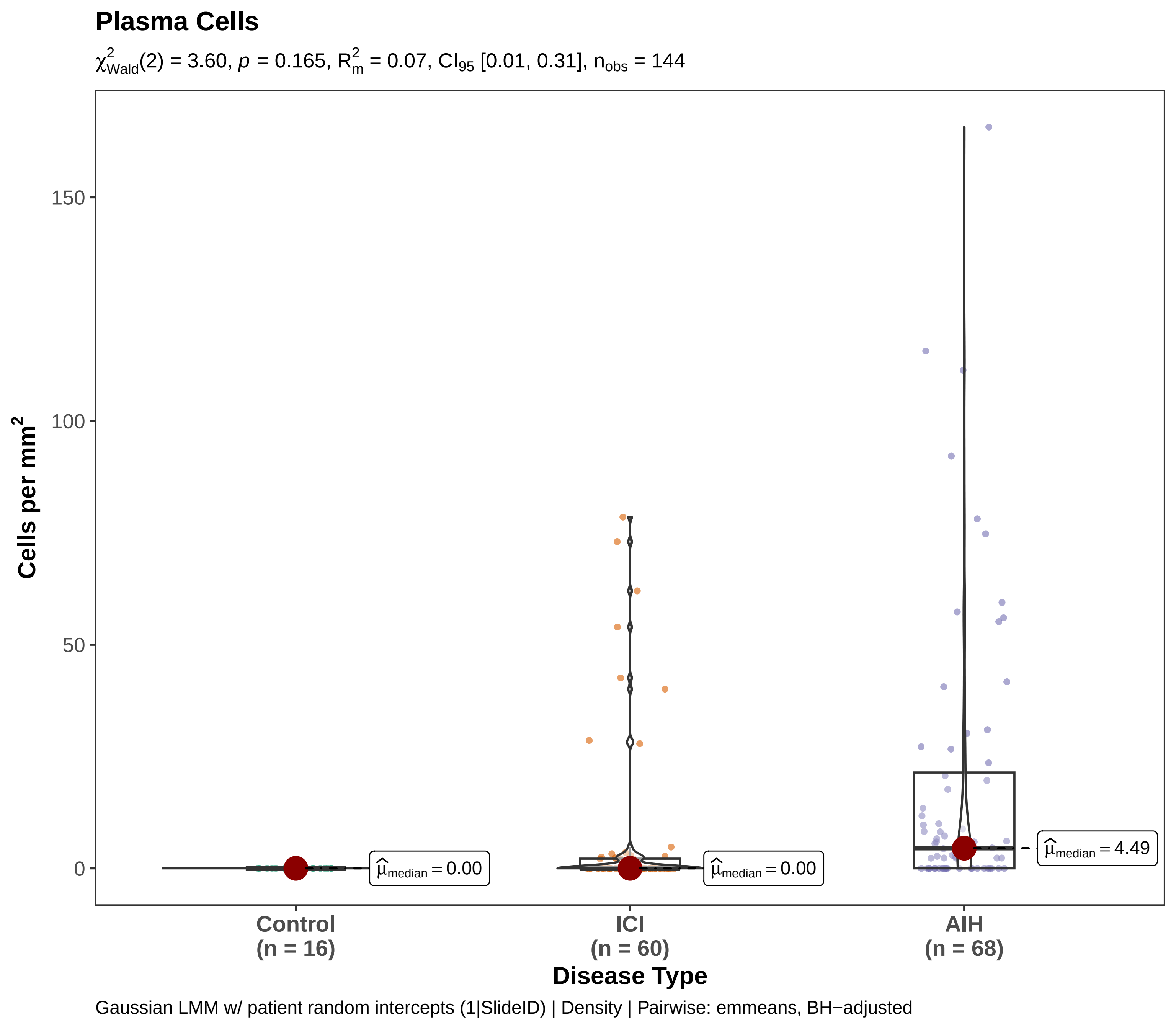
Plasma Cells
$\chi^2_{Wald}(2) = 3.60$, $p = 0.165$, $R^2_m = 0.07$, $CI_{95}$ [0.01, 0.31], $n_{obs} = 144$
Cells per mm$^2$
150
100
50
0
$\hat{\mu}_{median} = 0.00$
$\hat{\mu}_{median} = 0.00$
$\hat{\mu}_{median} = 4.49$
Control
(n = 16)
ICI
(n = 60)
AIH
(n = 68)
Disease Type
Gaussian LMM w/ patient random intercepts (1|SlideID) | Density | Pairwise: emmeans, BH−adjusted

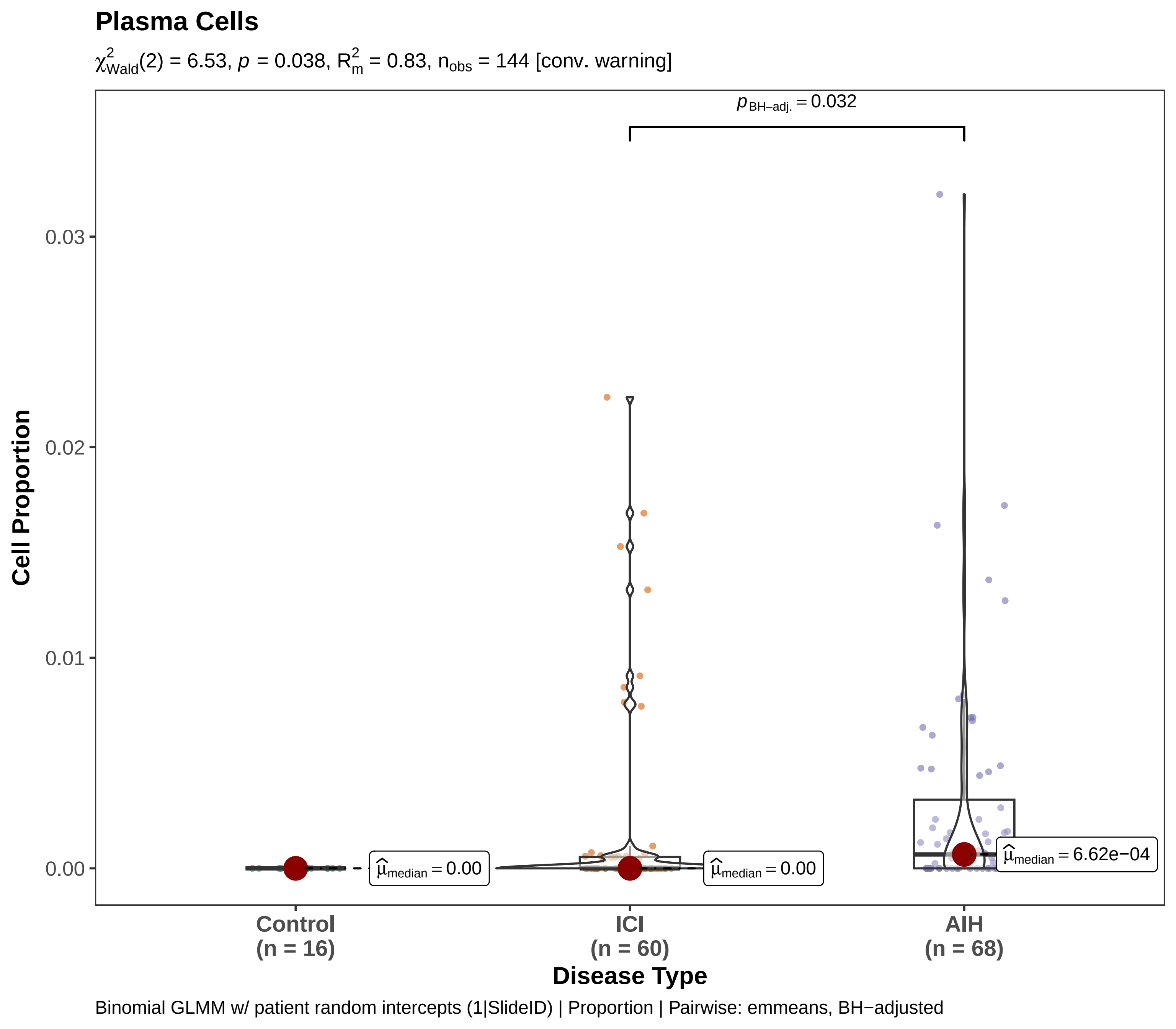

Plasma Cells
$\chi^2_{Wald}(2) = 6.53$, $p = 0.038$, $R^2_m = 0.83$, $n_{obs} = 144$ [conv. warning]
$p_{BH-adj.} = 0.032$
0.03
0.02
0.01
0.00
Cell Proportion
$\hat{\mu}_{median} = 0.00$
$\hat{\mu}_{median} = 0.00$
$\hat{\mu}_{median} = 6.62e-04$
Control (n = 16)
ICI (n = 60)
AIH (n = 68)
Disease Type
Binomial GLMM w/ patient random intercepts (1|SlideID) | Proportion | Pairwise: emmeans, BH−adjusted

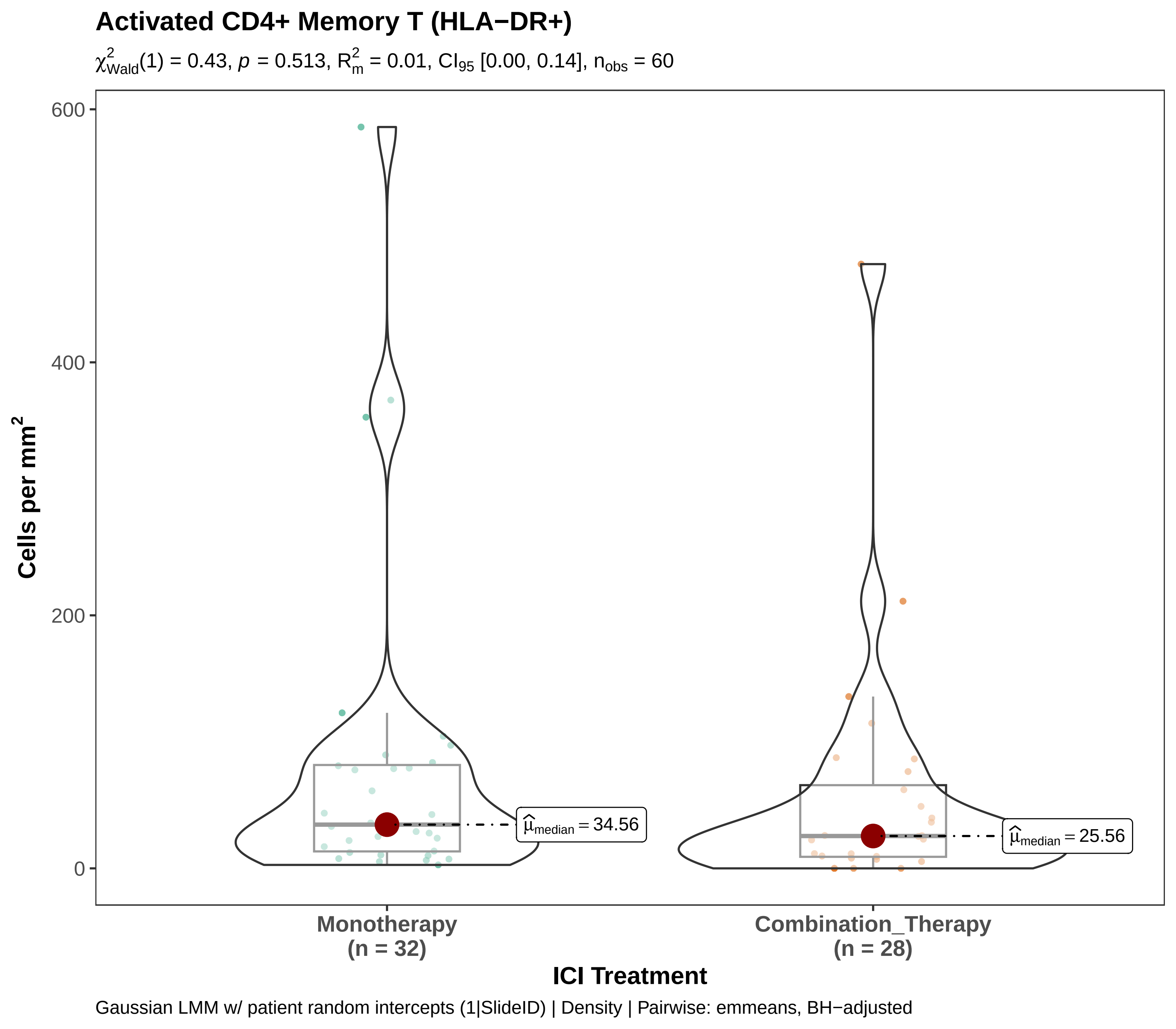

Activated CD4+ Memory T (HLA−DR+)
$\chi^2_{Wald}(1) = 0.43$, $p = 0.513$, $R^2_m = 0.01$, $CI_{95}$ [0.00, 0.14], $n_{obs} = 60$
600
400
200
0
Cells per mm²
$\widehat{\mu}_{median} = 34.56$
$\widehat{\mu}_{median} = 25.56$
Monotherapy
(n = 32)
Combination_Therapy
(n = 28)
ICI Treatment
Gaussian LMM w/ patient random intercepts (1|SlideID) | Density | Pairwise: emmeans, BH−adjusted

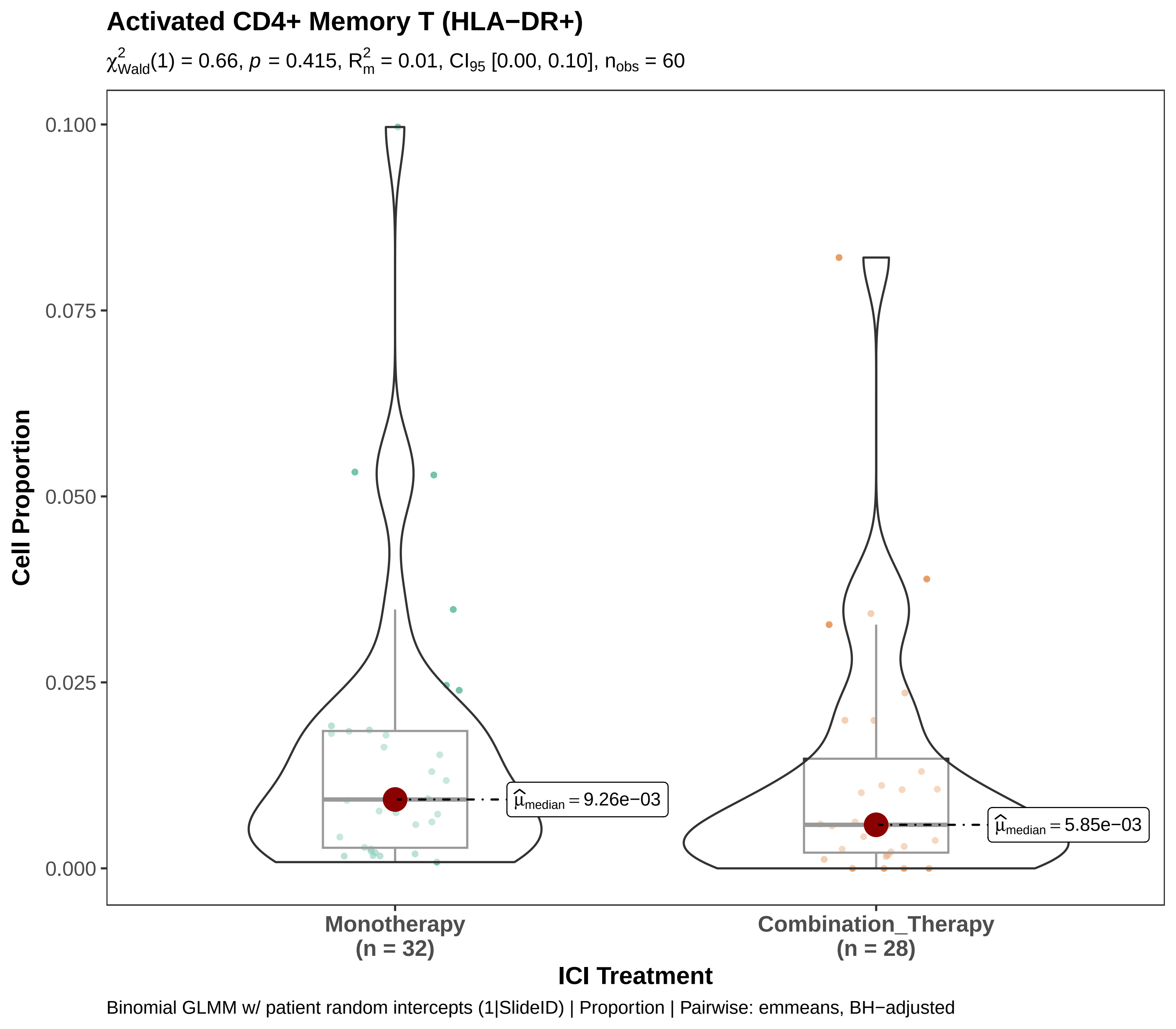

Activated CD4+ Memory T (HLA−DR+)
$\chi^2_{Wald}(1) = 0.66$, $p = 0.415$, $R^2_m = 0.01$, $CI_{95}$ [0.00, 0.10], $n_{obs} = 60$
Cell Proportion
0.100
0.075
0.050
0.025
0.000
$\widehat{\mu}_{median} = 9.26e{-}03$
$\widehat{\mu}_{median} = 5.85e{-}03$
Monotherapy
(n = 32)
Combination_Therapy
(n = 28)
ICI Treatment
Binomial GLMM w/ patient random intercepts (1|SlideID) | Proportion | Pairwise: emmeans, BH−adjusted

### Activated CD4+ Memory T (HLA−DR+)

$\chi^2_{Wald}(2) = 7.61$, $p = 0.022$, $R^2_m = 0.07$, $CI_{95}$ [0.01, 0.21], $n_{obs} = 144$

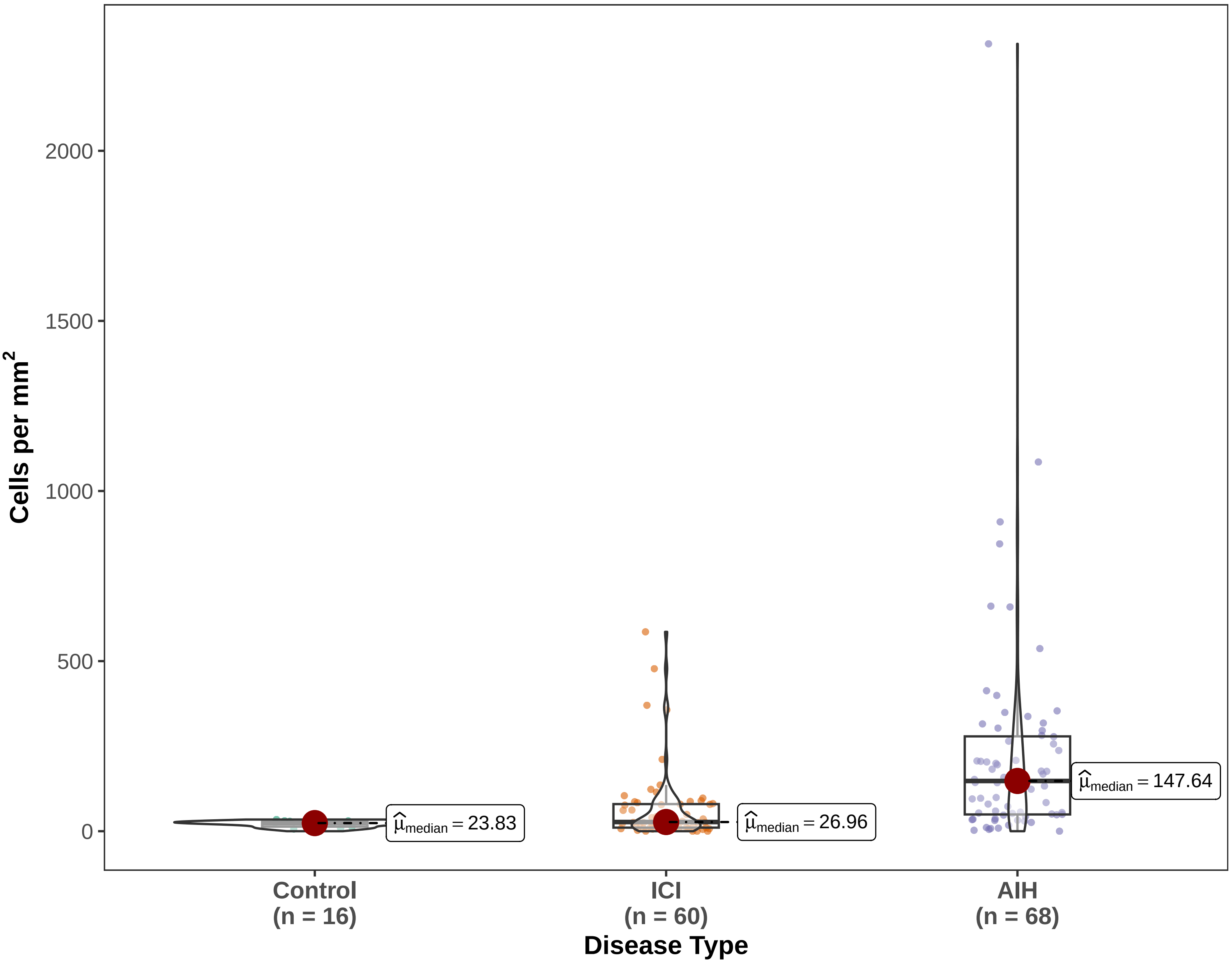


Gaussian LMM w/ patient random intercepts (1|SlideID) | Density | Pairwise: emmeans, BH−adjusted

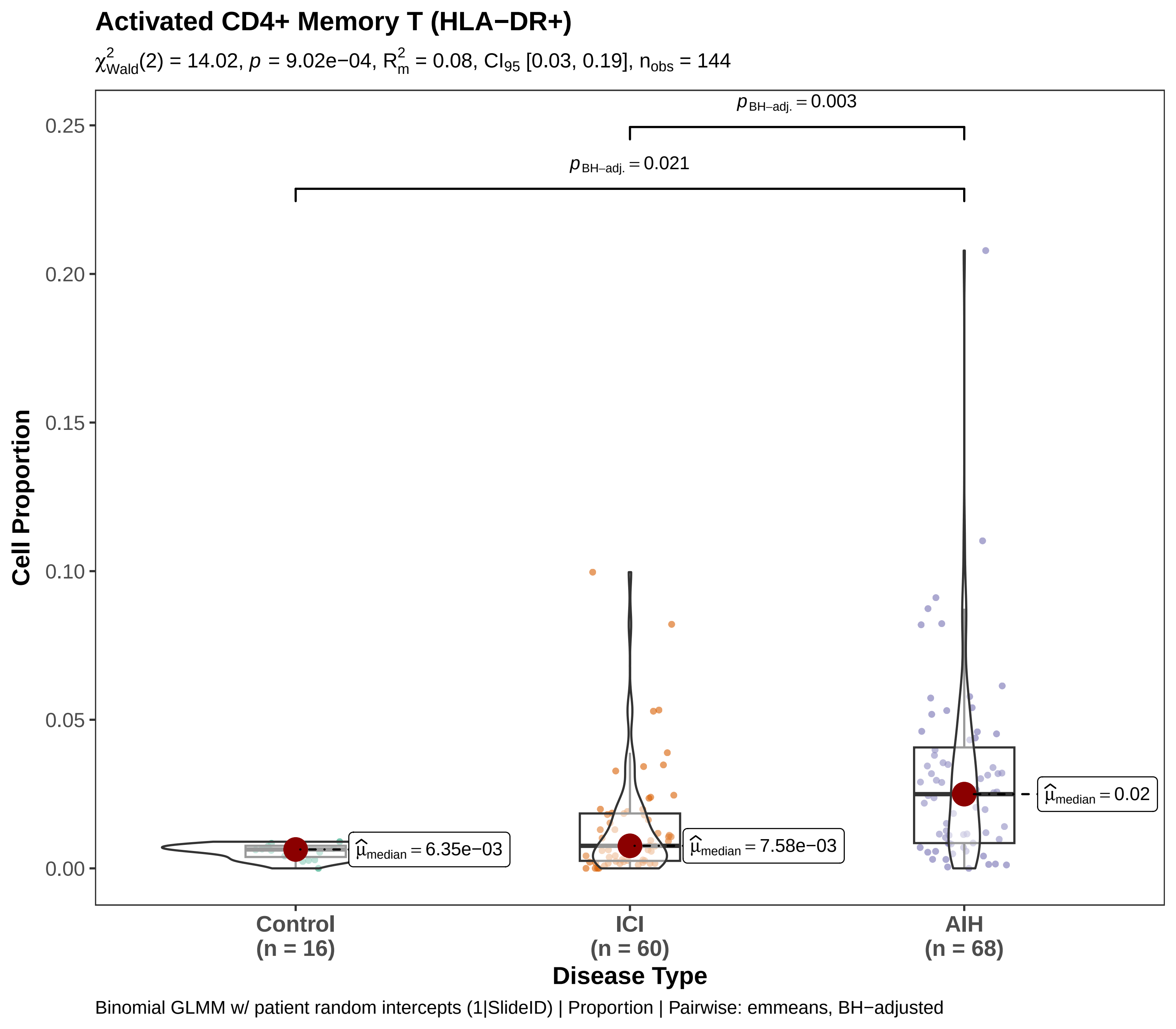
Activated CD4+ Memory T (HLA−DR+)
$\chi^2_{Wald}(2) = 14.02$, $p = 9.02e{-}04$, $R^2_m = 0.08$, $CI_{95}$ [0.03, 0.19], $n_{obs} = 144$
$p_{BH-adj.} = 0.003$
$p_{BH-adj.} = 0.021$
Cell Proportion
0.25
0.20
0.15
0.10
0.05
0.00
$\hat{\mu}_{median} = 6.35e{-}03$
$\hat{\mu}_{median} = 7.58e{-}03$
$\hat{\mu}_{median} = 0.02$
Control
(n = 16)
ICI
(n = 60)
AIH
(n = 68)
Disease Type
Binomial GLMM w/ patient random intercepts (1|SlideID) | Proportion | Pairwise: emmeans, BH−adjusted

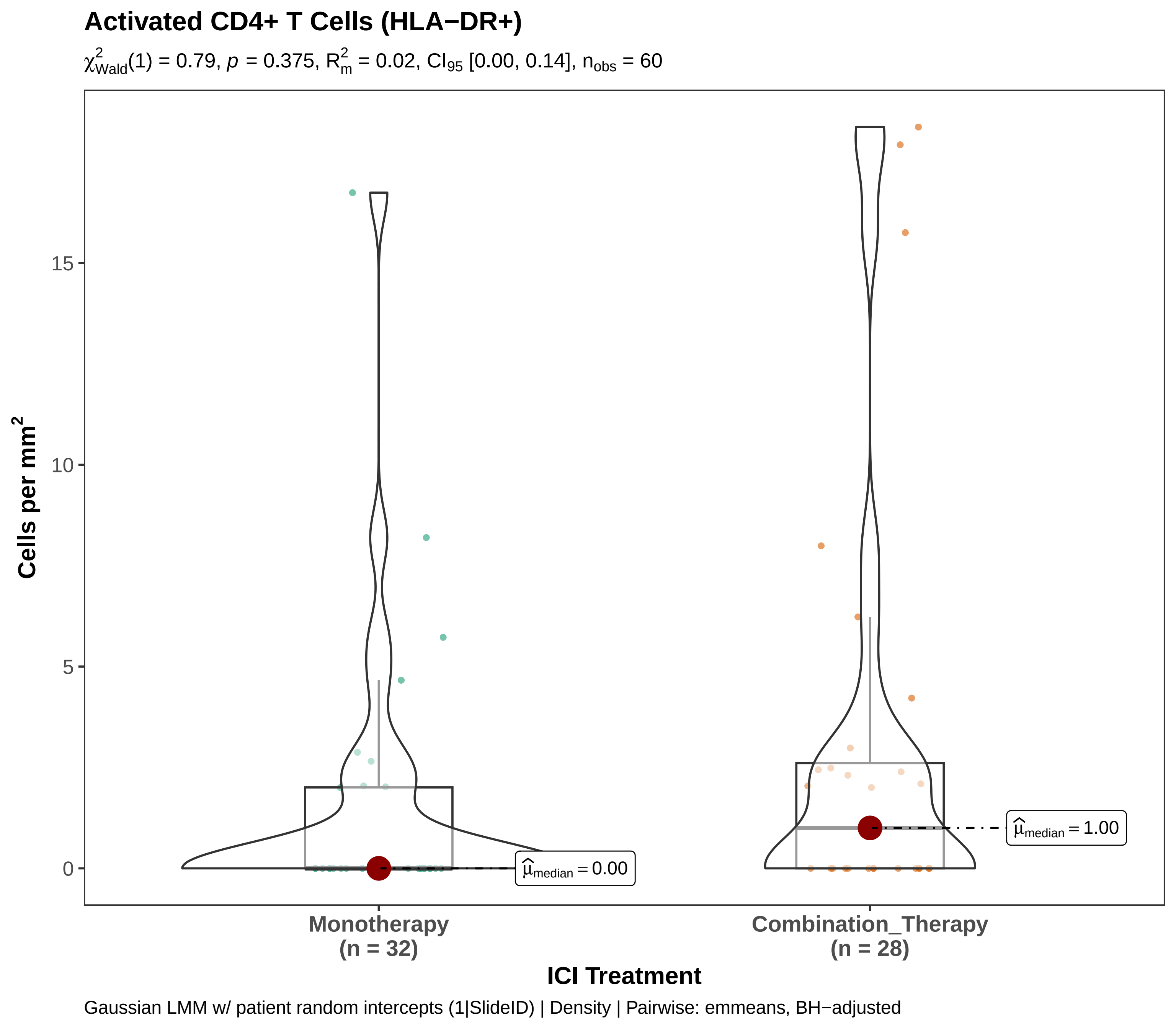

Activated CD4+ T Cells (HLA−DR+)
$\chi^2_{Wald}(1) = 0.79$, $p = 0.375$, $R^2_m = 0.02$, $CI_{95}$ [0.00, 0.14], $n_{obs} = 60$
Cells per mm$^2$
15
10
5
0
$\widehat{\mu}_{median} = 0.00$
$\widehat{\mu}_{median} = 1.00$
Monotherapy
(n = 32)
Combination_Therapy
(n = 28)
ICI Treatment
Gaussian LMM w/ patient random intercepts (1|SlideID) | Density | Pairwise: emmeans, BH−adjusted

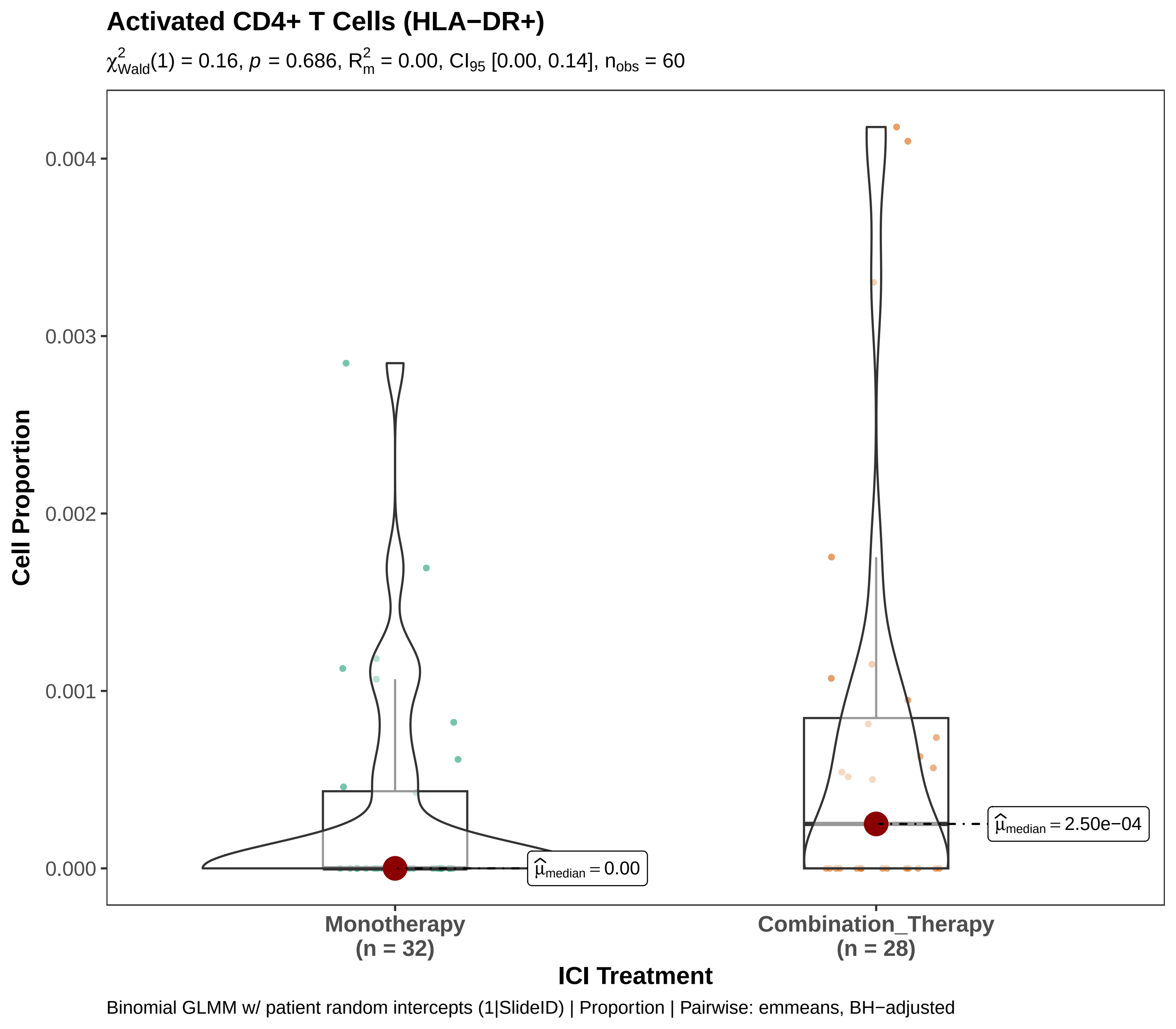

Activated CD4+ T Cells (HLA−DR+)
$\chi^2_{Wald}(1) = 0.16$, $p = 0.686$, $R^2_m = 0.00$, $CI_{95}$ [0.00, 0.14], $n_{obs} = 60$
Cell Proportion
0.004
0.003
0.002
0.001
0.000
$\widehat{\mu}_{median} = 0.00$
$\widehat{\mu}_{median} = 2.50e{-}04$
Monotherapy (n = 32)
Combination_Therapy (n = 28)
ICI Treatment
Binomial GLMM w/ patient random intercepts (1|SlideID) | Proportion | Pairwise: emmeans, BH−adjusted

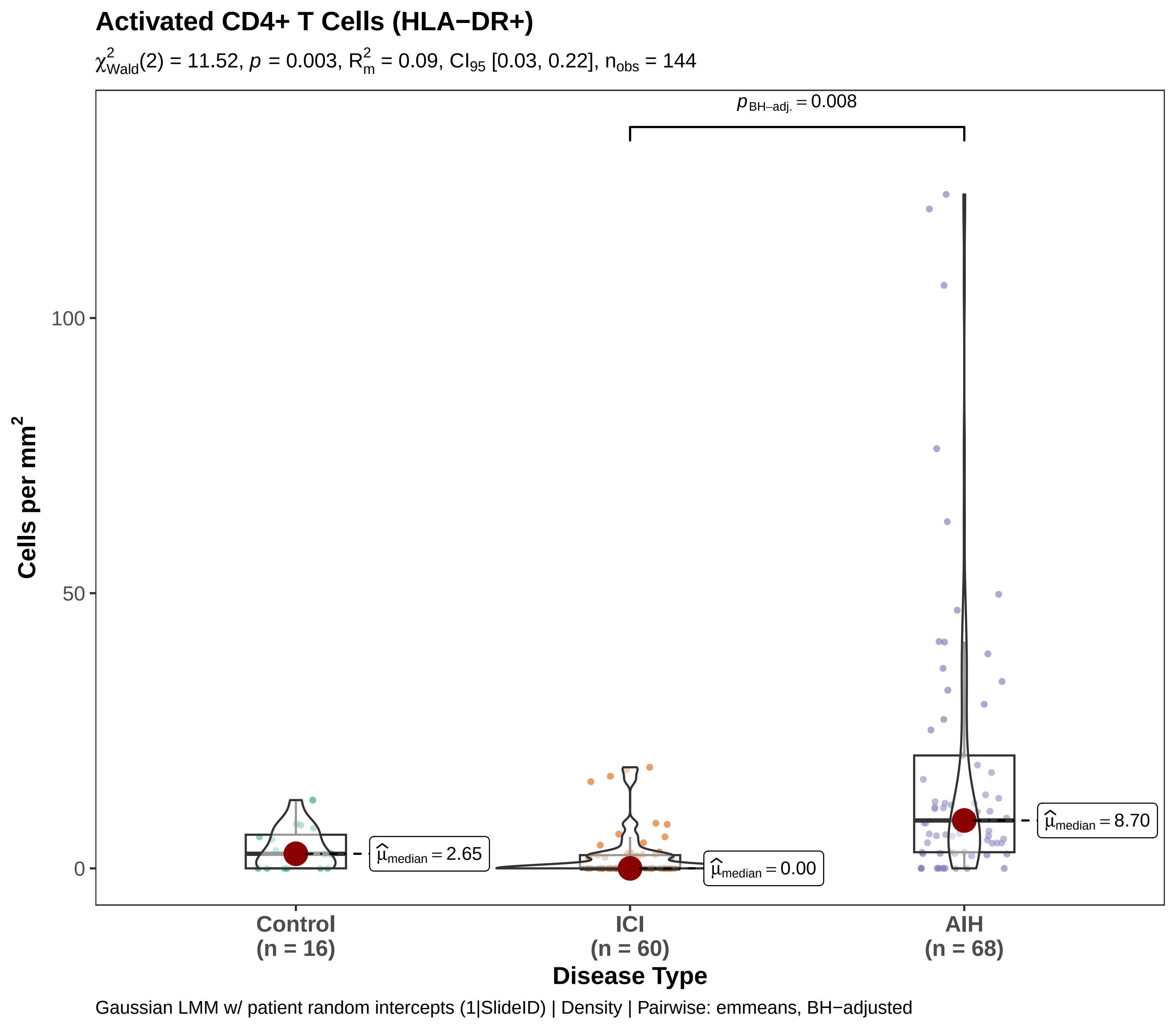
Activated CD4+ T Cells (HLA−DR+)
$\chi^2_{Wald}(2) = 11.52$, $p = 0.003$, $R^2_m = 0.09$, $CI_{95}$ [0.03, 0.22], $n_{obs} = 144$
$p_{BH-adj.} = 0.008$
$\hat{\mu}_{median} = 2.65$
$\hat{\mu}_{median} = 0.00$
$\hat{\mu}_{median} = 8.70$
Cells per mm$^2$
100
50
0
Control
(n = 16)
ICI
(n = 60)
AIH
(n = 68)
Disease Type
Gaussian LMM w/ patient random intercepts (1|SlideID) | Density | Pairwise: emmeans, BH−adjusted

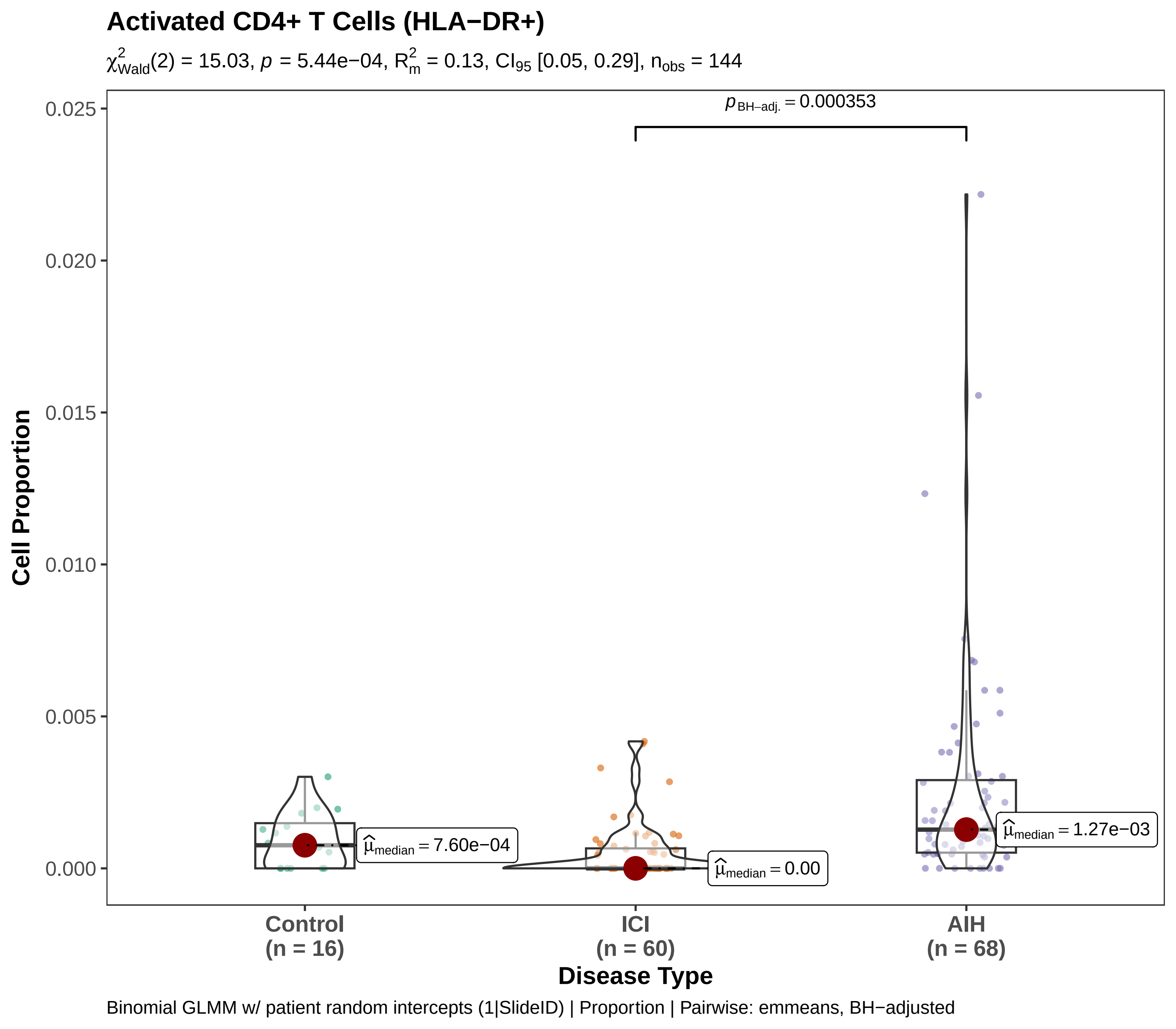

Activated CD4+ T Cells (HLA−DR+)
$\chi^2_{Wald}(2) = 15.03$, $p = 5.44e-04$, $R^2_m = 0.13$, $CI_{95}$ [0.05, 0.29], $n_{obs} = 144$
$p_{BH-adj.} = 0.000353$
0.025
0.020
0.015
0.010
0.005
0.000
Cell Proportion
$\widehat{\mu}_{median} = 7.60e-04$
$\widehat{\mu}_{median} = 0.00$
$\widehat{\mu}_{median} = 1.27e-03$
Control (n = 16)
ICI (n = 60)
AIH (n = 68)
Disease Type
Binomial GLMM w/ patient random intercepts (1|SlideID) | Proportion | Pairwise: emmeans, BH−adjusted

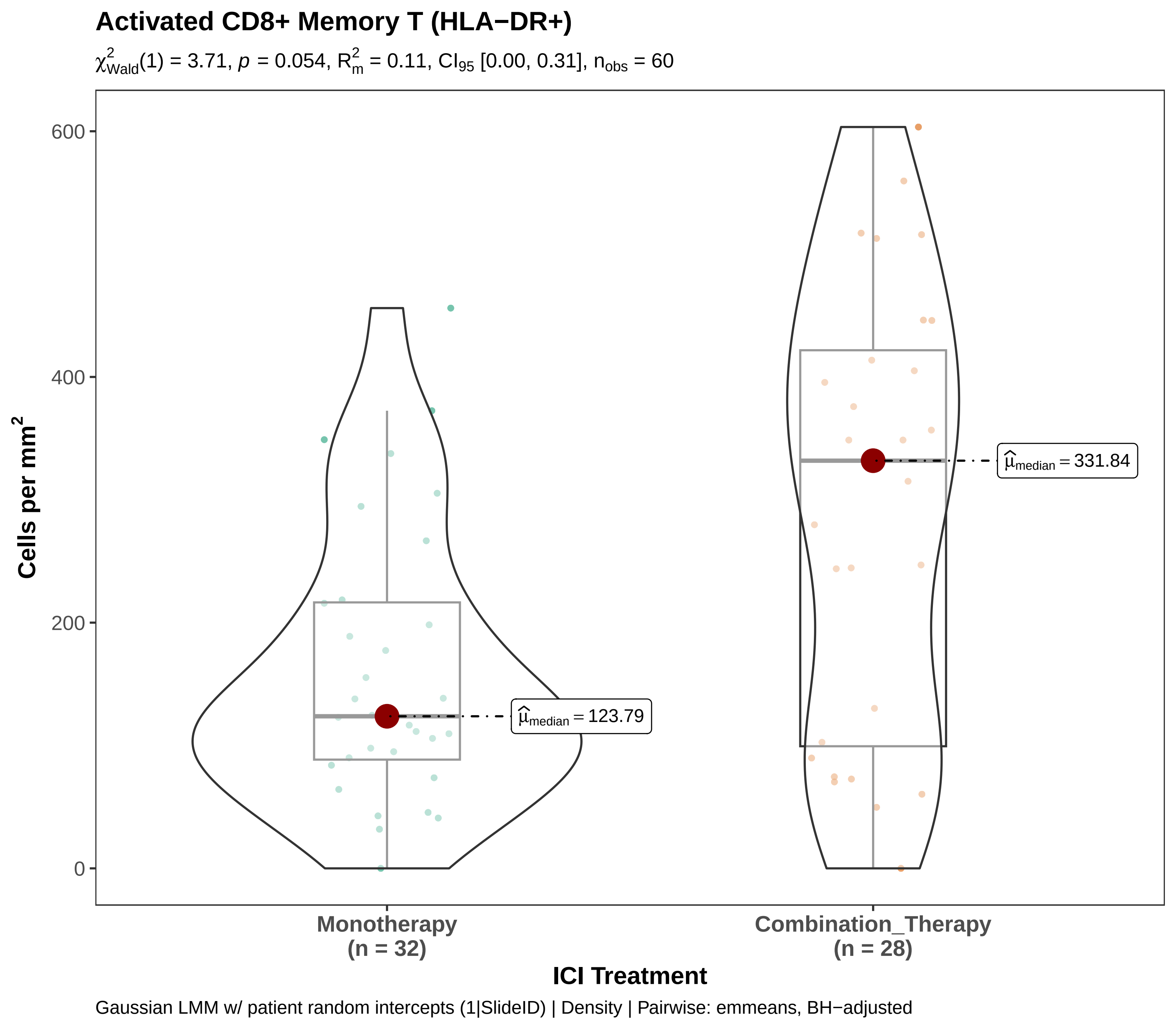

Activated CD8+ Memory T (HLA−DR+)
$\chi^2_{Wald}(1) = 3.71$, $p = 0.054$, $R^2_m = 0.11$, $CI_{95}$ [0.00, 0.31], $n_{obs} = 60$
Cells per mm$^2$
600
400
200
0
$\widehat{\mu}_{median} = 123.79$
$\widehat{\mu}_{median} = 331.84$
Monotherapy
(n = 32)
Combination_Therapy
(n = 28)
ICI Treatment
Gaussian LMM w/ patient random intercepts (1|SlideID) | Density | Pairwise: emmeans, BH−adjusted

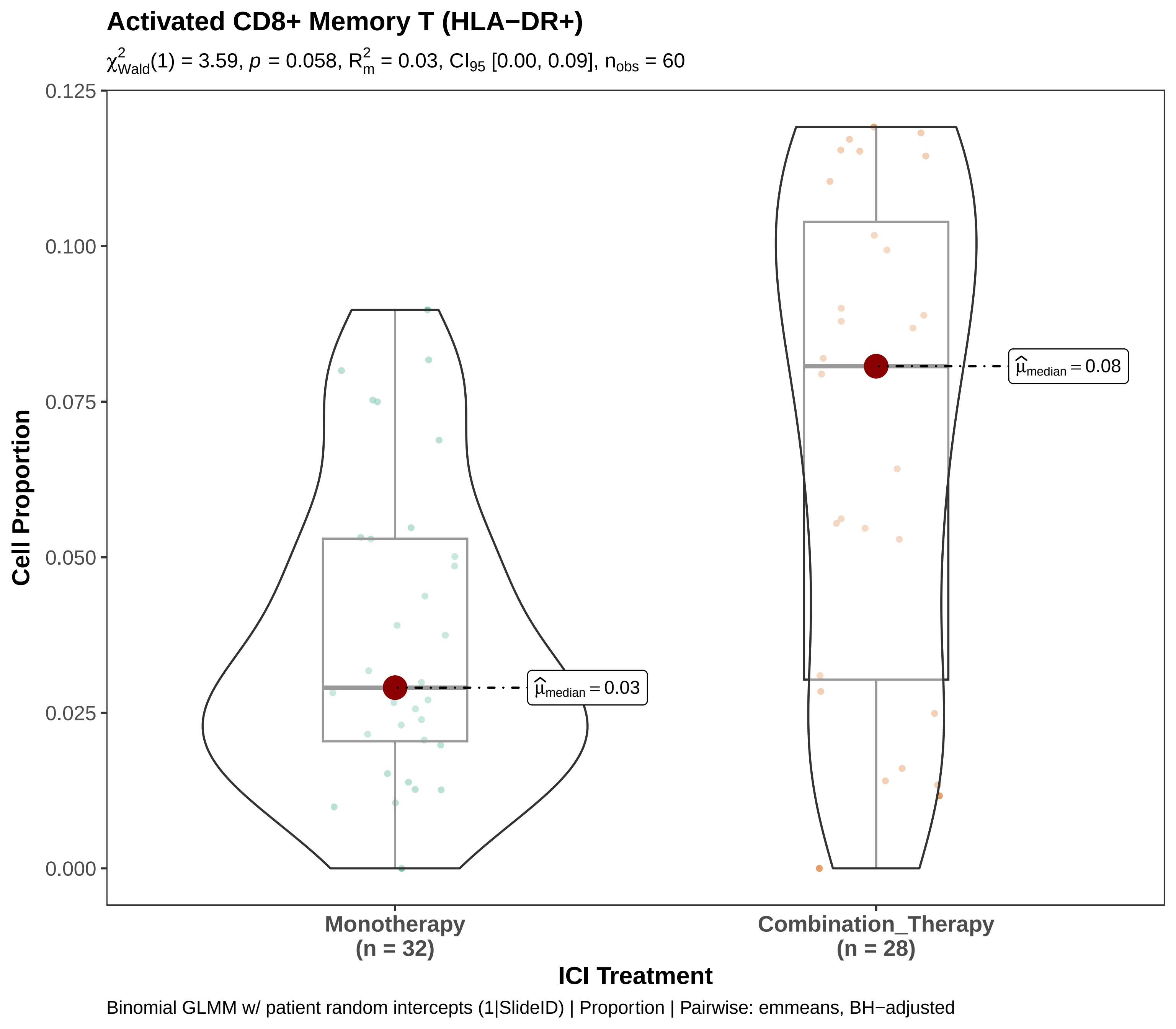

Activated CD8+ Memory T (HLA−DR+)
$\chi^2_{Wald}(1) = 3.59$, $p = 0.058$, $R^2_m = 0.03$, $CI_{95}$ [0.00, 0.09], $n_{obs} = 60$
Cell Proportion
0.125
0.100
0.075
0.050
0.025
0.000
$\widehat{\mu}_{median} = 0.03$
$\widehat{\mu}_{median} = 0.08$
Monotherapy
(n = 32)
Combination_Therapy
(n = 28)
ICI Treatment
Binomial GLMM w/ patient random intercepts (1|SlideID) | Proportion | Pairwise: emmeans, BH−adjusted

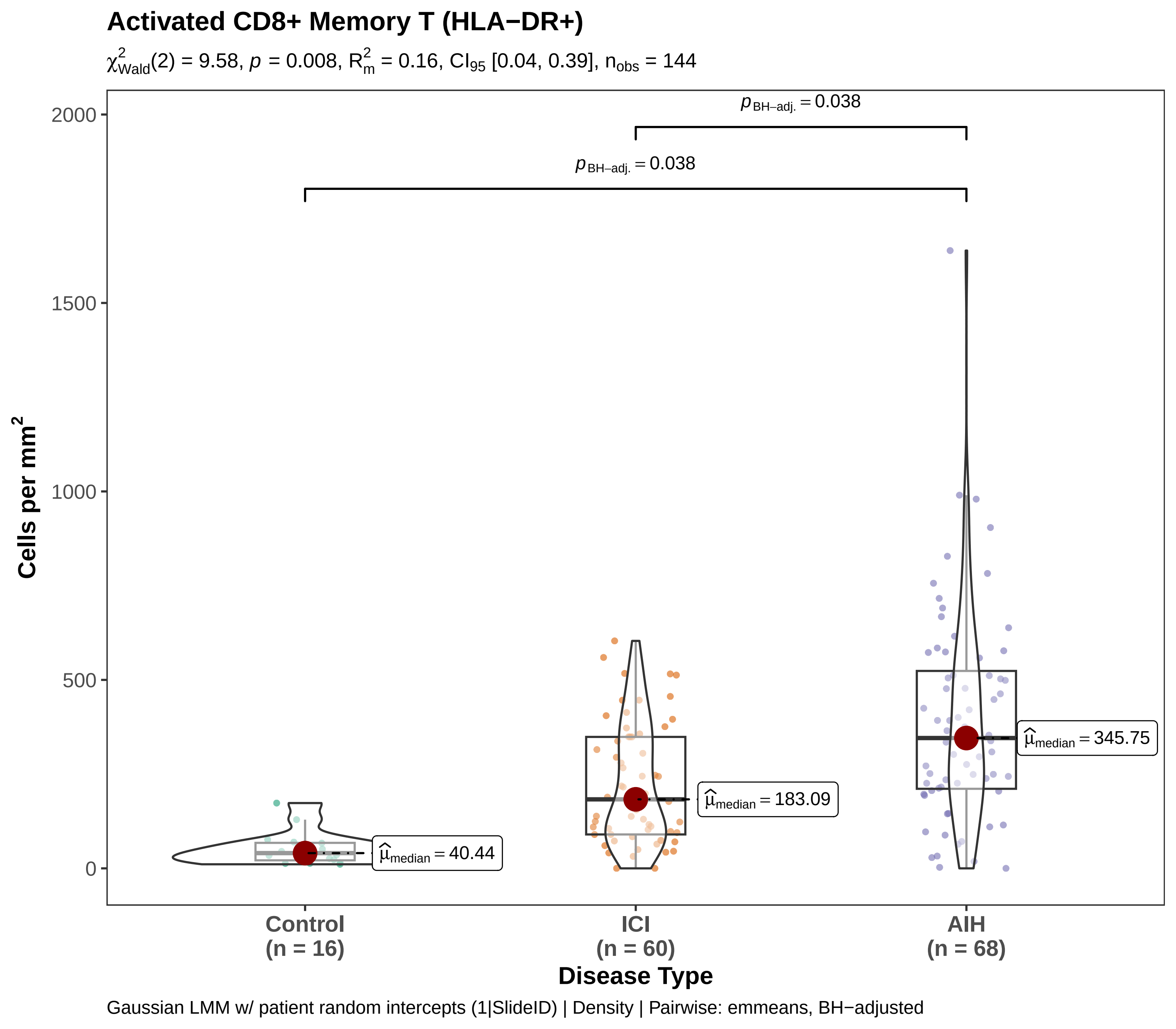

Activated CD8+ Memory T (HLA−DR+)
$\chi^2_{Wald}(2) = 9.58$, $p = 0.008$, $R^2_m = 0.16$, $CI_{95}$ [0.04, 0.39], $n_{obs} = 144$
$p_{BH-adj.} = 0.038$
$p_{BH-adj.} = 0.038$
$\widehat{\mu}_{median} = 40.44$
$\widehat{\mu}_{median} = 183.09$
$\widehat{\mu}_{median} = 345.75$
Cells per mm$^2$
2000
1500
1000
500
0
Control
(n = 16)
ICI
(n = 60)
AIH
(n = 68)
Disease Type
Gaussian LMM w/ patient random intercepts (1|SlideID) | Density | Pairwise: emmeans, BH−adjusted

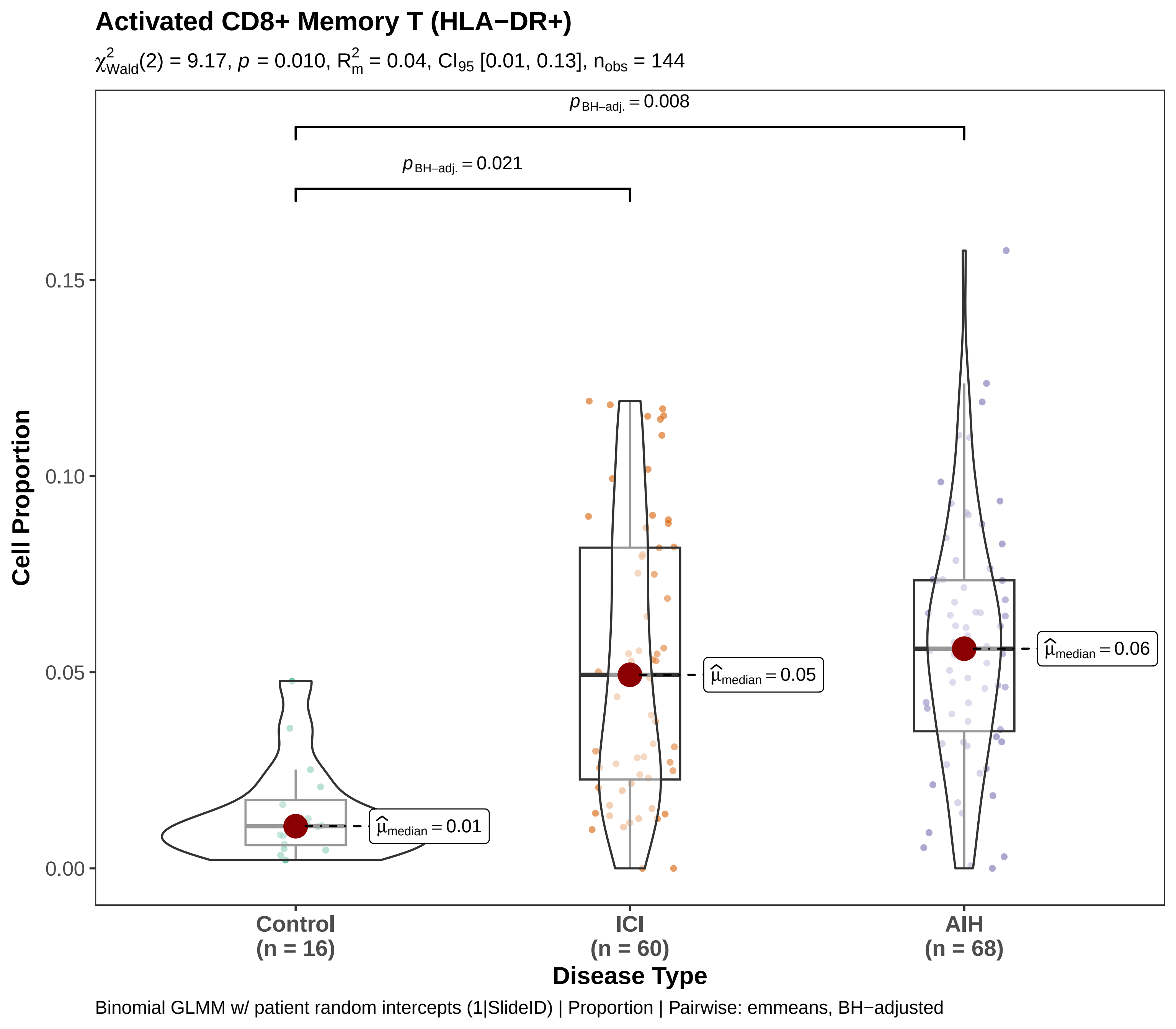

Activated CD8+ Memory T (HLA−DR+)
$\chi^2_{Wald}(2) = 9.17$, $p = 0.010$, $R^2_m = 0.04$, $CI_{95}$ [0.01, 0.13], $n_{obs} = 144$
$p_{BH-adj.} = 0.008$
$p_{BH-adj.} = 0.021$
$\hat{\mu}_{median} = 0.01$
$\hat{\mu}_{median} = 0.05$
$\hat{\mu}_{median} = 0.06$
Cell Proportion
0.15
0.10
0.05
0.00
Control (n = 16)
ICI (n = 60)
AIH (n = 68)
Disease Type
Binomial GLMM w/ patient random intercepts (1|SlideID) | Proportion | Pairwise: emmeans, BH−adjusted

**Activated CD8+ T cells (HLA−DR+)**

$\chi^2_{Wald}(1) = 2.05$, $p = 0.153$, $R^2_m = 0.05$, $CI_{95}$ [0.00, 0.20], $n_{obs} = 60$

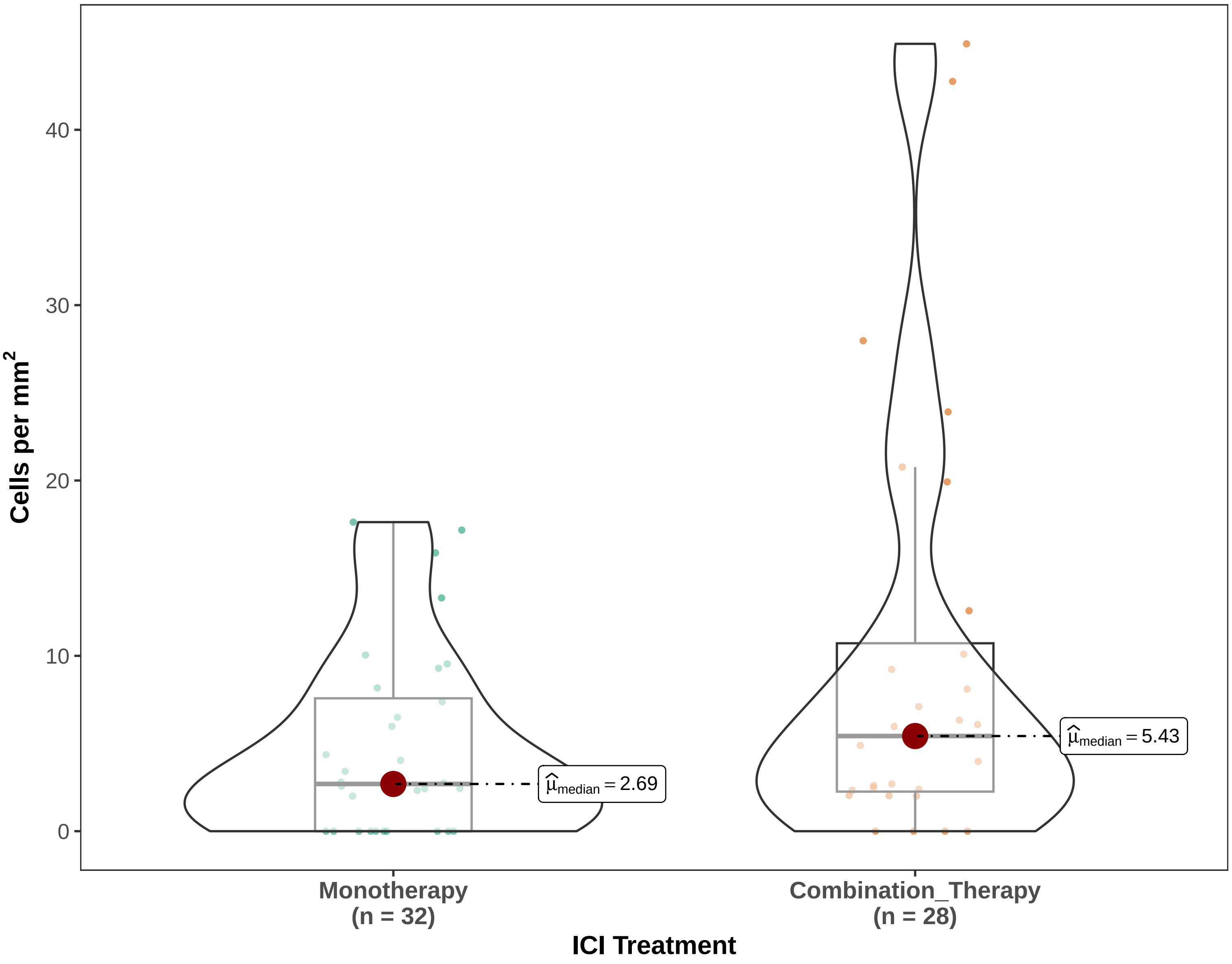


Gaussian LMM w/ patient random intercepts (1|SlideID) | Density | Pairwise: emmeans, BH−adjusted

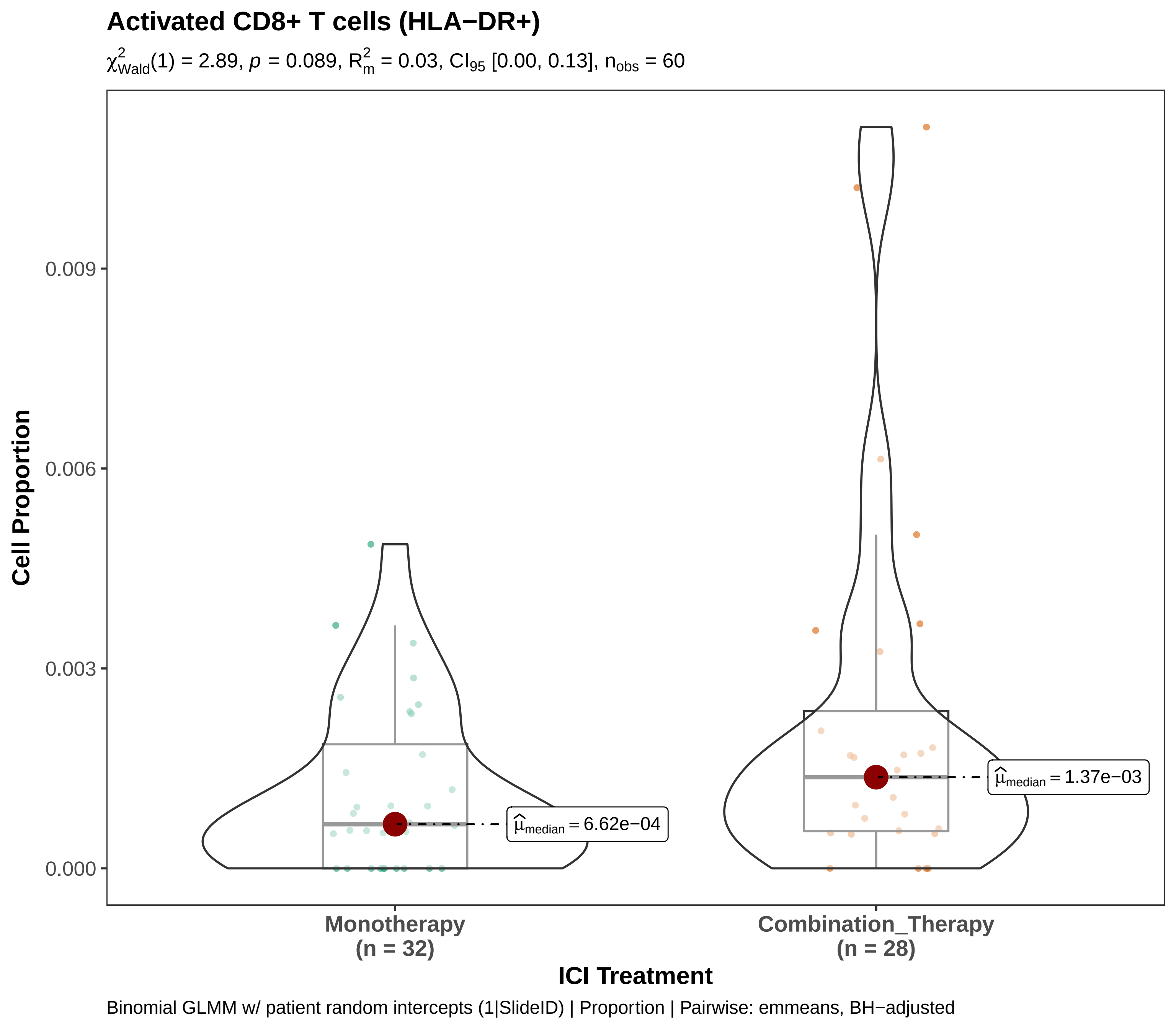
Activated CD8+ T cells (HLA−DR+)
χ²Wald(1) = 2.89, p = 0.089, R²m = 0.03, CI95 [0.00, 0.13], nobs = 60
Cell Proportion
0.009
0.006
0.003
0.000
μ̂median = 6.62e−04
μ̂median = 1.37e−03
Monotherapy
(n = 32)
Combination_Therapy
(n = 28)
ICI Treatment
Binomial GLMM w/ patient random intercepts (1|SlideID) | Proportion | Pairwise: emmeans, BH−adjusted

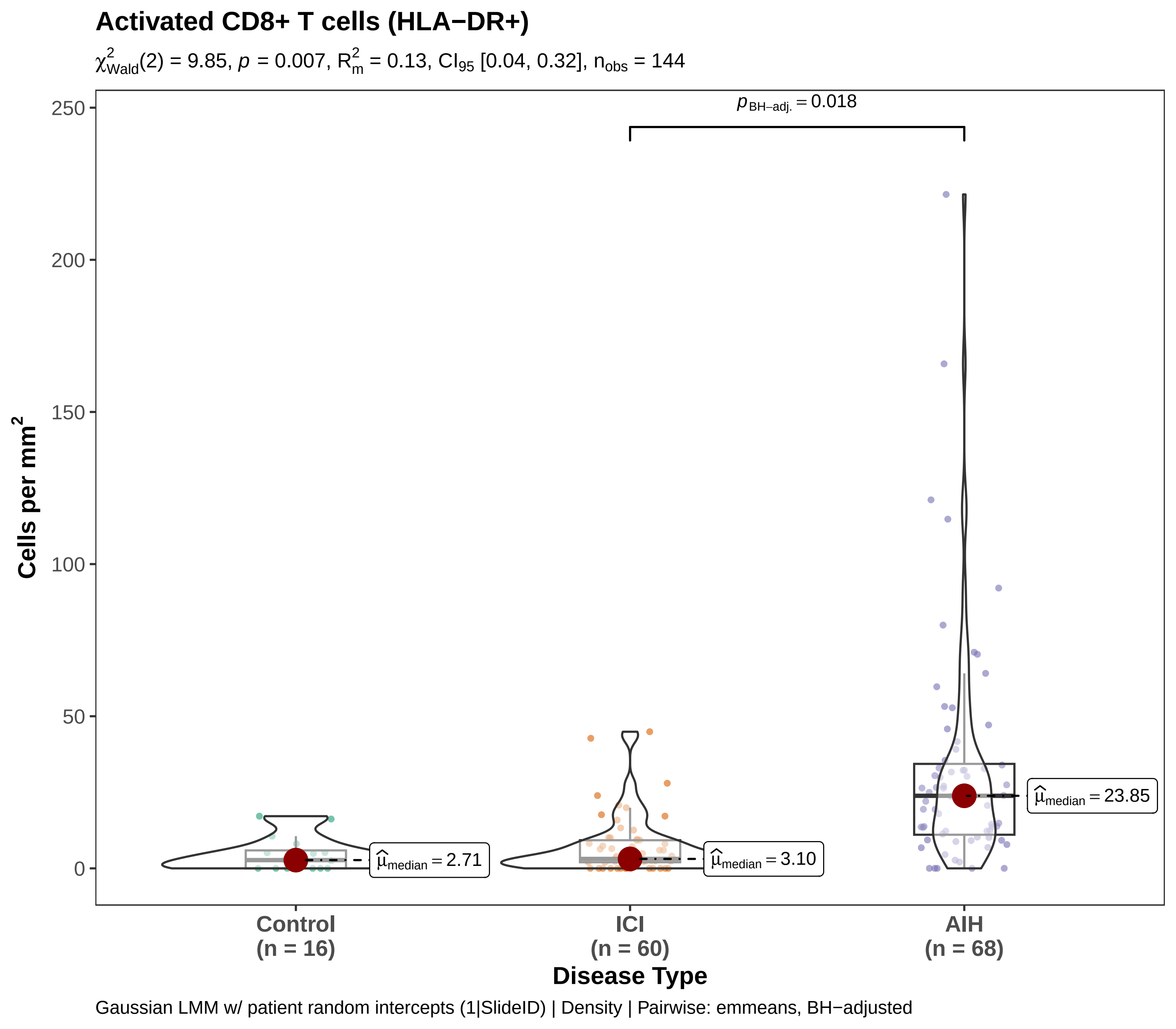
Activated CD8+ T cells (HLA−DR+)
χ²Wald(2) = 9.85, p = 0.007, R²m = 0.13, CI95 [0.04, 0.32], nobs = 144
pBH−adj. = 0.018
250
200
150
100
50
0
Cells per mm²
μ̂median = 2.71
μ̂median = 3.10
μ̂median = 23.85
Control
(n = 16)
ICI
(n = 60)
AIH
(n = 68)
Disease Type
Gaussian LMM w/ patient random intercepts (1|SlideID) | Density | Pairwise: emmeans, BH−adjusted

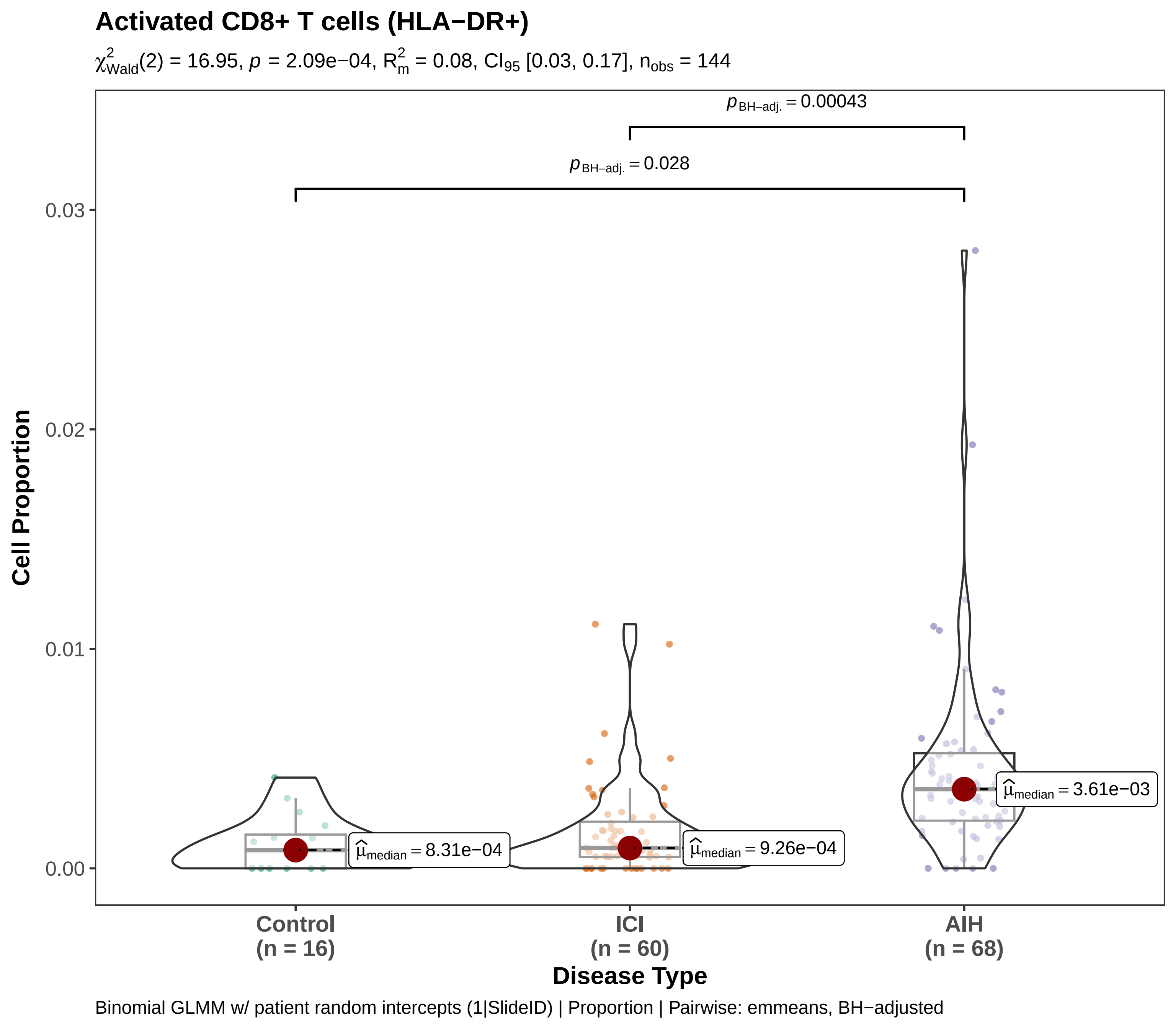
Activated CD8+ T cells (HLA−DR+)
$\chi^2_{Wald}(2) = 16.95$, $p = 2.09e{-}04$, $R^2_m = 0.08$, $CI_{95}$ [0.03, 0.17], $n_{obs} = 144$
$p_{BH-adj.} = 0.00043$
$p_{BH-adj.} = 0.028$
Cell Proportion
0.03
0.02
0.01
0.00
$\hat{\mu}_{median} = 8.31e{-}04$
$\hat{\mu}_{median} = 9.26e{-}04$
$\hat{\mu}_{median} = 3.61e{-}03$
Control (n = 16)
ICI (n = 60)
AIH (n = 68)
Disease Type
Binomial GLMM w/ patient random intercepts (1|SlideID) | Proportion | Pairwise: emmeans, BH−adjusted

#### CD4+ Memory T

$\chi^2_{\text{Wald}}(1) = 0.12$, $p = 0.731$, $R^2_{\text{m}} = 0.00$, $\text{CI}_{95}$ [0.00, 0.08], $n_{\text{obs}} = 60$

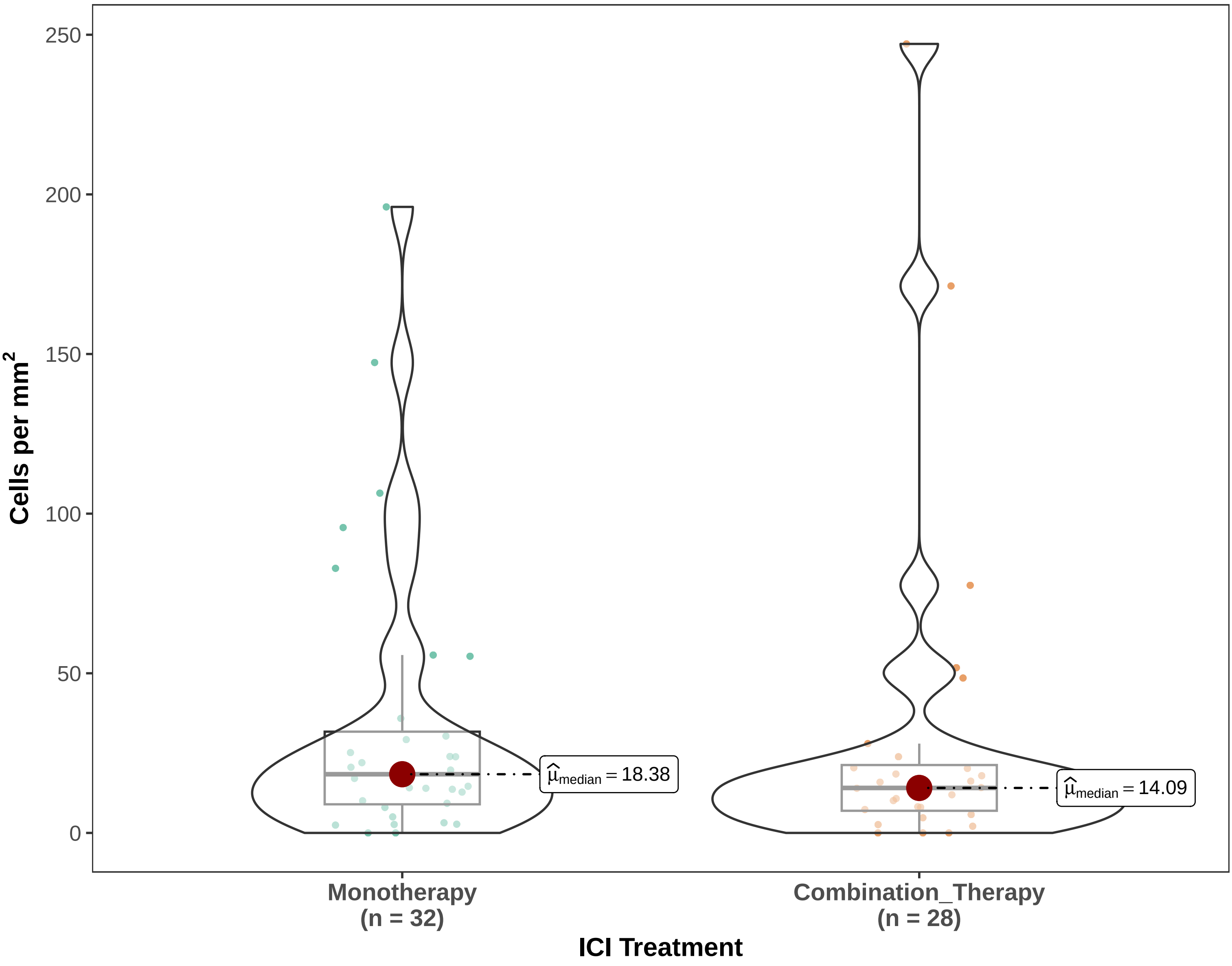


Gaussian LMM w/ patient random intercepts (1|SlideID) | Density | Pairwise: emmeans, BH−adjusted

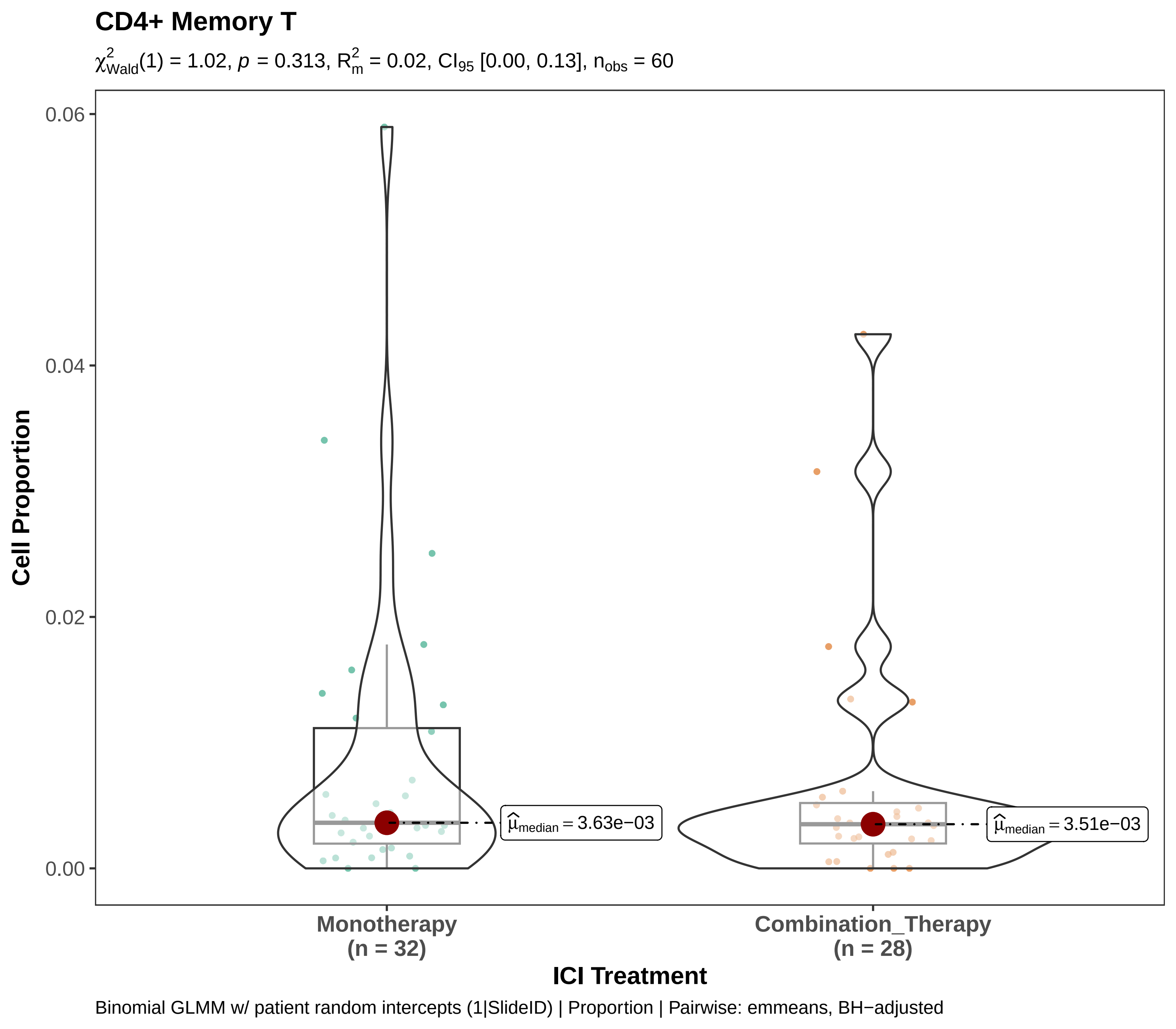

CD4+ Memory T
$\chi^2_{Wald}(1) = 1.02$, $p = 0.313$, $R^2_m = 0.02$, $CI_{95}$ [0.00, 0.13], $n_{obs} = 60$
Cell Proportion
0.06
0.04
0.02
0.00
$\widehat{\mu}_{median} = 3.63e{-}03$
$\widehat{\mu}_{median} = 3.51e{-}03$
Monotherapy
(n = 32)
Combination_Therapy
(n = 28)
ICI Treatment
Binomial GLMM w/ patient random intercepts (1|SlideID) | Proportion | Pairwise: emmeans, BH−adjusted

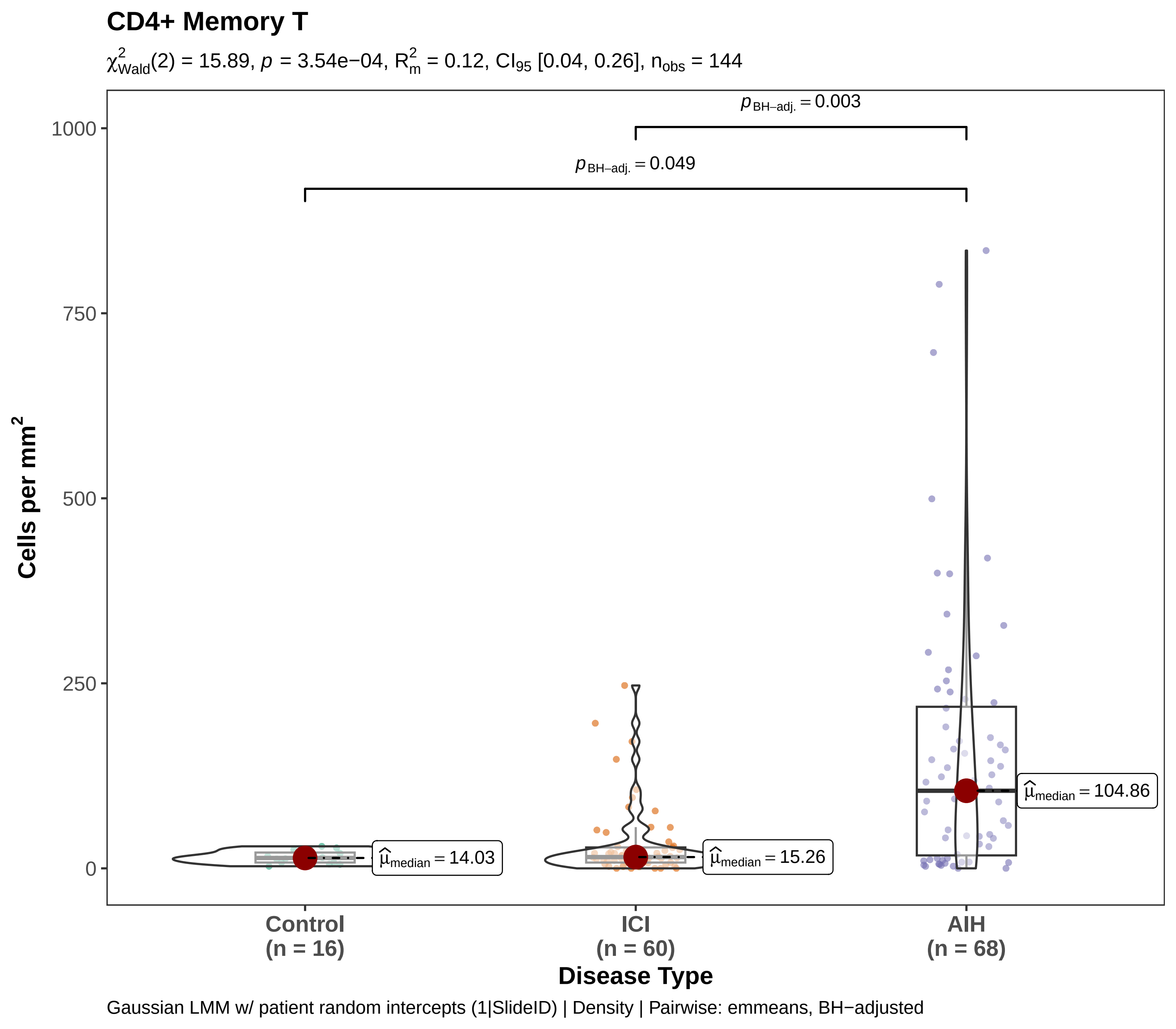
CD4+ Memory T
$\chi^2_{Wald}(2) = 15.89$, $p = 3.54e{-04}$, $R^2_m = 0.12$, $CI_{95}$ [0.04, 0.26], $n_{obs} = 144$
$p_{BH-adj.} = 0.003$
$p_{BH-adj.} = 0.049$
1000
750
500
250
0
Cells per mm²
$\hat{\mu}_{median} = 14.03$
$\hat{\mu}_{median} = 15.26$
$\hat{\mu}_{median} = 104.86$
Control
(n = 16)
ICI
(n = 60)
AIH
(n = 68)
Disease Type
Gaussian LMM w/ patient random intercepts (1|SlideID) | Density | Pairwise: emmeans, BH−adjusted

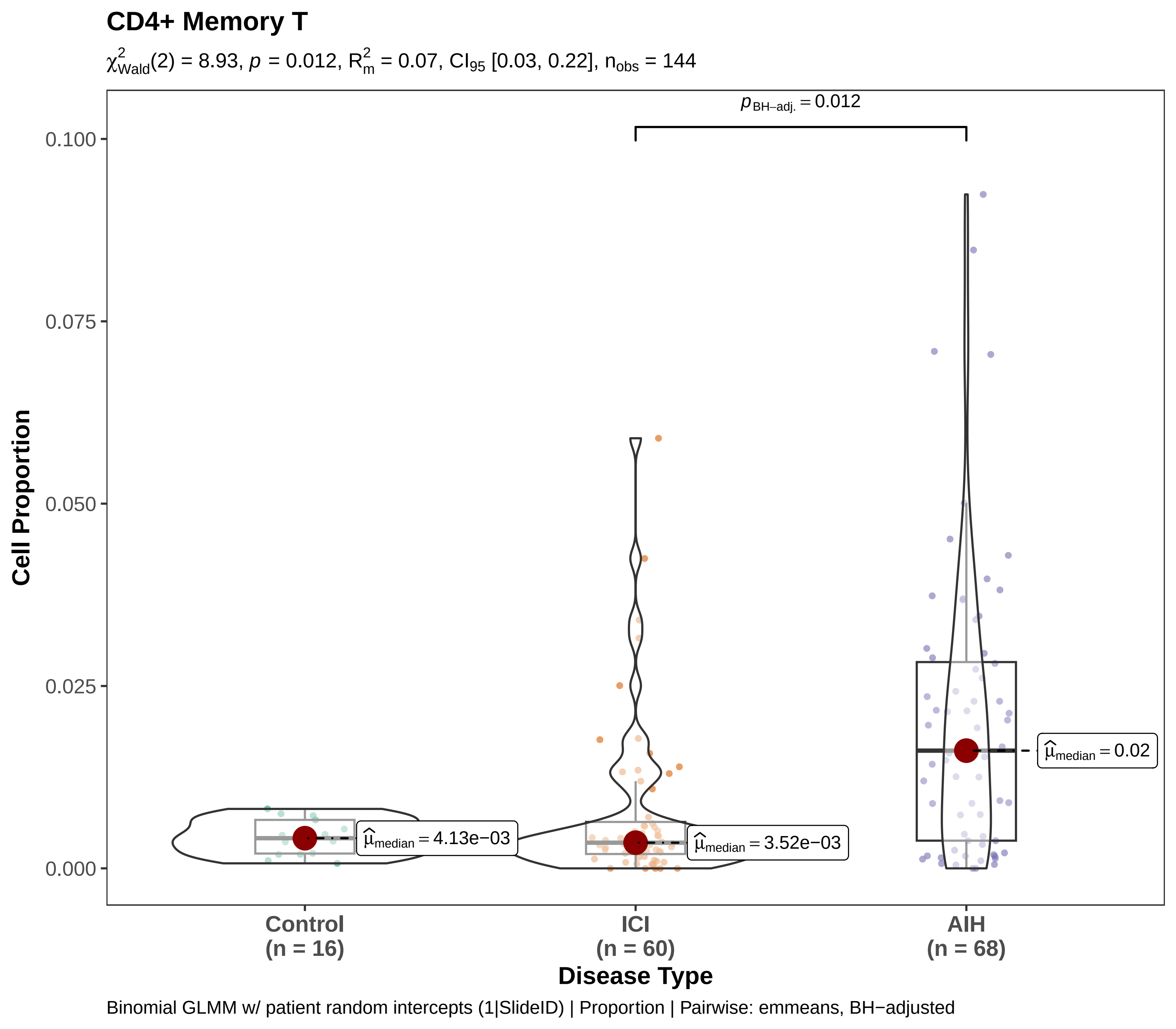
CD4+ Memory T
$\chi^2_{Wald}(2) = 8.93$, $p = 0.012$, $R^2_m = 0.07$, $CI_{95}$ [0.03, 0.22], $n_{obs} = 144$
$p_{BH-adj.} = 0.012$
0.100
0.075
0.050
0.025
0.000
Cell Proportion
$\hat{\mu}_{median} = 4.13e-03$
$\hat{\mu}_{median} = 3.52e-03$
$\hat{\mu}_{median} = 0.02$
Control
(n = 16)
ICI
(n = 60)
AIH
(n = 68)
Disease Type
Binomial GLMM w/ patient random intercepts (1|SlideID) | Proportion | Pairwise: emmeans, BH−adjusted

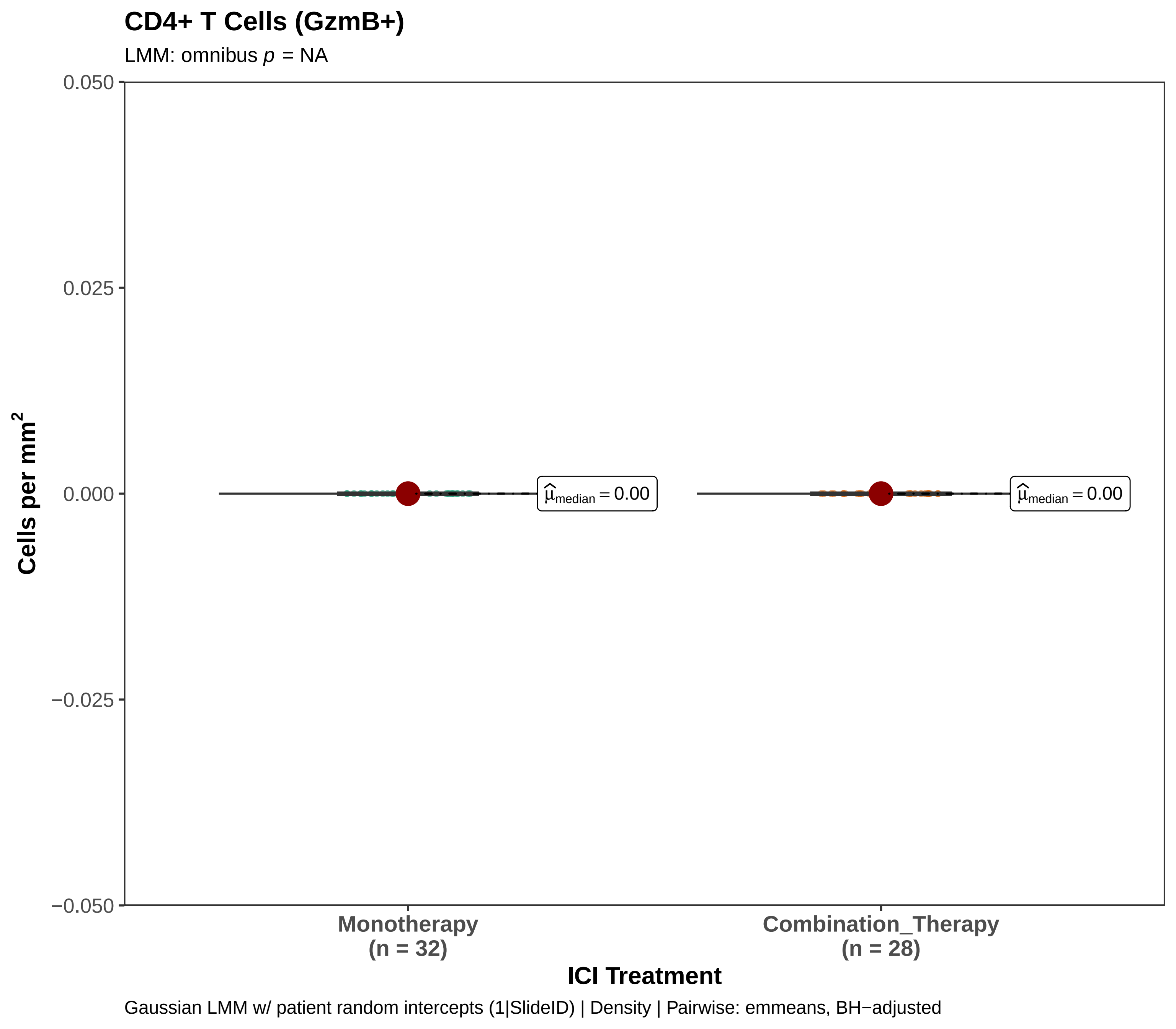
CD4+ T Cells (GzmB+)
LMM: omnibus p = NA
0.050
0.025
0.000
−0.025
−0.050
Cells per mm2
μ̂median = 0.00
μ̂median = 0.00
Monotherapy
(n = 32)
Combination_Therapy
(n = 28)
ICI Treatment
Gaussian LMM w/ patient random intercepts (1|SlideID) | Density | Pairwise: emmeans, BH−adjusted

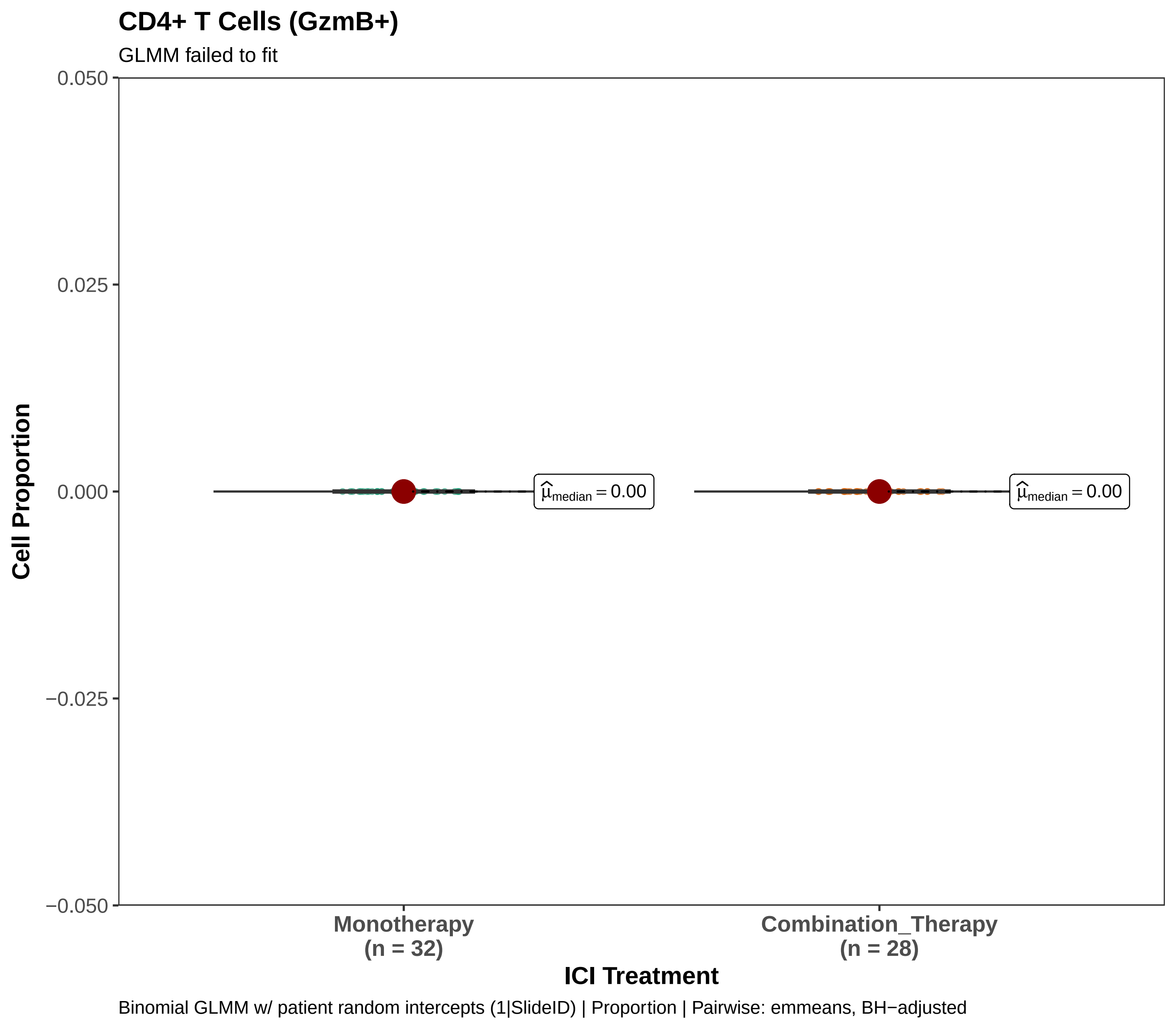

CD4+ T Cells (GzmB+)
GLMM failed to fit
0.050
0.025
0.000
−0.025
−0.050
Cell Proportion
$\widehat{\mu}_{median} = 0.00$
$\widehat{\mu}_{median} = 0.00$
Monotherapy
(n = 32)
Combination_Therapy
(n = 28)
ICI Treatment
Binomial GLMM w/ patient random intercepts (1|SlideID) | Proportion | Pairwise: emmeans, BH−adjusted

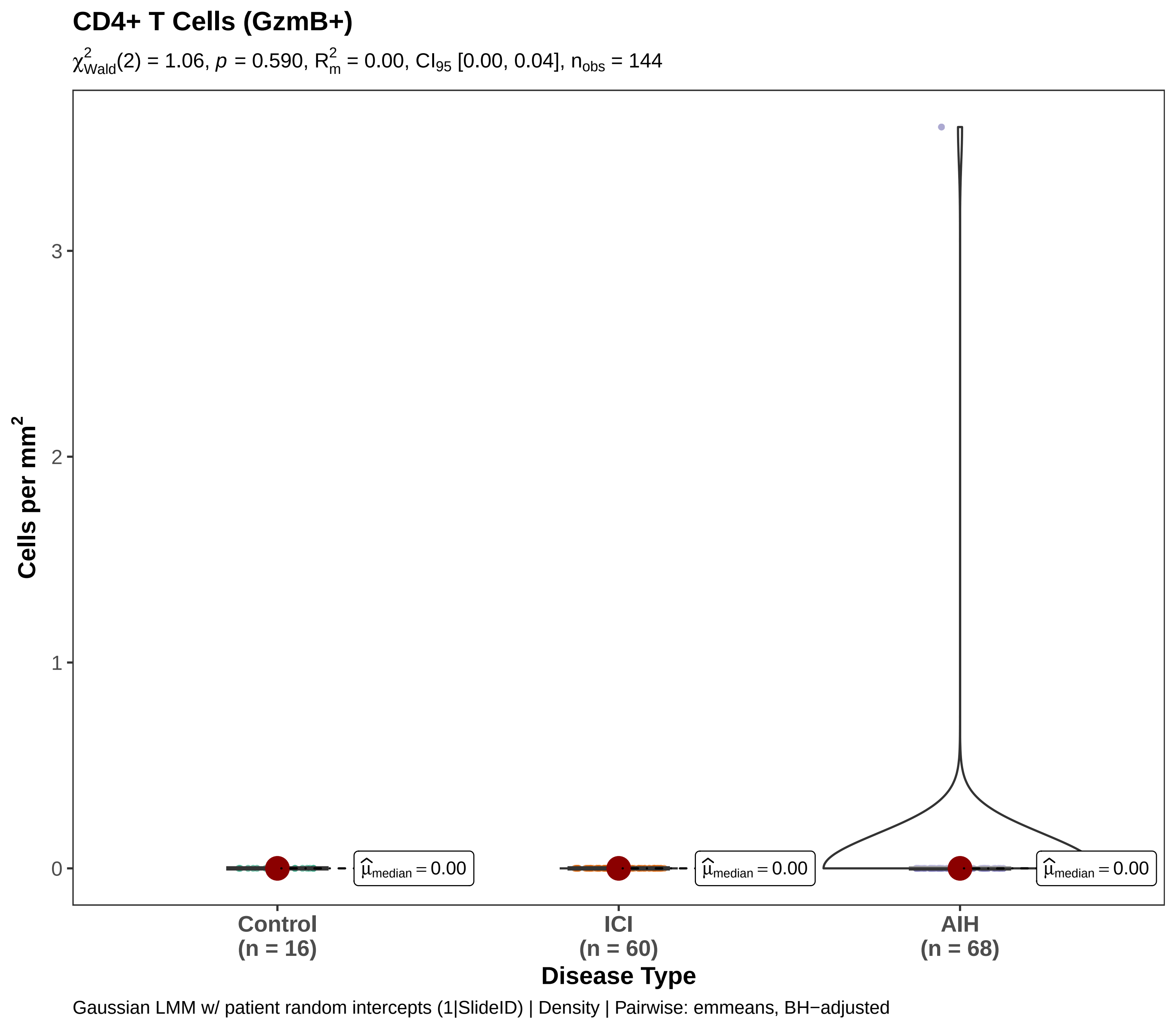
CD4+ T Cells (GzmB+)
$\chi^2_{Wald}(2) = 1.06$, $p = 0.590$, $R^2_m = 0.00$, $CI_{95}$ [0.00, 0.04], $n_{obs} = 144$
Cells per mm$^2$
3
2
1
0
$\hat{\mu}_{median} = 0.00$
$\hat{\mu}_{median} = 0.00$
$\hat{\mu}_{median} = 0.00$
Control
(n = 16)
ICI
(n = 60)
AIH
(n = 68)
Disease Type
Gaussian LMM w/ patient random intercepts (1|SlideID) | Density | Pairwise: emmeans, BH−adjusted

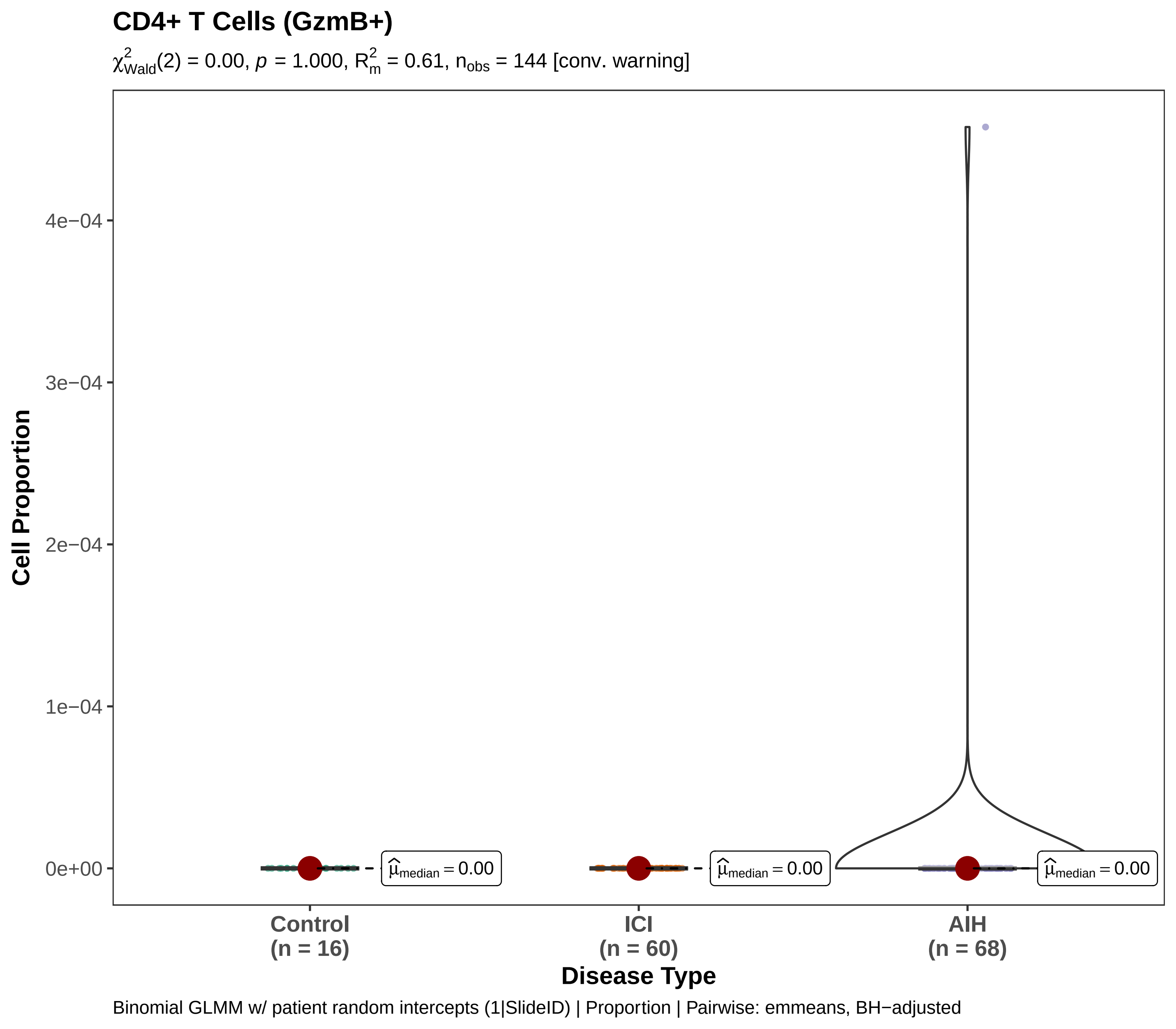

CD4+ T Cells (GzmB+)
$\chi^2_{Wald}(2) = 0.00$, $p = 1.000$, $R^2_m = 0.61$, $n_{obs} = 144$ [conv. warning]
Cell Proportion
4e−04
3e−04
2e−04
1e−04
0e+00
$\hat{\mu}_{median} = 0.00$
$\hat{\mu}_{median} = 0.00$
$\hat{\mu}_{median} = 0.00$
Control
(n = 16)
ICI
(n = 60)
AIH
(n = 68)
Disease Type
Binomial GLMM w/ patient random intercepts (1|SlideID) | Proportion | Pairwise: emmeans, BH−adjusted

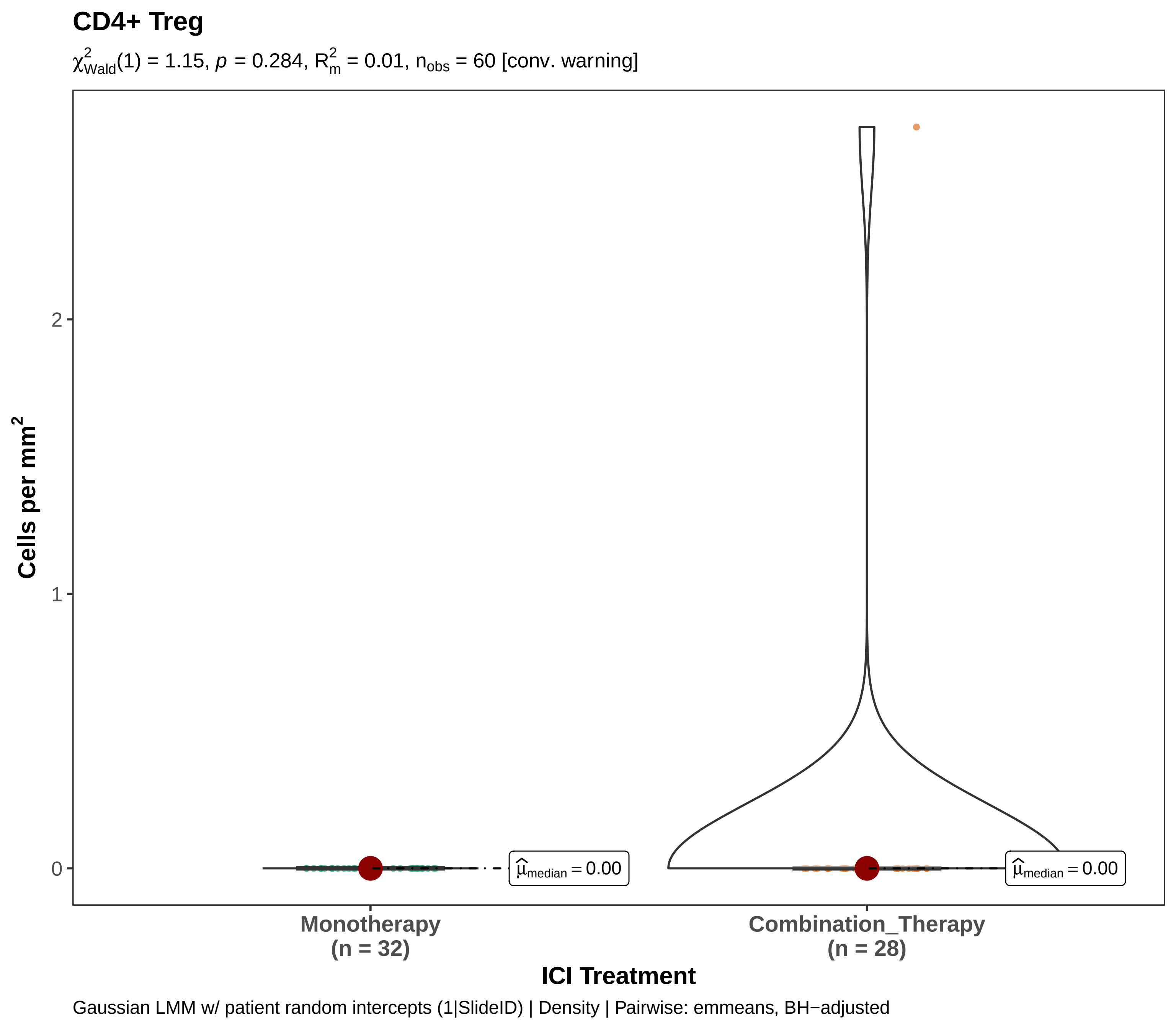

CD4+ Treg
$\chi^2_{Wald}(1) = 1.15$, $p = 0.284$, $R^2_m = 0.01$, $n_{obs} = 60$ [conv. warning]
Cells per mm$^2$
0
1
2
$\widehat{\mu}_{median} = 0.00$
$\widehat{\mu}_{median} = 0.00$
Monotherapy
(n = 32)
Combination_Therapy
(n = 28)
ICI Treatment
Gaussian LMM w/ patient random intercepts (1|SlideID) | Density | Pairwise: emmeans, BH−adjusted

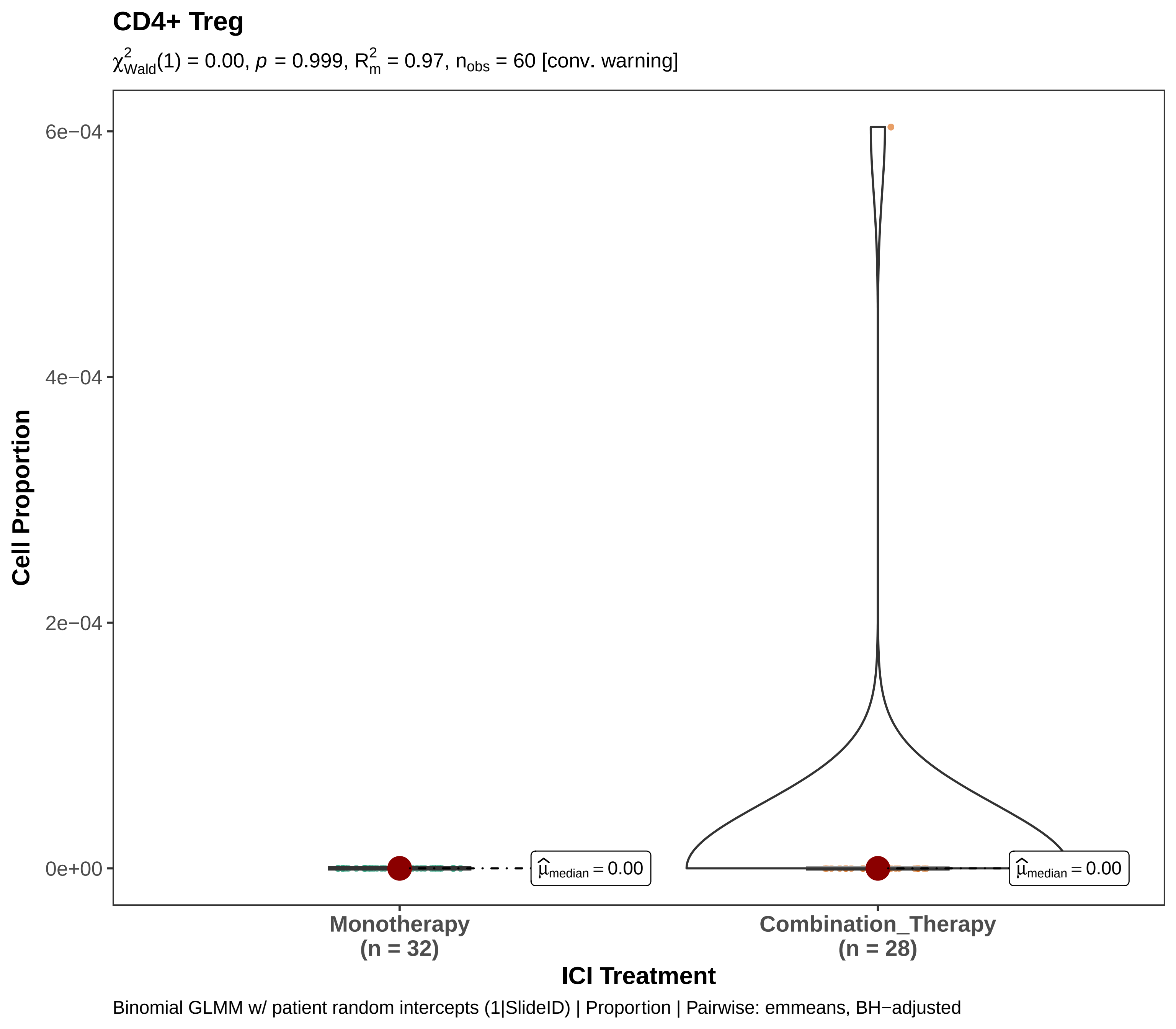
CD4+ Treg
χ²Wald(1) = 0.00, p = 0.999, R²m = 0.97, nobs = 60 [conv. warning]
6e−04
4e−04
2e−04
0e+00
Cell Proportion
μ̂median = 0.00
μ̂median = 0.00
Monotherapy
(n = 32)
Combination_Therapy
(n = 28)
ICI Treatment
Binomial GLMM w/ patient random intercepts (1|SlideID) | Proportion | Pairwise: emmeans, BH−adjusted

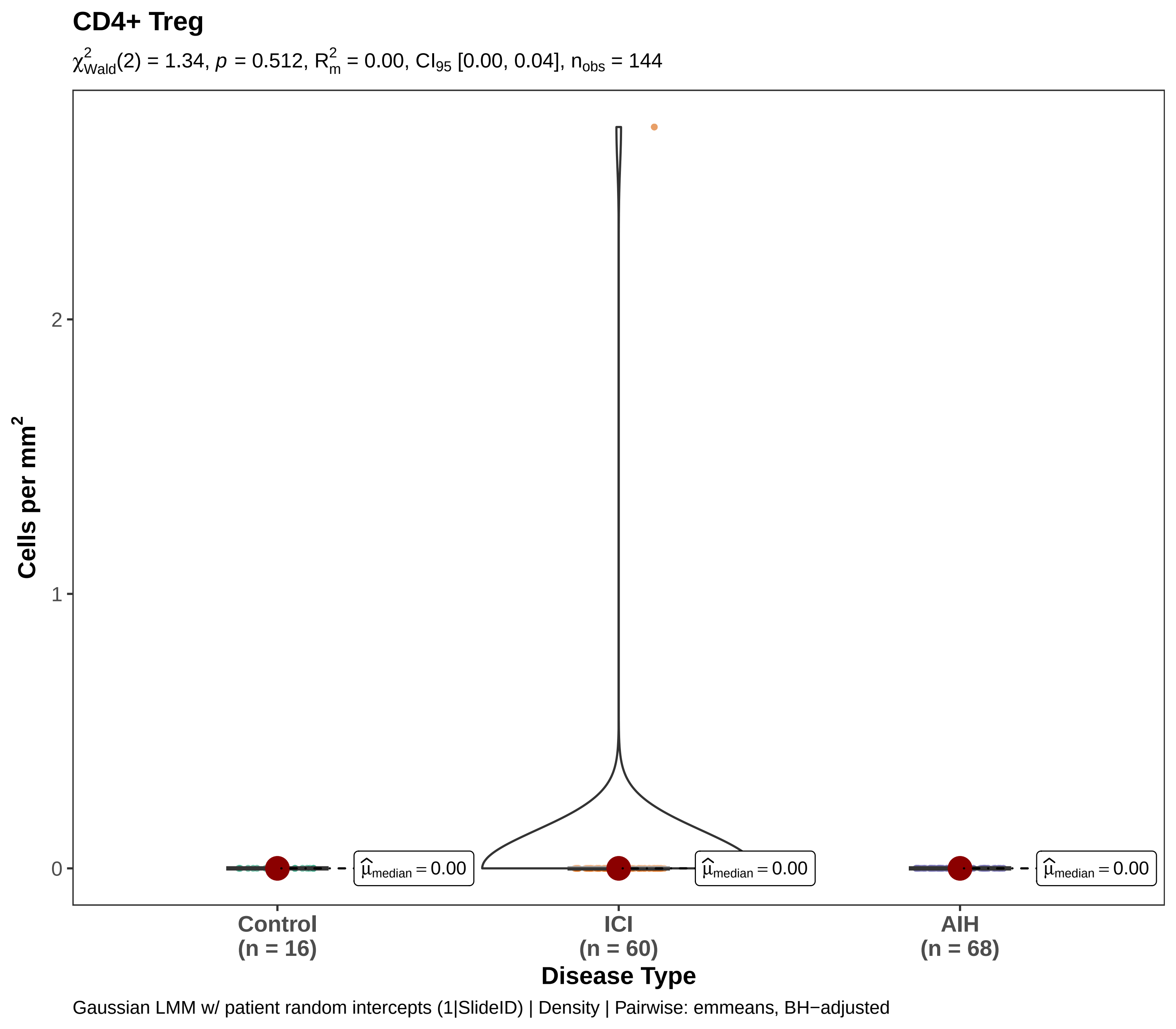

CD4+ Treg
$\chi^2_{Wald}(2) = 1.34$, $p = 0.512$, $R^2_m = 0.00$, $CI_{95}$ [0.00, 0.04], $n_{obs} = 144$
Cells per mm$^2$
0
1
2
$\widehat{\mu}_{median} = 0.00$
$\widehat{\mu}_{median} = 0.00$
$\widehat{\mu}_{median} = 0.00$
Control
(n = 16)
ICI
(n = 60)
AIH
(n = 68)
Disease Type
Gaussian LMM w/ patient random intercepts (1|SlideID) | Density | Pairwise: emmeans, BH−adjusted

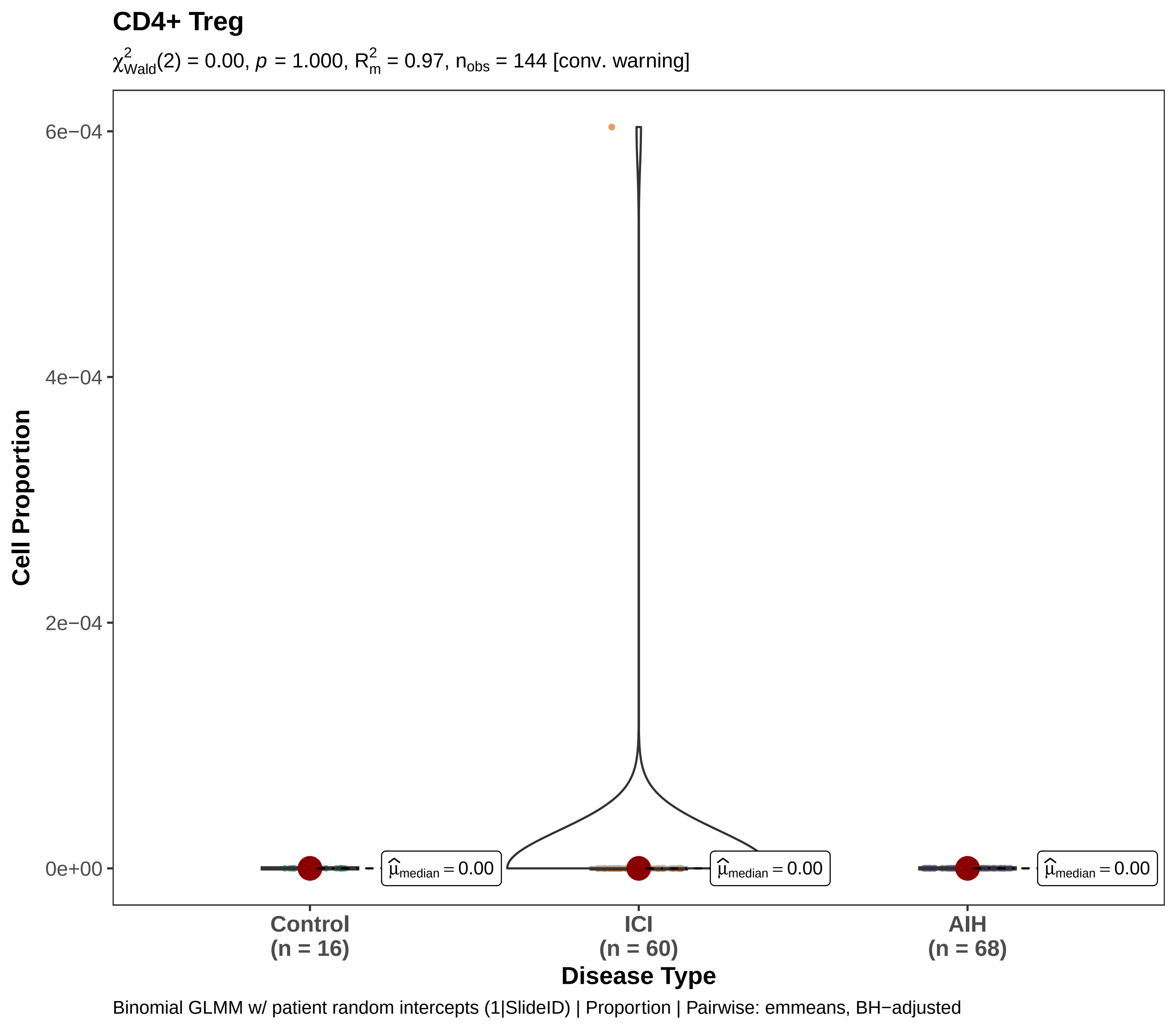
CD4+ Treg
$\chi^2_{Wald}(2) = 0.00$, $p = 1.000$, $R^2_m = 0.97$, $n_{obs} = 144$ [conv. warning]
Cell Proportion
6e−04
4e−04
2e−04
0e+00
$\widehat{\mu}_{median} = 0.00$
$\widehat{\mu}_{median} = 0.00$
$\widehat{\mu}_{median} = 0.00$
Control
(n = 16)
ICI
(n = 60)
AIH
(n = 68)
Disease Type
Binomial GLMM w/ patient random intercepts (1|SlideID) | Proportion | Pairwise: emmeans, BH−adjusted

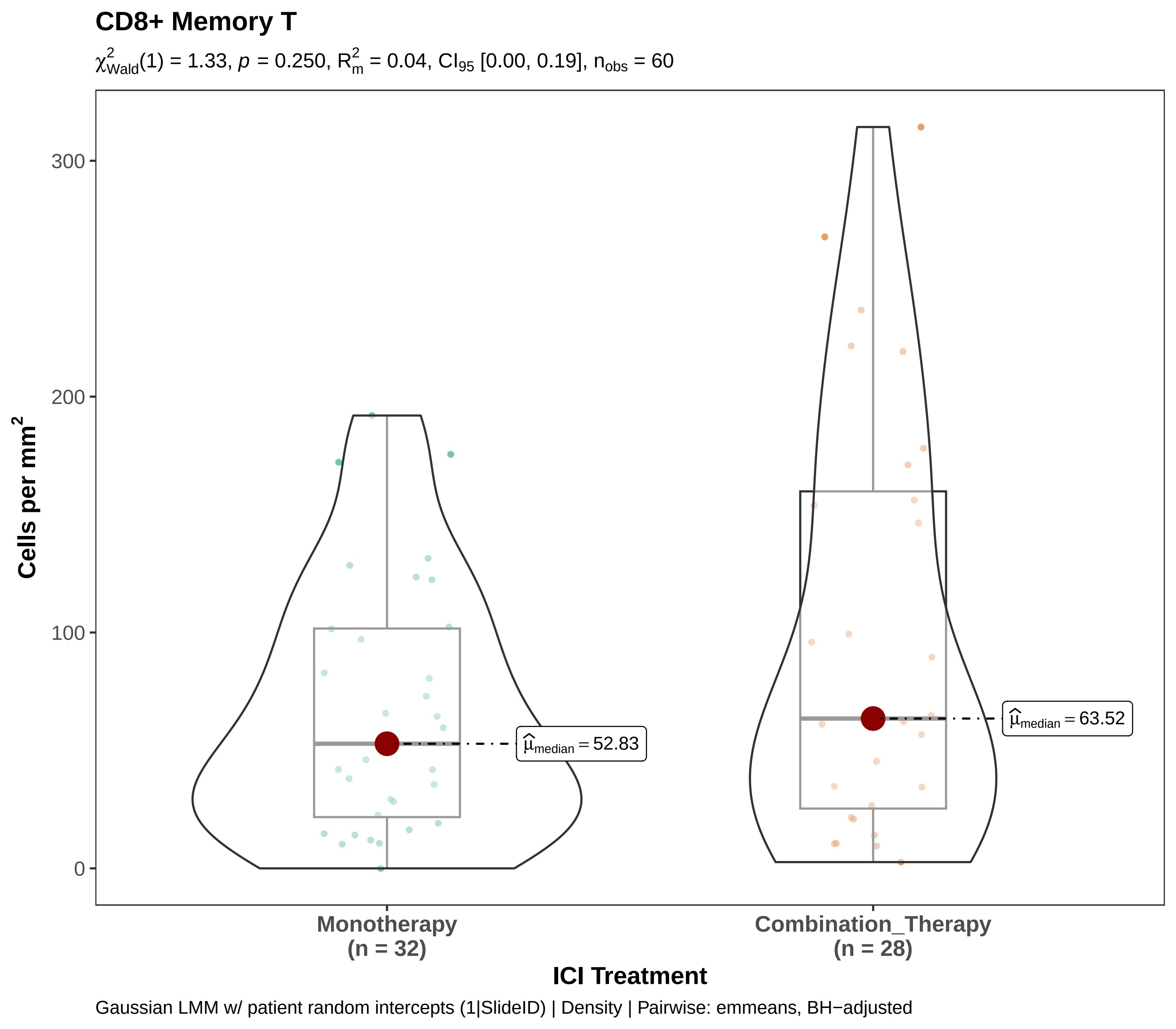
CD8+ Memory T
$\chi^2_{Wald}(1) = 1.33$, $p = 0.250$, $R^2_m = 0.04$, $CI_{95}$ [0.00, 0.19], $n_{obs} = 60$
Cells per mm$^2$
300
200
100
0
$\widehat{\mu}_{median} = 52.83$
$\widehat{\mu}_{median} = 63.52$
Monotherapy
(n = 32)
Combination_Therapy
(n = 28)
ICI Treatment
Gaussian LMM w/ patient random intercepts (1|SlideID) | Density | Pairwise: emmeans, BH−adjusted

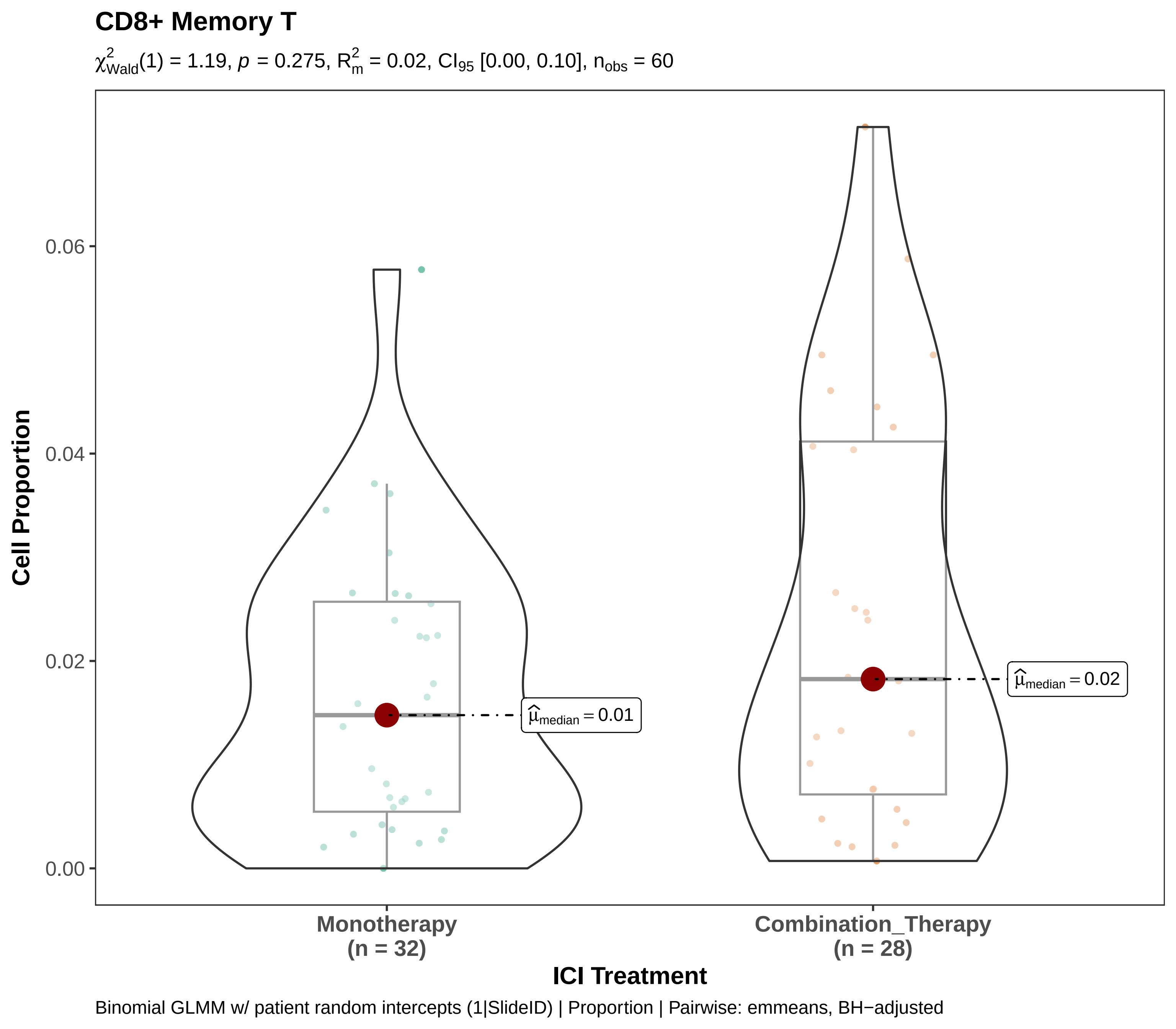

CD8+ Memory T
$\chi^2_{Wald}(1) = 1.19$, $p = 0.275$, $R^2_m = 0.02$, $CI_{95}$ [0.00, 0.10], $n_{obs} = 60$
Cell Proportion
0.06
0.04
0.02
0.00
$\hat{\mu}_{median} = 0.01$
$\hat{\mu}_{median} = 0.02$
Monotherapy
(n = 32)
Combination_Therapy
(n = 28)
ICI Treatment
Binomial GLMM w/ patient random intercepts (1|SlideID) | Proportion | Pairwise: emmeans, BH−adjusted

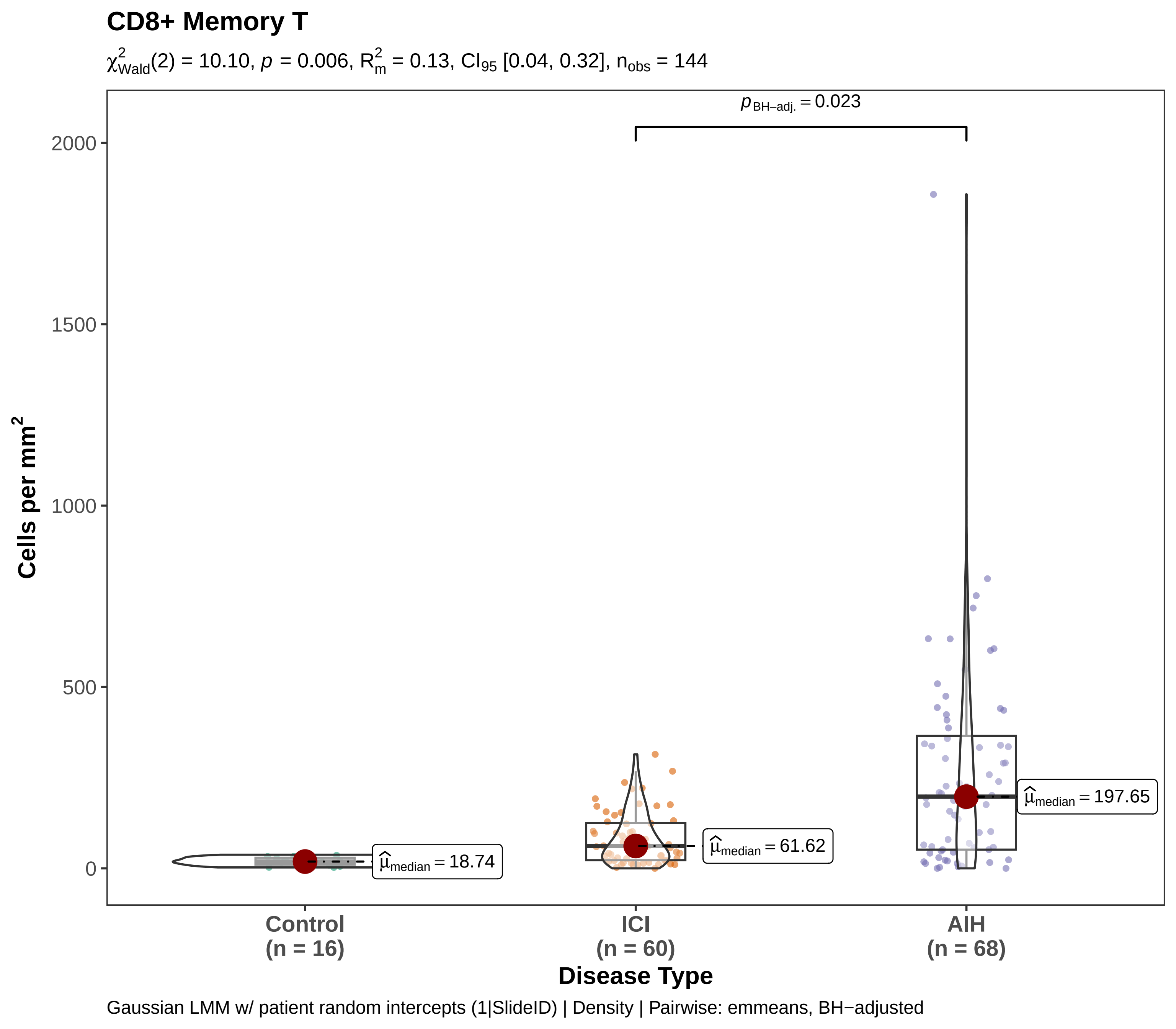

CD8+ Memory T
$\chi^2_{Wald}(2) = 10.10$, $p = 0.006$, $R^2_m = 0.13$, $CI_{95}$ [0.04, 0.32], $n_{obs} = 144$
$p_{BH-adj.} = 0.023$
Cells per mm²
2000
1500
1000
500
0
$\hat{\mu}_{median} = 18.74$
$\hat{\mu}_{median} = 61.62$
$\hat{\mu}_{median} = 197.65$
Control (n = 16)
ICI (n = 60)
AIH (n = 68)
Disease Type
Gaussian LMM w/ patient random intercepts (1|SlideID) | Density | Pairwise: emmeans, BH−adjusted

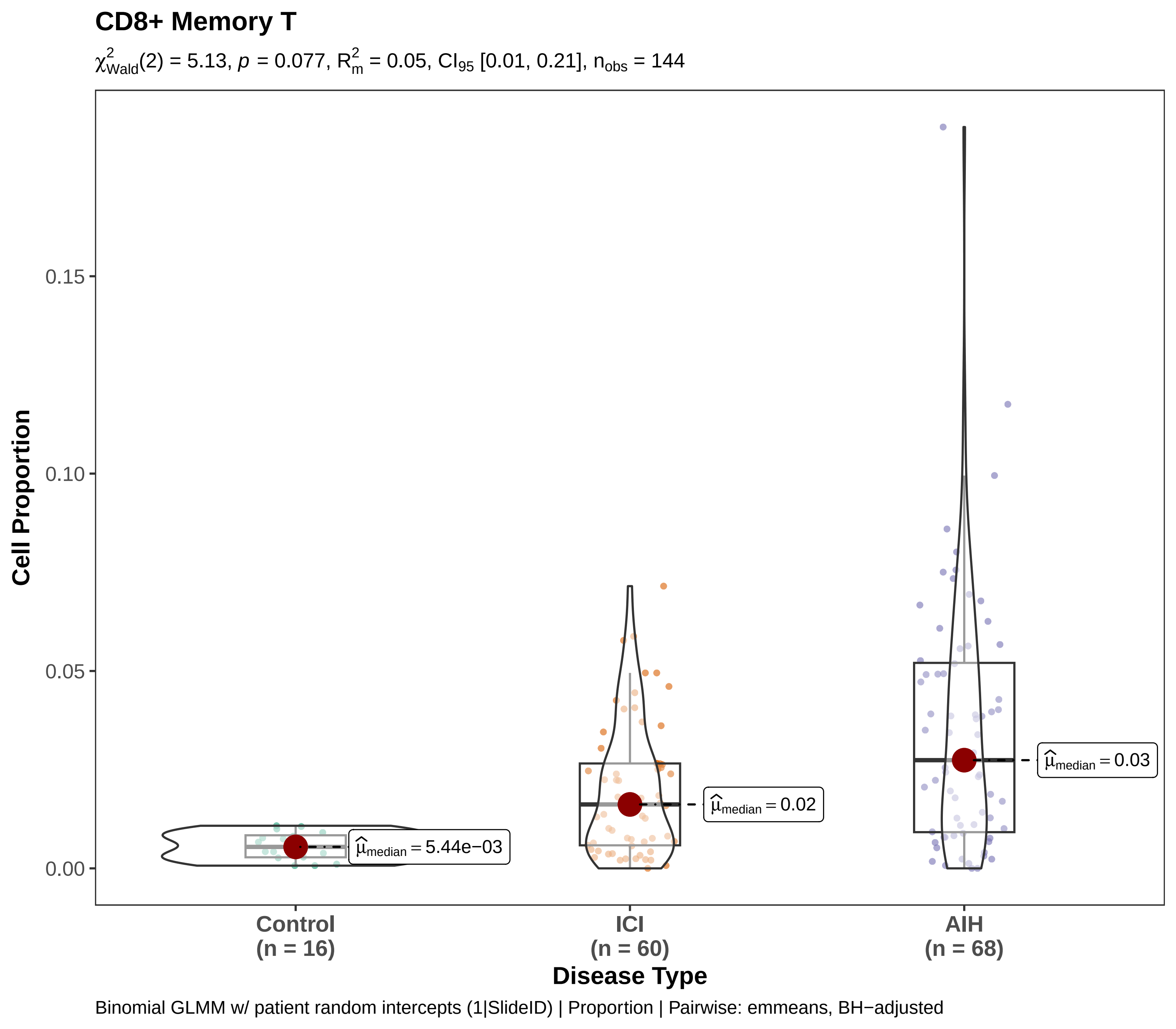
CD8+ Memory T
χ²Wald(2) = 5.13, p = 0.077, R²m = 0.05, CI95 [0.01, 0.21], nobs = 144
Cell Proportion
0.00
0.05
0.10
0.15
μ̂median = 5.44e−03
μ̂median = 0.02
μ̂median = 0.03
Control
(n = 16)
ICI
(n = 60)
AIH
(n = 68)
Disease Type
Binomial GLMM w/ patient random intercepts (1|SlideID) | Proportion | Pairwise: emmeans, BH−adjusted

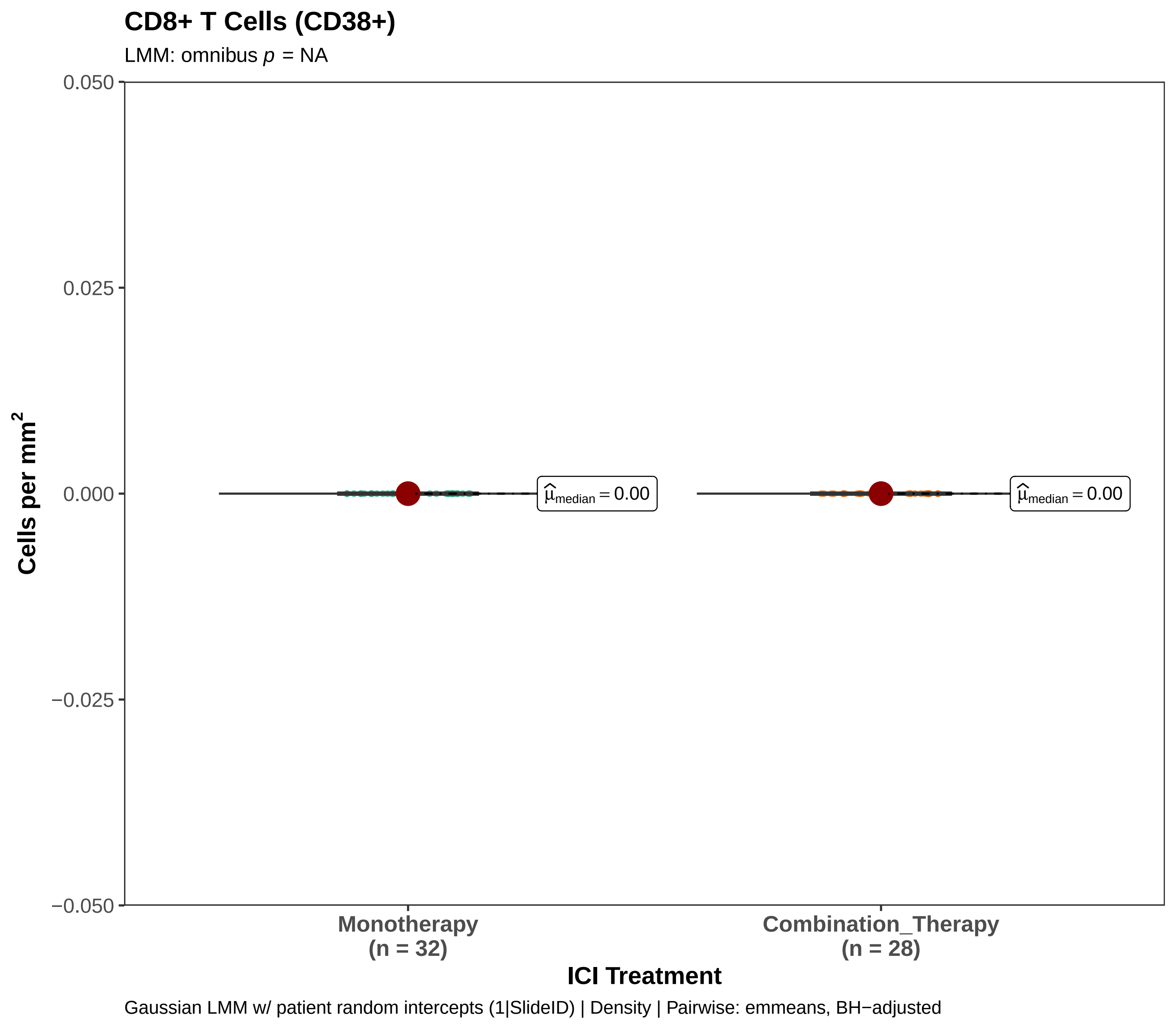
CD8+ T Cells (CD38+)
LMM: omnibus p = NA
0.050
0.025
0.000
−0.025
−0.050
Cells per mm2
$\widehat{\mu}_{median} = 0.00$
$\widehat{\mu}_{median} = 0.00$
Monotherapy
(n = 32)
Combination_Therapy
(n = 28)
ICI Treatment
Gaussian LMM w/ patient random intercepts (1|SlideID) | Density | Pairwise: emmeans, BH−adjusted

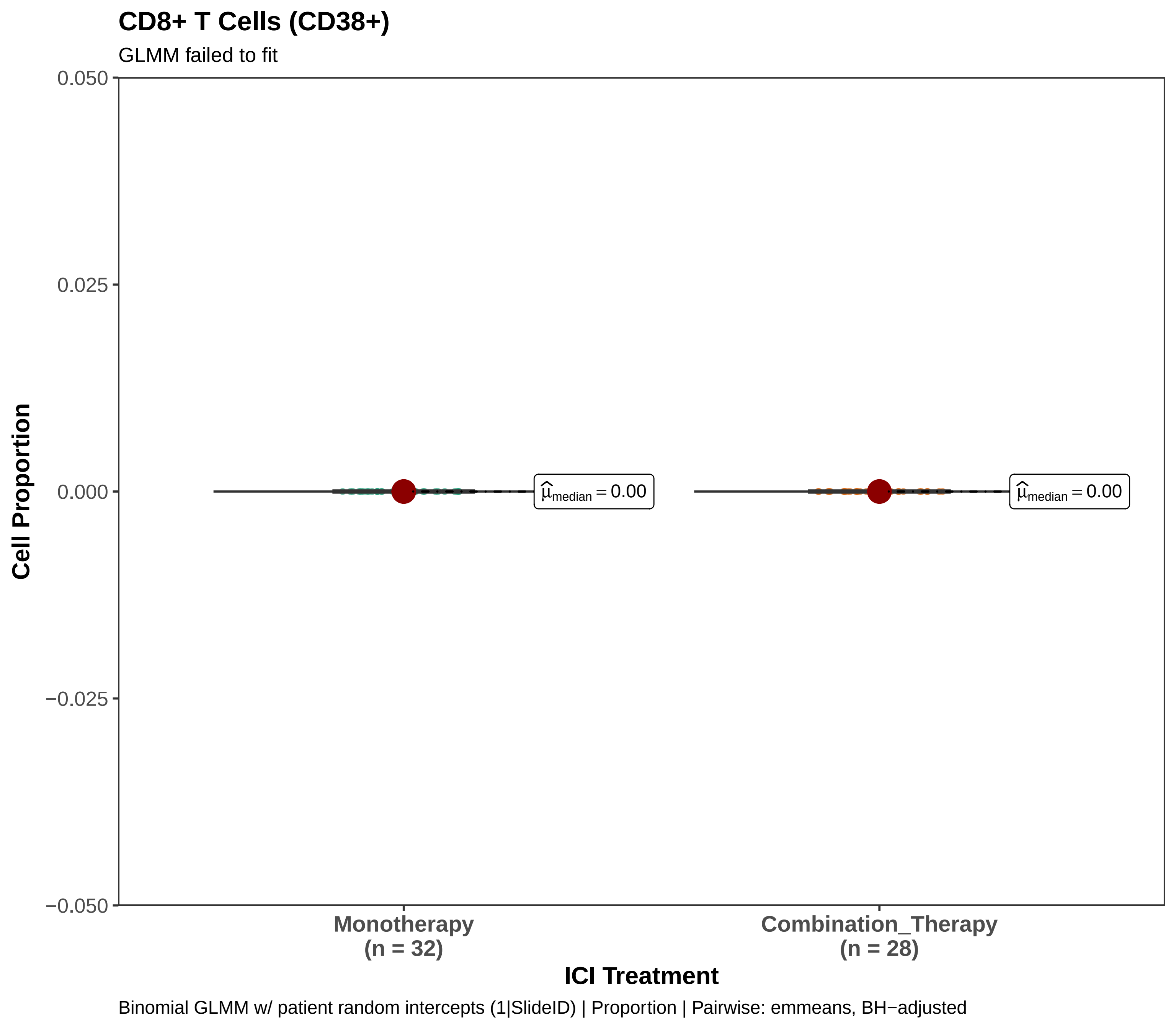
CD8+ T Cells (CD38+)
GLMM failed to fit
0.050
0.025
0.000
−0.025
−0.050
Cell Proportion
$\hat{\mu}_{median} = 0.00$
$\hat{\mu}_{median} = 0.00$
Monotherapy
(n = 32)
Combination_Therapy
(n = 28)
ICI Treatment
Binomial GLMM w/ patient random intercepts (1|SlideID) | Proportion | Pairwise: emmeans, BH−adjusted

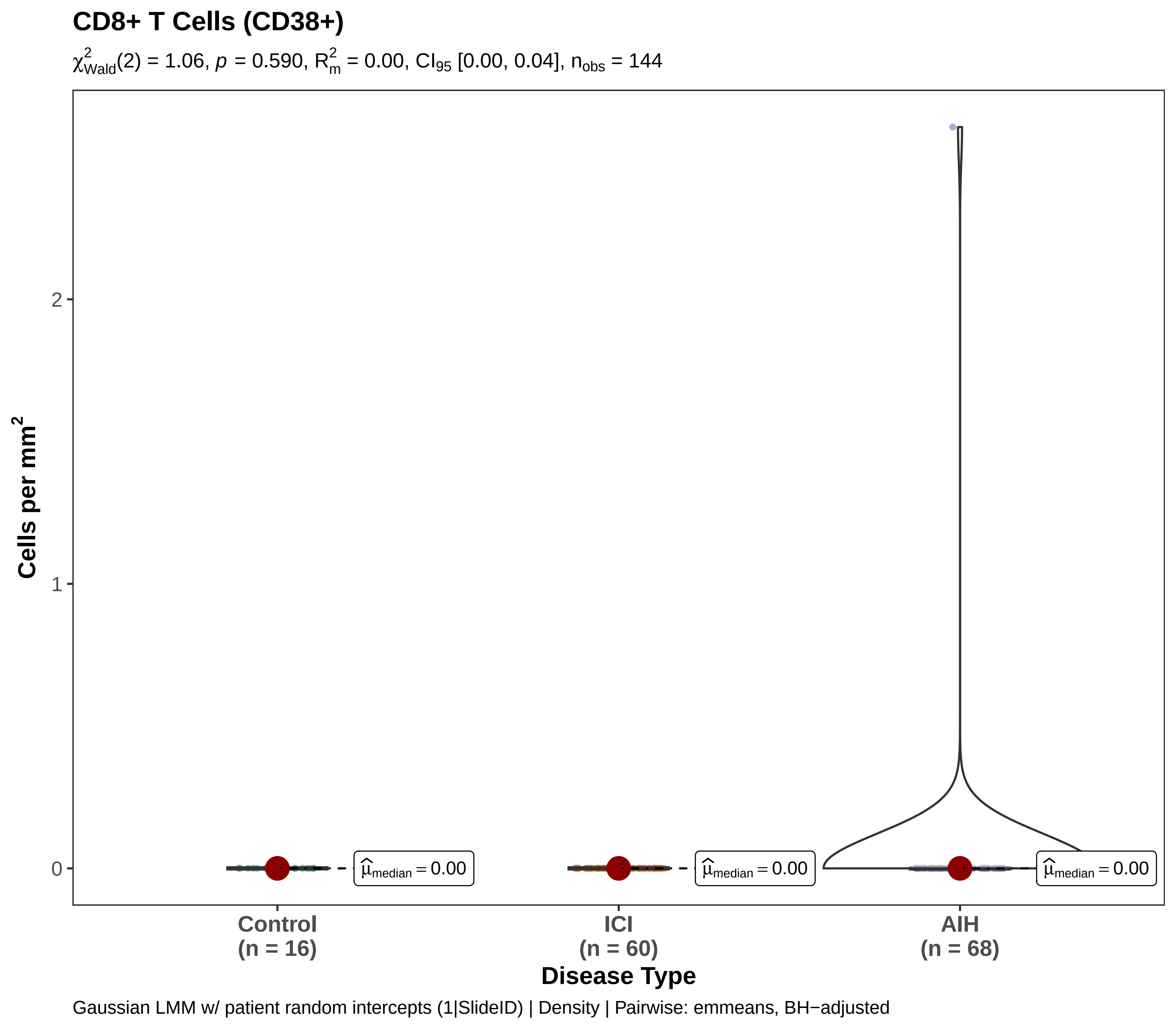
CD8+ T Cells (CD38+)
$\chi^2_{Wald}(2) = 1.06$, $p = 0.590$, $R^2_m = 0.00$, $CI_{95}$ [0.00, 0.04], $n_{obs} = 144$
Cells per mm$^2$
2
1
0
$\hat{\mu}_{median} = 0.00$
$\hat{\mu}_{median} = 0.00$
$\hat{\mu}_{median} = 0.00$
Control
(n = 16)
ICI
(n = 60)
AIH
(n = 68)
Disease Type
Gaussian LMM w/ patient random intercepts (1|SlideID) | Density | Pairwise: emmeans, BH−adjusted

#### CD8+ T Cells (CD38+)

$\chi^2_{Wald}(2) = 0.00$, $p = 1.000$, $R^2_m = 0.97$, $n_{obs} = 144$ [conv. warning]

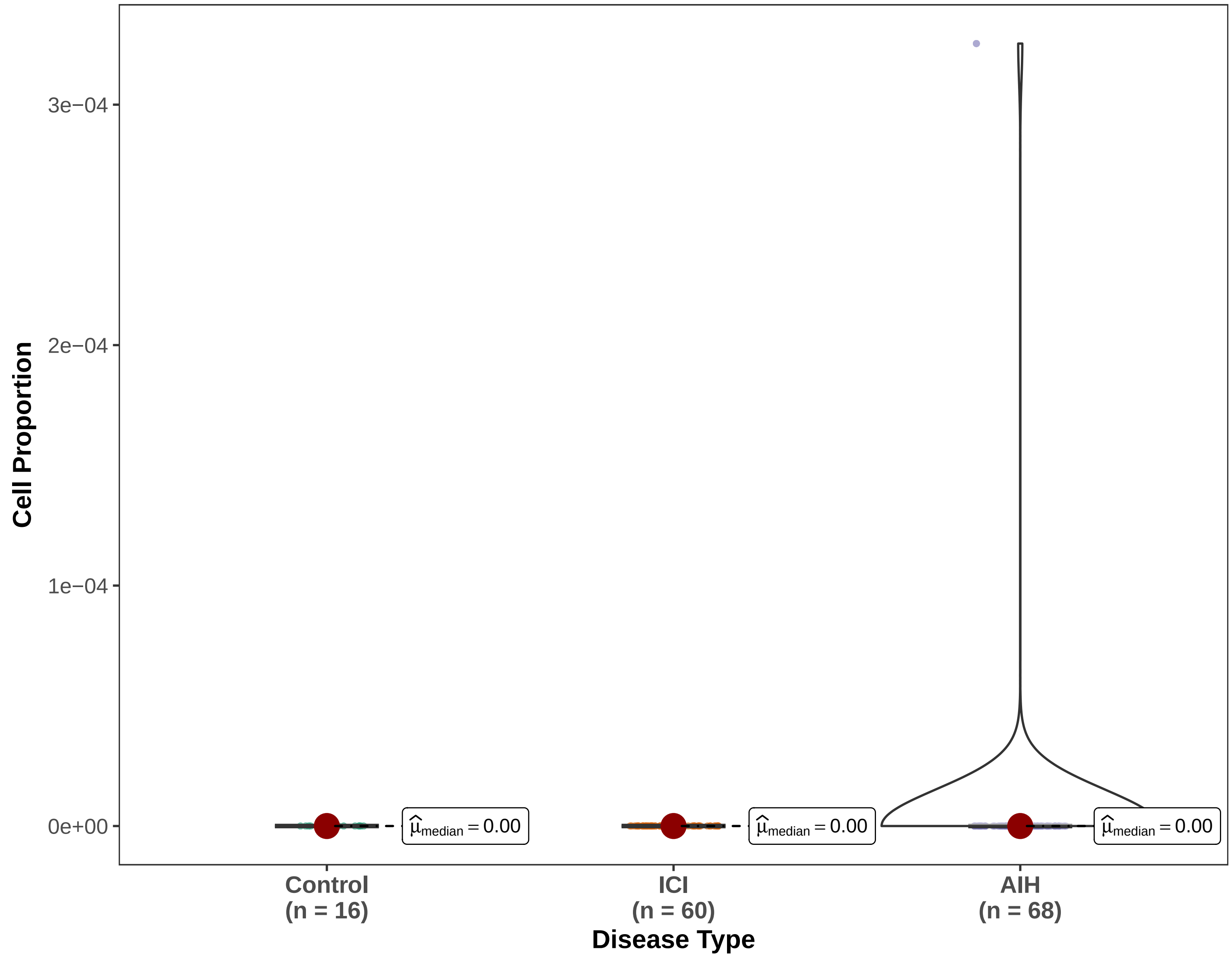


Binomial GLMM w/ patient random intercepts (1|SlideID) | Proportion | Pairwise: emmeans, BH−adjusted

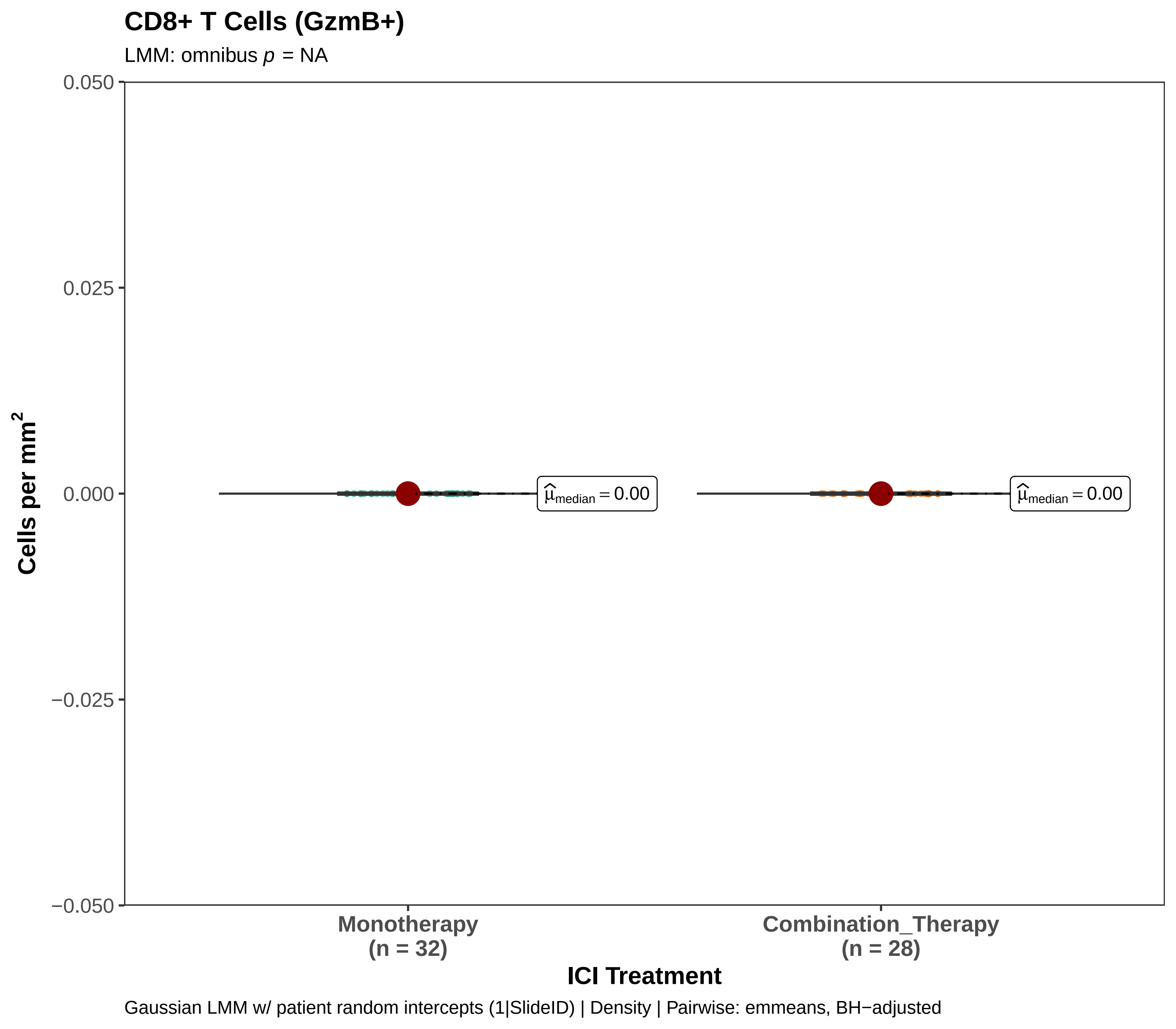
CD8+ T Cells (GzmB+)
LMM: omnibus p = NA
0.050
0.025
0.000
−0.025
−0.050
Cells per mm²
$\widehat{\mu}_{median} = 0.00$
$\widehat{\mu}_{median} = 0.00$
Monotherapy
(n = 32)
Combination_Therapy
(n = 28)
ICI Treatment
Gaussian LMM w/ patient random intercepts (1|SlideID) | Density | Pairwise: emmeans, BH−adjusted

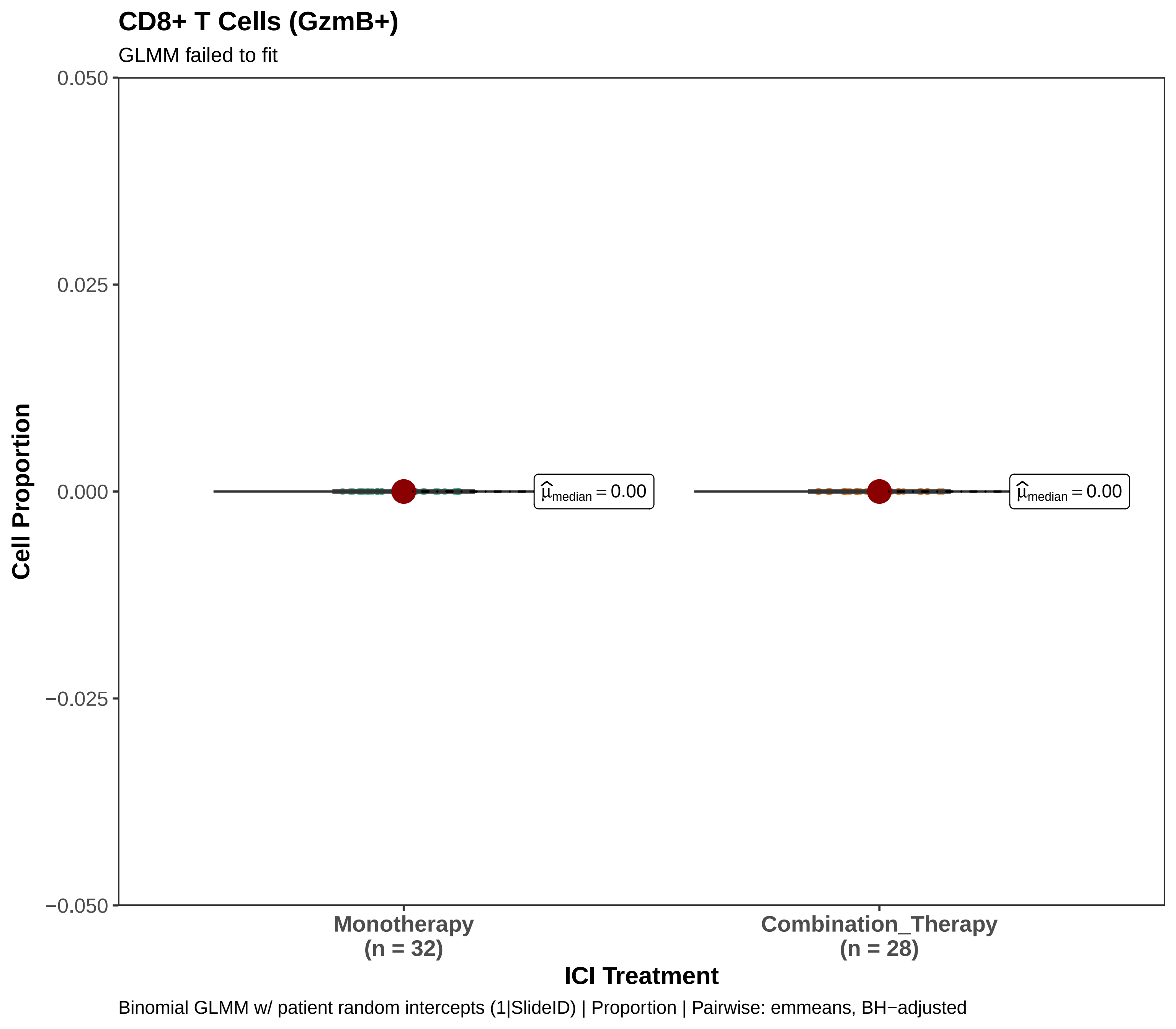
CD8+ T Cells (GzmB+)
GLMM failed to fit
0.050
0.025
0.000
−0.025
−0.050
Cell Proportion
$\widehat{\mu}_{median} = 0.00$
$\widehat{\mu}_{median} = 0.00$
Monotherapy
(n = 32)
Combination_Therapy
(n = 28)
ICI Treatment
Binomial GLMM w/ patient random intercepts (1|SlideID) | Proportion | Pairwise: emmeans, BH−adjusted

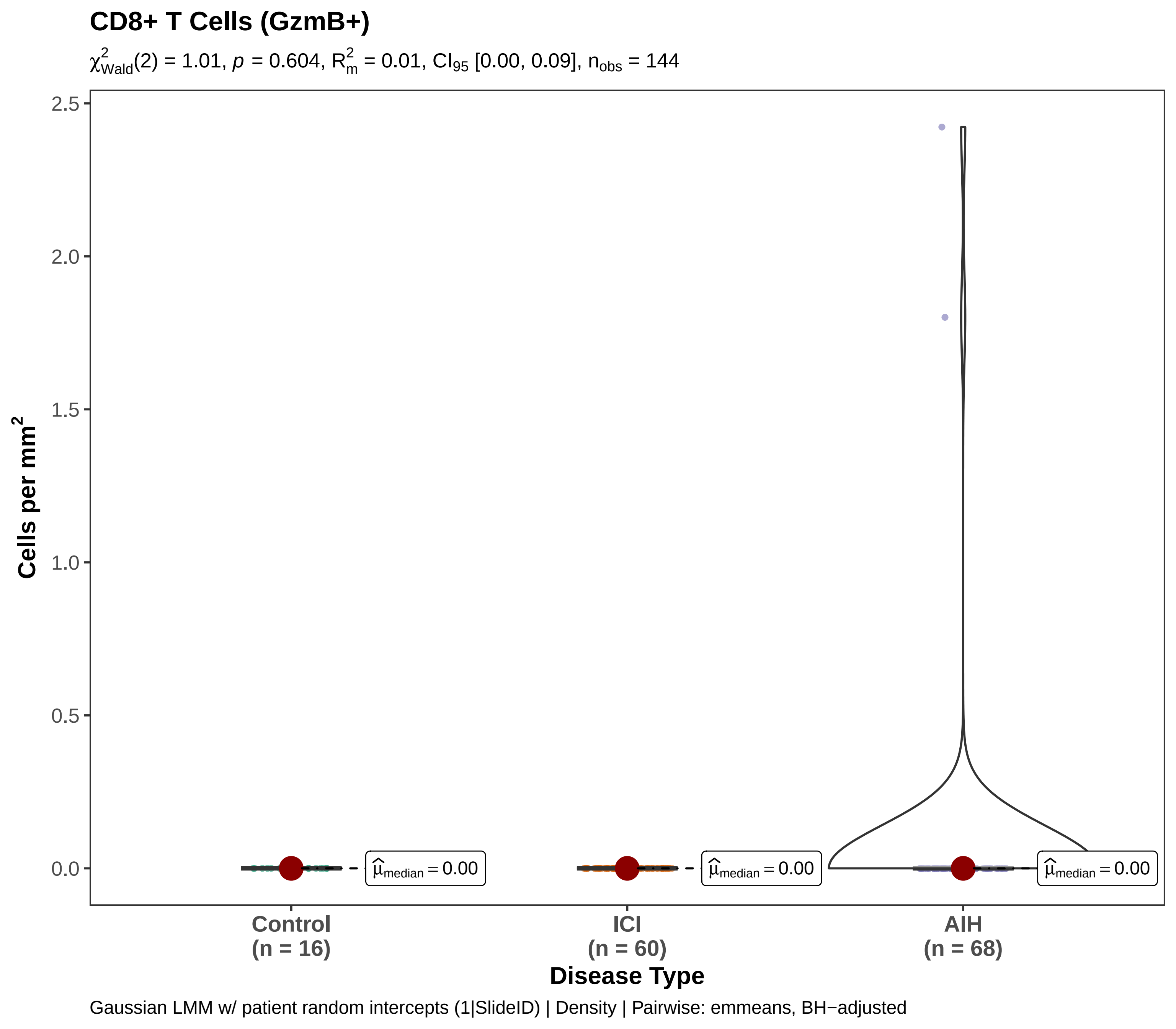

CD8+ T Cells (GzmB+)
$\chi^2_{Wald}(2) = 1.01$, $p = 0.604$, $R^2_m = 0.01$, $CI_{95}$ [0.00, 0.09], $n_{obs} = 144$
Cells per mm$^2$
2.5
2.0
1.5
1.0
0.5
0.0
$\widehat{\mu}_{median} = 0.00$
$\widehat{\mu}_{median} = 0.00$
$\widehat{\mu}_{median} = 0.00$
Control
(n = 16)
ICI
(n = 60)
AIH
(n = 68)
Disease Type
Gaussian LMM w/ patient random intercepts (1|SlideID) | Density | Pairwise: emmeans, BH−adjusted

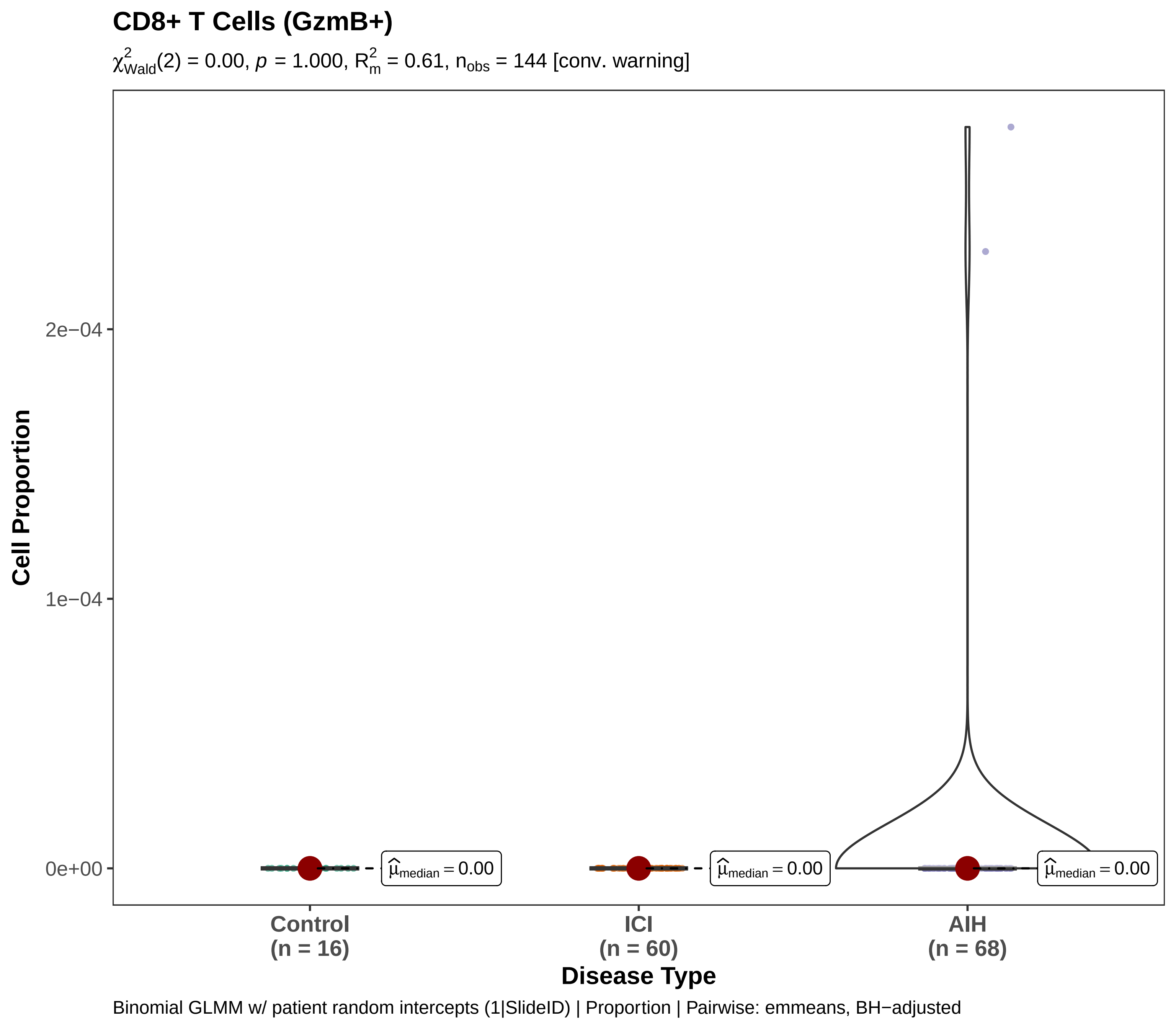

CD8+ T Cells (GzmB+)
$\chi^2_{Wald}(2) = 0.00$, $p = 1.000$, $R^2_m = 0.61$, $n_{obs} = 144$ [conv. warning]
Cell Proportion
2e−04
1e−04
0e+00
$\hat{\mu}_{median} = 0.00$
$\hat{\mu}_{median} = 0.00$
$\hat{\mu}_{median} = 0.00$
Control
(n = 16)
ICI
(n = 60)
AIH
(n = 68)
Disease Type
Binomial GLMM w/ patient random intercepts (1|SlideID) | Proportion | Pairwise: emmeans, BH−adjusted

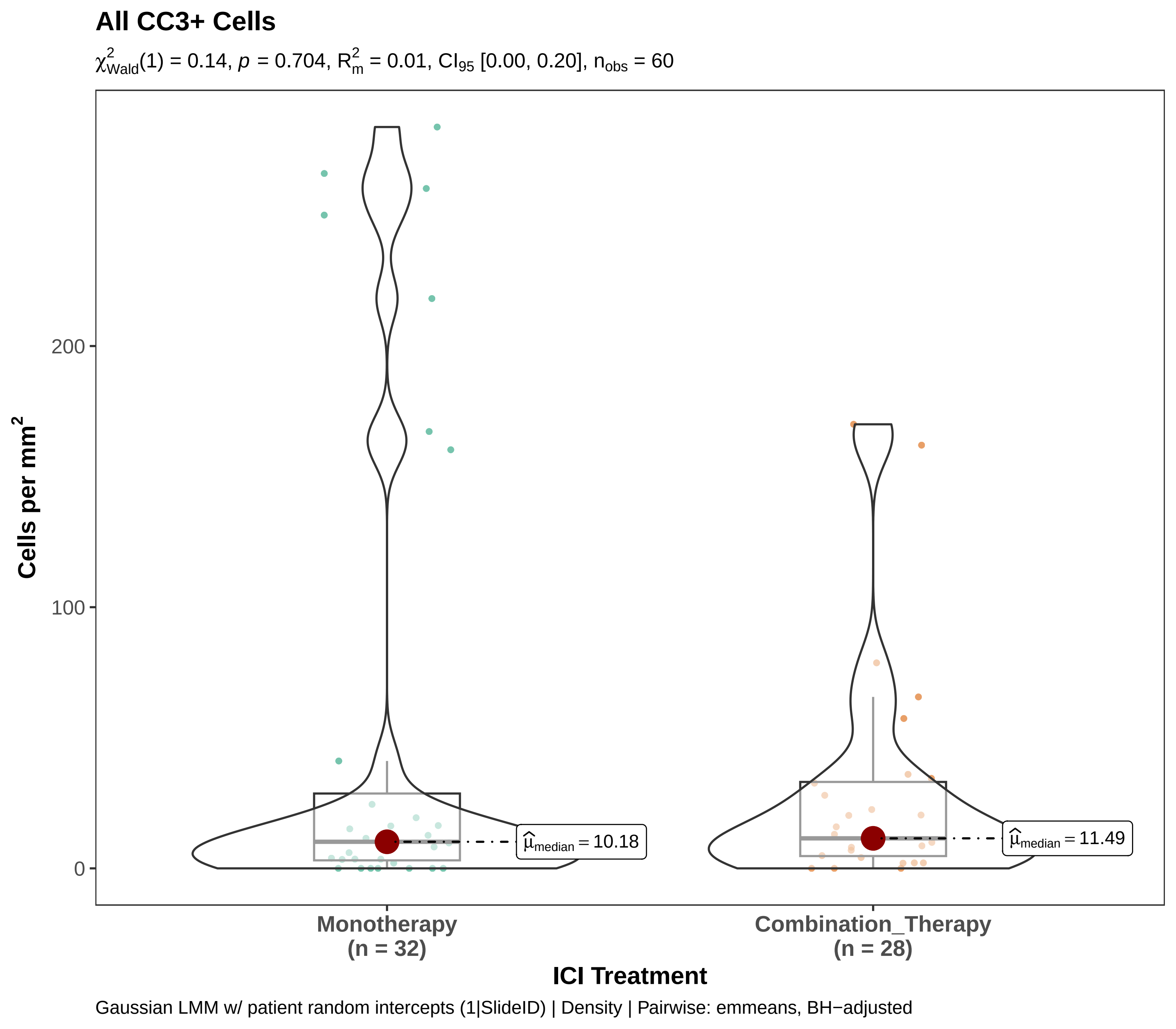
All CC3+ Cells
$\chi^2_{Wald}(1) = 0.14$, $p = 0.704$, $R^2_m = 0.01$, $CI_{95}$ [0.00, 0.20], $n_{obs} = 60$
Cells per mm$^2$
200
100
0
$\hat{\mu}_{median} = 10.18$
$\hat{\mu}_{median} = 11.49$
Monotherapy
(n = 32)
Combination_Therapy
(n = 28)
ICI Treatment
Gaussian LMM w/ patient random intercepts (1|SlideID) | Density | Pairwise: emmeans, BH−adjusted

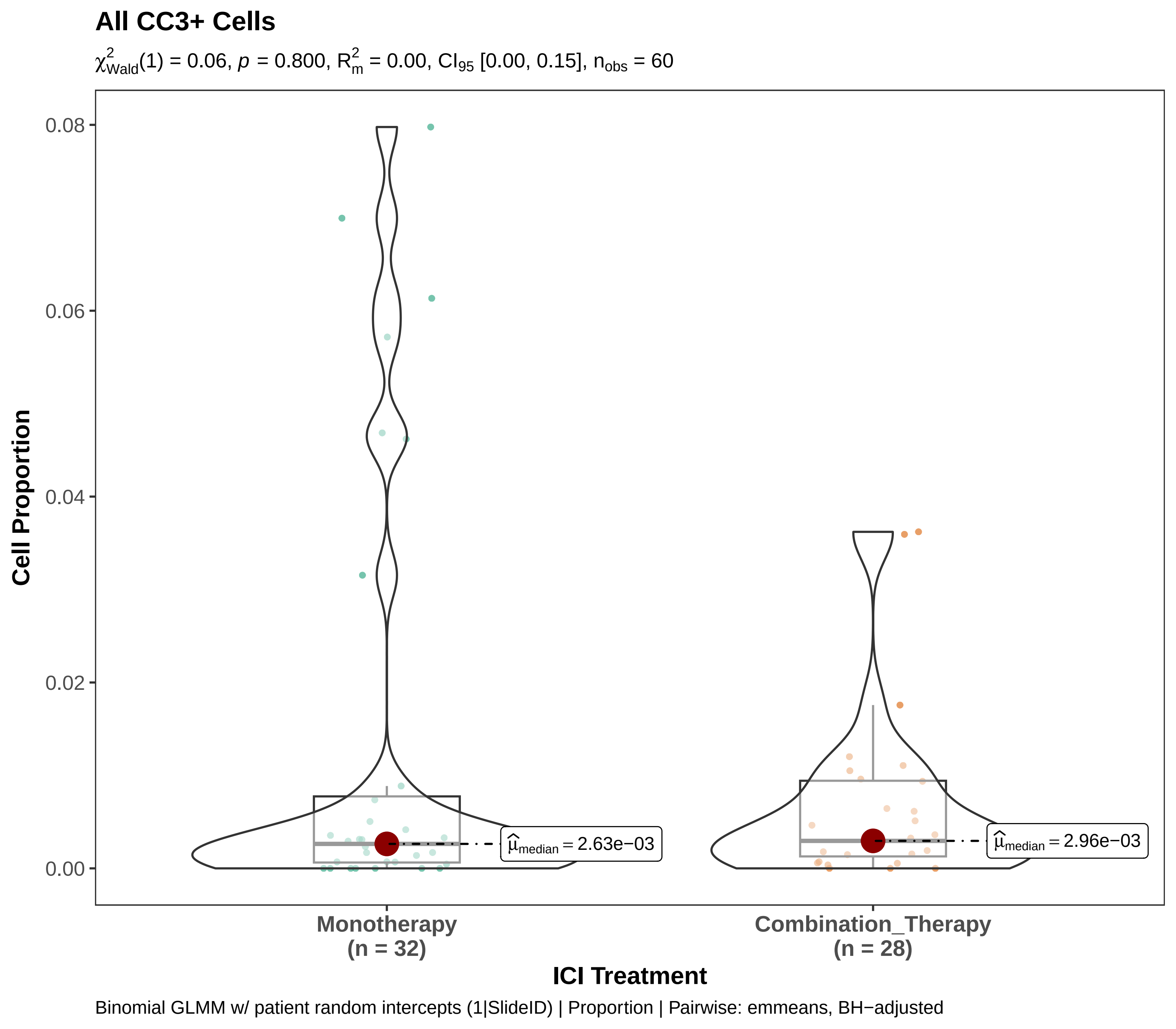
All CC3+ Cells
$\chi^2_{Wald}(1) = 0.06$, $p = 0.800$, $R^2_m = 0.00$, $CI_{95}$ [0.00, 0.15], $n_{obs} = 60$
Cell Proportion
0.08
0.06
0.04
0.02
0.00
$\widehat{\mu}_{median} = 2.63e{-}03$
$\widehat{\mu}_{median} = 2.96e{-}03$
Monotherapy
(n = 32)
Combination_Therapy
(n = 28)
ICI Treatment
Binomial GLMM w/ patient random intercepts (1|SlideID) | Proportion | Pairwise: emmeans, BH−adjusted

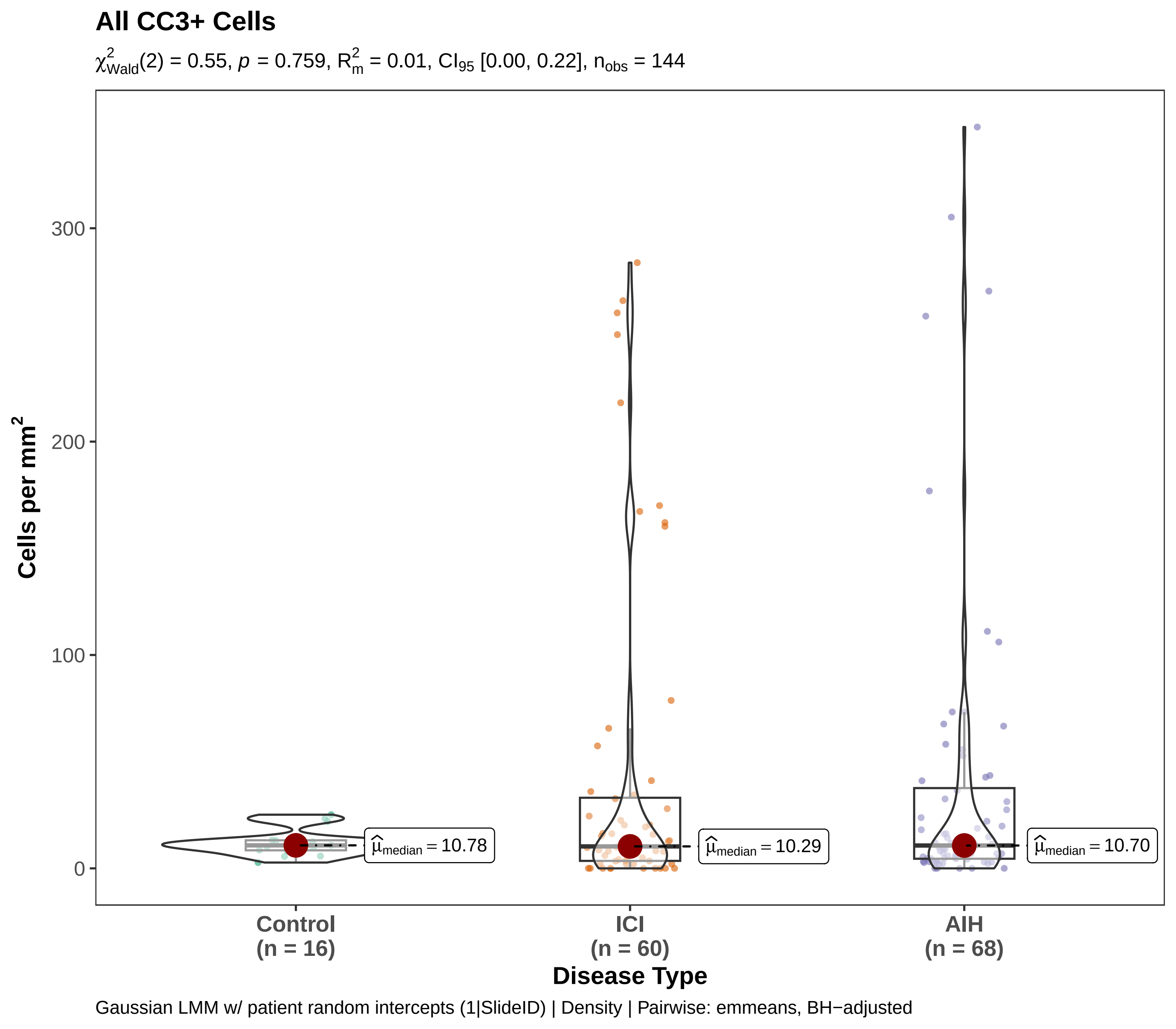

All CC3+ Cells
$\chi^2_{Wald}(2) = 0.55$, $p = 0.759$, $R^2_m = 0.01$, $CI_{95}$ [0.00, 0.22], $n_{obs} = 144$
Cells per mm²
300
200
100
0
$\hat{\mu}_{median} = 10.78$
$\hat{\mu}_{median} = 10.29$
$\hat{\mu}_{median} = 10.70$
Control
(n = 16)
ICI
(n = 60)
AIH
(n = 68)
Disease Type
Gaussian LMM w/ patient random intercepts (1|SlideID) | Density | Pairwise: emmeans, BH−adjusted

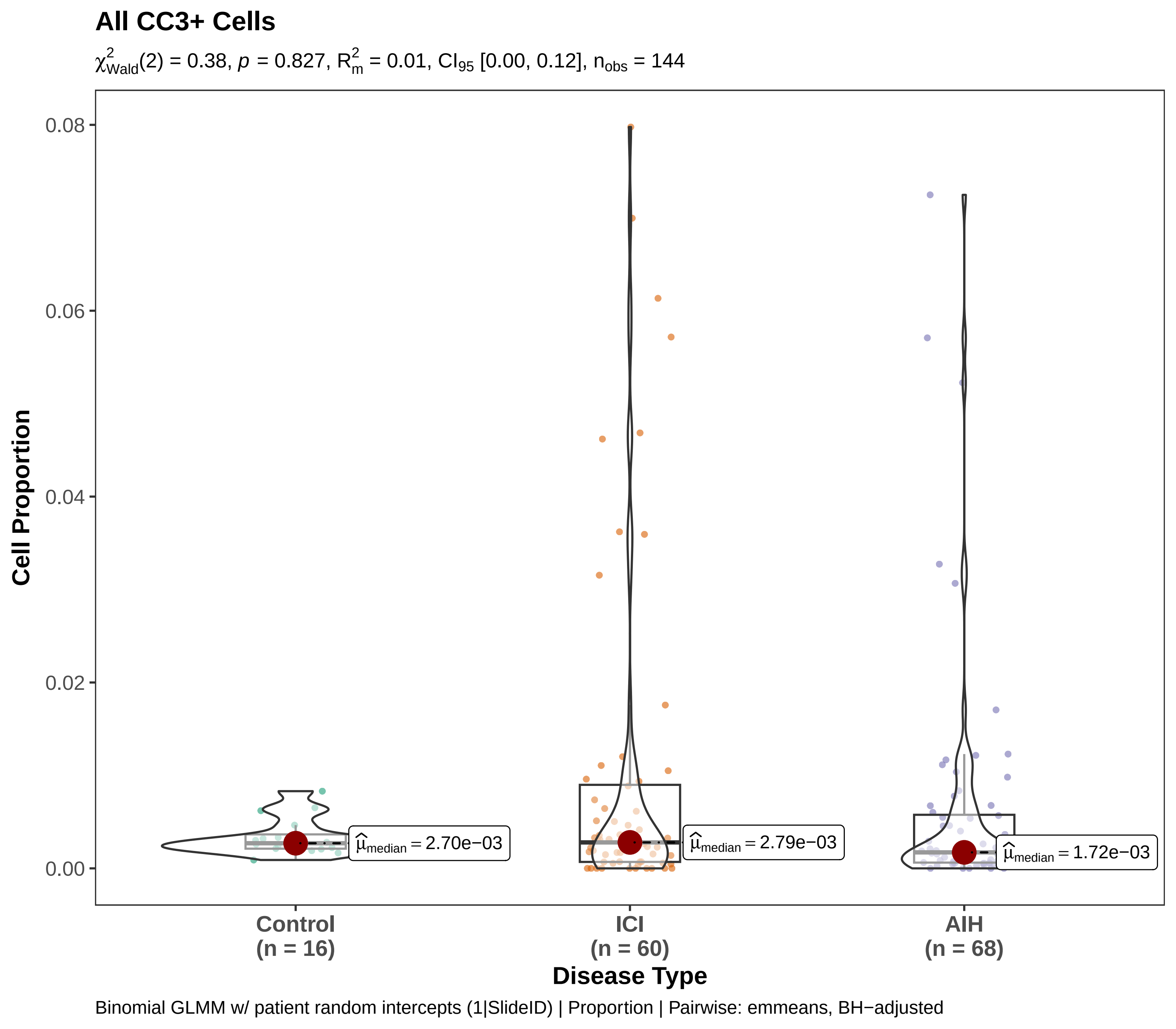
All CC3+ Cells
$\chi^2_{Wald}(2) = 0.38$, $p = 0.827$, $R^2_m = 0.01$, $CI_{95}$ [0.00, 0.12], $n_{obs} = 144$
Cell Proportion
0.08
0.06
0.04
0.02
0.00
$\hat{\mu}_{median} = 2.70e{-}03$
$\hat{\mu}_{median} = 2.79e{-}03$
$\hat{\mu}_{median} = 1.72e{-}03$
Control
(n = 16)
ICI
(n = 60)
AIH
(n = 68)
Disease Type
Binomial GLMM w/ patient random intercepts (1|SlideID) | Proportion | Pairwise: emmeans, BH−adjusted

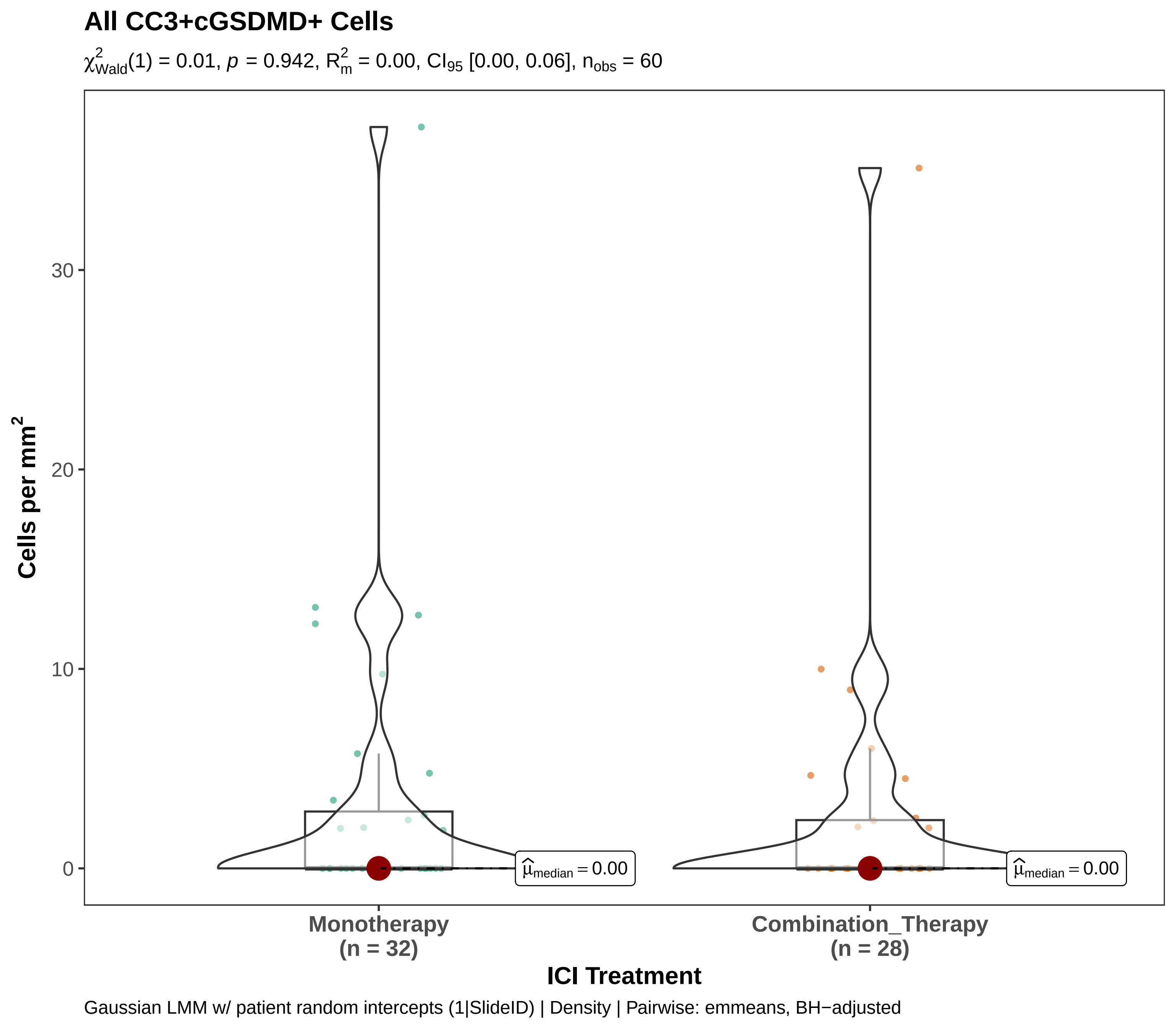
All CC3+cGSDMD+ Cells
$\chi^2_{Wald}(1) = 0.01$, $p = 0.942$, $R^2_m = 0.00$, $CI_{95}$ [0.00, 0.06], $n_{obs} = 60$
Cells per mm$^2$
30
20
10
0
$\widehat{\mu}_{median} = 0.00$
$\widehat{\mu}_{median} = 0.00$
Monotherapy
(n = 32)
Combination_Therapy
(n = 28)
ICI Treatment
Gaussian LMM w/ patient random intercepts (1|SlideID) | Density | Pairwise: emmeans, BH−adjusted

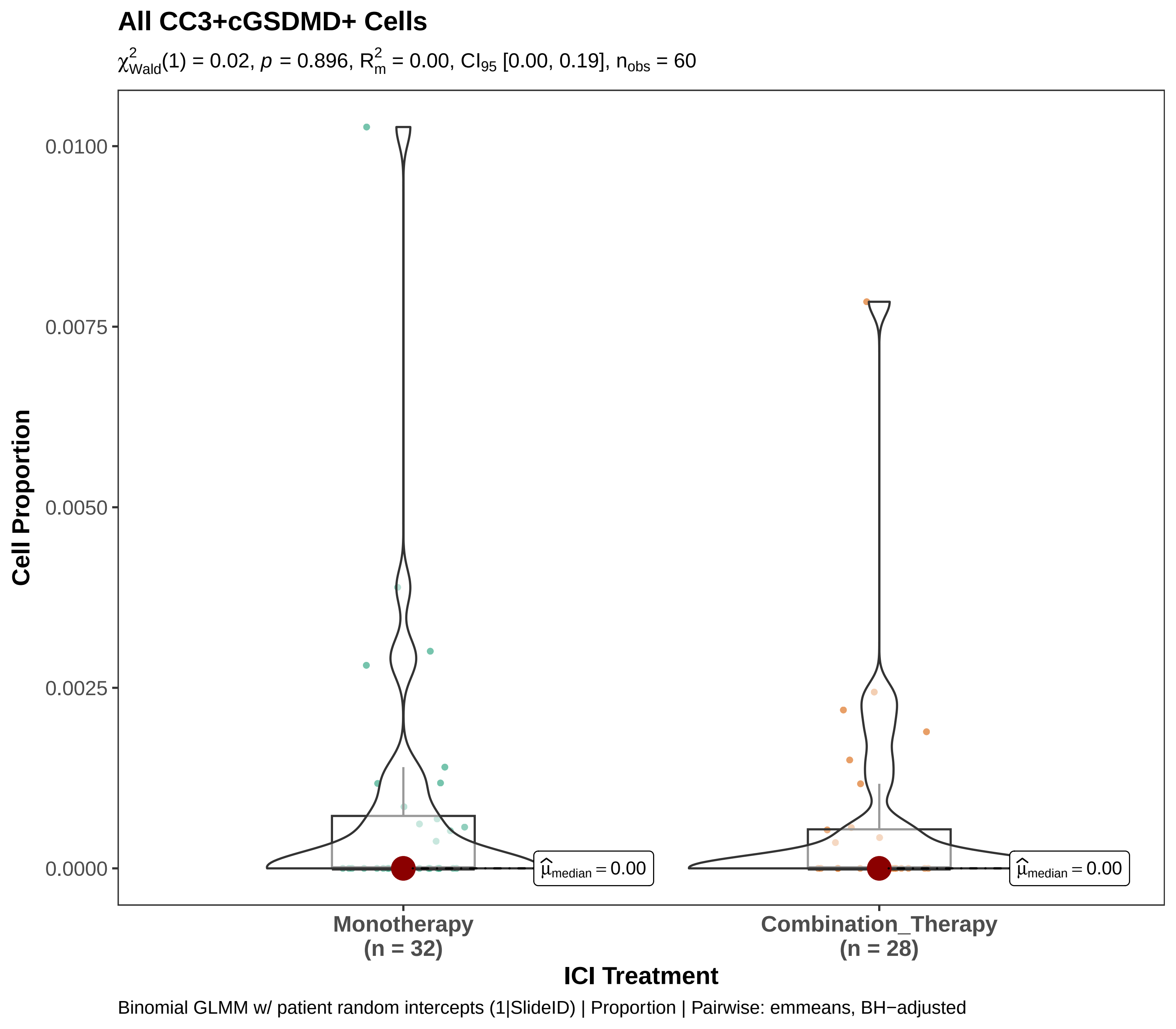
All CC3+cGSDMD+ Cells
$\chi^2_{Wald}(1) = 0.02$, $p = 0.896$, $R^2_m = 0.00$, $CI_{95}$ [0.00, 0.19], $n_{obs} = 60$
Cell Proportion
0.0100
0.0075
0.0050
0.0025
0.0000
$\hat{\mu}_{median} = 0.00$
$\hat{\mu}_{median} = 0.00$
Monotherapy
(n = 32)
Combination_Therapy
(n = 28)
ICI Treatment
Binomial GLMM w/ patient random intercepts (1|SlideID) | Proportion | Pairwise: emmeans, BH−adjusted

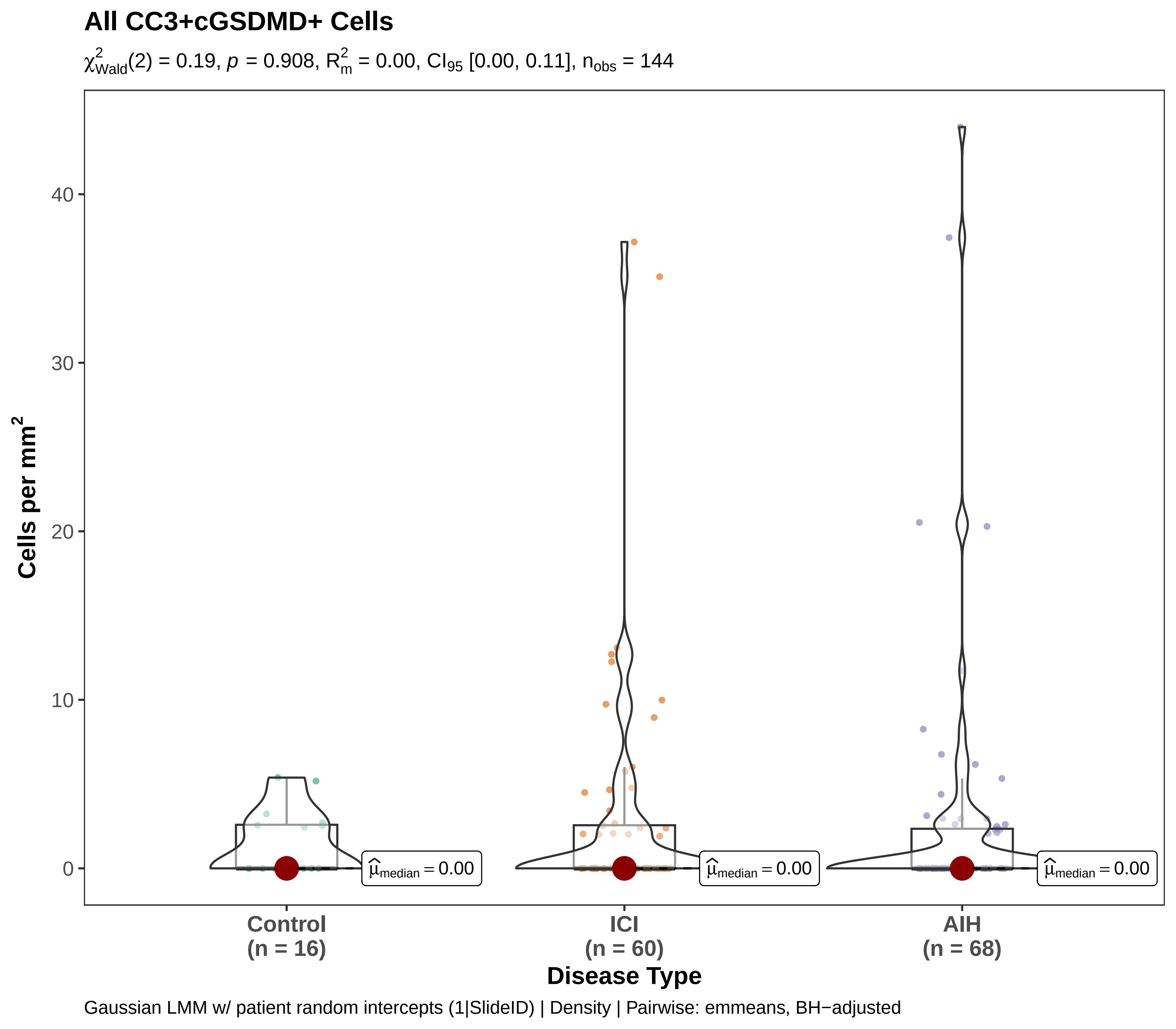
All CC3+cGSDMD+ Cells
$\chi^2_{Wald}(2) = 0.19$, $p = 0.908$, $R^2_m = 0.00$, $CI_{95}$ [0.00, 0.11], $n_{obs} = 144$
Cells per mm$^2$
40
30
20
10
0
$\hat{\mu}_{median} = 0.00$
$\hat{\mu}_{median} = 0.00$
$\hat{\mu}_{median} = 0.00$
Control
(n = 16)
ICI
(n = 60)
AIH
(n = 68)
Disease Type
Gaussian LMM w/ patient random intercepts (1|SlideID) | Density | Pairwise: emmeans, BH−adjusted

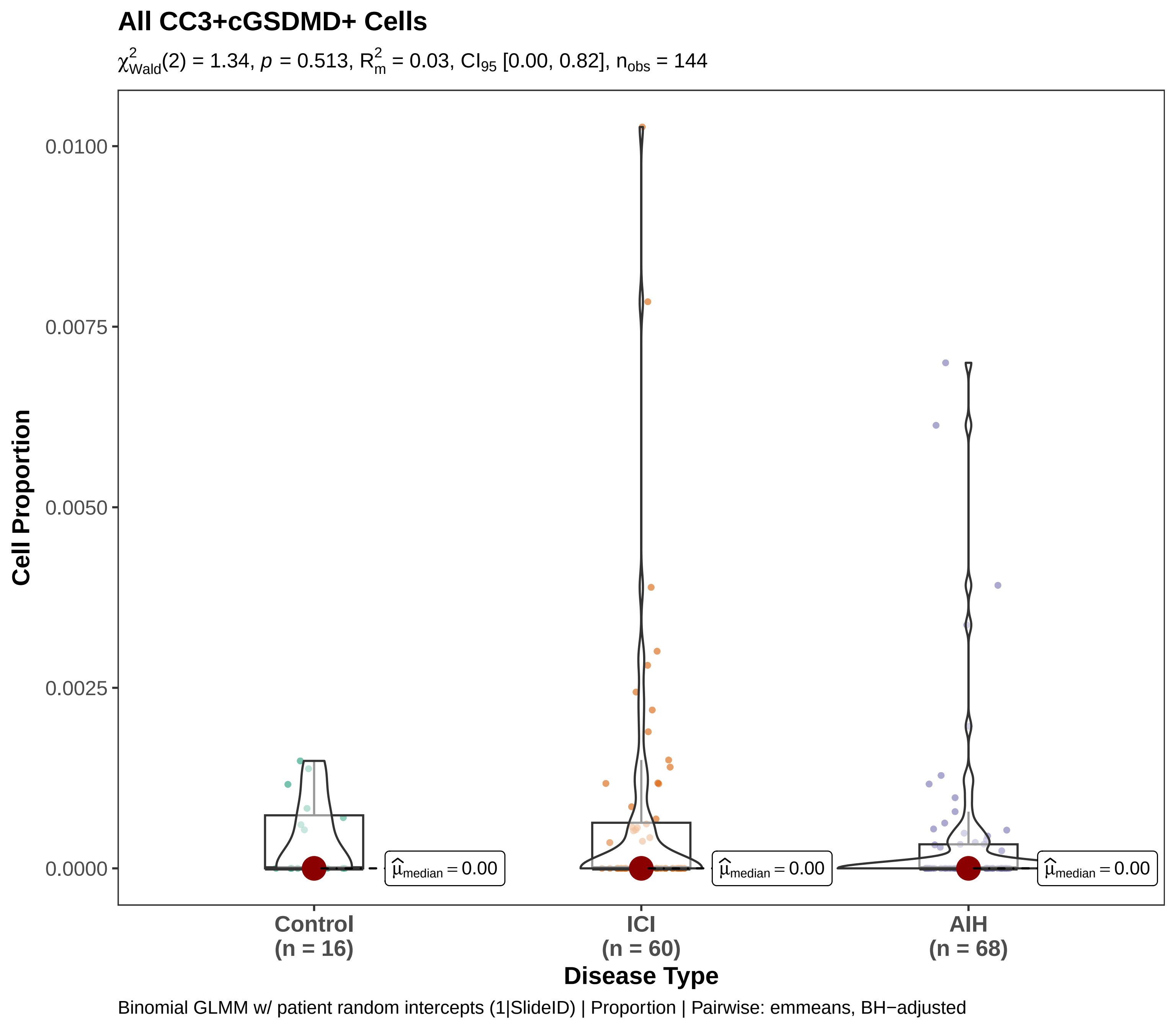

All CC3+cGSDMD+ Cells
$\chi^2_{Wald}(2) = 1.34$, $p = 0.513$, $R^2_m = 0.03$, $CI_{95}$ [0.00, 0.82], $n_{obs} = 144$
Cell Proportion
0.0100
0.0075
0.0050
0.0025
0.0000
$\hat{\mu}_{median} = 0.00$
$\hat{\mu}_{median} = 0.00$
$\hat{\mu}_{median} = 0.00$
Control (n = 16)
ICI (n = 60)
AIH (n = 68)
Disease Type
Binomial GLMM w/ patient random intercepts (1|SlideID) | Proportion | Pairwise: emmeans, BH−adjusted

#### All Cell Death Events

$\chi^2_{Wald}(1) = 0.16$, $p = 0.689$, $R^2_m = 0.01$, $CI_{95}$ [0.00, 0.19], $n_{obs} = 60$

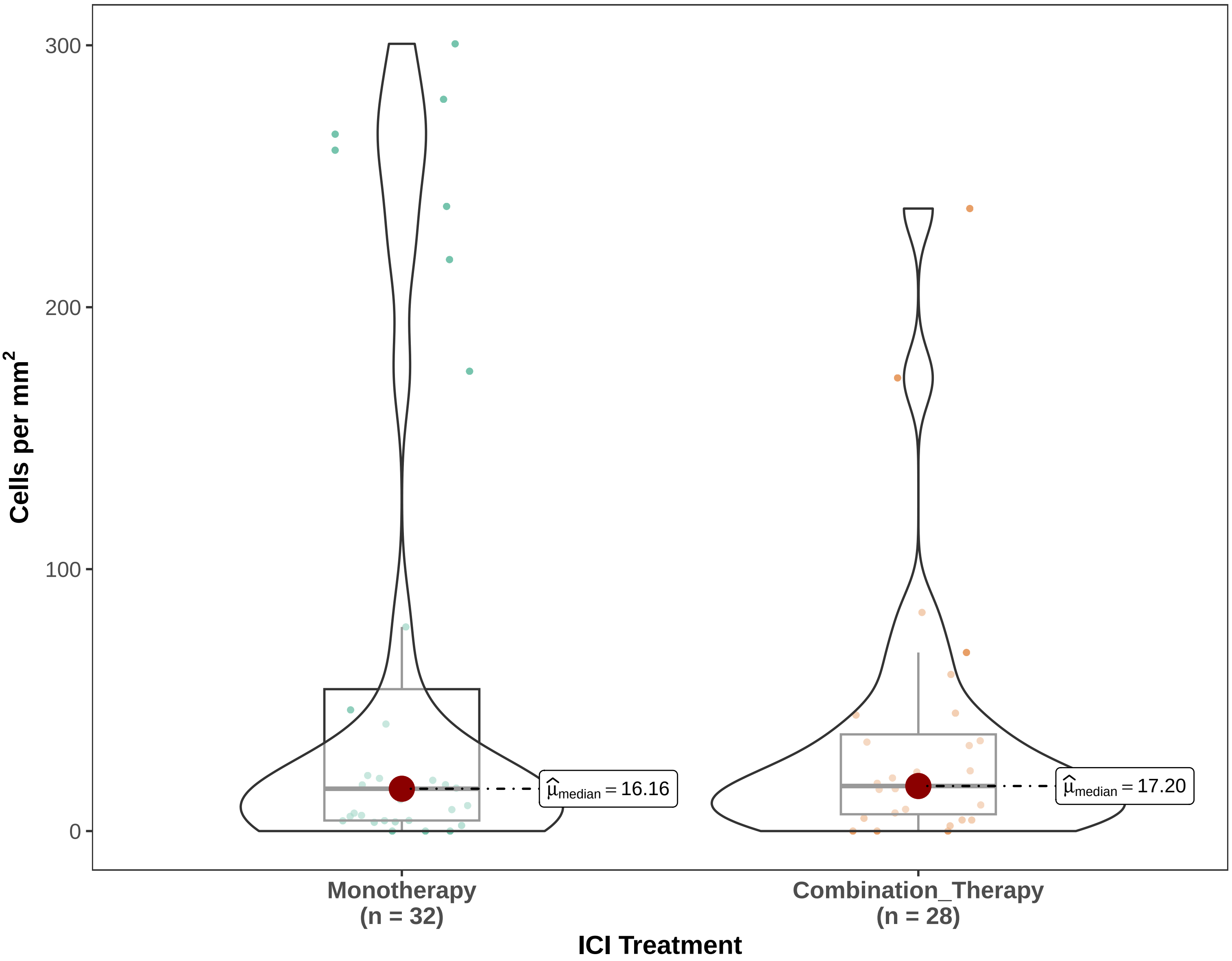


Gaussian LMM w/ patient random intercepts (1|SlideID) | Density | Pairwise: emmeans, BH−adjusted

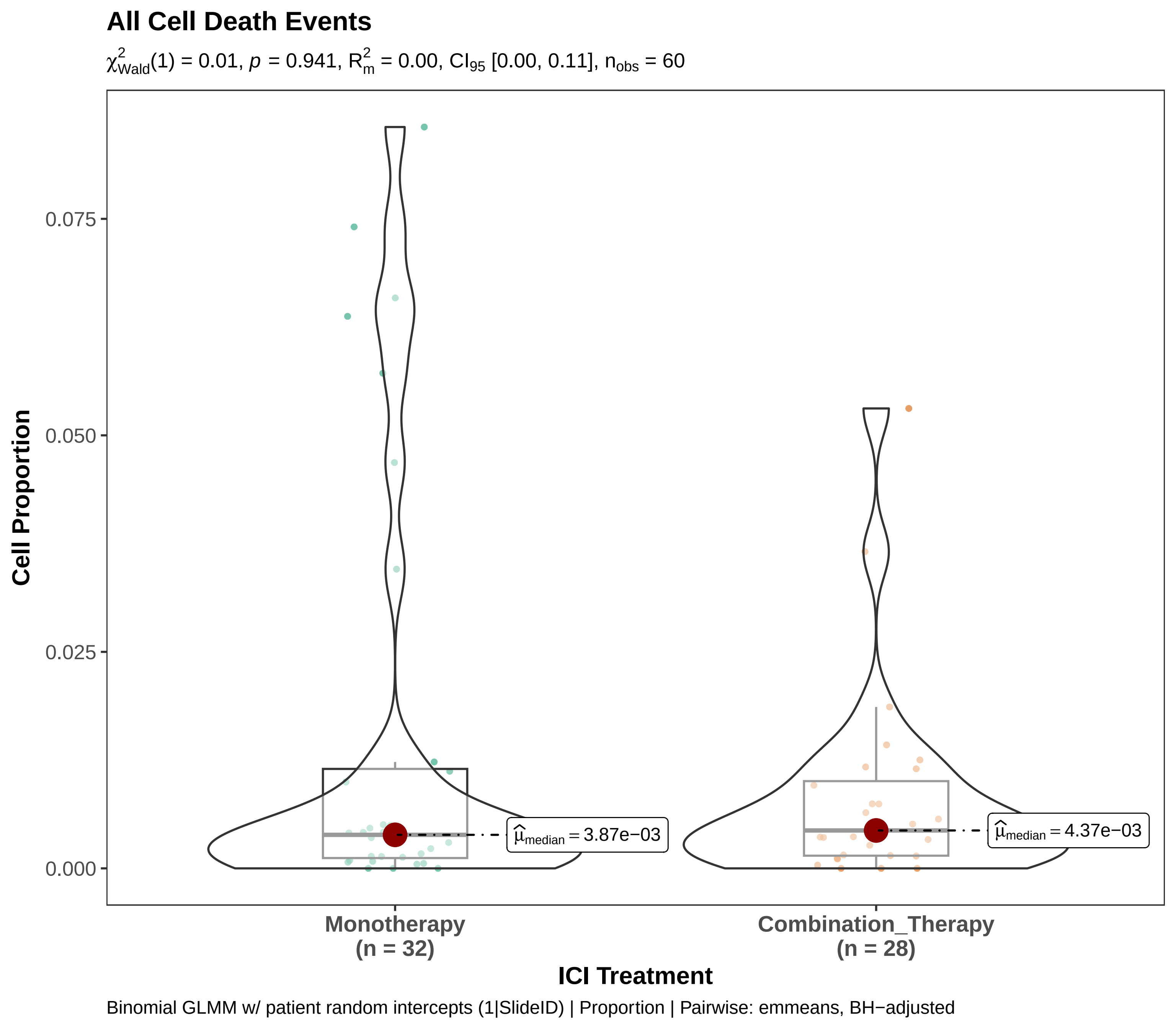

All Cell Death Events
$\chi^2_{Wald}(1) = 0.01$, $p = 0.941$, $R^2_m = 0.00$, $CI_{95}$ [0.00, 0.11], $n_{obs} = 60$
Cell Proportion
0.075
0.050
0.025
0.000
$\widehat{\mu}_{median} = 3.87e{-}03$
$\widehat{\mu}_{median} = 4.37e{-}03$
Monotherapy
(n = 32)
Combination_Therapy
(n = 28)
ICI Treatment
Binomial GLMM w/ patient random intercepts (1|SlideID) | Proportion | Pairwise: emmeans, BH−adjusted

**All Cell Death Events**

$\chi^2_{Wald}(2) = 0.33$, $p = 0.847$, $R^2_m = 0.01$, $CI_{95}$ [0.00, 0.19], $n_{obs} = 144$

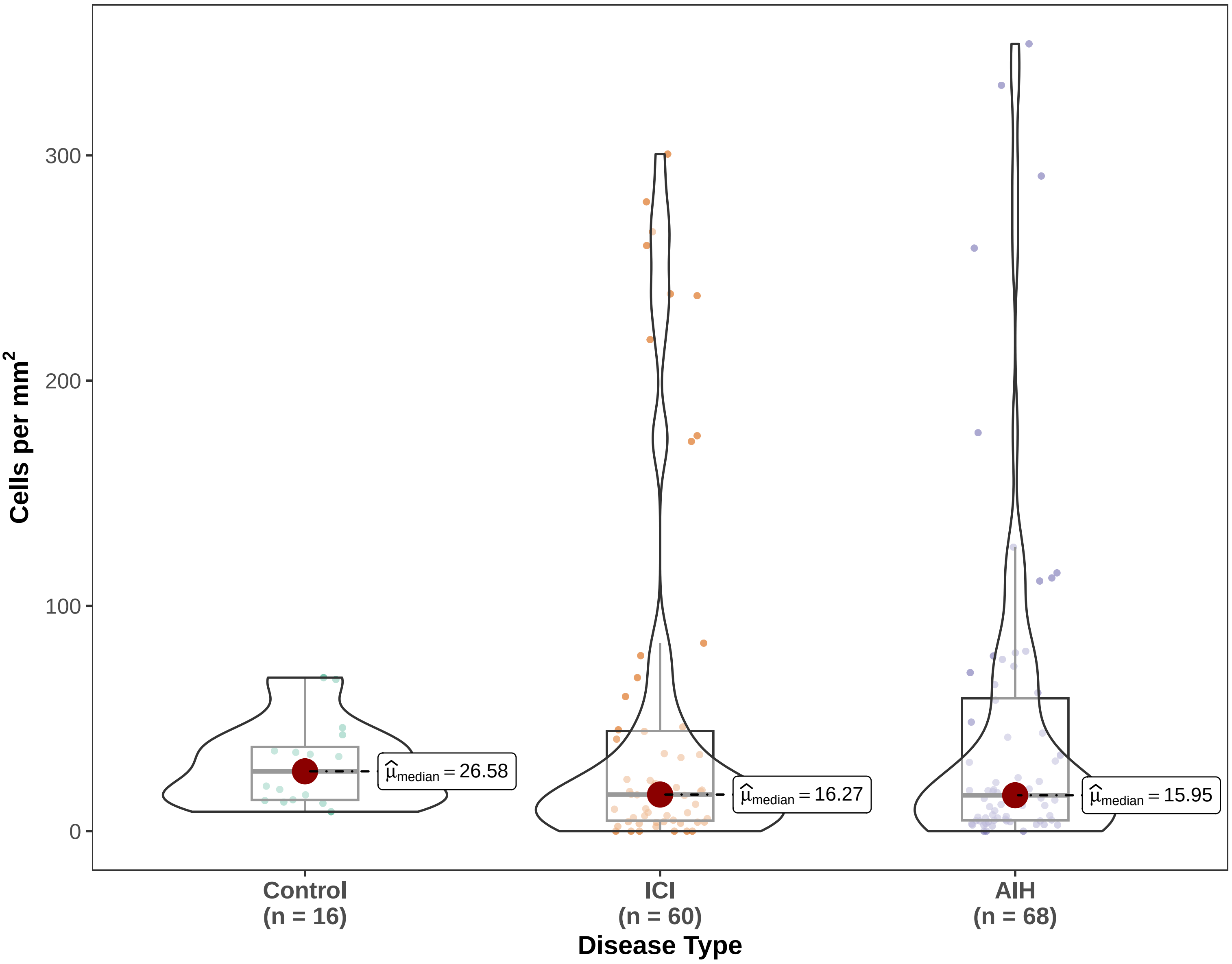


Gaussian LMM w/ patient random intercepts (1|SlideID) | Density | Pairwise: emmeans, BH−adjusted

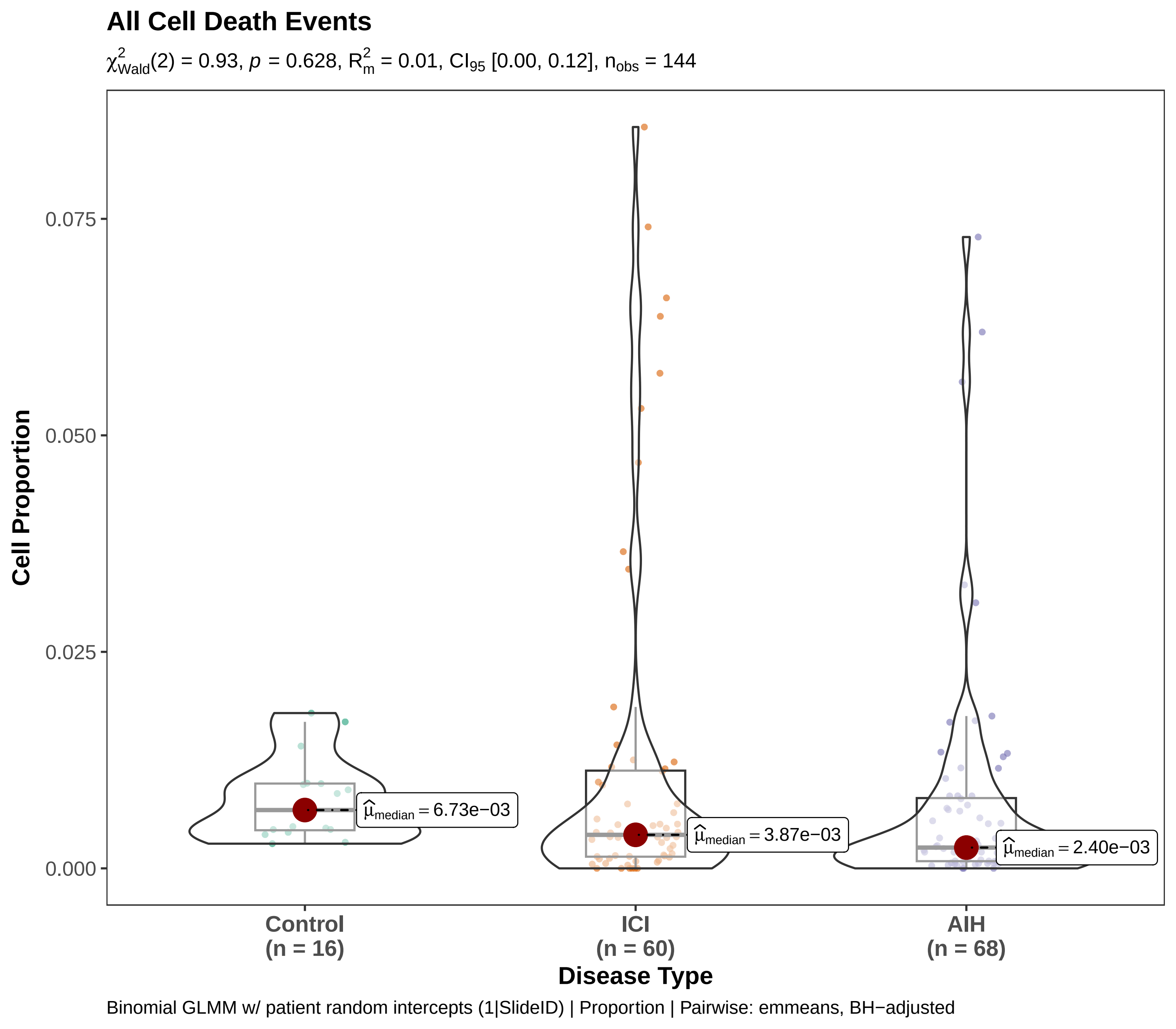

All Cell Death Events
$\chi^2_{Wald}(2) = 0.93$, $p = 0.628$, $R^2_m = 0.01$, $CI_{95}$ [0.00, 0.12], $n_{obs} = 144$
Cell Proportion
0.075
0.050
0.025
0.000
$\widehat{\mu}_{median} = 6.73e{-}03$
$\widehat{\mu}_{median} = 3.87e{-}03$
$\widehat{\mu}_{median} = 2.40e{-}03$
Control (n = 16)
ICI (n = 60)
AIH (n = 68)
Disease Type
Binomial GLMM w/ patient random intercepts (1|SlideID) | Proportion | Pairwise: emmeans, BH−adjusted

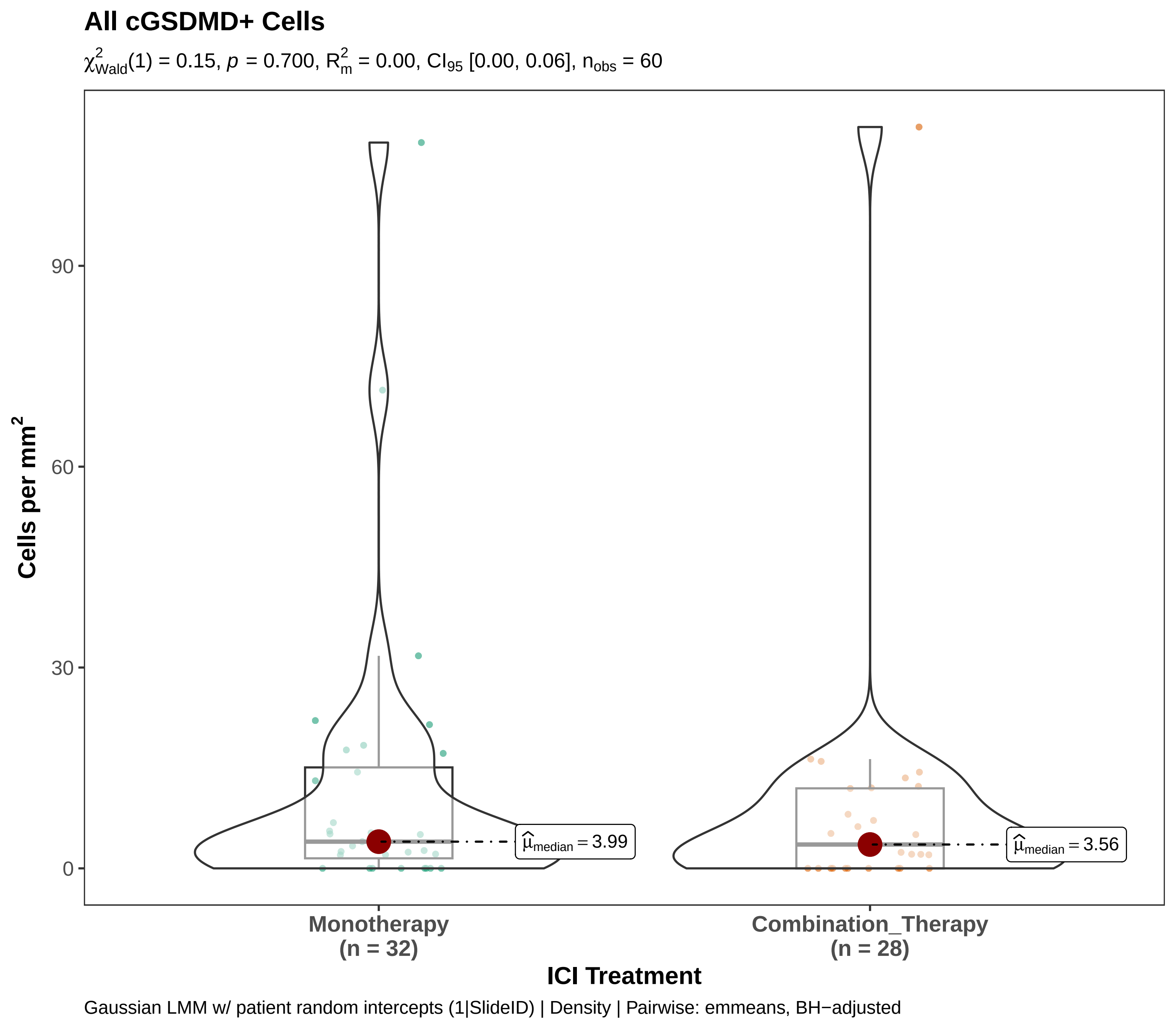
All cGSDMD+ Cells
$\chi^2_{Wald}(1) = 0.15$, $p = 0.700$, $R^2_m = 0.00$, $CI_{95}$ [0.00, 0.06], $n_{obs} = 60$
Cells per mm$^2$
90
60
30
0
$\widehat{\mu}_{median} = 3.99$
$\widehat{\mu}_{median} = 3.56$
Monotherapy
(n = 32)
Combination_Therapy
(n = 28)
ICI Treatment
Gaussian LMM w/ patient random intercepts (1|SlideID) | Density | Pairwise: emmeans, BH−adjusted

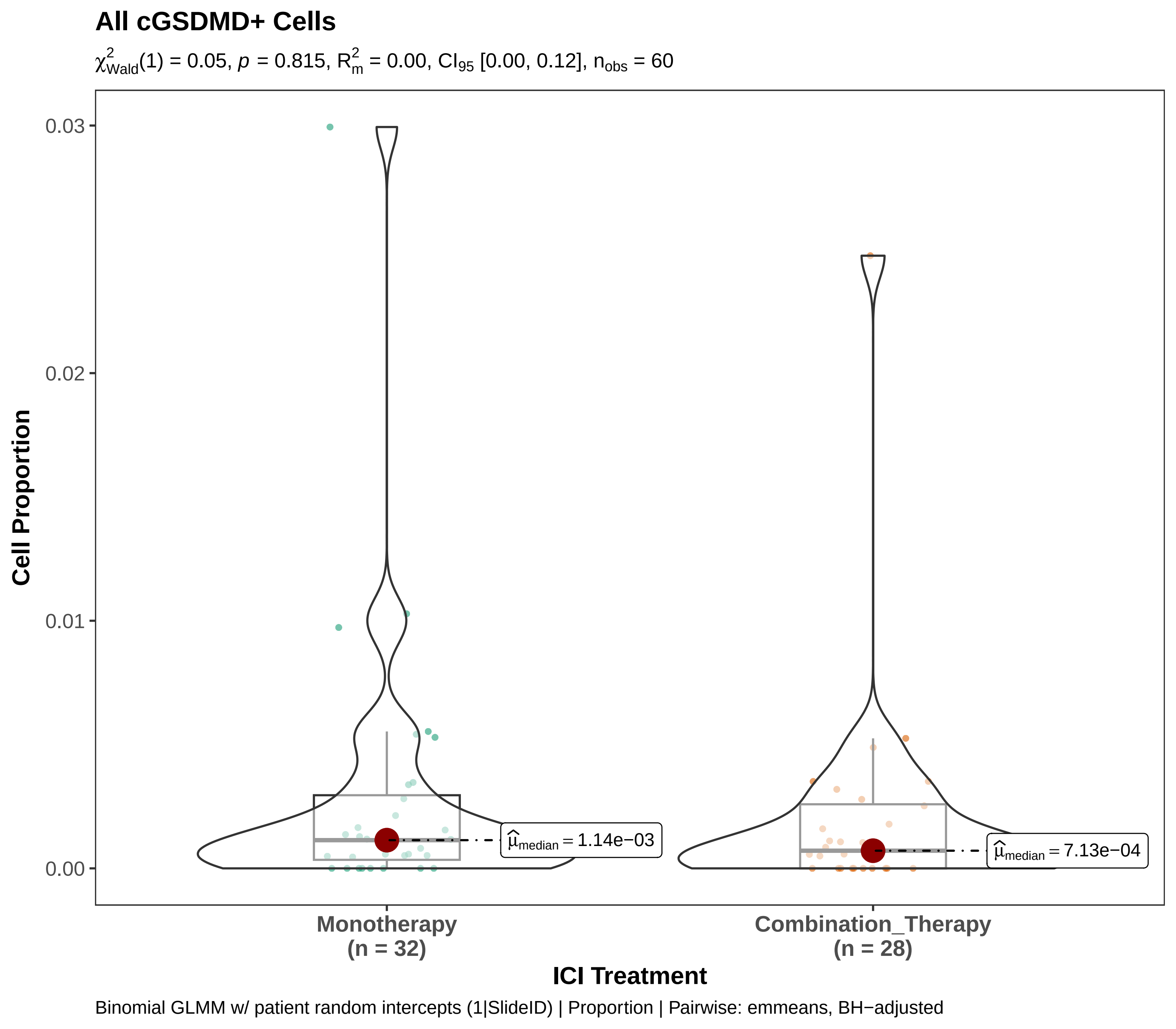
All cGSDMD+ Cells
$\chi^2_{Wald}(1) = 0.05$, $p = 0.815$, $R^2_m = 0.00$, $CI_{95}$ [0.00, 0.12], $n_{obs} = 60$
Cell Proportion
0.03
0.02
0.01
0.00
$\widehat{\mu}_{median} = 1.14e{-}03$
$\widehat{\mu}_{median} = 7.13e{-}04$
Monotherapy
(n = 32)
Combination_Therapy
(n = 28)
ICI Treatment
Binomial GLMM w/ patient random intercepts (1|SlideID) | Proportion | Pairwise: emmeans, BH−adjusted

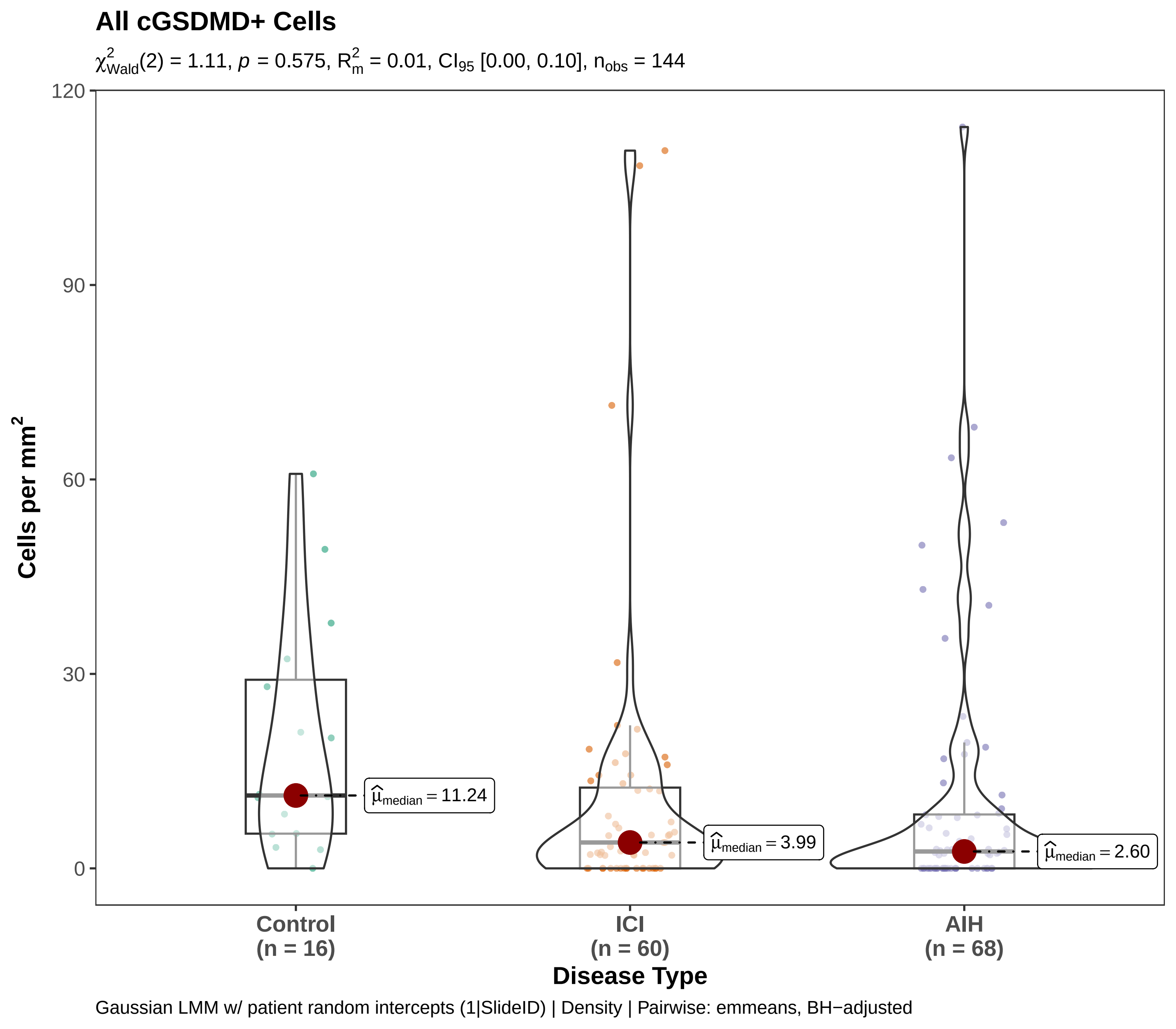

All cGSDMD+ Cells
$\chi^2_{Wald}(2) = 1.11$, $p = 0.575$, $R^2_m = 0.01$, $CI_{95}$ [0.00, 0.10], $n_{obs} = 144$
Cells per mm$^2$
120
90
60
30
0
$\widehat{\mu}_{median} = 11.24$
$\widehat{\mu}_{median} = 3.99$
$\widehat{\mu}_{median} = 2.60$
Control (n = 16)
ICI (n = 60)
AIH (n = 68)
Disease Type
Gaussian LMM w/ patient random intercepts (1|SlideID) | Density | Pairwise: emmeans, BH−adjusted

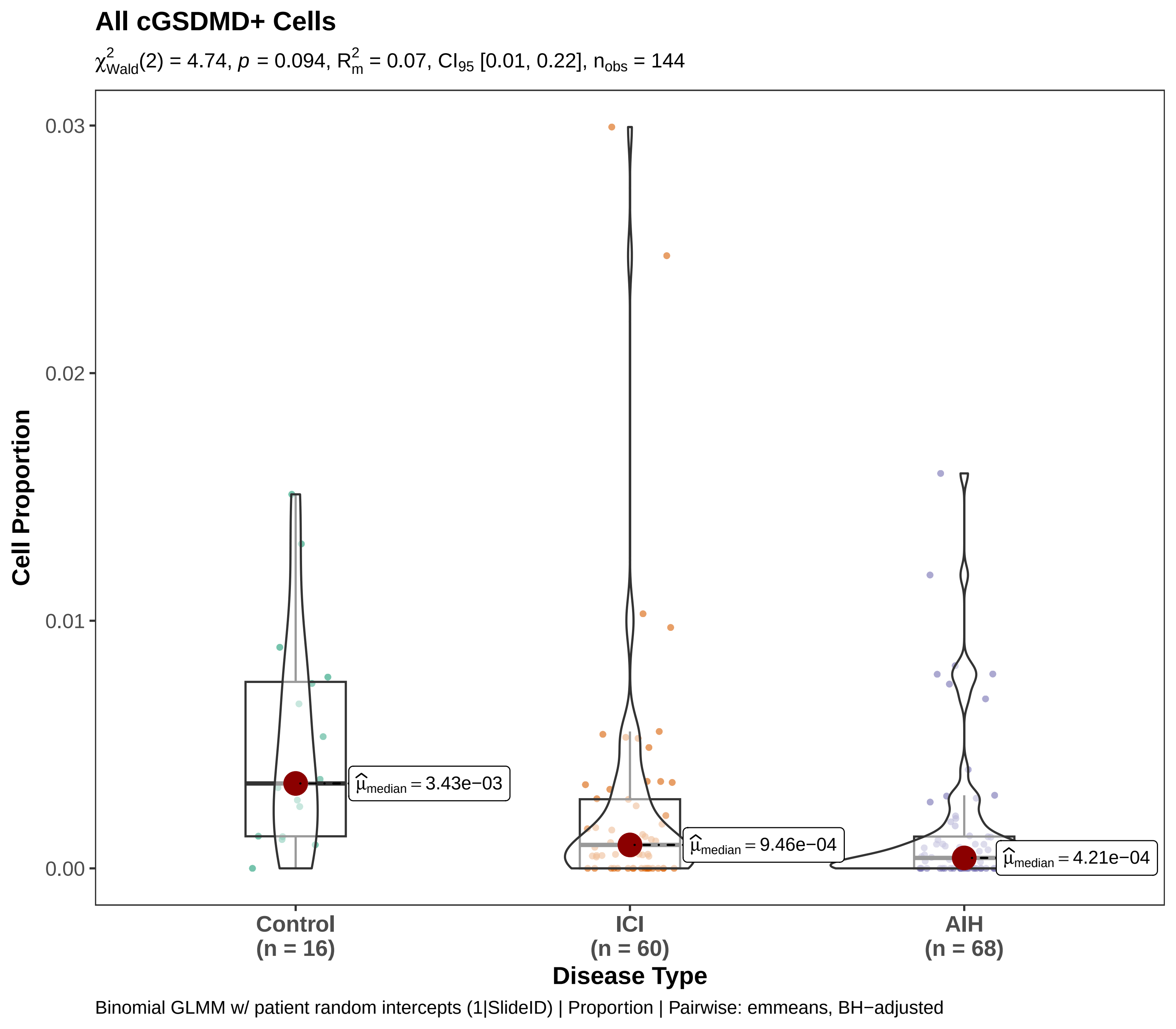
All cGSDMD+ Cells
$\chi^2_{\text{Wald}}(2) = 4.74$, $p = 0.094$, $R^2_m = 0.07$, $CI_{95}$ [0.01, 0.22], $n_{\text{obs}} = 144$
Cell Proportion
0.03
0.02
0.01
0.00
$\hat{\mu}_{\text{median}} = 3.43\text{e}{-03}$
$\hat{\mu}_{\text{median}} = 9.46\text{e}{-04}$
$\hat{\mu}_{\text{median}} = 4.21\text{e}{-04}$
Control (n = 16)
ICI (n = 60)
AIH (n = 68)
Disease Type
Binomial GLMM w/ patient random intercepts (1|SlideID) | Proportion | Pairwise: emmeans, BH−adjusted

#### Dying Hepatocytes (any)

$\chi^2_{Wald}(1) = 0.72$, $p = 0.397$, $R^2_m = 0.02$, $CI_{95}$ [0.00, 0.18], $n_{obs} = 60$

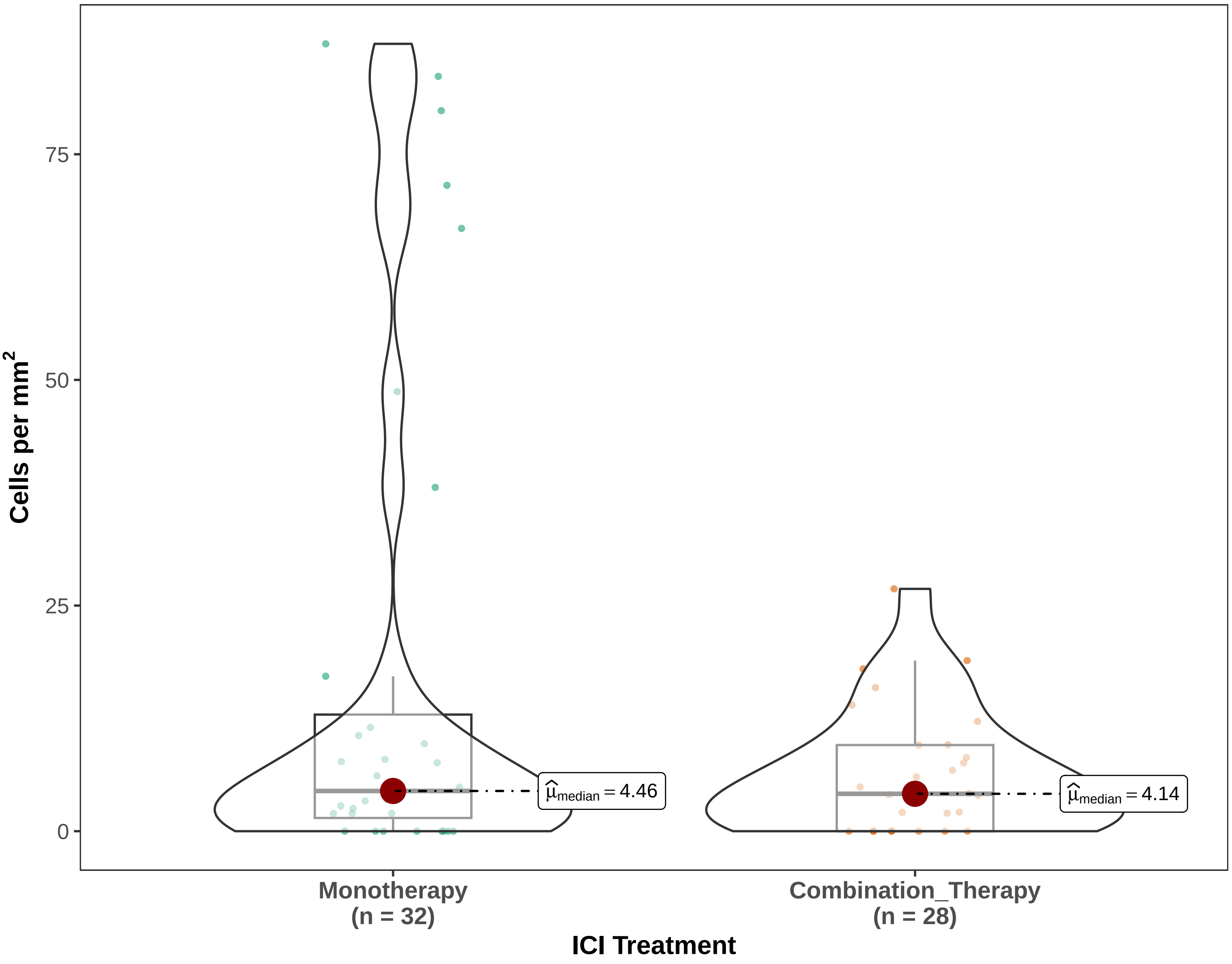


Gaussian LMM w/ patient random intercepts (1|SlideID) | Density | Pairwise: emmeans, BH−adjusted

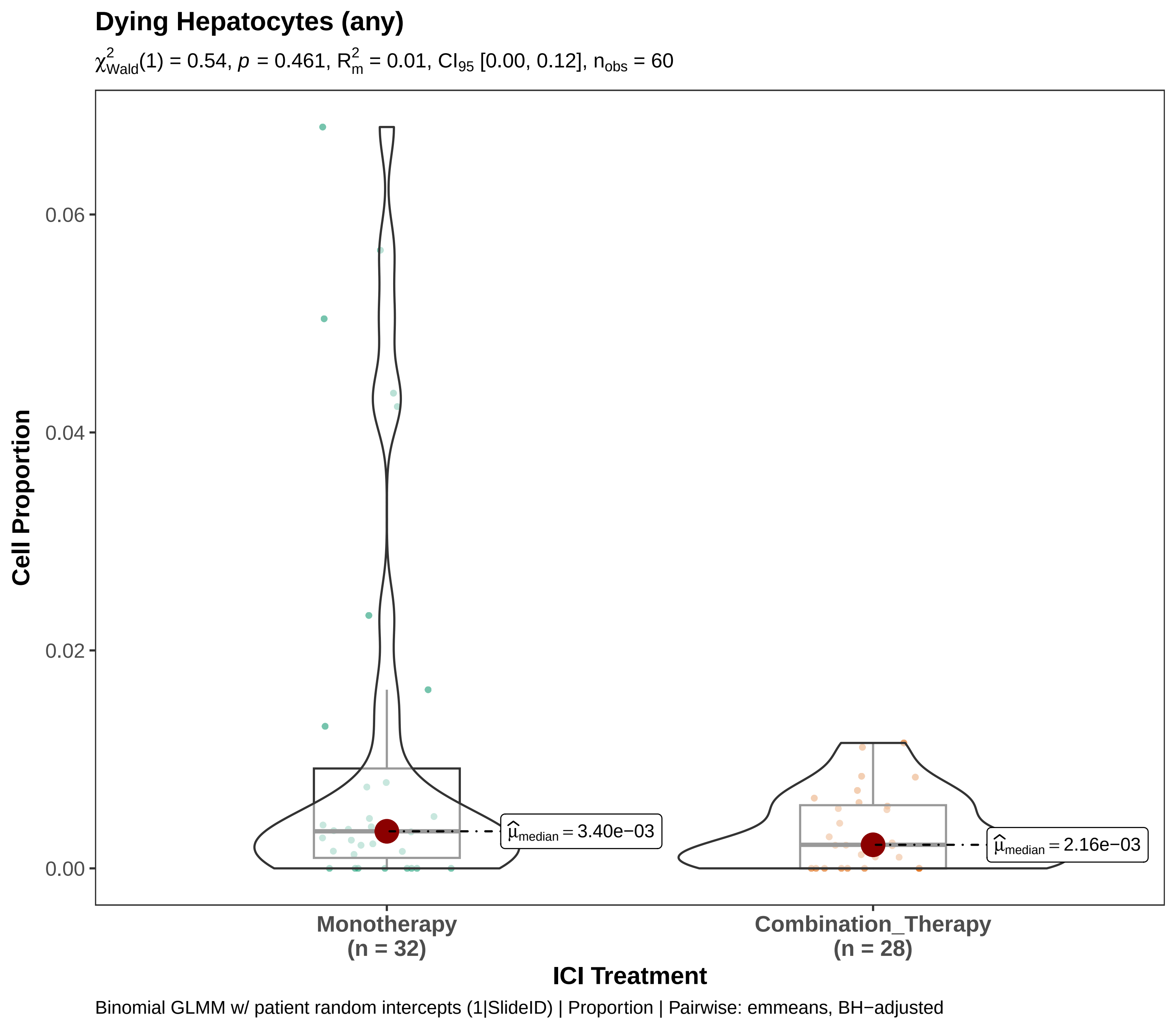
Dying Hepatocytes (any)
$\chi^2_{Wald}(1) = 0.54$, $p = 0.461$, $R^2_m = 0.01$, $CI_{95}$ [0.00, 0.12], $n_{obs} = 60$
Cell Proportion
0.00
0.02
0.04
0.06
$\widehat{\mu}_{median} = 3.40e{-}03$
$\widehat{\mu}_{median} = 2.16e{-}03$
Monotherapy
(n = 32)
Combination_Therapy
(n = 28)
ICI Treatment
Binomial GLMM w/ patient random intercepts (1|SlideID) | Proportion | Pairwise: emmeans, BH−adjusted

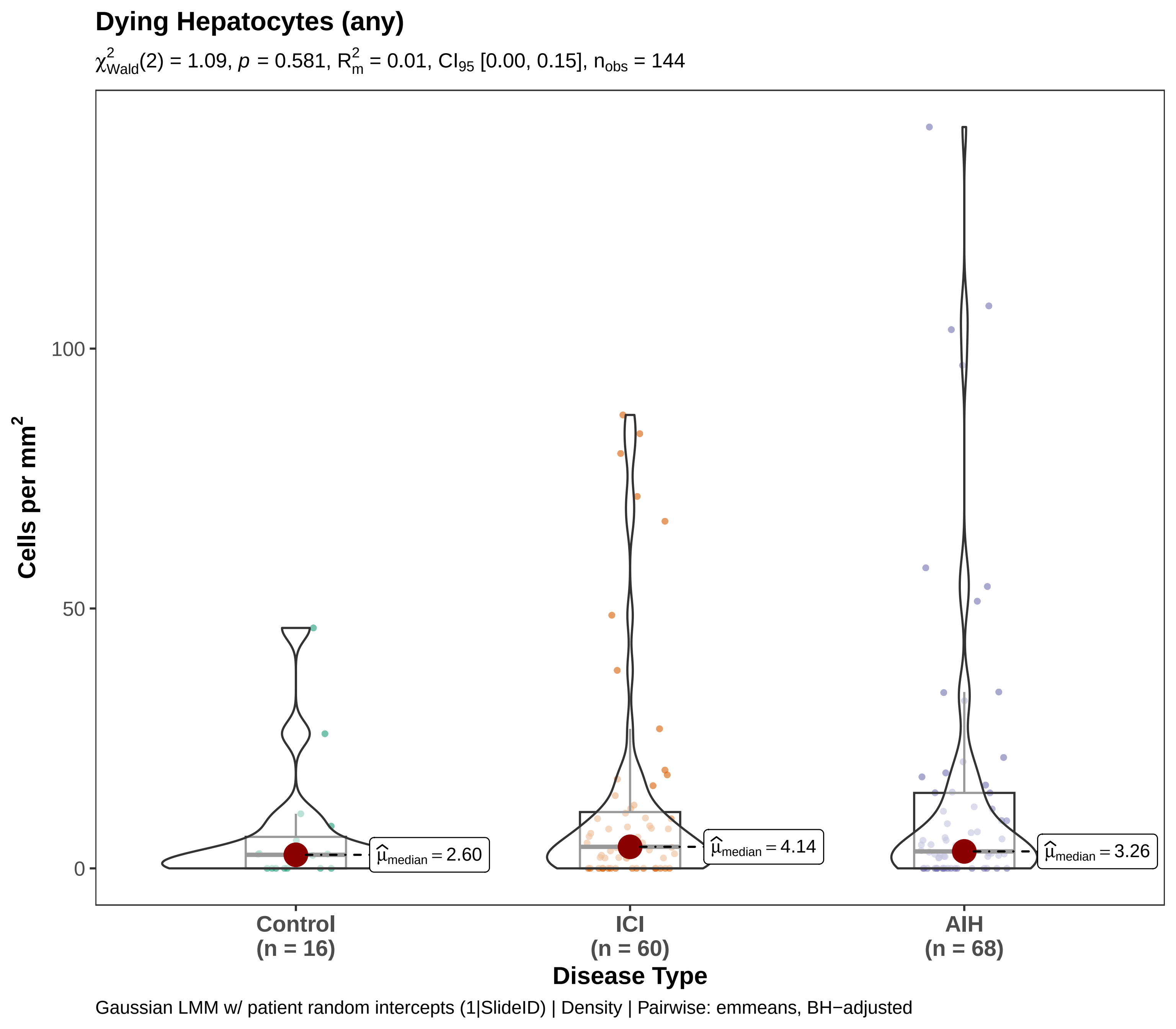
Dying Hepatocytes (any)
$\chi^2_{\mathrm{Wald}}(2) = 1.09$, $p = 0.581$, $R^2_m = 0.01$, $CI_{95}$ [0.00, 0.15], $n_{obs} = 144$
Cells per mm$^2$
100
50
0
$\hat{\mu}_{median} = 2.60$
$\hat{\mu}_{median} = 4.14$
$\hat{\mu}_{median} = 3.26$
Control
(n = 16)
ICI
(n = 60)
AIH
(n = 68)
Disease Type
Gaussian LMM w/ patient random intercepts (1|SlideID) | Density | Pairwise: emmeans, BH−adjusted

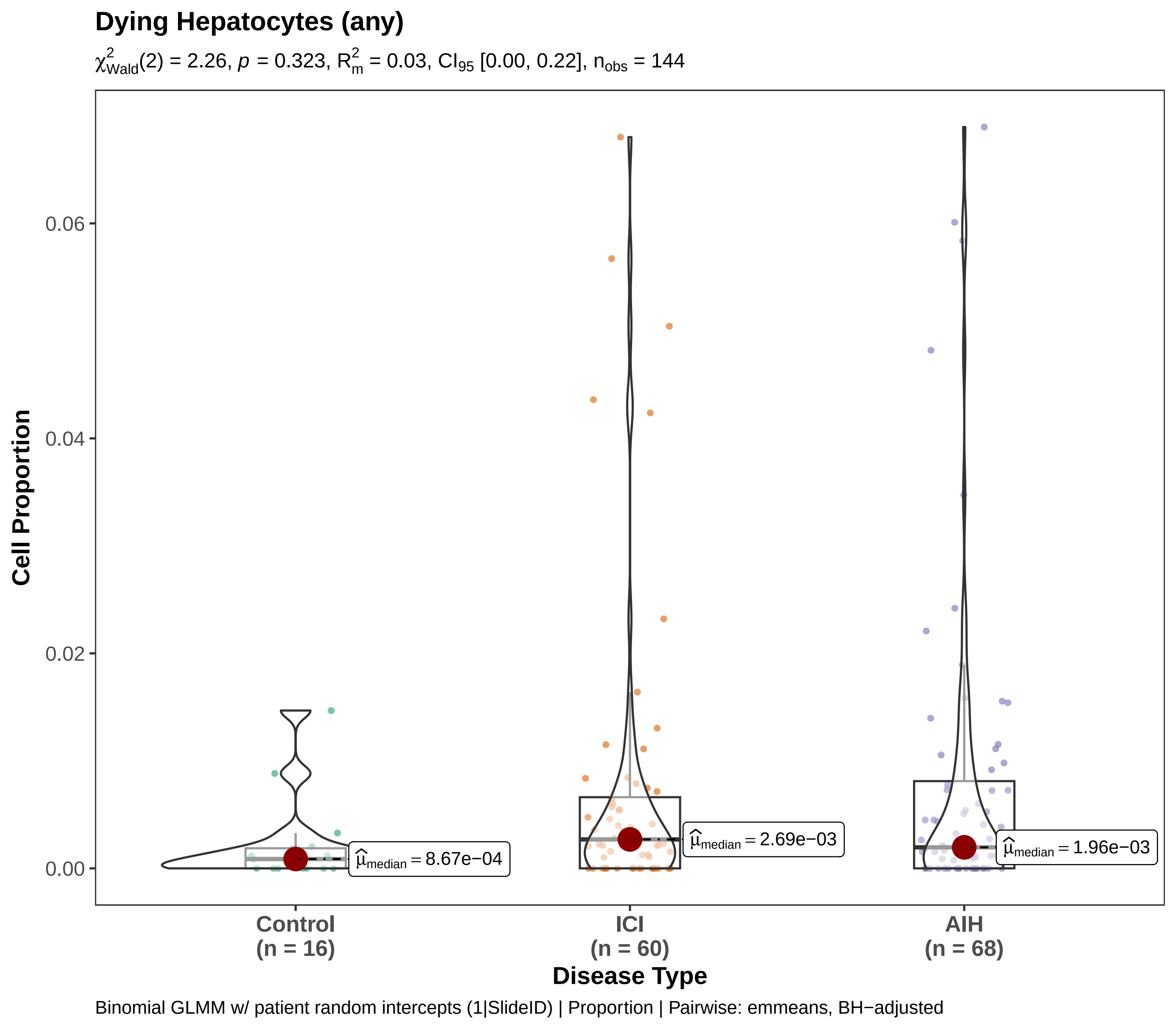

Dying Hepatocytes (any)
$\chi^2_{Wald}(2) = 2.26$, $p = 0.323$, $R^2_m = 0.03$, $CI_{95}$ [0.00, 0.22], $n_{obs} = 144$
Cell Proportion
0.06
0.04
0.02
0.00
$\hat{\mu}_{median} = 8.67e-04$
$\hat{\mu}_{median} = 2.69e-03$
$\hat{\mu}_{median} = 1.96e-03$
Control
(n = 16)
ICI
(n = 60)
AIH
(n = 68)
Disease Type
Binomial GLMM w/ patient random intercepts (1|SlideID) | Proportion | Pairwise: emmeans, BH−adjusted

**Dying Hepatocytes (CC3+)**

$\chi^2_{Wald}(1) = 0.40$, $p = 0.529$, $R^2_m = 0.01$, $CI_{95}$ [0.00, 0.18], $n_{obs} = 60$

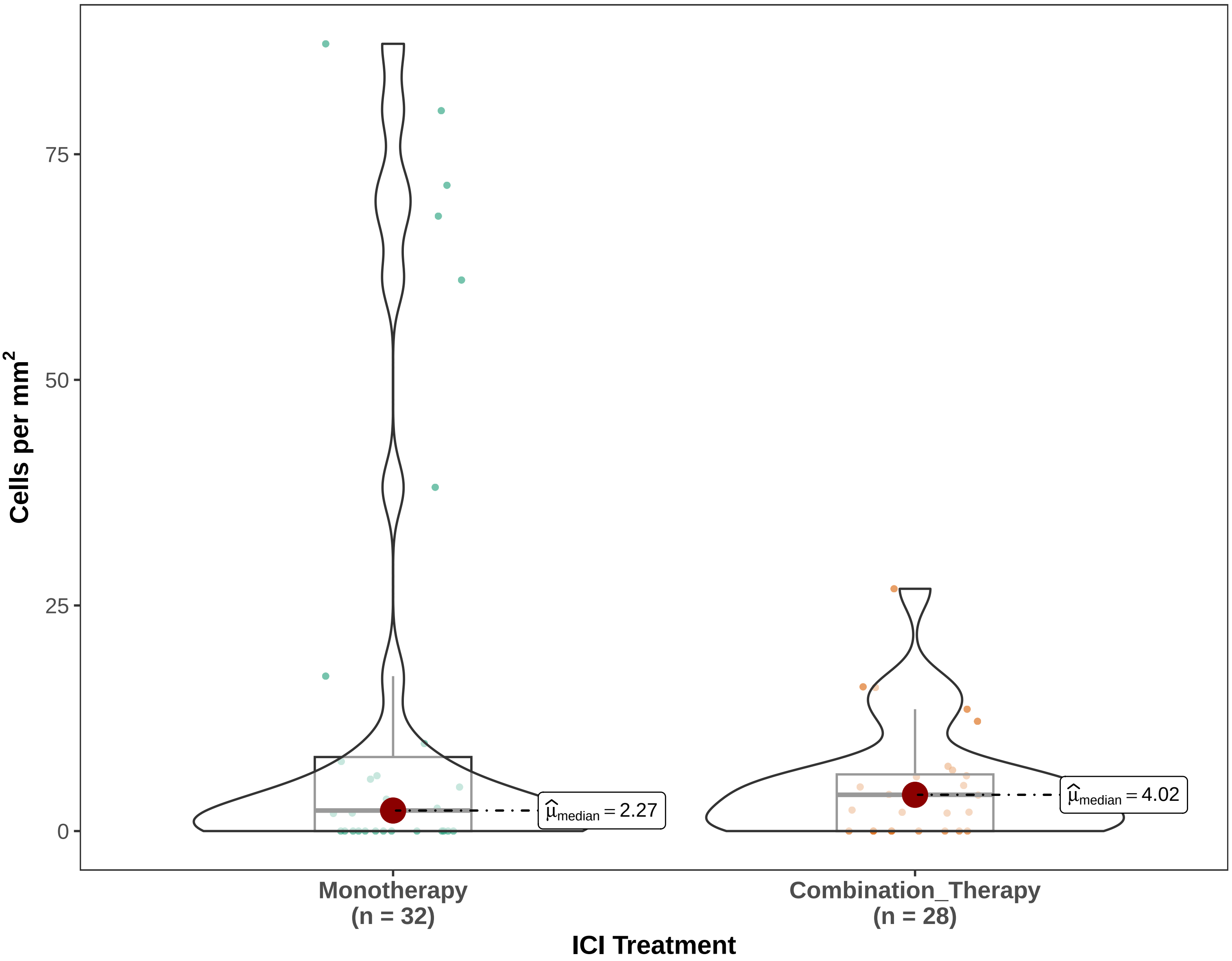


Gaussian LMM w/ patient random intercepts (1|SlideID) | Density | Pairwise: emmeans, BH−adjusted

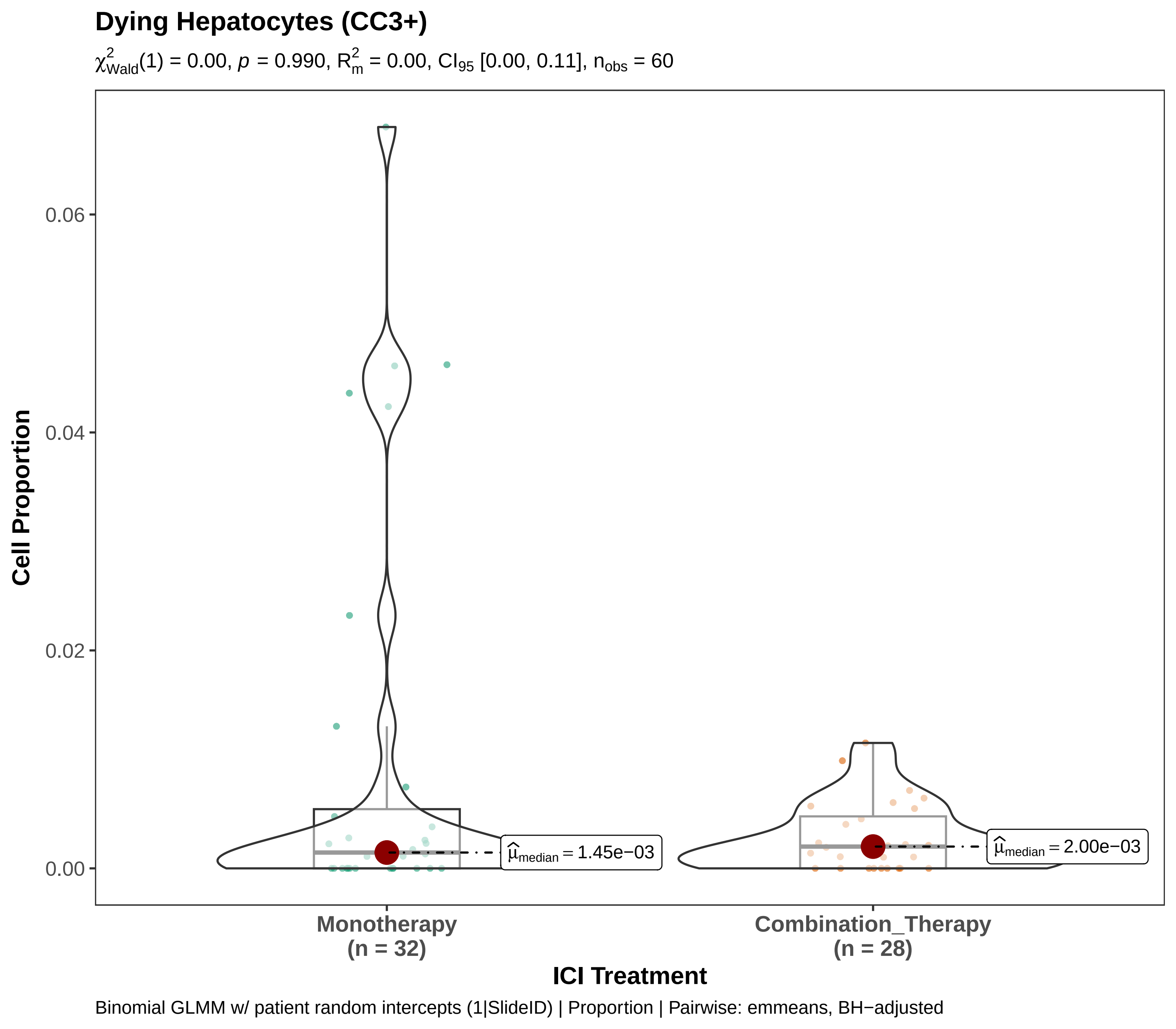

Dying Hepatocytes (CC3+)
$\chi^2_{Wald}(1) = 0.00$, $p = 0.990$, $R^2_m = 0.00$, $CI_{95}$ [0.00, 0.11], $n_{obs} = 60$
Cell Proportion
0.06
0.04
0.02
0.00
$\widehat{\mu}_{median} = 1.45e{-}03$
$\widehat{\mu}_{median} = 2.00e{-}03$
Monotherapy
(n = 32)
Combination_Therapy
(n = 28)
ICI Treatment
Binomial GLMM w/ patient random intercepts (1|SlideID) | Proportion | Pairwise: emmeans, BH−adjusted

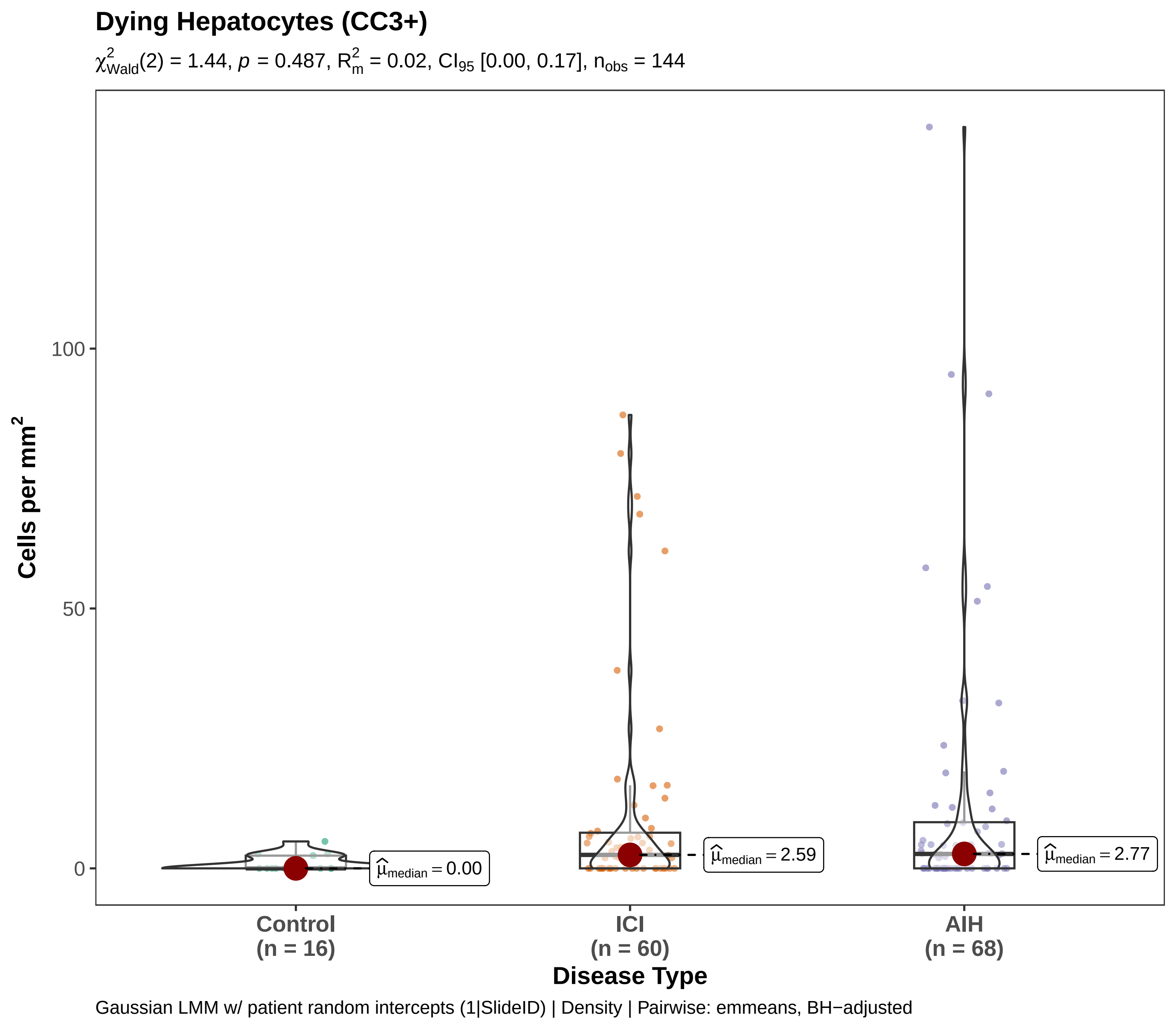
Dying Hepatocytes (CC3+)
$\chi^2_{Wald}(2) = 1.44$, $p = 0.487$, $R^2_m = 0.02$, $CI_{95}$ [0.00, 0.17], $n_{obs} = 144$
Cells per mm$^2$
100
50
0
$\widehat{\mu}_{median} = 0.00$
$\widehat{\mu}_{median} = 2.59$
$\widehat{\mu}_{median} = 2.77$
Control
(n = 16)
ICI
(n = 60)
AIH
(n = 68)
Disease Type
Gaussian LMM w/ patient random intercepts (1|SlideID) | Density | Pairwise: emmeans, BH−adjusted

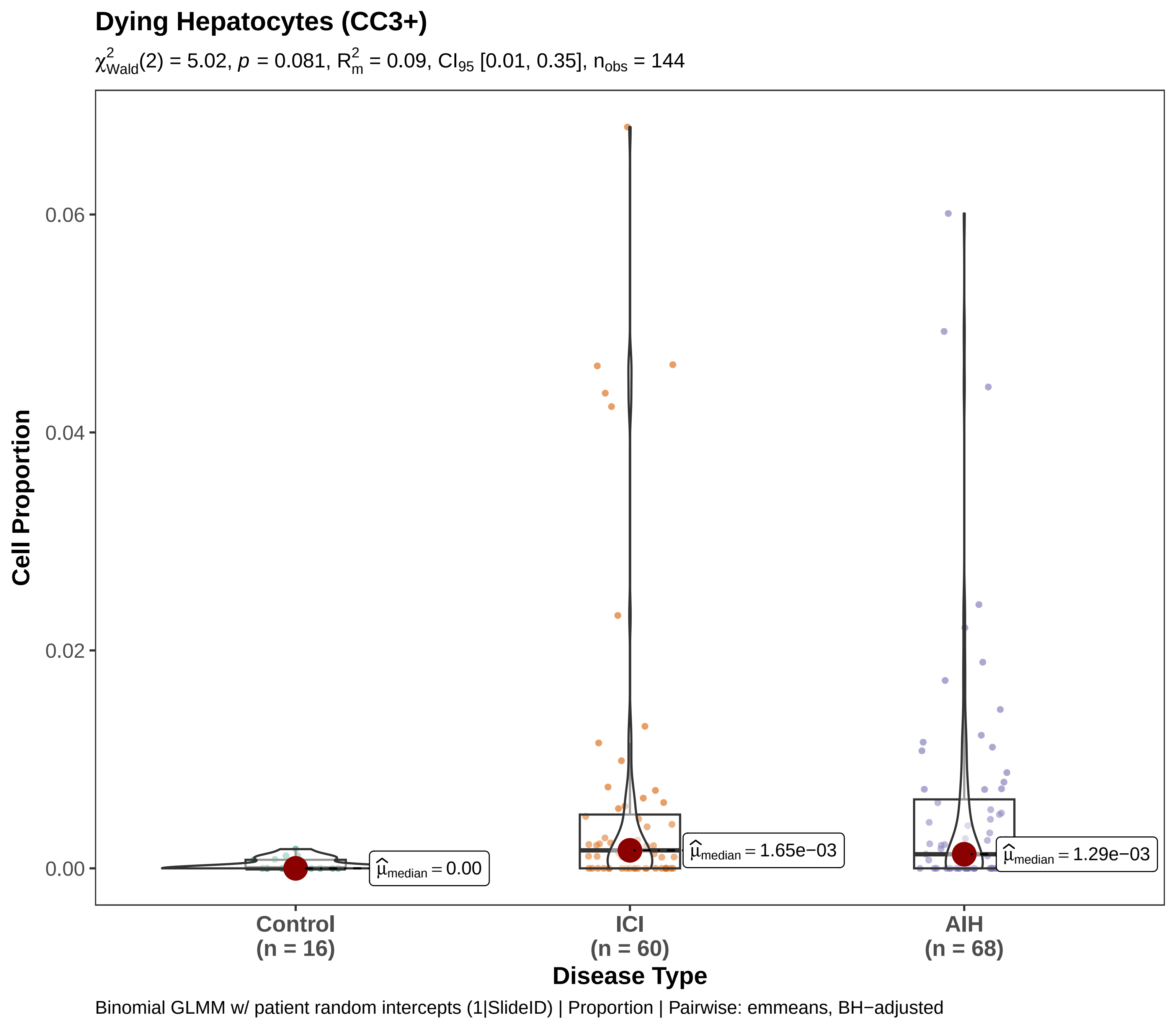
Dying Hepatocytes (CC3+)
$\chi^2_{\text{Wald}}(2) = 5.02$, $p = 0.081$, $R^2_m = 0.09$, $CI_{95}$ [0.01, 0.35], $n_{obs} = 144$
Cell Proportion
0.06
0.04
0.02
0.00
$\hat{\mu}_{median} = 0.00$
$\hat{\mu}_{median} = 1.65e{-03}$
$\hat{\mu}_{median} = 1.29e{-03}$
Control
(n = 16)
ICI
(n = 60)
AIH
(n = 68)
Disease Type
Binomial GLMM w/ patient random intercepts (1|SlideID) | Proportion | Pairwise: emmeans, BH−adjusted

#### Dying Hepatocytes (CC3+cGSDMD+)

$\chi^2_{Wald}(1) = 0.04$, $p = 0.840$, $R^2_m = 0.00$, $CI_{95}$ [0.00, 0.08], $n_{obs} = 60$

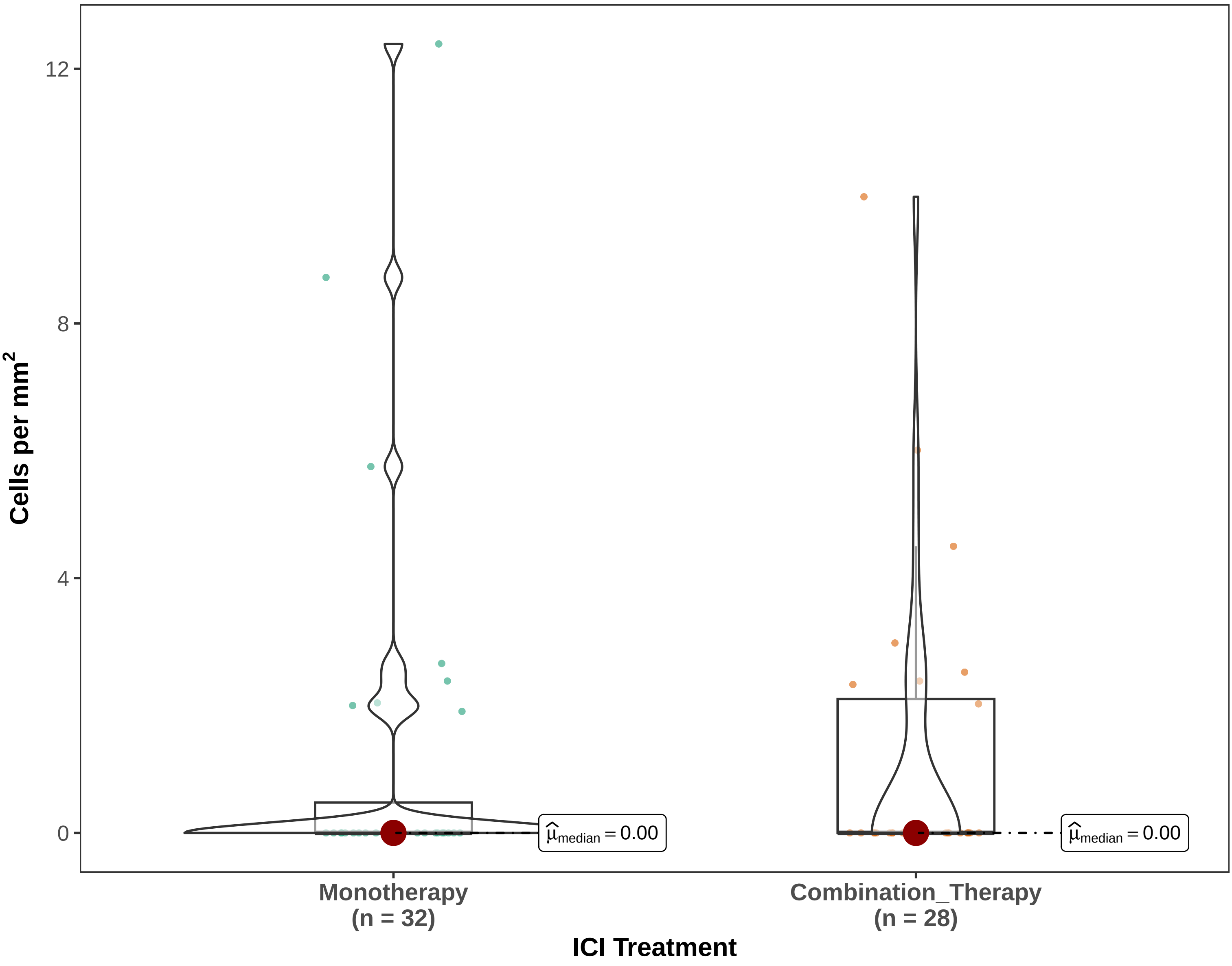


Gaussian LMM w/ patient random intercepts (1|SlideID) | Density | Pairwise: emmeans, BH−adjusted

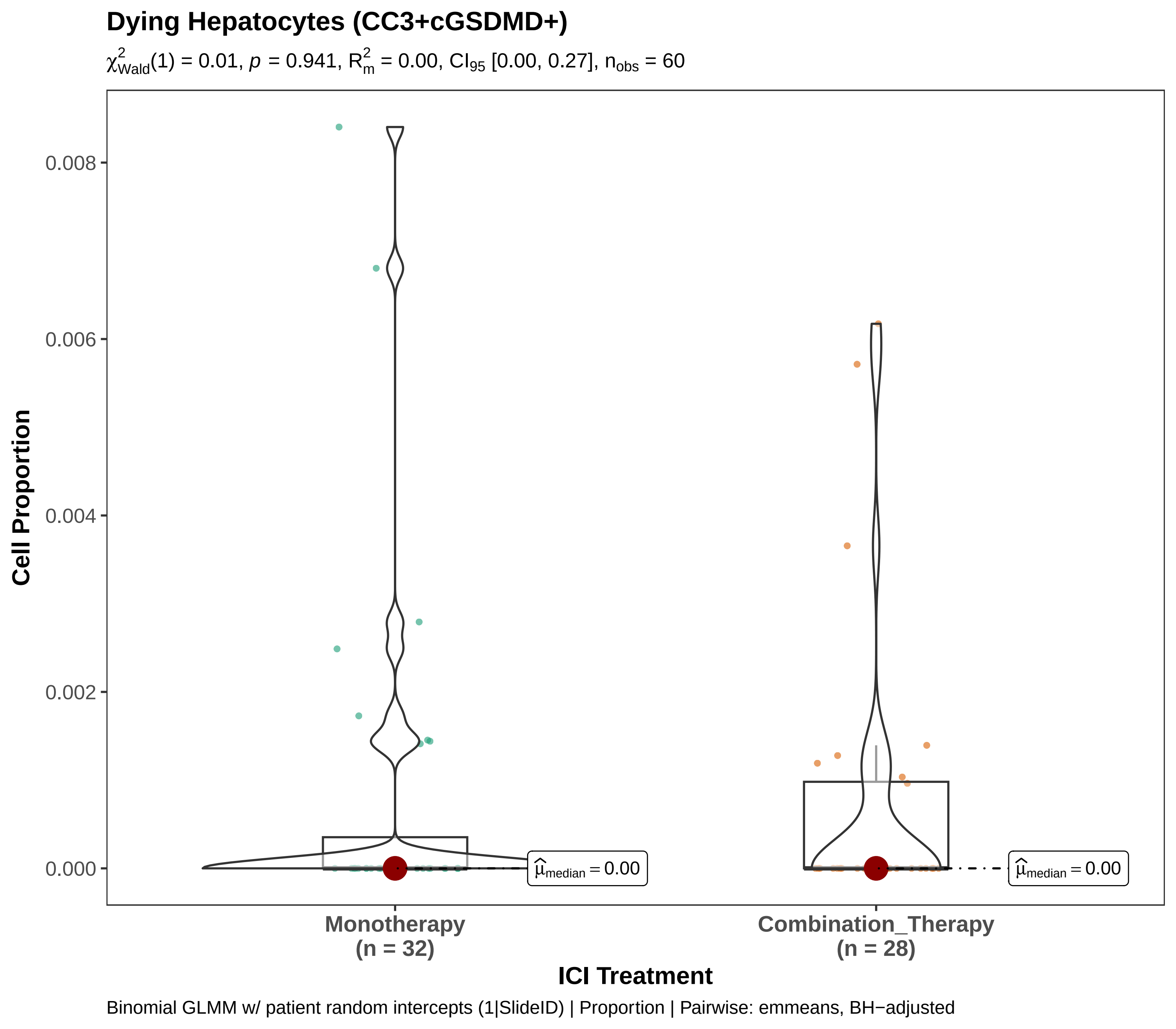
Dying Hepatocytes (CC3+cGSDMD+)
χ²Wald(1) = 0.01, p = 0.941, R²m = 0.00, CI95 [0.00, 0.27], nobs = 60
Cell Proportion
0.008
0.006
0.004
0.002
0.000
μ̂median = 0.00
μ̂median = 0.00
Monotherapy
(n = 32)
Combination_Therapy
(n = 28)
ICI Treatment
Binomial GLMM w/ patient random intercepts (1|SlideID) | Proportion | Pairwise: emmeans, BH−adjusted

### Dying Hepatocytes (CC3+cGSDMD+)

$\chi^2_{Wald}(2) = 0.72$, $p = 0.697$, $R^2_m = 0.01$, $CI_{95}$ [0.00, 0.11], $n_{obs} = 144$

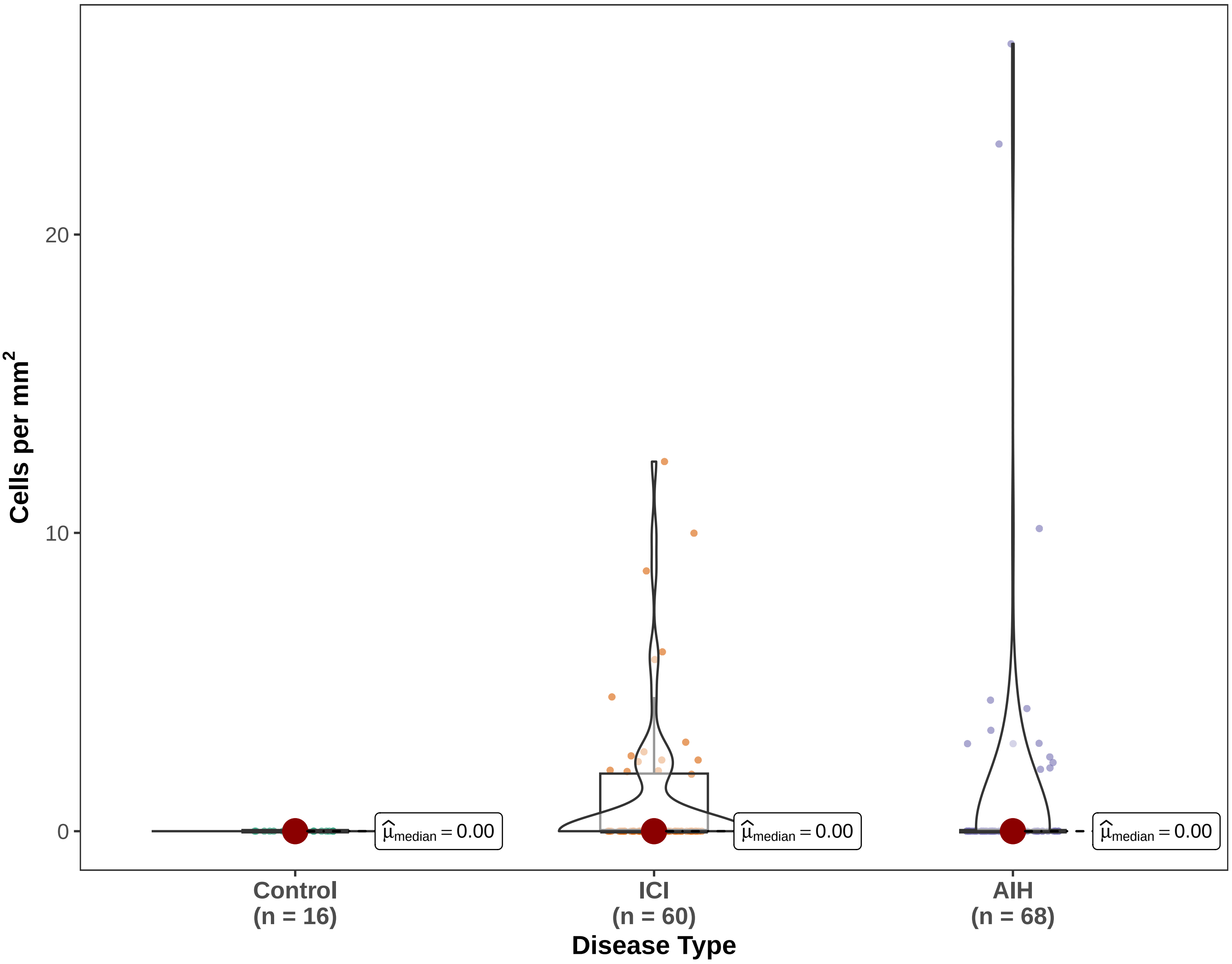


Gaussian LMM w/ patient random intercepts (1|SlideID) | Density | Pairwise: emmeans, BH−adjusted

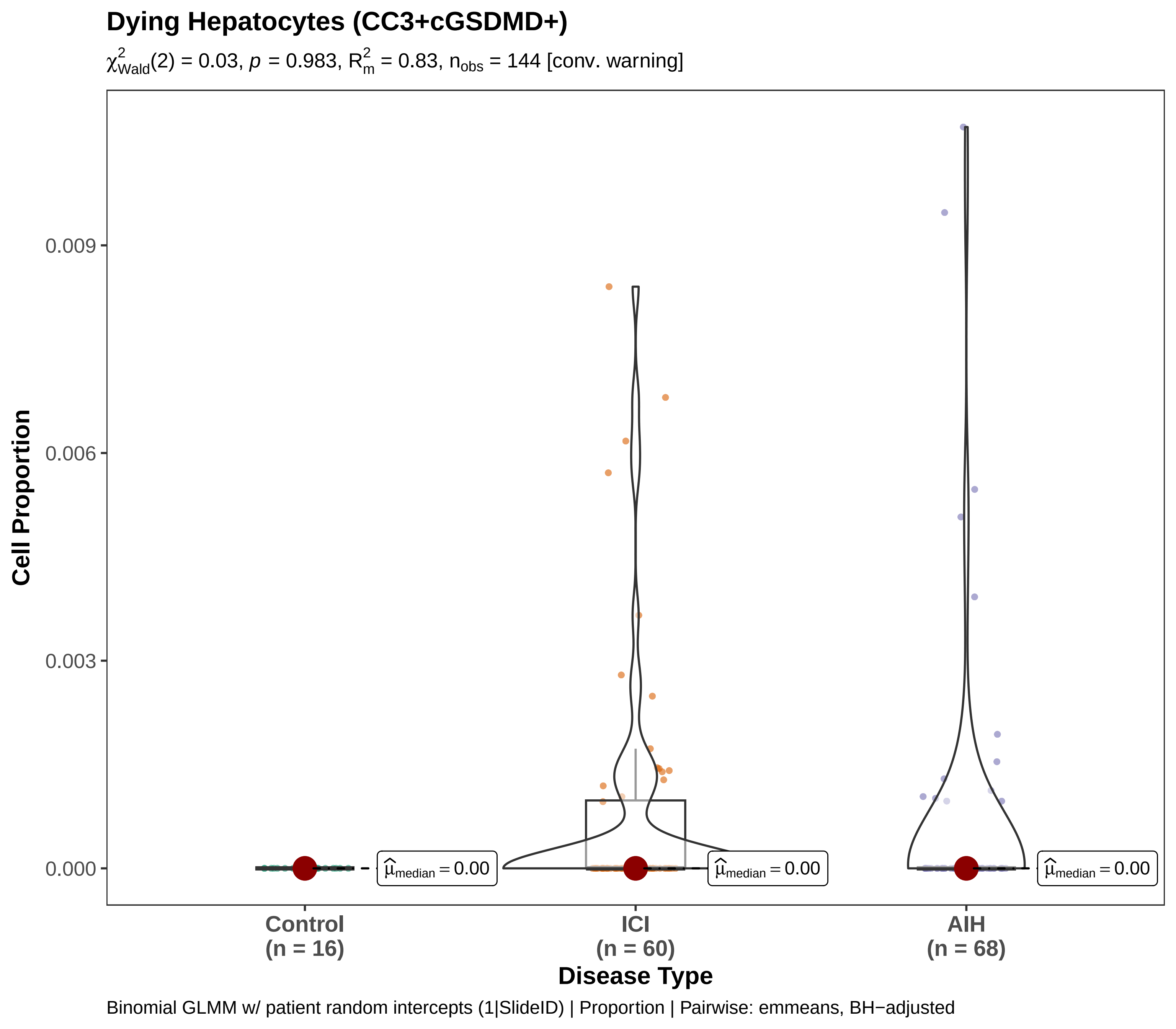

Dying Hepatocytes (CC3+cGSDMD+)
$\chi^2_{Wald}(2) = 0.03$, $p = 0.983$, $R^2_m = 0.83$, $n_{obs} = 144$ [conv. warning]
Cell Proportion
0.009
0.006
0.003
0.000
$\widehat{\mu}_{median} = 0.00$
$\widehat{\mu}_{median} = 0.00$
$\widehat{\mu}_{median} = 0.00$
Control
(n = 16)
ICI
(n = 60)
AIH
(n = 68)
Disease Type
Binomial GLMM w/ patient random intercepts (1|SlideID) | Proportion | Pairwise: emmeans, BH−adjusted

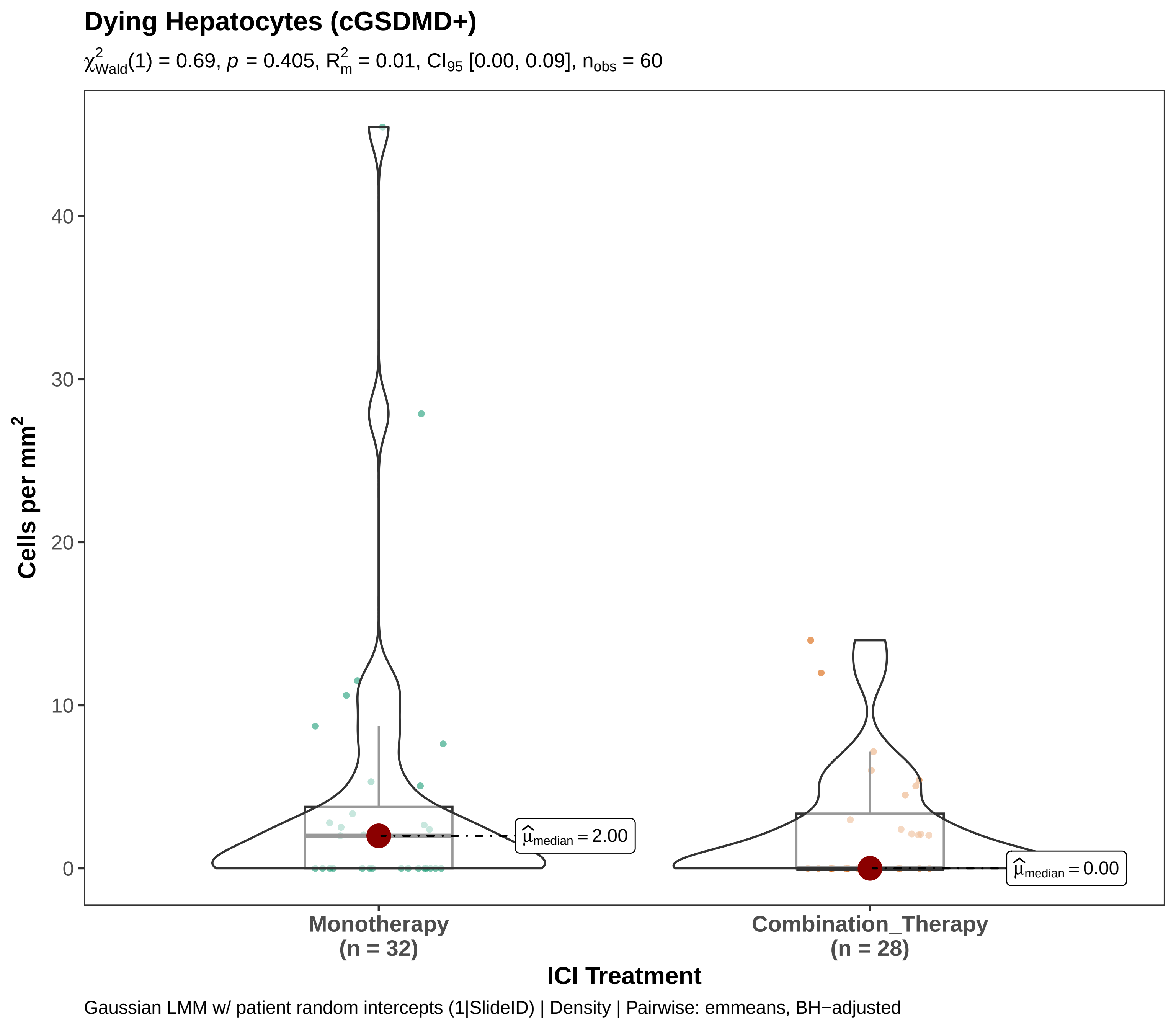
Dying Hepatocytes (cGSDMD+)
$\chi^2_{Wald}(1) = 0.69$, $p = 0.405$, $R^2_m = 0.01$, $CI_{95}$ [0.00, 0.09], $n_{obs} = 60$
Cells per mm$^2$
40
30
20
10
0
$\widehat{\mu}_{median} = 2.00$
$\widehat{\mu}_{median} = 0.00$
Monotherapy
(n = 32)
Combination_Therapy
(n = 28)
ICI Treatment
Gaussian LMM w/ patient random intercepts (1|SlideID) | Density | Pairwise: emmeans, BH−adjusted

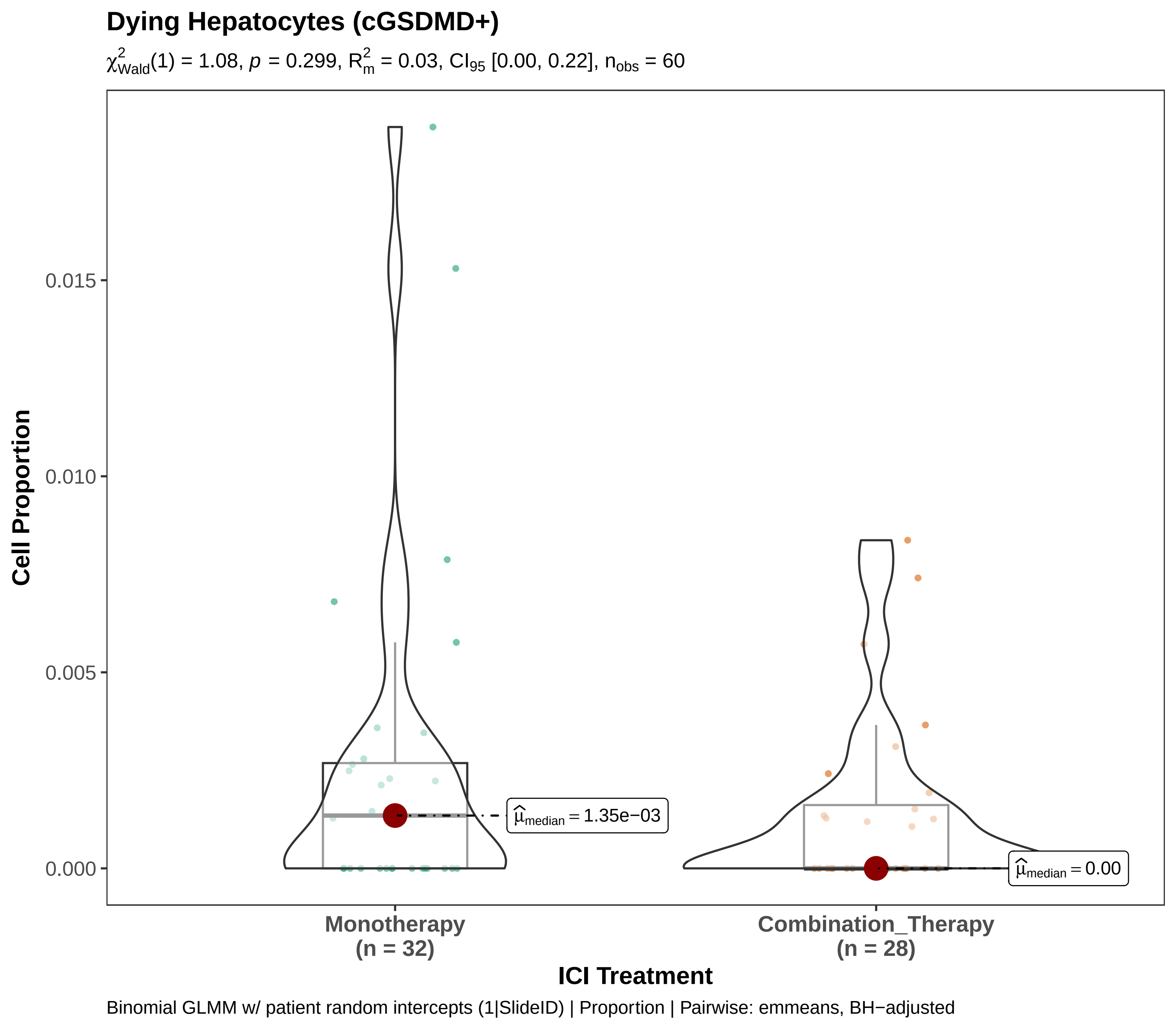

Dying Hepatocytes (cGSDMD+)
$\chi^2_{Wald}(1) = 1.08$, $p = 0.299$, $R^2_m = 0.03$, $CI_{95}$ [0.00, 0.22], $n_{obs} = 60$
Cell Proportion
0.015
0.010
0.005
0.000
$\widehat{\mu}_{median} = 1.35e-03$
$\widehat{\mu}_{median} = 0.00$
Monotherapy
(n = 32)
Combination_Therapy
(n = 28)
ICI Treatment
Binomial GLMM w/ patient random intercepts (1|SlideID) | Proportion | Pairwise: emmeans, BH−adjusted

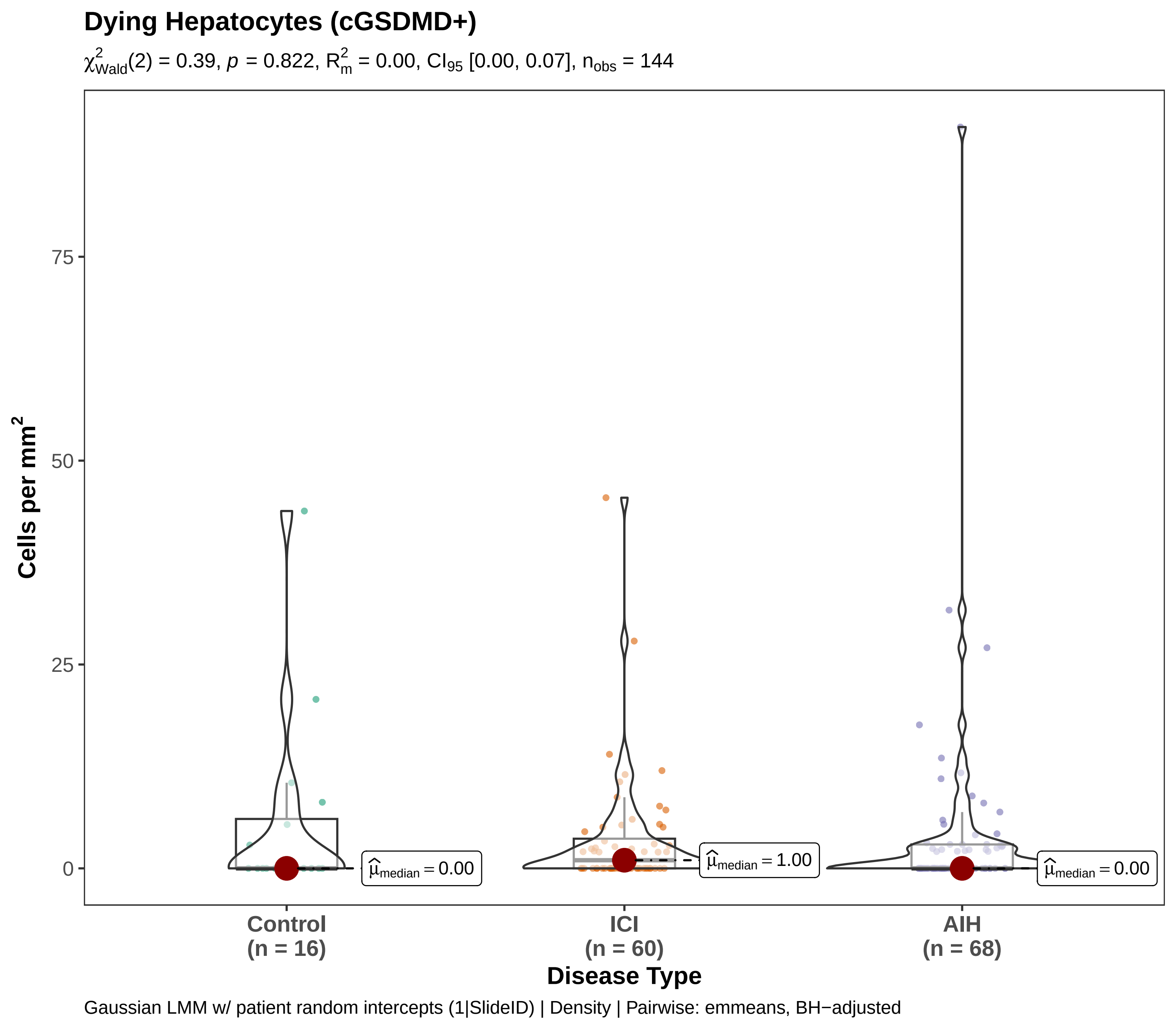
Dying Hepatocytes (cGSDMD+)
$\chi^2_{Wald}(2) = 0.39$, $p = 0.822$, $R^2_m = 0.00$, $CI_{95}$ [0.00, 0.07], $n_{obs} = 144$
Cells per mm$^2$
75
50
25
0
$\hat{\mu}_{median} = 0.00$
$\hat{\mu}_{median} = 1.00$
$\hat{\mu}_{median} = 0.00$
Control
(n = 16)
ICI
(n = 60)
AIH
(n = 68)
Disease Type
Gaussian LMM w/ patient random intercepts (1|SlideID) | Density | Pairwise: emmeans, BH−adjusted

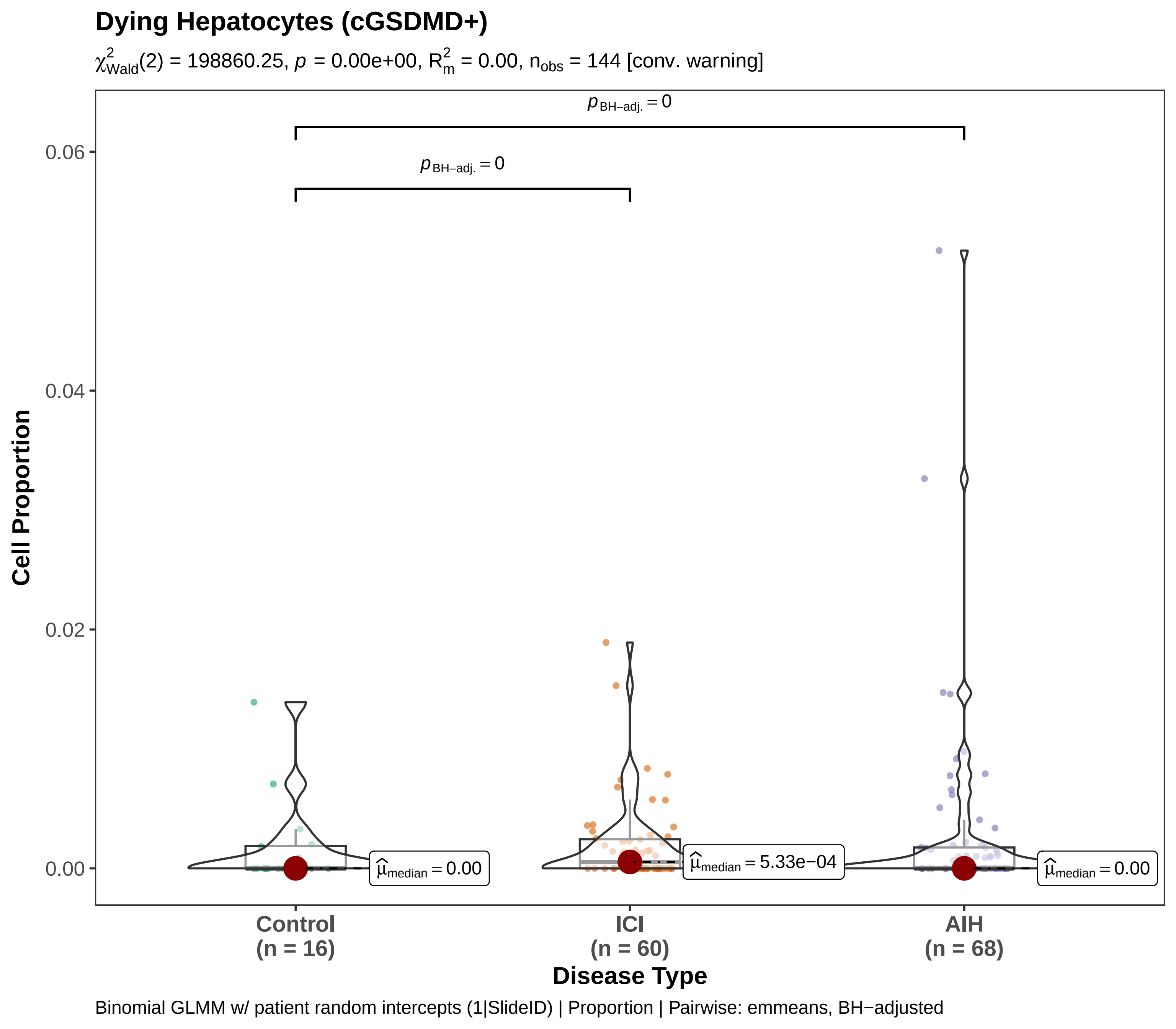

Dying Hepatocytes (cGSDMD+)
$\chi^2_{Wald}(2)$ = 198860.25, $p$ = 0.00e+00, $R^2_m$ = 0.00, $n_{obs}$ = 144 [conv. warning]
$p_{BH-adj.} = 0$
$p_{BH-adj.} = 0$
0.06
0.04
0.02
0.00
Cell Proportion
$\hat{\mu}_{median} = 0.00$
$\hat{\mu}_{median} = 5.33e-04$
$\hat{\mu}_{median} = 0.00$
Control
(n = 16)
ICI
(n = 60)
AIH
(n = 68)
Disease Type
Binomial GLMM w/ patient random intercepts (1|SlideID) | Proportion | Pairwise: emmeans, BH−adjusted

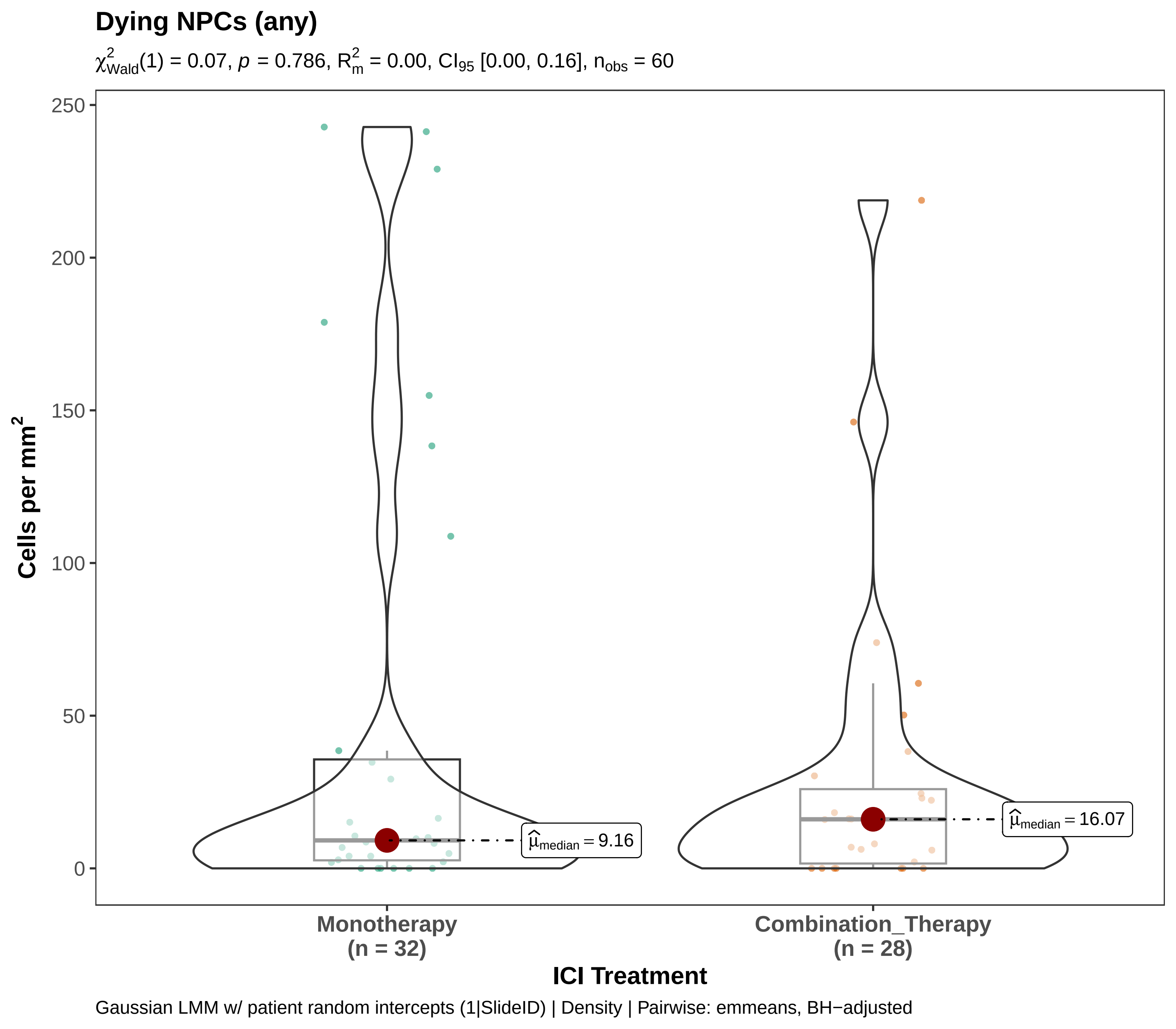
Dying NPCs (any)
$\chi^2_{Wald}(1) = 0.07$, $p = 0.786$, $R^2_m = 0.00$, $CI_{95}$ [0.00, 0.16], $n_{obs} = 60$
Cells per mm$^2$
250
200
150
100
50
0
$\widehat{\mu}_{median} = 9.16$
$\widehat{\mu}_{median} = 16.07$
Monotherapy
(n = 32)
Combination_Therapy
(n = 28)
ICI Treatment
Gaussian LMM w/ patient random intercepts (1|SlideID) | Density | Pairwise: emmeans, BH−adjusted

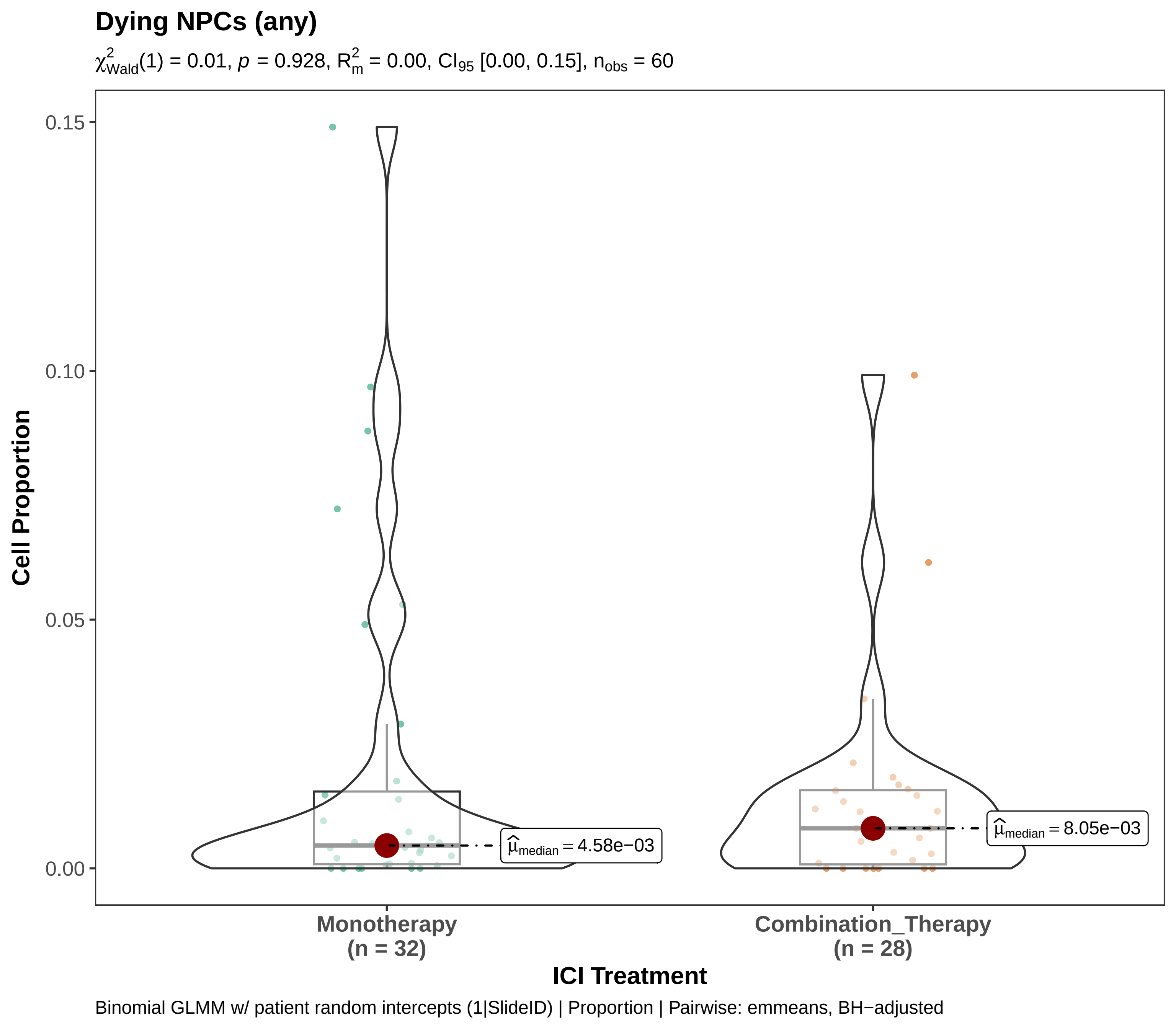

Dying NPCs (any)
$\chi^2_{Wald}(1) = 0.01$, $p = 0.928$, $R^2_m = 0.00$, $CI_{95}$ [0.00, 0.15], $n_{obs} = 60$
Cell Proportion
0.15
0.10
0.05
0.00
$\widehat{\mu}_{median} = 4.58e{-}03$
$\widehat{\mu}_{median} = 8.05e{-}03$
Monotherapy
(n = 32)
Combination_Therapy
(n = 28)
ICI Treatment
Binomial GLMM w/ patient random intercepts (1|SlideID) | Proportion | Pairwise: emmeans, BH−adjusted

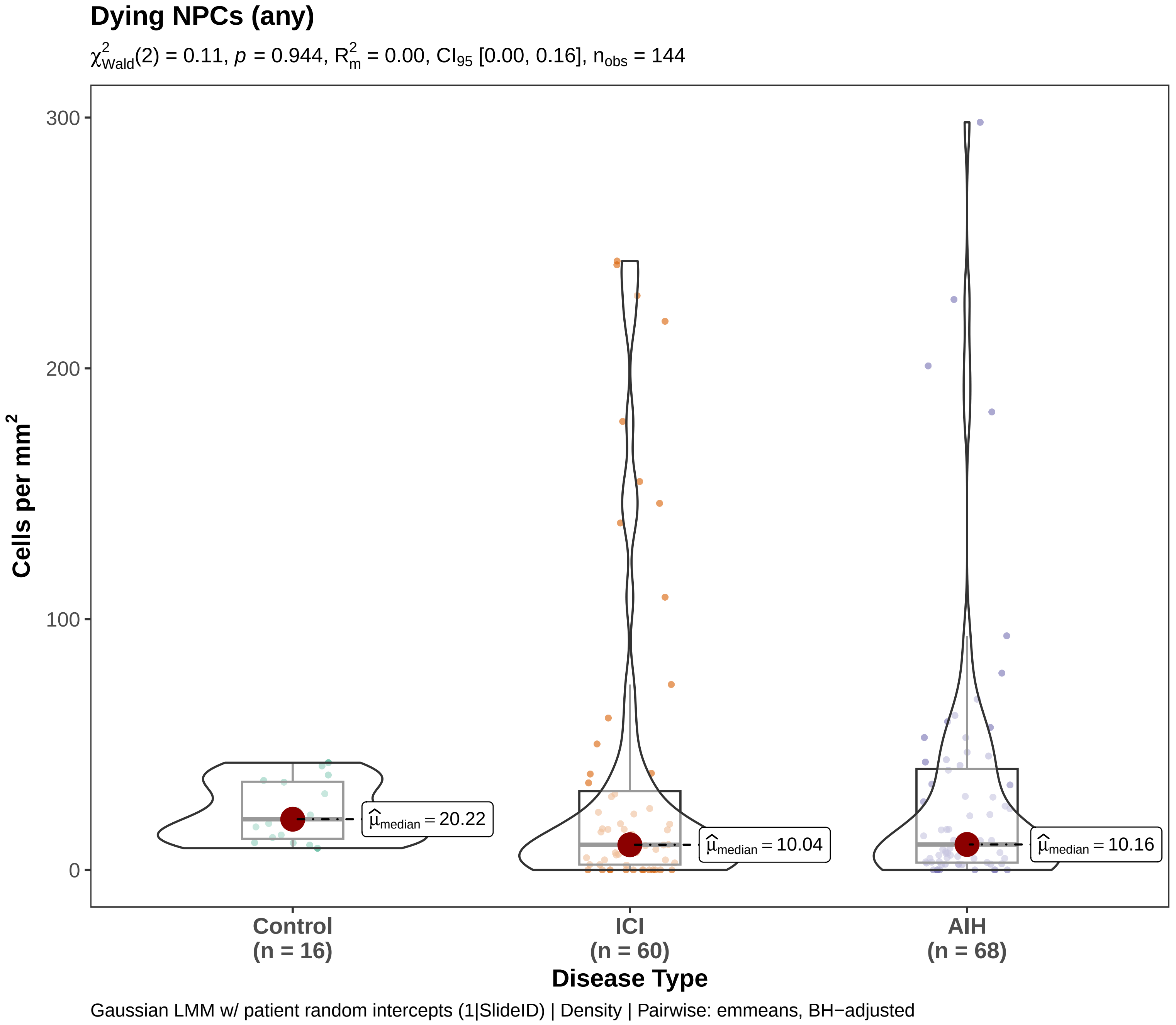

Dying NPCs (any)
$\chi^2_{Wald}(2) = 0.11$, $p = 0.944$, $R^2_m = 0.00$, $CI_{95}$ [0.00, 0.16], $n_{obs} = 144$
Cells per mm$^2$
300
200
100
0
$\hat{\mu}_{median} = 20.22$
$\hat{\mu}_{median} = 10.04$
$\hat{\mu}_{median} = 10.16$
Control
(n = 16)
ICI
(n = 60)
AIH
(n = 68)
Disease Type
Gaussian LMM w/ patient random intercepts (1|SlideID) | Density | Pairwise: emmeans, BH−adjusted

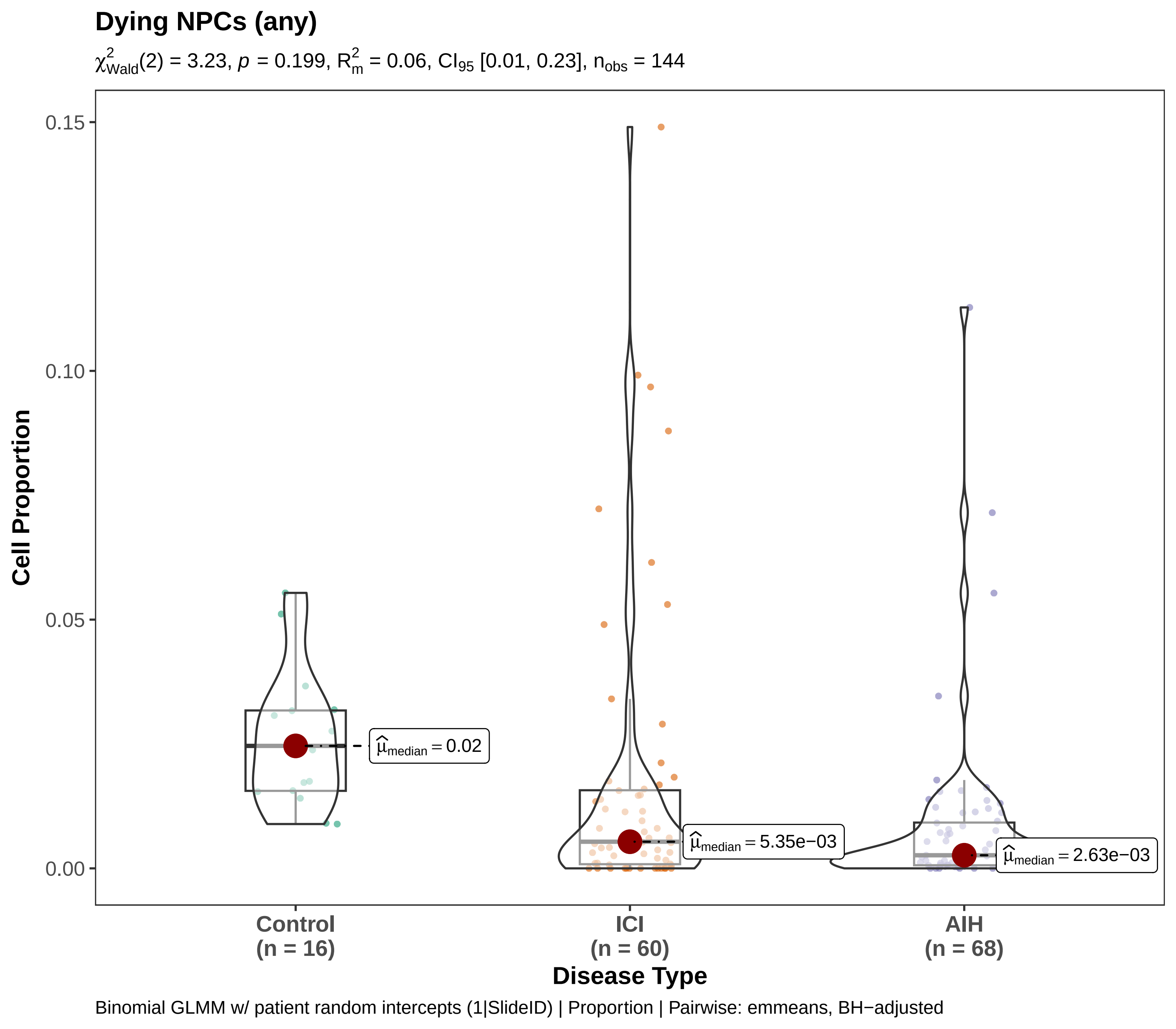
Dying NPCs (any)
$\chi^2_{Wald}(2) = 3.23$, $p = 0.199$, $R^2_m = 0.06$, $CI_{95}$ [0.01, 0.23], $n_{obs} = 144$
Cell Proportion
0.15
0.10
0.05
0.00
$\hat{\mu}_{median} = 0.02$
$\hat{\mu}_{median} = 5.35e-03$
$\hat{\mu}_{median} = 2.63e-03$
Control
(n = 16)
ICI
(n = 60)
AIH
(n = 68)
Disease Type
Binomial GLMM w/ patient random intercepts (1|SlideID) | Proportion | Pairwise: emmeans, BH−adjusted

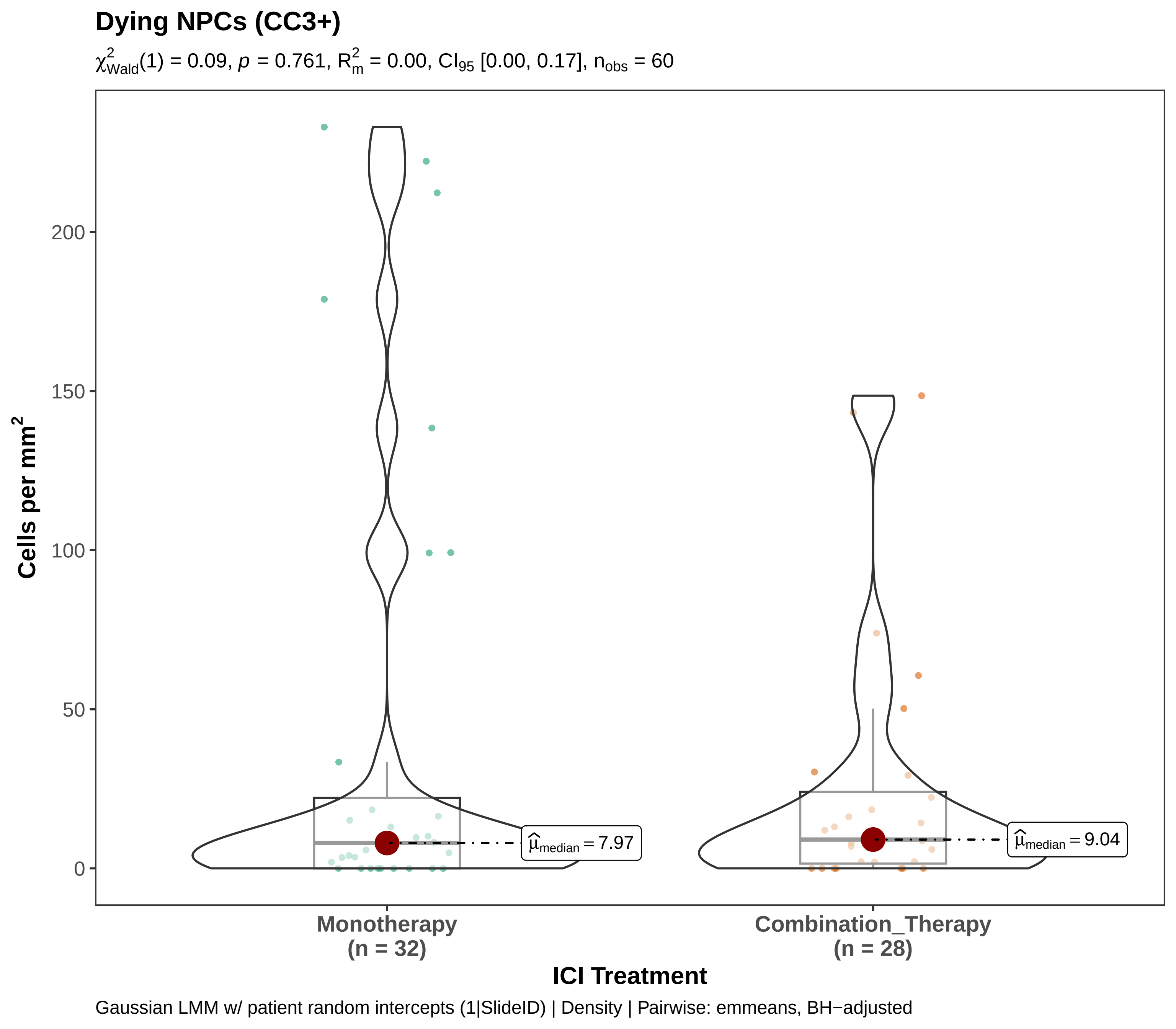

Dying NPCs (CC3+)
$\chi^2_{Wald}(1) = 0.09$, $p = 0.761$, $R^2_m = 0.00$, $CI_{95}$ [0.00, 0.17], $n_{obs} = 60$
Cells per mm$^2$
200
150
100
50
0
$\hat{\mu}_{median} = 7.97$
$\hat{\mu}_{median} = 9.04$
Monotherapy
(n = 32)
Combination_Therapy
(n = 28)
ICI Treatment
Gaussian LMM w/ patient random intercepts (1|SlideID) | Density | Pairwise: emmeans, BH−adjusted

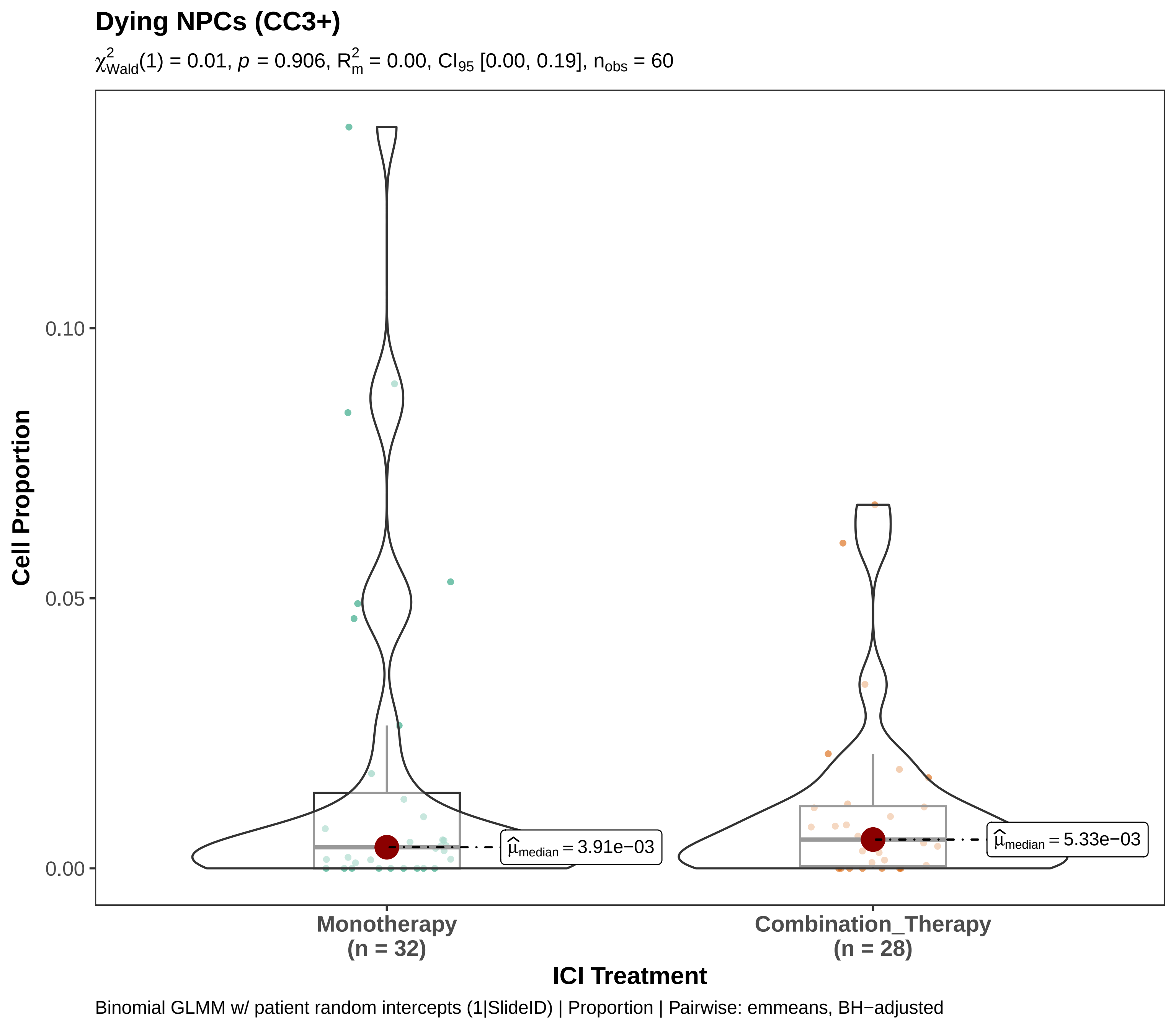
Dying NPCs (CC3+)
$\chi^2_{Wald}(1) = 0.01$, $p = 0.906$, $R^2_m = 0.00$, $CI_{95}$ [0.00, 0.19], $n_{obs} = 60$
Cell Proportion
0.10
0.05
0.00
$\widehat{\mu}_{median} = 3.91e{-}03$
$\widehat{\mu}_{median} = 5.33e{-}03$
Monotherapy
(n = 32)
Combination_Therapy
(n = 28)
ICI Treatment
Binomial GLMM w/ patient random intercepts (1|SlideID) | Proportion | Pairwise: emmeans, BH−adjusted

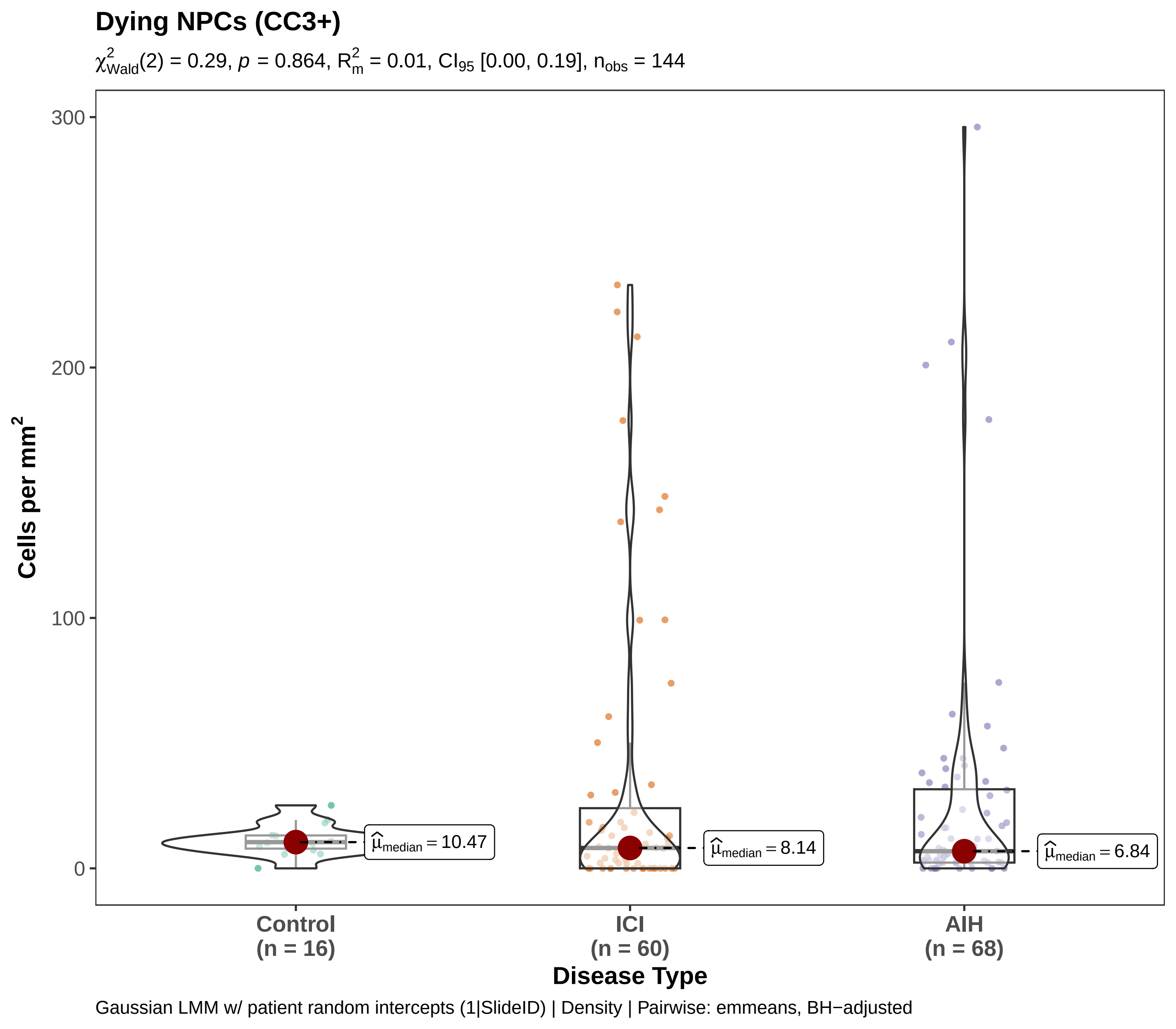

Dying NPCs (CC3+)
$\chi^2_{Wald}(2) = 0.29$, $p = 0.864$, $R^2_m = 0.01$, $CI_{95}$ [0.00, 0.19], $n_{obs} = 144$
Cells per mm$^2$
300
200
100
0
$\widehat{\mu}_{median} = 10.47$
$\widehat{\mu}_{median} = 8.14$
$\widehat{\mu}_{median} = 6.84$
Control
(n = 16)
ICI
(n = 60)
AIH
(n = 68)
Disease Type
Gaussian LMM w/ patient random intercepts (1|SlideID) | Density | Pairwise: emmeans, BH−adjusted

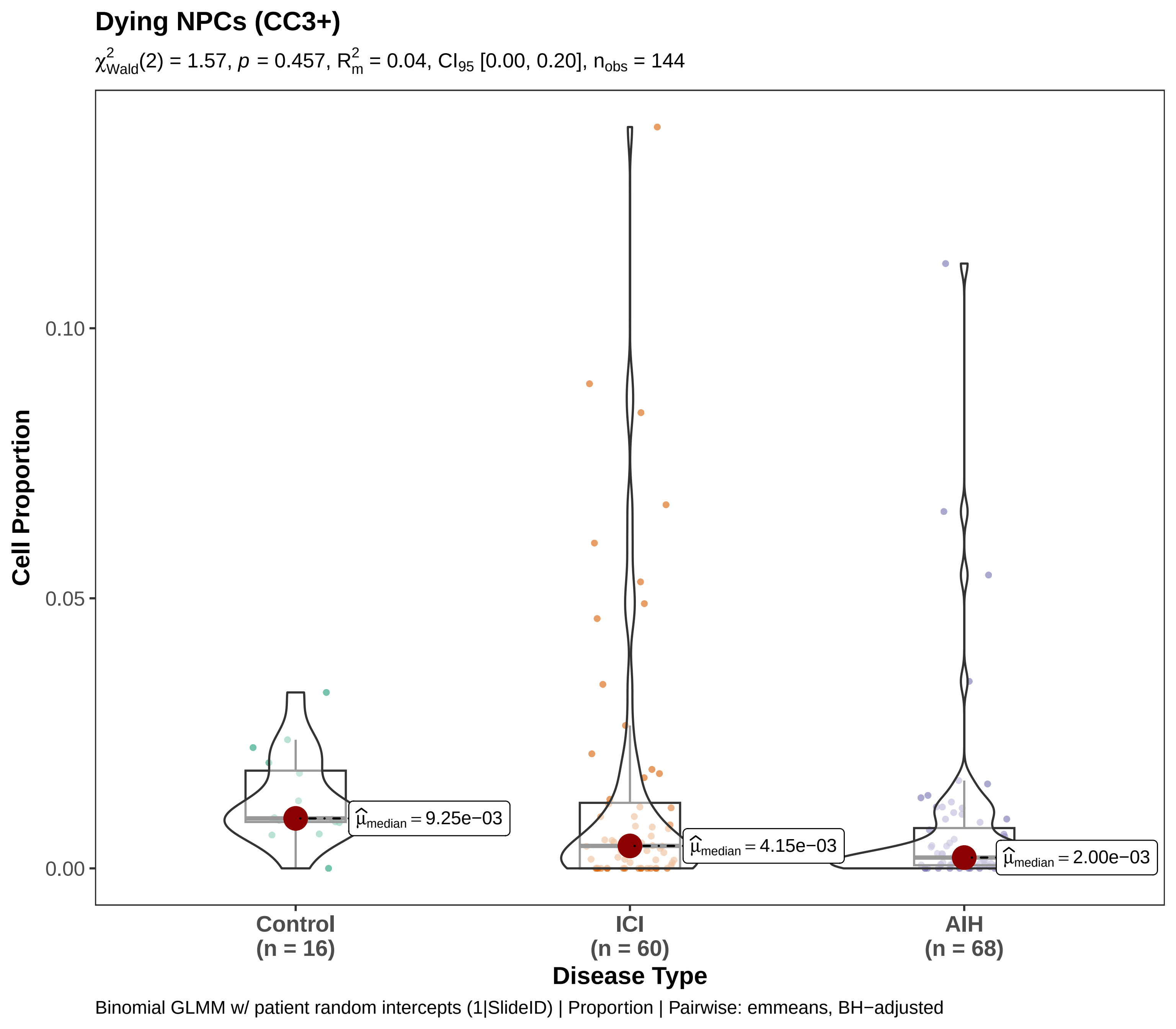
Dying NPCs (CC3+)
$\chi^2_{Wald}(2) = 1.57$, $p = 0.457$, $R^2_m = 0.04$, $CI_{95}$ [0.00, 0.20], $n_{obs} = 144$
Cell Proportion
0.10
0.05
0.00
$\hat{\mu}_{median} = 9.25e{-}03$
$\hat{\mu}_{median} = 4.15e{-}03$
$\hat{\mu}_{median} = 2.00e{-}03$
Control (n = 16)
ICI (n = 60)
AIH (n = 68)
Disease Type
Binomial GLMM w/ patient random intercepts (1|SlideID) | Proportion | Pairwise: emmeans, BH−adjusted

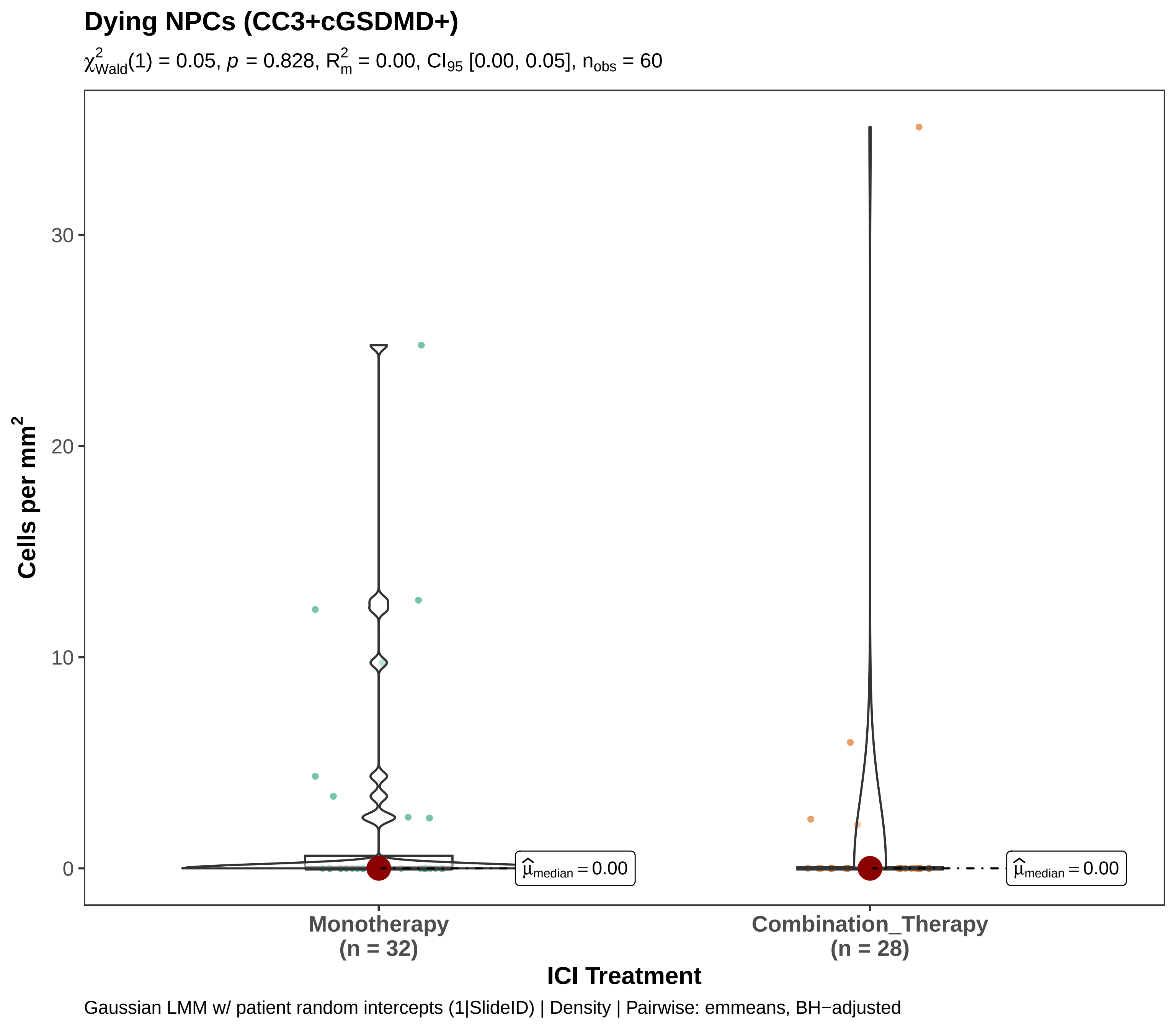

Dying NPCs (CC3+cGSDMD+)
$\chi^2_{Wald}(1) = 0.05$, $p = 0.828$, $R^2_m = 0.00$, $CI_{95}$ [0.00, 0.05], $n_{obs} = 60$
Cells per mm$^2$
30
20
10
0
$\hat{\mu}_{median} = 0.00$
$\hat{\mu}_{median} = 0.00$
Monotherapy
(n = 32)
Combination_Therapy
(n = 28)
ICI Treatment
Gaussian LMM w/ patient random intercepts (1|SlideID) | Density | Pairwise: emmeans, BH−adjusted

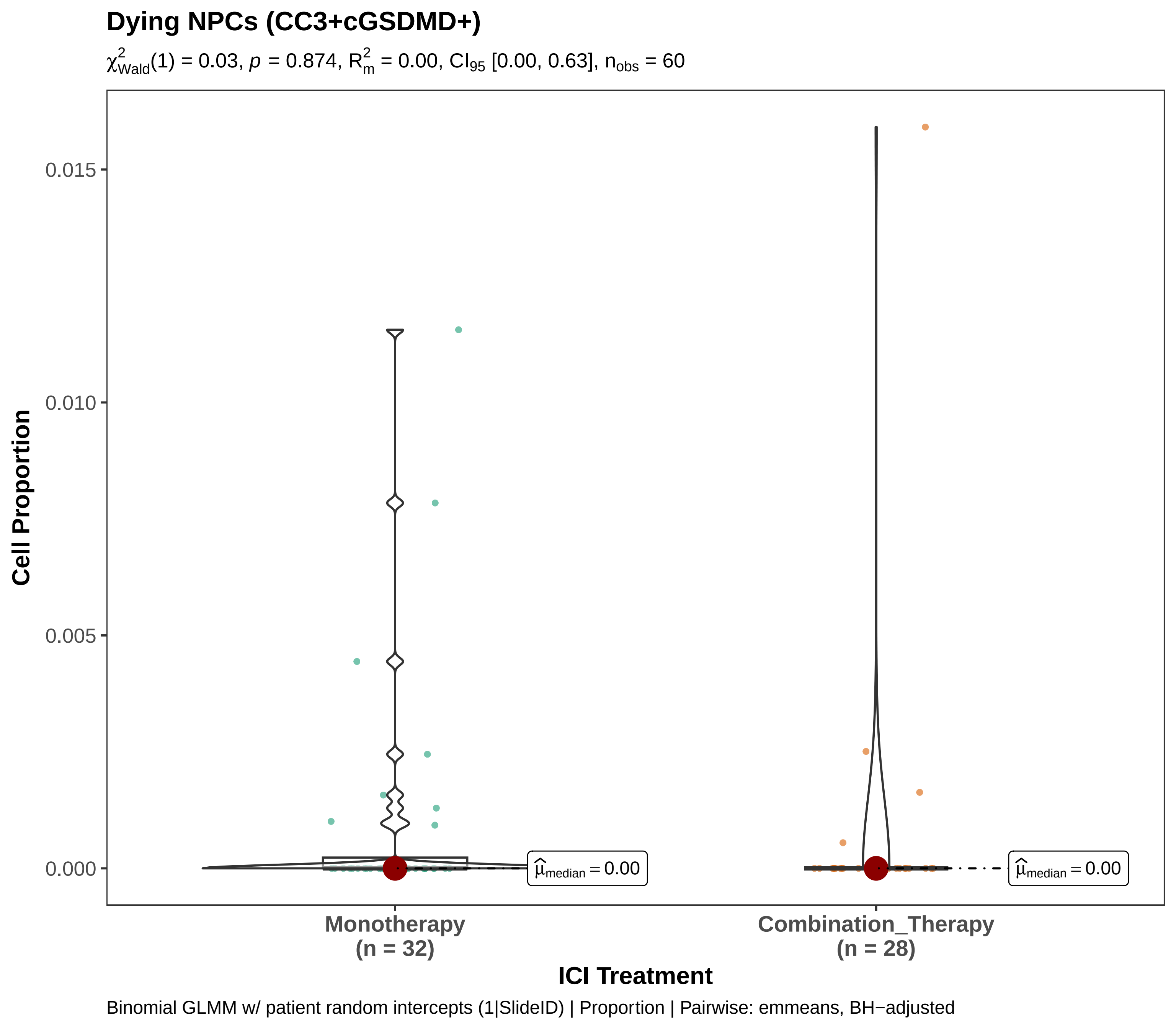
Dying NPCs (CC3+cGSDMD+)
$\chi^2_{Wald}(1) = 0.03$, $p = 0.874$, $R^2_m = 0.00$, $CI_{95}$ [0.00, 0.63], $n_{obs} = 60$
Cell Proportion
0.015
0.010
0.005
0.000
$\widehat{\mu}_{median} = 0.00$
$\widehat{\mu}_{median} = 0.00$
Monotherapy
(n = 32)
Combination_Therapy
(n = 28)
ICI Treatment
Binomial GLMM w/ patient random intercepts (1|SlideID) | Proportion | Pairwise: emmeans, BH−adjusted

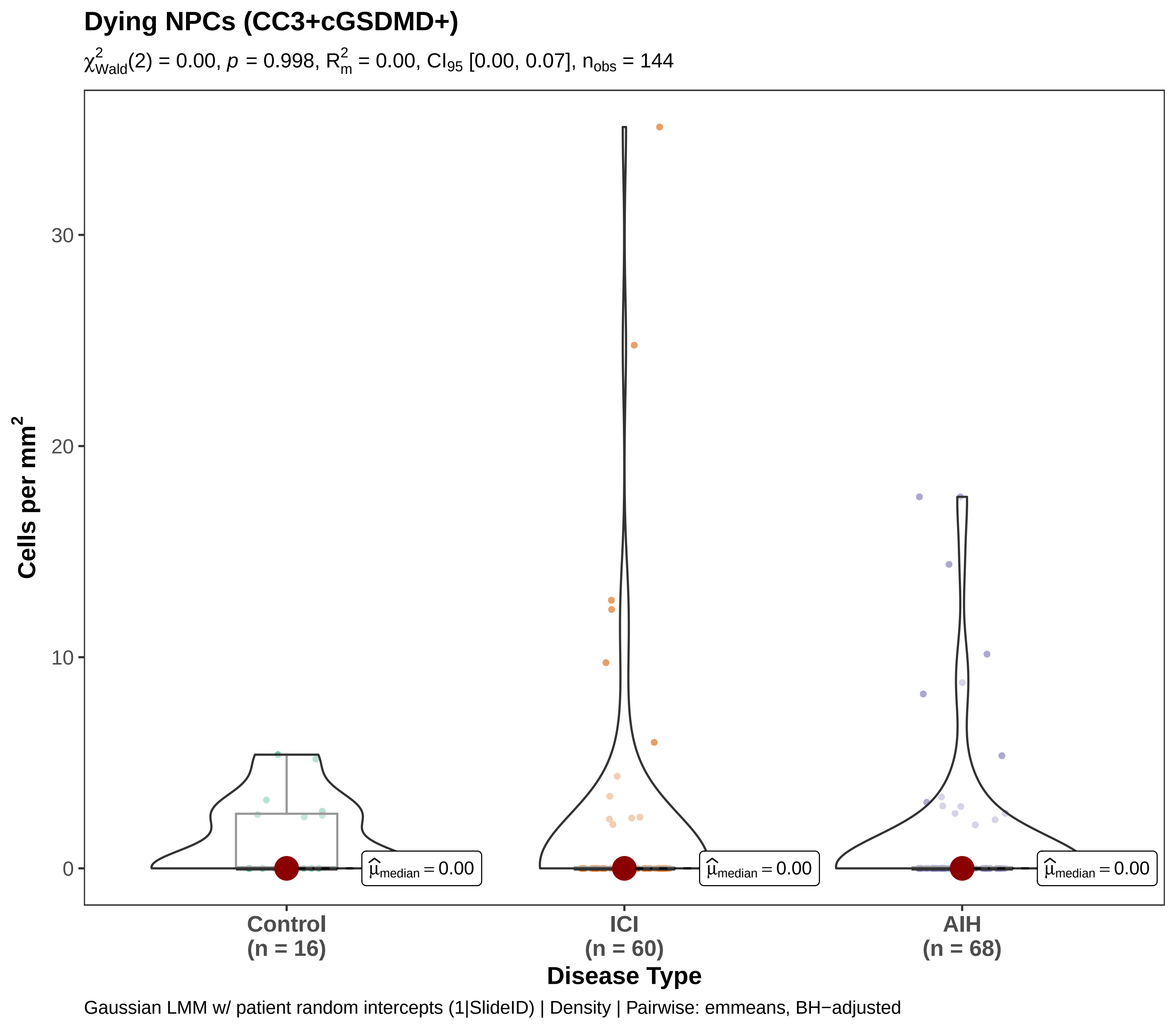

Dying NPCs (CC3+cGSDMD+)
$\chi^2_{Wald}(2) = 0.00$, $p = 0.998$, $R^2_m = 0.00$, $CI_{95}$ [0.00, 0.07], $n_{obs} = 144$
Cells per mm²
30
20
10
0
$\hat{\mu}_{median} = 0.00$
$\hat{\mu}_{median} = 0.00$
$\hat{\mu}_{median} = 0.00$
Control (n = 16)
ICI (n = 60)
AIH (n = 68)
Disease Type
Gaussian LMM w/ patient random intercepts (1|SlideID) | Density | Pairwise: emmeans, BH−adjusted

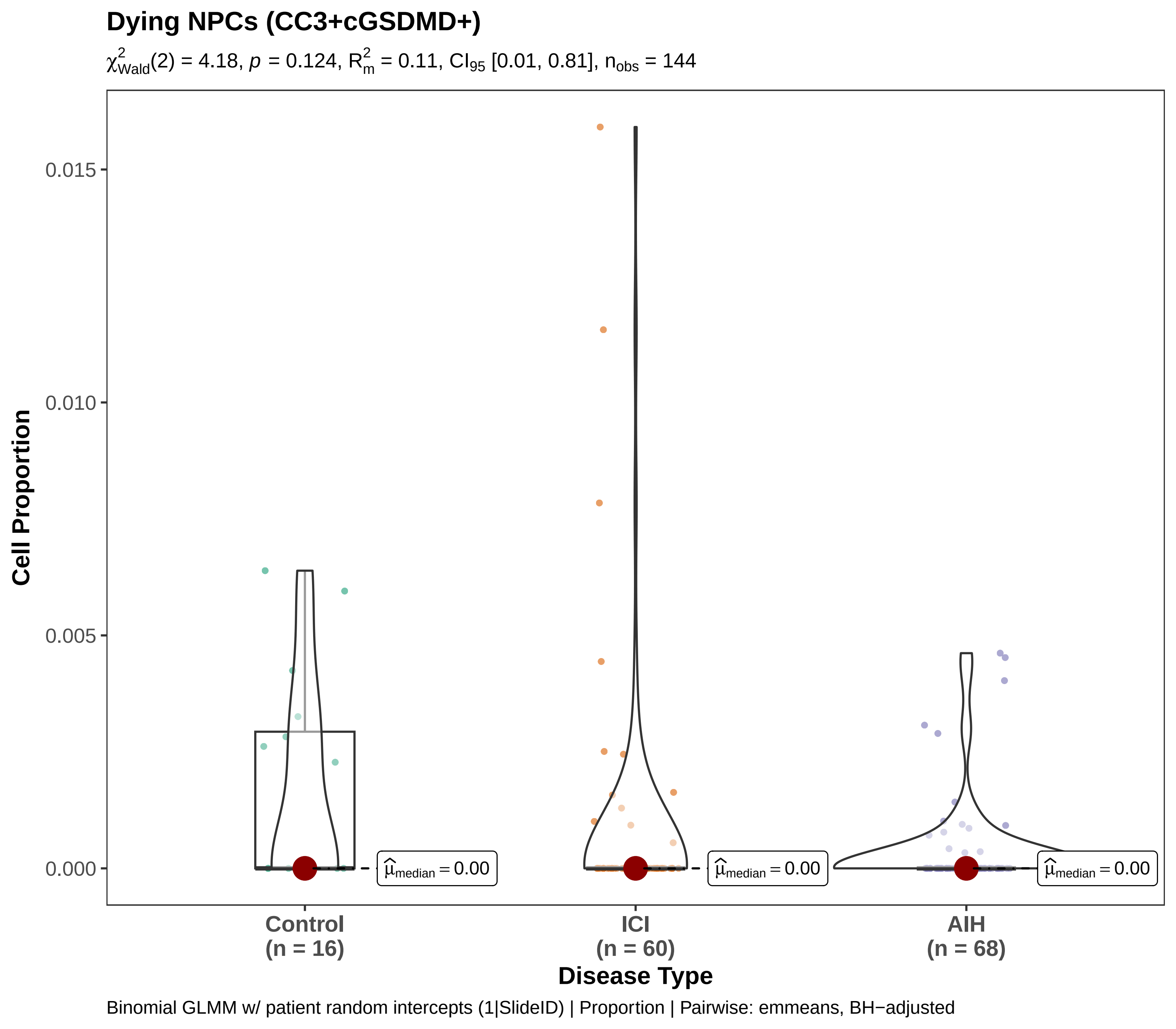
Dying NPCs (CC3+cGSDMD+)
$\chi^2_{Wald}(2) = 4.18$, $p = 0.124$, $R^2_m = 0.11$, $CI_{95}$ [0.01, 0.81], $n_{obs} = 144$
Cell Proportion
0.015
0.010
0.005
0.000
$\hat{\mu}_{median} = 0.00$
$\hat{\mu}_{median} = 0.00$
$\hat{\mu}_{median} = 0.00$
Control
(n = 16)
ICI
(n = 60)
AIH
(n = 68)
Disease Type
Binomial GLMM w/ patient random intercepts (1|SlideID) | Proportion | Pairwise: emmeans, BH−adjusted

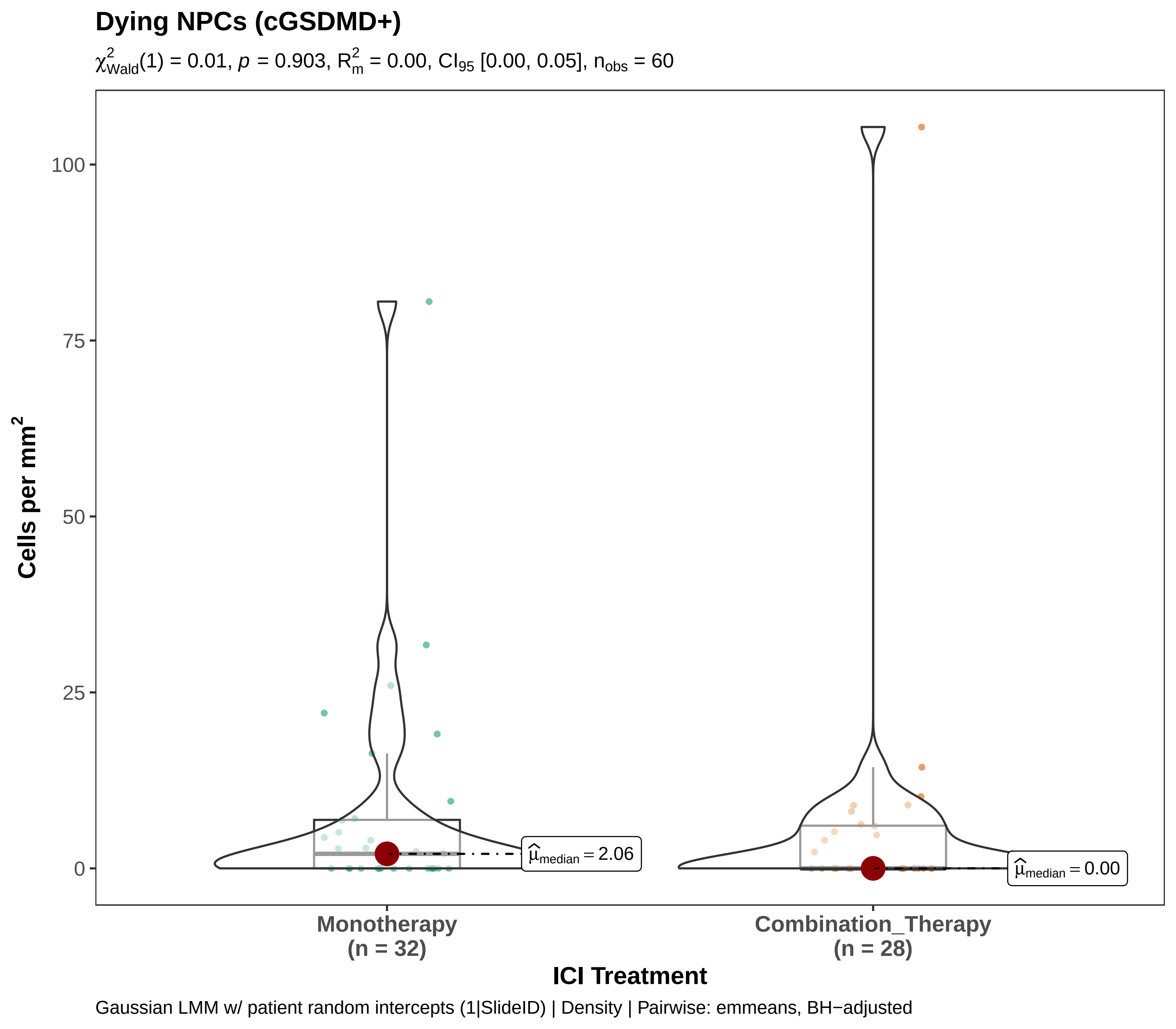

Dying NPCs (cGSDMD+)
$\chi^2_{Wald}(1) = 0.01$, $p = 0.903$, $R^2_m = 0.00$, $CI_{95}$ [0.00, 0.05], $n_{obs} = 60$
100
75
50
25
0
Cells per mm$^2$
$\hat{\mu}_{median} = 2.06$
$\hat{\mu}_{median} = 0.00$
Monotherapy
(n = 32)
Combination_Therapy
(n = 28)
ICI Treatment
Gaussian LMM w/ patient random intercepts (1|SlideID) | Density | Pairwise: emmeans, BH−adjusted

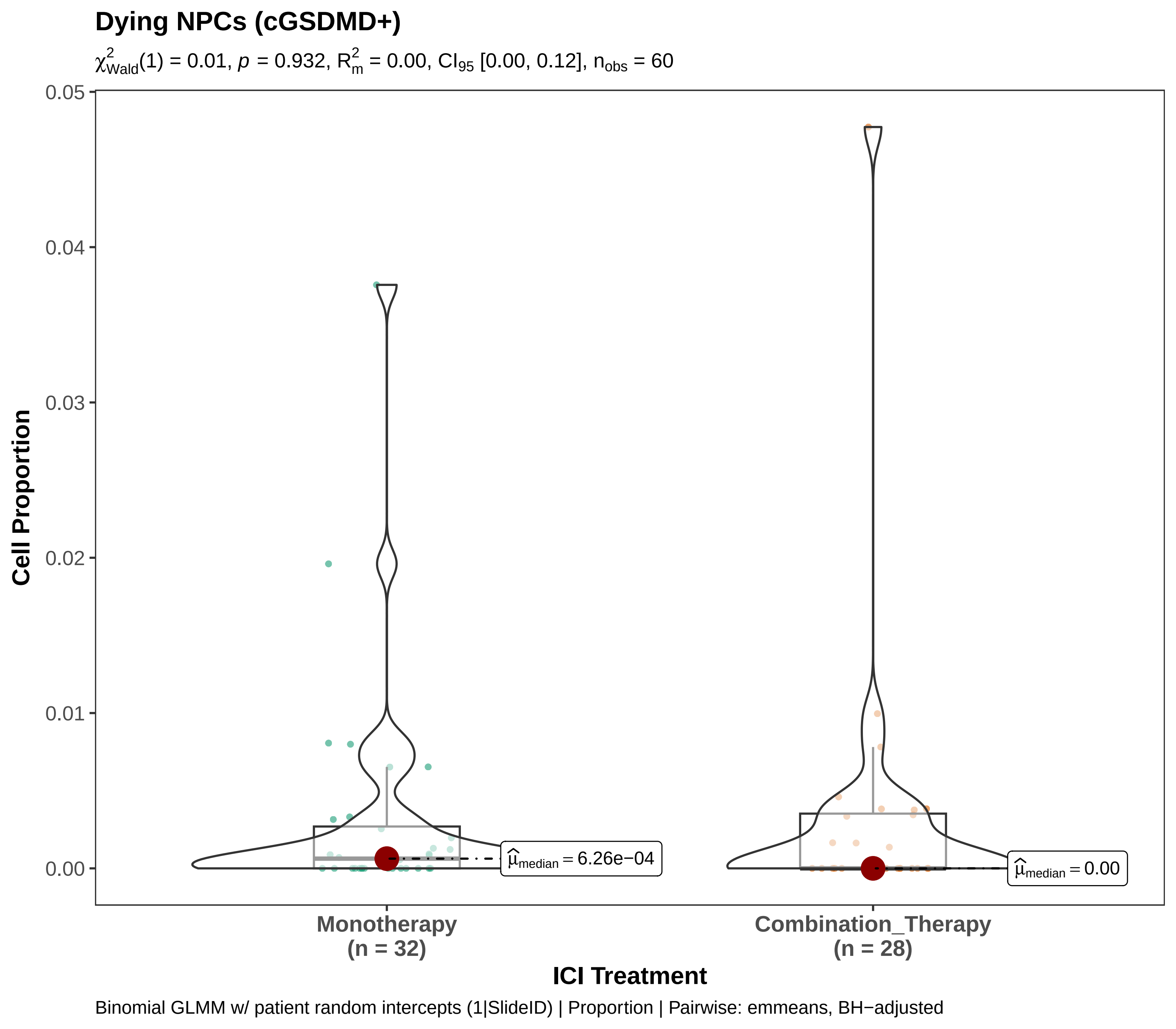
Dying NPCs (cGSDMD+)
$\chi^2_{Wald}(1) = 0.01$, $p = 0.932$, $R^2_m = 0.00$, $CI_{95}$ [0.00, 0.12], $n_{obs} = 60$
Cell Proportion
0.05
0.04
0.03
0.02
0.01
0.00
$\widehat{\mu}_{median} = 6.26e-04$
$\widehat{\mu}_{median} = 0.00$
Monotherapy
(n = 32)
Combination_Therapy
(n = 28)
ICI Treatment
Binomial GLMM w/ patient random intercepts (1|SlideID) | Proportion | Pairwise: emmeans, BH−adjusted

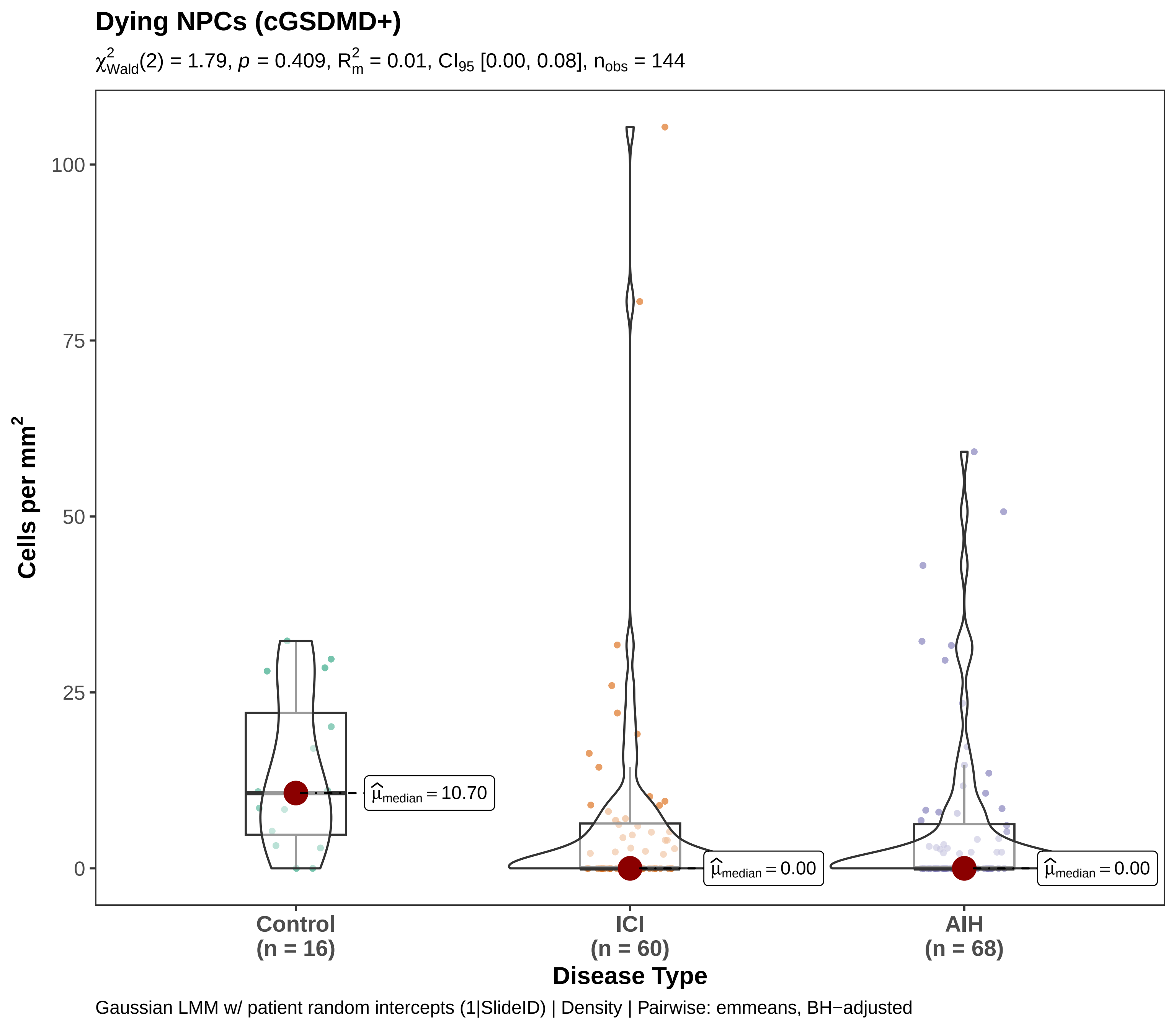
Dying NPCs (cGSDMD+)
$\chi^2_{Wald}(2) = 1.79$, $p = 0.409$, $R^2_m = 0.01$, $CI_{95}$ [0.00, 0.08], $n_{obs} = 144$
Cells per mm$^2$
100
75
50
25
0
$\widehat{\mu}_{median} = 10.70$
$\widehat{\mu}_{median} = 0.00$
$\widehat{\mu}_{median} = 0.00$
Control
(n = 16)
ICI
(n = 60)
AIH
(n = 68)
Disease Type
Gaussian LMM w/ patient random intercepts (1|SlideID) | Density | Pairwise: emmeans, BH−adjusted

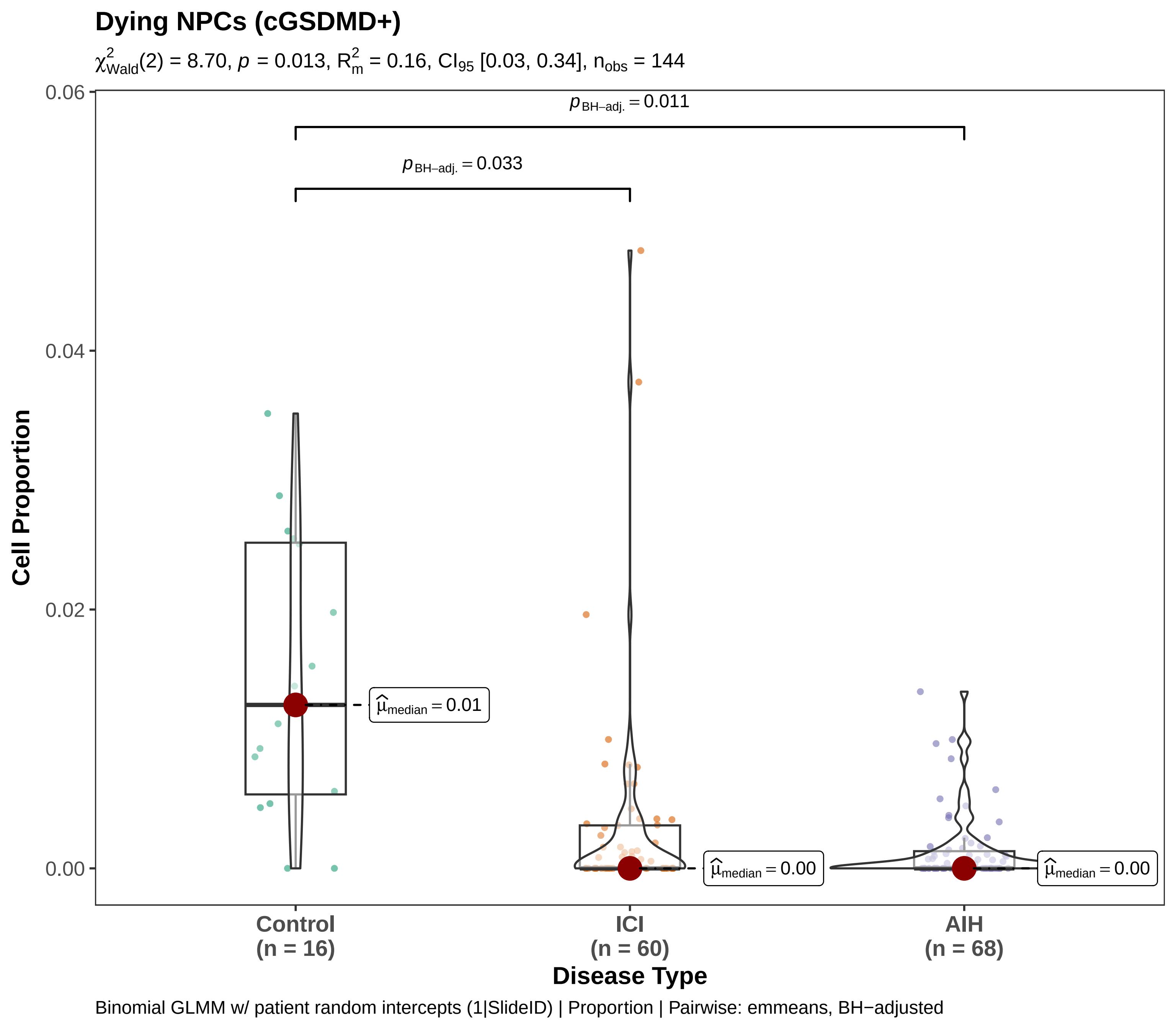
Dying NPCs (cGSDMD+)
$\chi^2_{Wald}(2) = 8.70$, $p = 0.013$, $R^2_m = 0.16$, $CI_{95}$ [0.03, 0.34], $n_{obs} = 144$
$p_{BH-adj.} = 0.011$
$p_{BH-adj.} = 0.033$
$\hat{\mu}_{median} = 0.01$
$\hat{\mu}_{median} = 0.00$
$\hat{\mu}_{median} = 0.00$
Cell Proportion
0.06
0.04
0.02
0.00
Control
(n = 16)
ICI
(n = 60)
AIH
(n = 68)
Disease Type
Binomial GLMM w/ patient random intercepts (1|SlideID) | Proportion | Pairwise: emmeans, BH−adjusted
