## Supplementary material for "Macrophage-CD8⁺ T Cell Spatial Coupling Defines an Innate-Adaptive Injury Niche in Human Checkpoint Inhibitor Hepatotoxicity": Figure S-7

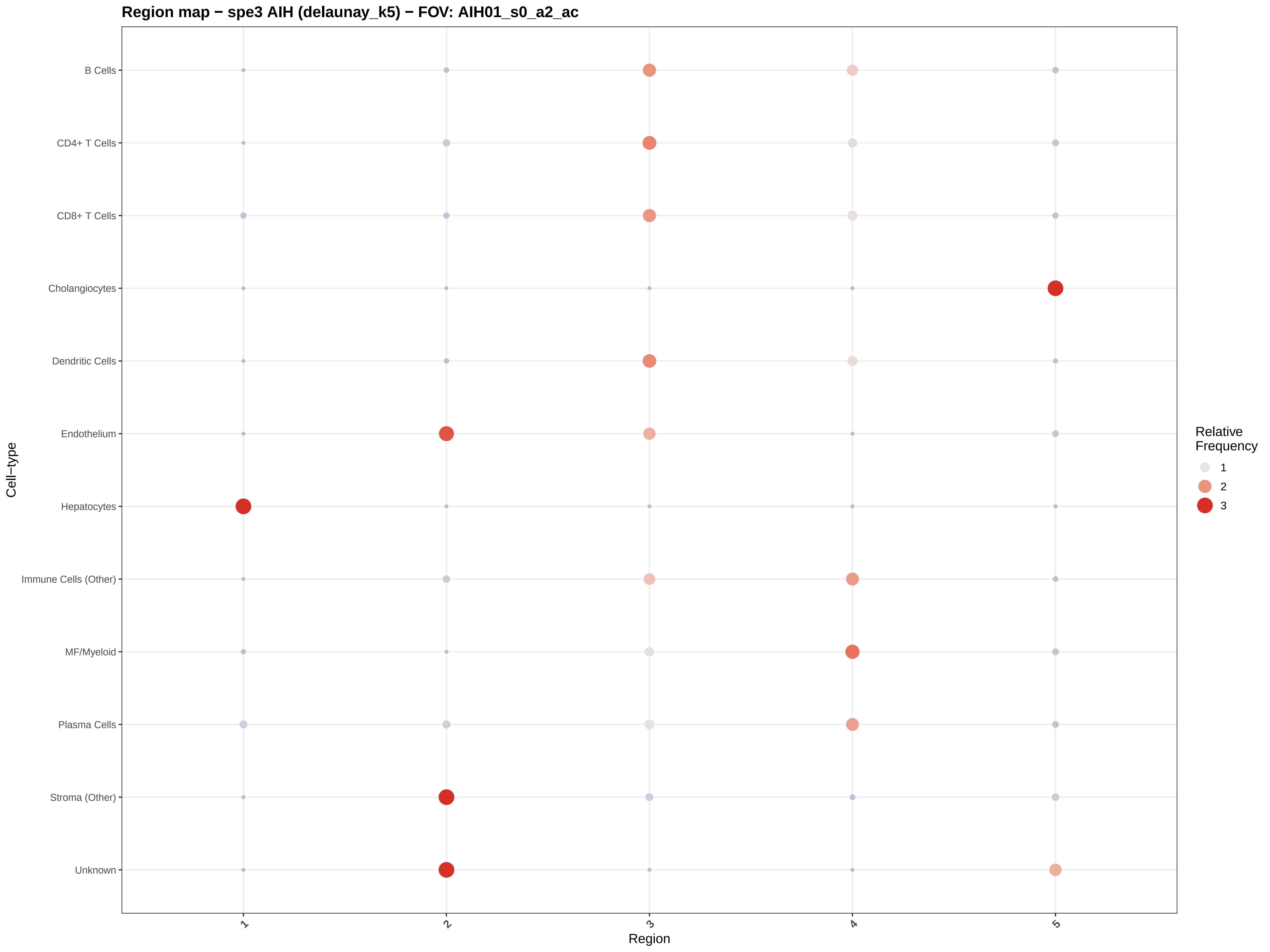
Region map − spe3 AIH (delaunay_k5) − FOV: AIH01_s0_a2_ac
B Cells
CD4+ T Cells
CD8+ T Cells
Cholangiocytes
Dendritic Cells
Endothelium
Hepatocytes
Immune Cells (Other)
MF/Myeloid
Plasma Cells
Stroma (Other)
Unknown
Cell−type
1
2
3
4
5
Region
Relative Frequency
1
2
3

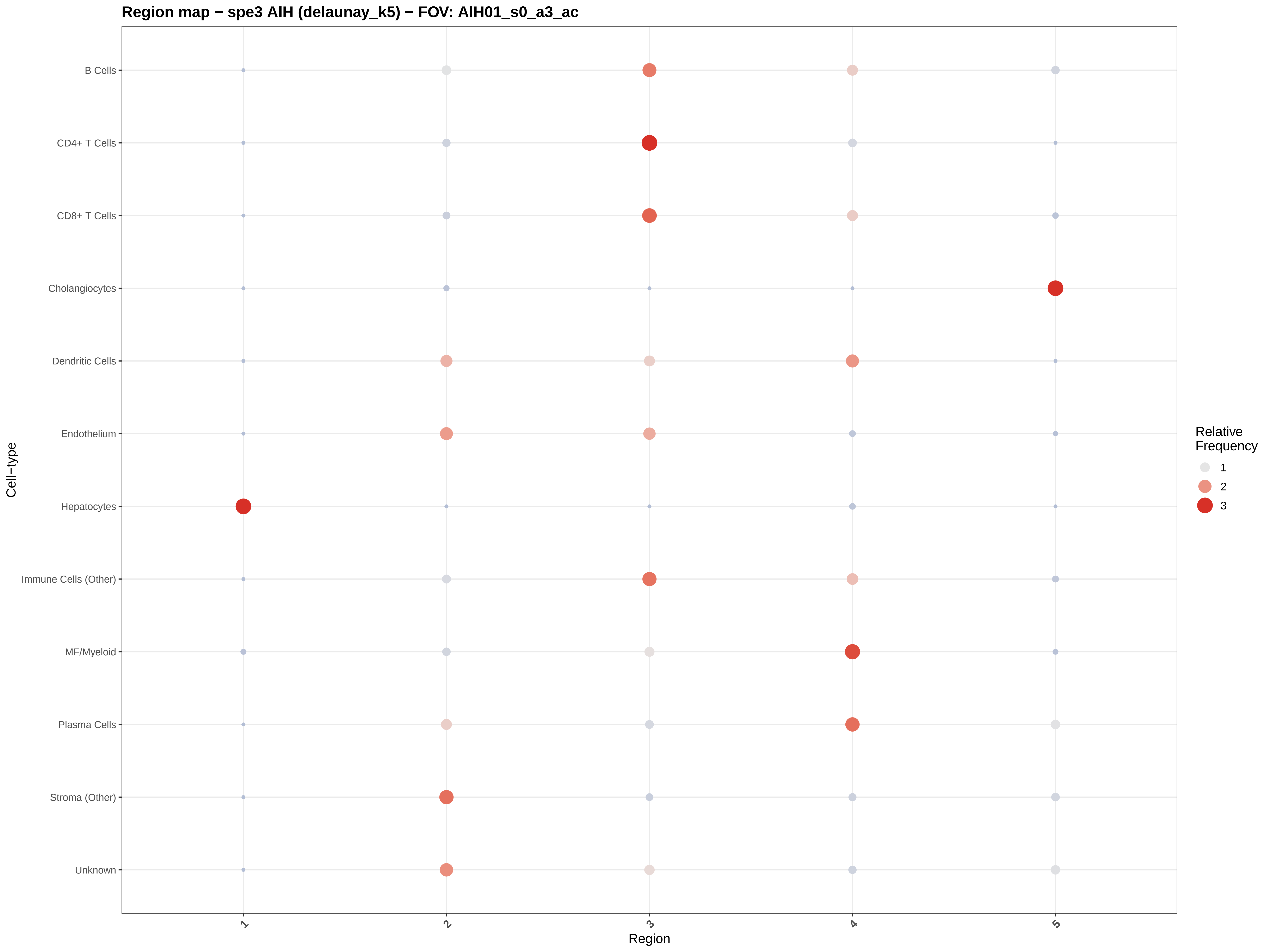

Region map − spe3 AIH (delaunay_k5) − FOV: AIH01_s0_a3_ac
B Cells
CD4+ T Cells
CD8+ T Cells
Cholangiocytes
Dendritic Cells
Endothelium
Hepatocytes
Immune Cells (Other)
MF/Myeloid
Plasma Cells
Stroma (Other)
Unknown
Cell−type
1
2
3
4
5
Region
Relative Frequency
1
2
3

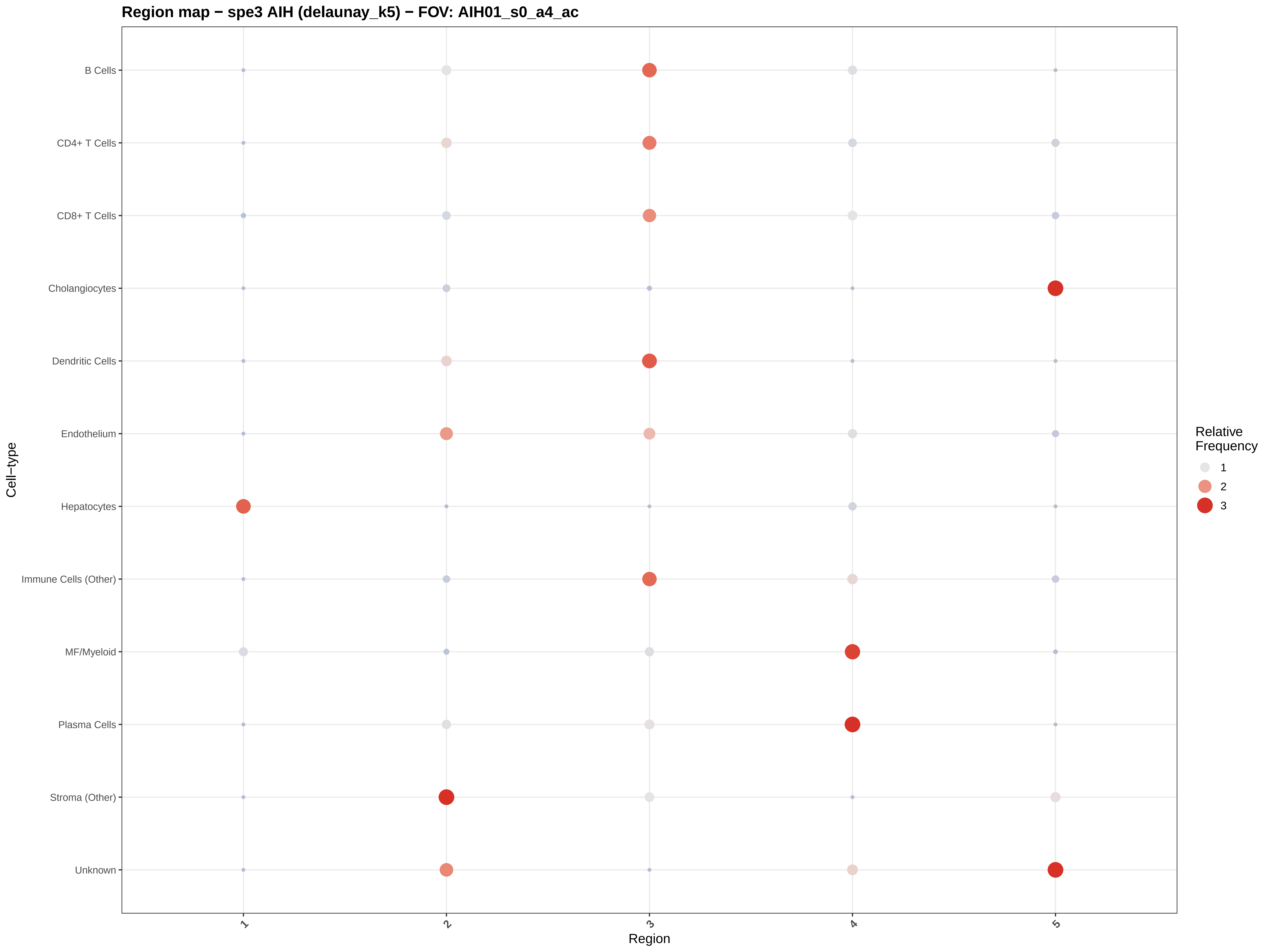

Region map − spe3 AIH (delaunay_k5) − FOV: AIH01_s0_a4_ac
B Cells
CD4+ T Cells
CD8+ T Cells
Cholangiocytes
Dendritic Cells
Endothelium
Hepatocytes
Immune Cells (Other)
MF/Myeloid
Plasma Cells
Stroma (Other)
Unknown
Cell−type
1
2
3
4
5
Region
Relative Frequency
1
2
3

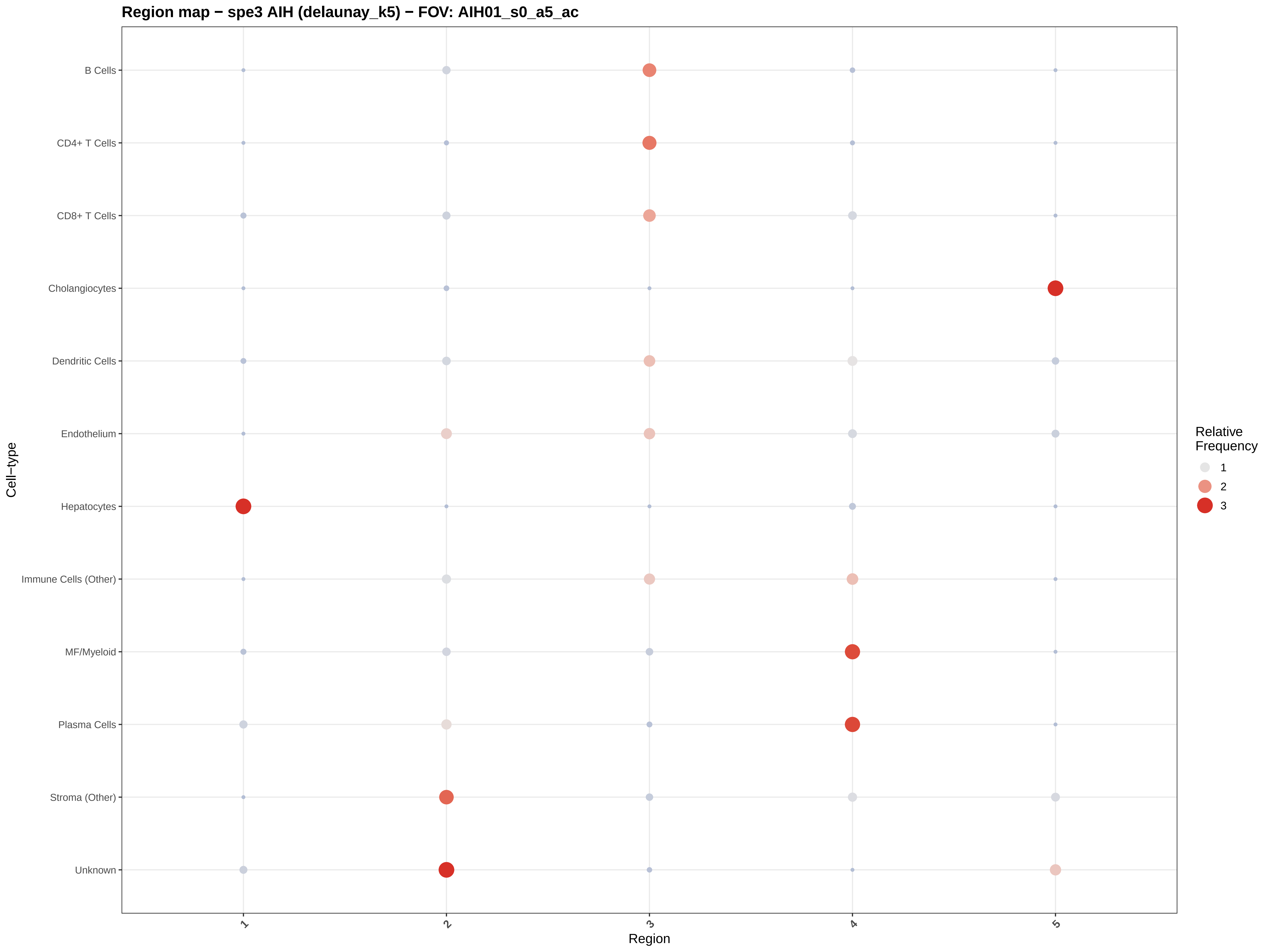
Region map − spe3 AIH (delaunay_k5) − FOV: AIH01_s0_a5_ac
B Cells
CD4+ T Cells
CD8+ T Cells
Cholangiocytes
Dendritic Cells
Endothelium
Hepatocytes
Immune Cells (Other)
MF/Myeloid
Plasma Cells
Stroma (Other)
Unknown
Cell−type
1
2
3
4
5
Region
Relative Frequency
1
2
3

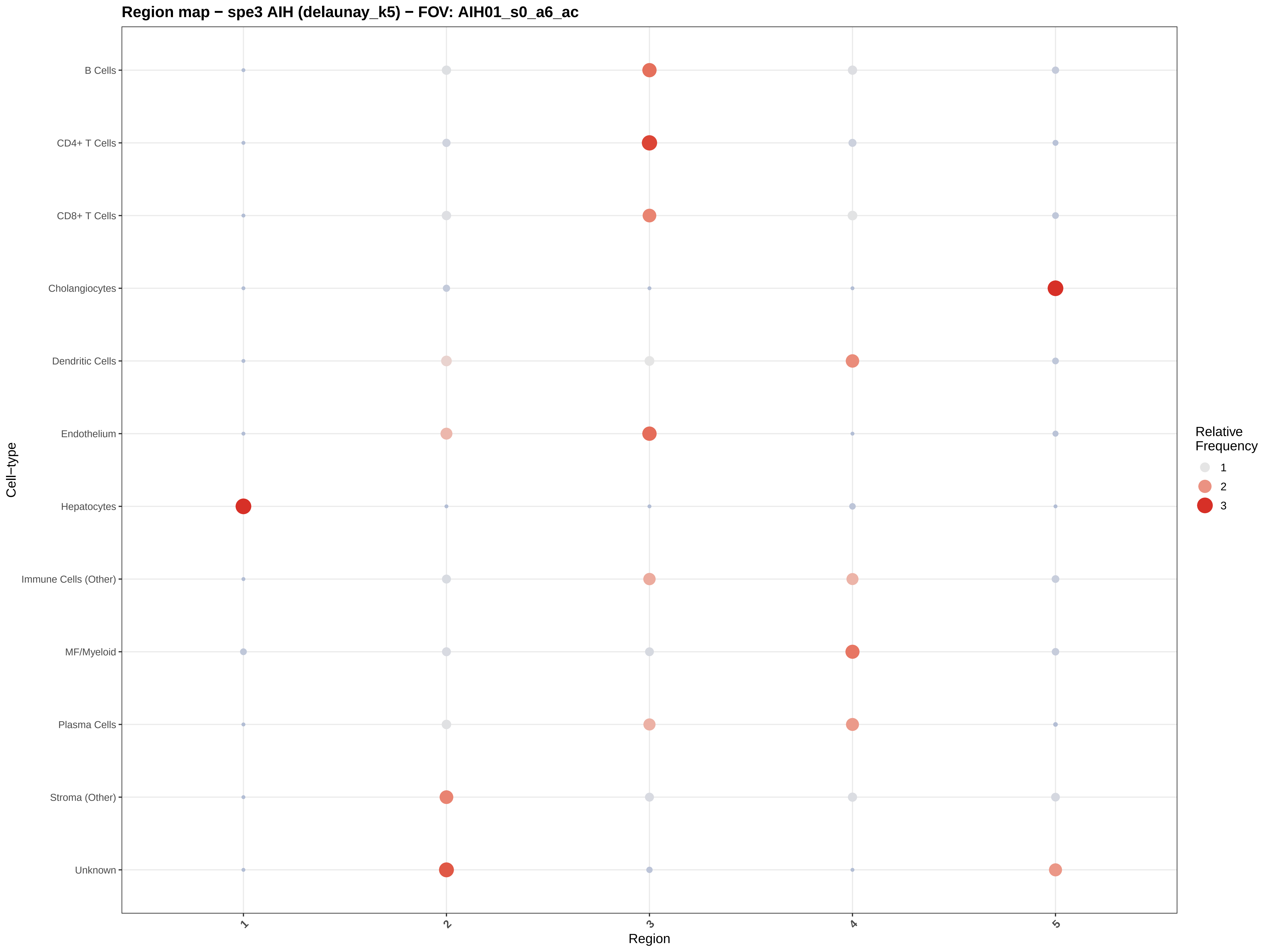

Region map − spe3 AIH (delaunay_k5) − FOV: AIH01_s0_a6_ac
B Cells
CD4+ T Cells
CD8+ T Cells
Cholangiocytes
Dendritic Cells
Endothelium
Hepatocytes
Immune Cells (Other)
MF/Myeloid
Plasma Cells
Stroma (Other)
Unknown
Cell−type
1
2
3
4
5
Region
Relative Frequency
1
2
3

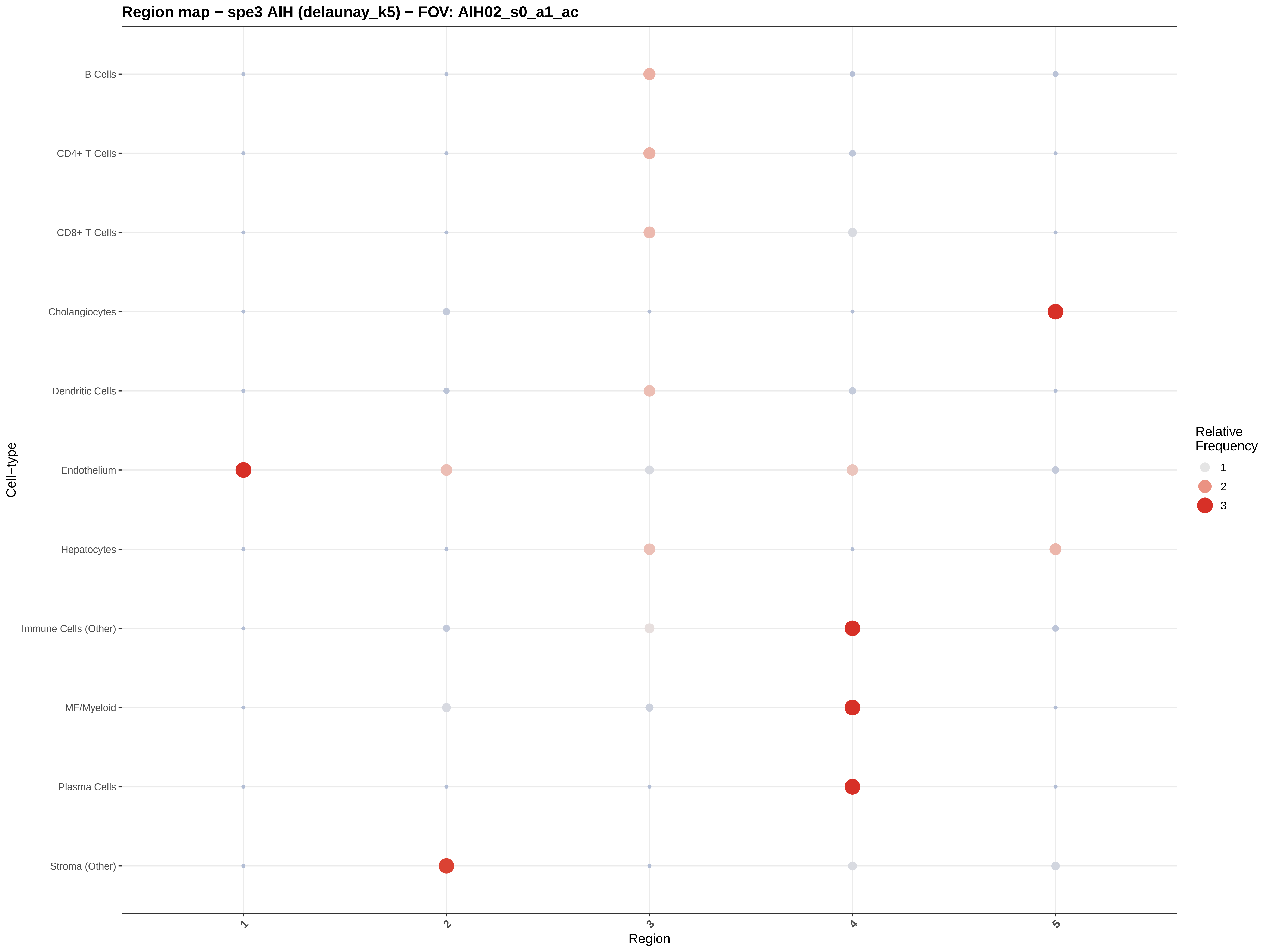
Region map − spe3 AIH (delaunay_k5) − FOV: AIH02_s0_a1_ac
B Cells
CD4+ T Cells
CD8+ T Cells
Cholangiocytes
Dendritic Cells
Endothelium
Hepatocytes
Immune Cells (Other)
MF/Myeloid
Plasma Cells
Stroma (Other)
Cell−type
1
2
3
4
5
Region
Relative Frequency
1
2
3

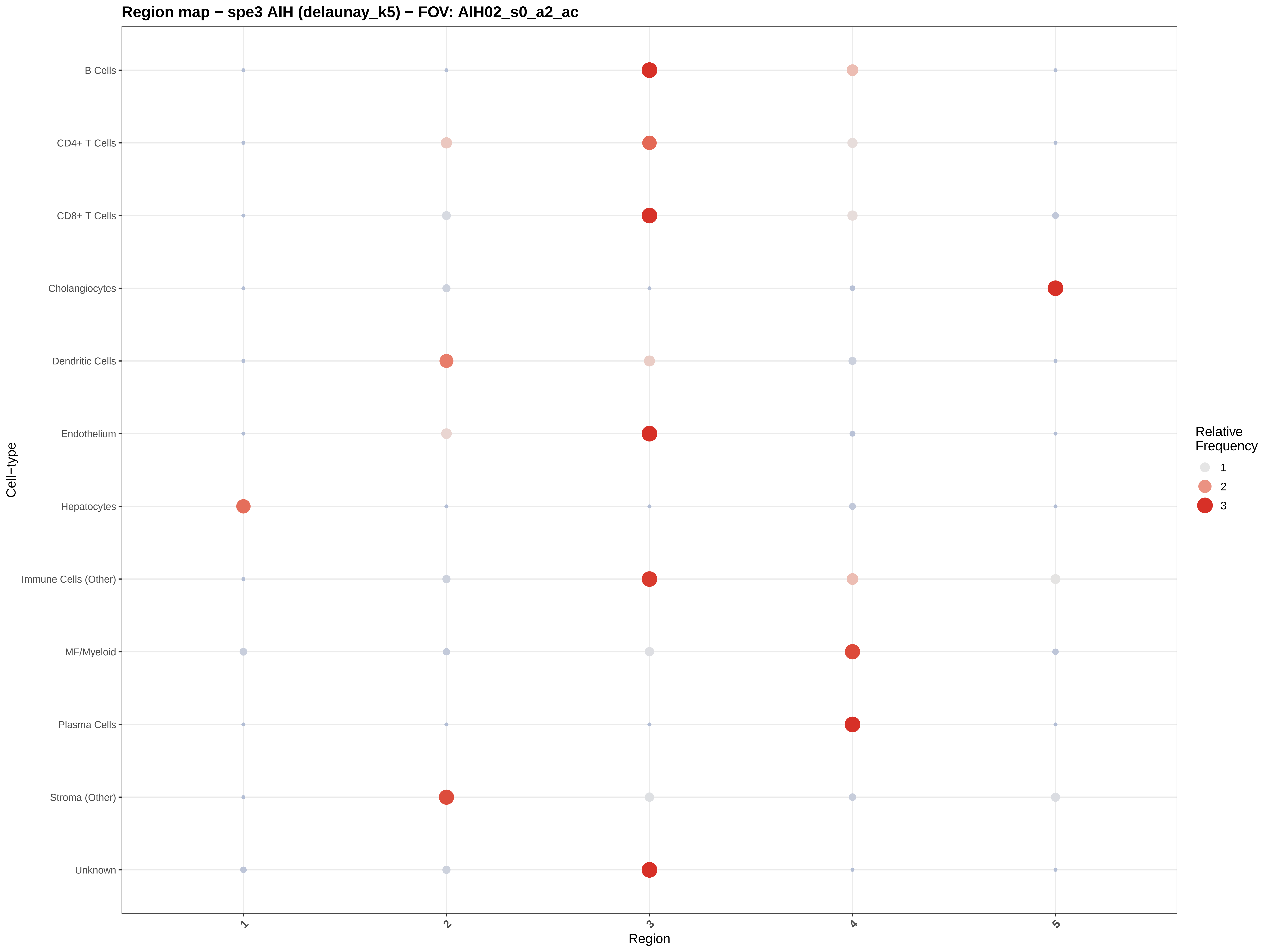

Region map − spe3 AIH (delaunay_k5) − FOV: AIH02_s0_a2_ac
B Cells
CD4+ T Cells
CD8+ T Cells
Cholangiocytes
Dendritic Cells
Endothelium
Hepatocytes
Immune Cells (Other)
MF/Myeloid
Plasma Cells
Stroma (Other)
Unknown
Cell−type
1
2
3
4
5
Region
Relative Frequency
1
2
3

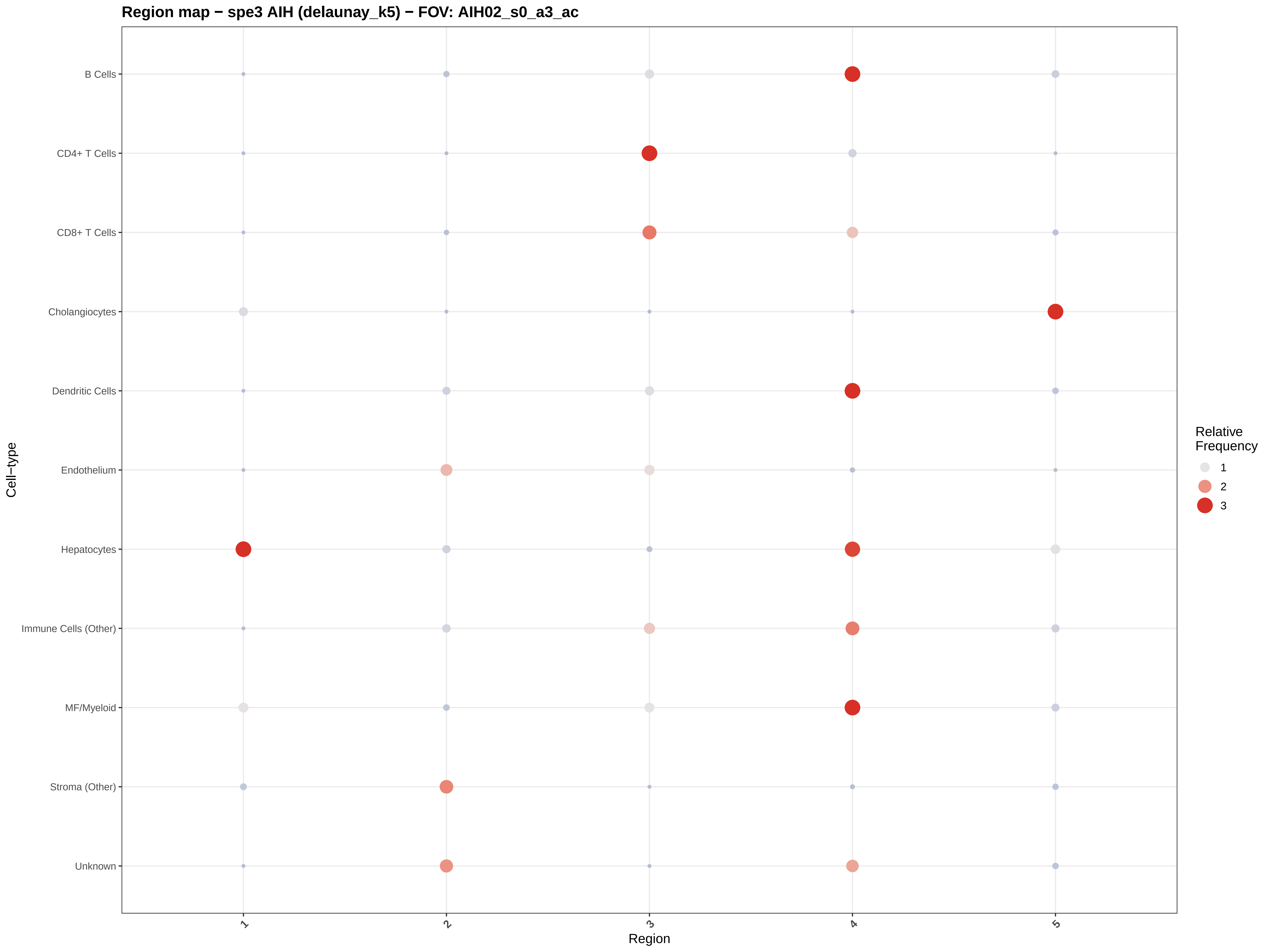

Region map − spe3 AIH (delaunay_k5) − FOV: AIH02_s0_a3_ac
B Cells
CD4+ T Cells
CD8+ T Cells
Cholangiocytes
Dendritic Cells
Endothelium
Hepatocytes
Immune Cells (Other)
MF/Myeloid
Stroma (Other)
Unknown
Cell−type
1
2
3
4
5
Region
Relative Frequency
1
2
3

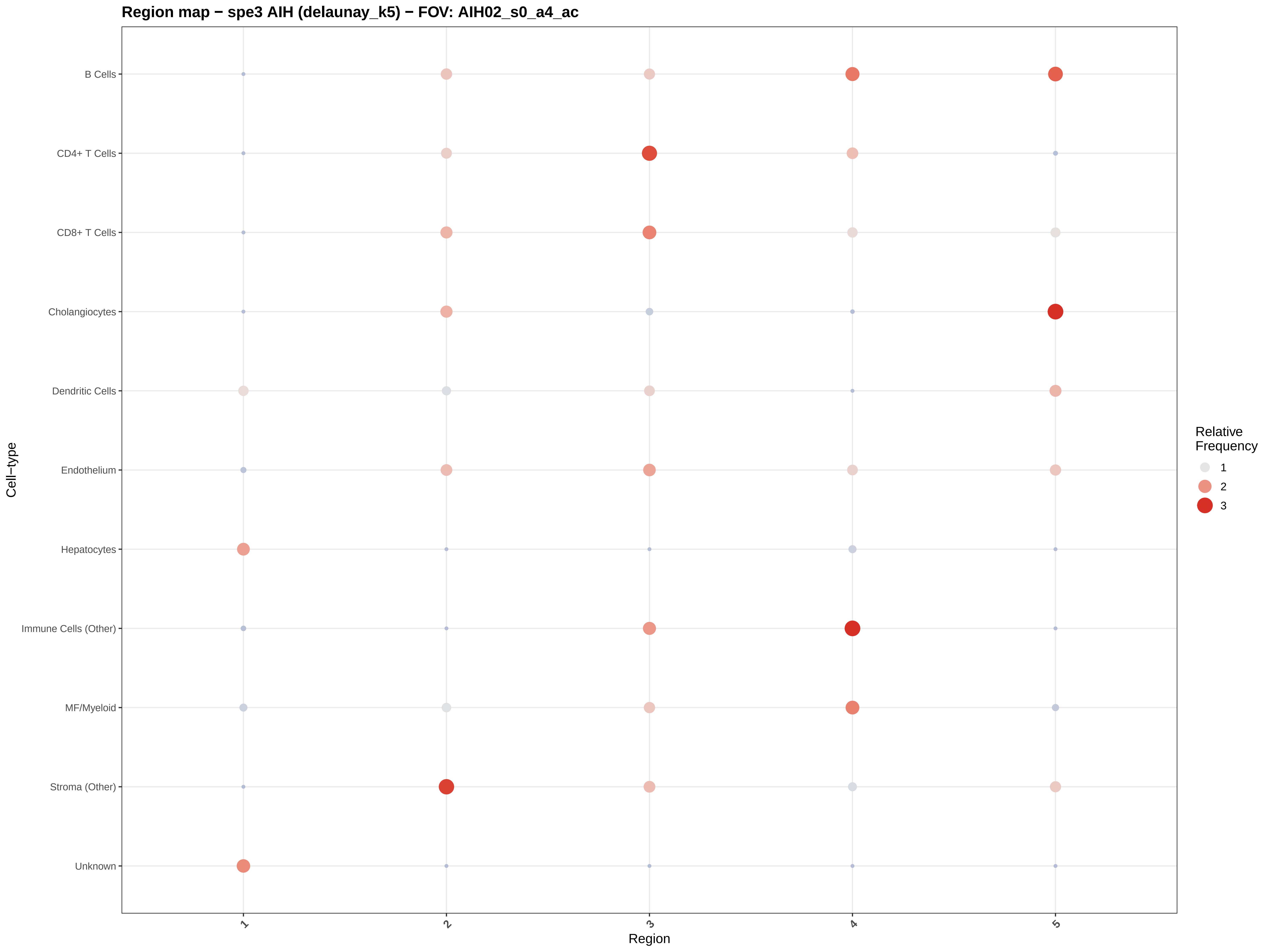

Region map − spe3 AIH (delaunay_k5) − FOV: AIH02_s0_a4_ac
B Cells
CD4+ T Cells
CD8+ T Cells
Cholangiocytes
Dendritic Cells
Endothelium
Hepatocytes
Immune Cells (Other)
MF/Myeloid
Stroma (Other)
Unknown
Cell−type
1
2
3
4
5
Region
Relative
Frequency
1
2
3

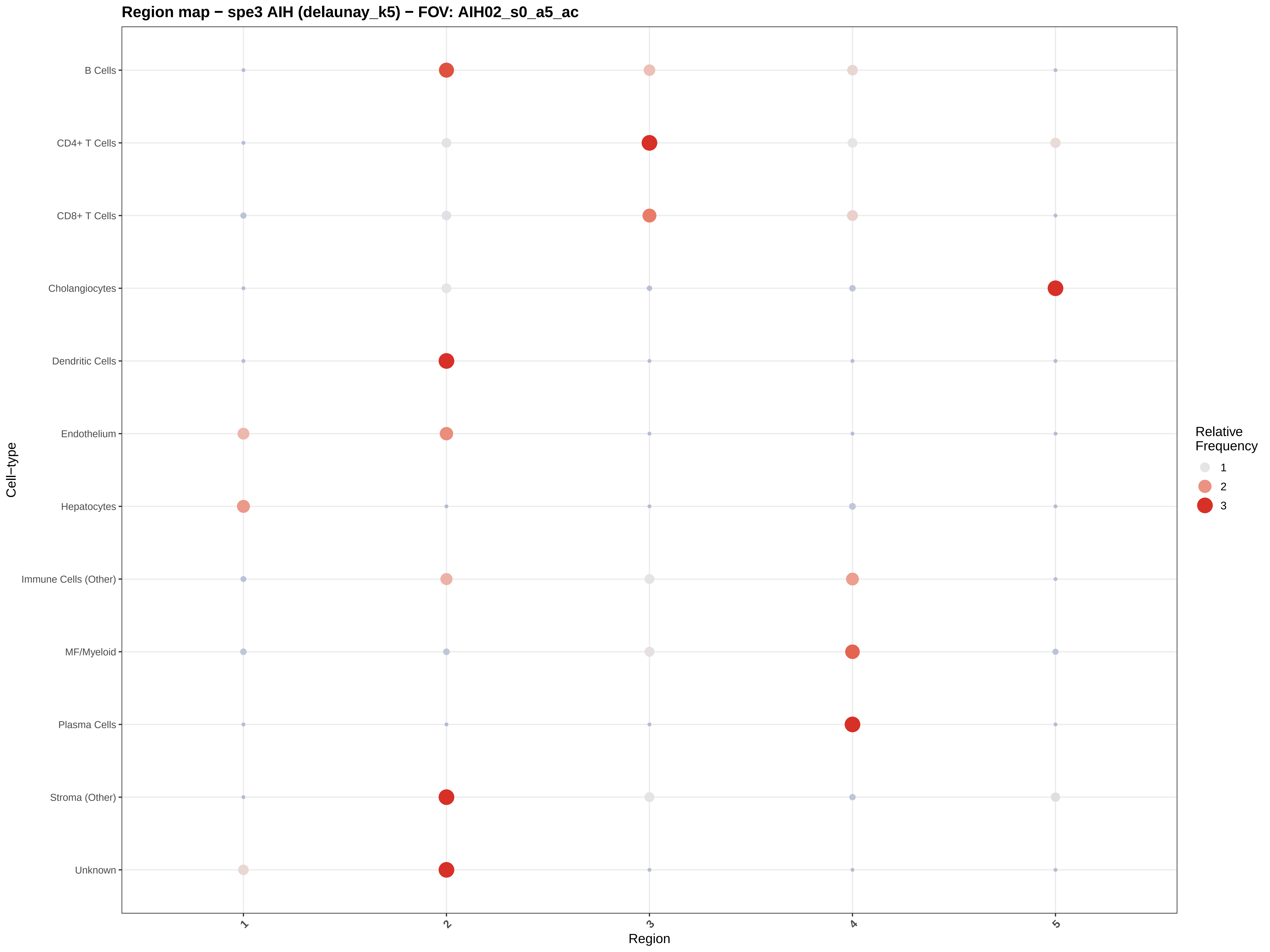

Region map − spe3 AIH (delaunay_k5) − FOV: AIH02_s0_a5_ac
B Cells
CD4+ T Cells
CD8+ T Cells
Cholangiocytes
Dendritic Cells
Endothelium
Hepatocytes
Immune Cells (Other)
MF/Myeloid
Plasma Cells
Stroma (Other)
Unknown
Cell−type
1
2
3
4
5
Region
Relative Frequency
1
2
3

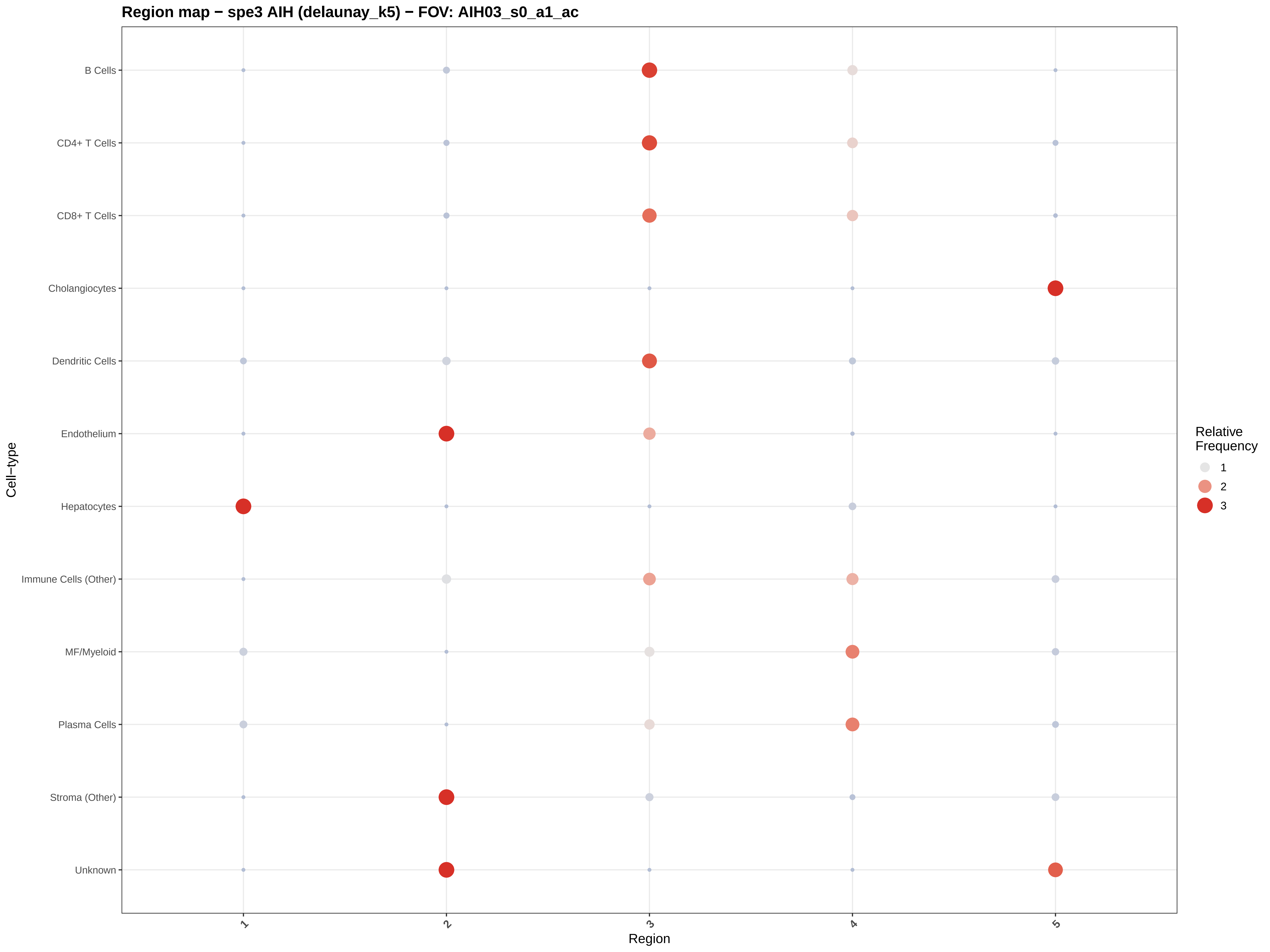
Region map − spe3 AIH (delaunay_k5) − FOV: AIH03_s0_a1_ac
B Cells
CD4+ T Cells
CD8+ T Cells
Cholangiocytes
Dendritic Cells
Endothelium
Hepatocytes
Immune Cells (Other)
MF/Myeloid
Plasma Cells
Stroma (Other)
Unknown
Cell−type
1
2
3
4
5
Region
Relative Frequency
1
2
3

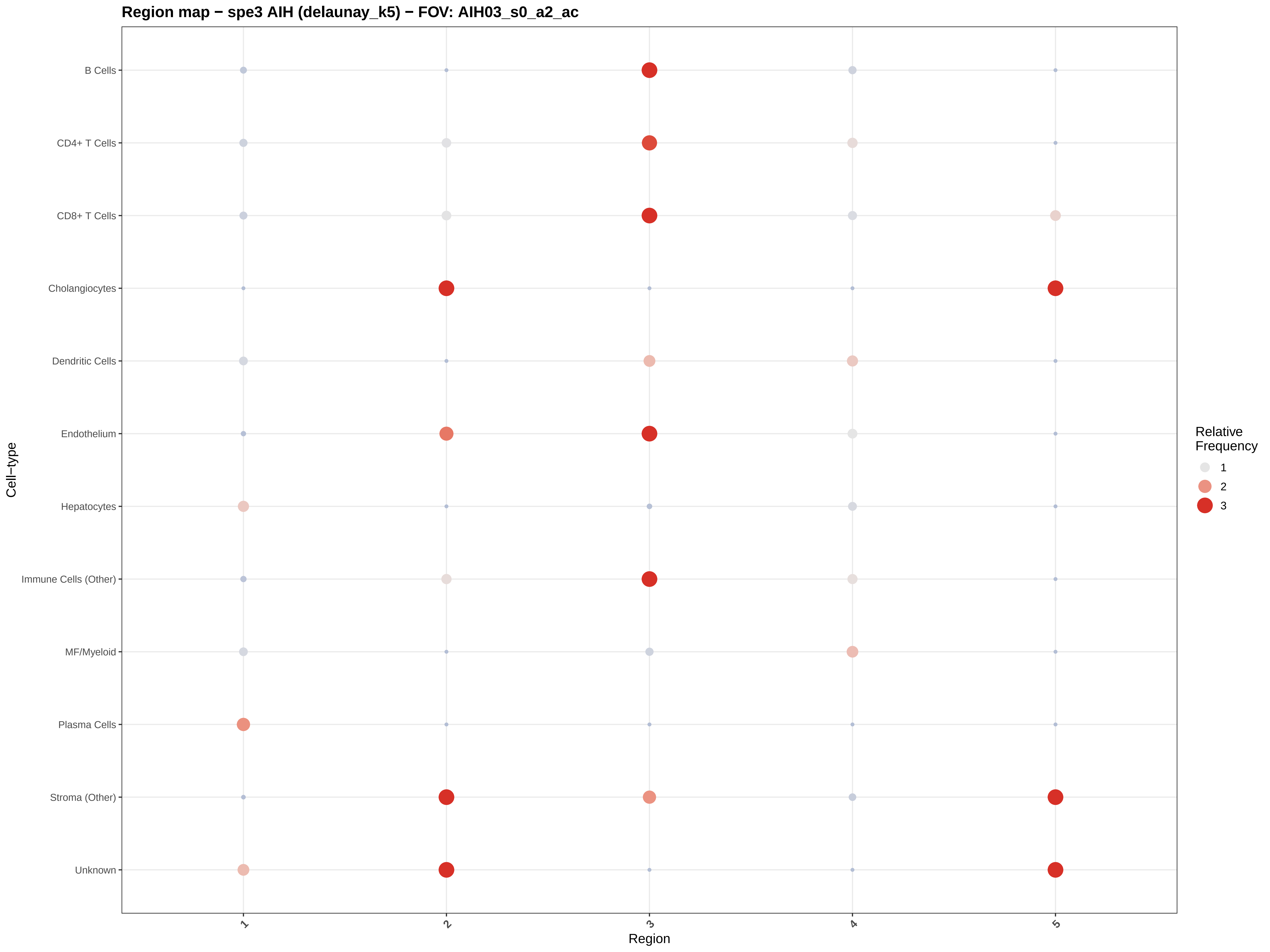
Region map − spe3 AIH (delaunay_k5) − FOV: AIH03_s0_a2_ac
B Cells
CD4+ T Cells
CD8+ T Cells
Cholangiocytes
Dendritic Cells
Endothelium
Hepatocytes
Immune Cells (Other)
MF/Myeloid
Plasma Cells
Stroma (Other)
Unknown
Cell−type
1
2
3
4
5
Region
Relative Frequency
1
2
3

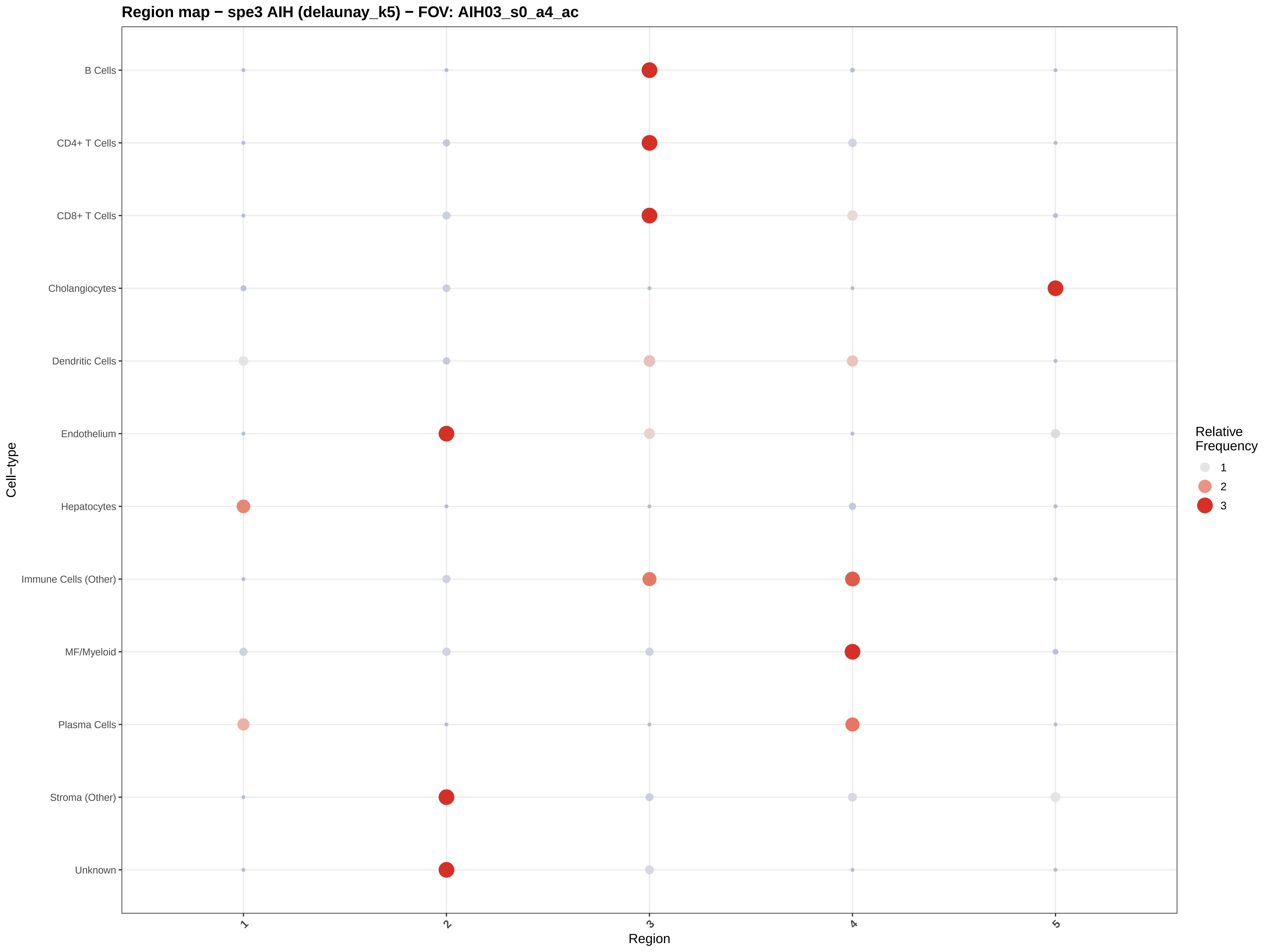
Region map − spe3 AIH (delaunay_k5) − FOV: AIH03_s0_a4_ac
B Cells
CD4+ T Cells
CD8+ T Cells
Cholangiocytes
Dendritic Cells
Endothelium
Hepatocytes
Immune Cells (Other)
MF/Myeloid
Plasma Cells
Stroma (Other)
Unknown
Cell−type
1
2
3
4
5
Region
Relative Frequency
1
2
3

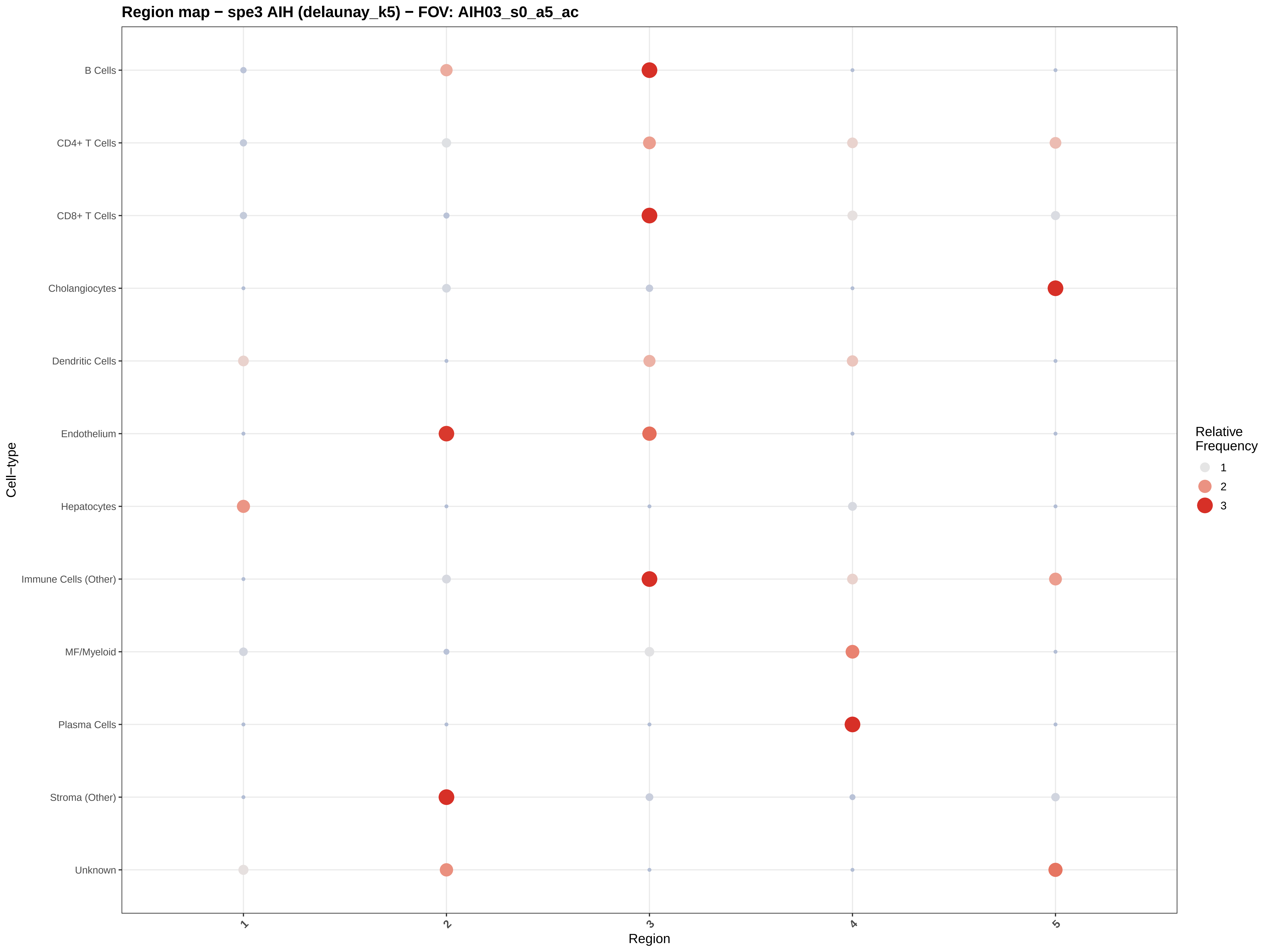

Region map − spe3 AIH (delaunay_k5) − FOV: AIH03_s0_a5_ac
B Cells
CD4+ T Cells
CD8+ T Cells
Cholangiocytes
Dendritic Cells
Endothelium
Hepatocytes
Immune Cells (Other)
MF/Myeloid
Plasma Cells
Stroma (Other)
Unknown
Cell−type
1
2
3
4
5
Region
Relative Frequency
1
2
3

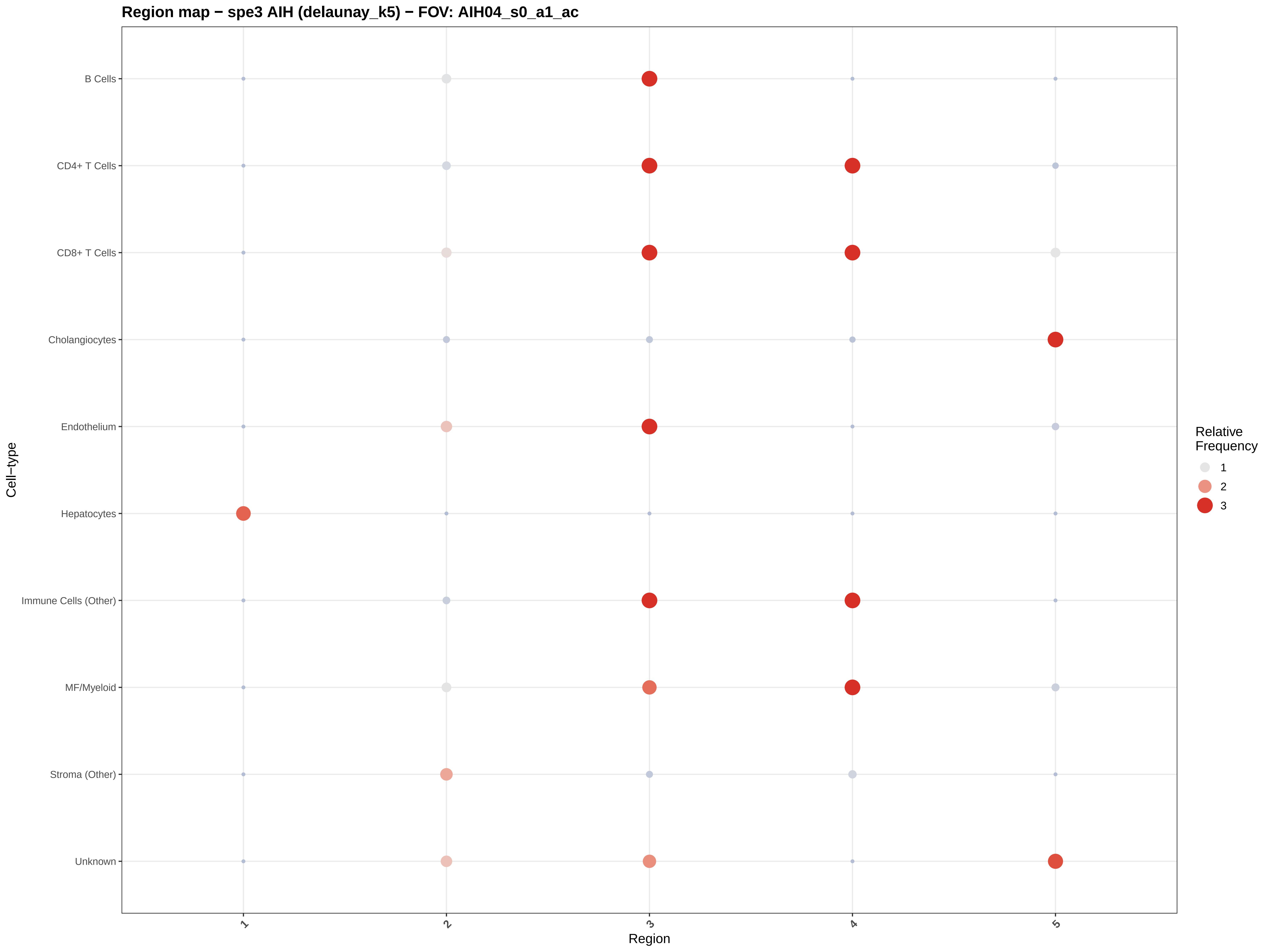

Region map − spe3 AIH (delaunay_k5) − FOV: AIH04_s0_a1_ac
B Cells
CD4+ T Cells
CD8+ T Cells
Cholangiocytes
Endothelium
Hepatocytes
Immune Cells (Other)
MF/Myeloid
Stroma (Other)
Unknown
Cell−type
1
2
3
4
5
Region
Relative Frequency
1
2
3

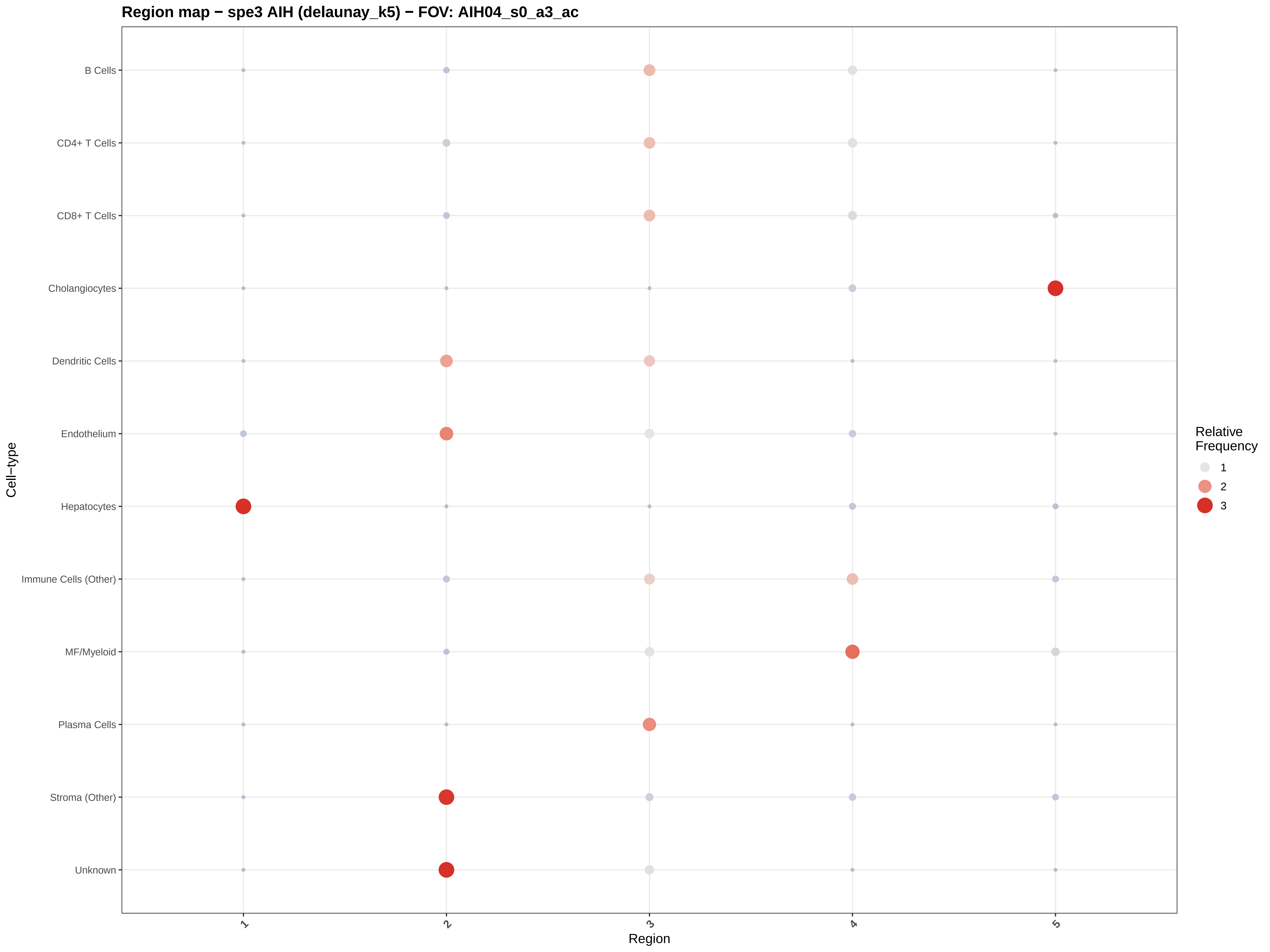
Region map − spe3 AIH (delaunay_k5) − FOV: AIH04_s0_a3_ac
B Cells
CD4+ T Cells
CD8+ T Cells
Cholangiocytes
Dendritic Cells
Endothelium
Hepatocytes
Immune Cells (Other)
MF/Myeloid
Plasma Cells
Stroma (Other)
Unknown
Cell−type
1
2
3
4
5
Region
Relative Frequency
1
2
3

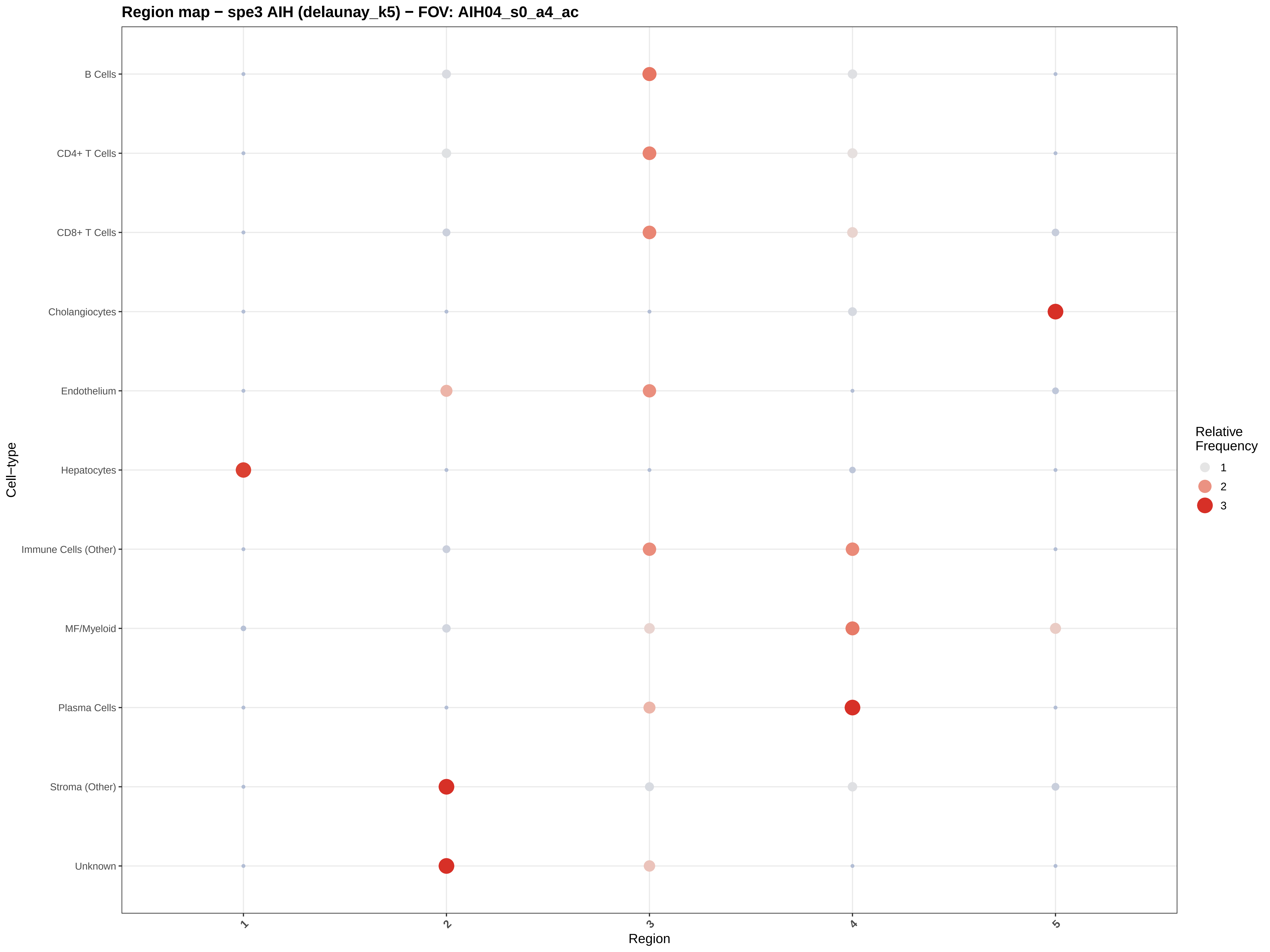

Region map − spe3 AIH (delaunay_k5) − FOV: AIH04_s0_a4_ac
B Cells
CD4+ T Cells
CD8+ T Cells
Cholangiocytes
Endothelium
Hepatocytes
Immune Cells (Other)
MF/Myeloid
Plasma Cells
Stroma (Other)
Unknown
Cell−type
1
2
3
4
5
Region
Relative Frequency
1
2
3

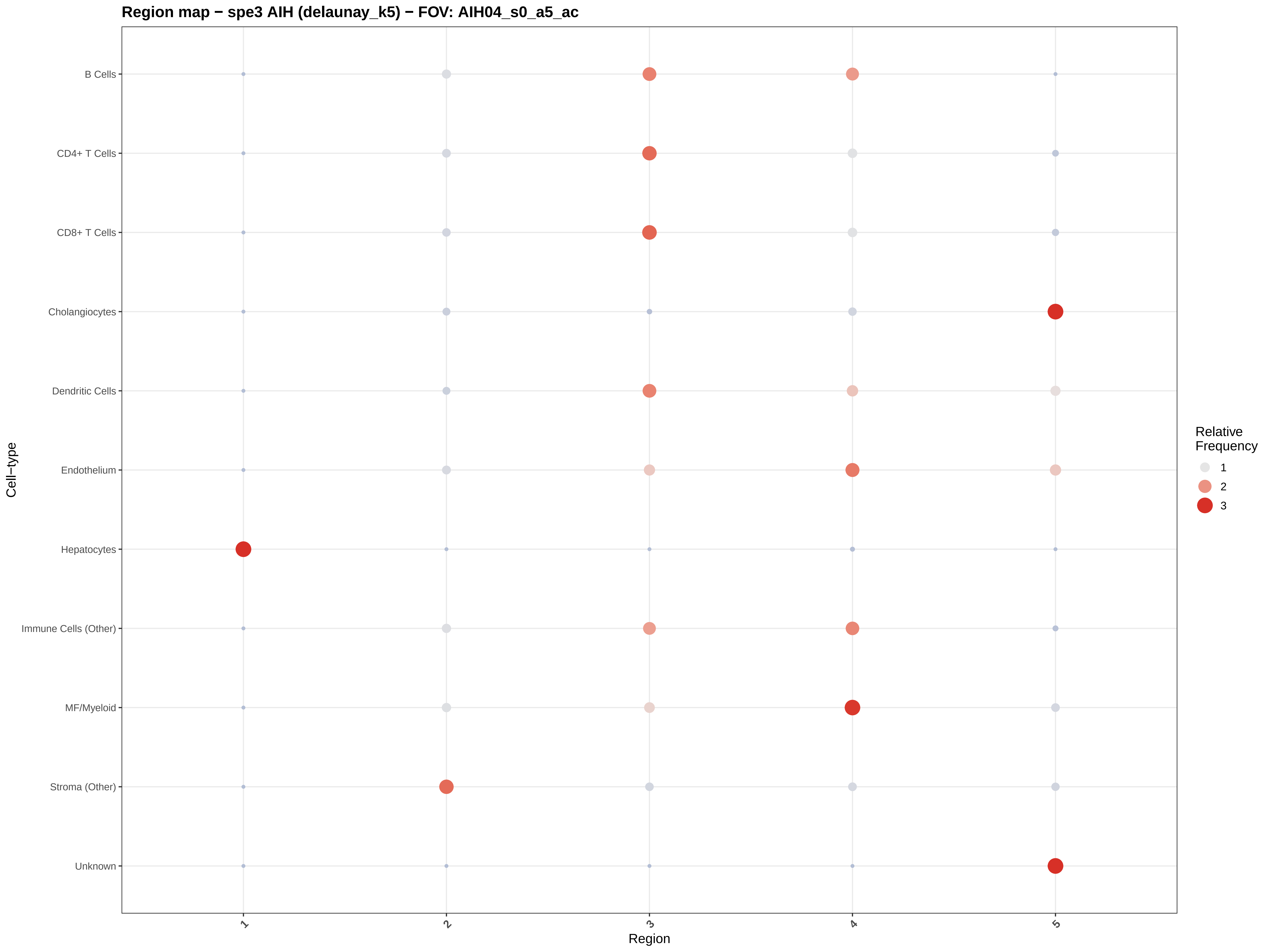
Region map − spe3 AIH (delaunay_k5) − FOV: AIH04_s0_a5_ac
B Cells
CD4+ T Cells
CD8+ T Cells
Cholangiocytes
Dendritic Cells
Endothelium
Hepatocytes
Immune Cells (Other)
MF/Myeloid
Stroma (Other)
Unknown
Cell−type
1
2
3
4
5
Region
Relative Frequency
1
2
3

#### Region map − spe3 AIH (delaunay_k5) − FOV: AIH04_s0_a6_ac

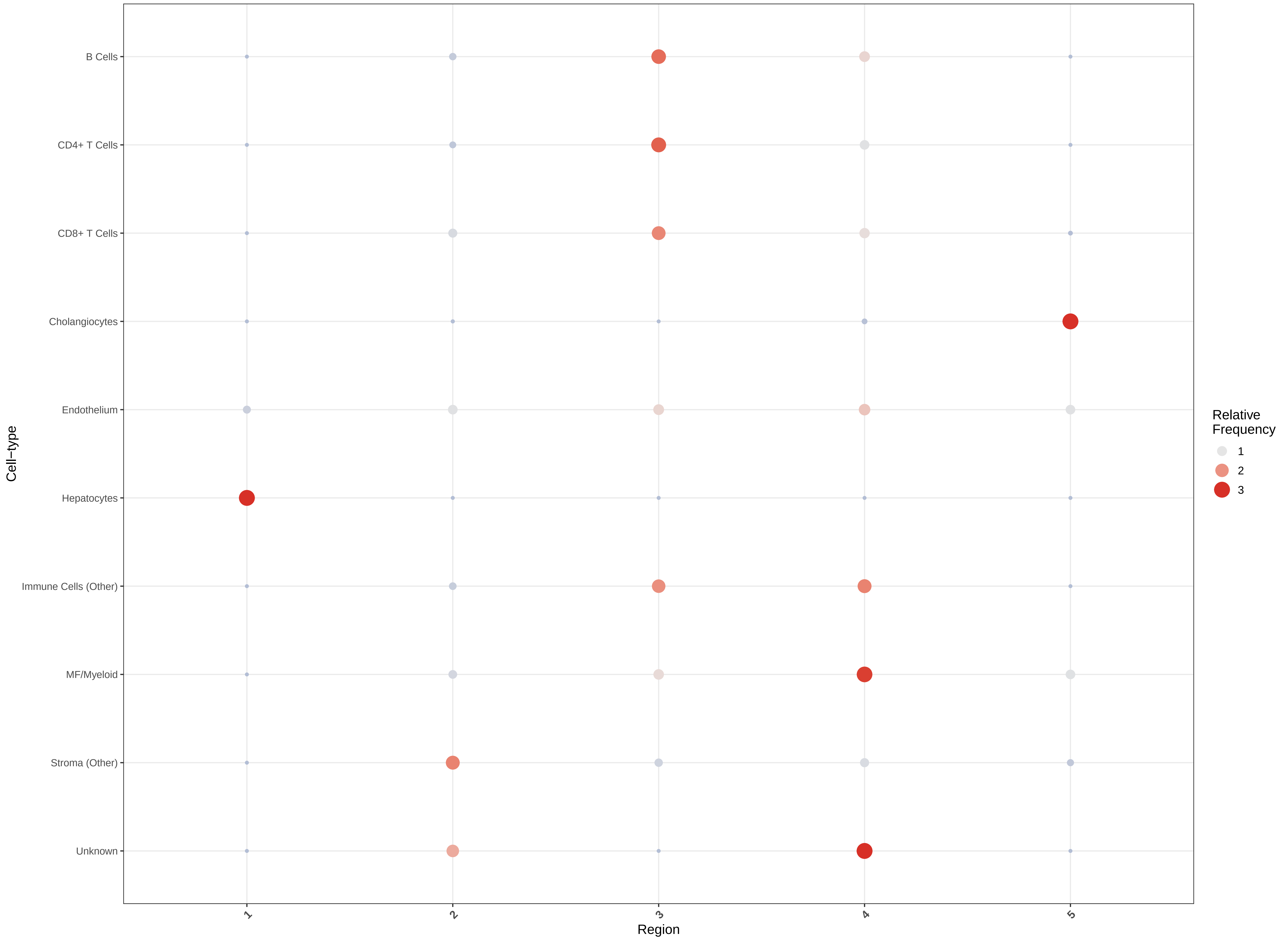

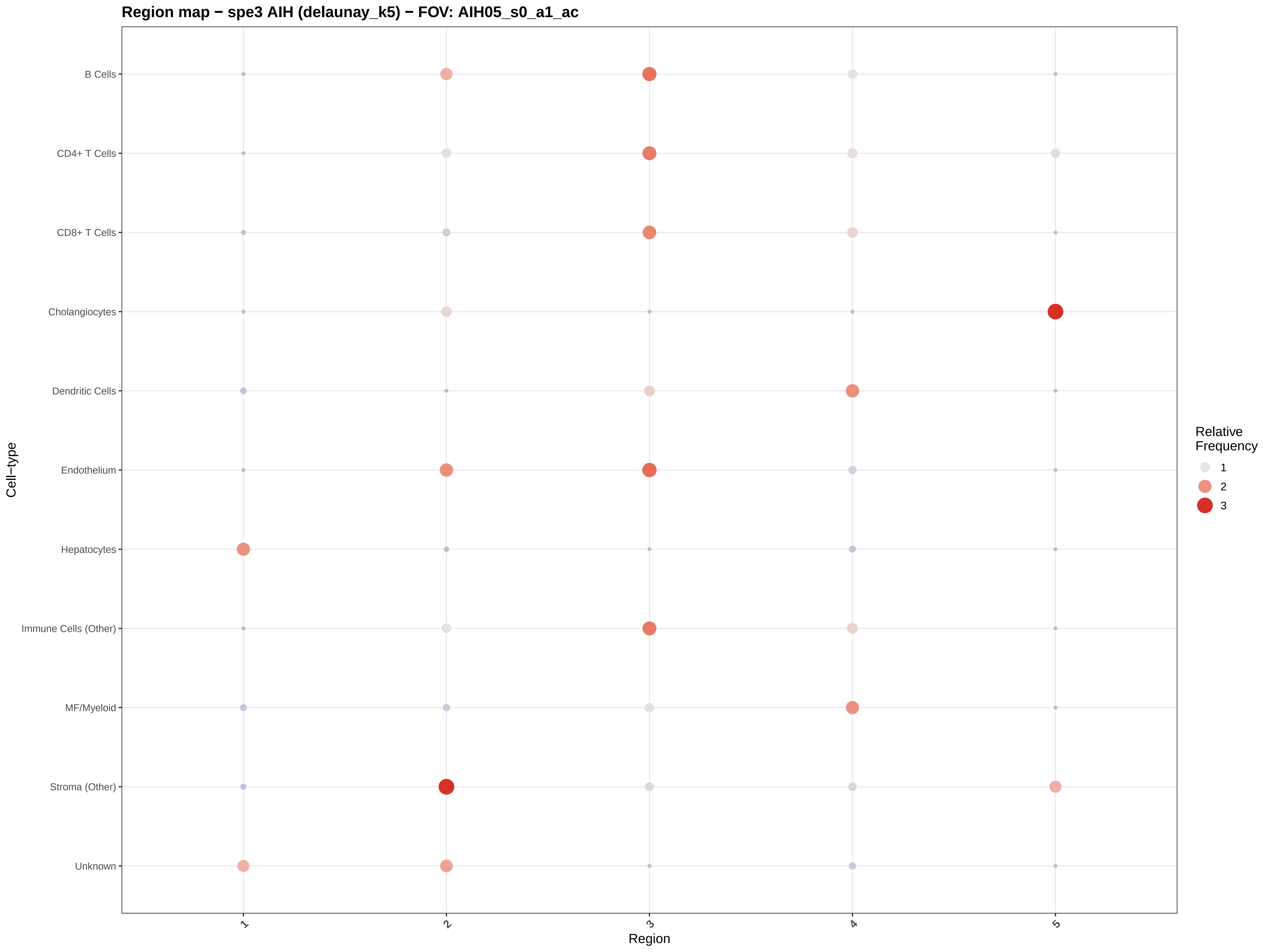

Region map − spe3 AIH (delaunay_k5) − FOV: AIH05_s0_a1_ac
B Cells
CD4+ T Cells
CD8+ T Cells
Cholangiocytes
Dendritic Cells
Endothelium
Hepatocytes
Immune Cells (Other)
MF/Myeloid
Stroma (Other)
Unknown
Cell−type
1
2
3
4
5
Region
Relative Frequency
1
2
3

Region map − spe3 AIH (delaunay_k5) − FOV: AIH05_s0_a2_ac

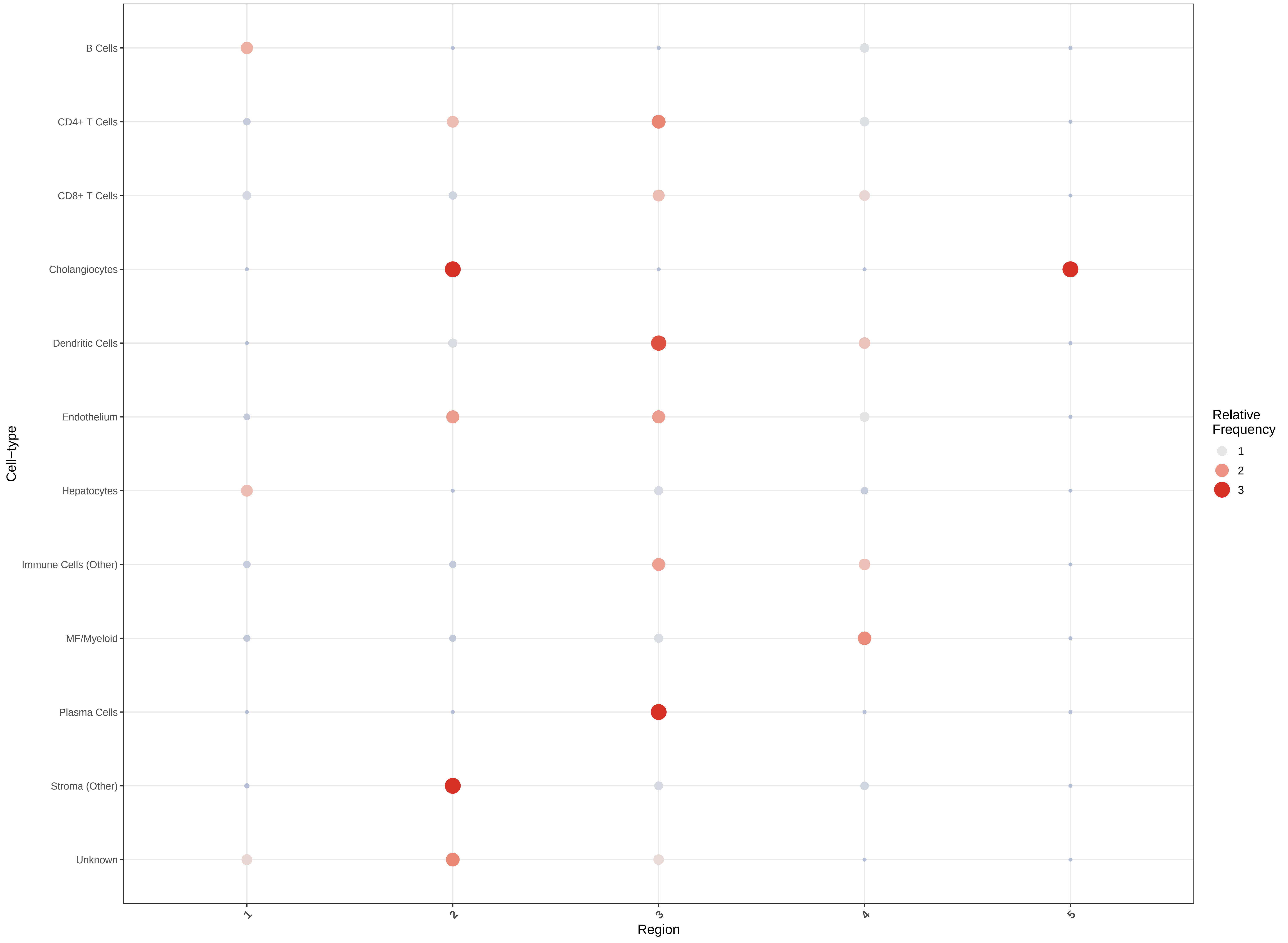

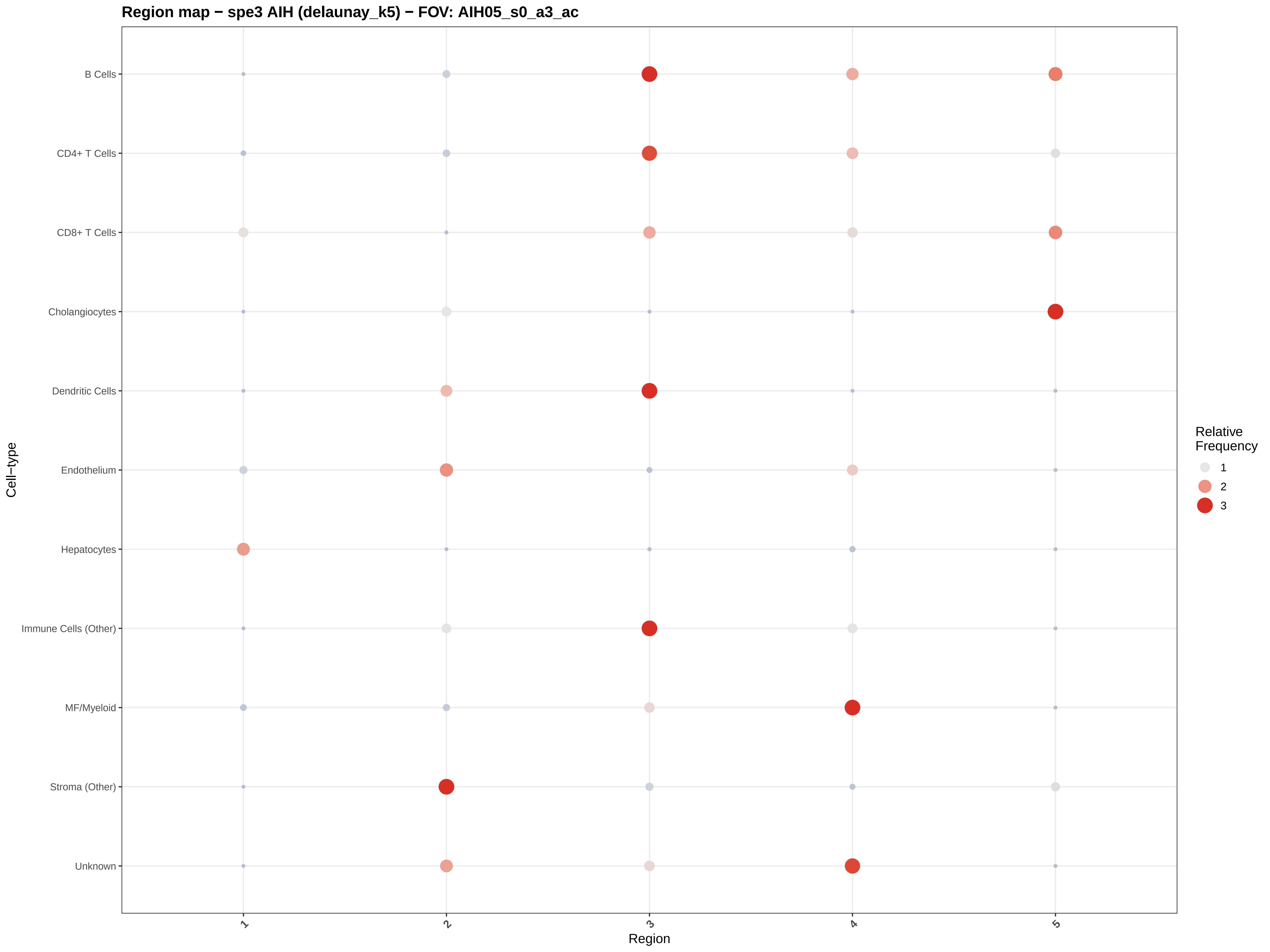
Region map − spe3 AIH (delaunay_k5) − FOV: AIH05_s0_a3_ac
B Cells
CD4+ T Cells
CD8+ T Cells
Cholangiocytes
Dendritic Cells
Endothelium
Hepatocytes
Immune Cells (Other)
MF/Myeloid
Stroma (Other)
Unknown
Cell−type
1
2
3
4
5
Region
Relative Frequency
1
2
3

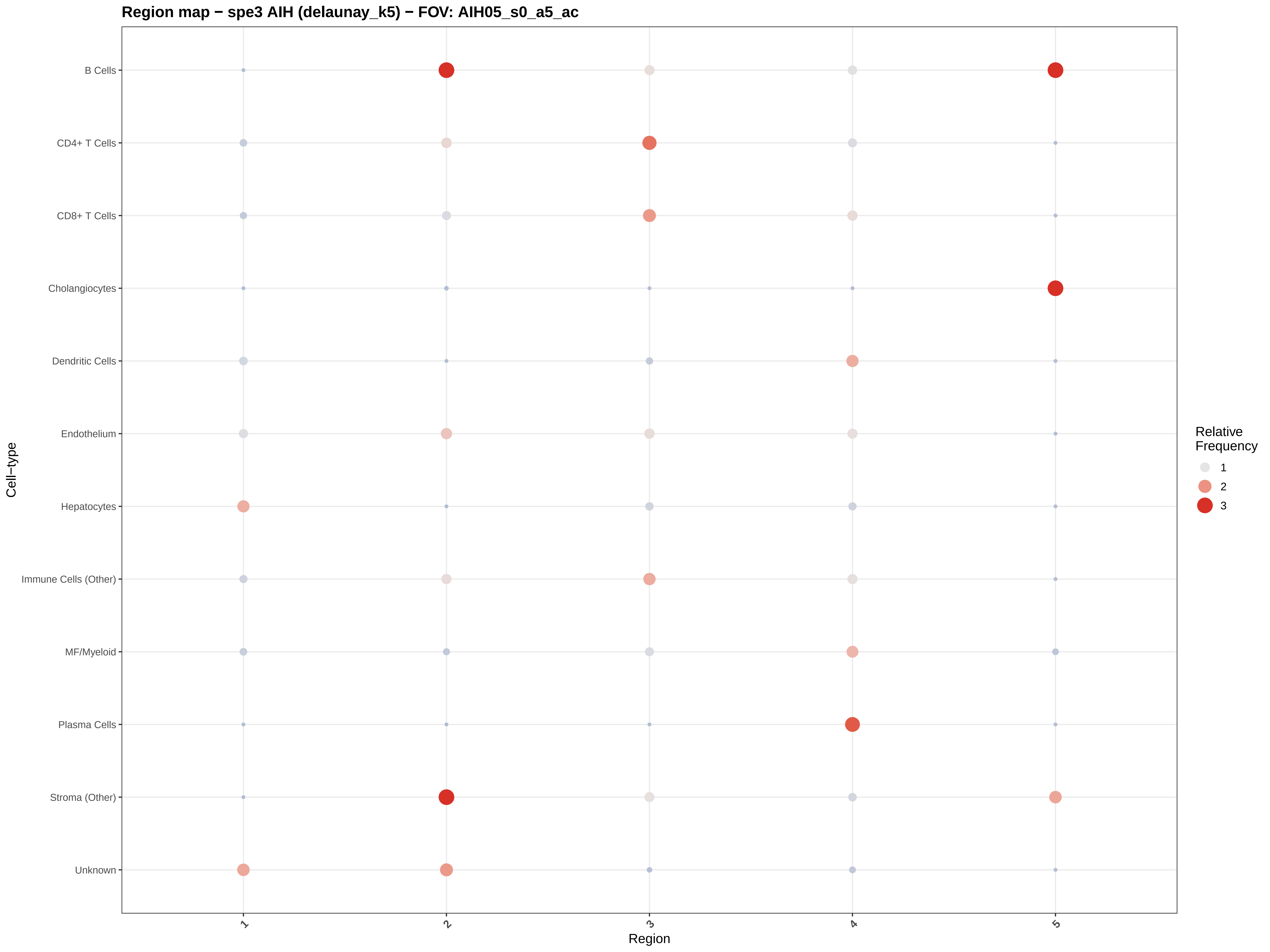

Region map − spe3 AIH (delaunay_k5) − FOV: AIH05_s0_a5_ac
B Cells
CD4+ T Cells
CD8+ T Cells
Cholangiocytes
Dendritic Cells
Endothelium
Hepatocytes
Immune Cells (Other)
MF/Myeloid
Plasma Cells
Stroma (Other)
Unknown
Cell−type
1
2
3
4
5
Region
Relative Frequency
1
2
3

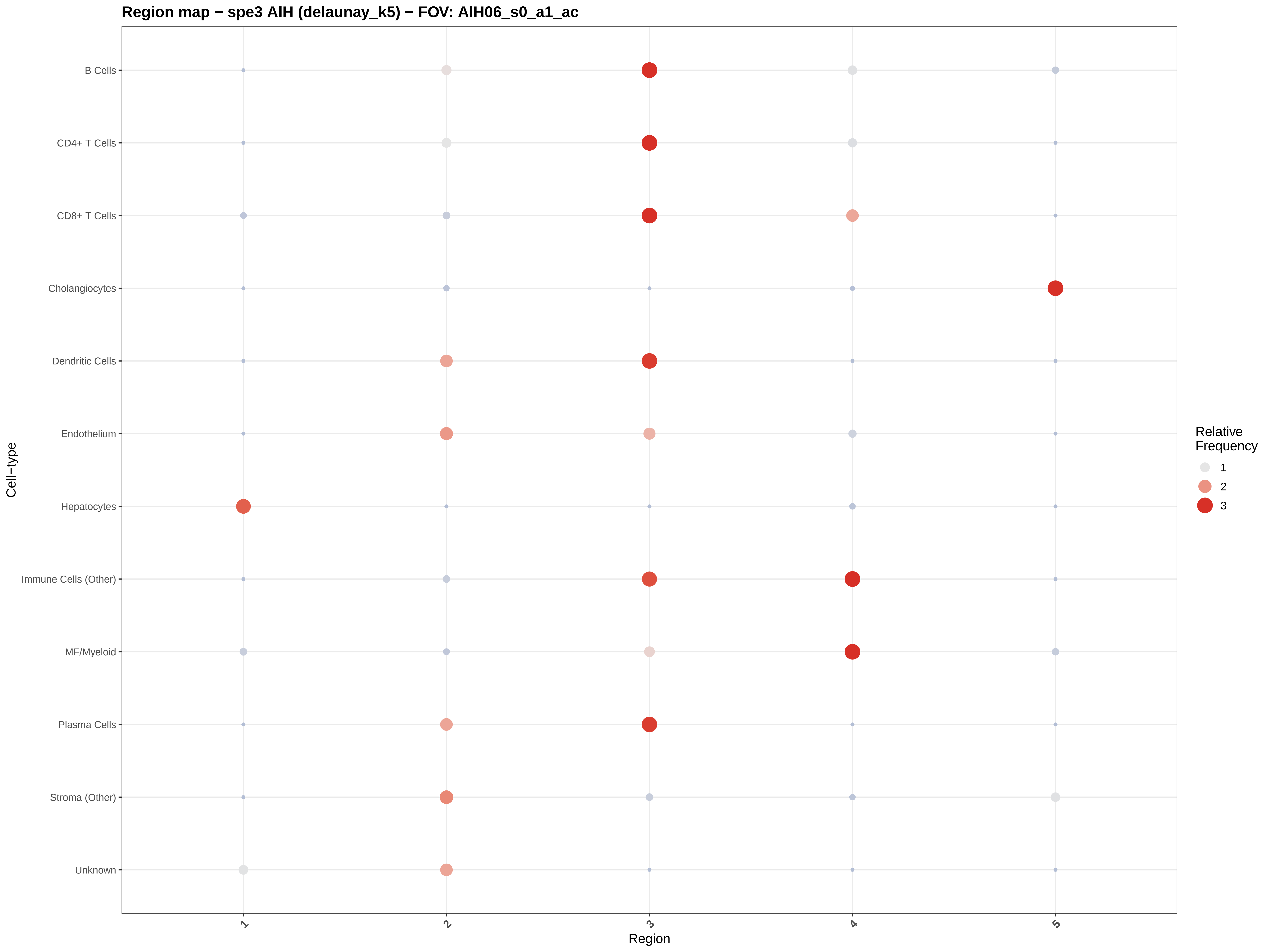

Region map − spe3 AIH (delaunay_k5) − FOV: AIH06_s0_a1_ac
B Cells
CD4+ T Cells
CD8+ T Cells
Cholangiocytes
Dendritic Cells
Endothelium
Hepatocytes
Immune Cells (Other)
MF/Myeloid
Plasma Cells
Stroma (Other)
Unknown
Cell−type
1
2
3
4
5
Region
Relative Frequency
1
2
3

Region map − spe3 AIH (delaunay_k5) − FOV: AIH06_s0_a7_ac

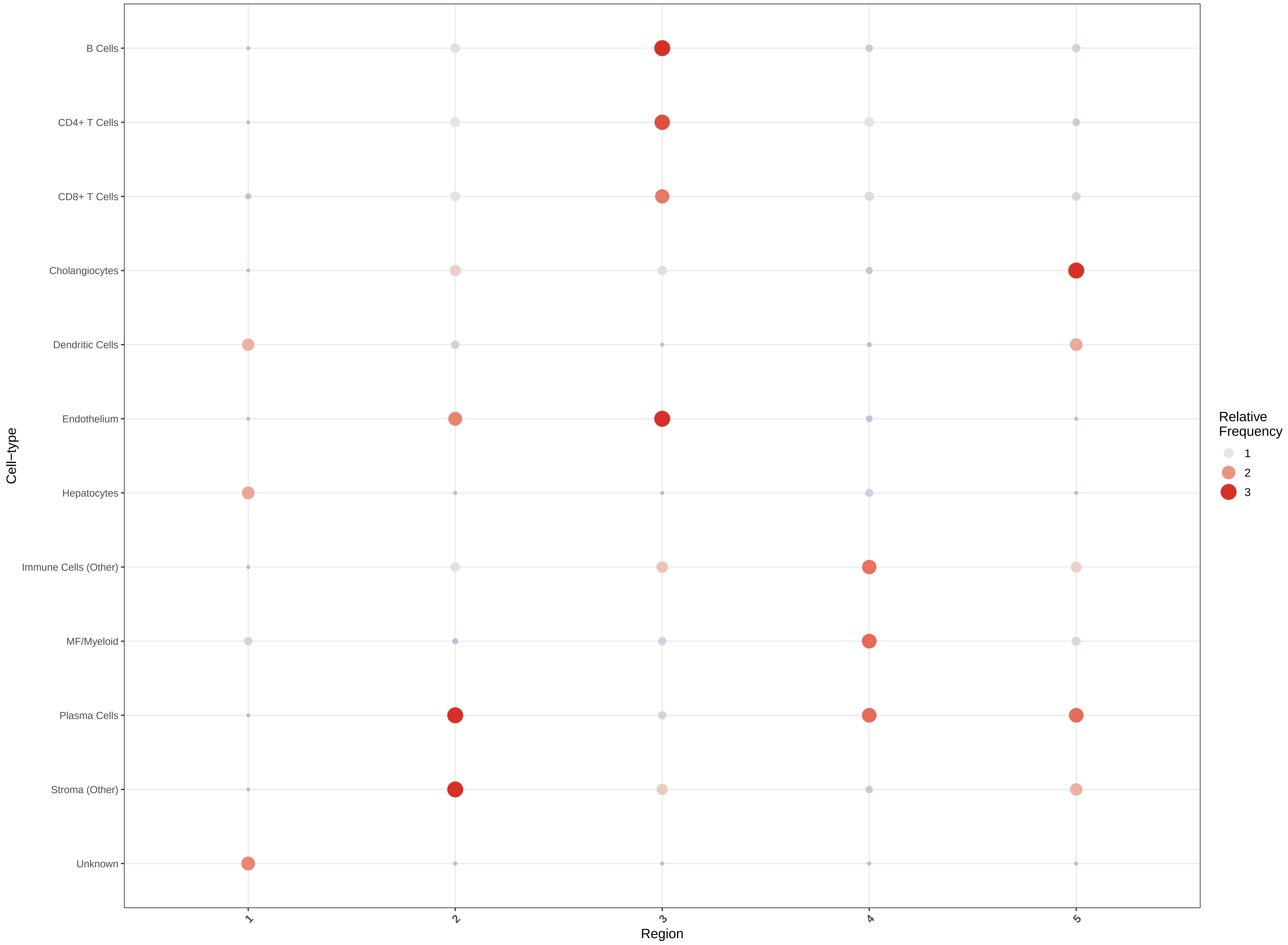

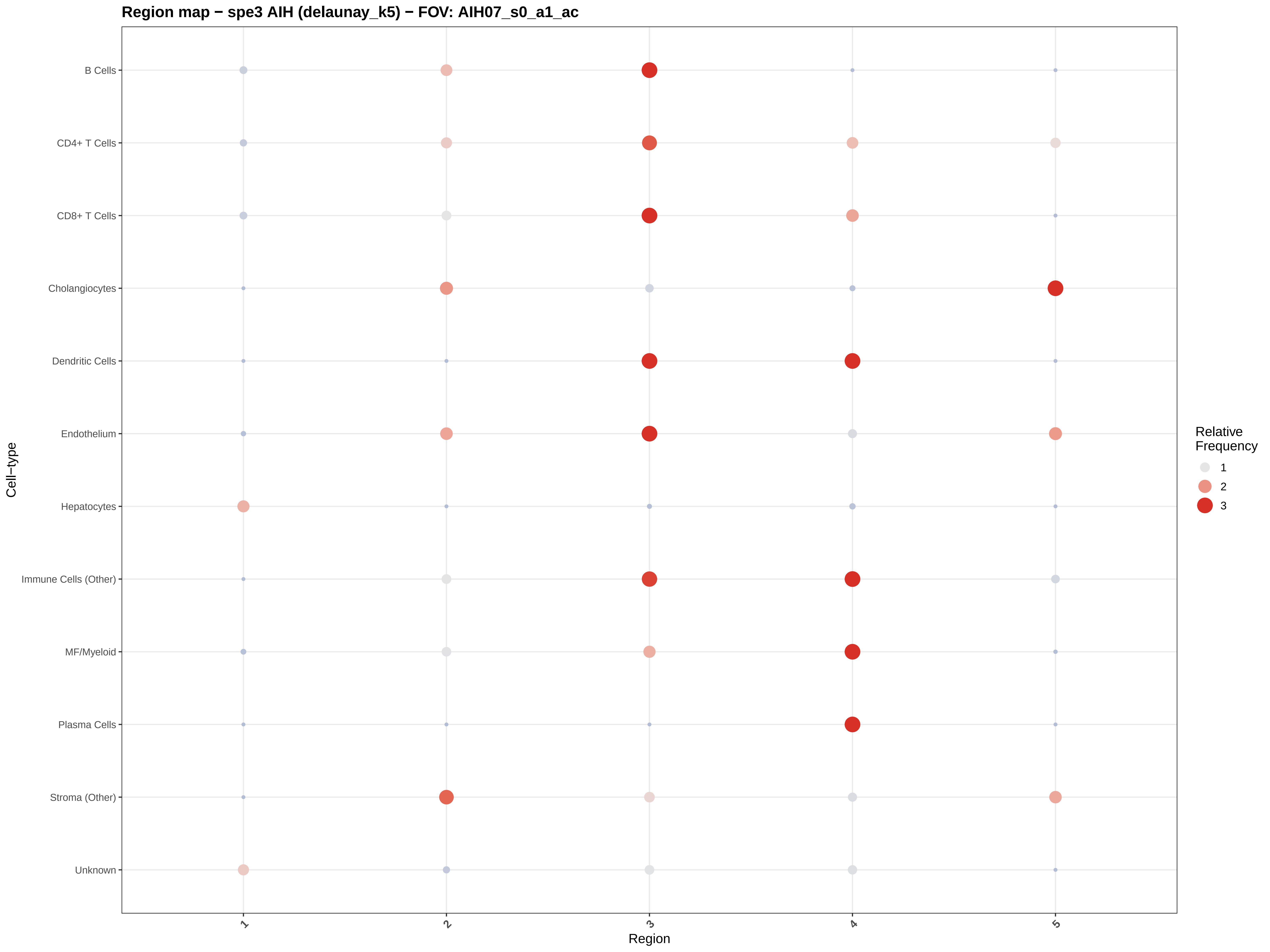
Region map − spe3 AIH (delaunay_k5) − FOV: AIH07_s0_a1_ac
B Cells
CD4+ T Cells
CD8+ T Cells
Cholangiocytes
Dendritic Cells
Endothelium
Hepatocytes
Immune Cells (Other)
MF/Myeloid
Plasma Cells
Stroma (Other)
Unknown
Cell−type
1
2
3
4
5
Region
Relative Frequency
1
2
3

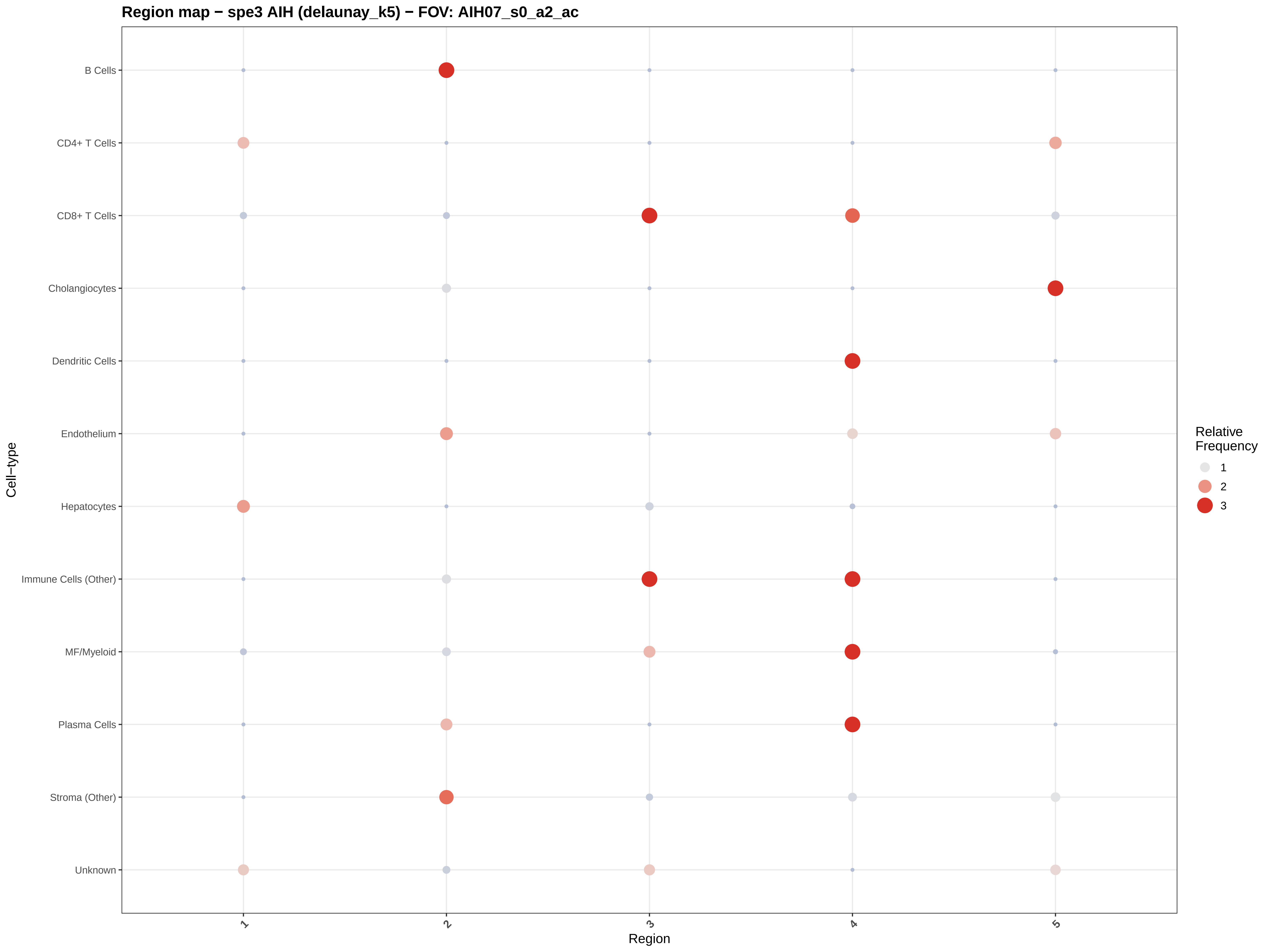

Region map − spe3 AIH (delaunay_k5) − FOV: AIH07_s0_a2_ac
B Cells
CD4+ T Cells
CD8+ T Cells
Cholangiocytes
Dendritic Cells
Endothelium
Hepatocytes
Immune Cells (Other)
MF/Myeloid
Plasma Cells
Stroma (Other)
Unknown
Cell−type
1
2
3
4
5
Region
Relative Frequency
1
2
3

Region map − spe3 AIH (delaunay_k5) − FOV: AIH07_s0_a3_ac

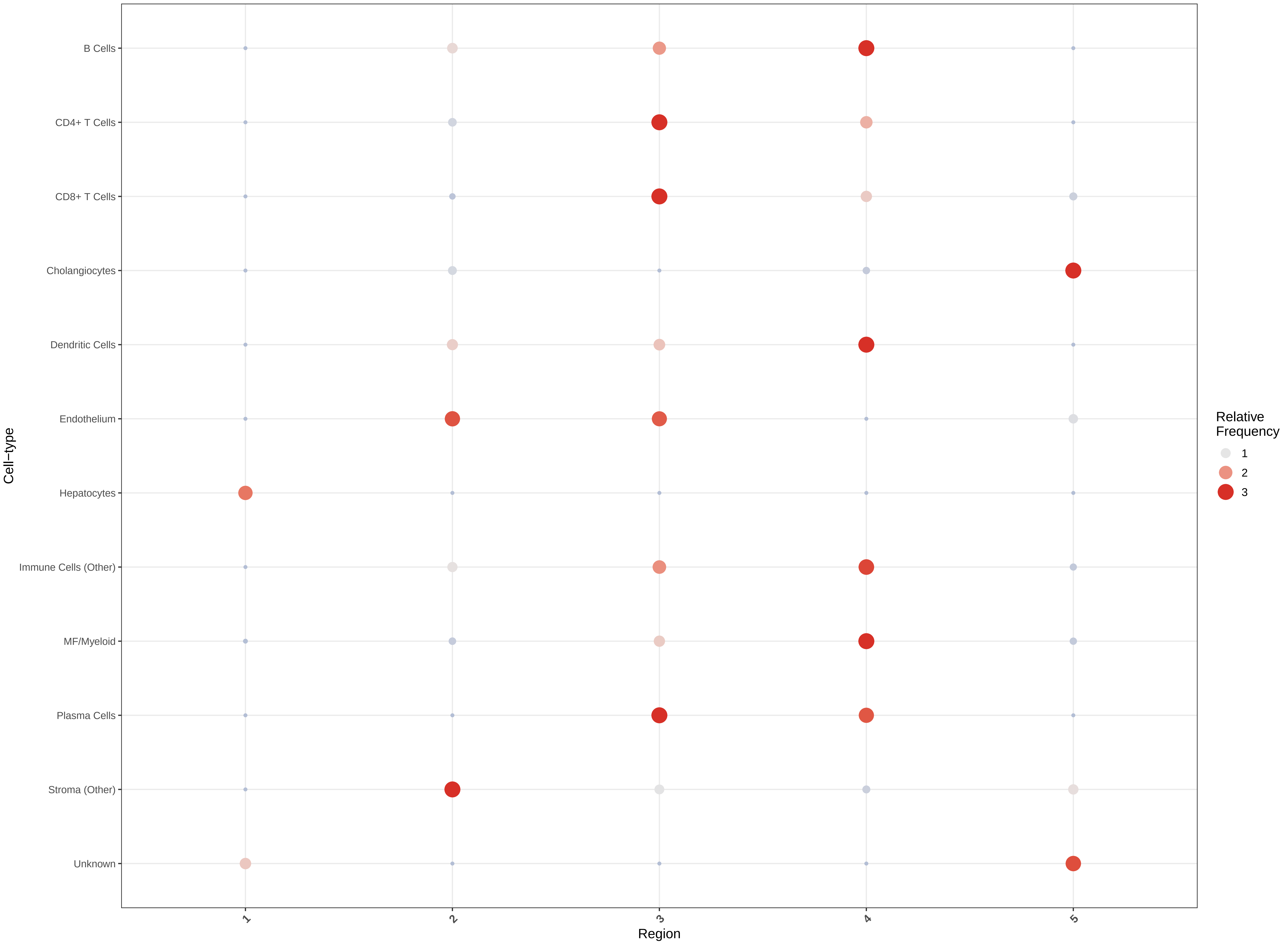

#### Region map − spe3 AIH (delaunay_k5) − FOV: AIH07_s0_a4_ac

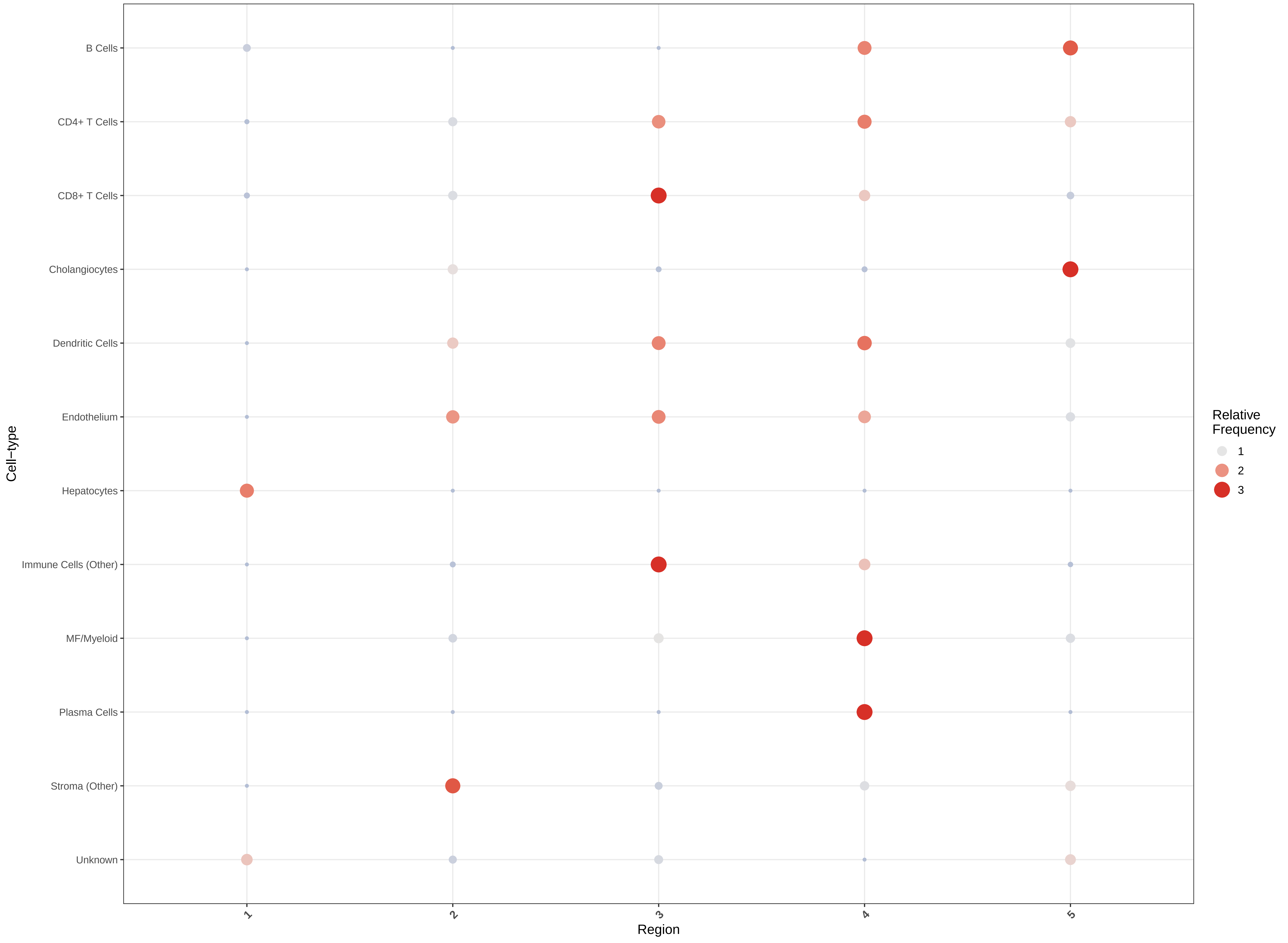

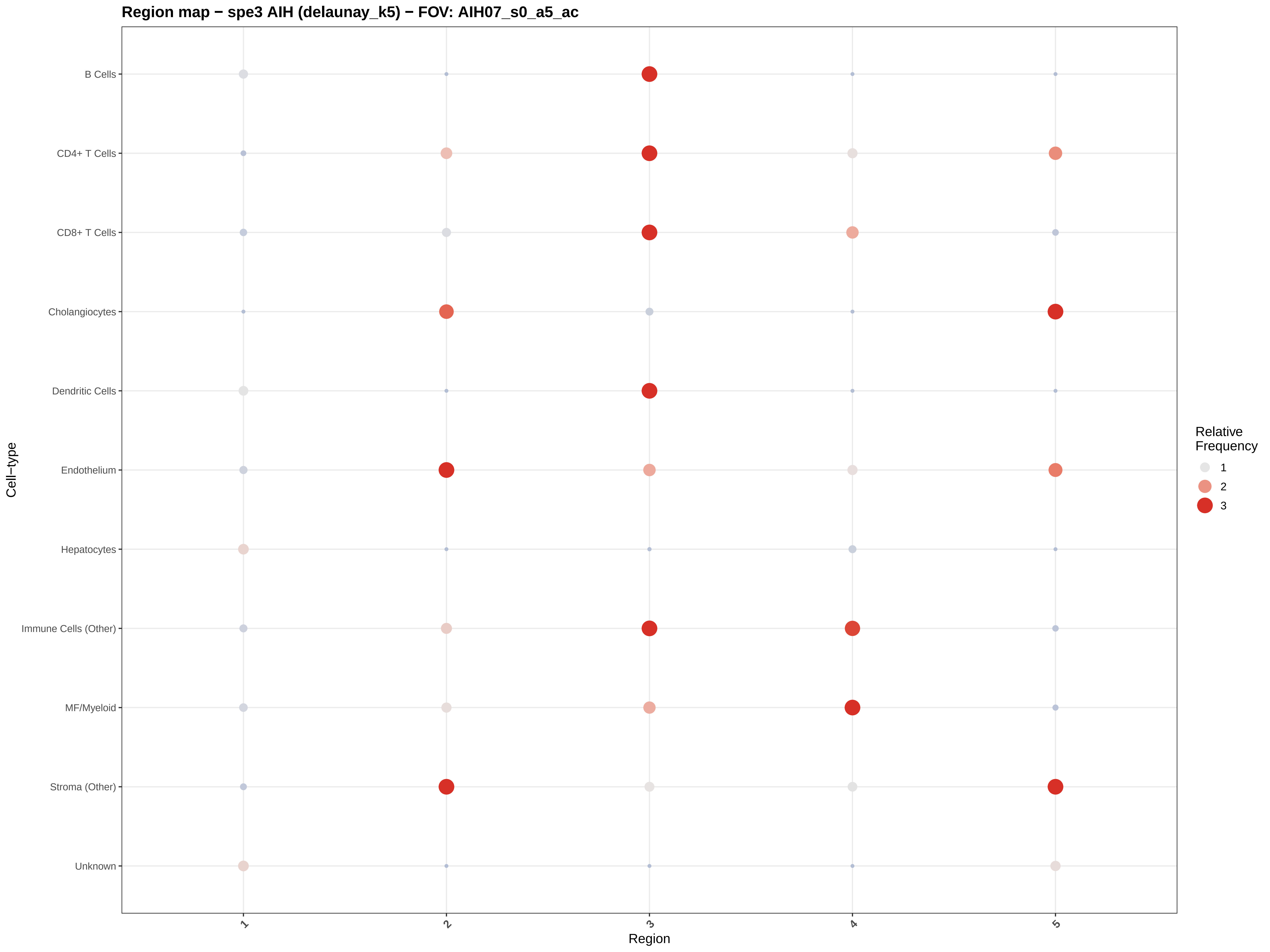

Region map − spe3 AIH (delaunay_k5) − FOV: AIH07_s0_a5_ac
B Cells
CD4+ T Cells
CD8+ T Cells
Cholangiocytes
Dendritic Cells
Endothelium
Hepatocytes
Immune Cells (Other)
MF/Myeloid
Stroma (Other)
Unknown
Cell−type
1
2
3
4
5
Region
Relative Frequency
1
2
3

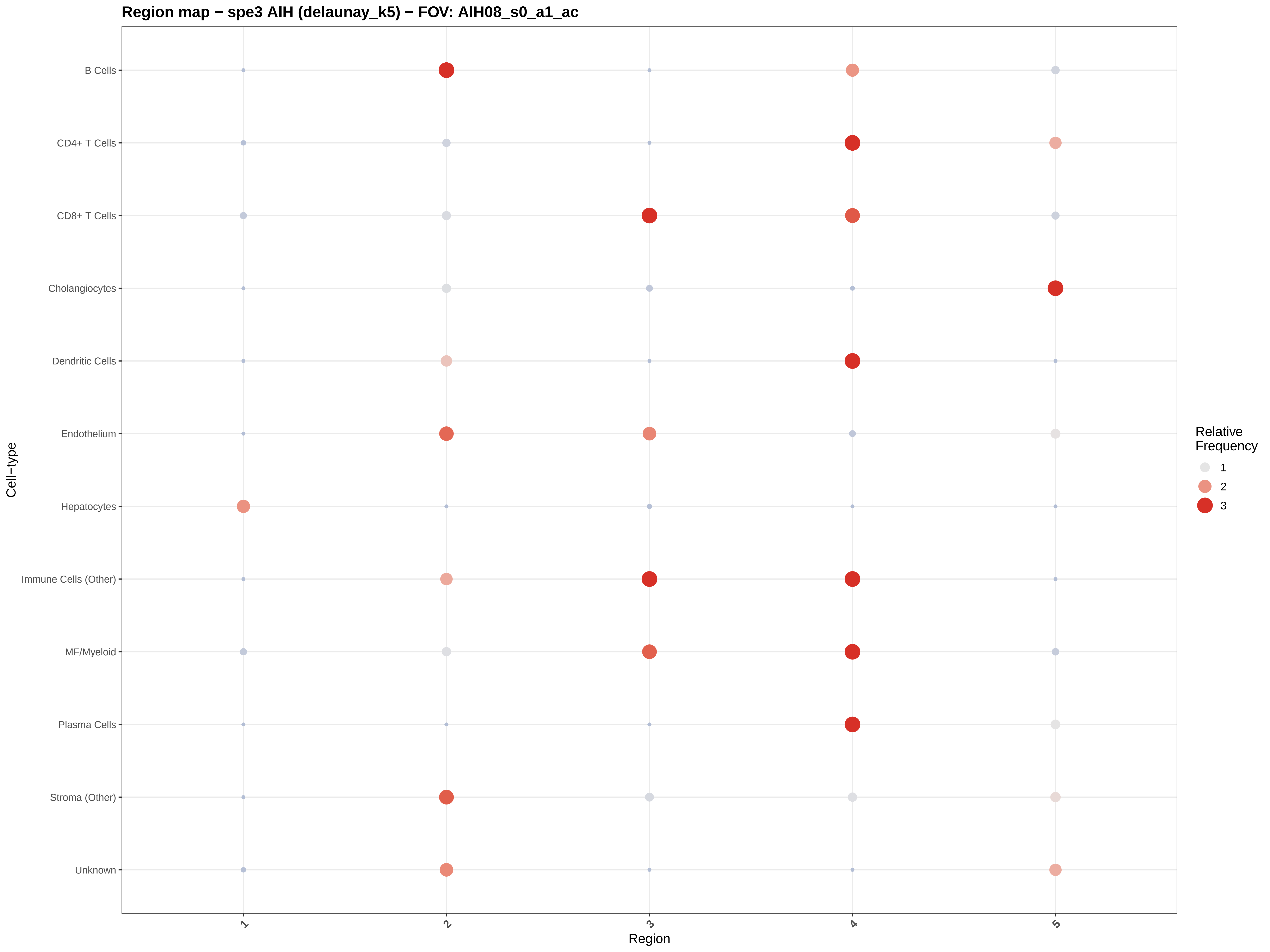

Region map − spe3 AIH (delaunay_k5) − FOV: AIH08_s0_a1_ac
B Cells
CD4+ T Cells
CD8+ T Cells
Cholangiocytes
Dendritic Cells
Endothelium
Hepatocytes
Immune Cells (Other)
MF/Myeloid
Plasma Cells
Stroma (Other)
Unknown
Cell−type
1
2
3
4
5
Region
Relative Frequency
1
2
3

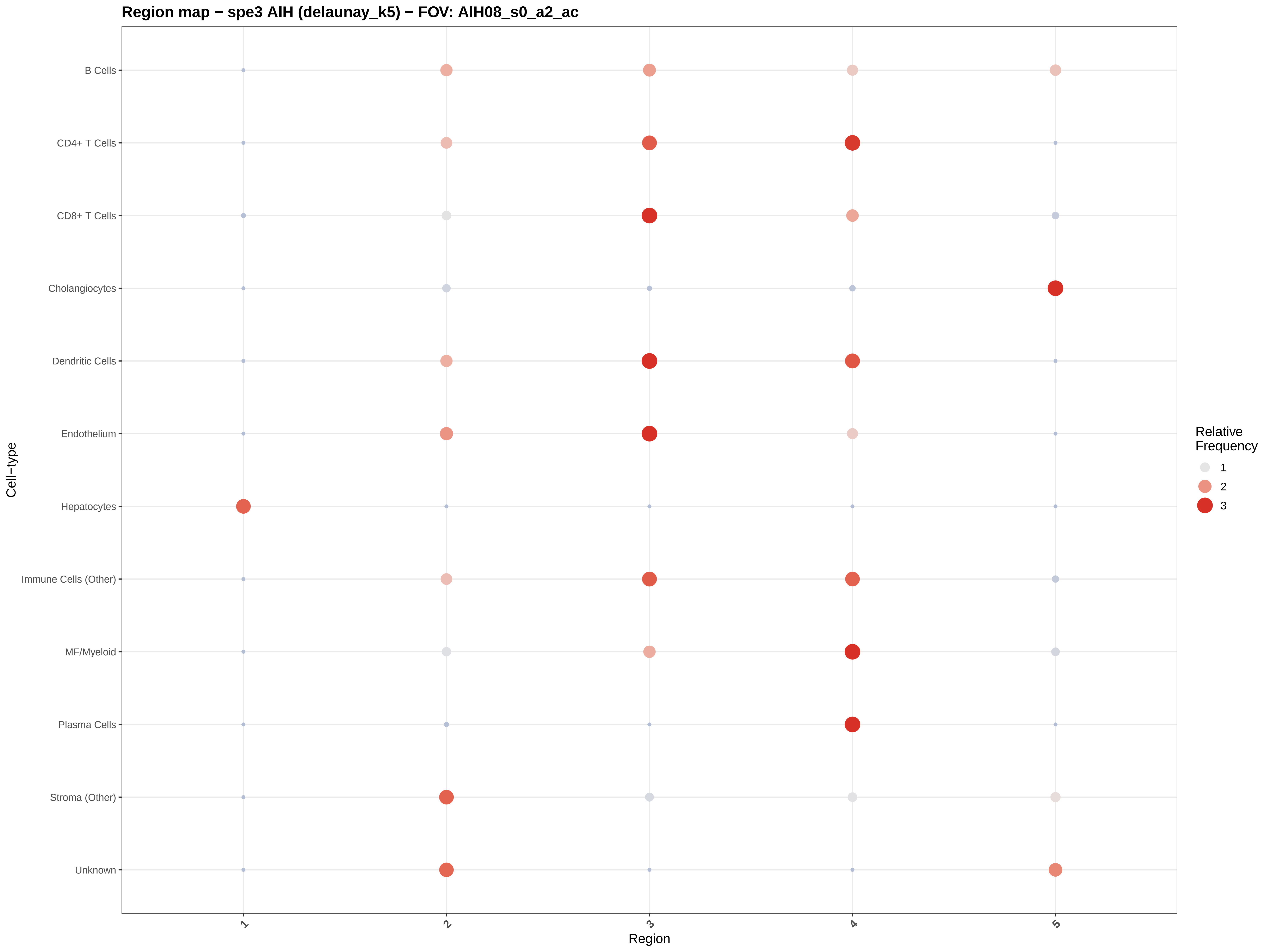

Region map − spe3 AIH (delaunay_k5) − FOV: AIH08_s0_a2_ac
B Cells
CD4+ T Cells
CD8+ T Cells
Cholangiocytes
Dendritic Cells
Endothelium
Hepatocytes
Immune Cells (Other)
MF/Myeloid
Plasma Cells
Stroma (Other)
Unknown
Cell−type
1
2
3
4
5
Region
Relative Frequency
1
2
3

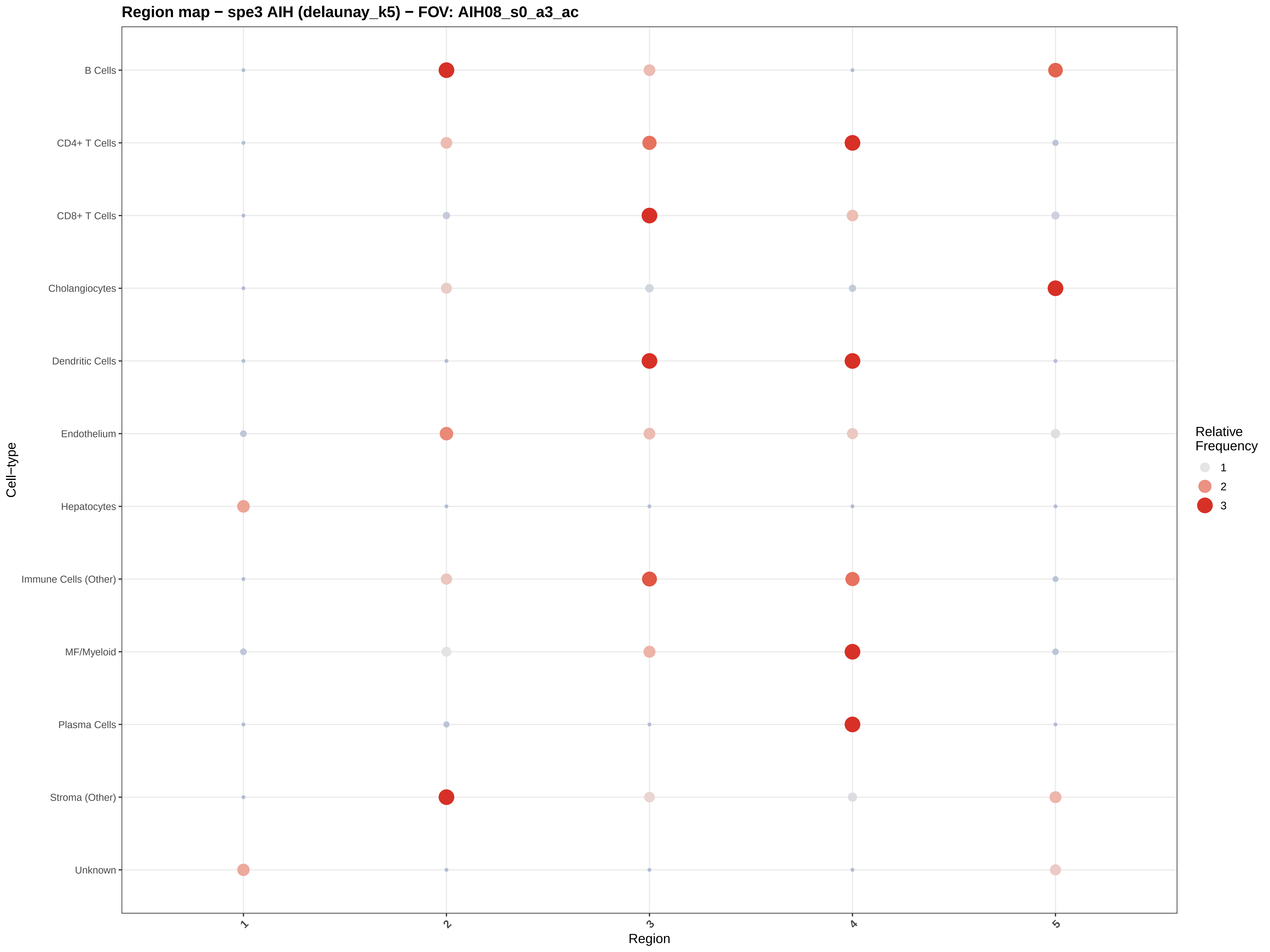
Region map − spe3 AIH (delaunay_k5) − FOV: AIH08_s0_a3_ac
B Cells
CD4+ T Cells
CD8+ T Cells
Cholangiocytes
Dendritic Cells
Endothelium
Hepatocytes
Immune Cells (Other)
MF/Myeloid
Plasma Cells
Stroma (Other)
Unknown
Cell−type
1
2
3
4
5
Region
Relative Frequency
1
2
3

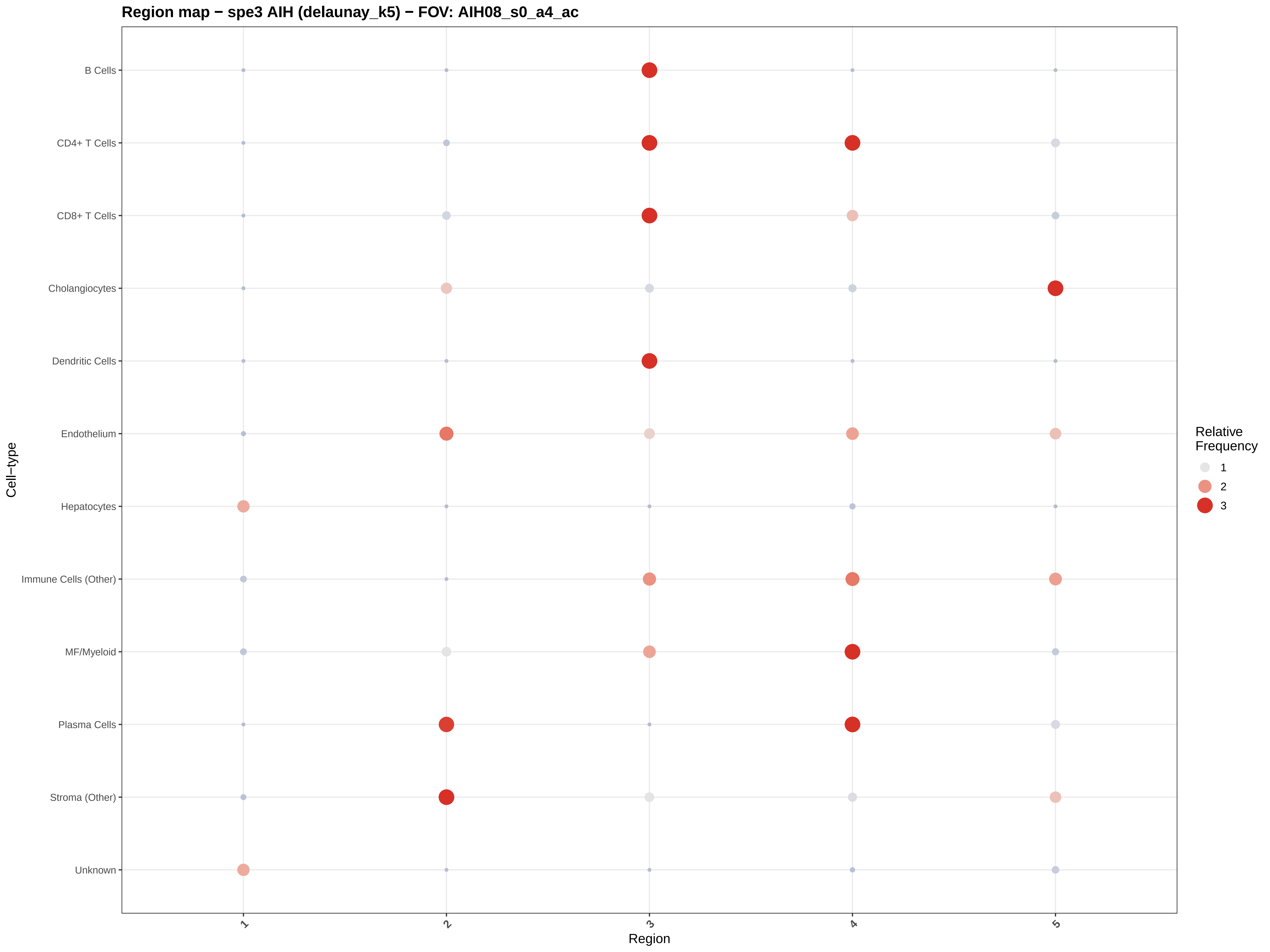

Region map − spe3 AIH (delaunay_k5) − FOV: AIH08_s0_a4_ac
B Cells
CD4+ T Cells
CD8+ T Cells
Cholangiocytes
Dendritic Cells
Endothelium
Hepatocytes
Immune Cells (Other)
MF/Myeloid
Plasma Cells
Stroma (Other)
Unknown
Cell−type
1
2
3
4
5
Region
Relative Frequency
1
2
3

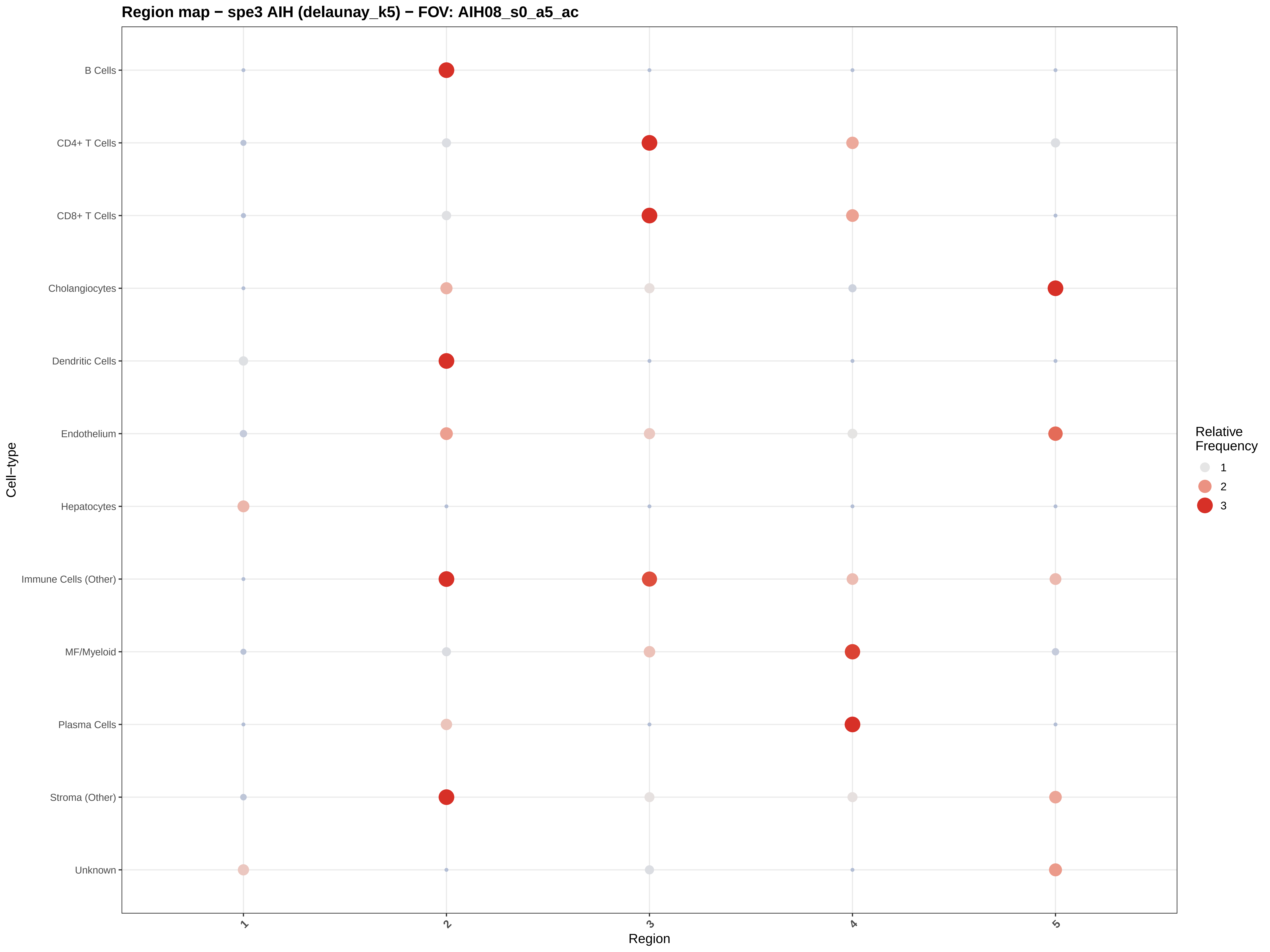

Region map − spe3 AIH (delaunay_k5) − FOV: AIH08_s0_a5_ac
B Cells
CD4+ T Cells
CD8+ T Cells
Cholangiocytes
Dendritic Cells
Endothelium
Hepatocytes
Immune Cells (Other)
MF/Myeloid
Plasma Cells
Stroma (Other)
Unknown
Cell−type
1
2
3
4
5
Region
Relative Frequency
1
2
3

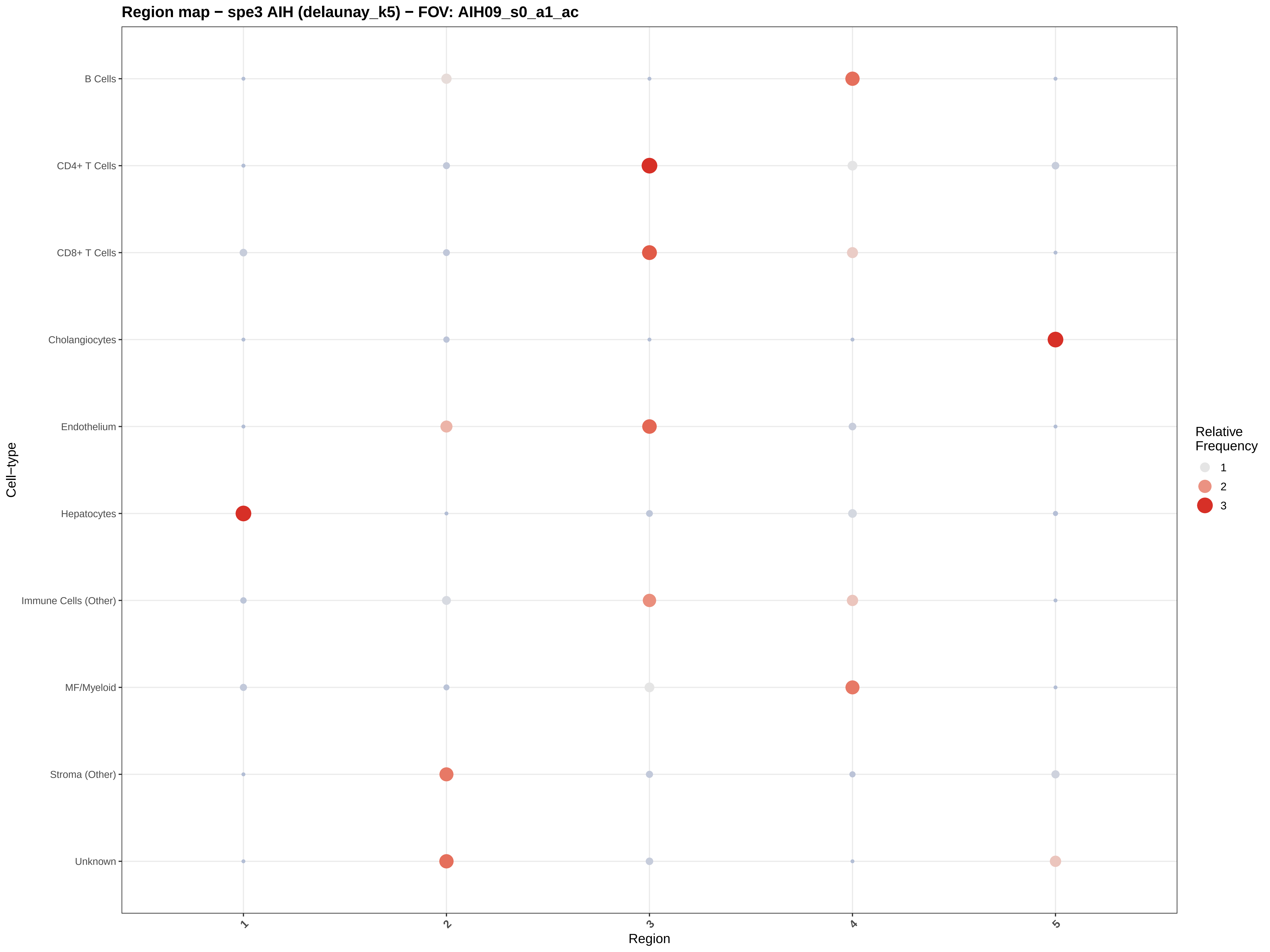

Region map − spe3 AIH (delaunay_k5) − FOV: AIH09_s0_a1_ac
B Cells
CD4+ T Cells
CD8+ T Cells
Cholangiocytes
Endothelium
Hepatocytes
Immune Cells (Other)
MF/Myeloid
Stroma (Other)
Unknown
Cell−type
1
2
3
4
5
Region
Relative Frequency
1
2
3

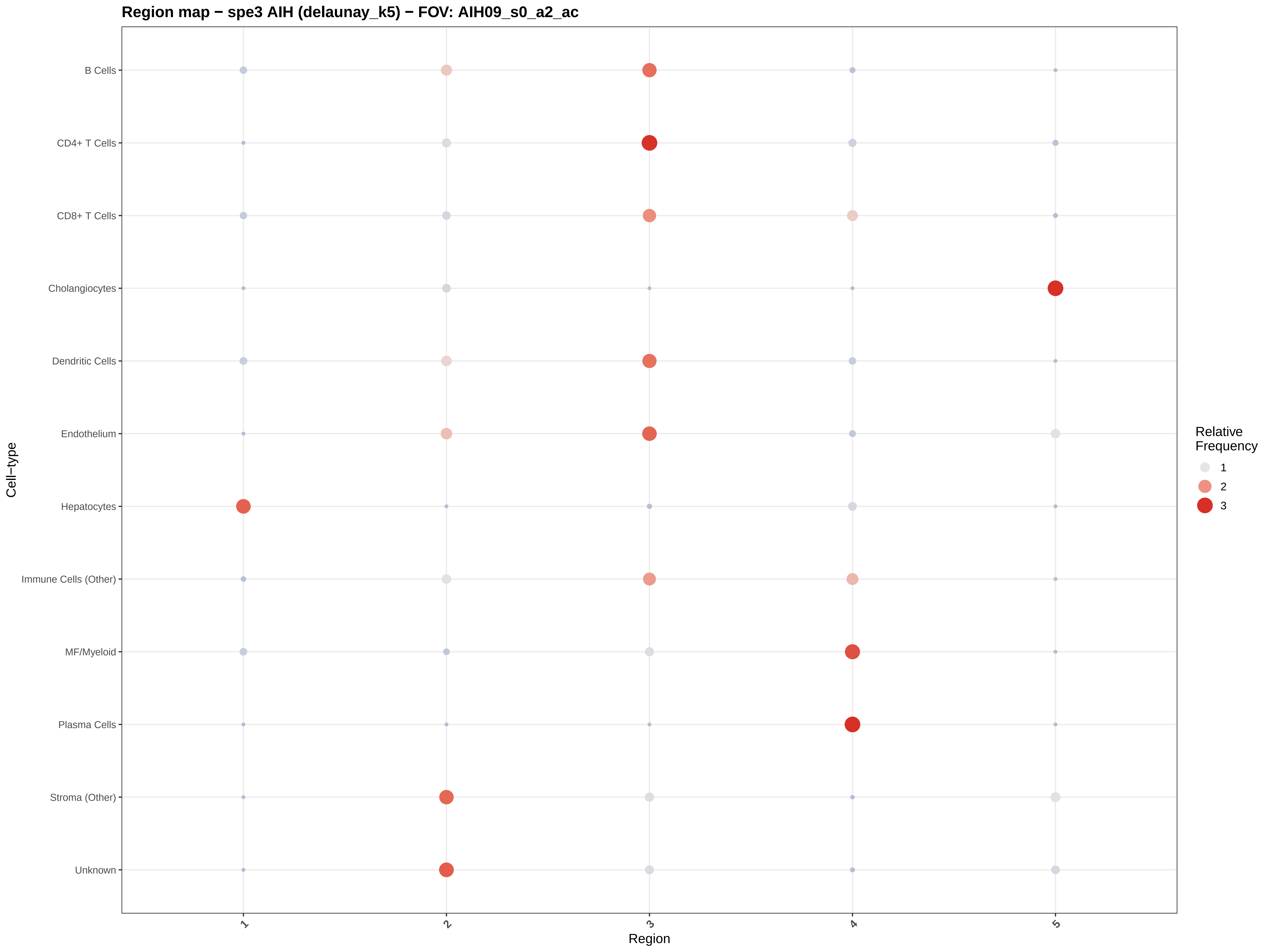

Region map − spe3 AIH (delaunay_k5) − FOV: AIH09_s0_a2_ac
B Cells
CD4+ T Cells
CD8+ T Cells
Cholangiocytes
Dendritic Cells
Endothelium
Hepatocytes
Immune Cells (Other)
MF/Myeloid
Plasma Cells
Stroma (Other)
Unknown
Cell−type
1
2
3
4
5
Region
Relative Frequency
1
2
3

#### Region map − spe3 AIH (delaunay_k5) − FOV: AIH09_s0_a3_ac

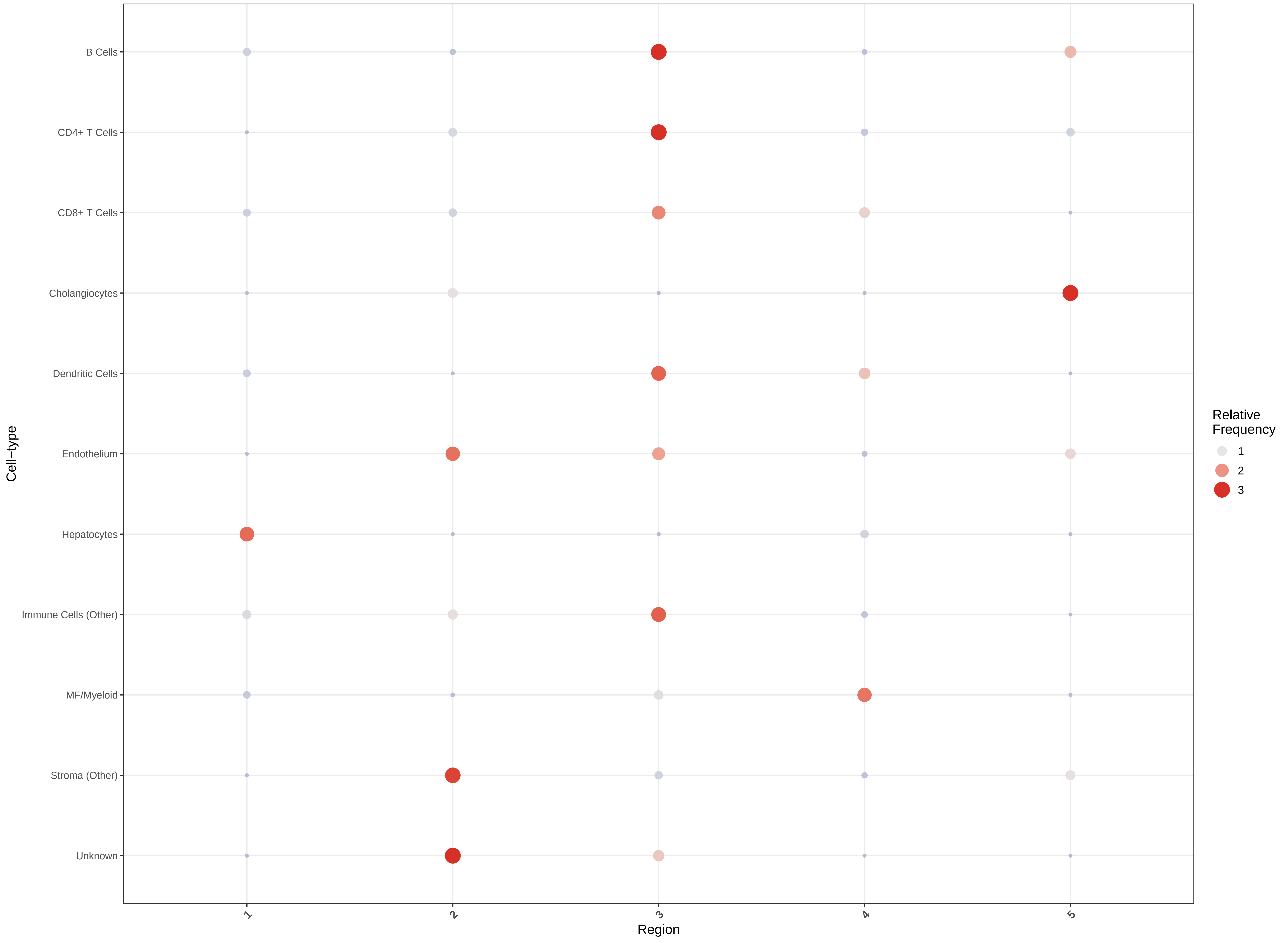

Region map − spe3 AIH (delaunay_k5) − FOV: AIH09_s0_a4_ac

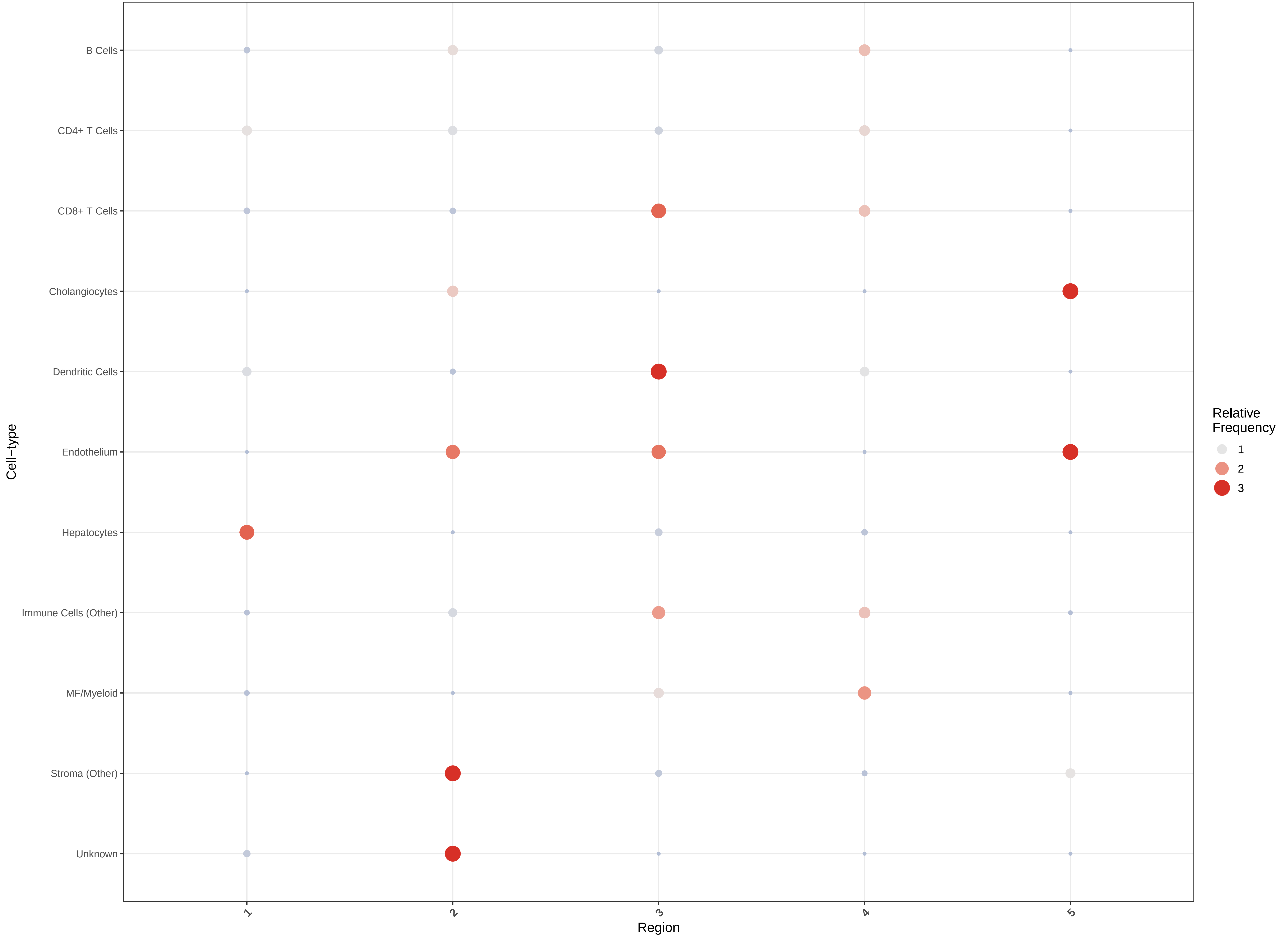

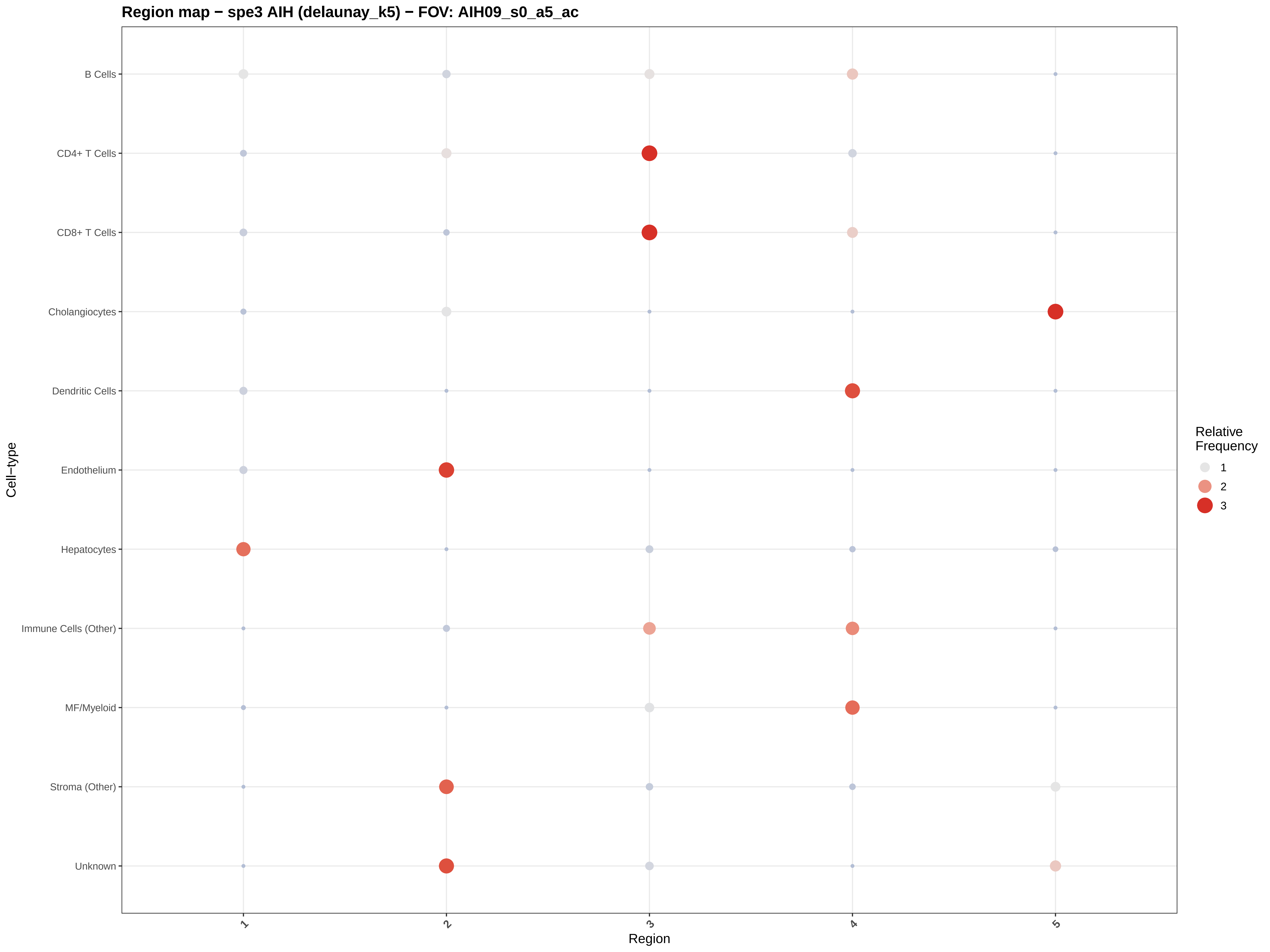
Region map − spe3 AIH (delaunay_k5) − FOV: AIH09_s0_a5_ac
B Cells
CD4+ T Cells
CD8+ T Cells
Cholangiocytes
Dendritic Cells
Endothelium
Hepatocytes
Immune Cells (Other)
MF/Myeloid
Stroma (Other)
Unknown
Cell−type
1
2
3
4
5
Region
Relative
Frequency
1
2
3

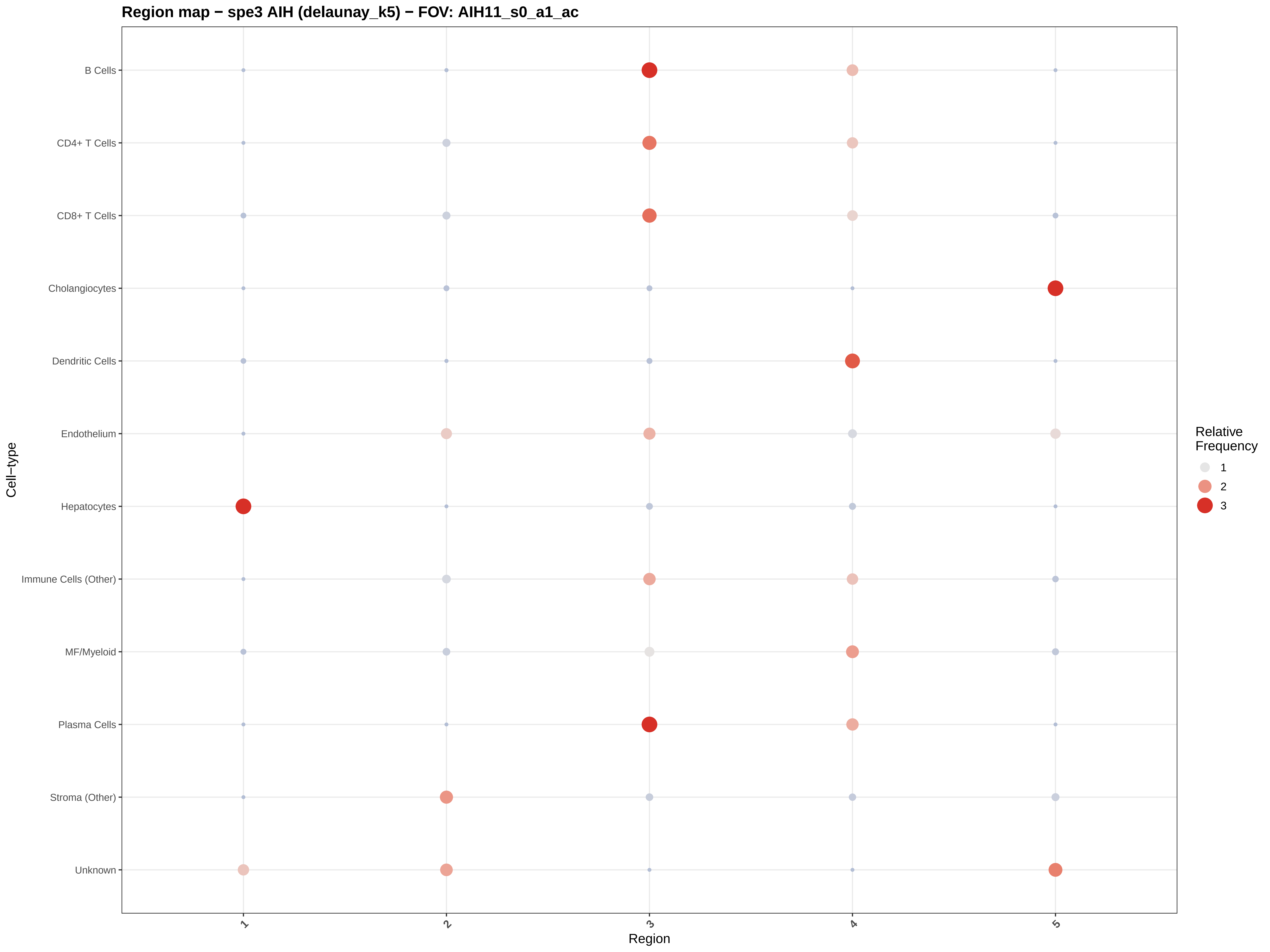

Region map − spe3 AIH (delaunay_k5) − FOV: AIH11_s0_a1_ac
B Cells
CD4+ T Cells
CD8+ T Cells
Cholangiocytes
Dendritic Cells
Endothelium
Hepatocytes
Immune Cells (Other)
MF/Myeloid
Plasma Cells
Stroma (Other)
Unknown
Cell−type
1
2
3
4
5
Region
Relative Frequency
1
2
3

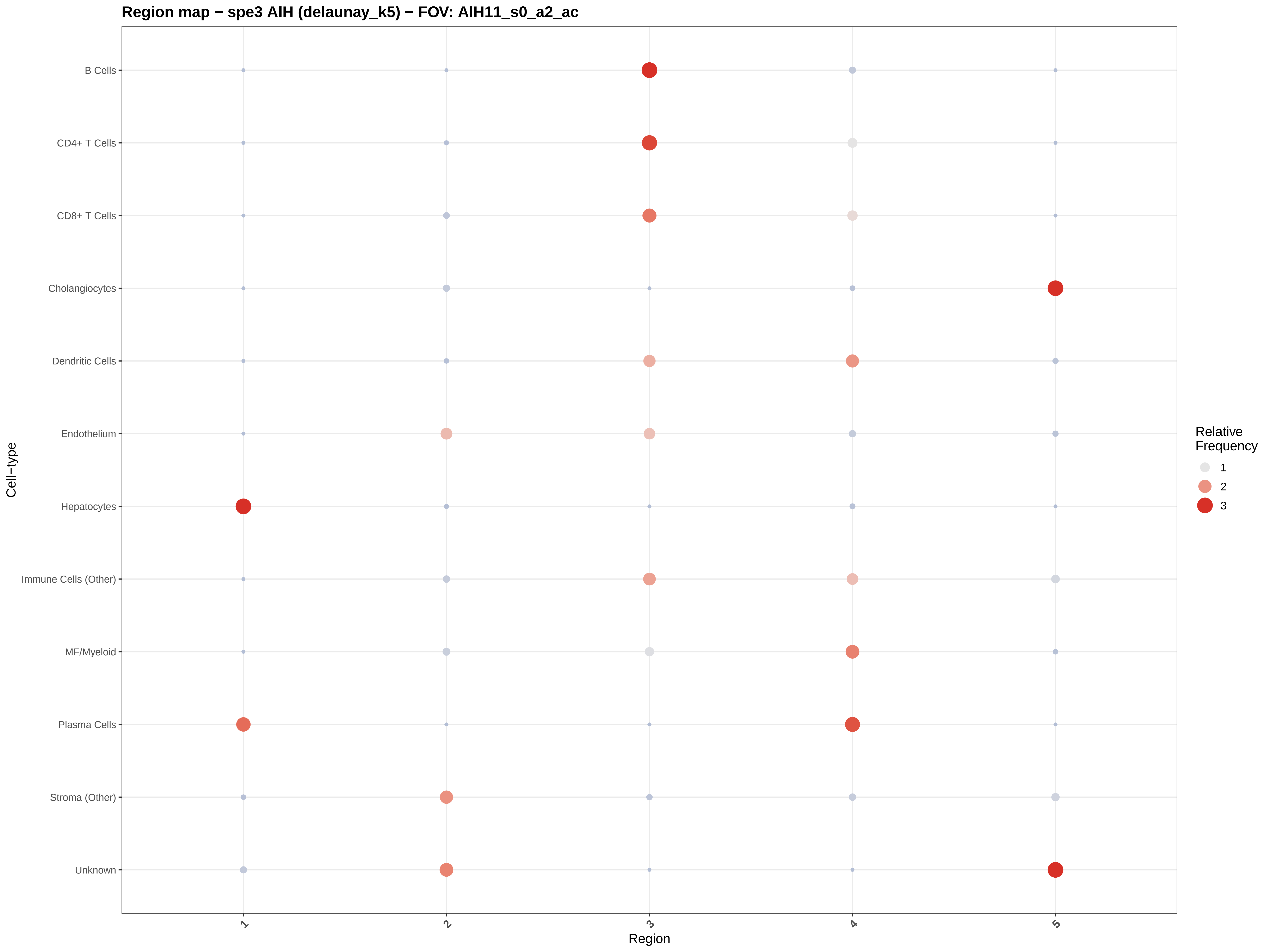
Region map − spe3 AIH (delaunay_k5) − FOV: AIH11_s0_a2_ac
B Cells
CD4+ T Cells
CD8+ T Cells
Cholangiocytes
Dendritic Cells
Endothelium
Hepatocytes
Immune Cells (Other)
MF/Myeloid
Plasma Cells
Stroma (Other)
Unknown
Cell−type
1
2
3
4
5
Region
Relative Frequency
1
2
3

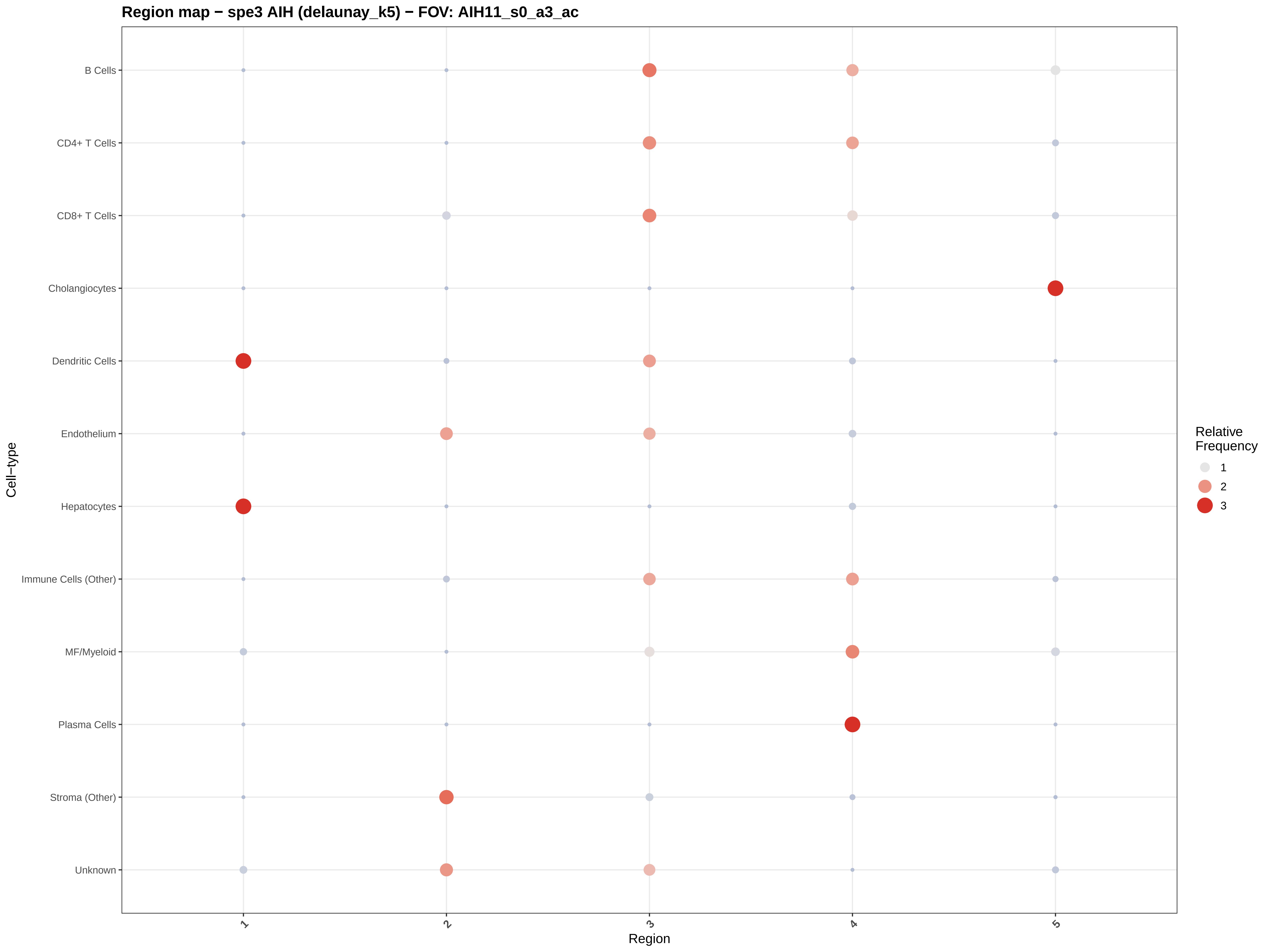

Region map − spe3 AIH (delaunay_k5) − FOV: AIH11_s0_a3_ac
B Cells
CD4+ T Cells
CD8+ T Cells
Cholangiocytes
Dendritic Cells
Endothelium
Hepatocytes
Immune Cells (Other)
MF/Myeloid
Plasma Cells
Stroma (Other)
Unknown
Cell−type
1
2
3
4
5
Region
Relative Frequency
1
2
3

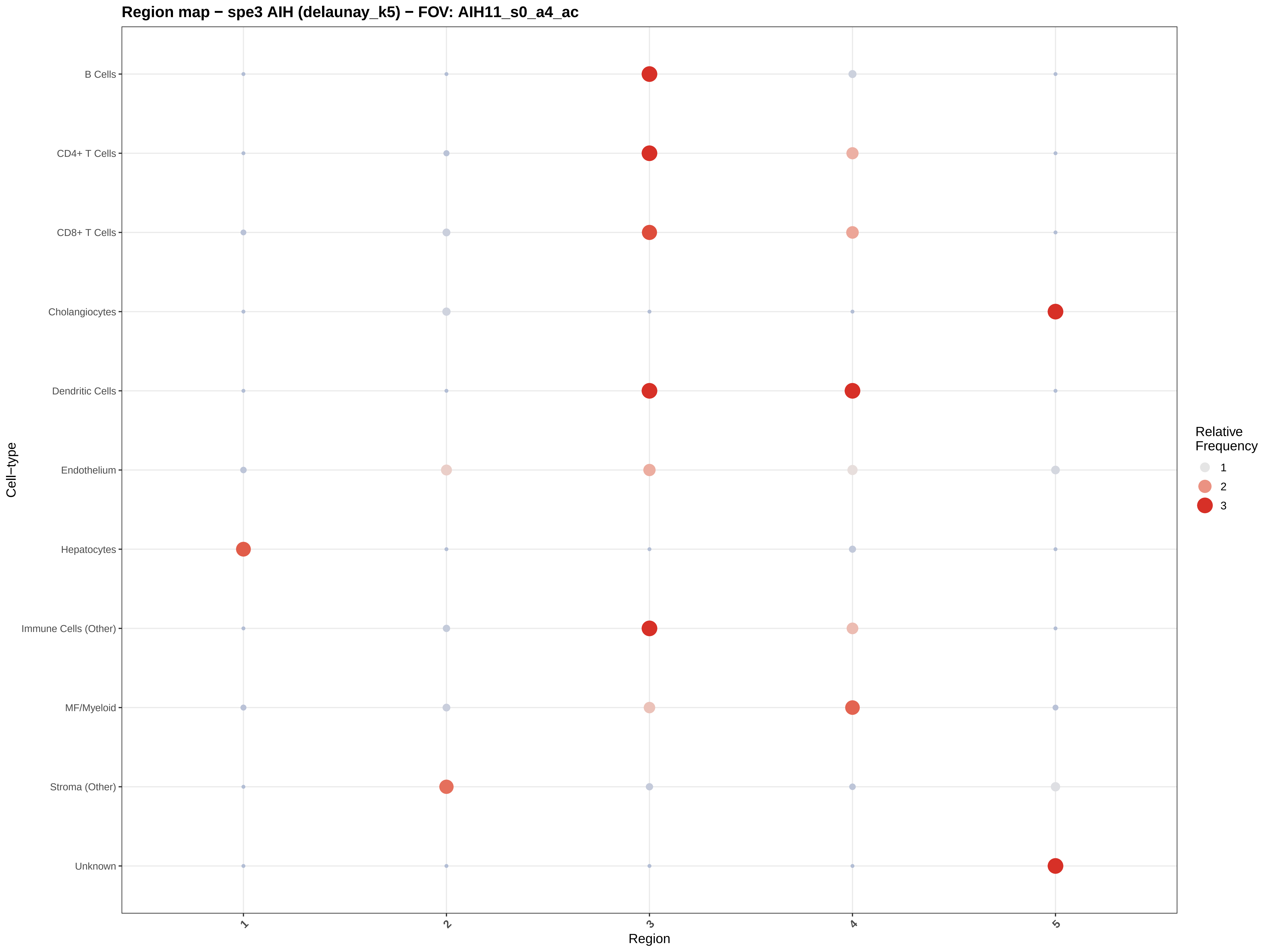

Region map − spe3 AIH (delaunay_k5) − FOV: AIH11_s0_a4_ac
B Cells
CD4+ T Cells
CD8+ T Cells
Cholangiocytes
Dendritic Cells
Endothelium
Hepatocytes
Immune Cells (Other)
MF/Myeloid
Stroma (Other)
Unknown
Cell−type
1
2
3
4
5
Region
Relative Frequency
1
2
3

#### Region map − spe3 AIH (delaunay_k5) − FOV: AIH11_s0_a5_ac

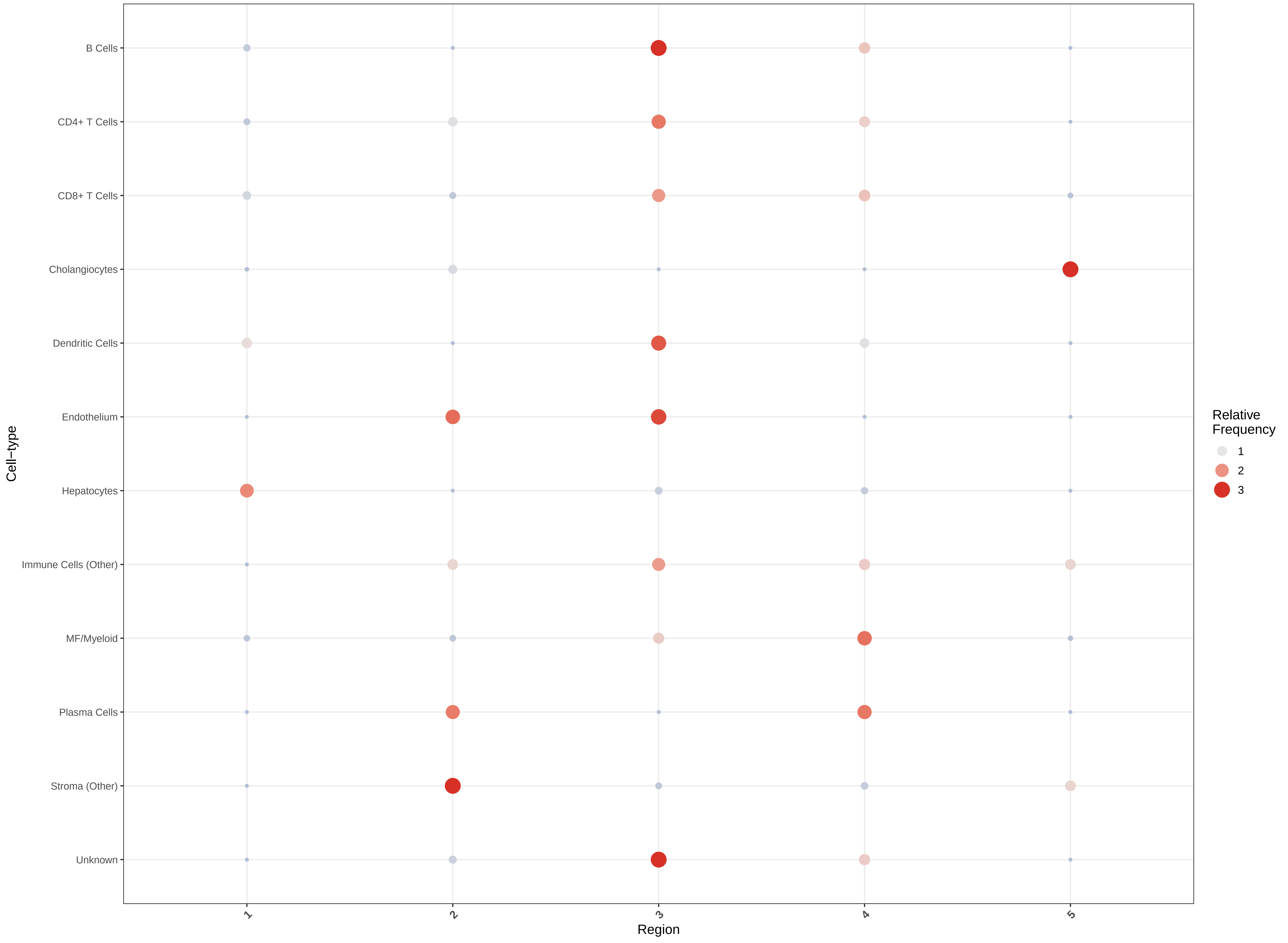

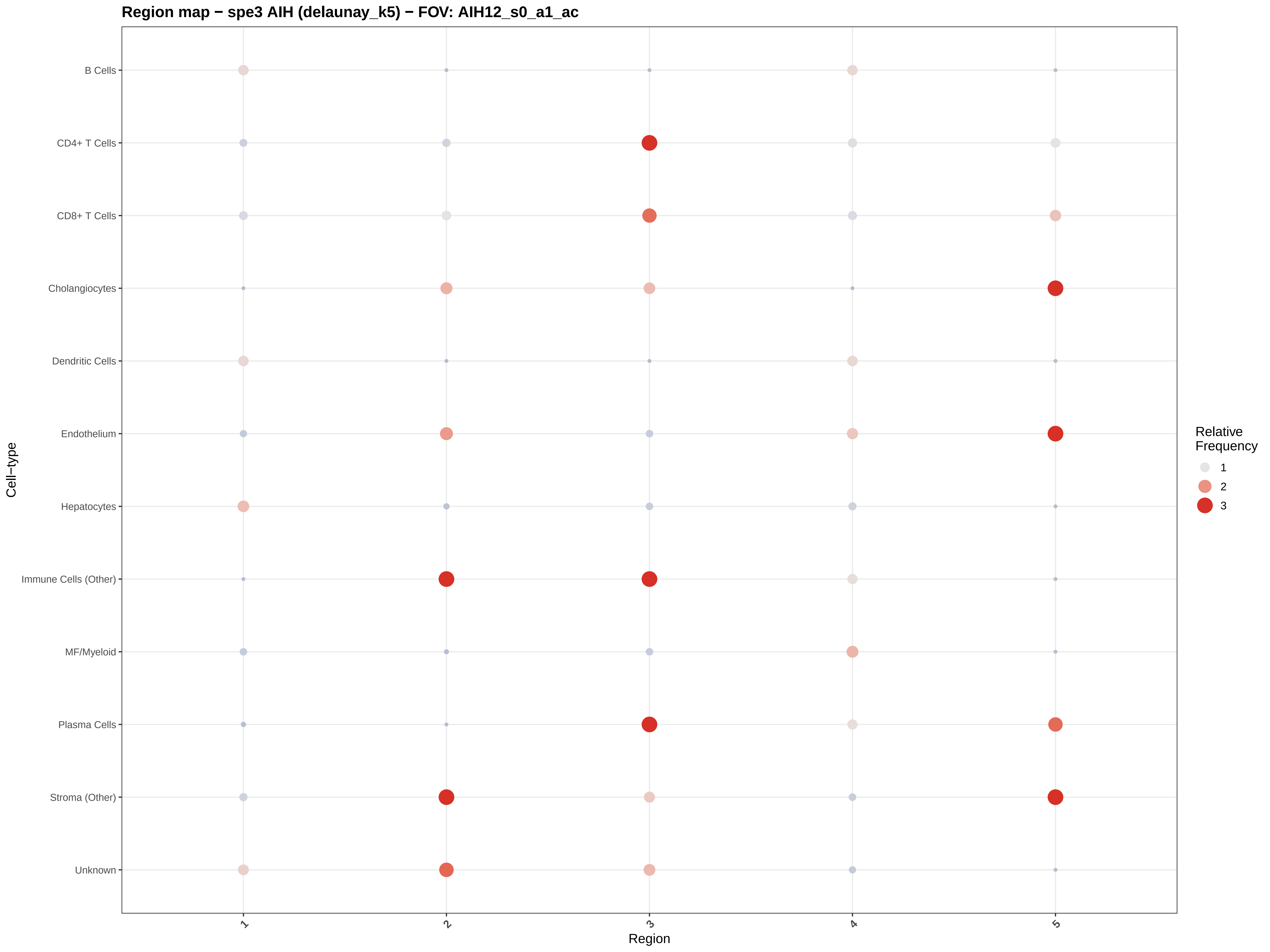
Region map − spe3 AIH (delaunay_k5) − FOV: AIH12_s0_a1_ac
B Cells
CD4+ T Cells
CD8+ T Cells
Cholangiocytes
Dendritic Cells
Endothelium
Hepatocytes
Immune Cells (Other)
MF/Myeloid
Plasma Cells
Stroma (Other)
Unknown
Cell−type
1
2
3
4
5
Region
Relative Frequency
1
2
3

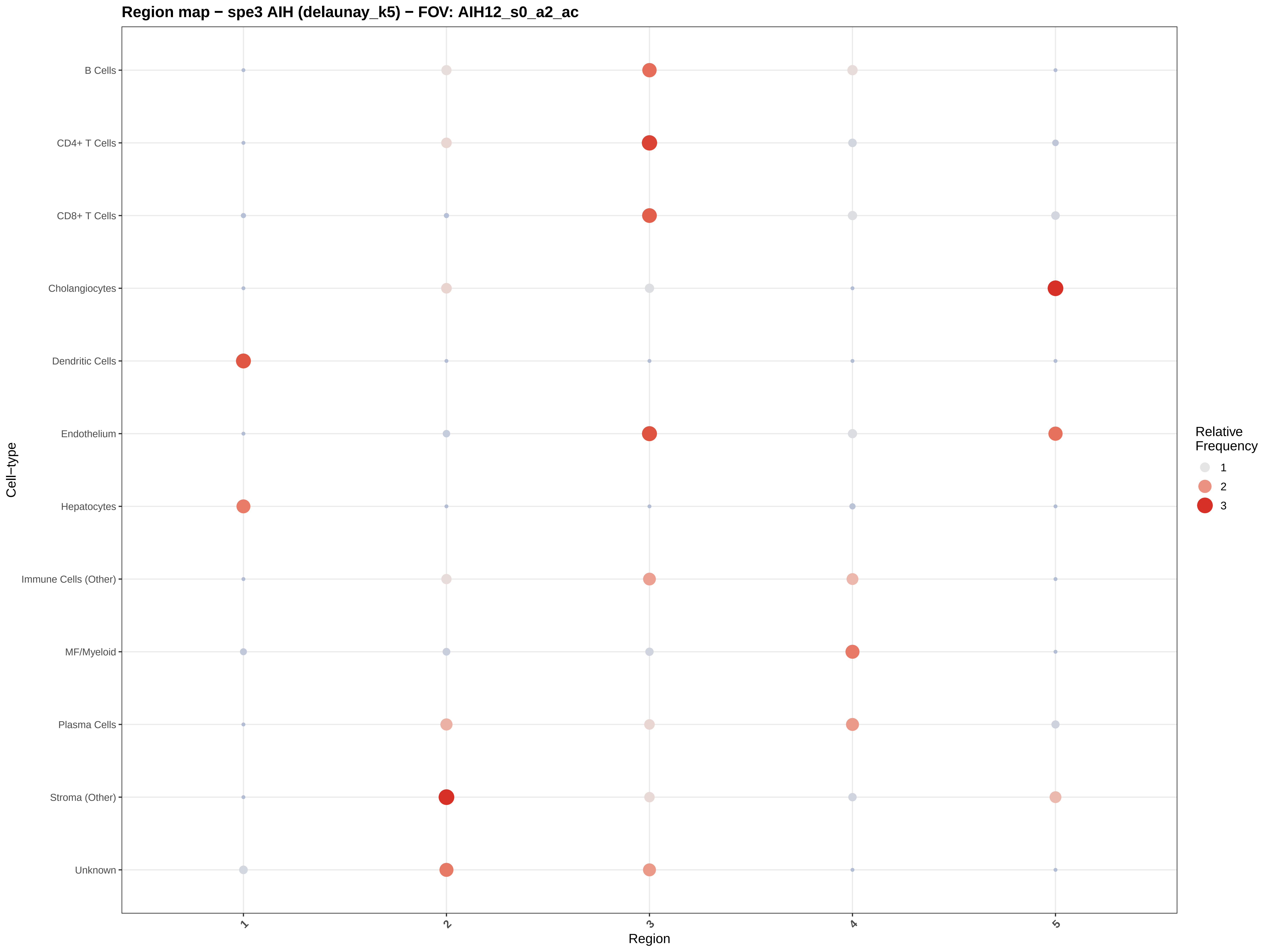
Region map − spe3 AIH (delaunay_k5) − FOV: AIH12_s0_a2_ac
B Cells
CD4+ T Cells
CD8+ T Cells
Cholangiocytes
Dendritic Cells
Endothelium
Hepatocytes
Immune Cells (Other)
MF/Myeloid
Plasma Cells
Stroma (Other)
Unknown
Cell−type
1
2
3
4
5
Region
Relative Frequency
1
2
3

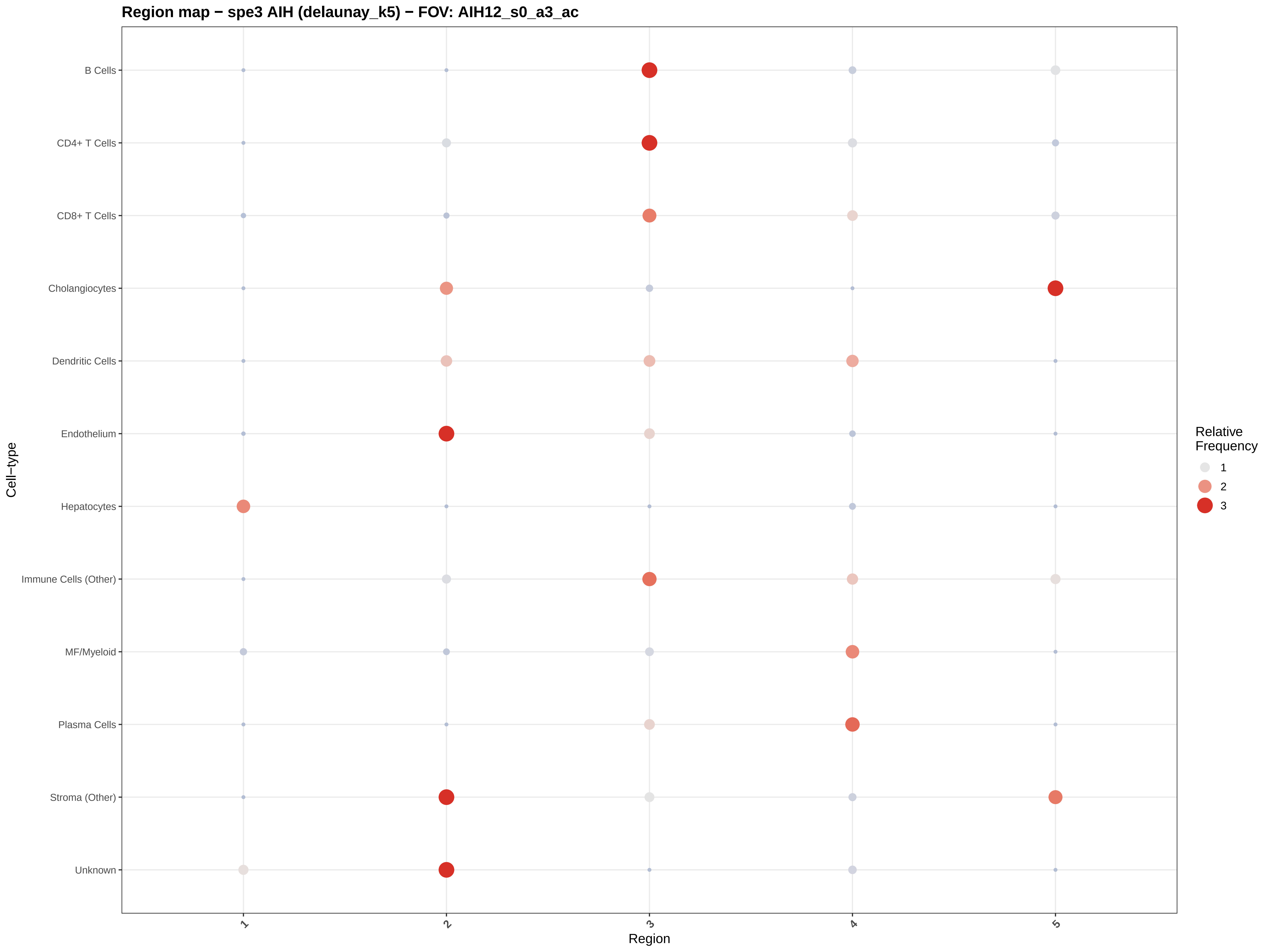

Region map − spe3 AIH (delaunay_k5) − FOV: AIH12_s0_a3_ac
B Cells
CD4+ T Cells
CD8+ T Cells
Cholangiocytes
Dendritic Cells
Endothelium
Hepatocytes
Immune Cells (Other)
MF/Myeloid
Plasma Cells
Stroma (Other)
Unknown
Cell−type
1
2
3
4
5
Region
Relative Frequency
1
2
3

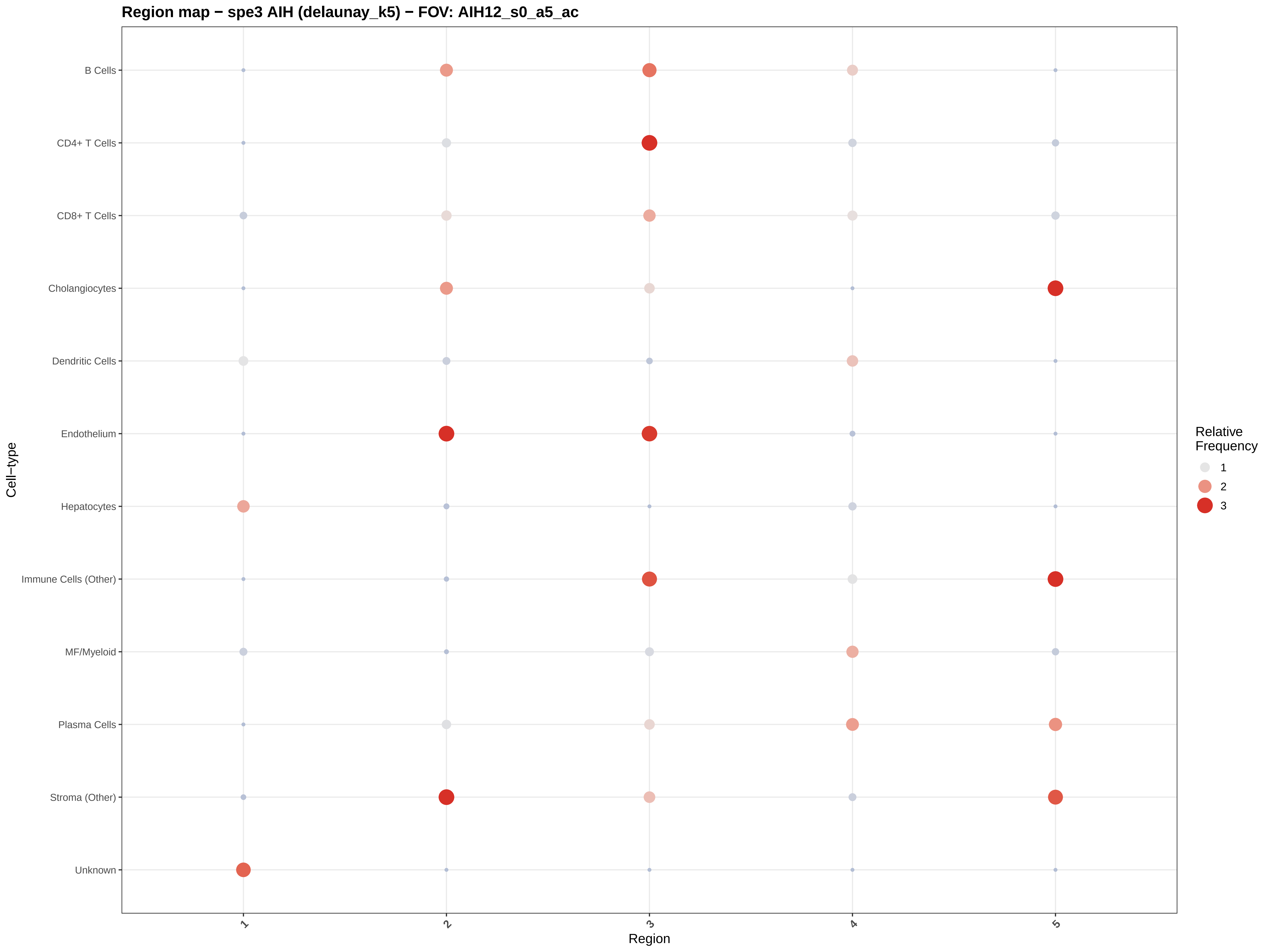

Region map − spe3 AIH (delaunay_k5) − FOV: AIH12_s0_a5_ac
B Cells
CD4+ T Cells
CD8+ T Cells
Cholangiocytes
Dendritic Cells
Endothelium
Hepatocytes
Immune Cells (Other)
MF/Myeloid
Plasma Cells
Stroma (Other)
Unknown
Cell−type
1
2
3
4
5
Region
Relative Frequency
1
2
3

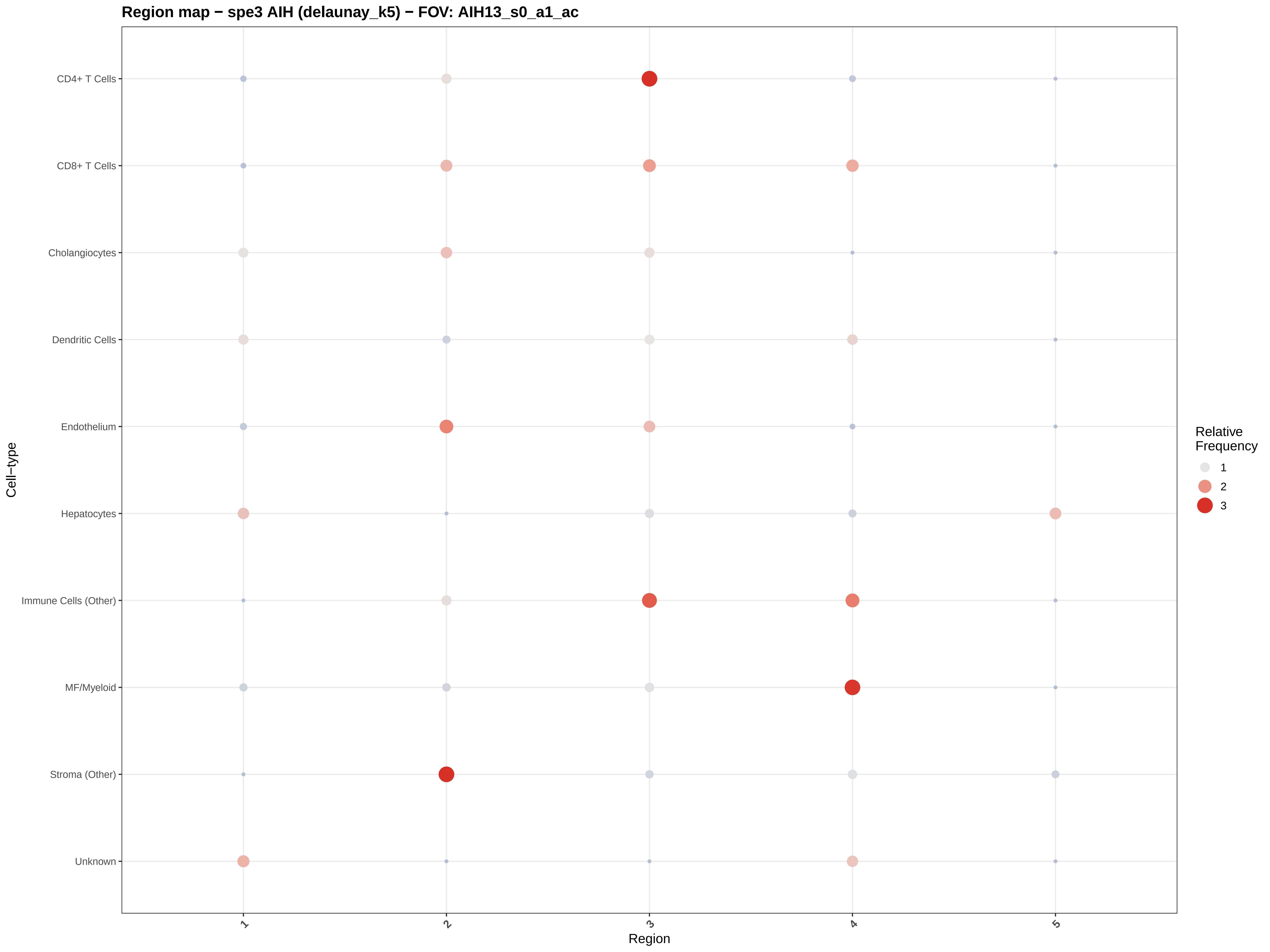
Region map − spe3 AIH (delaunay_k5) − FOV: AIH13_s0_a1_ac
CD4+ T Cells
CD8+ T Cells
Cholangiocytes
Dendritic Cells
Endothelium
Hepatocytes
Immune Cells (Other)
MF/Myeloid
Stroma (Other)
Unknown
Cell−type
1
2
3
4
5
Region
Relative Frequency
1
2
3

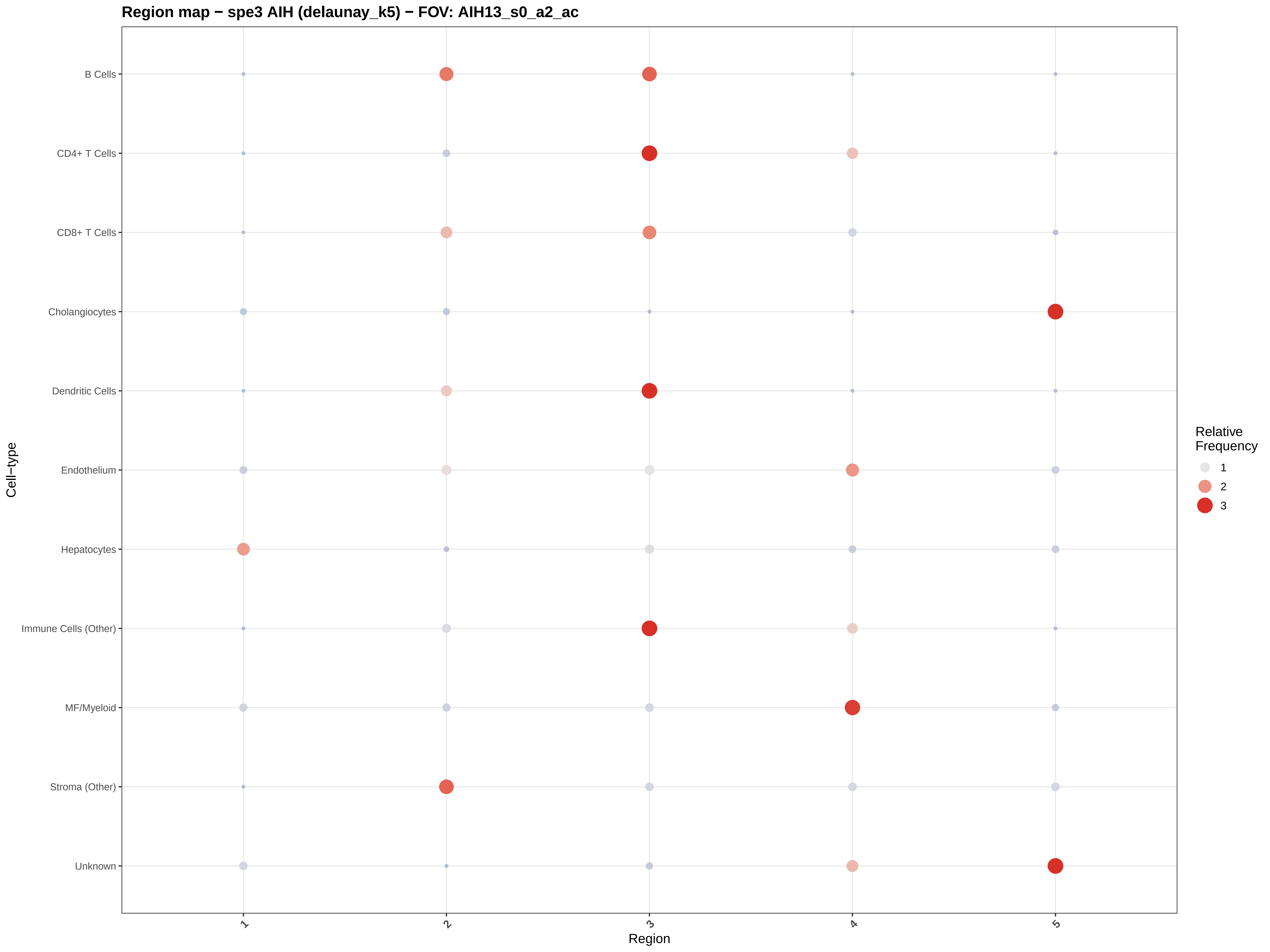

Region map − spe3 AIH (delaunay_k5) − FOV: AIH13_s0_a2_ac
B Cells
CD4+ T Cells
CD8+ T Cells
Cholangiocytes
Dendritic Cells
Endothelium
Hepatocytes
Immune Cells (Other)
MF/Myeloid
Stroma (Other)
Unknown
Cell−type
1
2
3
4
5
Region
Relative Frequency
1
2
3

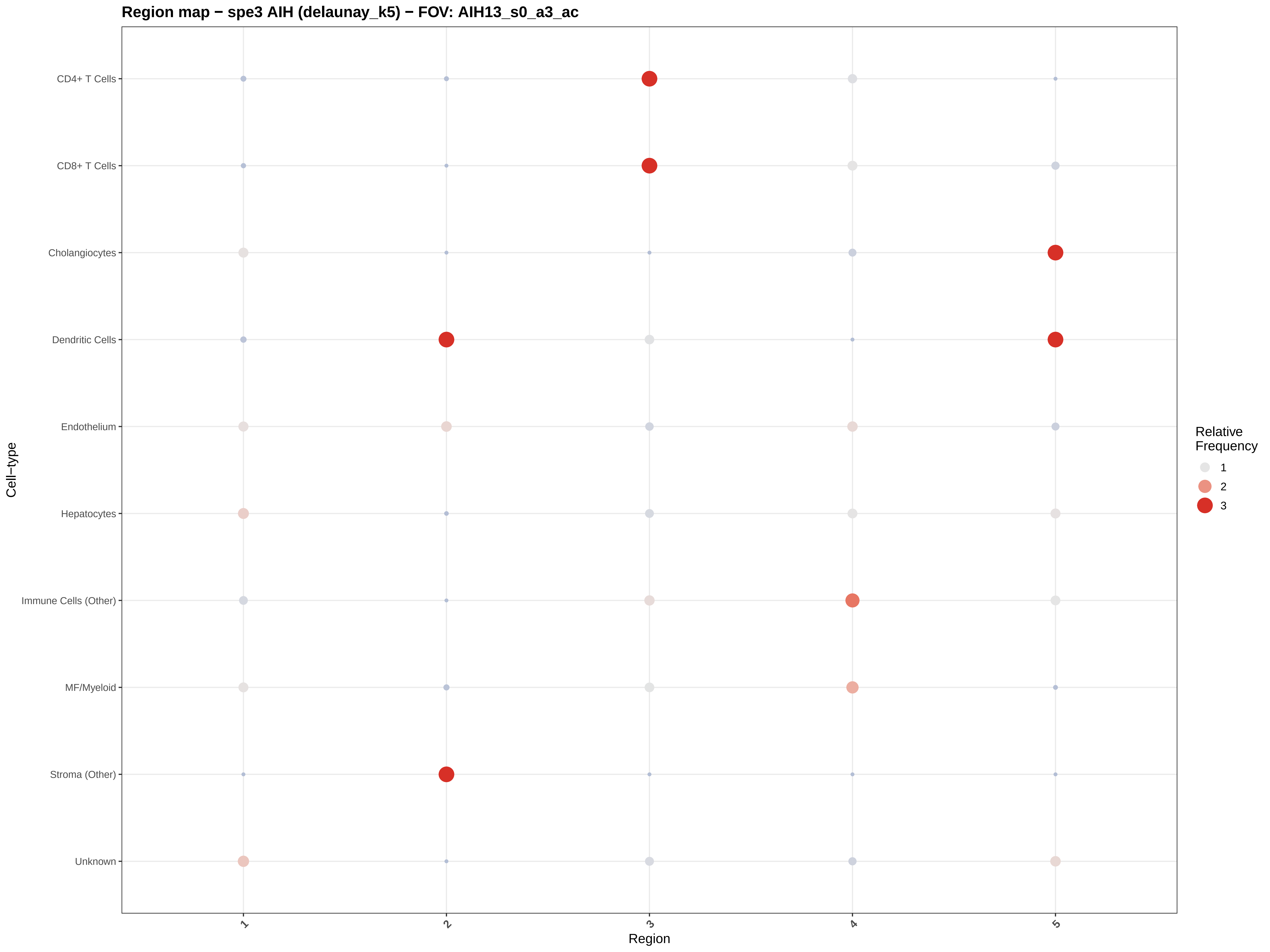
Region map − spe3 AIH (delaunay_k5) − FOV: AIH13_s0_a3_ac
CD4+ T Cells
CD8+ T Cells
Cholangiocytes
Dendritic Cells
Endothelium
Hepatocytes
Immune Cells (Other)
MF/Myeloid
Stroma (Other)
Unknown
Cell−type
1
2
3
4
5
Region
Relative Frequency
1
2
3

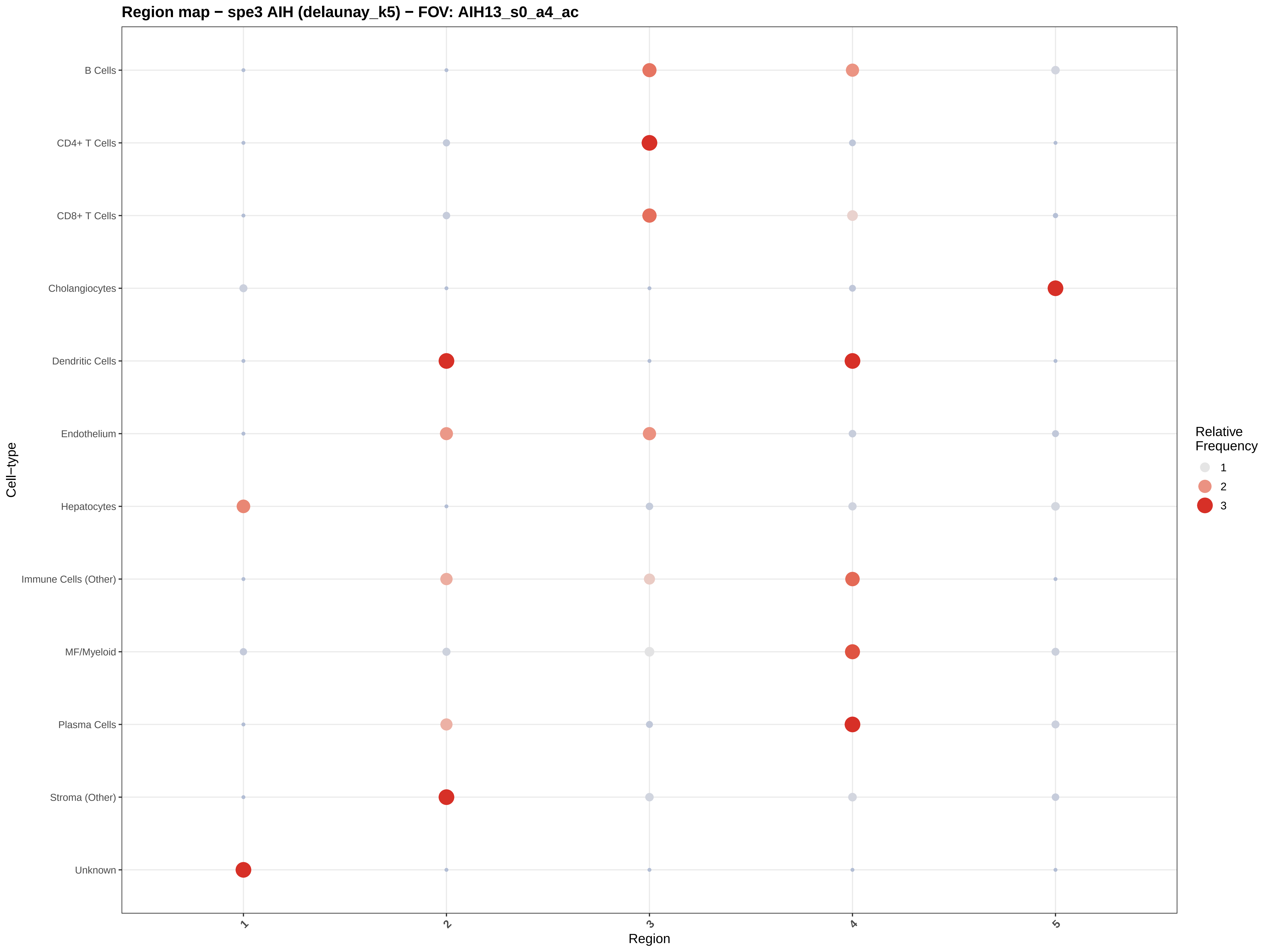
Region map − spe3 AIH (delaunay_k5) − FOV: AIH13_s0_a4_ac
B Cells
CD4+ T Cells
CD8+ T Cells
Cholangiocytes
Dendritic Cells
Endothelium
Hepatocytes
Immune Cells (Other)
MF/Myeloid
Plasma Cells
Stroma (Other)
Unknown
Cell−type
1
2
3
4
5
Region
Relative
Frequency
1
2
3

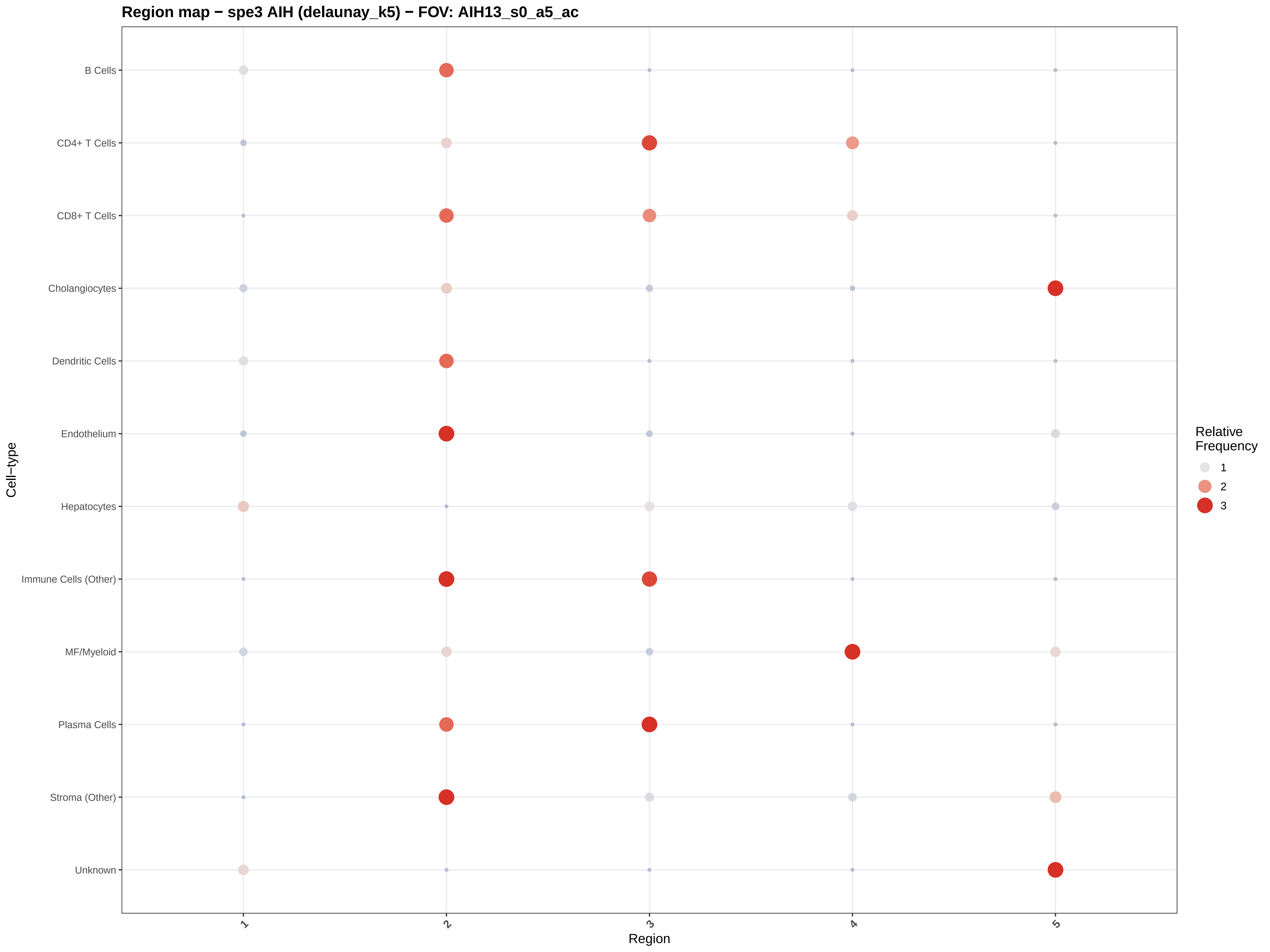
Region map − spe3 AIH (delaunay_k5) − FOV: AIH13_s0_a5_ac
B Cells
CD4+ T Cells
CD8+ T Cells
Cholangiocytes
Dendritic Cells
Endothelium
Hepatocytes
Immune Cells (Other)
MF/Myeloid
Plasma Cells
Stroma (Other)
Unknown
Cell−type
1
2
3
4
5
Region
Relative Frequency
1
2
3

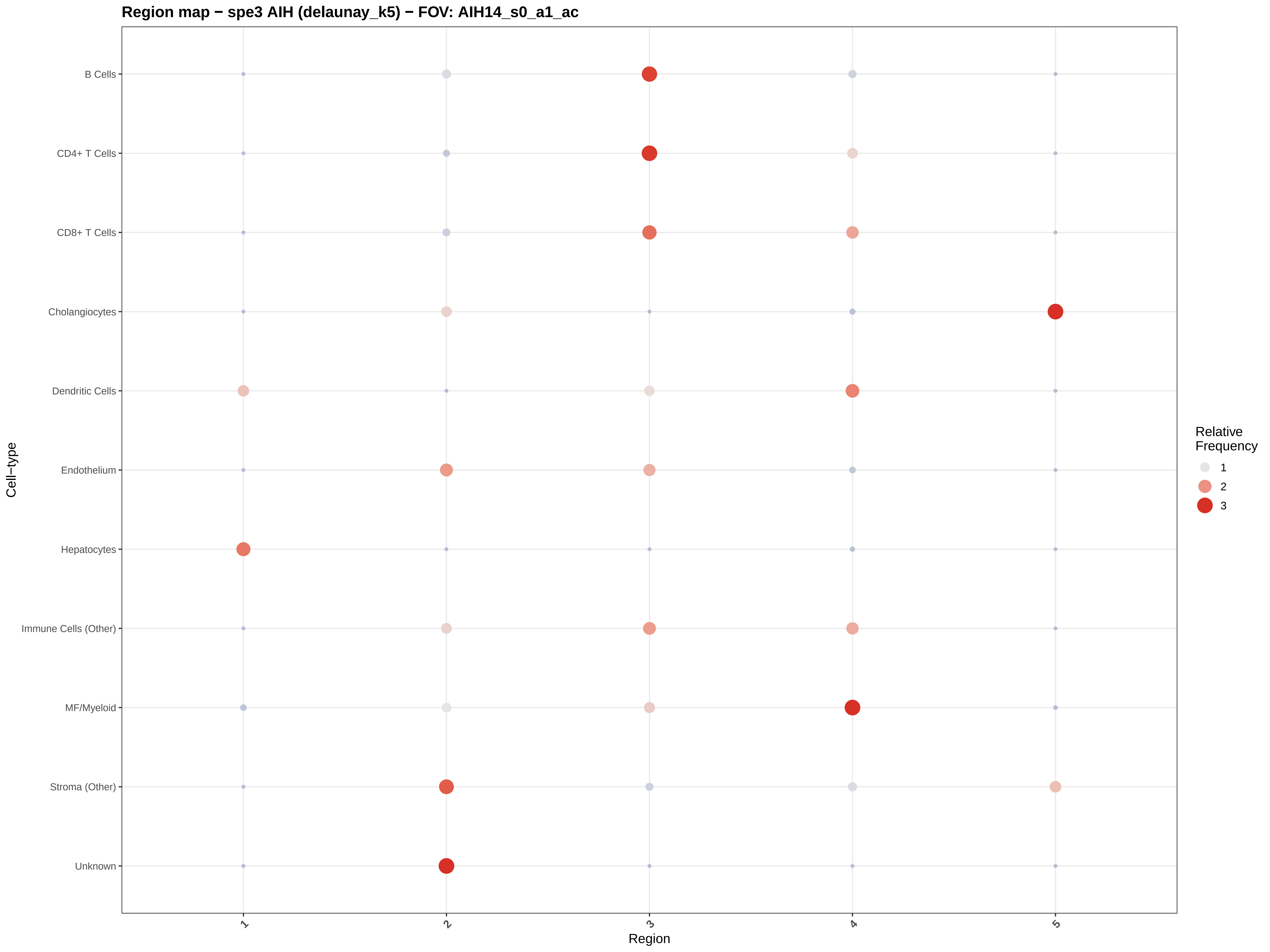

Region map − spe3 AIH (delaunay_k5) − FOV: AIH14_s0_a1_ac
B Cells
CD4+ T Cells
CD8+ T Cells
Cholangiocytes
Dendritic Cells
Endothelium
Hepatocytes
Immune Cells (Other)
MF/Myeloid
Stroma (Other)
Unknown
Cell−type
1
2
3
4
5
Region
Relative Frequency
1
2
3

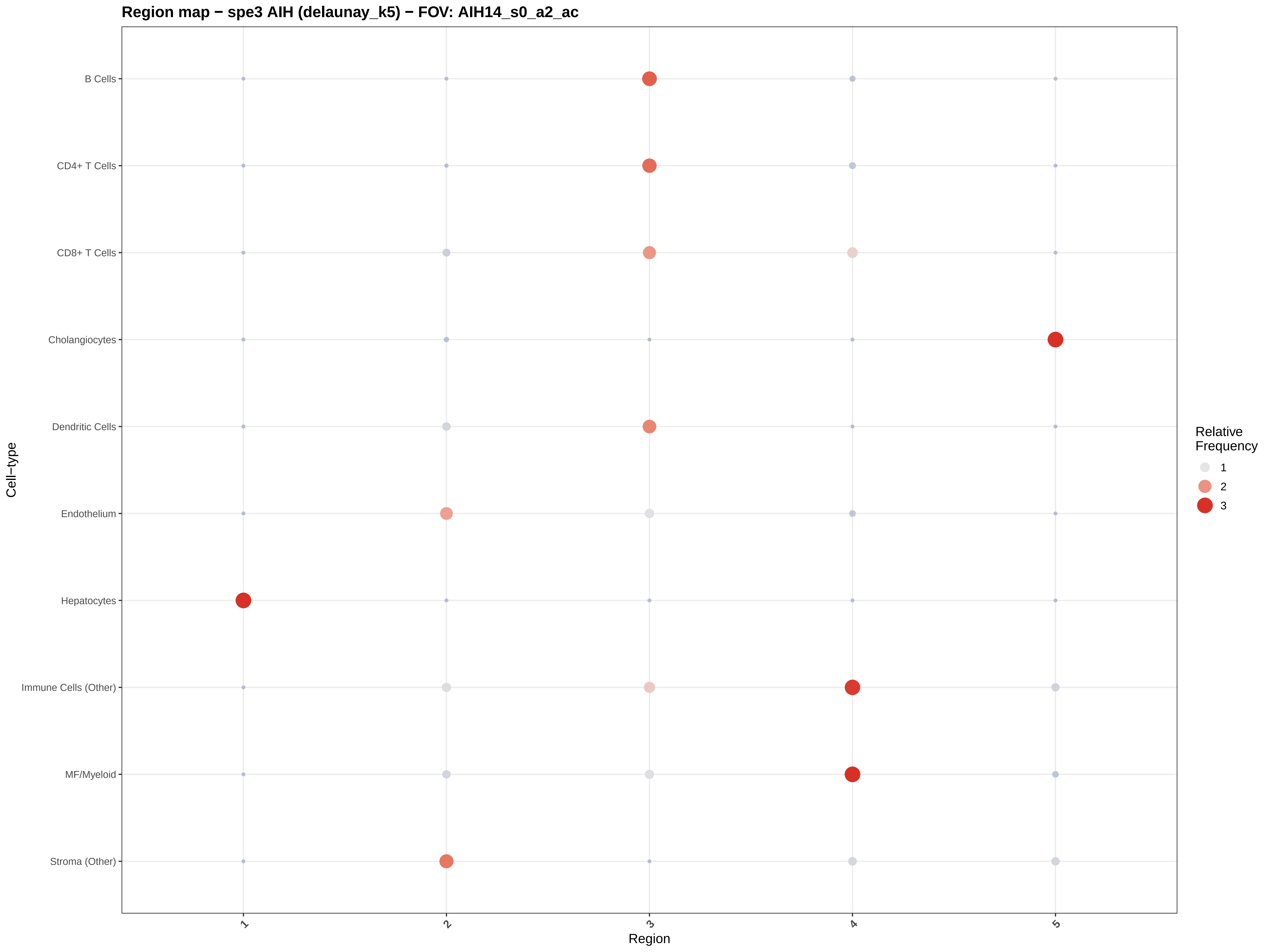

Region map − spe3 AIH (delaunay_k5) − FOV: AIH14_s0_a2_ac
B Cells
CD4+ T Cells
CD8+ T Cells
Cholangiocytes
Dendritic Cells
Endothelium
Hepatocytes
Immune Cells (Other)
MF/Myeloid
Stroma (Other)
Cell−type
1
2
3
4
5
Region
Relative Frequency
1
2
3

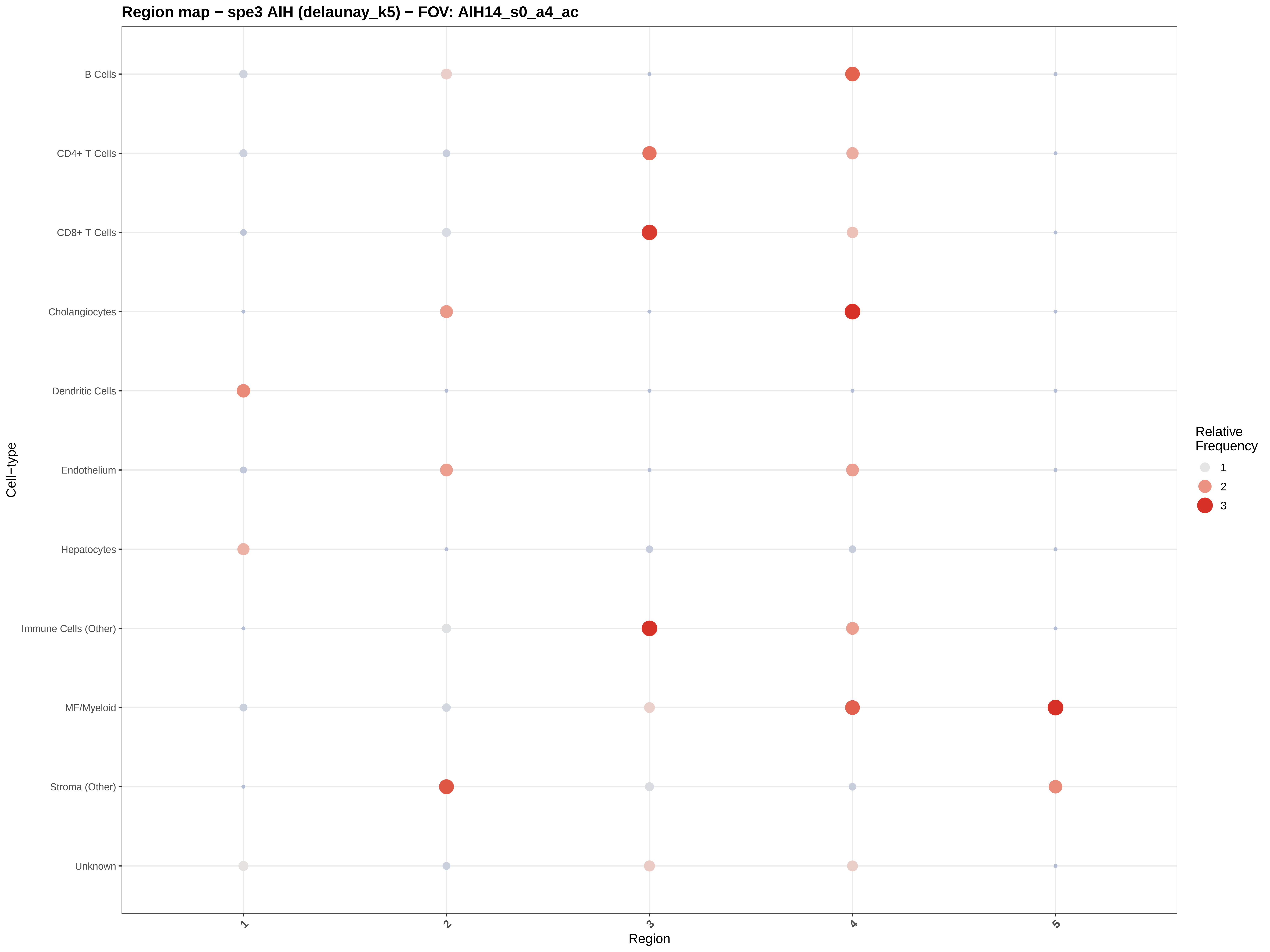
Region map − spe3 AIH (delaunay_k5) − FOV: AIH14_s0_a4_ac
B Cells
CD4+ T Cells
CD8+ T Cells
Cholangiocytes
Dendritic Cells
Endothelium
Hepatocytes
Immune Cells (Other)
MF/Myeloid
Stroma (Other)
Unknown
Cell−type
1
2
3
4
5
Region
Relative Frequency
1
2
3

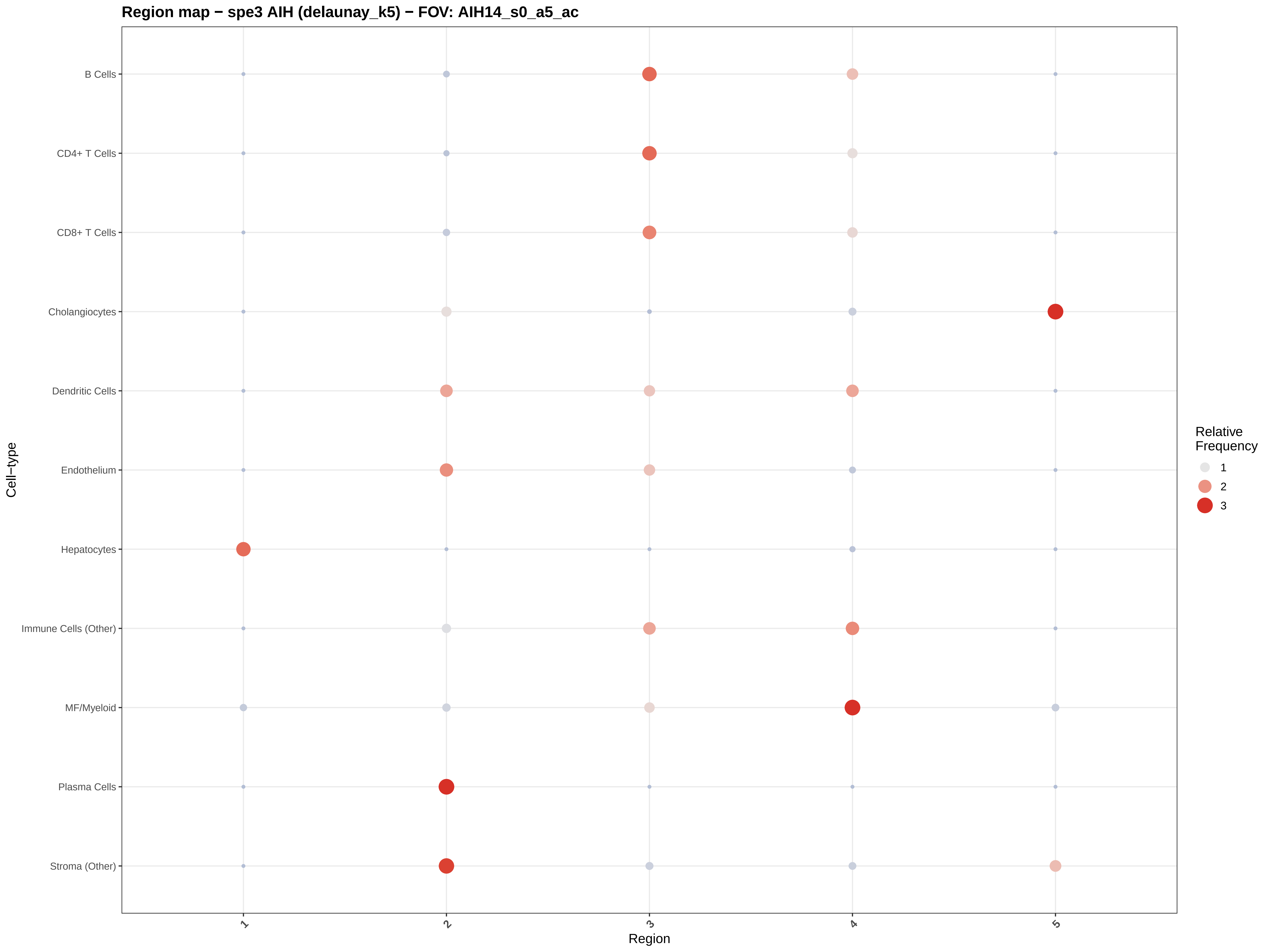

Region map − spe3 AIH (delaunay_k5) − FOV: AIH14_s0_a5_ac
B Cells
CD4+ T Cells
CD8+ T Cells
Cholangiocytes
Dendritic Cells
Endothelium
Hepatocytes
Immune Cells (Other)
MF/Myeloid
Plasma Cells
Stroma (Other)
Cell−type
1
2
3
4
5
Region
Relative Frequency
1
2
3

#### Region map − spe3 AIH (delaunay_k5) − FOV: AIH15_s0_a1_ac

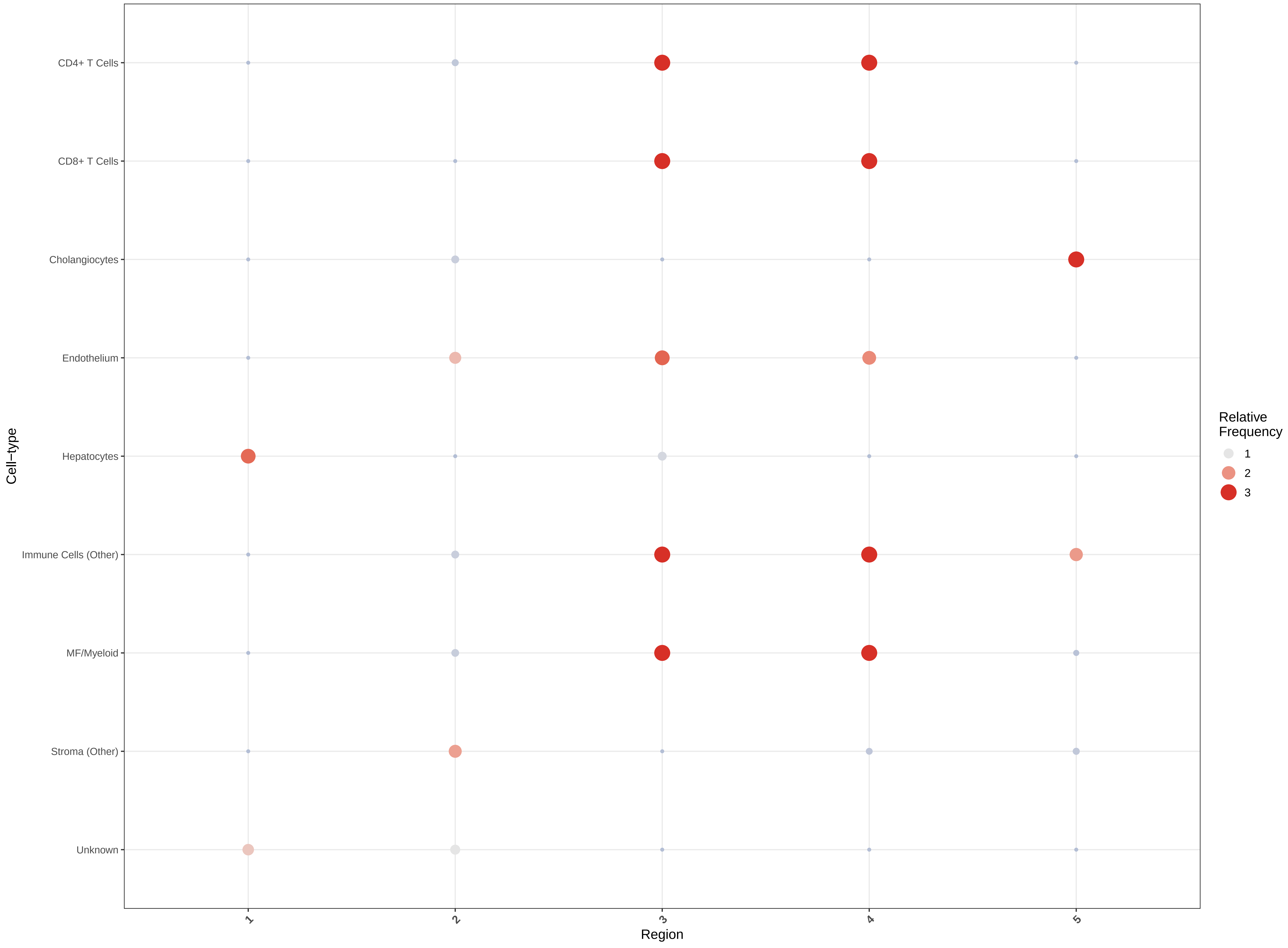

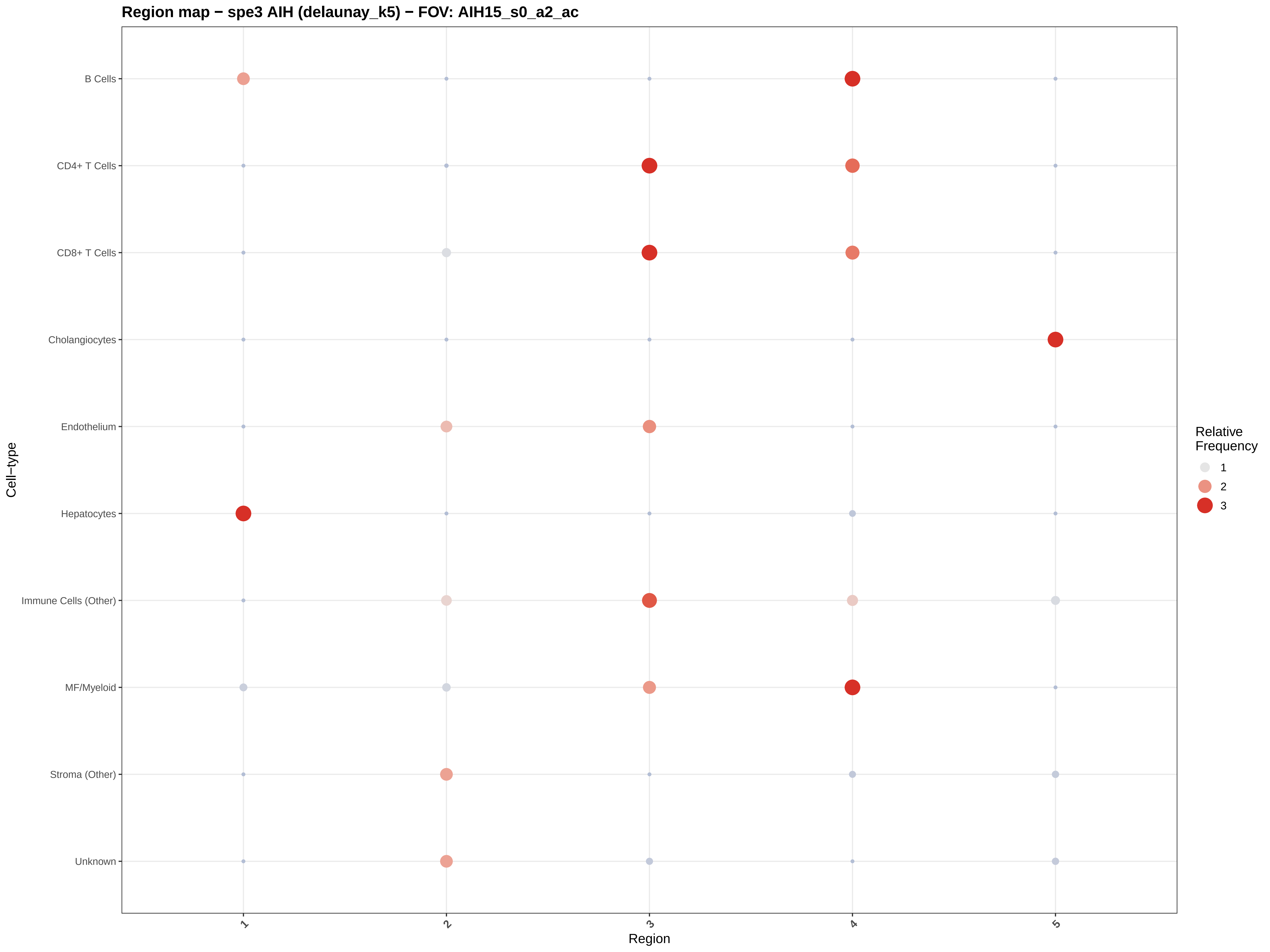
Region map − spe3 AIH (delaunay_k5) − FOV: AIH15_s0_a2_ac
B Cells
CD4+ T Cells
CD8+ T Cells
Cholangiocytes
Endothelium
Hepatocytes
Immune Cells (Other)
MF/Myeloid
Stroma (Other)
Unknown
Cell−type
1
2
3
4
5
Region
Relative Frequency
1
2
3

#### Region map − spe3 AIH (delaunay_k5) − FOV: AIH15_s0_a3_ac

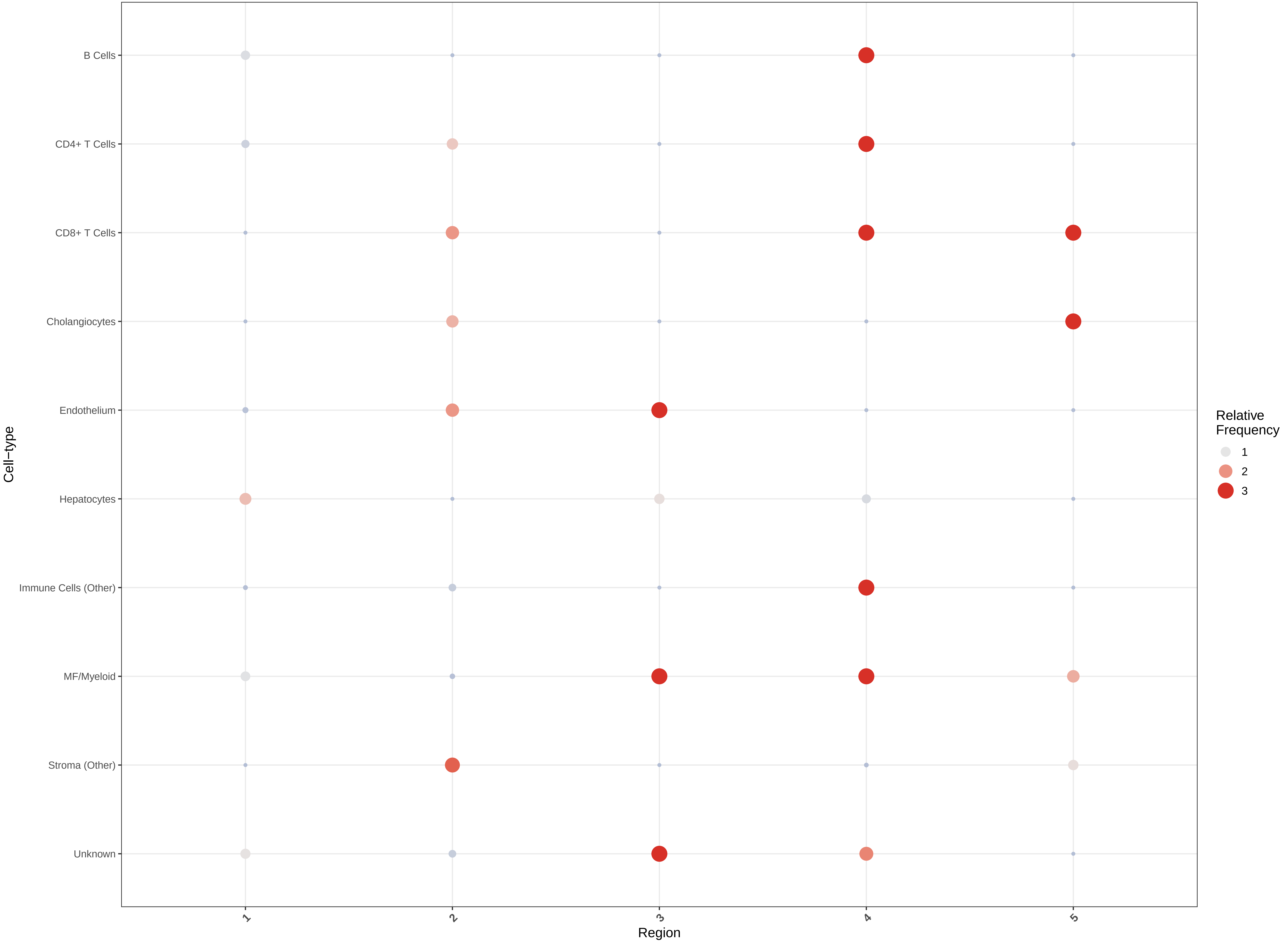

**Region map − spe3 AIH (delaunay_k5) − FOV: AIH15_s0_a4_ac**

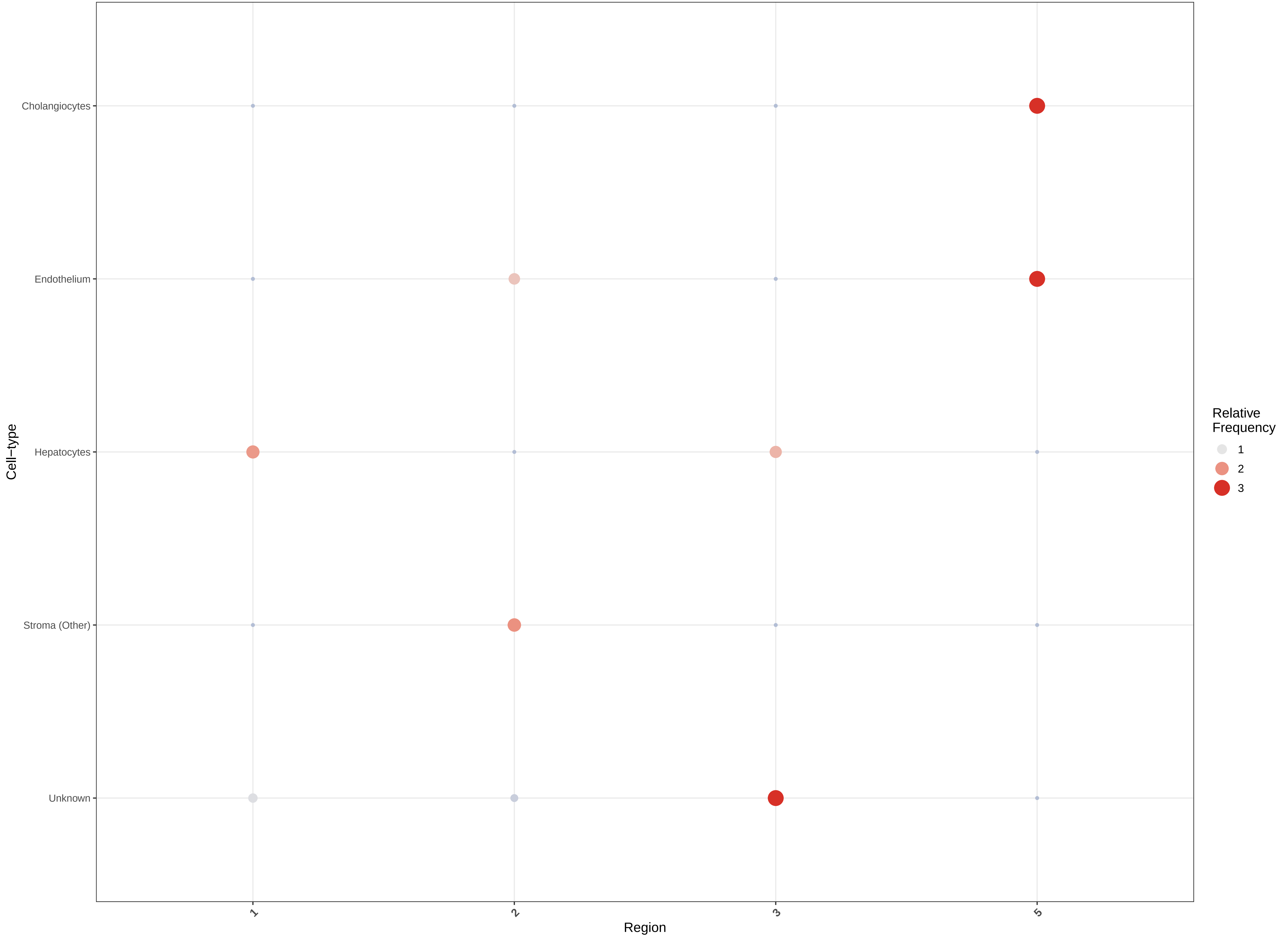

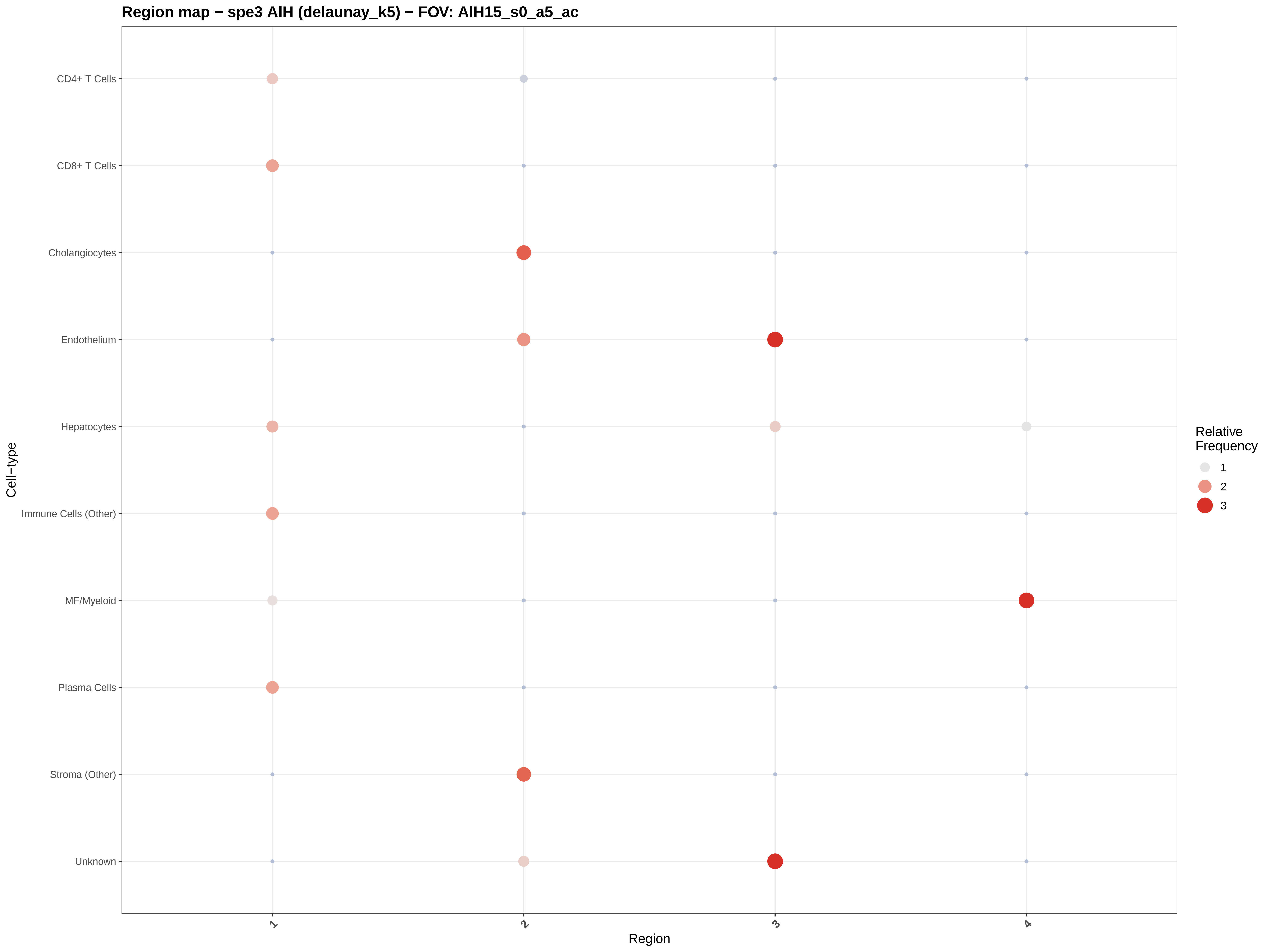
Region map − spe3 AIH (delaunay_k5) − FOV: AIH15_s0_a5_ac
CD4+ T Cells
CD8+ T Cells
Cholangiocytes
Endothelium
Hepatocytes
Immune Cells (Other)
MF/Myeloid
Plasma Cells
Stroma (Other)
Unknown
Cell−type
1
2
3
4
Region
Relative Frequency
1
2
3

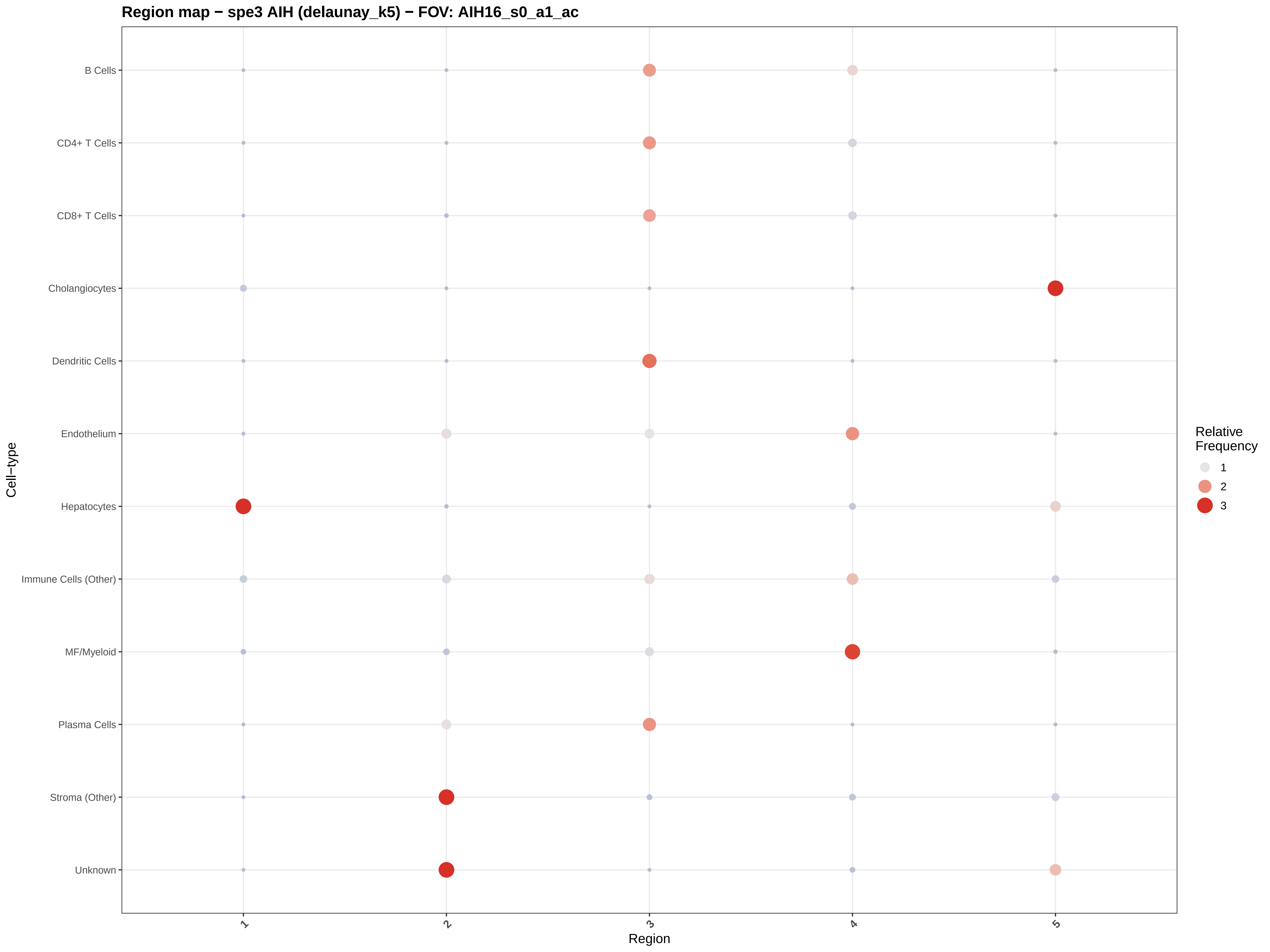
Region map − spe3 AIH (delaunay_k5) − FOV: AIH16_s0_a1_ac
B Cells
CD4+ T Cells
CD8+ T Cells
Cholangiocytes
Dendritic Cells
Endothelium
Hepatocytes
Immune Cells (Other)
MF/Myeloid
Plasma Cells
Stroma (Other)
Unknown
Cell−type
1
2
3
4
5
Region
Relative Frequency
1
2
3

#### Region map − spe3 AIH (delaunay_k5) − FOV: AIH16_s0_a2_ac

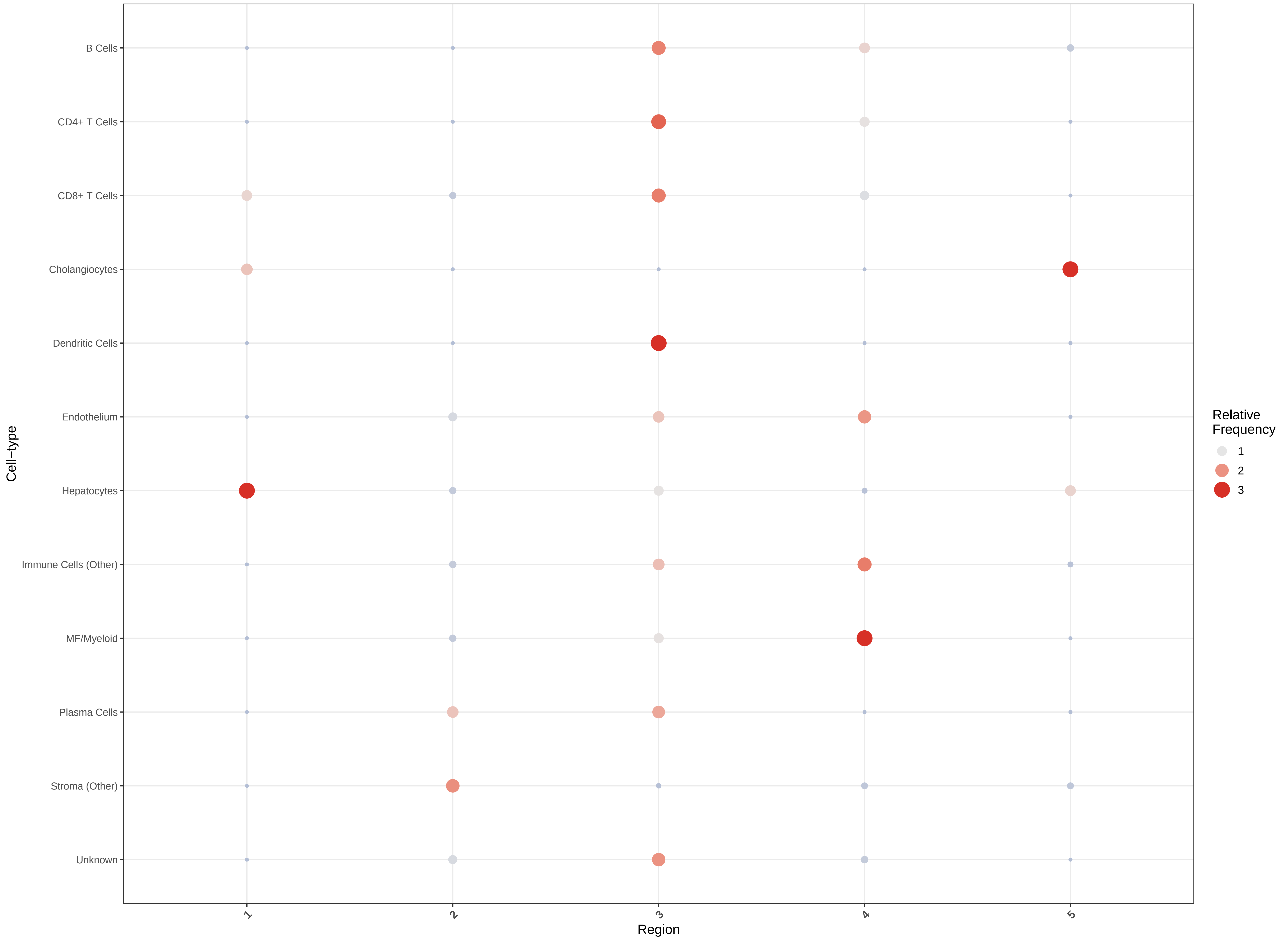

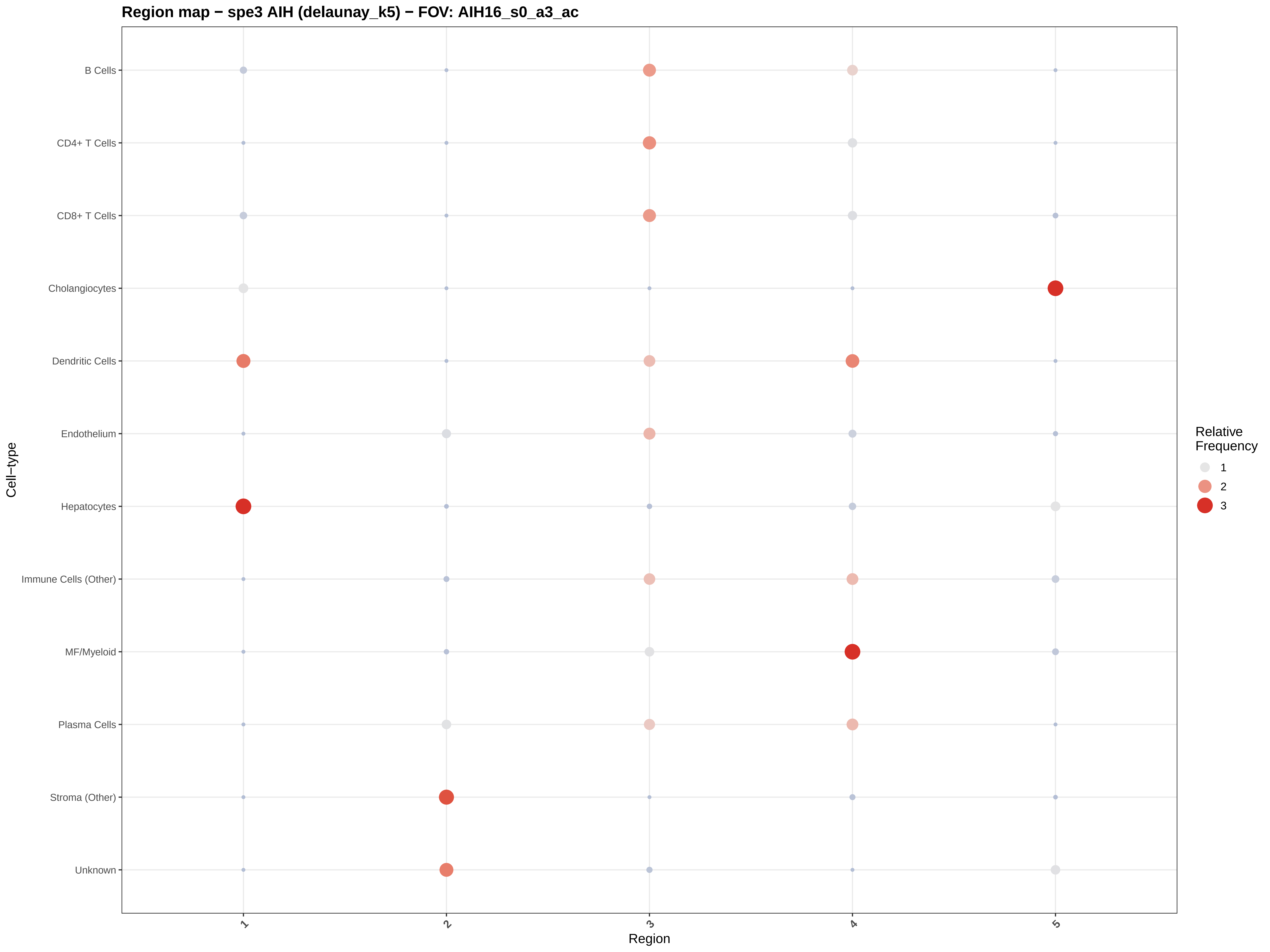
Region map − spe3 AIH (delaunay_k5) − FOV: AIH16_s0_a3_ac
B Cells
CD4+ T Cells
CD8+ T Cells
Cholangiocytes
Dendritic Cells
Endothelium
Hepatocytes
Immune Cells (Other)
MF/Myeloid
Plasma Cells
Stroma (Other)
Unknown
Cell−type
1
2
3
4
5
Region
Relative Frequency
1
2
3

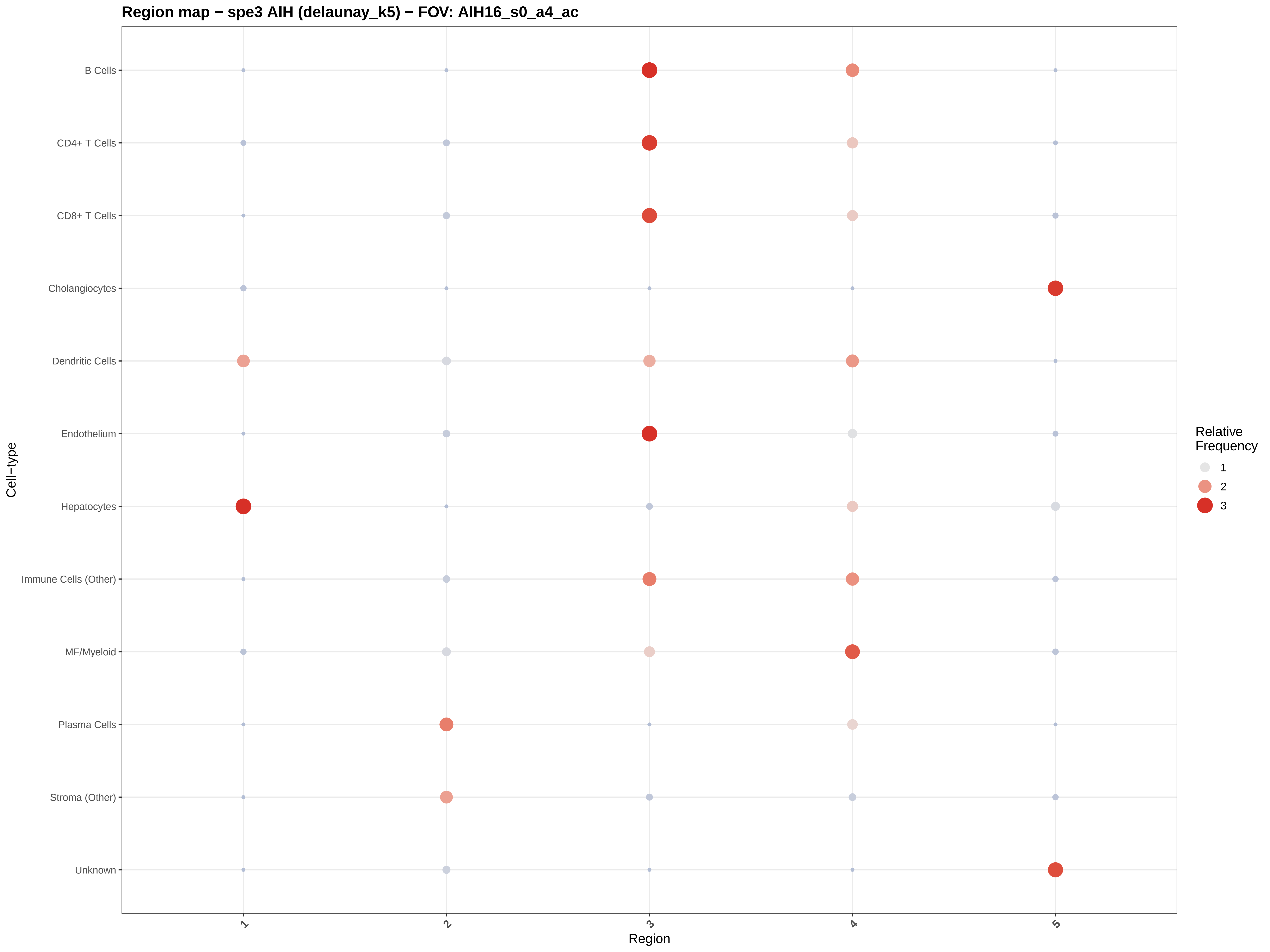
Region map − spe3 AIH (delaunay_k5) − FOV: AIH16_s0_a4_ac
B Cells
CD4+ T Cells
CD8+ T Cells
Cholangiocytes
Dendritic Cells
Endothelium
Hepatocytes
Immune Cells (Other)
MF/Myeloid
Plasma Cells
Stroma (Other)
Unknown
Cell−type
1
2
3
4
5
Region
Relative Frequency
1
2
3

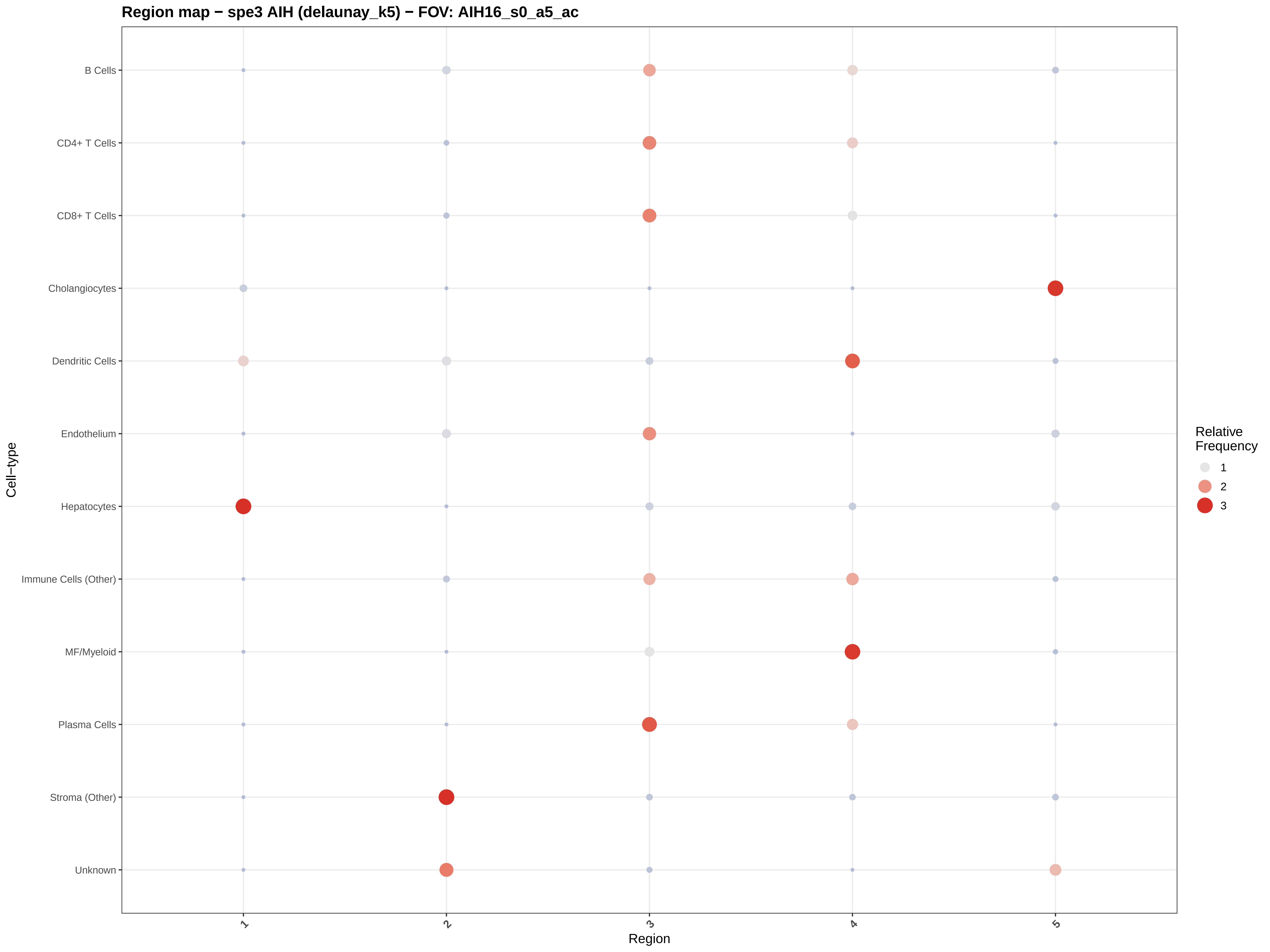

Region map − spe3 AIH (delaunay_k5) − FOV: AIH16_s0_a5_ac
B Cells
CD4+ T Cells
CD8+ T Cells
Cholangiocytes
Dendritic Cells
Endothelium
Hepatocytes
Immune Cells (Other)
MF/Myeloid
Plasma Cells
Stroma (Other)
Unknown
Cell−type
1
2
3
4
5
Region
Relative Frequency
1
2
3

#### CN (delaunay, k=5) − spe3 AIH − FOV: AIH01_s0_a2_ac

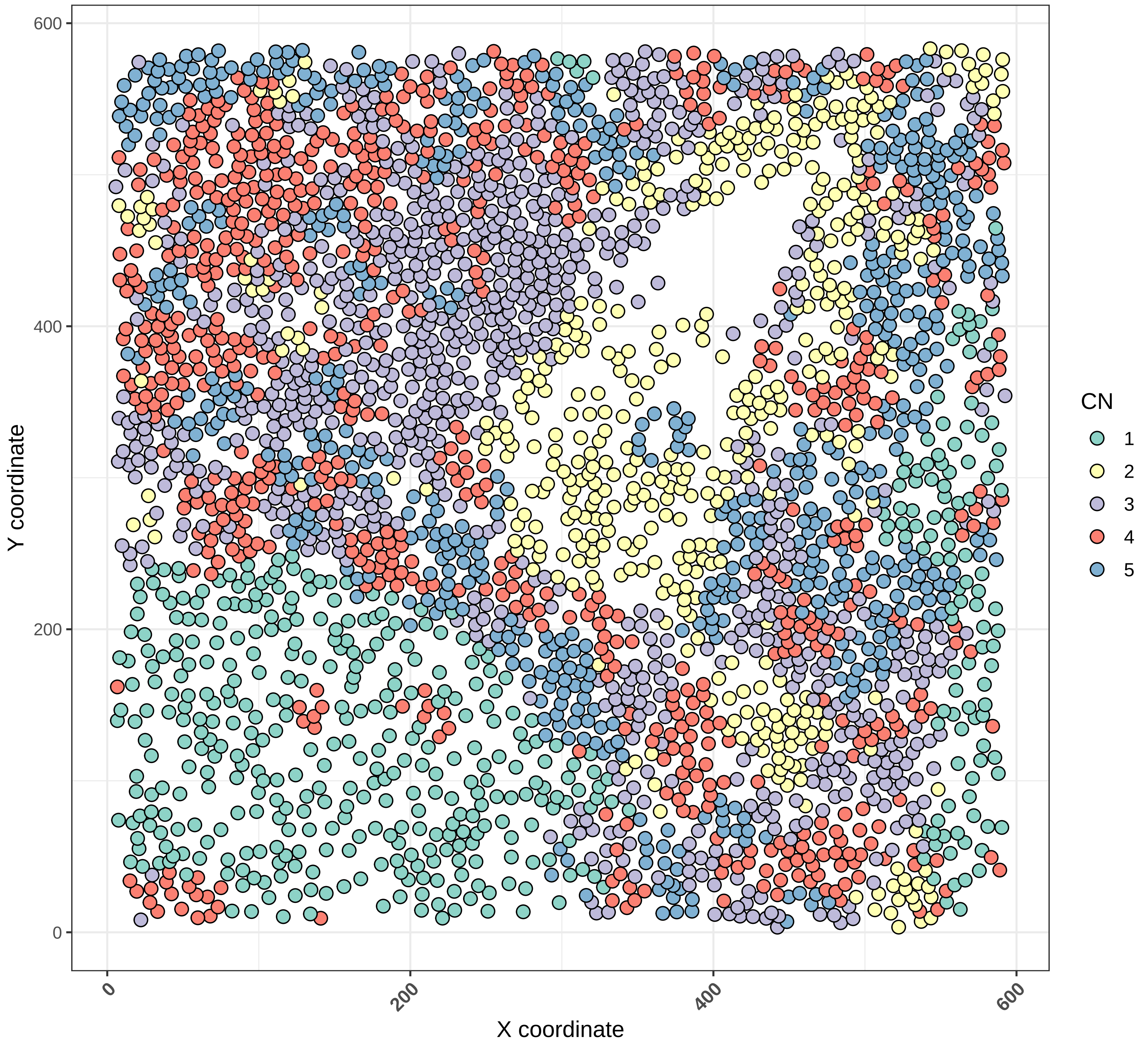

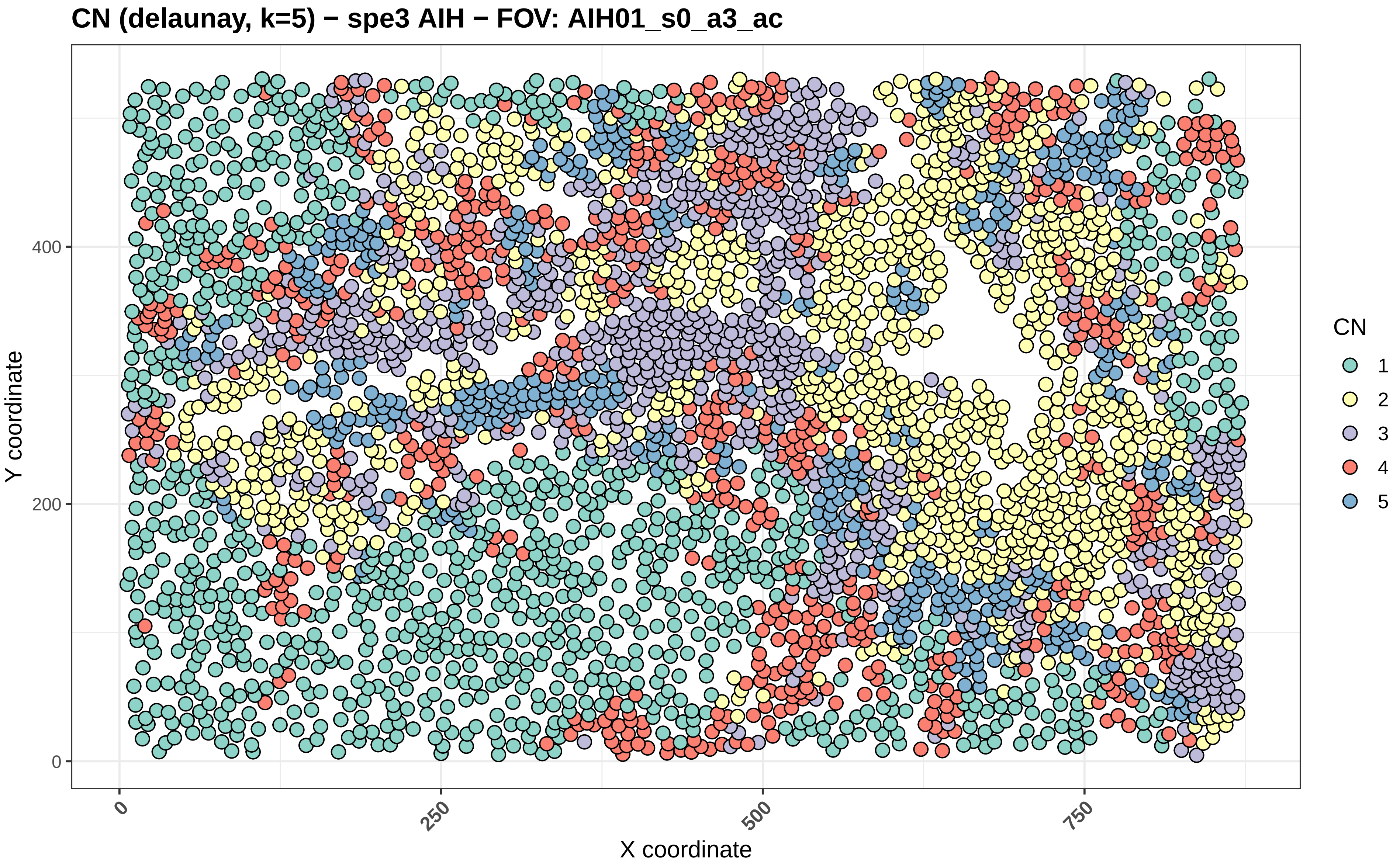
CN (delaunay, k=5) − spe3 AIH − FOV: AIH01_s0_a3_ac
Y coordinate
X coordinate
0
200
400
0
250
500
750
CN
1
2
3
4
5

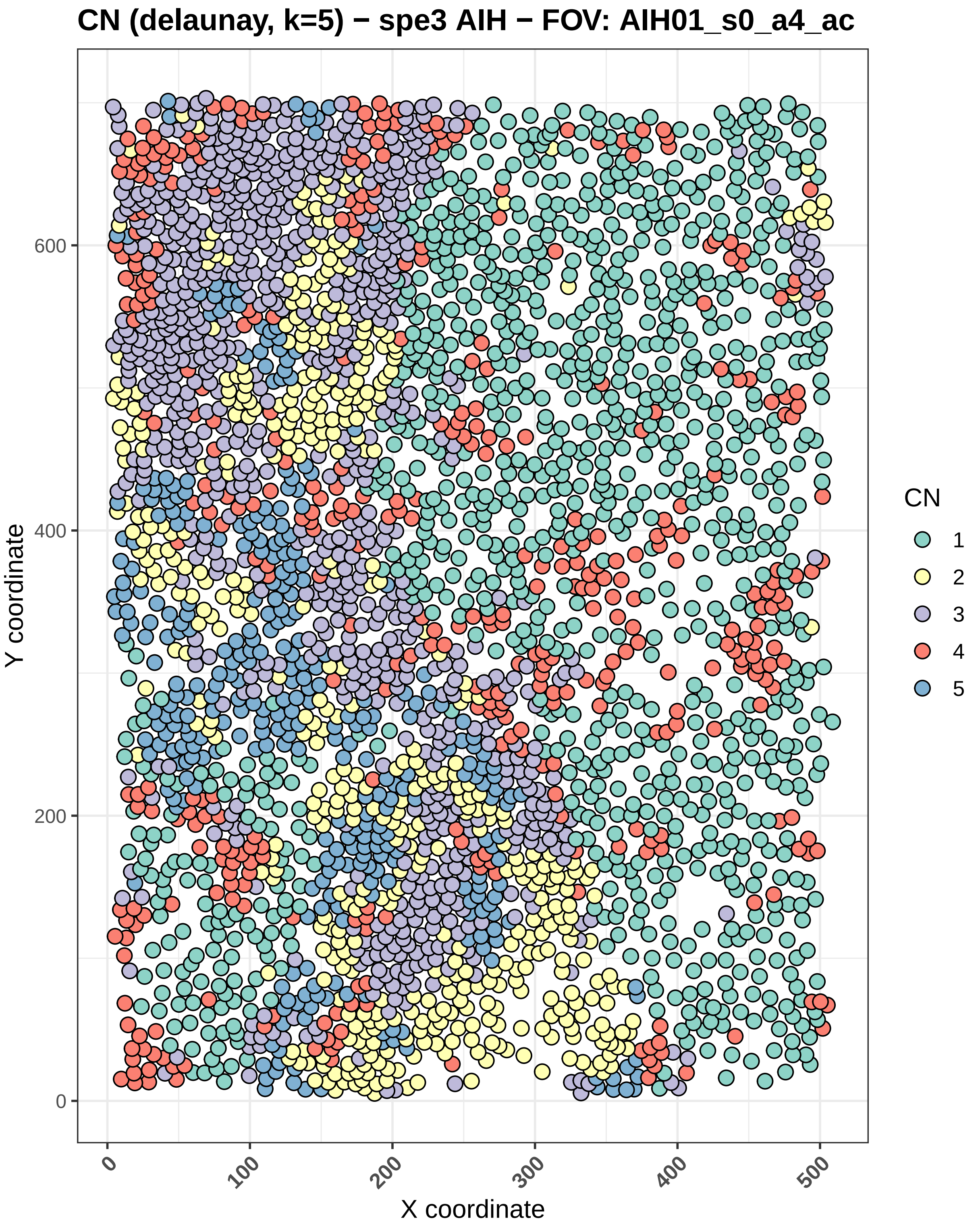

CN (delaunay, k=5) − spe3 AIH − FOV: AIH01_s0_a4_ac
Y coordinate
0
200
400
600
X coordinate
0
100
200
300
400
500
CN
1
2
3
4
5

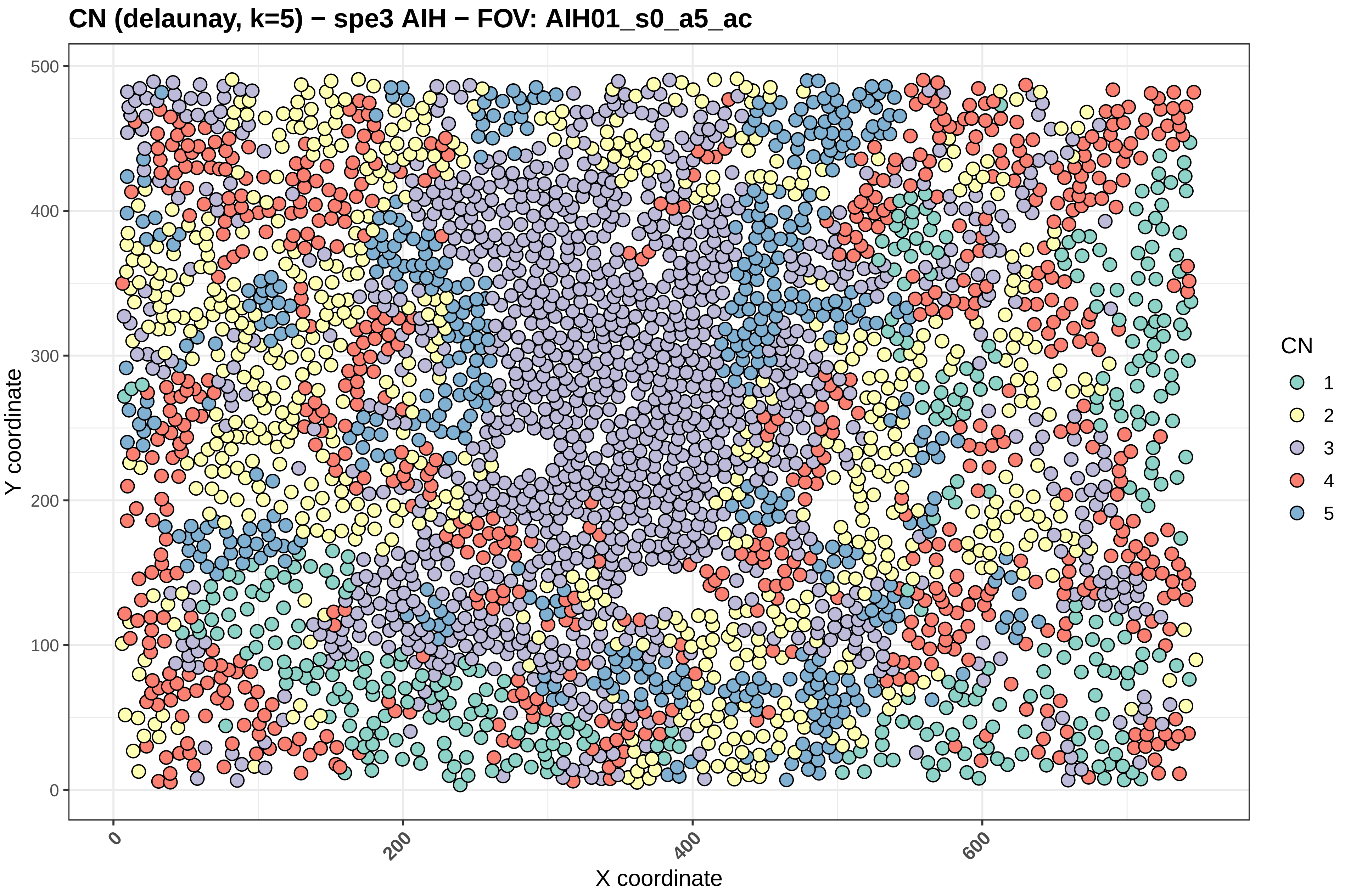
CN (delaunay, k=5) − spe3 AIH − FOV: AIH01_s0_a5_ac
Y coordinate
X coordinate
0
100
200
300
400
500
0
200
400
600
CN
1
2
3
4
5

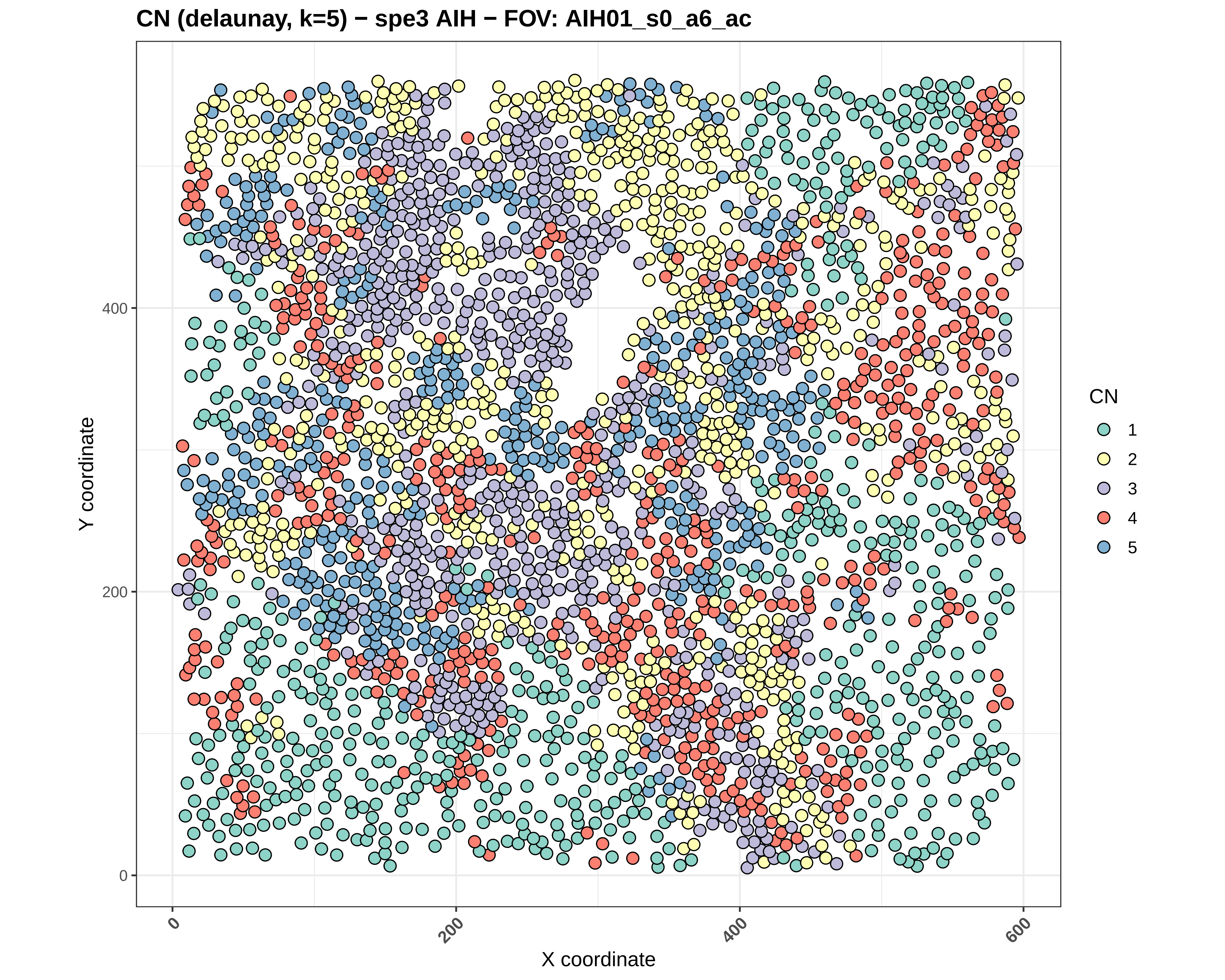
CN (delaunay, k=5) − spe3 AIH − FOV: AIH01_s0_a6_ac
Y coordinate
X coordinate
0
200
400
600
0
200
400
CN
1
2
3
4
5

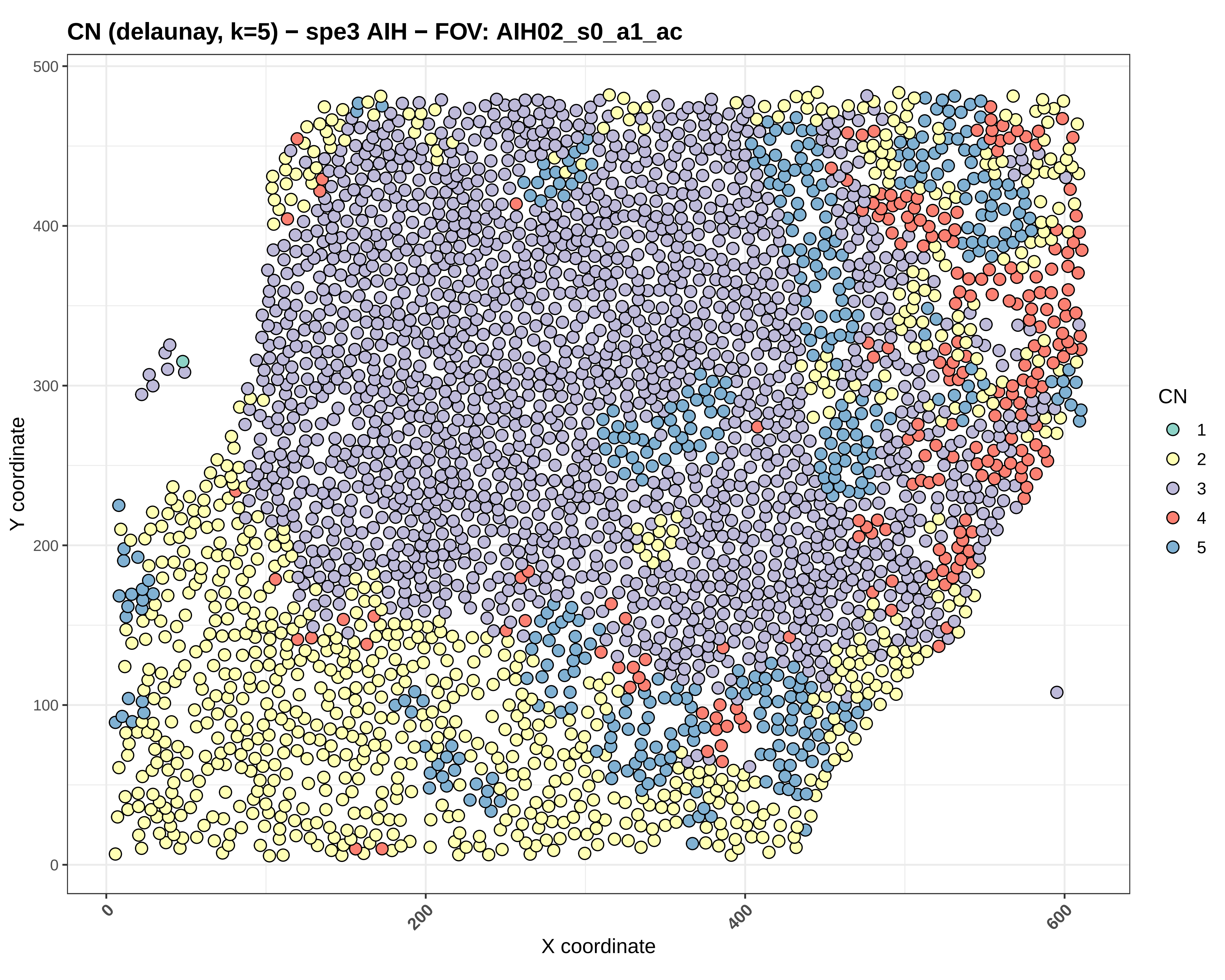

CN (delaunay, k=5) − spe3 AIH − FOV: AIH02_s0_a1_ac
Y coordinate
X coordinate
0
100
200
300
400
500
0
200
400
600
CN
1
2
3
4
5

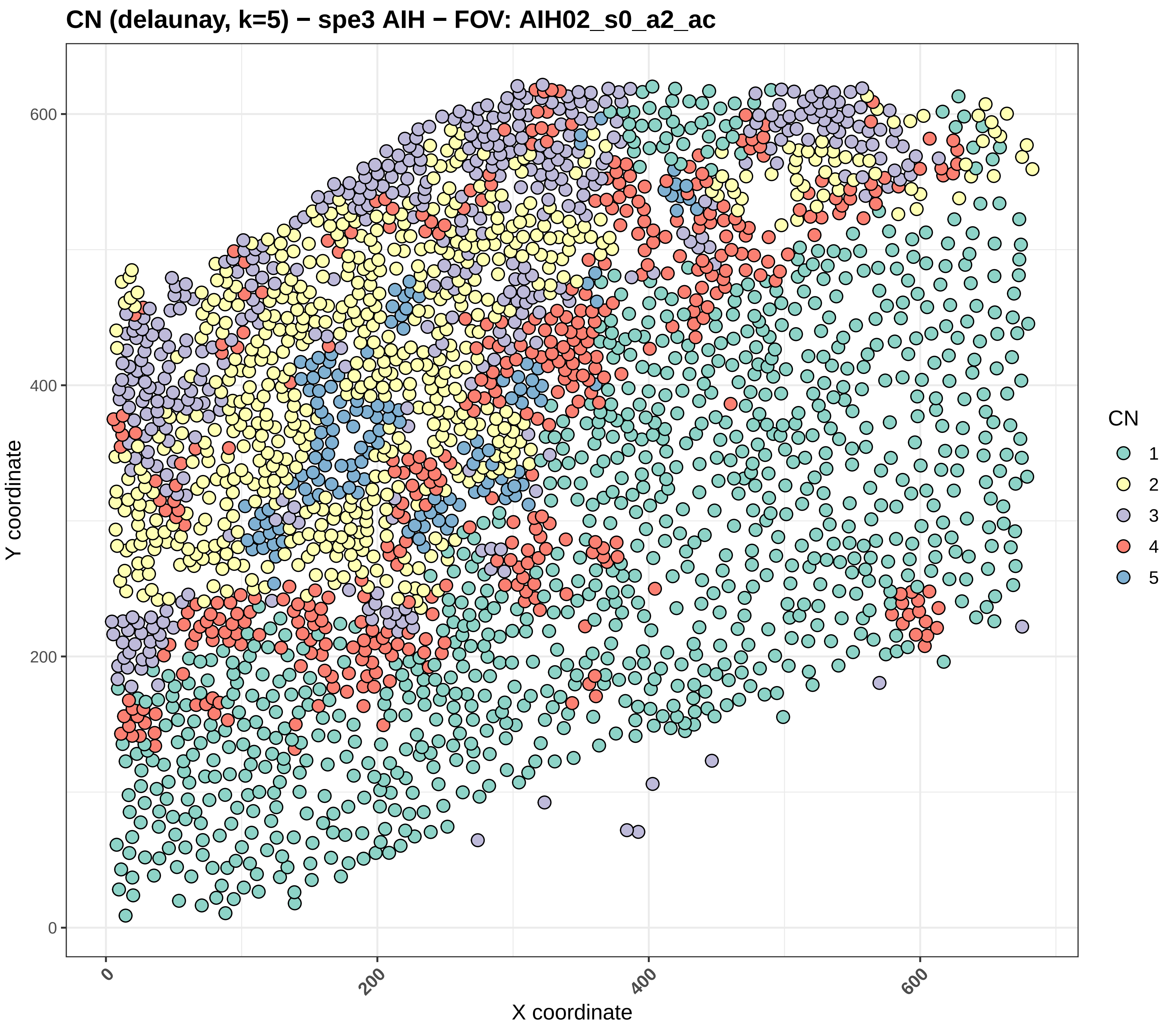
CN (delaunay, k=5) − spe3 AIH − FOV: AIH02_s0_a2_ac
Y coordinate
0
200
400
600
X coordinate
0
200
400
600
CN
1
2
3
4
5

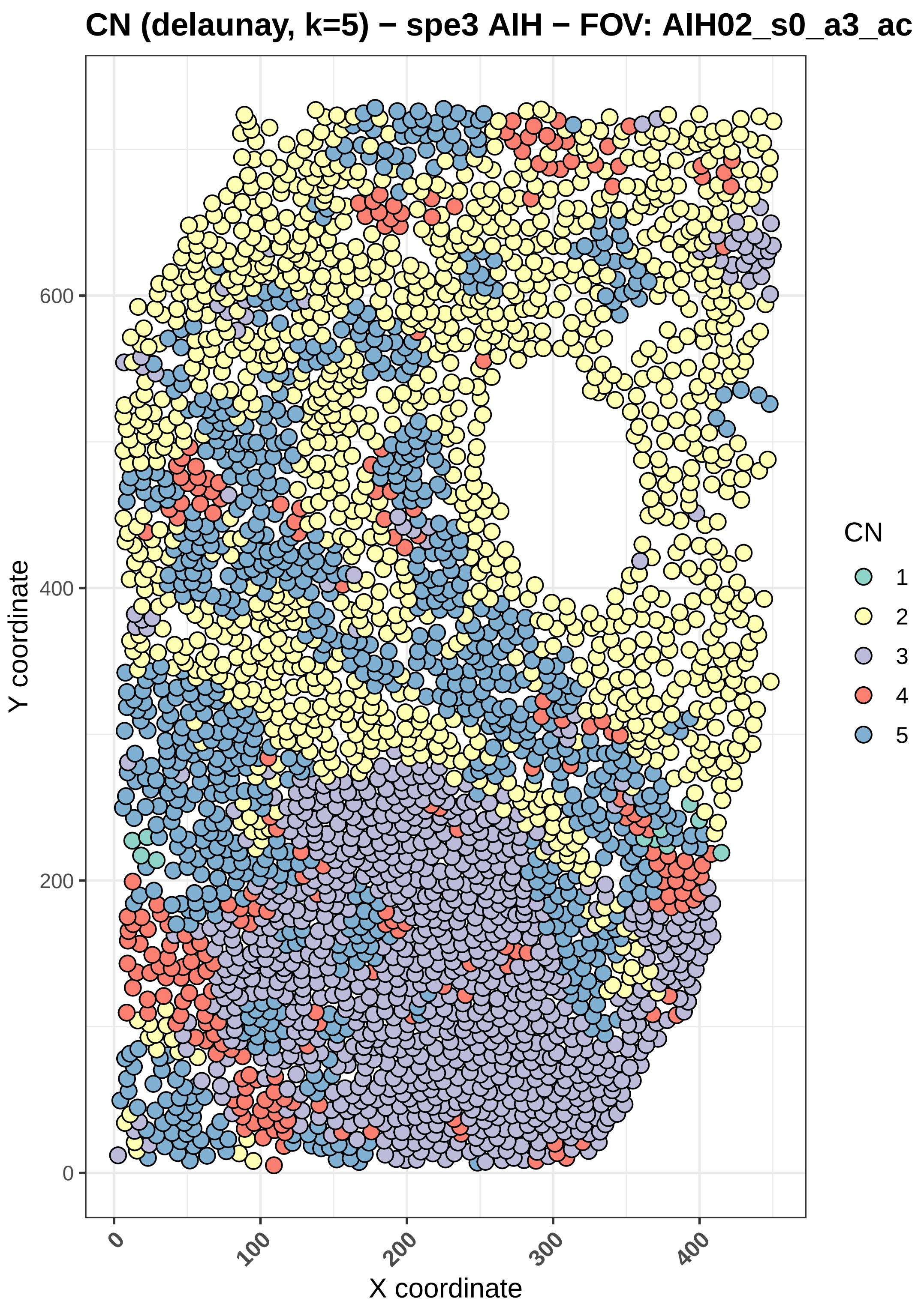
CN (delaunay, k=5) − spe3 AIH − FOV: AIH02_s0_a3_ac
Y coordinate
X coordinate
0
200
400
600
100
300
CN
1
2
3
4
5

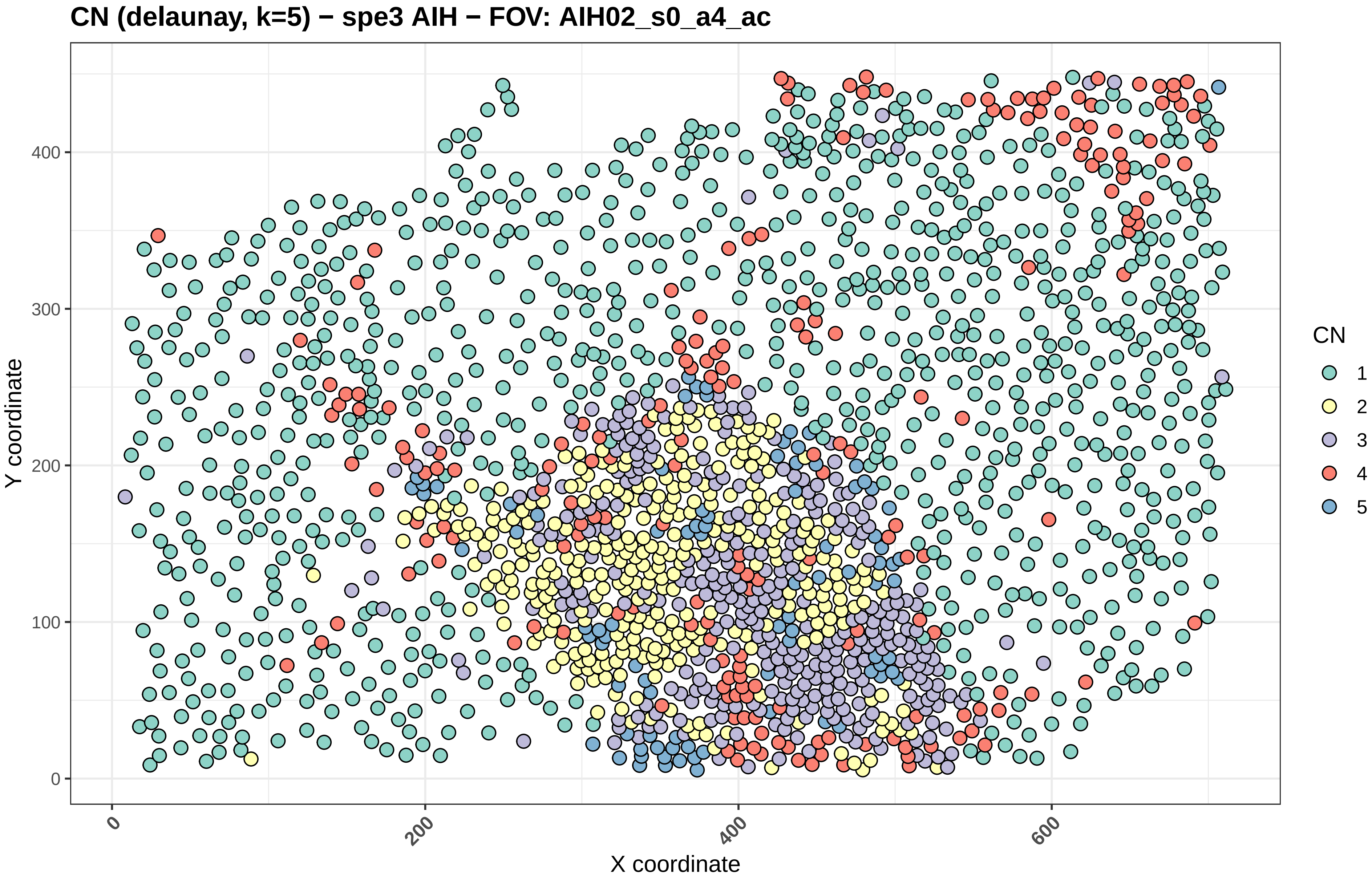

CN (delaunay, k=5) − spe3 AIH − FOV: AIH02_s0_a4_ac
Y coordinate
X coordinate
0
100
200
300
400
0
200
400
600
CN
1
2
3
4
5

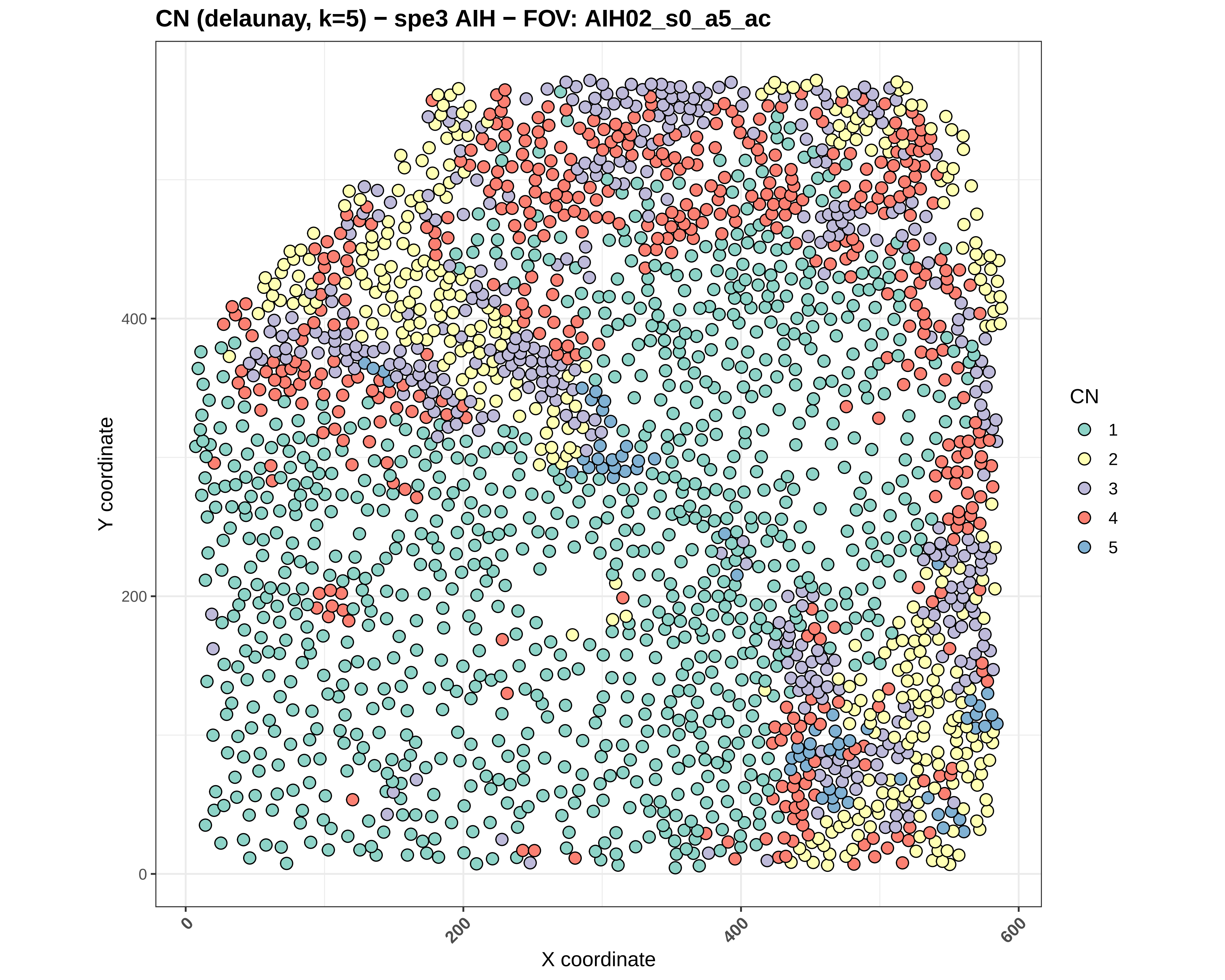

CN (delaunay, k=5) − spe3 AIH − FOV: AIH02_s0_a5_ac
Y coordinate
X coordinate
0
200
400
0
200
400
600
CN
1
2
3
4
5

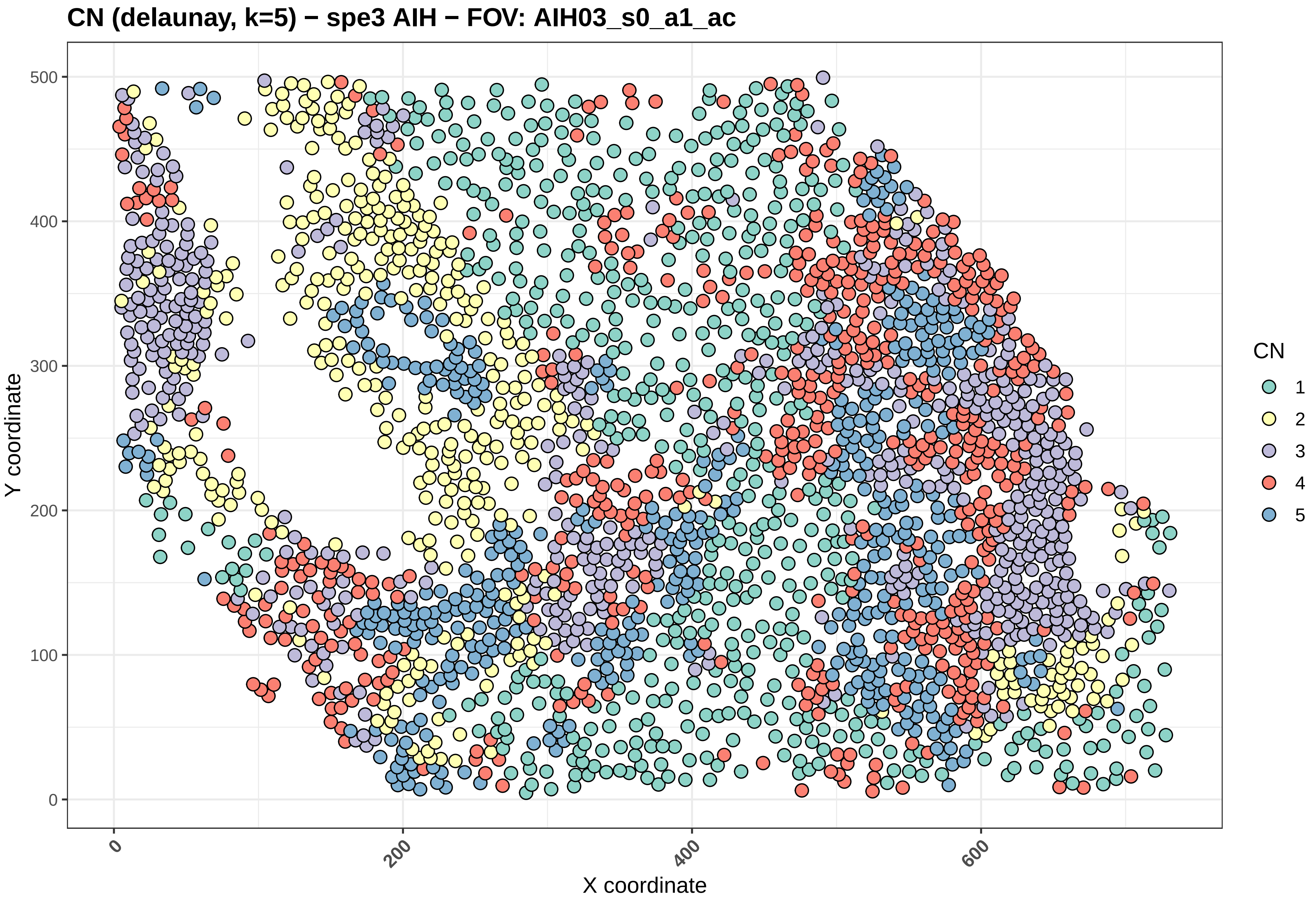
CN (delaunay, k=5) − spe3 AIH − FOV: AIH03_s0_a1_ac
Y coordinate
X coordinate
0
100
200
300
400
500
0
200
400
600
CN
1
2
3
4
5

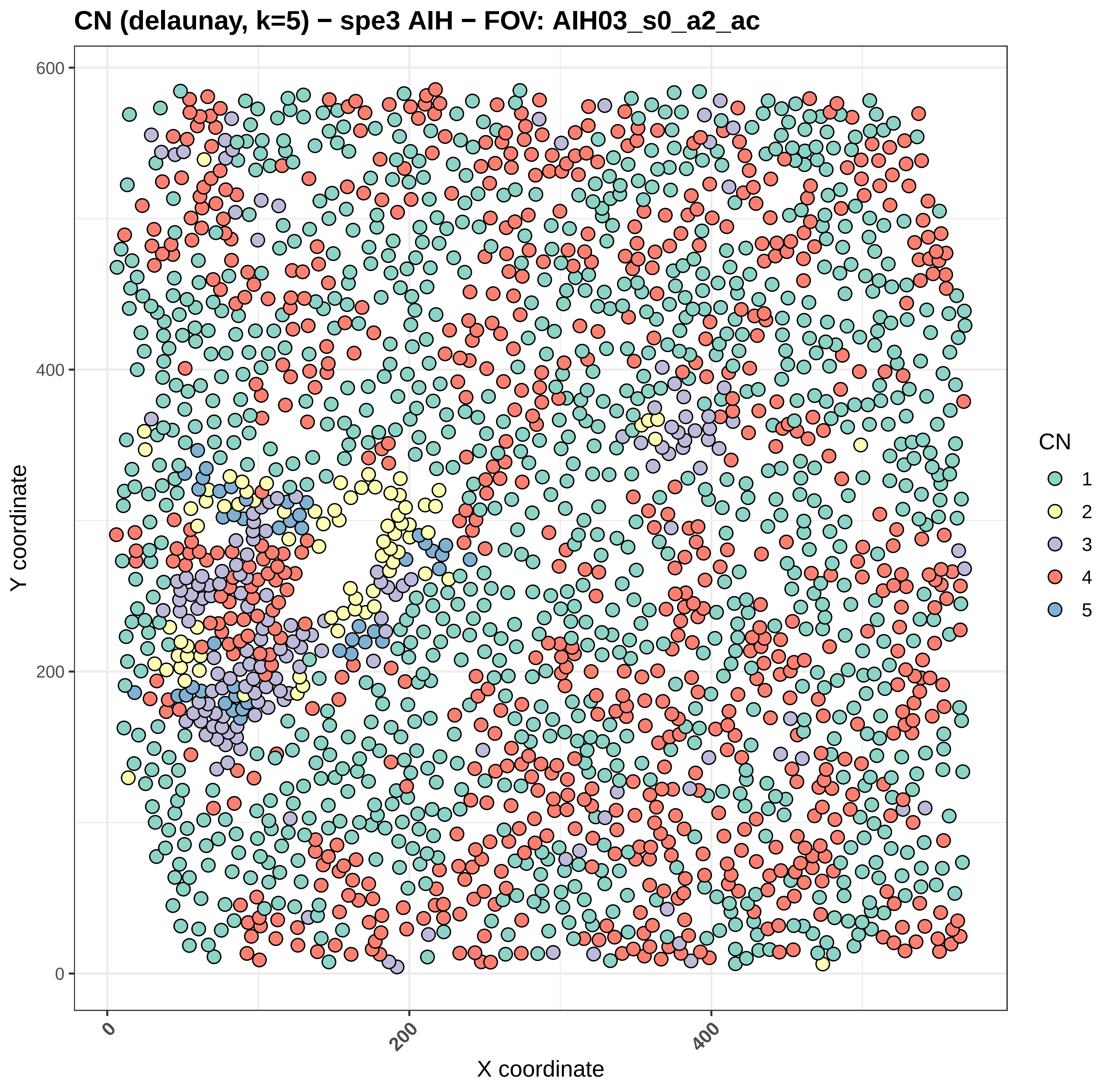
CN (delaunay, k=5) − spe3 AIH − FOV: AIH03_s0_a2_ac
Y coordinate
X coordinate
0
200
400
600
CN
1
2
3
4
5

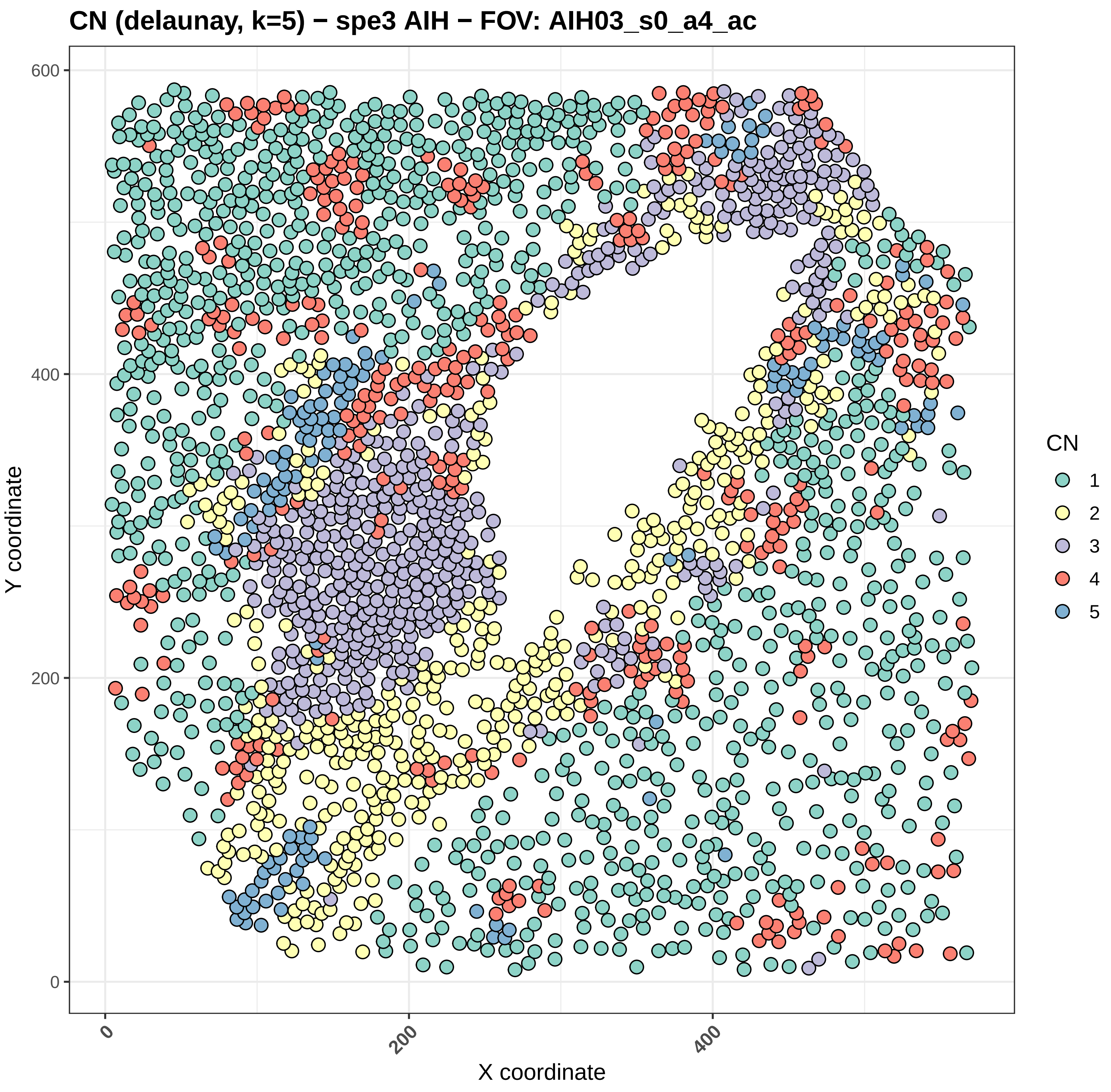
CN (delaunay, k=5) − spe3 AIH − FOV: AIH03_s0_a4_ac
Y coordinate
X coordinate
0
200
400
600
0
200
400
CN
1
2
3
4
5

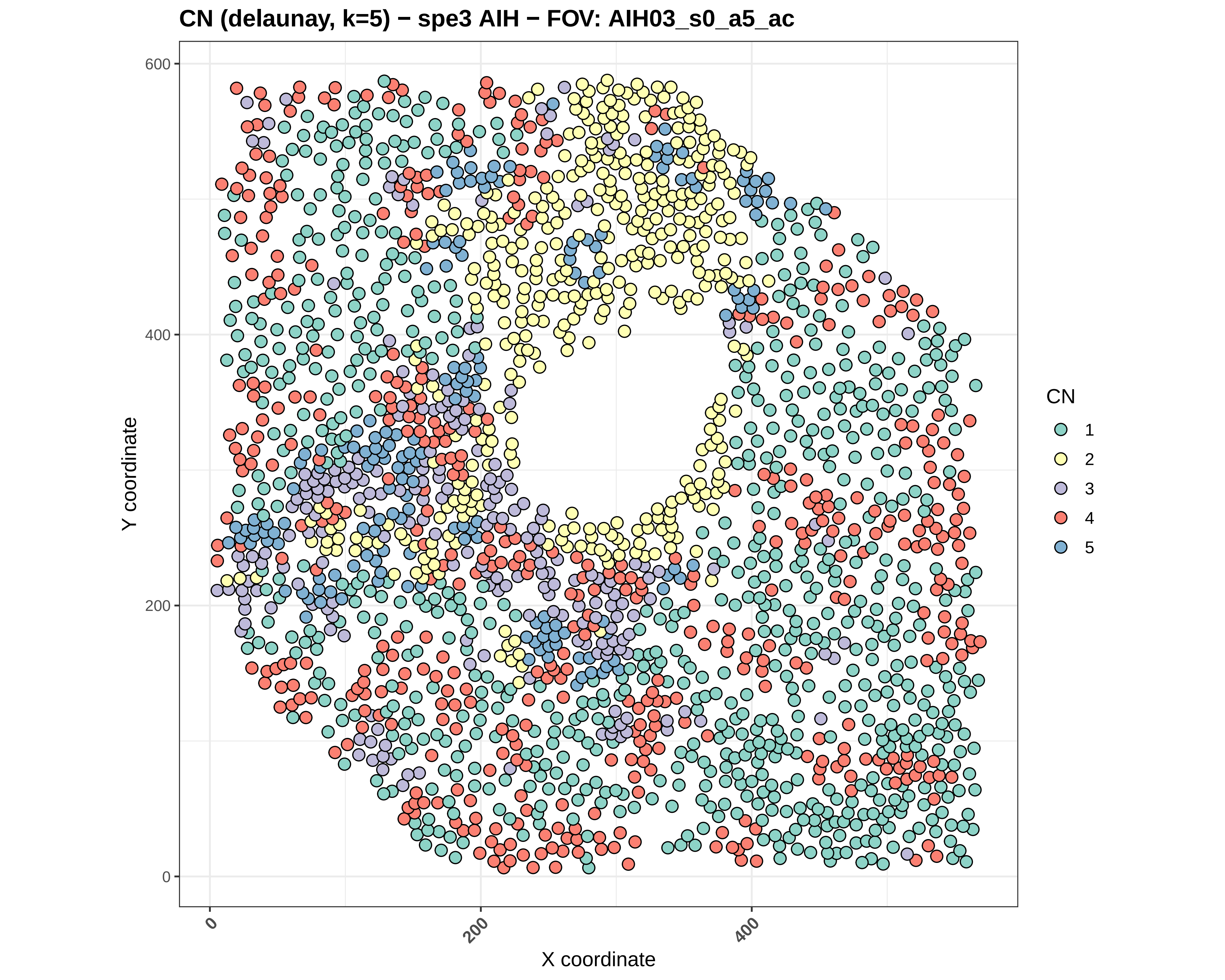

CN (delaunay, k=5) − spe3 AIH − FOV: AIH03_s0_a5_ac
Y coordinate
X coordinate
0
200
400
600
CN
1
2
3
4
5

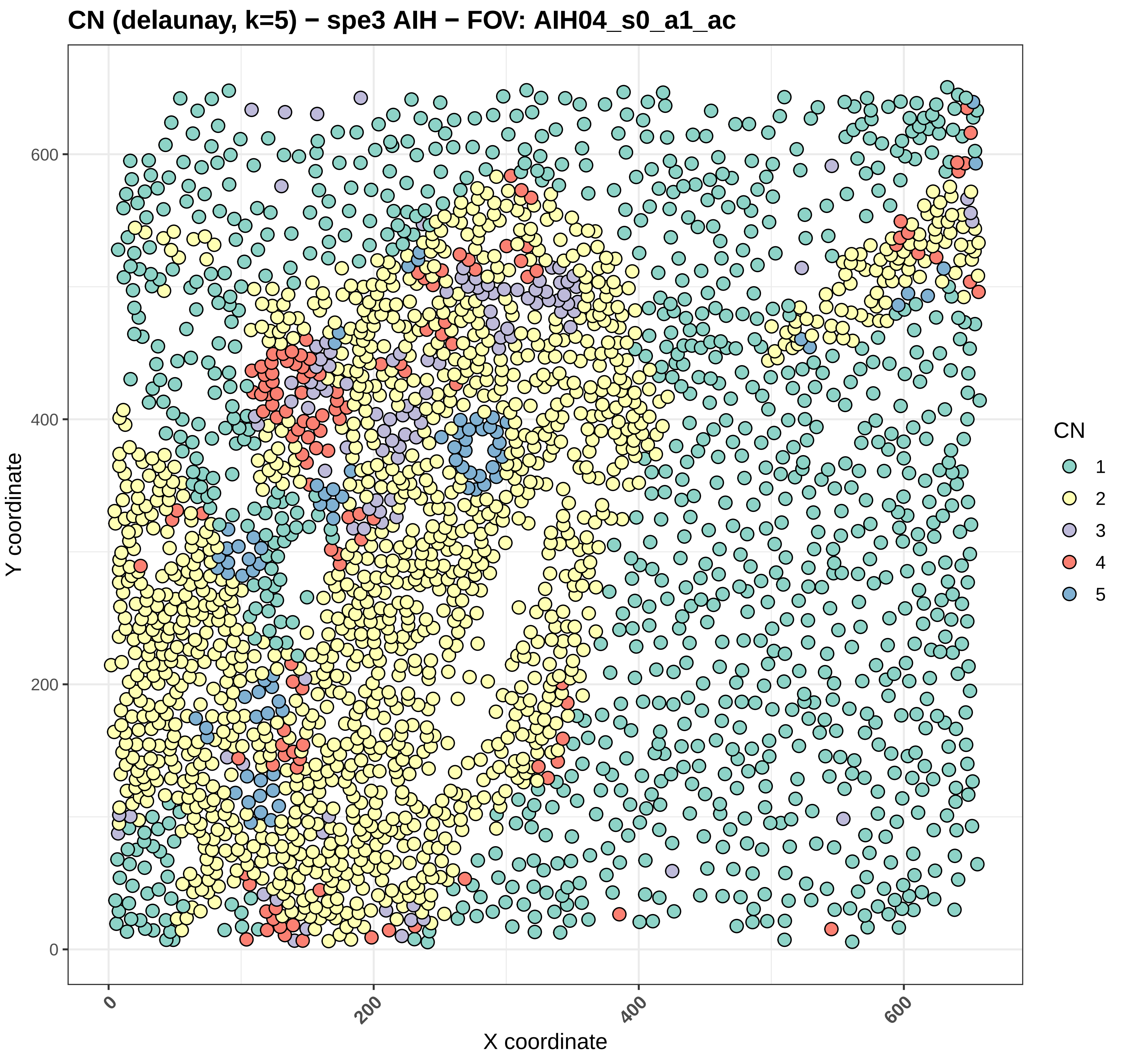
CN (delaunay, k=5) − spe3 AIH − FOV: AIH04_s0_a1_ac
Y coordinate
X coordinate
0
200
400
600
CN
1
2
3
4
5

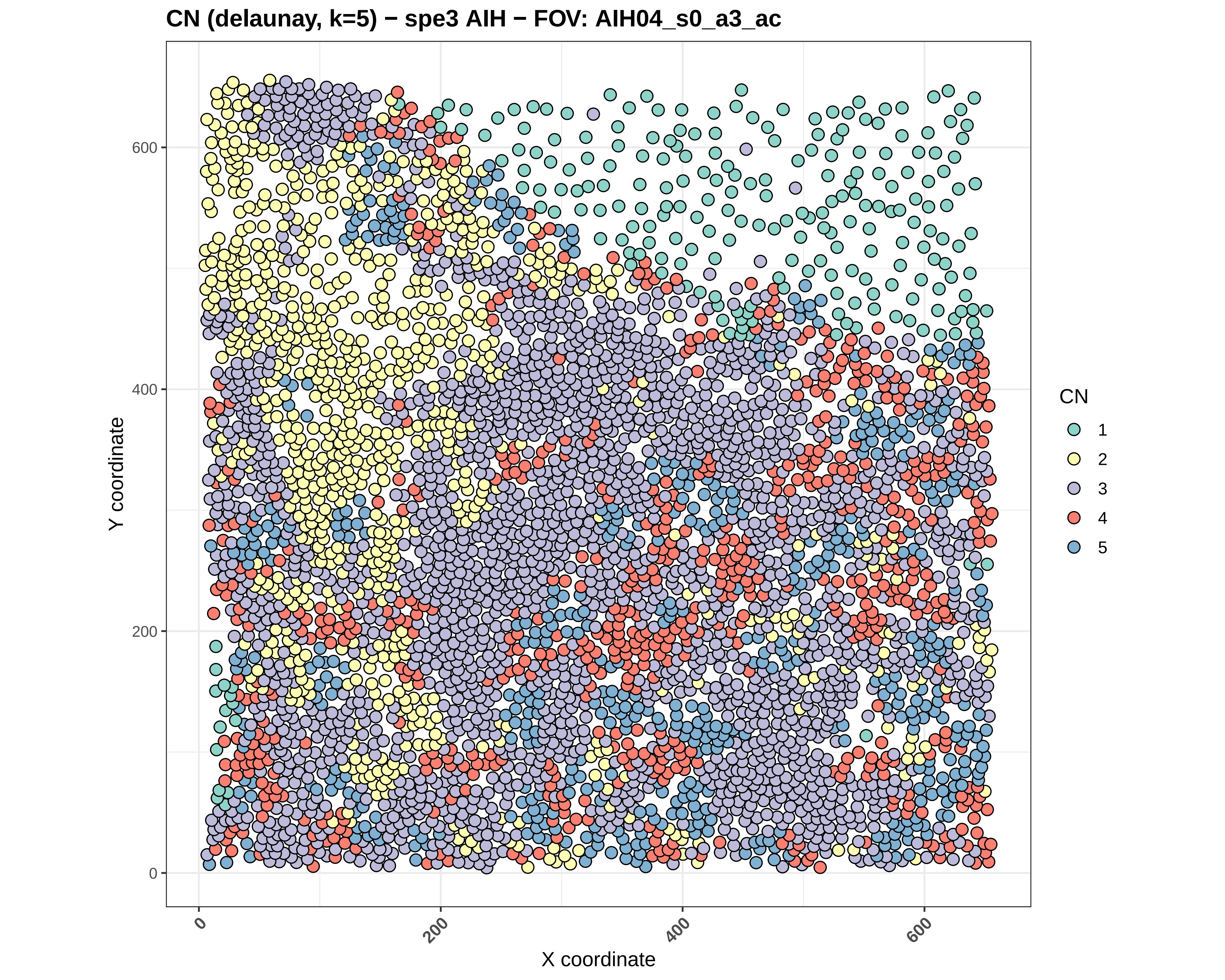
CN (delaunay, k=5) − spe3 AIH − FOV: AIH04_s0_a3_ac
Y coordinate
X coordinate
0
200
400
600
CN
1
2
3
4
5

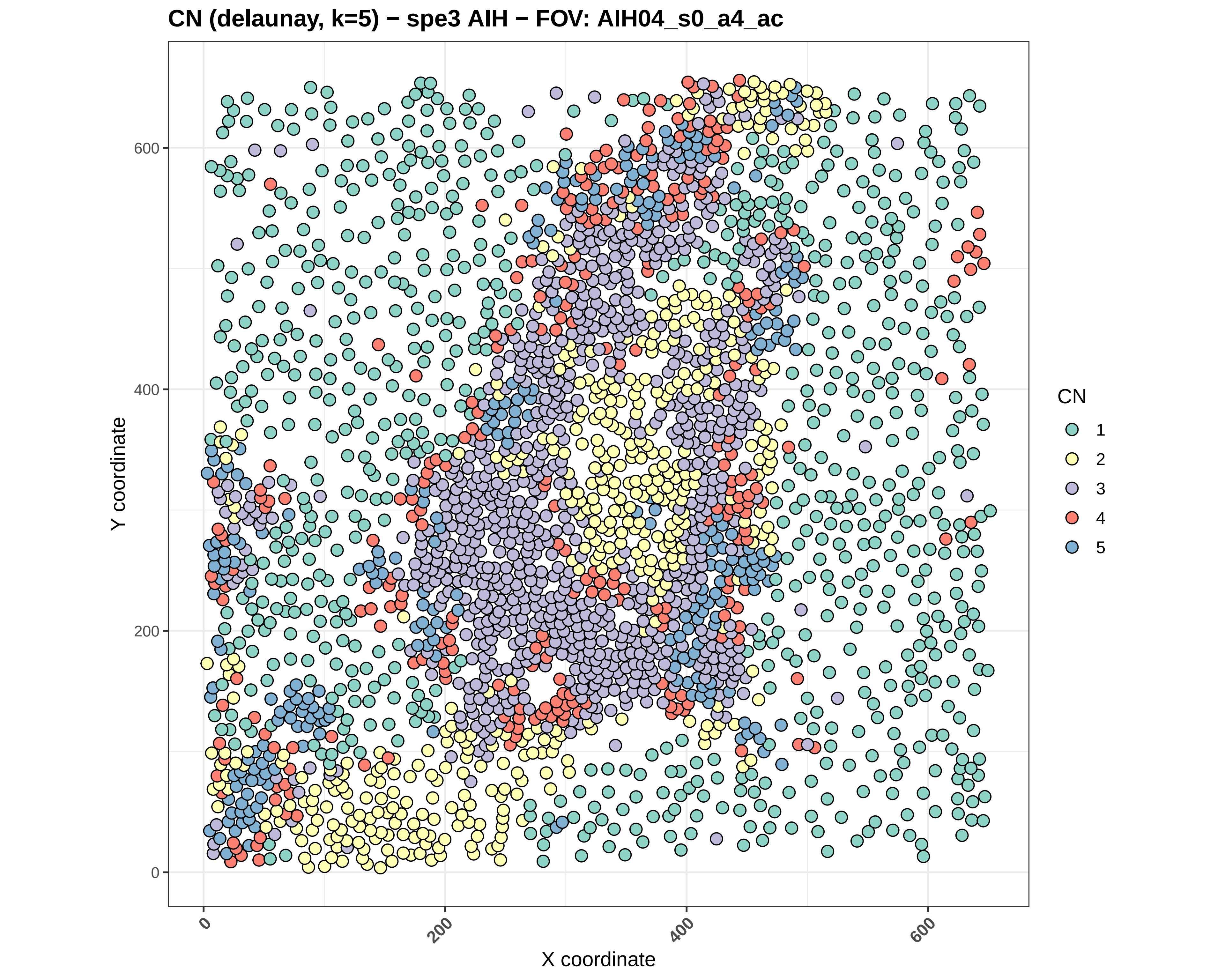
CN (delaunay, k=5) − spe3 AIH − FOV: AIH04_s0_a4_ac
Y coordinate
X coordinate
0
200
400
600
CN
1
2
3
4
5

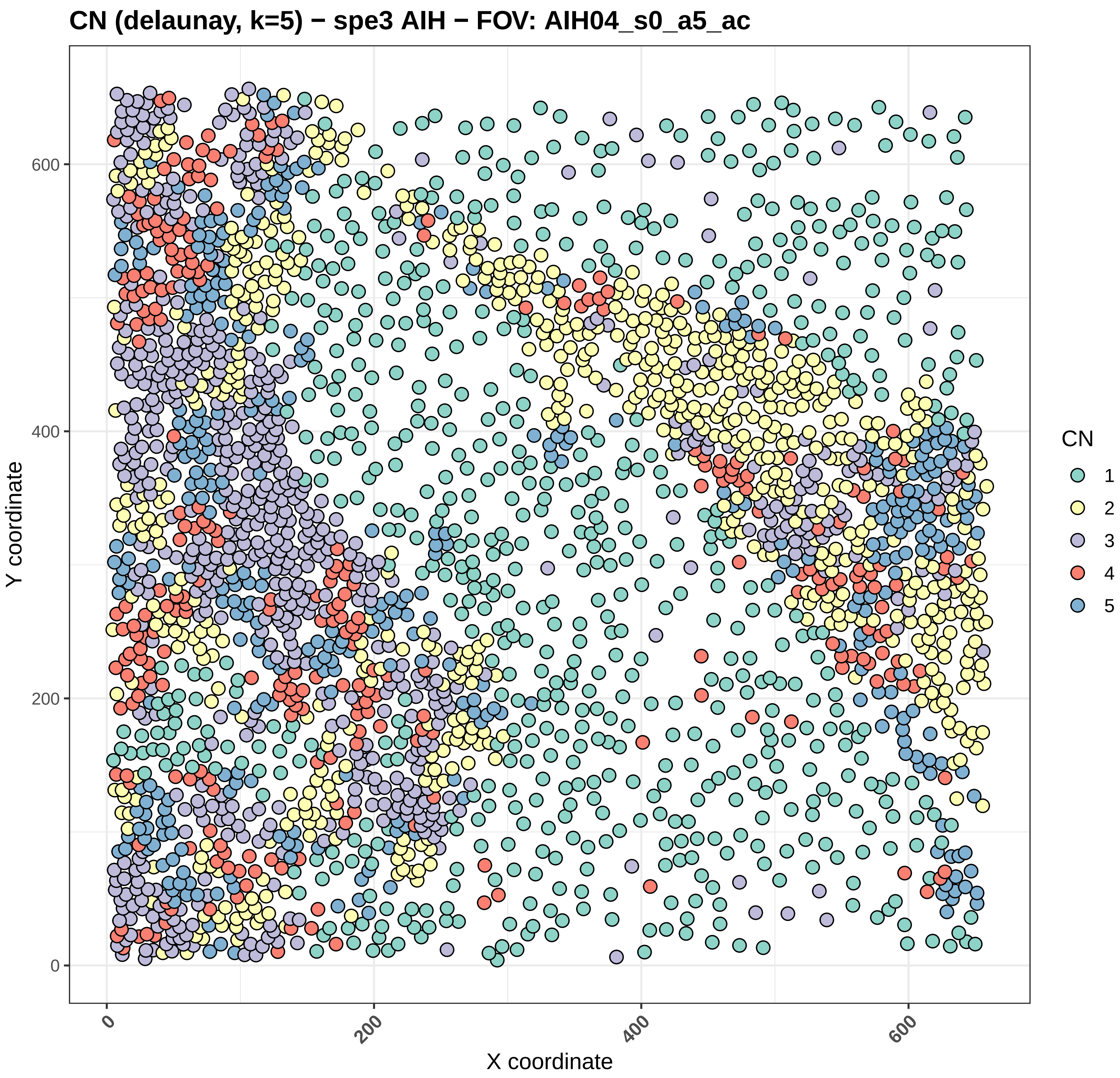

CN (delaunay, k=5) − spe3 AIH − FOV: AIH04_s0_a5_ac
Y coordinate
X coordinate
0
200
400
600
CN
1
2
3
4
5

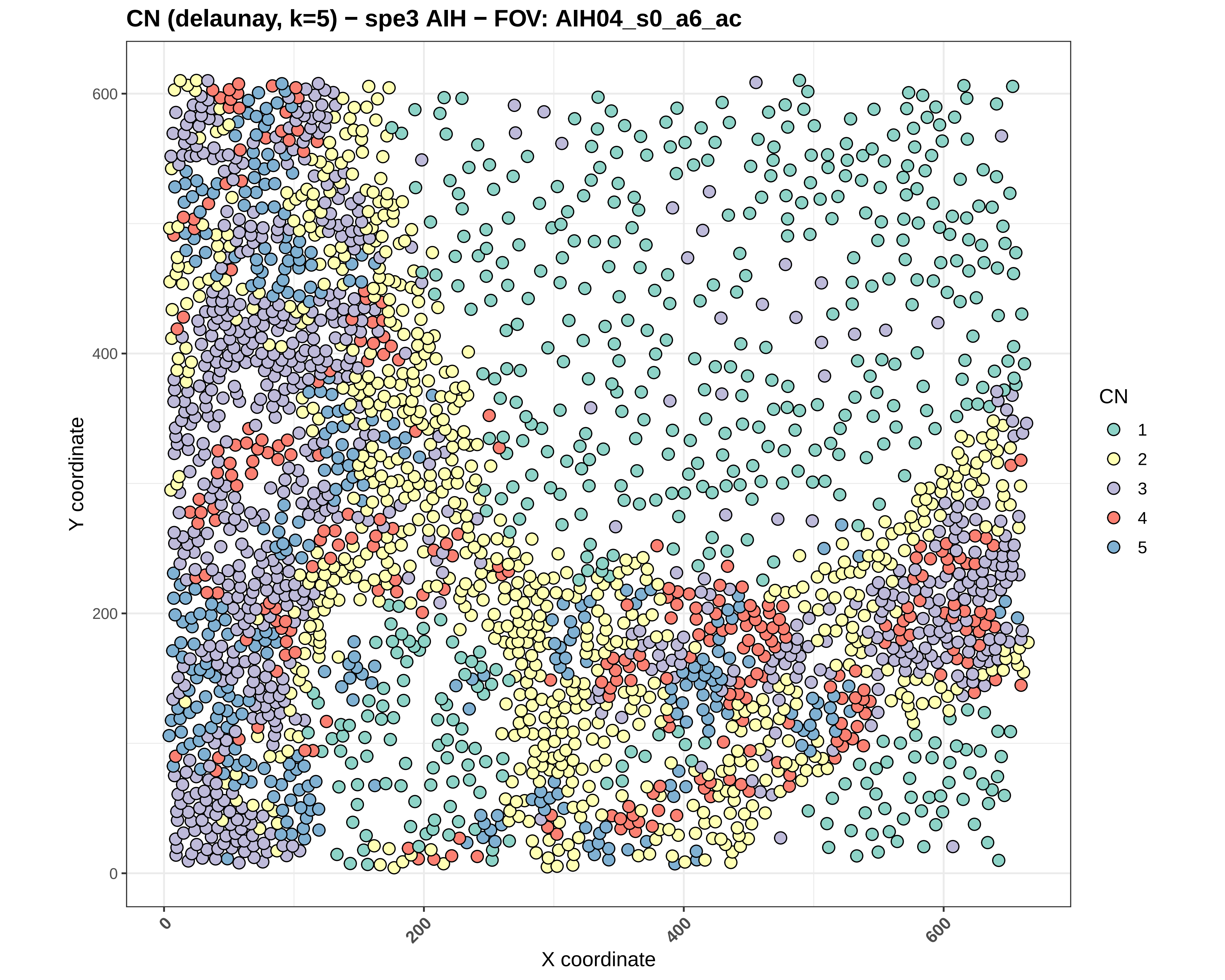
CN (delaunay, k=5) − spe3 AIH − FOV: AIH04_s0_a6_ac
Y coordinate
X coordinate
0
200
400
600
CN
1
2
3
4
5

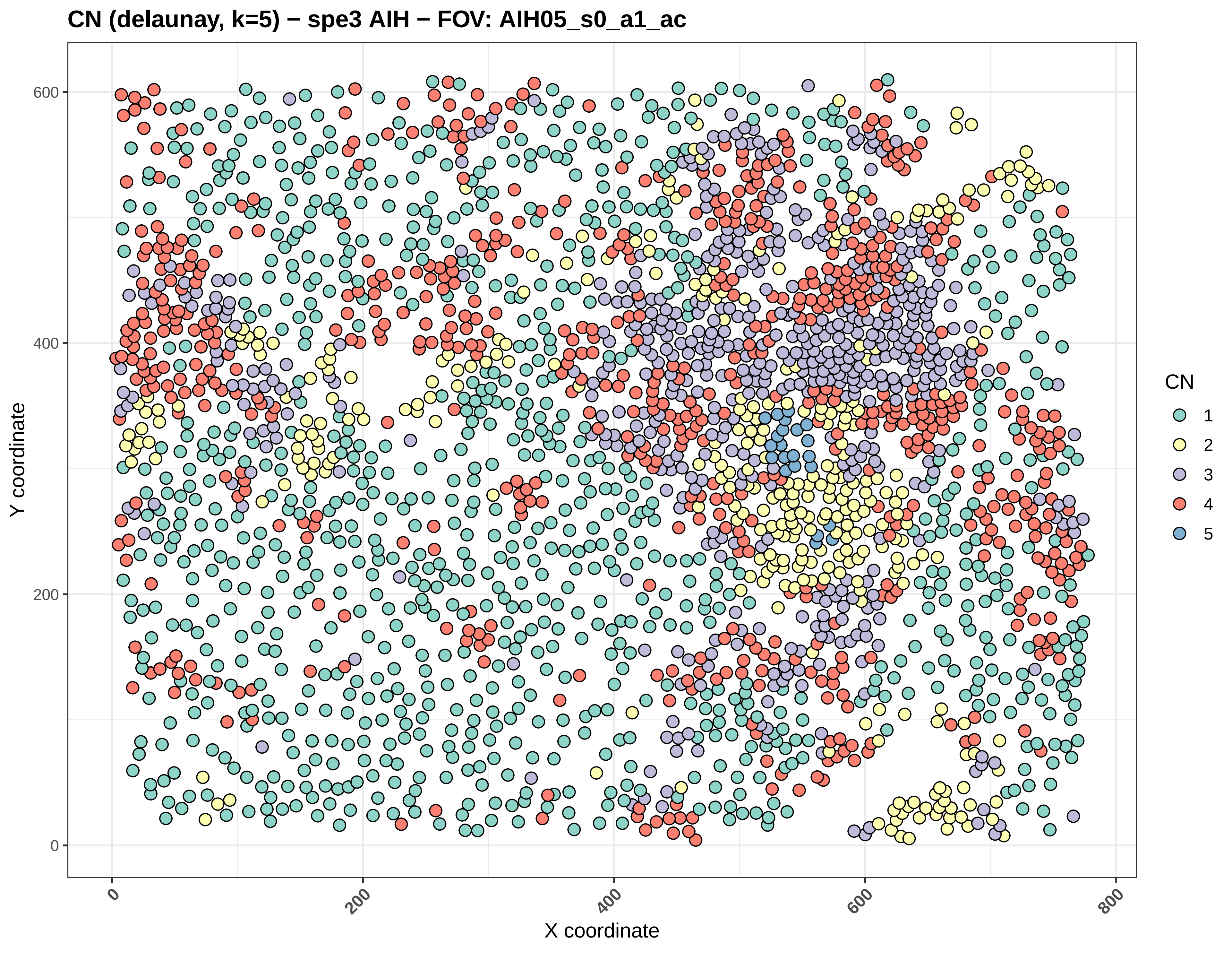

CN (delaunay, k=5) − spe3 AIH − FOV: AIH05_s0_a1_ac
Y coordinate
X coordinate
0
200
400
600
800
CN
1
2
3
4
5

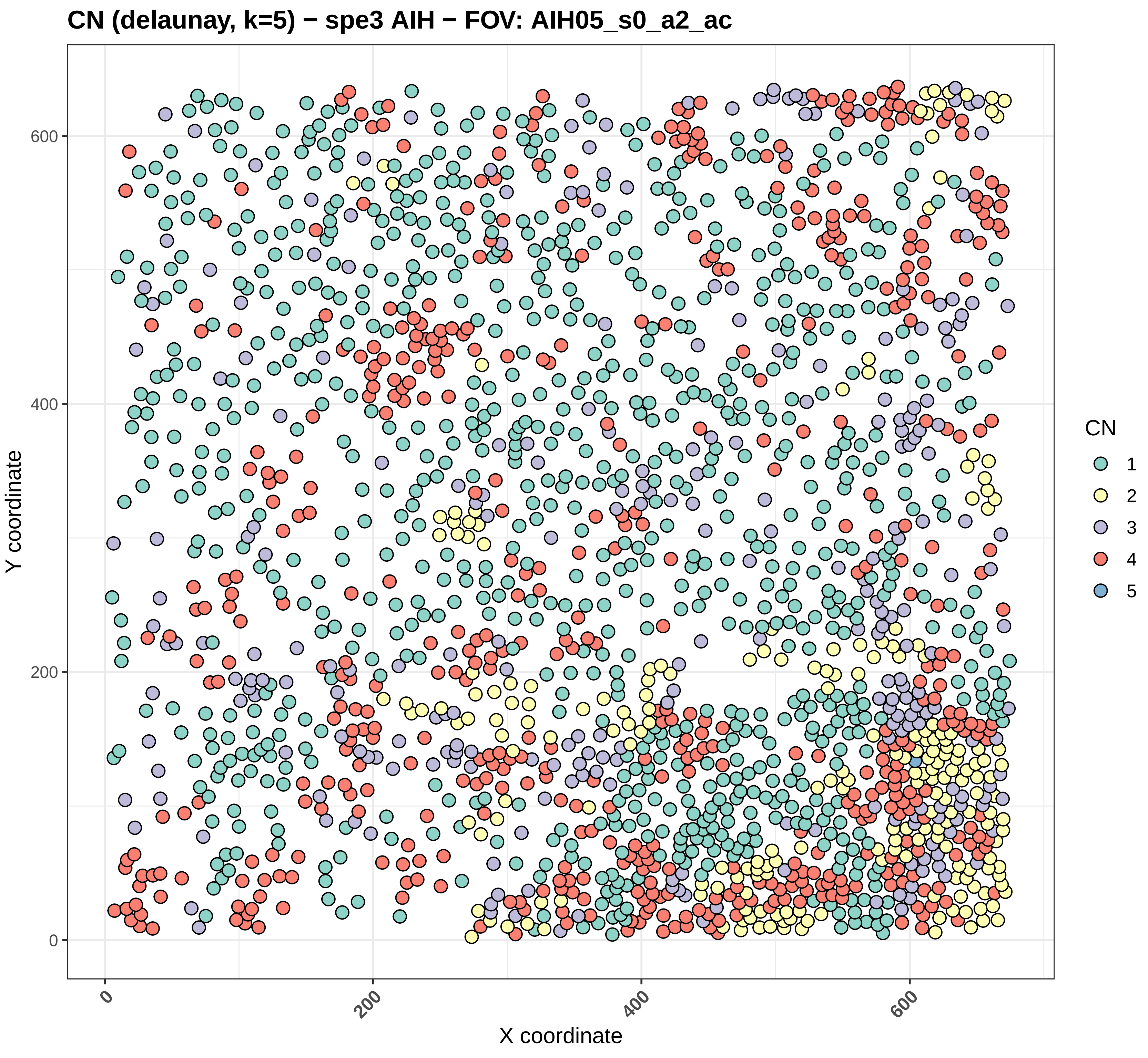

CN (delaunay, k=5) − spe3 AIH − FOV: AIH05_s0_a2_ac
Y coordinate
X coordinate
0
200
400
600
CN
1
2
3
4
5

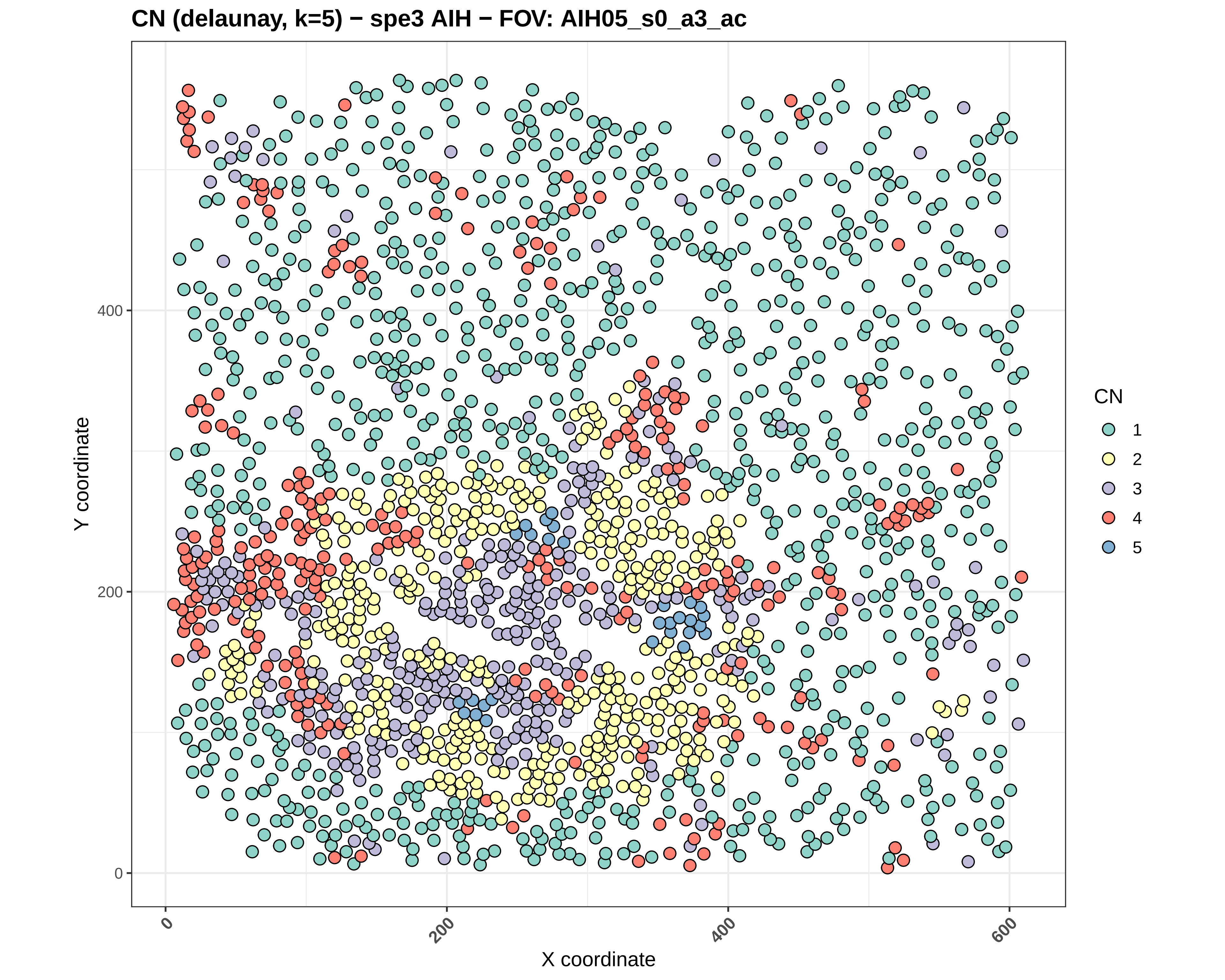

CN (delaunay, k=5) − spe3 AIH − FOV: AIH05_s0_a3_ac
Y coordinate
X coordinate
0
200
400
600
CN
1
2
3
4
5

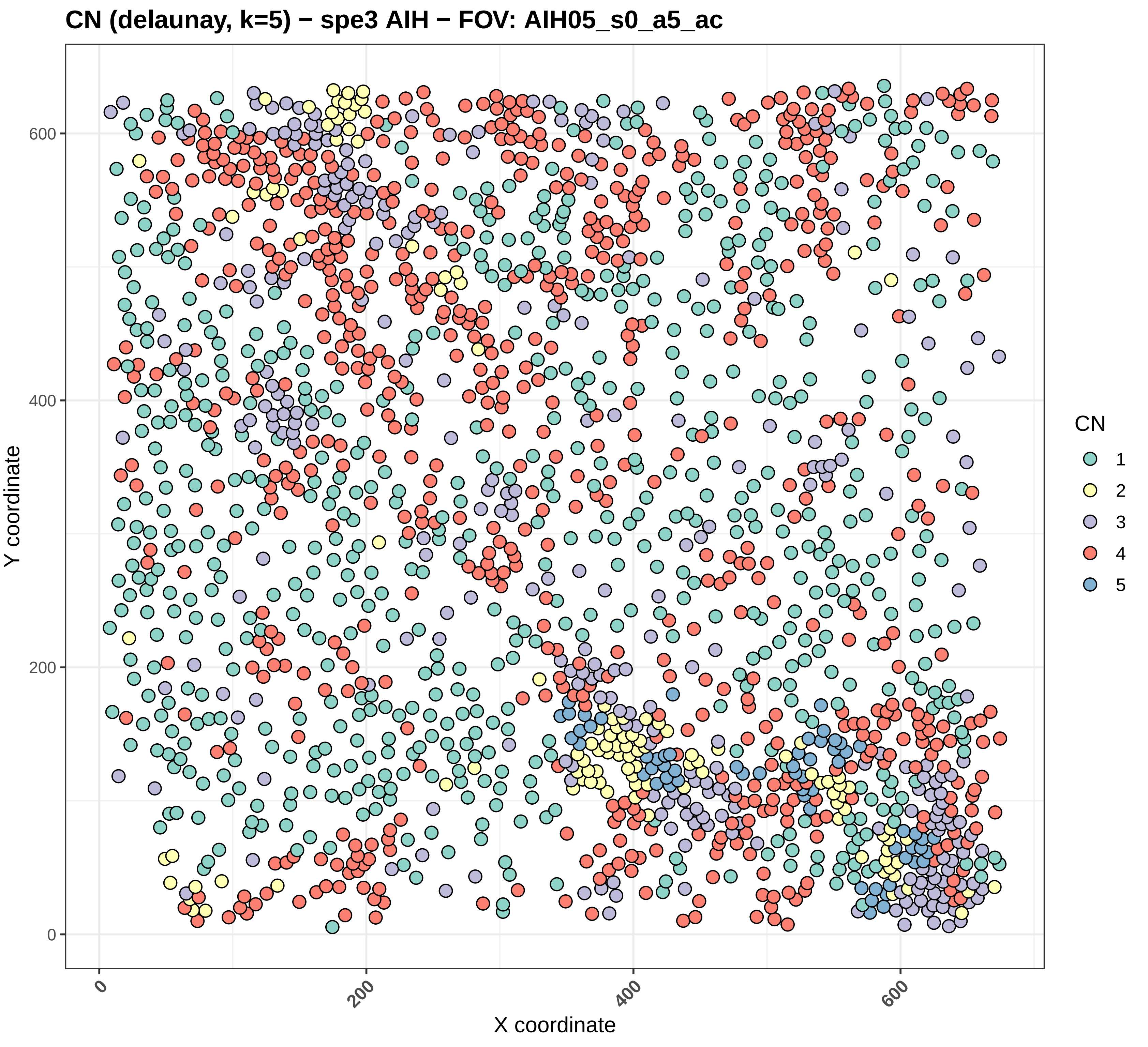
CN (delaunay, k=5) − spe3 AIH − FOV: AIH05_s0_a5_ac
Y coordinate
X coordinate
0
200
400
600
CN
1
2
3
4
5

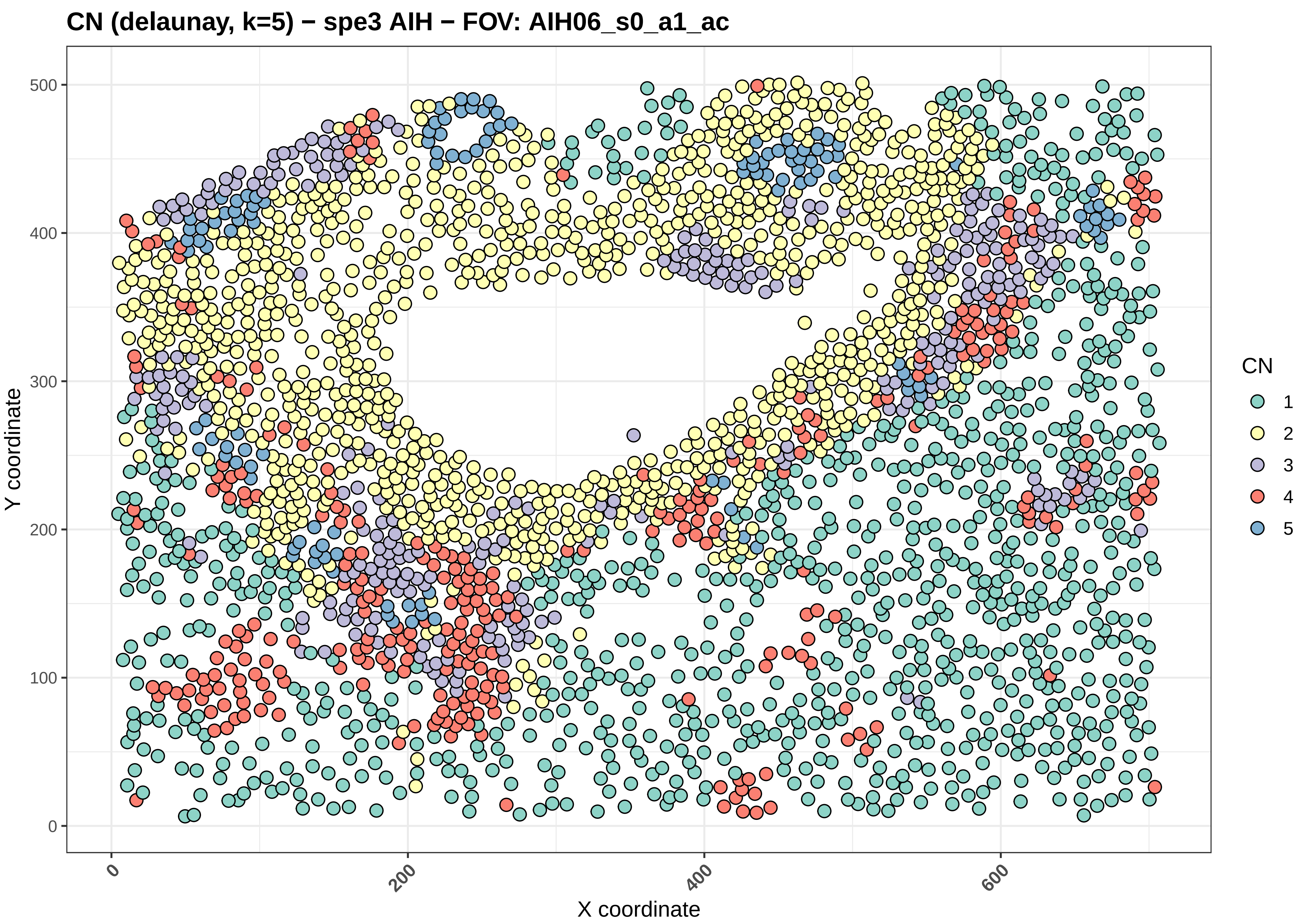

CN (delaunay, k=5) − spe3 AIH − FOV: AIH06_s0_a1_ac
Y coordinate
X coordinate
0
100
200
300
400
500
0
200
400
600
CN
1
2
3
4
5

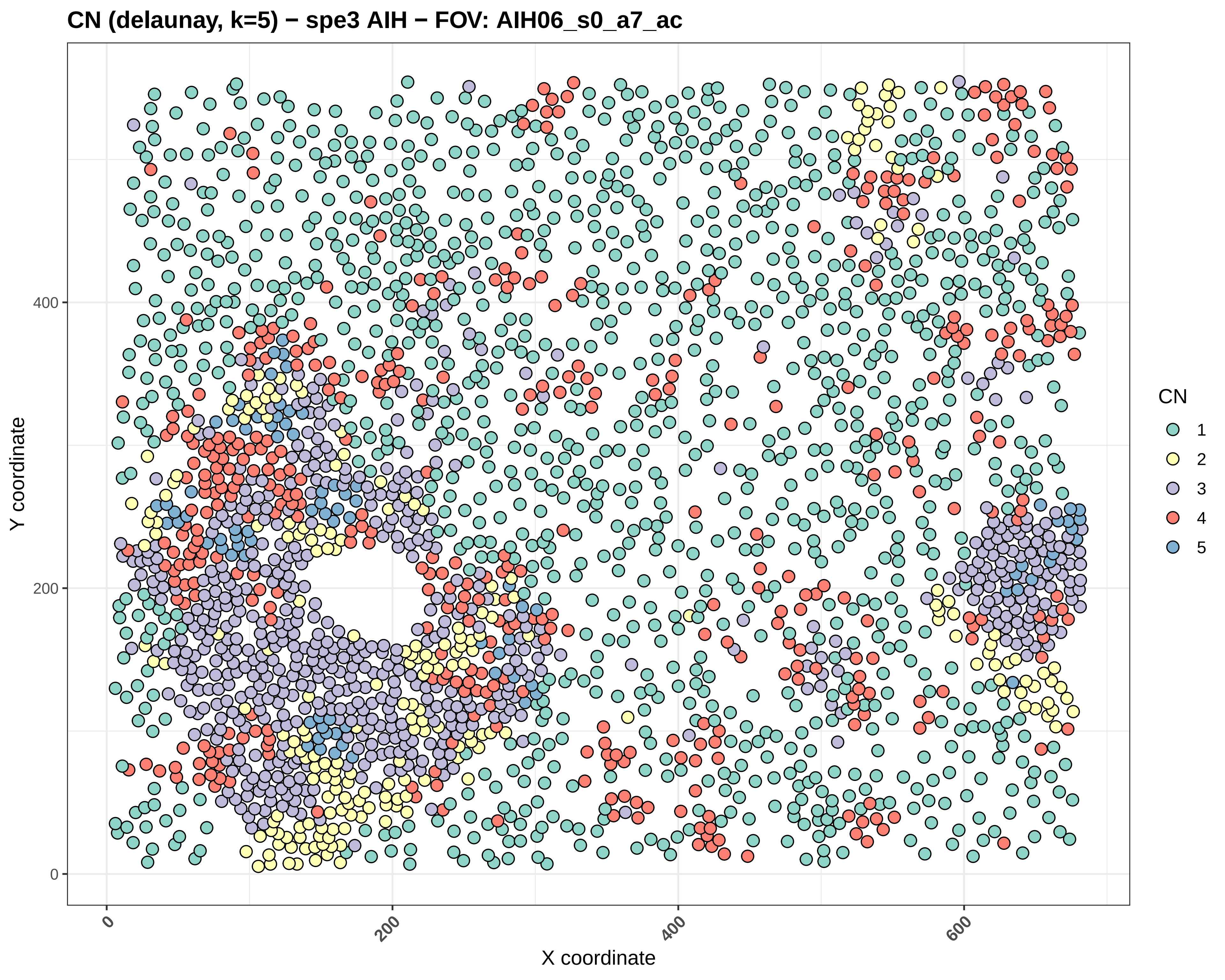
CN (delaunay, k=5) − spe3 AIH − FOV: AIH06_s0_a7_ac
Y coordinate
X coordinate
0
200
400
0
200
400
600
CN
1
2
3
4
5

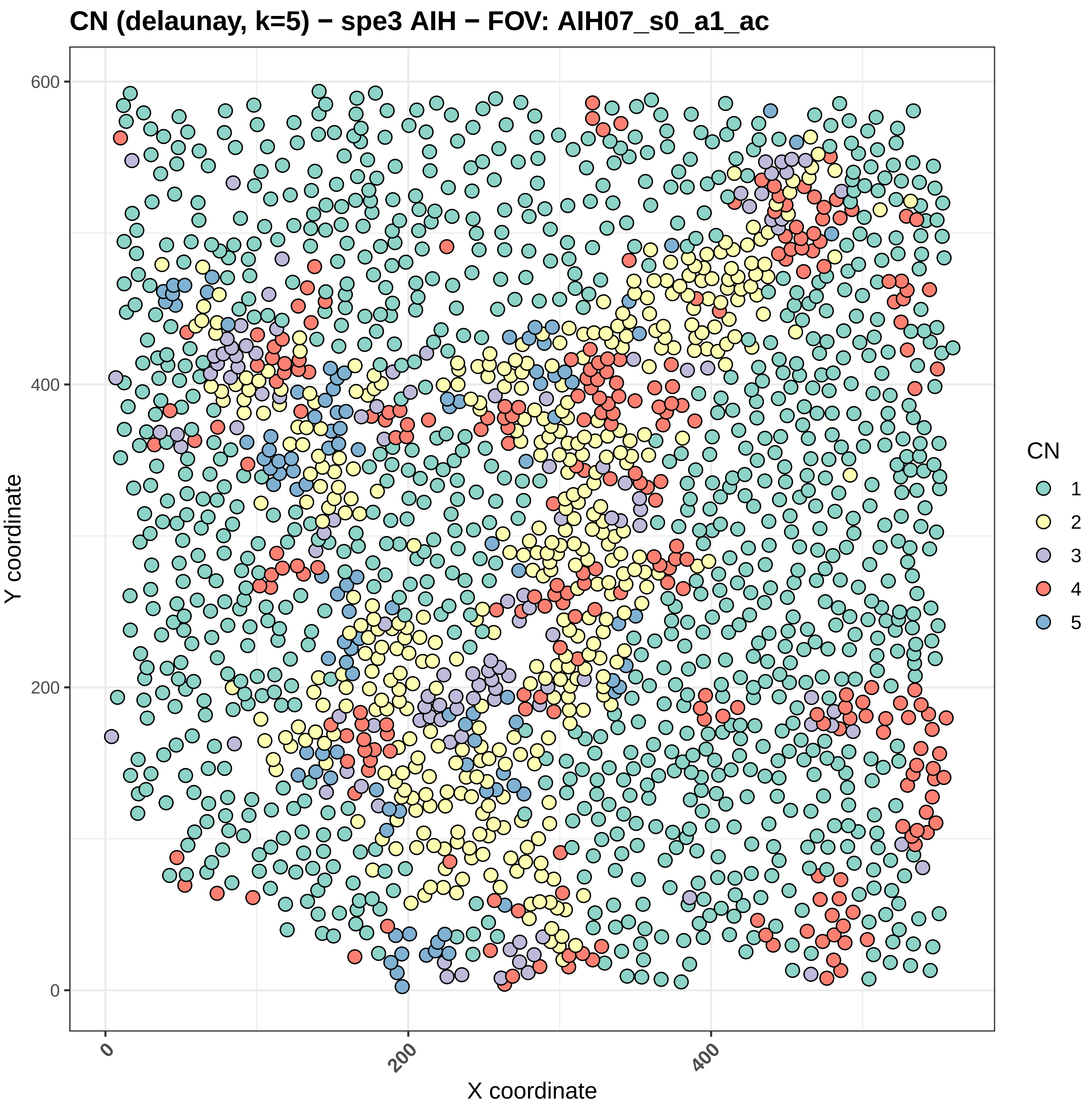
CN (delaunay, k=5) − spe3 AIH − FOV: AIH07_s0_a1_ac
Y coordinate
X coordinate
0
200
400
600
0
200
400
CN
1
2
3
4
5

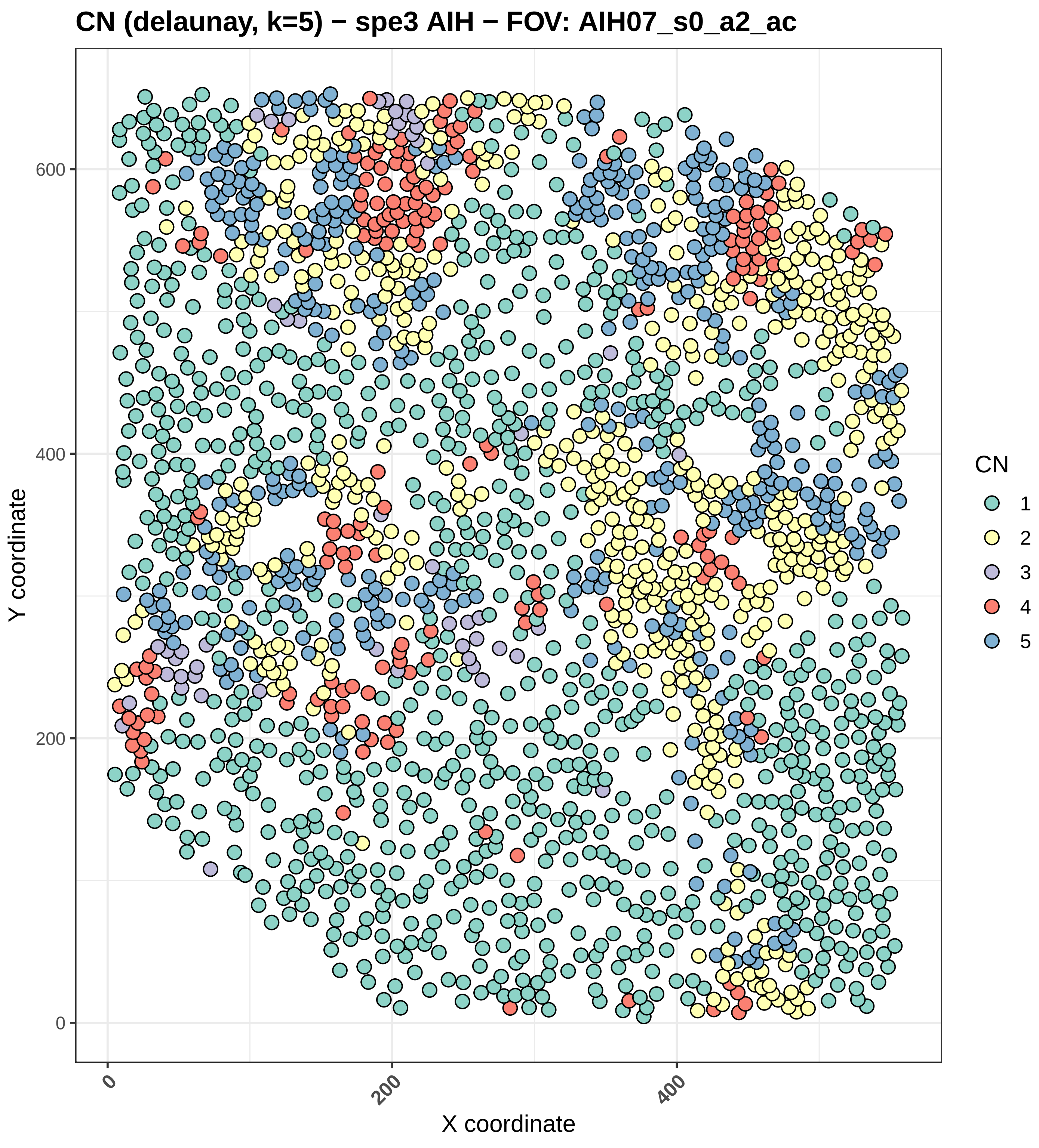
CN (delaunay, k=5) − spe3 AIH − FOV: AIH07_s0_a2_ac
Y coordinate
X coordinate
0
200
400
600
CN
1
2
3
4
5

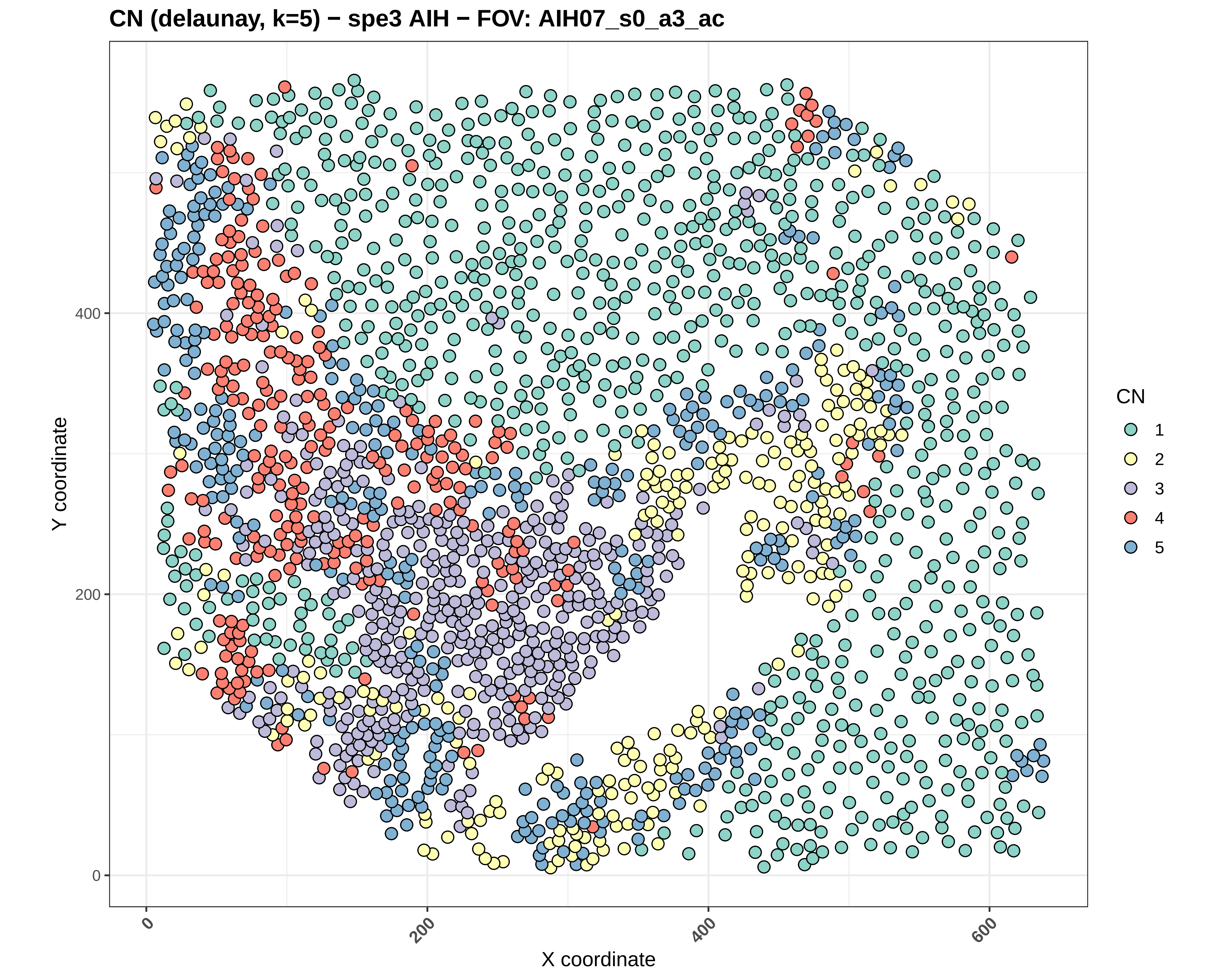
CN (delaunay, k=5) − spe3 AIH − FOV: AIH07_s0_a3_ac
Y coordinate
X coordinate
0
200
400
600
CN
1
2
3
4
5

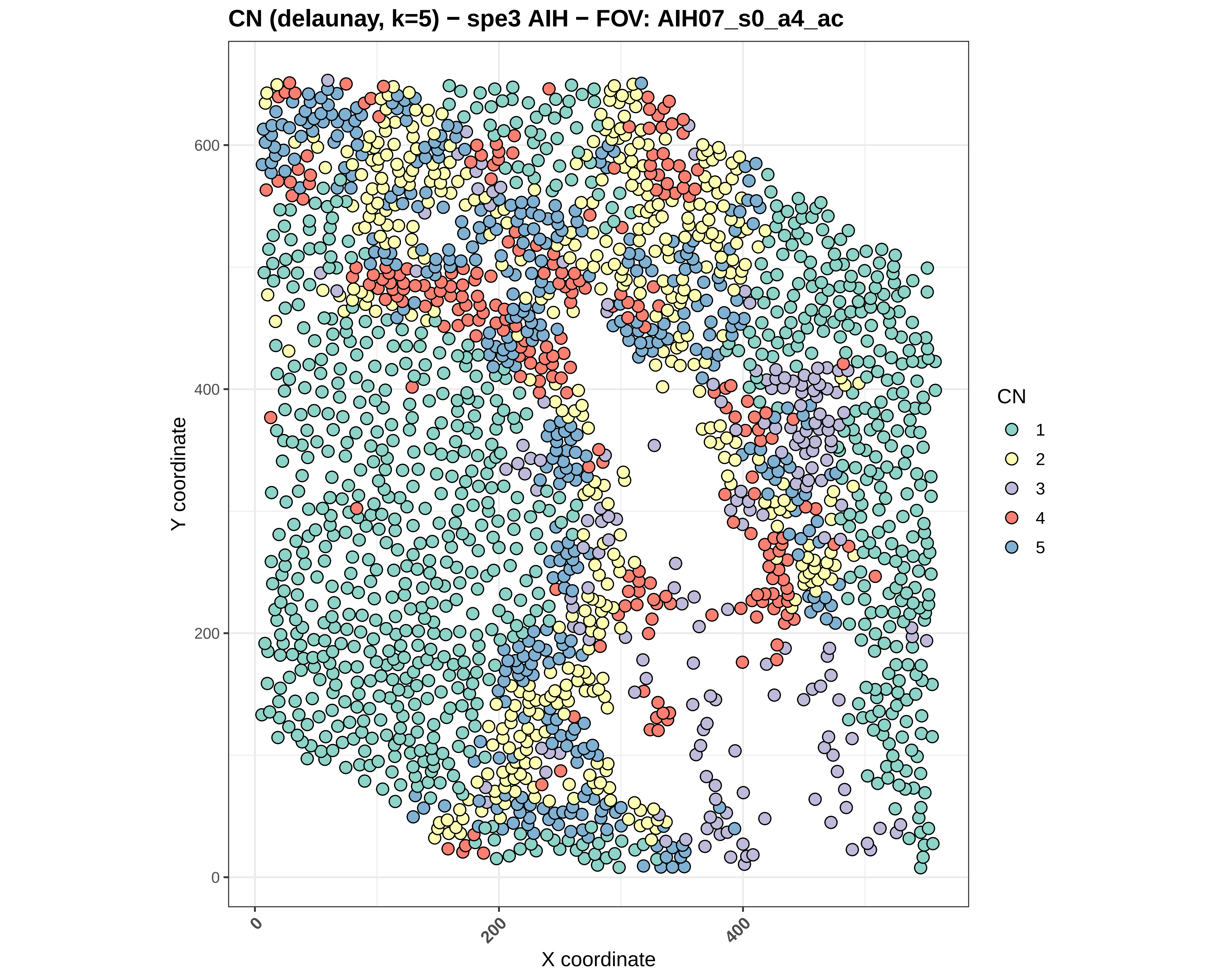
CN (delaunay, k=5) − spe3 AIH − FOV: AIH07_s0_a4_ac
Y coordinate
X coordinate
0
200
400
600
CN
1
2
3
4
5

#### CN (delaunay, k=5) − spe3 AIH − FOV: AIH07_s0_a5_ac

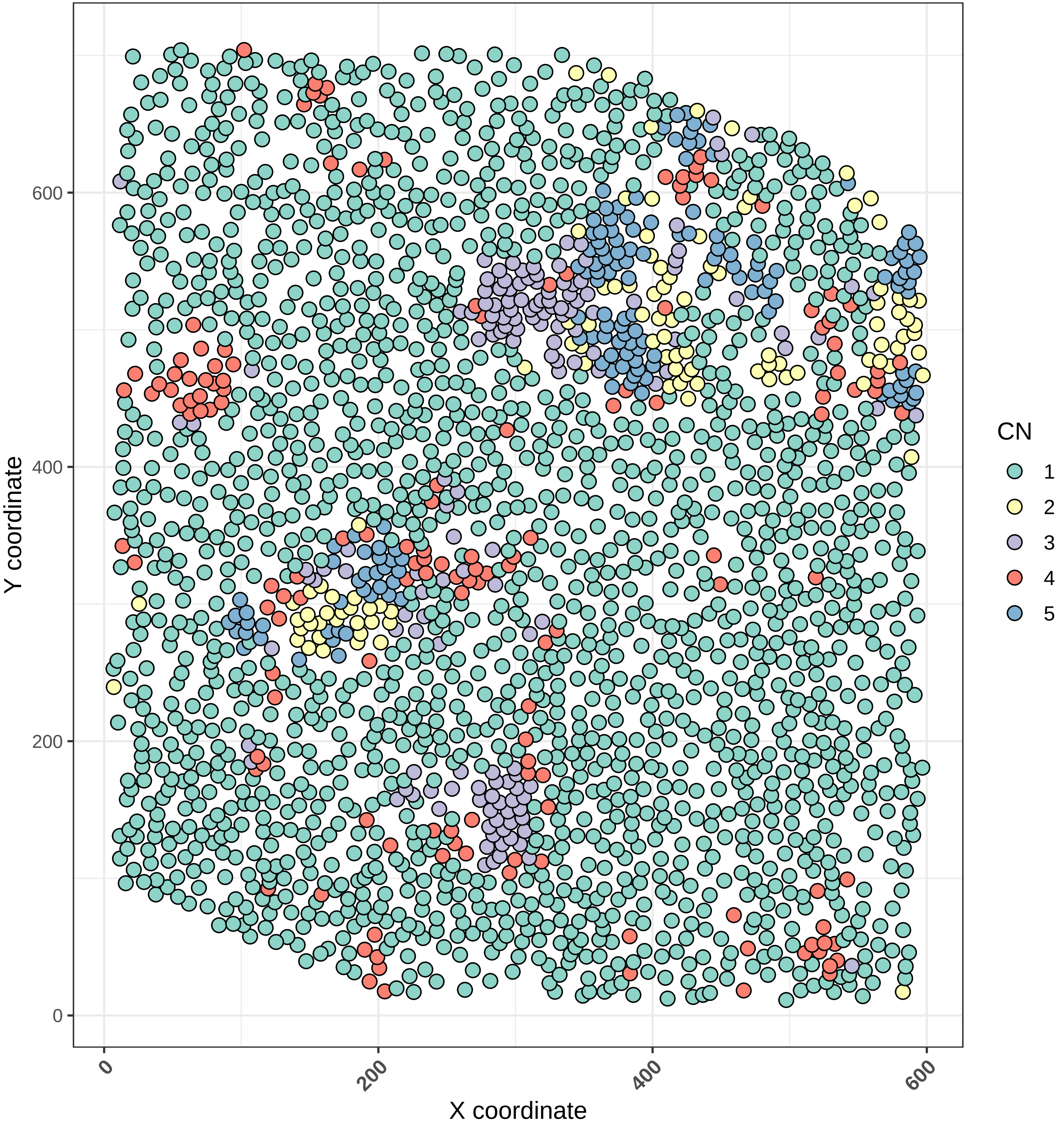

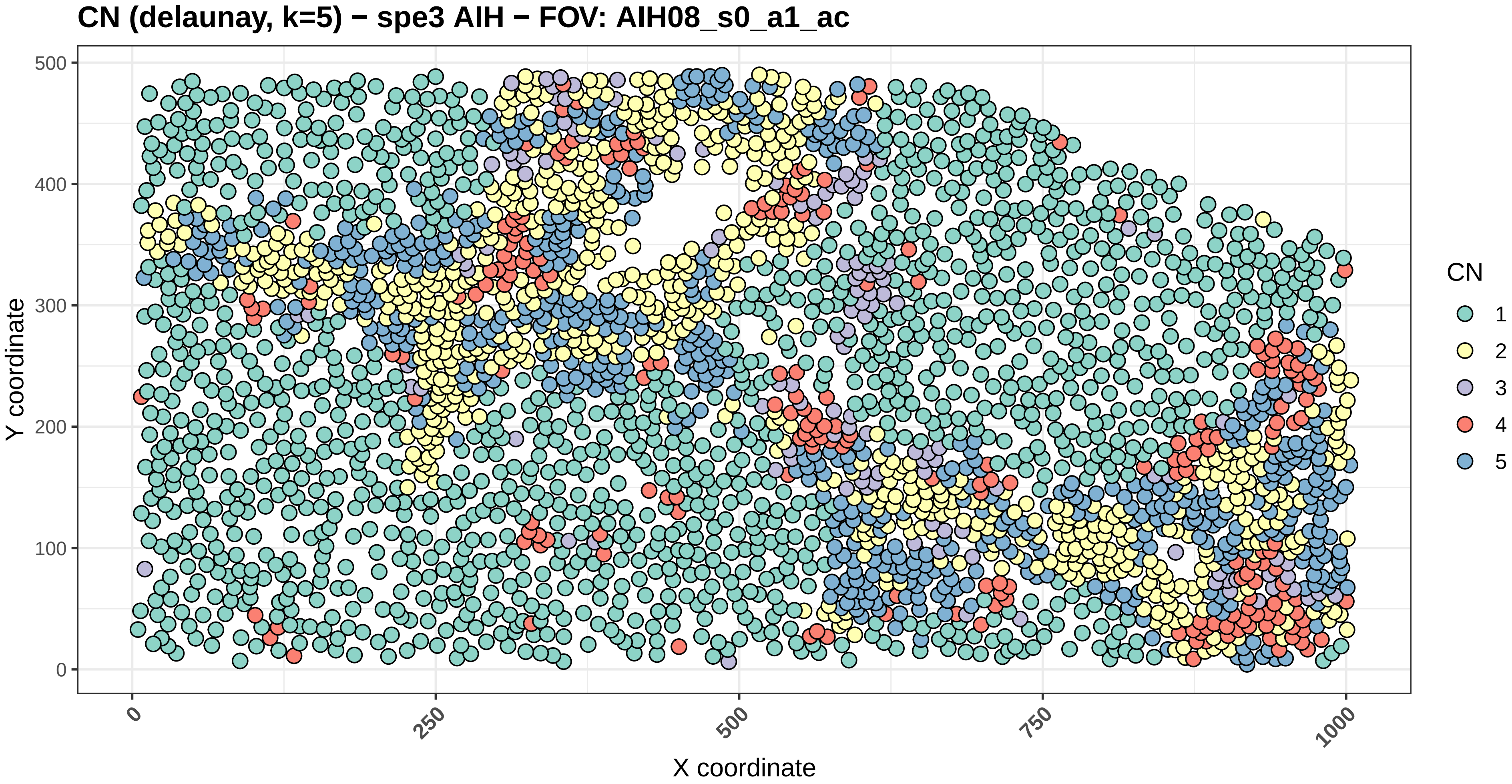

CN (delaunay, k=5) − spe3 AIH − FOV: AIH08_s0_a1_ac
Y coordinate
0
100
200
300
400
500
X coordinate
0
250
500
750
1000
CN
1
2
3
4
5

#### CN (delaunay, k=5) − spe3 AIH − FOV: AIH08_s0_a2_ac

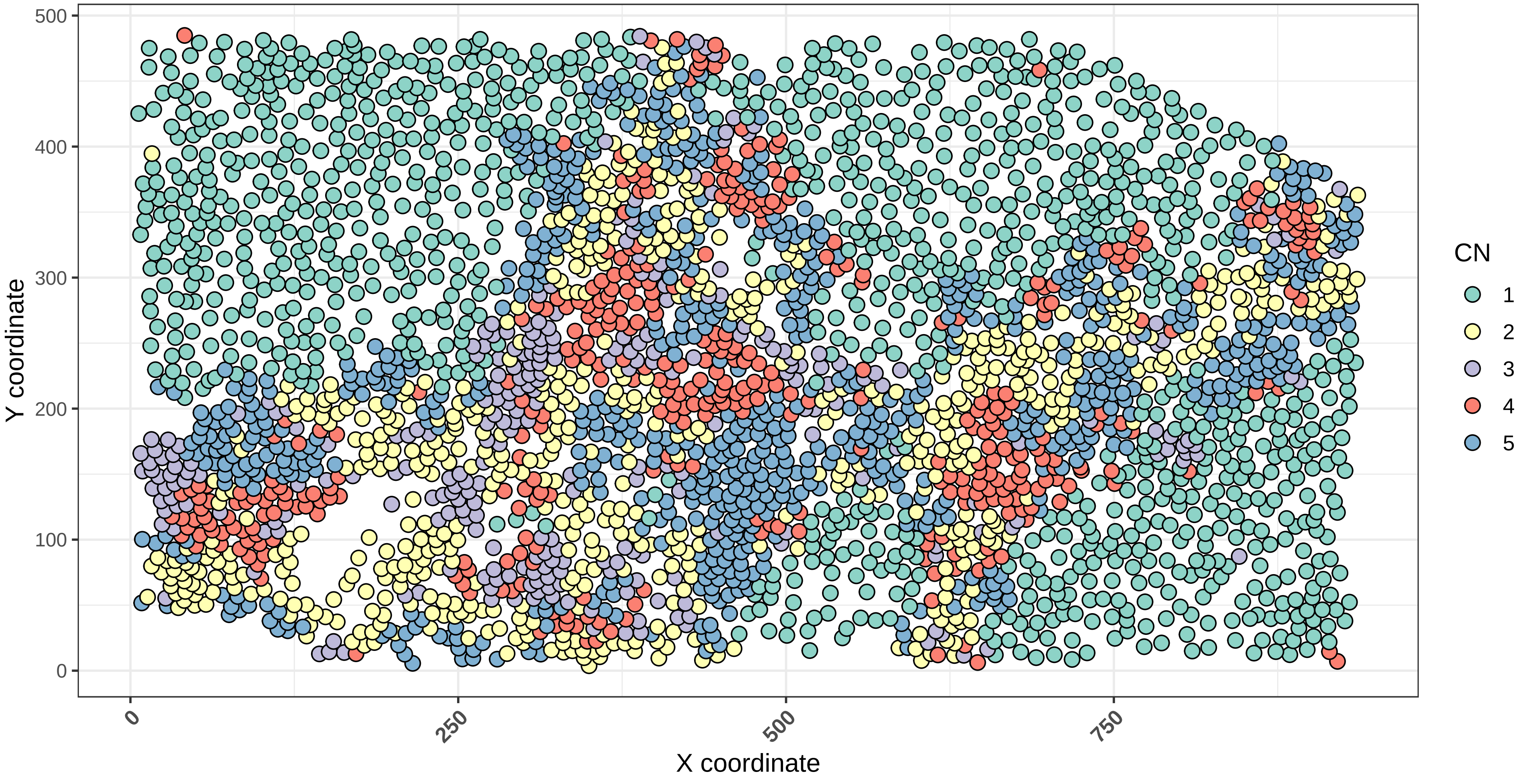

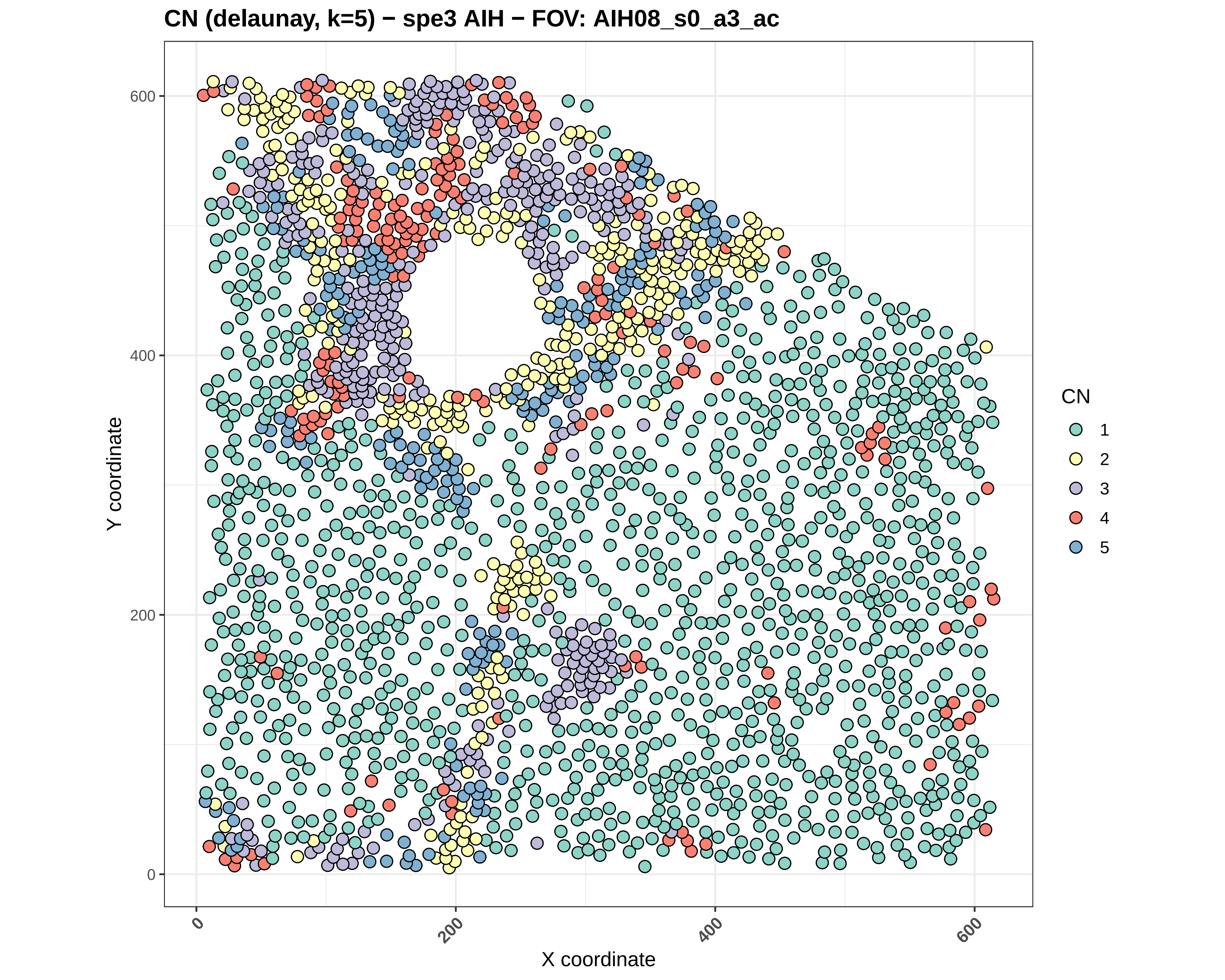
CN (delaunay, k=5) − spe3 AIH − FOV: AIH08_s0_a3_ac
Y coordinate
X coordinate
0
200
400
600
CN
1
2
3
4
5

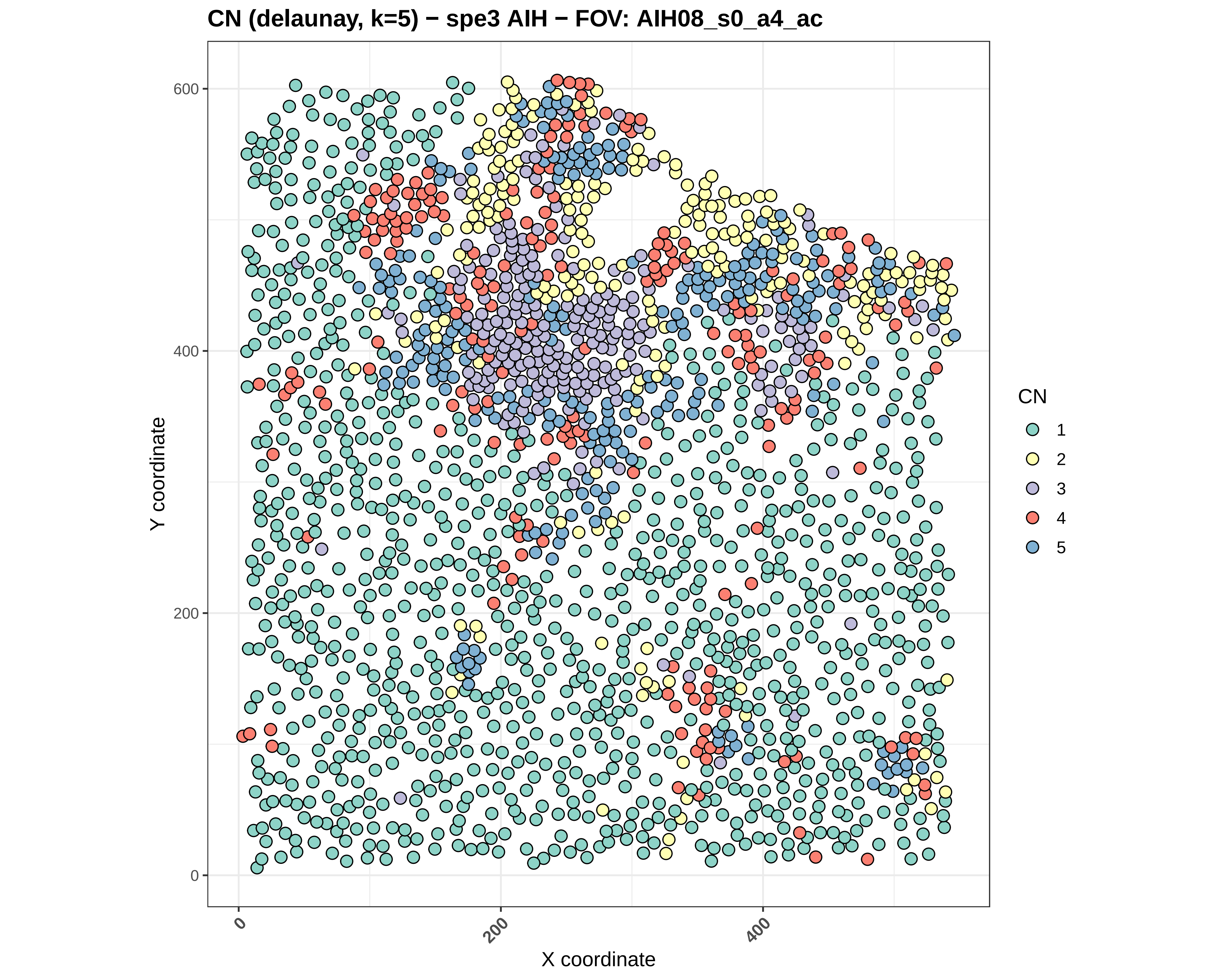

CN (delaunay, k=5) − spe3 AIH − FOV: AIH08_s0_a4_ac
Y coordinate
X coordinate
0
200
400
600
CN
1
2
3
4
5

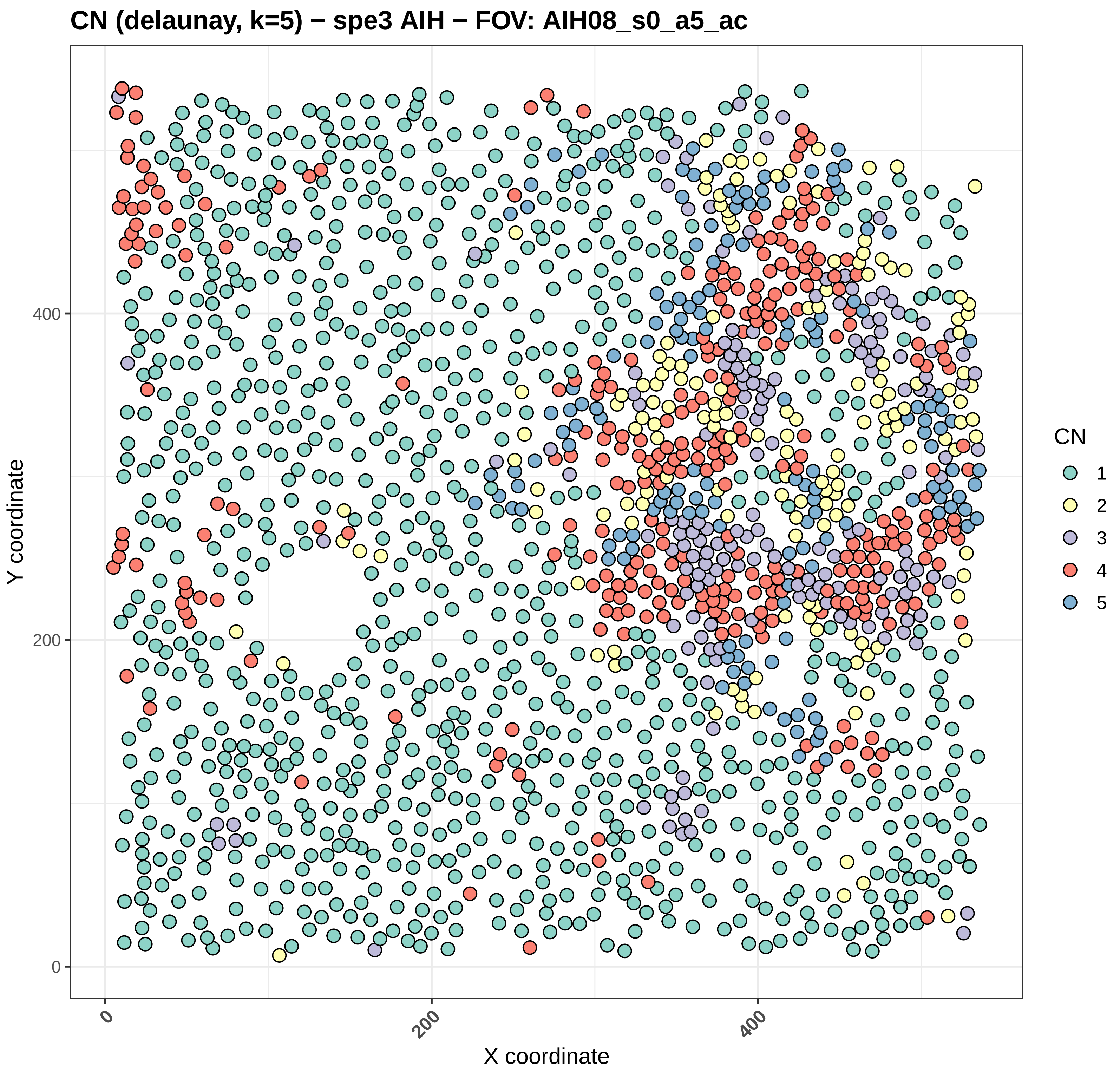
CN (delaunay, k=5) − spe3 AIH − FOV: AIH08_s0_a5_ac
Y coordinate
X coordinate
0
200
400
CN
1
2
3
4
5

CN (delaunay, k=5) − spe3 AIH − FOV: AIH09_s0_a1_ac

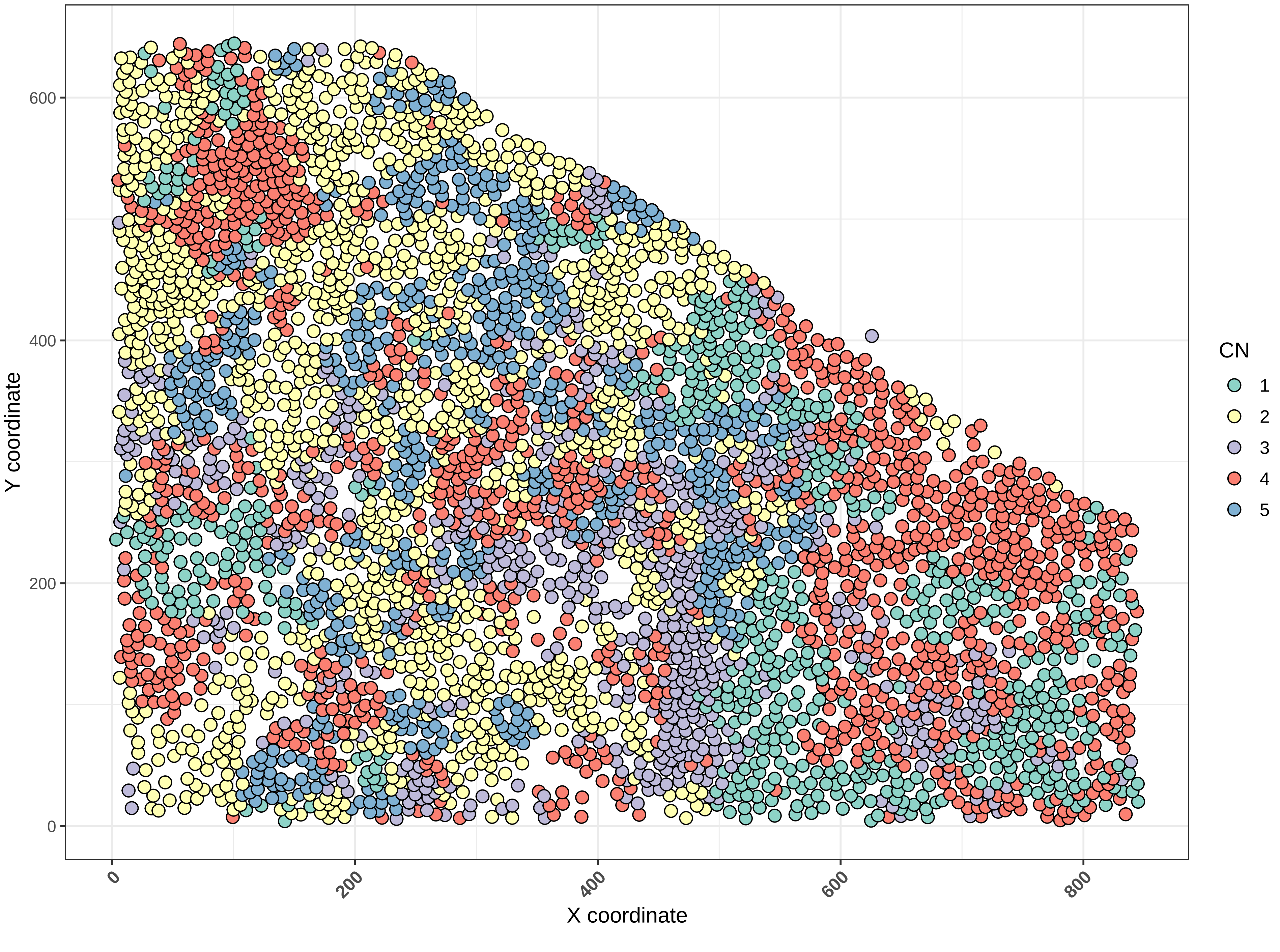

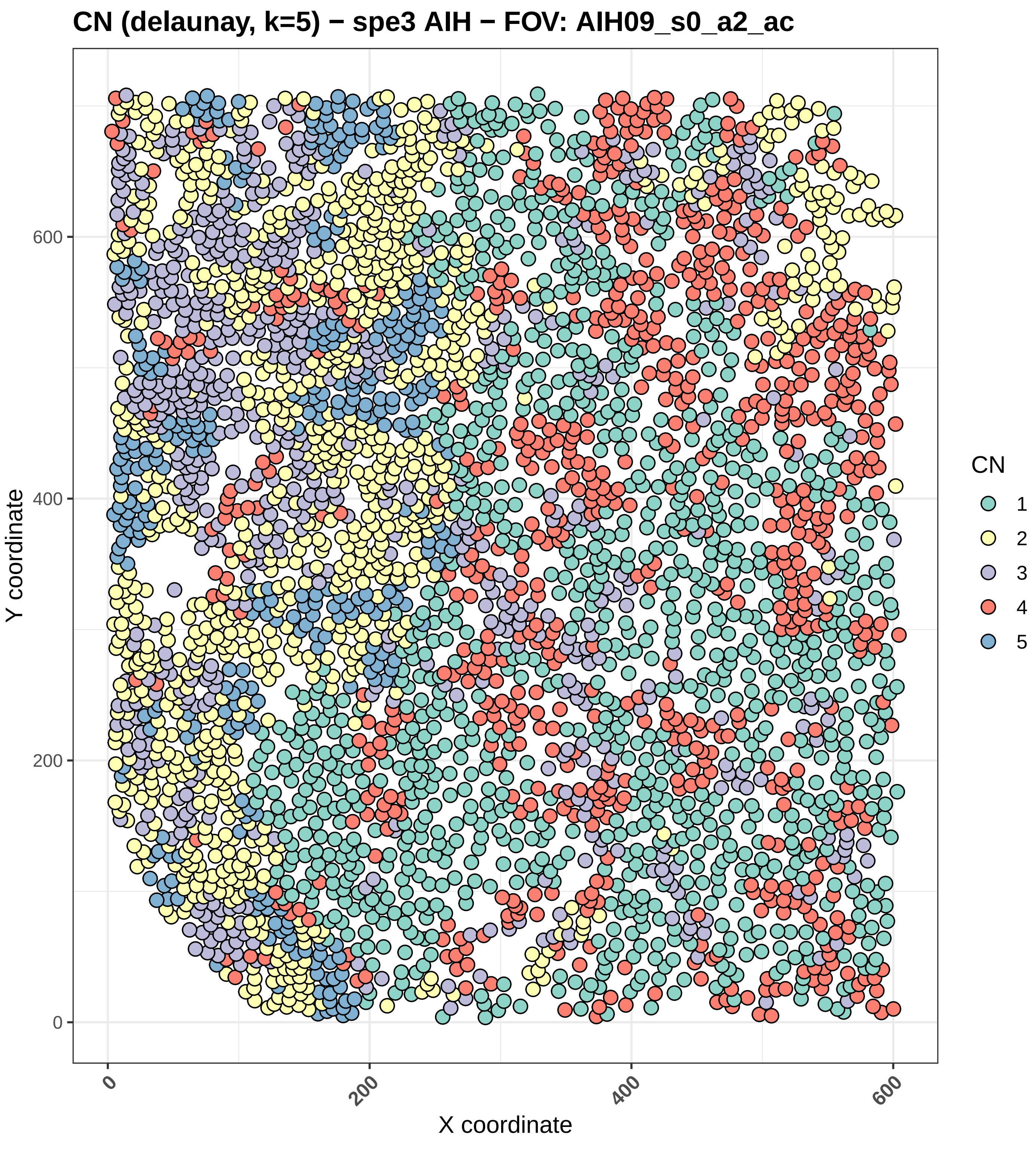
CN (delaunay, k=5) − spe3 AIH − FOV: AIH09_s0_a2_ac
Y coordinate
X coordinate
0
200
400
600
CN
1
2
3
4
5

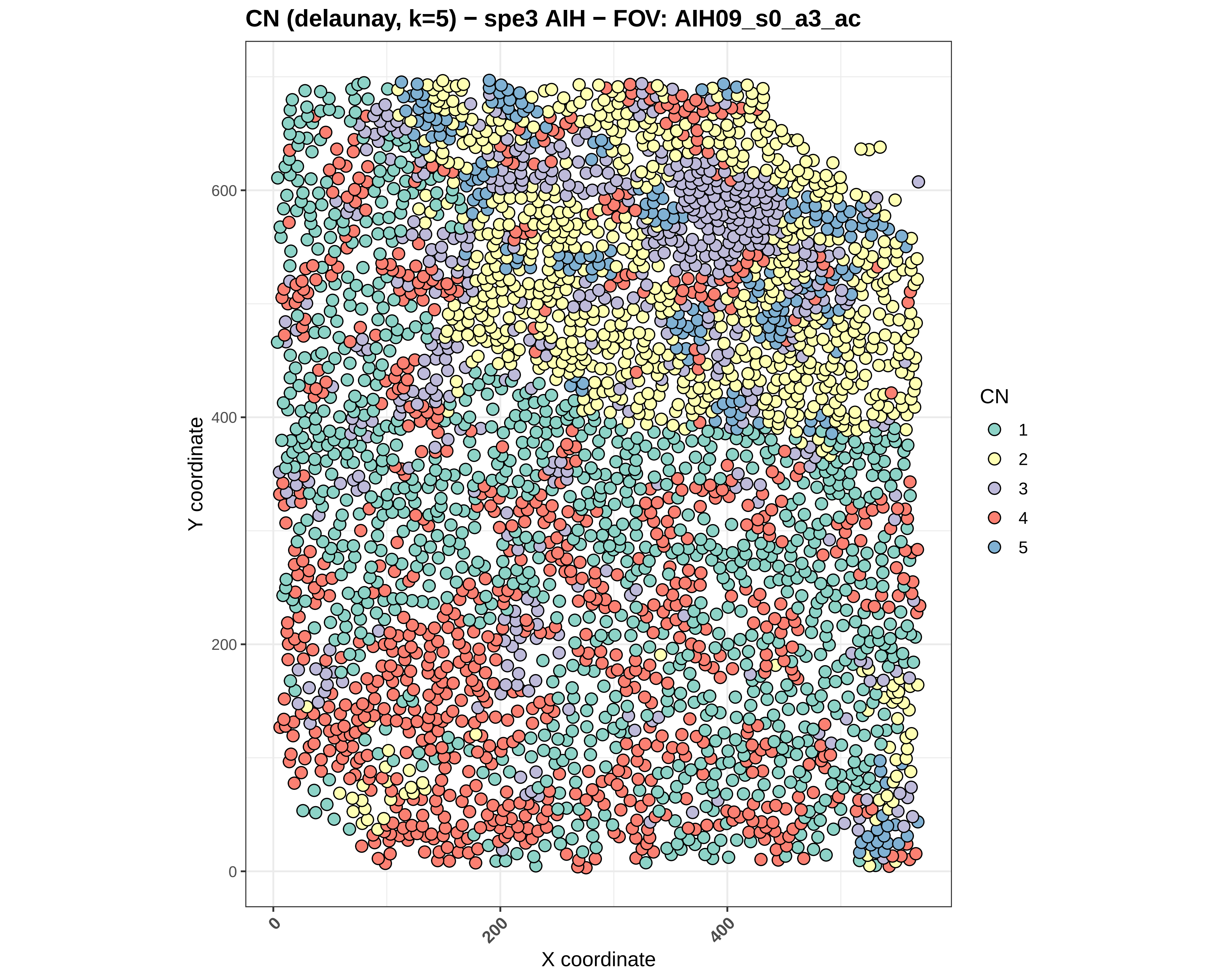
CN (delaunay, k=5) − spe3 AIH − FOV: AIH09_s0_a3_ac
Y coordinate
0
200
400
600
X coordinate
0
200
400
CN
1
2
3
4
5

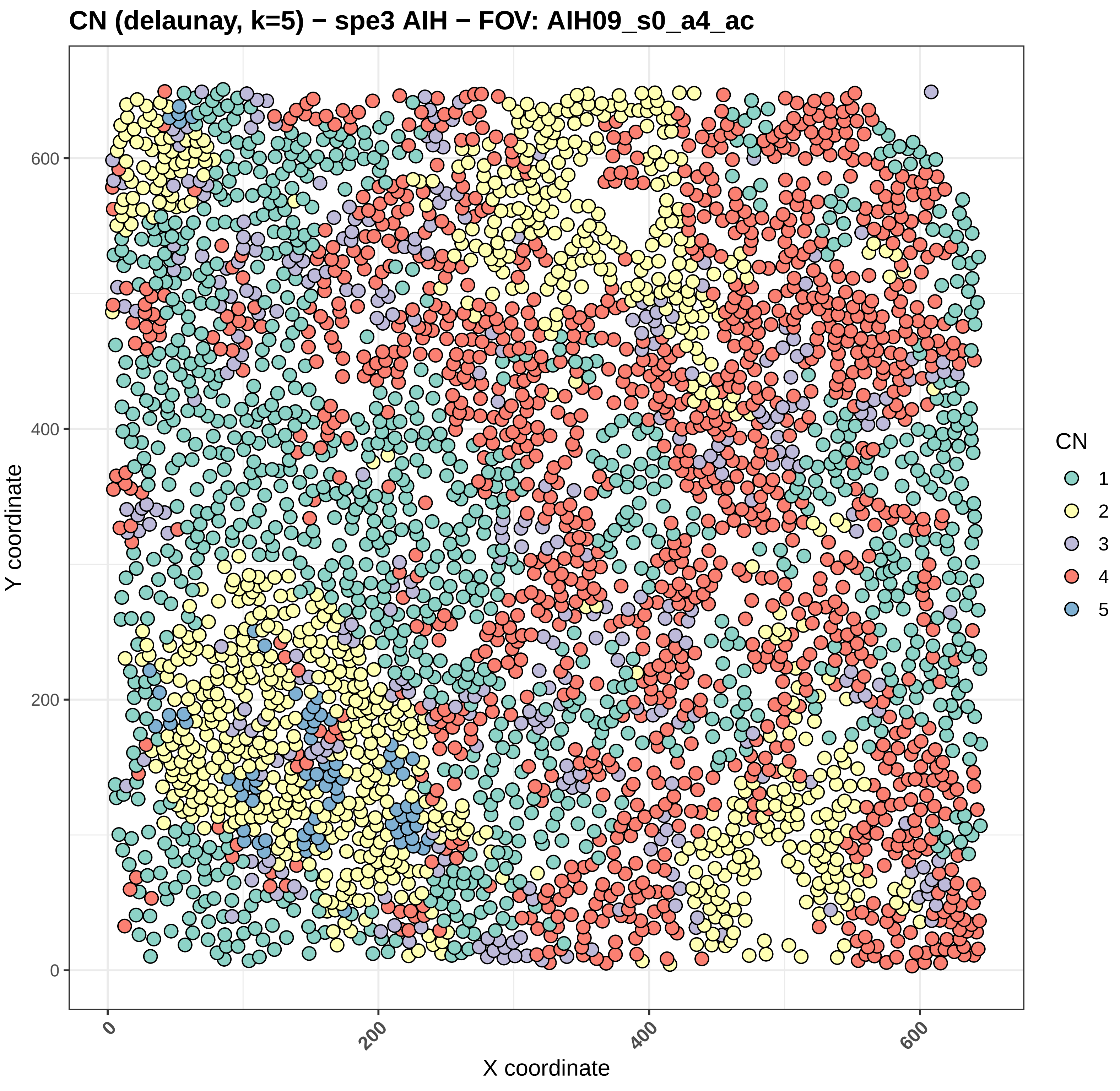
CN (delaunay, k=5) − spe3 AIH − FOV: AIH09_s0_a4_ac
Y coordinate
X coordinate
0
200
400
600
CN
1
2
3
4
5

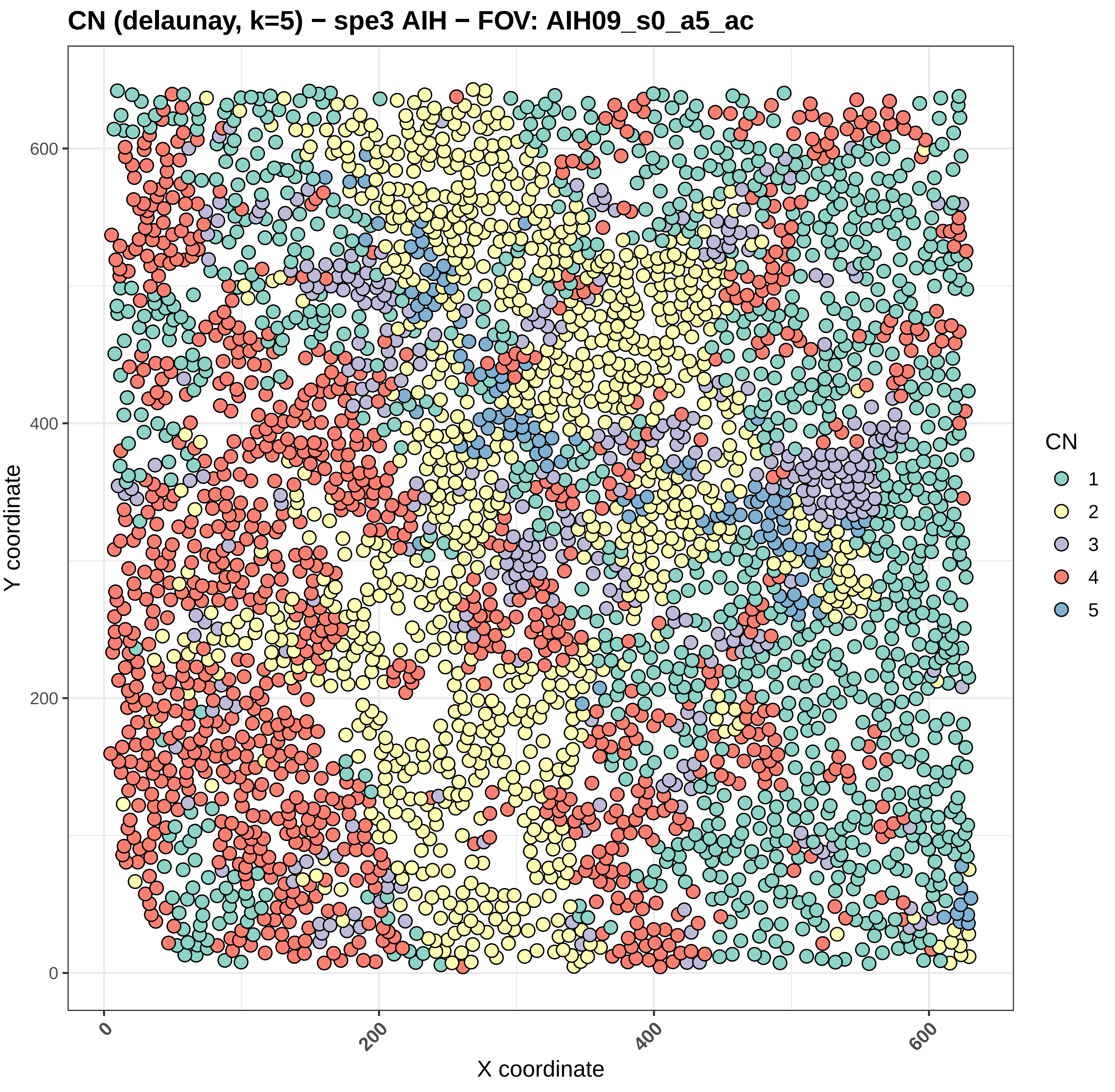

CN (delaunay, k=5) − spe3 AIH − FOV: AIH09_s0_a5_ac
Y coordinate
X coordinate
0
200
400
600
CN
1
2
3
4
5

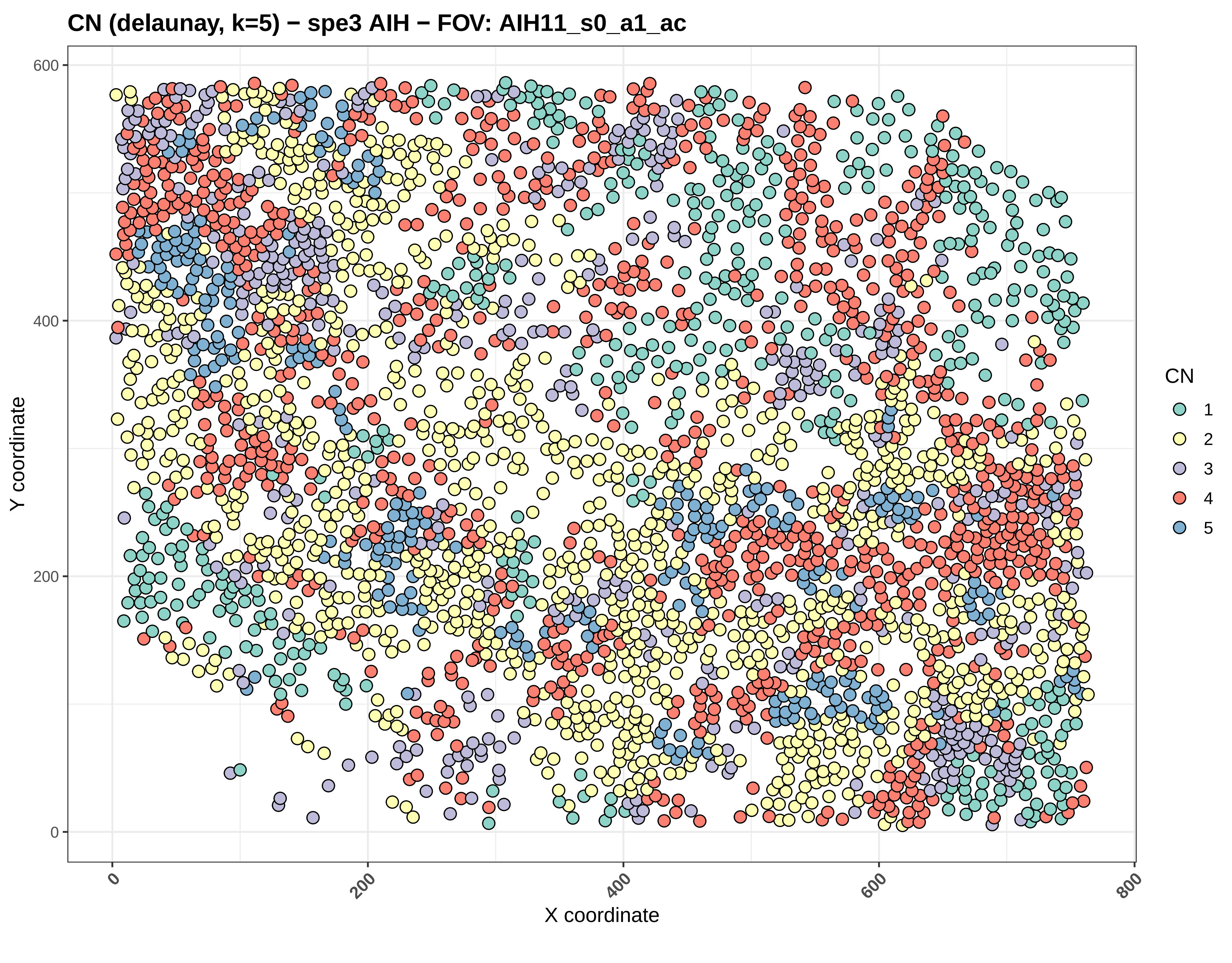
CN (delaunay, k=5) − spe3 AIH − FOV: AIH11_s0_a1_ac
Y coordinate
X coordinate
0
200
400
600
0
200
400
600
800
CN
1
2
3
4
5

CN (delaunay, k=5) − spe3 AIH − FOV: AIH11_s0_a2_ac

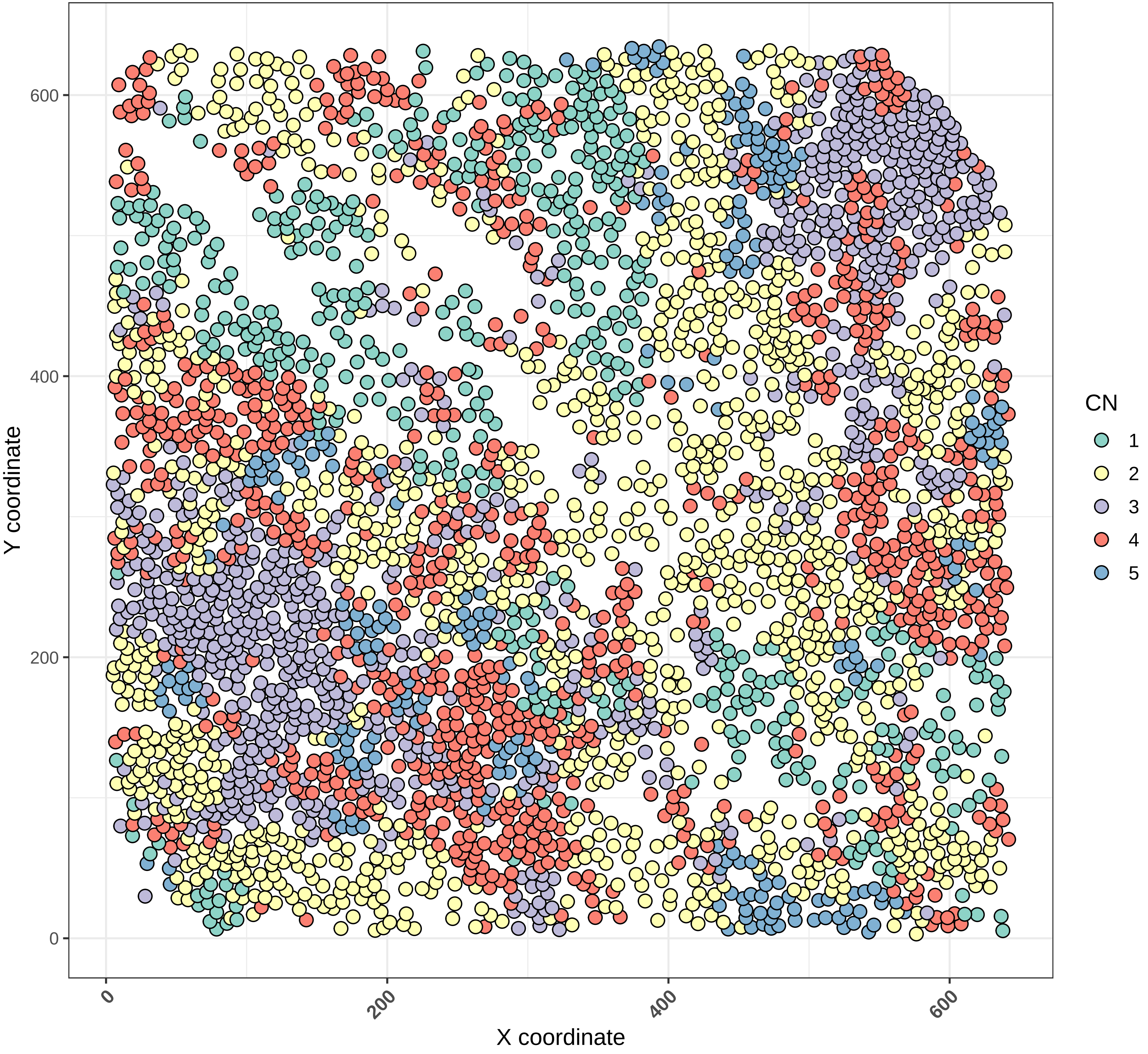

#### CN (delaunay, k=5) − spe3 AIH − FOV: AIH11_s0_a3_ac

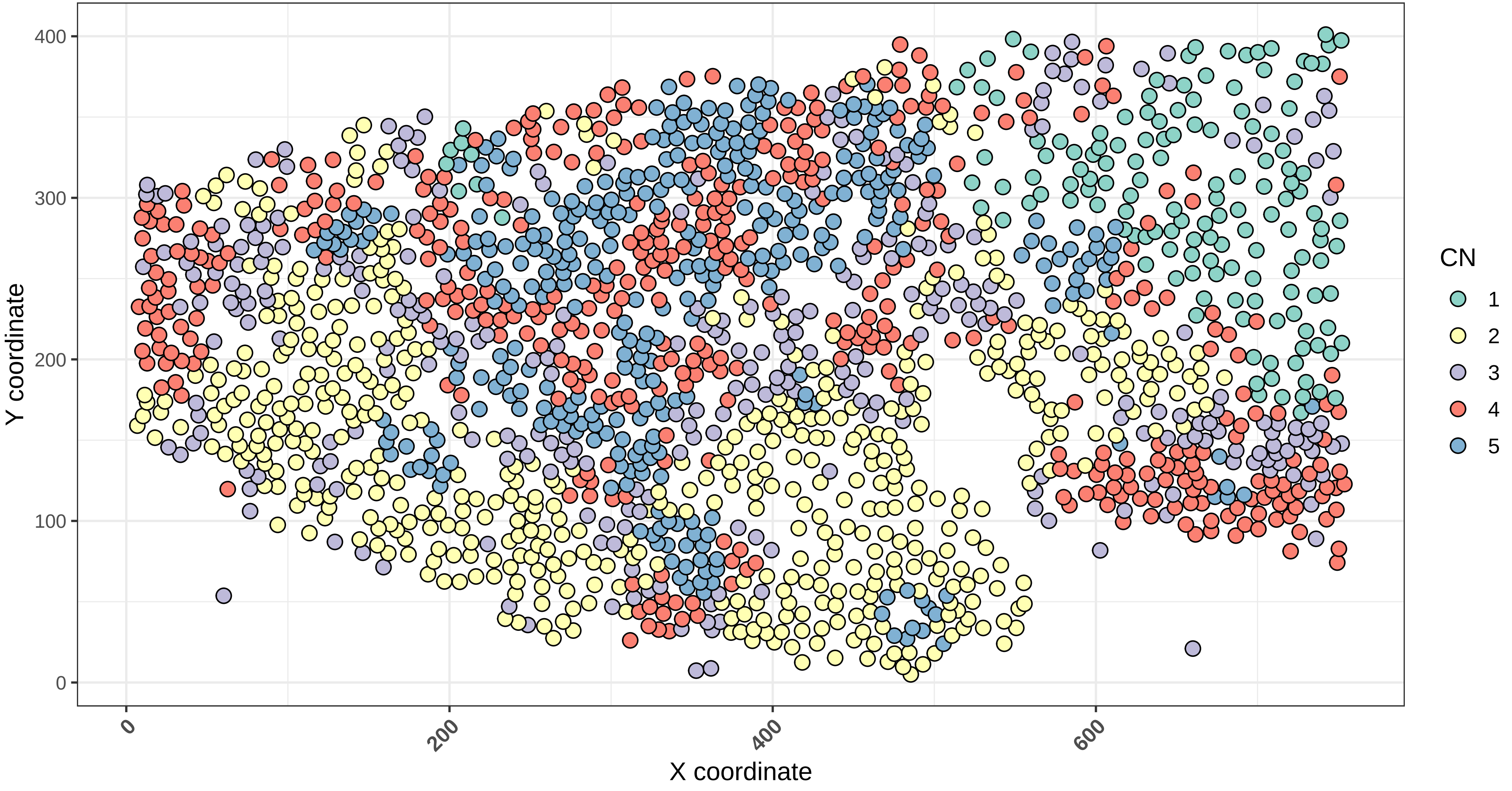

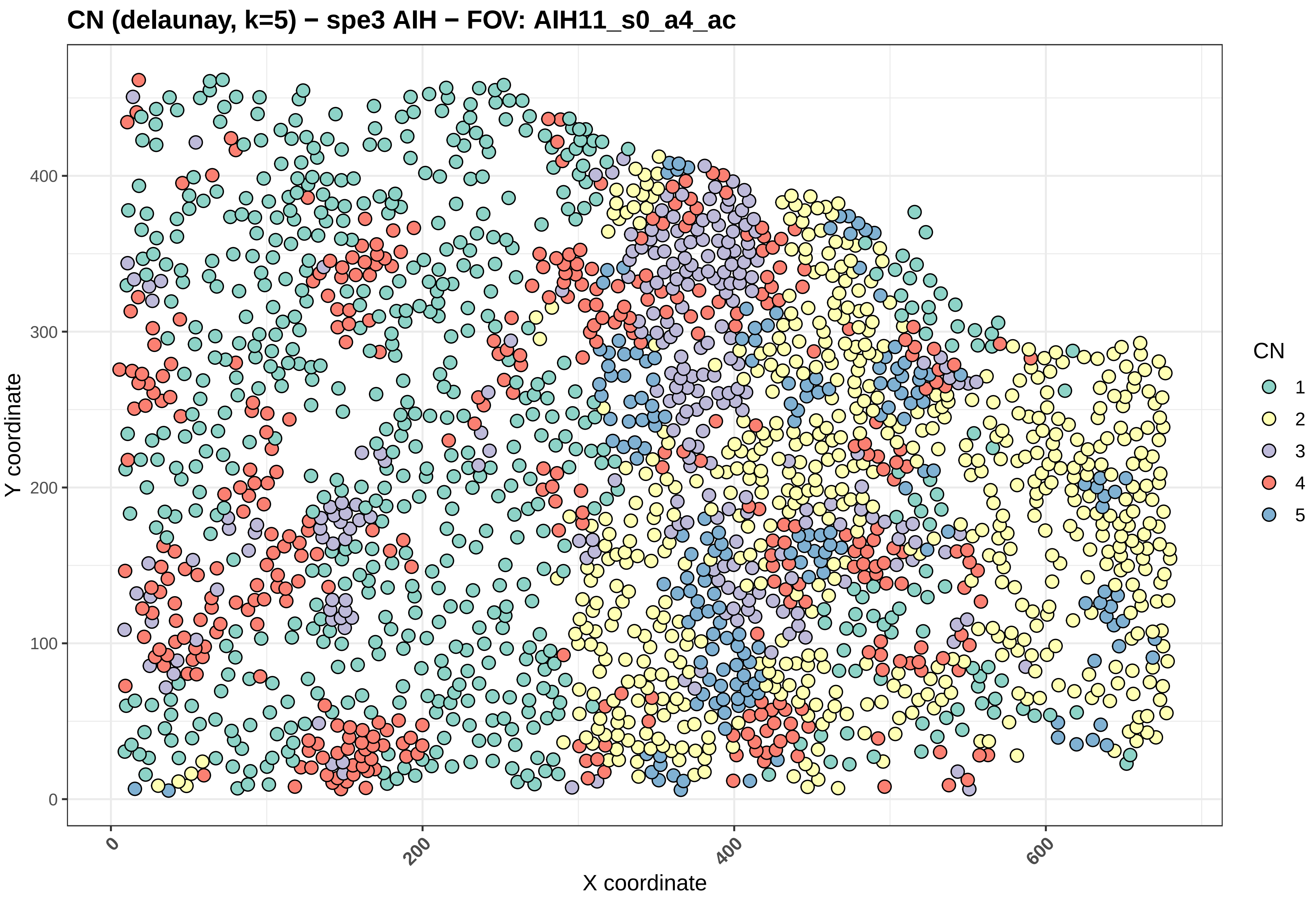
CN (delaunay, k=5) − spe3 AIH − FOV: AIH11_s0_a4_ac
Y coordinate
X coordinate
0
100
200
300
400
600
CN
1
2
3
4
5

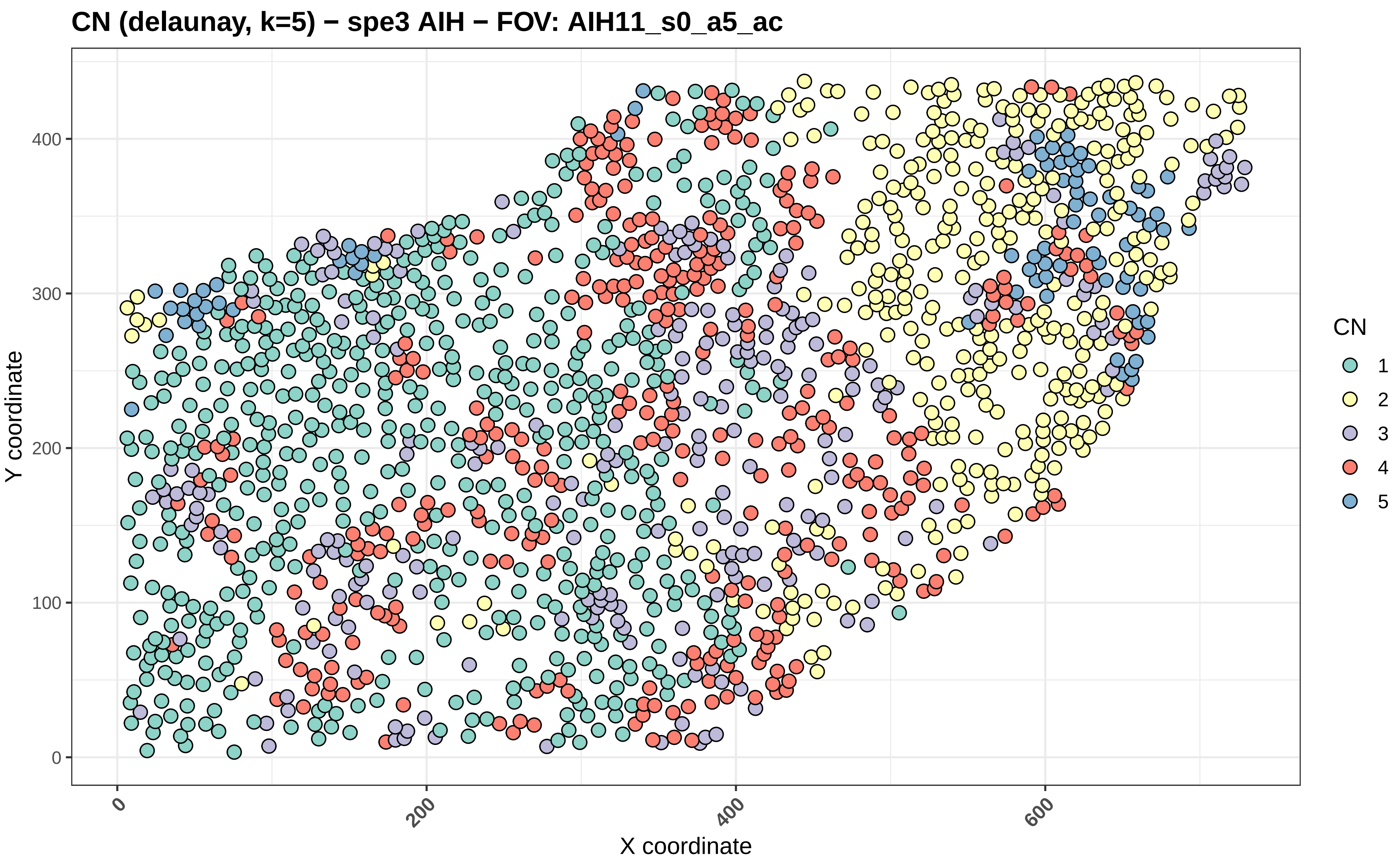

CN (delaunay, k=5) − spe3 AIH − FOV: AIH11_s0_a5_ac
Y coordinate
X coordinate
0
100
200
300
400
0
200
400
600
CN
1
2
3
4
5

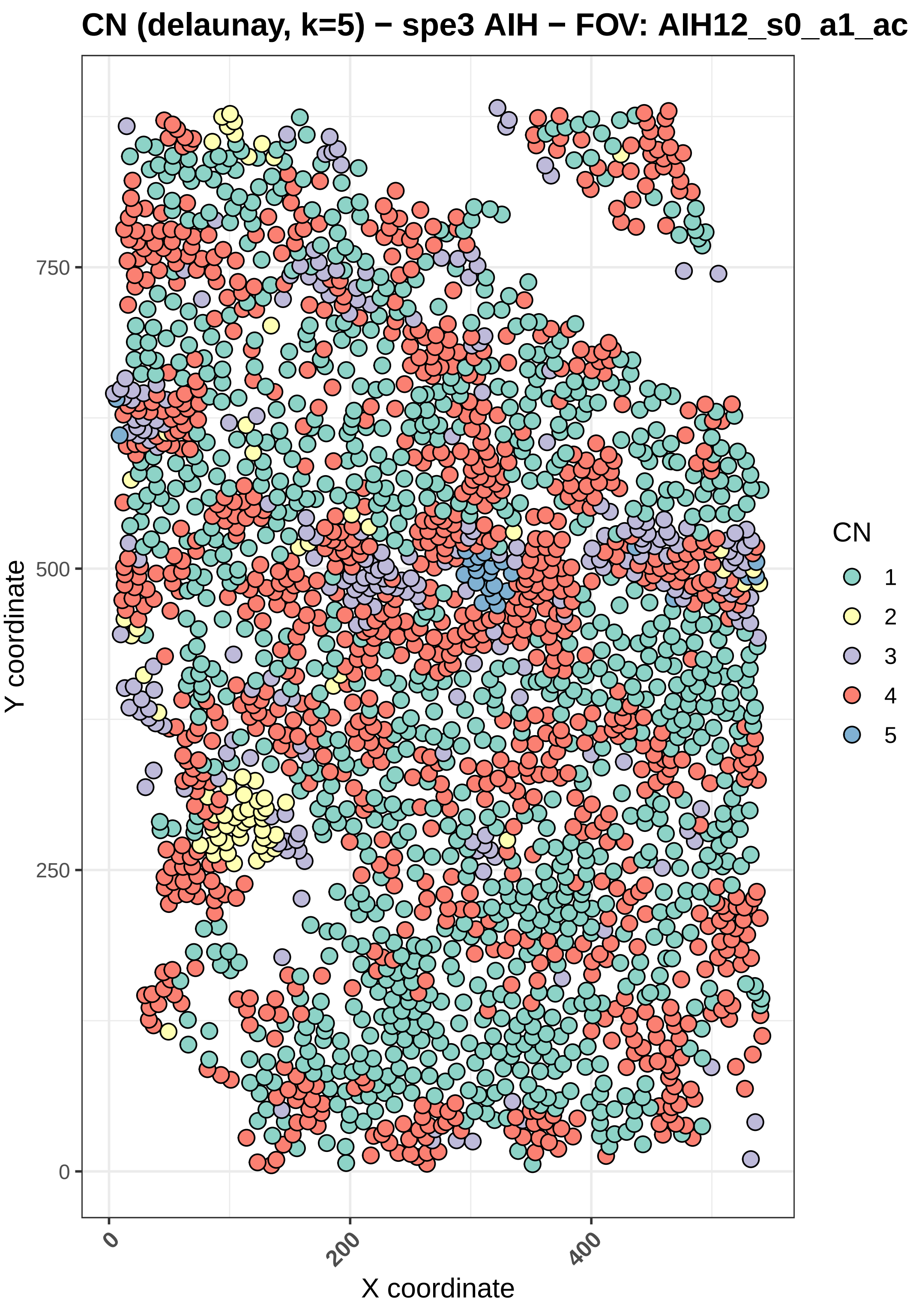
CN (delaunay, k=5) − spe3 AIH − FOV: AIH12_s0_a1_ac
Y coordinate
X coordinate
0
250
500
750
0
200
400
CN
1
2
3
4
5

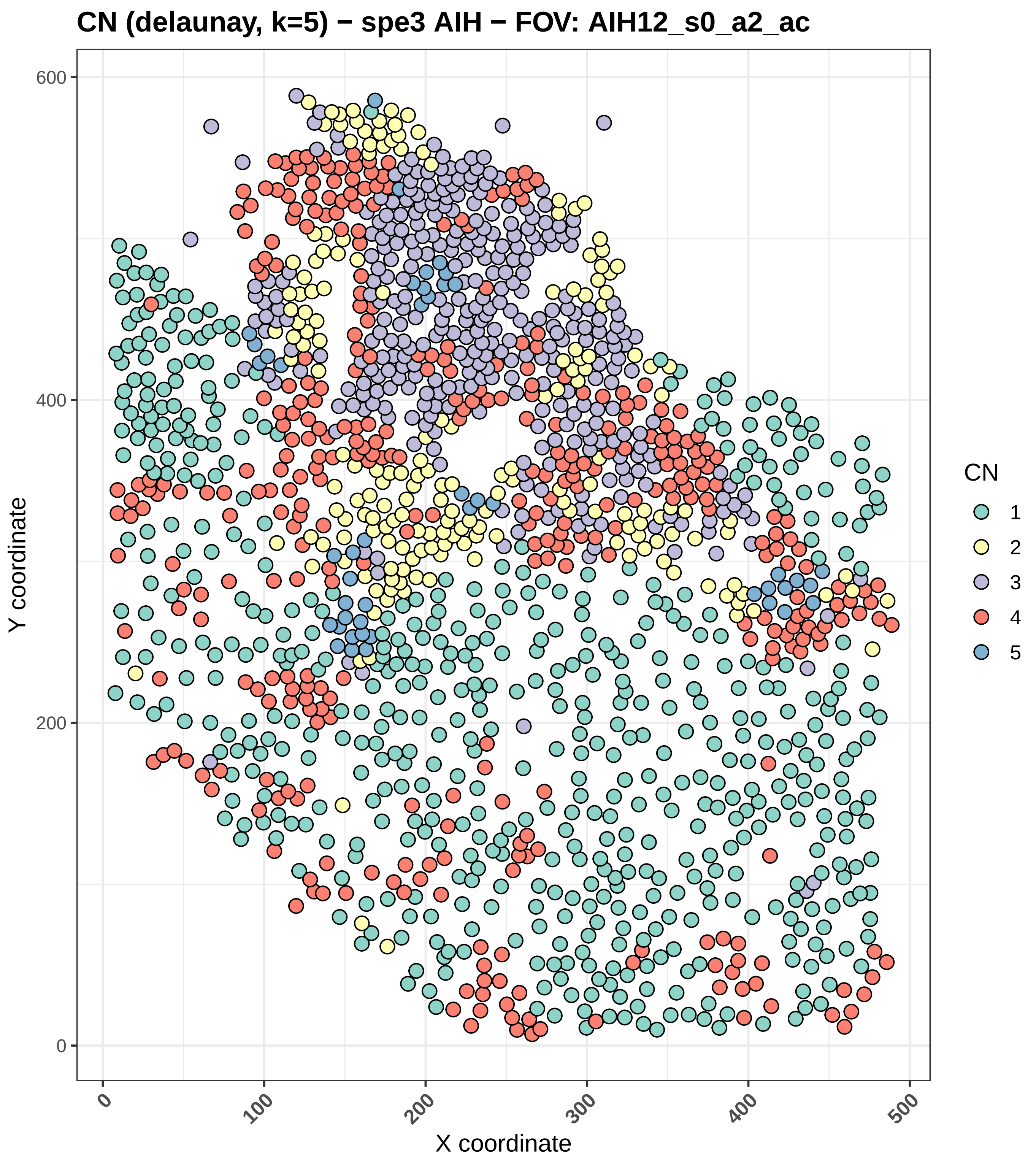
CN (delaunay, k=5) − spe3 AIH − FOV: AIH12_s0_a2_ac
Y coordinate
X coordinate
0
200
400
600
0
100
200
300
400
500
CN
1
2
3
4
5

#### CN (delaunay, k=5) − spe3 AIH − FOV: AIH12_s0_a3_ac

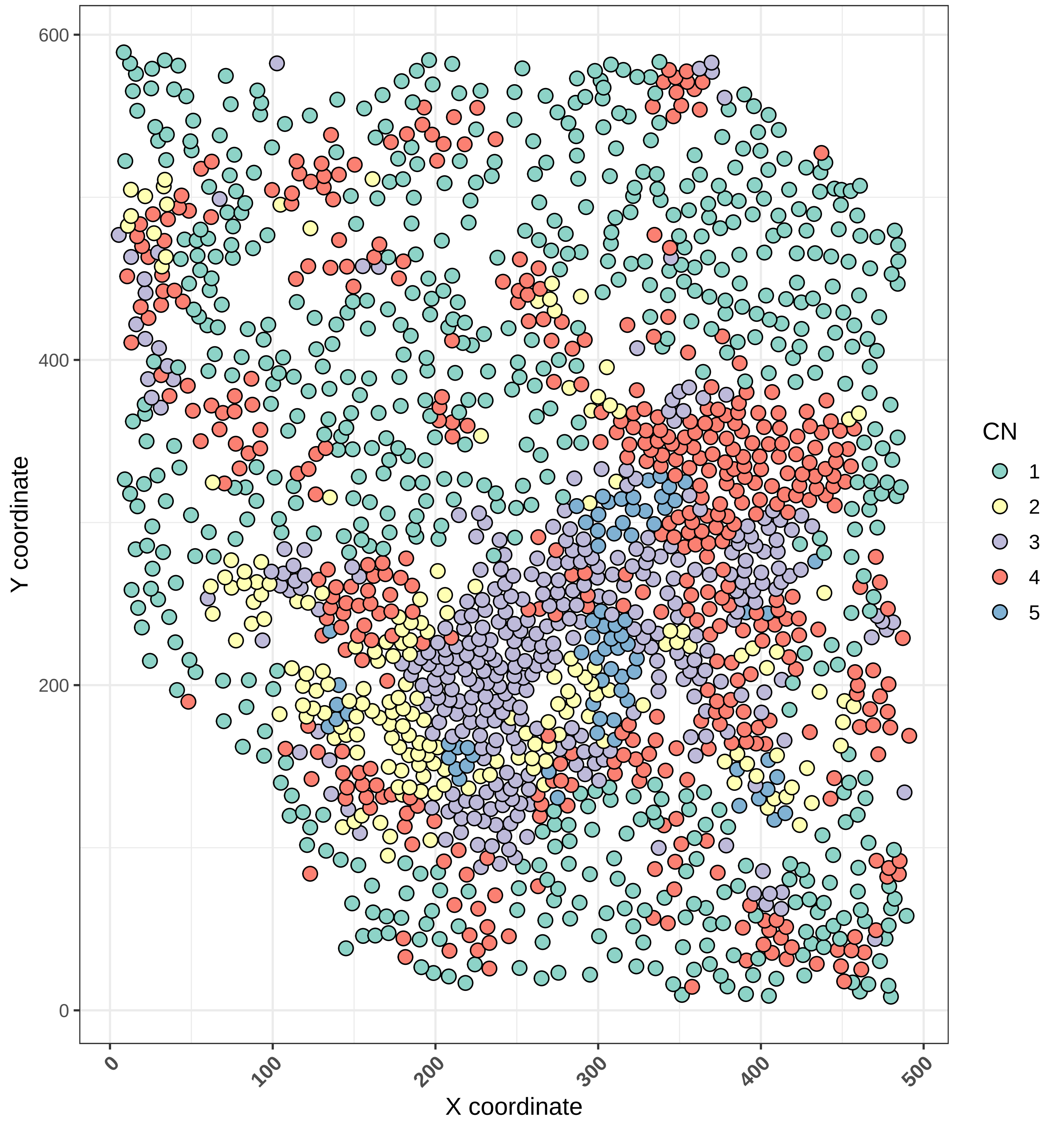

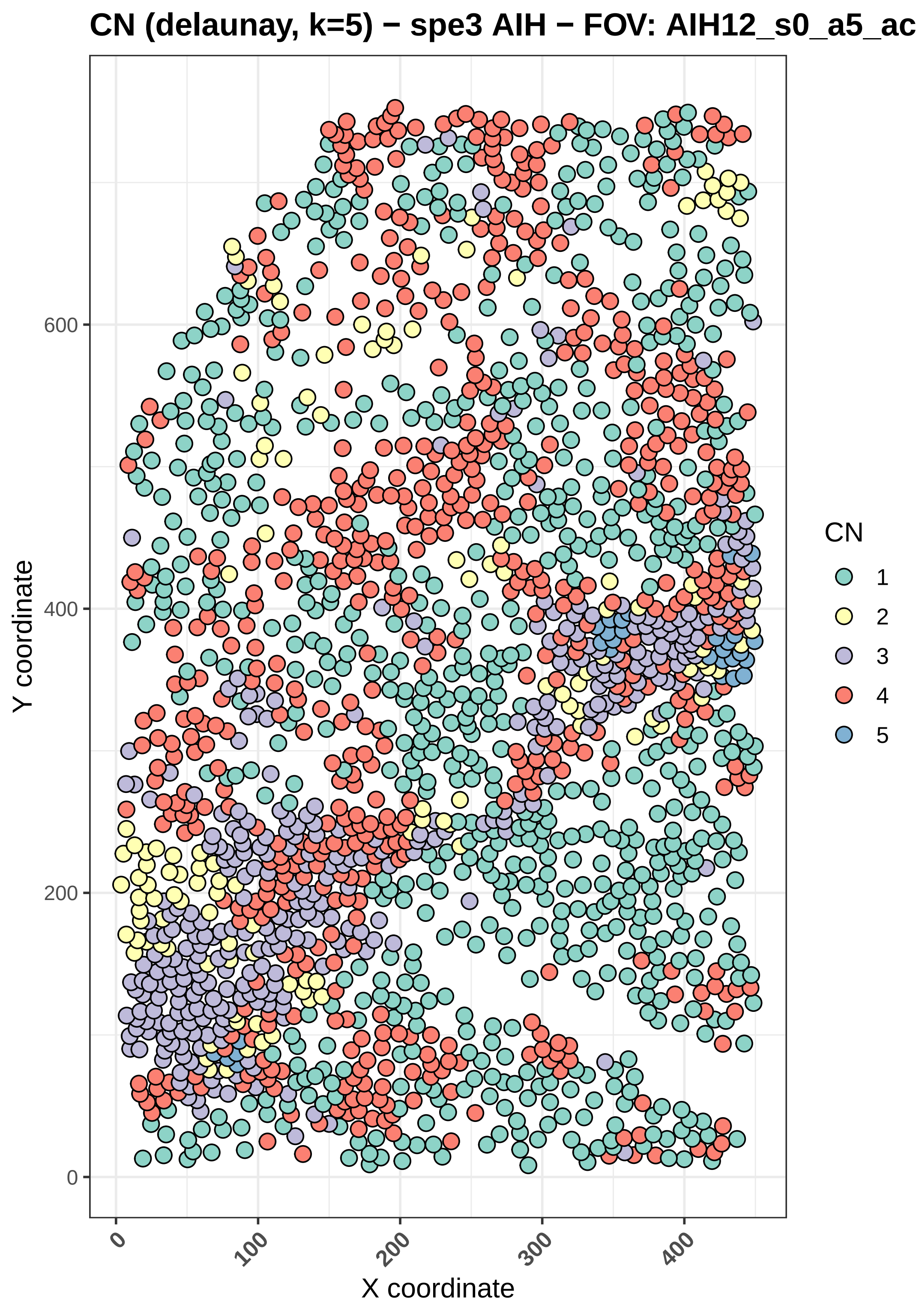
CN (delaunay, k=5) − spe3 AIH − FOV: AIH12_s0_a5_ac
Y coordinate
X coordinate
0
200
400
600
0
100
200
300
400
CN
1
2
3
4
5

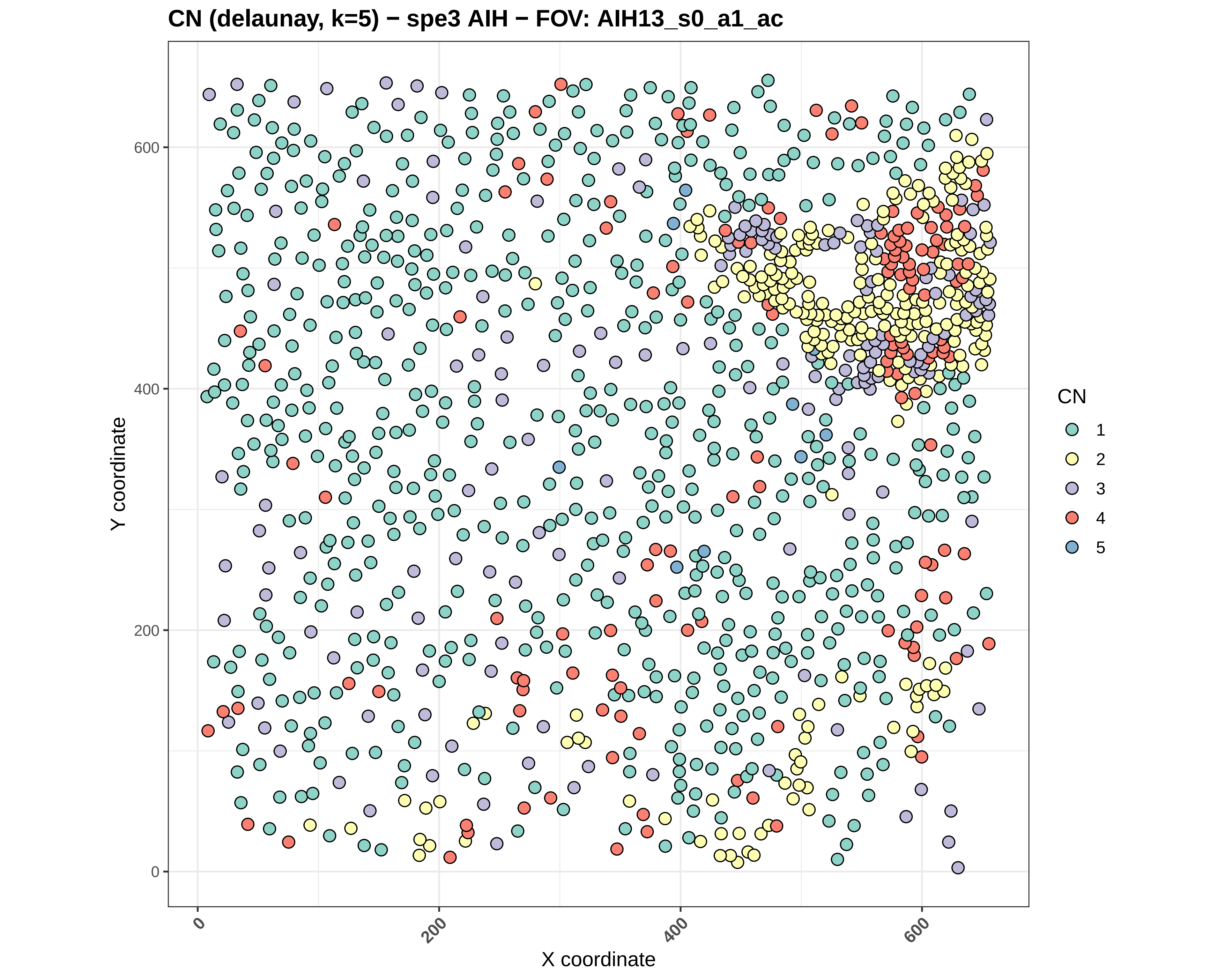
CN (delaunay, k=5) − spe3 AIH − FOV: AIH13_s0_a1_ac
Y coordinate
X coordinate
0
200
400
600
CN
1
2
3
4
5

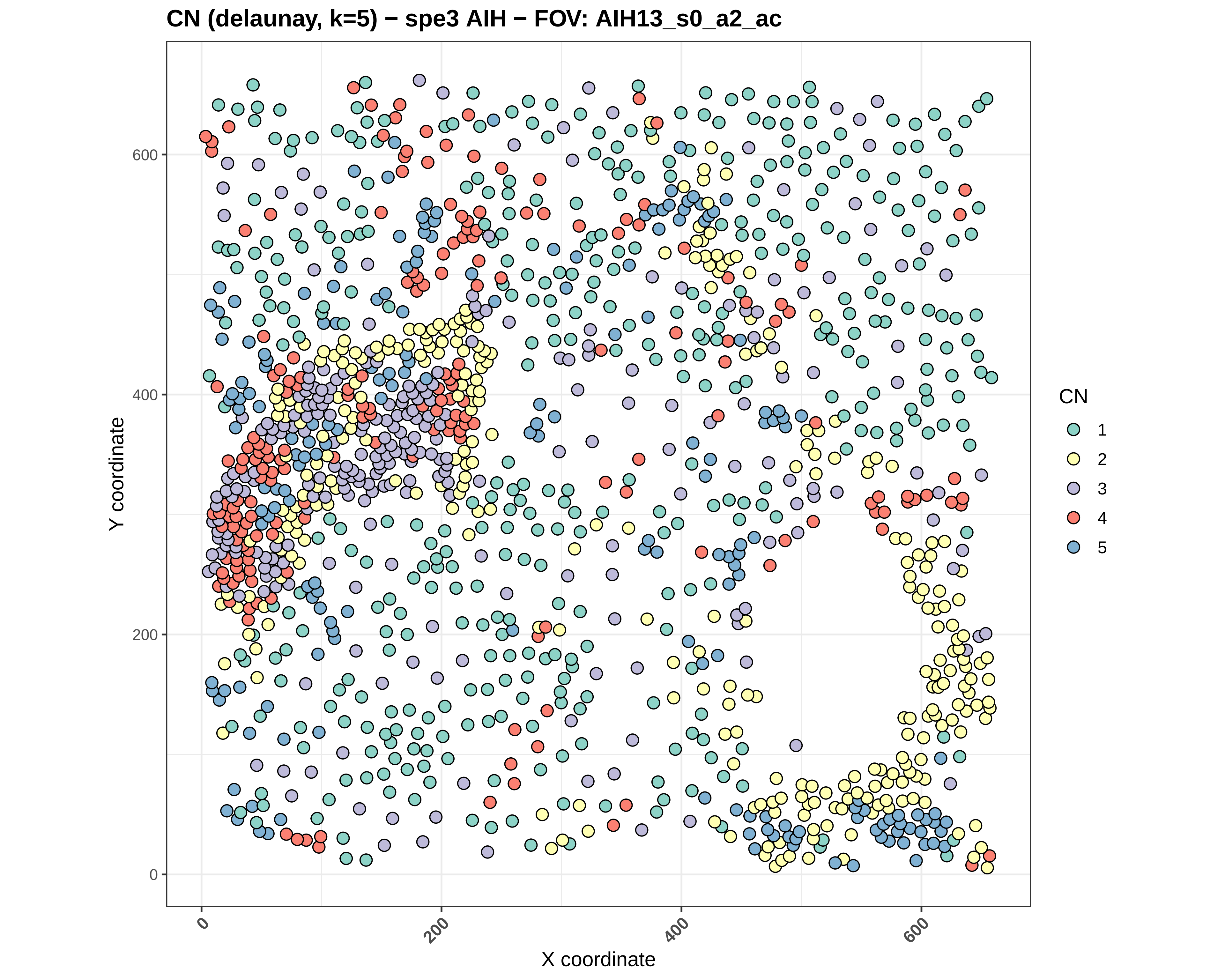

CN (delaunay, k=5) − spe3 AIH − FOV: AIH13_s0_a2_ac
Y coordinate
X coordinate
0
200
400
600
CN
1
2
3
4
5

#### CN (delaunay, k=5) − spe3 AIH − FOV: AIH13_s0_a3_ac

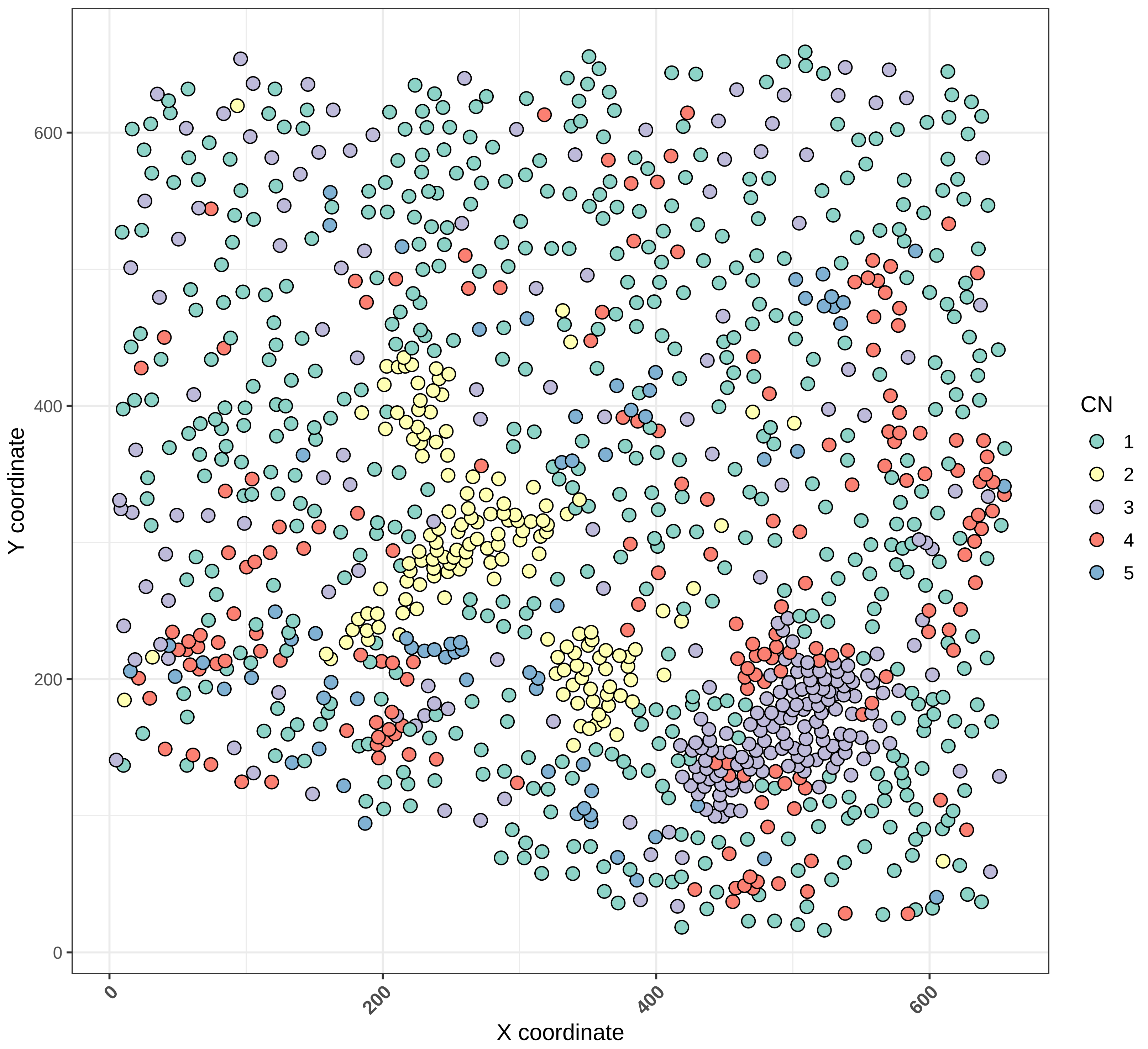

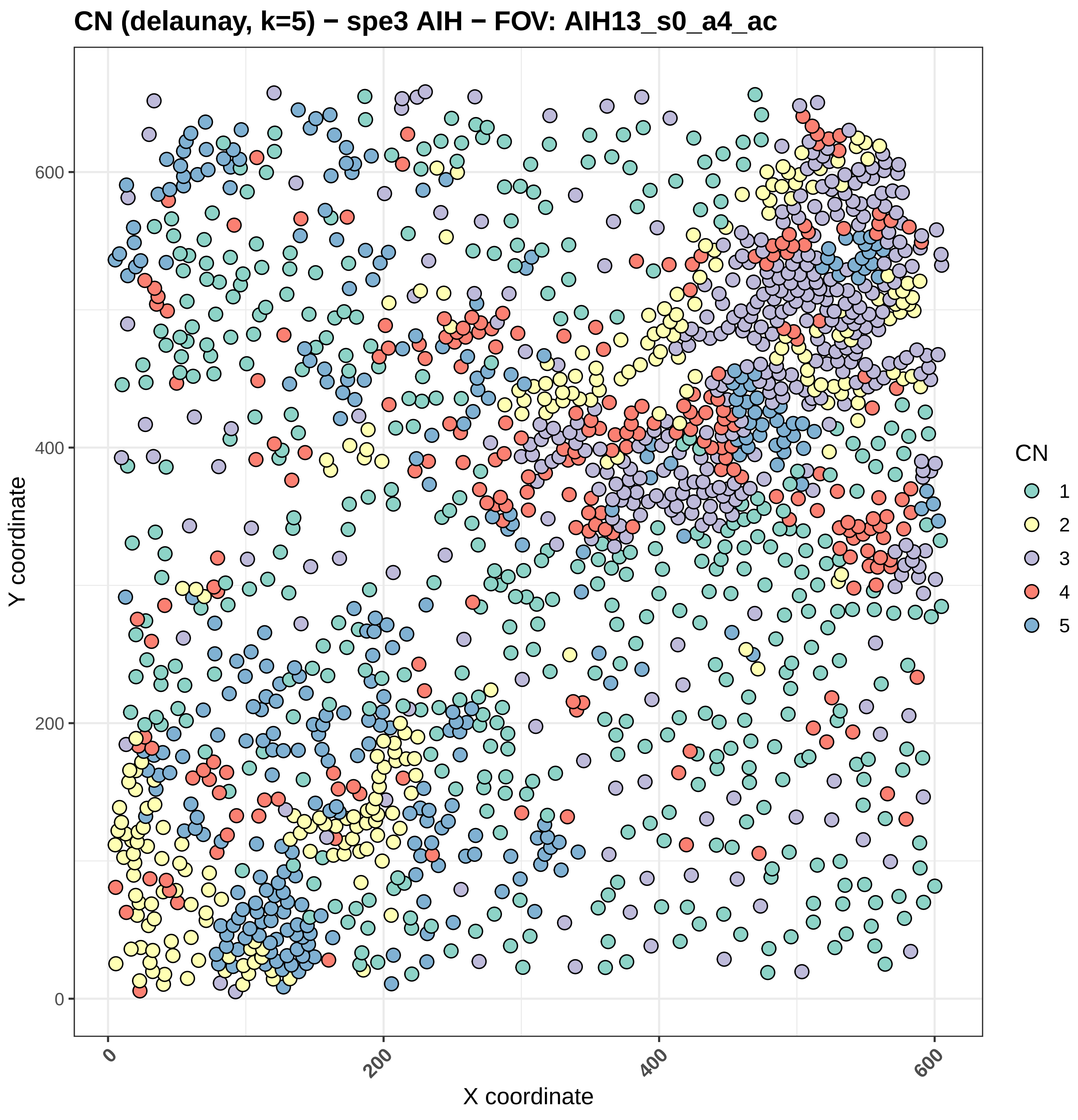
CN (delaunay, k=5) − spe3 AIH − FOV: AIH13_s0_a4_ac
Y coordinate
X coordinate
0
200
400
600
CN
1
2
3
4
5

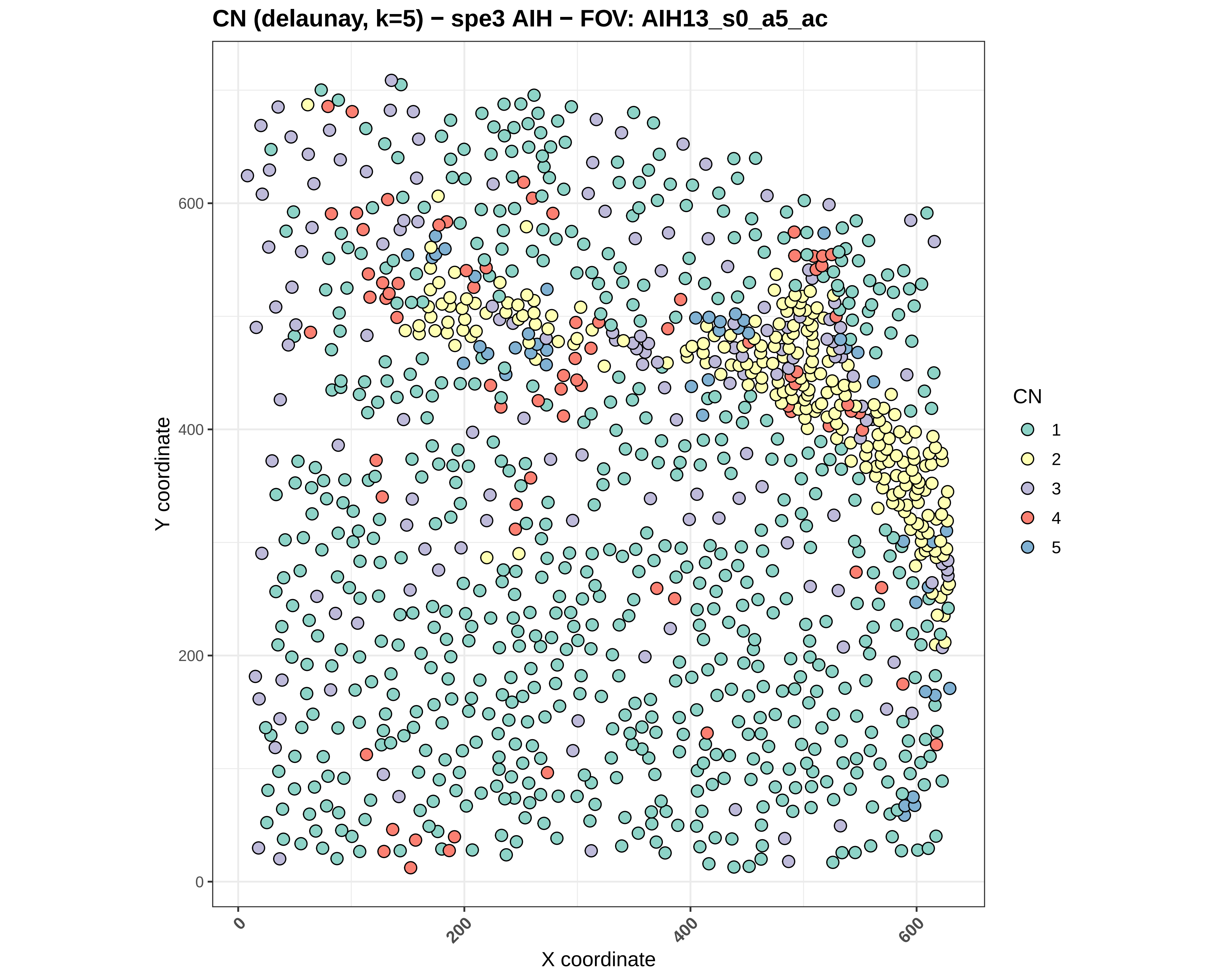
CN (delaunay, k=5) − spe3 AIH − FOV: AIH13_s0_a5_ac
Y coordinate
X coordinate
0
200
400
600
CN
1
2
3
4
5

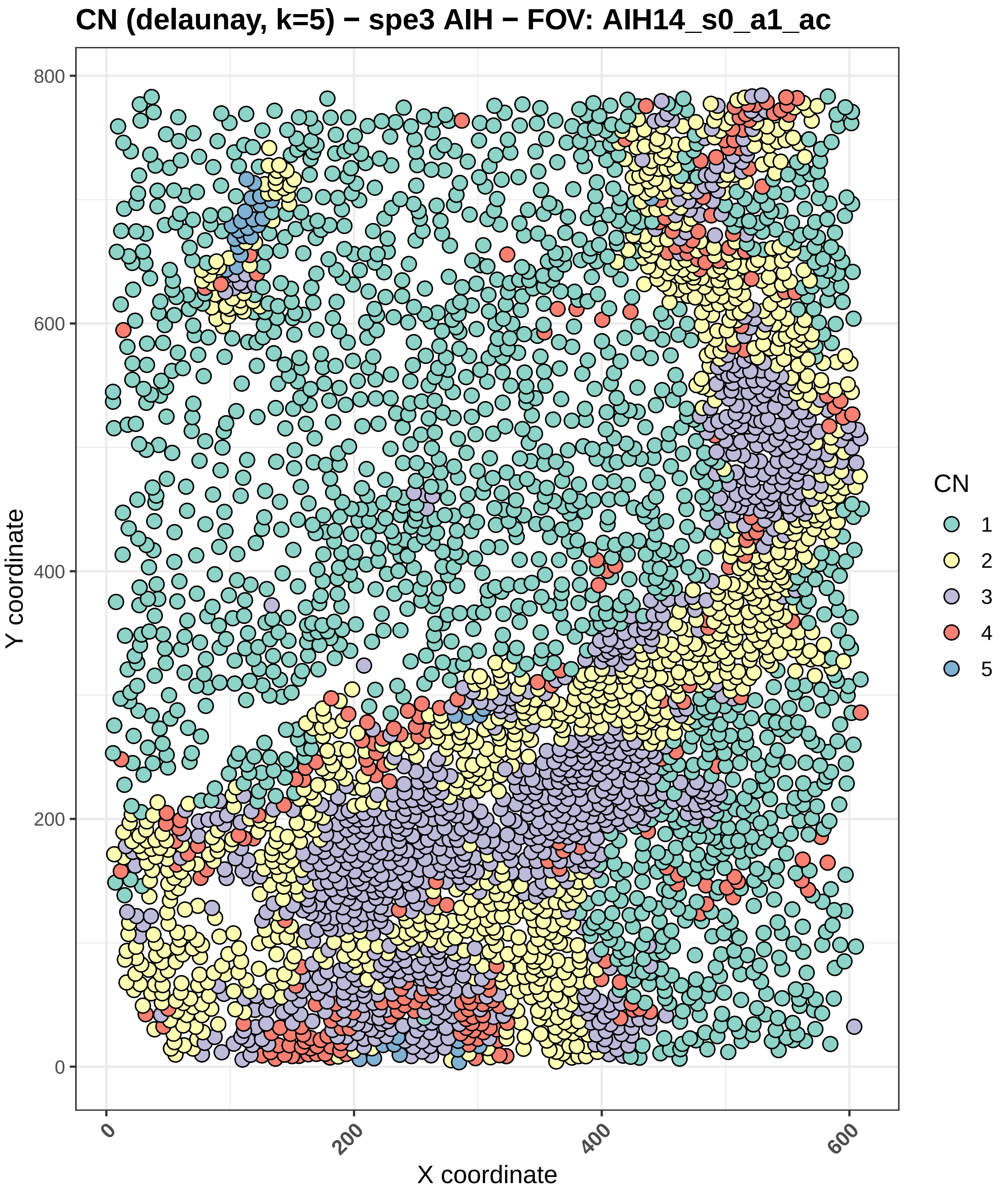
CN (delaunay, k=5) − spe3 AIH − FOV: AIH14_s0_a1_ac
Y coordinate
X coordinate
0
200
400
600
800
CN
1
2
3
4
5

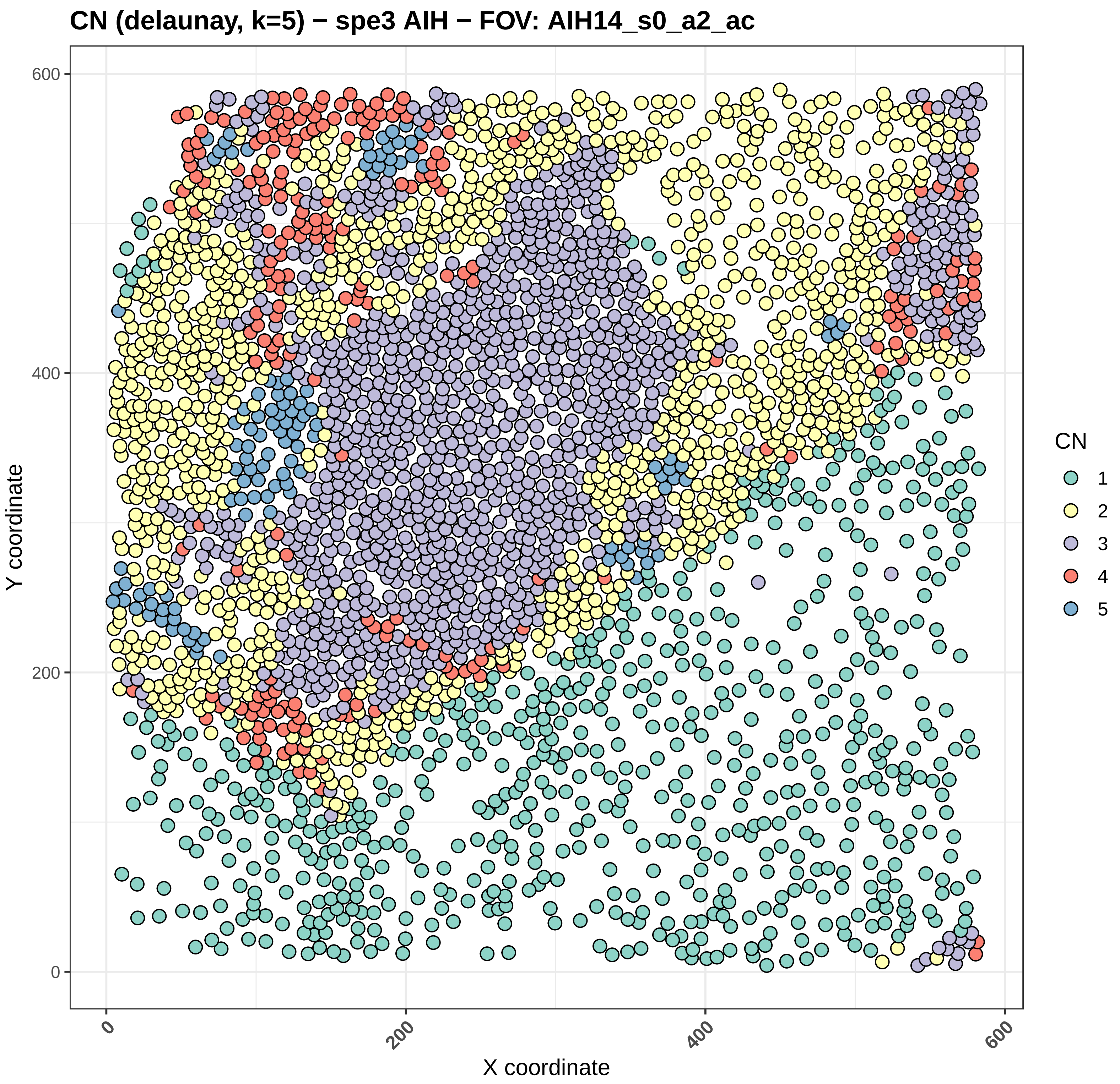
CN (delaunay, k=5) − spe3 AIH − FOV: AIH14_s0_a2_ac
Y coordinate
X coordinate
0
200
400
600
CN
1
2
3
4
5

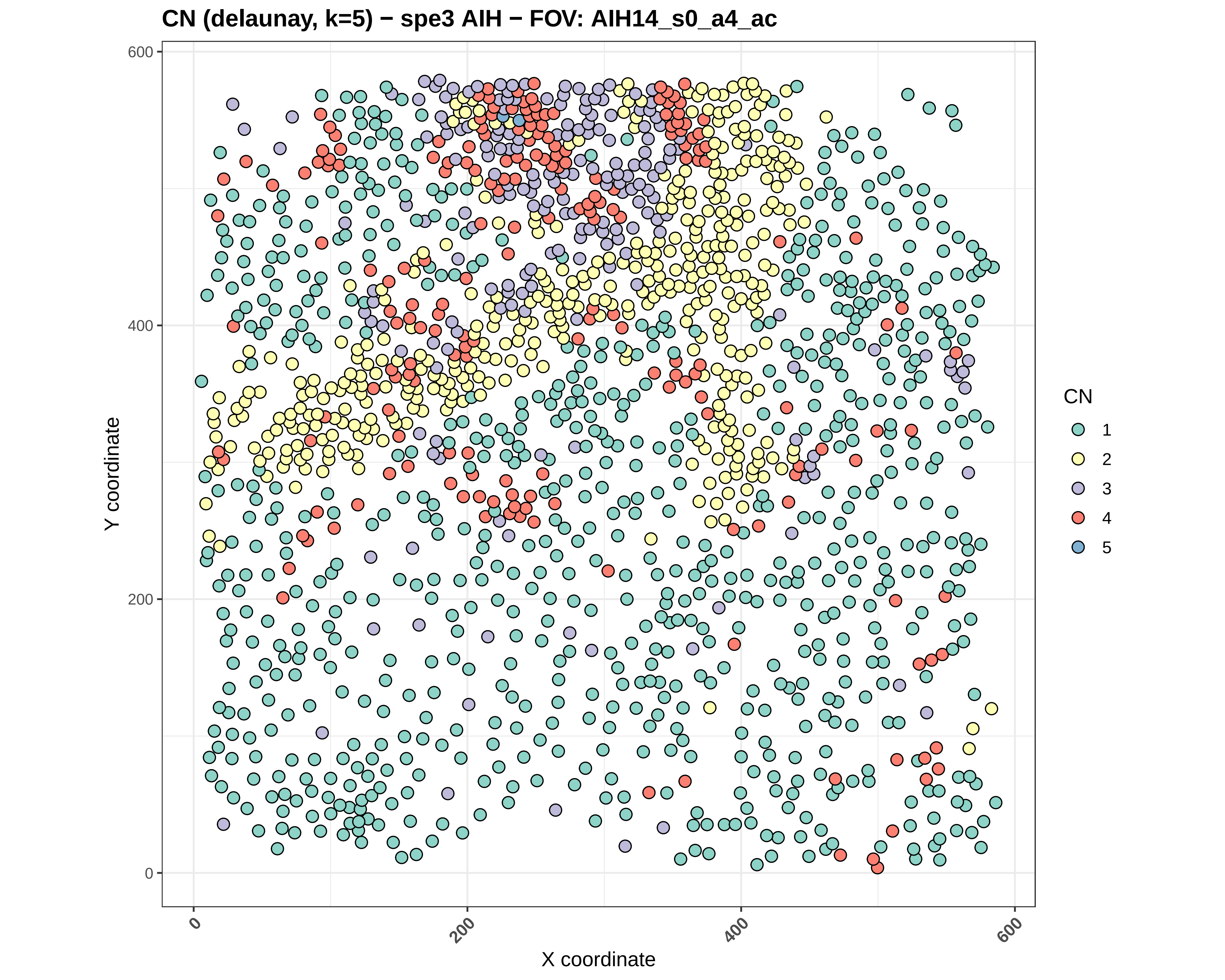
CN (delaunay, k=5) − spe3 AIH − FOV: AIH14_s0_a4_ac
Y coordinate
X coordinate
0
200
400
600
CN
1
2
3
4
5

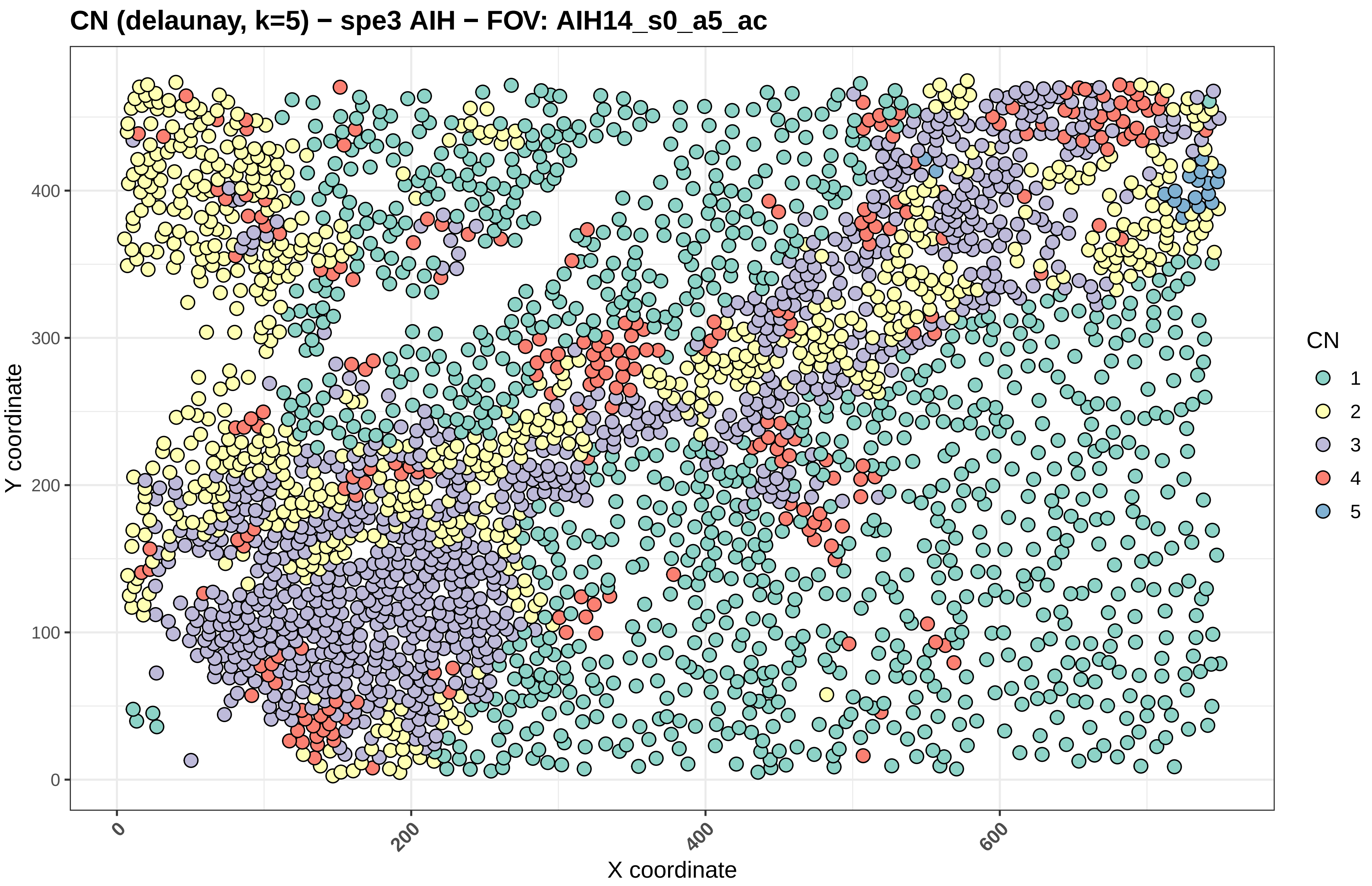
CN (delaunay, k=5) − spe3 AIH − FOV: AIH14_s0_a5_ac
Y coordinate
X coordinate
0
100
200
300
400
0
200
400
600
CN
1
2
3
4
5

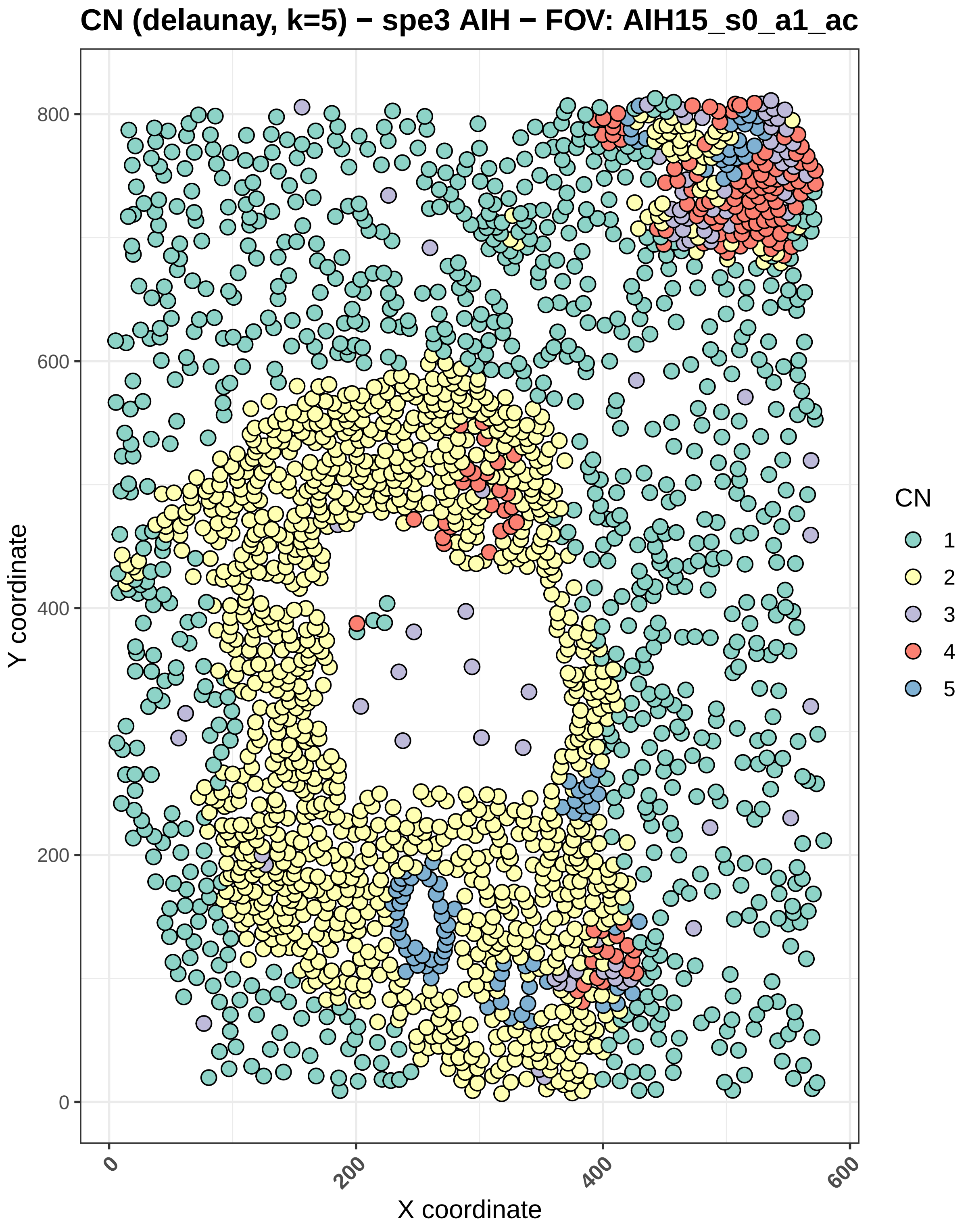

CN (delaunay, k=5) − spe3 AIH − FOV: AIH15_s0_a1_ac
Y coordinate
X coordinate
0
200
400
600
800
CN
1
2
3
4
5

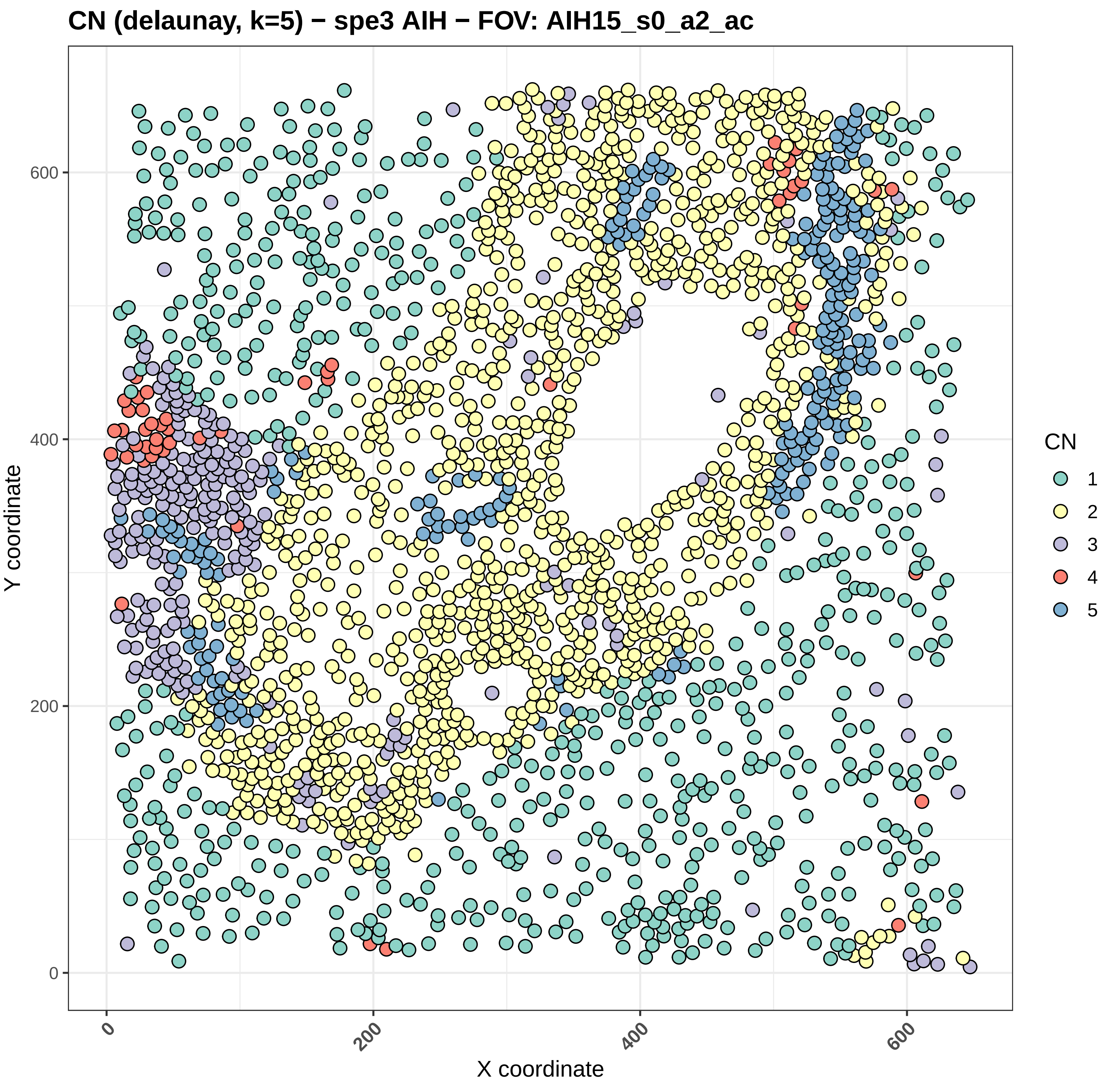

CN (delaunay, k=5) − spe3 AIH − FOV: AIH15_s0_a2_ac
Y coordinate
X coordinate
0
200
400
600
CN
1
2
3
4
5

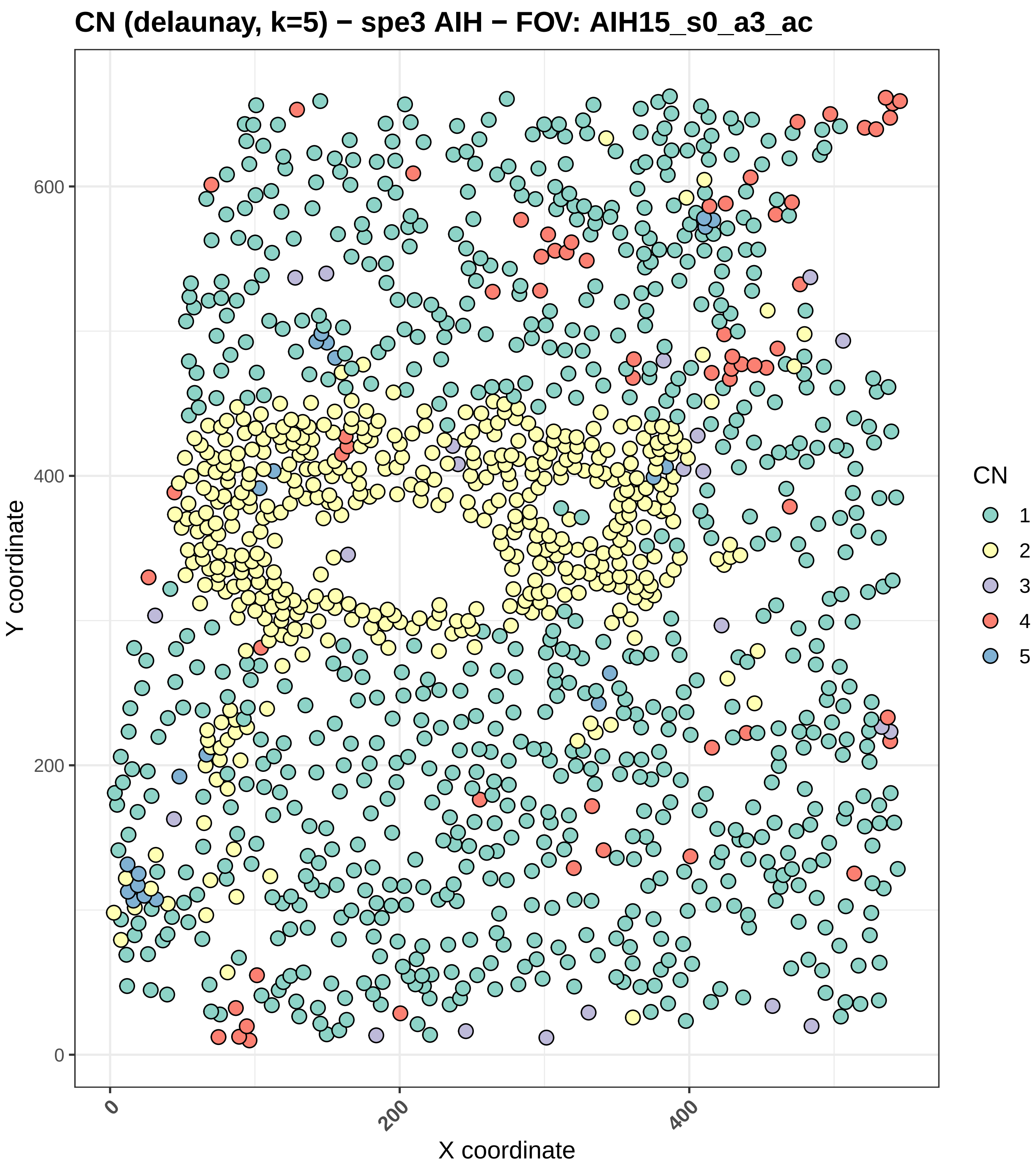

CN (delaunay, k=5) − spe3 AIH − FOV: AIH15_s0_a3_ac
Y coordinate
X coordinate
0
200
400
600
0
200
400
CN
1
2
3
4
5

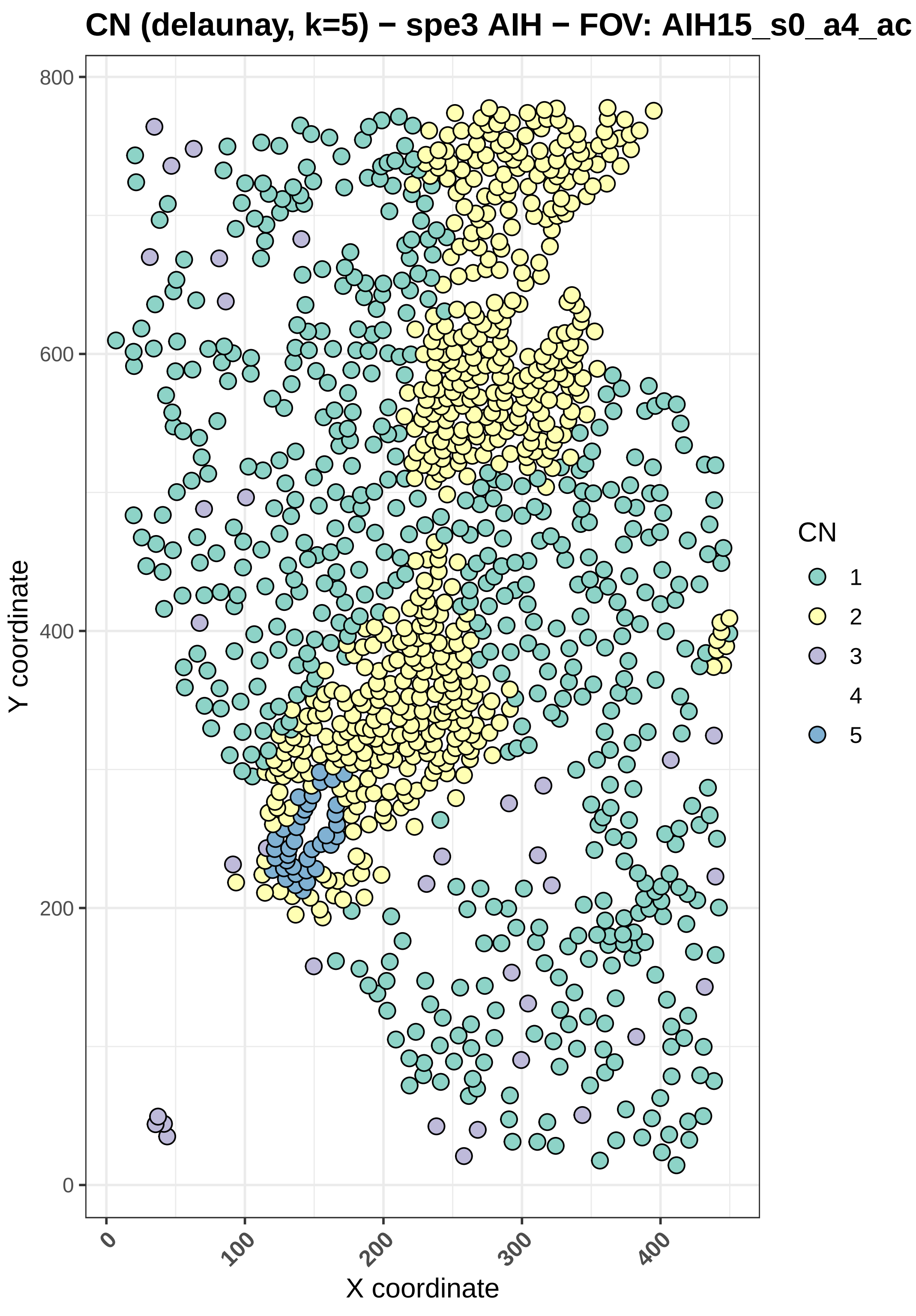
CN (delaunay, k=5) − spe3 AIH − FOV: AIH15_s0_a4_ac
Y coordinate
X coordinate
0
200
400
600
800
0
100
200
300
400
CN
1
2
3
4
5

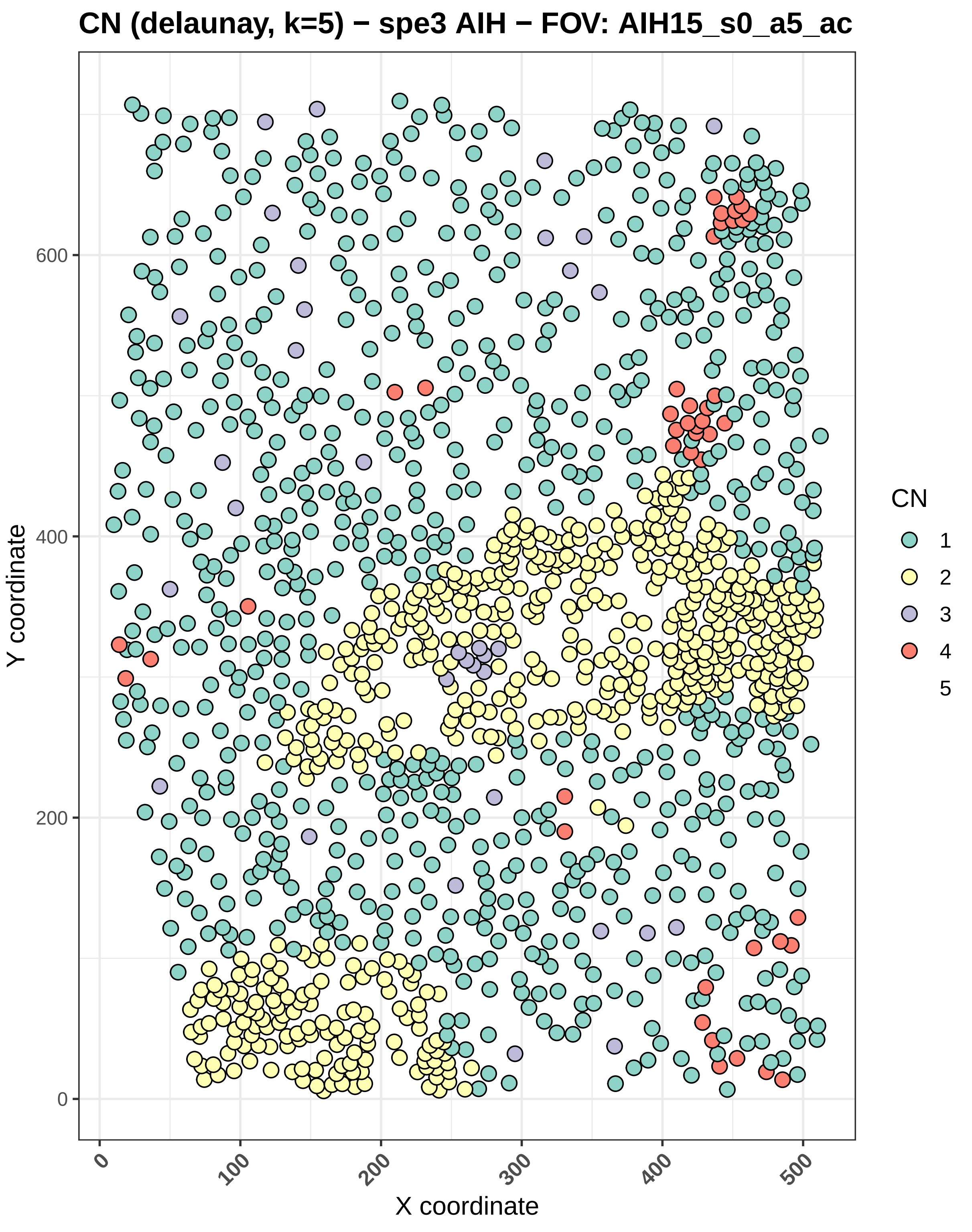

CN (delaunay, k=5) − spe3 AIH − FOV: AIH15_s0_a5_ac
Y coordinate
X coordinate
0
200
400
600
0
100
200
300
400
500
CN
1
2
3
4
5

#### CN (delaunay, k=5) − spe3 AIH − FOV: AIH16_s0_a1_ac

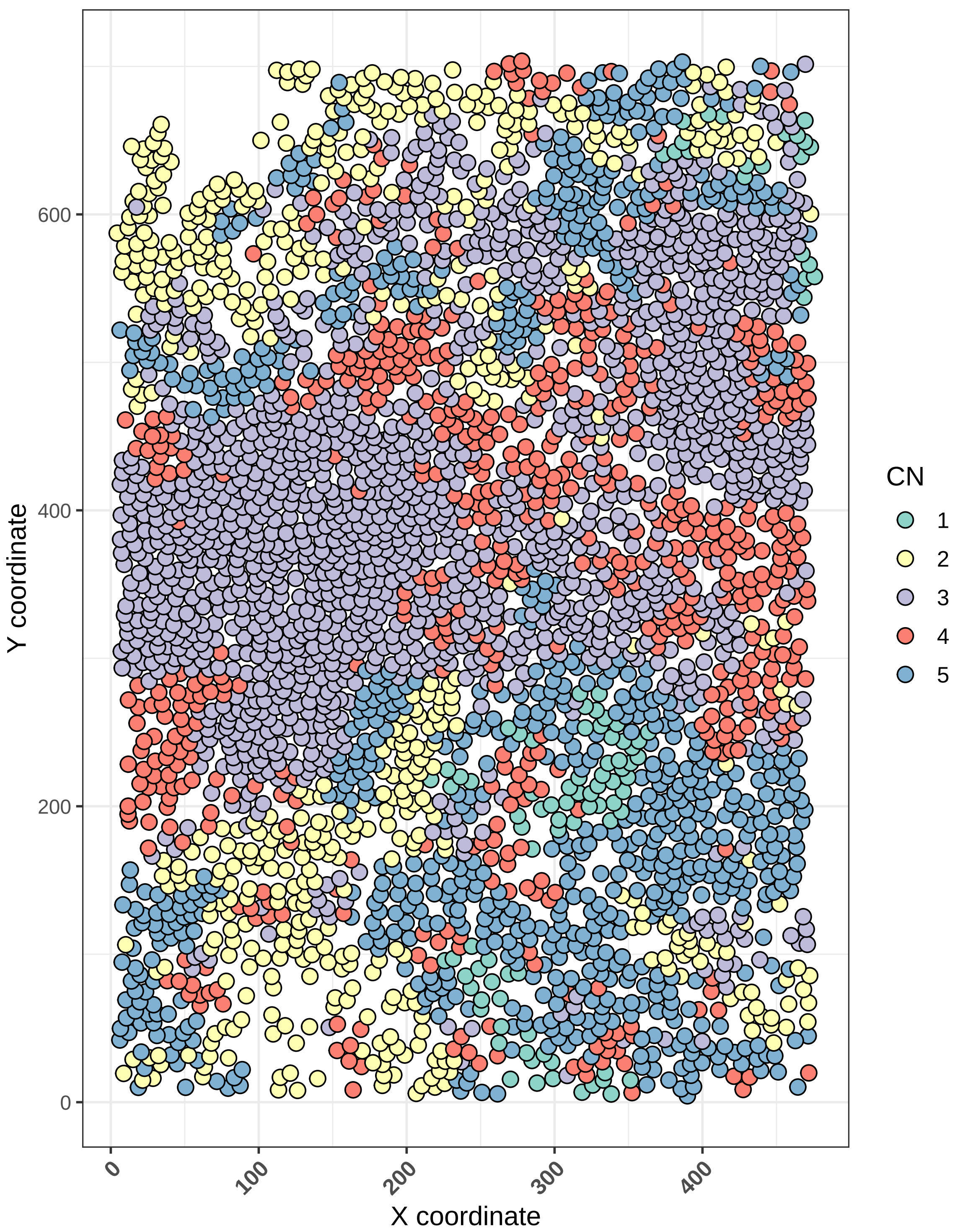

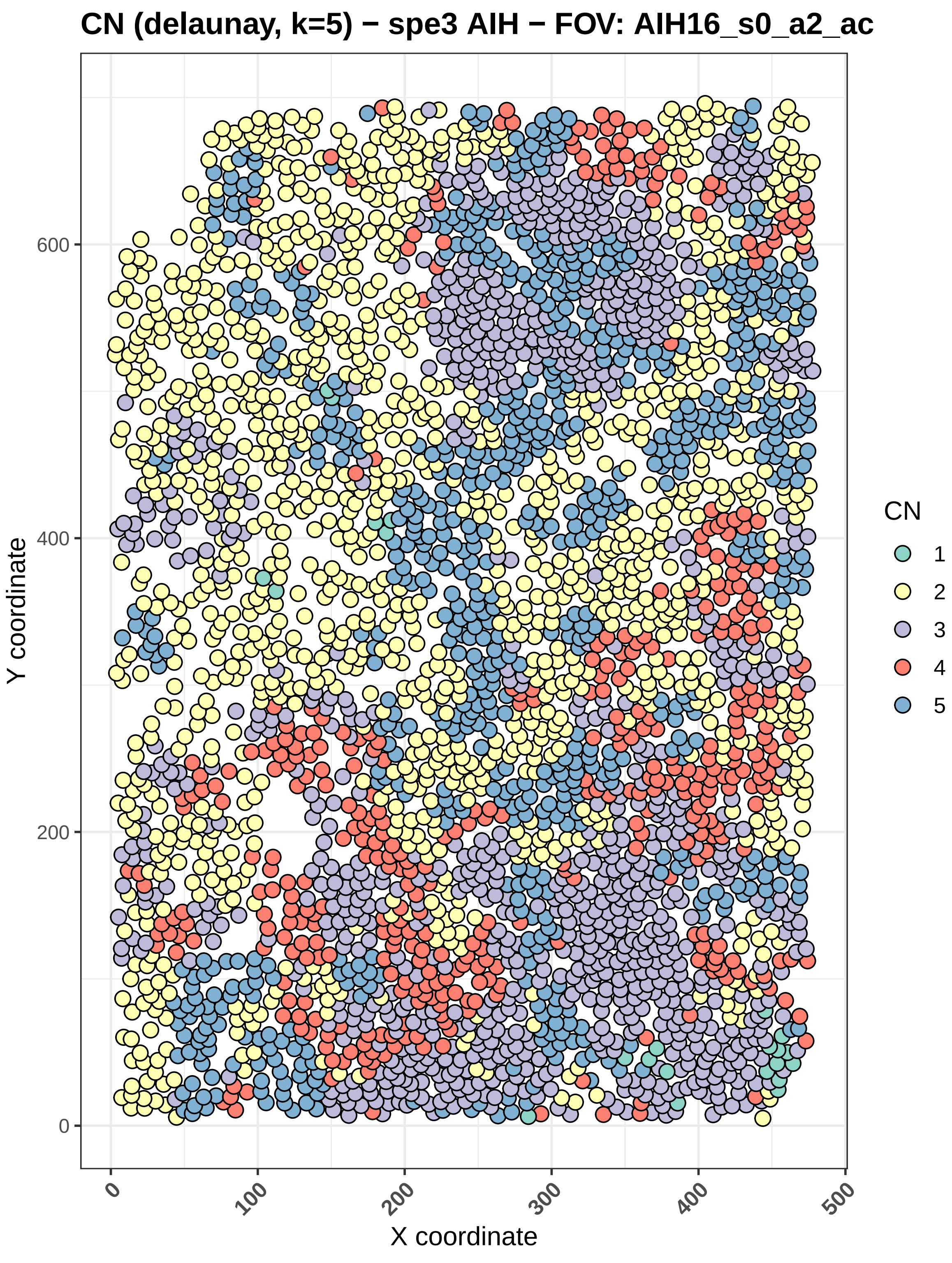

CN (delaunay, k=5) − spe3 AIH − FOV: AIH16_s0_a2_ac
Y coordinate
X coordinate
0
200
400
600
0
100
200
300
400
500
CN
1
2
3
4
5

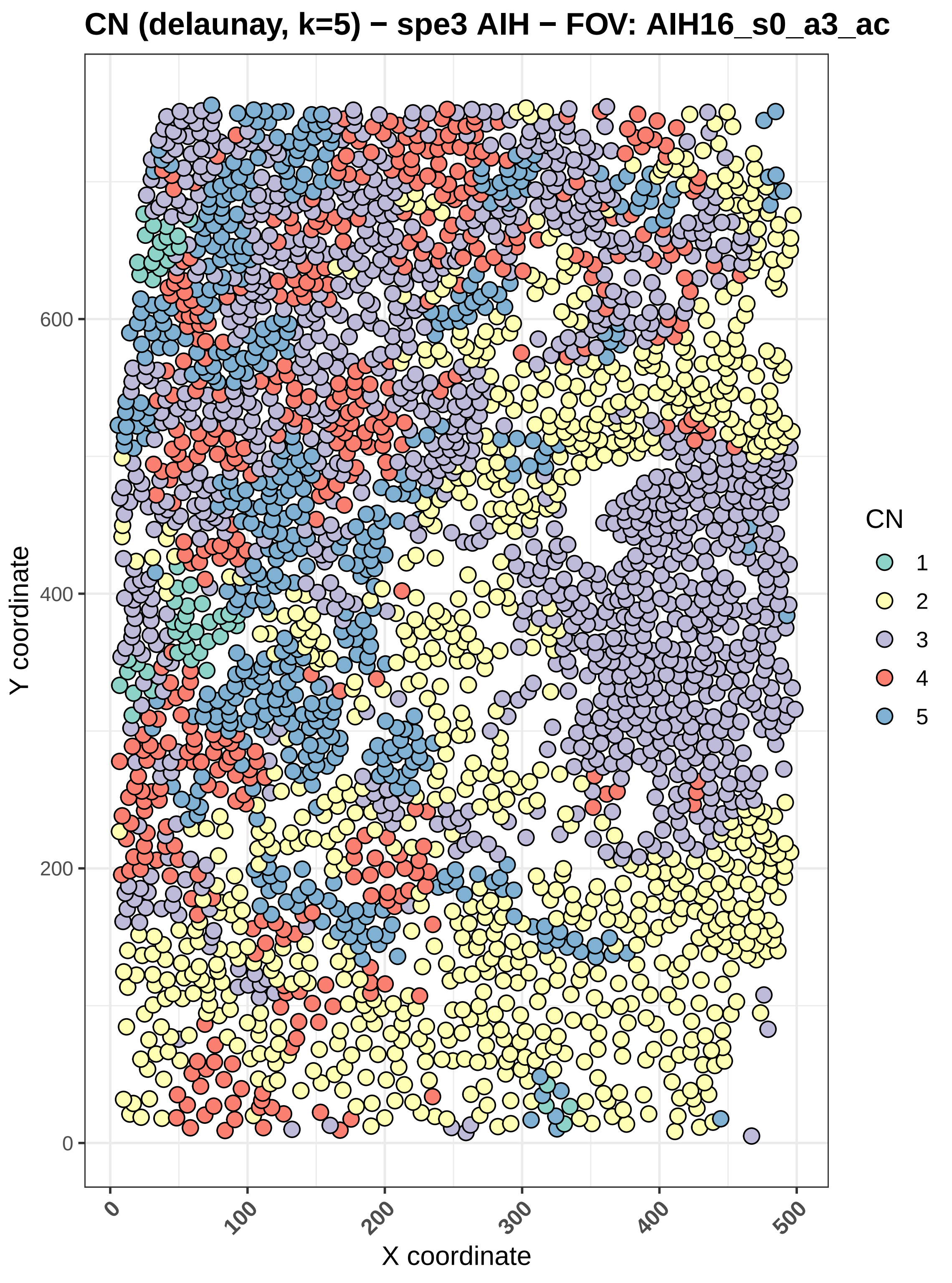

CN (delaunay, k=5) − spe3 AIH − FOV: AIH16_s0_a3_ac
Y coordinate
X coordinate
0
200
400
600
0
100
200
300
400
500
CN
1
2
3
4
5

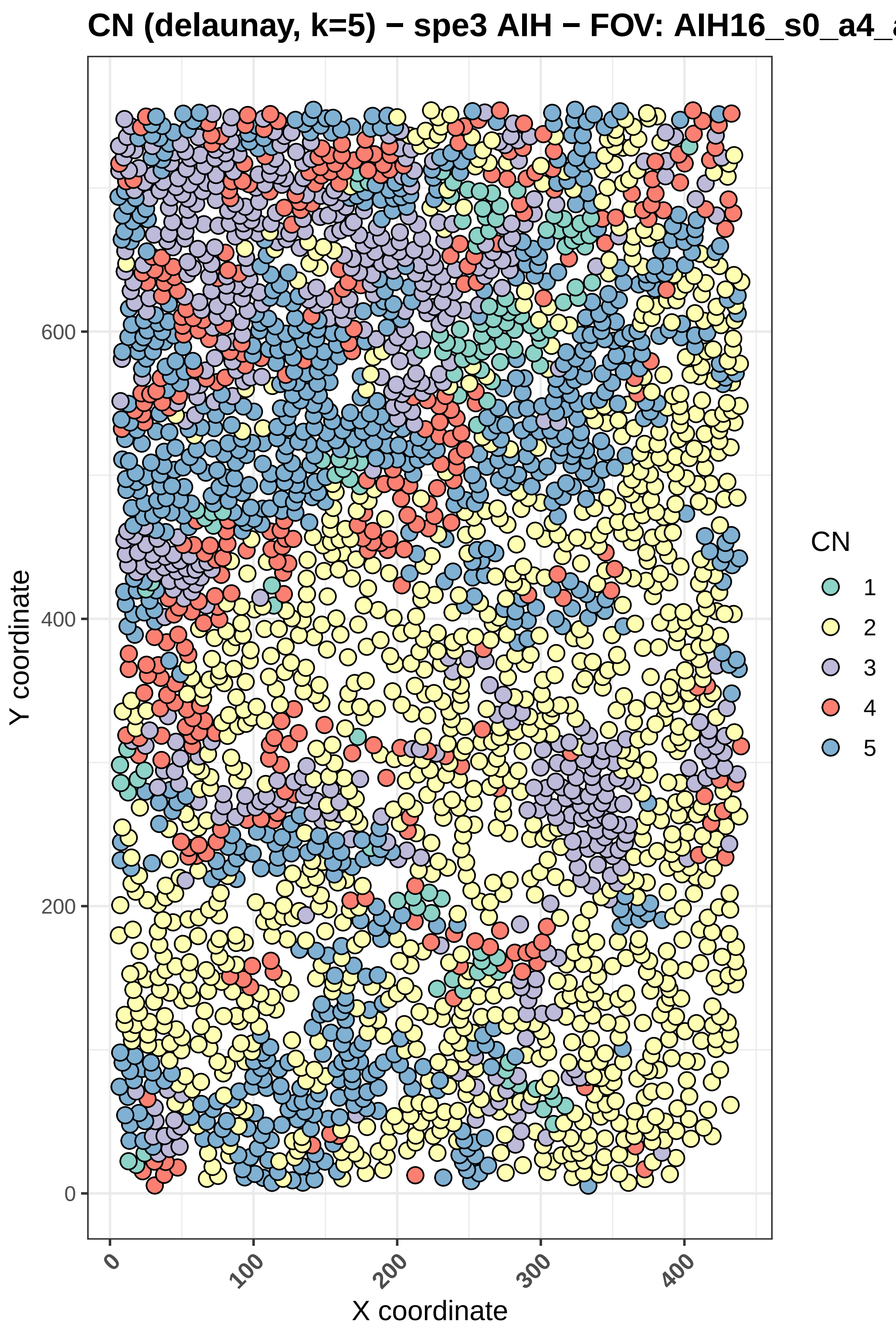

CN (delaunay, k=5) − spe3 AIH − FOV: AIH16_s0_a4_ac
Y coordinate
X coordinate
0
200
400
600
0
100
200
300
400
CN
1
2
3
4
5

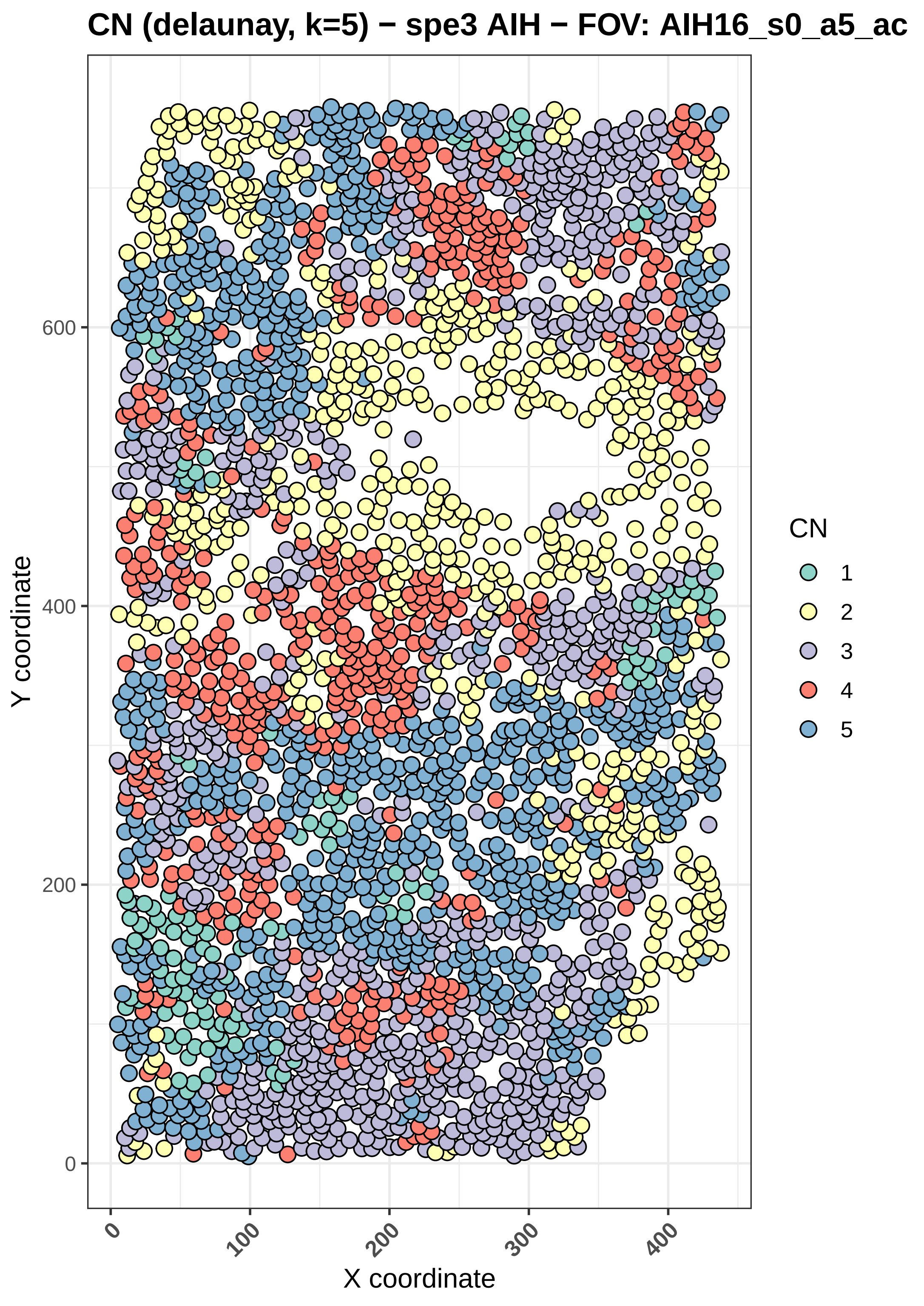
CN (delaunay, k=5) − spe3 AIH − FOV: AIH16_s0_a5_ac
Y coordinate
X coordinate
0
200
400
600
0
100
200
300
400
CN
1
2
3
4
5

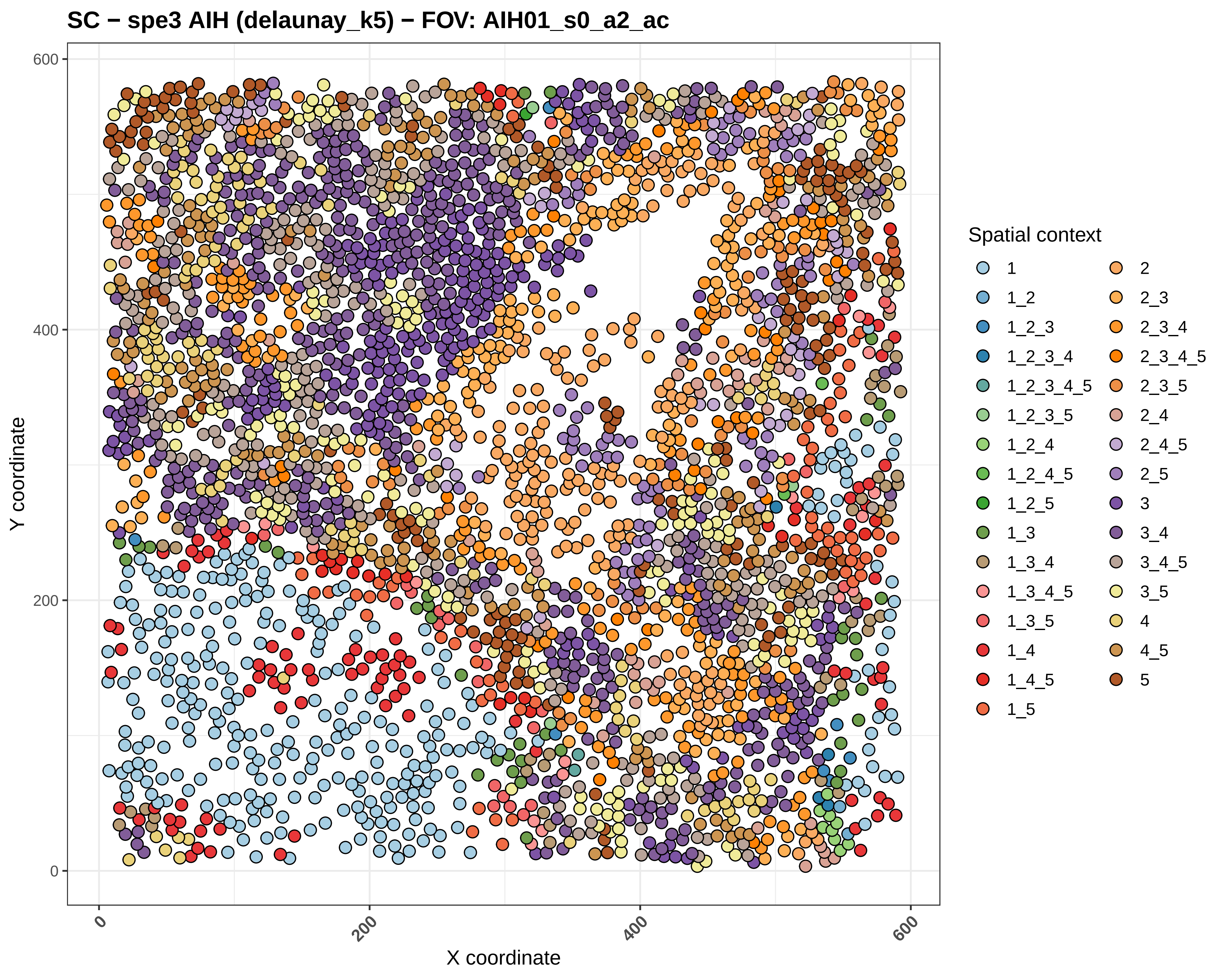
SC − spe3 AIH (delaunay_k5) − FOV: AIH01_s0_a2_ac
Y coordinate
X coordinate
0
200
400
600
Spatial context
1
1_2
1_2_3
1_2_3_4
1_2_3_4_5
1_2_3_5
1_2_4
1_2_4_5
1_2_5
1_3
1_3_4
1_3_4_5
1_3_5
1_4
1_4_5
1_5
2
2_3
2_3_4
2_3_4_5
2_3_5
2_4
2_4_5
2_5
3
3_4
3_4_5
3_5
4
4_5
5

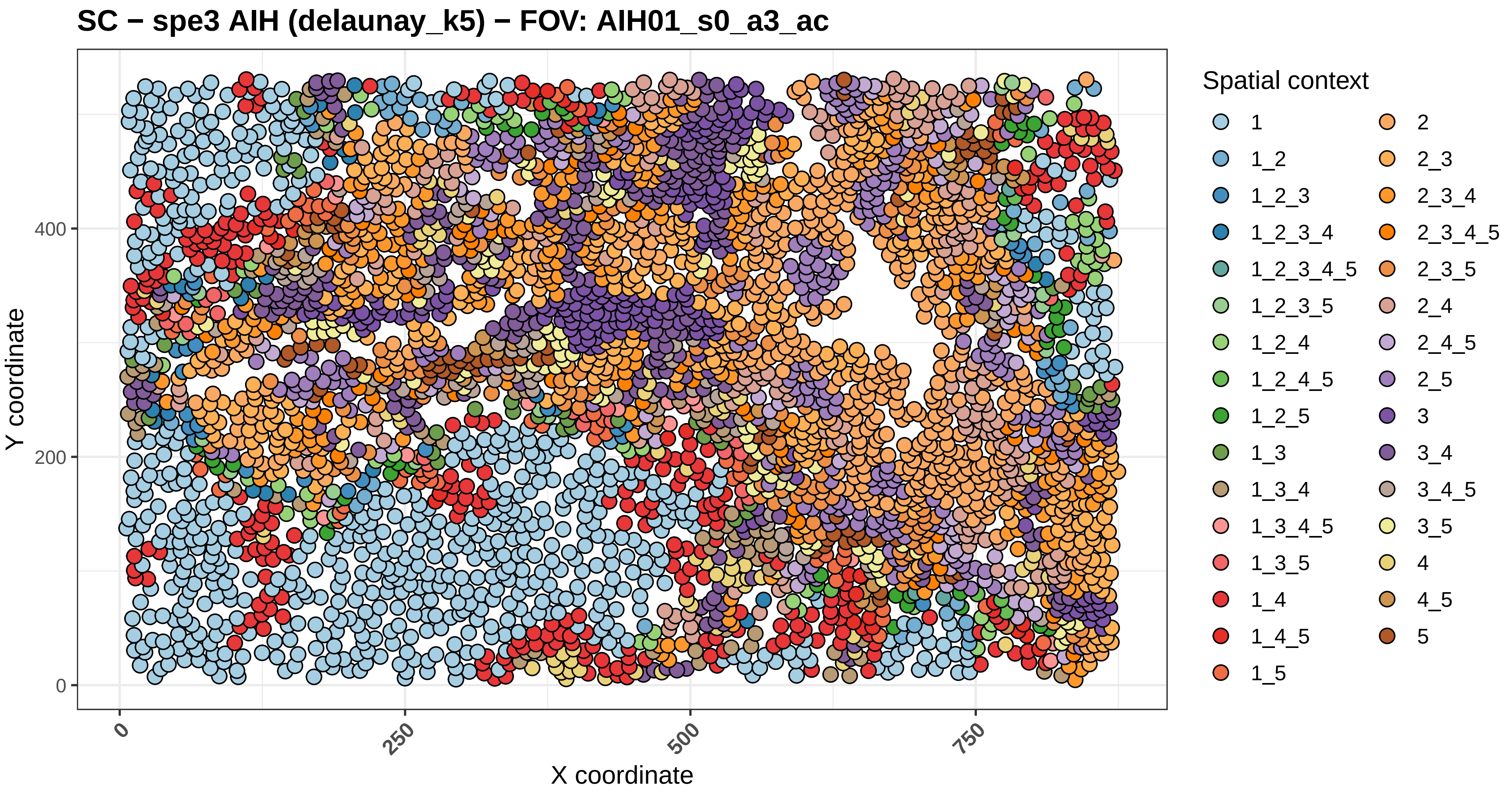
SC − spe3 AIH (delaunay_k5) − FOV: AIH01_s0_a3_ac
Y coordinate
X coordinate
0
200
400
0
250
500
750
Spatial context
1
1_2
1_2_3
1_2_3_4
1_2_3_4_5
1_2_3_5
1_2_4
1_2_4_5
1_2_5
1_3
1_3_4
1_3_4_5
1_3_5
1_4
1_4_5
1_5
2
2_3
2_3_4
2_3_4_5
2_3_5
2_4
2_4_5
2_5
3
3_4
3_4_5
3_5
4
4_5
5

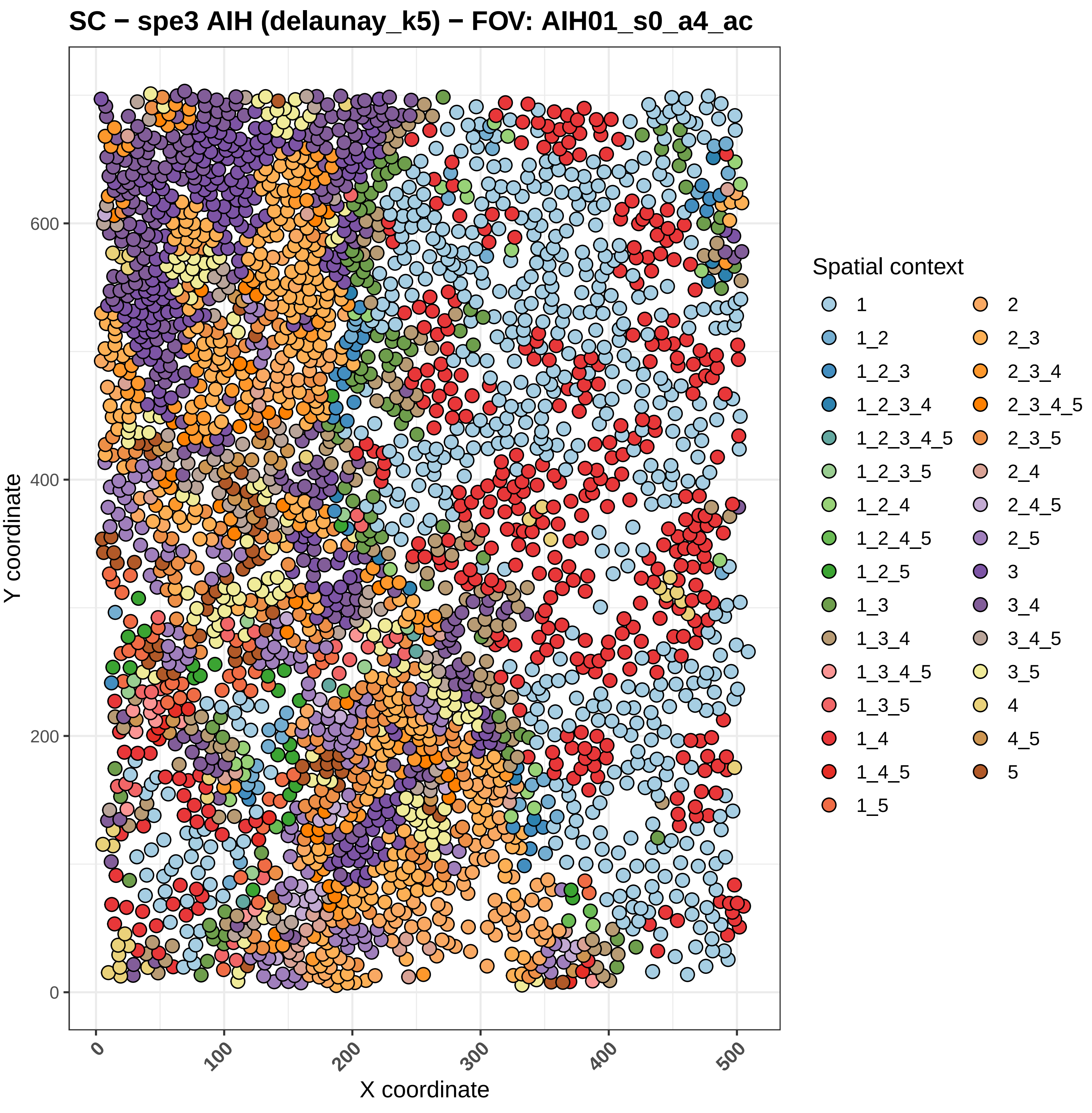

SC − spe3 AIH (delaunay_k5) − FOV: AIH01_s0_a4_ac
Y coordinate
X coordinate
0
200
400
600
0
100
200
300
400
500
Spatial context
1
1_2
1_2_3
1_2_3_4
1_2_3_4_5
1_2_3_5
1_2_4
1_2_4_5
1_2_5
1_3
1_3_4
1_3_4_5
1_3_5
1_4
1_4_5
1_5
2
2_3
2_3_4
2_3_4_5
2_3_5
2_4
2_4_5
2_5
3
3_4
3_4_5
3_5
4
4_5
5

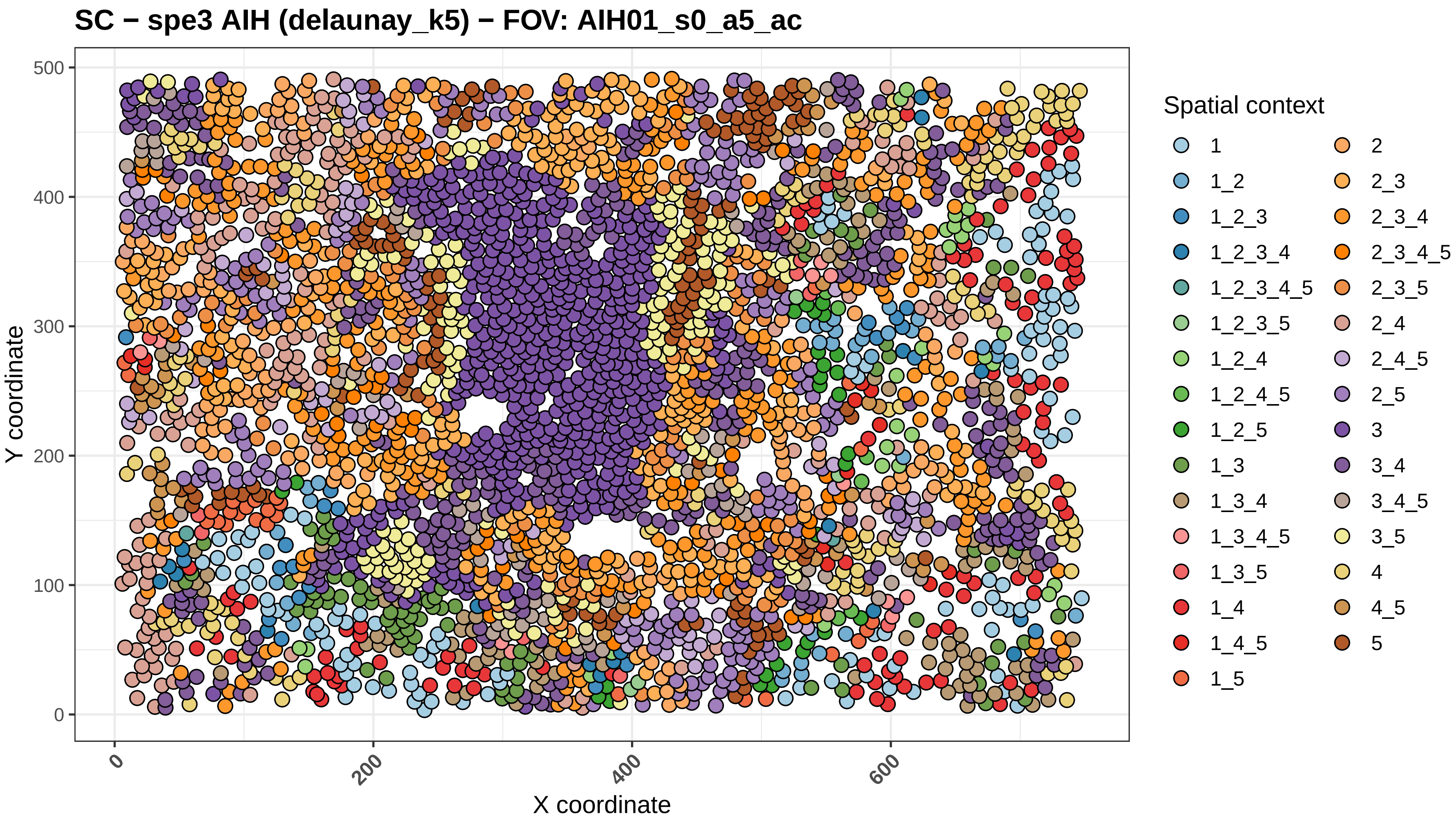
SC − spe3 AIH (delaunay_k5) − FOV: AIH01_s0_a5_ac
Y coordinate
X coordinate
0
100
200
300
400
500
0
200
400
600
Spatial context
1
1_2
1_2_3
1_2_3_4
1_2_3_4_5
1_2_3_5
1_2_4
1_2_4_5
1_2_5
1_3
1_3_4
1_3_4_5
1_3_5
1_4
1_4_5
1_5
2
2_3
2_3_4
2_3_4_5
2_3_5
2_4
2_4_5
2_5
3
3_4
3_4_5
3_5
4
4_5
5

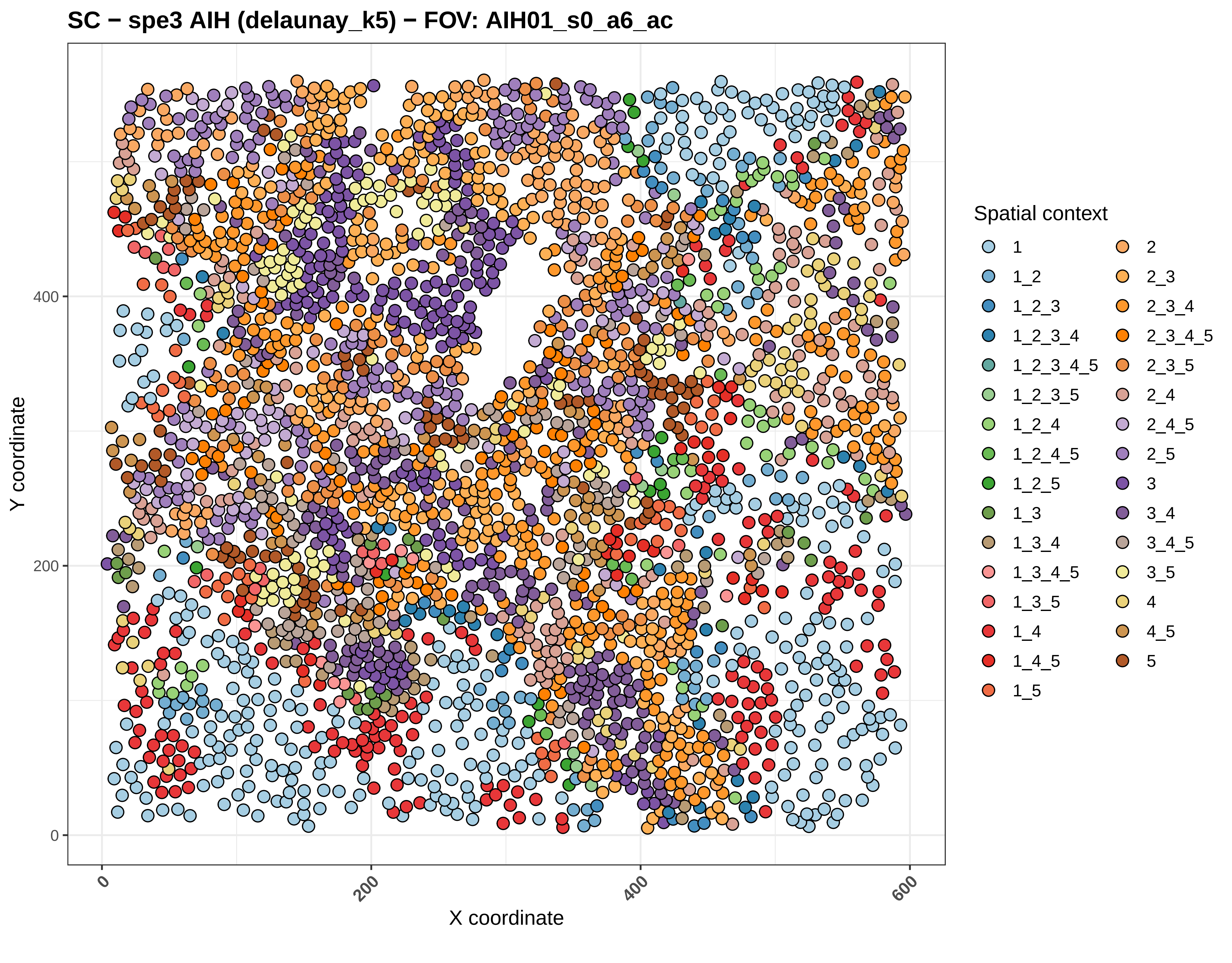
SC − spe3 AIH (delaunay_k5) − FOV: AIH01_s0_a6_ac
Y coordinate
X coordinate
0
200
400
600
Spatial context
1
1_2
1_2_3
1_2_3_4
1_2_3_4_5
1_2_3_5
1_2_4
1_2_4_5
1_2_5
1_3
1_3_4
1_3_4_5
1_3_5
1_4
1_4_5
1_5
2
2_3
2_3_4
2_3_4_5
2_3_5
2_4
2_4_5
2_5
3
3_4
3_4_5
3_5
4
4_5
5

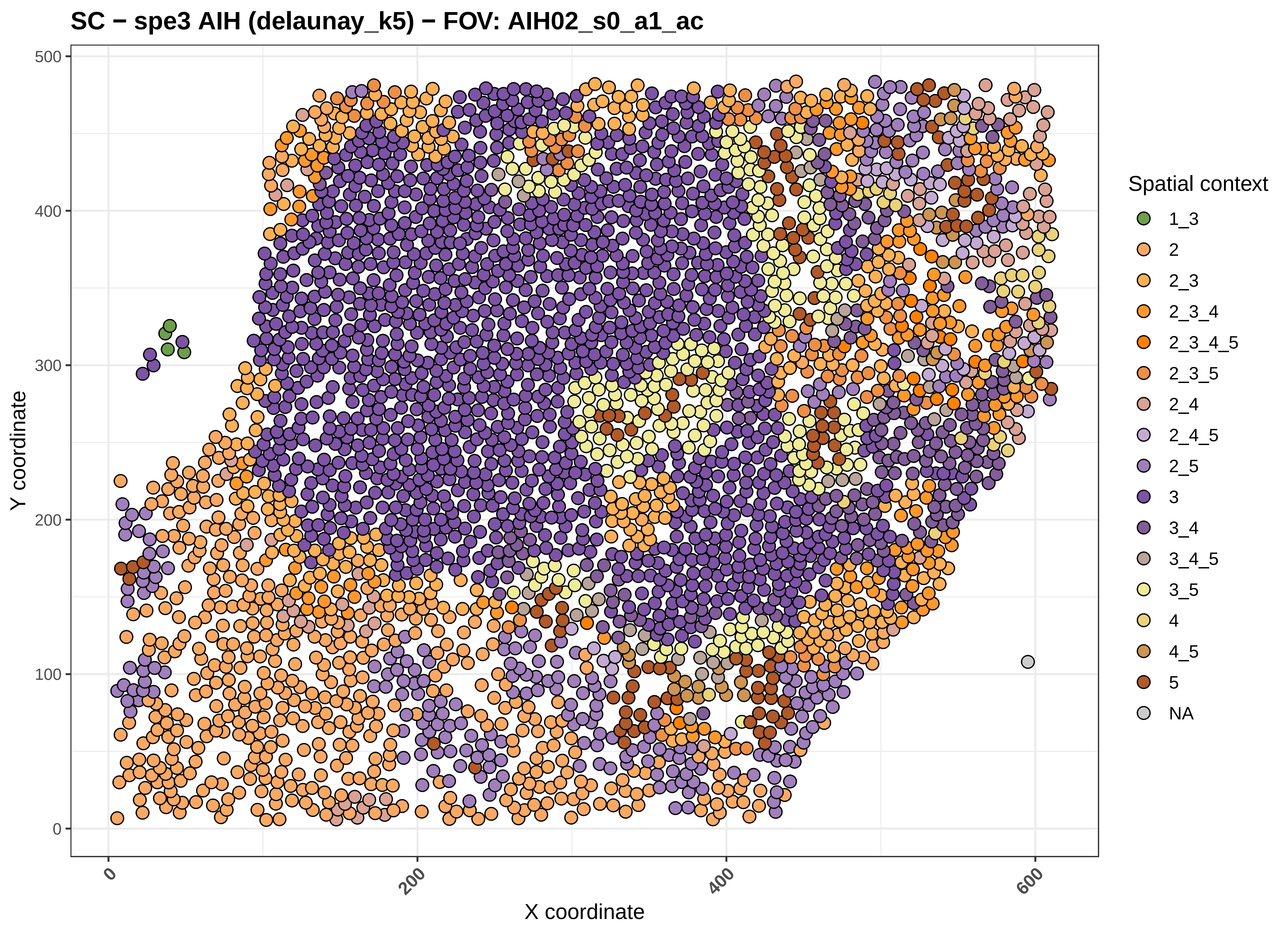

SC − spe3 AIH (delaunay_k5) − FOV: AIH02_s0_a1_ac
Y coordinate
X coordinate
0
100
200
300
400
500
0
200
400
600
Spatial context
1_3
2
2_3
2_3_4
2_3_4_5
2_3_5
2_4
2_4_5
2_5
3
3_4
3_4_5
3_5
4
4_5
5
NA

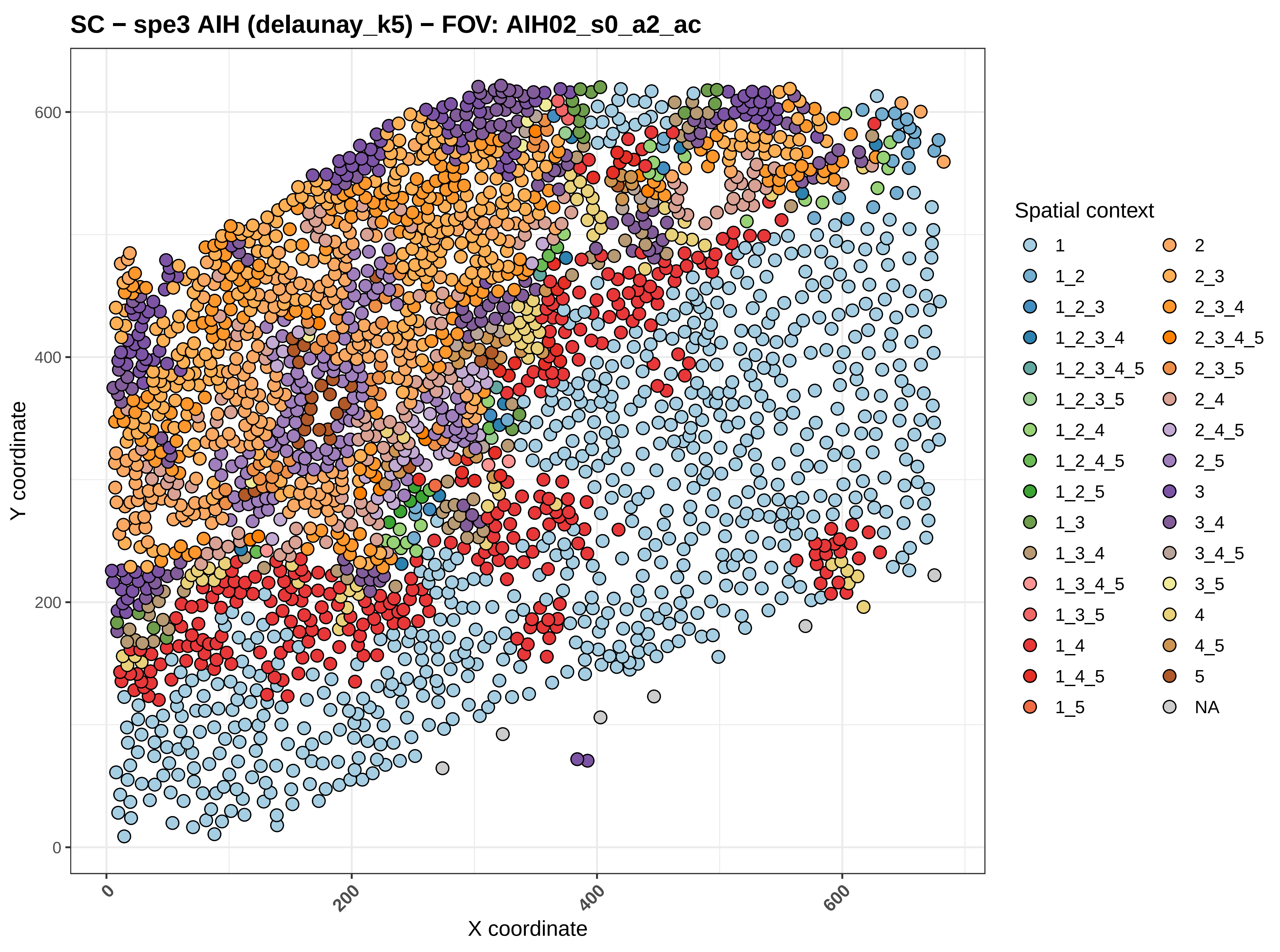

SC − spe3 AIH (delaunay_k5) − FOV: AIH02_s0_a2_ac
Y coordinate
X coordinate
0
200
400
600
Spatial context
1
1_2
1_2_3
1_2_3_4
1_2_3_4_5
1_2_3_5
1_2_4
1_2_4_5
1_2_5
1_3
1_3_4
1_3_4_5
1_3_5
1_4
1_4_5
1_5
2
2_3
2_3_4
2_3_4_5
2_3_5
2_4
2_4_5
2_5
3
3_4
3_4_5
3_5
4
4_5
5
NA

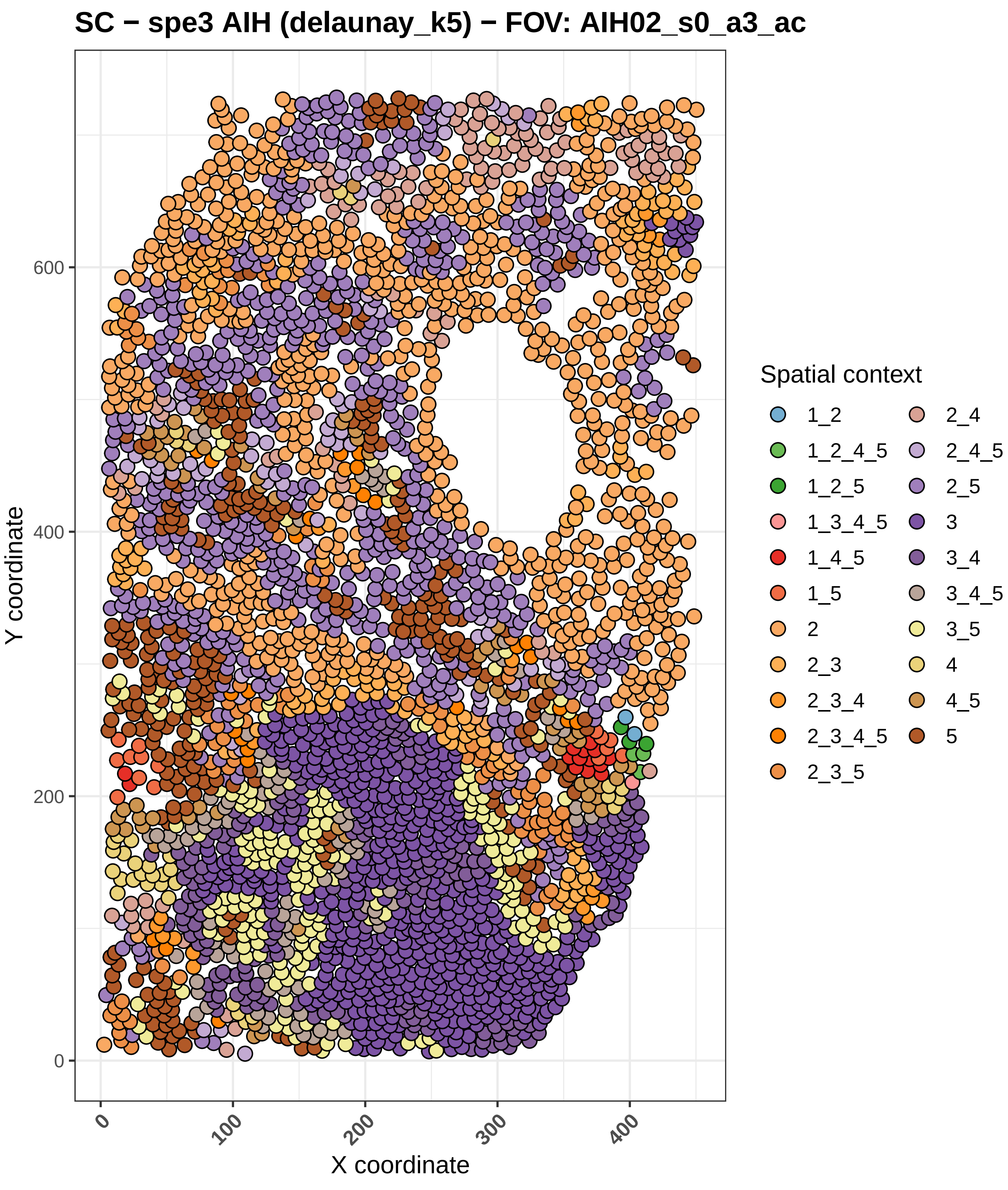
SC − spe3 AIH (delaunay_k5) − FOV: AIH02_s0_a3_ac
Y coordinate
X coordinate
0
200
400
600
0
100
200
300
400
Spatial context
1_2
1_2_4_5
1_2_5
1_3_4_5
1_4_5
1_5
2
2_3
2_3_4
2_3_4_5
2_3_5
2_4
2_4_5
2_5
3
3_4
3_4_5
3_5
4
4_5
5

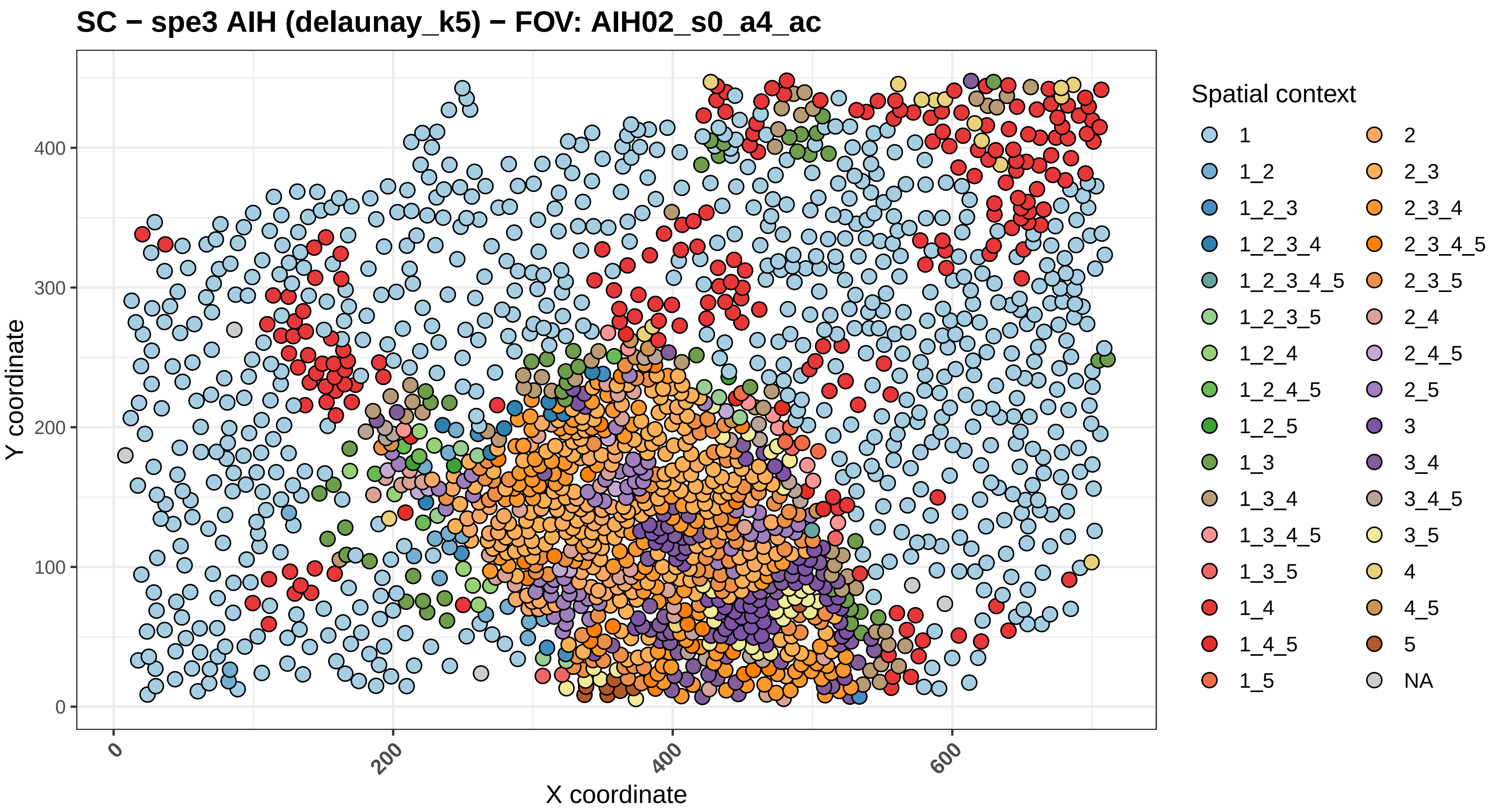

SC − spe3 AIH (delaunay_k5) − FOV: AIH02_s0_a4_ac
Y coordinate
X coordinate
0
100
200
300
400
0
200
400
600
Spatial context
1
1_2
1_2_3
1_2_3_4
1_2_3_4_5
1_2_3_5
1_2_4
1_2_4_5
1_2_5
1_3
1_3_4
1_3_4_5
1_3_5
1_4
1_4_5
1_5
2
2_3
2_3_4
2_3_4_5
2_3_5
2_4
2_4_5
2_5
3
3_4
3_4_5
3_5
4
4_5
5
NA

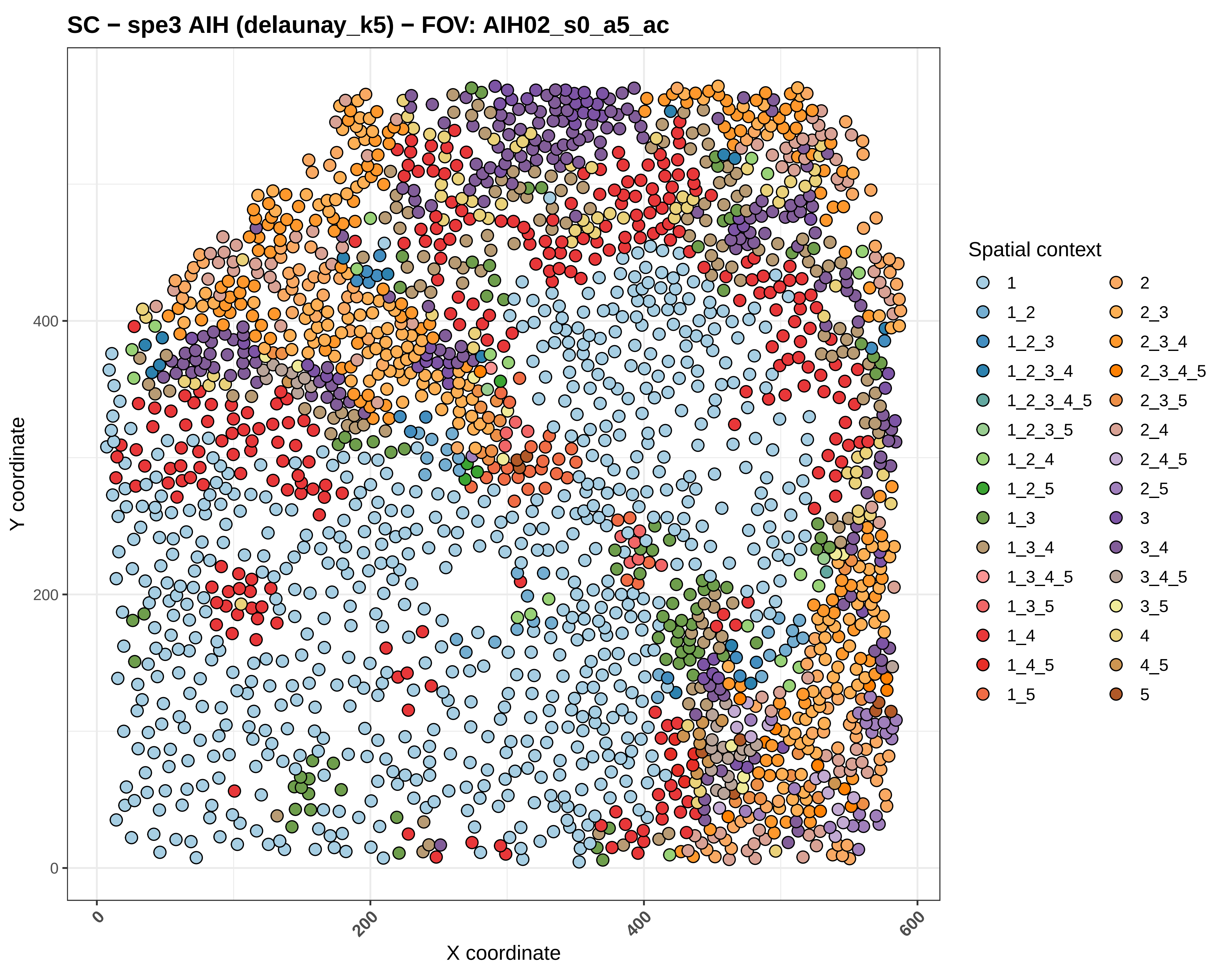
SC − spe3 AIH (delaunay_k5) − FOV: AIH02_s0_a5_ac
Y coordinate
X coordinate
0
200
400
600
Spatial context
1
1_2
1_2_3
1_2_3_4
1_2_3_4_5
1_2_3_5
1_2_4
1_2_5
1_3
1_3_4
1_3_4_5
1_3_5
1_4
1_4_5
1_5
2
2_3
2_3_4
2_3_4_5
2_3_5
2_4
2_4_5
2_5
3
3_4
3_4_5
3_5
4
4_5
5

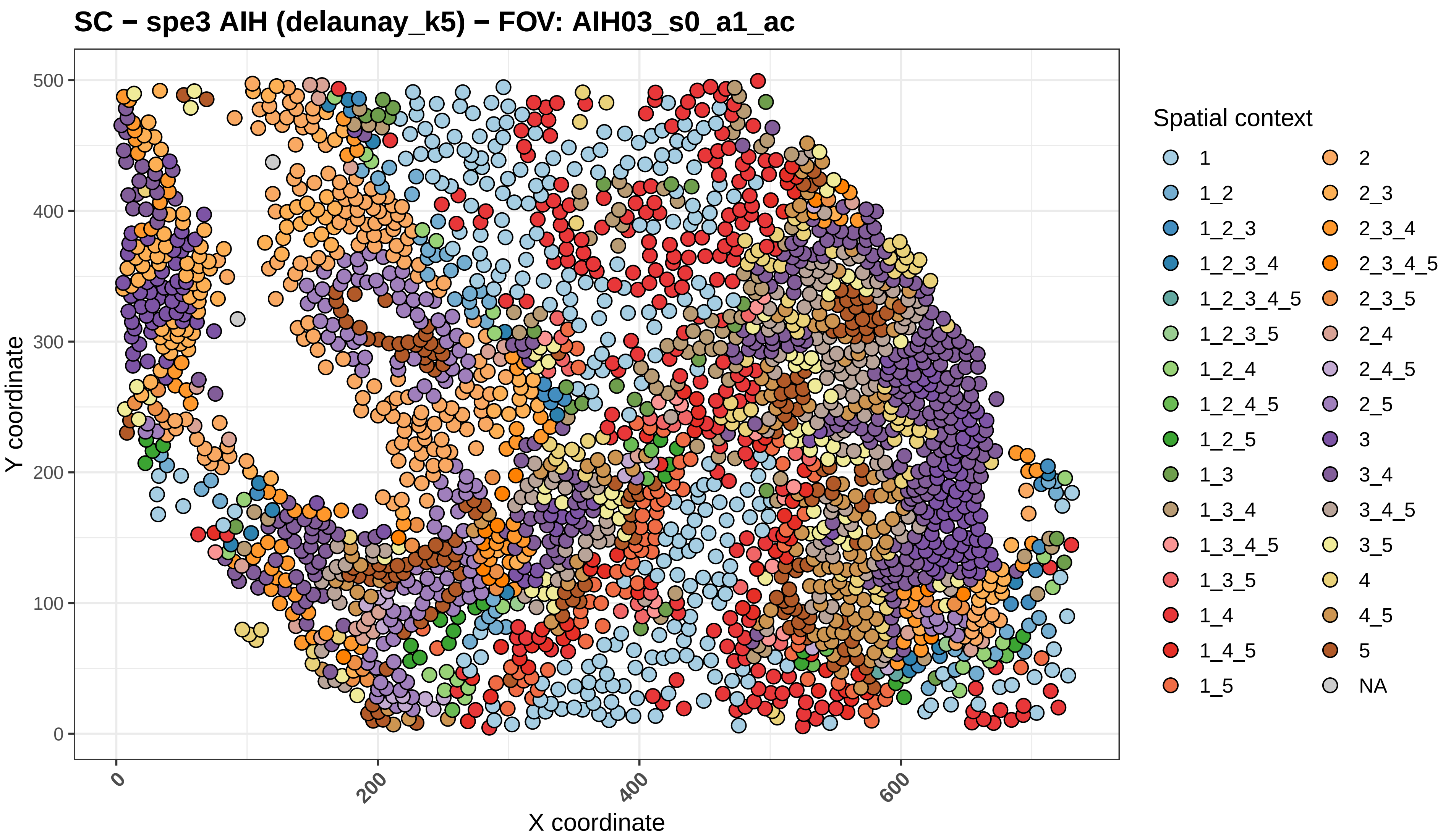
SC − spe3 AIH (delaunay_k5) − FOV: AIH03_s0_a1_ac
Y coordinate
X coordinate
0
100
200
300
400
500
0
200
400
600
Spatial context
1
1_2
1_2_3
1_2_3_4
1_2_3_4_5
1_2_3_5
1_2_4
1_2_4_5
1_2_5
1_3
1_3_4
1_3_4_5
1_3_5
1_4
1_4_5
1_5
2
2_3
2_3_4
2_3_4_5
2_3_5
2_4
2_4_5
2_5
3
3_4
3_4_5
3_5
4
4_5
5
NA

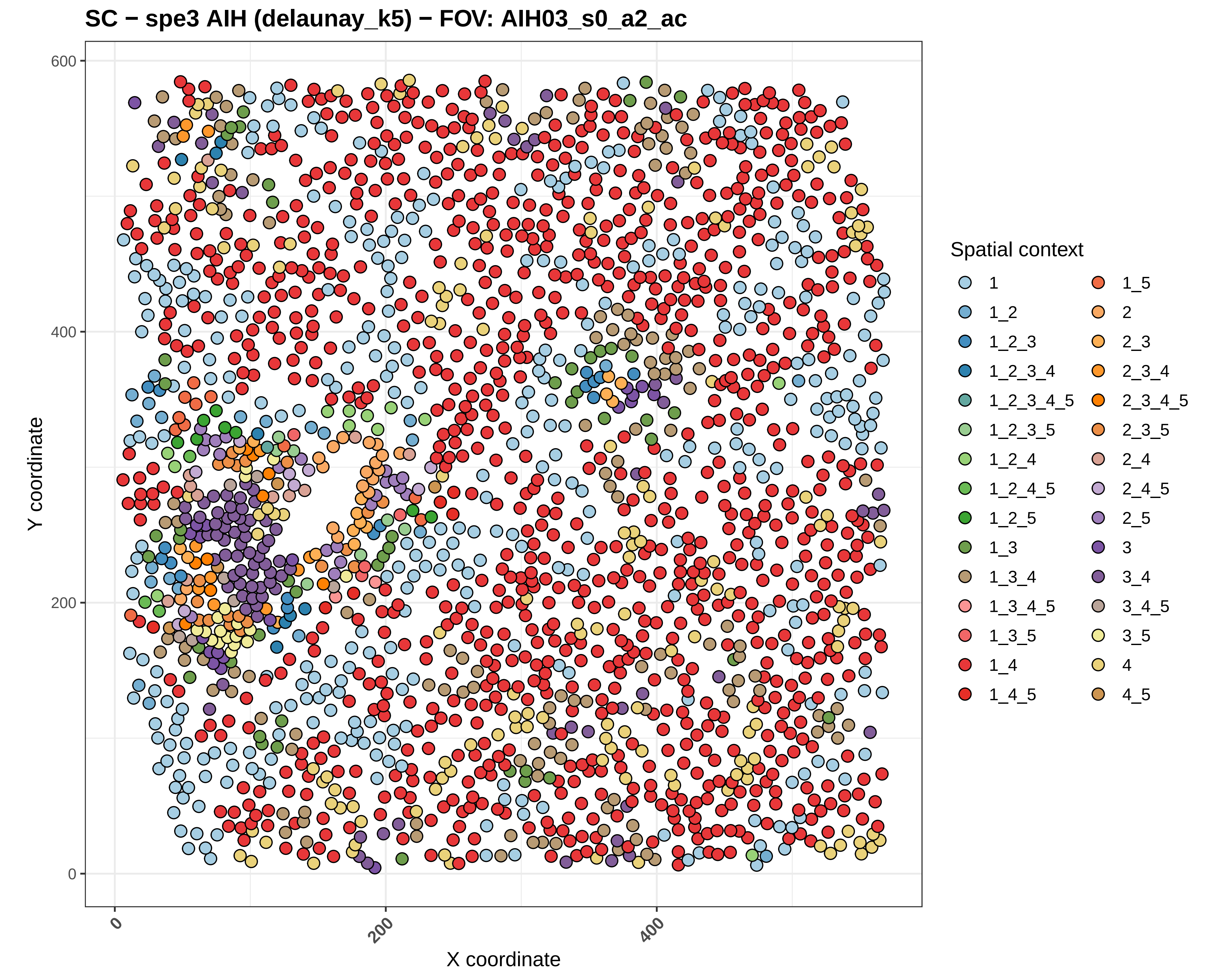

SC − spe3 AIH (delaunay_k5) − FOV: AIH03_s0_a2_ac
Y coordinate
X coordinate
0
200
400
600
Spatial context
1
1_2
1_2_3
1_2_3_4
1_2_3_4_5
1_2_3_5
1_2_4
1_2_4_5
1_2_5
1_3
1_3_4
1_3_4_5
1_3_5
1_4
1_4_5
1_5
2
2_3
2_3_4
2_3_4_5
2_3_5
2_4
2_4_5
2_5
3
3_4
3_4_5
3_5
4
4_5

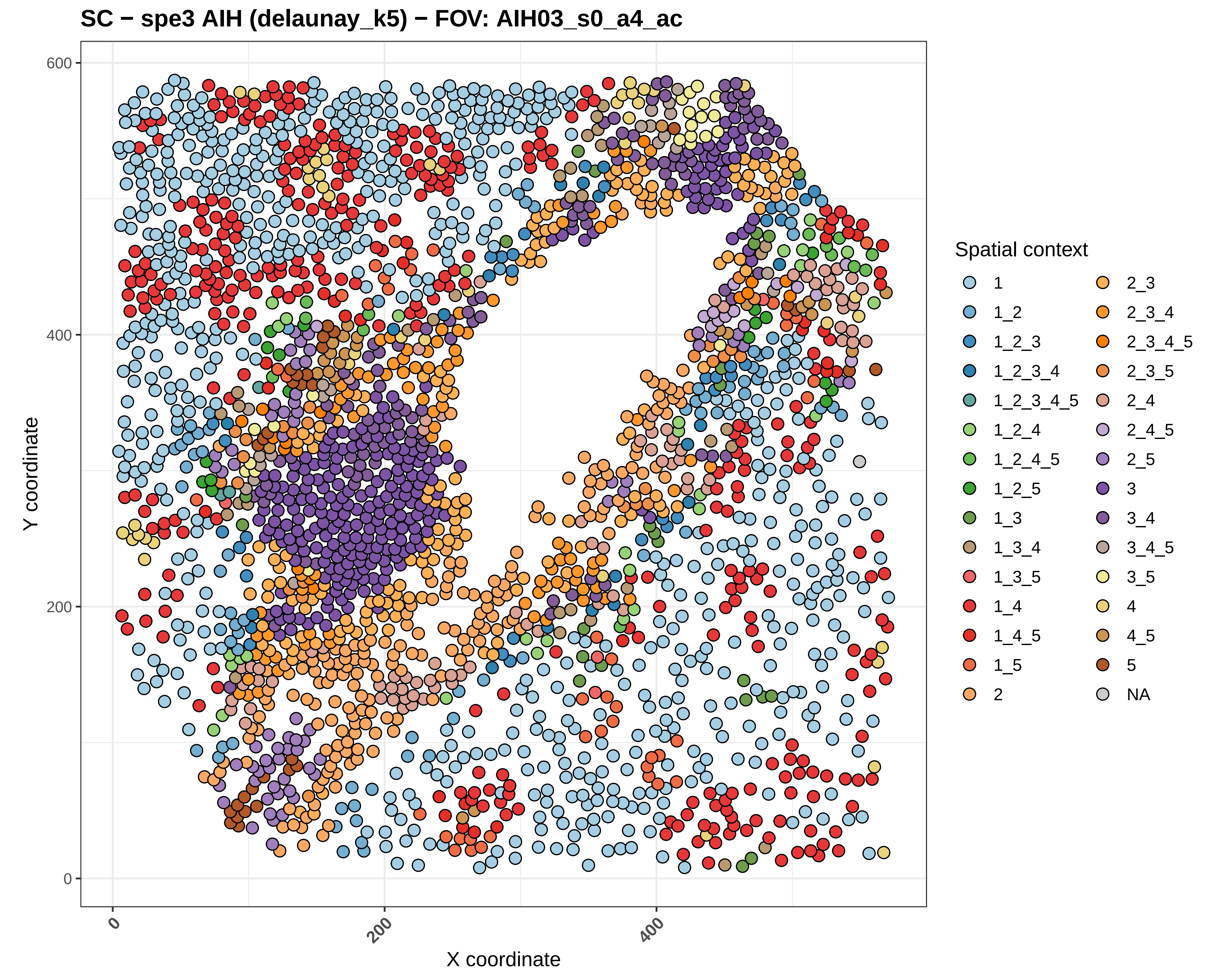
SC − spe3 AIH (delaunay_k5) − FOV: AIH03_s0_a4_ac
Y coordinate
X coordinate
0
200
400
600
Spatial context
1
1_2
1_2_3
1_2_3_4
1_2_3_4_5
1_2_4
1_2_4_5
1_2_5
1_3
1_3_4
1_3_5
1_4
1_4_5
1_5
2
2_3
2_3_4
2_3_4_5
2_3_5
2_4
2_4_5
2_5
3
3_4
3_4_5
3_5
4
4_5
5
NA

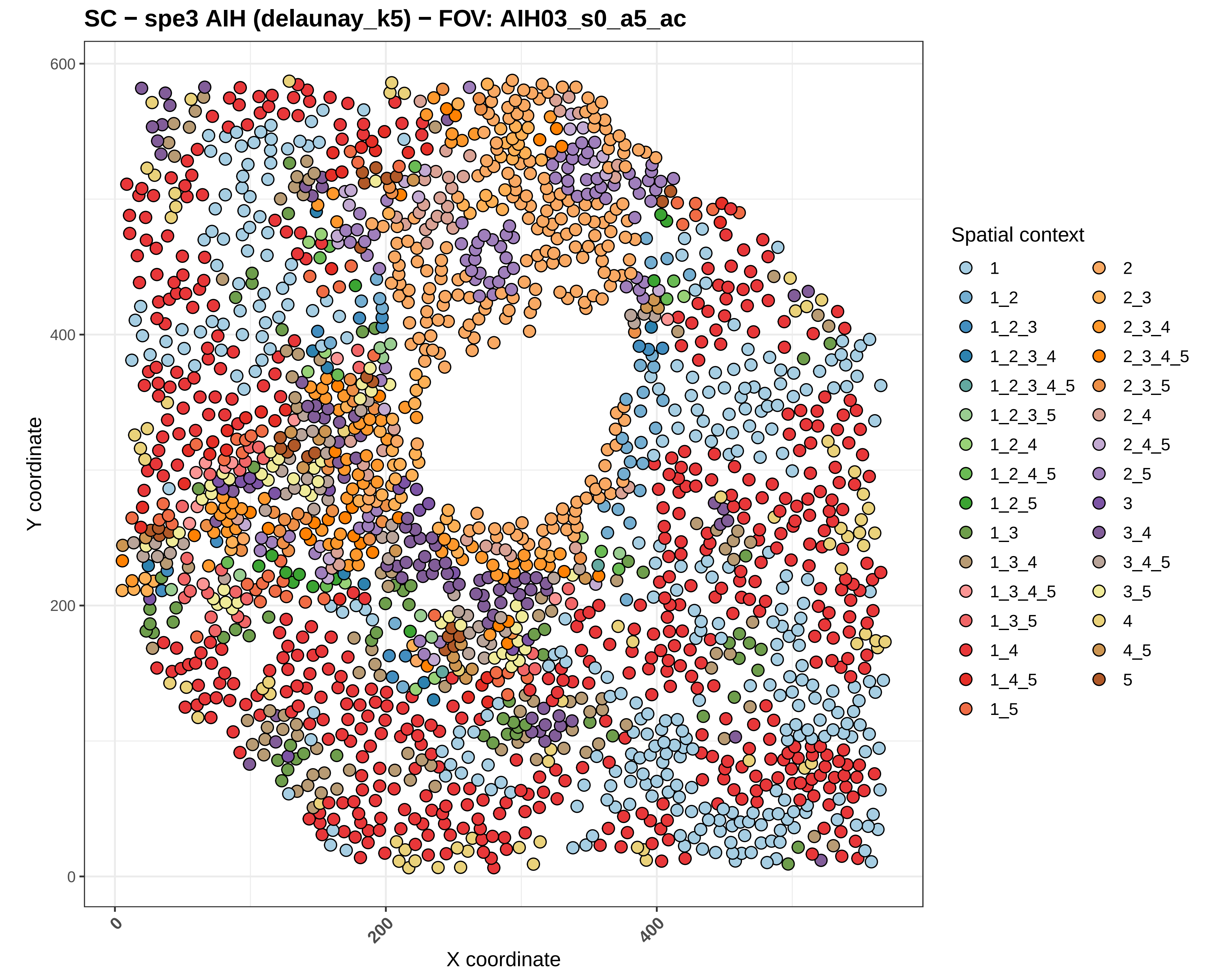
SC − spe3 AIH (delaunay_k5) − FOV: AIH03_s0_a5_ac
Y coordinate
X coordinate
0
200
400
600
Spatial context
1
1_2
1_2_3
1_2_3_4
1_2_3_4_5
1_2_3_5
1_2_4
1_2_4_5
1_2_5
1_3
1_3_4
1_3_4_5
1_3_5
1_4
1_4_5
1_5
2
2_3
2_3_4
2_3_4_5
2_3_5
2_4
2_4_5
2_5
3
3_4
3_4_5
3_5
4
4_5
5

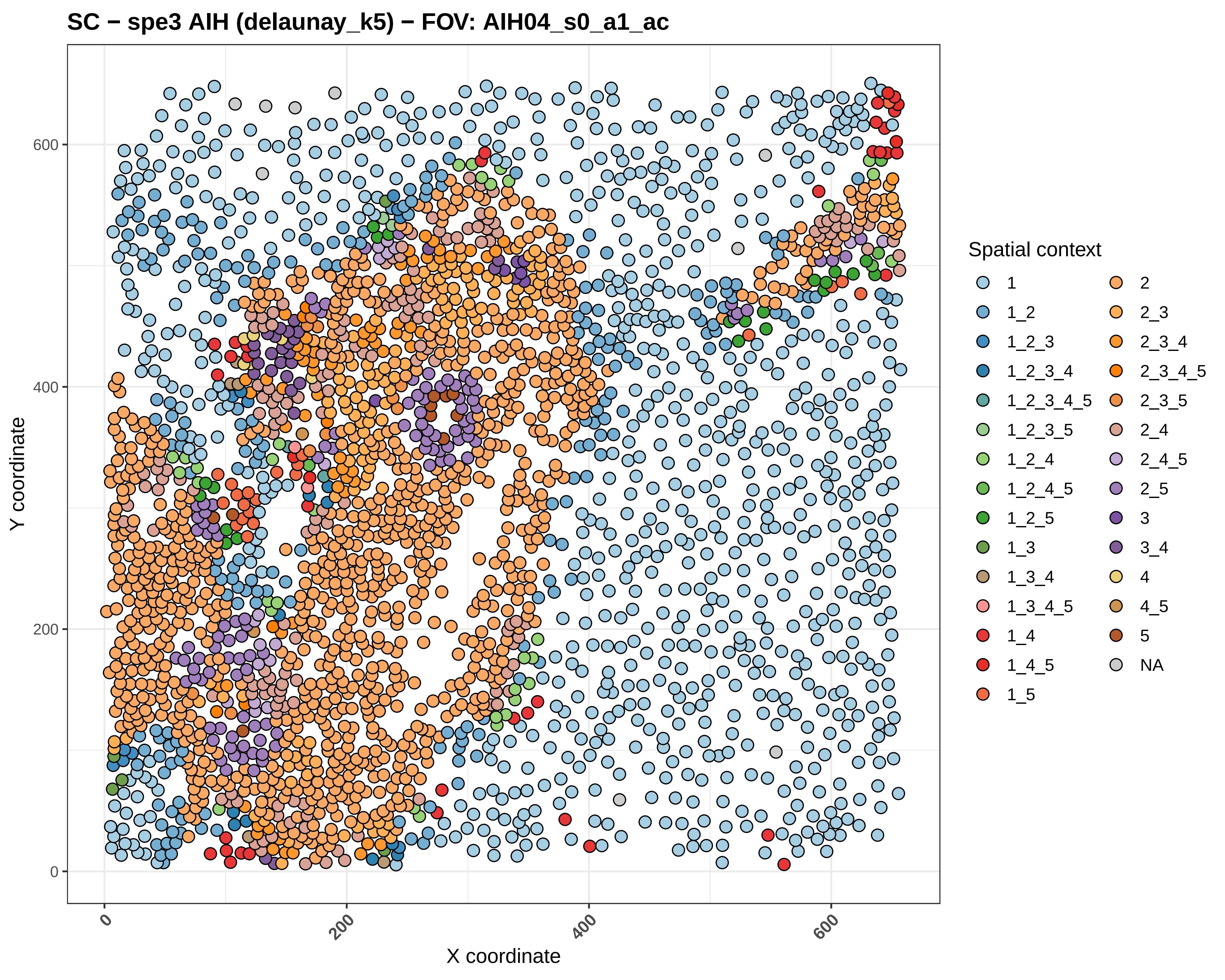

SC − spe3 AIH (delaunay_k5) − FOV: AIH04_s0_a1_ac
Y coordinate
X coordinate
0
200
400
600
Spatial context
1
1_2
1_2_3
1_2_3_4
1_2_3_4_5
1_2_3_5
1_2_4
1_2_4_5
1_2_5
1_3
1_3_4
1_3_4_5
1_4
1_4_5
1_5
2
2_3
2_3_4
2_3_4_5
2_3_5
2_4
2_4_5
2_5
3
3_4
4
4_5
5
NA

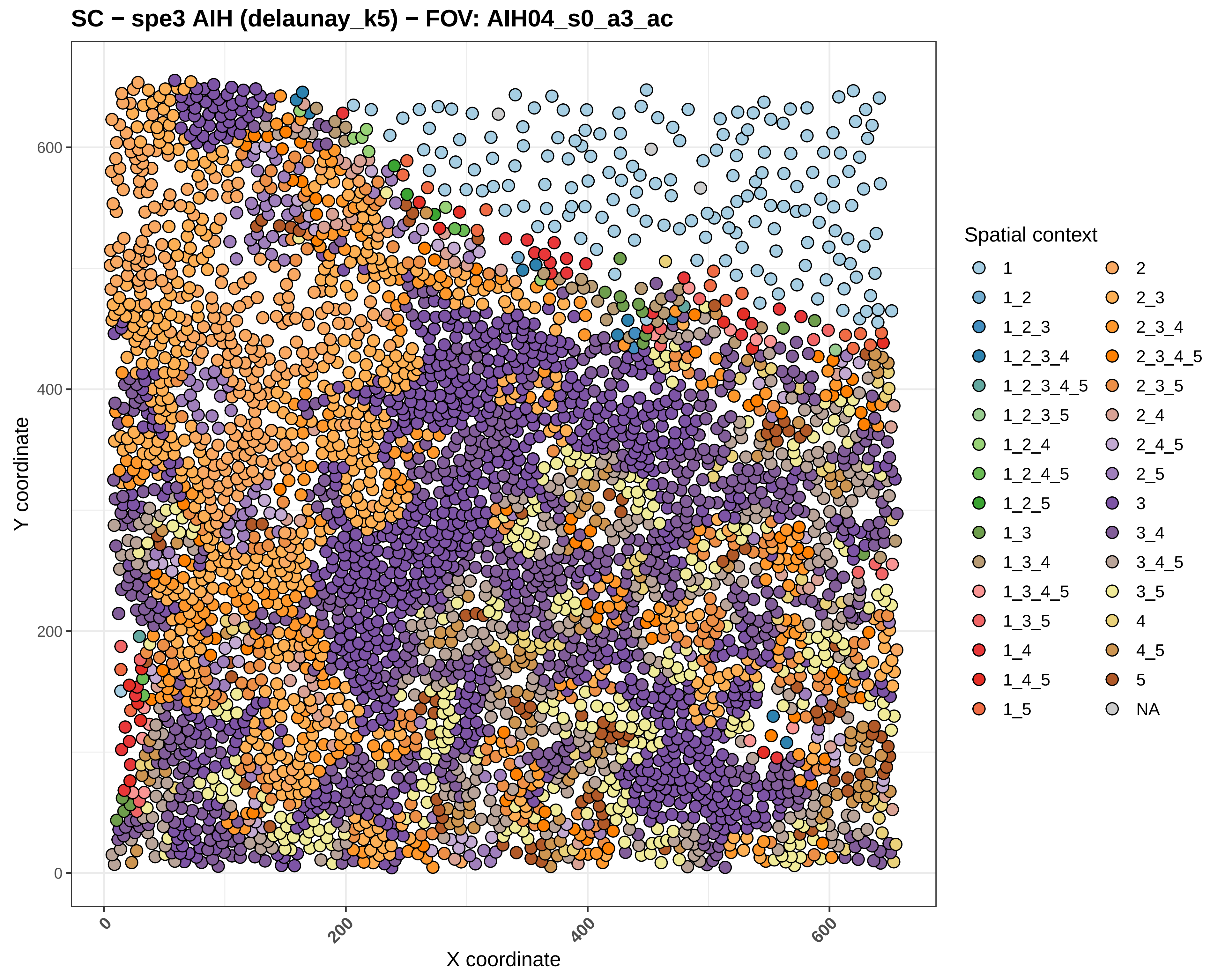
SC − spe3 AIH (delaunay_k5) − FOV: AIH04_s0_a3_ac
Y coordinate
X coordinate
0
200
400
600
Spatial context
1
1_2
1_2_3
1_2_3_4
1_2_3_4_5
1_2_3_5
1_2_4
1_2_4_5
1_2_5
1_3
1_3_4
1_3_4_5
1_3_5
1_4
1_4_5
1_5
2
2_3
2_3_4
2_3_4_5
2_3_5
2_4
2_4_5
2_5
3
3_4
3_4_5
3_5
4
4_5
5
NA

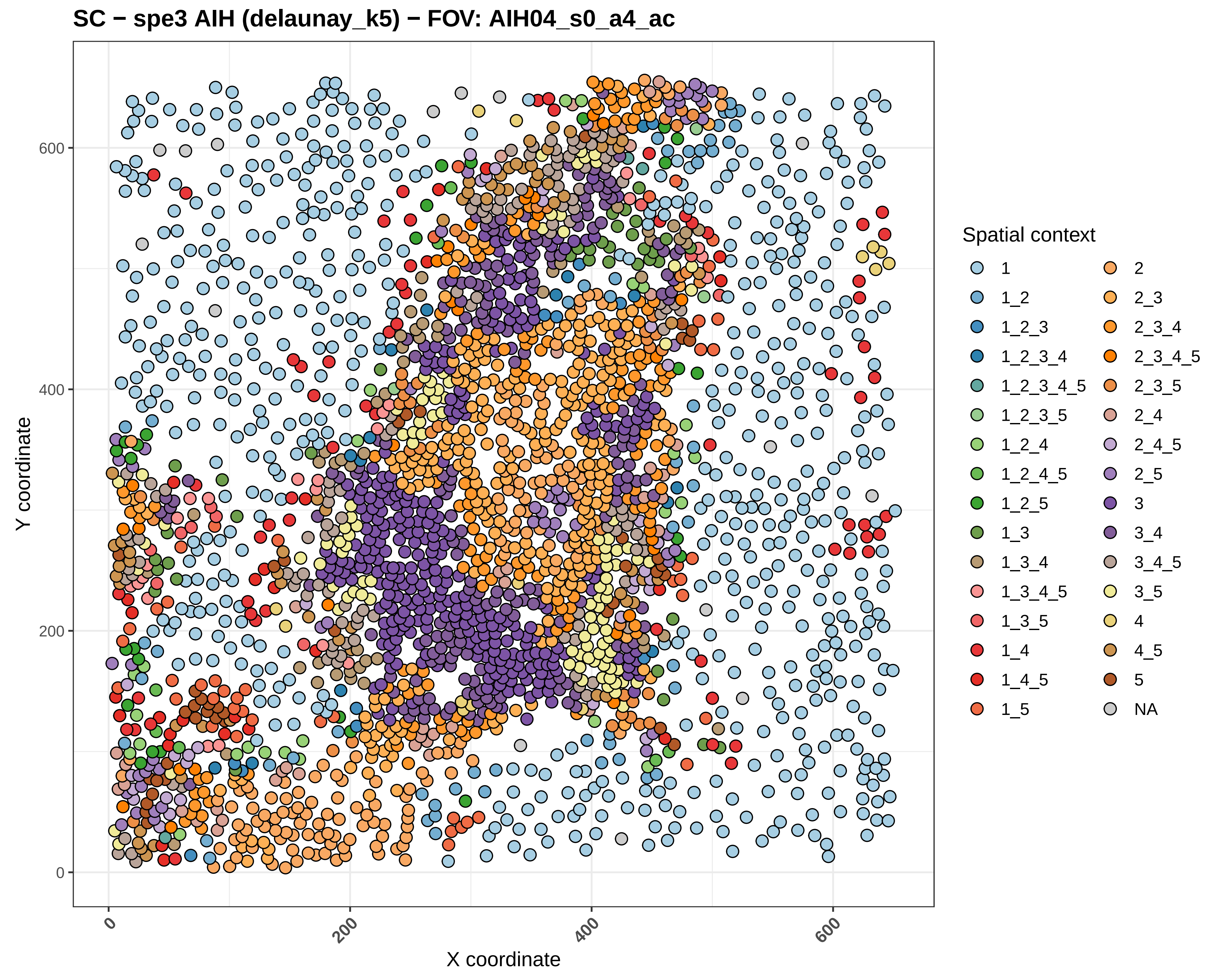
SC − spe3 AIH (delaunay_k5) − FOV: AIH04_s0_a4_ac
Y coordinate
X coordinate
0
200
400
600
Spatial context
1
1_2
1_2_3
1_2_3_4
1_2_3_4_5
1_2_3_5
1_2_4
1_2_4_5
1_2_5
1_3
1_3_4
1_3_4_5
1_3_5
1_4
1_4_5
1_5
2
2_3
2_3_4
2_3_4_5
2_3_5
2_4
2_4_5
2_5
3
3_4
3_4_5
3_5
4
4_5
5
NA

#### SC − spe3 AIH (delaunay_k5) − FOV: AIH04_s0_a5_ac

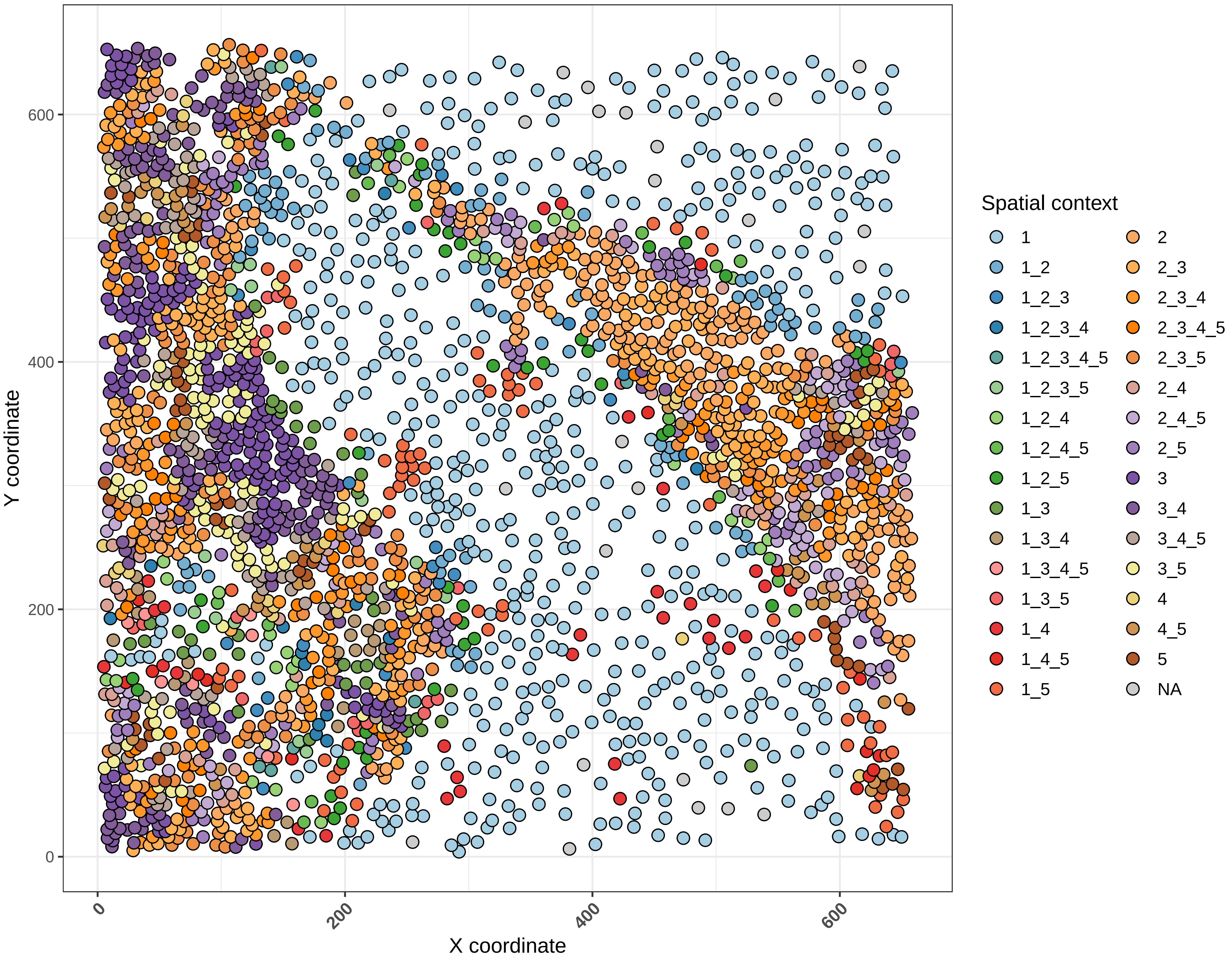

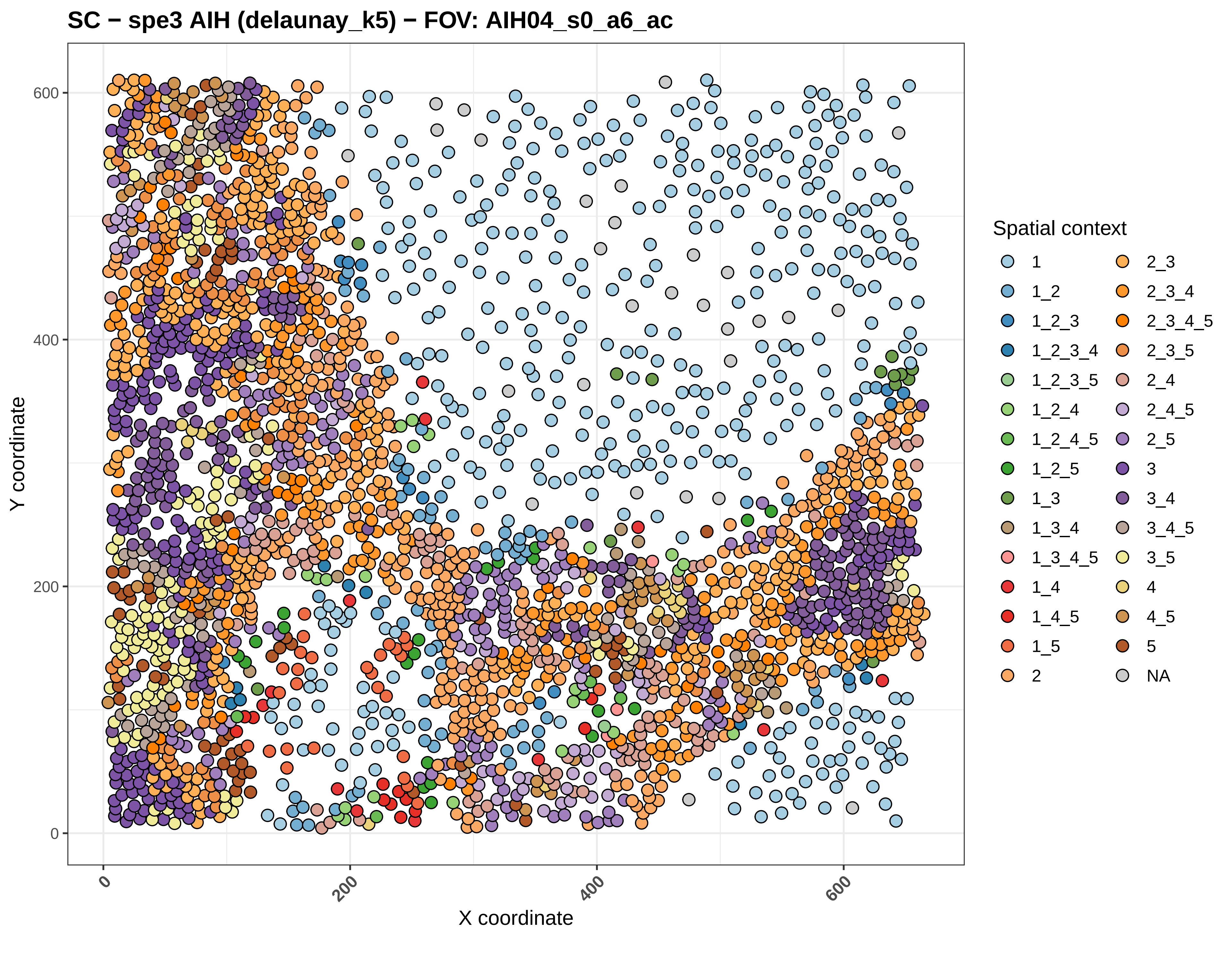

SC − spe3 AIH (delaunay_k5) − FOV: AIH04_s0_a6_ac
Y coordinate
X coordinate
0
200
400
600
Spatial context
1
1_2
1_2_3
1_2_3_4
1_2_3_5
1_2_4
1_2_4_5
1_2_5
1_3
1_3_4
1_3_4_5
1_4
1_4_5
1_5
2
2_3
2_3_4
2_3_4_5
2_3_5
2_4
2_4_5
2_5
3
3_4
3_4_5
3_5
4
4_5
5
NA

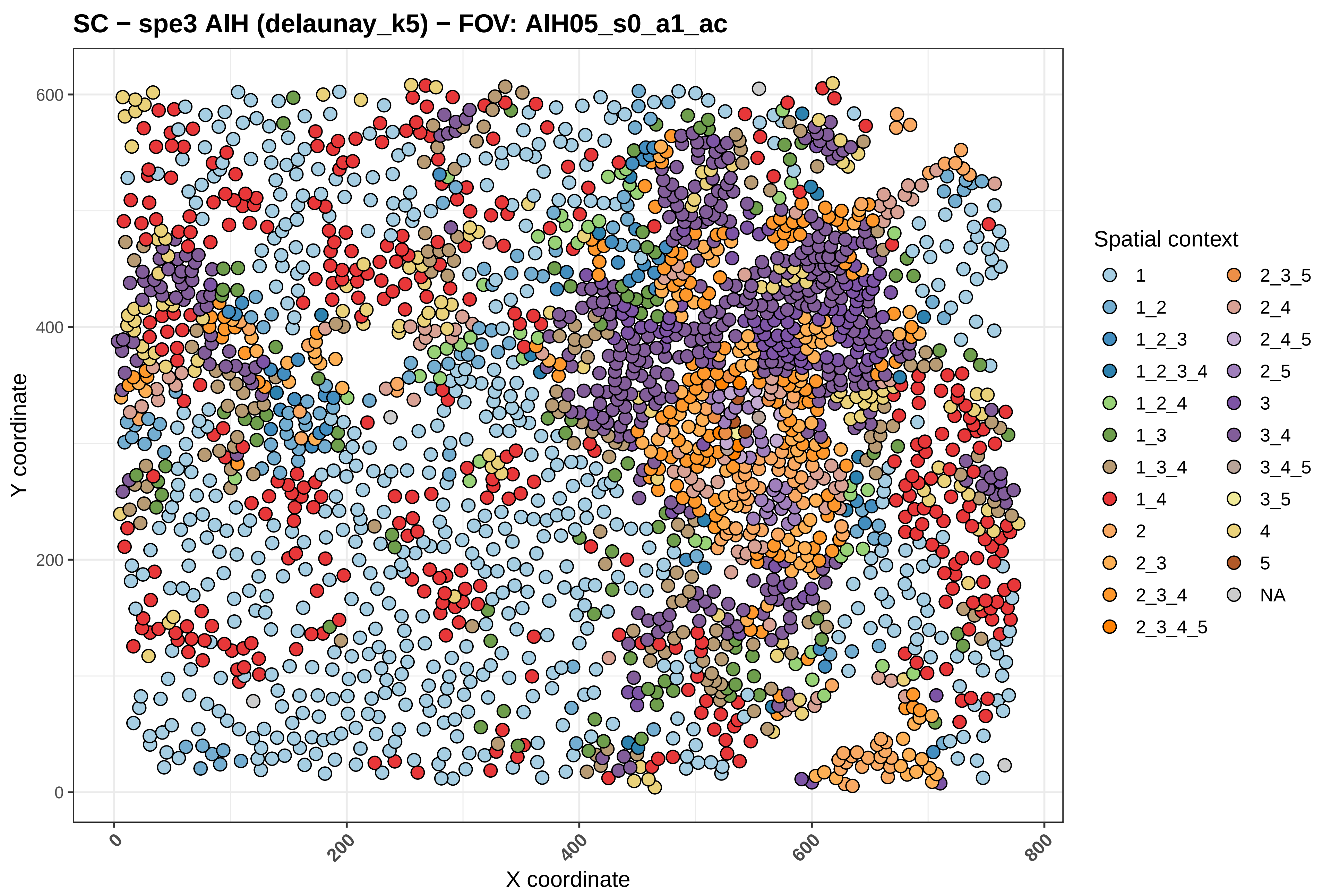

SC − spe3 AIH (delaunay_k5) − FOV: AIH05_s0_a1_ac
Y coordinate
X coordinate
0
200
400
600
800
Spatial context
1
1_2
1_2_3
1_2_3_4
1_2_4
1_3
1_3_4
1_4
2
2_3
2_3_4
2_3_4_5
2_3_5
2_4
2_4_5
2_5
3
3_4
3_4_5
3_5
4
5
NA

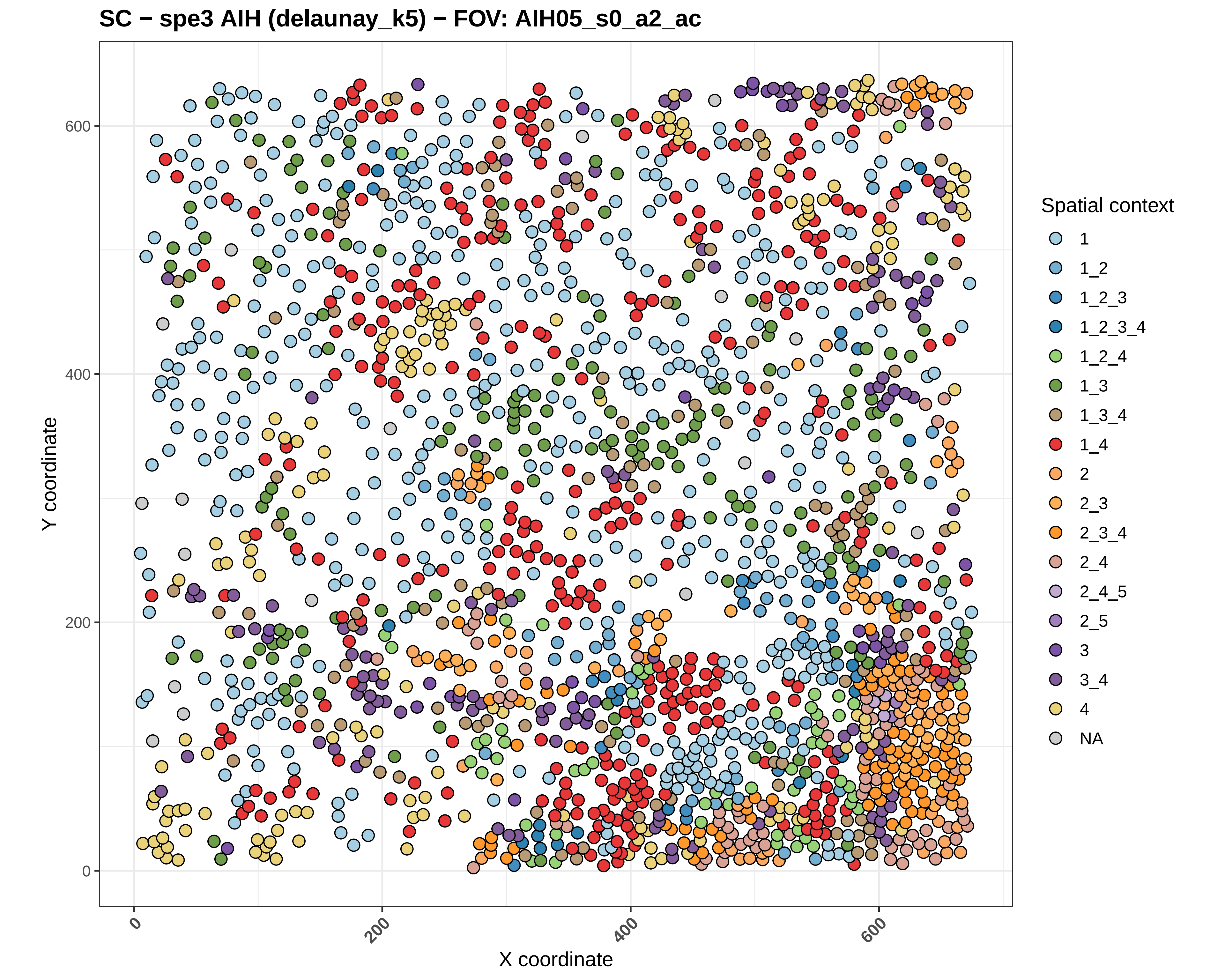
SC − spe3 AIH (delaunay_k5) − FOV: AIH05_s0_a2_ac
Y coordinate
X coordinate
0
200
400
600
Spatial context
1
1_2
1_2_3
1_2_3_4
1_2_4
1_3
1_3_4
1_4
2
2_3
2_3_4
2_4
2_4_5
2_5
3
3_4
4
NA

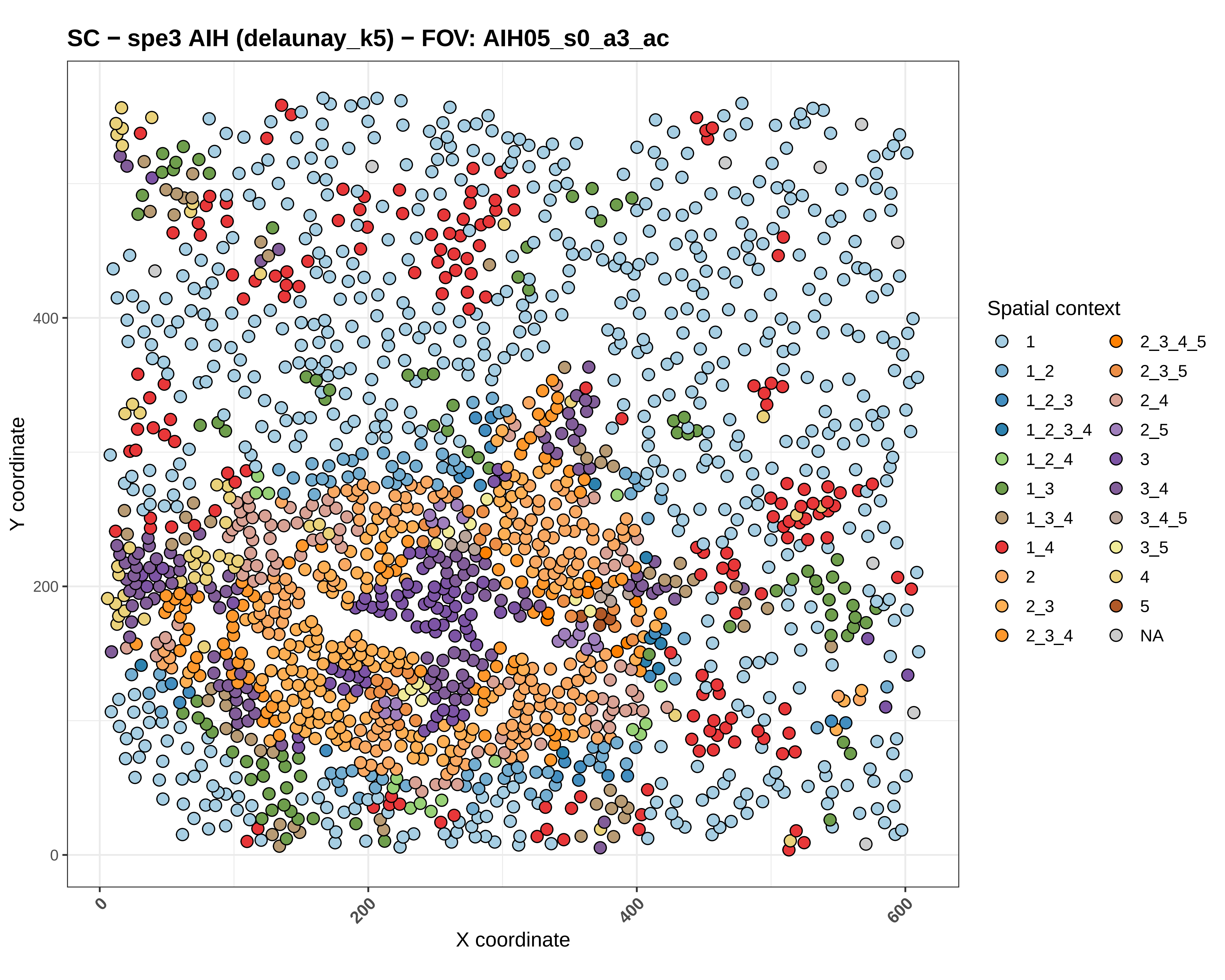
SC − spe3 AIH (delaunay_k5) − FOV: AIH05_s0_a3_ac
Y coordinate
X coordinate
0
200
400
600
Spatial context
1
1_2
1_2_3
1_2_3_4
1_2_4
1_3
1_3_4
1_4
2
2_3
2_3_4
2_3_4_5
2_3_5
2_4
2_5
3
3_4
3_4_5
3_5
4
5
NA

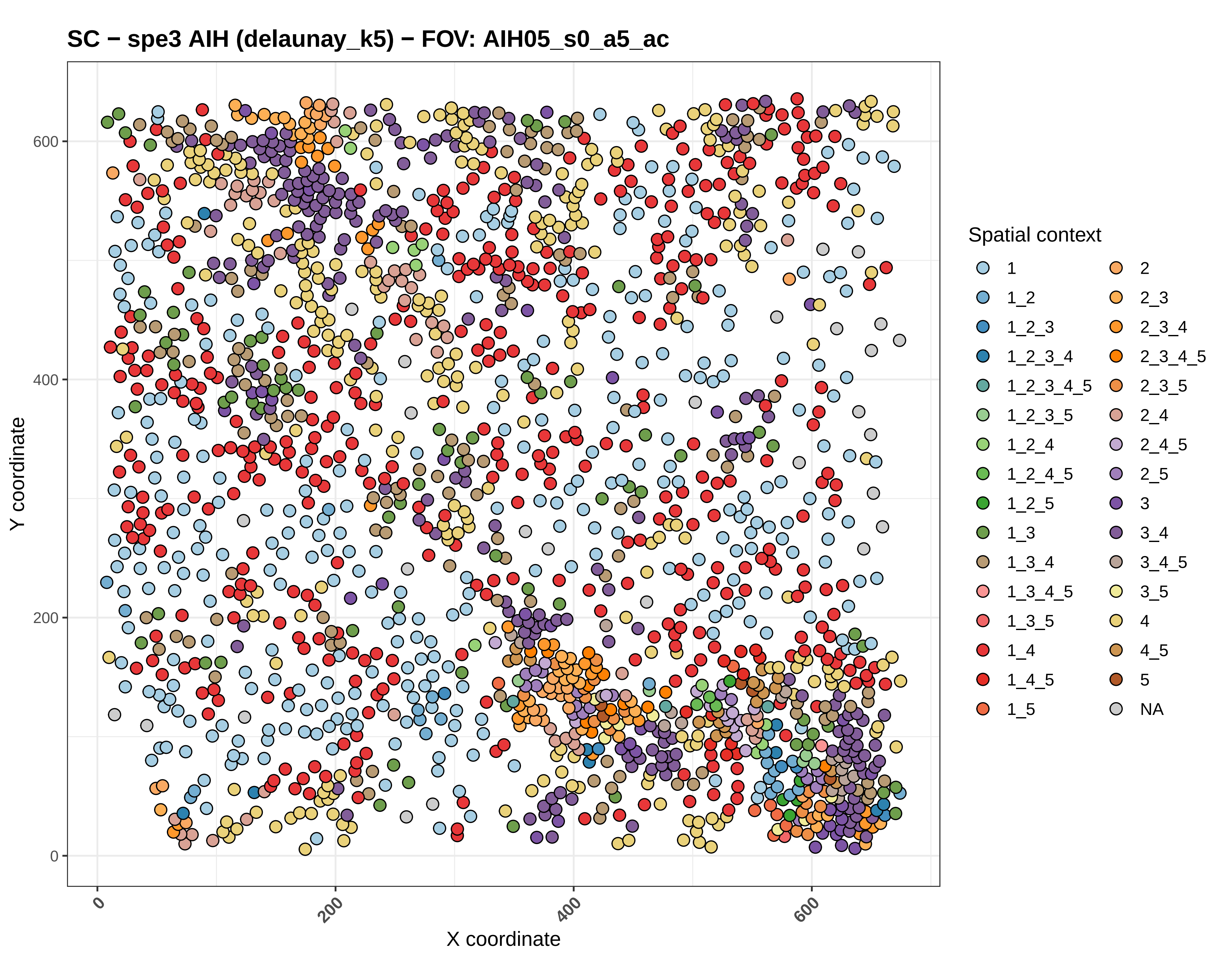

SC − spe3 AIH (delaunay_k5) − FOV: AIH05_s0_a5_ac
Y coordinate
X coordinate
0
200
400
600
Spatial context
1
1_2
1_2_3
1_2_3_4
1_2_3_4_5
1_2_3_5
1_2_4
1_2_4_5
1_2_5
1_3
1_3_4
1_3_4_5
1_3_5
1_4
1_4_5
1_5
2
2_3
2_3_4
2_3_4_5
2_3_5
2_4
2_4_5
2_5
3
3_4
3_4_5
3_5
4
4_5
5
NA

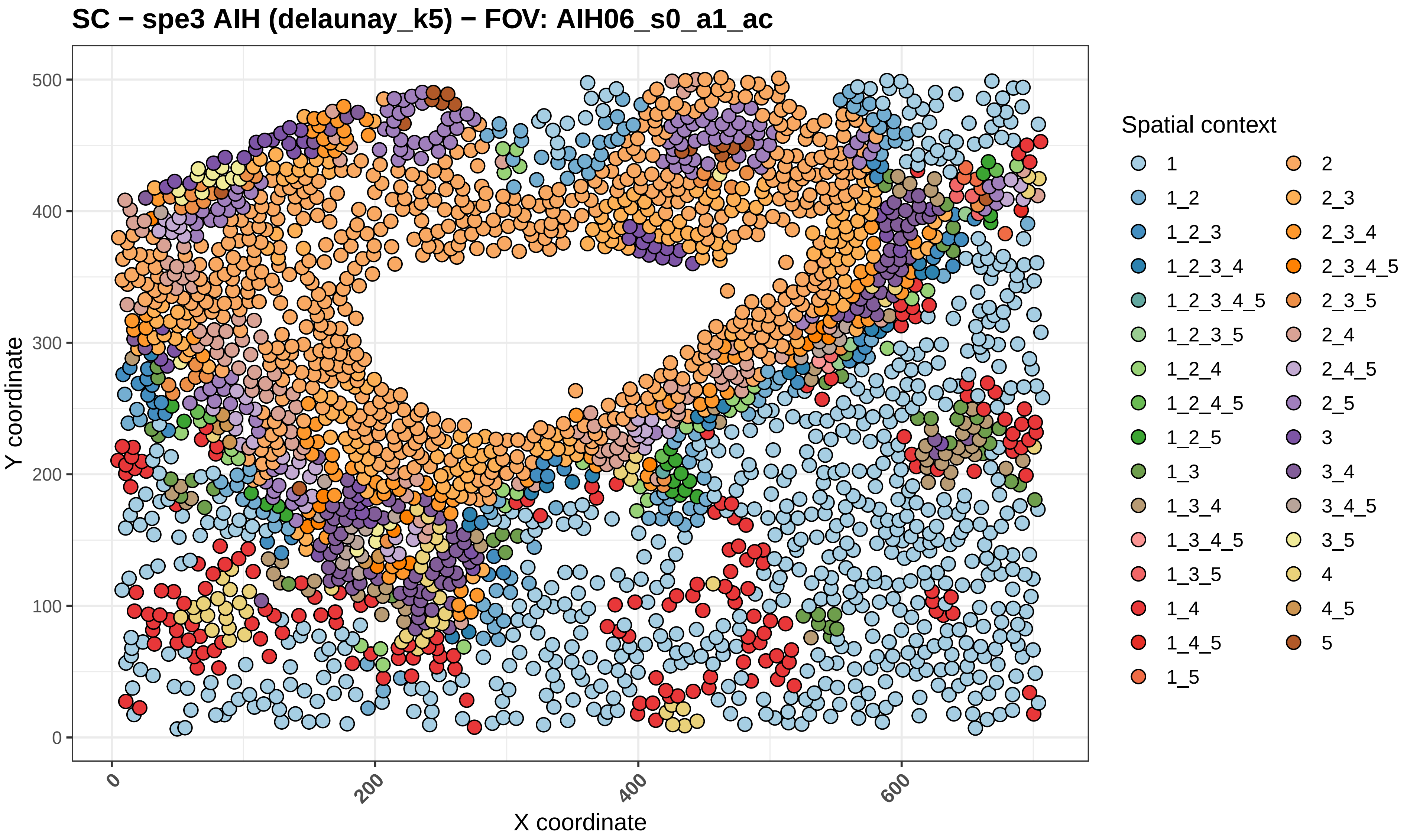
SC − spe3 AIH (delaunay_k5) − FOV: AIH06_s0_a1_ac
Y coordinate
0
100
200
300
400
500
X coordinate
0
200
400
600
Spatial context
1
1_2
1_2_3
1_2_3_4
1_2_3_4_5
1_2_3_5
1_2_4
1_2_4_5
1_2_5
1_3
1_3_4
1_3_4_5
1_3_5
1_4
1_4_5
1_5
2
2_3
2_3_4
2_3_4_5
2_3_5
2_4
2_4_5
2_5
3
3_4
3_4_5
3_5
4
4_5
5

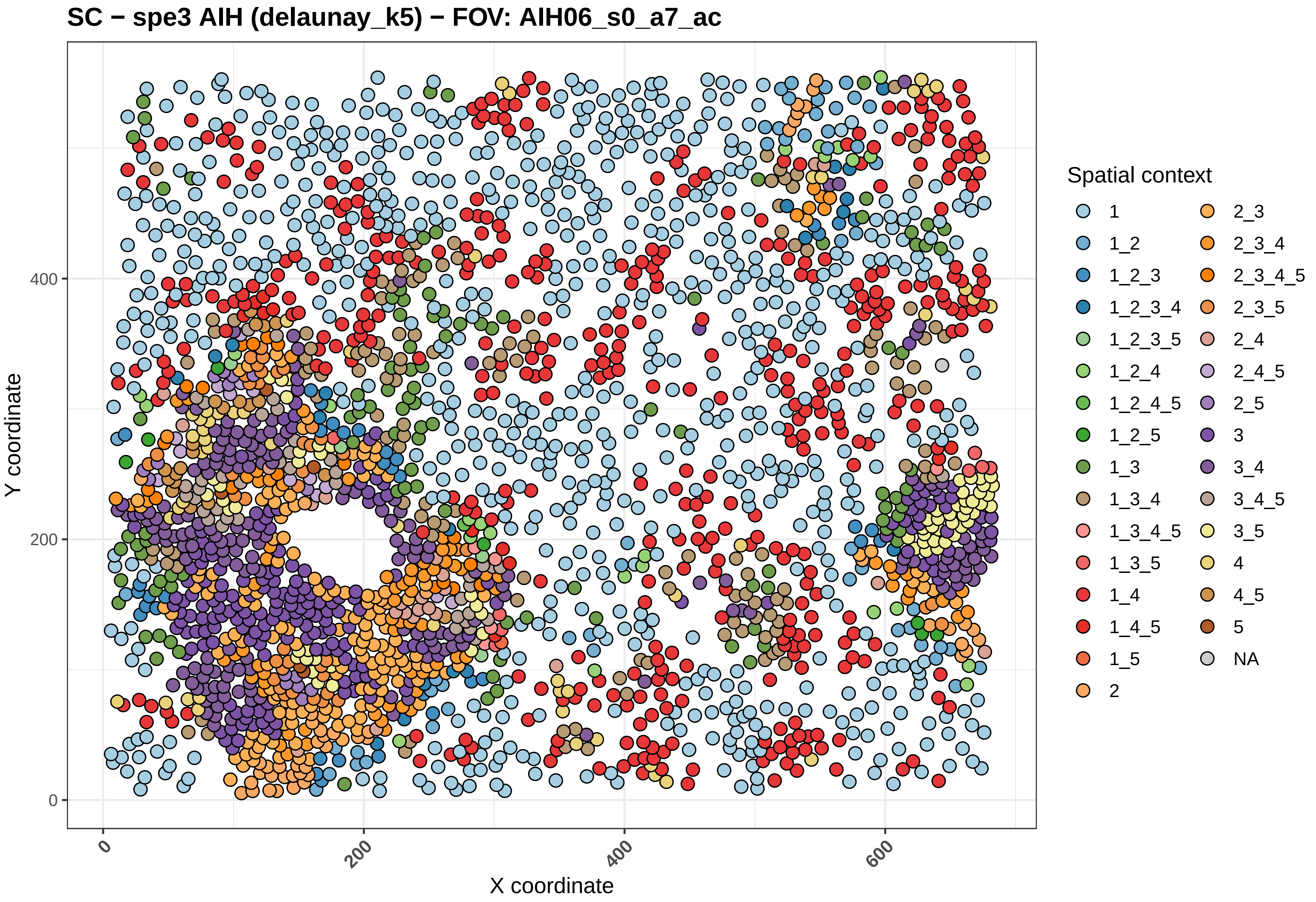

SC − spe3 AIH (delaunay_k5) − FOV: AIH06_s0_a7_ac
Y coordinate
X coordinate
0
200
400
600
Spatial context
1
1_2
1_2_3
1_2_3_4
1_2_3_5
1_2_4
1_2_4_5
1_2_5
1_3
1_3_4
1_3_4_5
1_3_5
1_4
1_4_5
1_5
2
2_3
2_3_4
2_3_4_5
2_3_5
2_4
2_4_5
2_5
3
3_4
3_4_5
3_5
4
4_5
5
NA

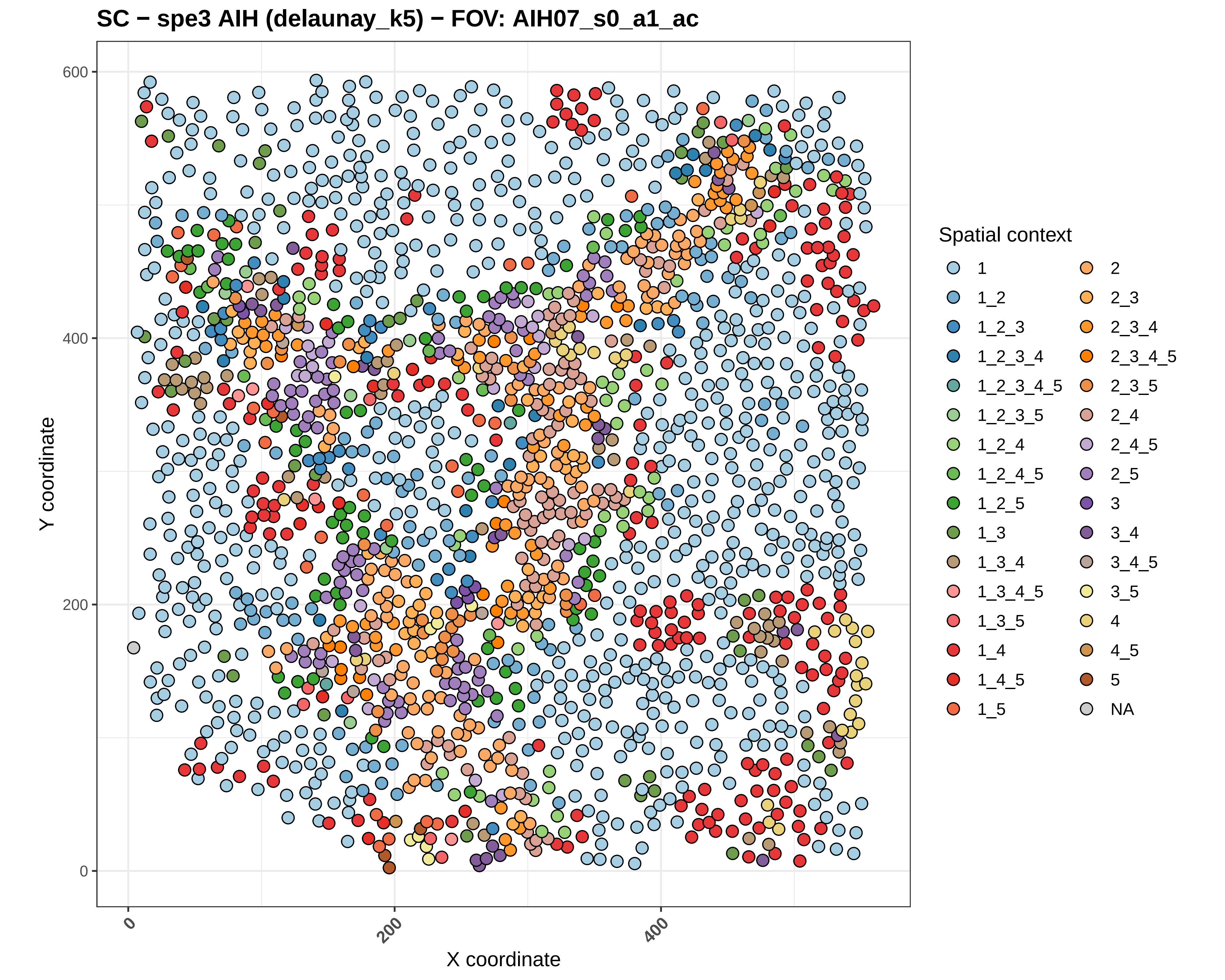
SC − spe3 AIH (delaunay_k5) − FOV: AIH07_s0_a1_ac
Y coordinate
X coordinate
0
200
400
600
Spatial context
1
1_2
1_2_3
1_2_3_4
1_2_3_4_5
1_2_3_5
1_2_4
1_2_4_5
1_2_5
1_3
1_3_4
1_3_4_5
1_3_5
1_4
1_4_5
1_5
2
2_3
2_3_4
2_3_4_5
2_3_5
2_4
2_4_5
2_5
3
3_4
3_4_5
3_5
4
4_5
5
NA

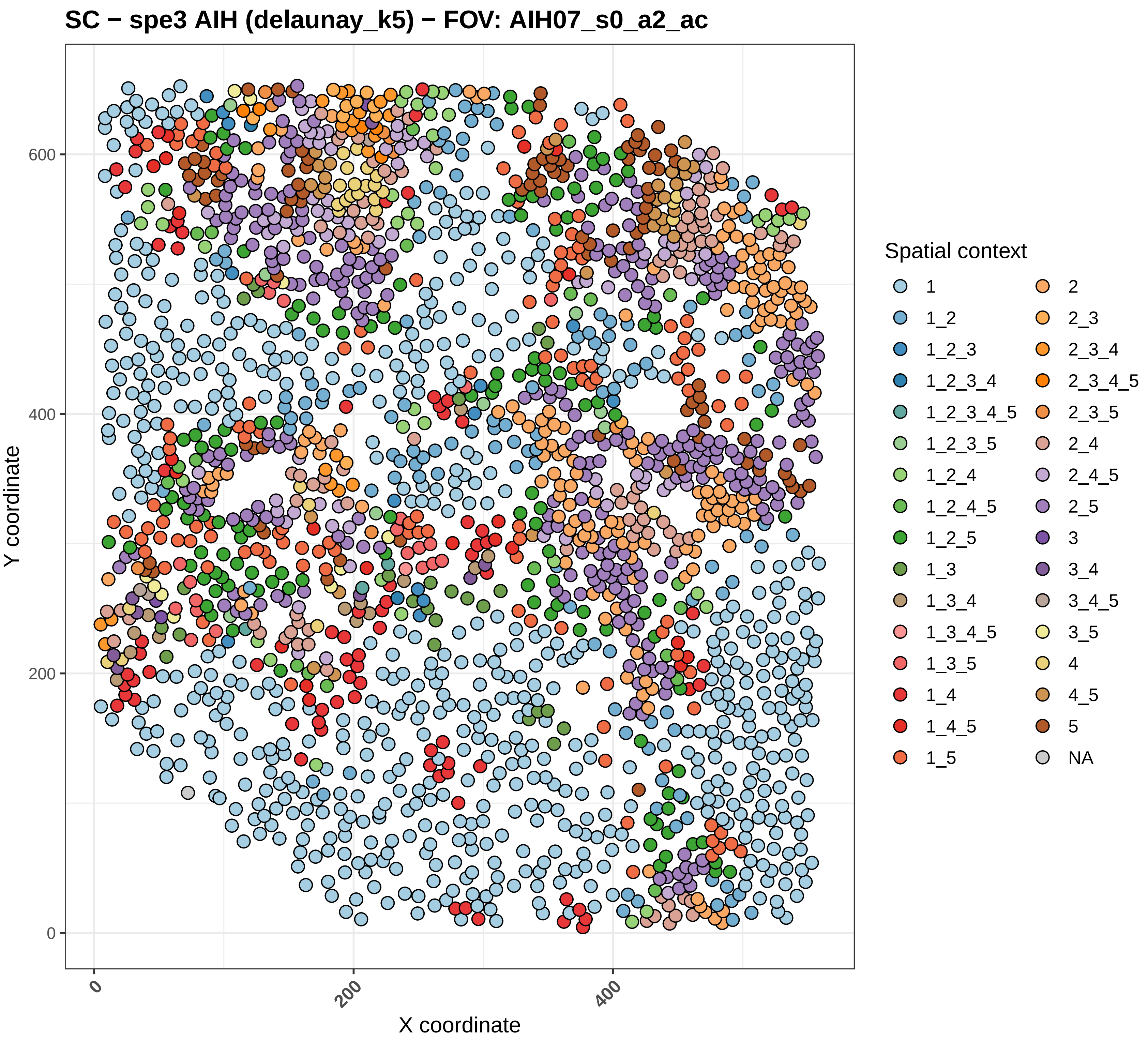
SC − spe3 AIH (delaunay_k5) − FOV: AIH07_s0_a2_ac
Y coordinate
X coordinate
0
200
400
600
0
200
400
Spatial context
1
1_2
1_2_3
1_2_3_4
1_2_3_4_5
1_2_3_5
1_2_4
1_2_4_5
1_2_5
1_3
1_3_4
1_3_4_5
1_3_5
1_4
1_4_5
1_5
2
2_3
2_3_4
2_3_4_5
2_3_5
2_4
2_4_5
2_5
3
3_4
3_4_5
3_5
4
4_5
5
NA

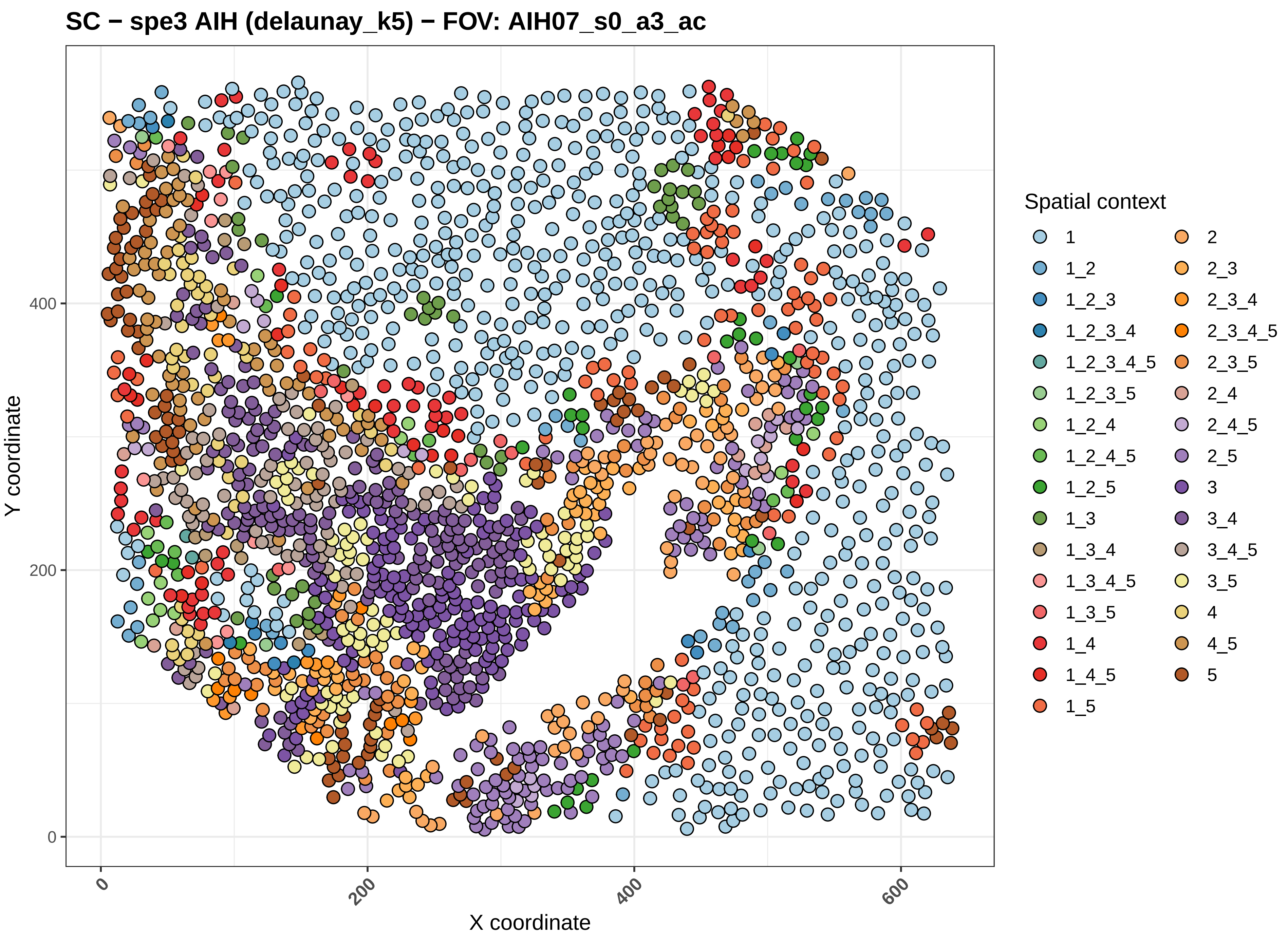

SC − spe3 AIH (delaunay_k5) − FOV: AIH07_s0_a3_ac
Y coordinate
X coordinate
0
200
400
600
Spatial context
1
1_2
1_2_3
1_2_3_4
1_2_3_4_5
1_2_3_5
1_2_4
1_2_4_5
1_2_5
1_3
1_3_4
1_3_4_5
1_3_5
1_4
1_4_5
1_5
2
2_3
2_3_4
2_3_4_5
2_3_5
2_4
2_4_5
2_5
3
3_4
3_4_5
3_5
4
4_5
5

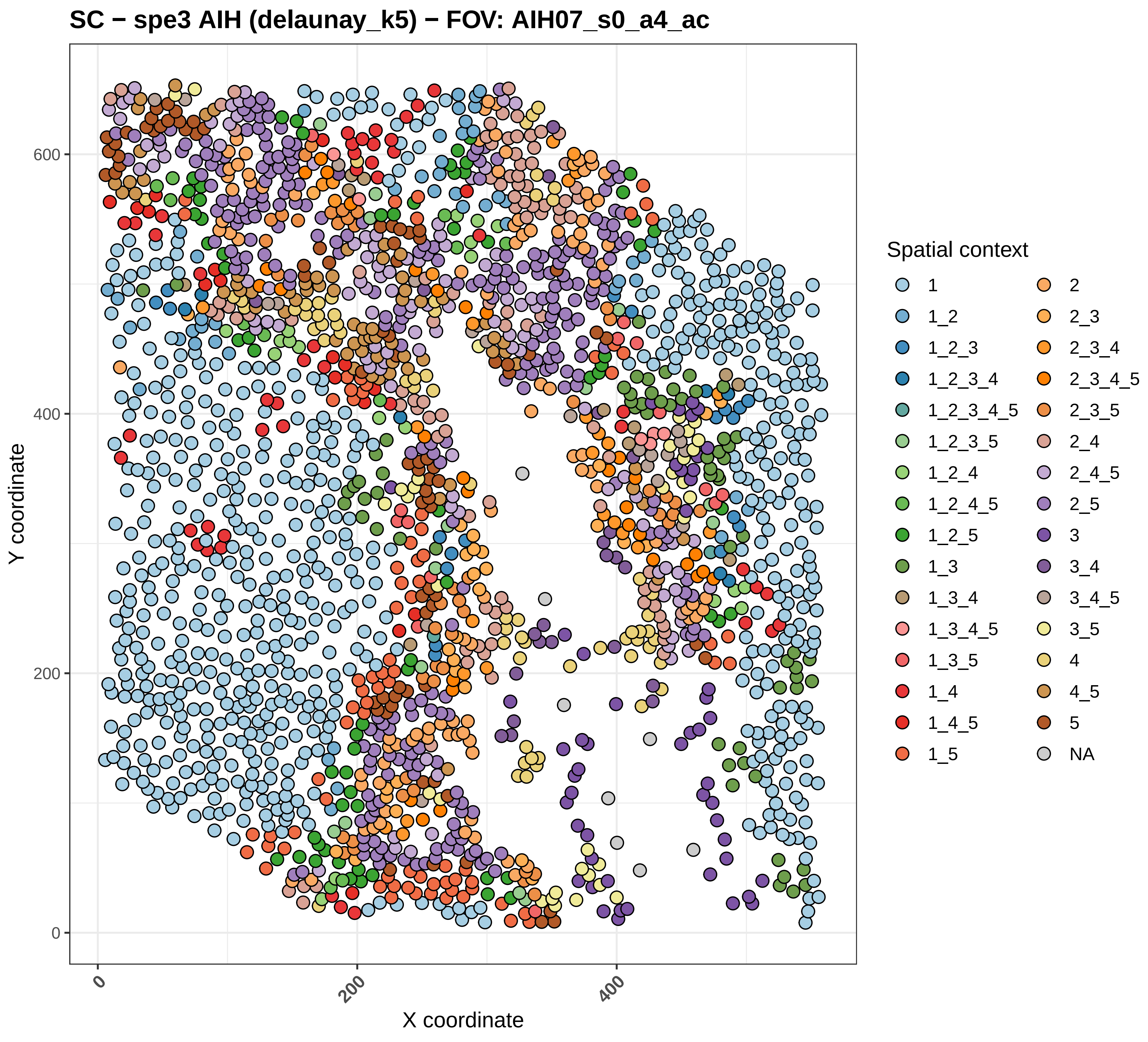
SC − spe3 AIH (delaunay_k5) − FOV: AIH07_s0_a4_ac
Y coordinate
X coordinate
0
200
400
600
Spatial context
1
1_2
1_2_3
1_2_3_4
1_2_3_4_5
1_2_3_5
1_2_4
1_2_4_5
1_2_5
1_3
1_3_4
1_3_4_5
1_3_5
1_4
1_4_5
1_5
2
2_3
2_3_4
2_3_4_5
2_3_5
2_4
2_4_5
2_5
3
3_4
3_4_5
3_5
4
4_5
5
NA

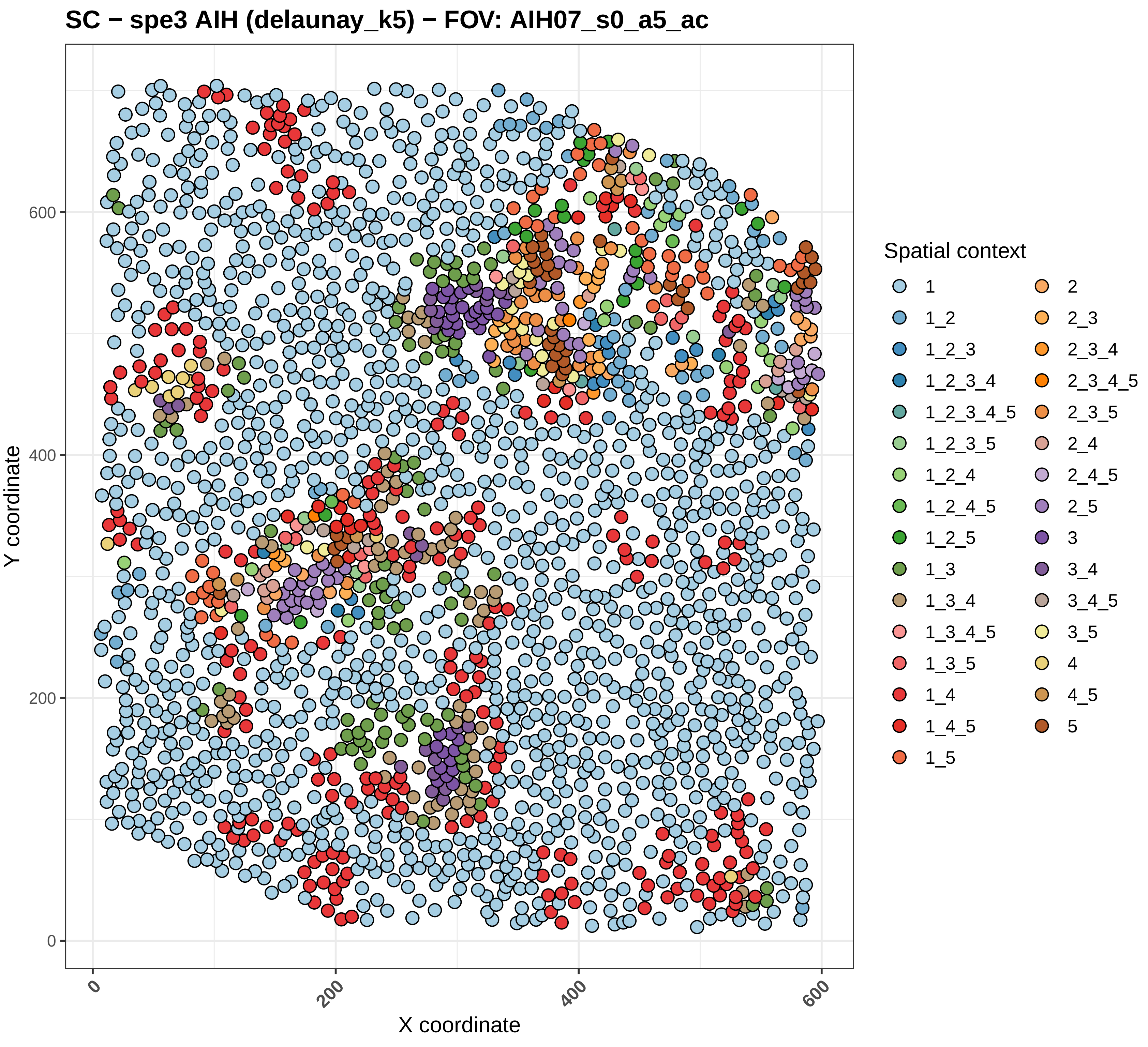
SC − spe3 AIH (delaunay_k5) − FOV: AIH07_s0_a5_ac
Y coordinate
X coordinate
0
200
400
600
Spatial context
1
1_2
1_2_3
1_2_3_4
1_2_3_4_5
1_2_3_5
1_2_4
1_2_4_5
1_2_5
1_3
1_3_4
1_3_4_5
1_3_5
1_4
1_4_5
1_5
2
2_3
2_3_4
2_3_4_5
2_3_5
2_4
2_4_5
2_5
3
3_4
3_4_5
3_5
4
4_5
5

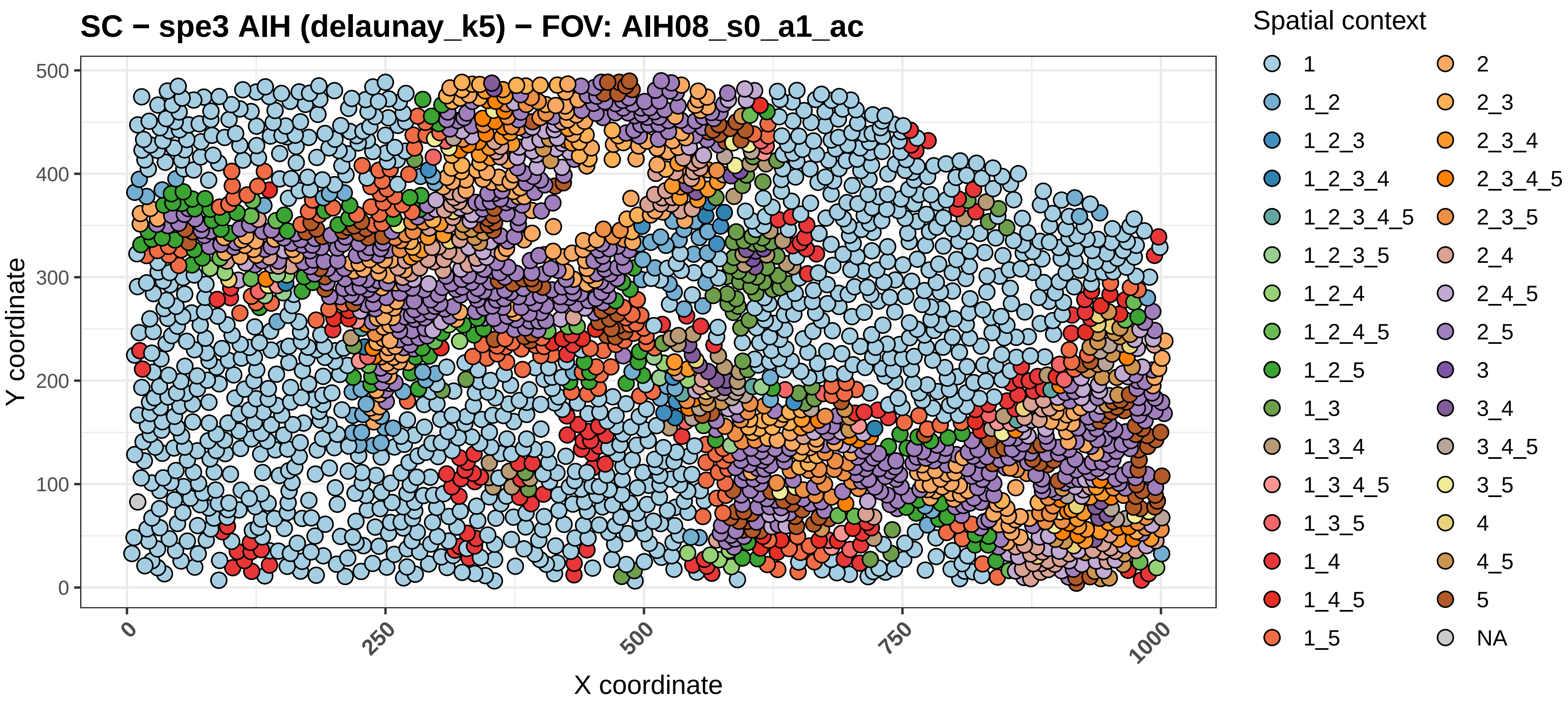

SC − spe3 AIH (delaunay_k5) − FOV: AIH08_s0_a1_ac
Y coordinate
0
100
200
300
400
500
X coordinate
0
250
500
750
1000
Spatial context
1
1_2
1_2_3
1_2_3_4
1_2_3_4_5
1_2_3_5
1_2_4
1_2_4_5
1_2_5
1_3
1_3_4
1_3_4_5
1_3_5
1_4
1_4_5
1_5
2
2_3
2_3_4
2_3_4_5
2_3_5
2_4
2_4_5
2_5
3
3_4
3_4_5
3_5
4
4_5
5
NA

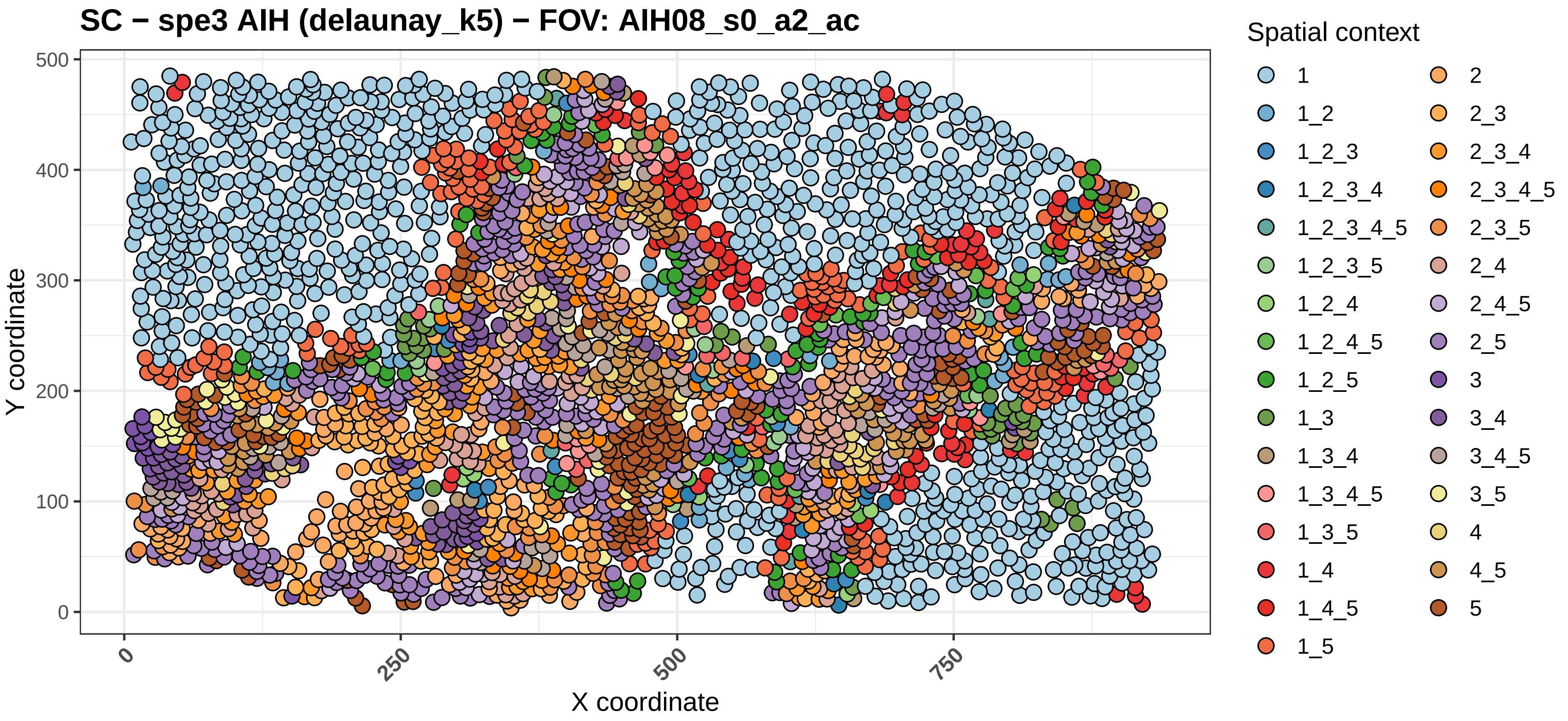
SC − spe3 AIH (delaunay_k5) − FOV: AIH08_s0_a2_ac
Y coordinate
X coordinate
0
100
200
300
400
500
0
250
500
750
Spatial context
1
1_2
1_2_3
1_2_3_4
1_2_3_4_5
1_2_3_5
1_2_4
1_2_4_5
1_2_5
1_3
1_3_4
1_3_4_5
1_3_5
1_4
1_4_5
1_5
2
2_3
2_3_4
2_3_4_5
2_3_5
2_4
2_4_5
2_5
3
3_4
3_4_5
3_5
4
4_5
5

SC − spe3 AIH (delaunay_k5) − FOV: AIH08_s0_a3_ac

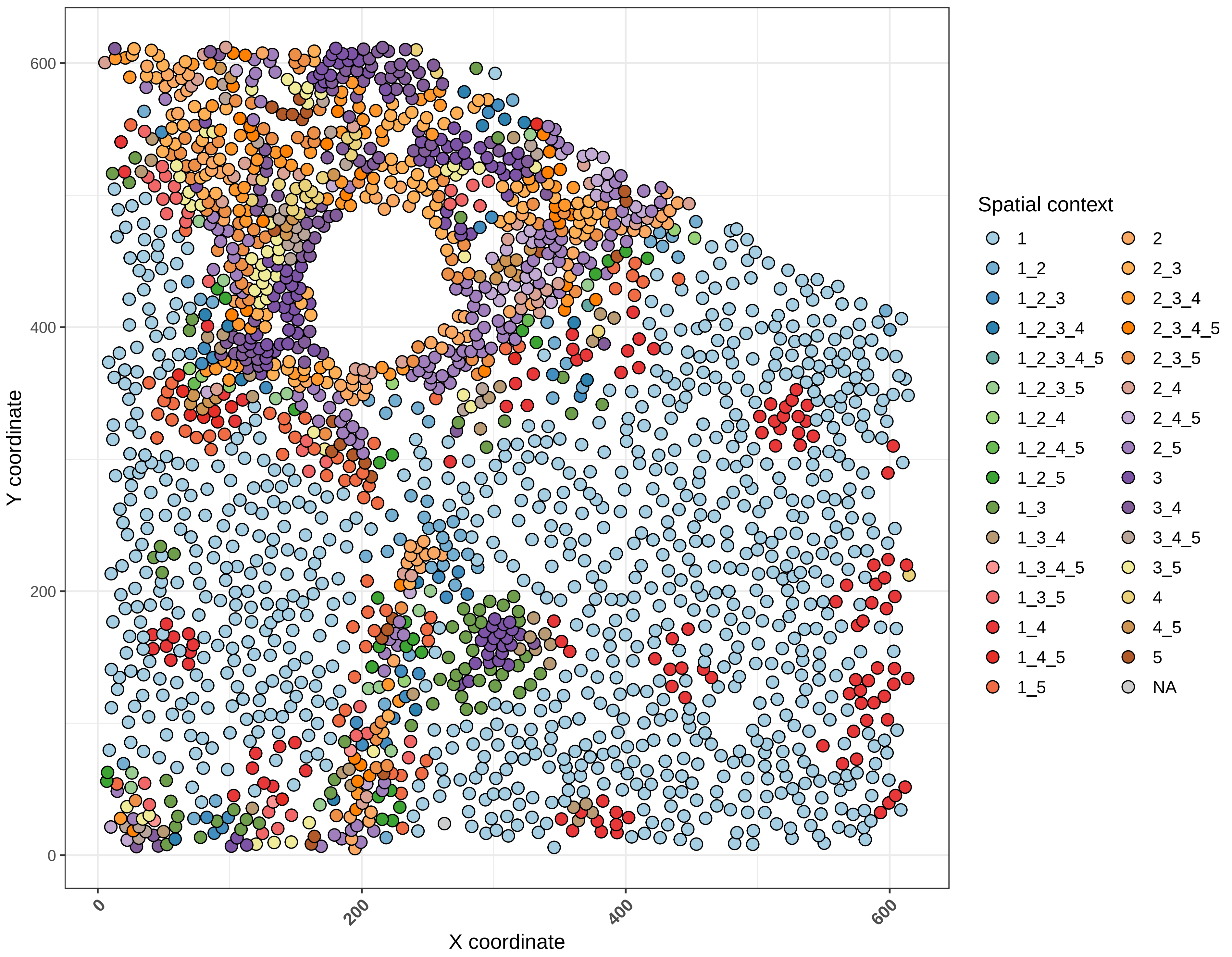

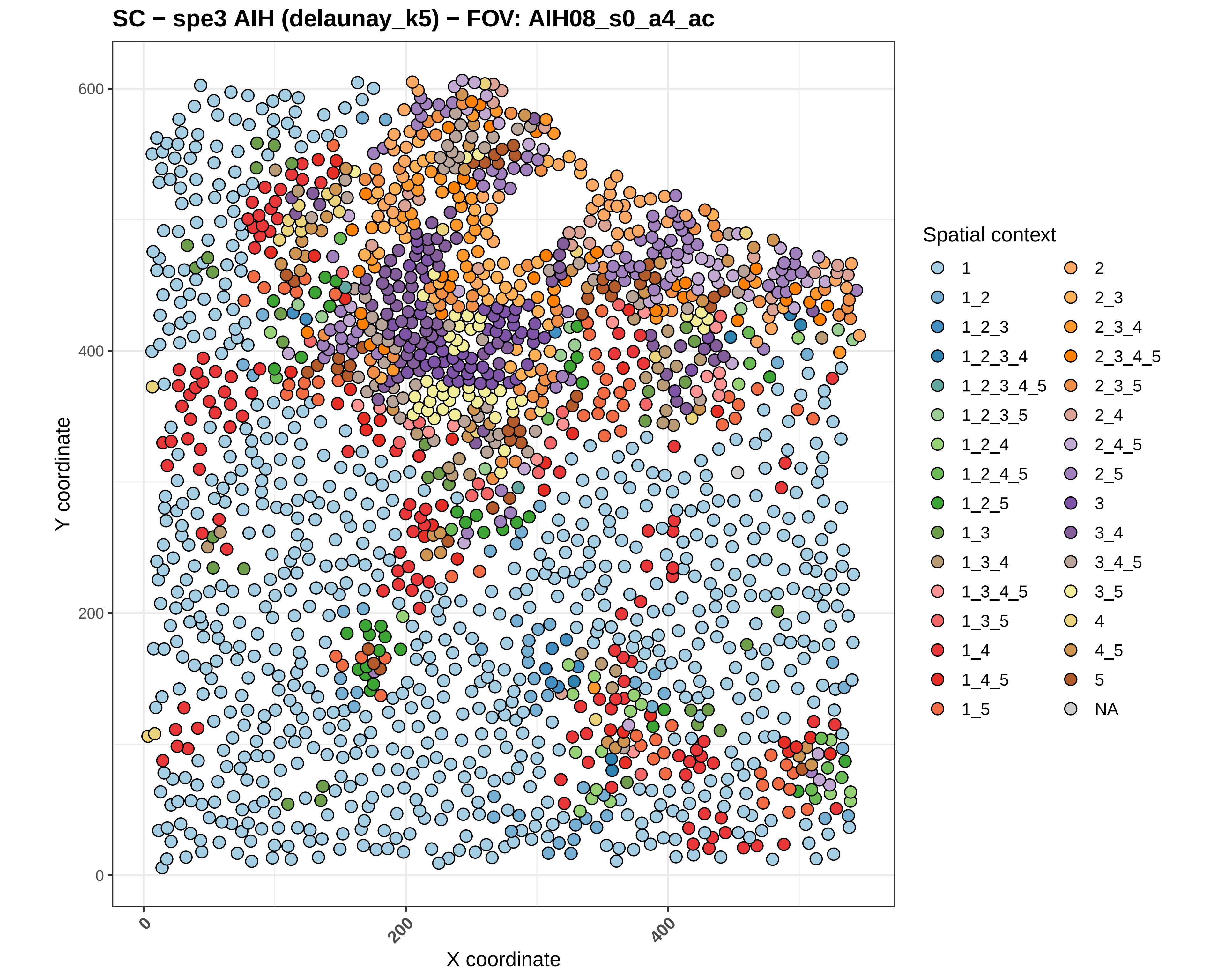
SC − spe3 AIH (delaunay_k5) − FOV: AIH08_s0_a4_ac
Y coordinate
X coordinate
0
200
400
600
0
200
400
Spatial context
1
1_2
1_2_3
1_2_3_4
1_2_3_4_5
1_2_3_5
1_2_4
1_2_4_5
1_2_5
1_3
1_3_4
1_3_4_5
1_3_5
1_4
1_4_5
1_5
2
2_3
2_3_4
2_3_4_5
2_3_5
2_4
2_4_5
2_5
3
3_4
3_4_5
3_5
4
4_5
5
NA

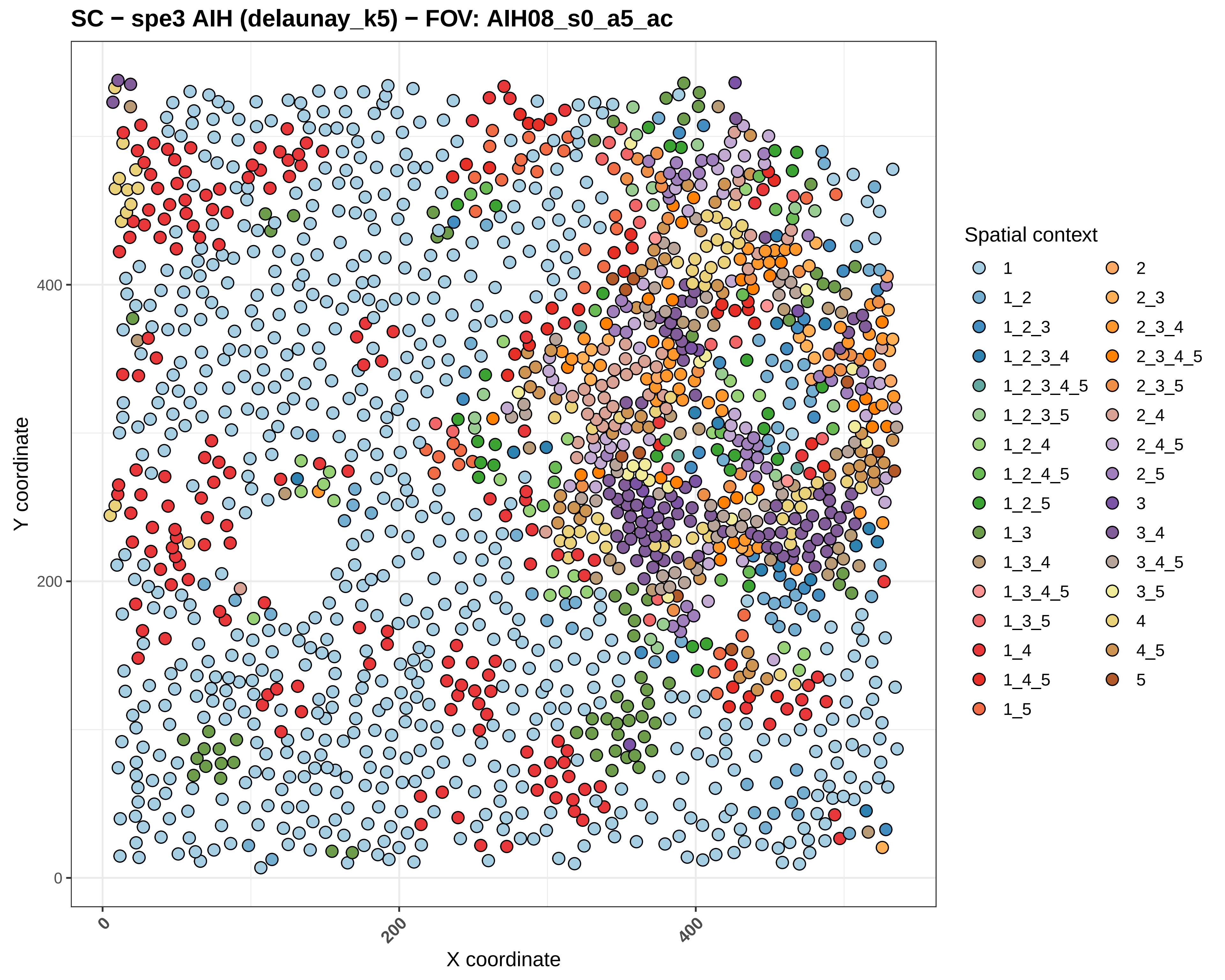
SC − spe3 AIH (delaunay_k5) − FOV: AIH08_s0_a5_ac
Y coordinate
X coordinate
0
200
400
Spatial context
1
1_2
1_2_3
1_2_3_4
1_2_3_4_5
1_2_3_5
1_2_4
1_2_4_5
1_2_5
1_3
1_3_4
1_3_4_5
1_3_5
1_4
1_4_5
1_5
2
2_3
2_3_4
2_3_4_5
2_3_5
2_4
2_4_5
2_5
3
3_4
3_4_5
3_5
4
4_5
5

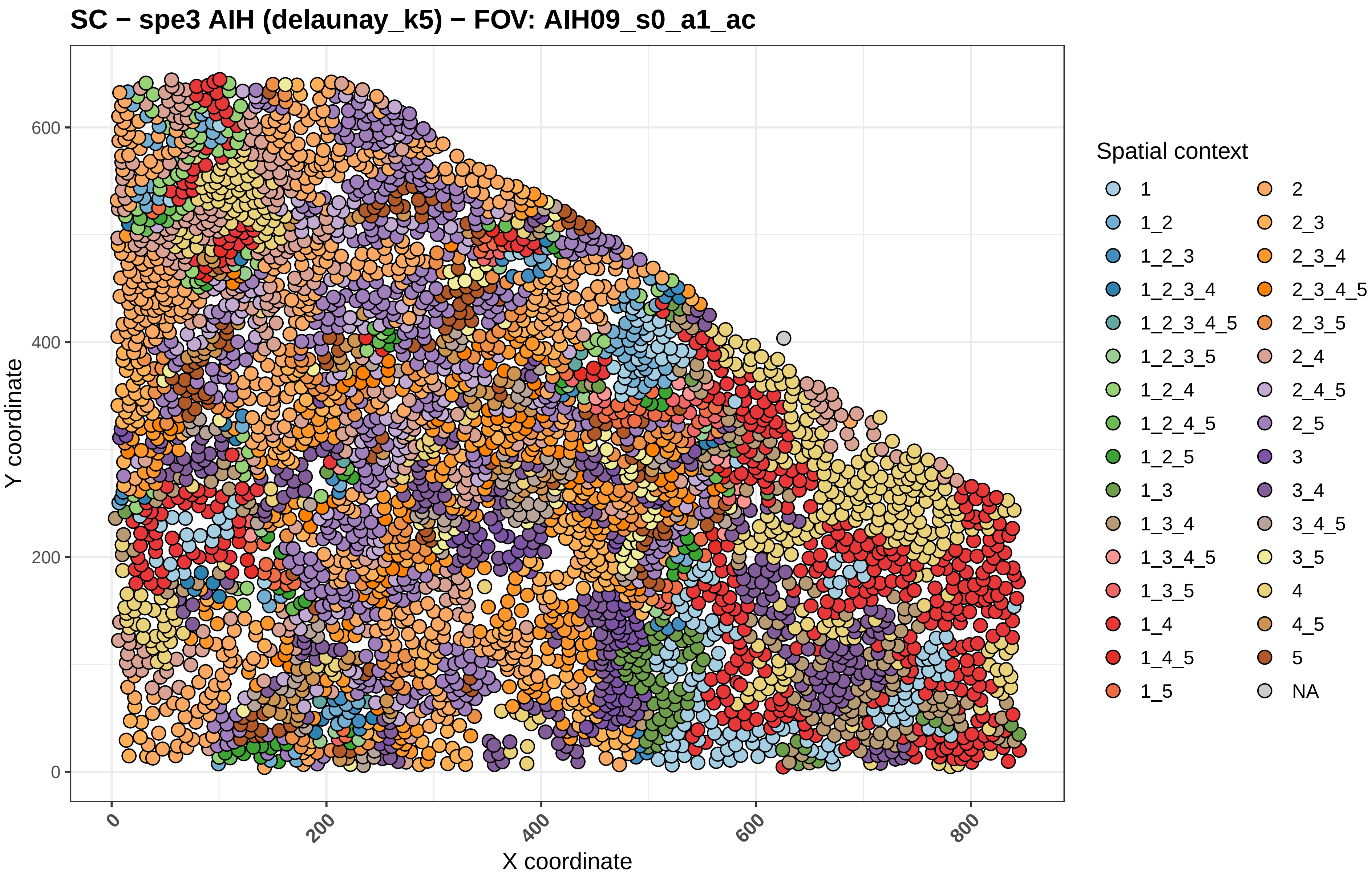
SC − spe3 AIH (delaunay_k5) − FOV: AIH09_s0_a1_ac
Y coordinate
X coordinate
0
200
400
600
800
Spatial context
1
1_2
1_2_3
1_2_3_4
1_2_3_4_5
1_2_3_5
1_2_4
1_2_4_5
1_2_5
1_3
1_3_4
1_3_4_5
1_3_5
1_4
1_4_5
1_5
2
2_3
2_3_4
2_3_4_5
2_3_5
2_4
2_4_5
2_5
3
3_4
3_4_5
3_5
4
4_5
5
NA

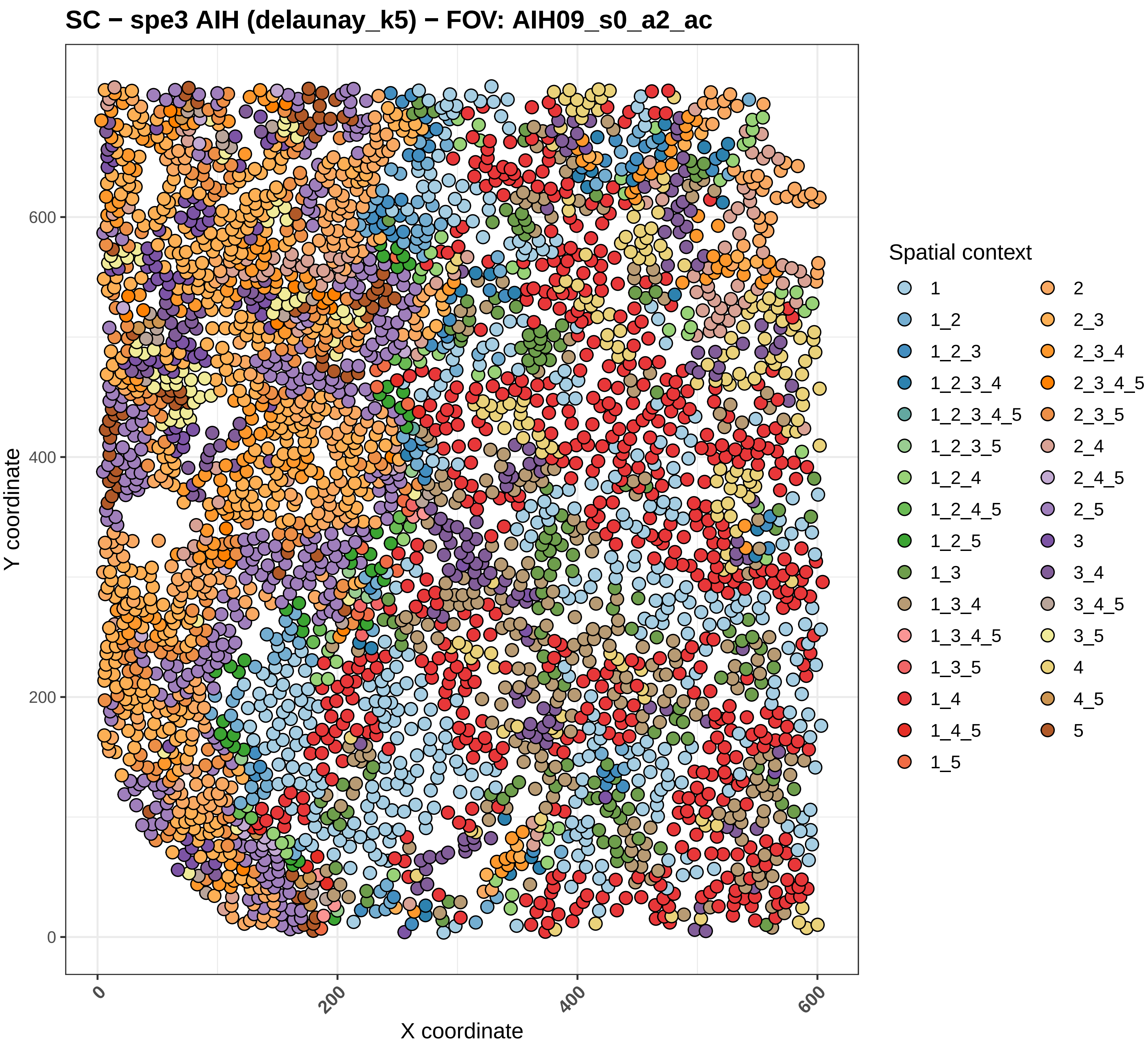

SC − spe3 AIH (delaunay_k5) − FOV: AIH09_s0_a2_ac
Y coordinate
X coordinate
0
200
400
600
Spatial context
1
1_2
1_2_3
1_2_3_4
1_2_3_4_5
1_2_3_5
1_2_4
1_2_4_5
1_2_5
1_3
1_3_4
1_3_4_5
1_3_5
1_4
1_4_5
1_5
2
2_3
2_3_4
2_3_4_5
2_3_5
2_4
2_4_5
2_5
3
3_4
3_4_5
3_5
4
4_5
5

#### SC − spe3 AIH (delaunay_k5) − FOV: AIH09_s0_a3_ac

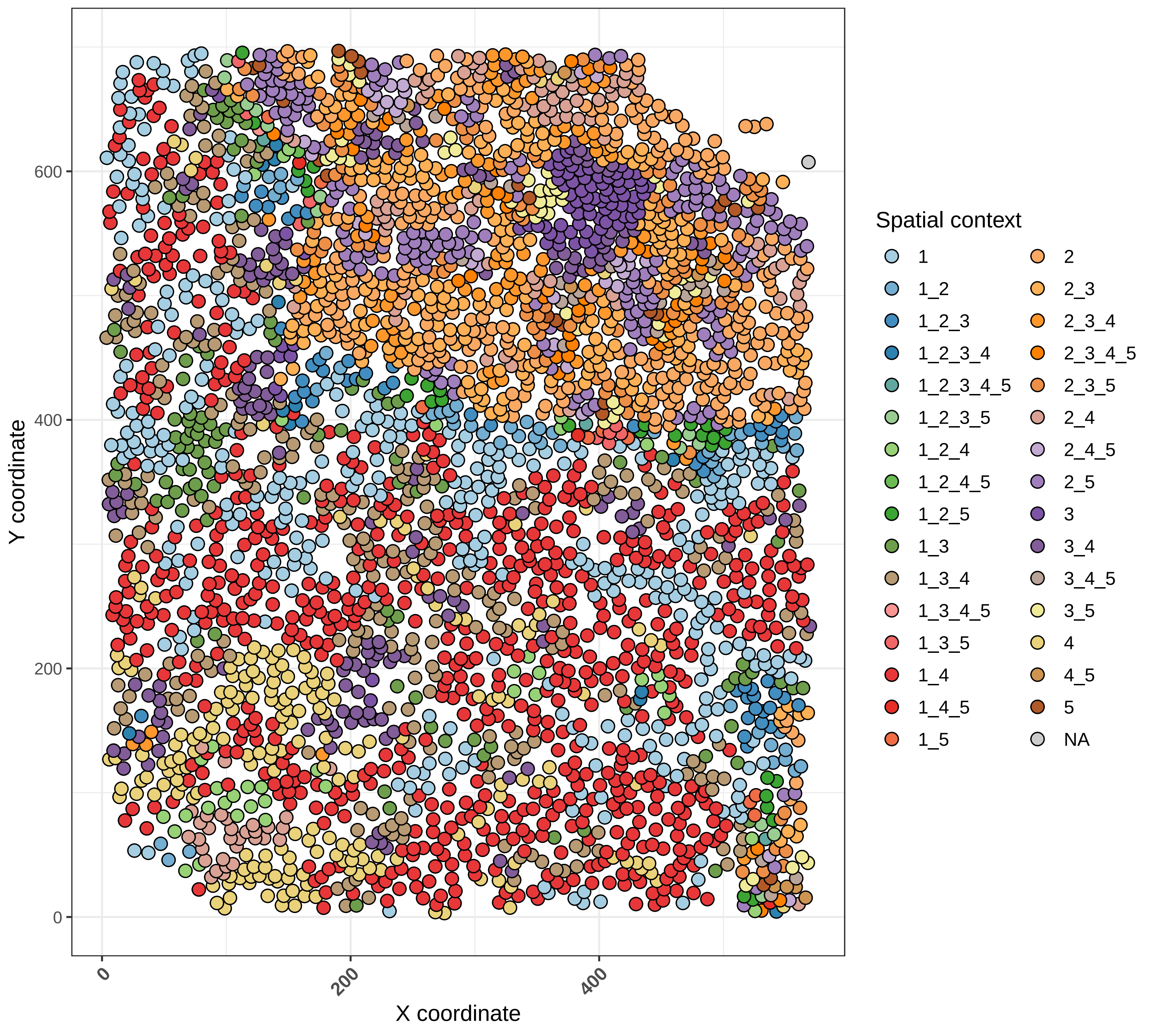

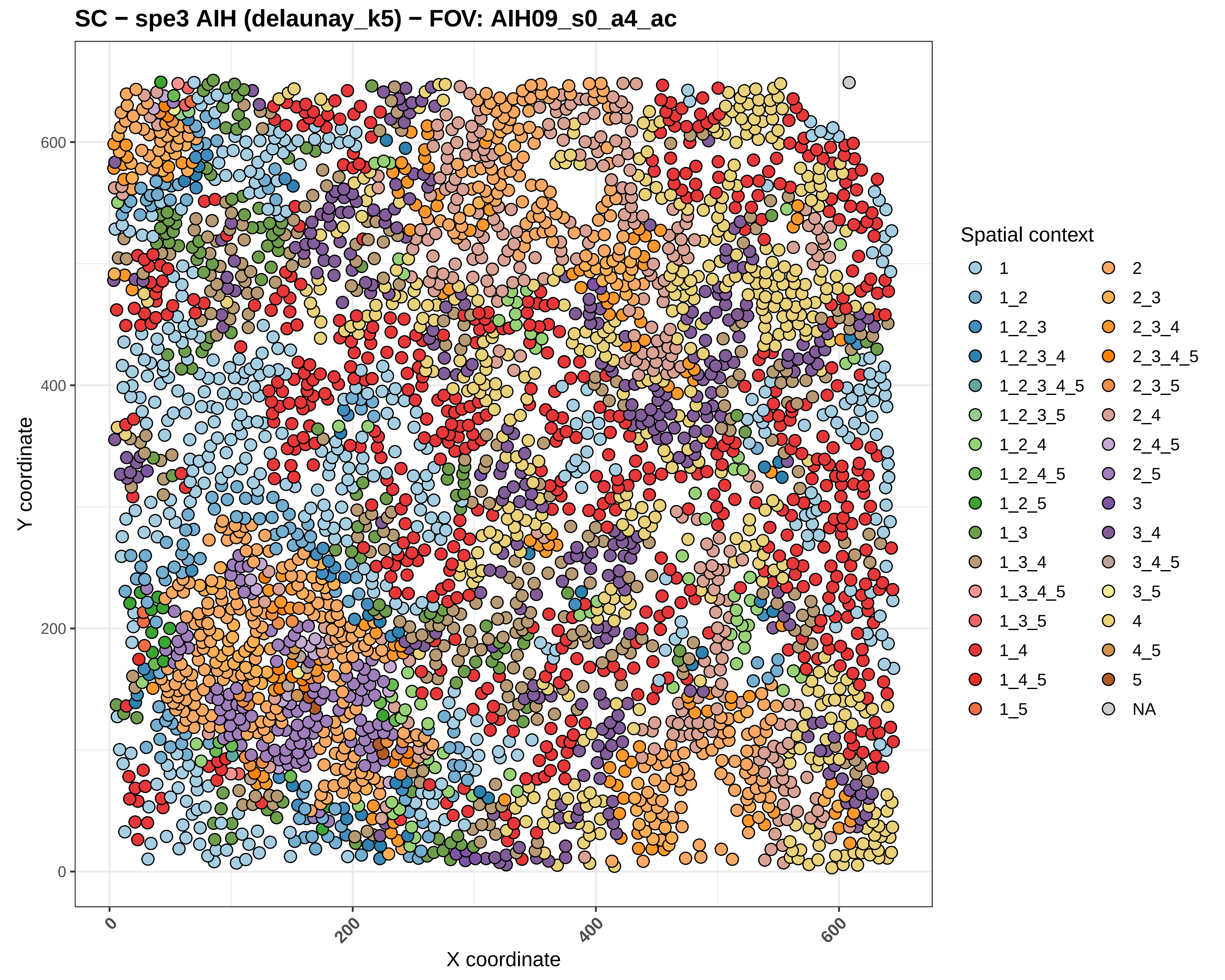

SC − spe3 AIH (delaunay_k5) − FOV: AIH09_s0_a4_ac
Y coordinate
X coordinate
0
200
400
600
Spatial context
1
1_2
1_2_3
1_2_3_4
1_2_3_4_5
1_2_3_5
1_2_4
1_2_4_5
1_2_5
1_3
1_3_4
1_3_4_5
1_3_5
1_4
1_4_5
1_5
2
2_3
2_3_4
2_3_4_5
2_3_5
2_4
2_4_5
2_5
3
3_4
3_4_5
3_5
4
4_5
5
NA

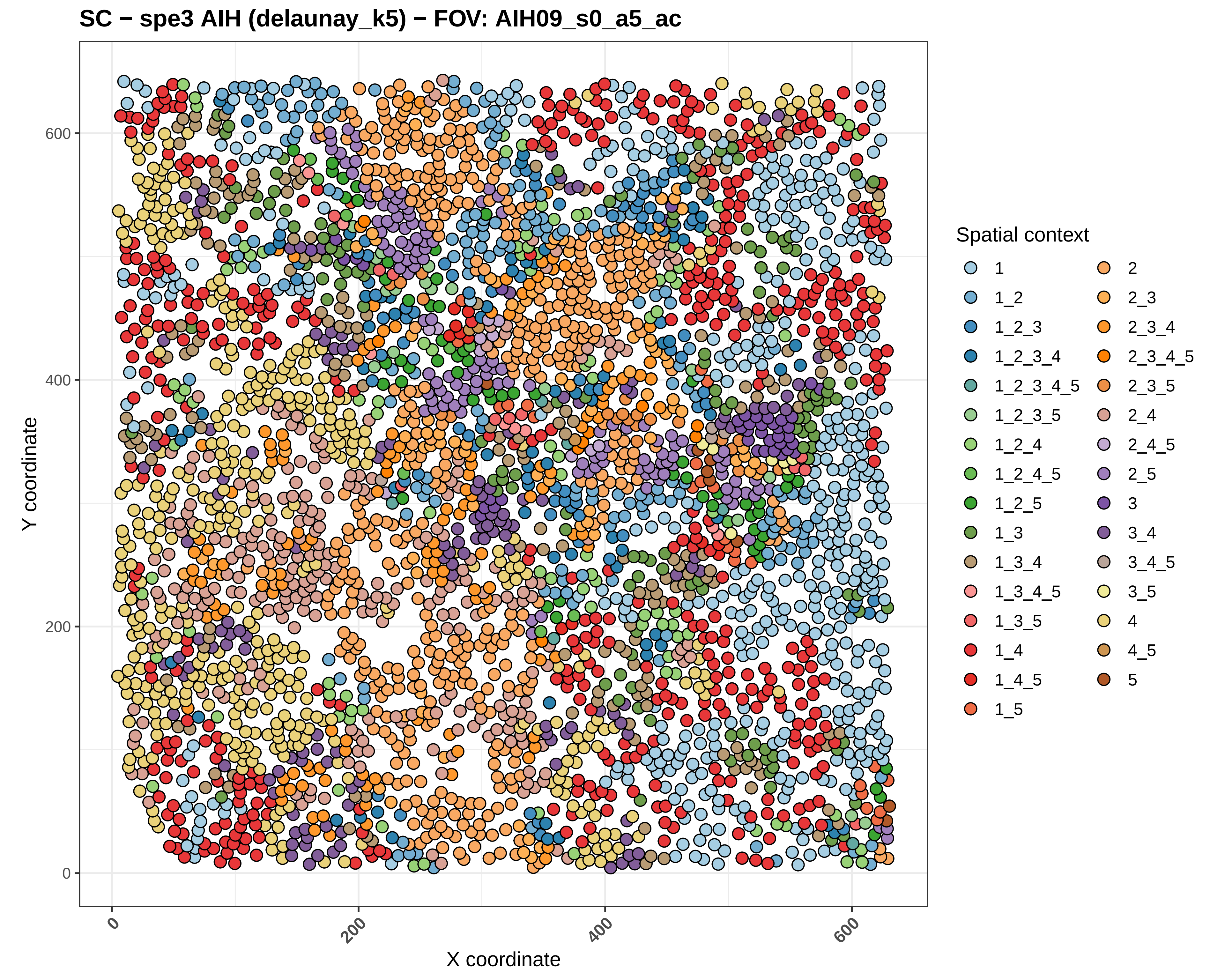
SC − spe3 AIH (delaunay_k5) − FOV: AIH09_s0_a5_ac
Y coordinate
X coordinate
0
200
400
600
Spatial context
1
1_2
1_2_3
1_2_3_4
1_2_3_4_5
1_2_3_5
1_2_4
1_2_4_5
1_2_5
1_3
1_3_4
1_3_4_5
1_3_5
1_4
1_4_5
1_5
2
2_3
2_3_4
2_3_4_5
2_3_5
2_4
2_4_5
2_5
3
3_4
3_4_5
3_5
4
4_5
5

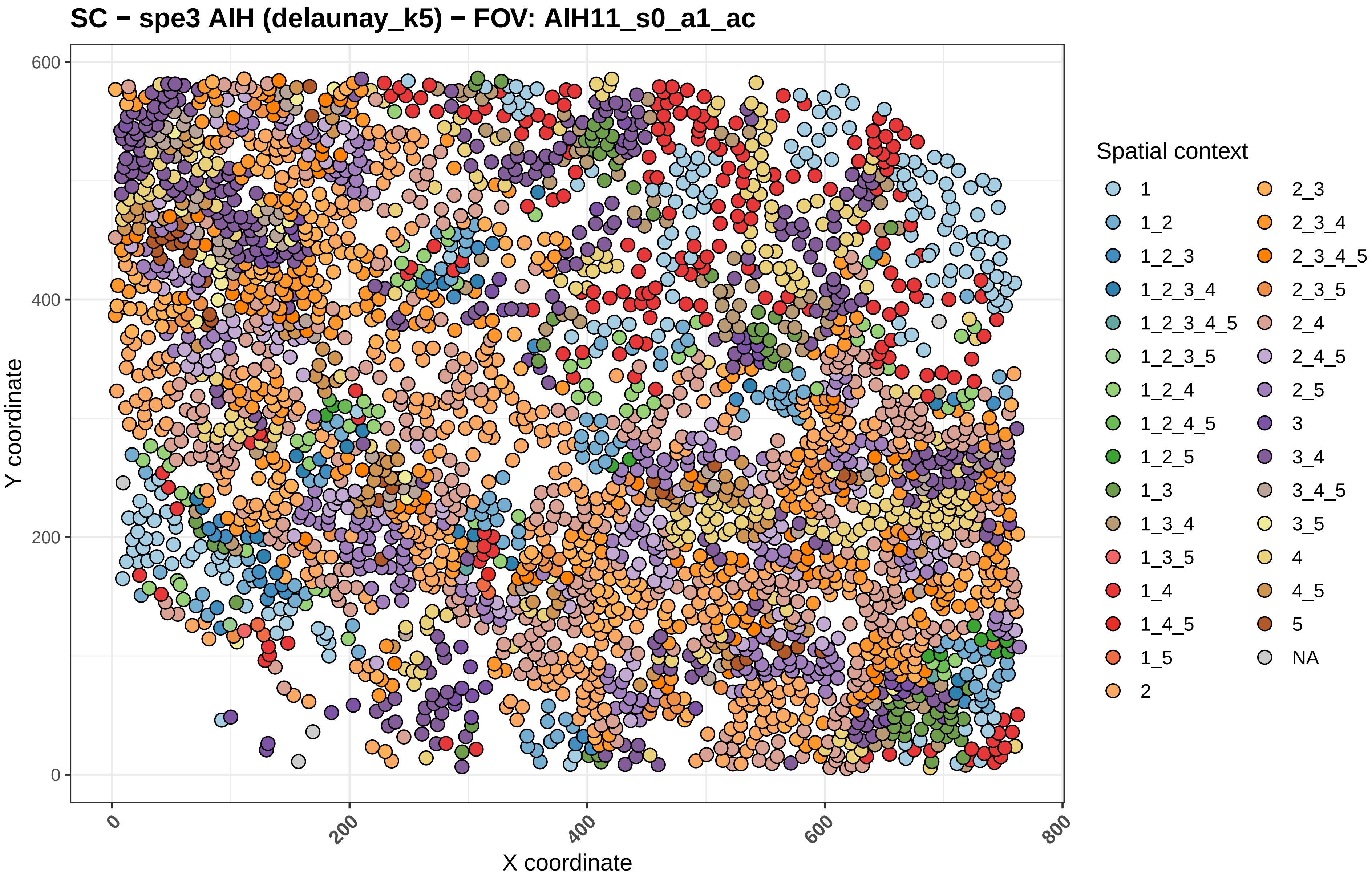

SC − spe3 AIH (delaunay_k5) − FOV: AIH11_s0_a1_ac
Y coordinate
X coordinate
0
200
400
600
800
Spatial context
1
1_2
1_2_3
1_2_3_4
1_2_3_4_5
1_2_3_5
1_2_4
1_2_4_5
1_2_5
1_3
1_3_4
1_3_5
1_4
1_4_5
1_5
2
2_3
2_3_4
2_3_4_5
2_3_5
2_4
2_4_5
2_5
3
3_4
3_4_5
3_5
4
4_5
5
NA

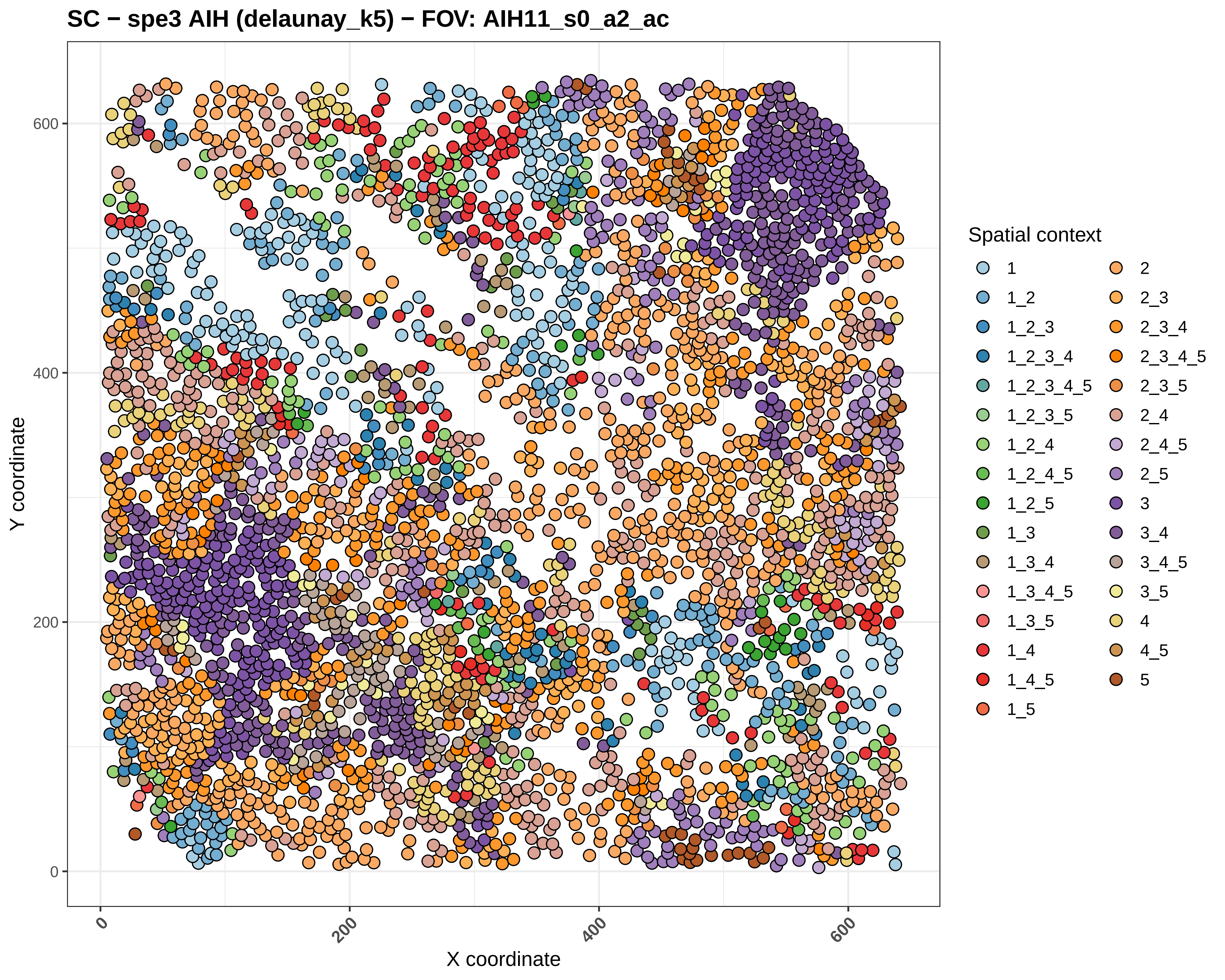
SC − spe3 AIH (delaunay_k5) − FOV: AIH11_s0_a2_ac
Y coordinate
X coordinate
0
200
400
600
Spatial context
1
1_2
1_2_3
1_2_3_4
1_2_3_4_5
1_2_3_5
1_2_4
1_2_4_5
1_2_5
1_3
1_3_4
1_3_4_5
1_3_5
1_4
1_4_5
1_5
2
2_3
2_3_4
2_3_4_5
2_3_5
2_4
2_4_5
2_5
3
3_4
3_4_5
3_5
4
4_5
5

#### SC − spe3 AIH (delaunay_k5) − FOV: AIH11_s0_a3_ac

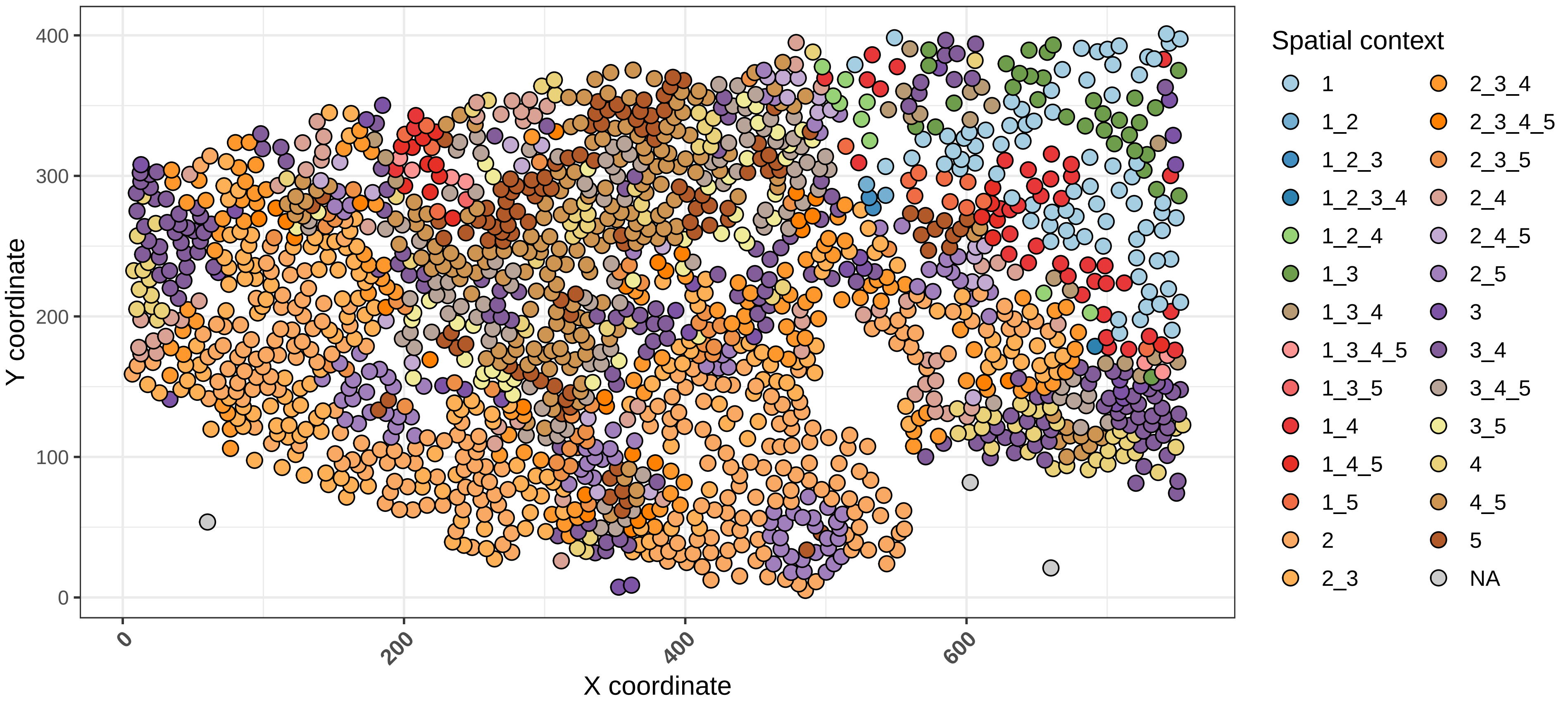

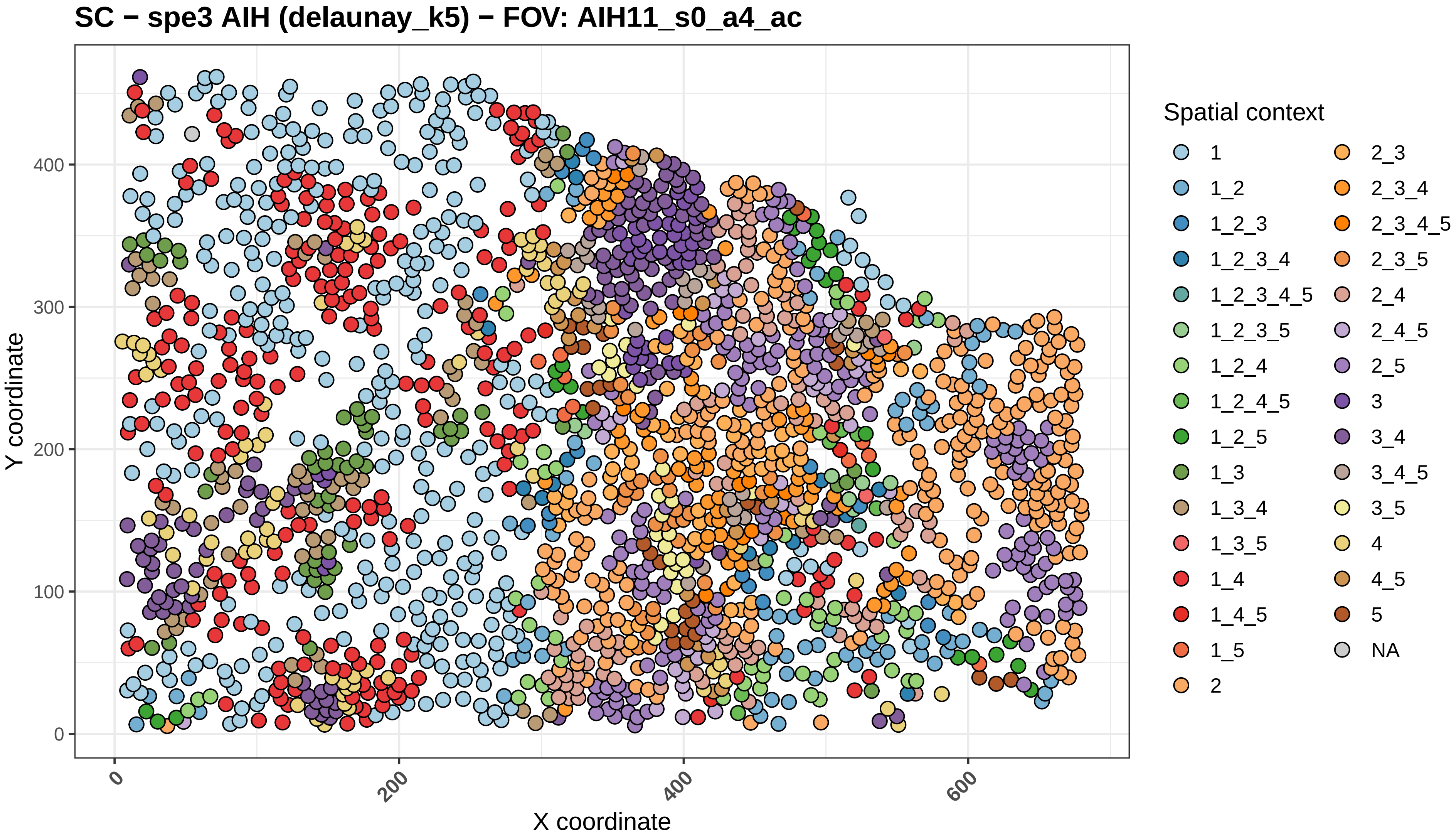

SC − spe3 AIH (delaunay_k5) − FOV: AIH11_s0_a4_ac
Y coordinate
X coordinate
0
100
200
300
400
0
200
400
600
Spatial context
1
1_2
1_2_3
1_2_3_4
1_2_3_4_5
1_2_3_5
1_2_4
1_2_4_5
1_2_5
1_3
1_3_4
1_3_5
1_4
1_4_5
1_5
2
2_3
2_3_4
2_3_4_5
2_3_5
2_4
2_4_5
2_5
3
3_4
3_4_5
3_5
4
4_5
5
NA

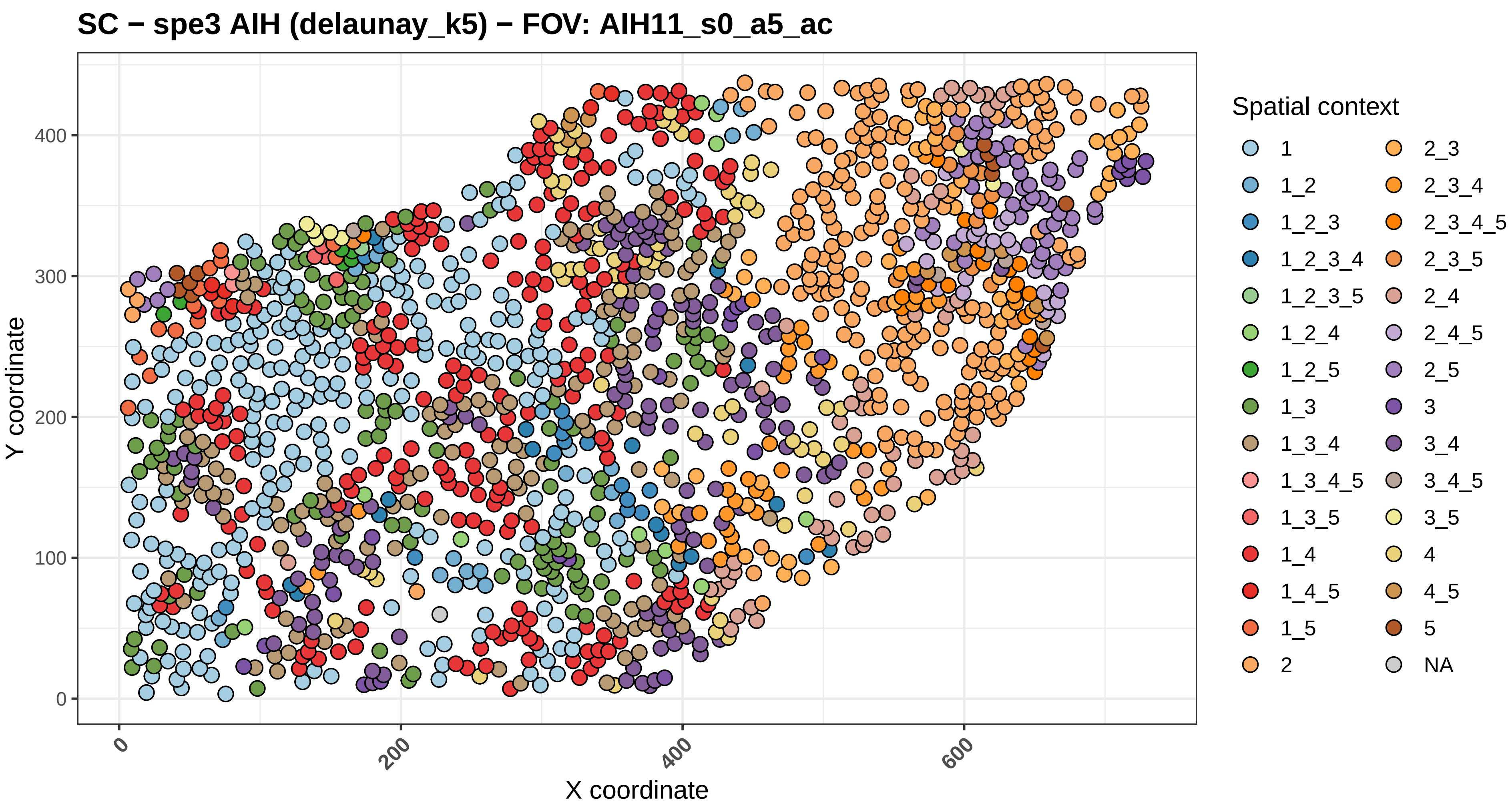

SC − spe3 AIH (delaunay_k5) − FOV: AIH11_s0_a5_ac
Y coordinate
X coordinate
0
100
200
300
400
0
200
400
600
Spatial context
1
1_2
1_2_3
1_2_3_4
1_2_3_5
1_2_4
1_2_5
1_3
1_3_4
1_3_4_5
1_3_5
1_4
1_4_5
1_5
2
2_3
2_3_4
2_3_4_5
2_3_5
2_4
2_4_5
2_5
3
3_4
3_4_5
3_5
4
4_5
5
NA

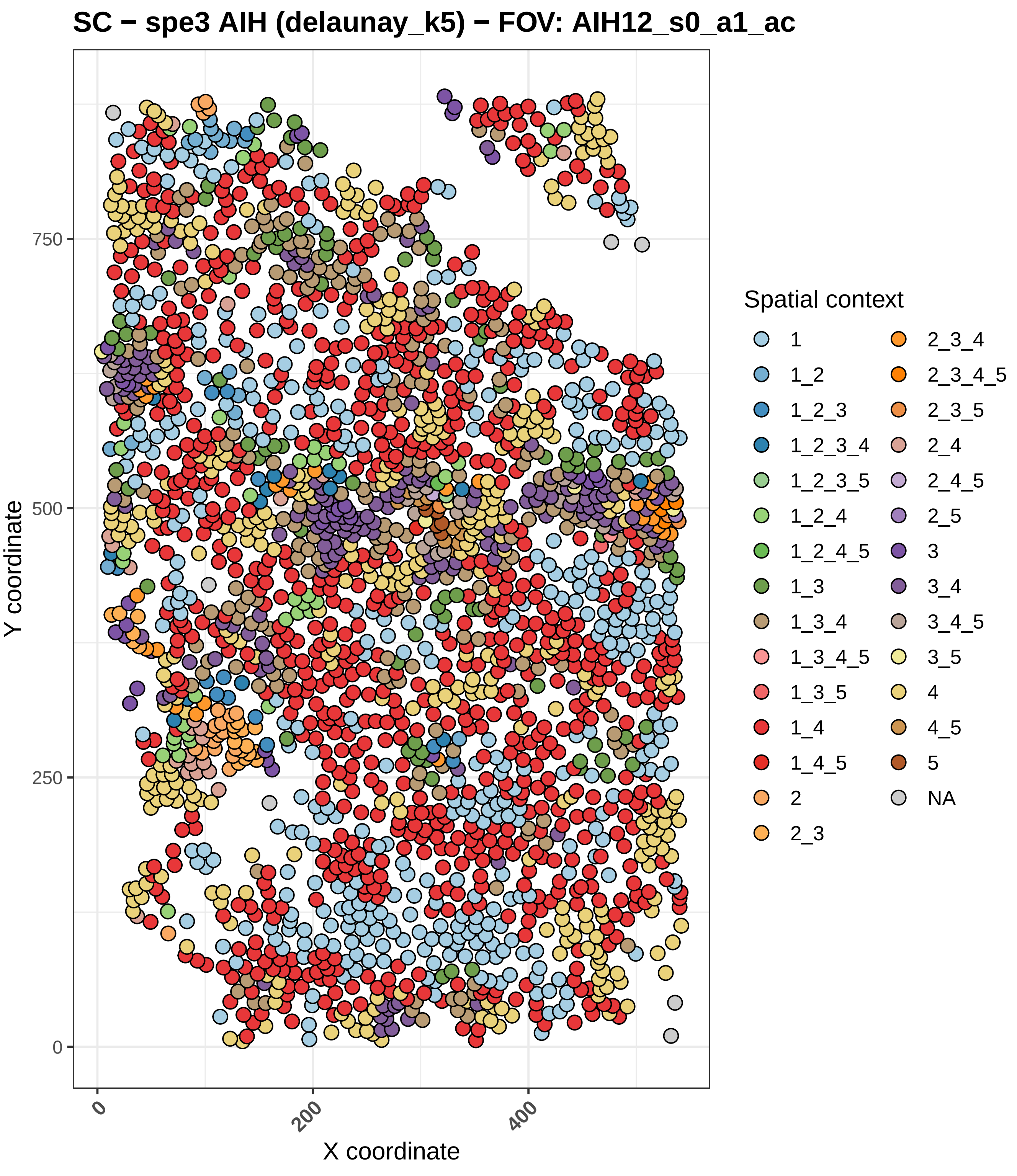

SC − spe3 AIH (delaunay_k5) − FOV: AIH12_s0_a1_ac
Y coordinate
X coordinate
0
250
500
750
0
200
400
Spatial context
1
1_2
1_2_3
1_2_3_4
1_2_3_5
1_2_4
1_2_4_5
1_3
1_3_4
1_3_4_5
1_3_5
1_4
1_4_5
2
2_3
2_3_4
2_3_4_5
2_3_5
2_4
2_4_5
2_5
3
3_4
3_4_5
3_5
4
4_5
5
NA

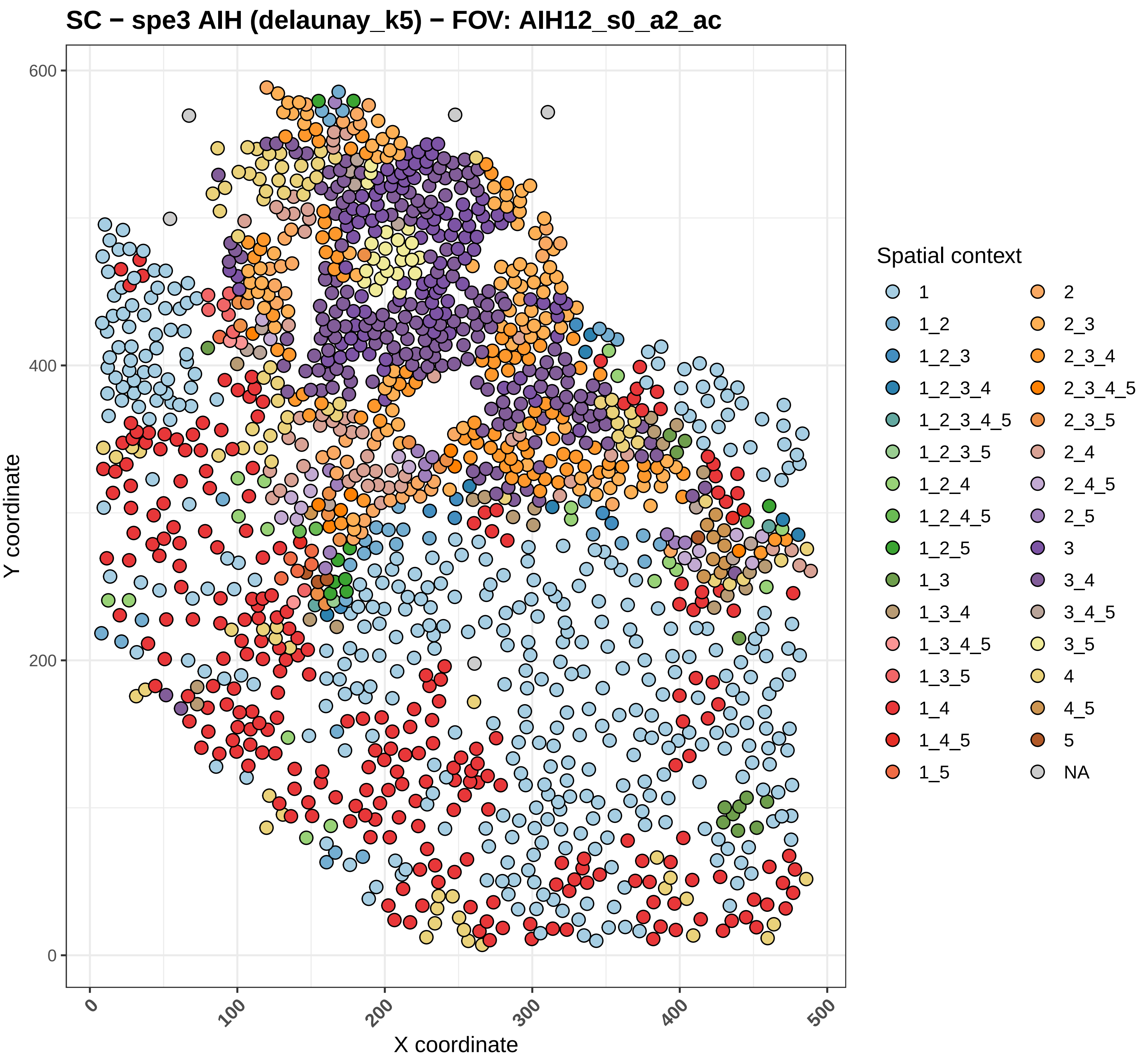

SC − spe3 AIH (delaunay_k5) − FOV: AIH12_s0_a2_ac
Y coordinate
X coordinate
0
200
400
600
0
100
200
300
400
500
Spatial context
1
1_2
1_2_3
1_2_3_4
1_2_3_4_5
1_2_3_5
1_2_4
1_2_4_5
1_2_5
1_3
1_3_4
1_3_4_5
1_3_5
1_4
1_4_5
1_5
2
2_3
2_3_4
2_3_4_5
2_3_5
2_4
2_4_5
2_5
3
3_4
3_4_5
3_5
4
4_5
5
NA

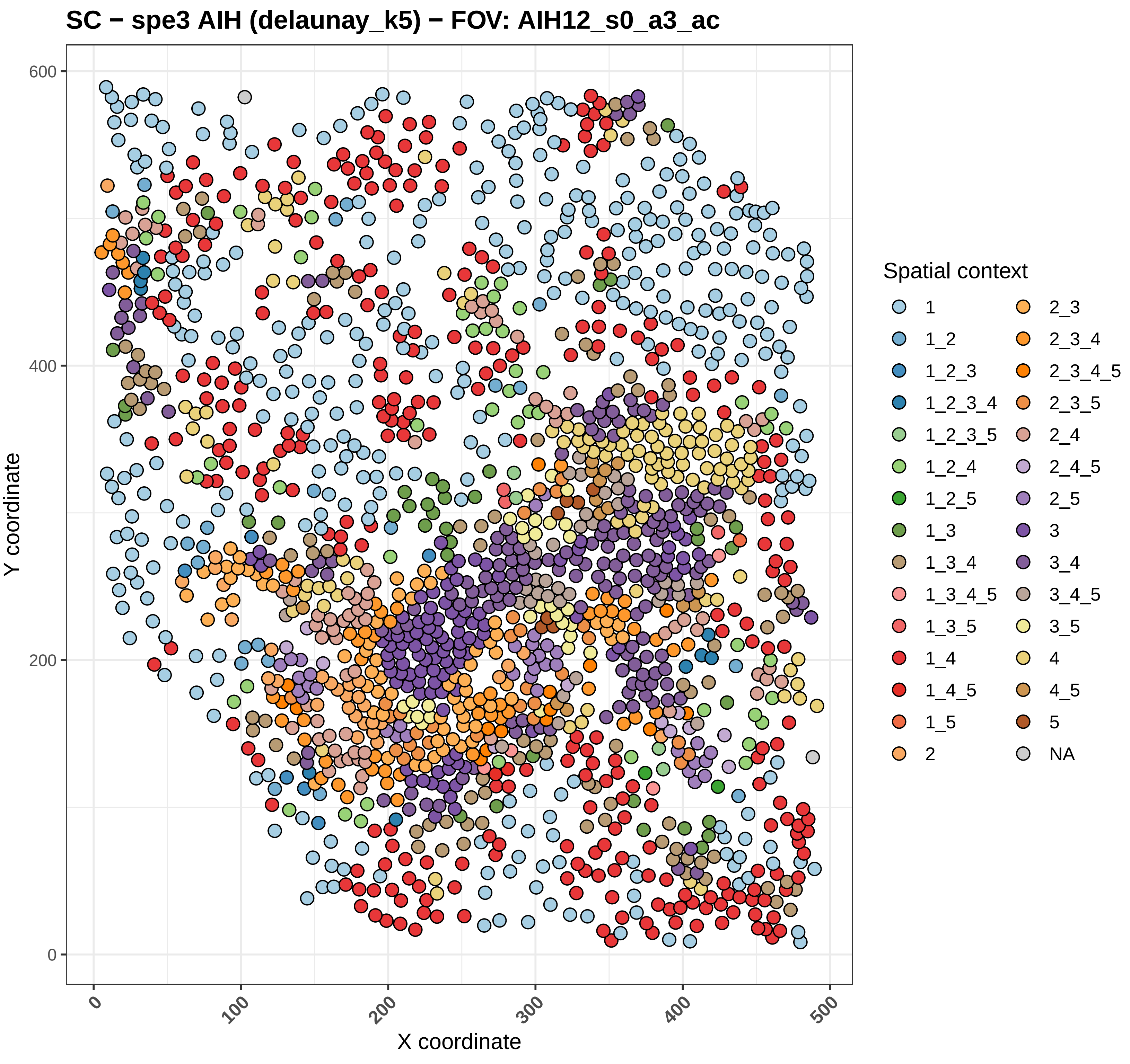
SC − spe3 AIH (delaunay_k5) − FOV: AIH12_s0_a3_ac
Y coordinate
X coordinate
0
100
200
300
400
500
0
200
400
600
Spatial context
1
1_2
1_2_3
1_2_3_4
1_2_3_5
1_2_4
1_2_5
1_3
1_3_4
1_3_4_5
1_3_5
1_4
1_4_5
1_5
2
2_3
2_3_4
2_3_4_5
2_3_5
2_4
2_4_5
2_5
3
3_4
3_4_5
3_5
4
4_5
5
NA

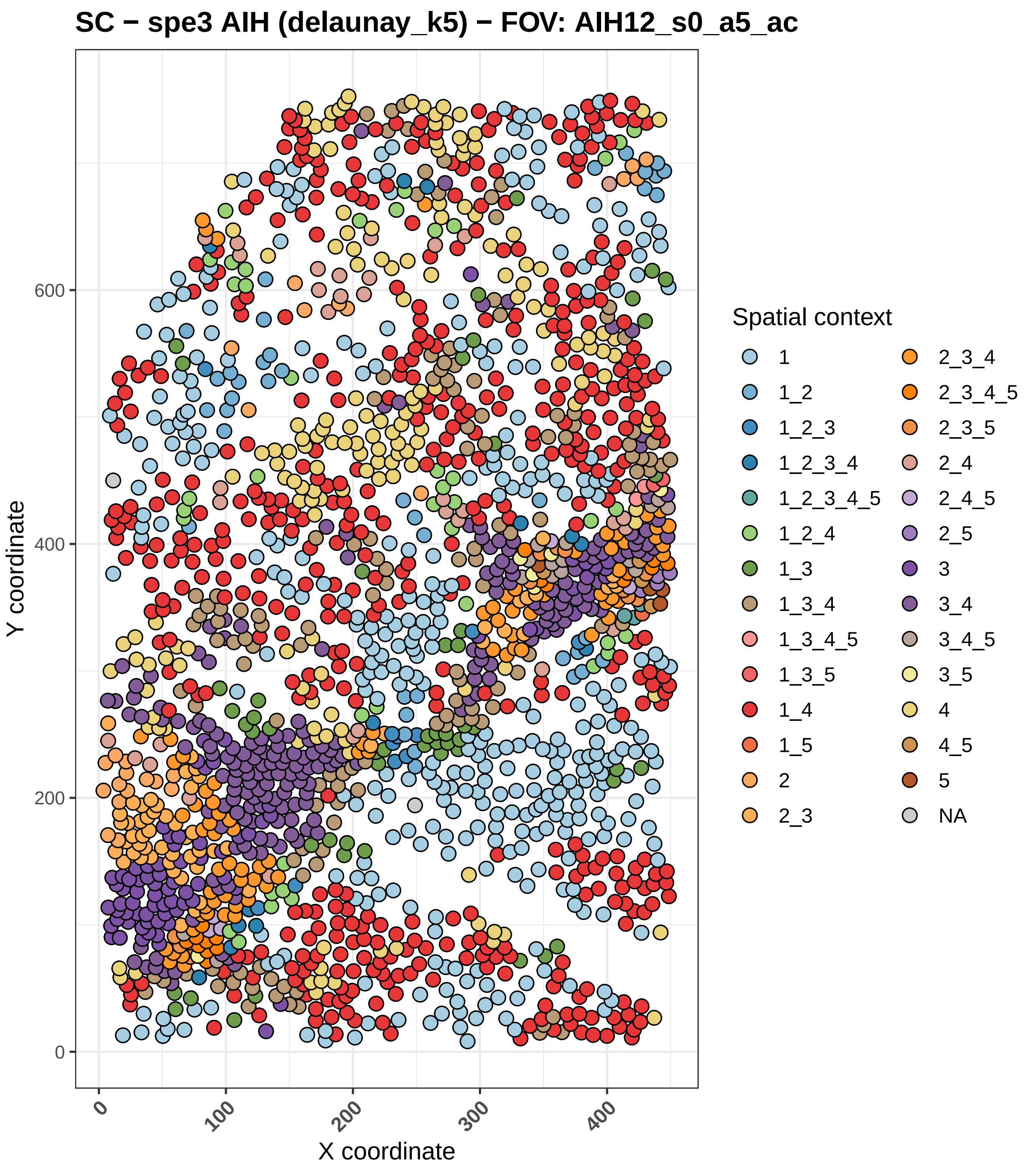
SC − spe3 AIH (delaunay_k5) − FOV: AIH12_s0_a5_ac
Y coordinate
X coordinate
0
200
400
600
0
100
200
300
400
Spatial context
1
1_2
1_2_3
1_2_3_4
1_2_3_4_5
1_2_4
1_3
1_3_4
1_3_4_5
1_3_5
1_4
1_5
2
2_3
2_3_4
2_3_4_5
2_3_5
2_4
2_4_5
2_5
3
3_4
3_4_5
3_5
4
4_5
5
NA

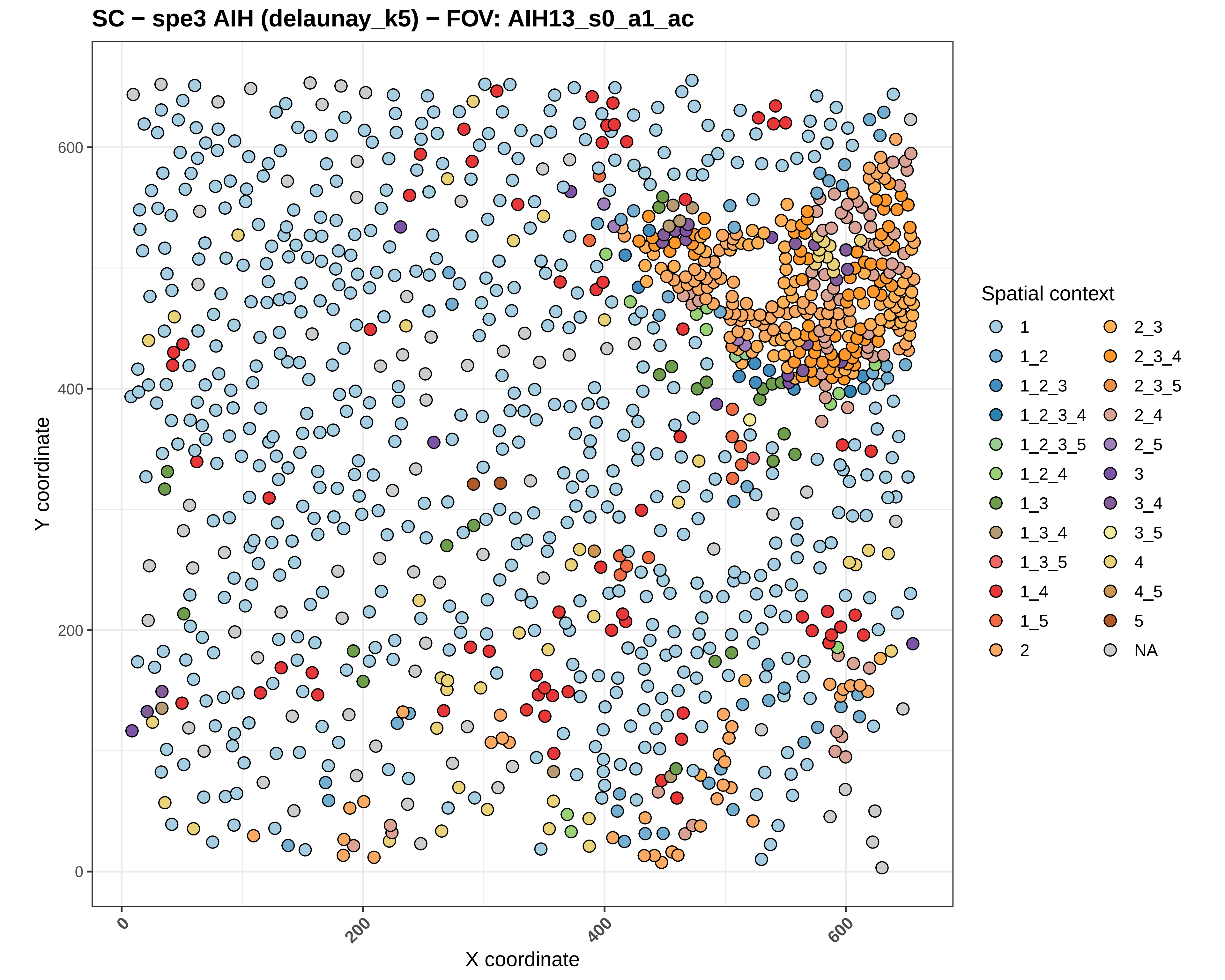
SC − spe3 AIH (delaunay_k5) − FOV: AIH13_s0_a1_ac
Y coordinate
X coordinate
0
200
400
600
Spatial context
1
1_2
1_2_3
1_2_3_4
1_2_3_5
1_2_4
1_3
1_3_4
1_3_5
1_4
1_5
2
2_3
2_3_4
2_3_5
2_4
2_5
3
3_4
3_5
4
4_5
5
NA

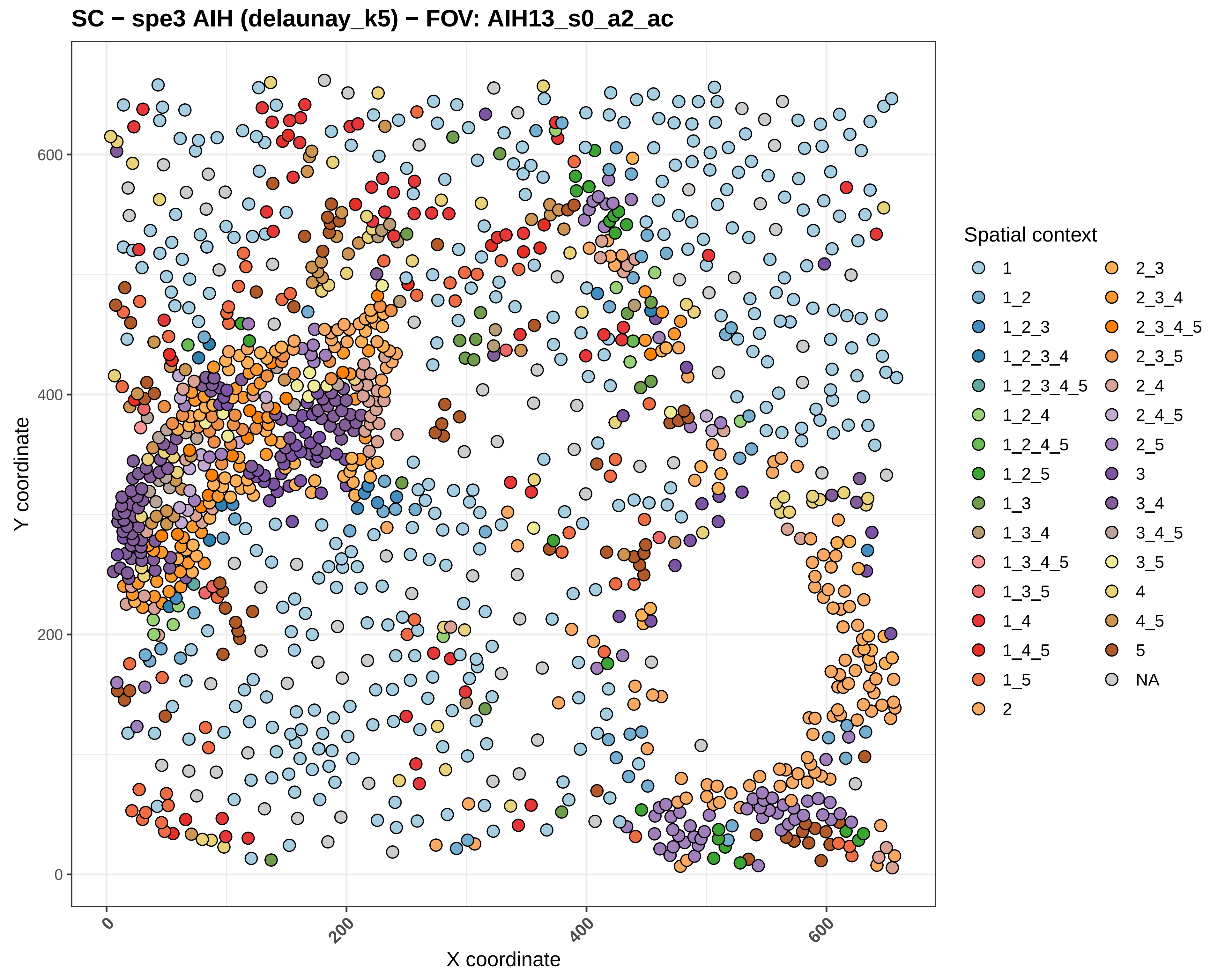

SC − spe3 AIH (delaunay_k5) − FOV: AIH13_s0_a2_ac
Y coordinate
X coordinate
0
200
400
600
Spatial context
1
1_2
1_2_3
1_2_3_4
1_2_3_4_5
1_2_4
1_2_4_5
1_2_5
1_3
1_3_4
1_3_4_5
1_3_5
1_4
1_4_5
1_5
2
2_3
2_3_4
2_3_4_5
2_3_5
2_4
2_4_5
2_5
3
3_4
3_4_5
3_5
4
4_5
5
NA

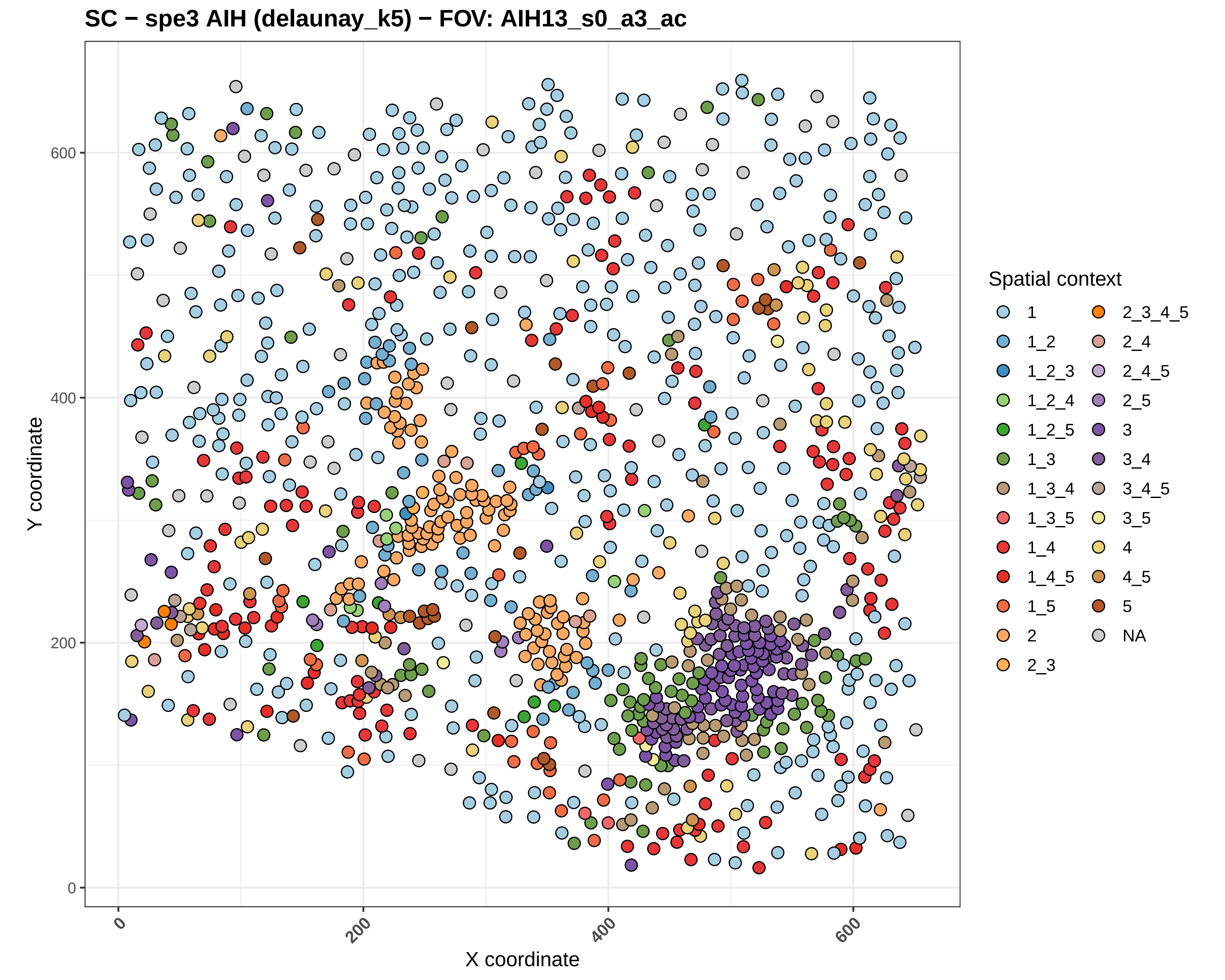
SC − spe3 AIH (delaunay_k5) − FOV: AIH13_s0_a3_ac
Y coordinate
X coordinate
0
200
400
600
Spatial context
1
1_2
1_2_3
1_2_4
1_2_5
1_3
1_3_4
1_3_5
1_4
1_4_5
1_5
2
2_3
2_3_4_5
2_4
2_4_5
2_5
3
3_4
3_4_5
3_5
4
4_5
5
NA

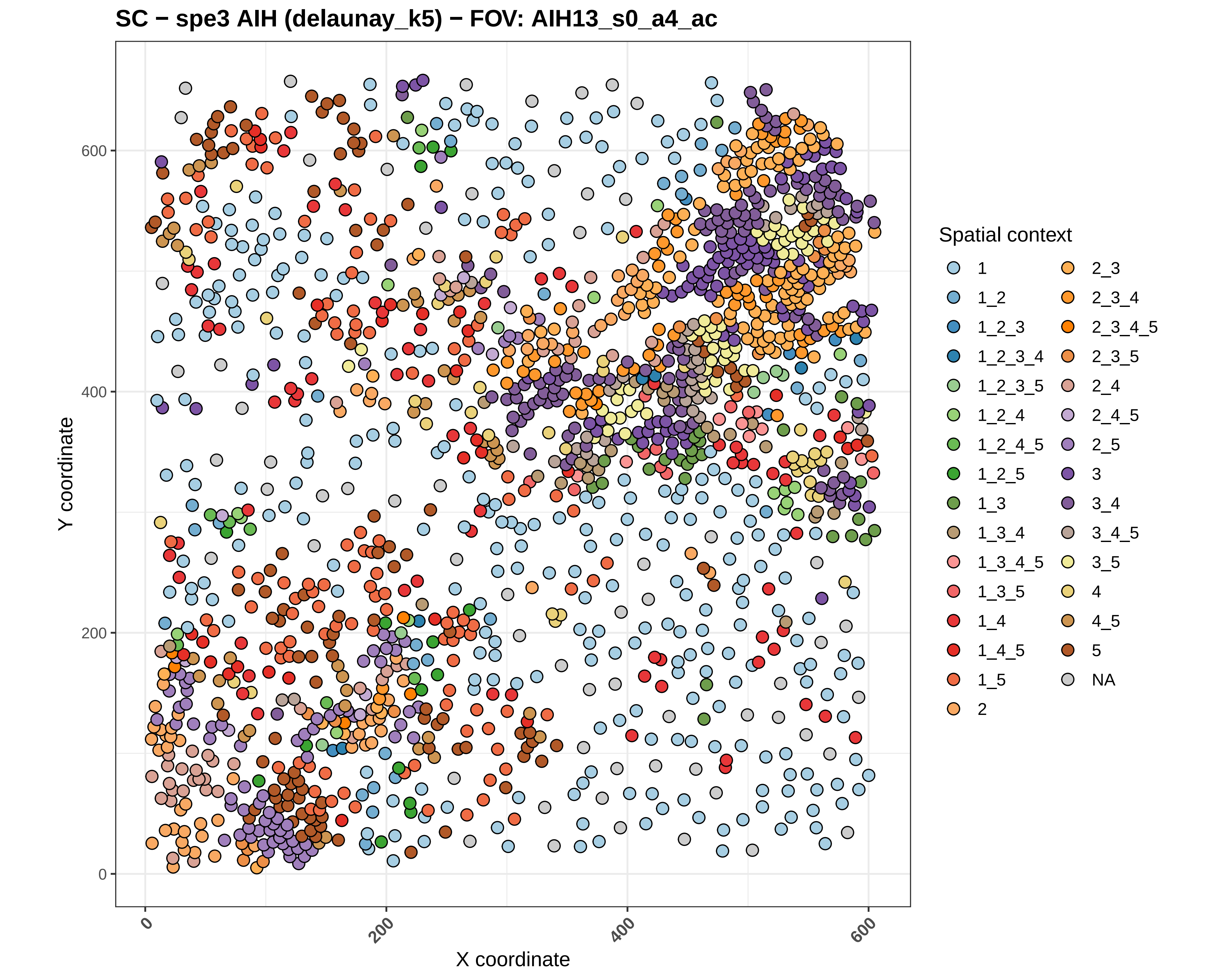
SC − spe3 AIH (delaunay_k5) − FOV: AIH13_s0_a4_ac
Y coordinate
X coordinate
0
200
400
600
Spatial context
1
1_2
1_2_3
1_2_3_4
1_2_3_5
1_2_4
1_2_4_5
1_2_5
1_3
1_3_4
1_3_4_5
1_3_5
1_4
1_4_5
1_5
2
2_3
2_3_4
2_3_4_5
2_3_5
2_4
2_4_5
2_5
3
3_4
3_4_5
3_5
4
4_5
5
NA

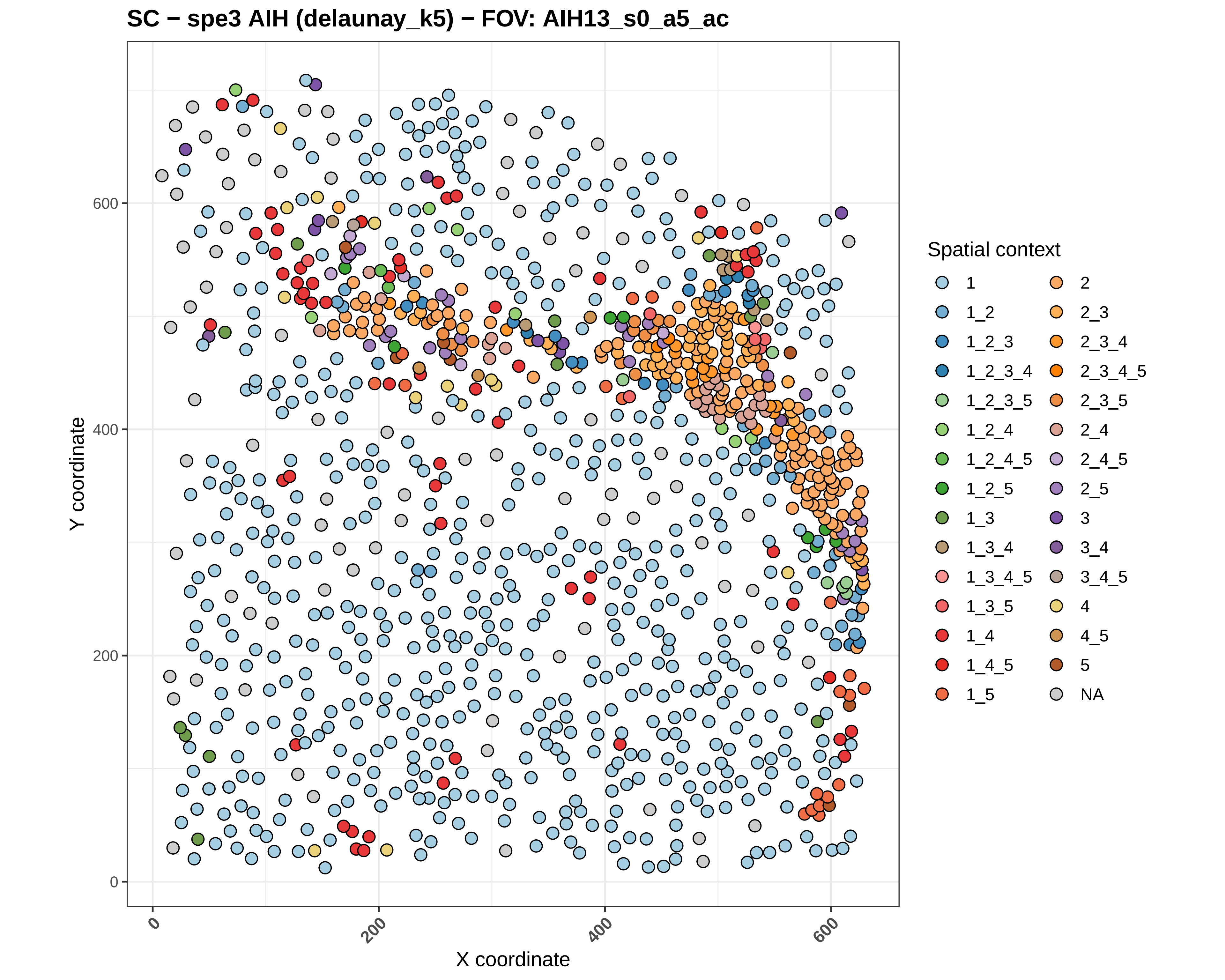
SC − spe3 AIH (delaunay_k5) − FOV: AIH13_s0_a5_ac
Y coordinate
X coordinate
0
200
400
600
Spatial context
1
1_2
1_2_3
1_2_3_4
1_2_3_5
1_2_4
1_2_4_5
1_2_5
1_3
1_3_4
1_3_4_5
1_3_5
1_4
1_4_5
1_5
2
2_3
2_3_4
2_3_4_5
2_3_5
2_4
2_4_5
2_5
3
3_4
3_4_5
4
4_5
5
NA

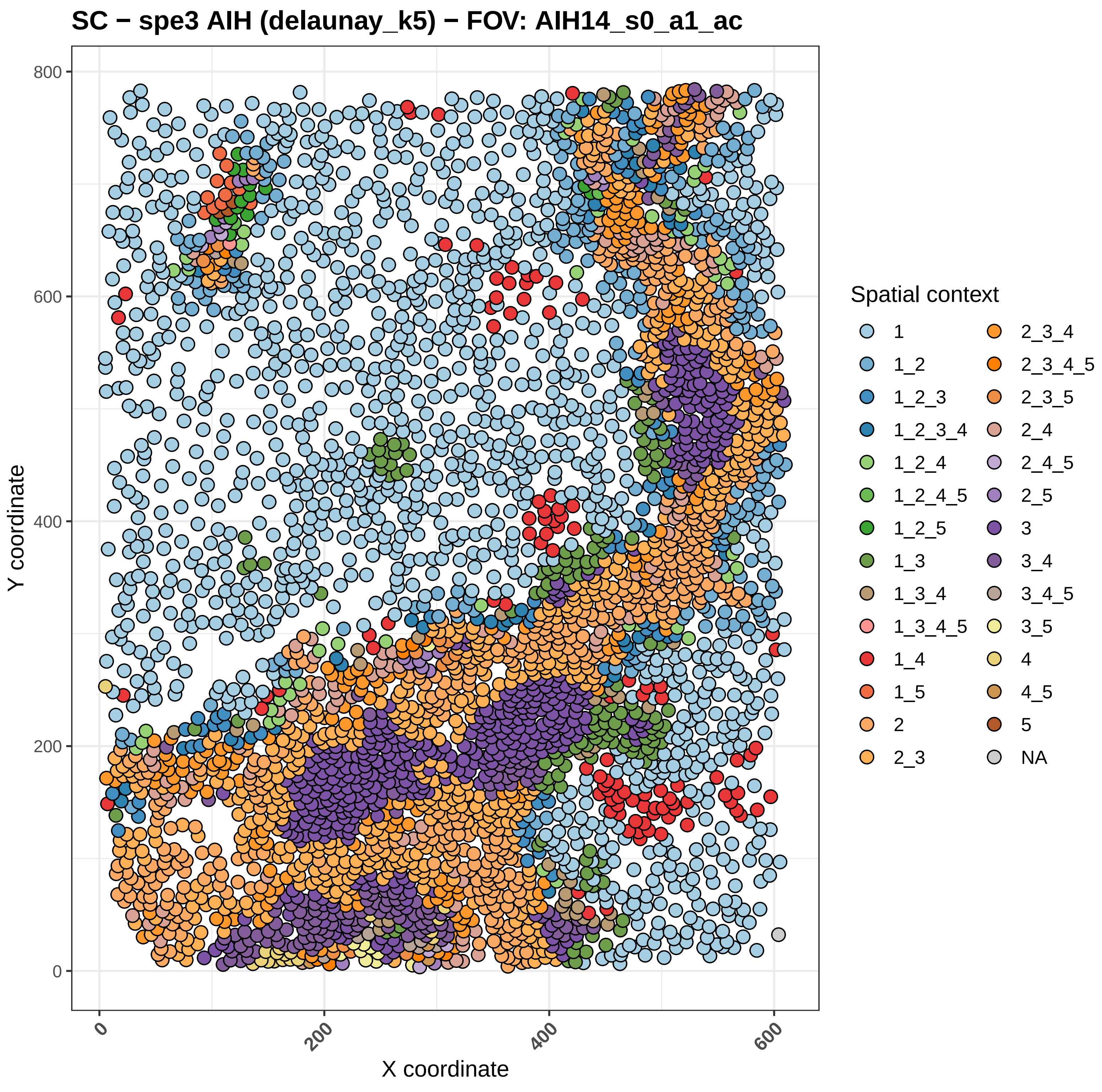

SC − spe3 AIH (delaunay_k5) − FOV: AIH14_s0_a1_ac
Y coordinate
X coordinate
0
200
400
600
800
Spatial context
1
1_2
1_2_3
1_2_3_4
1_2_4
1_2_4_5
1_2_5
1_3
1_3_4
1_3_4_5
1_4
1_5
2
2_3
2_3_4
2_3_4_5
2_3_5
2_4
2_4_5
2_5
3
3_4
3_4_5
3_5
4
4_5
5
NA

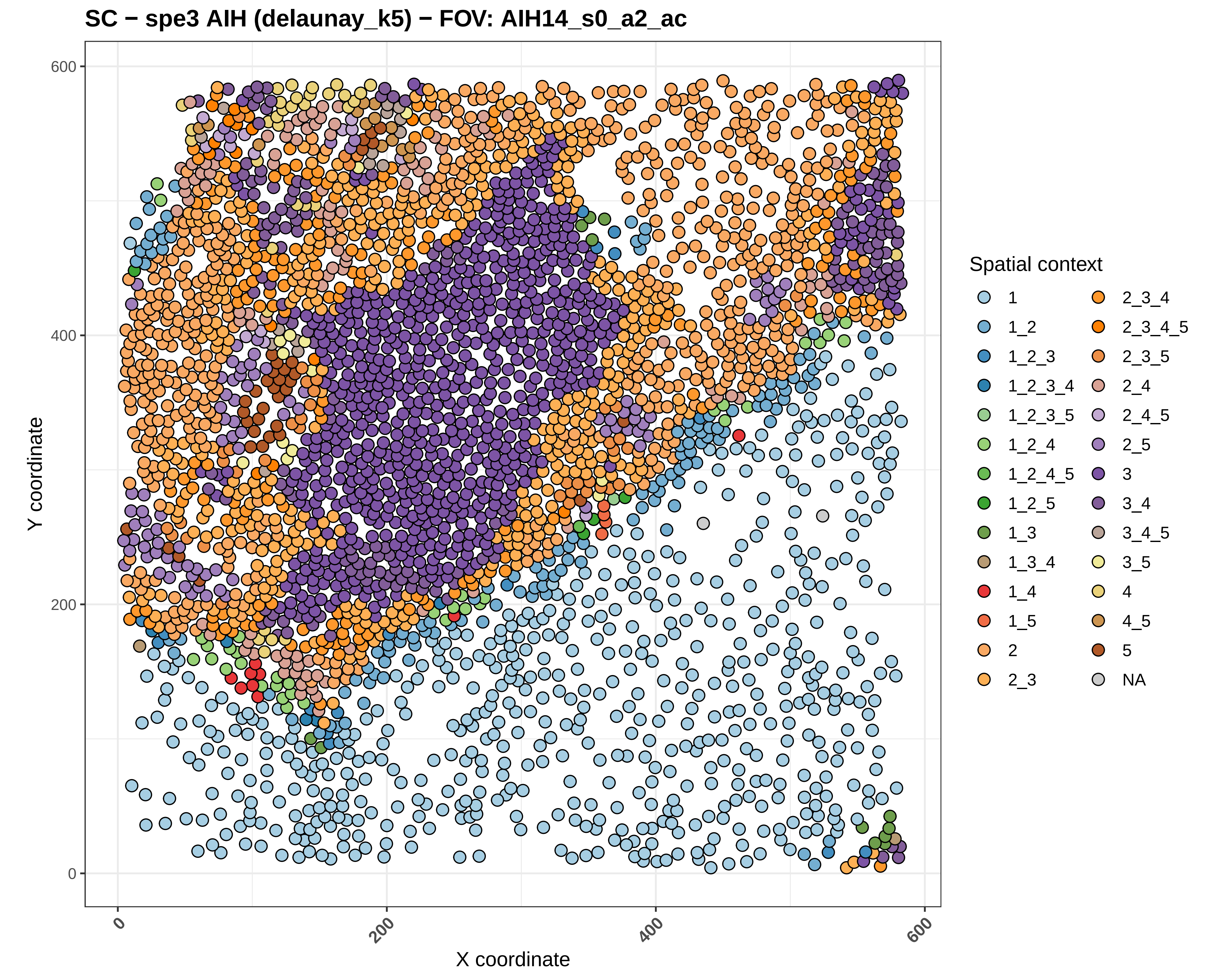

SC − spe3 AIH (delaunay_k5) − FOV: AIH14_s0_a2_ac
Y coordinate
X coordinate
0
200
400
600
0
200
400
600
Spatial context
1
1_2
1_2_3
1_2_3_4
1_2_3_5
1_2_4
1_2_4_5
1_2_5
1_3
1_3_4
1_4
1_5
2
2_3
2_3_4
2_3_4_5
2_3_5
2_4
2_4_5
2_5
3
3_4
3_4_5
3_5
4
4_5
5
NA

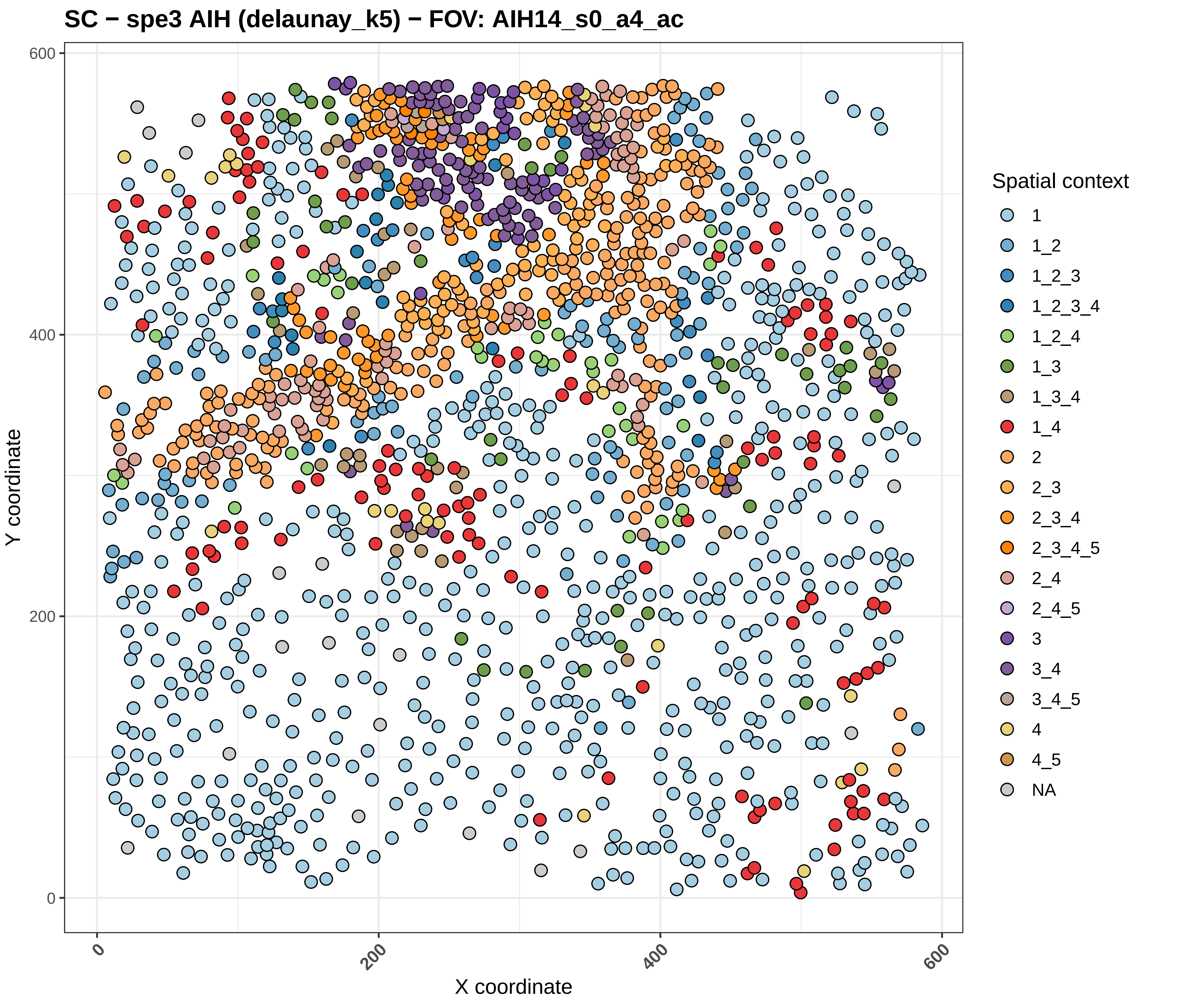

SC − spe3 AIH (delaunay_k5) − FOV: AIH14_s0_a4_ac
Y coordinate
X coordinate
0
200
400
600
Spatial context
1
1_2
1_2_3
1_2_3_4
1_2_4
1_3
1_3_4
1_4
2
2_3
2_3_4
2_3_4_5
2_4
2_4_5
3
3_4
3_4_5
4
4_5
NA

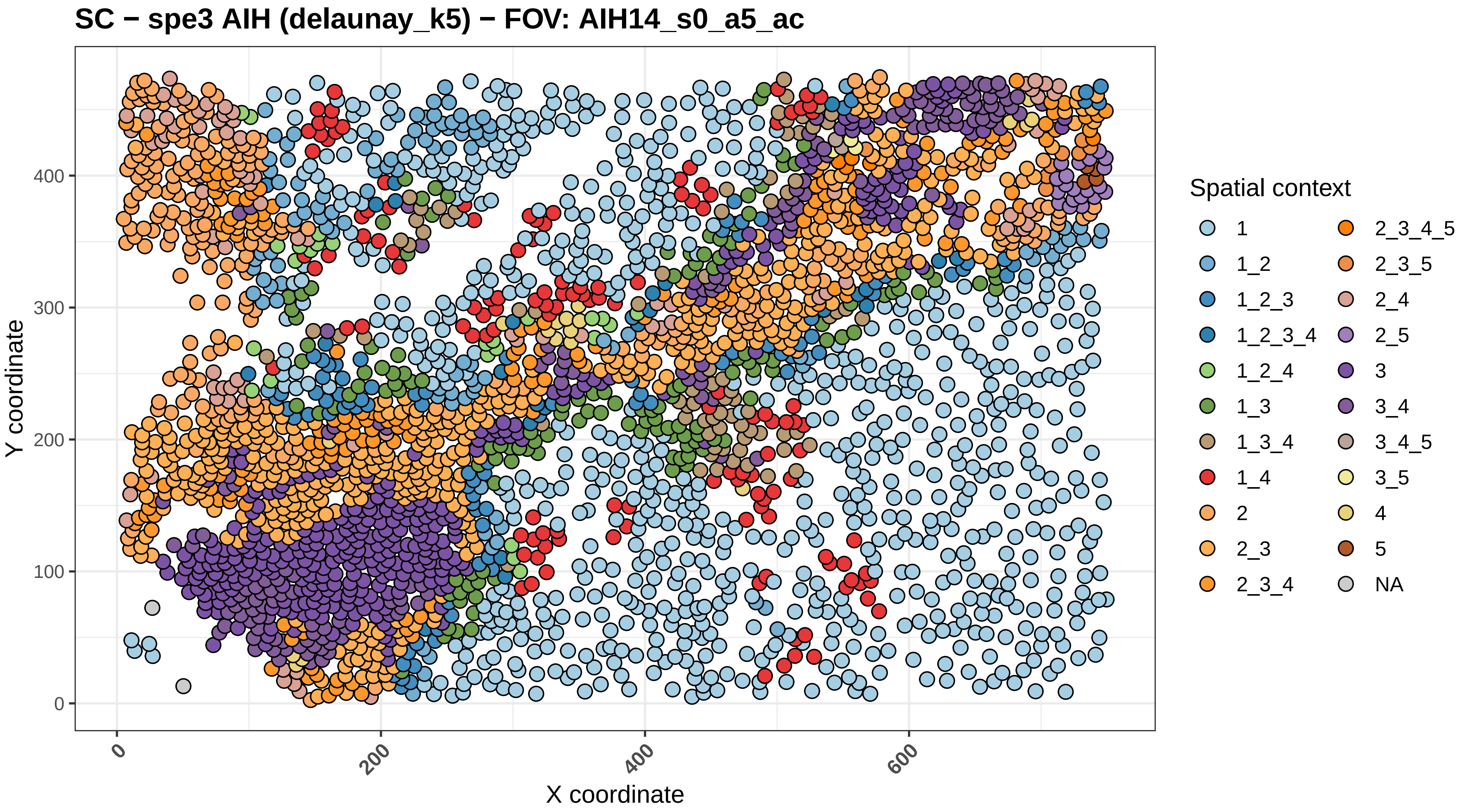
SC − spe3 AIH (delaunay_k5) − FOV: AIH14_s0_a5_ac
Y coordinate
X coordinate
0
100
200
300
400
0
200
400
600
Spatial context
1
1_2
1_2_3
1_2_3_4
1_2_4
1_3
1_3_4
1_4
2
2_3
2_3_4
2_3_4_5
2_3_5
2_4
2_5
3
3_4
3_4_5
3_5
4
5
NA

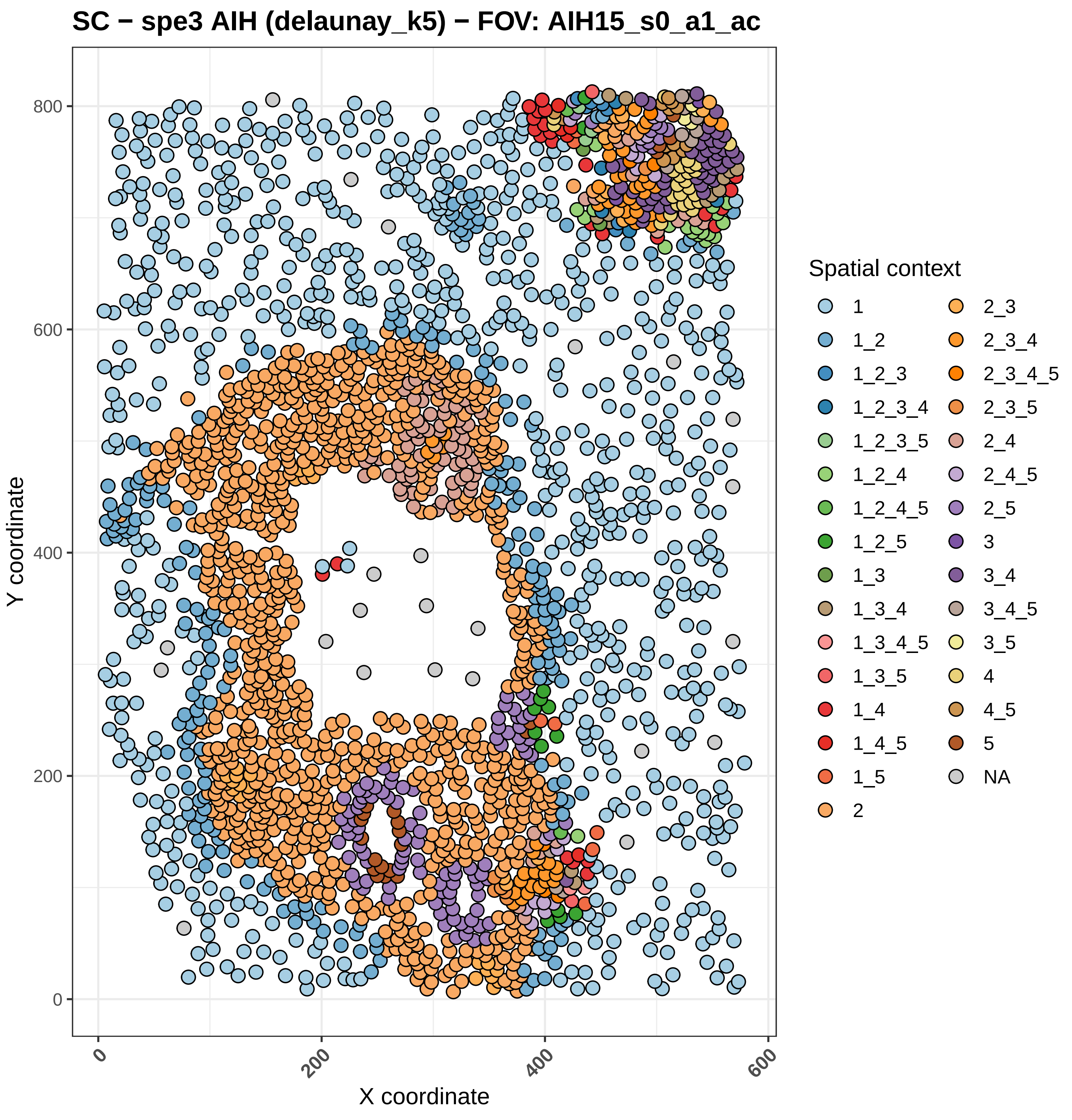
SC − spe3 AIH (delaunay_k5) − FOV: AIH15_s0_a1_ac
Y coordinate
X coordinate
0
200
400
600
800
0
200
400
600
Spatial context
1
1_2
1_2_3
1_2_3_4
1_2_3_5
1_2_4
1_2_4_5
1_2_5
1_3
1_3_4
1_3_4_5
1_3_5
1_4
1_4_5
1_5
2
2_3
2_3_4
2_3_4_5
2_3_5
2_4
2_4_5
2_5
3
3_4
3_4_5
3_5
4
4_5
5
NA

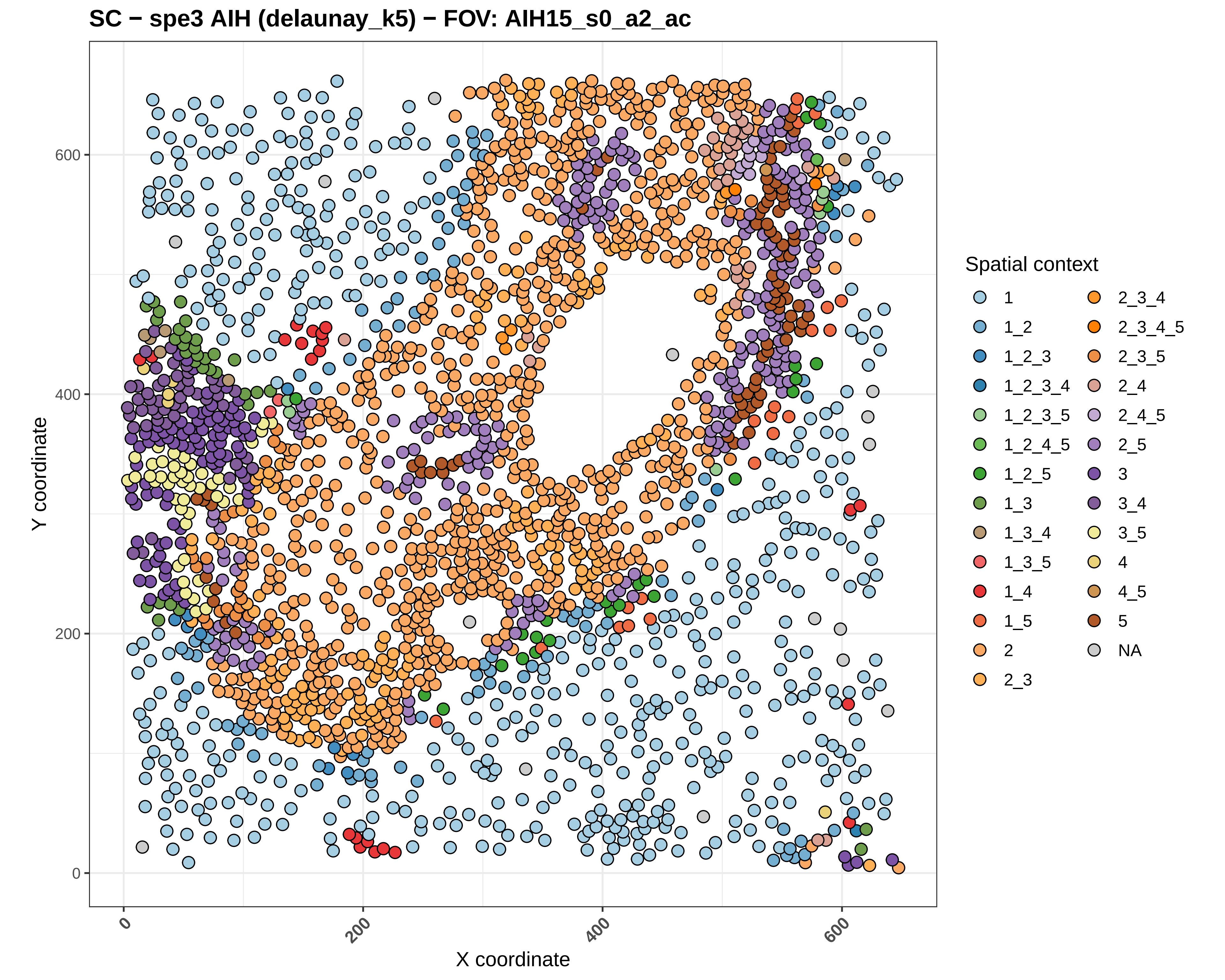
SC − spe3 AIH (delaunay_k5) − FOV: AIH15_s0_a2_ac
Y coordinate
X coordinate
0
200
400
600
Spatial context
1
1_2
1_2_3
1_2_3_4
1_2_3_5
1_2_4_5
1_2_5
1_3
1_3_4
1_3_5
1_4
1_5
2
2_3
2_3_4
2_3_4_5
2_3_5
2_4
2_4_5
2_5
3
3_4
3_5
4
4_5
5
NA

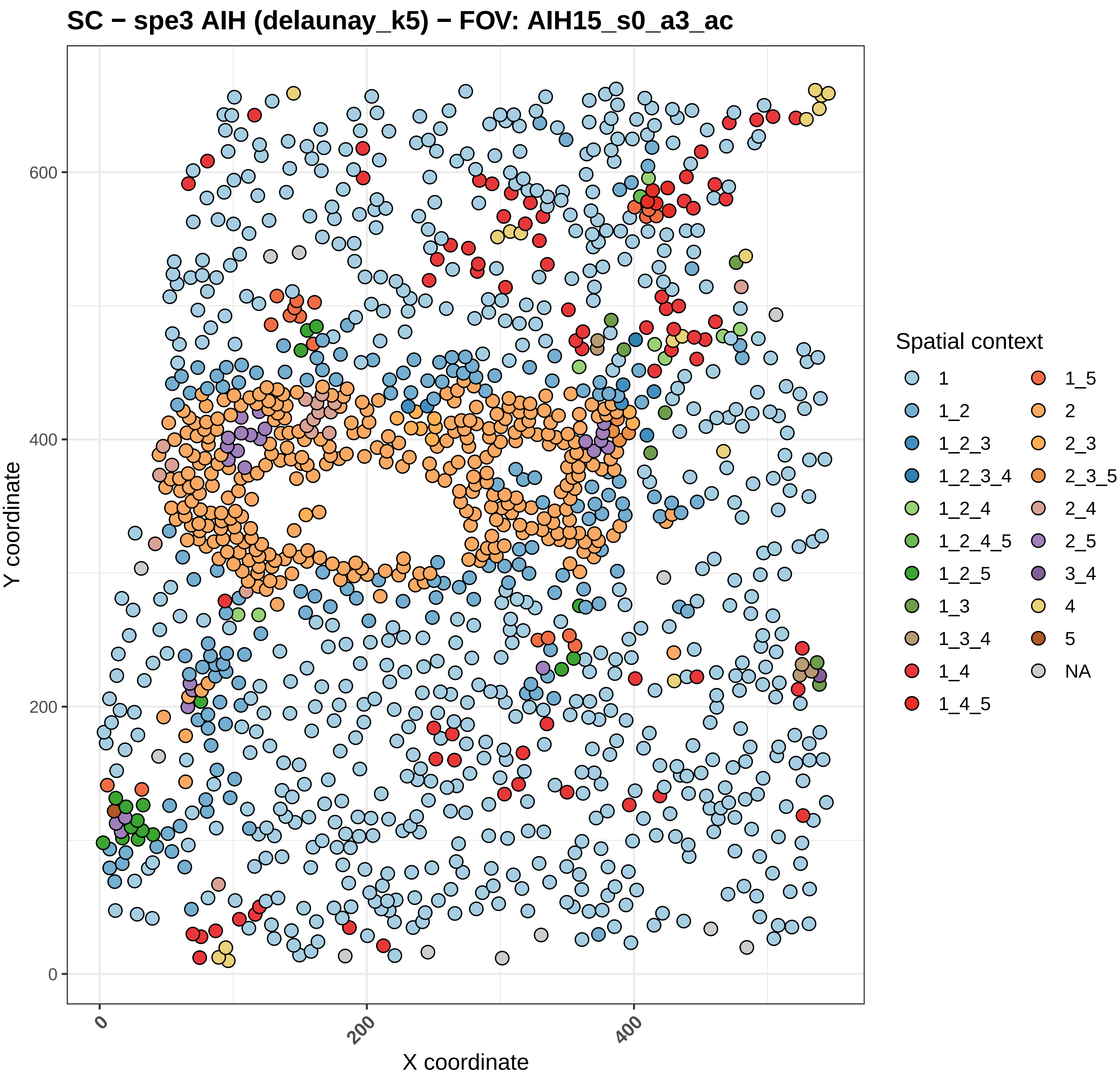
SC − spe3 AIH (delaunay_k5) − FOV: AIH15_s0_a3_ac
Y coordinate
X coordinate
0
200
400
600
Spatial context
1
1_2
1_2_3
1_2_3_4
1_2_4
1_2_4_5
1_2_5
1_3
1_3_4
1_4
1_4_5
1_5
2
2_3
2_3_5
2_4
2_5
3_4
4
5
NA

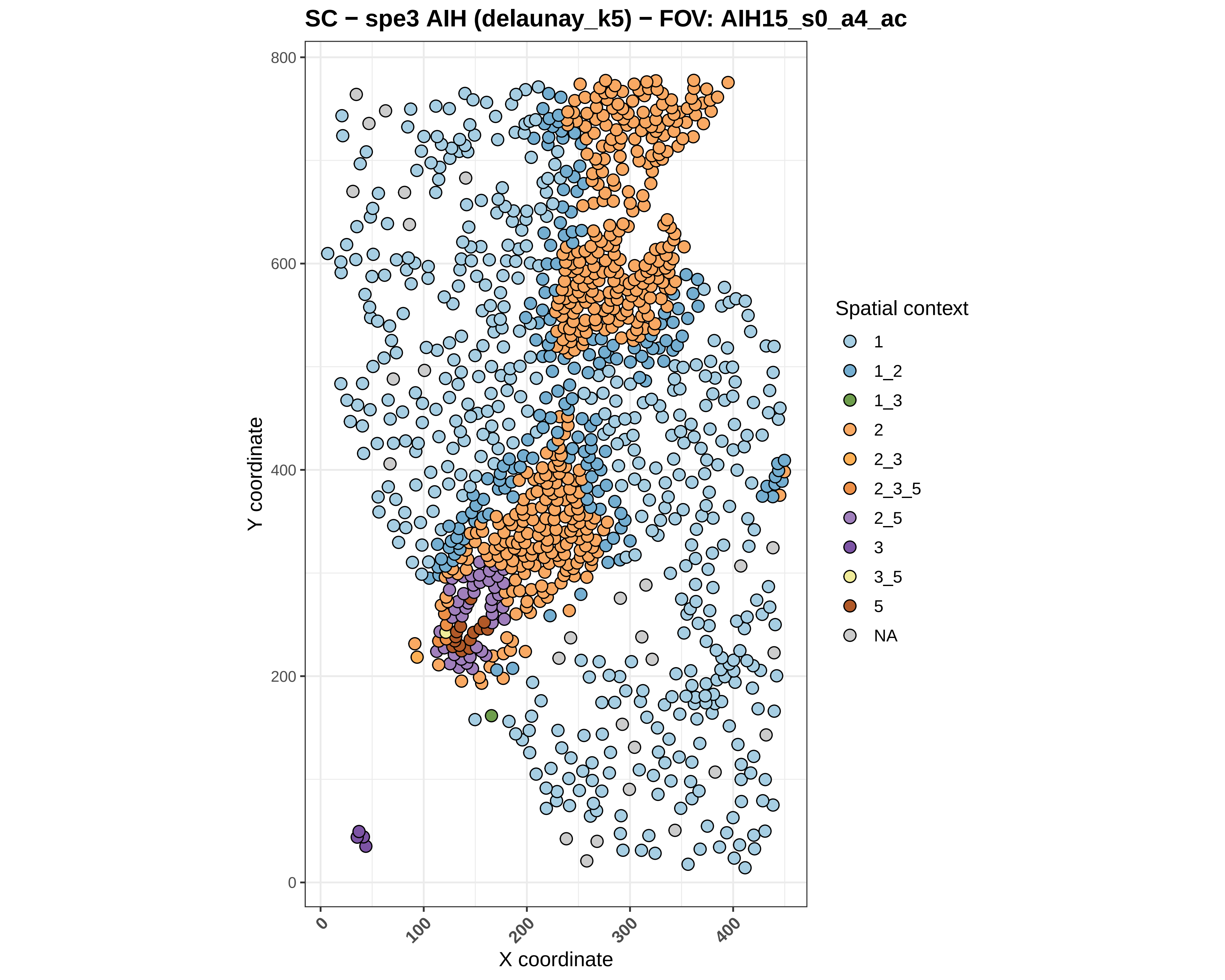
SC − spe3 AIH (delaunay_k5) − FOV: AIH15_s0_a4_ac
Y coordinate
X coordinate
0
200
400
600
800
0
100
200
300
400
Spatial context
1
1_2
1_3
2
2_3
2_3_5
2_5
3
3_5
5
NA

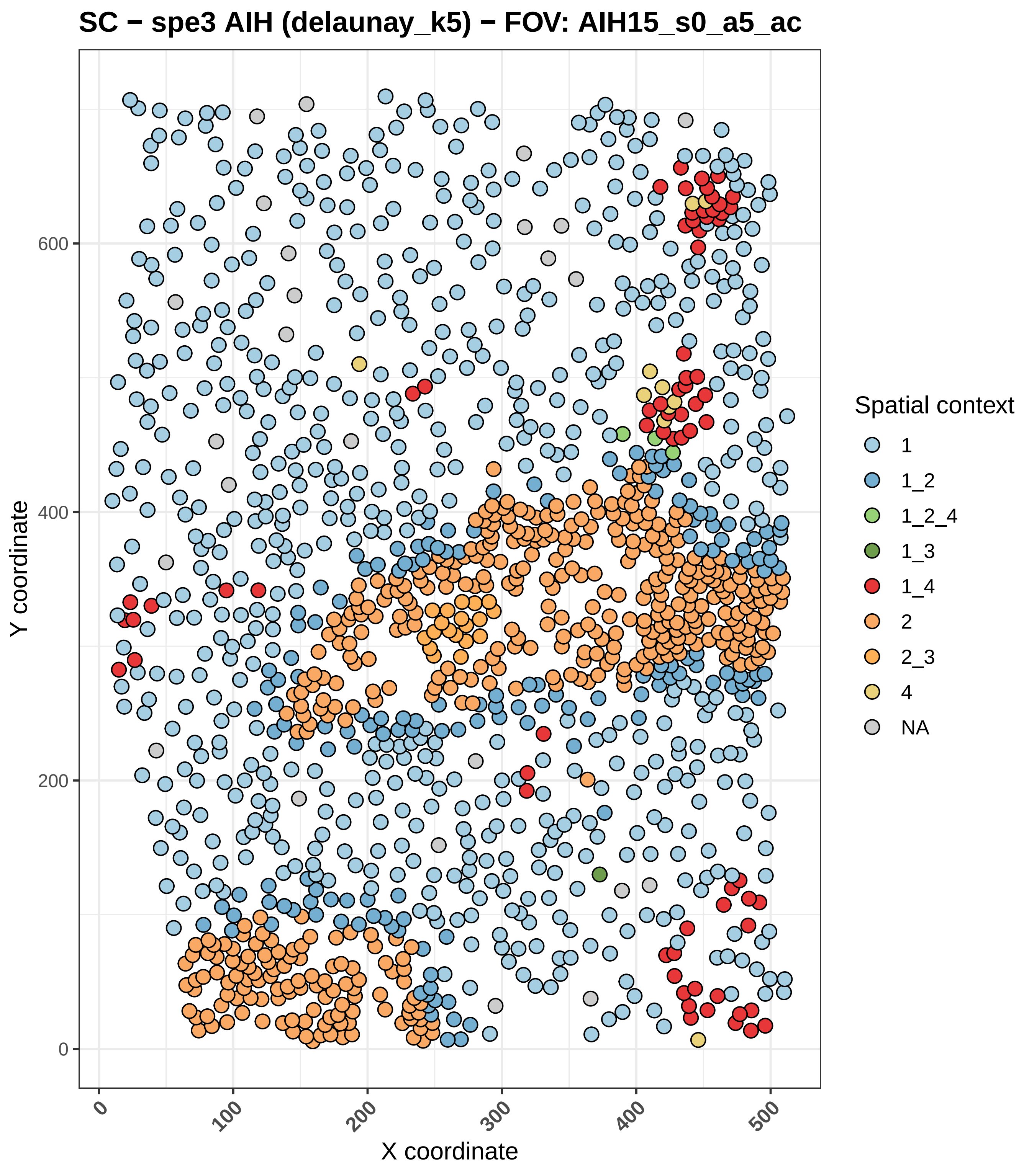
SC − spe3 AIH (delaunay_k5) − FOV: AIH15_s0_a5_ac
Y coordinate
X coordinate
0
100
200
300
400
500
600
Spatial context
1
1_2
1_2_4
1_3
1_4
2
2_3
4
NA

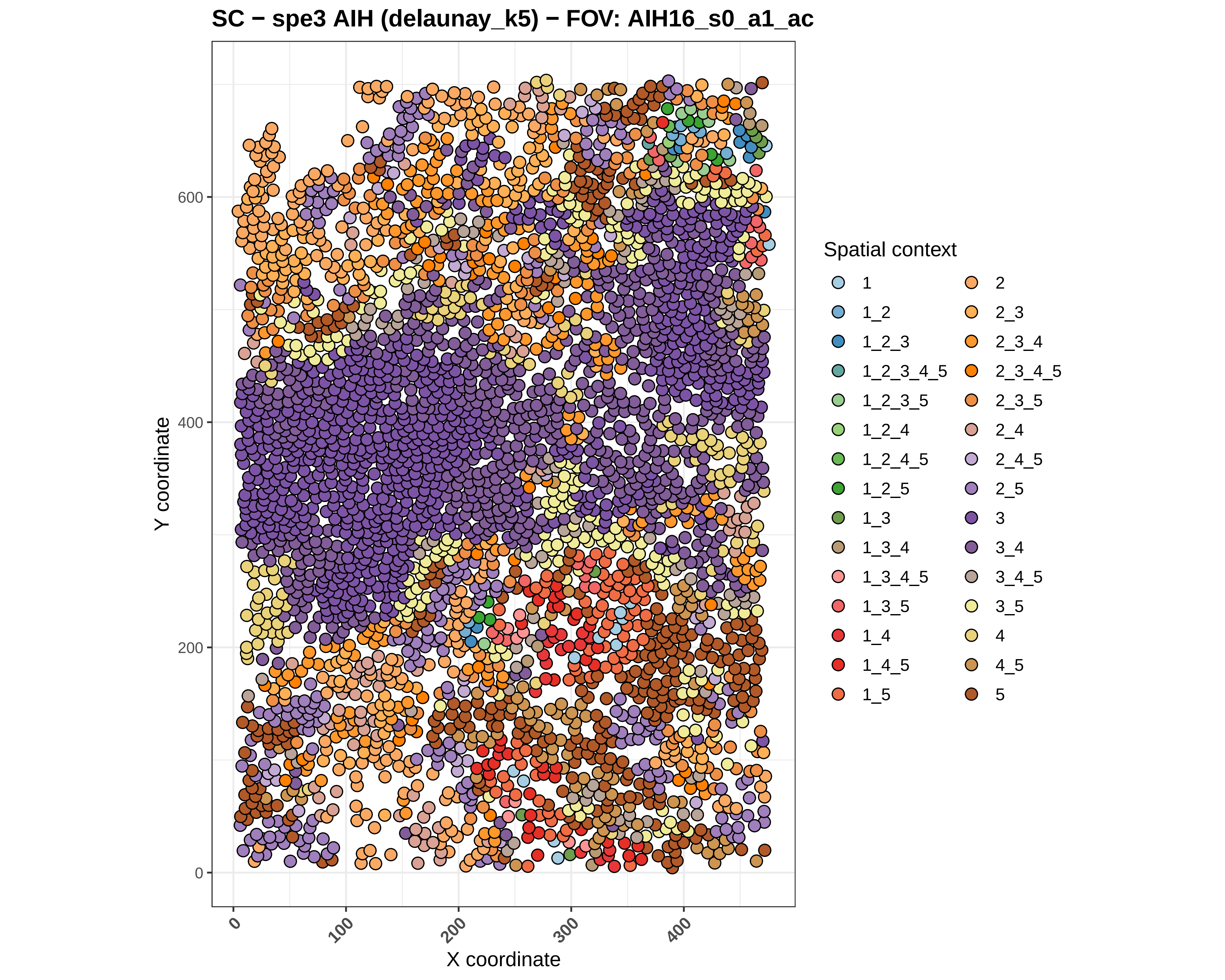
SC − spe3 AIH (delaunay_k5) − FOV: AIH16_s0_a1_ac
Y coordinate
0
200
400
600
X coordinate
0
100
200
300
400
Spatial context
1
1_2
1_2_3
1_2_3_4_5
1_2_3_5
1_2_4
1_2_4_5
1_2_5
1_3
1_3_4
1_3_4_5
1_3_5
1_4
1_4_5
1_5
2
2_3
2_3_4
2_3_4_5
2_3_5
2_4
2_4_5
2_5
3
3_4
3_4_5
3_5
4
4_5
5

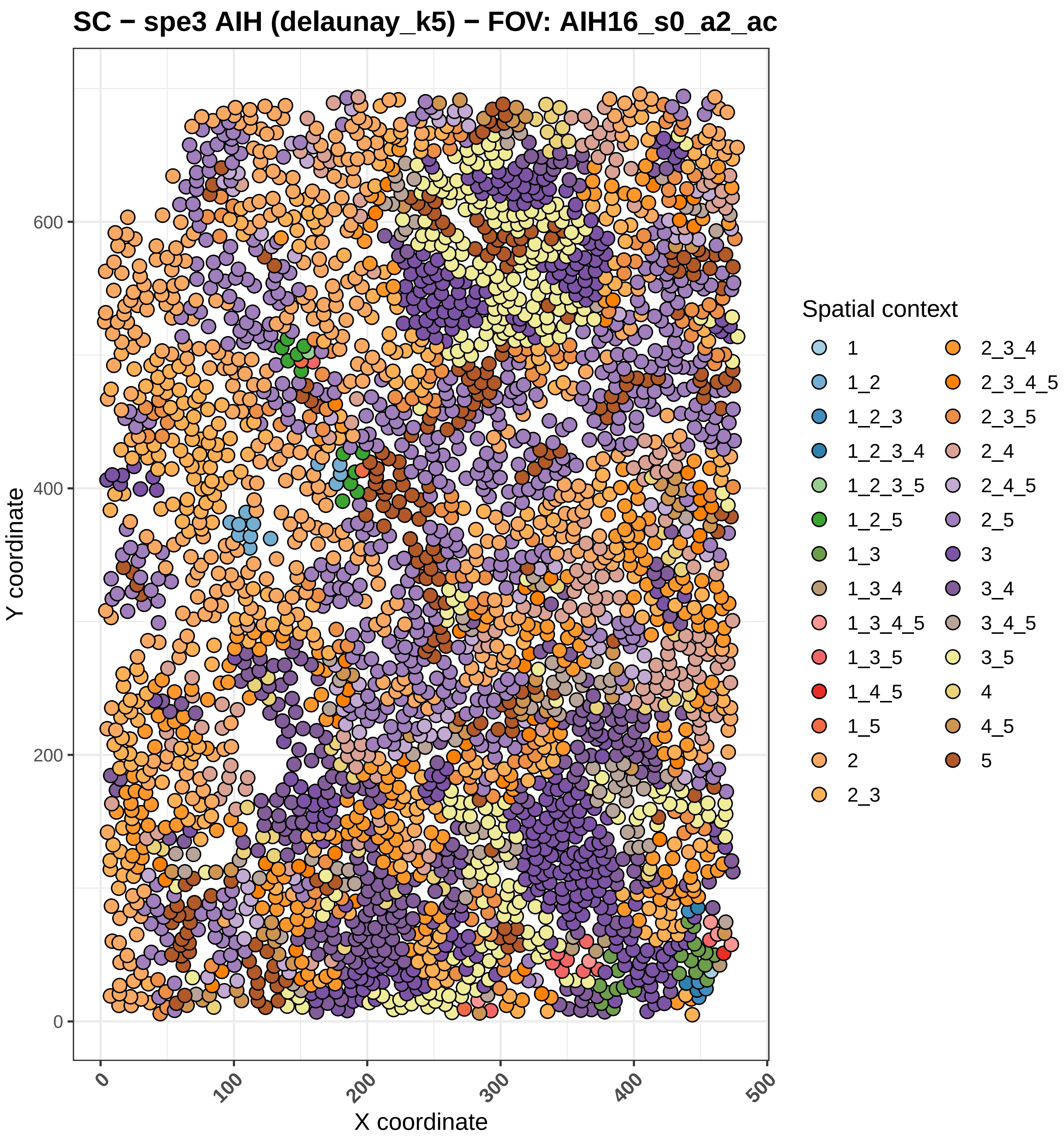
SC − spe3 AIH (delaunay_k5) − FOV: AIH16_s0_a2_ac
Y coordinate
X coordinate
0
200
400
600
0
100
200
300
400
500
Spatial context
1
1_2
1_2_3
1_2_3_4
1_2_3_5
1_2_5
1_3
1_3_4
1_3_4_5
1_3_5
1_4_5
1_5
2
2_3
2_3_4
2_3_4_5
2_3_5
2_4
2_4_5
2_5
3
3_4
3_4_5
3_5
4
4_5
5

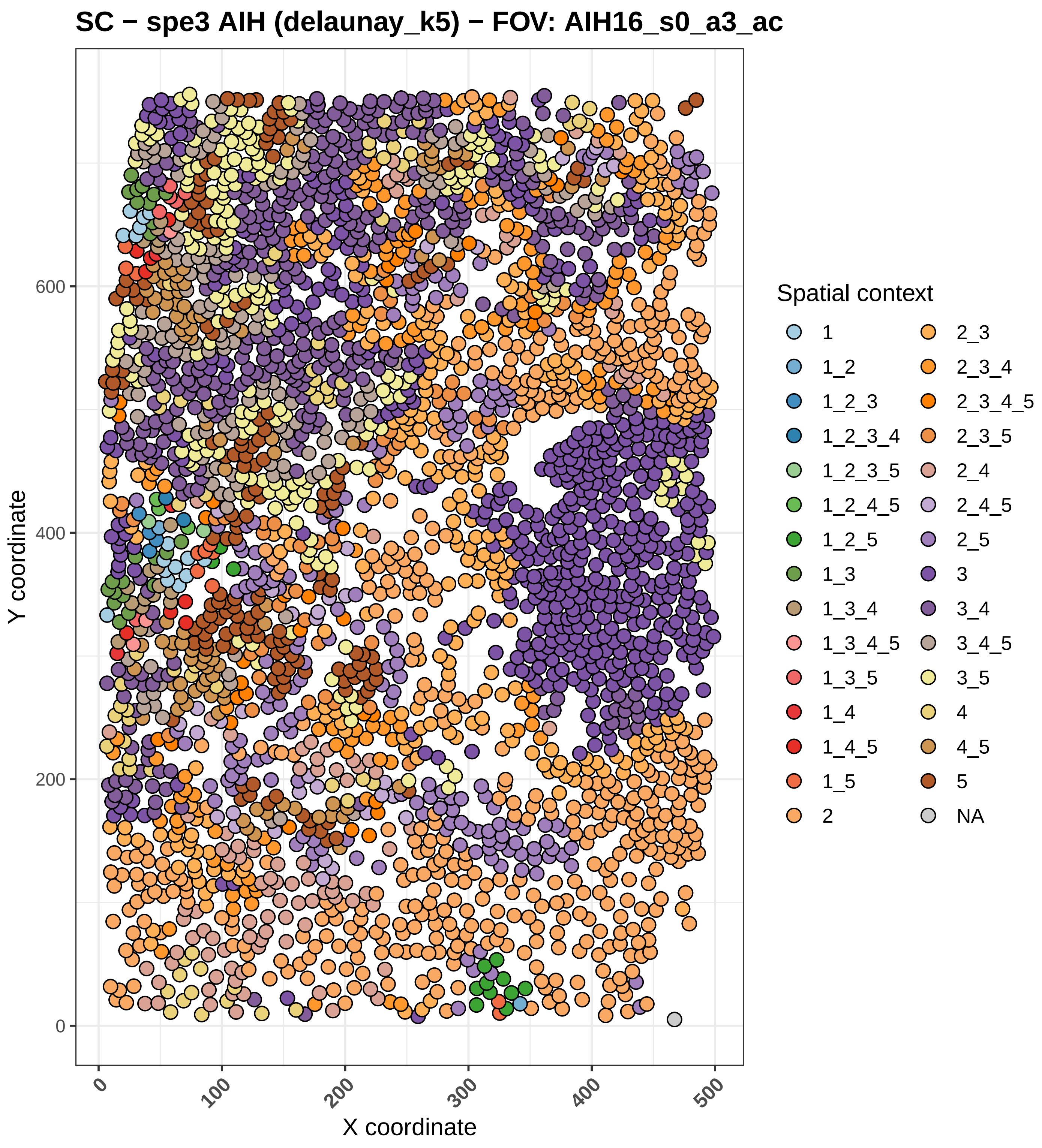

SC − spe3 AIH (delaunay_k5) − FOV: AIH16_s0_a3_ac
Y coordinate
X coordinate
0
200
400
600
0
100
200
300
400
500
Spatial context
1
1_2
1_2_3
1_2_3_4
1_2_3_5
1_2_4_5
1_2_5
1_3
1_3_4
1_3_4_5
1_3_5
1_4
1_4_5
1_5
2
2_3
2_3_4
2_3_4_5
2_3_5
2_4
2_4_5
2_5
3
3_4
3_4_5
3_5
4
4_5
5
NA

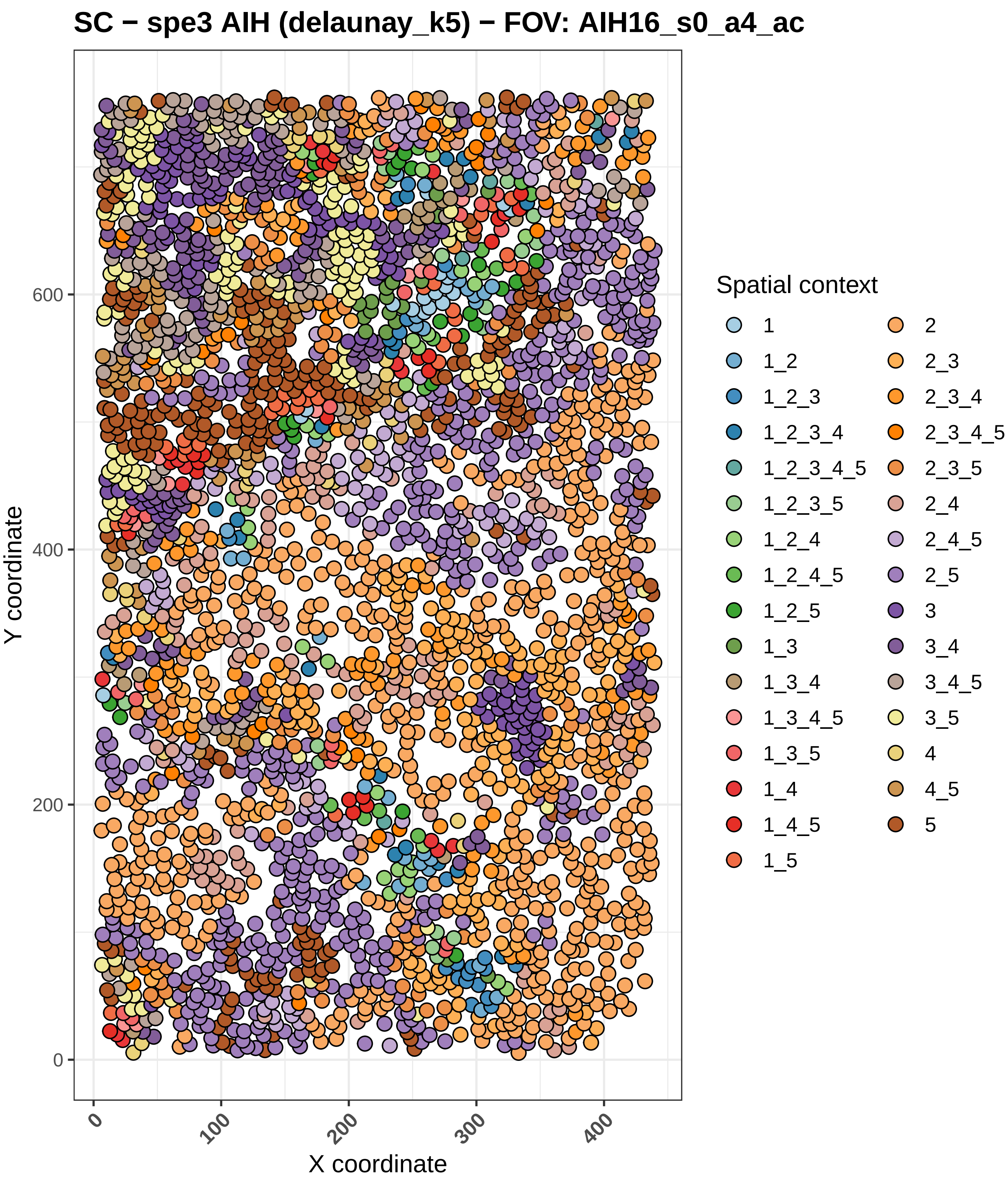

SC − spe3 AIH (delaunay_k5) − FOV: AIH16_s0_a4_ac
Y coordinate
X coordinate
0
200
400
600
100
300
Spatial context
1
1_2
1_2_3
1_2_3_4
1_2_3_4_5
1_2_3_5
1_2_4
1_2_4_5
1_2_5
1_3
1_3_4
1_3_4_5
1_3_5
1_4
1_4_5
1_5
2
2_3
2_3_4
2_3_4_5
2_3_5
2_4
2_4_5
2_5
3
3_4
3_4_5
3_5
4
4_5
5

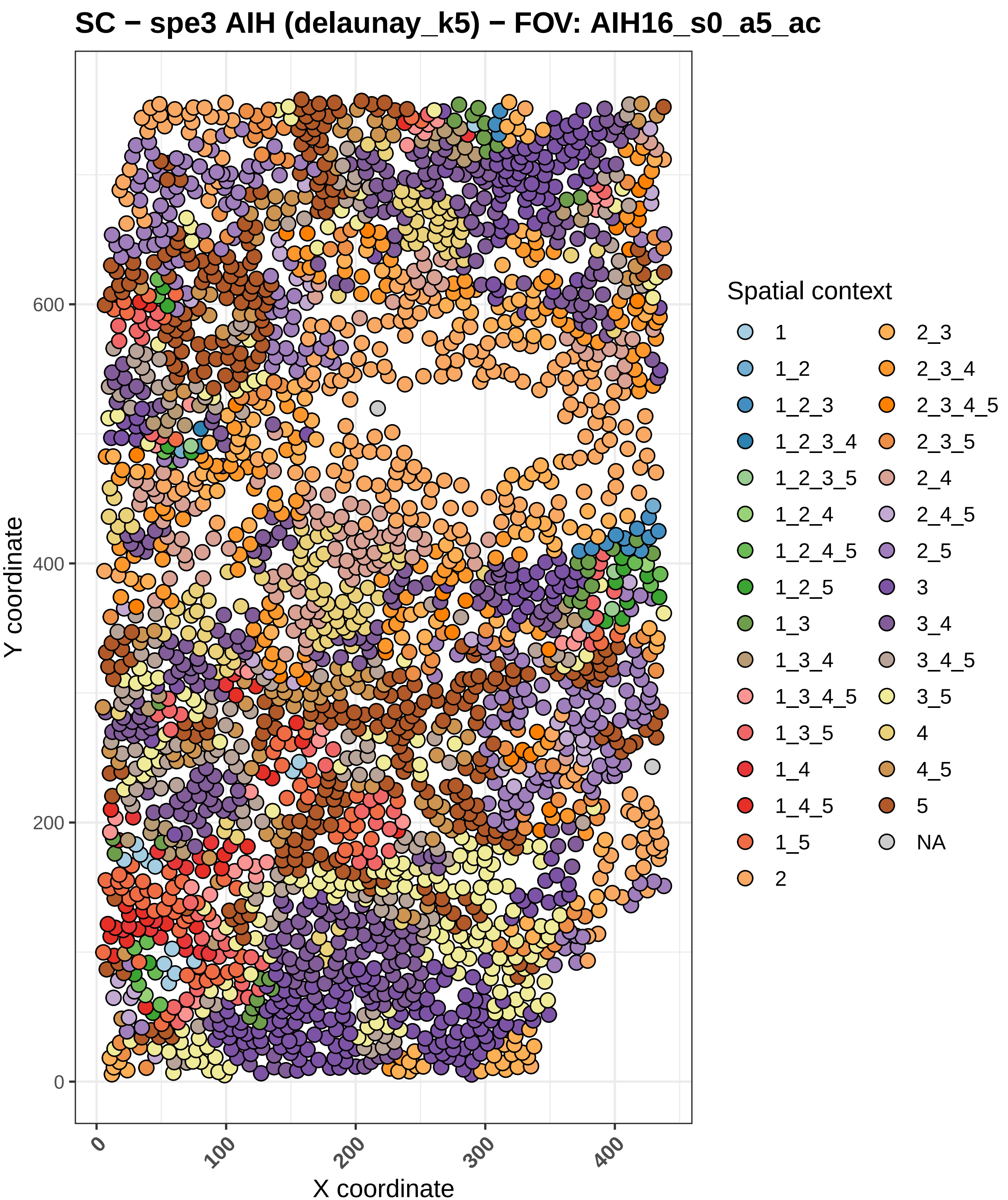

SC − spe3 AIH (delaunay_k5) − FOV: AIH16_s0_a5_ac
Y coordinate
X coordinate
0
200
400
600
0
100
200
300
400
Spatial context
1
1_2
1_2_3
1_2_3_4
1_2_3_5
1_2_4
1_2_4_5
1_2_5
1_3
1_3_4
1_3_4_5
1_3_5
1_4
1_4_5
1_5
2
2_3
2_3_4
2_3_4_5
2_3_5
2_4
2_4_5
2_5
3
3_4
3_4_5
3_5
4
4_5
5
NA

#### Region map − spe3 ICI (delaunay_k5) − FOV: ICI01_s0_a1_ac

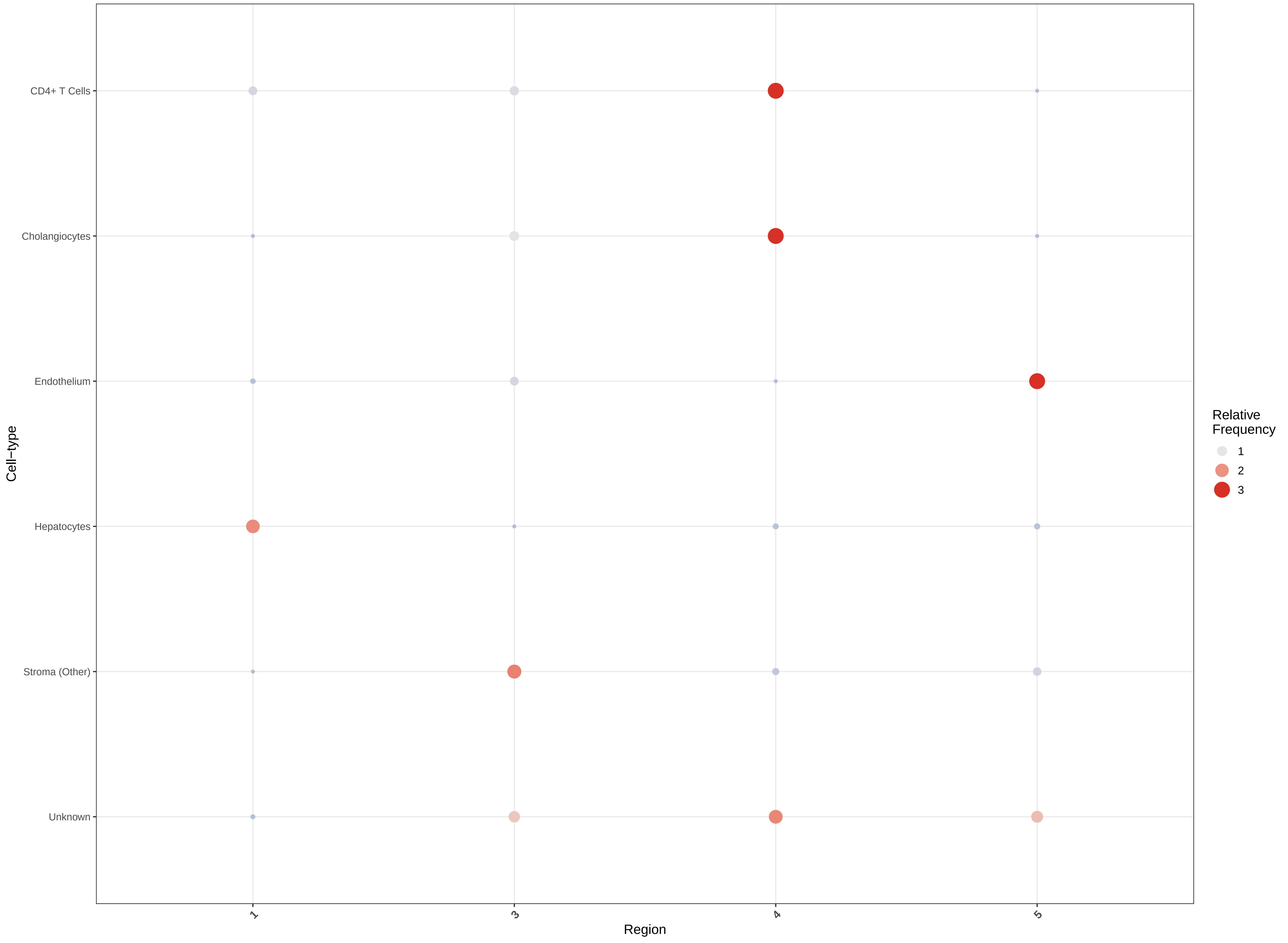

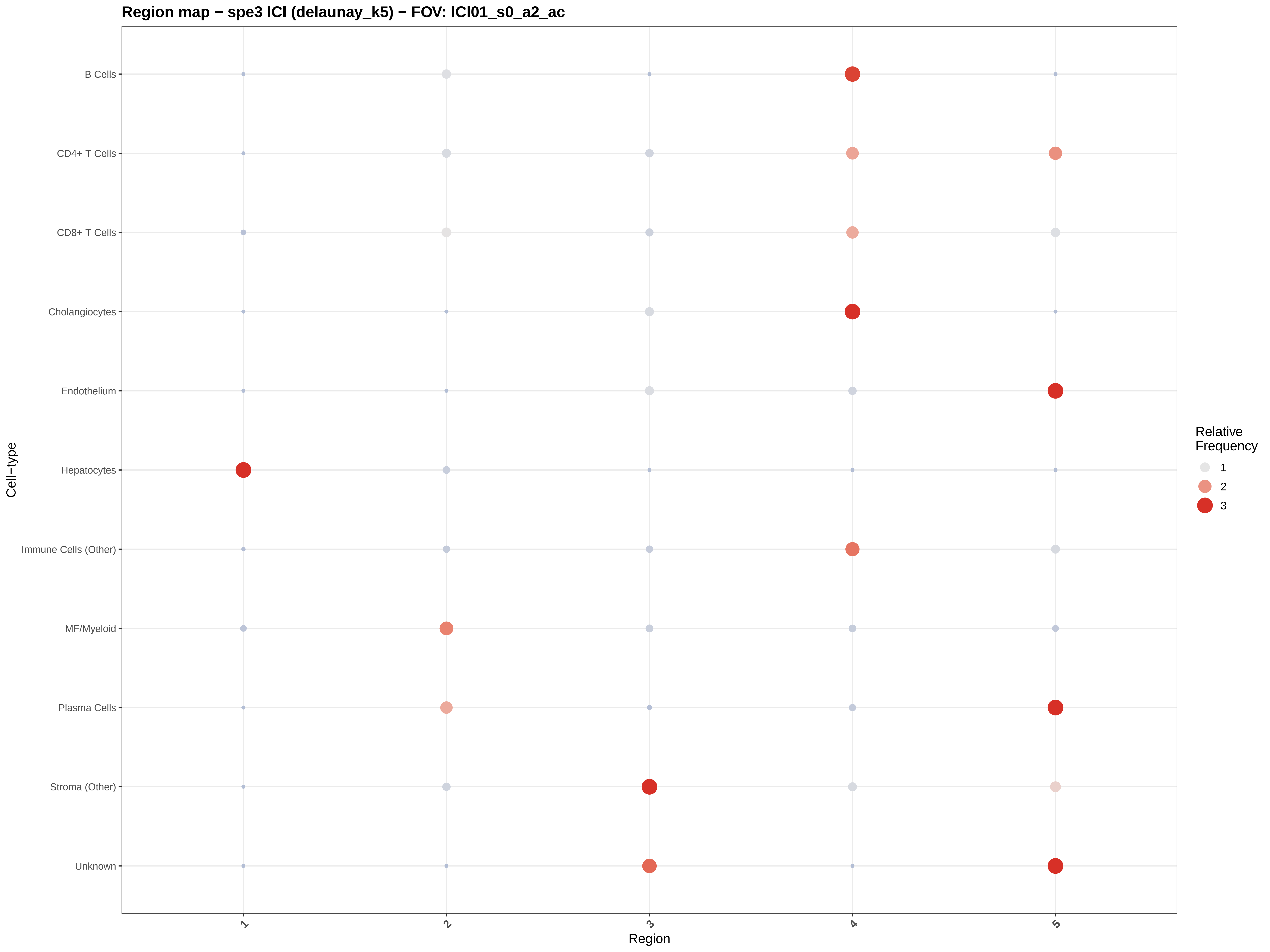

Region map − spe3 ICI (delaunay_k5) − FOV: ICI01_s0_a2_ac
B Cells
CD4+ T Cells
CD8+ T Cells
Cholangiocytes
Endothelium
Hepatocytes
Immune Cells (Other)
MF/Myeloid
Plasma Cells
Stroma (Other)
Unknown
Cell−type
1
2
3
4
5
Region
Relative Frequency
1
2
3

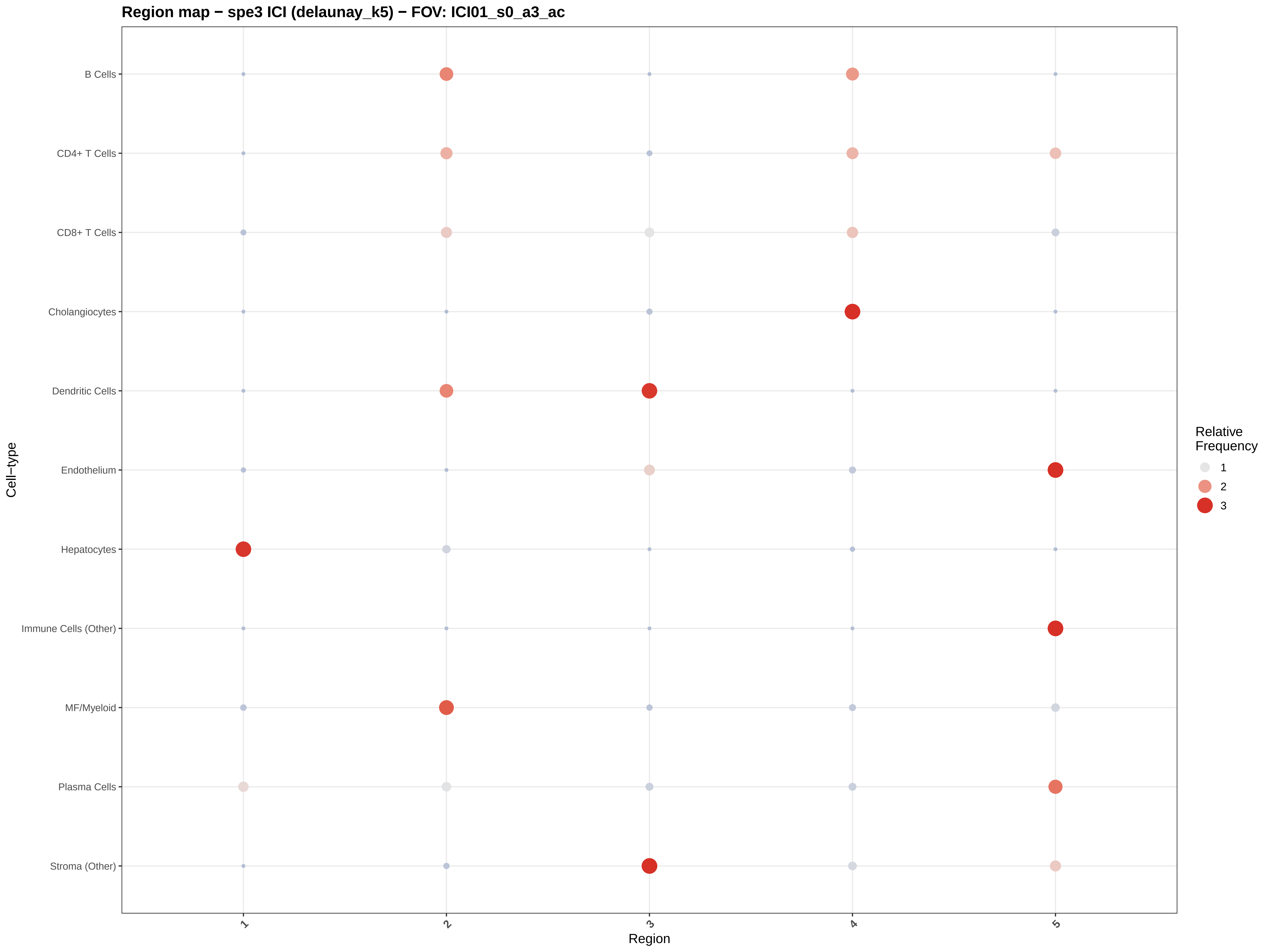

Region map − spe3 ICI (delaunay_k5) − FOV: ICI01_s0_a3_ac
B Cells
CD4+ T Cells
CD8+ T Cells
Cholangiocytes
Dendritic Cells
Endothelium
Hepatocytes
Immune Cells (Other)
MF/Myeloid
Plasma Cells
Stroma (Other)
Cell−type
1
2
3
4
5
Region
Relative Frequency
1
2
3

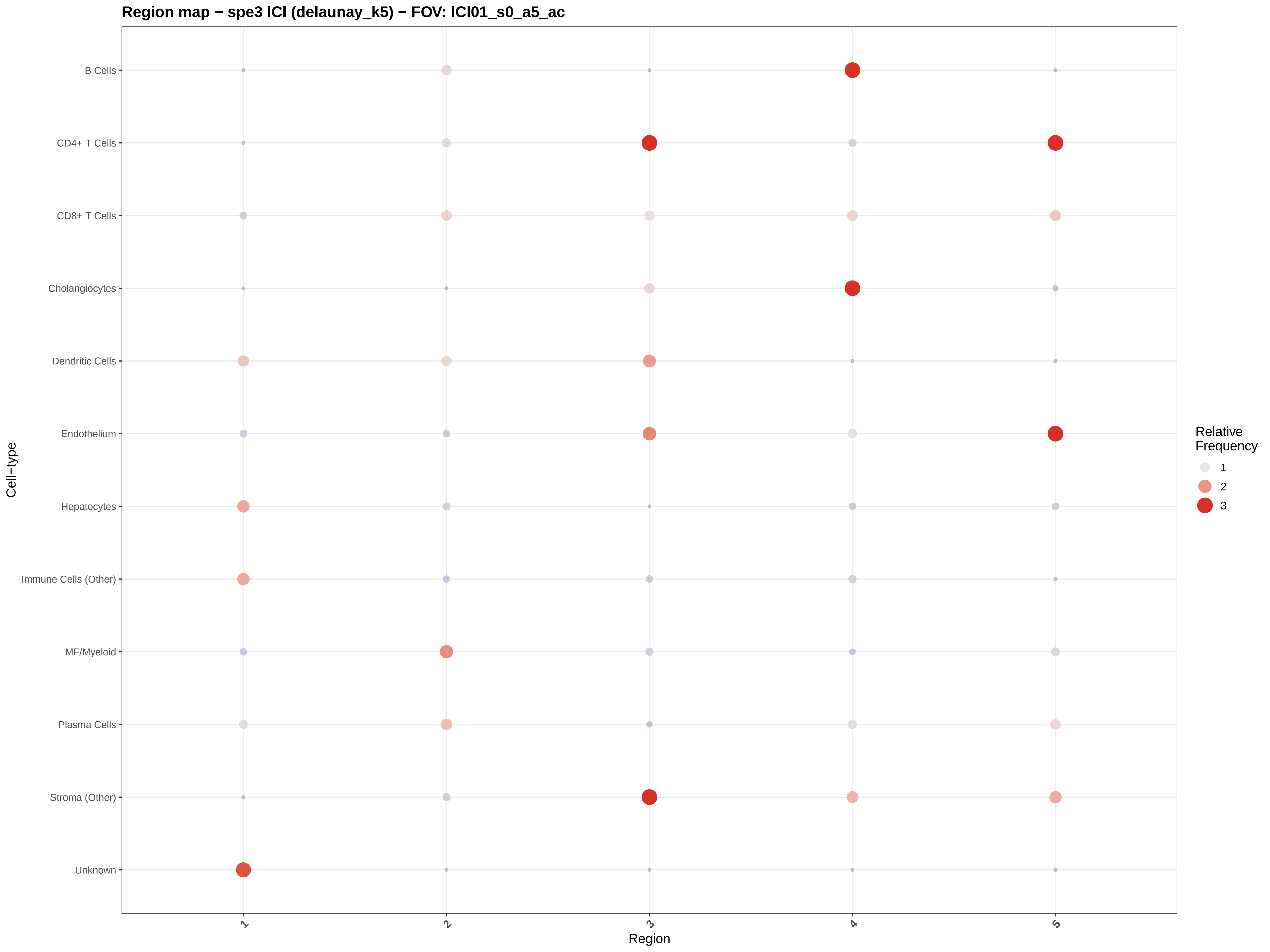

Region map − spe3 ICI (delaunay_k5) − FOV: ICI01_s0_a5_ac
B Cells
CD4+ T Cells
CD8+ T Cells
Cholangiocytes
Dendritic Cells
Endothelium
Hepatocytes
Immune Cells (Other)
MF/Myeloid
Plasma Cells
Stroma (Other)
Unknown
Cell−type
1
2
3
4
5
Region
Relative Frequency
1
2
3

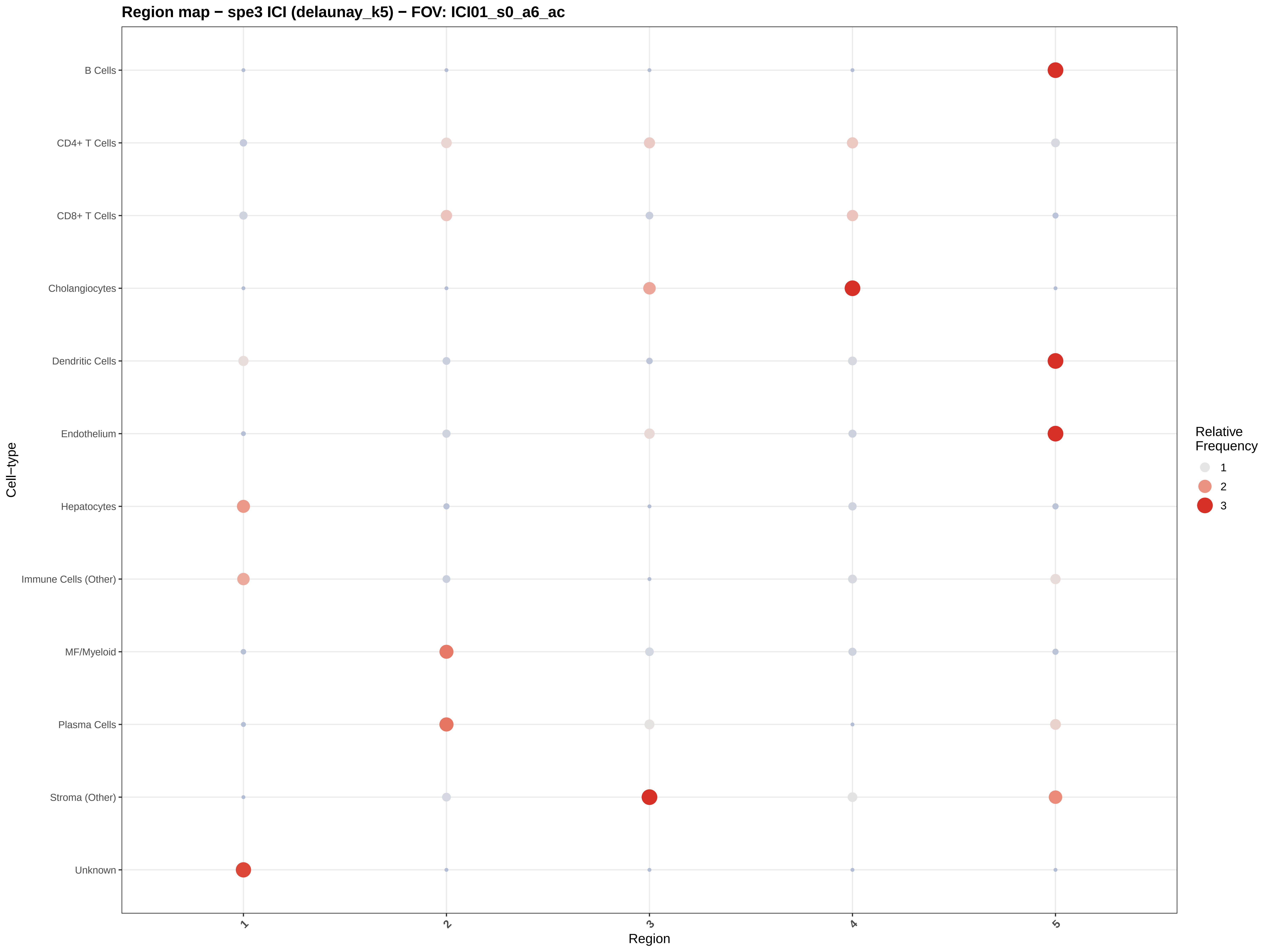
Region map − spe3 ICI (delaunay_k5) − FOV: ICI01_s0_a6_ac
B Cells
CD4+ T Cells
CD8+ T Cells
Cholangiocytes
Dendritic Cells
Endothelium
Hepatocytes
Immune Cells (Other)
MF/Myeloid
Plasma Cells
Stroma (Other)
Unknown
Cell−type
1
2
3
4
5
Region
Relative Frequency
1
2
3

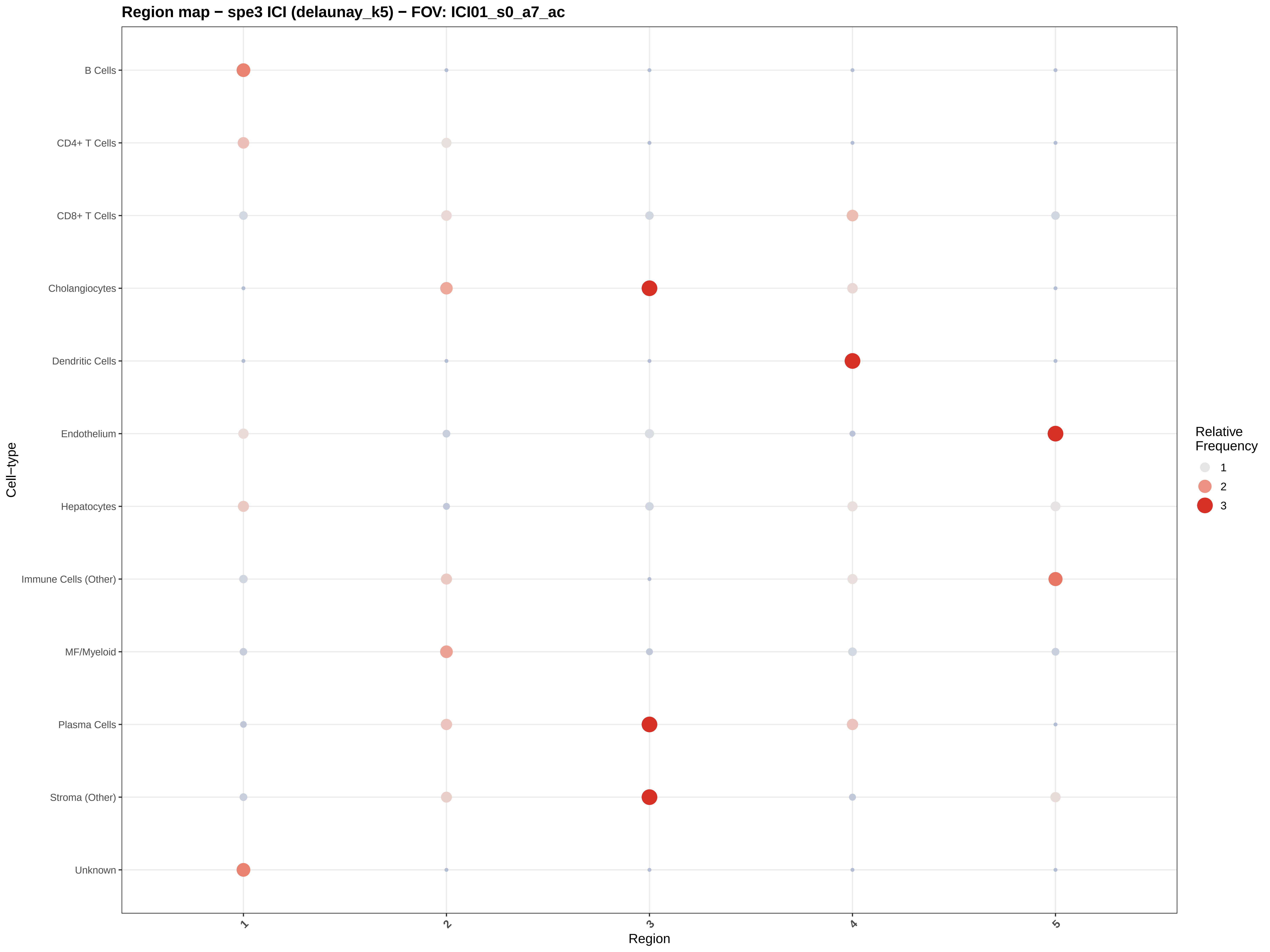

Region map − spe3 ICI (delaunay_k5) − FOV: ICI01_s0_a7_ac
B Cells
CD4+ T Cells
CD8+ T Cells
Cholangiocytes
Dendritic Cells
Endothelium
Hepatocytes
Immune Cells (Other)
MF/Myeloid
Plasma Cells
Stroma (Other)
Unknown
Cell−type
1
2
3
4
5
Region
Relative Frequency
1
2
3

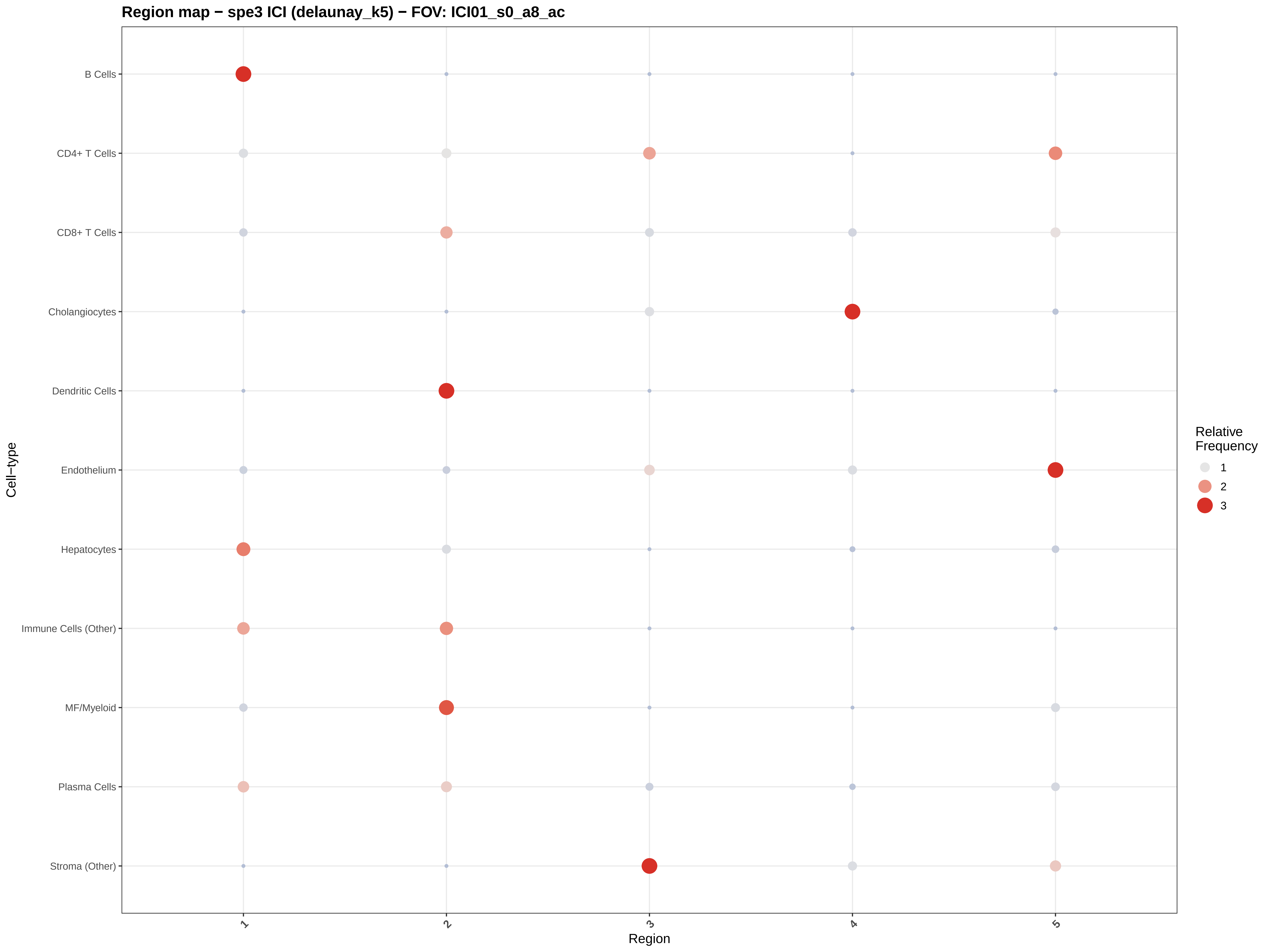

Region map − spe3 ICI (delaunay_k5) − FOV: ICI01_s0_a8_ac
B Cells
CD4+ T Cells
CD8+ T Cells
Cholangiocytes
Dendritic Cells
Endothelium
Hepatocytes
Immune Cells (Other)
MF/Myeloid
Plasma Cells
Stroma (Other)
Cell−type
1
2
3
4
5
Region
Relative Frequency
1
2
3

#### Region map − spe3 ICI (delaunay_k5) − FOV: ICI01_s0_a9_ac

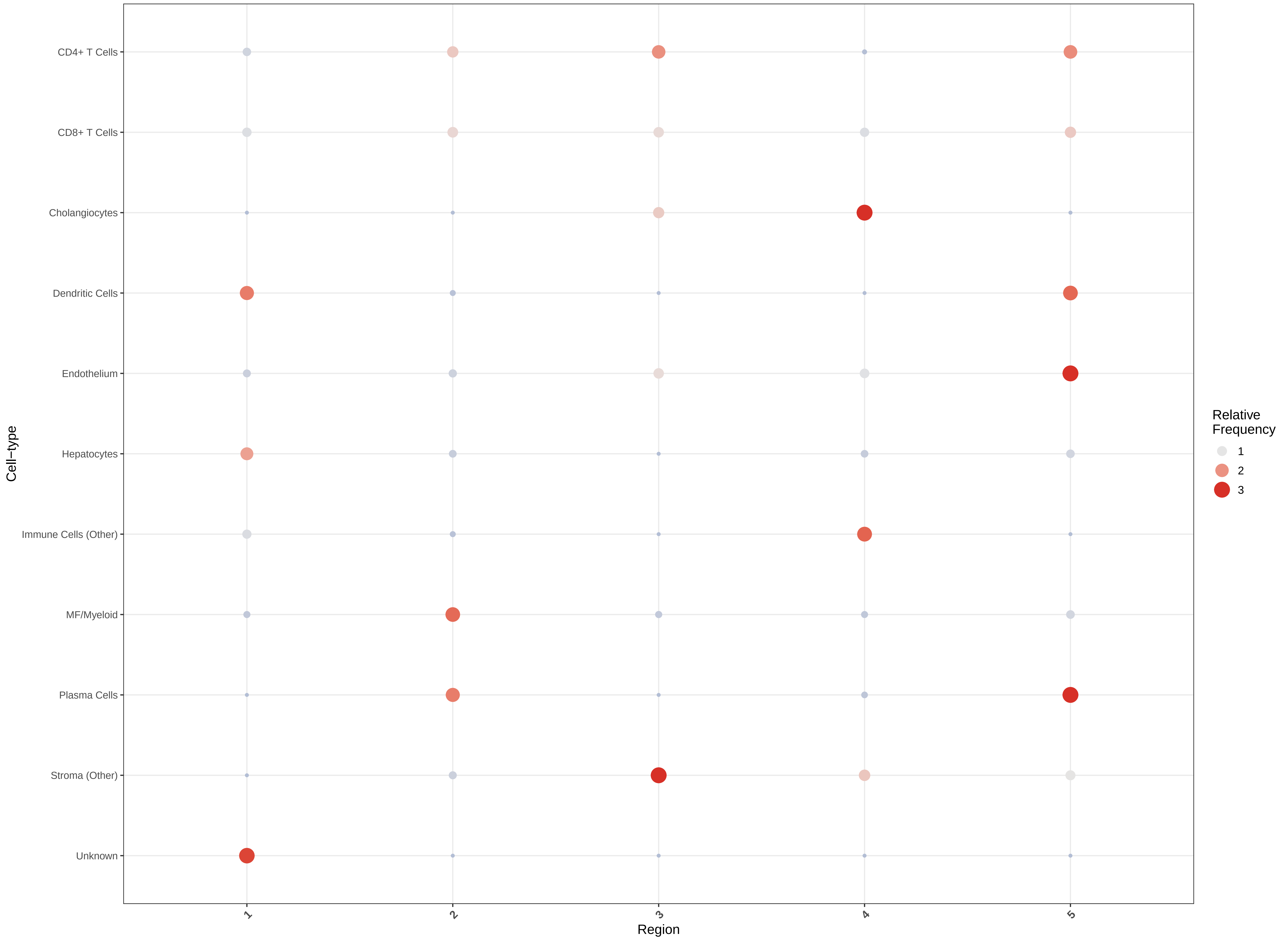

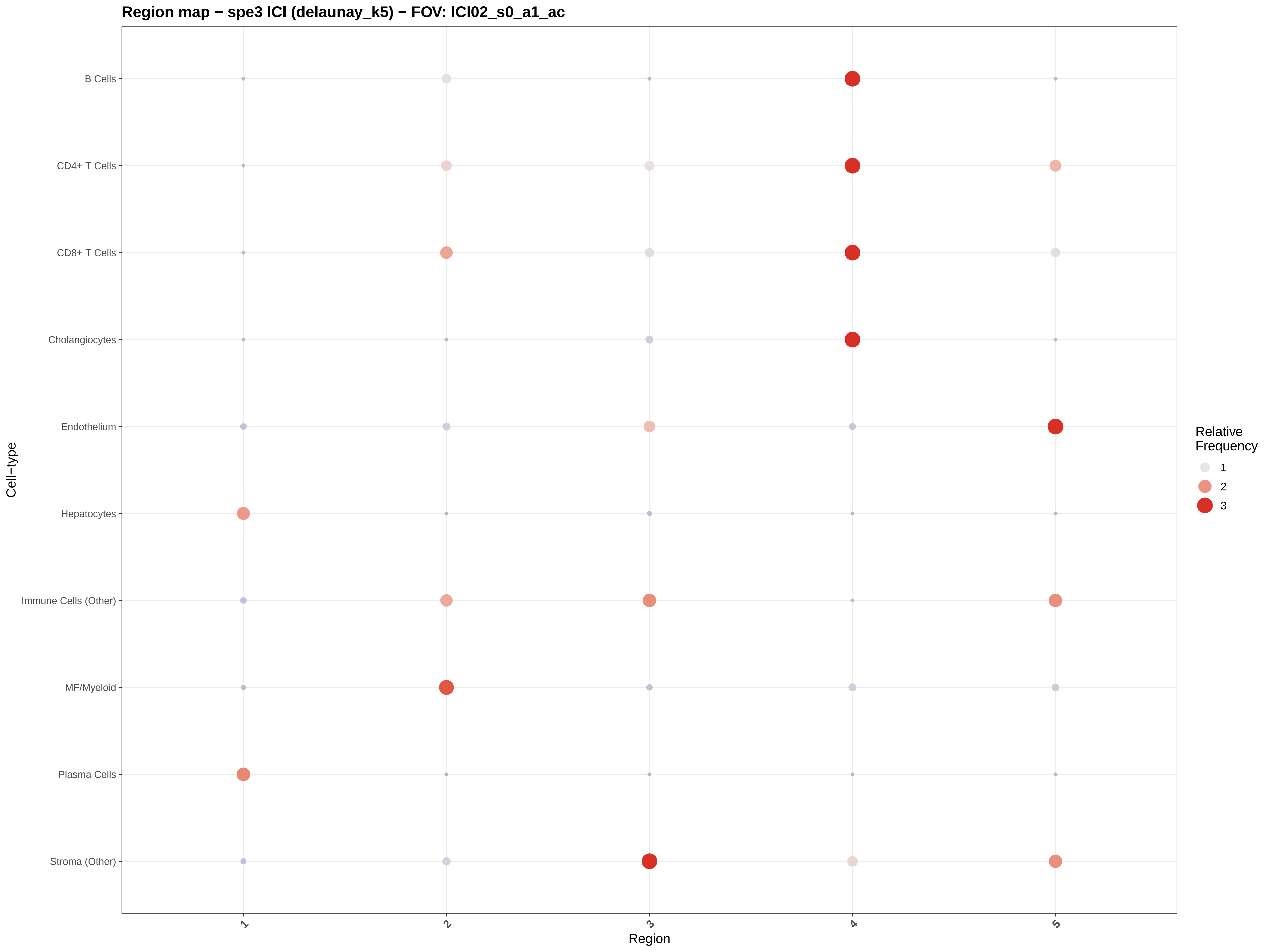

Region map − spe3 ICI (delaunay_k5) − FOV: ICI02_s0_a1_ac
B Cells
CD4+ T Cells
CD8+ T Cells
Cholangiocytes
Endothelium
Hepatocytes
Immune Cells (Other)
MF/Myeloid
Plasma Cells
Stroma (Other)
Cell−type
1
2
3
4
5
Region
Relative Frequency
1
2
3

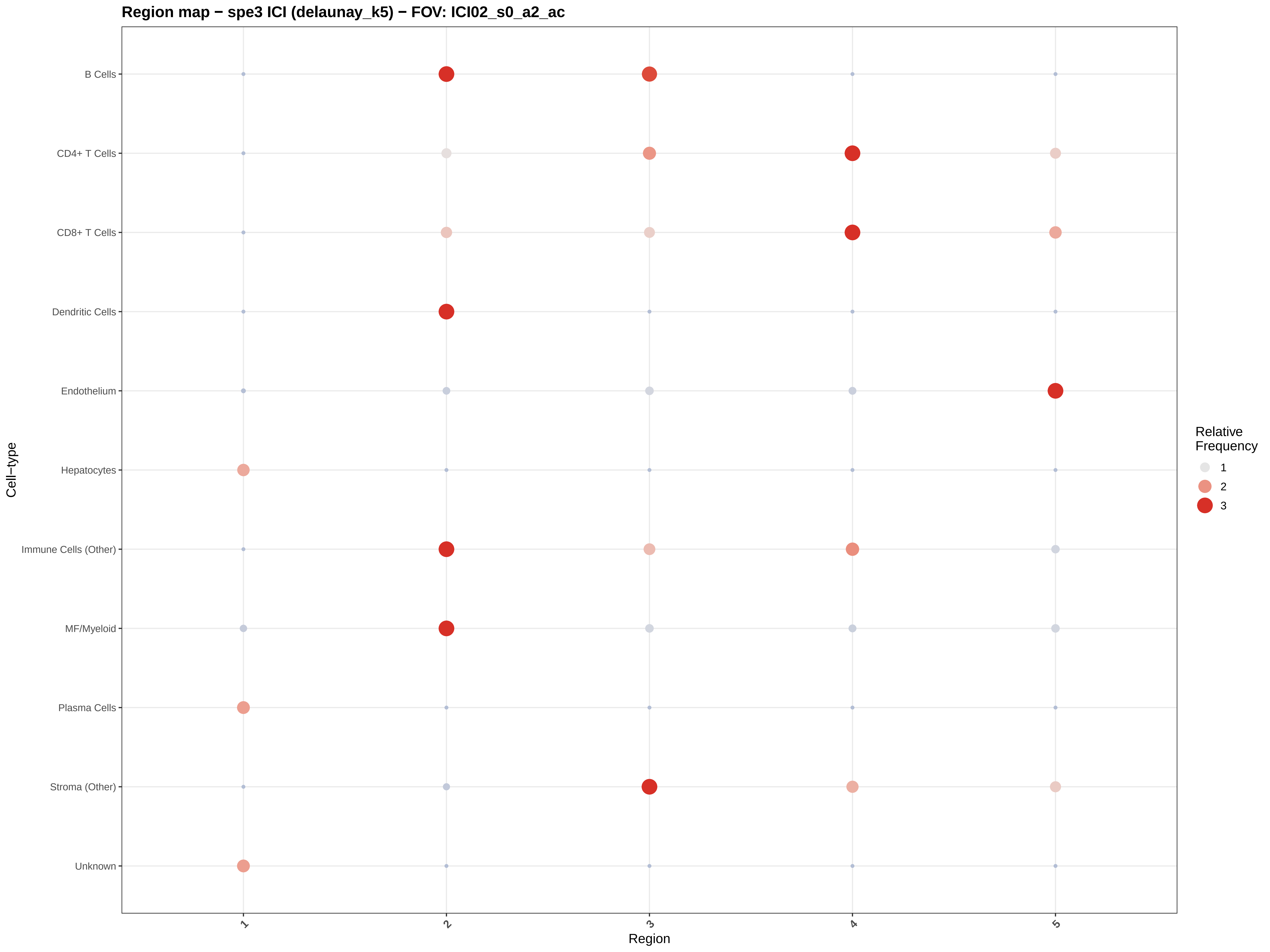

Region map − spe3 ICI (delaunay_k5) − FOV: ICI02_s0_a2_ac
B Cells
CD4+ T Cells
CD8+ T Cells
Dendritic Cells
Endothelium
Hepatocytes
Immune Cells (Other)
MF/Myeloid
Plasma Cells
Stroma (Other)
Unknown
Cell−type
1
2
3
4
5
Region
Relative Frequency
1
2
3

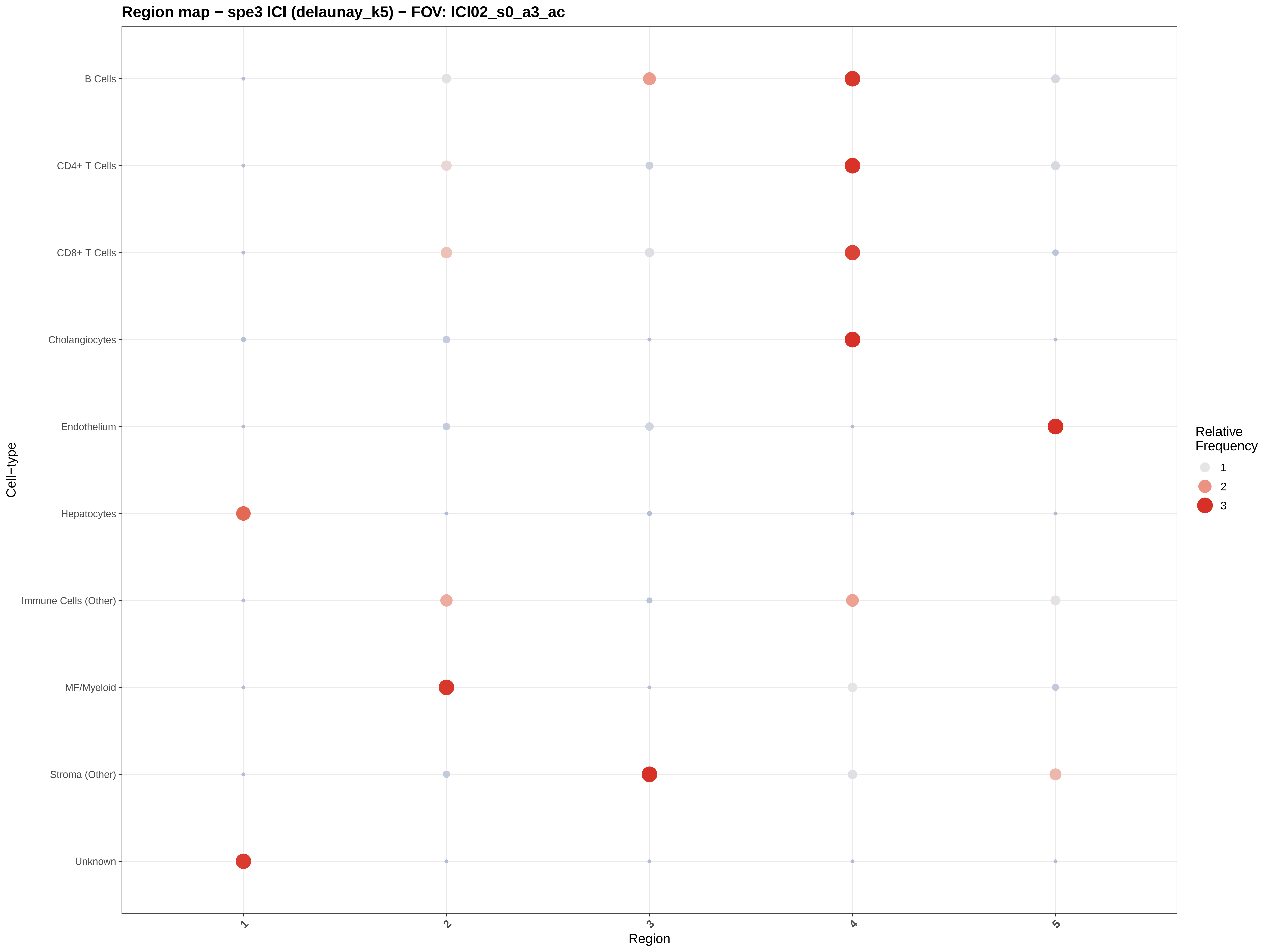

Region map − spe3 ICI (delaunay_k5) − FOV: ICI02_s0_a3_ac
B Cells
CD4+ T Cells
CD8+ T Cells
Cholangiocytes
Endothelium
Hepatocytes
Immune Cells (Other)
MF/Myeloid
Stroma (Other)
Unknown
Cell−type
1
2
3
4
5
Region
Relative
Frequency
1
2
3

Region map − spe3 ICI (delaunay_k5) − FOV: ICI02_s0_a4_ac

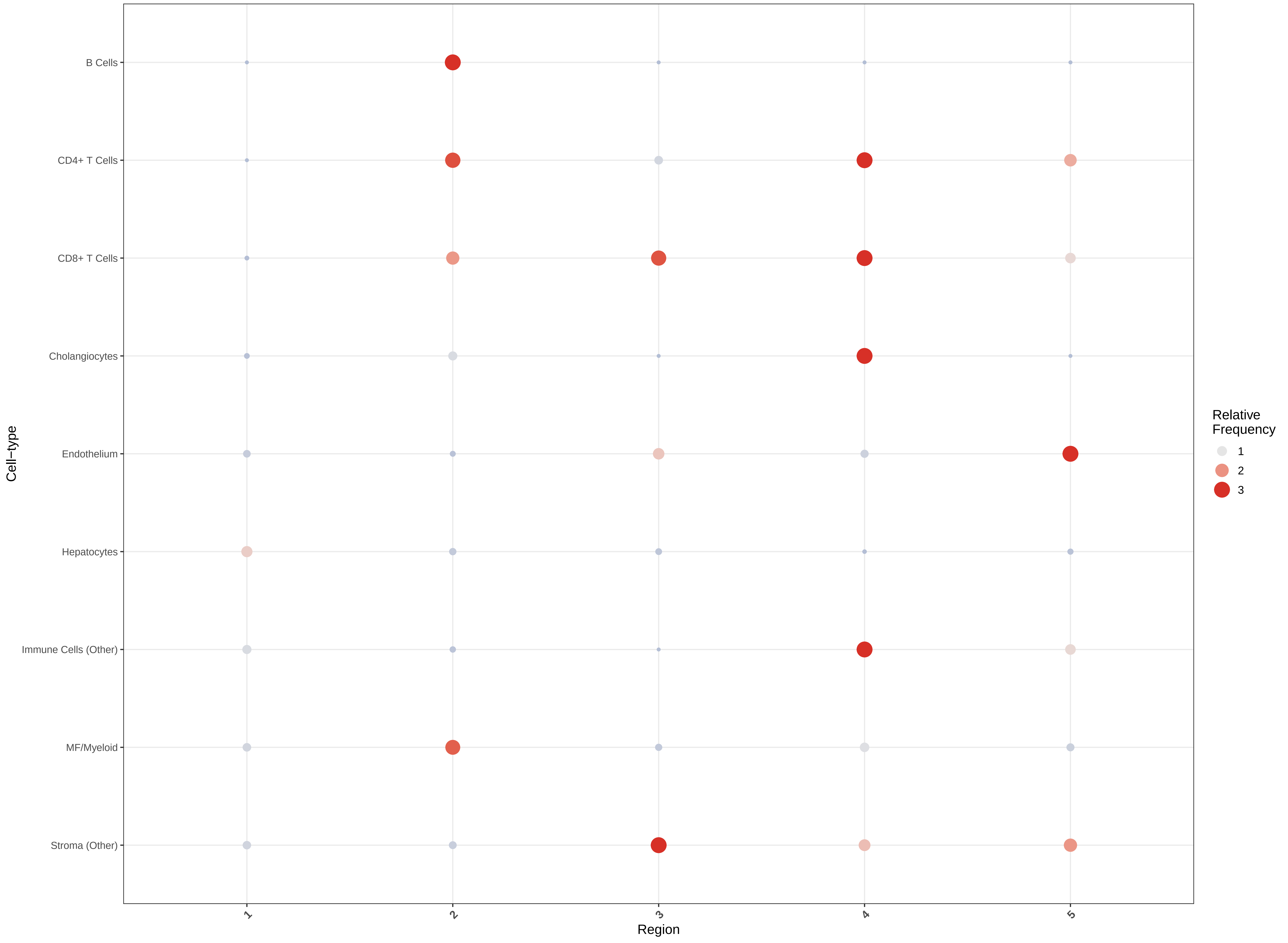

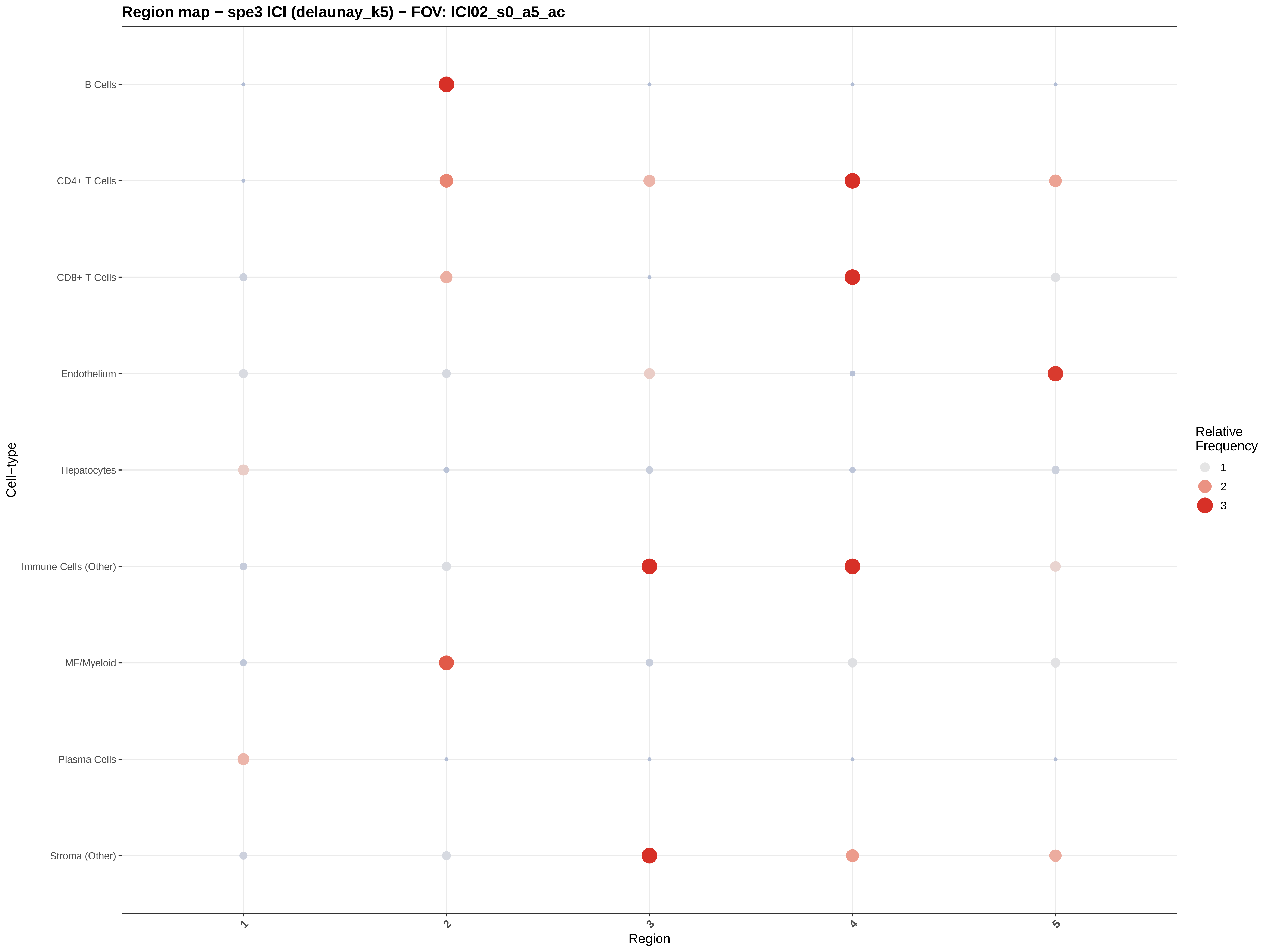
Region map − spe3 ICI (delaunay_k5) − FOV: ICI02_s0_a5_ac
B Cells
CD4+ T Cells
CD8+ T Cells
Endothelium
Hepatocytes
Immune Cells (Other)
MF/Myeloid
Plasma Cells
Stroma (Other)
Cell−type
1
2
3
4
5
Region
Relative Frequency
1
2
3

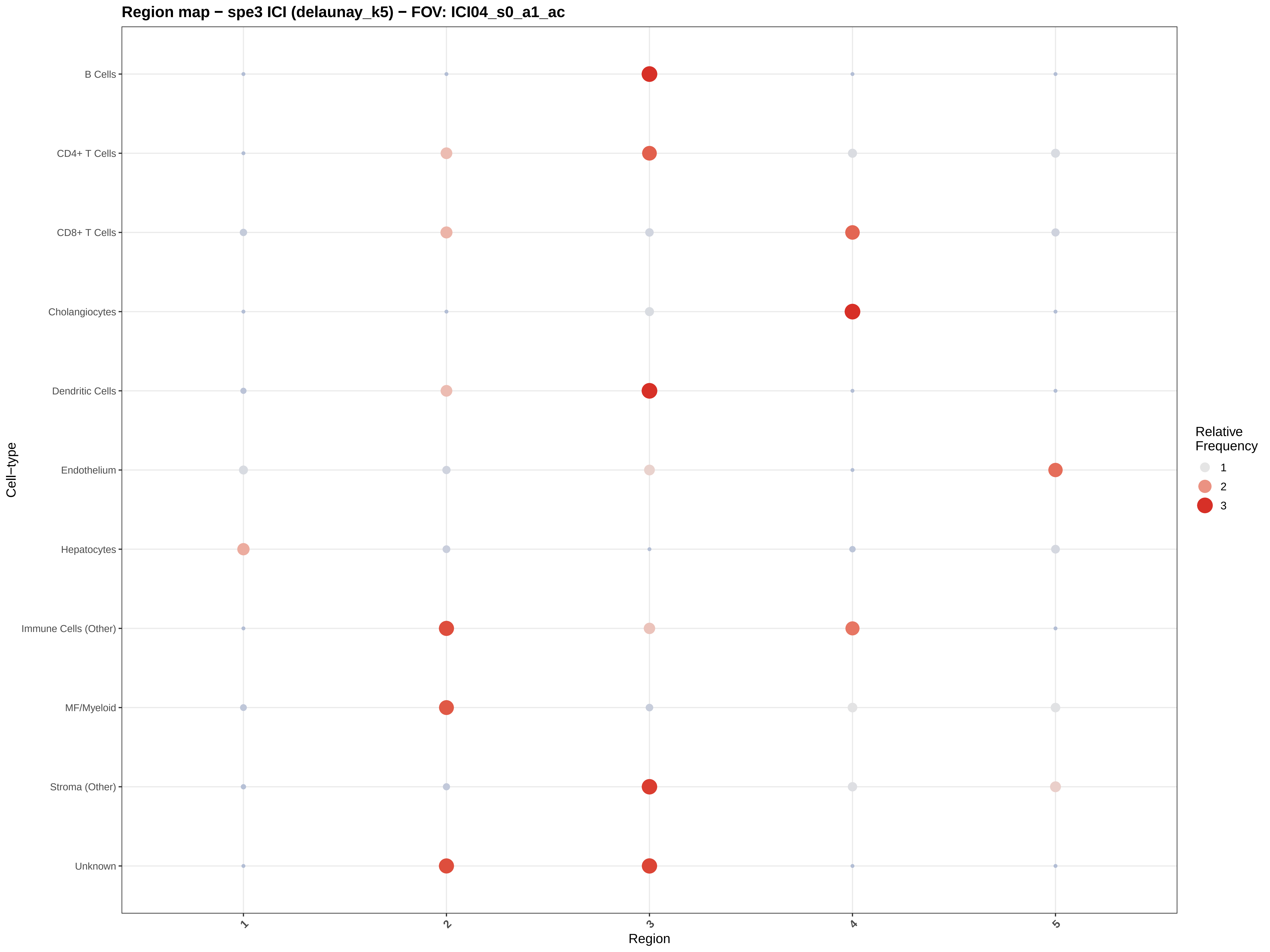

Region map − spe3 ICI (delaunay_k5) − FOV: ICI04_s0_a1_ac
B Cells
CD4+ T Cells
CD8+ T Cells
Cholangiocytes
Dendritic Cells
Endothelium
Hepatocytes
Immune Cells (Other)
MF/Myeloid
Stroma (Other)
Unknown
Cell−type
1
2
3
4
5
Region
Relative Frequency
1
2
3

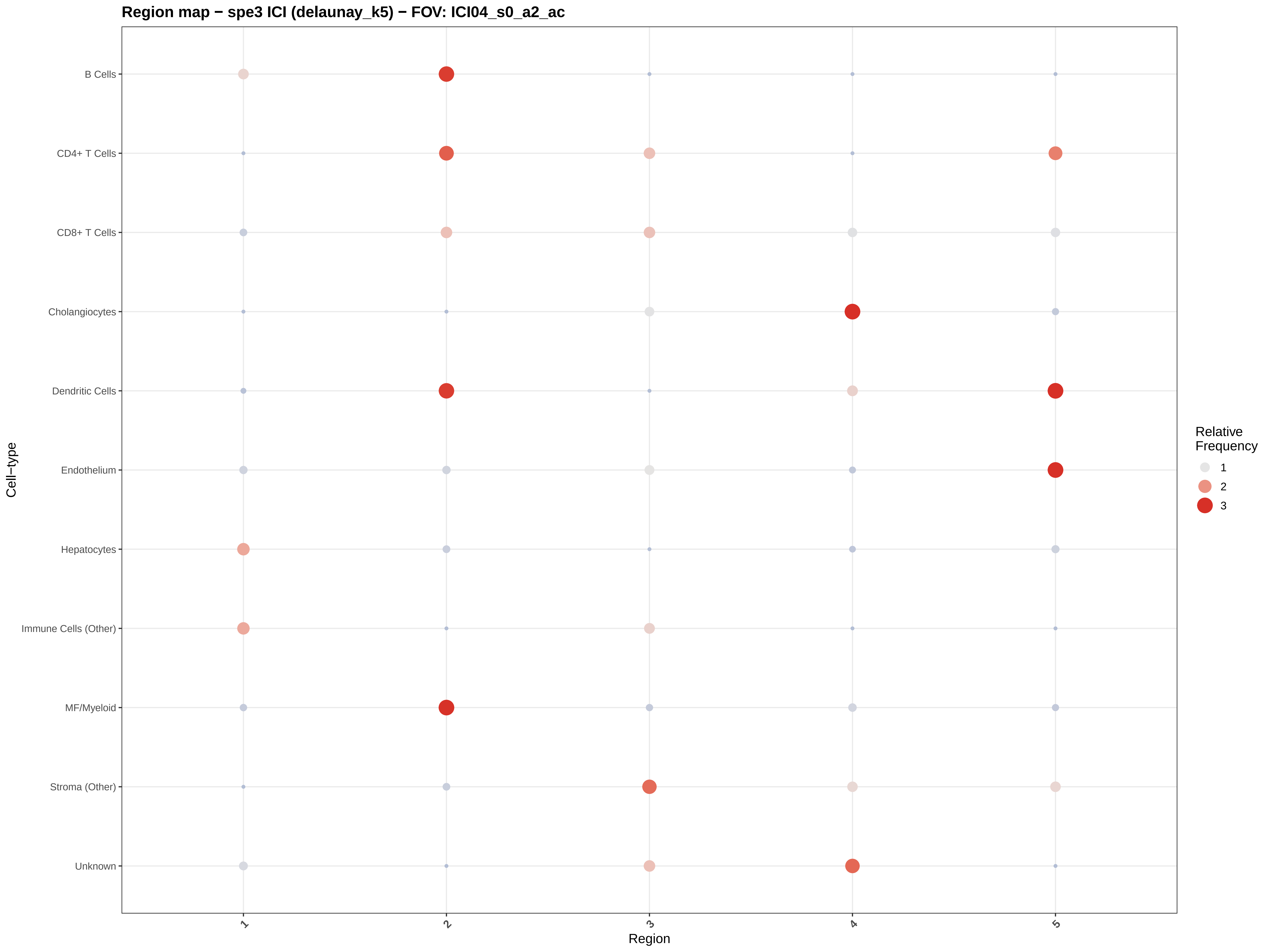

Region map − spe3 ICI (delaunay_k5) − FOV: ICI04_s0_a2_ac
B Cells
CD4+ T Cells
CD8+ T Cells
Cholangiocytes
Dendritic Cells
Endothelium
Hepatocytes
Immune Cells (Other)
MF/Myeloid
Stroma (Other)
Unknown
Cell−type
1
2
3
4
5
Region
Relative Frequency
1
2
3

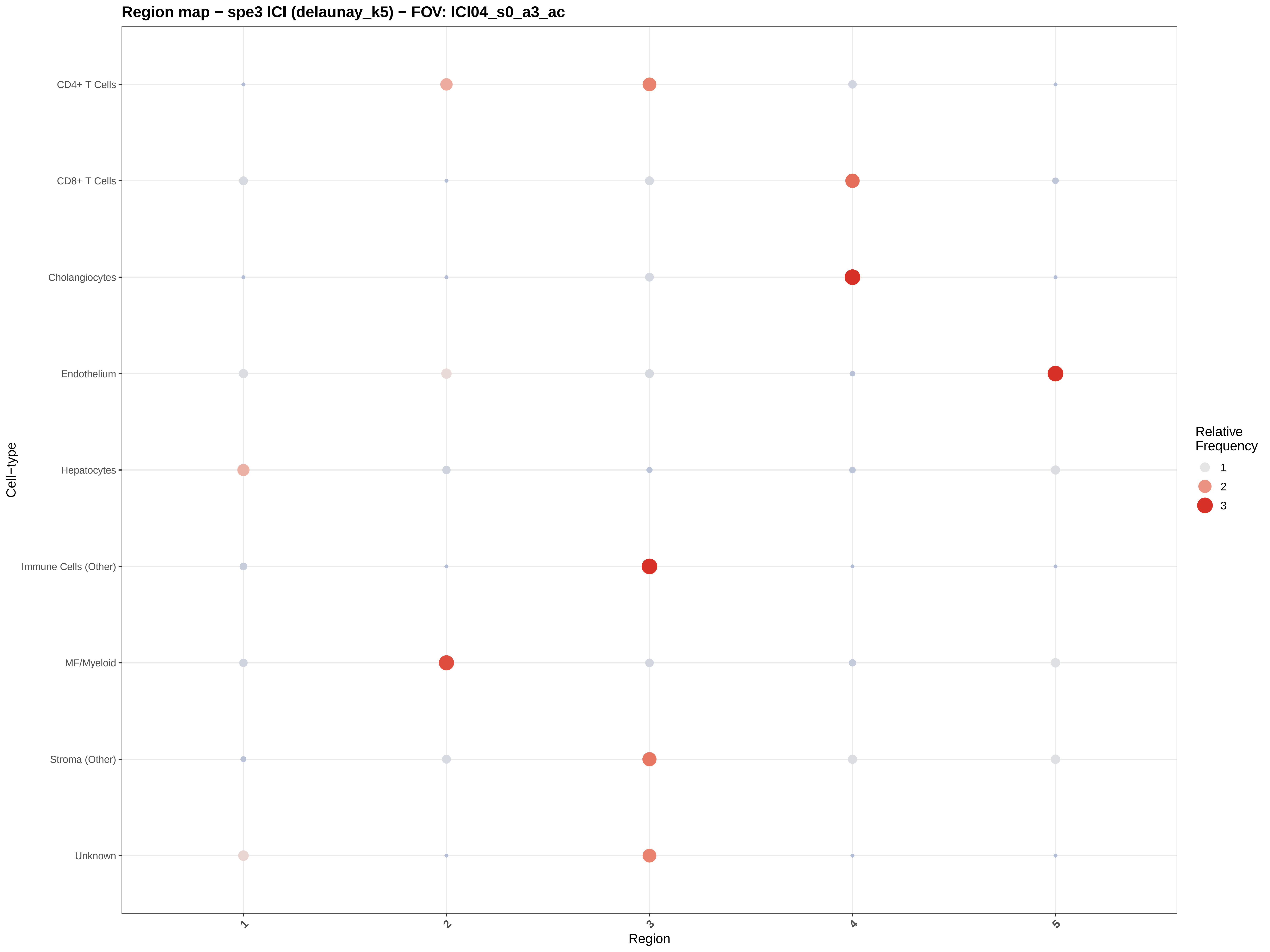

Region map − spe3 ICI (delaunay_k5) − FOV: ICI04_s0_a3_ac
CD4+ T Cells
CD8+ T Cells
Cholangiocytes
Endothelium
Hepatocytes
Immune Cells (Other)
MF/Myeloid
Stroma (Other)
Unknown
Cell−type
1
2
3
4
5
Region
Relative Frequency
1
2
3

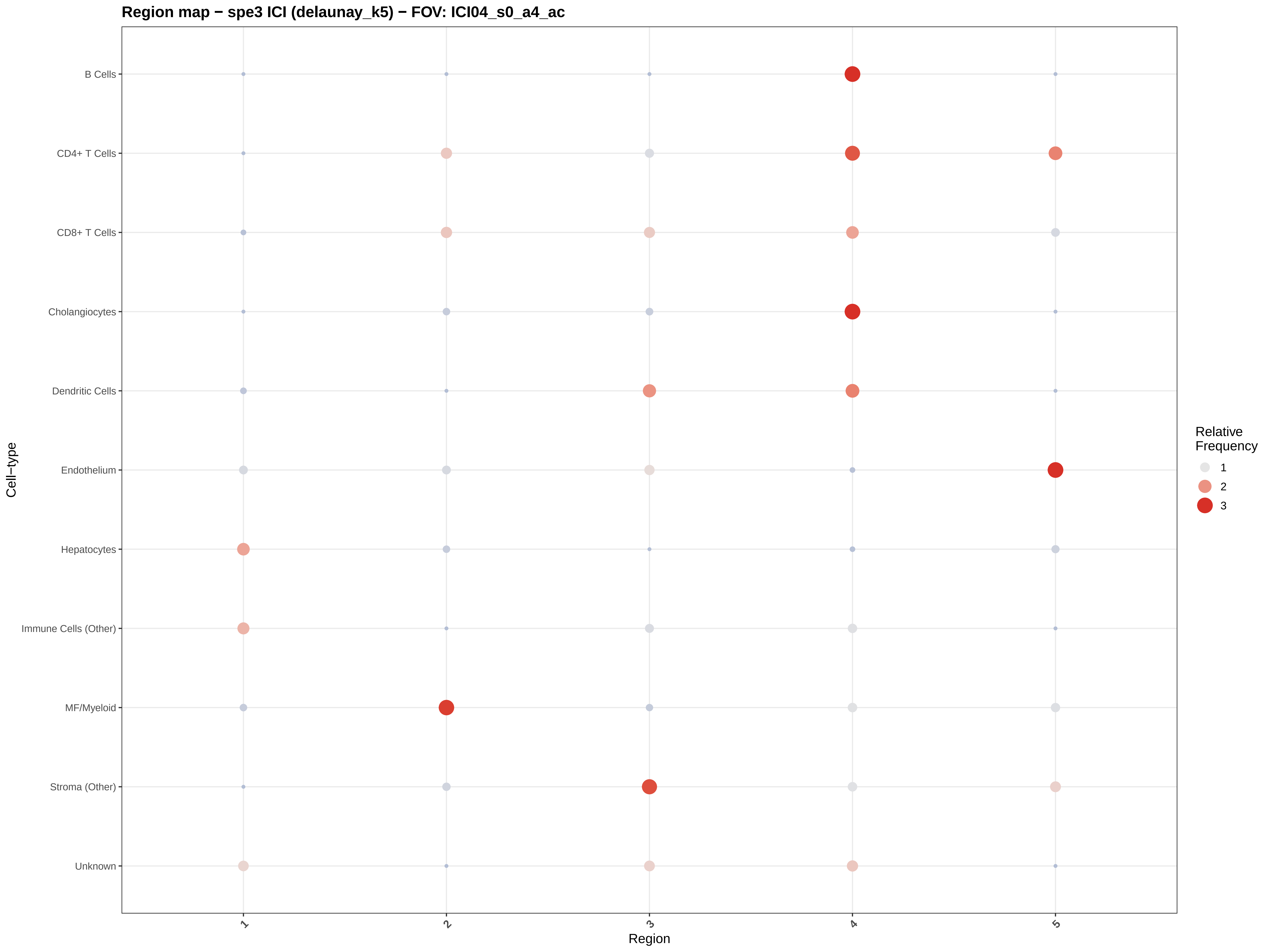

Region map − spe3 ICI (delaunay_k5) − FOV: ICI04_s0_a4_ac
B Cells
CD4+ T Cells
CD8+ T Cells
Cholangiocytes
Dendritic Cells
Endothelium
Hepatocytes
Immune Cells (Other)
MF/Myeloid
Stroma (Other)
Unknown
Cell−type
1
2
3
4
5
Region
Relative Frequency
1
2
3

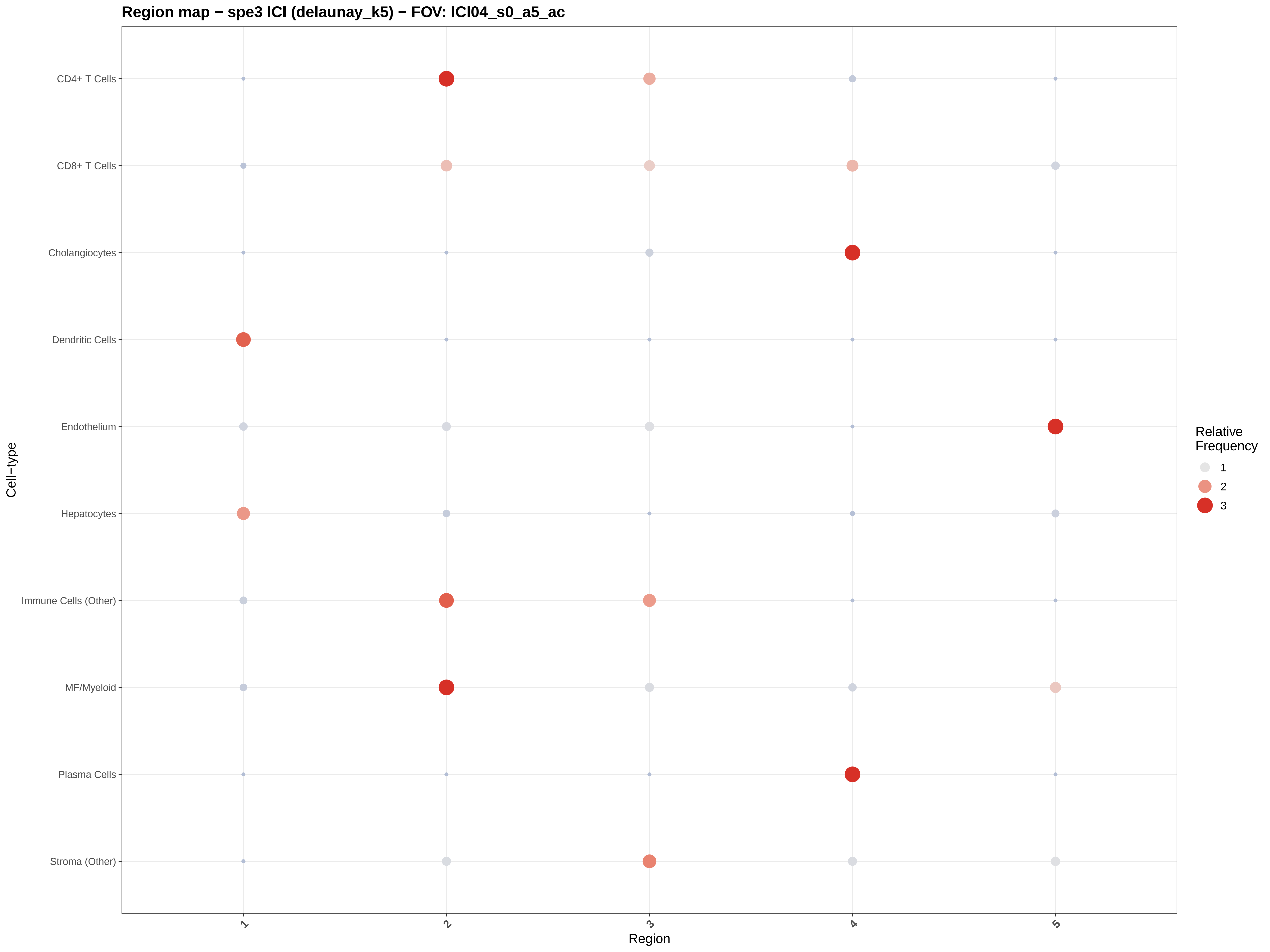
Region map − spe3 ICI (delaunay_k5) − FOV: ICI04_s0_a5_ac
CD4+ T Cells
CD8+ T Cells
Cholangiocytes
Dendritic Cells
Endothelium
Hepatocytes
Immune Cells (Other)
MF/Myeloid
Plasma Cells
Stroma (Other)
Cell−type
1
2
3
4
5
Region
Relative Frequency
1
2
3

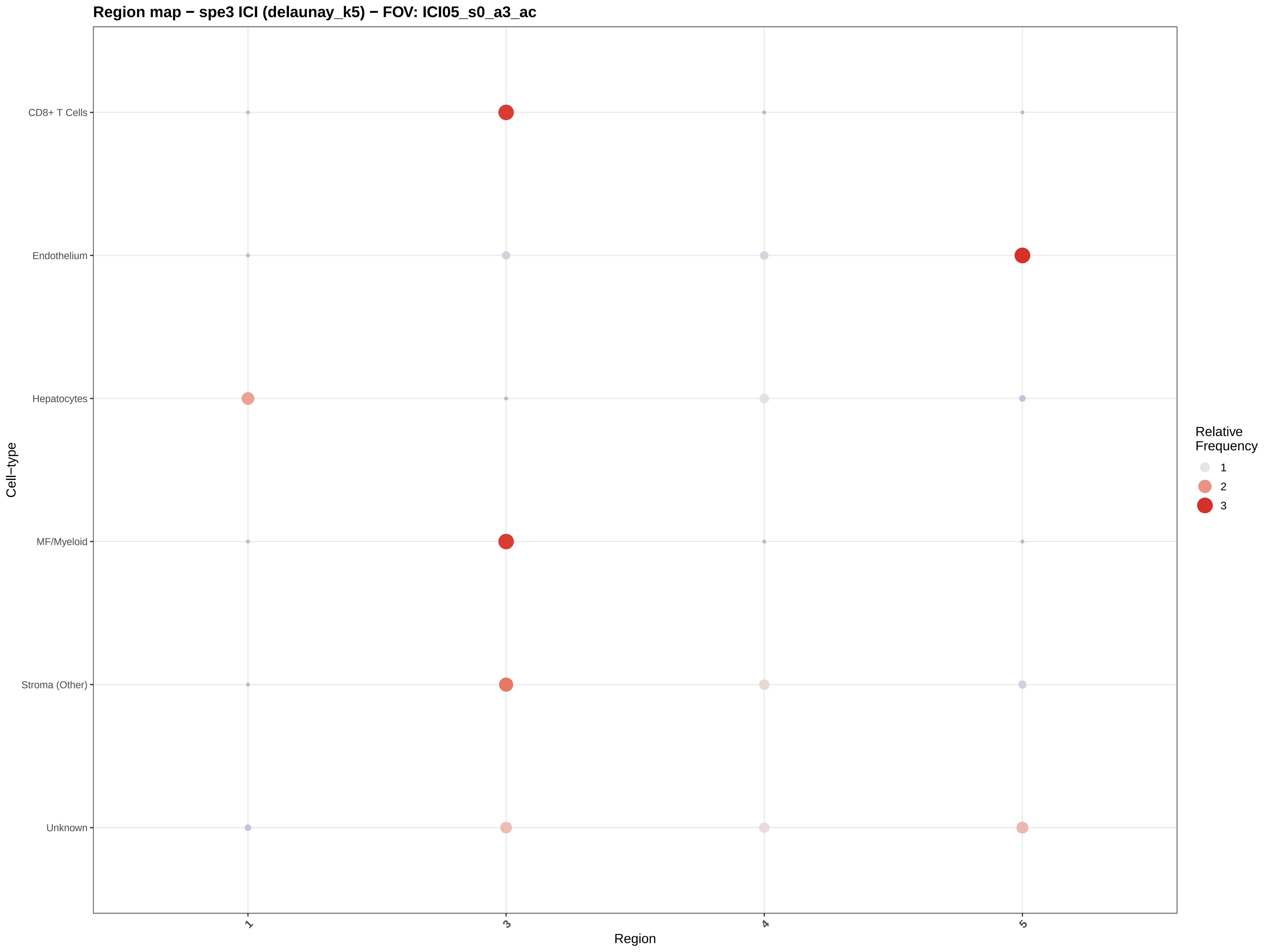

Region map − spe3 ICI (delaunay_k5) − FOV: ICI05_s0_a3_ac
CD8+ T Cells
Endothelium
Hepatocytes
MF/Myeloid
Stroma (Other)
Unknown
Cell−type
1
3
4
5
Region
Relative Frequency
1
2
3

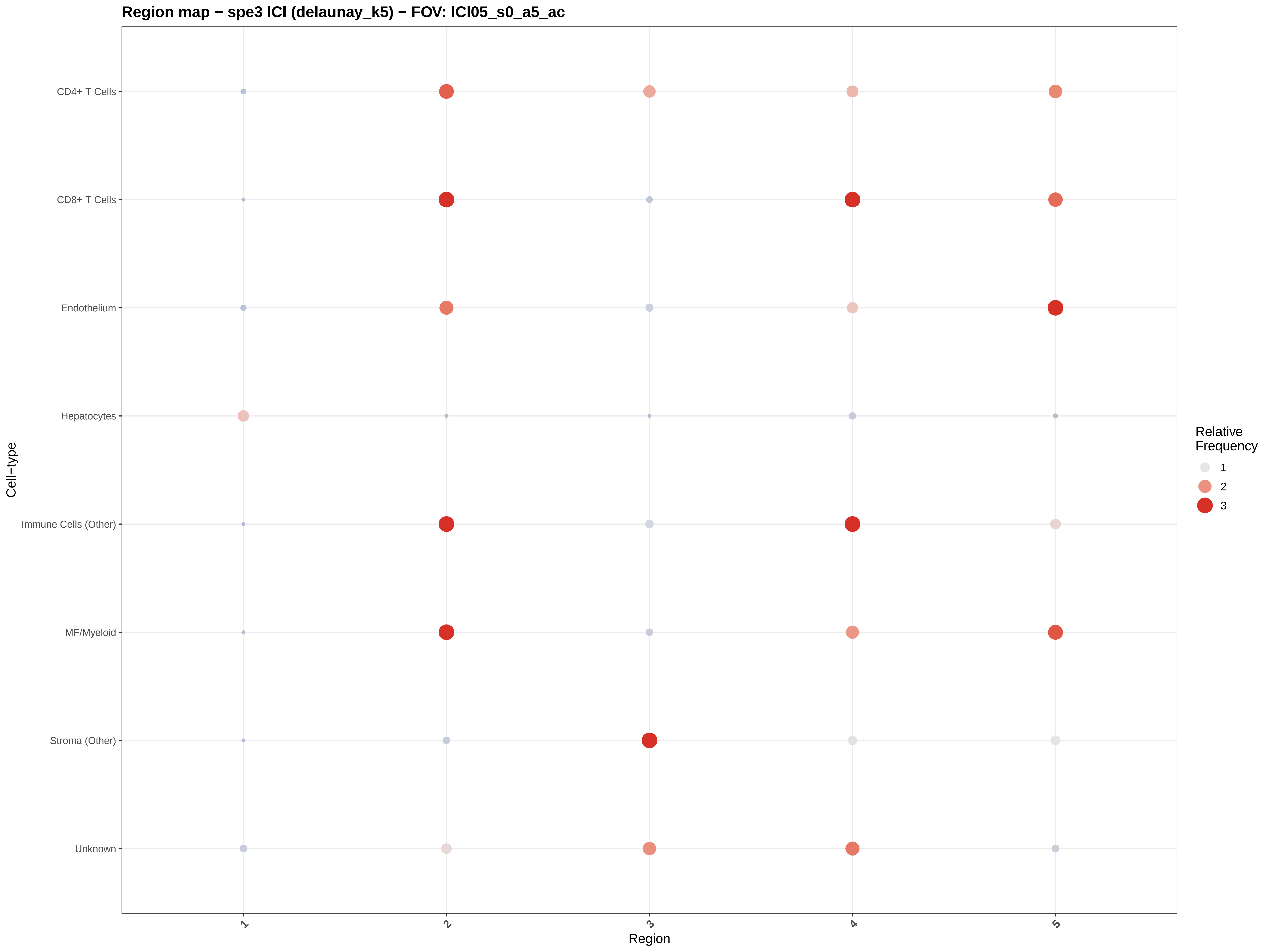

Region map − spe3 ICI (delaunay_k5) − FOV: ICI05_s0_a5_ac
CD4+ T Cells
CD8+ T Cells
Endothelium
Hepatocytes
Immune Cells (Other)
MF/Myeloid
Stroma (Other)
Unknown
Cell−type
1
2
3
4
5
Region
Relative Frequency
1
2
3

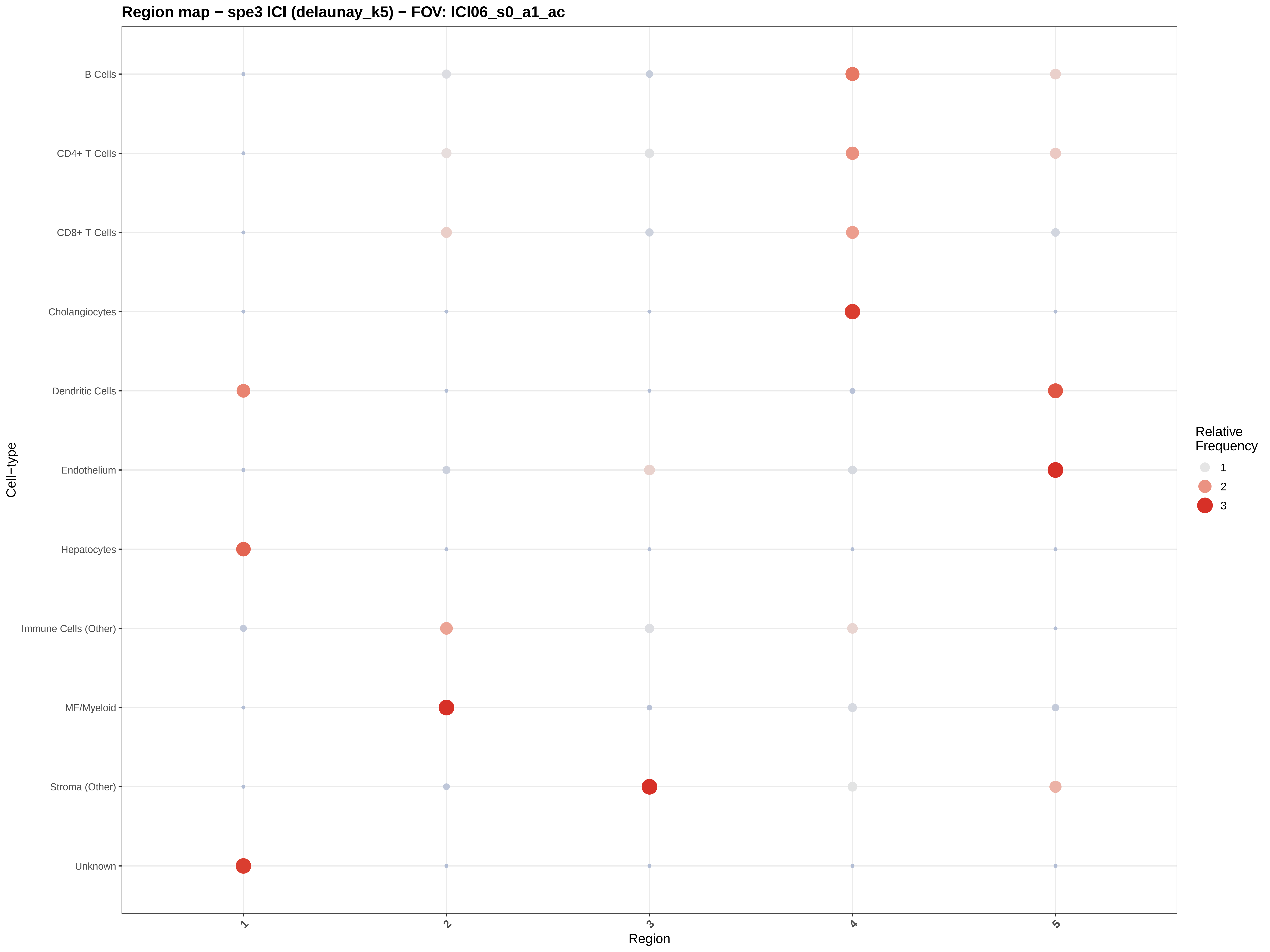
Region map − spe3 ICI (delaunay_k5) − FOV: ICI06_s0_a1_ac
B Cells
CD4+ T Cells
CD8+ T Cells
Cholangiocytes
Dendritic Cells
Endothelium
Hepatocytes
Immune Cells (Other)
MF/Myeloid
Stroma (Other)
Unknown
Cell−type
1
2
3
4
5
Region
Relative
Frequency
1
2
3

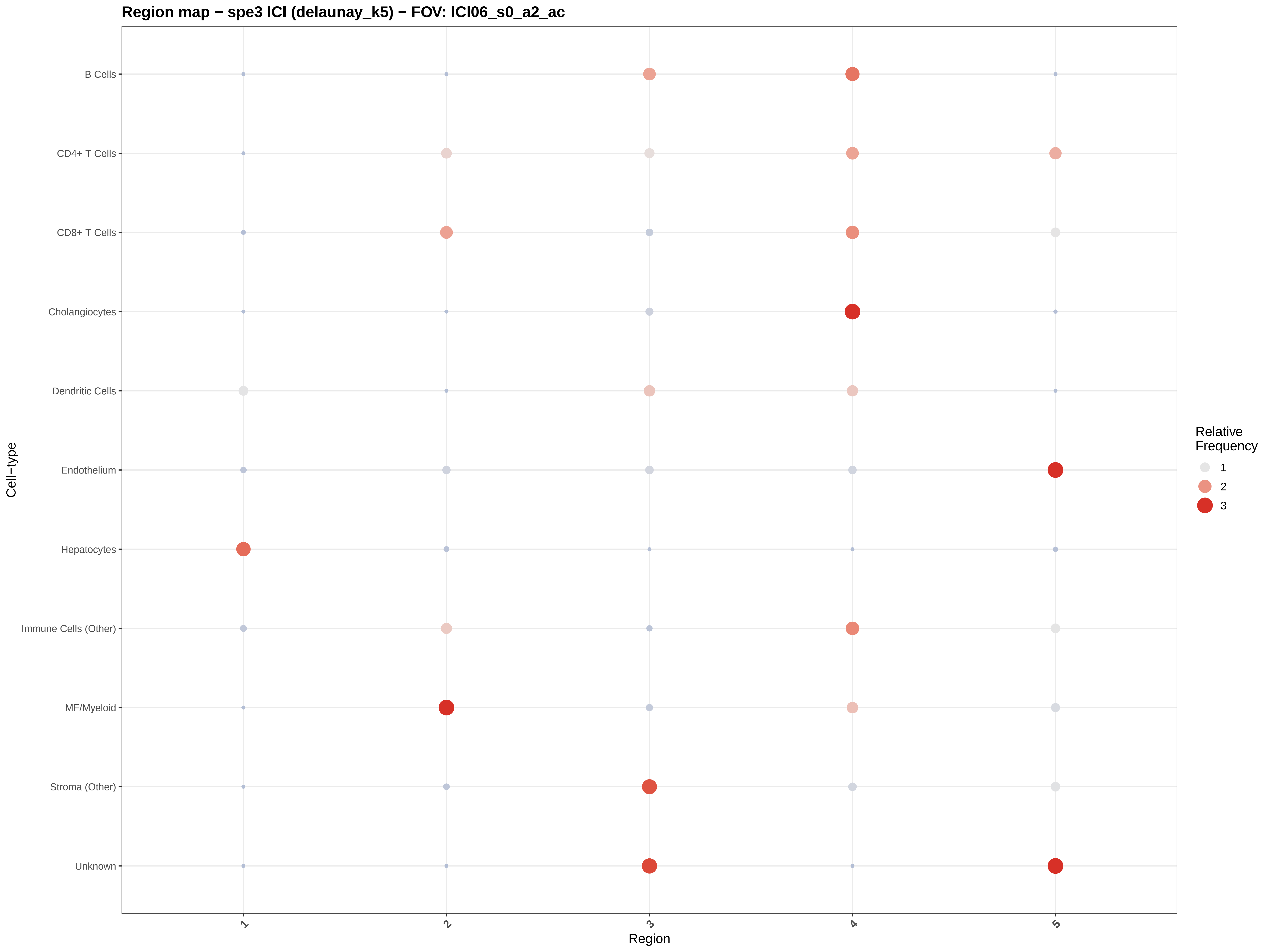

Region map − spe3 ICI (delaunay_k5) − FOV: ICI06_s0_a2_ac
B Cells
CD4+ T Cells
CD8+ T Cells
Cholangiocytes
Dendritic Cells
Endothelium
Hepatocytes
Immune Cells (Other)
MF/Myeloid
Stroma (Other)
Unknown
Cell−type
1
2
3
4
5
Region
Relative Frequency
1
2
3

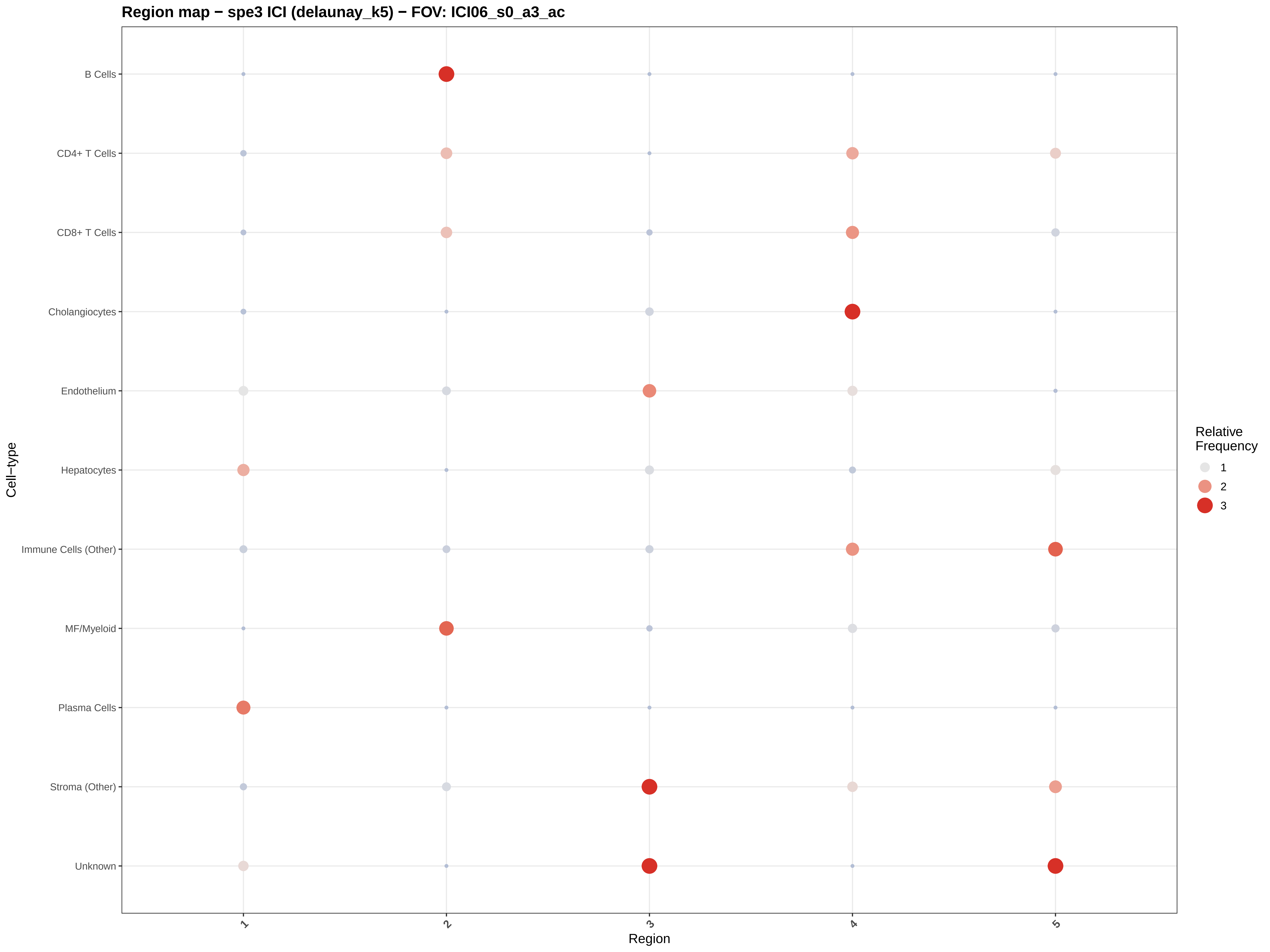

Region map − spe3 ICI (delaunay_k5) − FOV: ICI06_s0_a3_ac
B Cells
CD4+ T Cells
CD8+ T Cells
Cholangiocytes
Endothelium
Hepatocytes
Immune Cells (Other)
MF/Myeloid
Plasma Cells
Stroma (Other)
Unknown
Cell−type
1
2
3
4
5
Region
Relative Frequency
1
2
3

Region map − spe3 ICI (delaunay_k5) − FOV: ICI06_s0_a4_ac

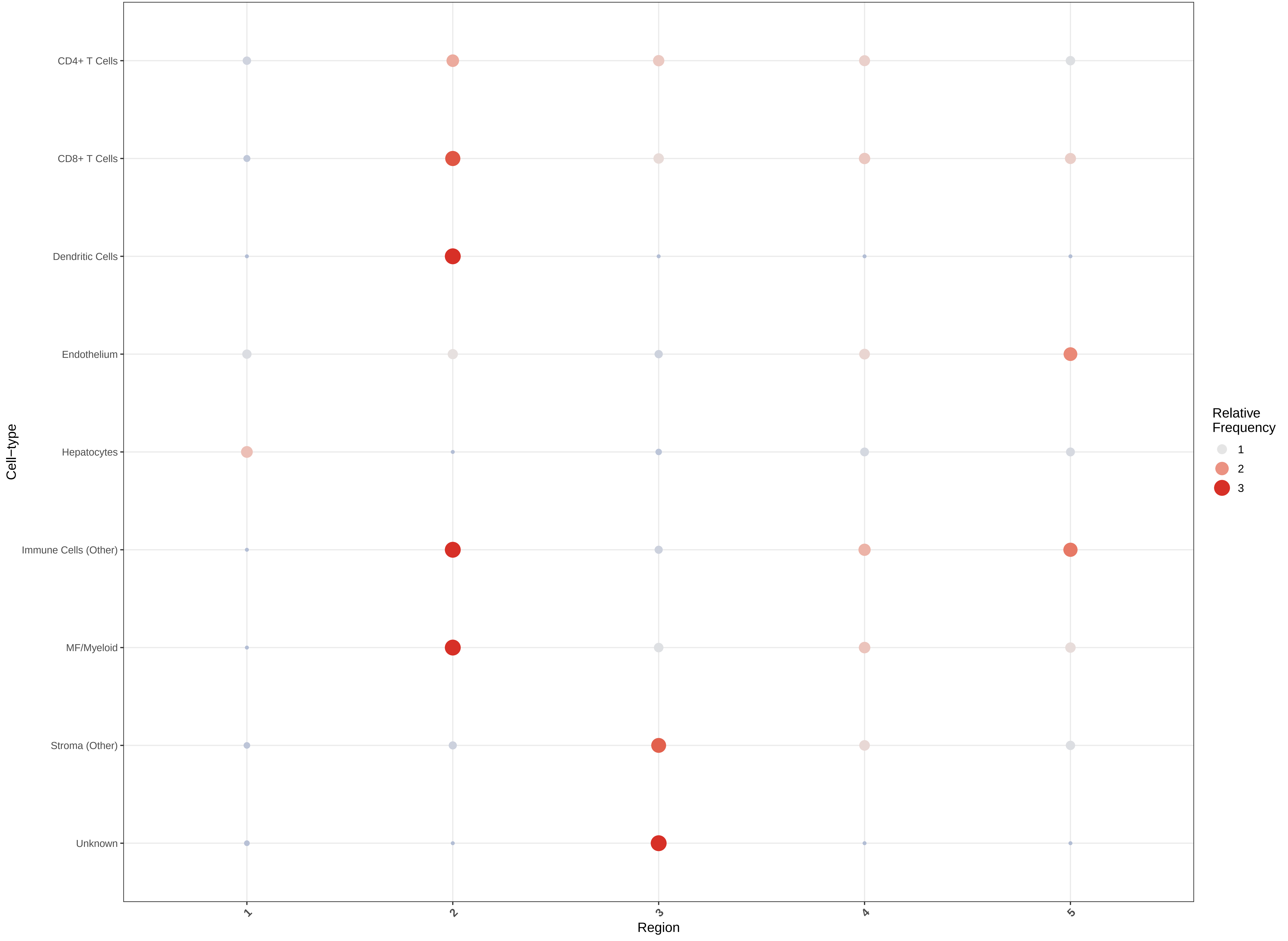

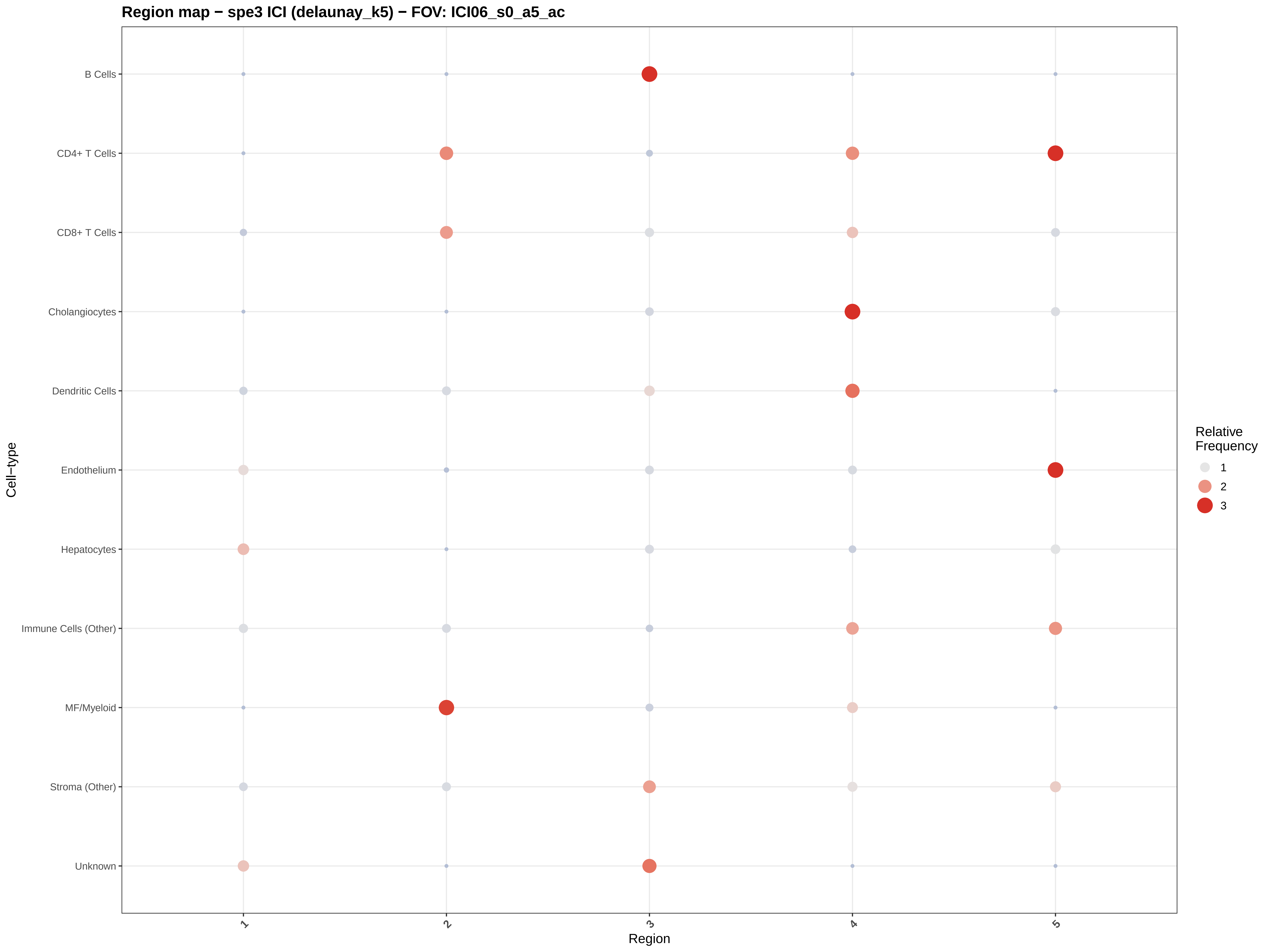
Region map − spe3 ICI (delaunay_k5) − FOV: ICI06_s0_a5_ac
B Cells
CD4+ T Cells
CD8+ T Cells
Cholangiocytes
Dendritic Cells
Endothelium
Hepatocytes
Immune Cells (Other)
MF/Myeloid
Stroma (Other)
Unknown
Cell−type
1
2
3
4
5
Region
Relative Frequency
1
2
3

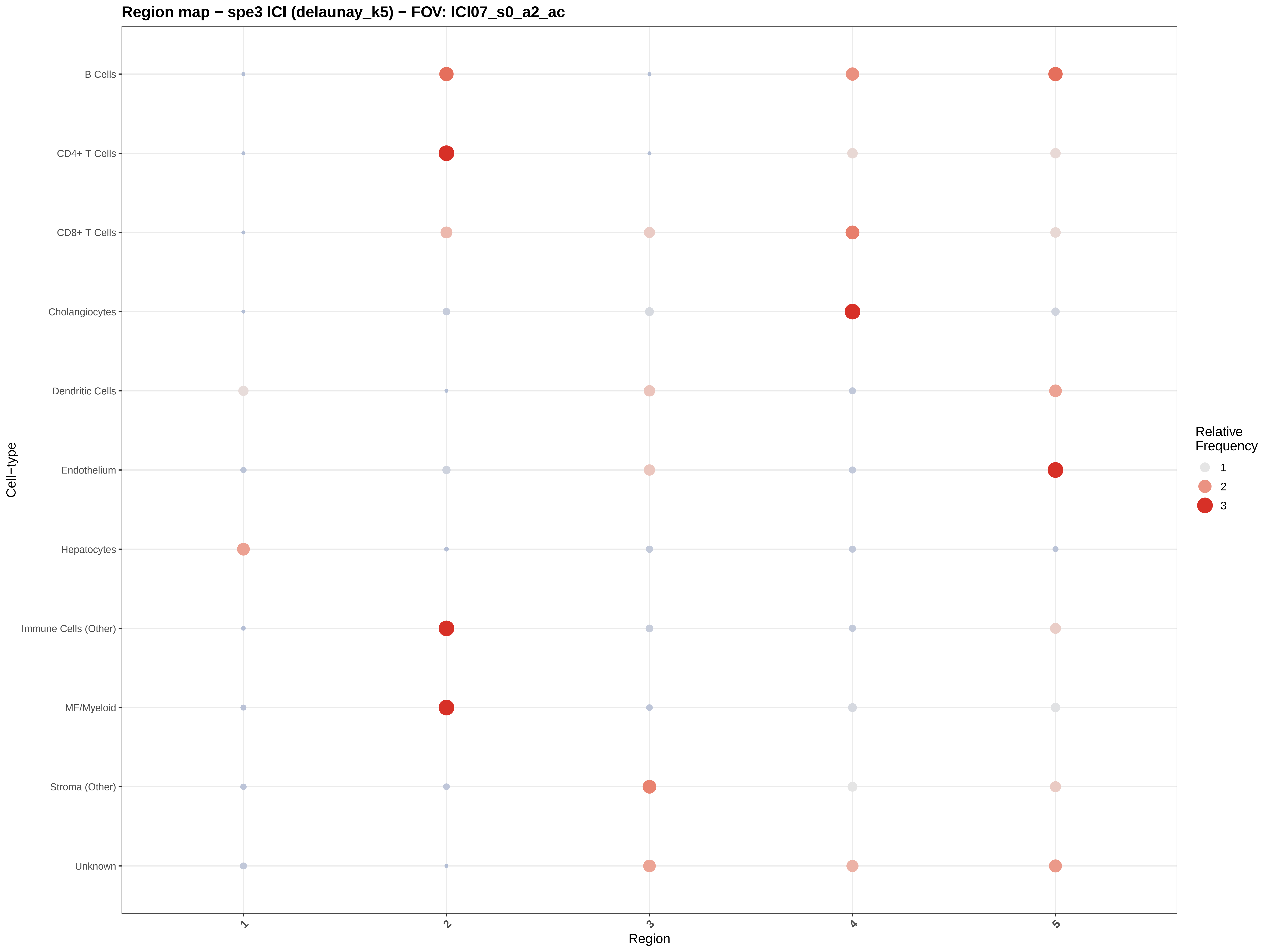

Region map − spe3 ICI (delaunay_k5) − FOV: ICI07_s0_a2_ac
B Cells
CD4+ T Cells
CD8+ T Cells
Cholangiocytes
Dendritic Cells
Endothelium
Hepatocytes
Immune Cells (Other)
MF/Myeloid
Stroma (Other)
Unknown
Cell−type
1
2
3
4
5
Region
Relative Frequency
1
2
3

#### Region map − spe3 ICI (delaunay_k5) − FOV: ICI08_s0_a1_ac

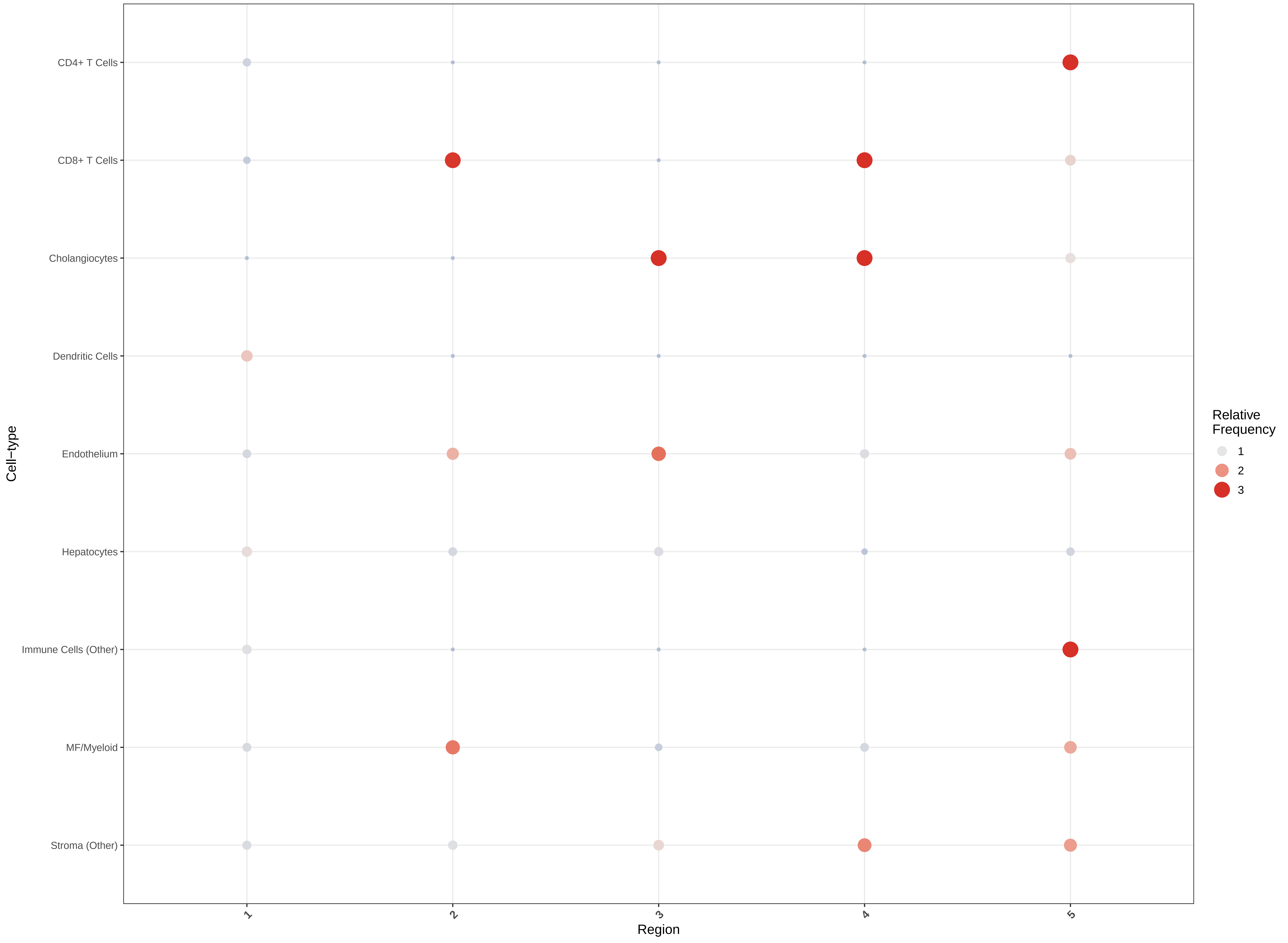

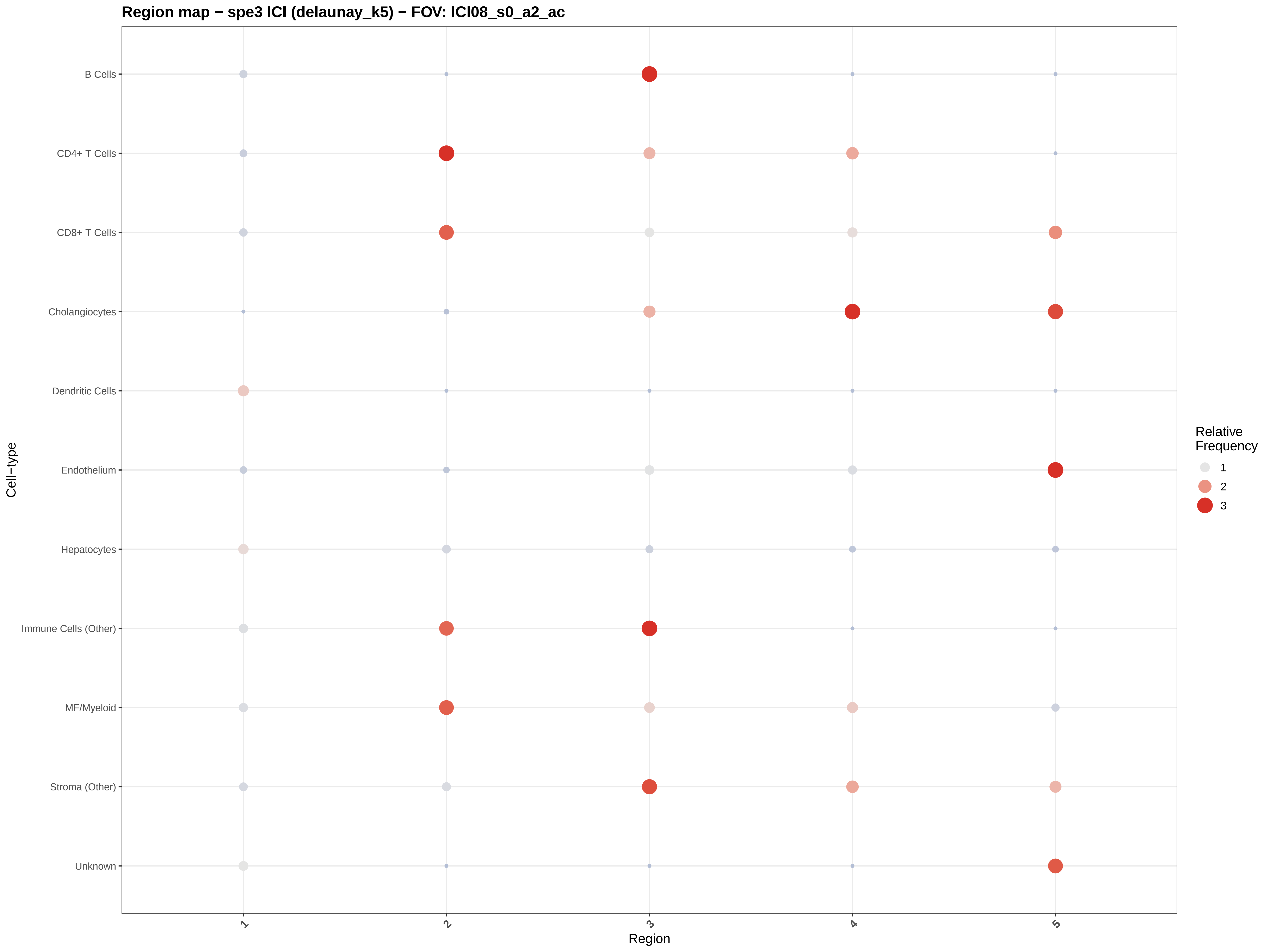

Region map − spe3 ICI (delaunay_k5) − FOV: ICI08_s0_a2_ac
B Cells
CD4+ T Cells
CD8+ T Cells
Cholangiocytes
Dendritic Cells
Endothelium
Hepatocytes
Immune Cells (Other)
MF/Myeloid
Stroma (Other)
Unknown
Cell−type
1
2
3
4
5
Region
Relative Frequency
1
2
3

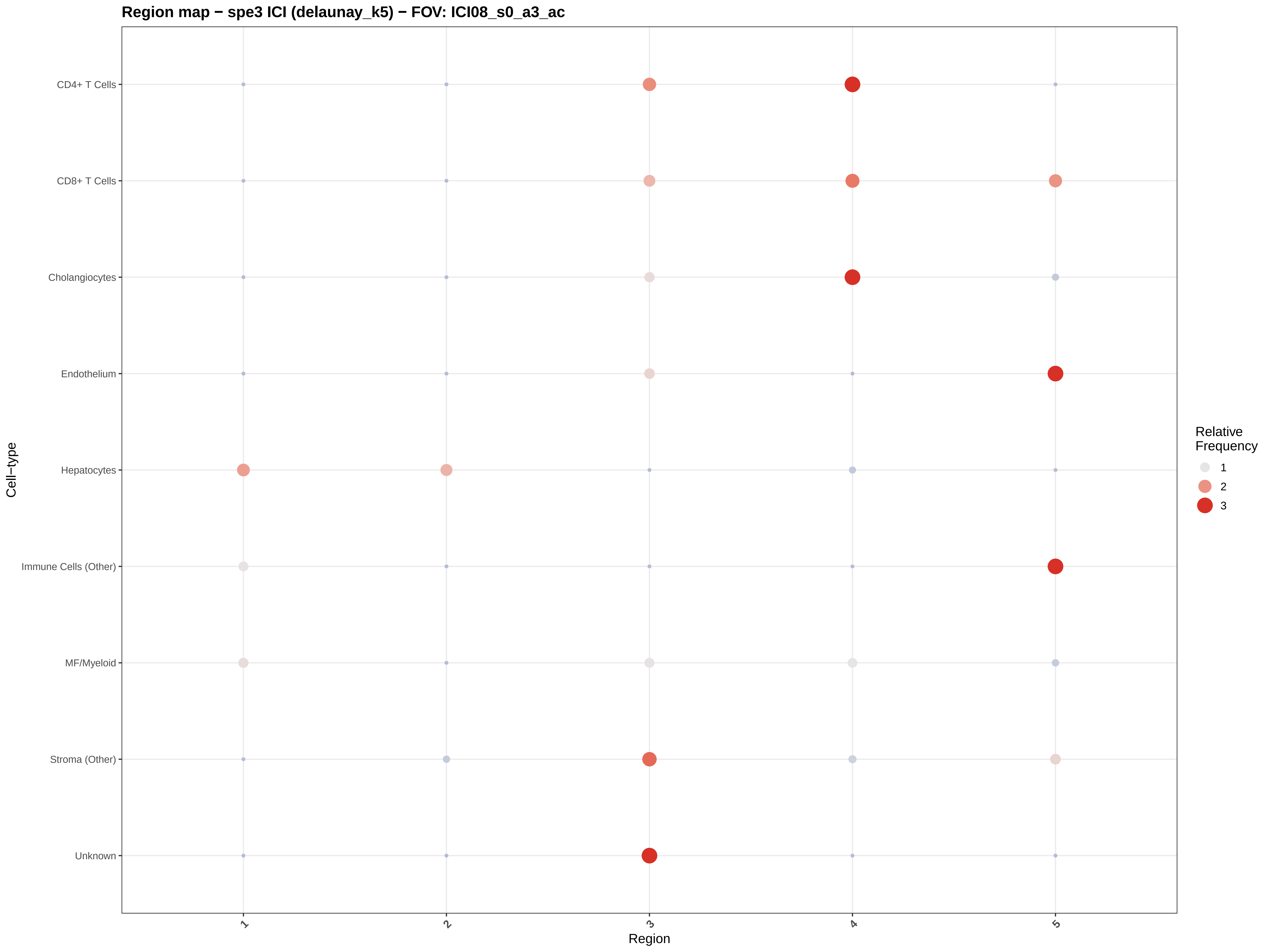
Region map − spe3 ICI (delaunay_k5) − FOV: ICI08_s0_a3_ac
CD4+ T Cells
CD8+ T Cells
Cholangiocytes
Endothelium
Hepatocytes
Immune Cells (Other)
MF/Myeloid
Stroma (Other)
Unknown
Cell−type
1
2
3
4
5
Region
Relative Frequency
1
2
3

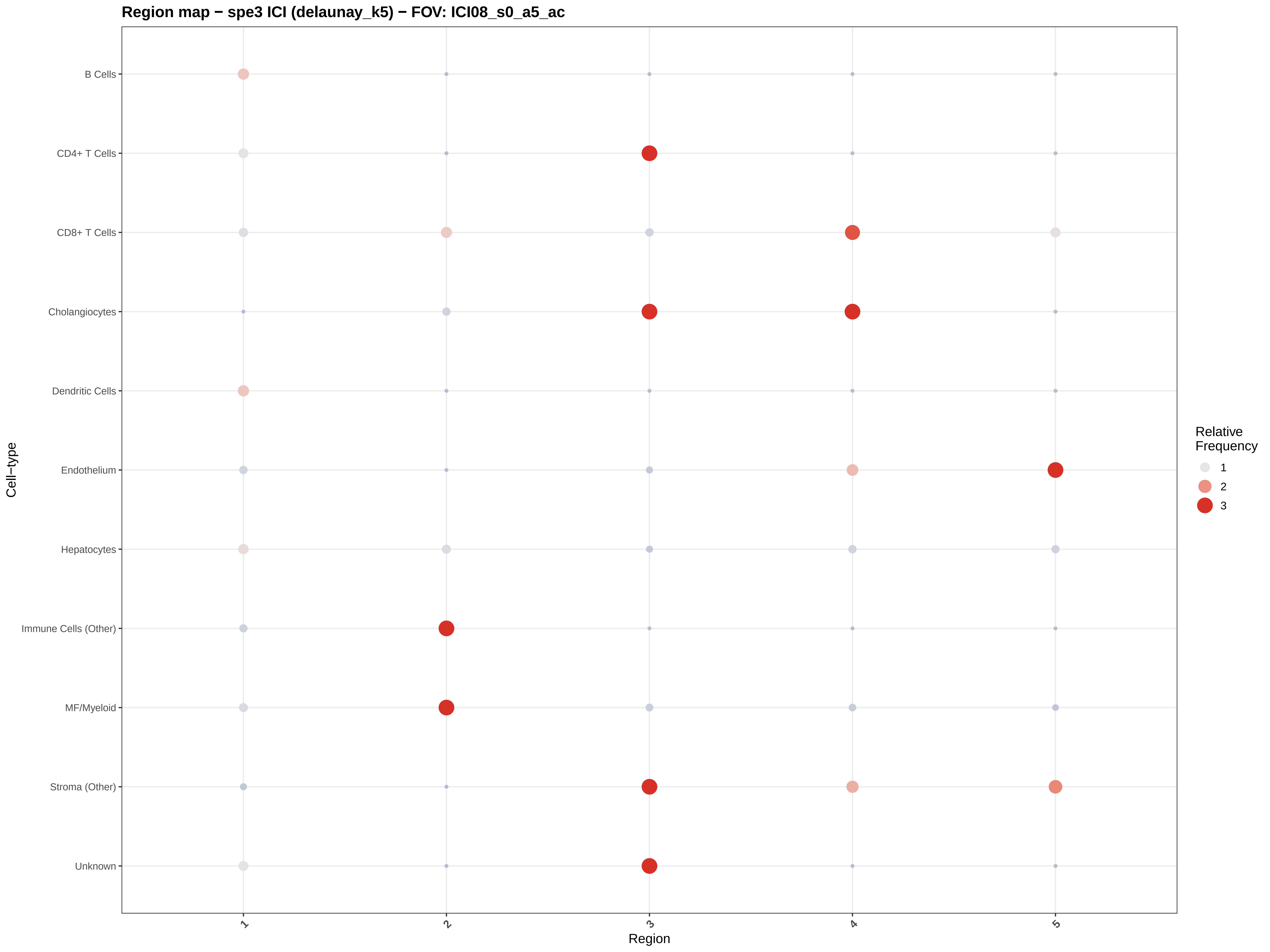
Region map − spe3 ICI (delaunay_k5) − FOV: ICI08_s0_a5_ac
B Cells
CD4+ T Cells
CD8+ T Cells
Cholangiocytes
Dendritic Cells
Endothelium
Hepatocytes
Immune Cells (Other)
MF/Myeloid
Stroma (Other)
Unknown
Cell−type
1
2
3
4
5
Region
Relative Frequency
1
2
3

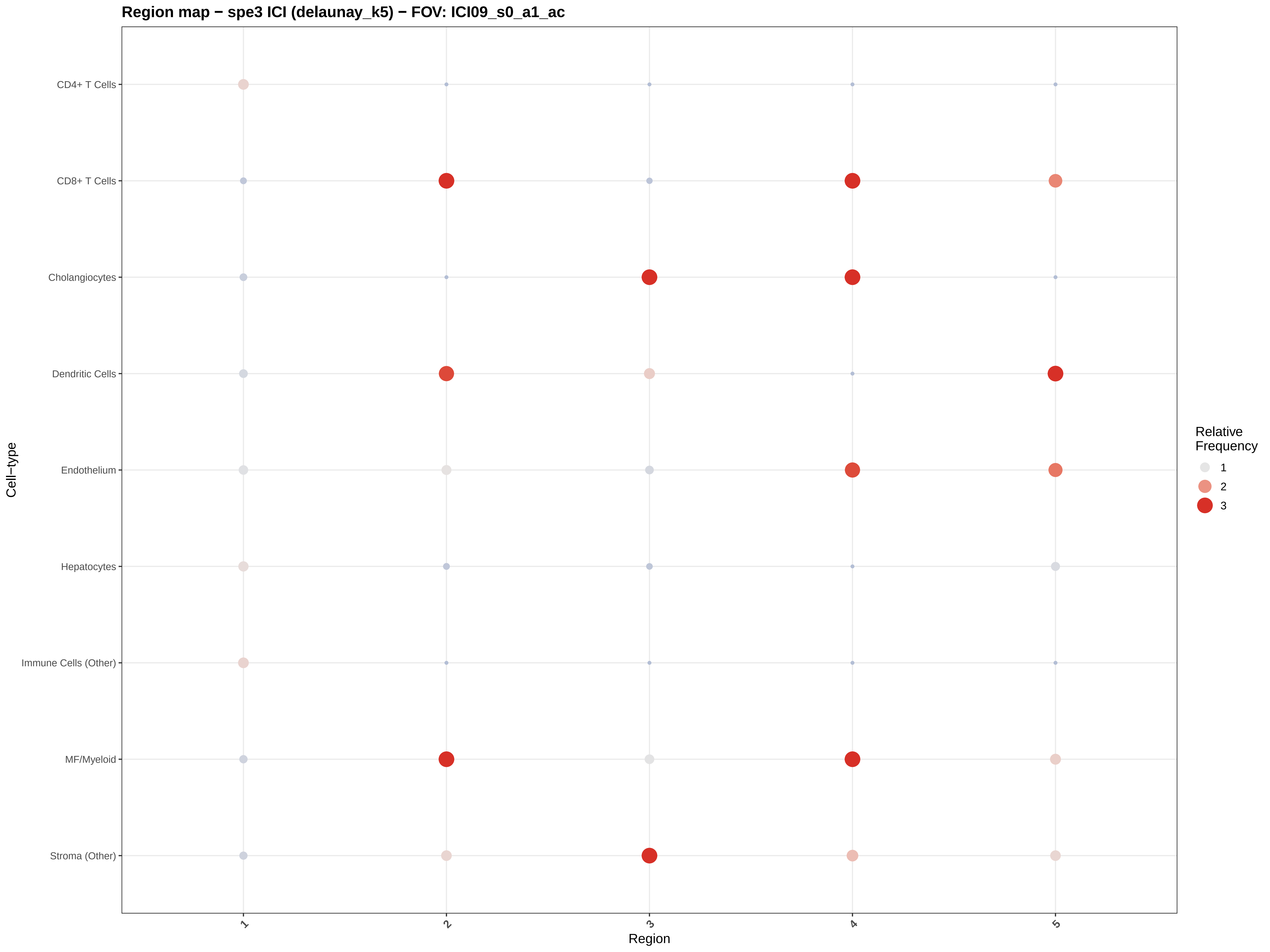

Region map − spe3 ICI (delaunay_k5) − FOV: ICI09_s0_a1_ac
CD4+ T Cells
CD8+ T Cells
Cholangiocytes
Dendritic Cells
Endothelium
Hepatocytes
Immune Cells (Other)
MF/Myeloid
Stroma (Other)
Cell−type
1
2
3
4
5
Region
Relative
Frequency
1
2
3

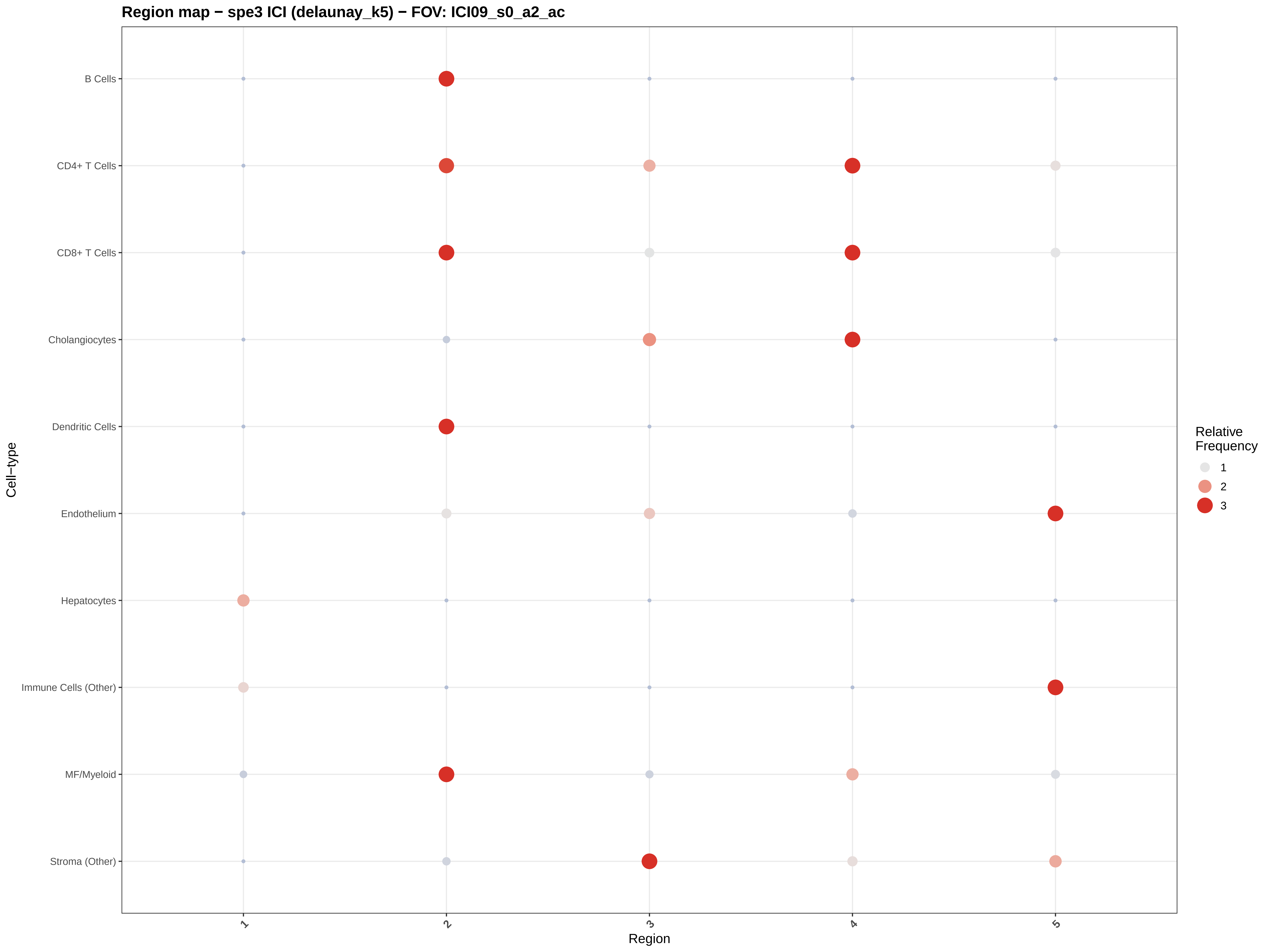

Region map − spe3 ICI (delaunay_k5) − FOV: ICI09_s0_a2_ac
B Cells
CD4+ T Cells
CD8+ T Cells
Cholangiocytes
Dendritic Cells
Endothelium
Hepatocytes
Immune Cells (Other)
MF/Myeloid
Stroma (Other)
Cell−type
1
2
3
4
5
Region
Relative Frequency
1
2
3

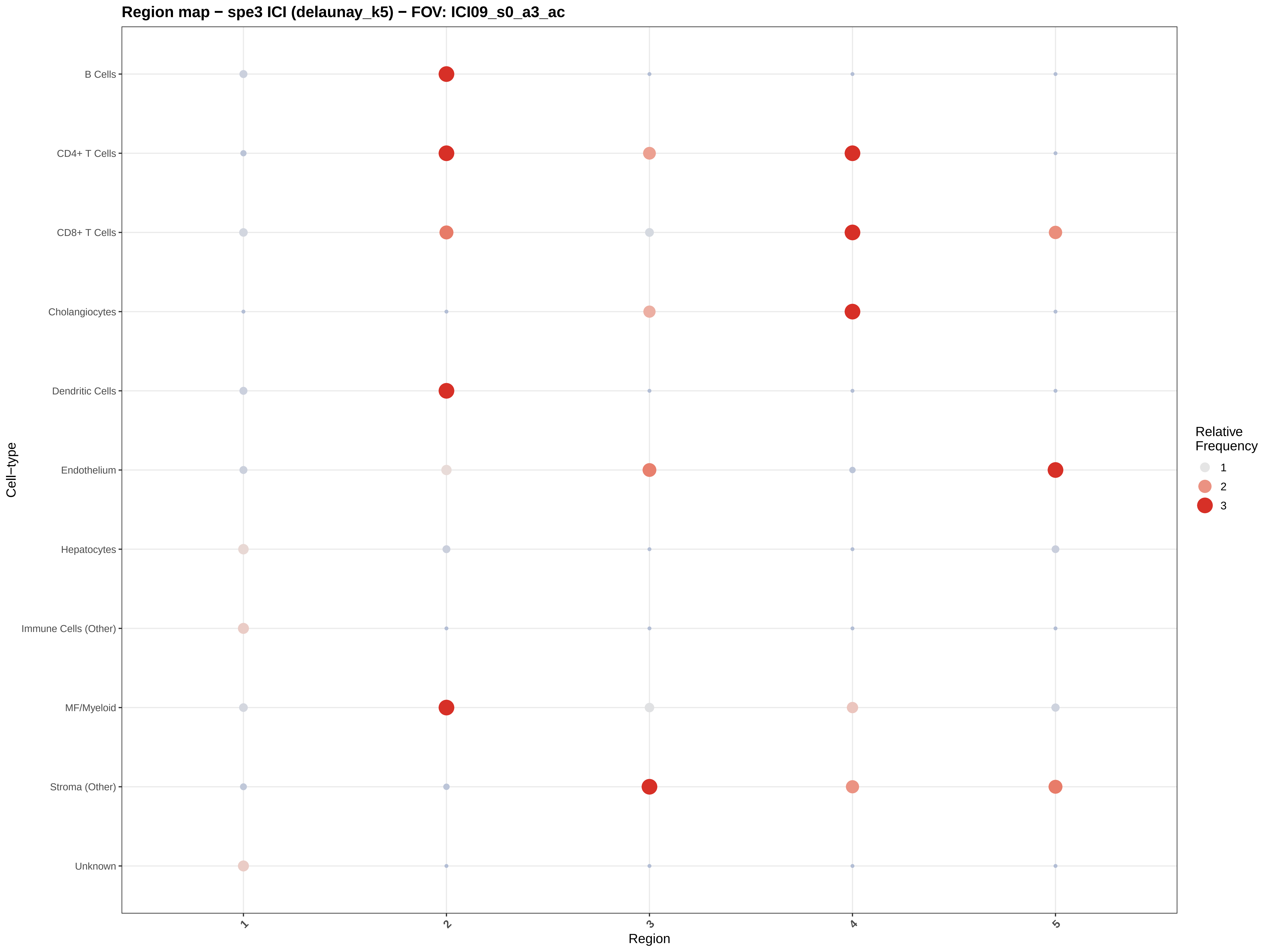
Region map − spe3 ICI (delaunay_k5) − FOV: ICI09_s0_a3_ac
B Cells
CD4+ T Cells
CD8+ T Cells
Cholangiocytes
Dendritic Cells
Endothelium
Hepatocytes
Immune Cells (Other)
MF/Myeloid
Stroma (Other)
Unknown
Cell−type
1
2
3
4
5
Region
Relative Frequency
1
2
3

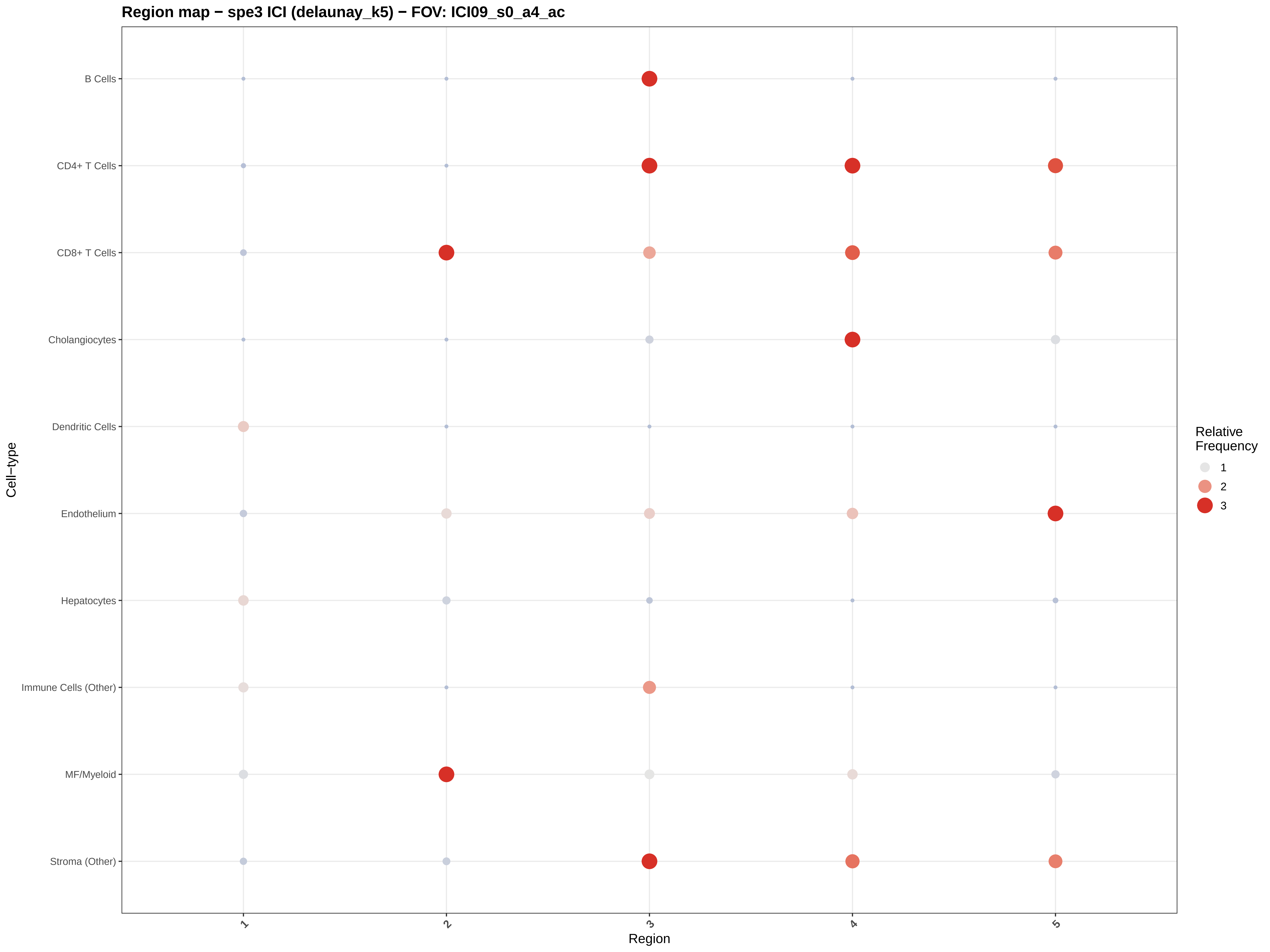
Region map − spe3 ICI (delaunay_k5) − FOV: ICI09_s0_a4_ac
B Cells
CD4+ T Cells
CD8+ T Cells
Cholangiocytes
Dendritic Cells
Endothelium
Hepatocytes
Immune Cells (Other)
MF/Myeloid
Stroma (Other)
Cell−type
1
2
3
4
5
Region
Relative Frequency
1
2
3

#### Region map − spe3 ICI (delaunay_k5) − FOV: ICI09_s0_a5_ac

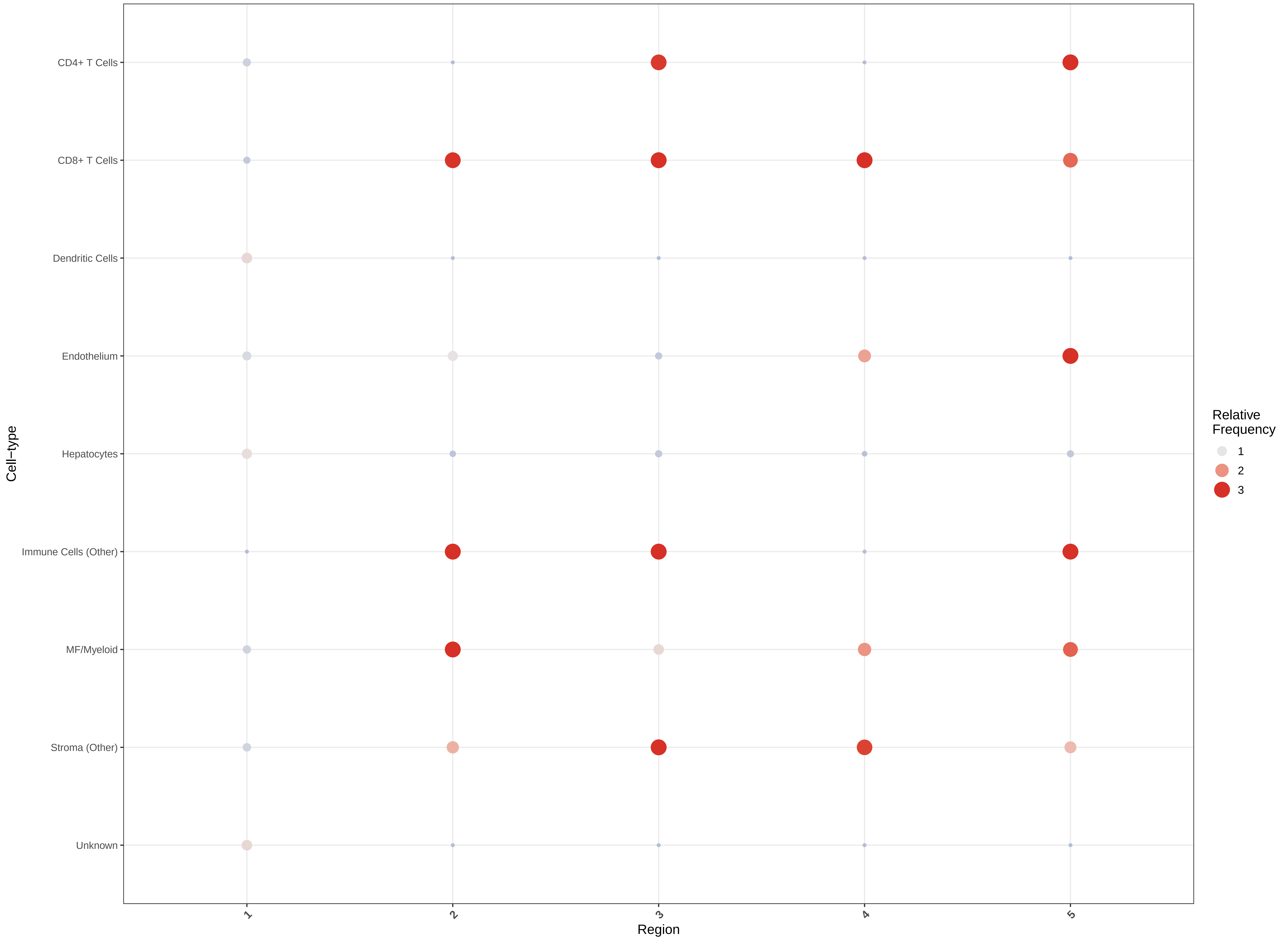

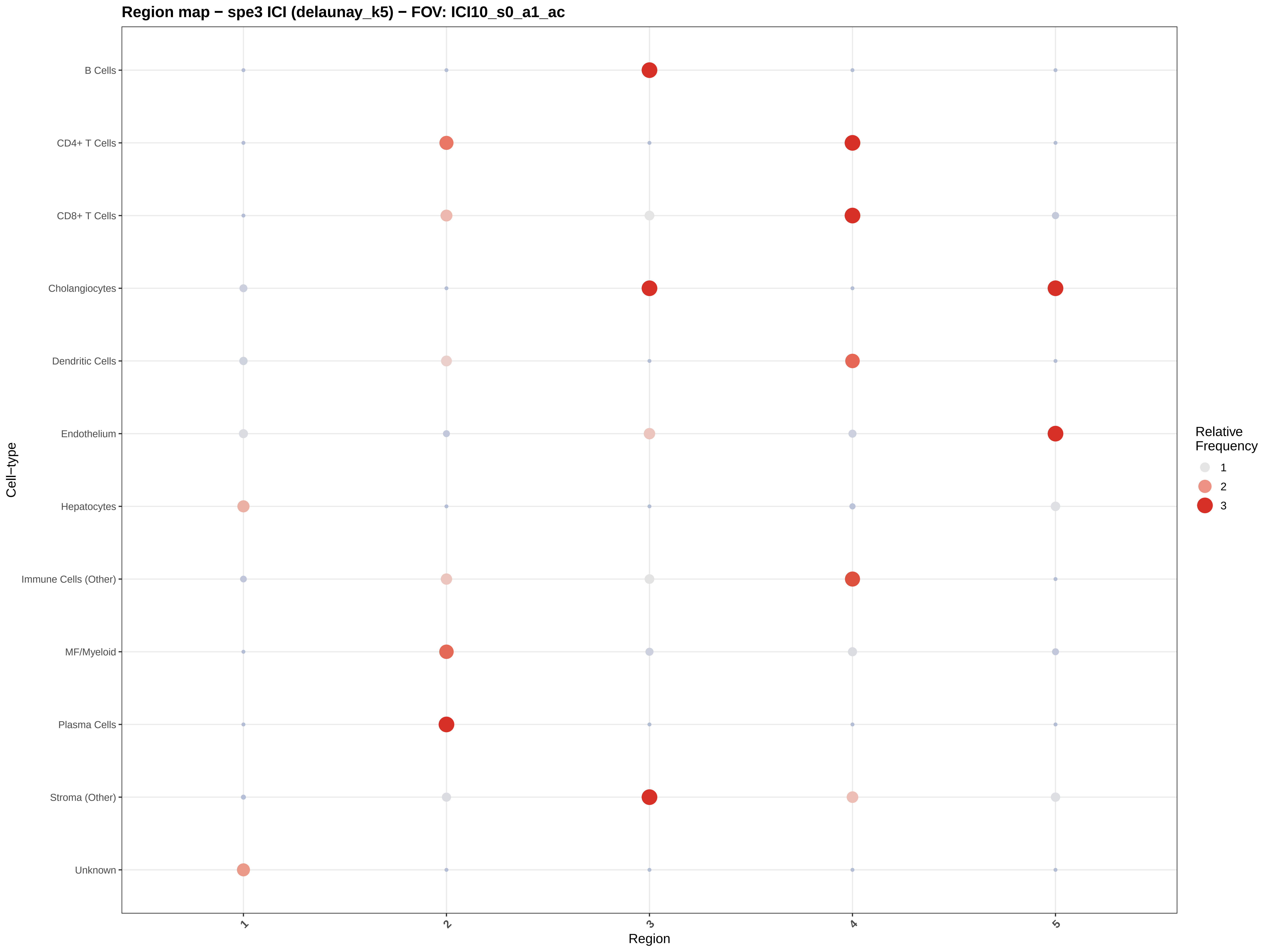

Region map − spe3 ICI (delaunay_k5) − FOV: ICI10_s0_a1_ac
B Cells
CD4+ T Cells
CD8+ T Cells
Cholangiocytes
Dendritic Cells
Endothelium
Hepatocytes
Immune Cells (Other)
MF/Myeloid
Plasma Cells
Stroma (Other)
Unknown
Cell−type
1
2
3
4
5
Region
Relative Frequency
1
2
3

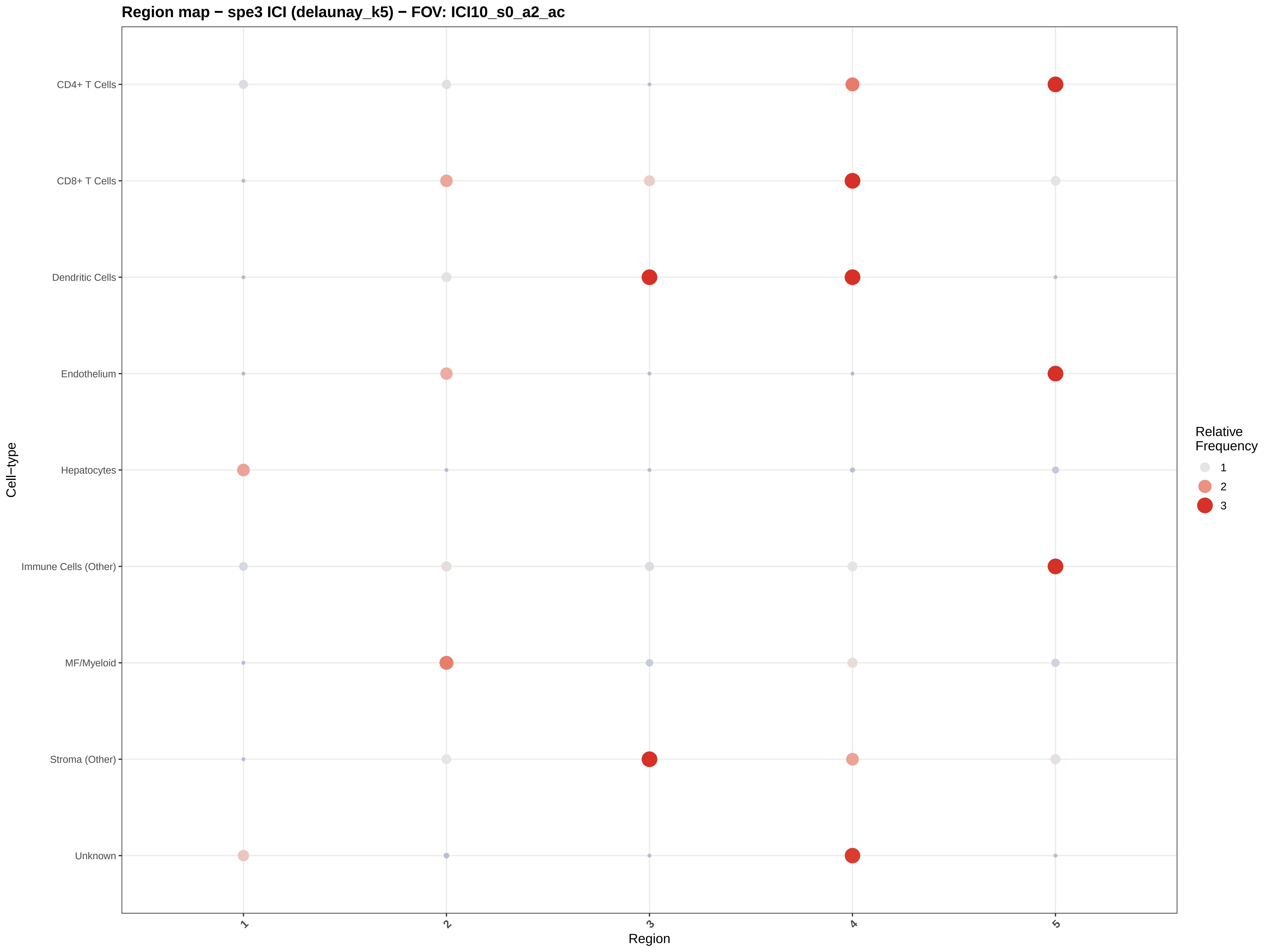

Region map − spe3 ICI (delaunay_k5) − FOV: ICI10_s0_a2_ac
CD4+ T Cells
CD8+ T Cells
Dendritic Cells
Endothelium
Hepatocytes
Immune Cells (Other)
MF/Myeloid
Stroma (Other)
Unknown
Cell−type
1
2
3
4
5
Region
Relative Frequency
1
2
3

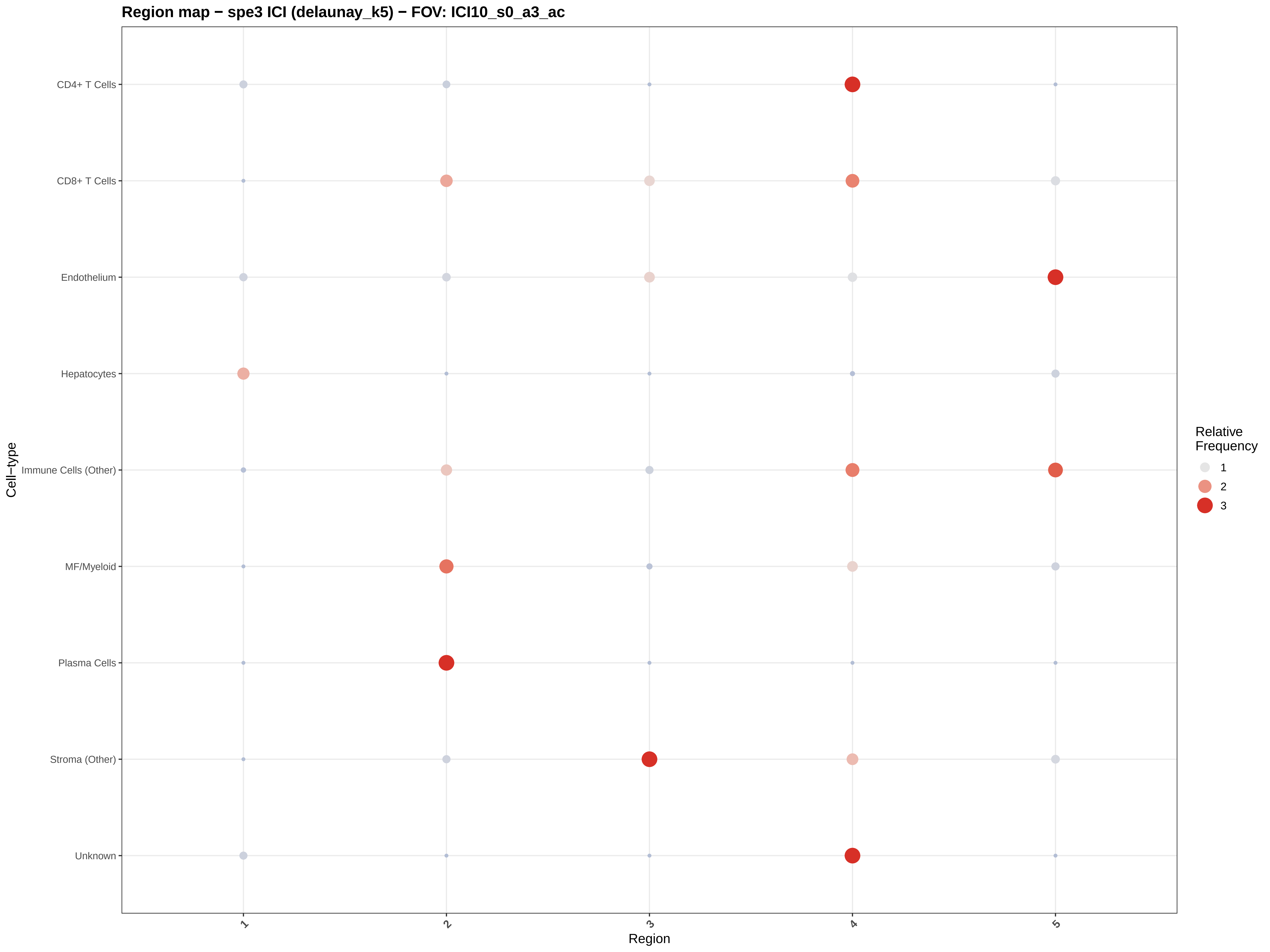

Region map − spe3 ICI (delaunay_k5) − FOV: ICI10_s0_a3_ac
CD4+ T Cells
CD8+ T Cells
Endothelium
Hepatocytes
Immune Cells (Other)
MF/Myeloid
Plasma Cells
Stroma (Other)
Unknown
Cell−type
1
2
3
4
5
Region
Relative Frequency
1
2
3

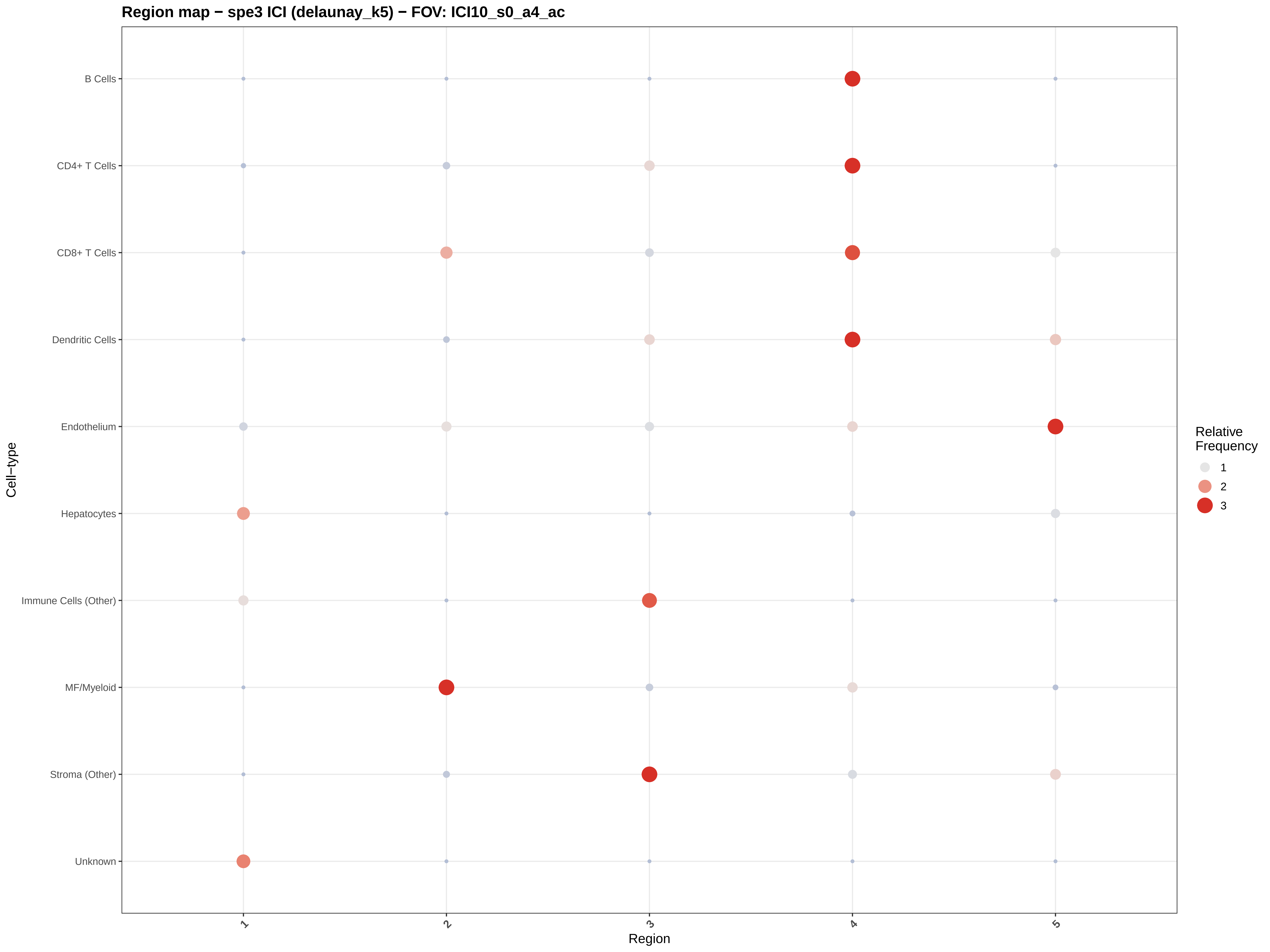

Region map − spe3 ICI (delaunay_k5) − FOV: ICI10_s0_a4_ac
B Cells
CD4+ T Cells
CD8+ T Cells
Dendritic Cells
Endothelium
Hepatocytes
Immune Cells (Other)
MF/Myeloid
Stroma (Other)
Unknown
Cell−type
1
2
3
4
5
Region
Relative Frequency
1
2
3

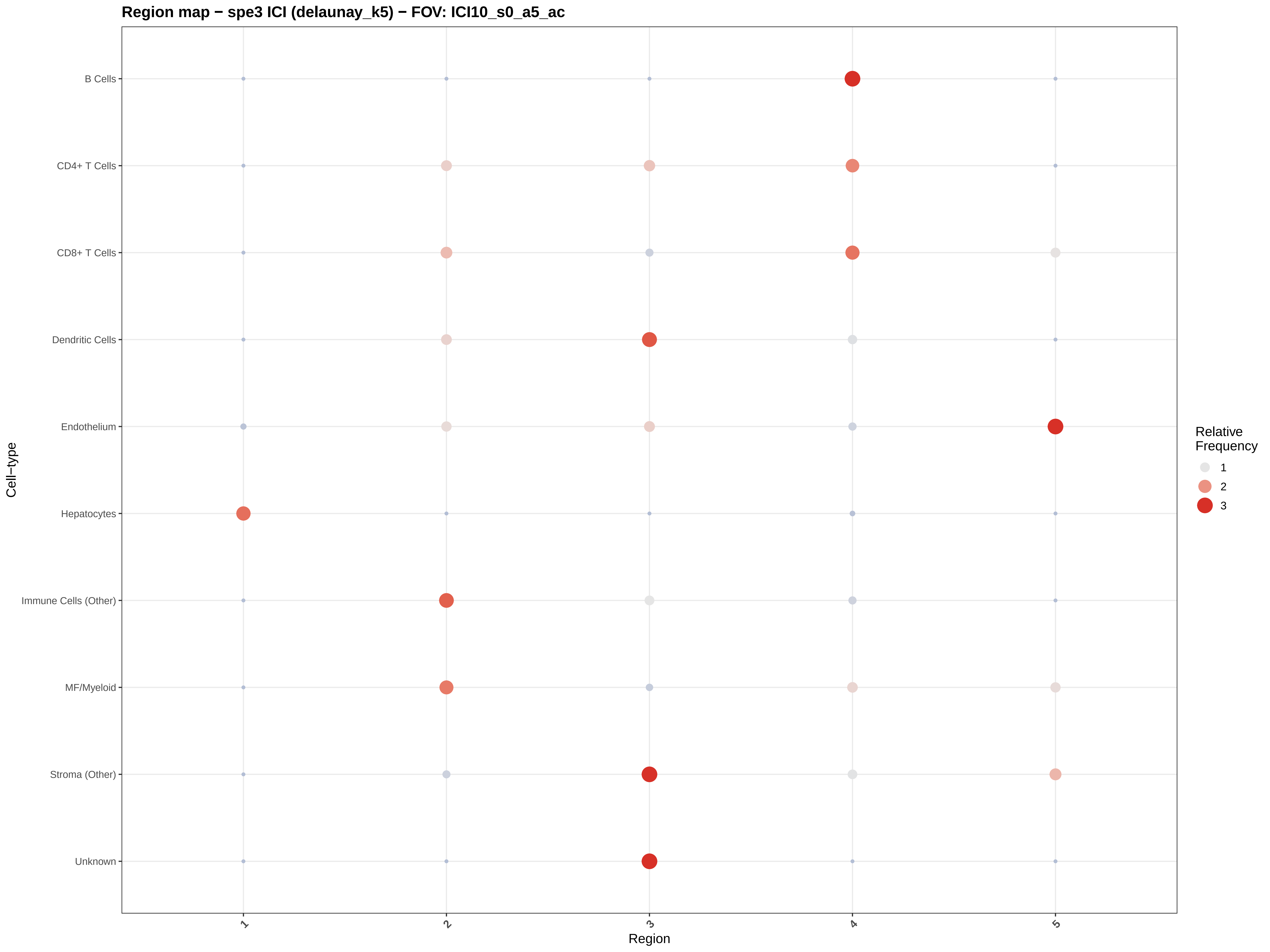

Region map − spe3 ICI (delaunay_k5) − FOV: ICI10_s0_a5_ac
B Cells
CD4+ T Cells
CD8+ T Cells
Dendritic Cells
Endothelium
Hepatocytes
Immune Cells (Other)
MF/Myeloid
Stroma (Other)
Unknown
Cell−type
1
2
3
4
5
Region
Relative Frequency
1
2
3

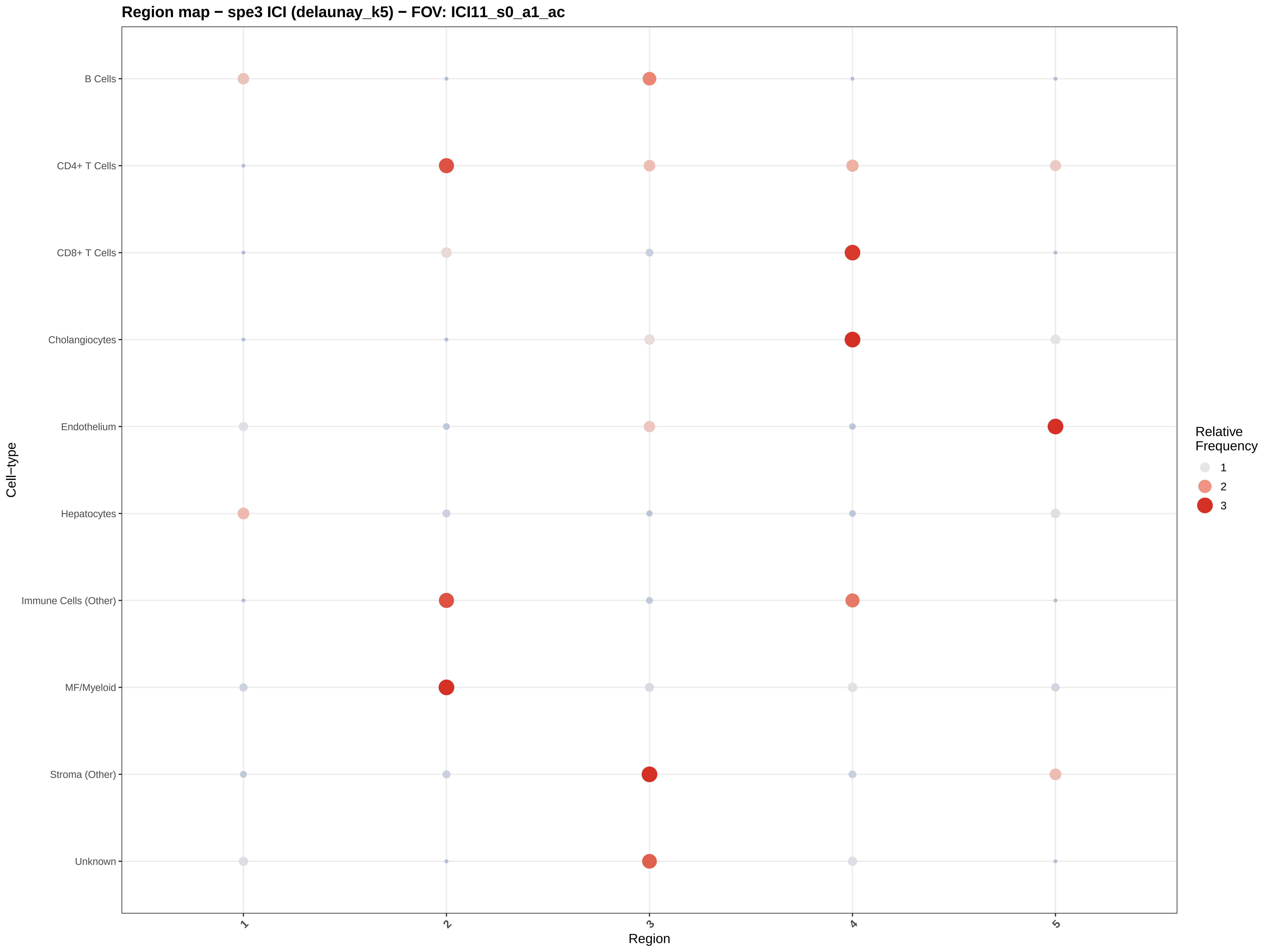
Region map − spe3 ICI (delaunay_k5) − FOV: ICI11_s0_a1_ac
B Cells
CD4+ T Cells
CD8+ T Cells
Cholangiocytes
Endothelium
Hepatocytes
Immune Cells (Other)
MF/Myeloid
Stroma (Other)
Unknown
Cell−type
1
2
3
4
5
Region
Relative Frequency
1
2
3

Region map − spe3 ICI (delaunay_k5) − FOV: ICI11_s0_a4_ac

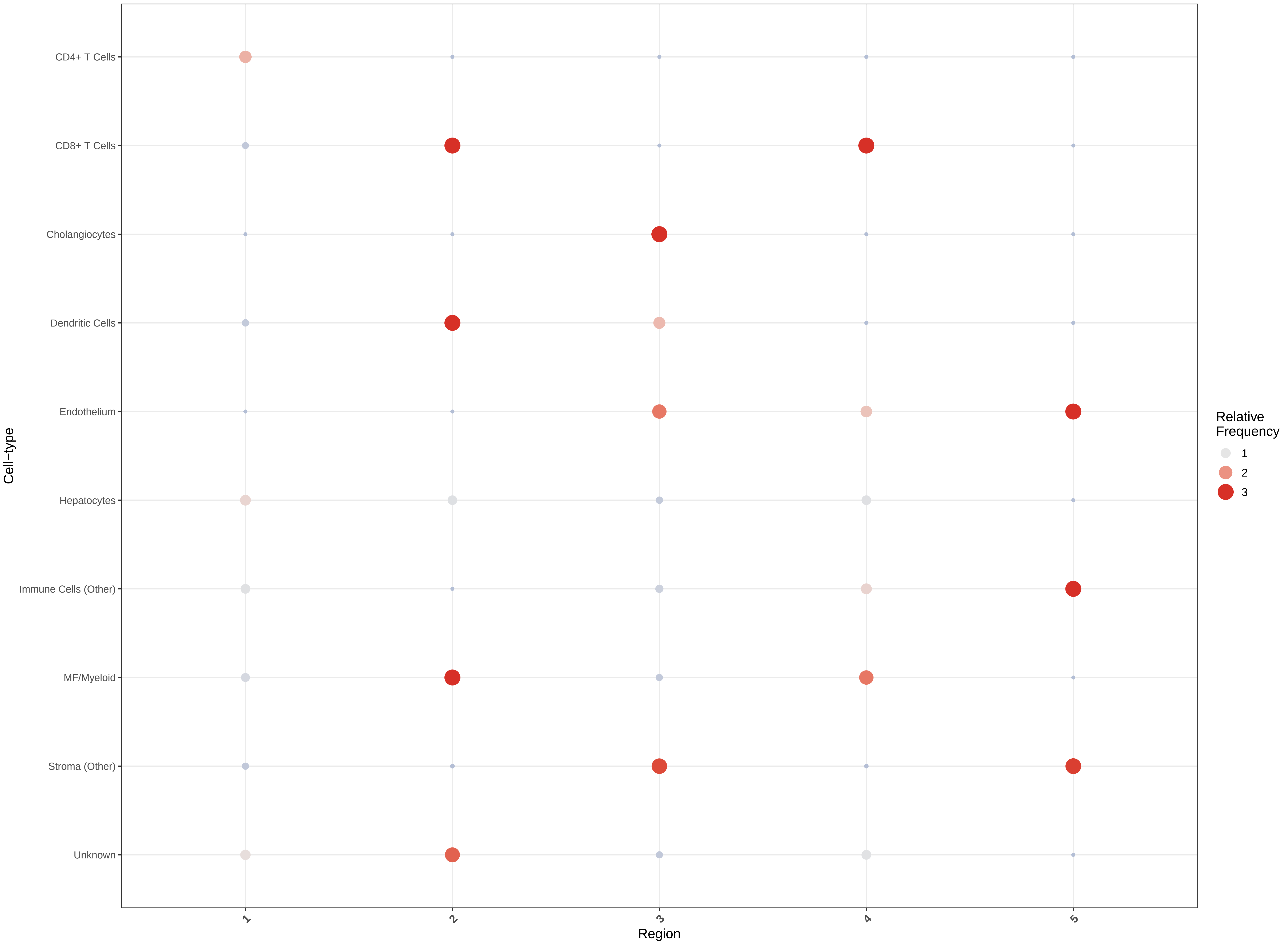

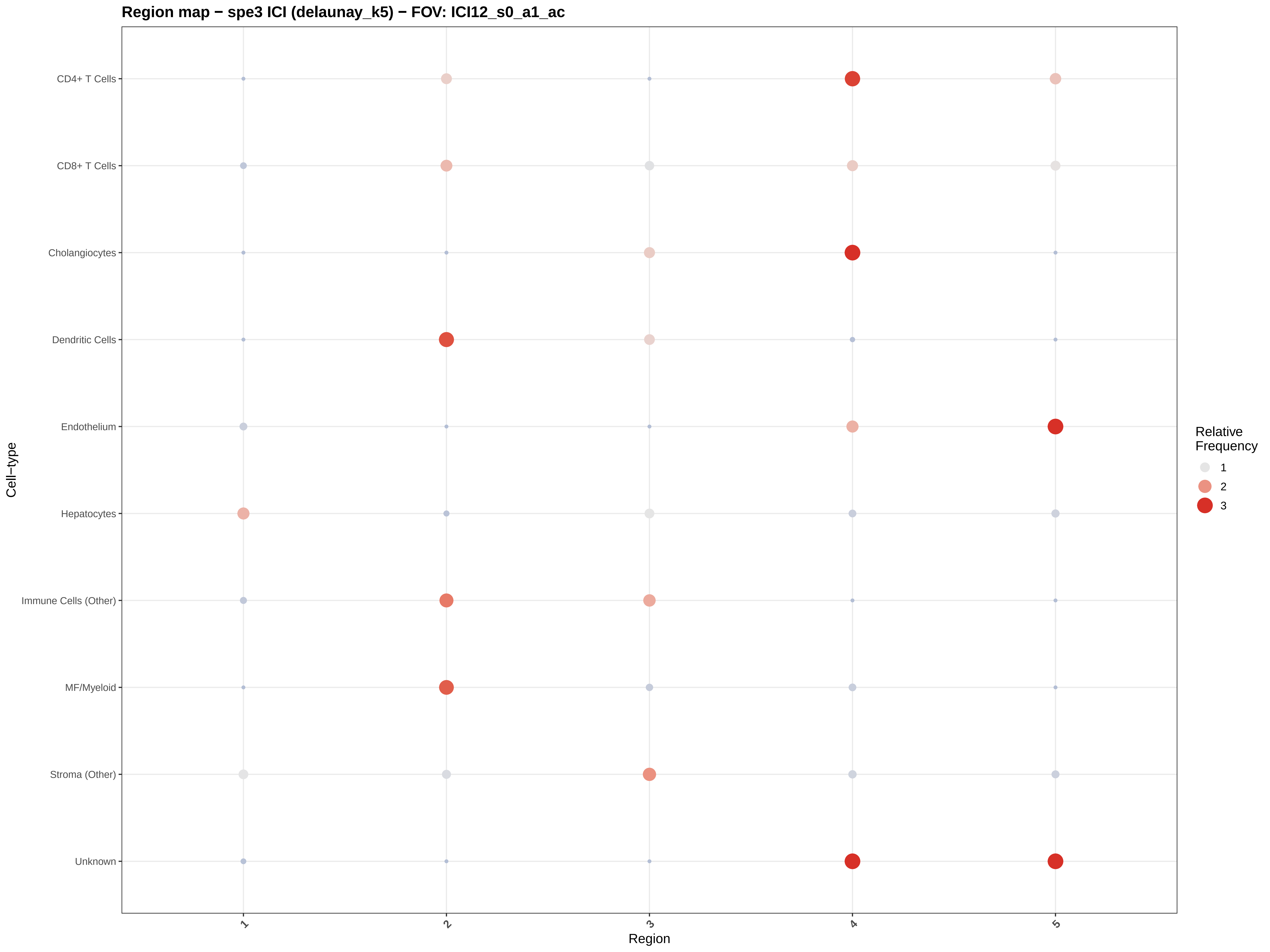
Region map − spe3 ICI (delaunay_k5) − FOV: ICI12_s0_a1_ac
CD4+ T Cells
CD8+ T Cells
Cholangiocytes
Dendritic Cells
Endothelium
Hepatocytes
Immune Cells (Other)
MF/Myeloid
Stroma (Other)
Unknown
Cell−type
1
2
3
4
5
Region
Relative Frequency
1
2
3

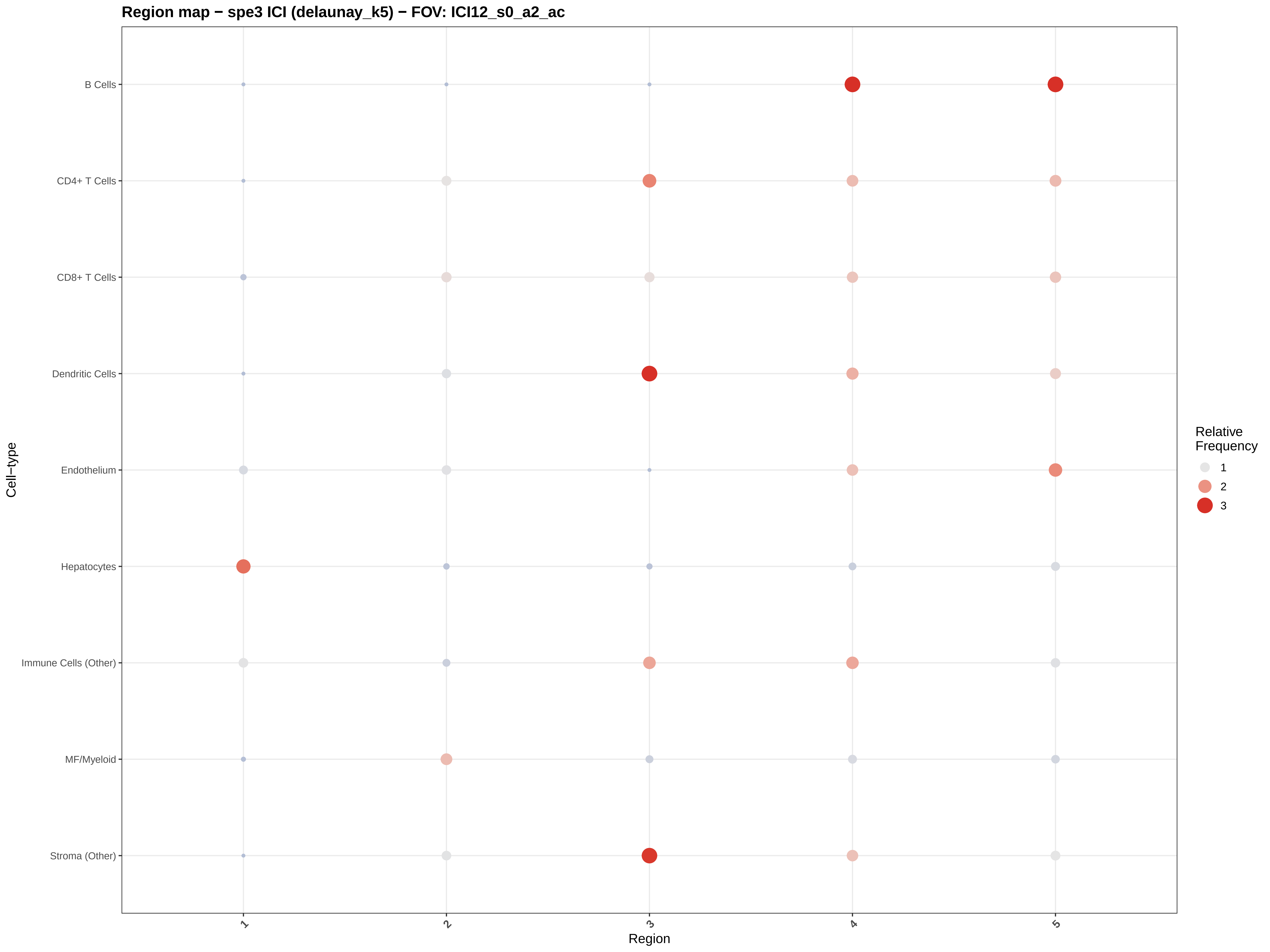

Region map − spe3 ICI (delaunay_k5) − FOV: ICI12_s0_a2_ac
B Cells
CD4+ T Cells
CD8+ T Cells
Dendritic Cells
Endothelium
Hepatocytes
Immune Cells (Other)
MF/Myeloid
Stroma (Other)
Cell−type
1
2
3
4
5
Region
Relative Frequency
1
2
3

#### Region map − spe3 ICI (delaunay_k5) − FOV: ICI12_s0_a3_ac

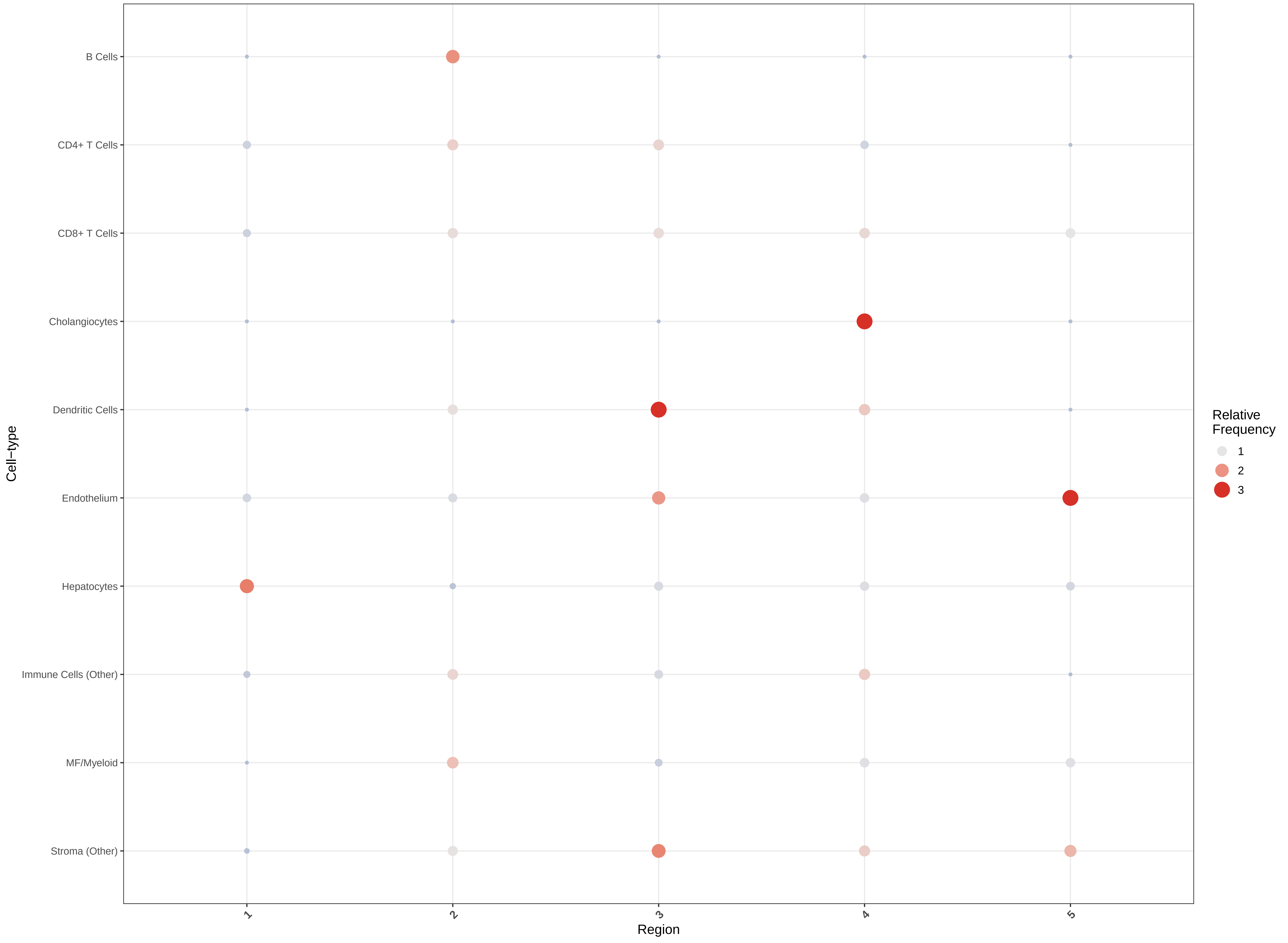

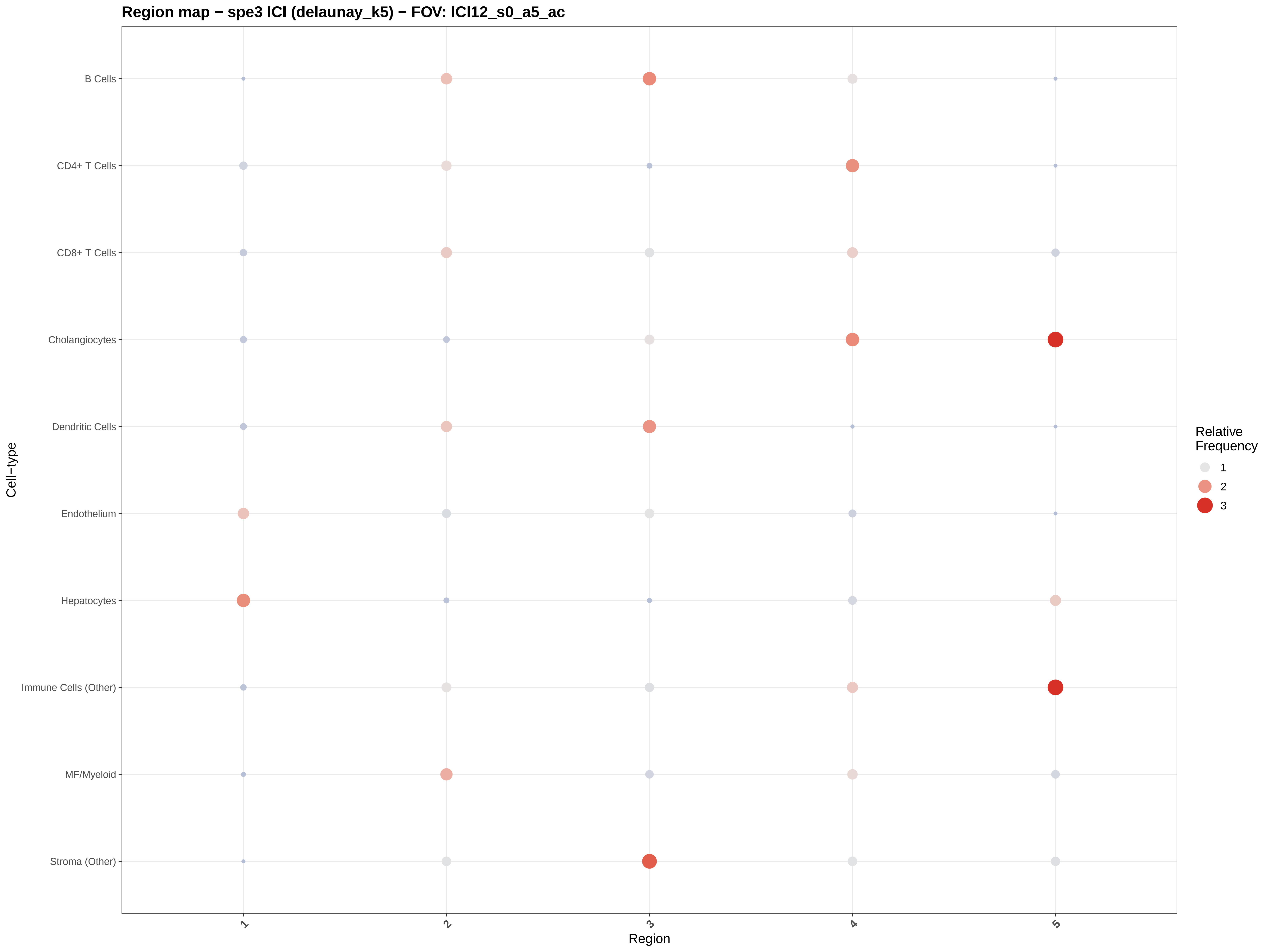

Region map − spe3 ICI (delaunay_k5) − FOV: ICI12_s0_a5_ac
B Cells
CD4+ T Cells
CD8+ T Cells
Cholangiocytes
Dendritic Cells
Endothelium
Hepatocytes
Immune Cells (Other)
MF/Myeloid
Stroma (Other)
Cell−type
1
2
3
4
5
Region
Relative Frequency
1
2
3

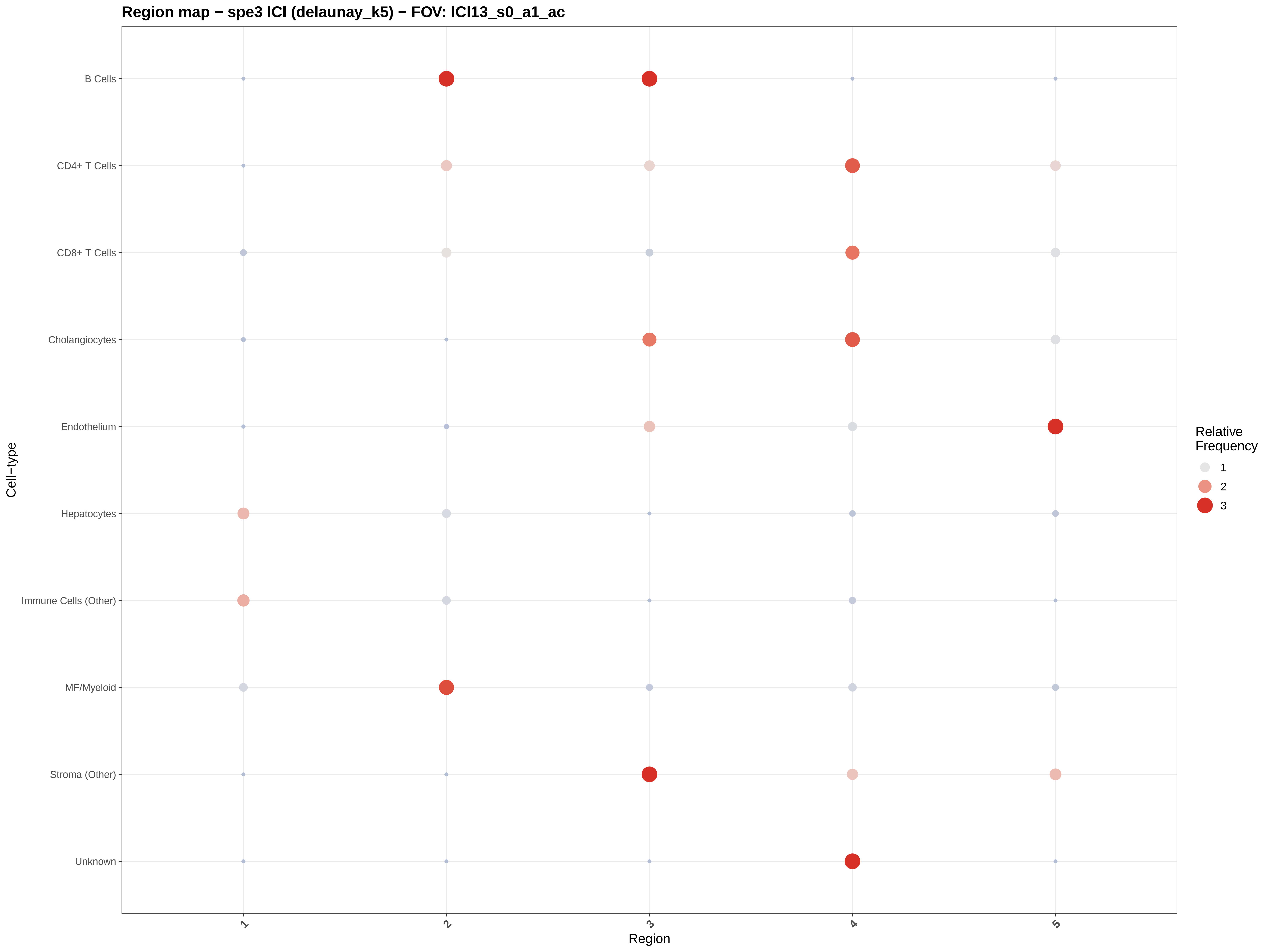

Region map − spe3 ICI (delaunay_k5) − FOV: ICI13_s0_a1_ac
B Cells
CD4+ T Cells
CD8+ T Cells
Cholangiocytes
Endothelium
Hepatocytes
Immune Cells (Other)
MF/Myeloid
Stroma (Other)
Unknown
Cell−type
1
2
3
4
5
Region
Relative Frequency
1
2
3

#### Region map − spe3 ICI (delaunay_k5) − FOV: ICI13_s0_a2_ac

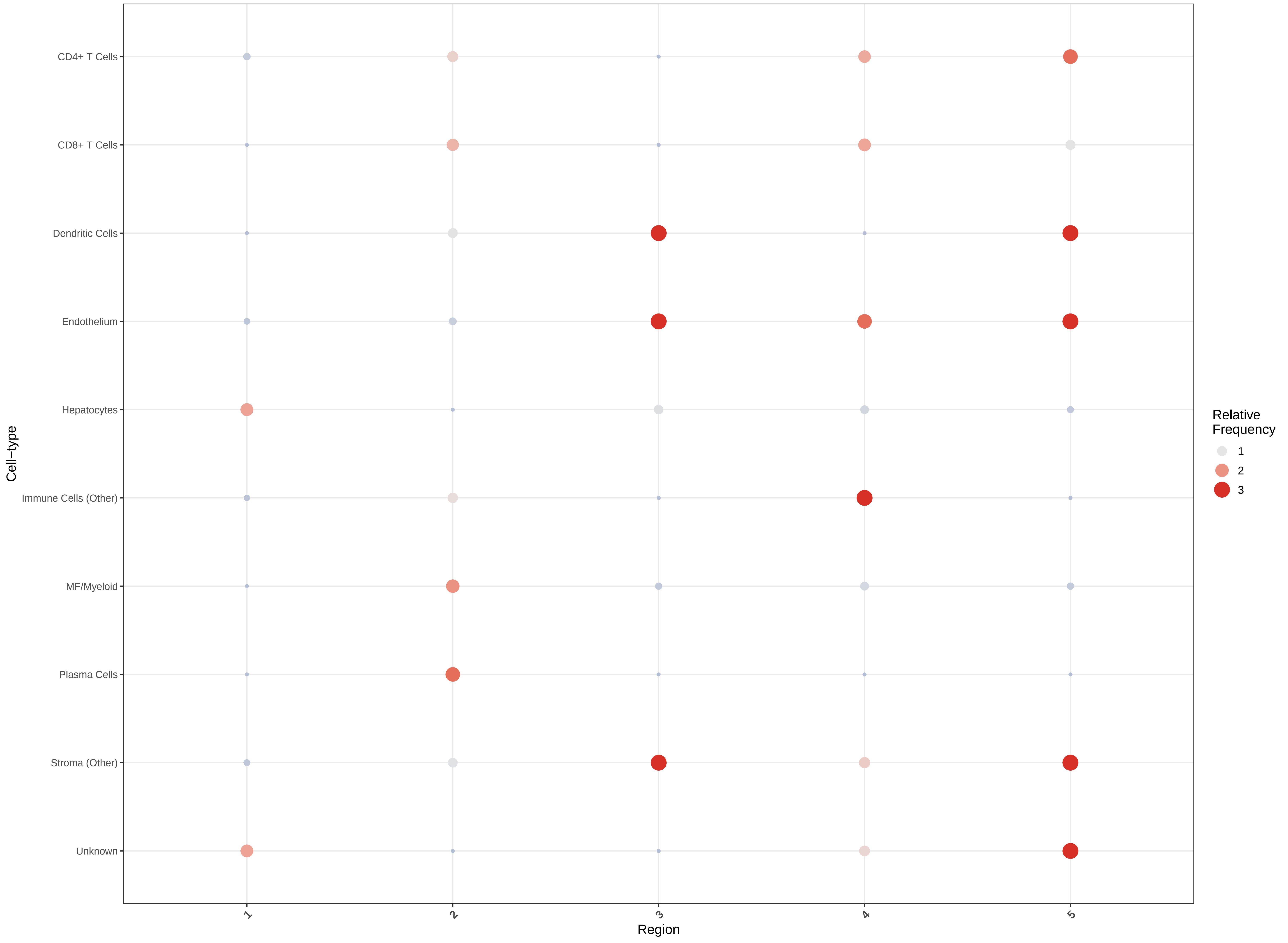

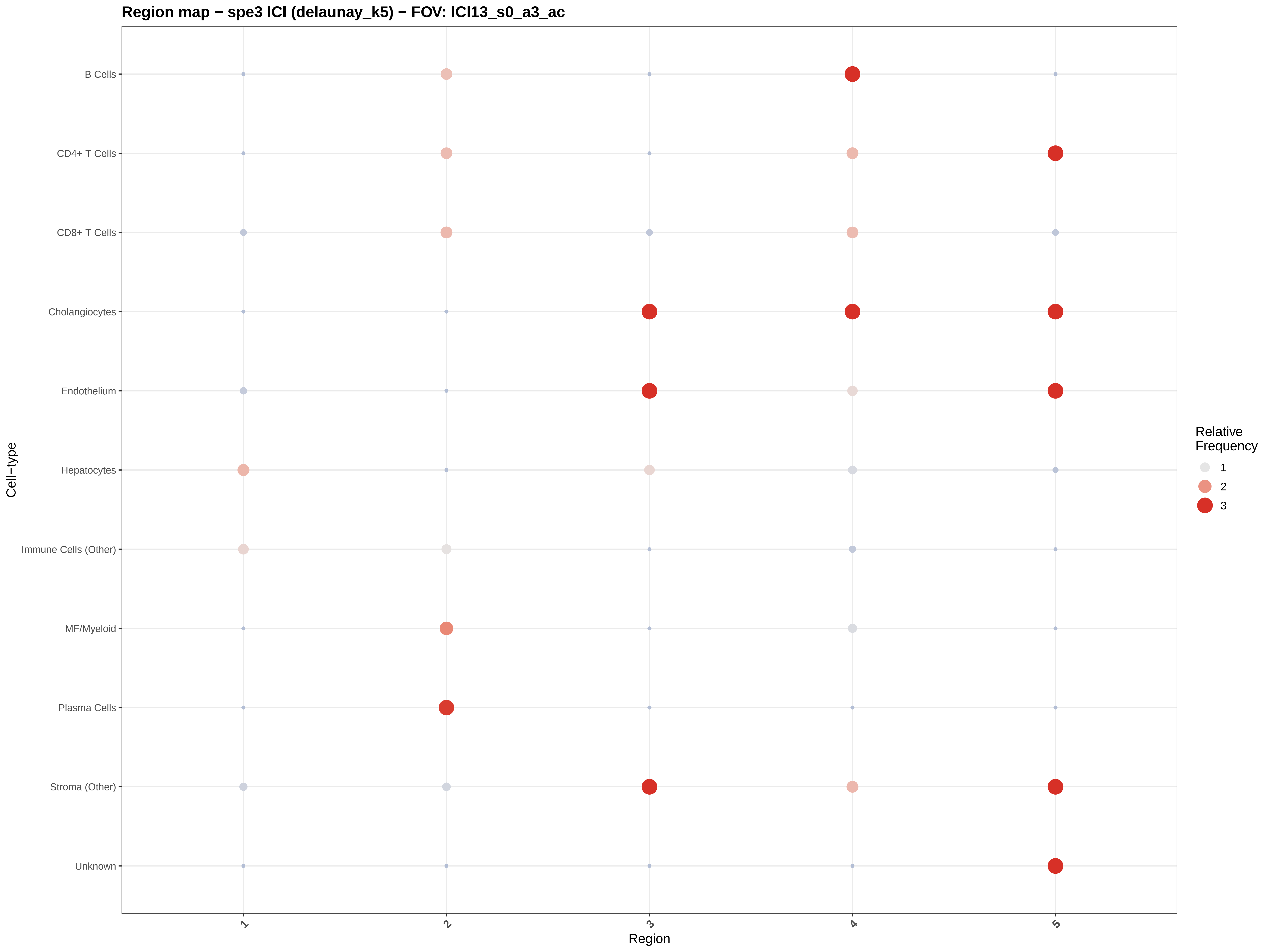
Region map − spe3 ICI (delaunay_k5) − FOV: ICI13_s0_a3_ac
B Cells
CD4+ T Cells
CD8+ T Cells
Cholangiocytes
Endothelium
Hepatocytes
Immune Cells (Other)
MF/Myeloid
Plasma Cells
Stroma (Other)
Unknown
Cell−type
1
2
3
4
5
Region
Relative Frequency
1
2
3

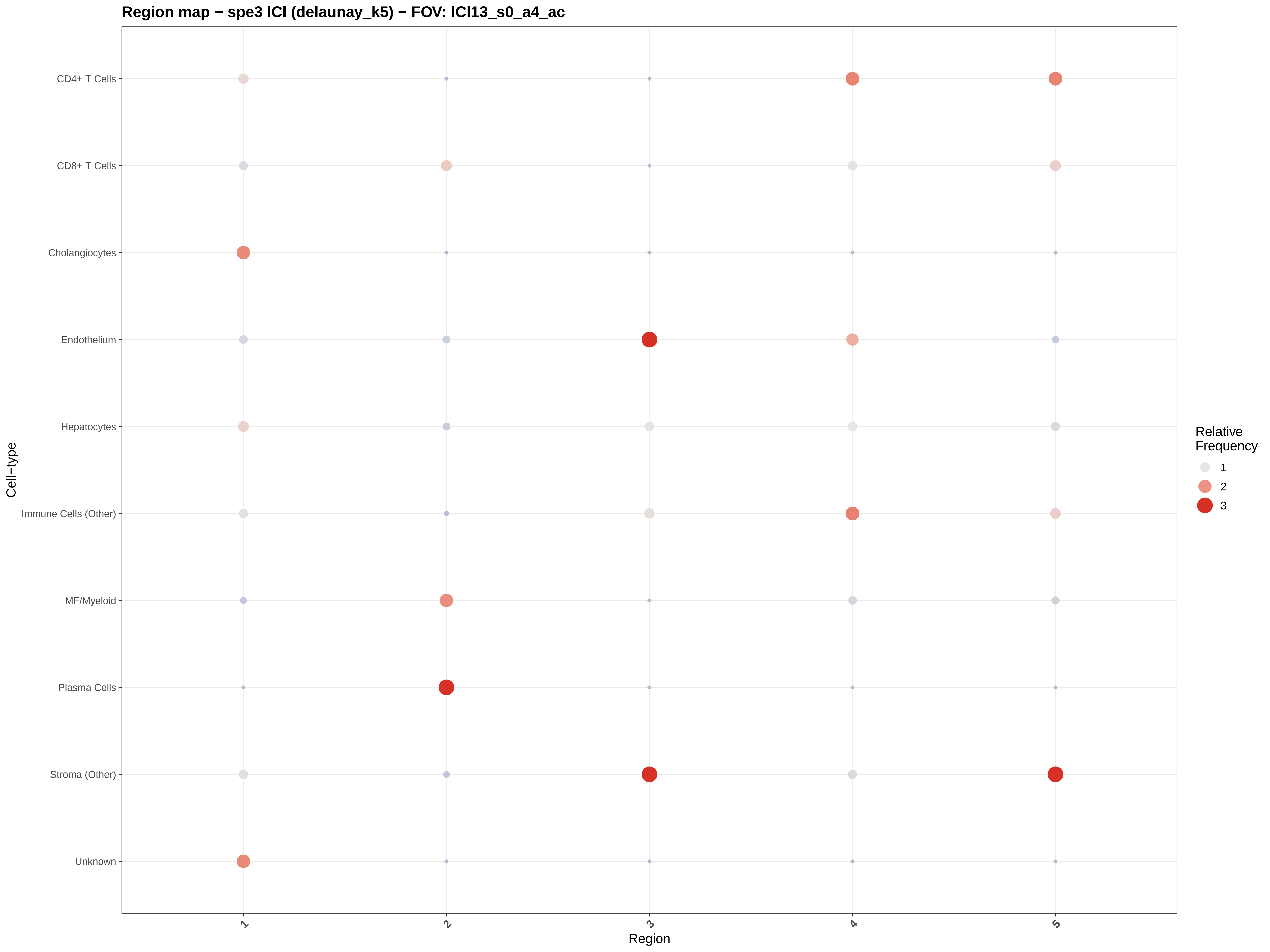

Region map − spe3 ICI (delaunay_k5) − FOV: ICI13_s0_a4_ac
CD4+ T Cells
CD8+ T Cells
Cholangiocytes
Endothelium
Hepatocytes
Immune Cells (Other)
MF/Myeloid
Plasma Cells
Stroma (Other)
Unknown
Cell−type
1
2
3
4
5
Region
Relative Frequency
1
2
3

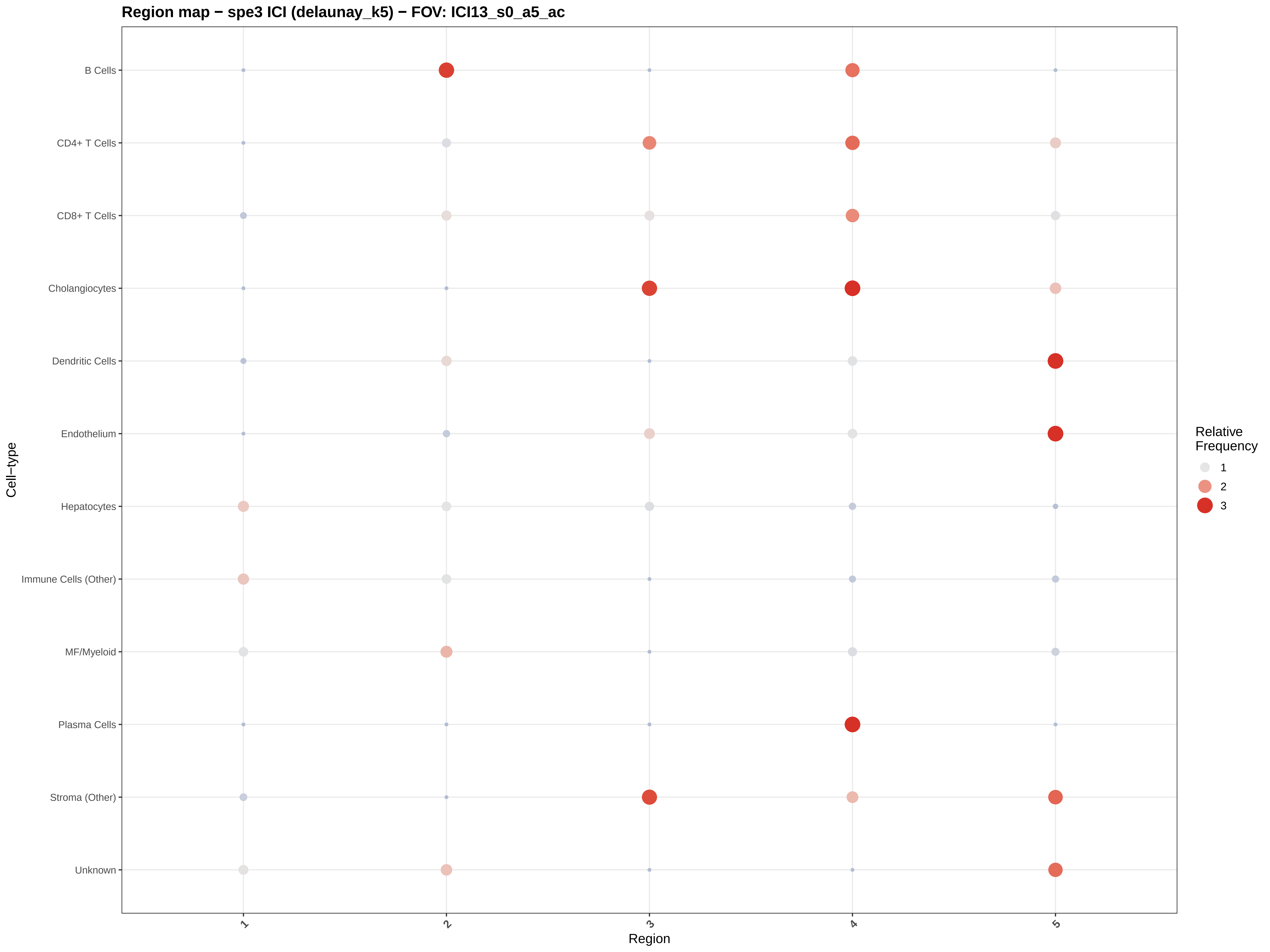

Region map − spe3 ICI (delaunay_k5) − FOV: ICI13_s0_a5_ac
B Cells
CD4+ T Cells
CD8+ T Cells
Cholangiocytes
Dendritic Cells
Endothelium
Hepatocytes
Immune Cells (Other)
MF/Myeloid
Plasma Cells
Stroma (Other)
Unknown
Cell−type
1
2
3
4
5
Region
Relative Frequency
1
2
3

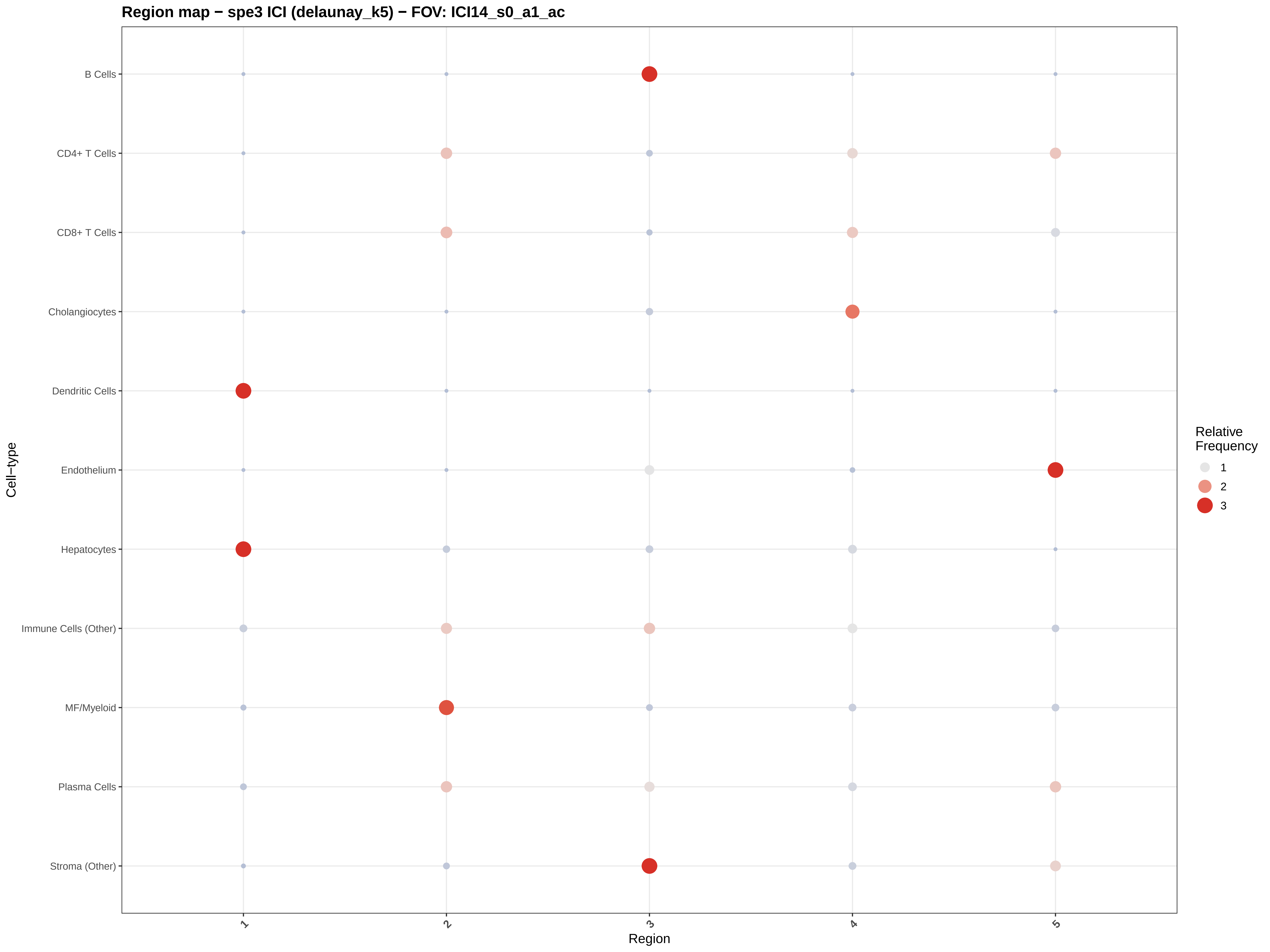
Region map − spe3 ICI (delaunay_k5) − FOV: ICI14_s0_a1_ac
B Cells
CD4+ T Cells
CD8+ T Cells
Cholangiocytes
Dendritic Cells
Endothelium
Hepatocytes
Immune Cells (Other)
MF/Myeloid
Plasma Cells
Stroma (Other)
Cell−type
1
2
3
4
5
Region
Relative Frequency
1
2
3

#### Region map − spe3 ICI (delaunay_k5) − FOV: ICI14_s0_a3_ac

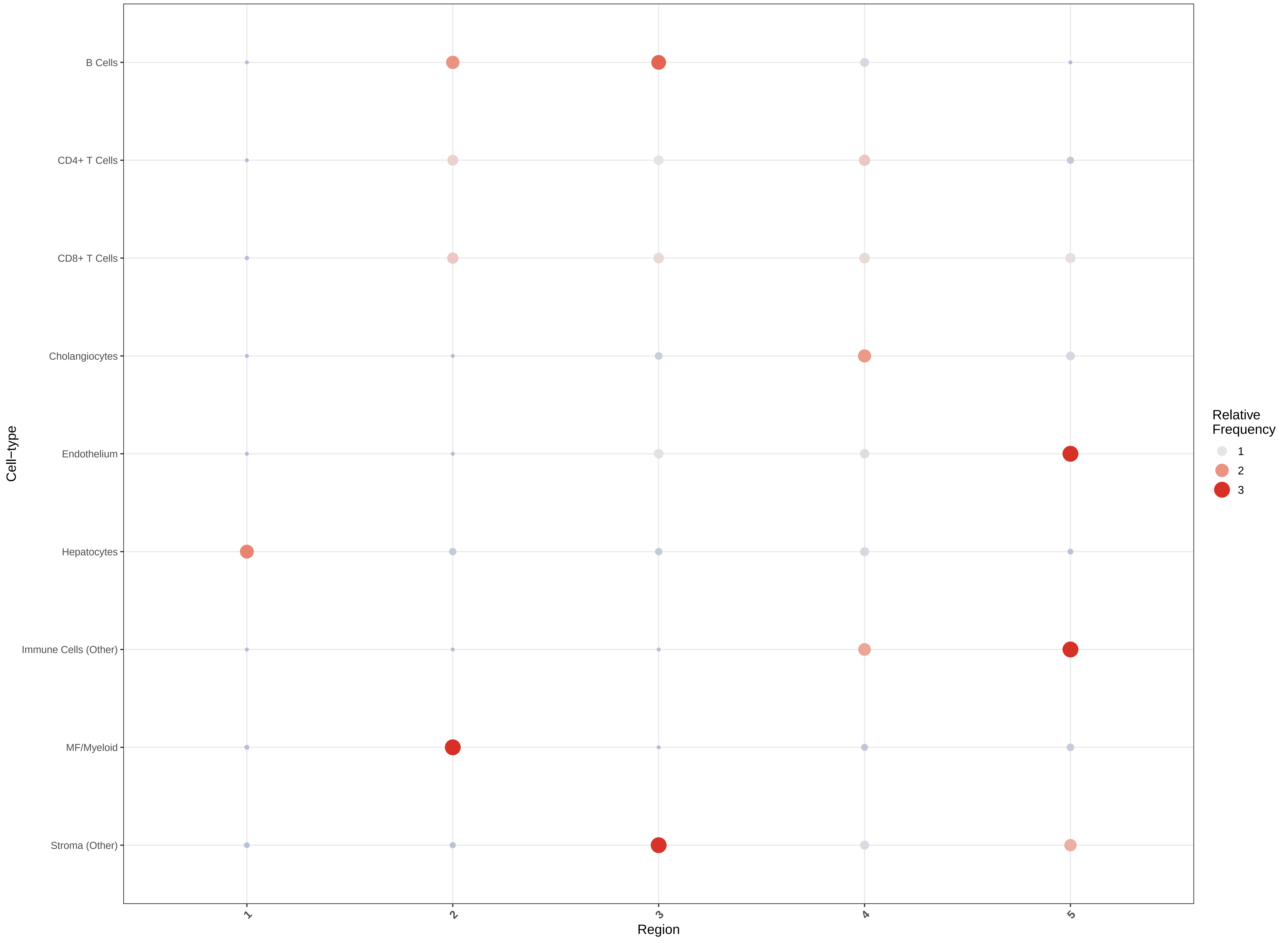

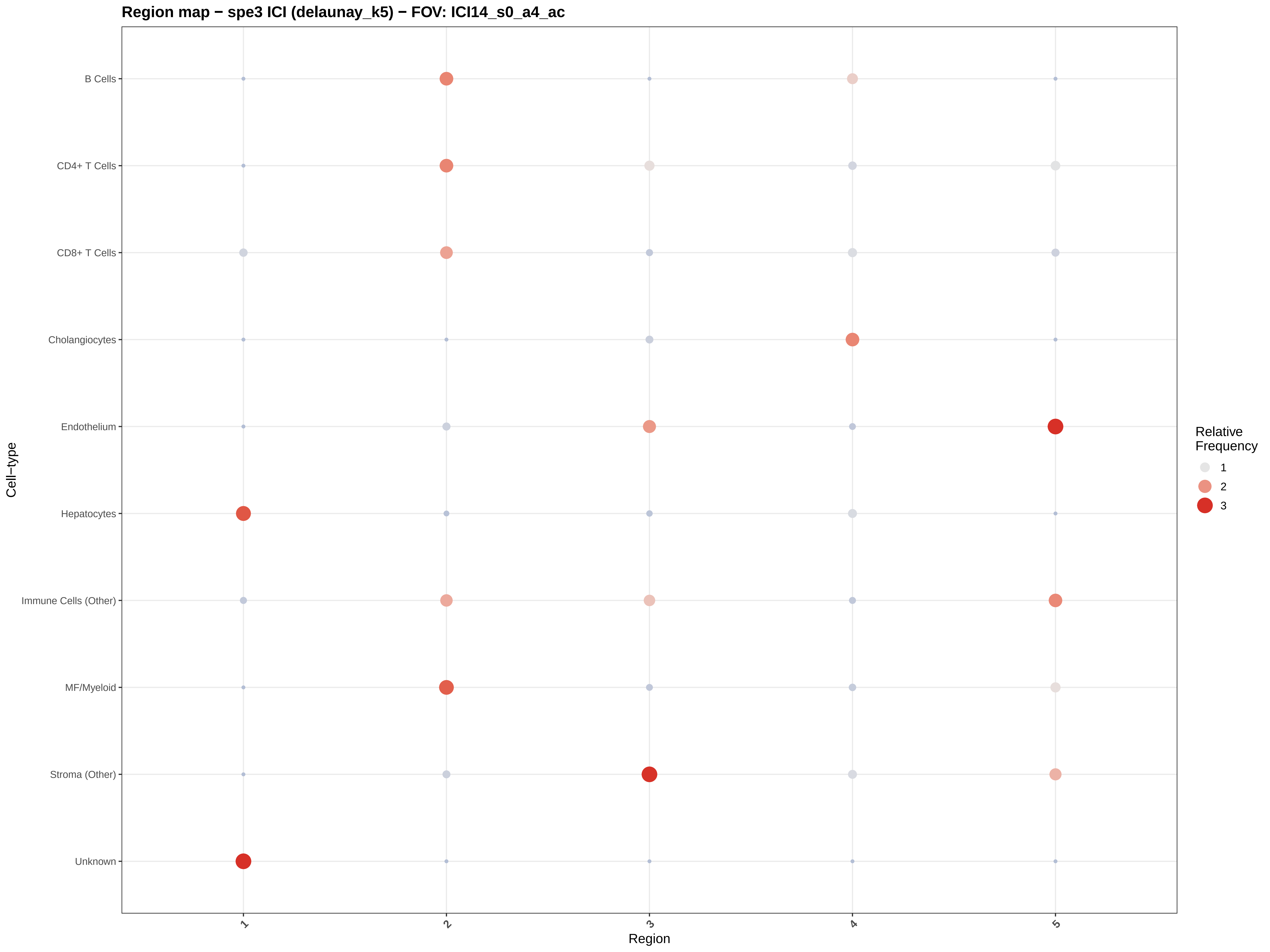

Region map − spe3 ICI (delaunay_k5) − FOV: ICI14_s0_a4_ac
B Cells
CD4+ T Cells
CD8+ T Cells
Cholangiocytes
Endothelium
Hepatocytes
Immune Cells (Other)
MF/Myeloid
Stroma (Other)
Unknown
Cell−type
1
2
3
4
5
Region
Relative Frequency
1
2
3

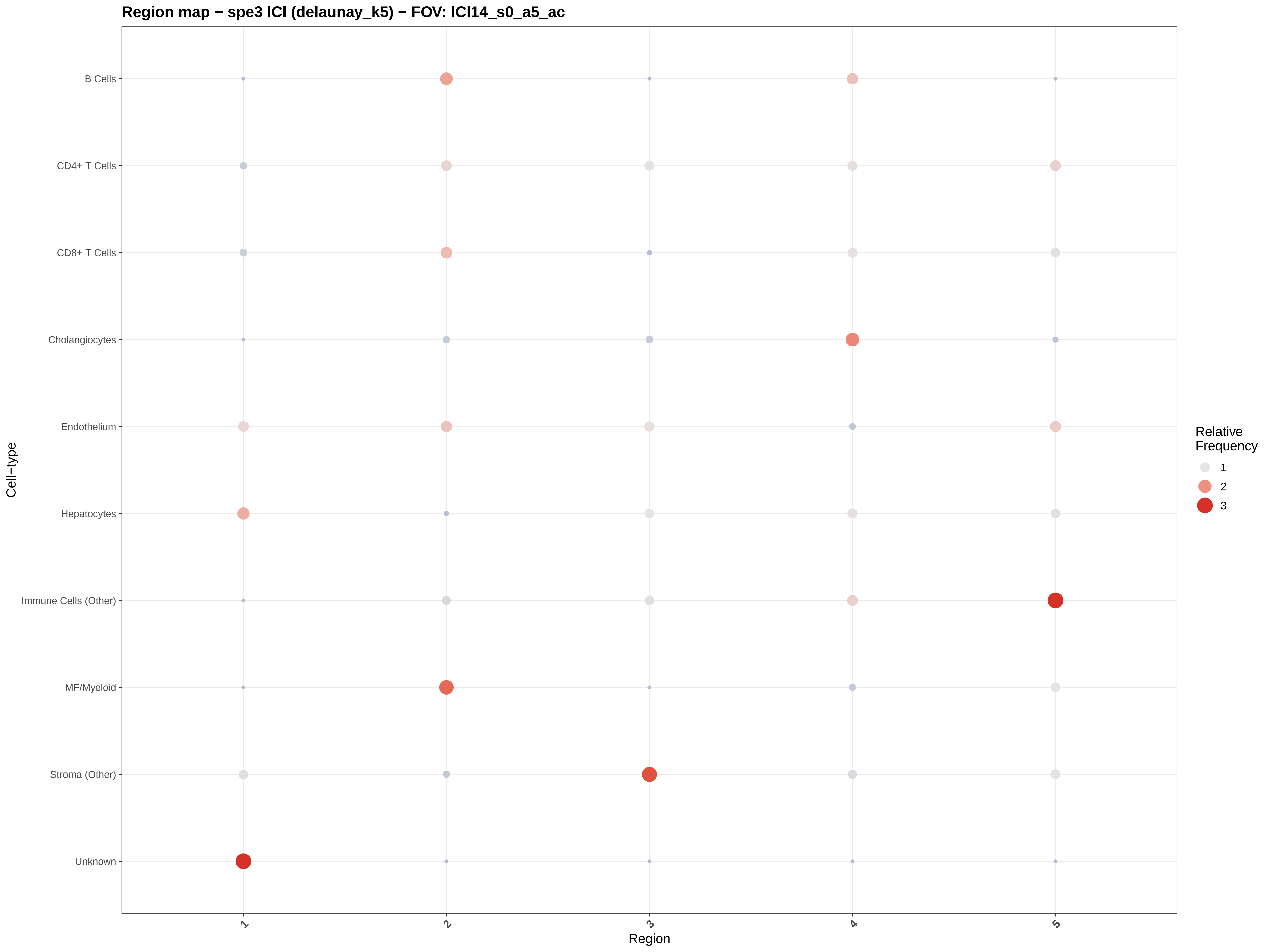
Region map − spe3 ICI (delaunay_k5) − FOV: ICI14_s0_a5_ac
B Cells
CD4+ T Cells
CD8+ T Cells
Cholangiocytes
Endothelium
Hepatocytes
Immune Cells (Other)
MF/Myeloid
Stroma (Other)
Unknown
Cell−type
1
2
3
4
5
Region
Relative
Frequency
1
2
3

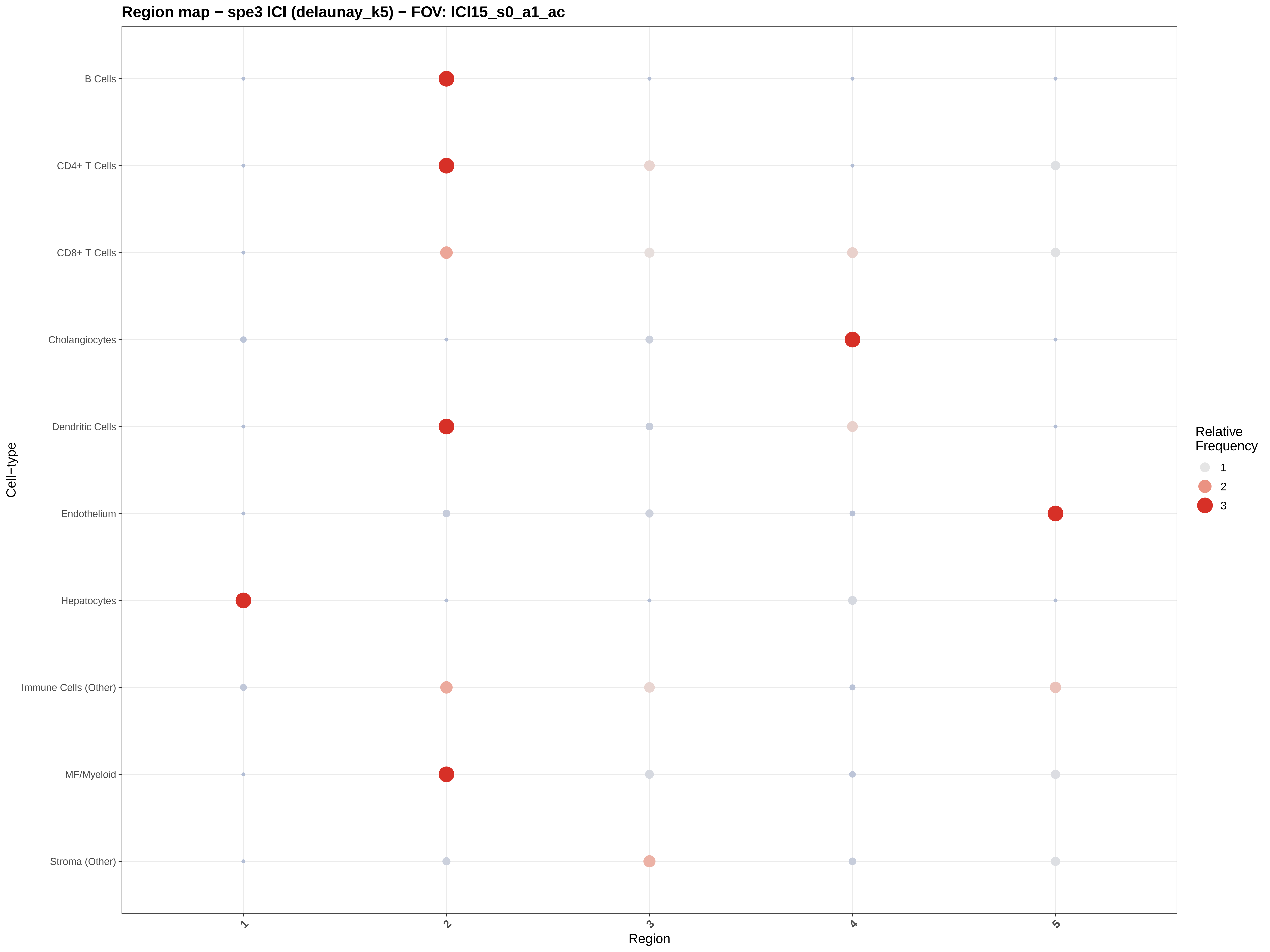
Region map − spe3 ICI (delaunay_k5) − FOV: ICI15_s0_a1_ac
B Cells
CD4+ T Cells
CD8+ T Cells
Cholangiocytes
Dendritic Cells
Endothelium
Hepatocytes
Immune Cells (Other)
MF/Myeloid
Stroma (Other)
Cell−type
1
2
3
4
5
Region
Relative Frequency
1
2
3

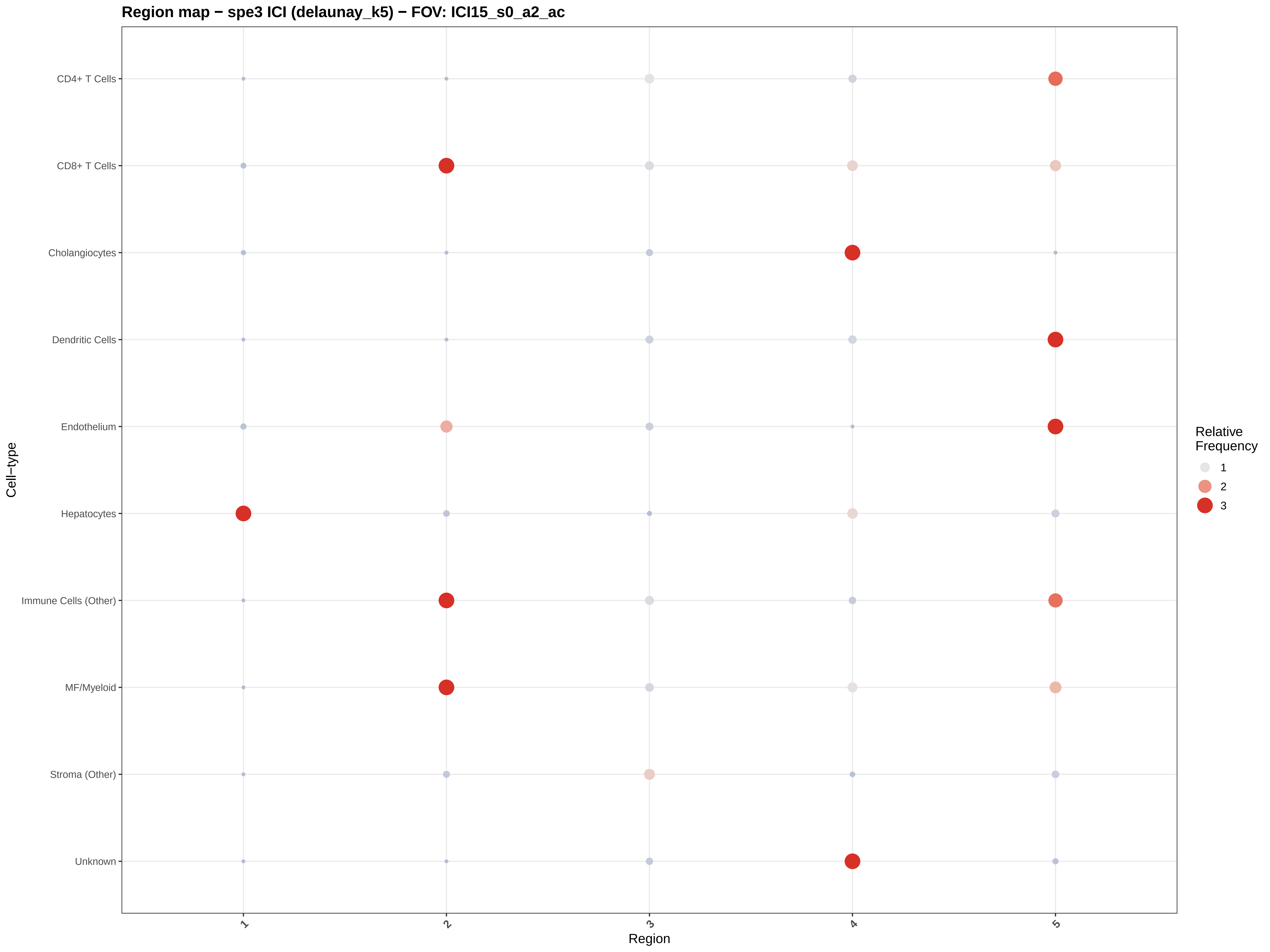

Region map − spe3 ICI (delaunay_k5) − FOV: ICI15_s0_a2_ac
CD4+ T Cells
CD8+ T Cells
Cholangiocytes
Dendritic Cells
Endothelium
Hepatocytes
Immune Cells (Other)
MF/Myeloid
Stroma (Other)
Unknown
Cell−type
1
2
3
4
5
Region
Relative Frequency
1
2
3

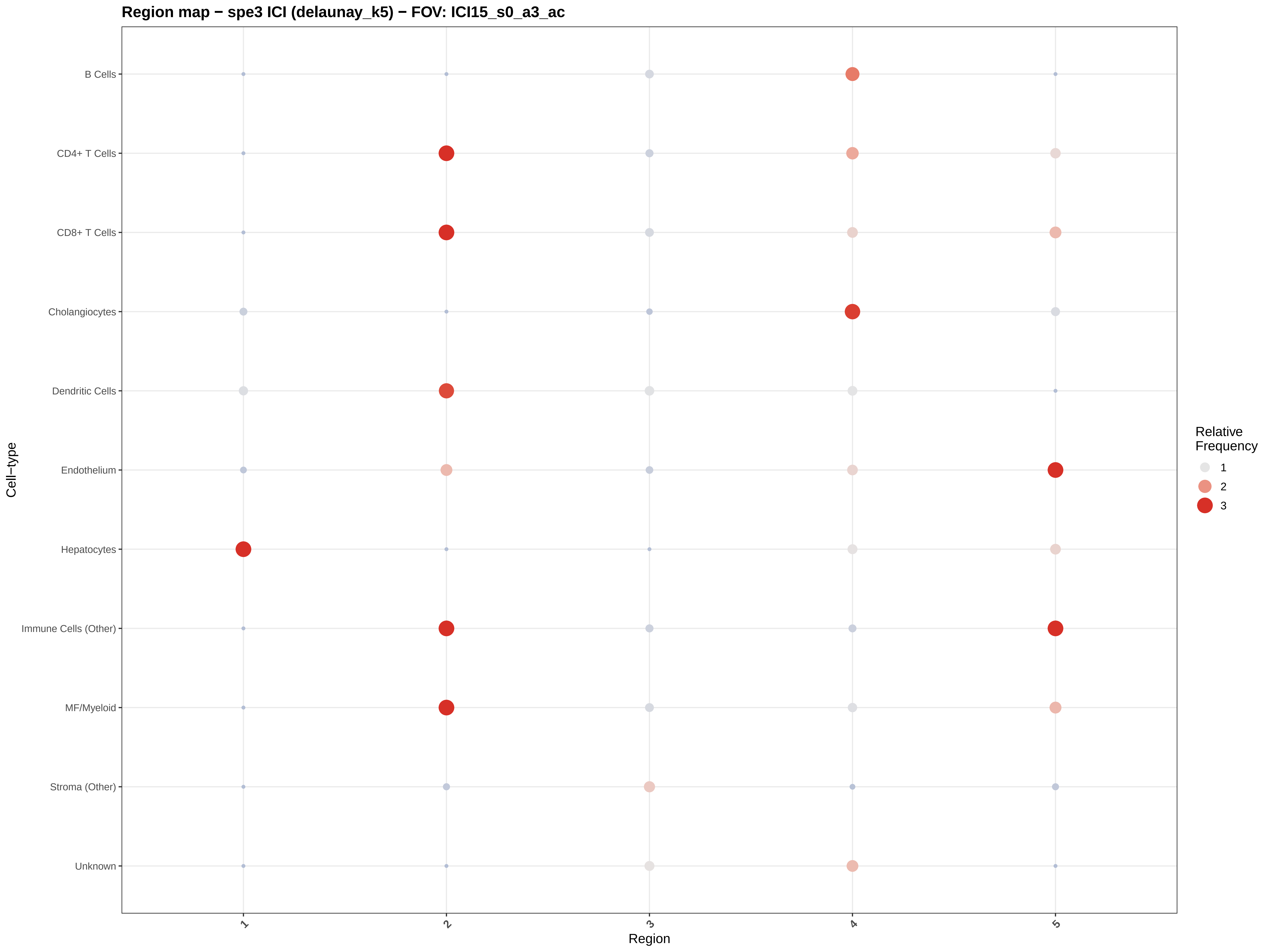
Region map − spe3 ICI (delaunay_k5) − FOV: ICI15_s0_a3_ac
B Cells
CD4+ T Cells
CD8+ T Cells
Cholangiocytes
Dendritic Cells
Endothelium
Hepatocytes
Immune Cells (Other)
MF/Myeloid
Stroma (Other)
Unknown
Cell−type
1
2
3
4
5
Region
Relative Frequency
1
2
3

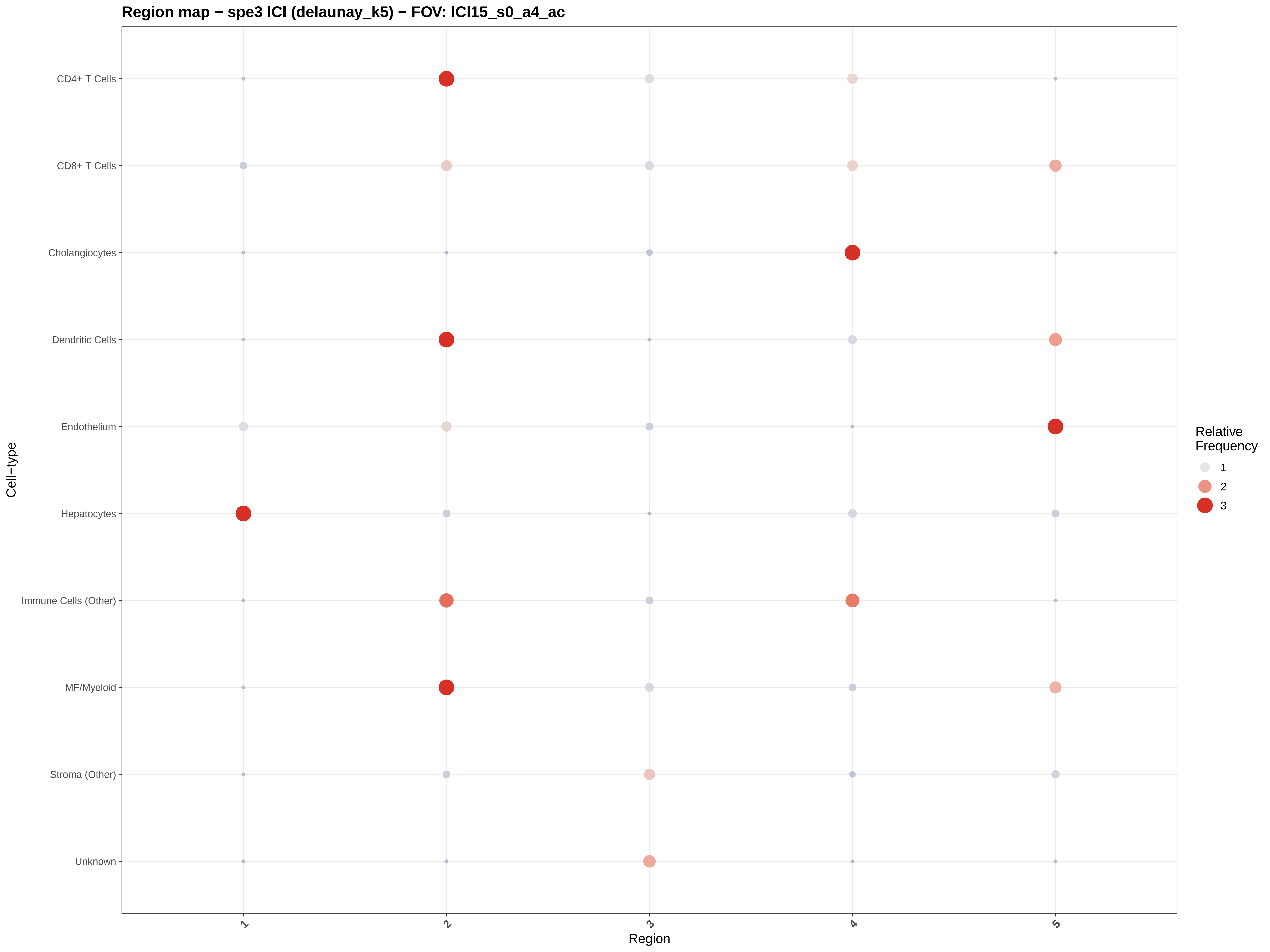

Region map − spe3 ICI (delaunay_k5) − FOV: ICI15_s0_a4_ac
CD4+ T Cells
CD8+ T Cells
Cholangiocytes
Dendritic Cells
Endothelium
Hepatocytes
Immune Cells (Other)
MF/Myeloid
Stroma (Other)
Unknown
Cell−type
1
2
3
4
5
Region
Relative Frequency
1
2
3

#### Region map − spe3 ICI (delaunay_k5) − FOV: ICI15_s0_a5_ac

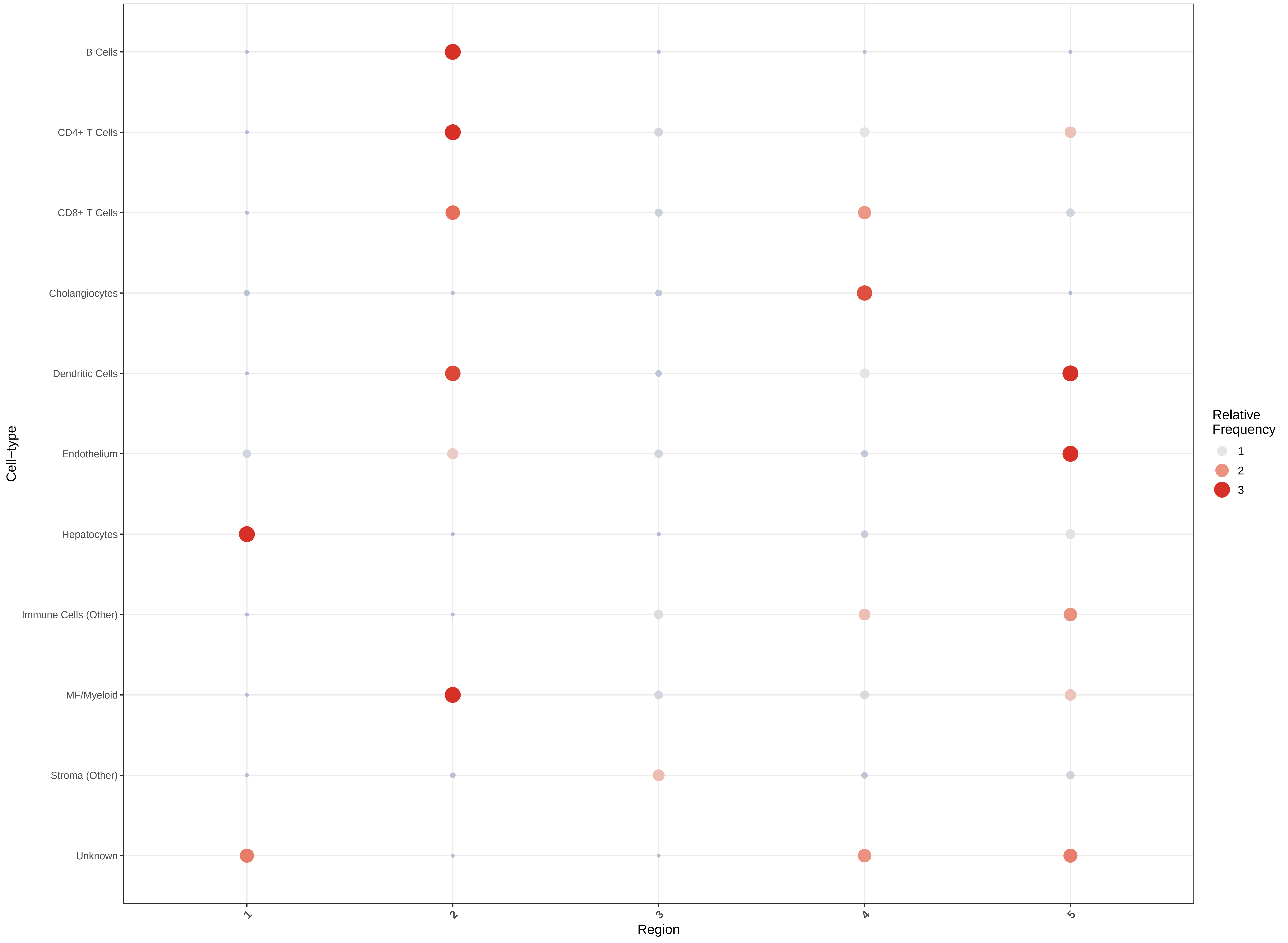

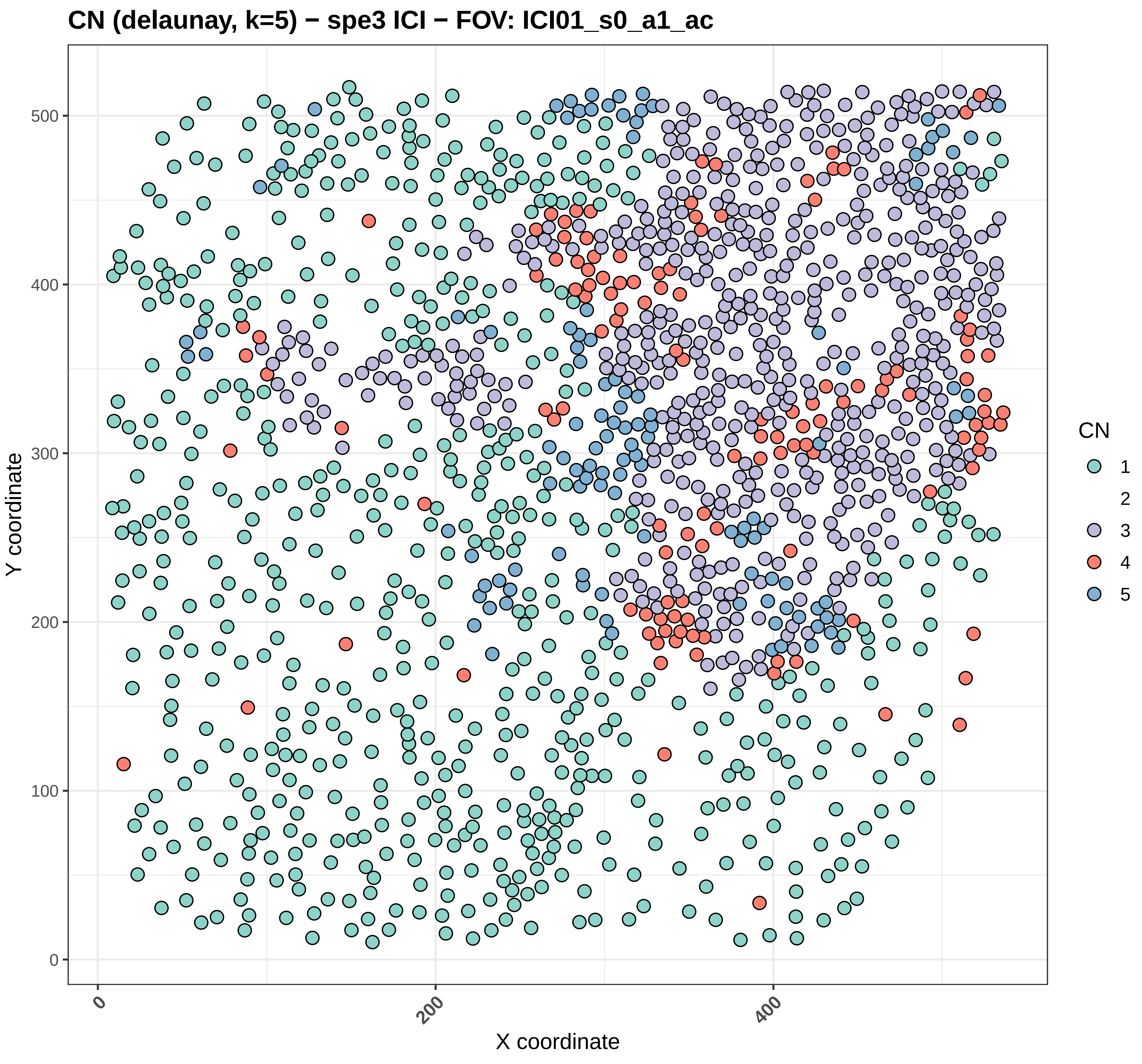
CN (delaunay, k=5) − spe3 ICI − FOV: ICI01_s0_a1_ac
Y coordinate
X coordinate
0
100
200
300
400
500
0
200
400
CN
1
2
3
4
5

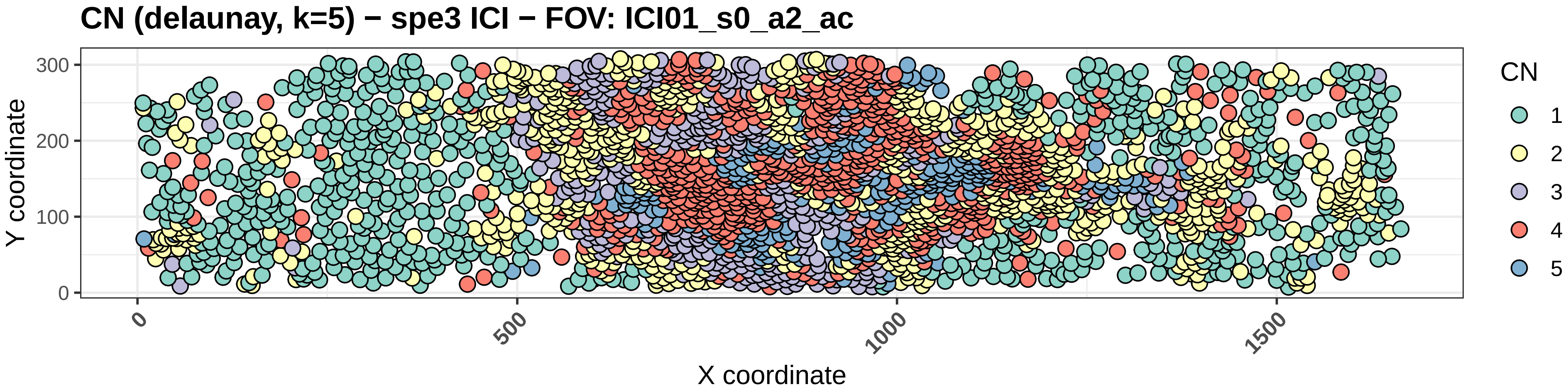
CN (delaunay, k=5) − spe3 ICI − FOV: ICI01_s0_a2_ac
Y coordinate
0
100
200
300
X coordinate
0
500
1000
1500
CN
1
2
3
4
5

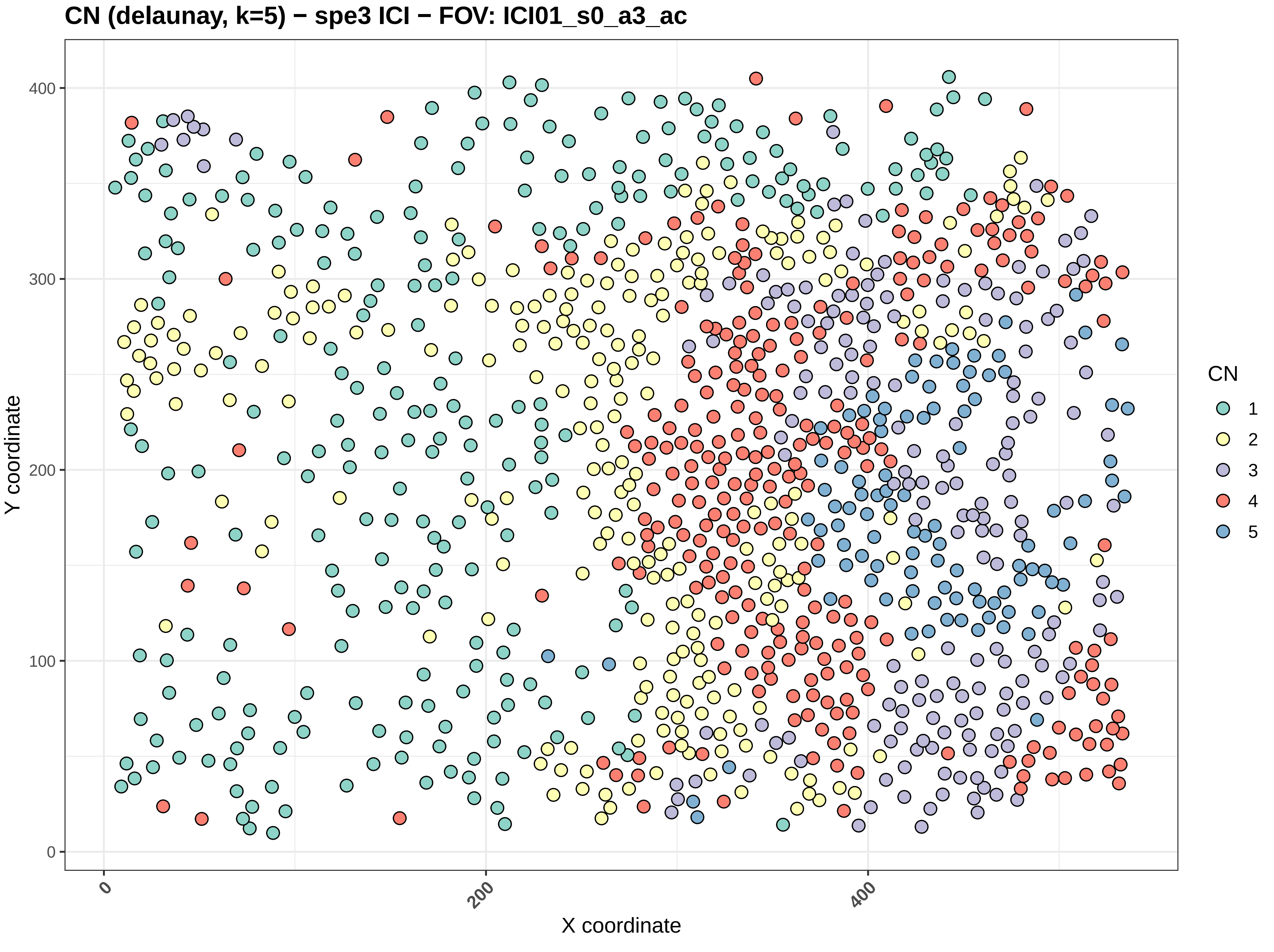
CN (delaunay, k=5) − spe3 ICI − FOV: ICI01_s0_a3_ac
Y coordinate
X coordinate
0
100
200
300
400
0
200
400
CN
1
2
3
4
5

#### CN (delaunay, k=5) − spe3 ICI − FOV: ICI01_s0_a5_ac

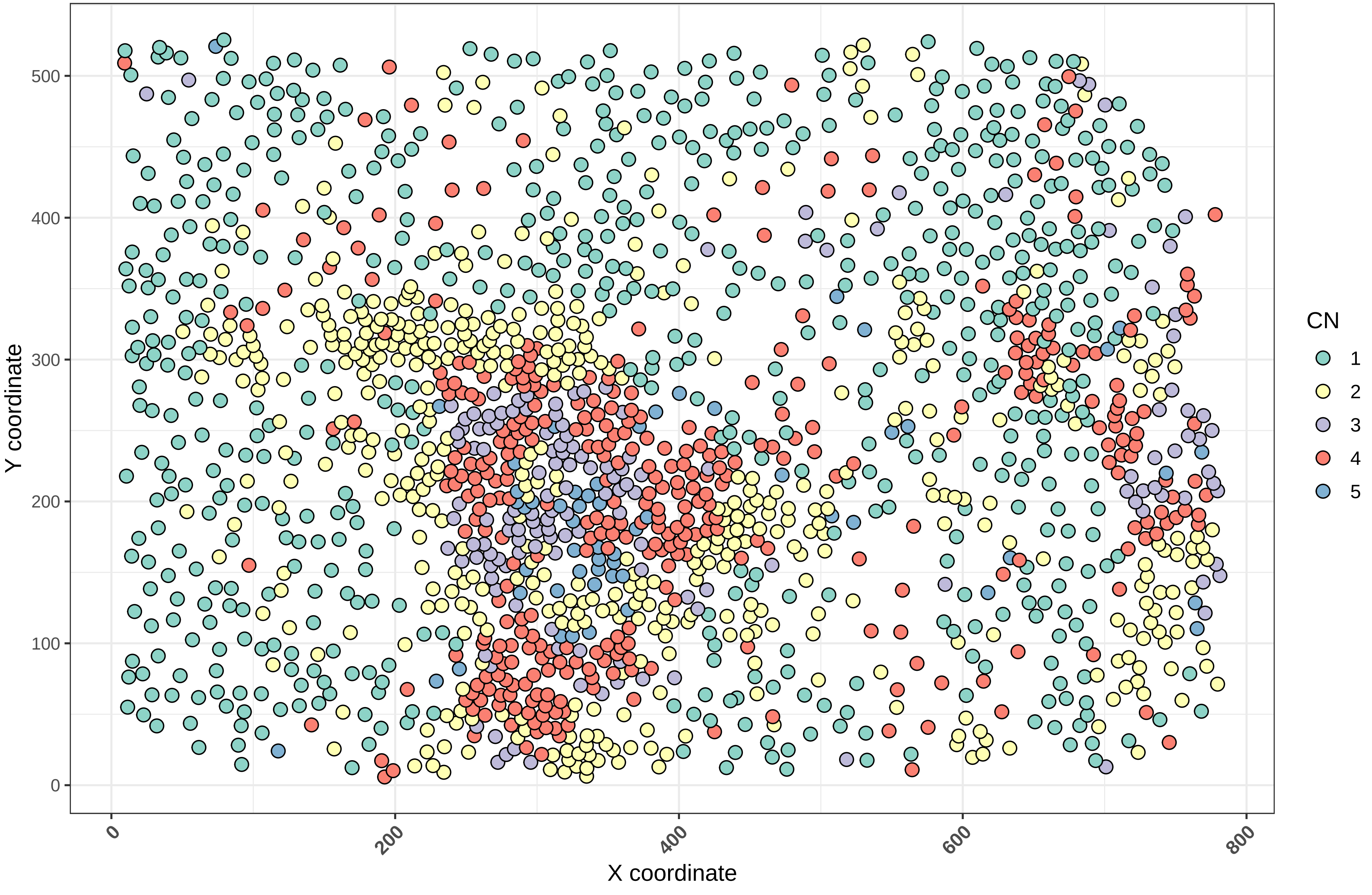

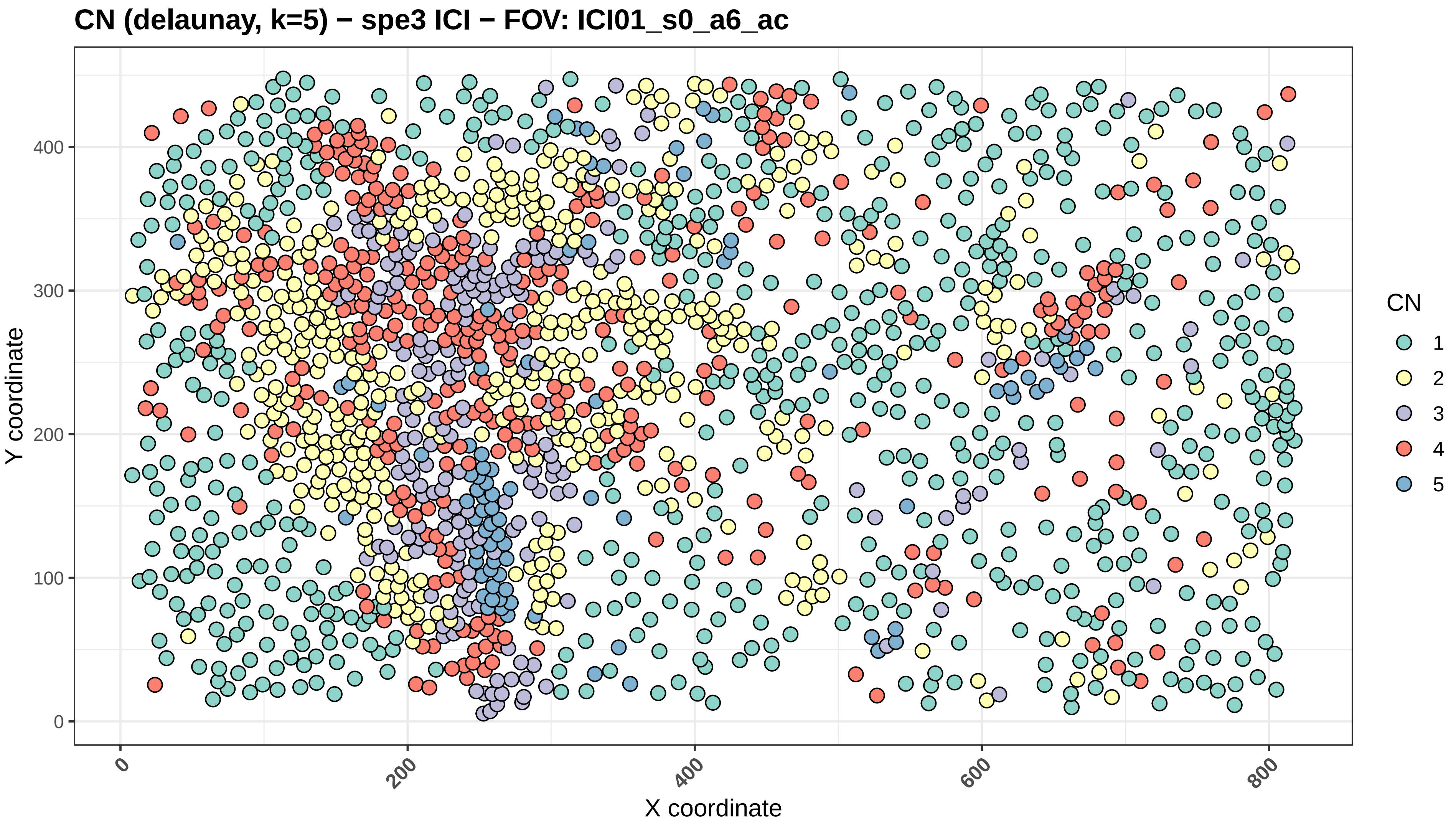
CN (delaunay, k=5) − spe3 ICI − FOV: ICI01_s0_a6_ac
Y coordinate
X coordinate
0
100
200
300
400
0
200
400
600
800
CN
1
2
3
4
5

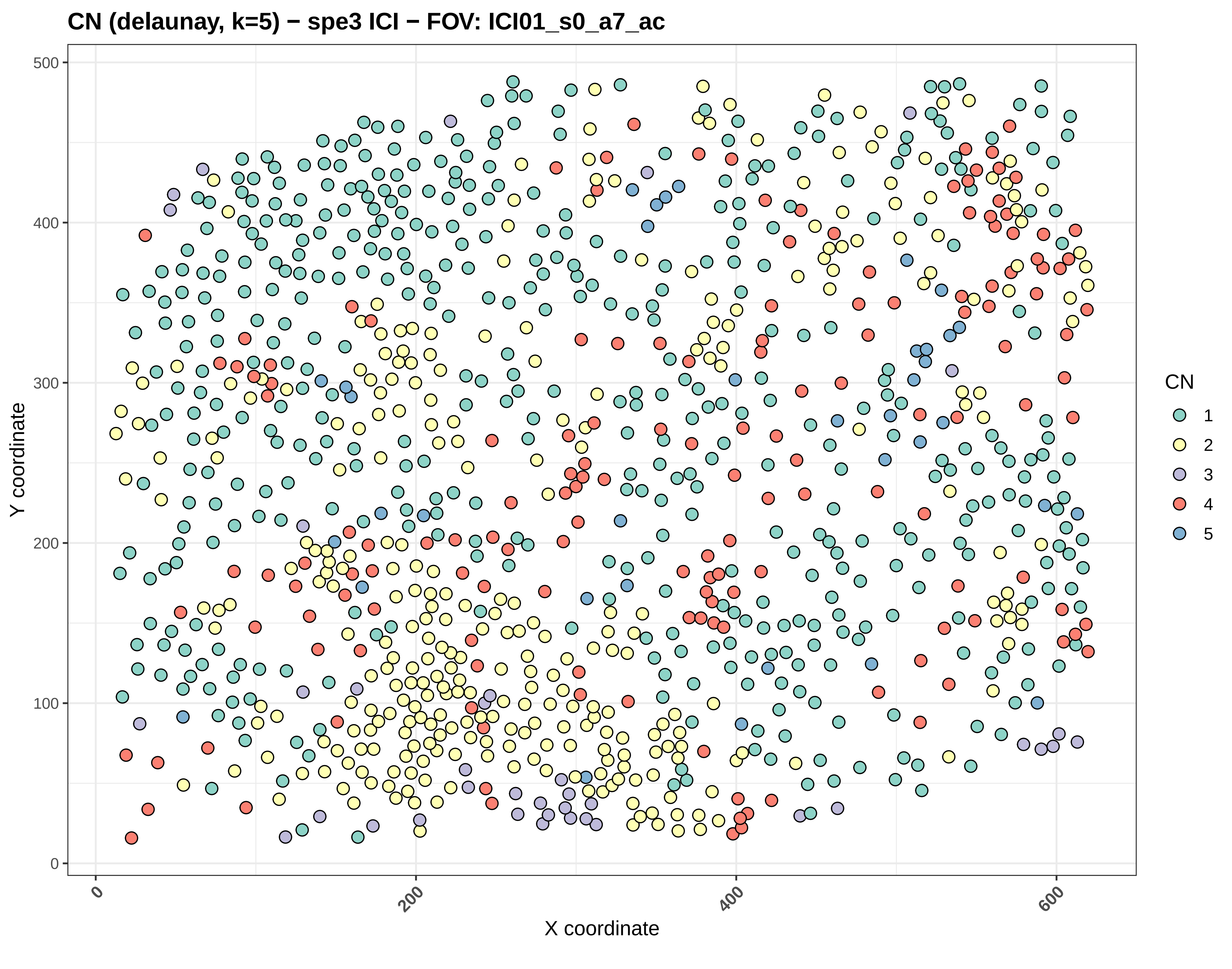
CN (delaunay, k=5) − spe3 ICI − FOV: ICI01_s0_a7_ac
Y coordinate
X coordinate
0
100
200
300
400
500
0
200
400
600
CN
1
2
3
4
5

CN (delaunay, k=5) − spe3 ICI − FOV: ICI01_s0_a8_ac

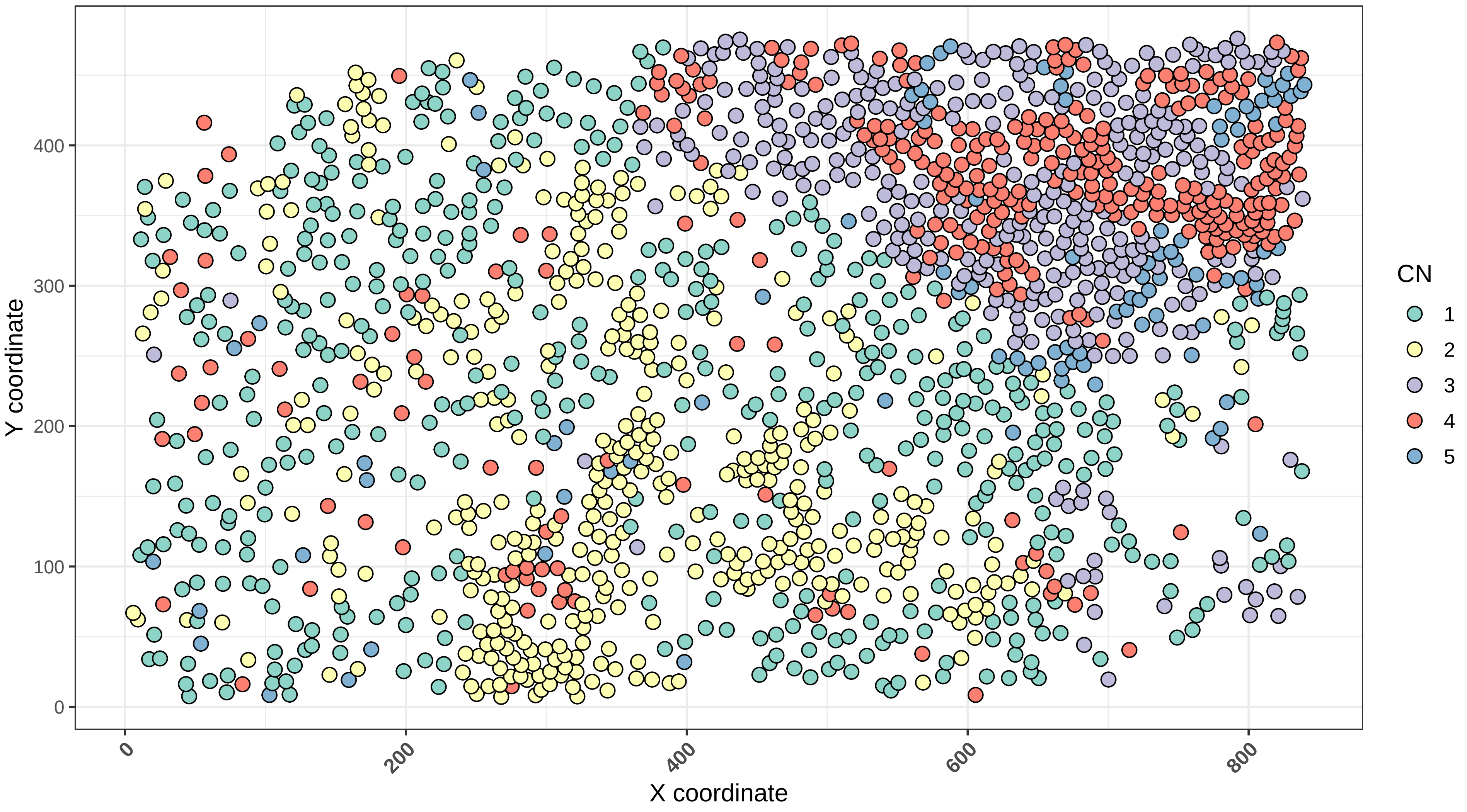

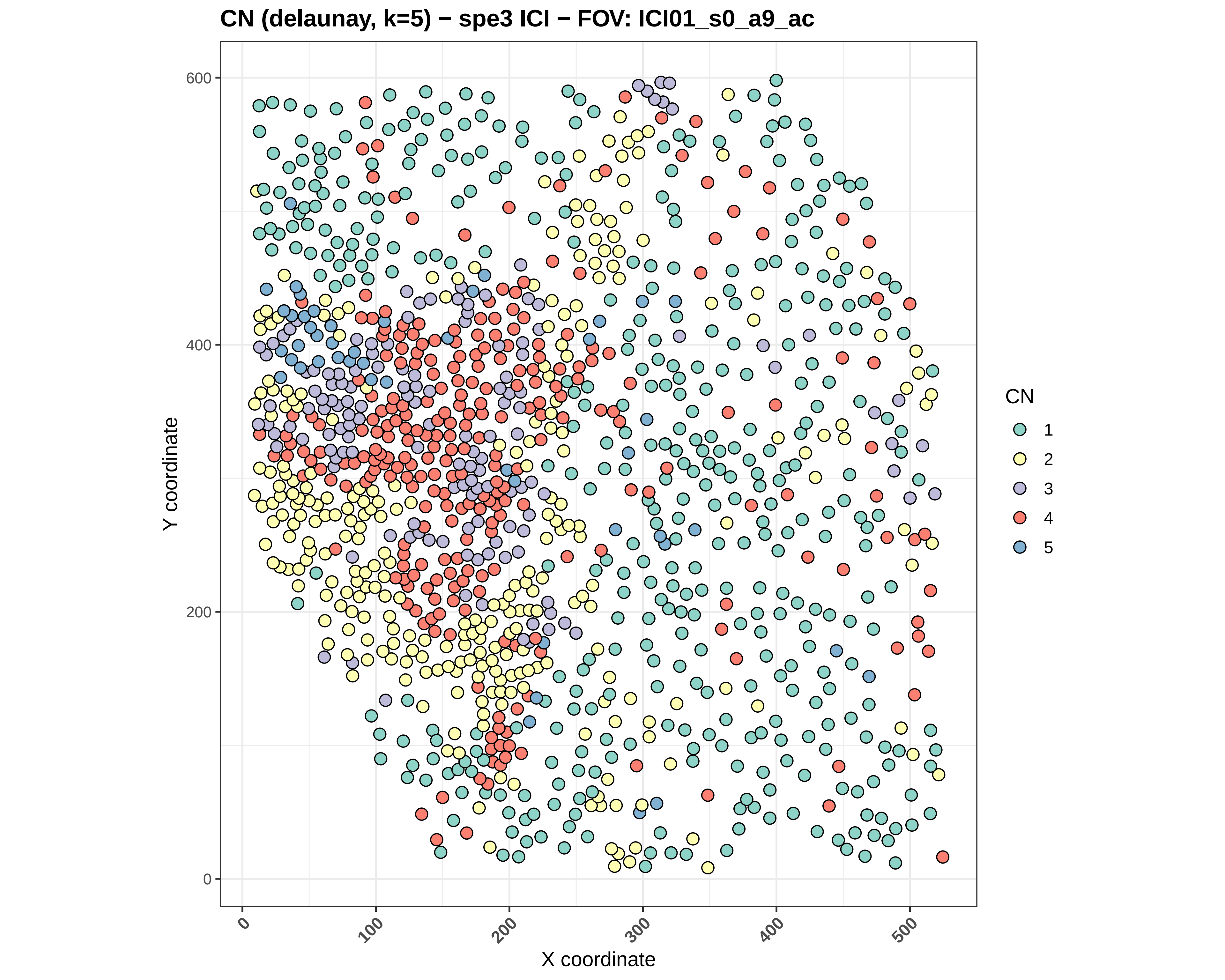
CN (delaunay, k=5) − spe3 ICI − FOV: ICI01_s0_a9_ac
Y coordinate
600
400
200
0
X coordinate
0
100
200
300
400
500
CN
1
2
3
4
5

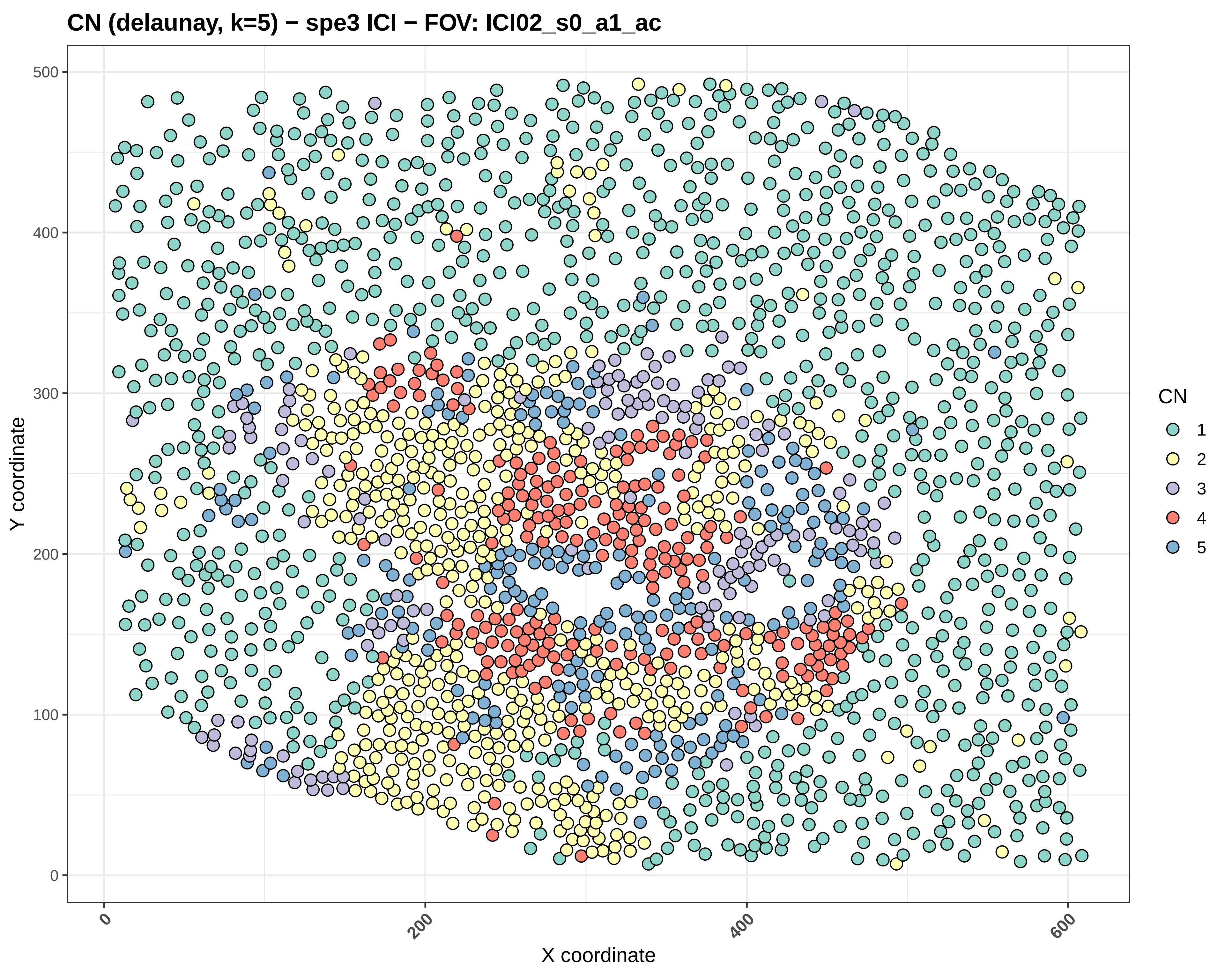

CN (delaunay, k=5) − spe3 ICI − FOV: ICI02_s0_a1_ac
Y coordinate
X coordinate
0
100
200
300
400
500
0
200
400
600
CN
1
2
3
4
5

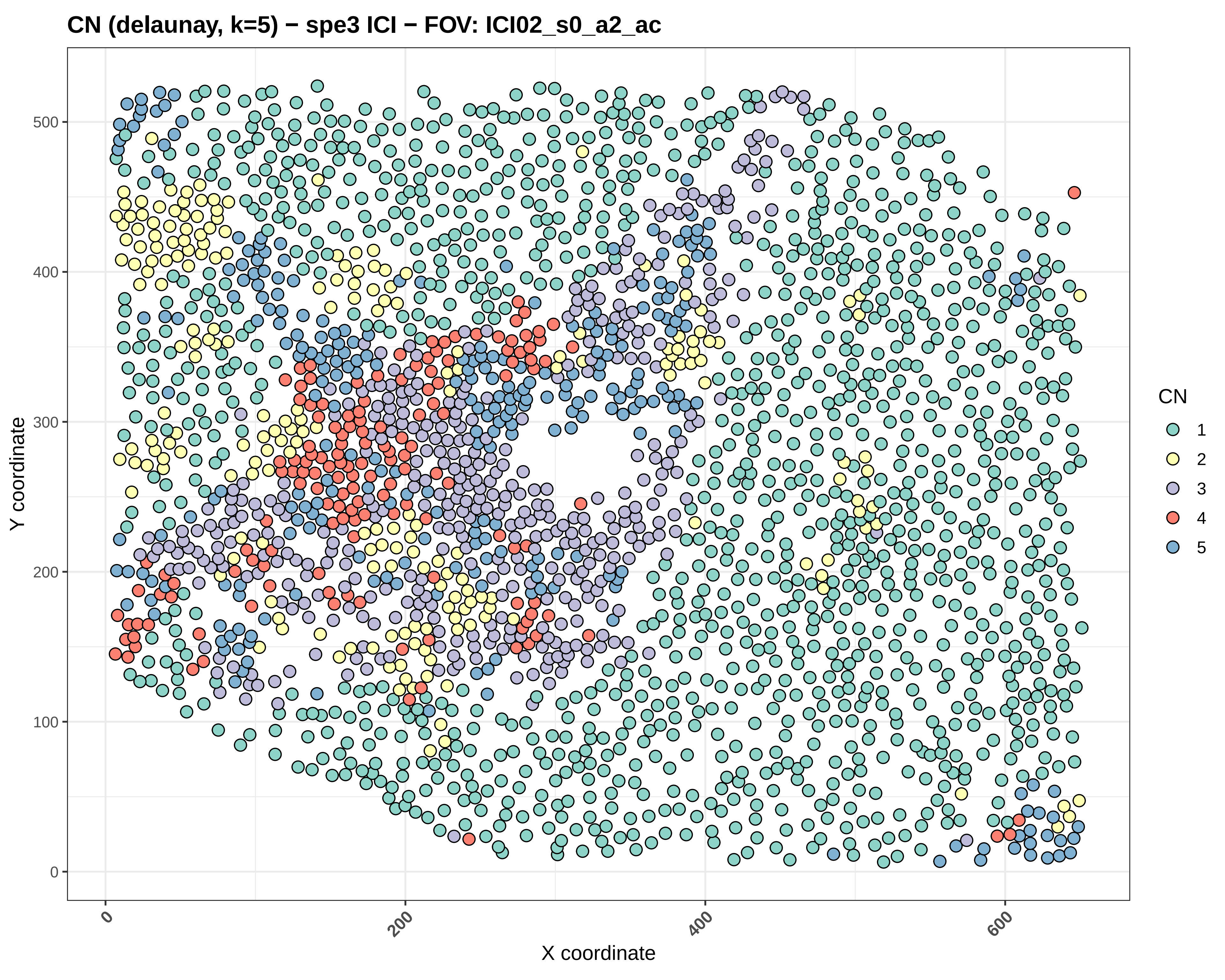
CN (delaunay, k=5) − spe3 ICI − FOV: ICI02_s0_a2_ac
Y coordinate
X coordinate
0
100
200
300
400
500
0
200
400
600
CN
1
2
3
4
5

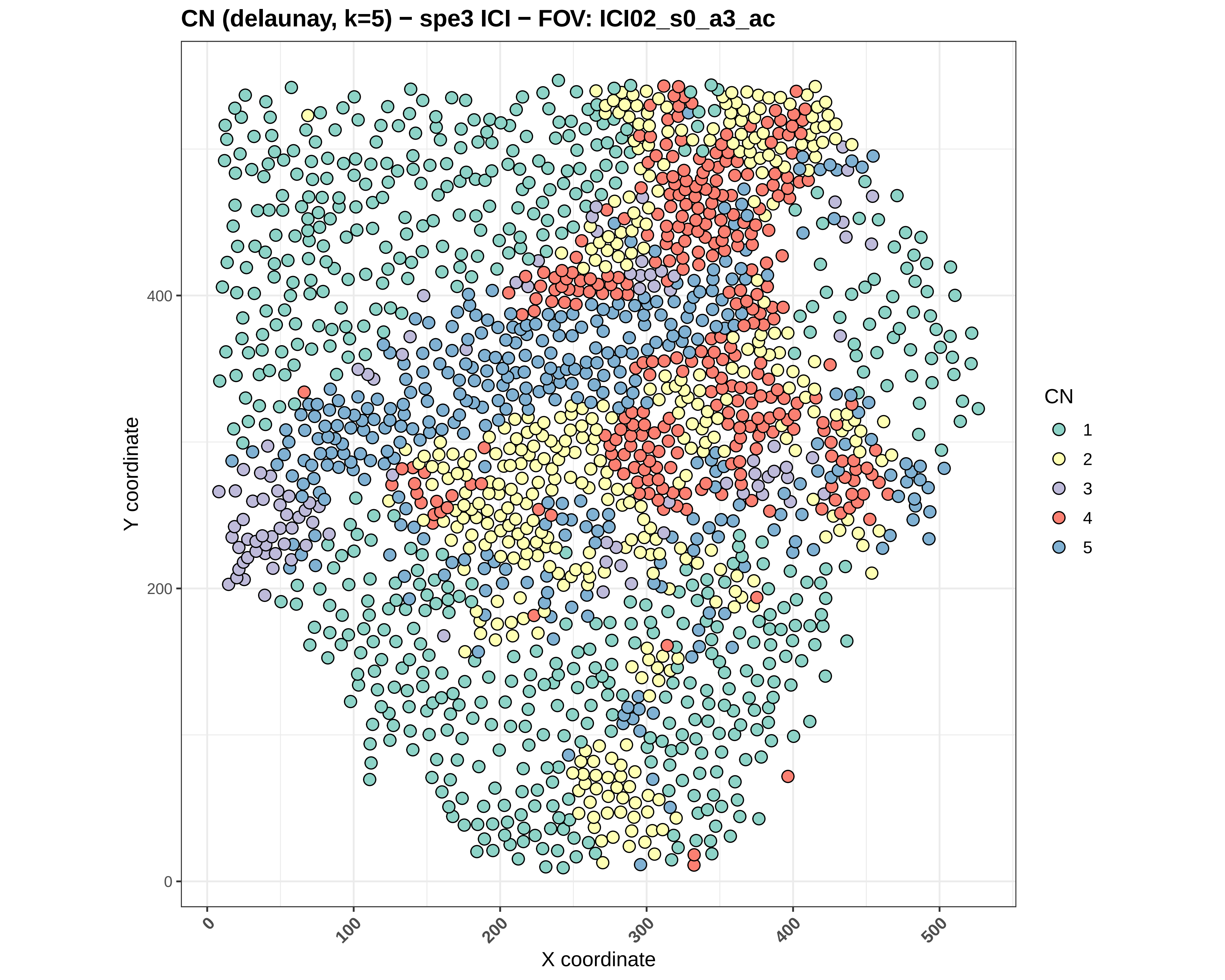

CN (delaunay, k=5) − spe3 ICI − FOV: ICI02_s0_a3_ac
Y coordinate
X coordinate
0
200
400
0
100
200
300
400
500
CN
1
2
3
4
5

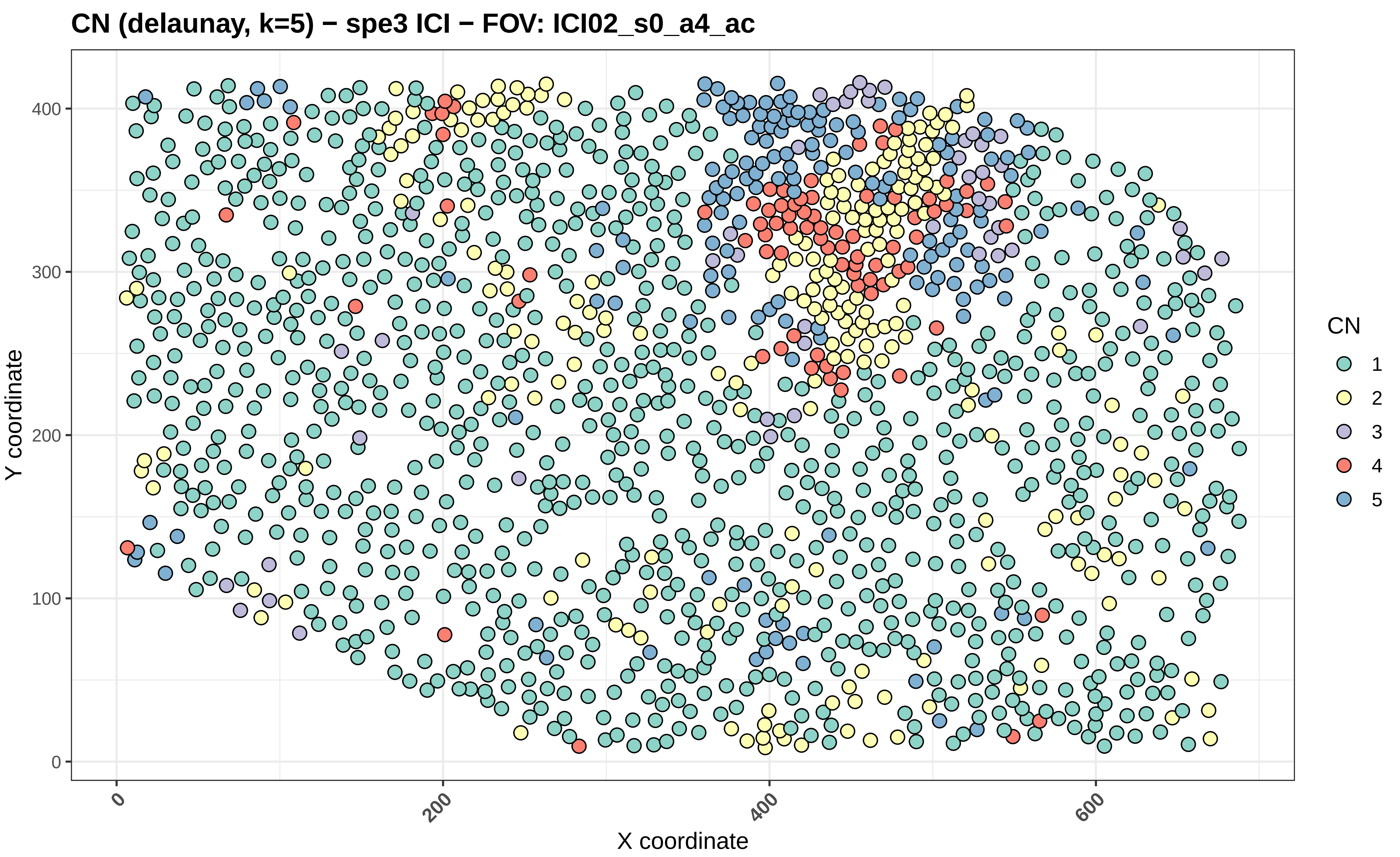
CN (delaunay, k=5) − spe3 ICI − FOV: ICI02_s0_a4_ac
Y coordinate
X coordinate
0
100
200
300
400
0
200
400
600
CN
1
2
3
4
5

#### CN (delaunay, k=5) − spe3 ICI − FOV: ICI02_s0_a5_ac

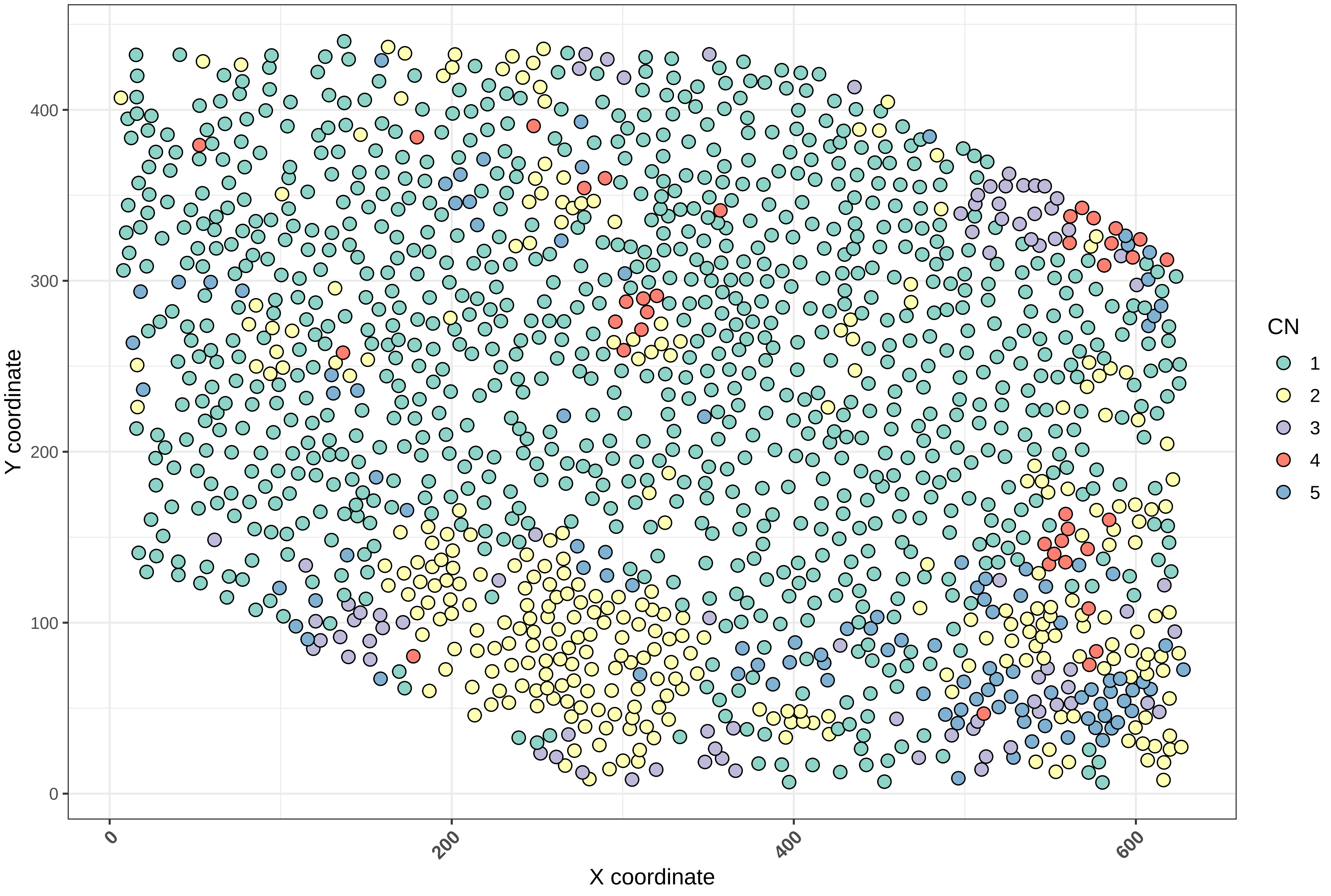

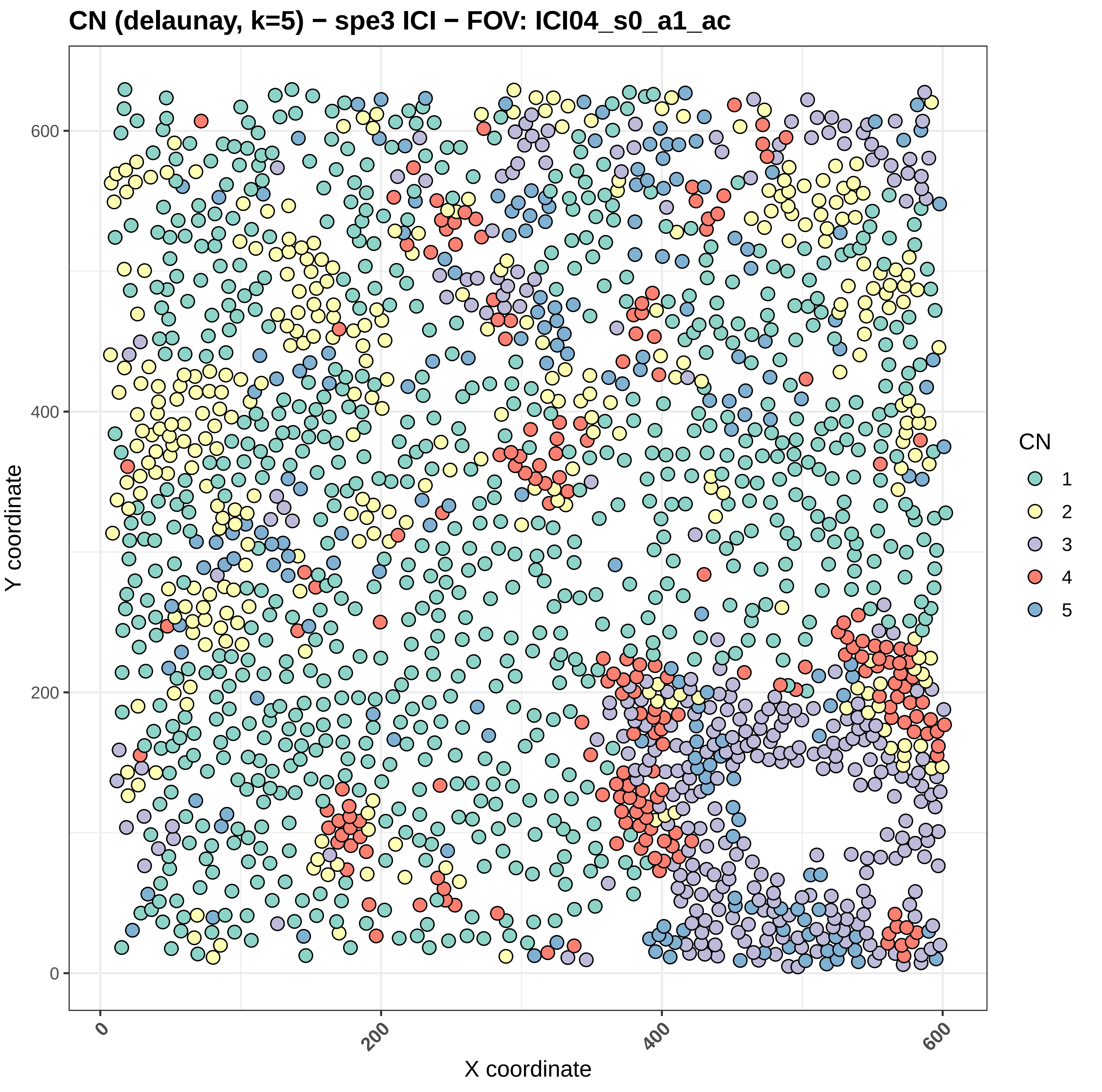
CN (delaunay, k=5) − spe3 ICI − FOV: ICI04_s0_a1_ac
Y coordinate
X coordinate
0
200
400
600
CN
1
2
3
4
5

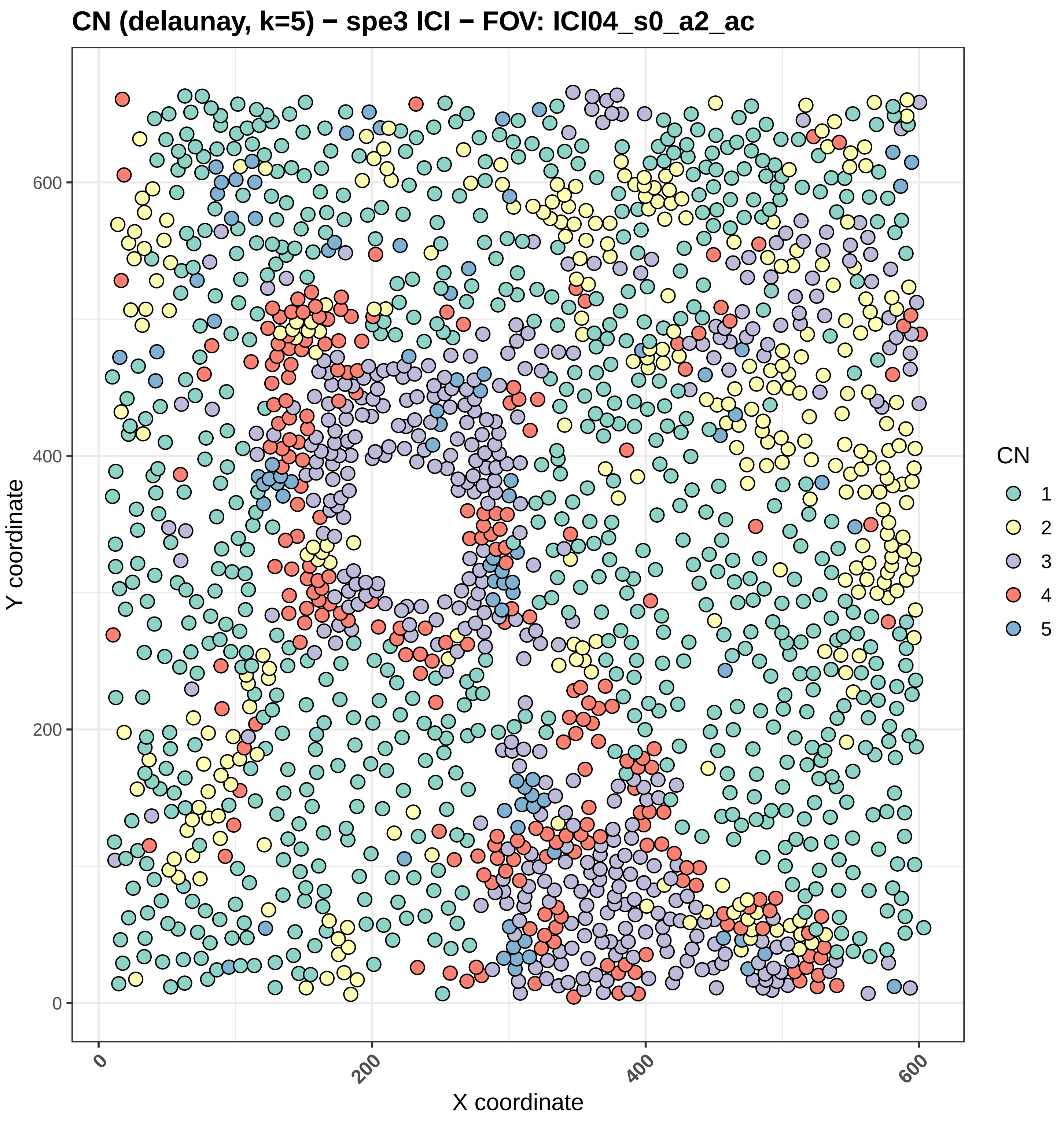

CN (delaunay, k=5) − spe3 ICI − FOV: ICI04_s0_a2_ac
Y coordinate
X coordinate
0
200
400
600
CN
1
2
3
4
5

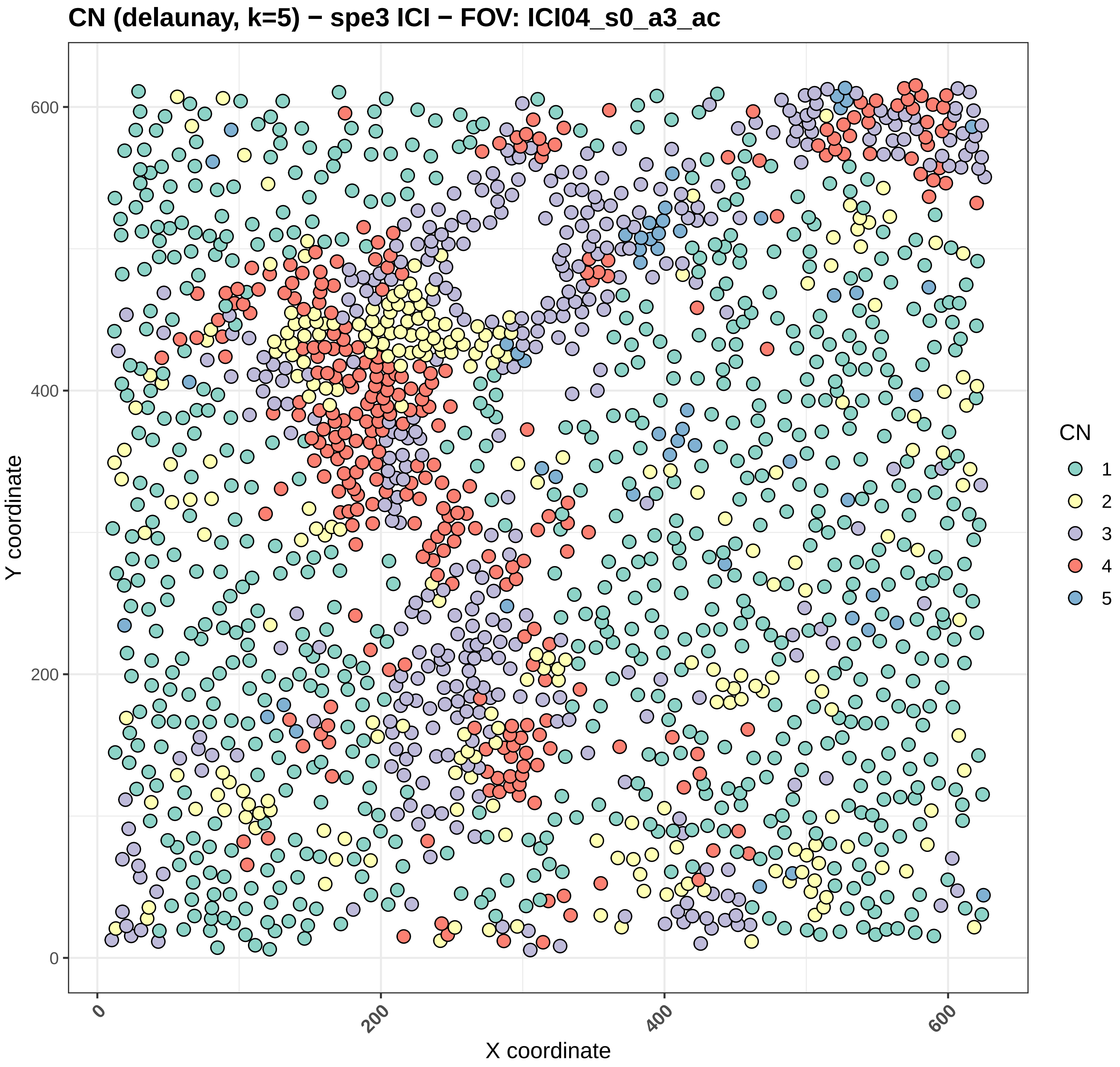

CN (delaunay, k=5) − spe3 ICI − FOV: ICI04_s0_a3_ac
Y coordinate
X coordinate
0
200
400
600
CN
1
2
3
4
5

### CN (delaunay, k=5) − spe3 ICI − FOV: ICI04_s0_a4_ac

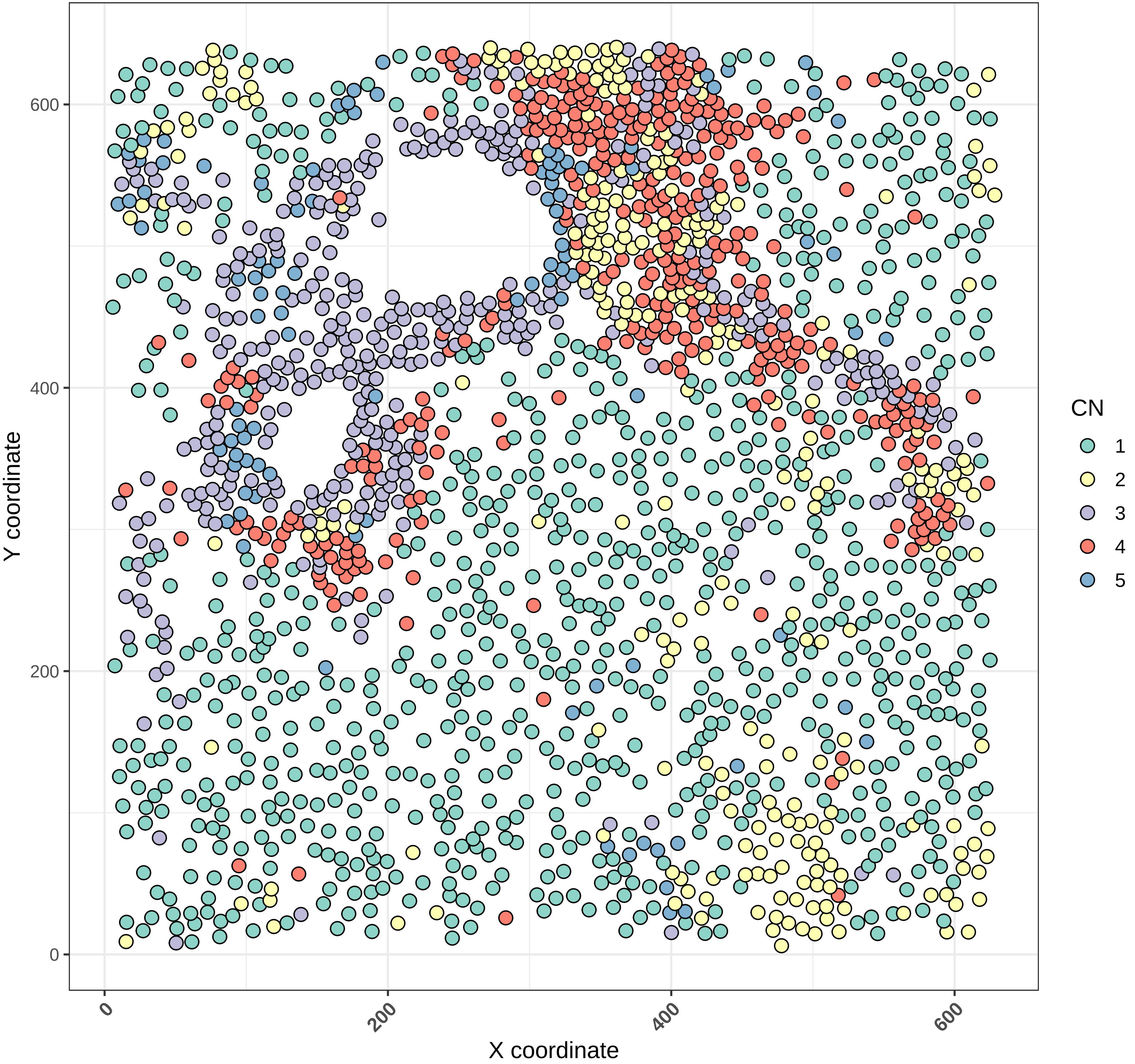

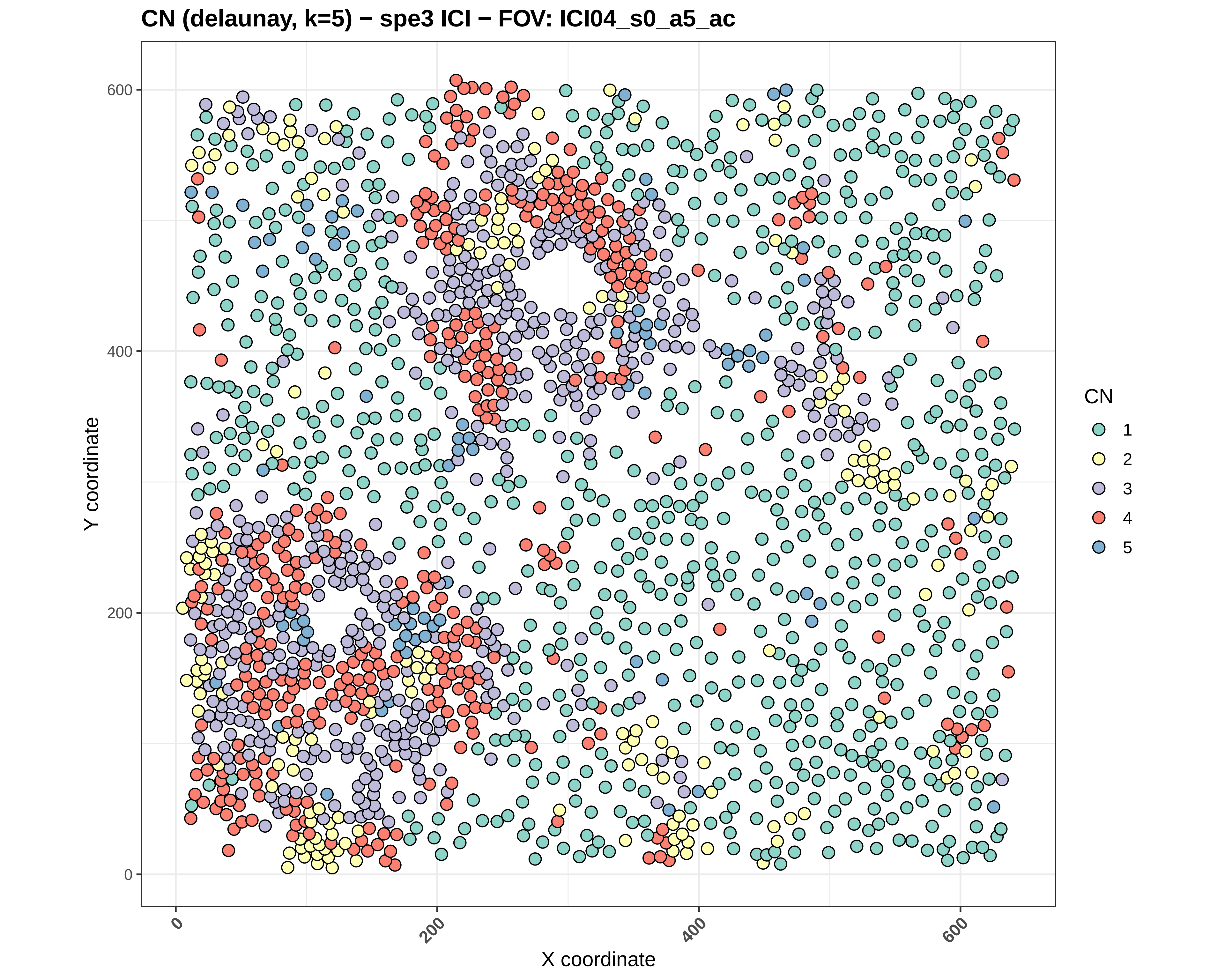
CN (delaunay, k=5) − spe3 ICI − FOV: ICI04_s0_a5_ac
Y coordinate
X coordinate
0
200
400
600
0
200
400
600
CN
1
2
3
4
5

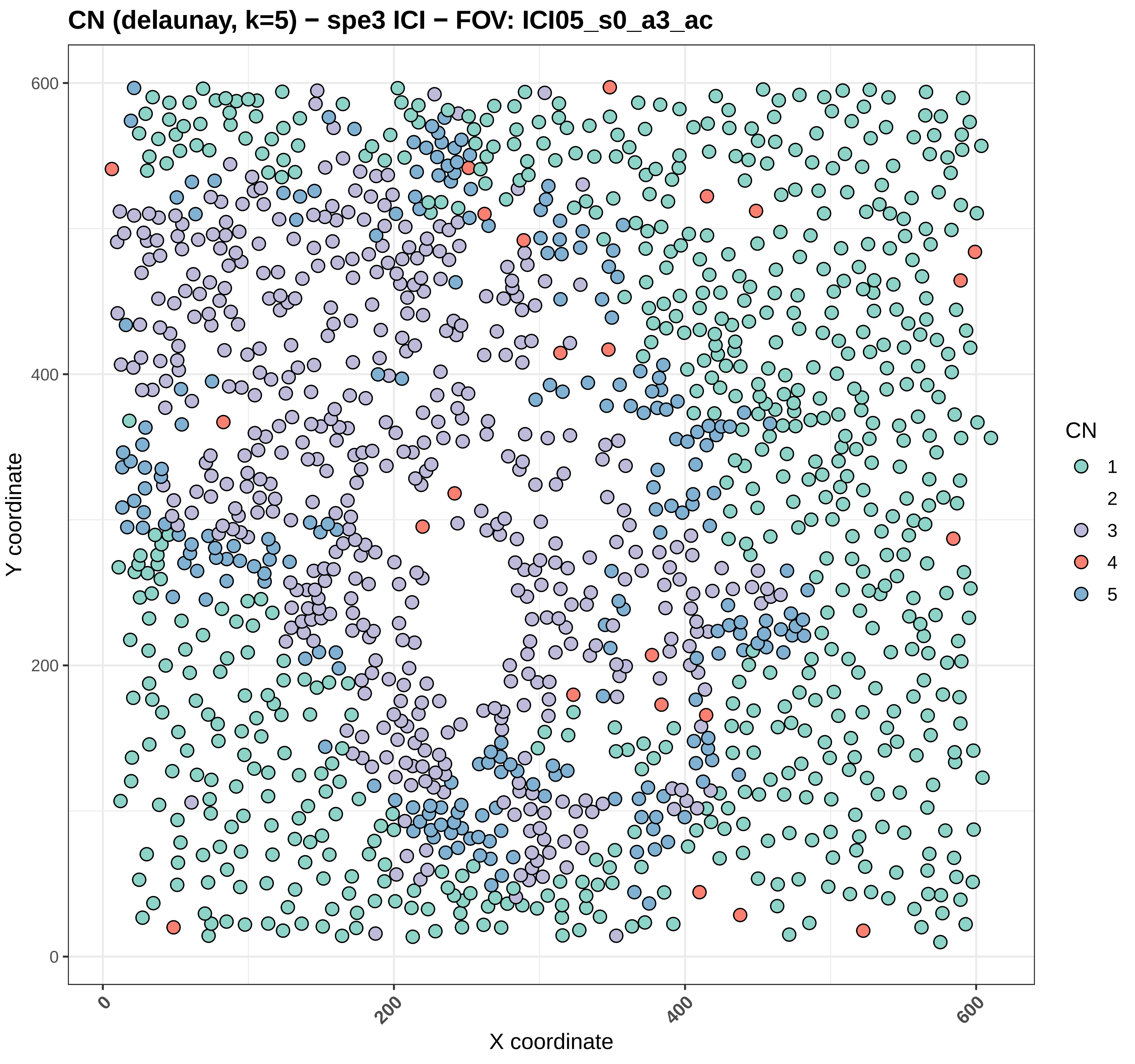
CN (delaunay, k=5) − spe3 ICI − FOV: ICI05_s0_a3_ac
Y coordinate
X coordinate
0
200
400
600
CN
1
2
3
4
5

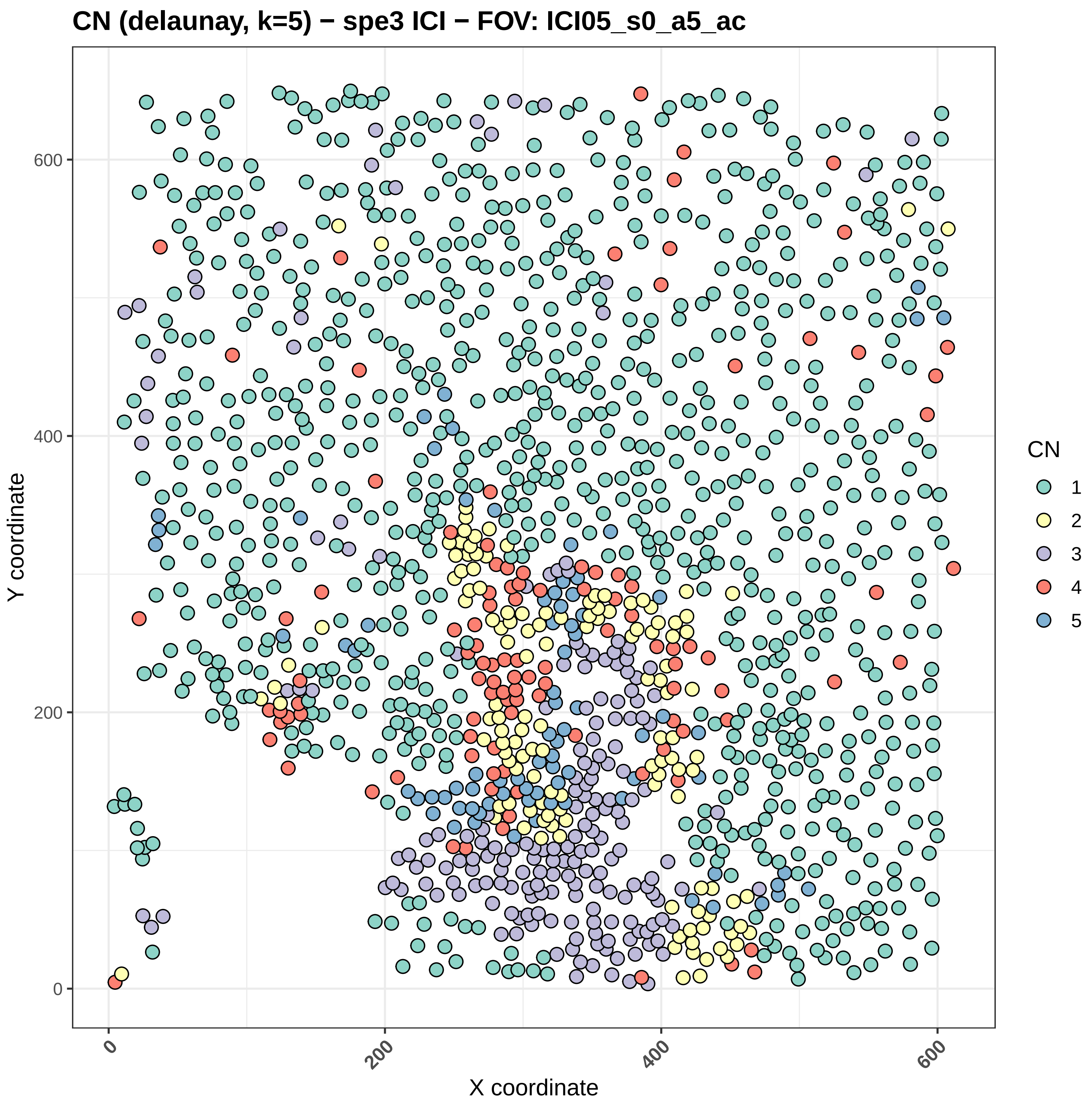
CN (delaunay, k=5) − spe3 ICI − FOV: ICI05_s0_a5_ac
Y coordinate
X coordinate
0
200
400
600
CN
1
2
3
4
5

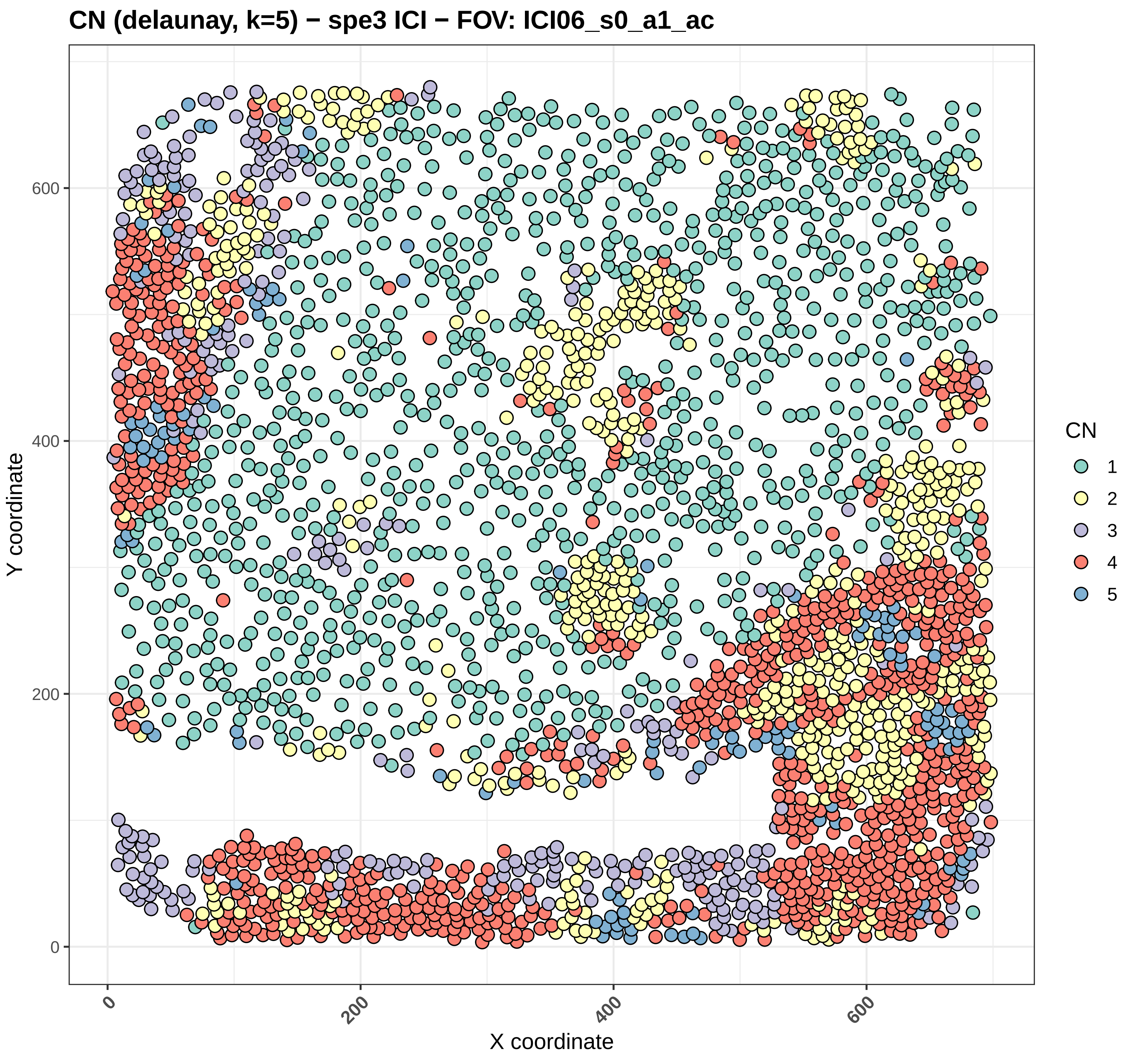
CN (delaunay, k=5) − spe3 ICI − FOV: ICI06_s0_a1_ac
Y coordinate
X coordinate
0
200
400
600
0
200
400
600
CN
1
2
3
4
5

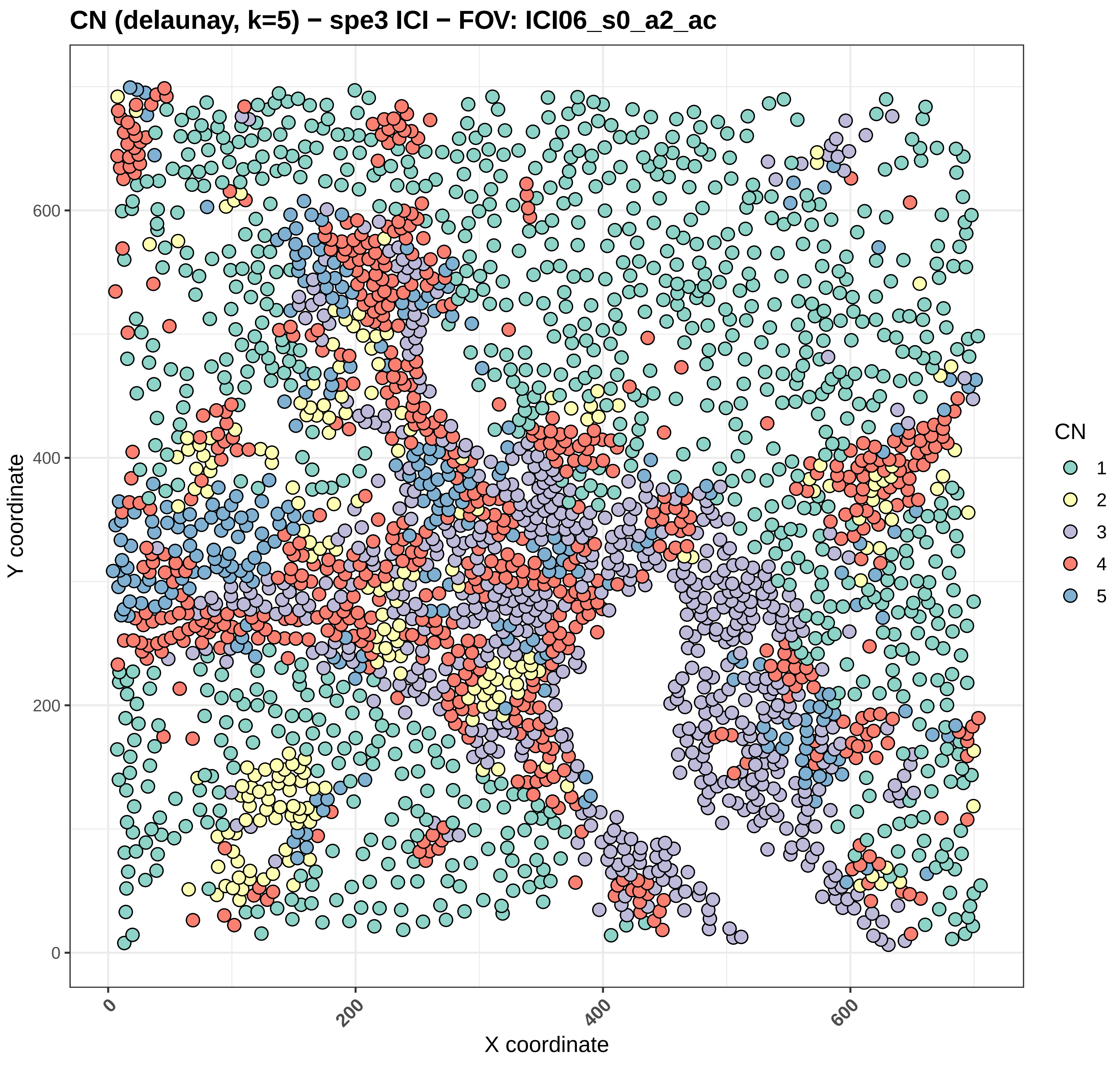
CN (delaunay, k=5) − spe3 ICI − FOV: ICI06_s0_a2_ac
Y coordinate
X coordinate
0
200
400
600
CN
1
2
3
4
5

CN (delaunay, k=5) − spe3 ICI − FOV: ICI06_s0_a3_ac

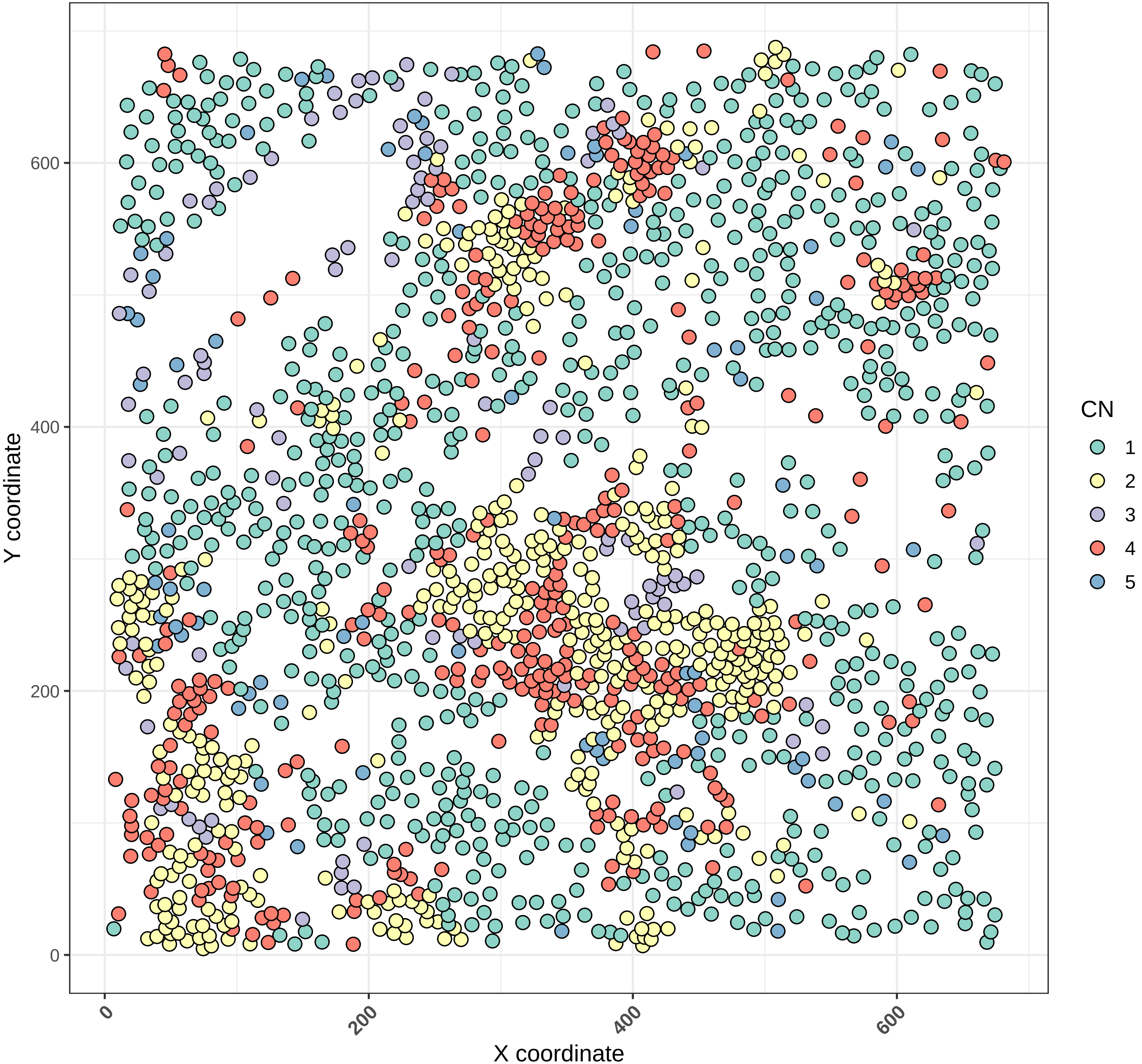

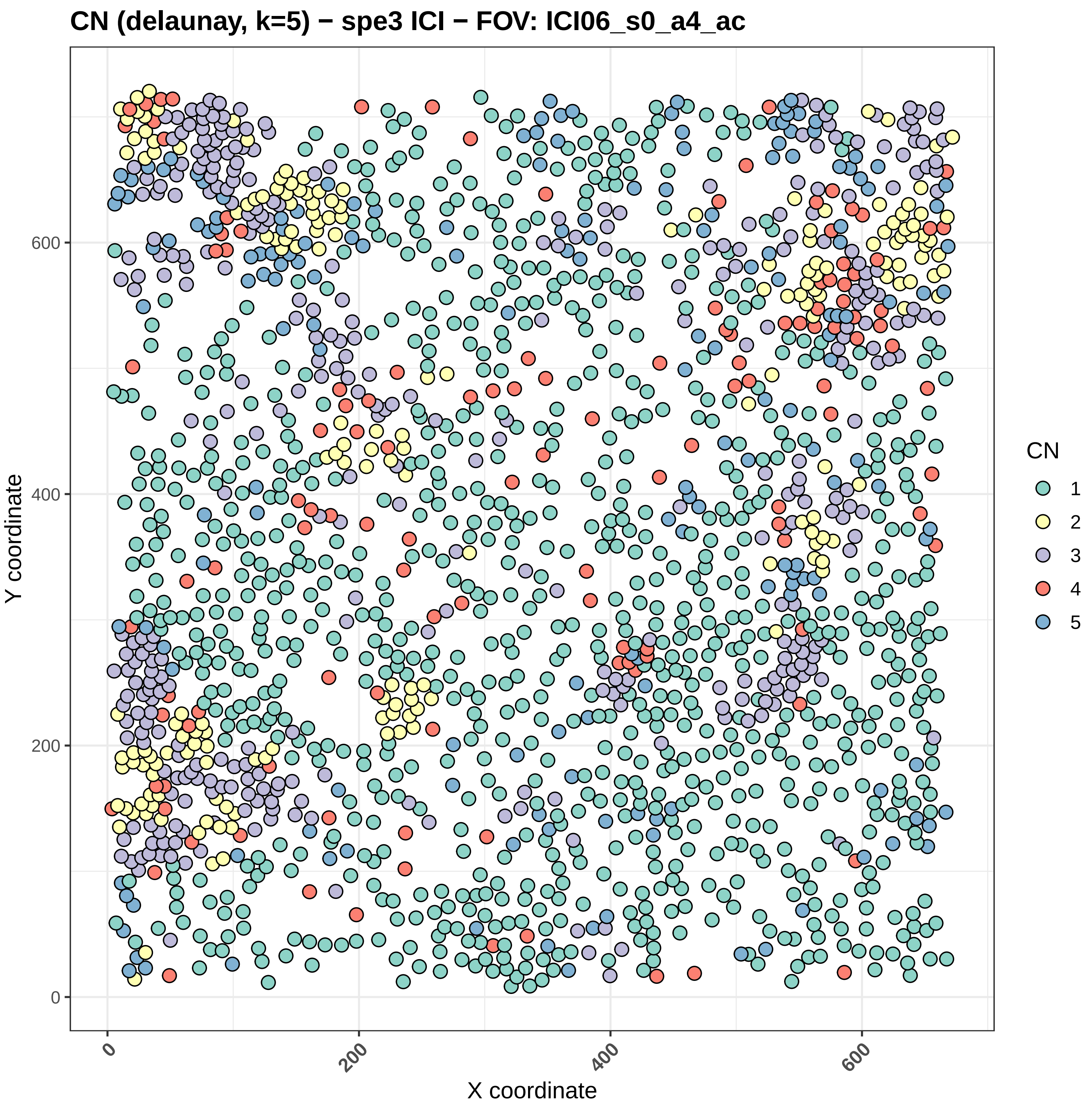
CN (delaunay, k=5) − spe3 ICI − FOV: ICI06_s0_a4_ac
Y coordinate
X coordinate
0
200
400
600
CN
1
2
3
4
5

#### CN (delaunay, k=5) − spe3 ICI − FOV: ICI06_s0_a5_ac

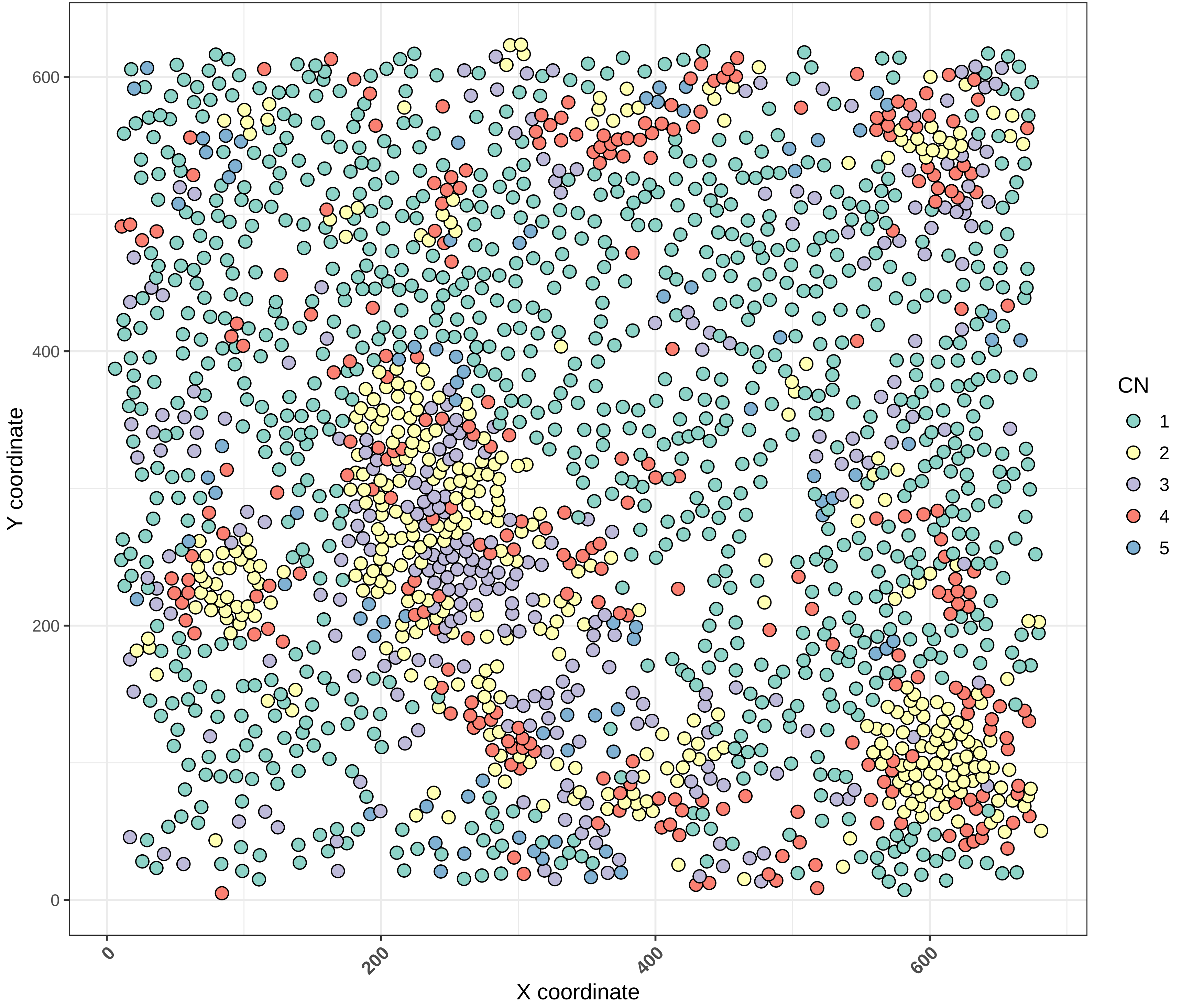

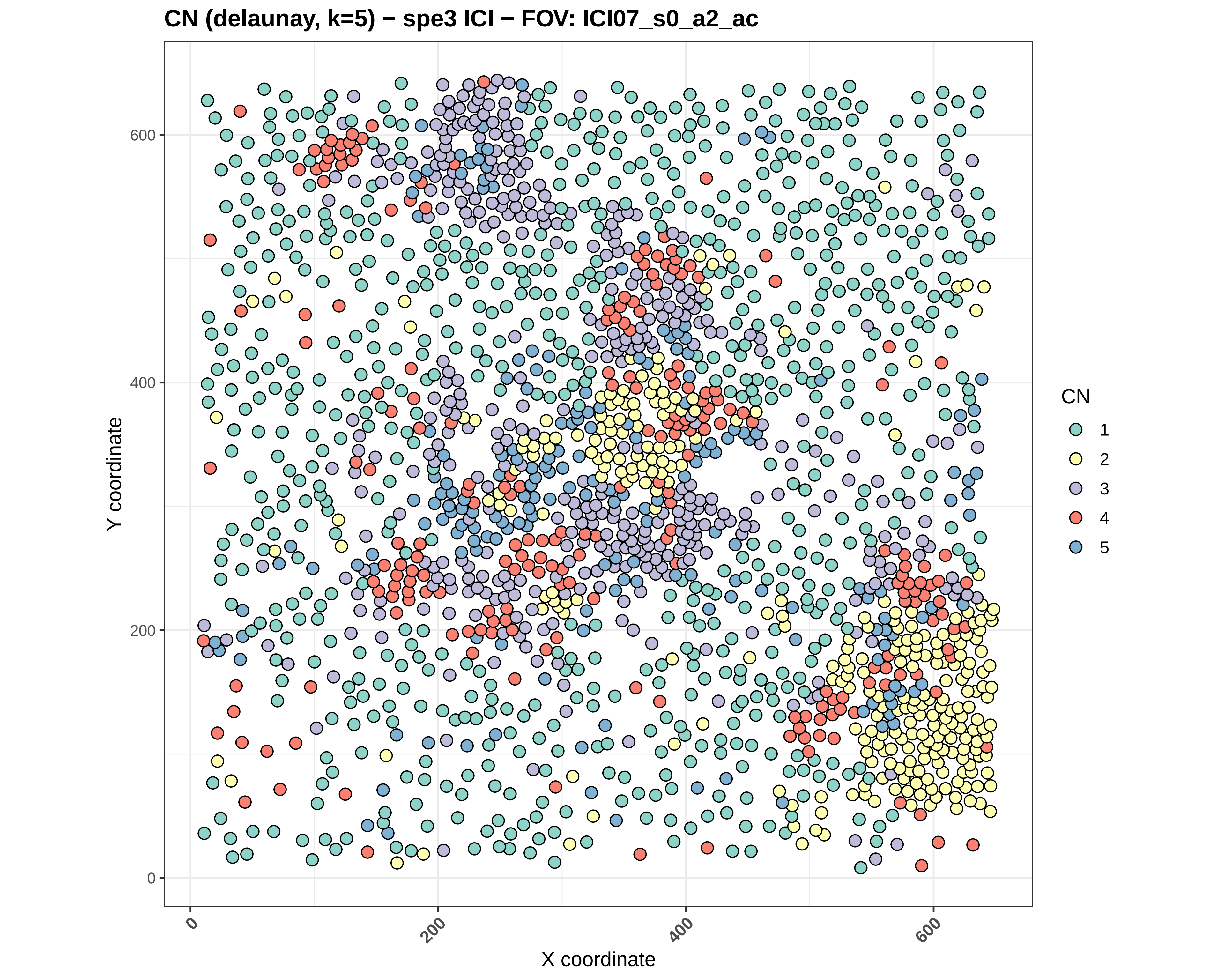
CN (delaunay, k=5) − spe3 ICI − FOV: ICI07_s0_a2_ac
Y coordinate
X coordinate
0
200
400
600
CN
1
2
3
4
5

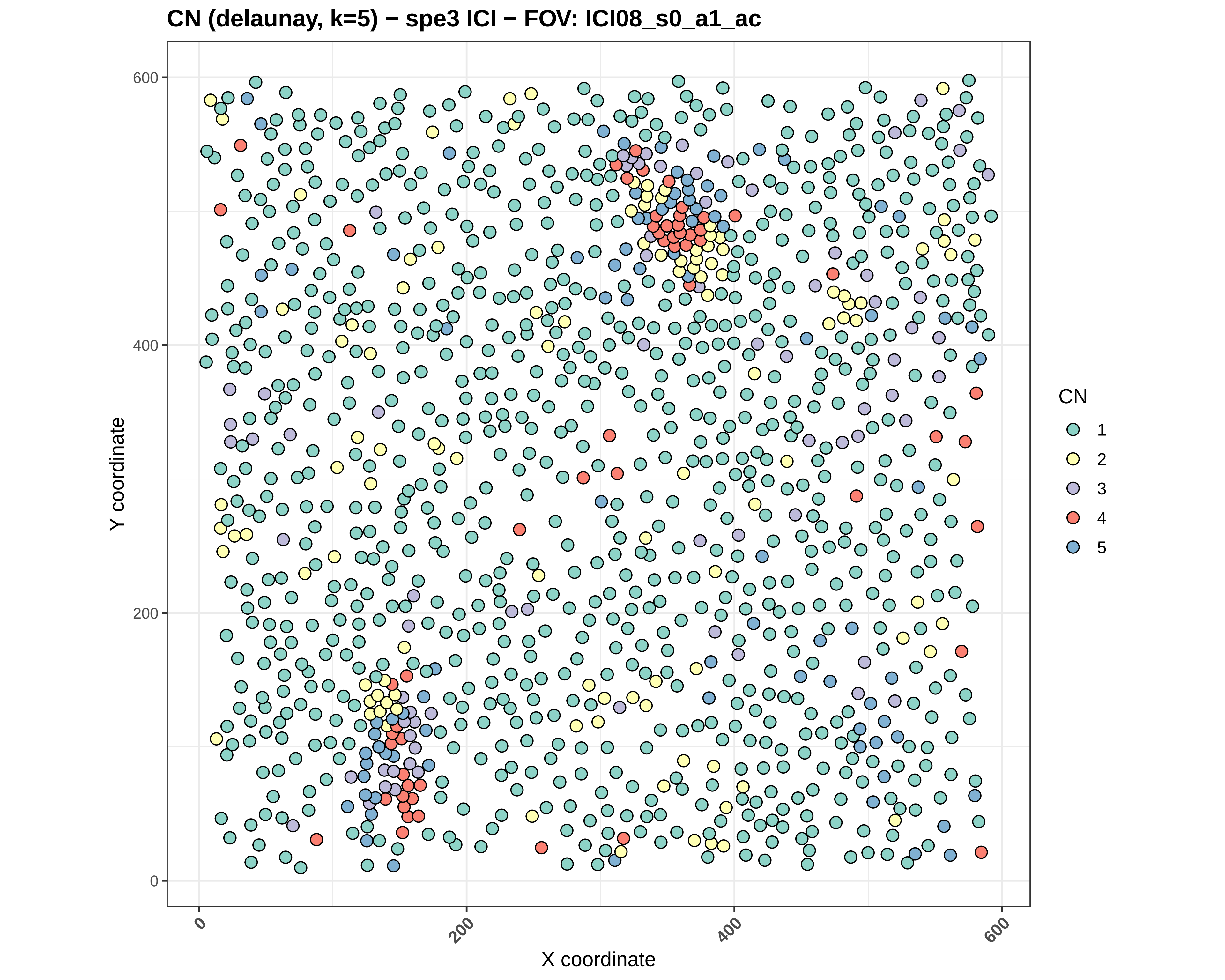
CN (delaunay, k=5) − spe3 ICI − FOV: ICI08_s0_a1_ac
Y coordinate
X coordinate
0
200
400
600
0
200
400
600
CN
1
2
3
4
5

#### CN (delaunay, k=5) − spe3 ICI − FOV: ICI08_s0_a2_ac

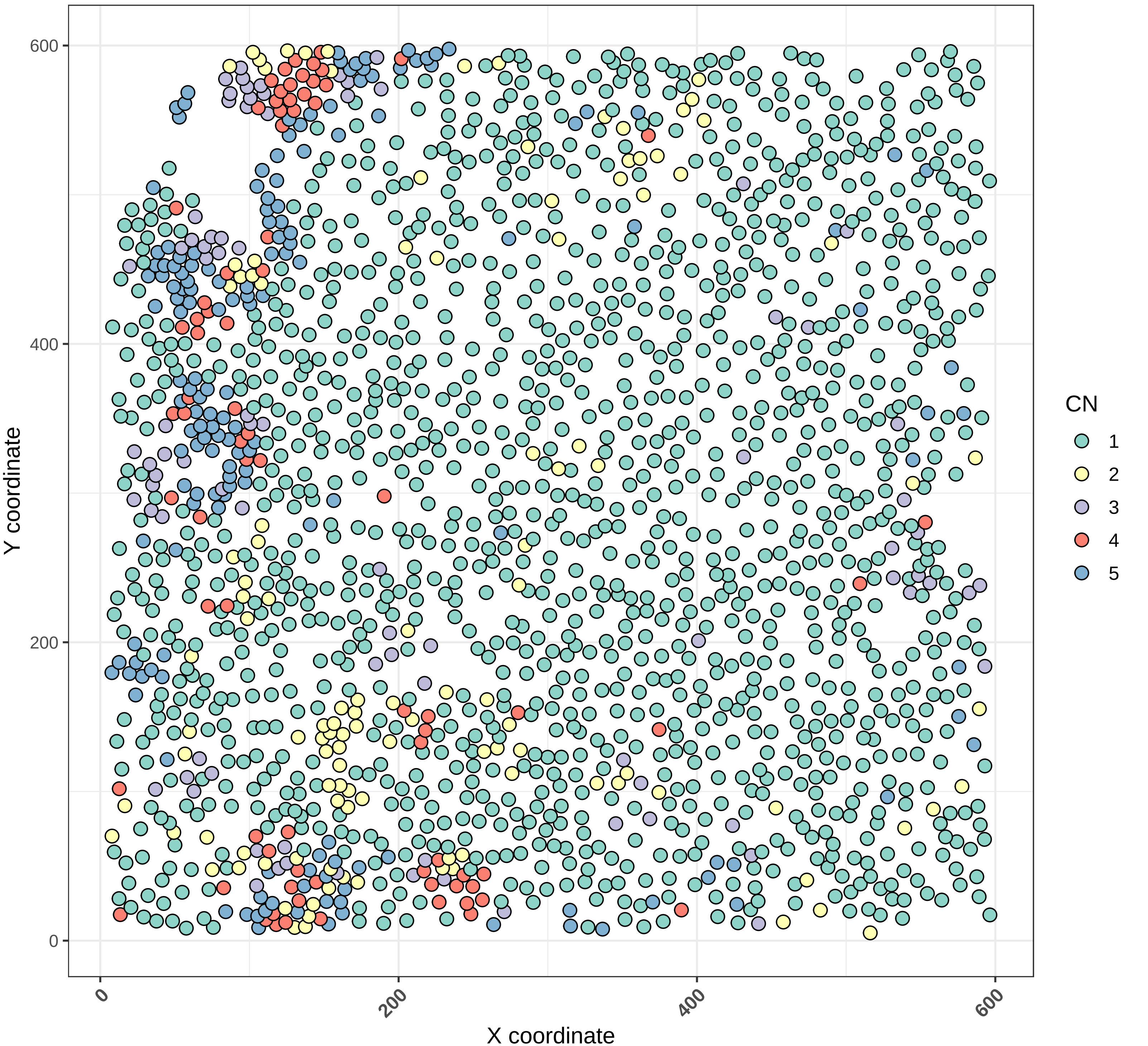

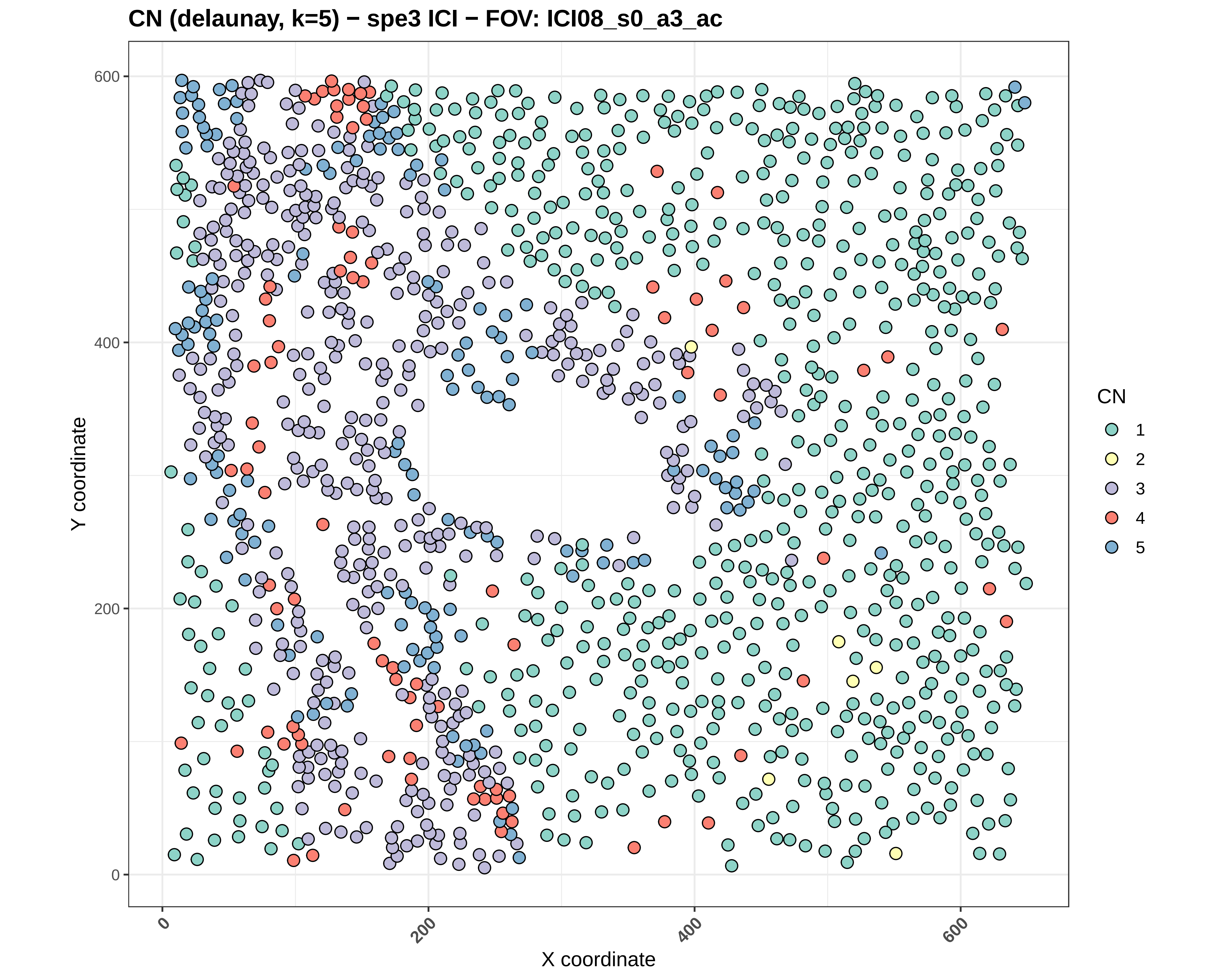

CN (delaunay, k=5) − spe3 ICI − FOV: ICI08_s0_a3_ac
Y coordinate
X coordinate
0
200
400
600
CN
1
2
3
4
5

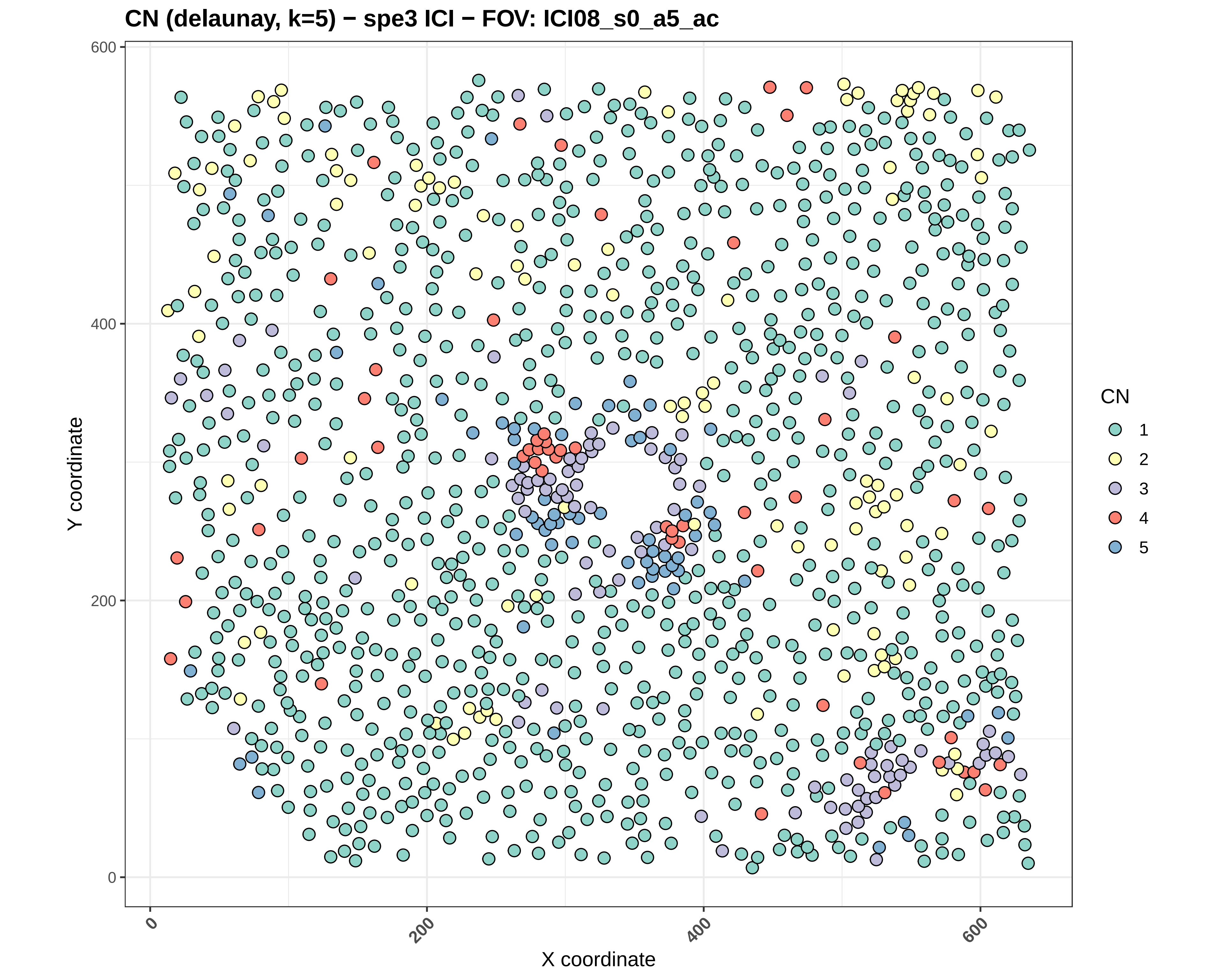

CN (delaunay, k=5) − spe3 ICI − FOV: ICI08_s0_a5_ac
Y coordinate
X coordinate
0
200
400
600
0
200
400
600
CN
1
2
3
4
5

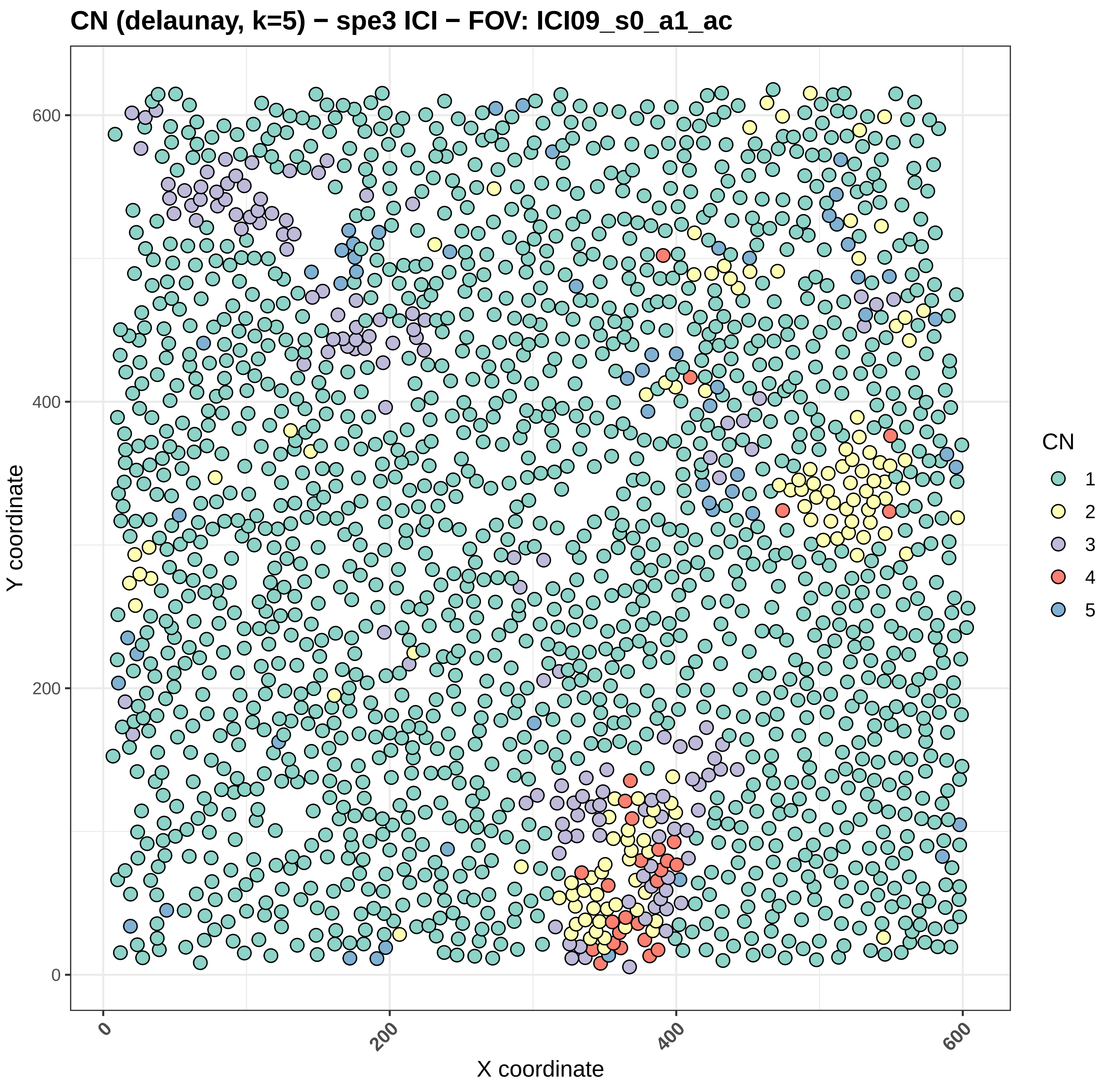

CN (delaunay, k=5) − spe3 ICI − FOV: ICI09_s0_a1_ac
Y coordinate
X coordinate
0
200
400
600
CN
1
2
3
4
5

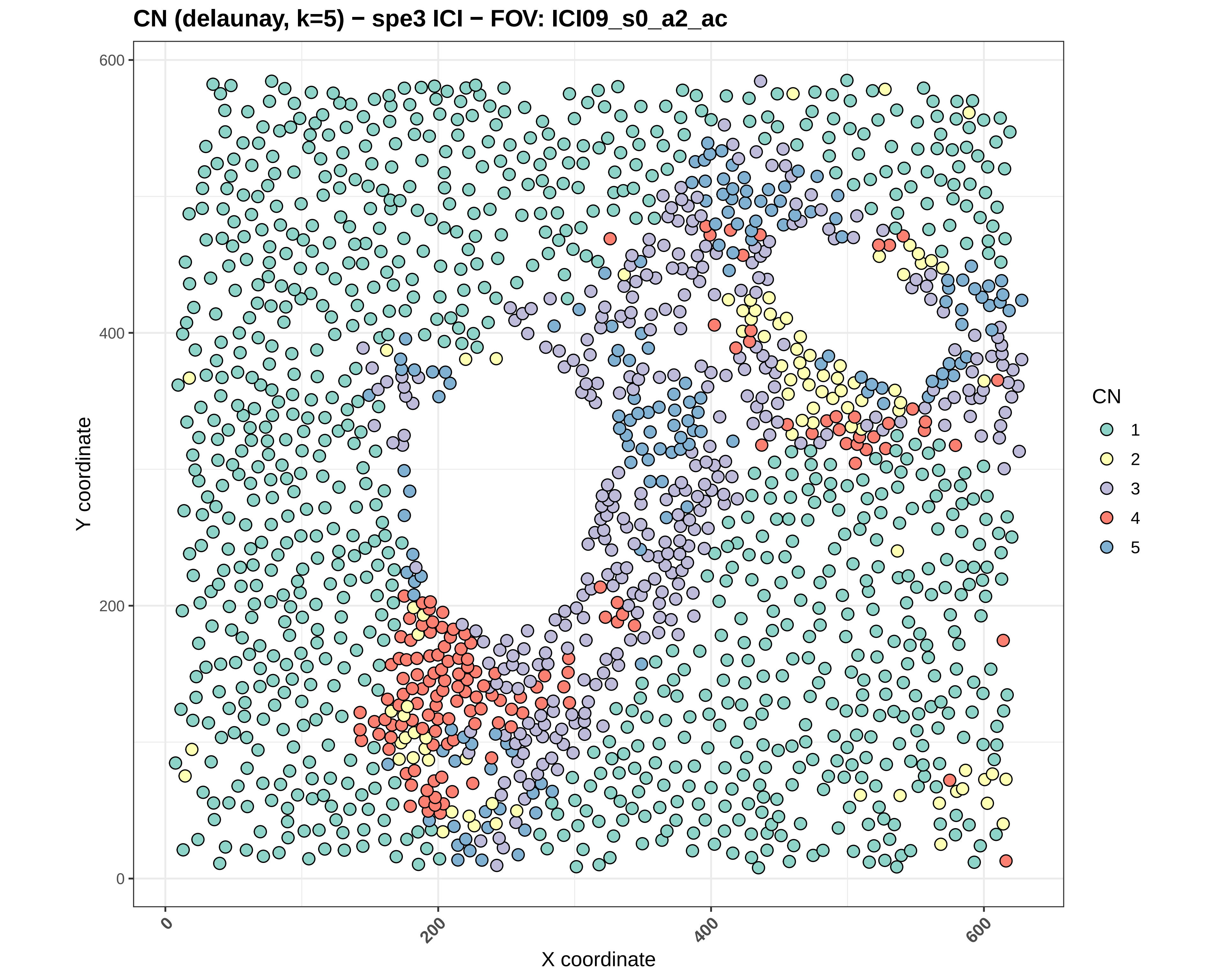
CN (delaunay, k=5) − spe3 ICI − FOV: ICI09_s0_a2_ac
Y coordinate
X coordinate
0
200
400
600
CN
1
2
3
4
5

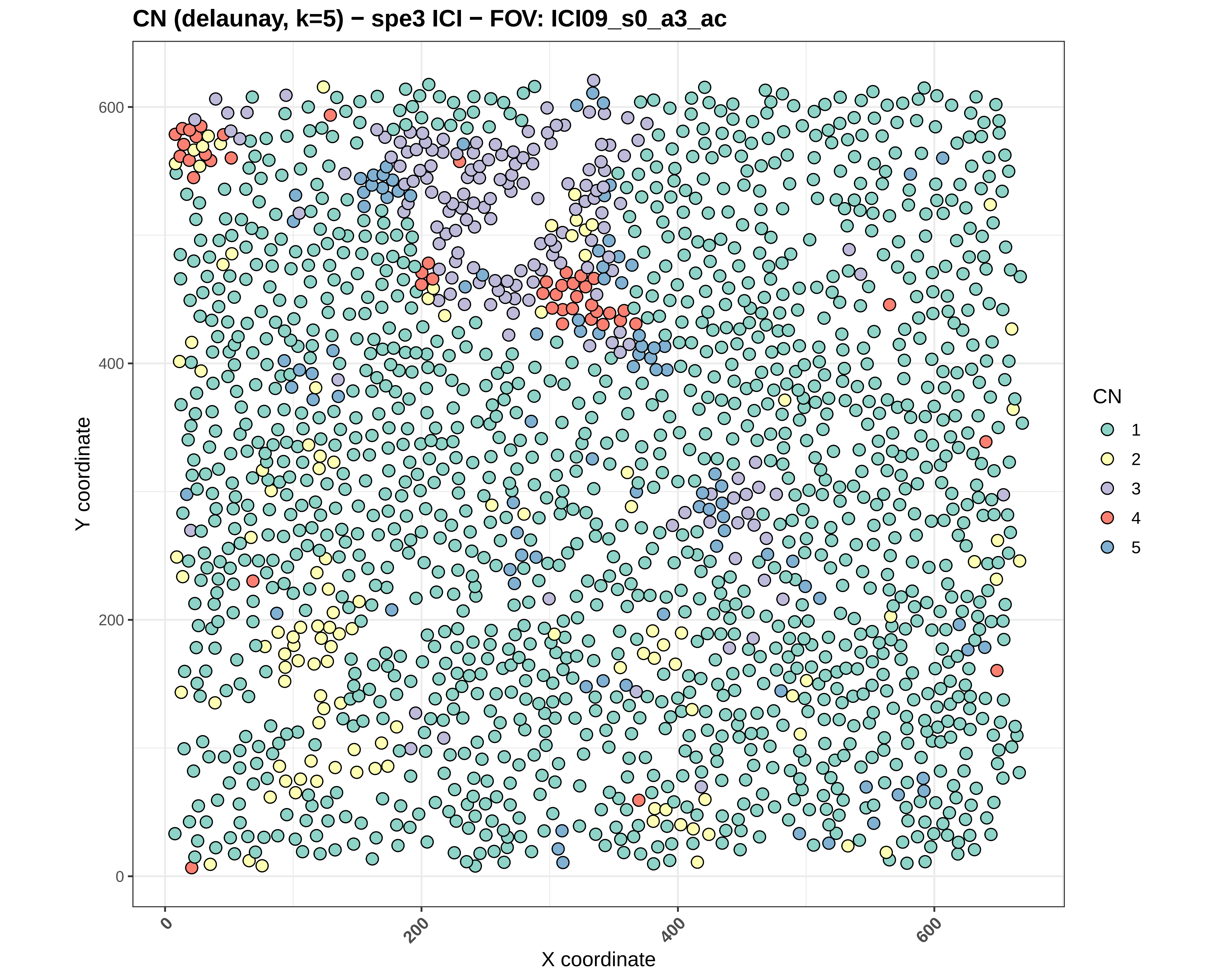

CN (delaunay, k=5) − spe3 ICI − FOV: ICI09_s0_a3_ac
Y coordinate
X coordinate
0
200
400
600
0
200
400
600
CN
1
2
3
4
5

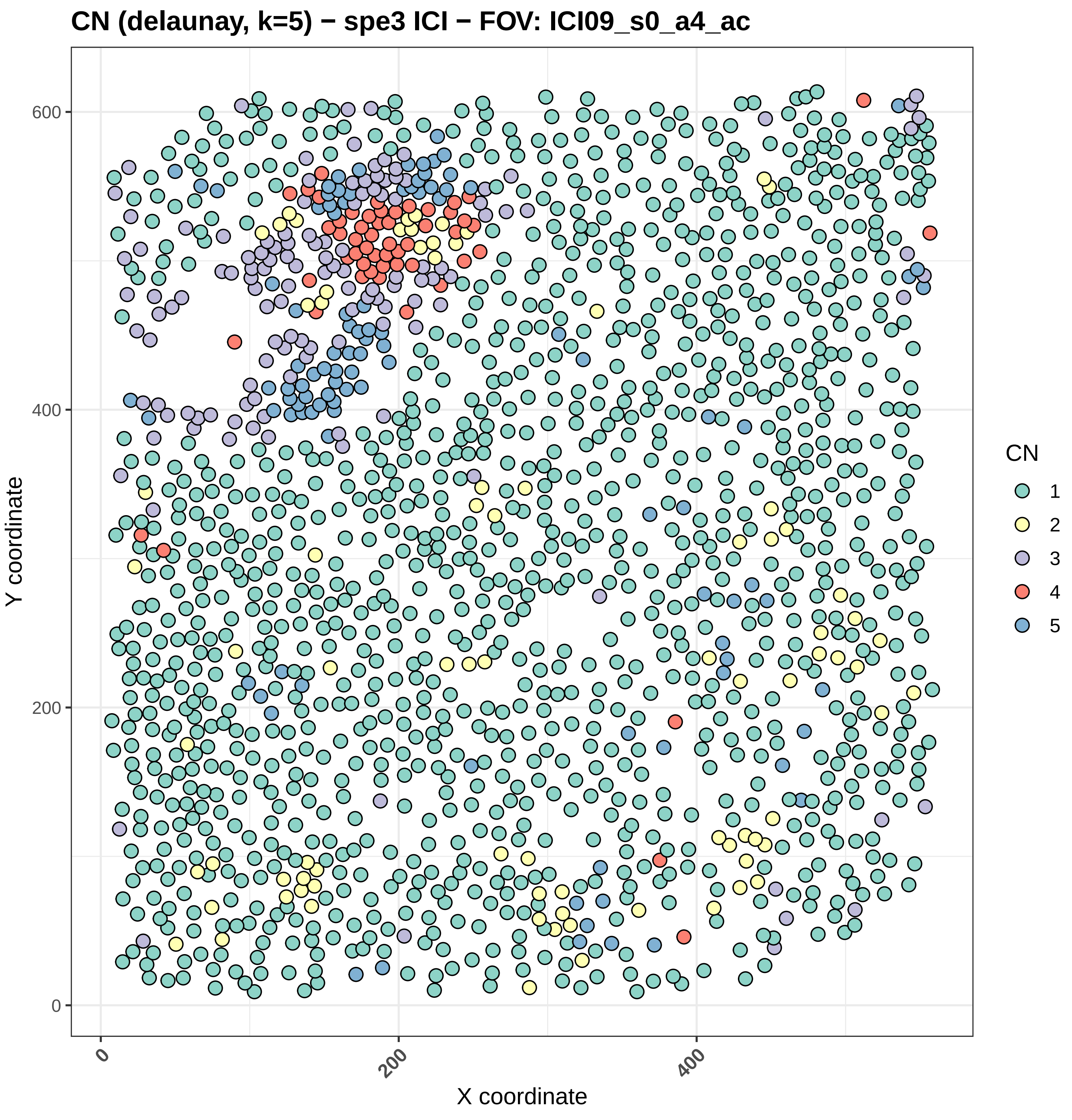
CN (delaunay, k=5) − spe3 ICI − FOV: ICI09_s0_a4_ac
Y coordinate
0
200
400
600
X coordinate
0
200
400
CN
1
2
3
4
5

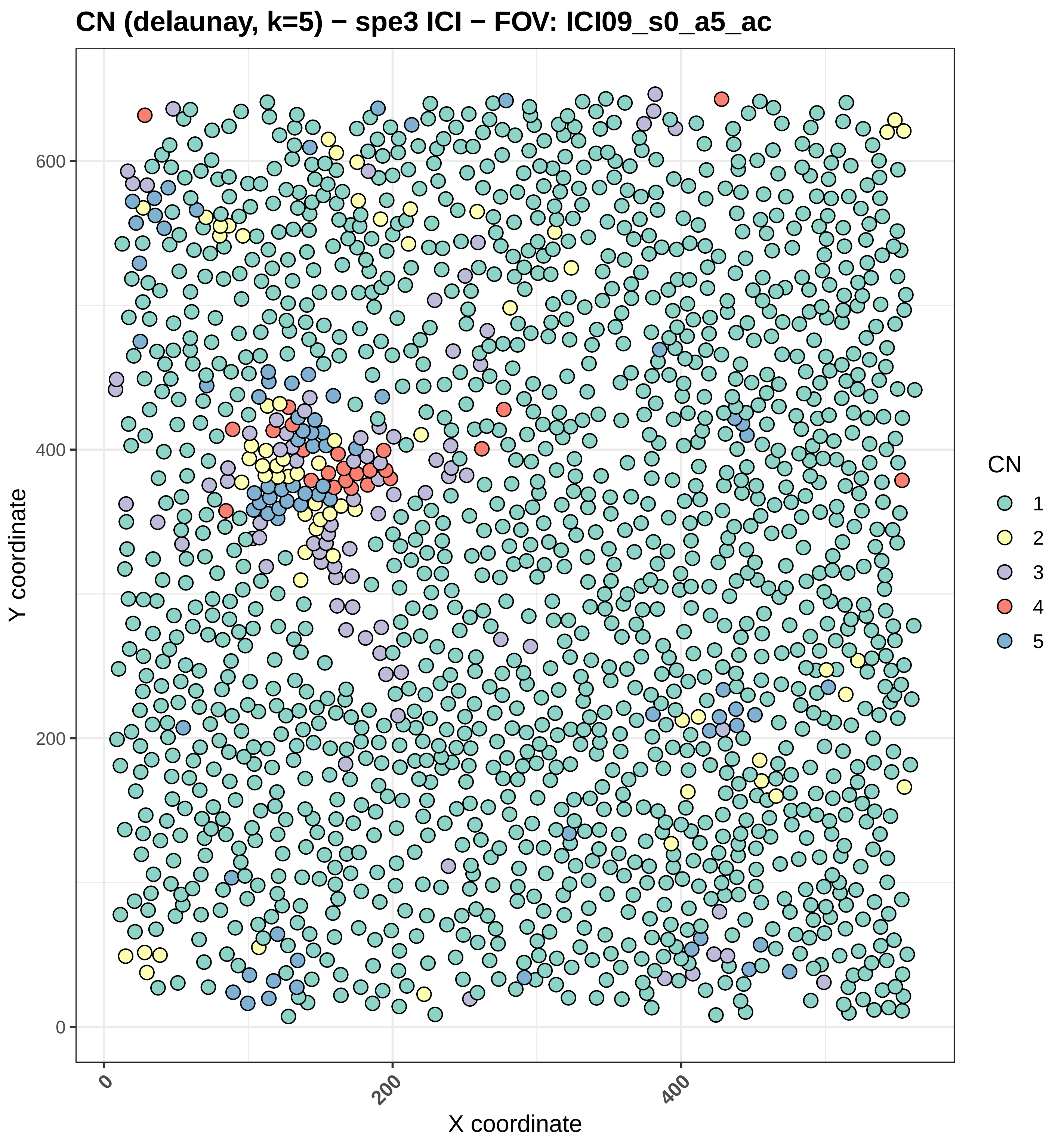

CN (delaunay, k=5) − spe3 ICI − FOV: ICI09_s0_a5_ac
Y coordinate
X coordinate
0
200
400
600
0
200
400
CN
1
2
3
4
5

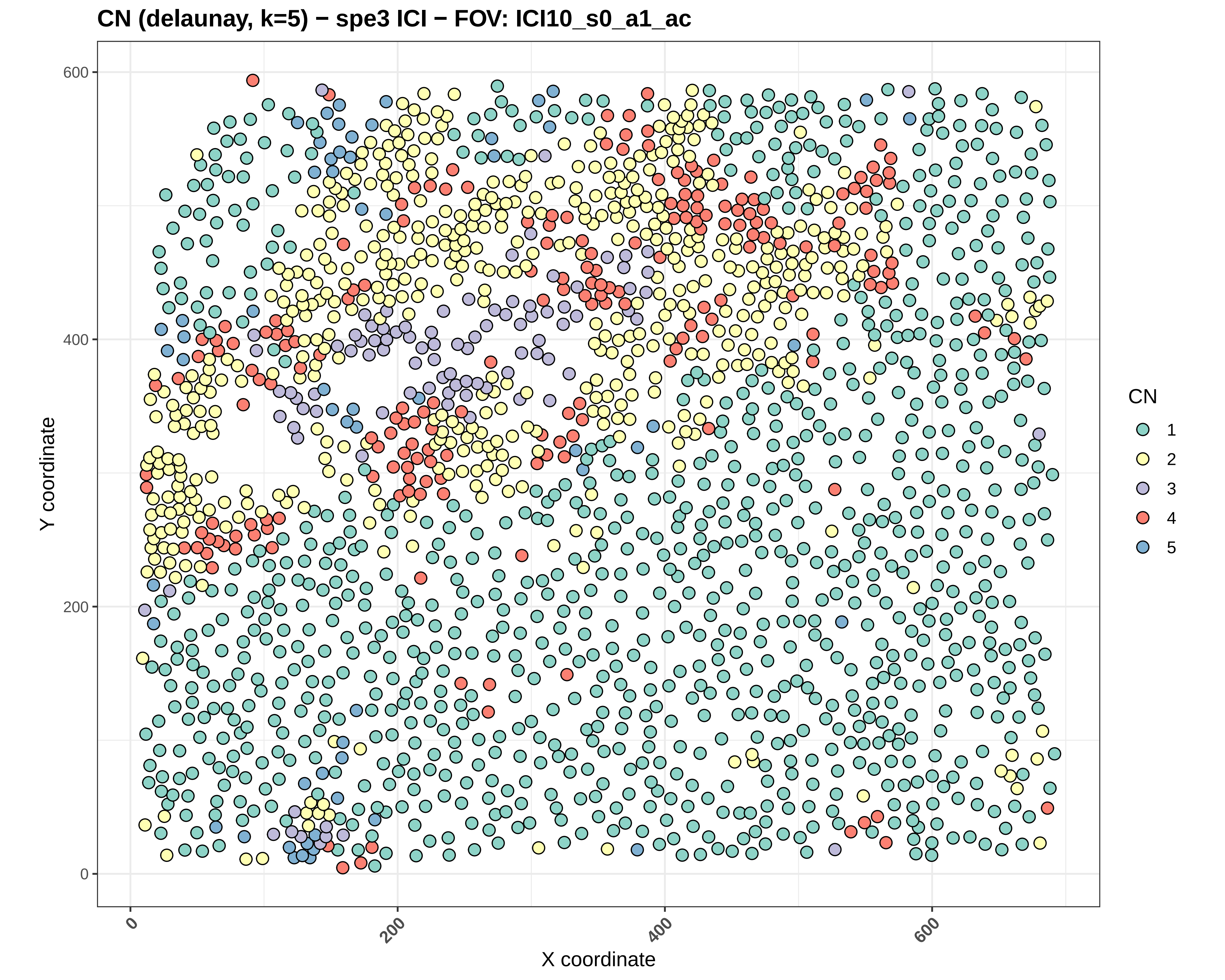
CN (delaunay, k=5) − spe3 ICI − FOV: ICI10_s0_a1_ac
Y coordinate
X coordinate
0
200
400
600
CN
1
2
3
4
5

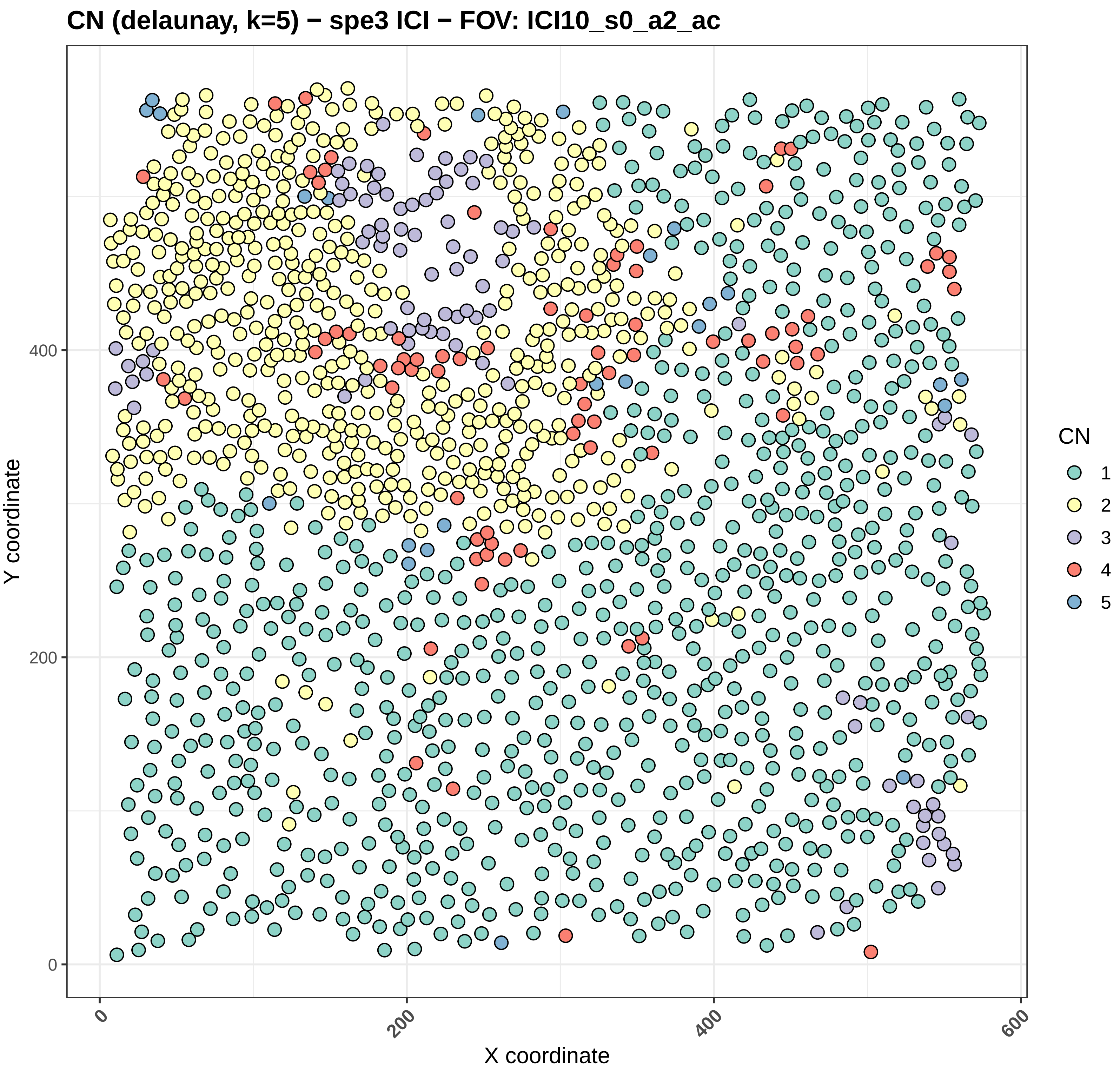
CN (delaunay, k=5) − spe3 ICI − FOV: ICI10_s0_a2_ac
Y coordinate
X coordinate
0
200
400
600
CN
1
2
3
4
5

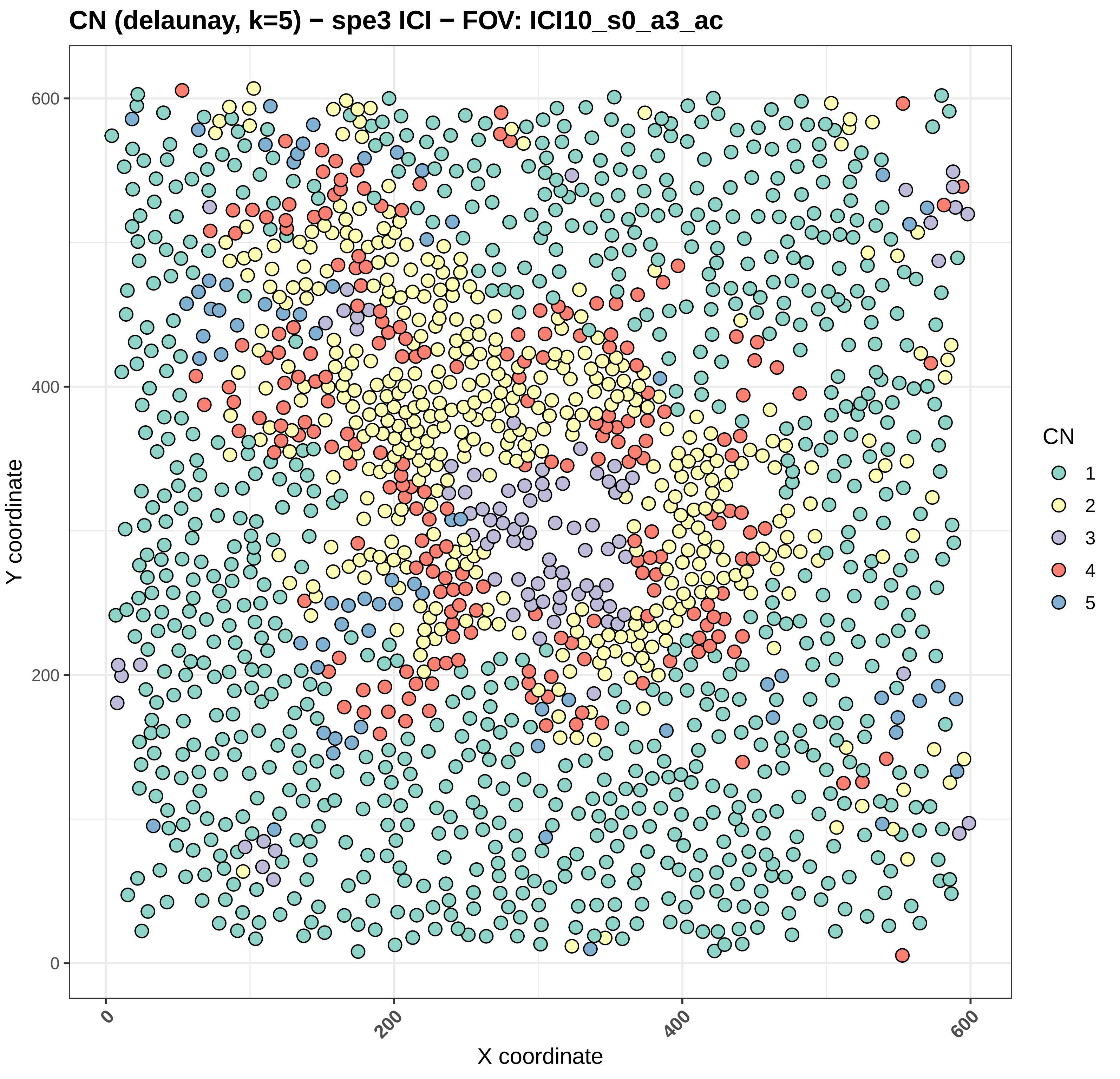
CN (delaunay, k=5) − spe3 ICI − FOV: ICI10_s0_a3_ac
Y coordinate
X coordinate
0
200
400
600
CN
1
2
3
4
5

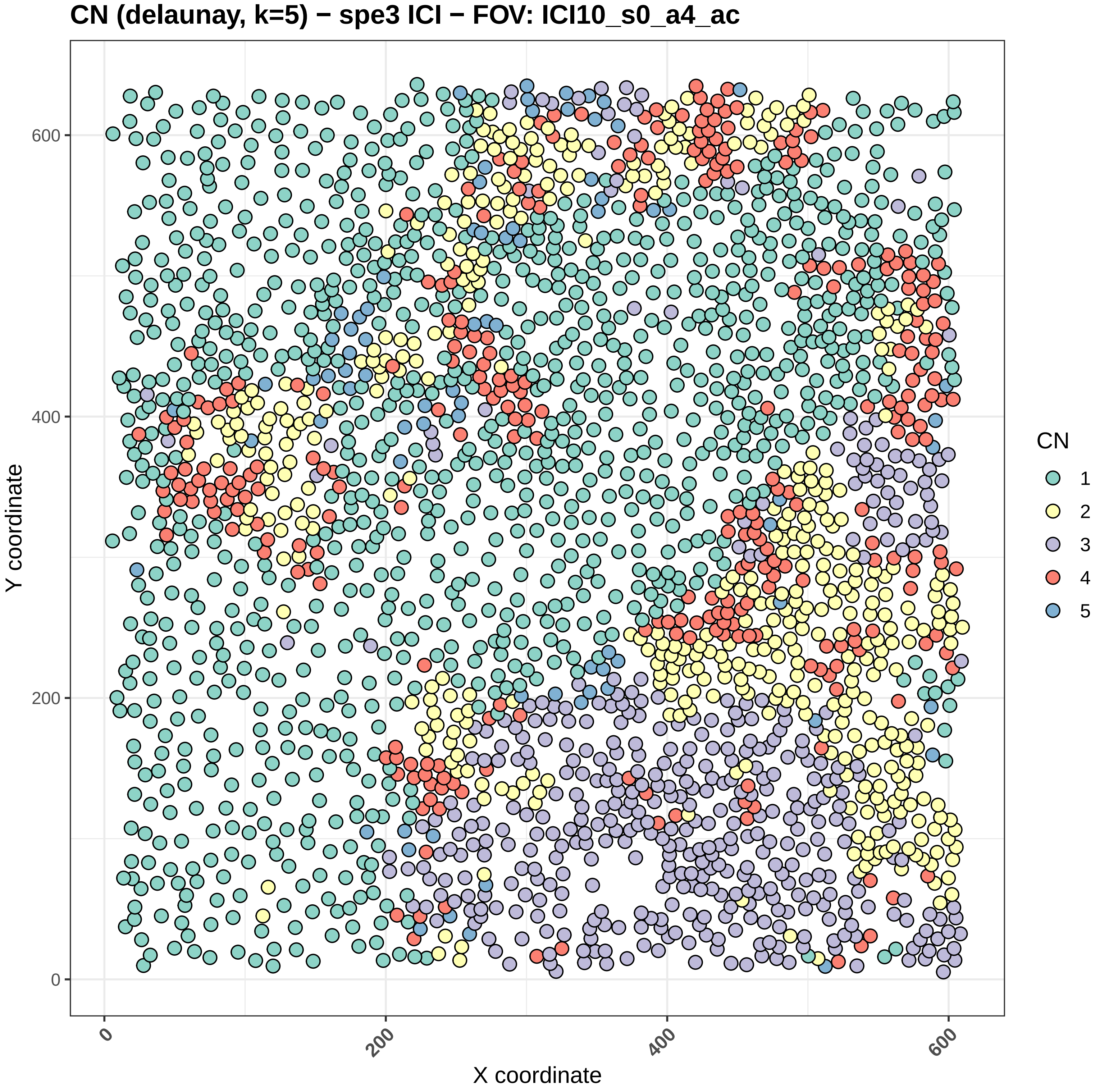
CN (delaunay, k=5) − spe3 ICI − FOV: ICI10_s0_a4_ac
Y coordinate
X coordinate
0
200
400
600
CN
1
2
3
4
5

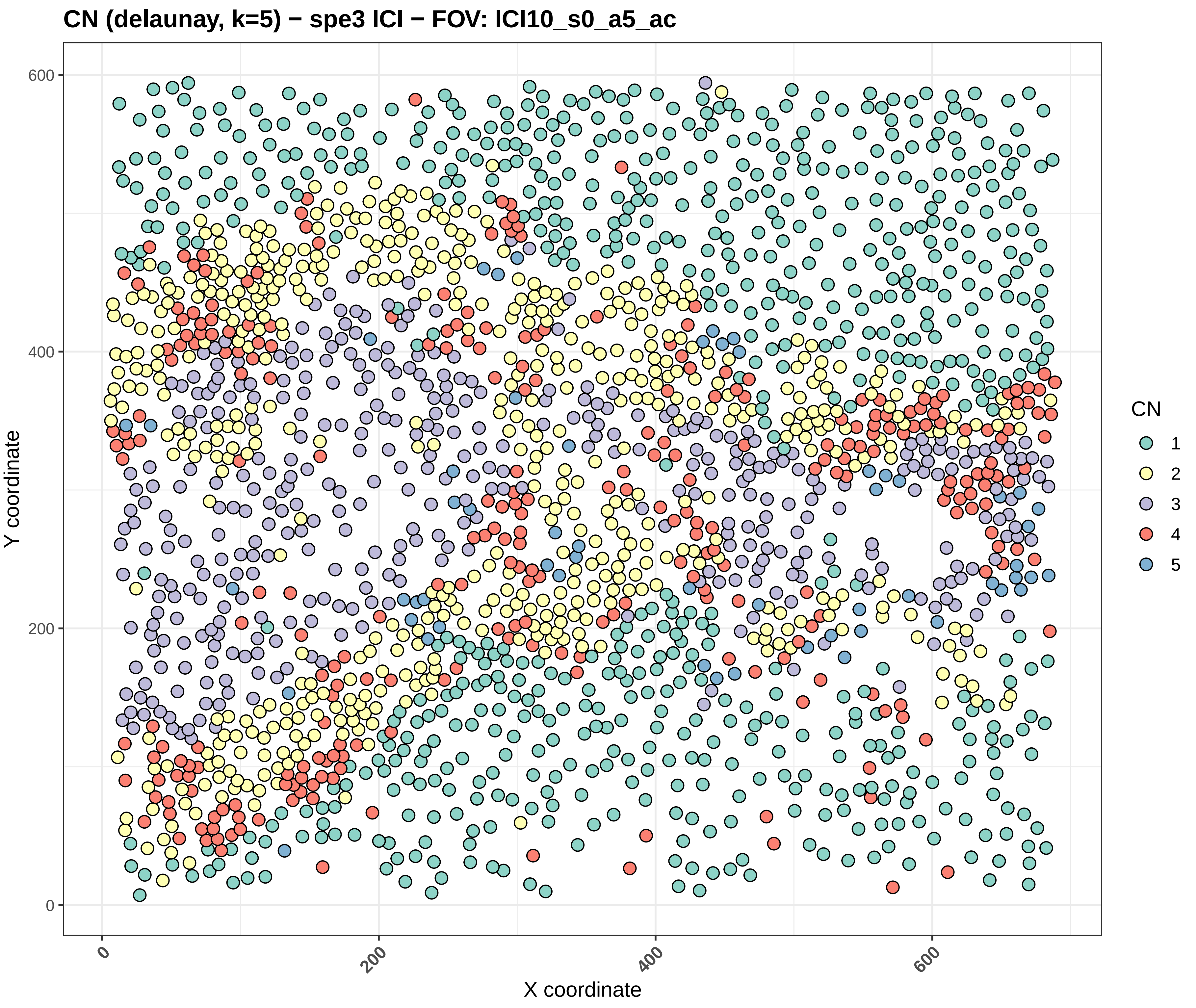
CN (delaunay, k=5) − spe3 ICI − FOV: ICI10_s0_a5_ac
Y coordinate
X coordinate
0
200
400
600
CN
1
2
3
4
5

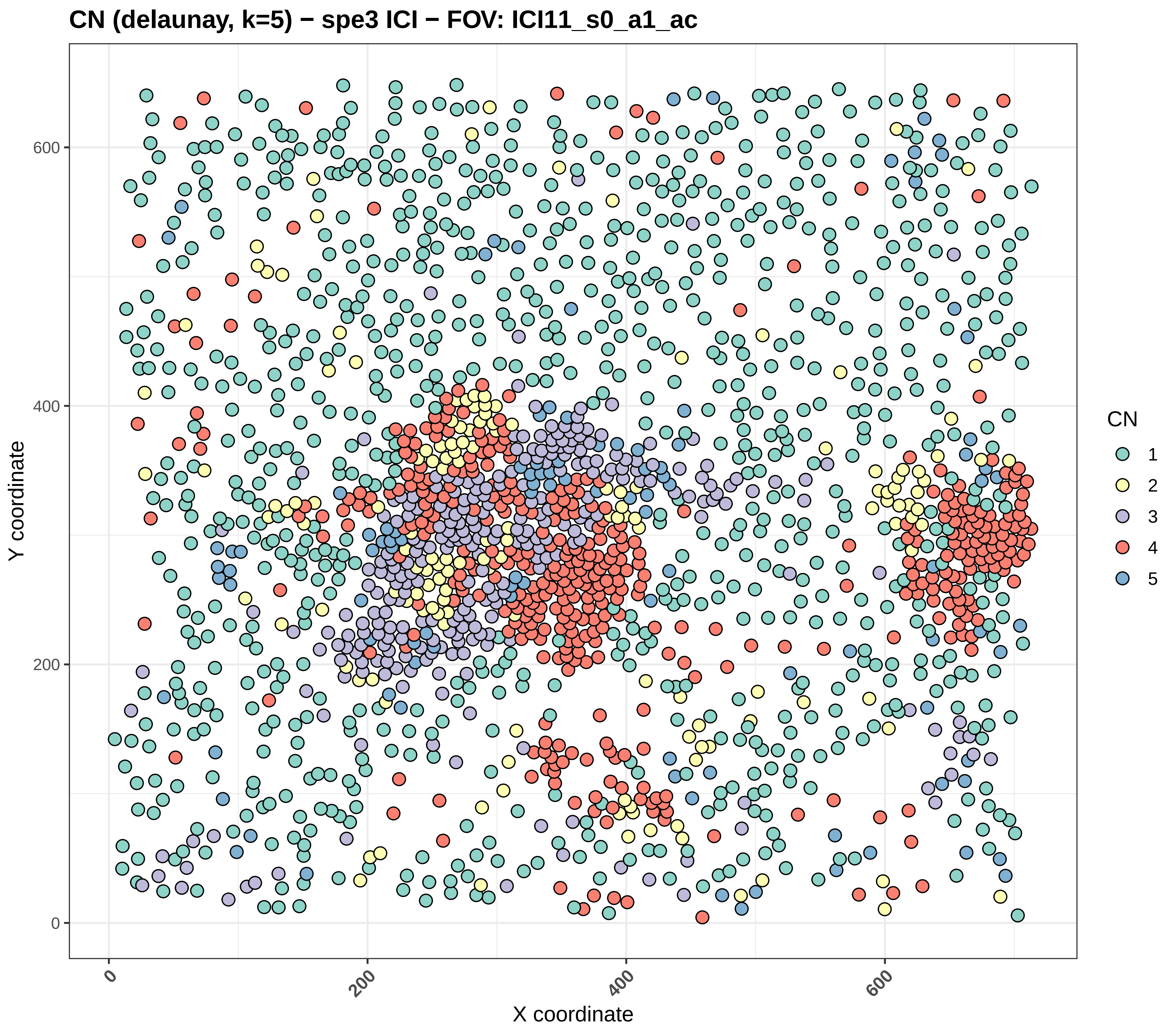

CN (delaunay, k=5) − spe3 ICI − FOV: ICI11_s0_a1_ac
Y coordinate
X coordinate
0
200
400
600
0
200
400
600
CN
1
2
3
4
5

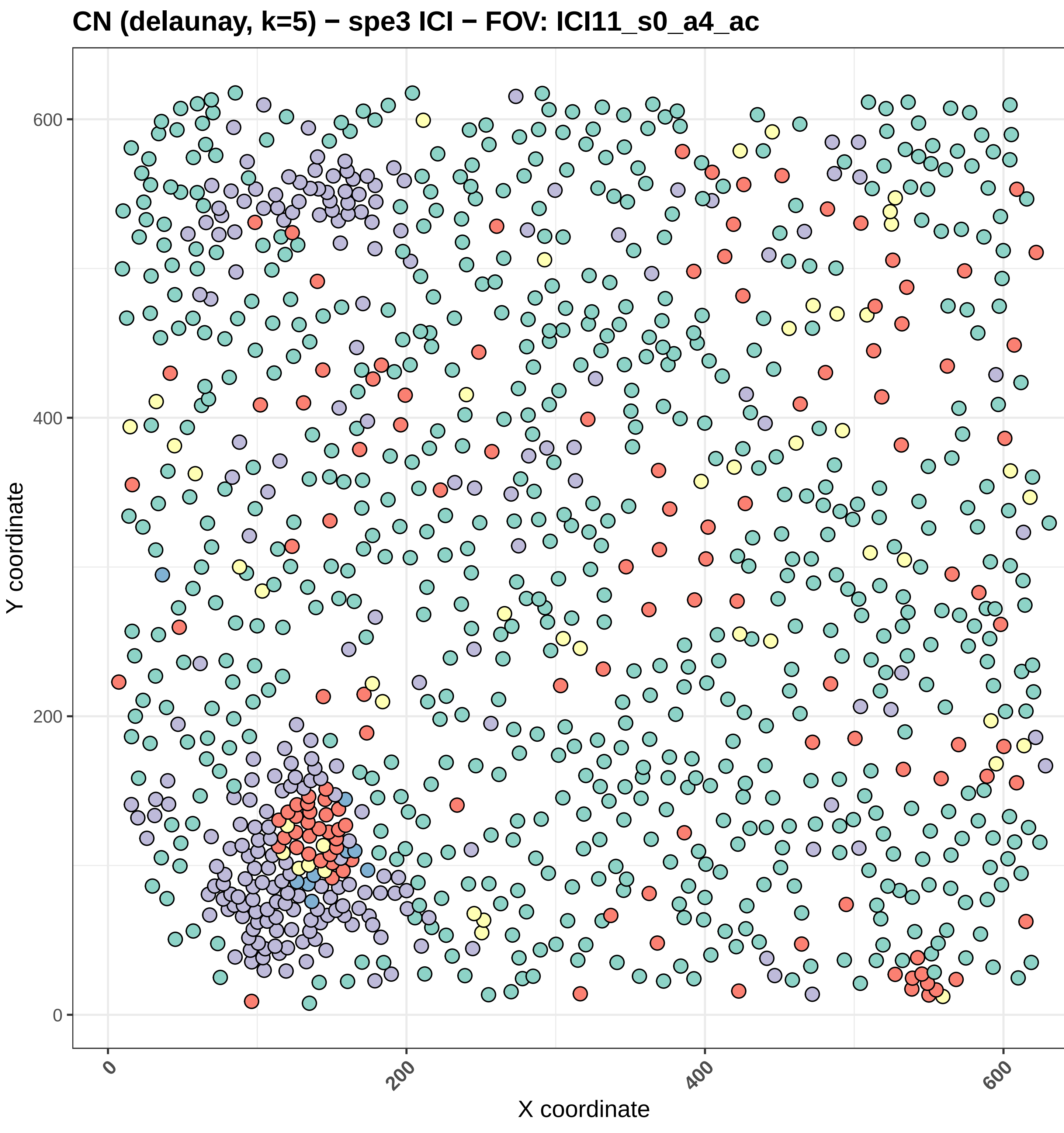
CN (delaunay, k=5) − spe3 ICI − FOV: ICI11_s0_a4_ac
Y coordinate
X coordinate
0
200
400
600
CN
1
2
3
4
5

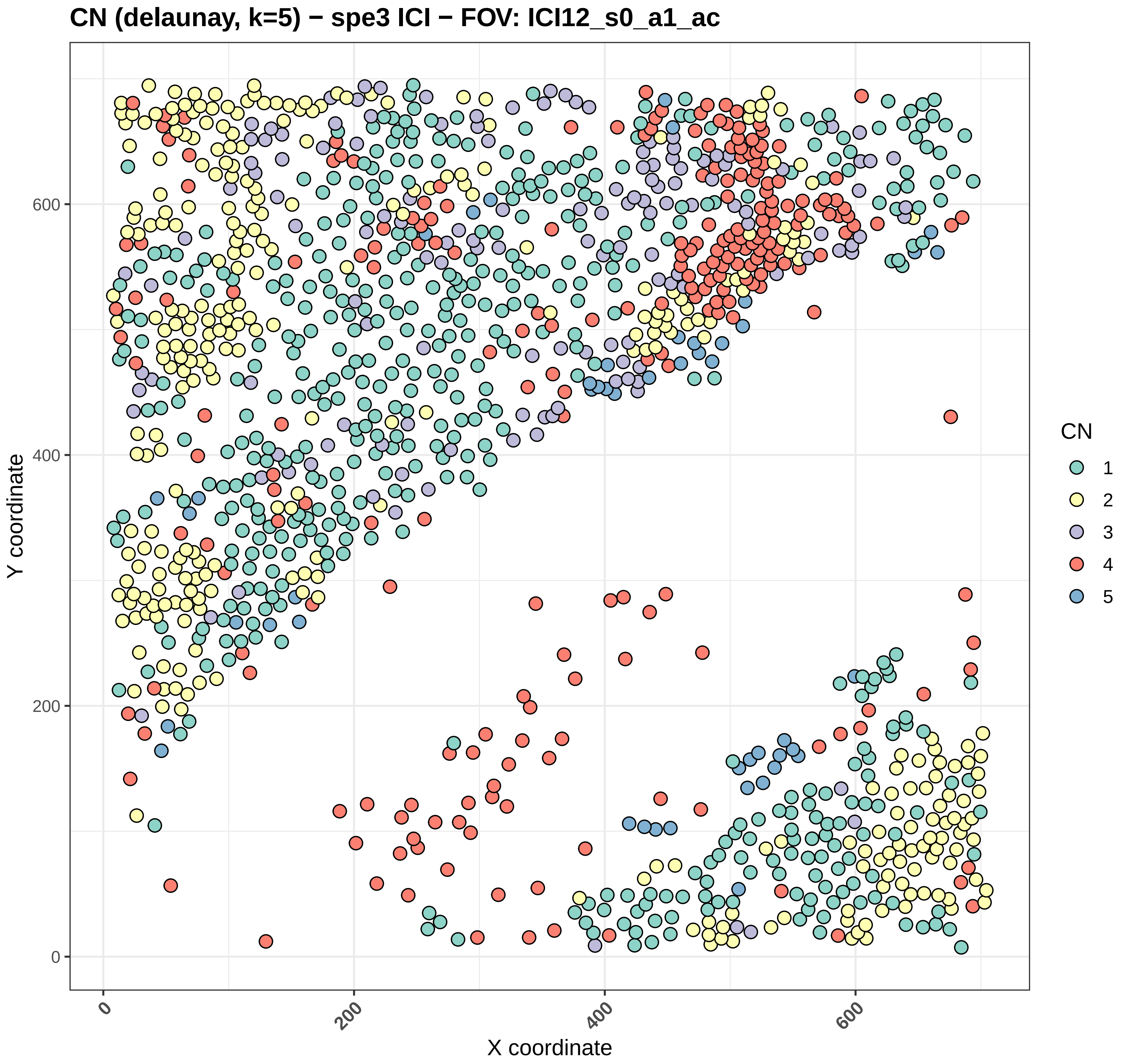
CN (delaunay, k=5) − spe3 ICI − FOV: ICI12_s0_a1_ac
Y coordinate
0
200
400
600
X coordinate
0
200
400
600
CN
1
2
3
4
5

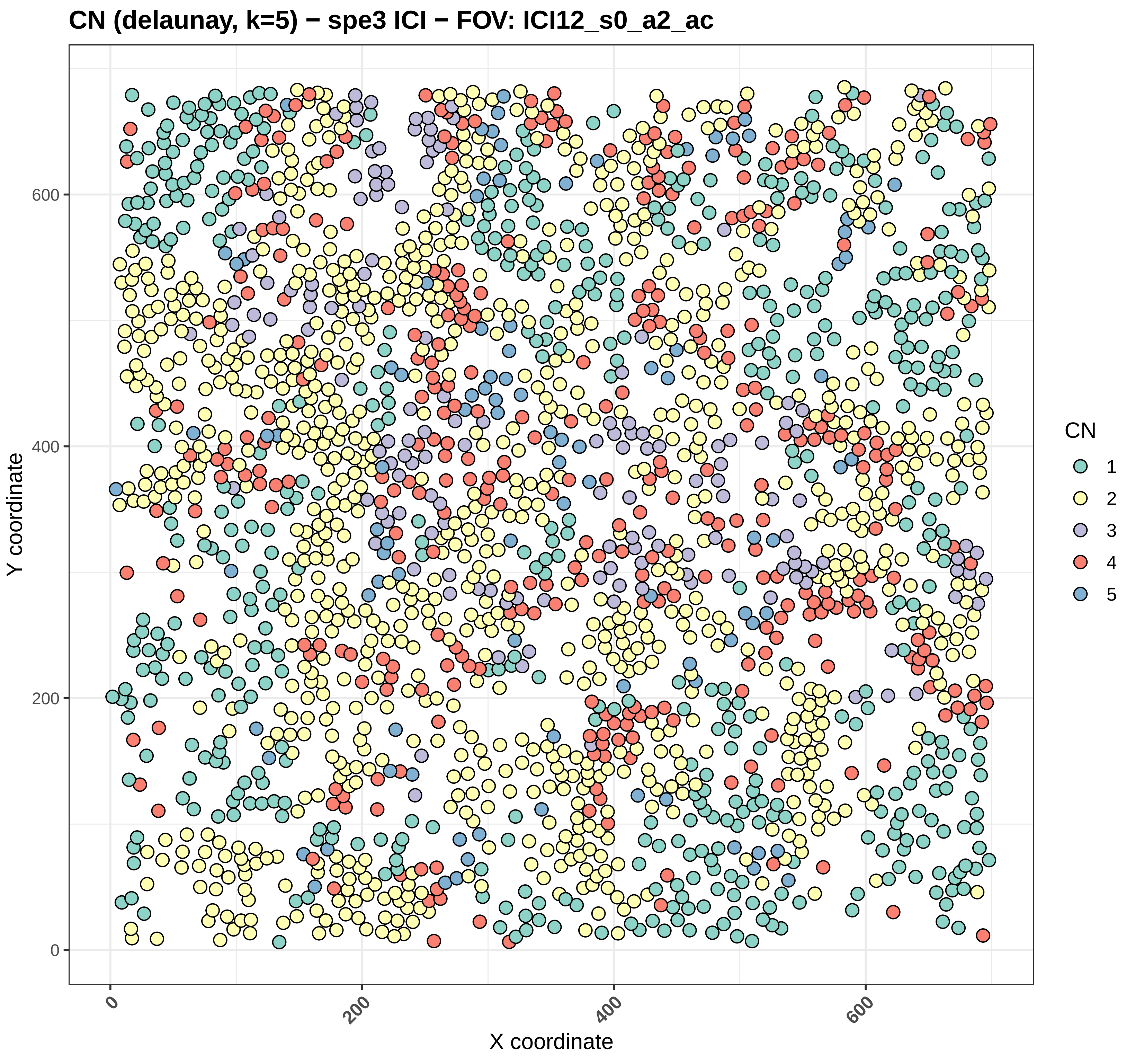

CN (delaunay, k=5) − spe3 ICI − FOV: ICI12_s0_a2_ac
Y coordinate
X coordinate
0
200
400
600
0
200
400
600
CN
1
2
3
4
5

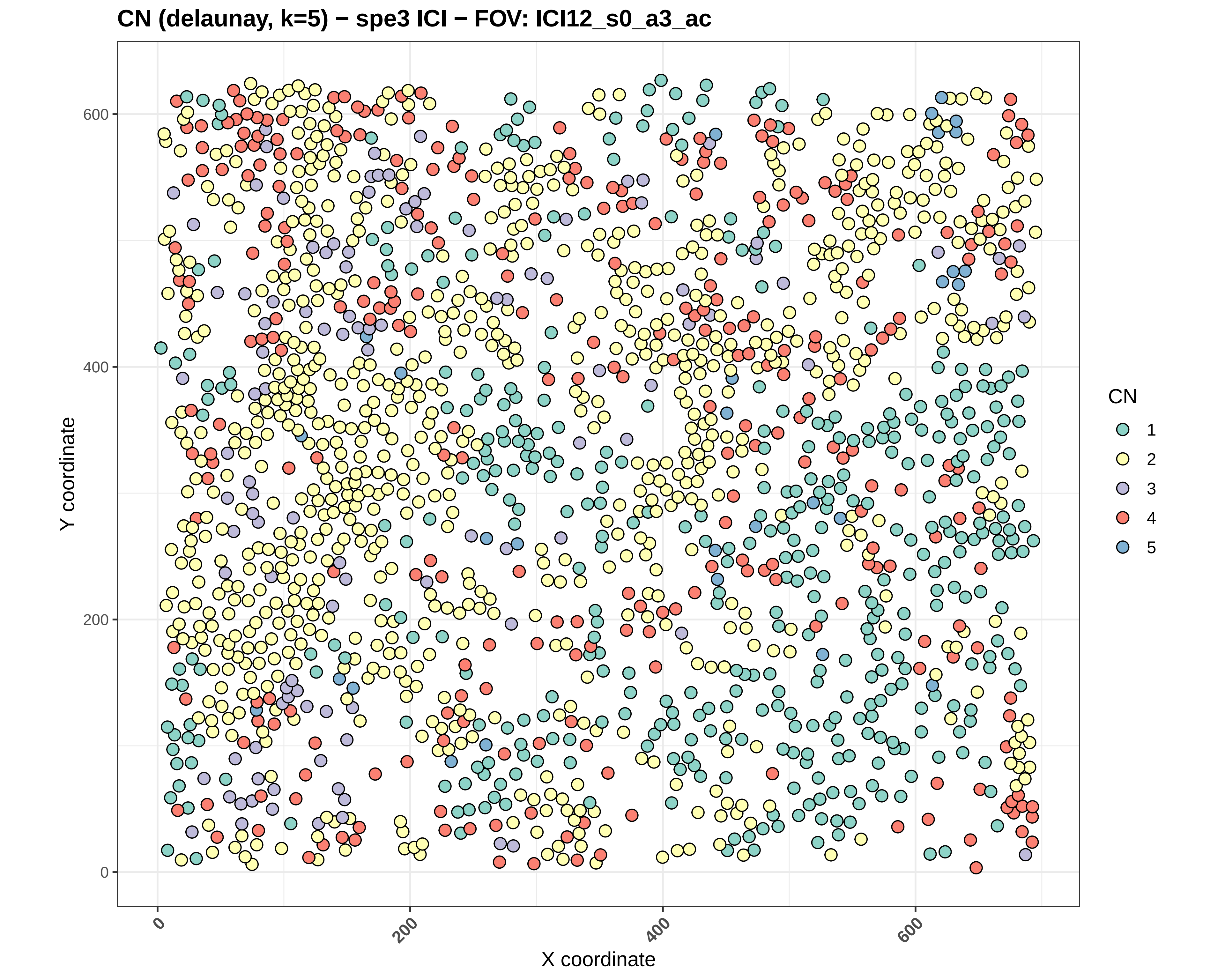
CN (delaunay, k=5) − spe3 ICI − FOV: ICI12_s0_a3_ac
Y coordinate
X coordinate
0
200
400
600
CN
1
2
3
4
5

CN (delaunay, k=5) − spe3 ICI − FOV: ICI12_s0_a5_ac

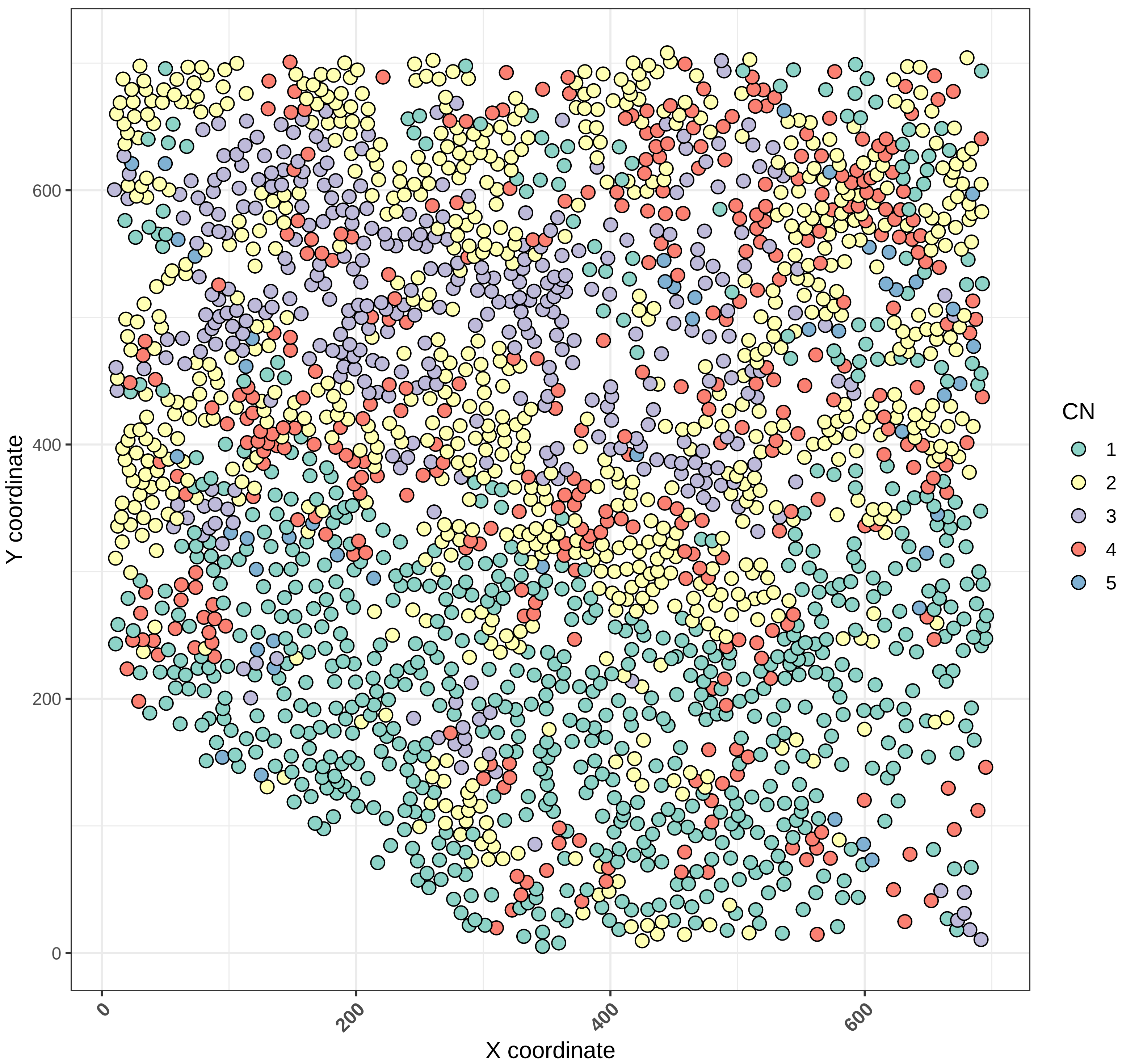

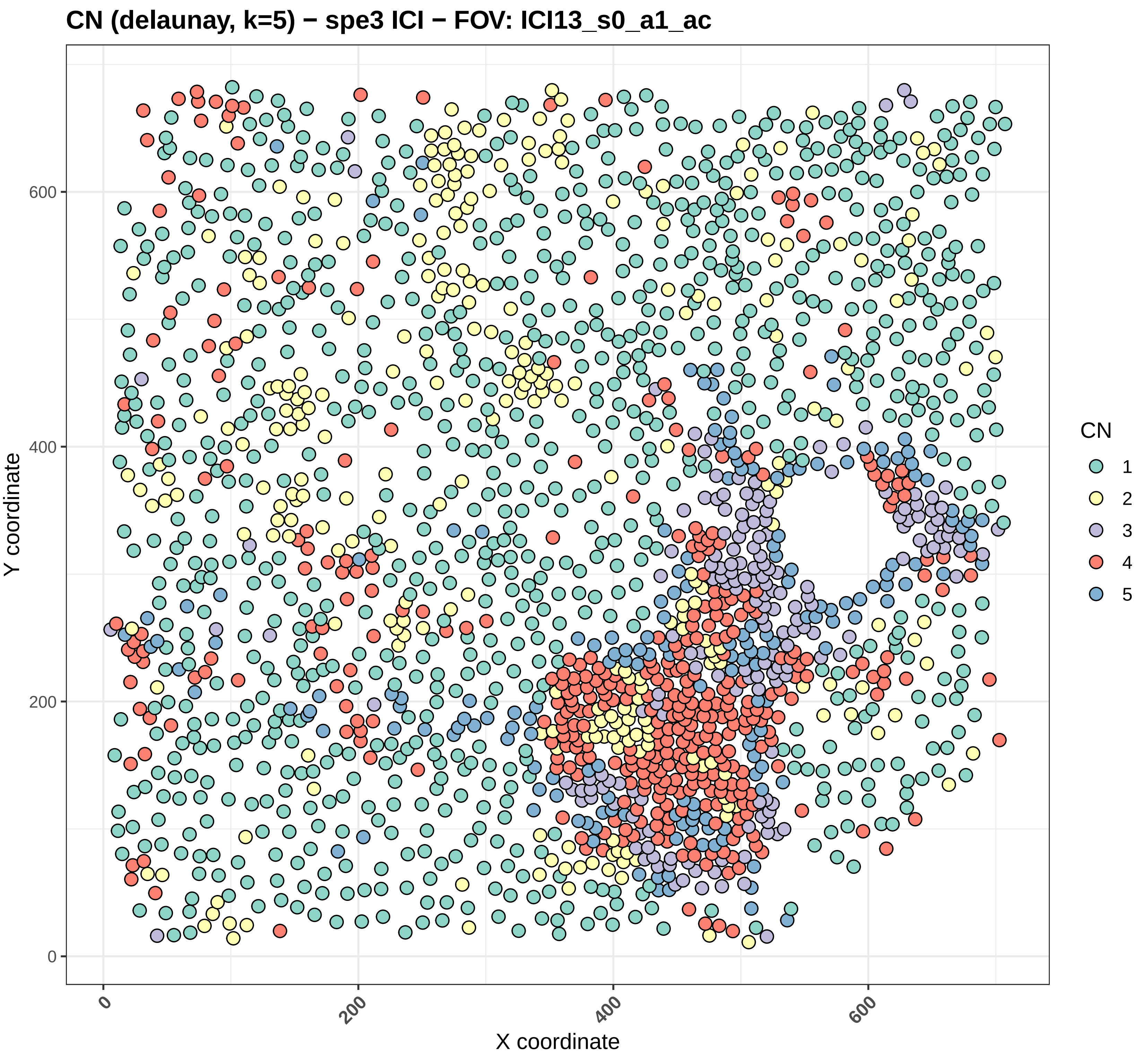

CN (delaunay, k=5) − spe3 ICI − FOV: ICI13_s0_a1_ac
Y coordinate
X coordinate
0
200
400
600
CN
1
2
3
4
5

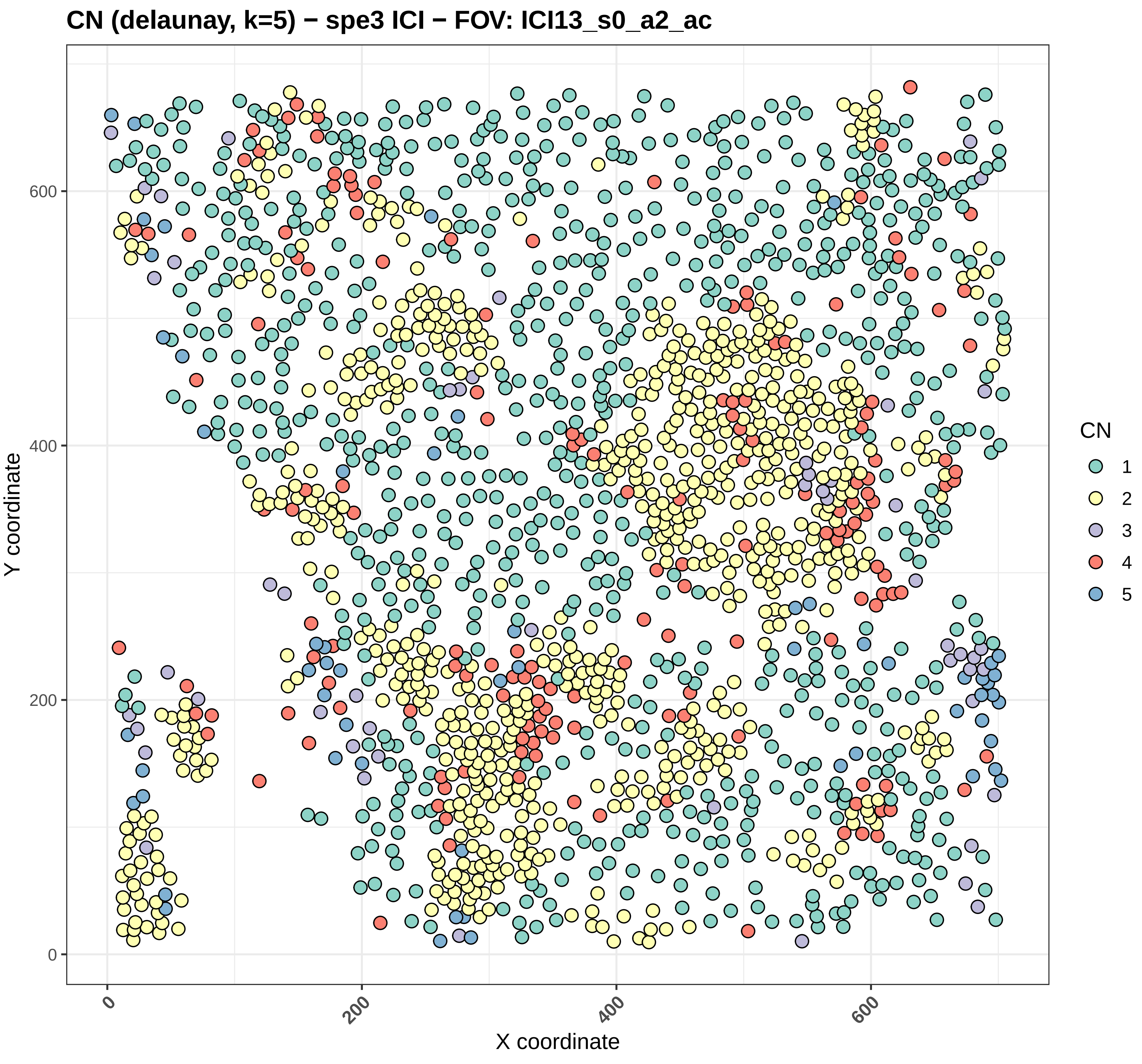

CN (delaunay, k=5) − spe3 ICI − FOV: ICI13_s0_a2_ac
Y coordinate
X coordinate
0
200
400
600
CN
1
2
3
4
5

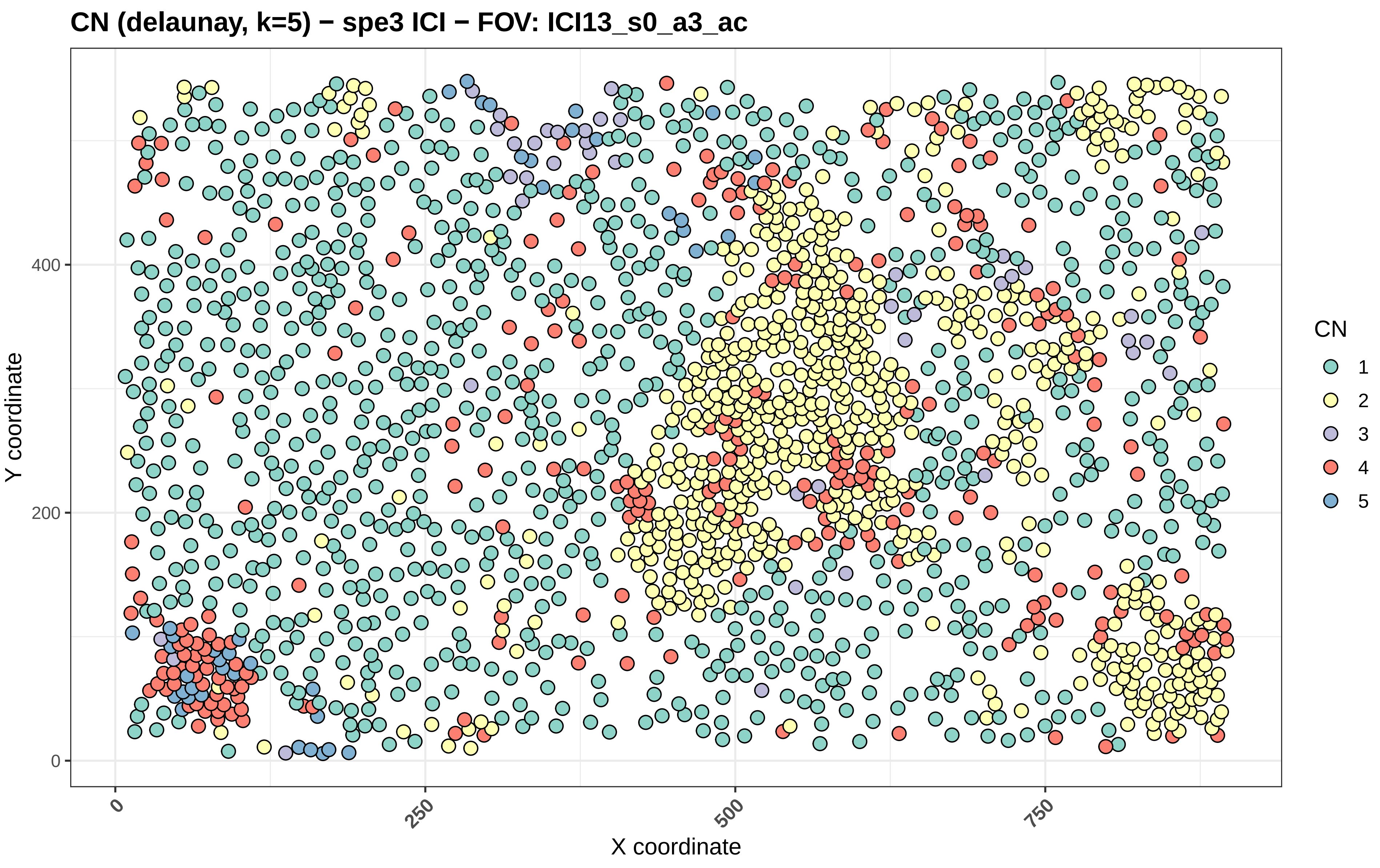
CN (delaunay, k=5) − spe3 ICI − FOV: ICI13_s0_a3_ac
Y coordinate
X coordinate
0
200
400
0
250
500
750
CN
1
2
3
4
5

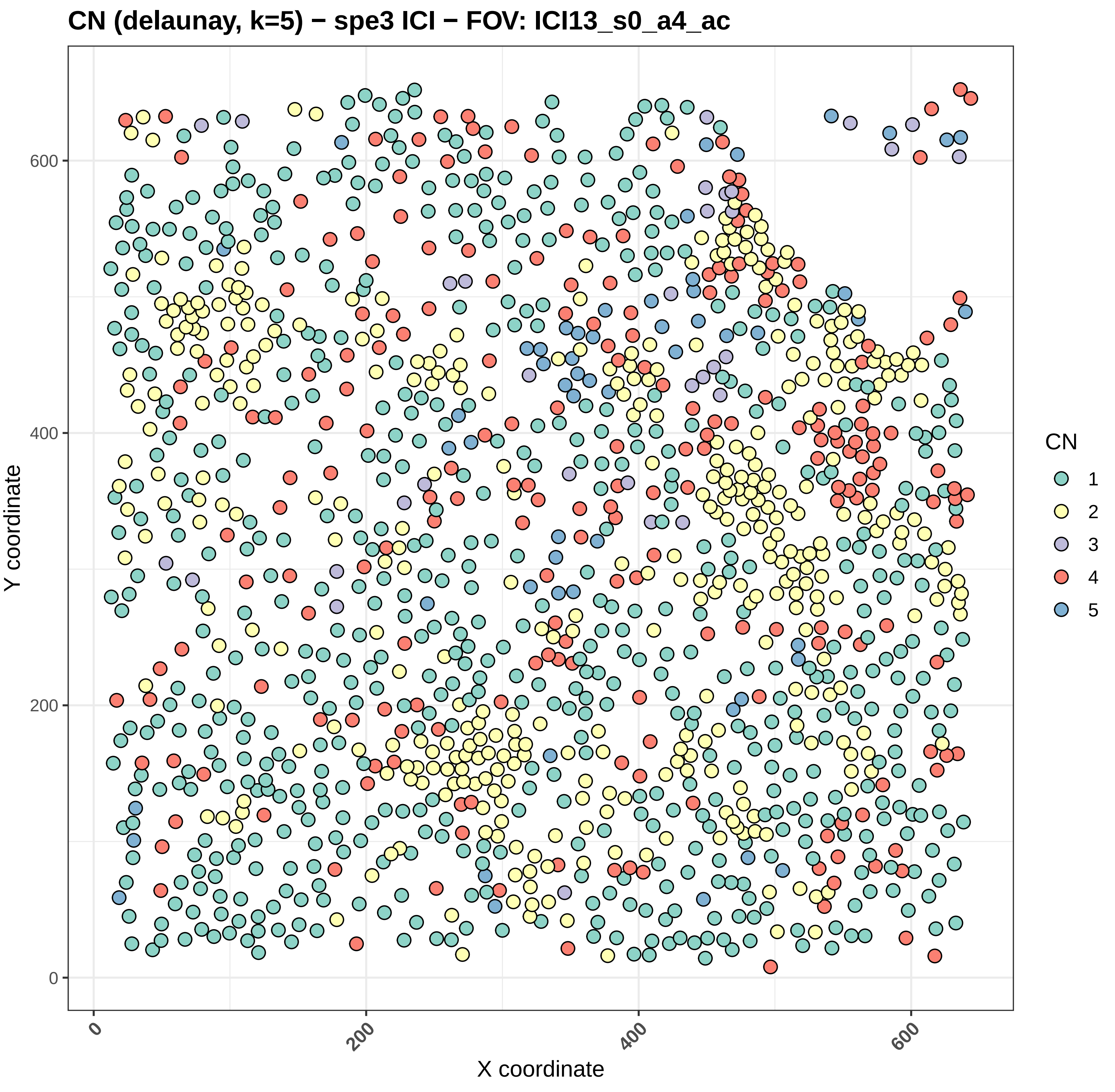
CN (delaunay, k=5) − spe3 ICI − FOV: ICI13_s0_a4_ac
Y coordinate
X coordinate
0
200
400
600
CN
1
2
3
4
5

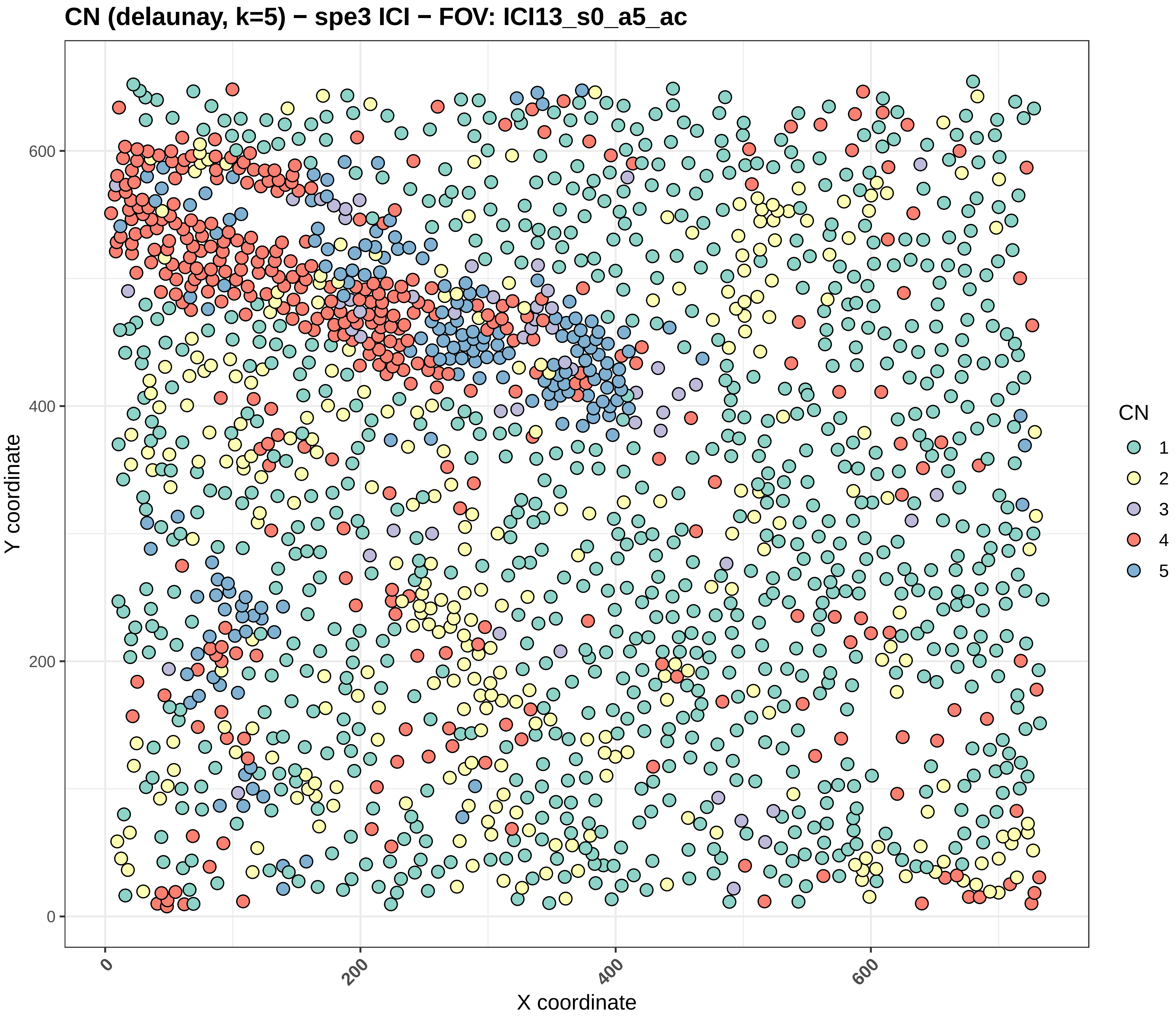

CN (delaunay, k=5) − spe3 ICI − FOV: ICI13_s0_a5_ac
Y coordinate
X coordinate
0
200
400
600
CN
1
2
3
4
5

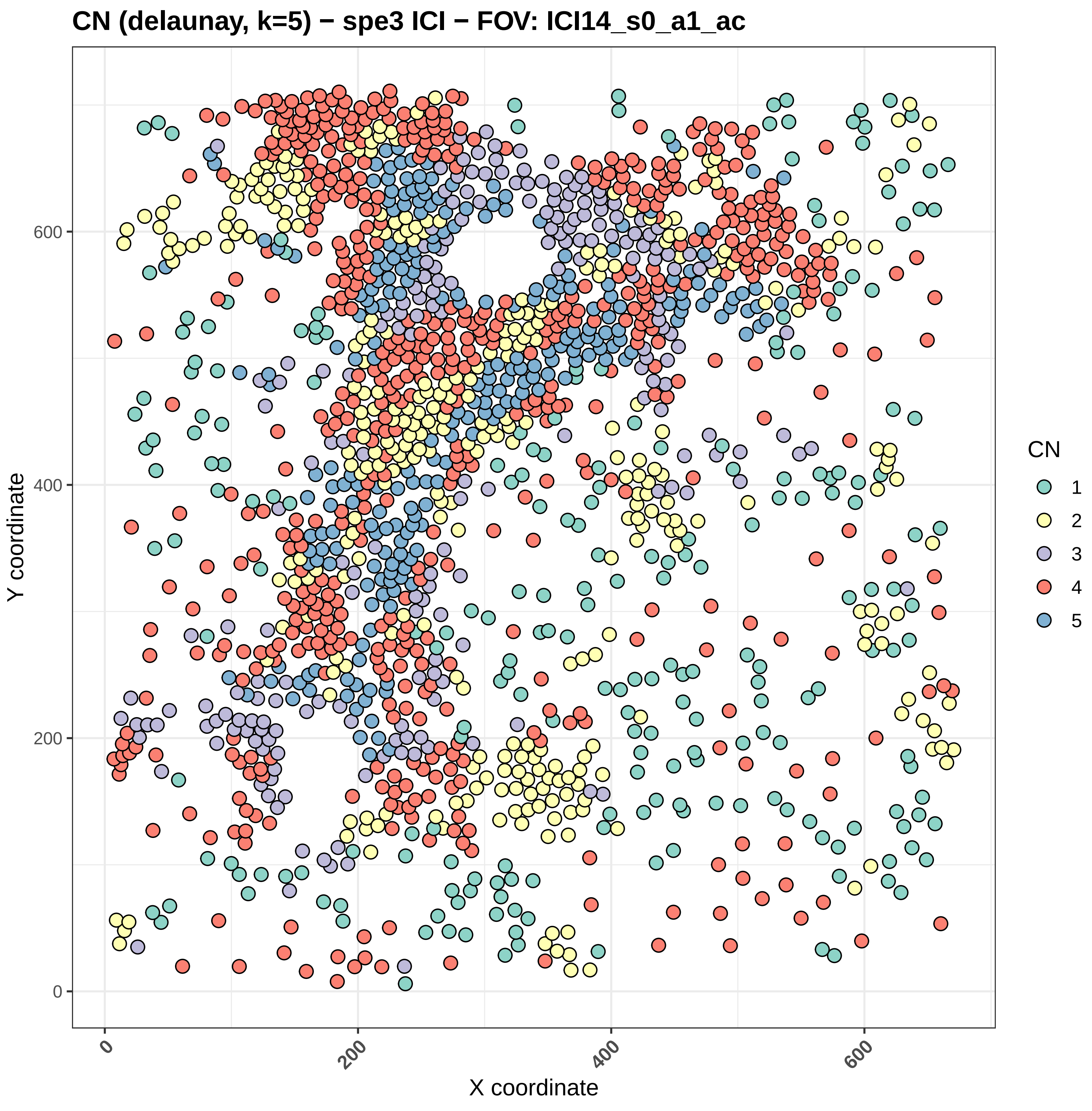
CN (delaunay, k=5) − spe3 ICI − FOV: ICI14_s0_a1_ac
Y coordinate
X coordinate
0
200
400
600
CN
1
2
3
4
5

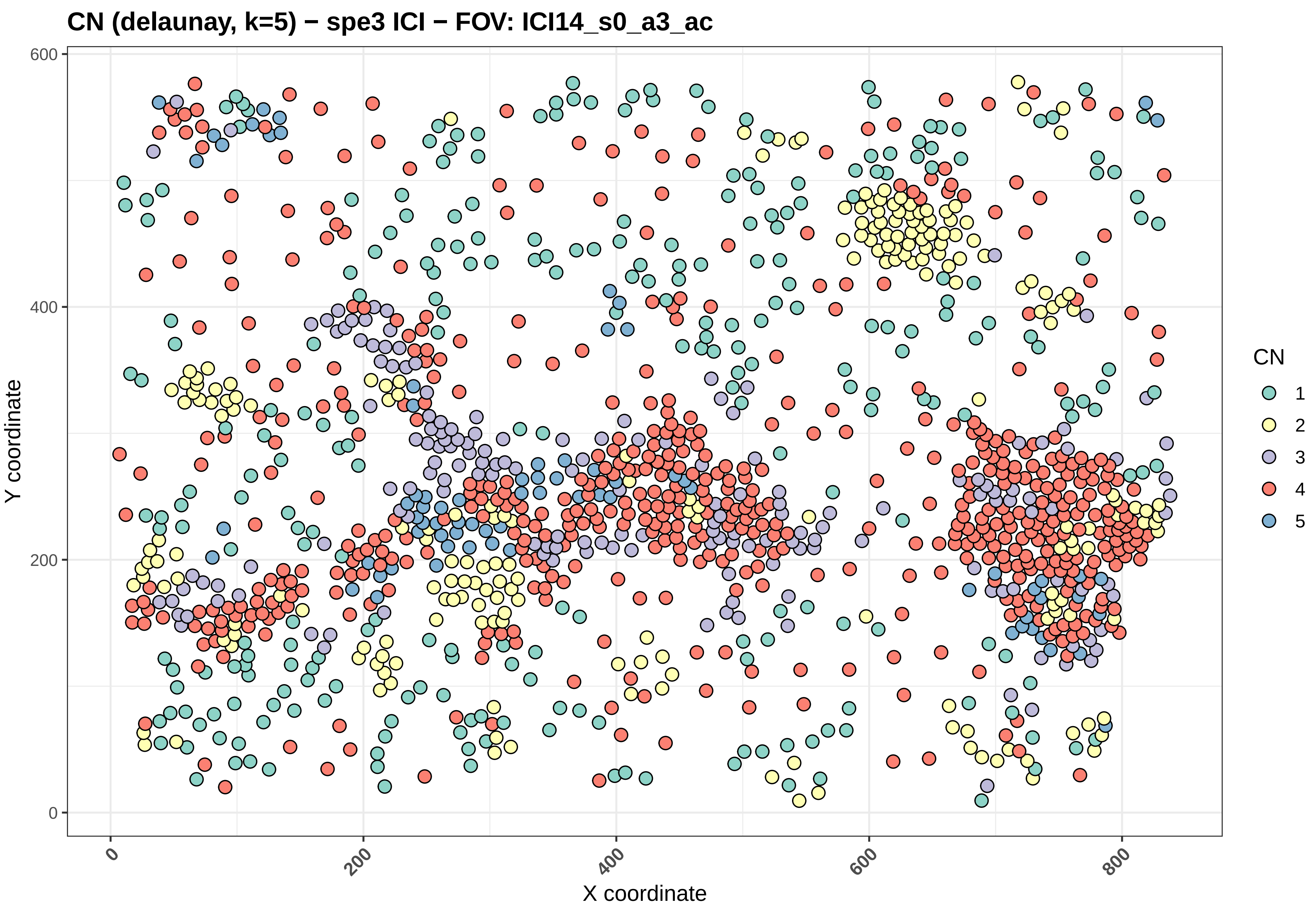

CN (delaunay, k=5) − spe3 ICI − FOV: ICI14_s0_a3_ac
Y coordinate
X coordinate
0
200
400
600
800
CN
1
2
3
4
5

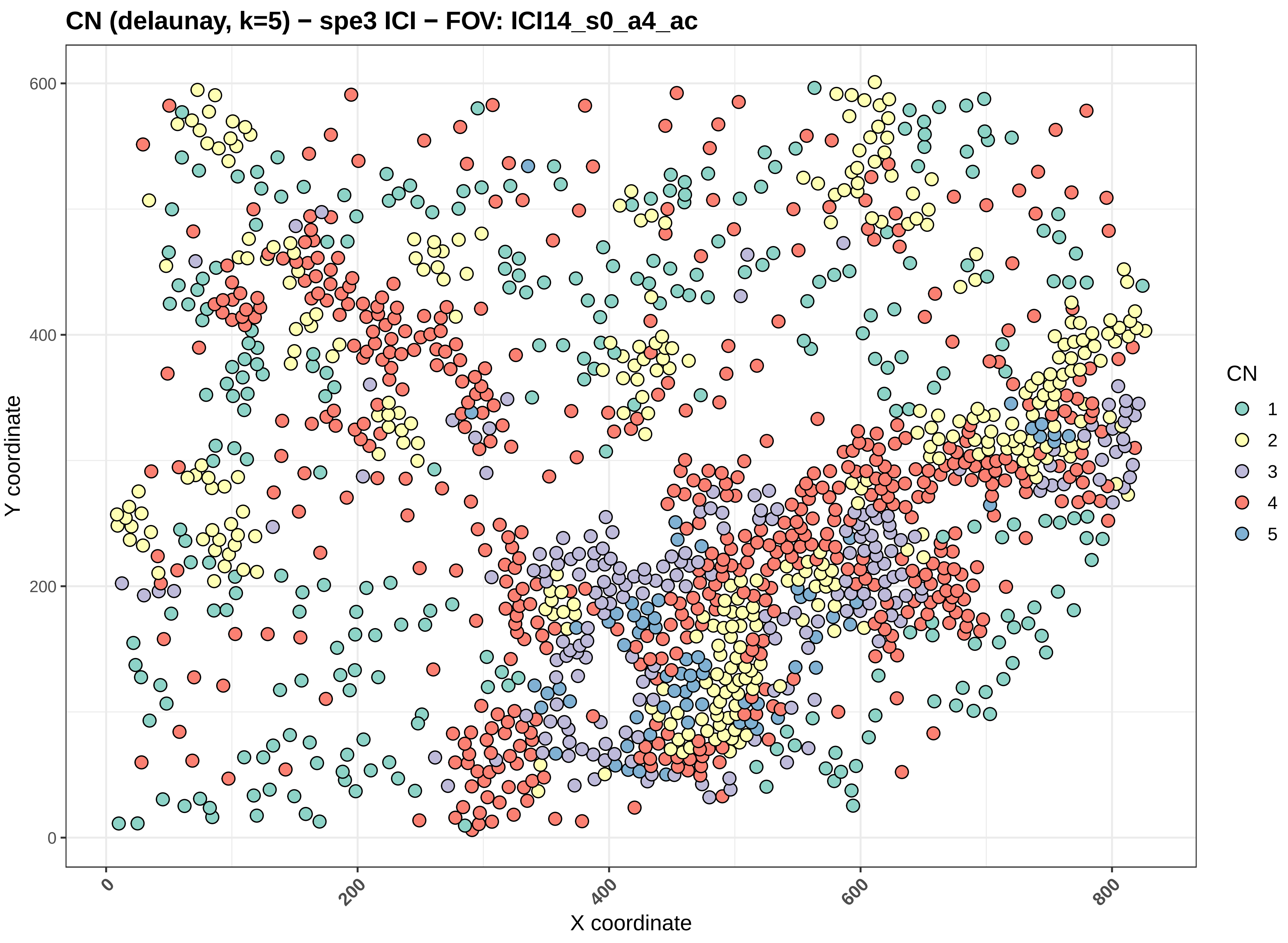
CN (delaunay, k=5) − spe3 ICI − FOV: ICI14_s0_a4_ac
Y coordinate
X coordinate
0
200
400
600
800
CN
1
2
3
4
5

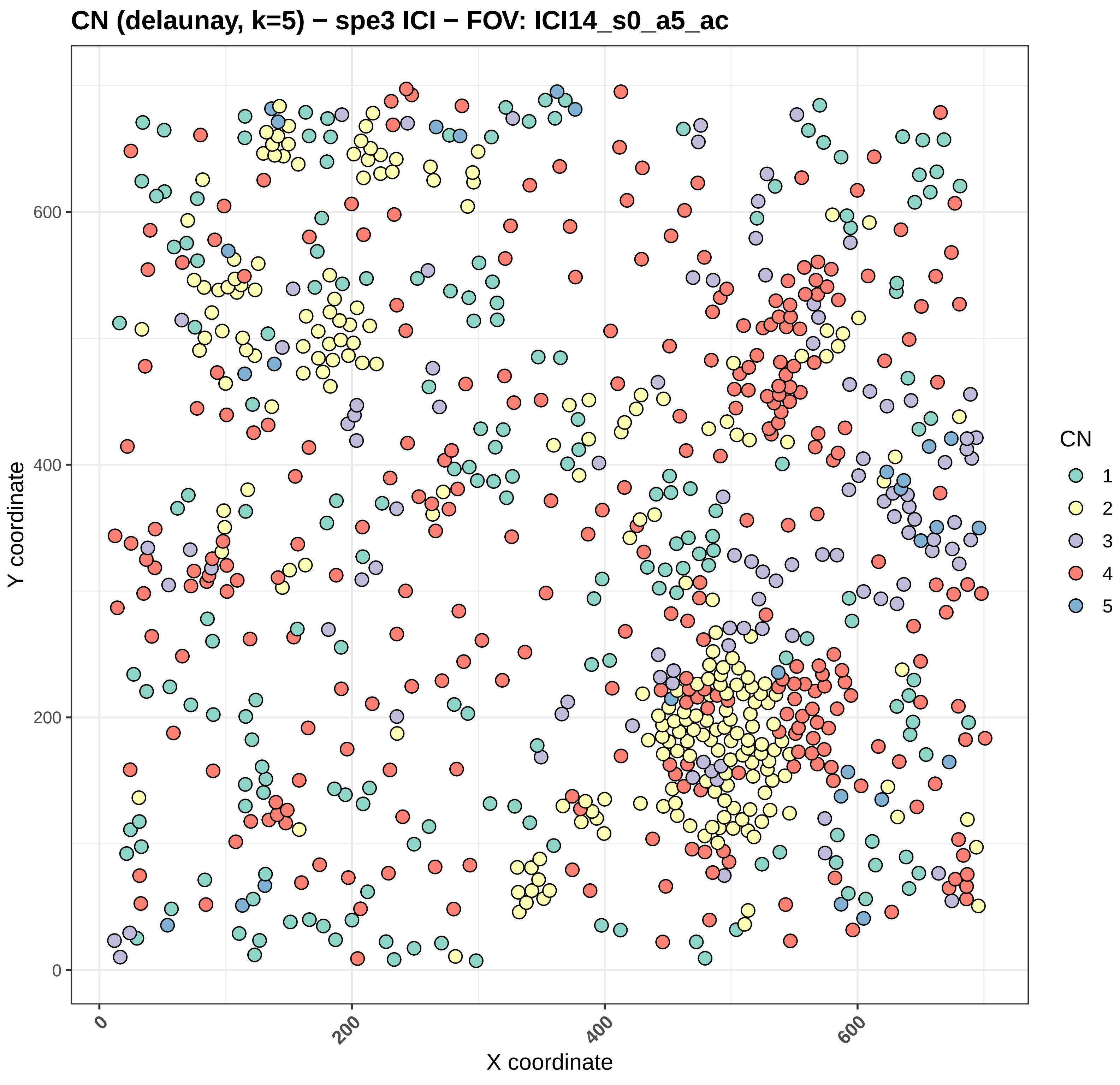

CN (delaunay, k=5) − spe3 ICI − FOV: ICI14_s0_a5_ac
Y coordinate
X coordinate
0
200
400
600
CN
1
2
3
4
5

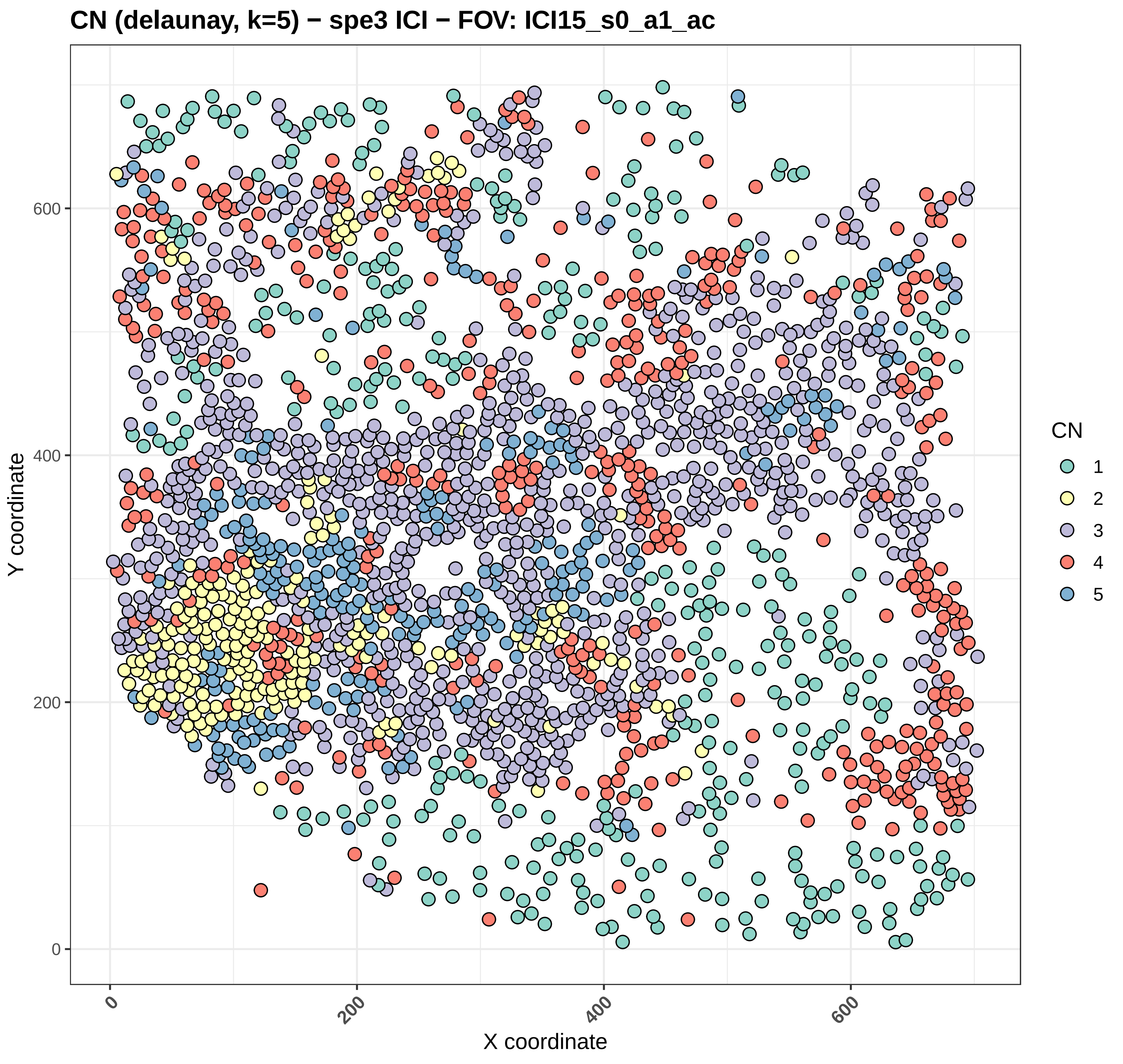
CN (delaunay, k=5) − spe3 ICI − FOV: ICI15_s0_a1_ac
Y coordinate
X coordinate
0
200
400
600
CN
1
2
3
4
5

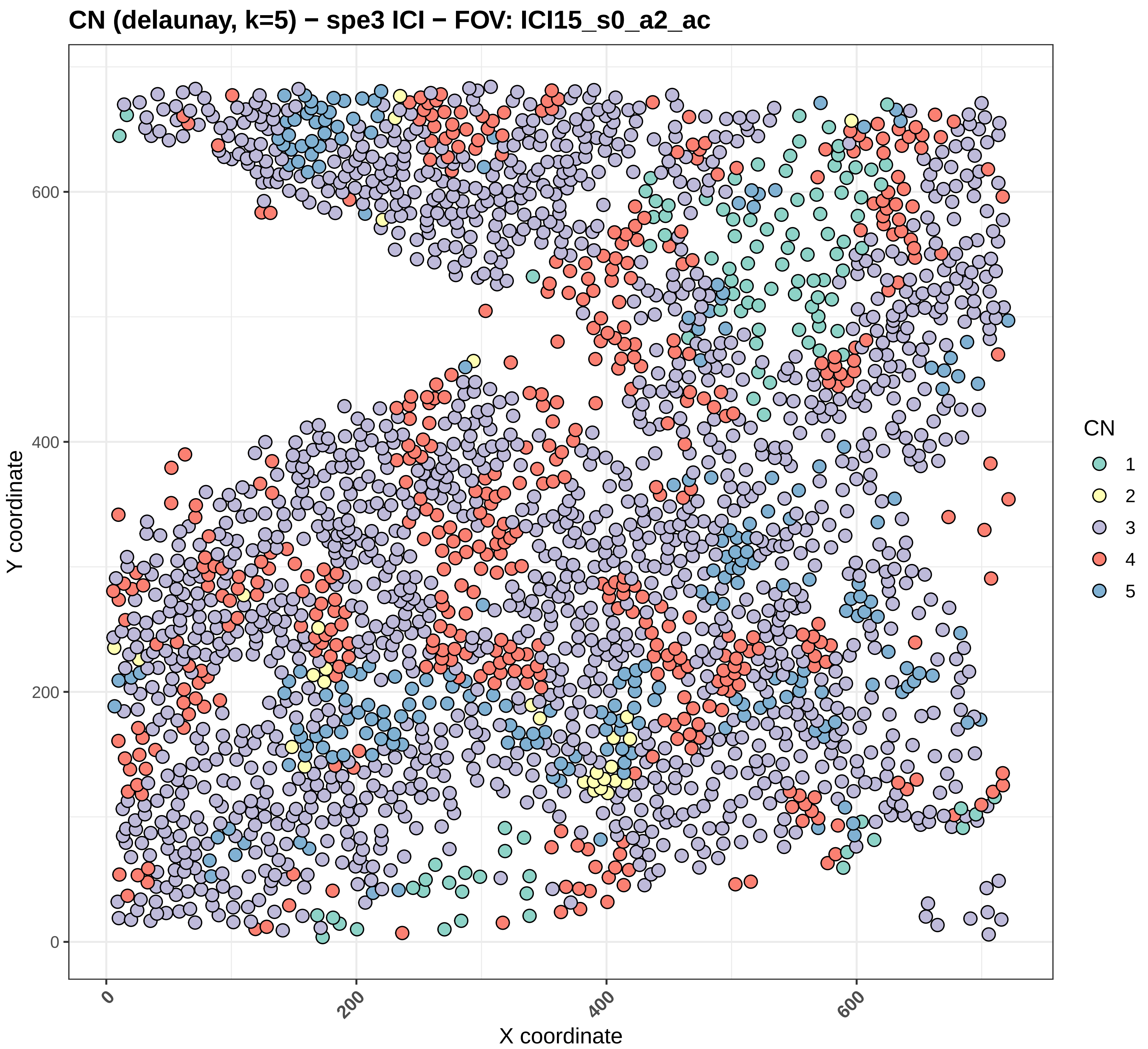
CN (delaunay, k=5) − spe3 ICI − FOV: ICI15_s0_a2_ac
Y coordinate
X coordinate
0
200
400
600
CN
1
2
3
4
5

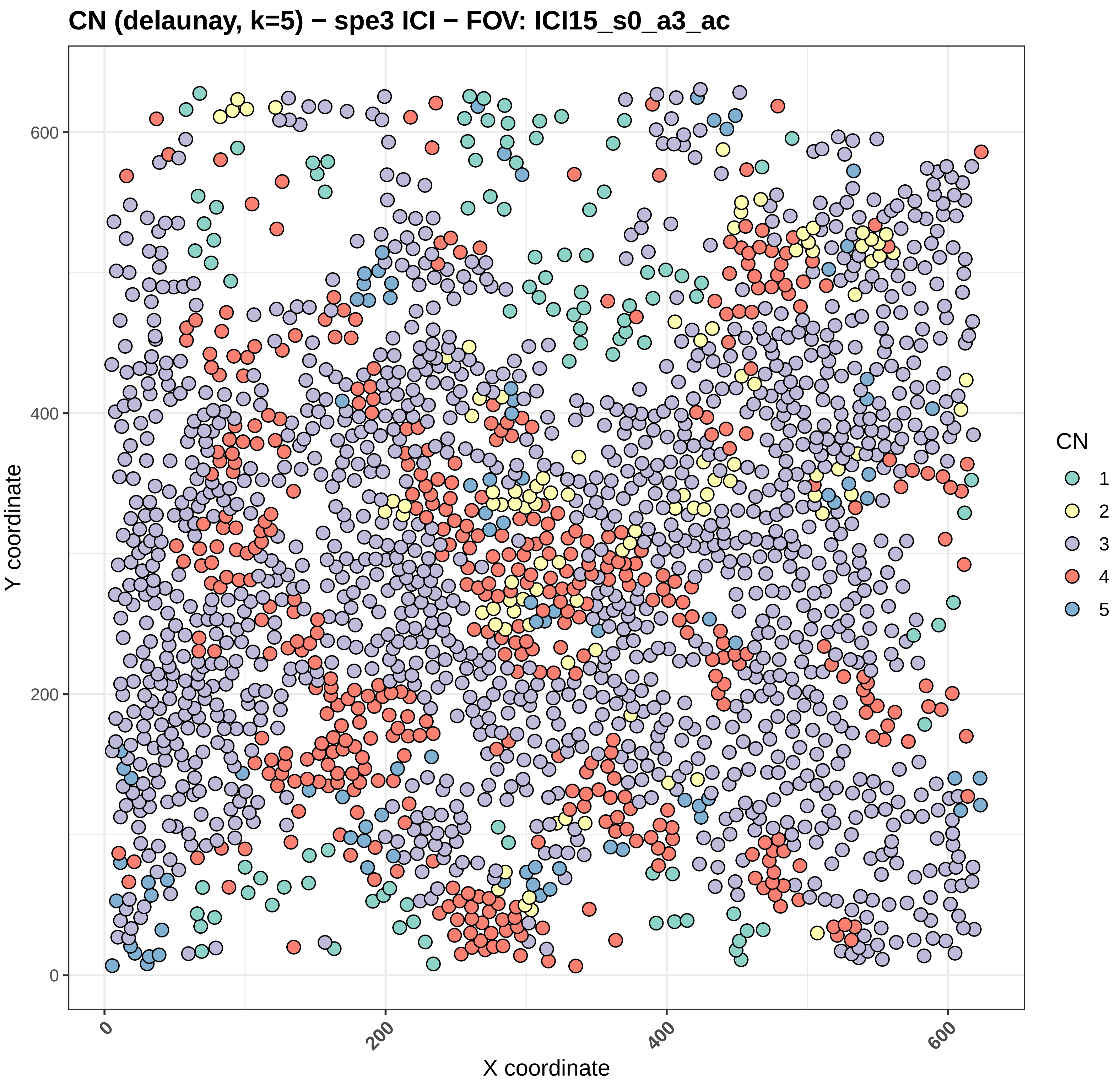
CN (delaunay, k=5) − spe3 ICI − FOV: ICI15_s0_a3_ac
Y coordinate
X coordinate
0
200
400
600
CN
1
2
3
4
5

#### CN (delaunay, k=5) − spe3 ICI − FOV: ICI15_s0_a4_ac

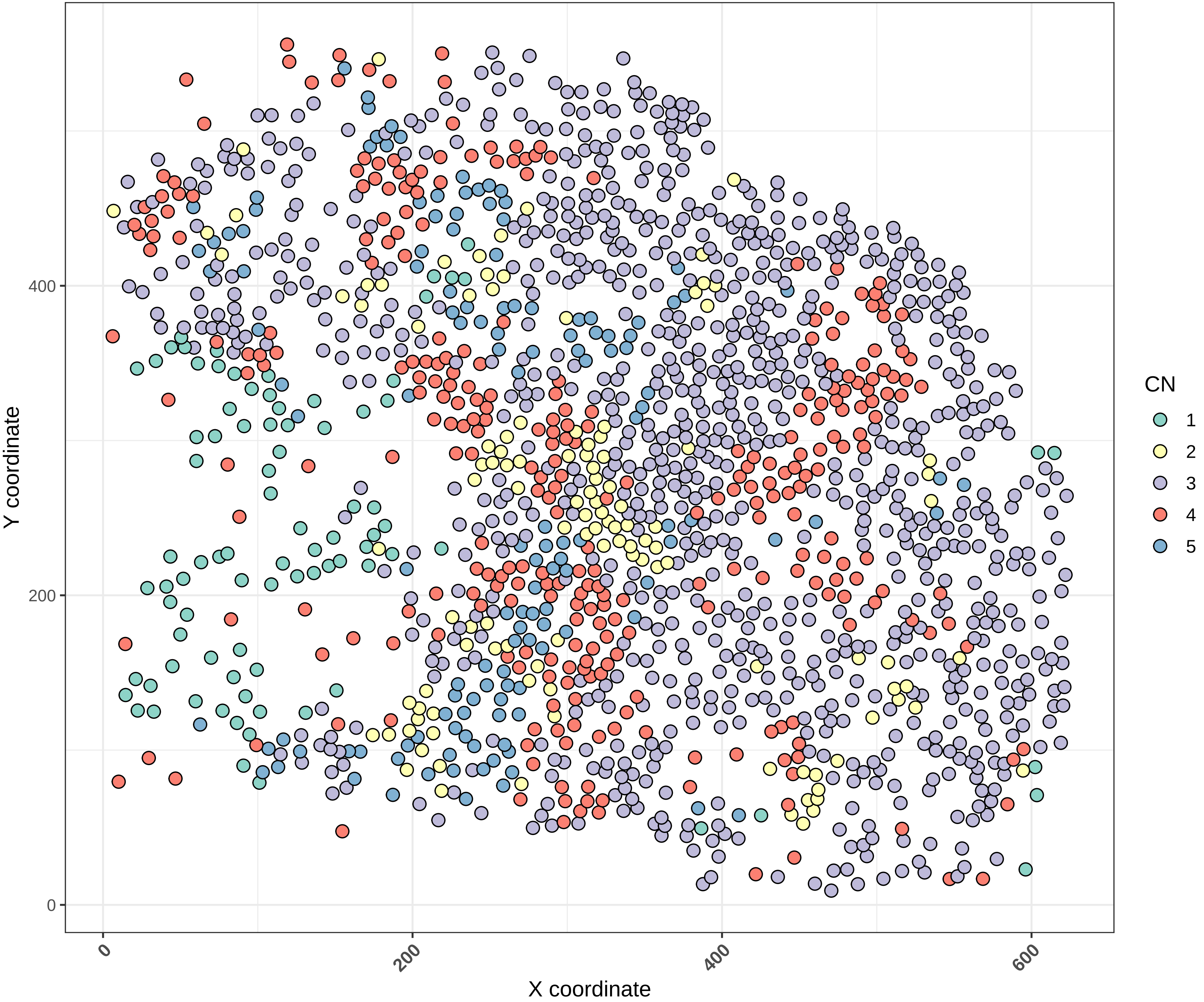

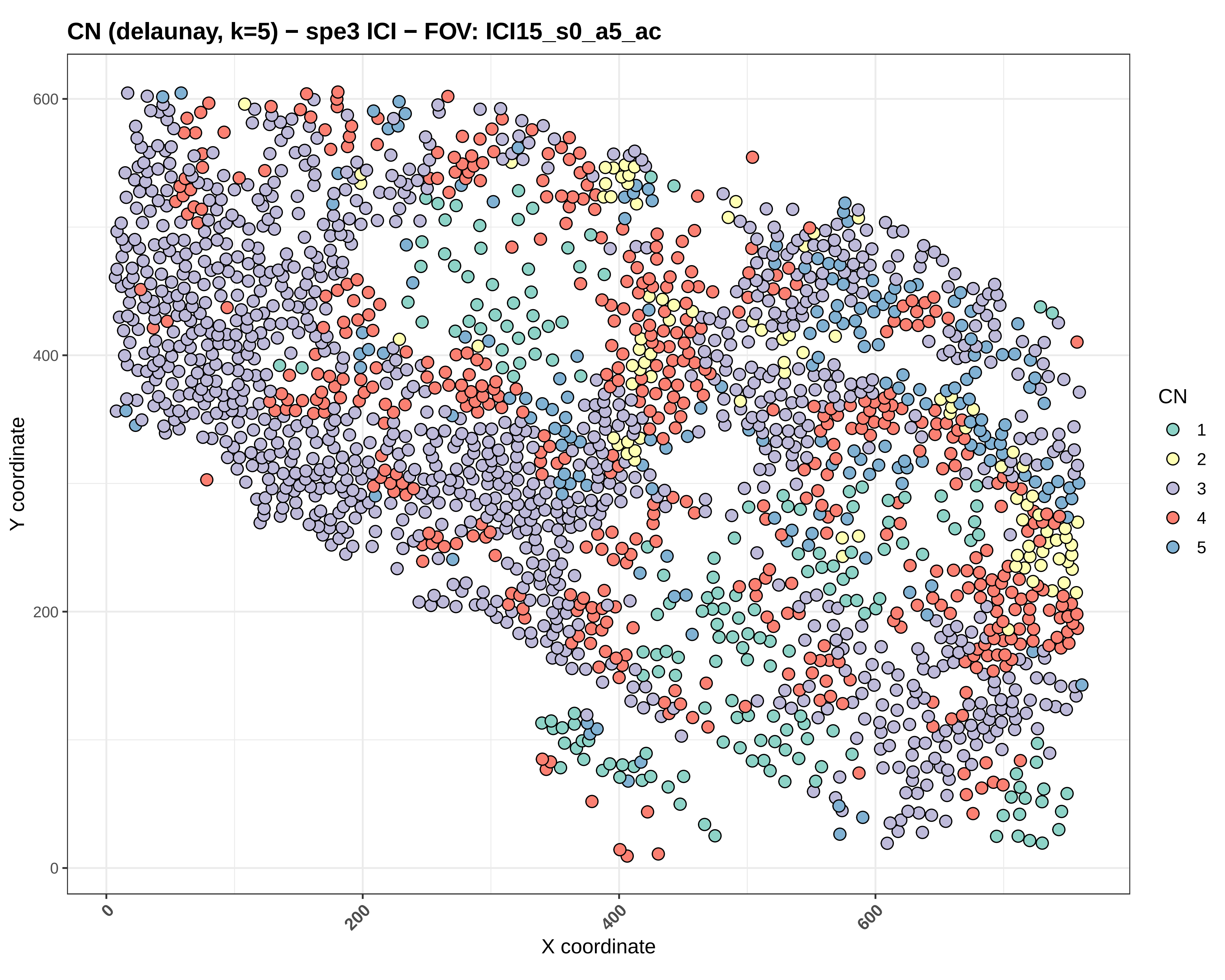

CN (delaunay, k=5) − spe3 ICI − FOV: ICI15_s0_a5_ac
Y coordinate
X coordinate
0
200
400
600
CN
1
2
3
4
5

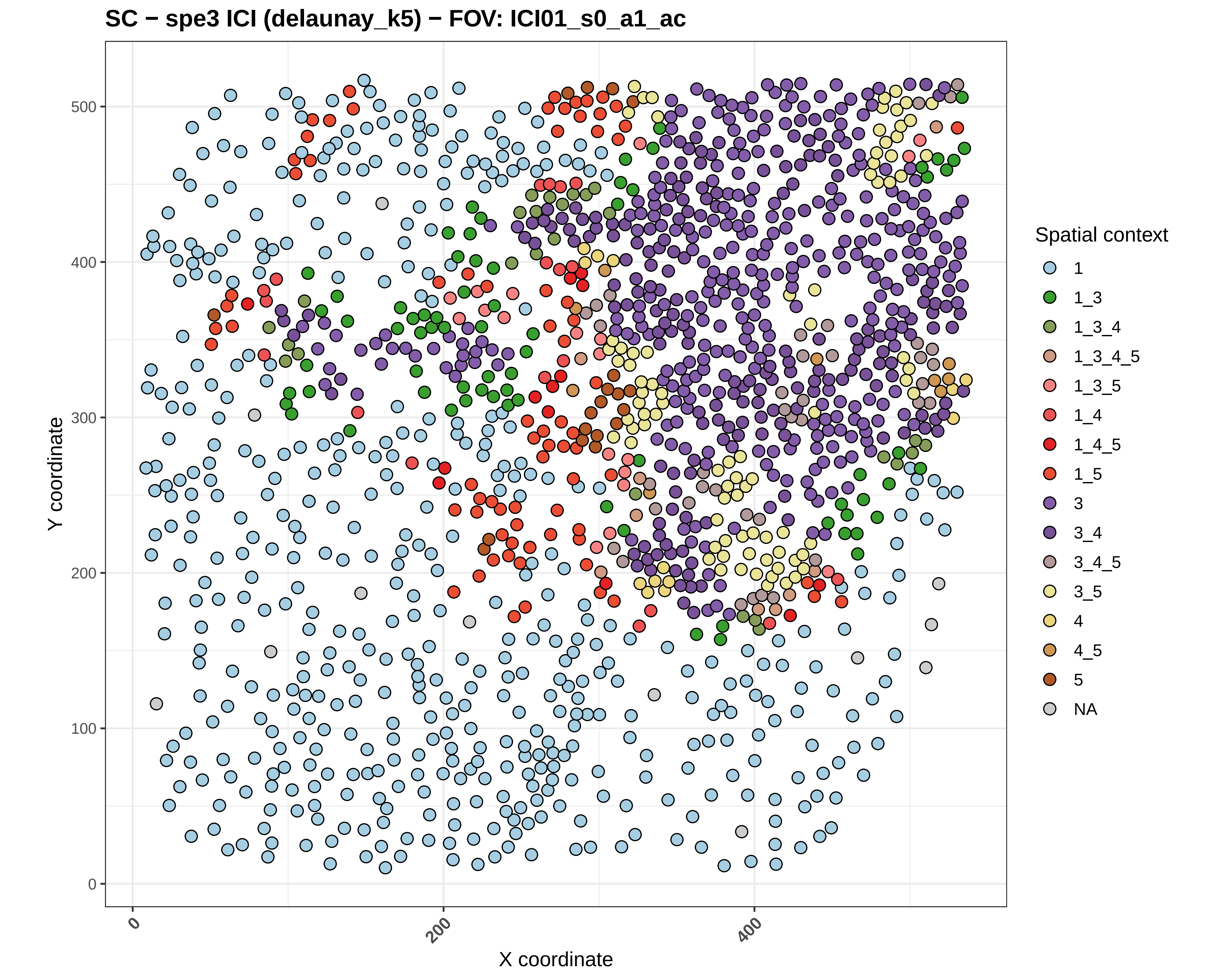
SC − spe3 ICI (delaunay_k5) − FOV: ICI01_s0_a1_ac
Y coordinate
X coordinate
0
100
200
300
400
500
0
200
400
Spatial context
1
1_3
1_3_4
1_3_4_5
1_3_5
1_4
1_4_5
1_5
3
3_4
3_4_5
3_5
4
4_5
5
NA

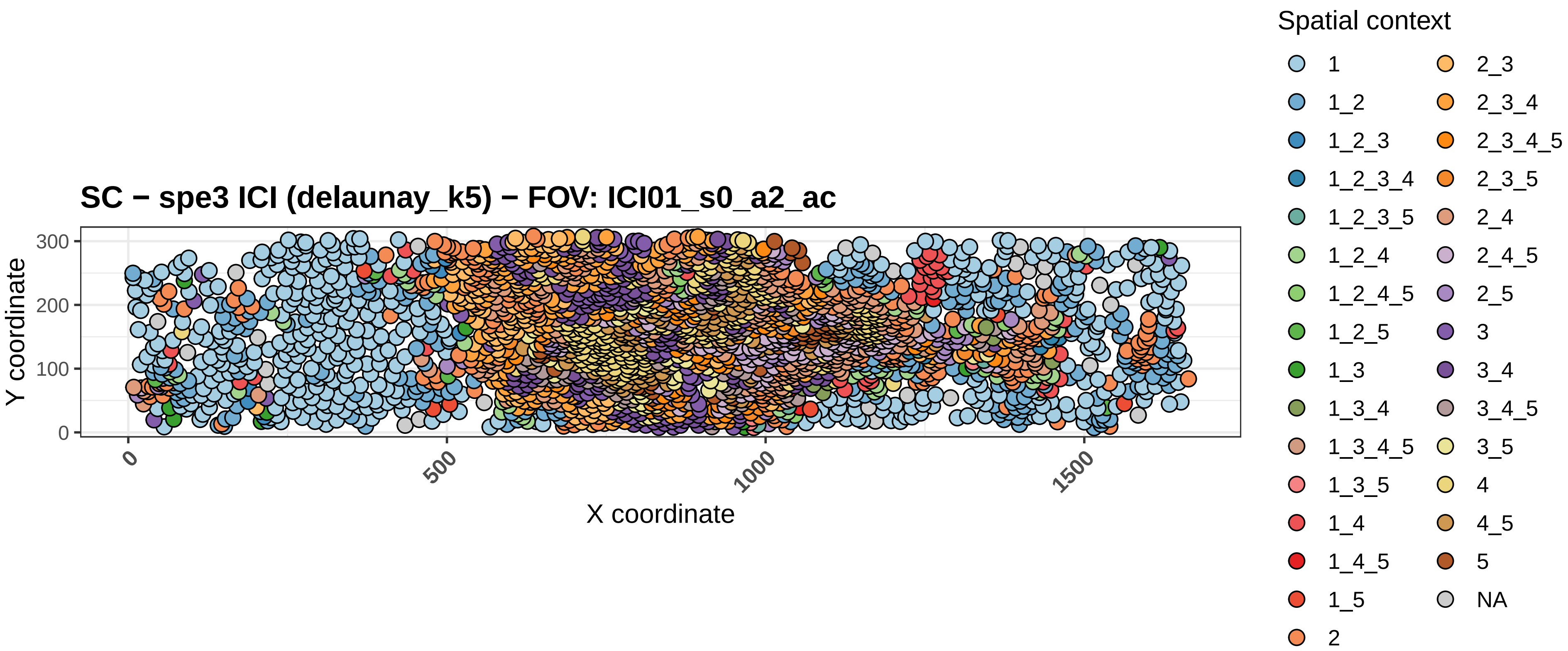

SC − spe3 ICI (delaunay_k5) − FOV: ICI01_s0_a2_ac
Y coordinate
X coordinate
0
100
200
300
0
500
1000
1500
Spatial context
1
1_2
1_2_3
1_2_3_4
1_2_3_5
1_2_4
1_2_4_5
1_2_5
1_3
1_3_4
1_3_4_5
1_3_5
1_4
1_4_5
1_5
2
2_3
2_3_4
2_3_4_5
2_3_5
2_4
2_4_5
2_5
3
3_4
3_4_5
3_5
4
4_5
5
NA

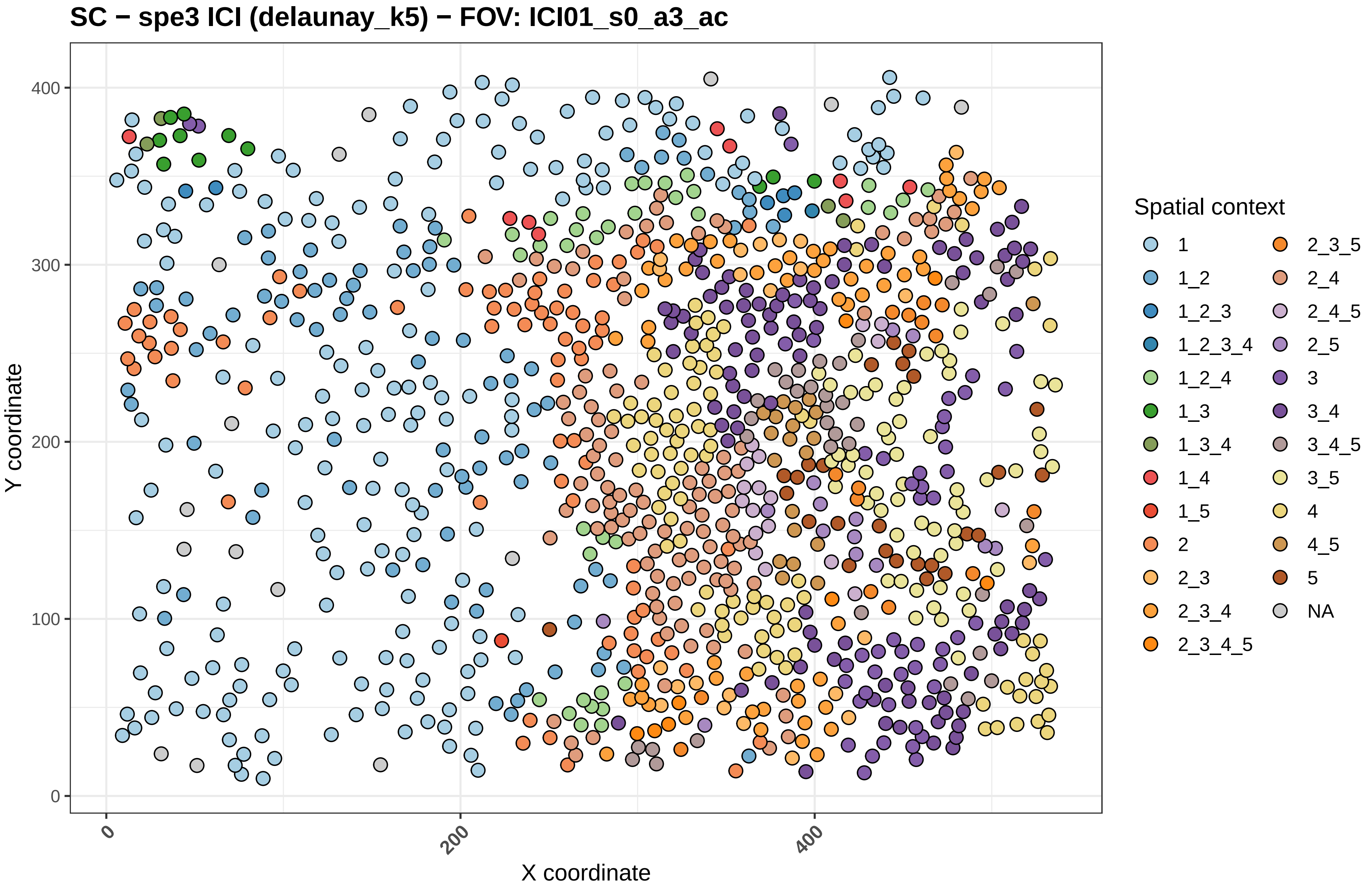

SC − spe3 ICI (delaunay_k5) − FOV: ICI01_s0_a3_ac
Y coordinate
X coordinate
0
100
200
300
400
0
200
400
Spatial context
1
1_2
1_2_3
1_2_3_4
1_2_4
1_3
1_3_4
1_4
1_5
2
2_3
2_3_4
2_3_4_5
2_3_5
2_4
2_4_5
2_5
3
3_4
3_4_5
3_5
4
4_5
5
NA

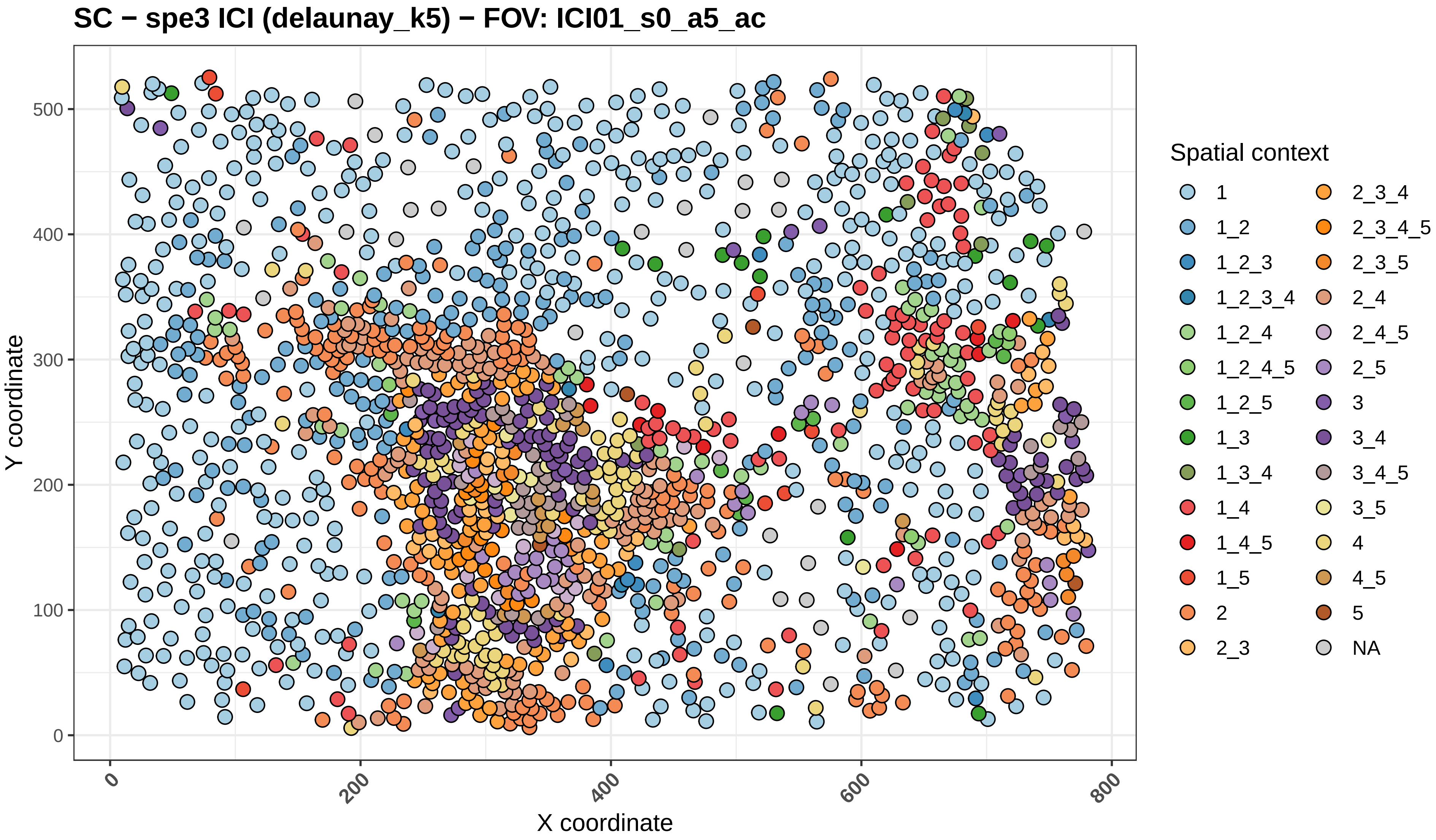
SC − spe3 ICI (delaunay_k5) − FOV: ICI01_s0_a5_ac
Y coordinate
X coordinate
0
100
200
300
400
500
0
200
400
600
800
Spatial context
1
1_2
1_2_3
1_2_3_4
1_2_4
1_2_4_5
1_2_5
1_3
1_3_4
1_4
1_4_5
1_5
2
2_3
2_3_4
2_3_4_5
2_3_5
2_4
2_4_5
2_5
3
3_4
3_4_5
3_5
4
4_5
5
NA

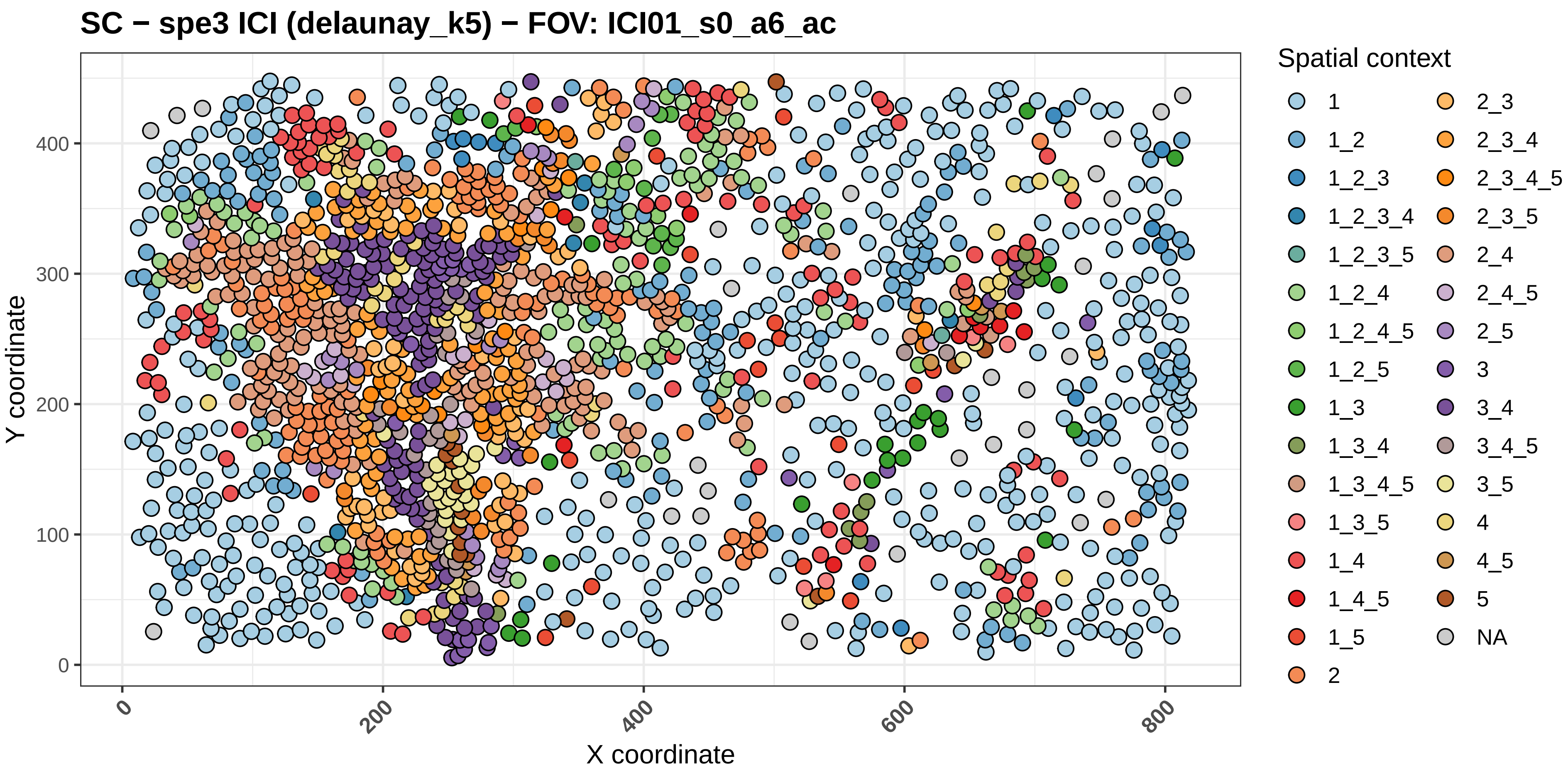

SC − spe3 ICI (delaunay_k5) − FOV: ICI01_s0_a6_ac
Y coordinate
X coordinate
0
100
200
300
400
0
200
400
600
800
Spatial context
1
1_2
1_2_3
1_2_3_4
1_2_3_5
1_2_4
1_2_4_5
1_2_5
1_3
1_3_4
1_3_4_5
1_3_5
1_4
1_4_5
1_5
2
2_3
2_3_4
2_3_4_5
2_3_5
2_4
2_4_5
2_5
3
3_4
3_4_5
3_5
4
4_5
5
NA

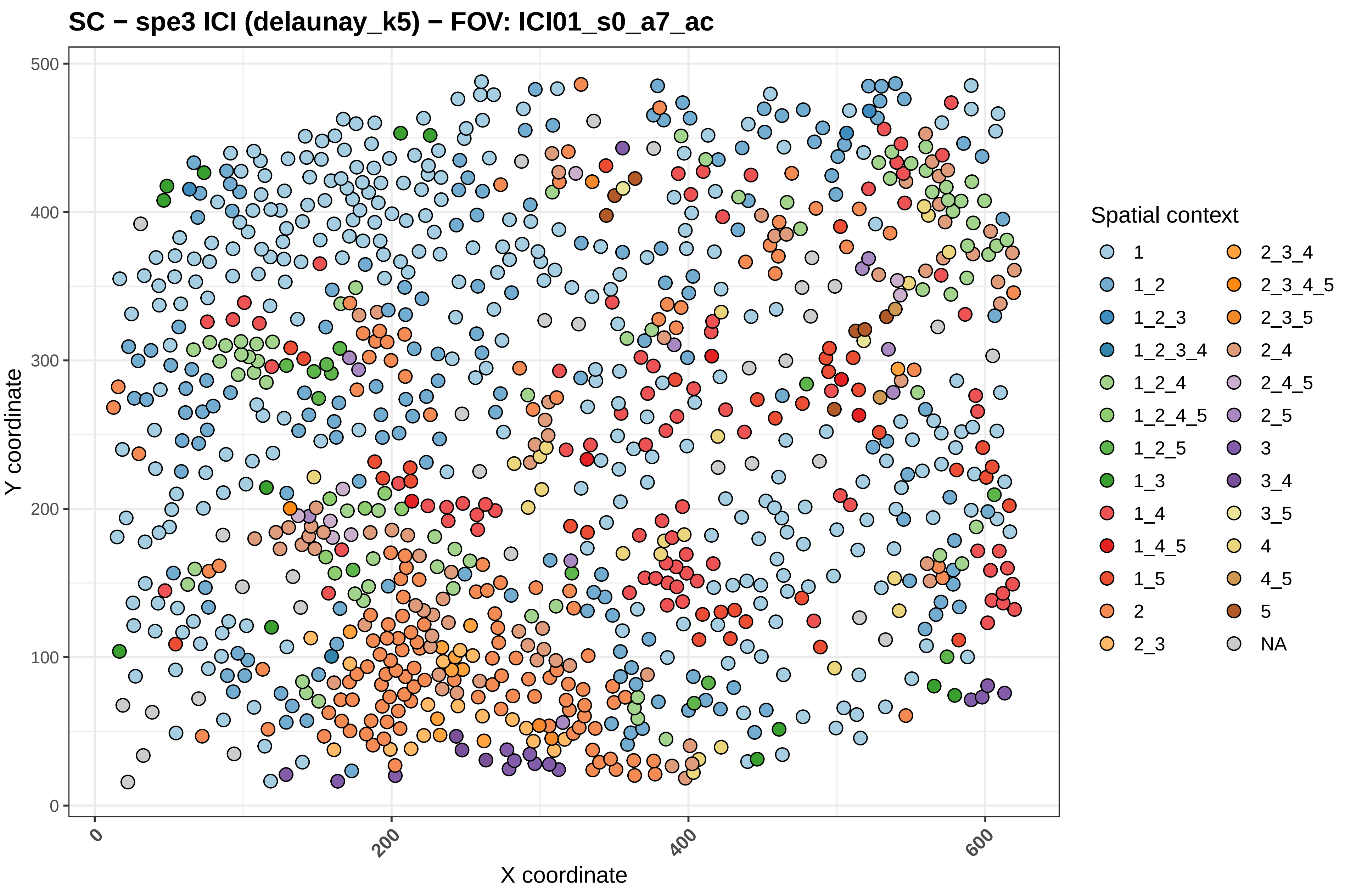

SC − spe3 ICI (delaunay_k5) − FOV: ICI01_s0_a7_ac
Y coordinate
X coordinate
0
100
200
300
400
500
0
200
400
600
Spatial context
1
1_2
1_2_3
1_2_3_4
1_2_4
1_2_4_5
1_2_5
1_3
1_4
1_4_5
1_5
2
2_3
2_3_4
2_3_4_5
2_3_5
2_4
2_4_5
2_5
3
3_4
3_5
4
4_5
5
NA

SC − spe3 ICI (delaunay_k5) − FOV: ICI01_s0_a8_ac

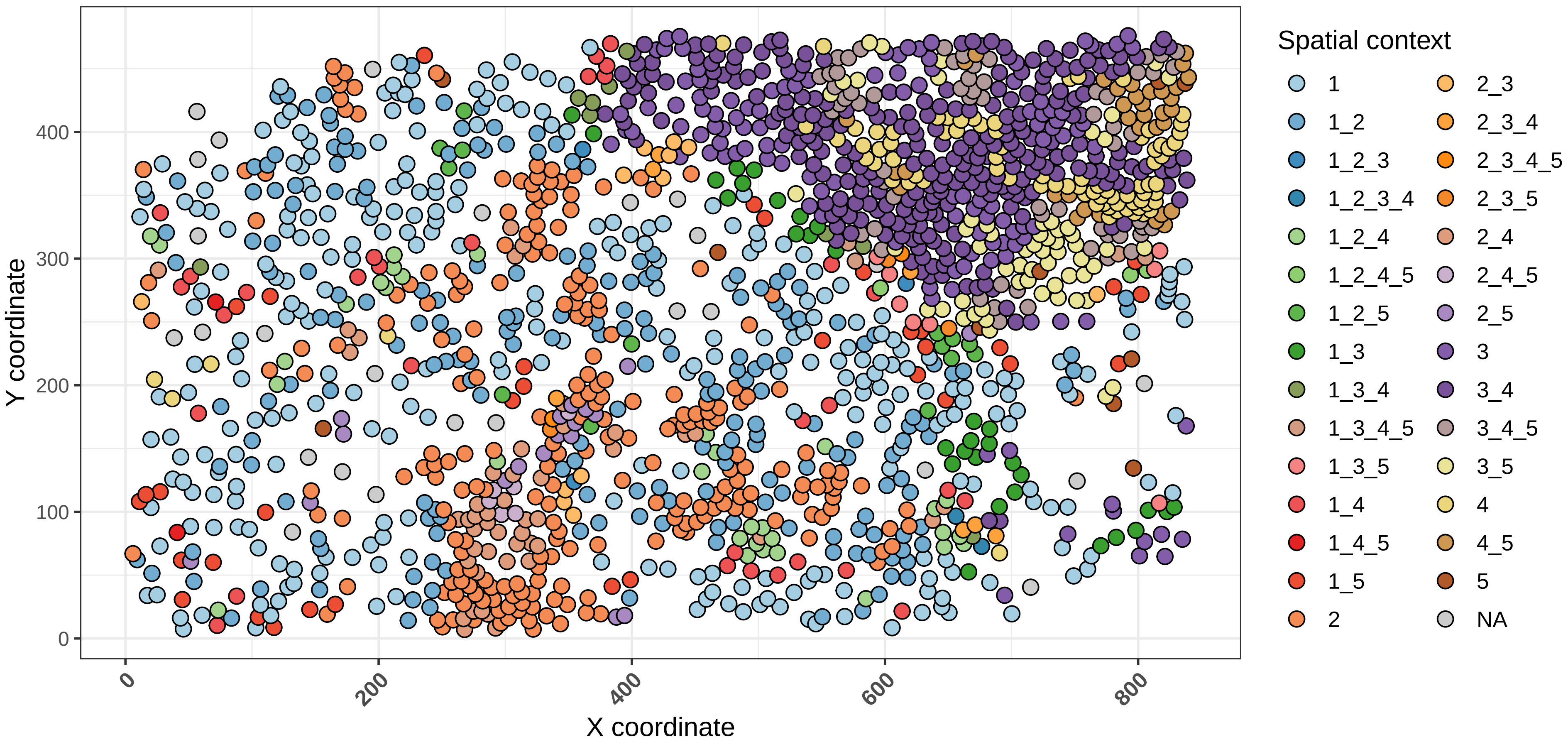

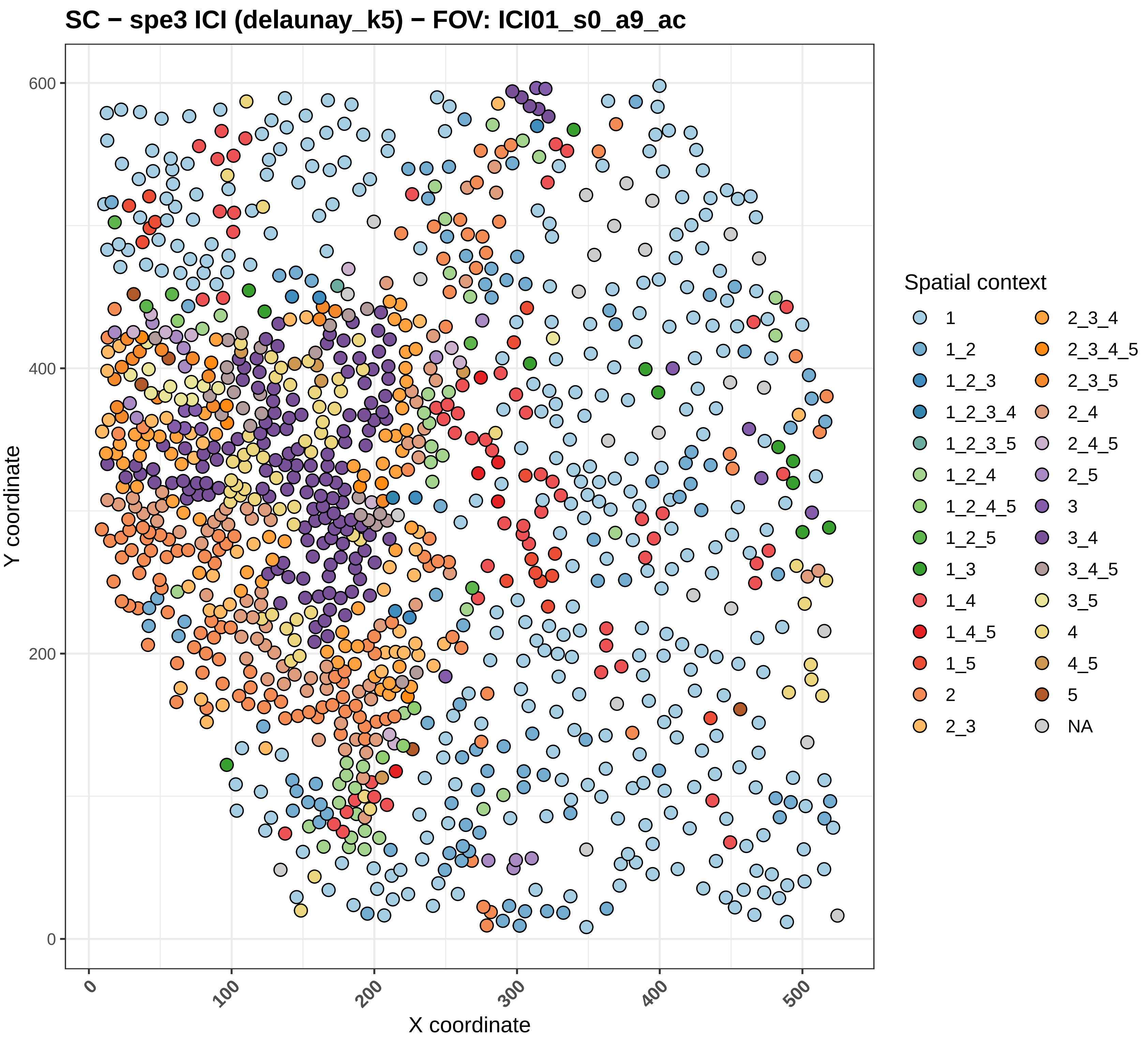

SC − spe3 ICI (delaunay_k5) − FOV: ICI01_s0_a9_ac
Y coordinate
X coordinate
0
200
400
600
0
100
200
300
400
500
Spatial context
1
1_2
1_2_3
1_2_3_4
1_2_3_5
1_2_4
1_2_4_5
1_2_5
1_3
1_4
1_4_5
1_5
2
2_3
2_3_4
2_3_4_5
2_3_5
2_4
2_4_5
2_5
3
3_4
3_4_5
3_5
4
4_5
5
NA

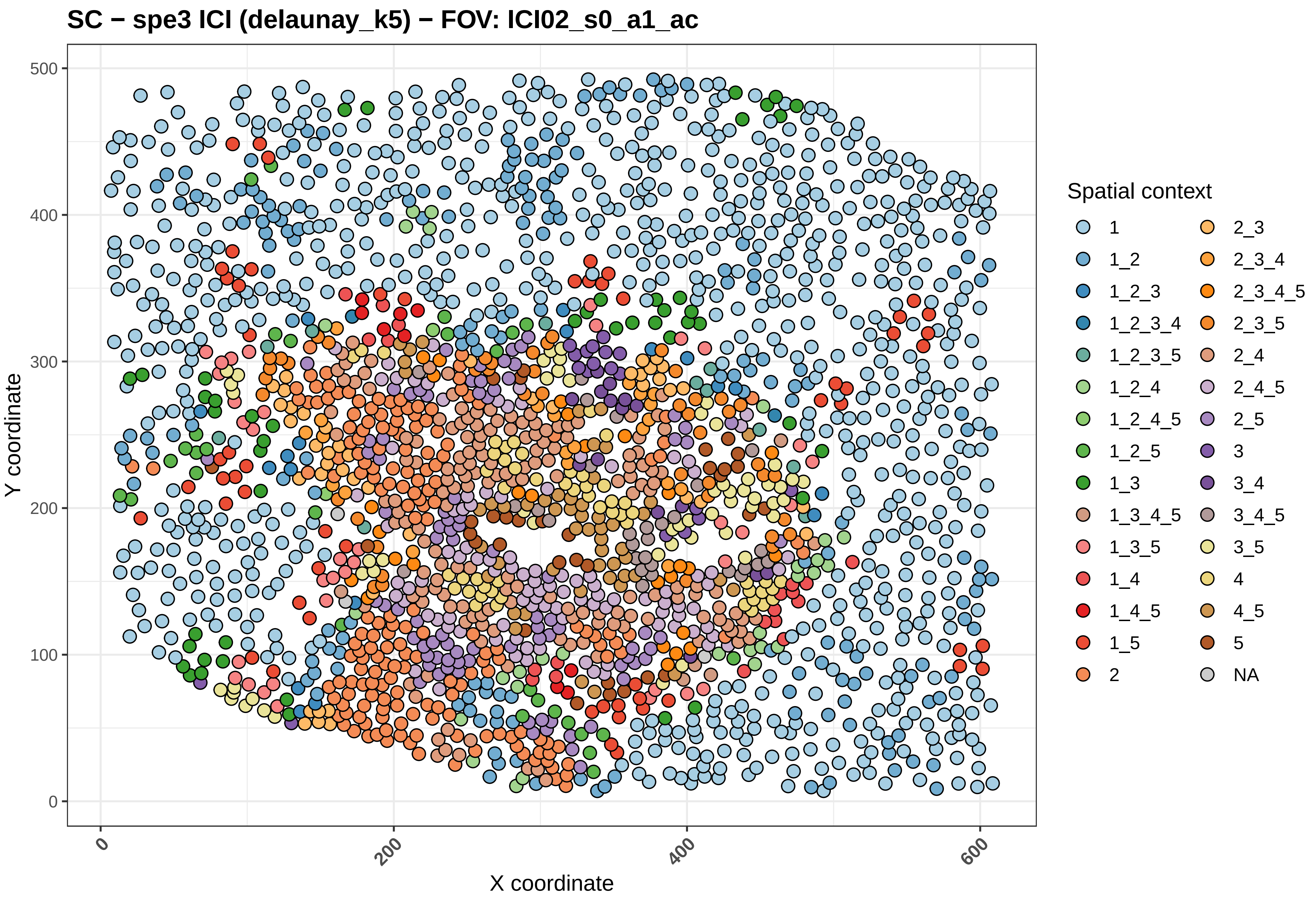

SC − spe3 ICI (delaunay_k5) − FOV: ICI02_s0_a1_ac
Y coordinate
X coordinate
0
100
200
300
400
500
600
Spatial context
1
1_2
1_2_3
1_2_3_4
1_2_3_5
1_2_4
1_2_4_5
1_2_5
1_3
1_3_4_5
1_3_5
1_4
1_4_5
1_5
2
2_3
2_3_4
2_3_4_5
2_3_5
2_4
2_4_5
2_5
3
3_4
3_4_5
3_5
4
4_5
5
NA

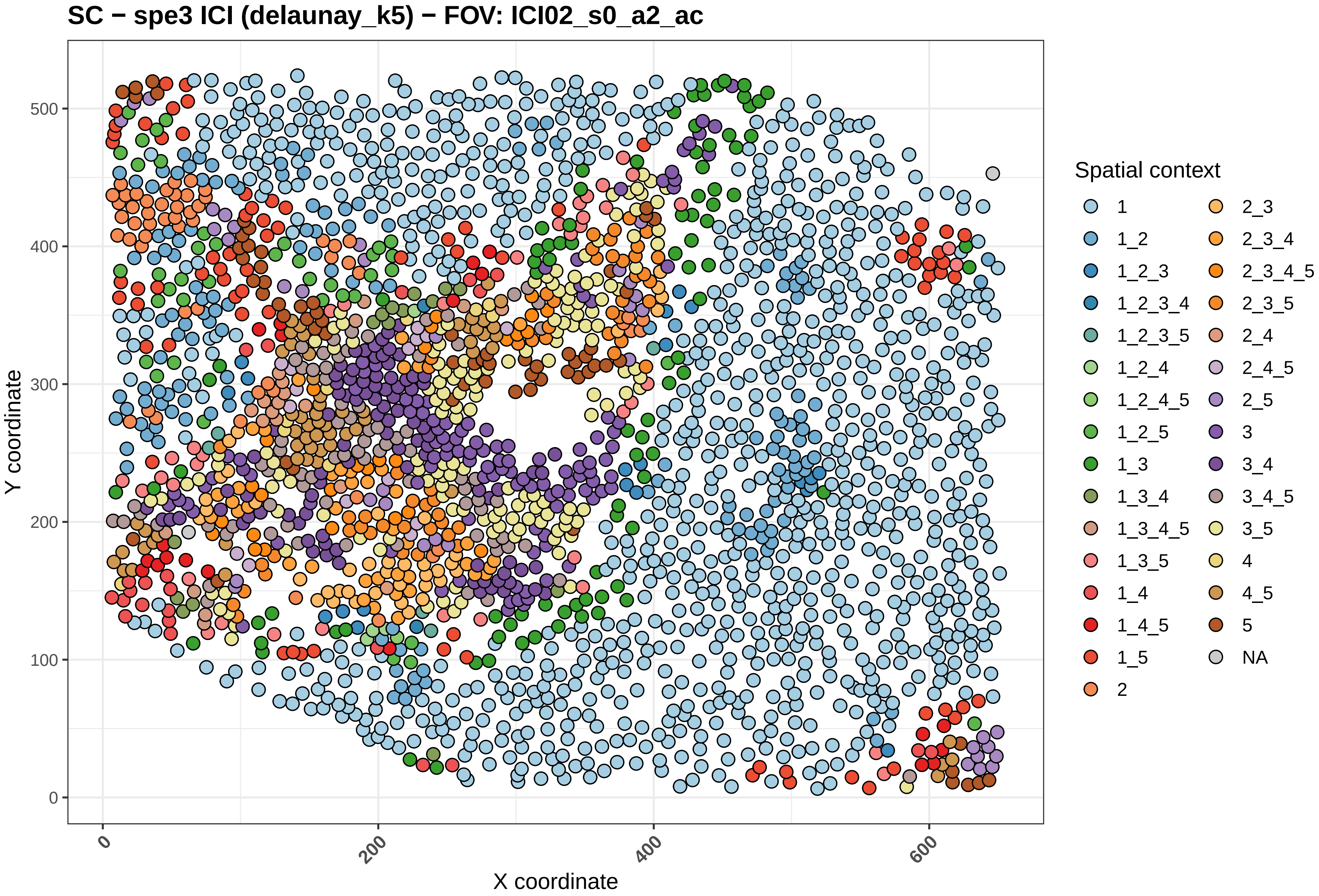
SC − spe3 ICI (delaunay_k5) − FOV: ICI02_s0_a2_ac
Y coordinate
X coordinate
0
100
200
300
400
500
0
200
400
600
Spatial context
1
1_2
1_2_3
1_2_3_4
1_2_3_5
1_2_4
1_2_4_5
1_2_5
1_3
1_3_4
1_3_4_5
1_3_5
1_4
1_4_5
1_5
2
2_3
2_3_4
2_3_4_5
2_3_5
2_4
2_4_5
2_5
3
3_4
3_4_5
3_5
4
4_5
5
NA

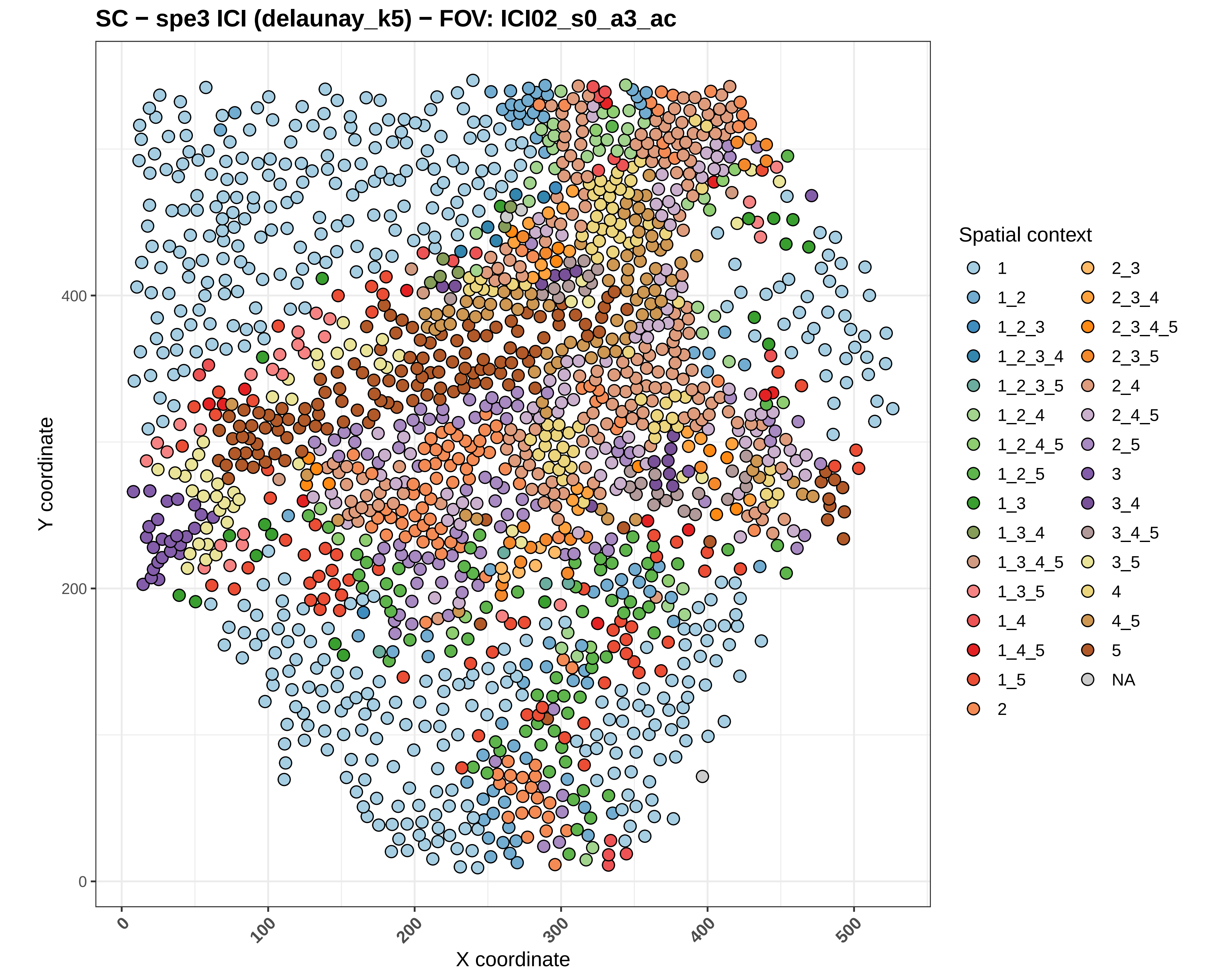

SC − spe3 ICI (delaunay_k5) − FOV: ICI02_s0_a3_ac
Y coordinate
X coordinate
0
200
400
0
100
200
300
400
500
Spatial context
1
1_2
1_2_3
1_2_3_4
1_2_3_5
1_2_4
1_2_4_5
1_2_5
1_3
1_3_4
1_3_4_5
1_3_5
1_4
1_4_5
1_5
2
2_3
2_3_4
2_3_4_5
2_3_5
2_4
2_4_5
2_5
3
3_4
3_4_5
3_5
4
4_5
5
NA

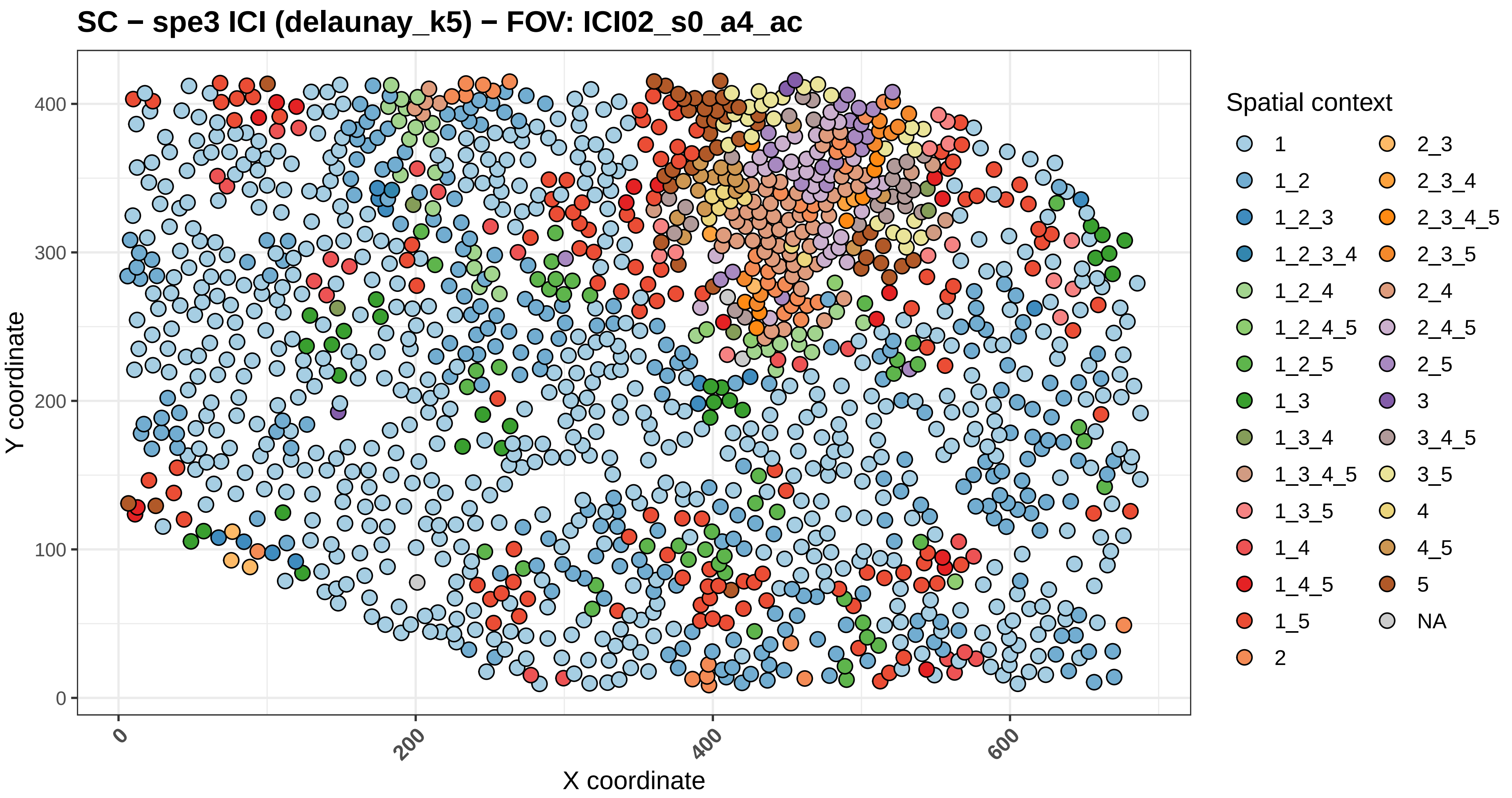

SC − spe3 ICI (delaunay_k5) − FOV: ICI02_s0_a4_ac
Y coordinate
X coordinate
0
100
200
300
400
0
200
400
600
Spatial context
1
1_2
1_2_3
1_2_3_4
1_2_4
1_2_4_5
1_2_5
1_3
1_3_4
1_3_4_5
1_3_5
1_4
1_4_5
1_5
2
2_3
2_3_4
2_3_4_5
2_3_5
2_4
2_4_5
2_5
3
3_4_5
3_5
4
4_5
5
NA

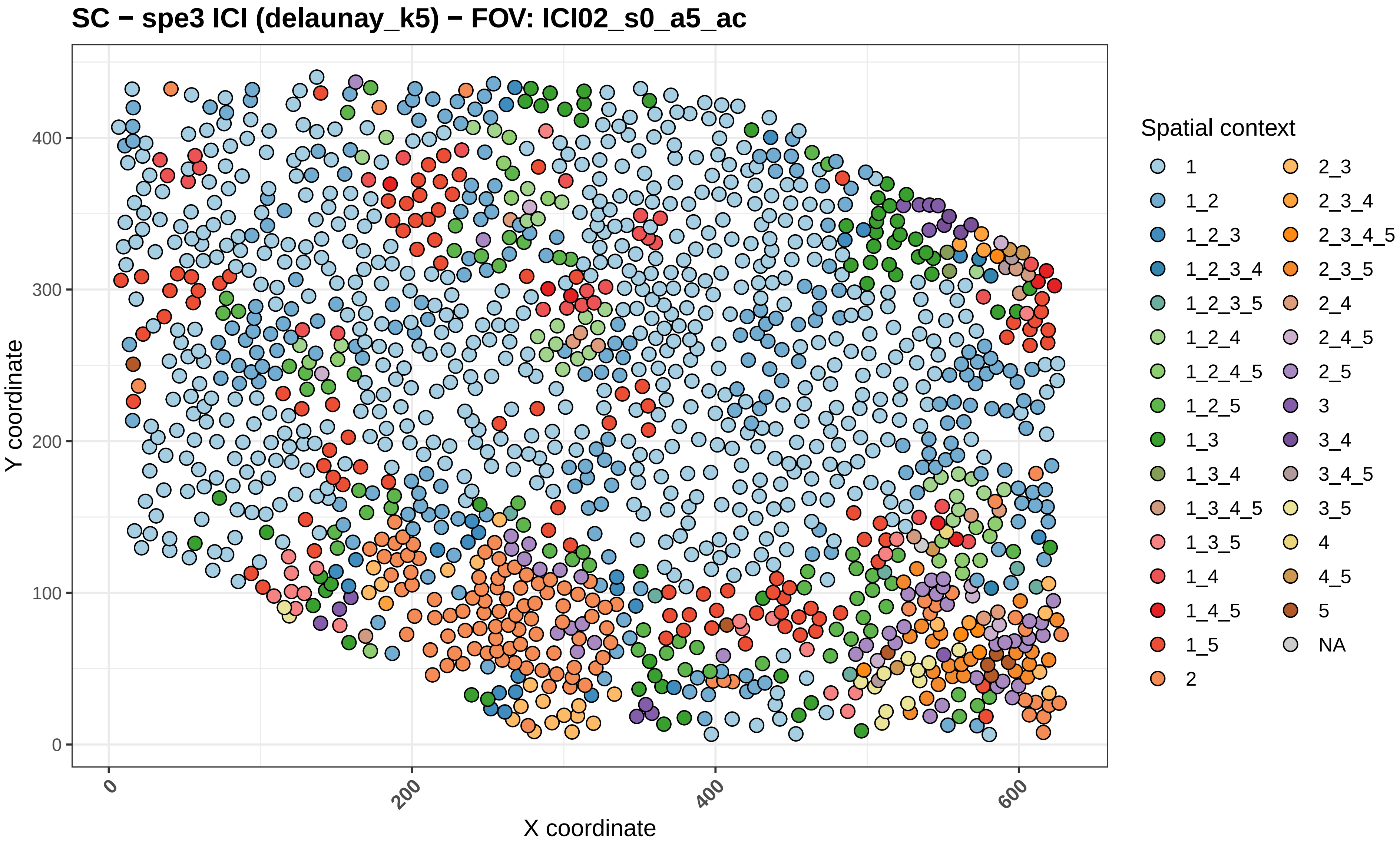

SC − spe3 ICI (delaunay_k5) − FOV: ICI02_s0_a5_ac
Y coordinate
X coordinate
0
100
200
300
400
0
200
400
600
Spatial context
1
1_2
1_2_3
1_2_3_4
1_2_3_5
1_2_4
1_2_4_5
1_2_5
1_3
1_3_4
1_3_4_5
1_3_5
1_4
1_4_5
1_5
2
2_3
2_3_4
2_3_4_5
2_3_5
2_4
2_4_5
2_5
3
3_4
3_4_5
3_5
4
4_5
5
NA

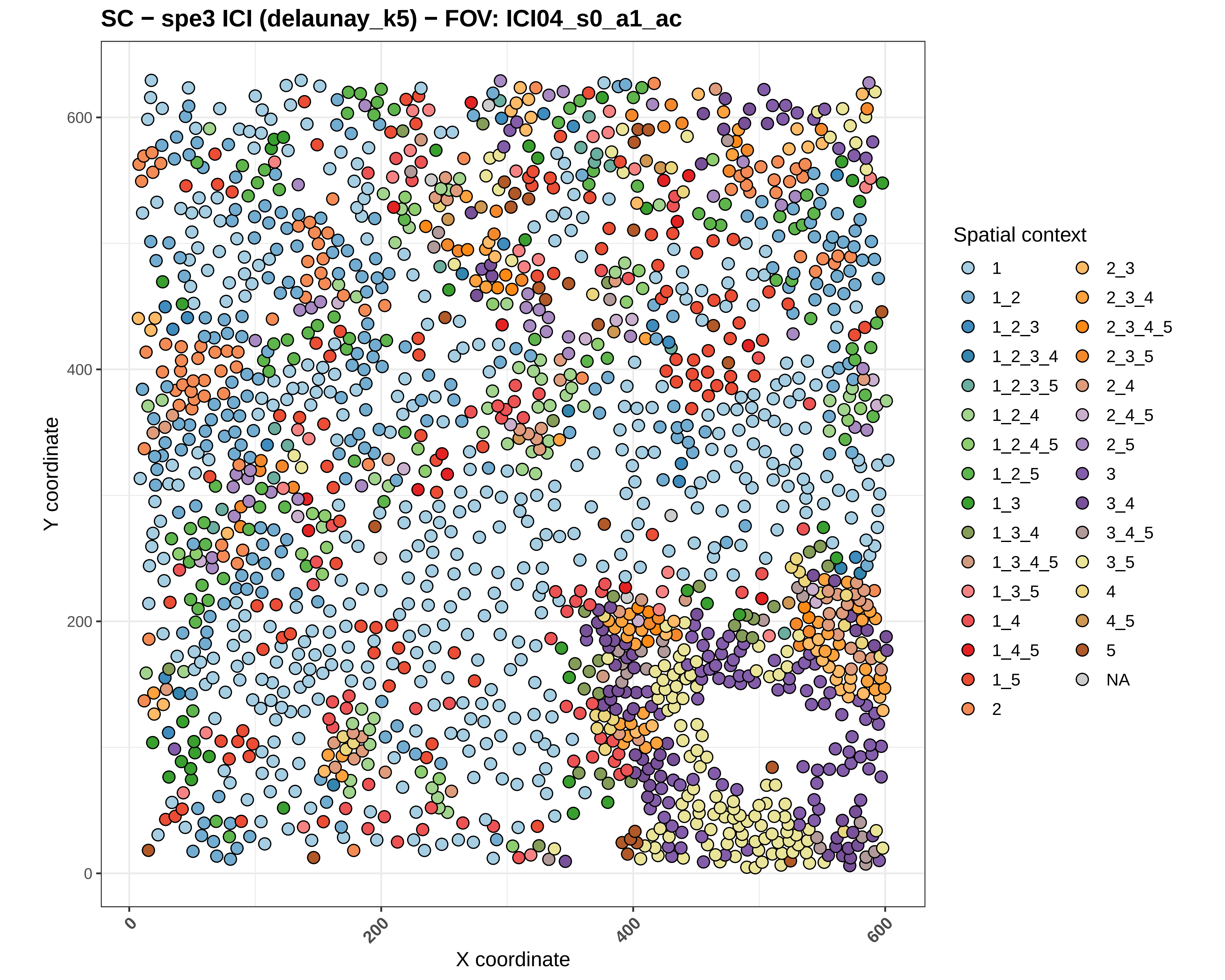

SC − spe3 ICI (delaunay_k5) − FOV: ICI04_s0_a1_ac
Y coordinate
X coordinate
0
200
400
600
Spatial context
1
1_2
1_2_3
1_2_3_4
1_2_3_5
1_2_4
1_2_4_5
1_2_5
1_3
1_3_4
1_3_4_5
1_3_5
1_4
1_4_5
1_5
2
2_3
2_3_4
2_3_4_5
2_3_5
2_4
2_4_5
2_5
3
3_4
3_4_5
3_5
4
4_5
5
NA

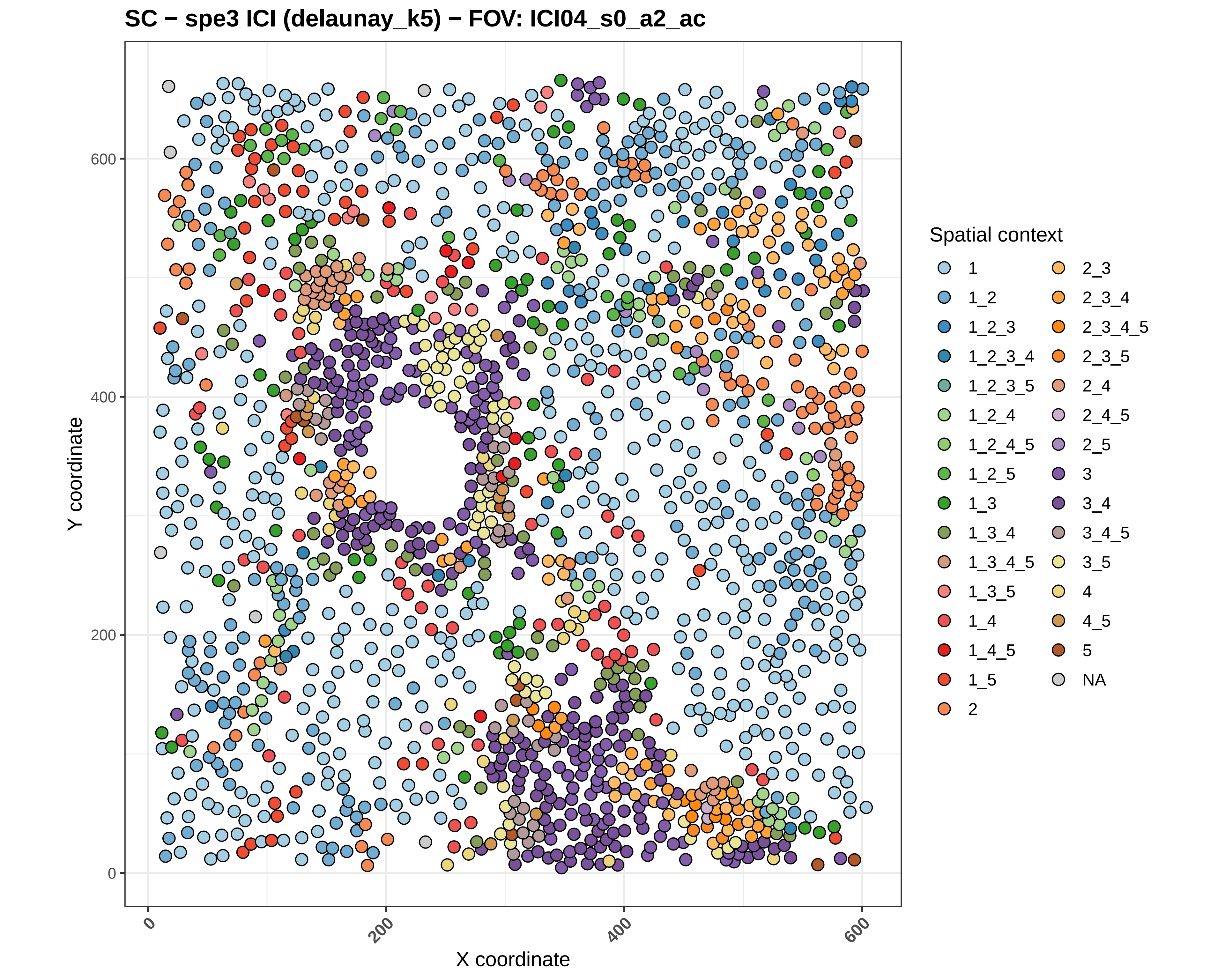

SC − spe3 ICI (delaunay_k5) − FOV: ICI04_s0_a2_ac
Y coordinate
X coordinate
0
200
400
600
Spatial context
1
1_2
1_2_3
1_2_3_4
1_2_3_5
1_2_4
1_2_4_5
1_2_5
1_3
1_3_4
1_3_4_5
1_3_5
1_4
1_4_5
1_5
2
2_3
2_3_4
2_3_4_5
2_3_5
2_4
2_4_5
2_5
3
3_4
3_4_5
3_5
4
4_5
5
NA

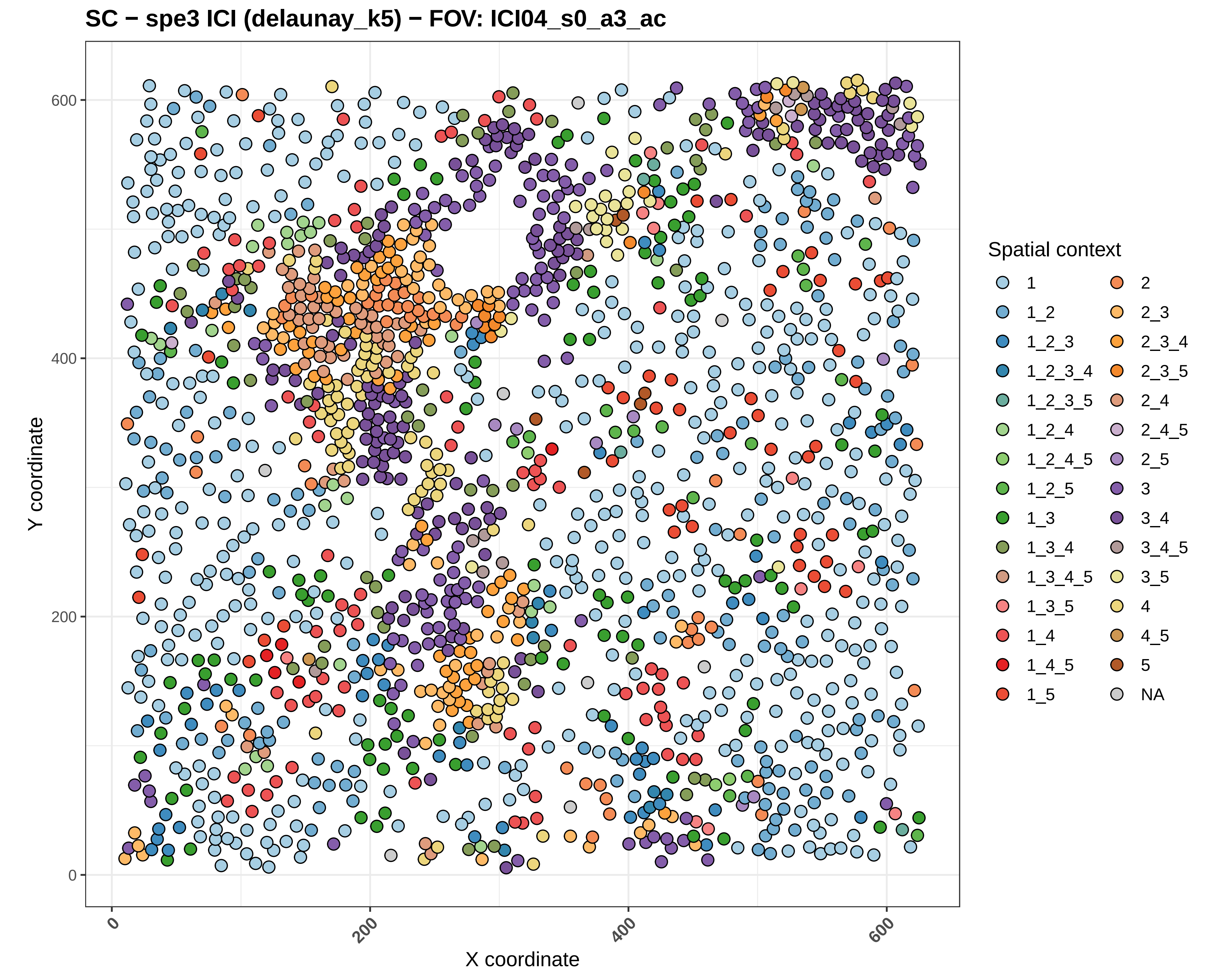
SC − spe3 ICI (delaunay_k5) − FOV: ICI04_s0_a3_ac
Y coordinate
X coordinate
0
200
400
600
Spatial context
1
1_2
1_2_3
1_2_3_4
1_2_3_5
1_2_4
1_2_4_5
1_2_5
1_3
1_3_4
1_3_4_5
1_3_5
1_4
1_4_5
1_5
2
2_3
2_3_4
2_3_5
2_4
2_4_5
2_5
3
3_4
3_4_5
3_5
4
4_5
5
NA

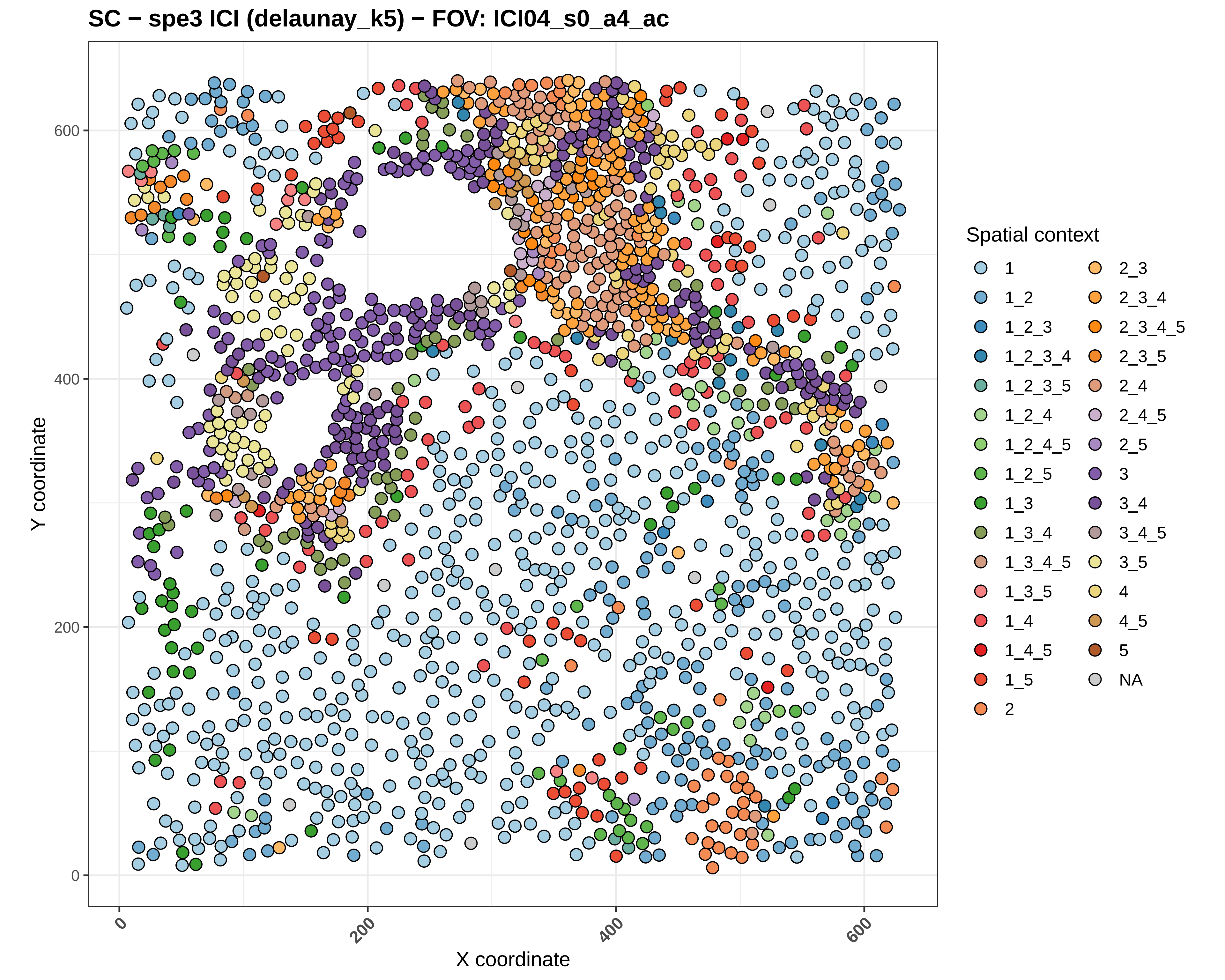
SC − spe3 ICI (delaunay_k5) − FOV: ICI04_s0_a4_ac
Y coordinate
X coordinate
0
200
400
600
Spatial context
1
1_2
1_2_3
1_2_3_4
1_2_3_5
1_2_4
1_2_4_5
1_2_5
1_3
1_3_4
1_3_4_5
1_3_5
1_4
1_4_5
1_5
2
2_3
2_3_4
2_3_4_5
2_3_5
2_4
2_4_5
2_5
3
3_4
3_4_5
3_5
4
4_5
5
NA

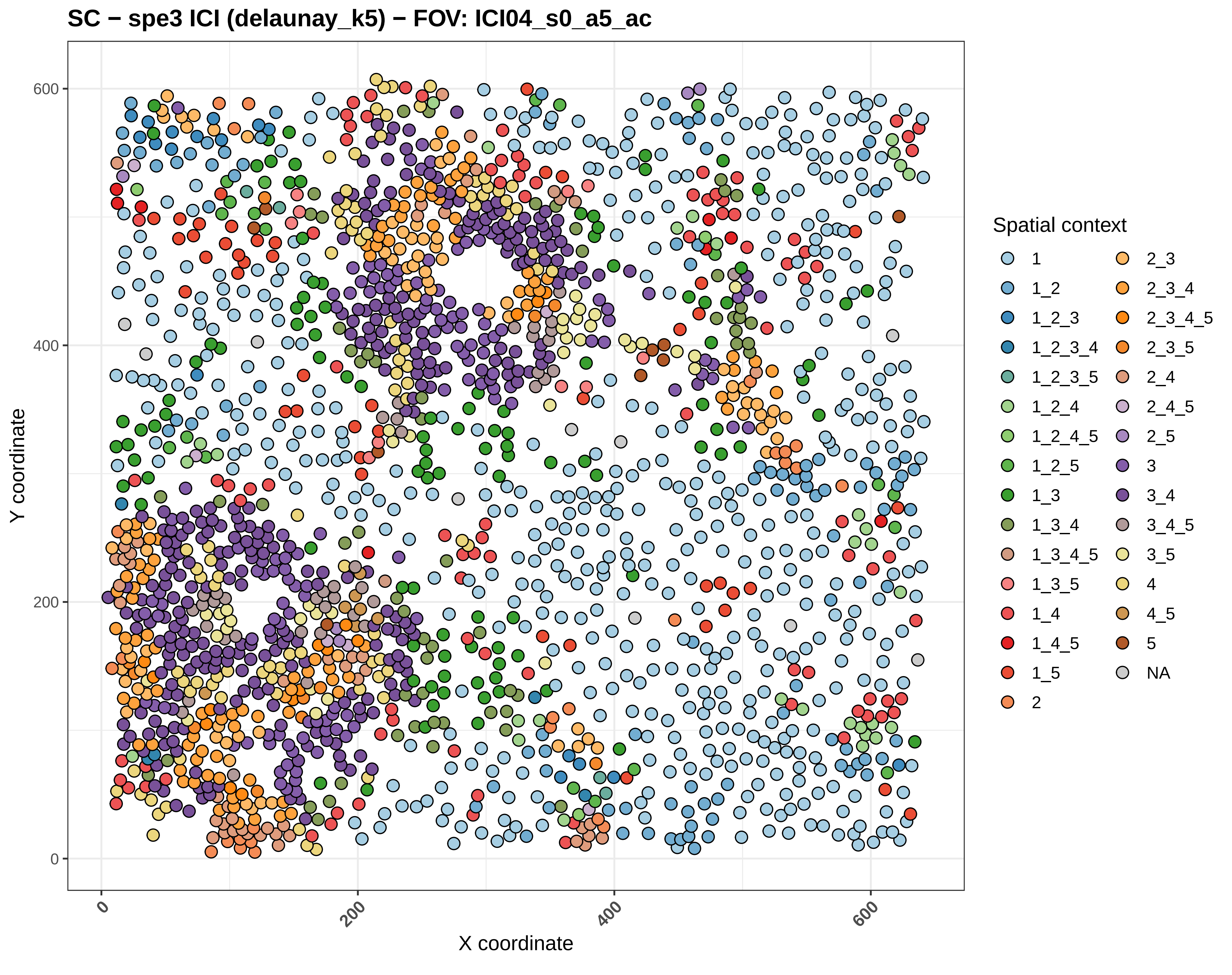

SC − spe3 ICI (delaunay_k5) − FOV: ICI04_s0_a5_ac
Y coordinate
X coordinate
0
200
400
600
Spatial context
1
1_2
1_2_3
1_2_3_4
1_2_3_5
1_2_4
1_2_4_5
1_2_5
1_3
1_3_4
1_3_4_5
1_3_5
1_4
1_4_5
1_5
2
2_3
2_3_4
2_3_4_5
2_3_5
2_4
2_4_5
2_5
3
3_4
3_4_5
3_5
4
4_5
5
NA

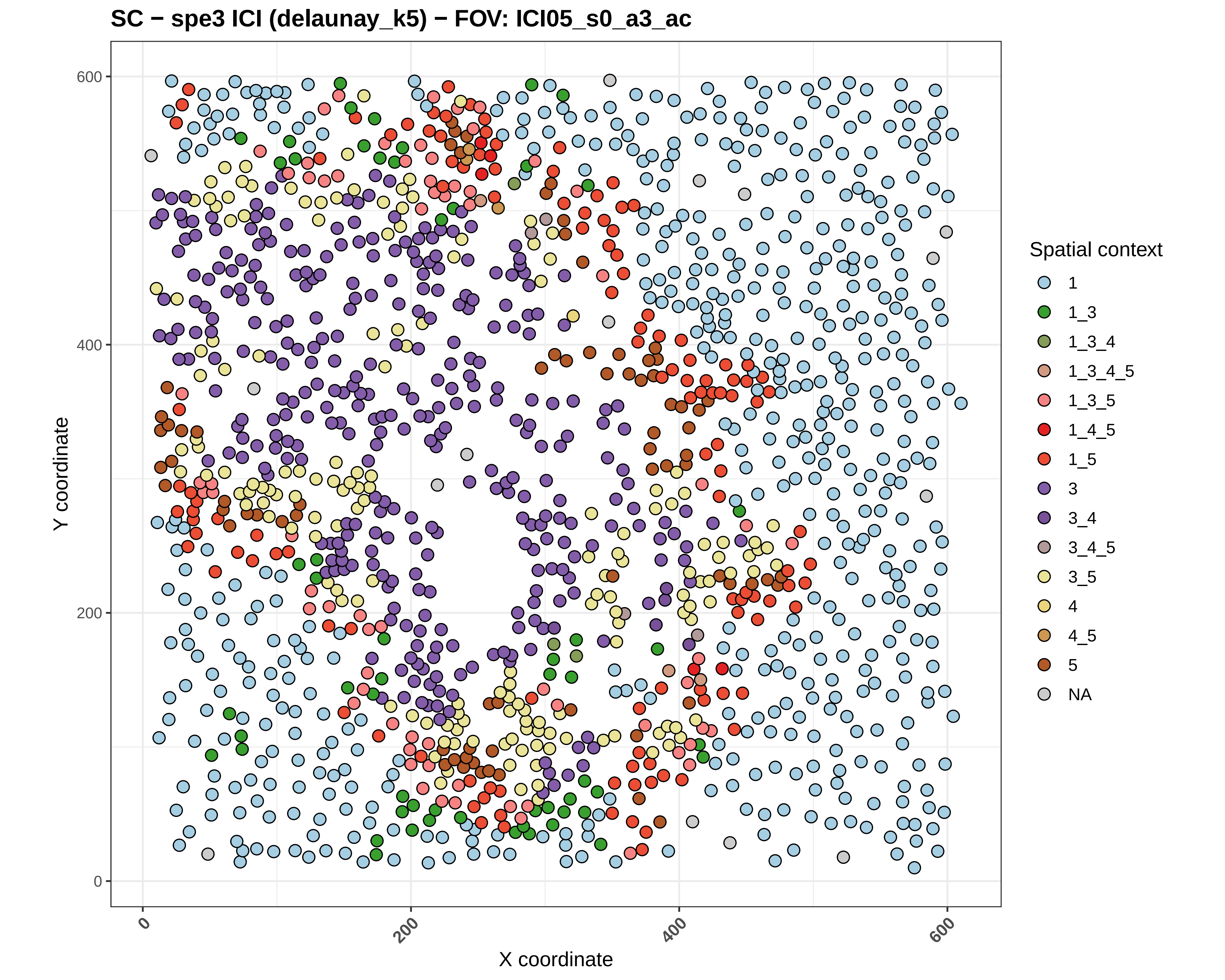

SC − spe3 ICI (delaunay_k5) − FOV: ICI05_s0_a3_ac
Y coordinate
X coordinate
0
200
400
600
0
200
400
600
Spatial context
1
1_3
1_3_4
1_3_4_5
1_3_5
1_4_5
1_5
3
3_4
3_4_5
3_5
4
4_5
5
NA

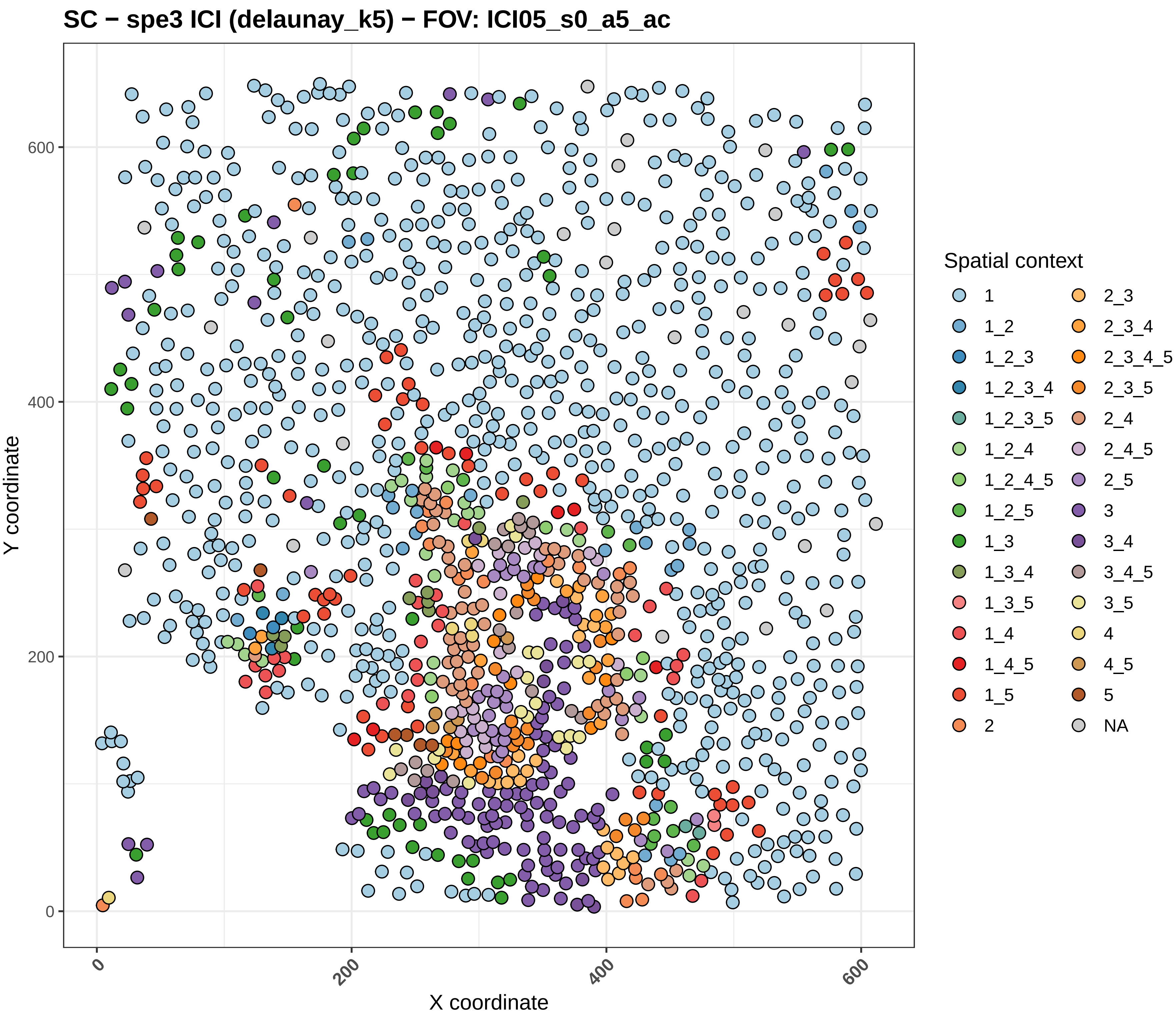
SC − spe3 ICI (delaunay_k5) − FOV: ICI05_s0_a5_ac
Y coordinate
X coordinate
0
200
400
600
Spatial context
1
1_2
1_2_3
1_2_3_4
1_2_3_5
1_2_4
1_2_4_5
1_2_5
1_3
1_3_4
1_3_5
1_4
1_4_5
1_5
2
2_3
2_3_4
2_3_4_5
2_3_5
2_4
2_4_5
2_5
3
3_4
3_4_5
3_5
4
4_5
5
NA

#### SC − spe3 ICI (delaunay_k5) − FOV: ICI06_s0_a1_ac

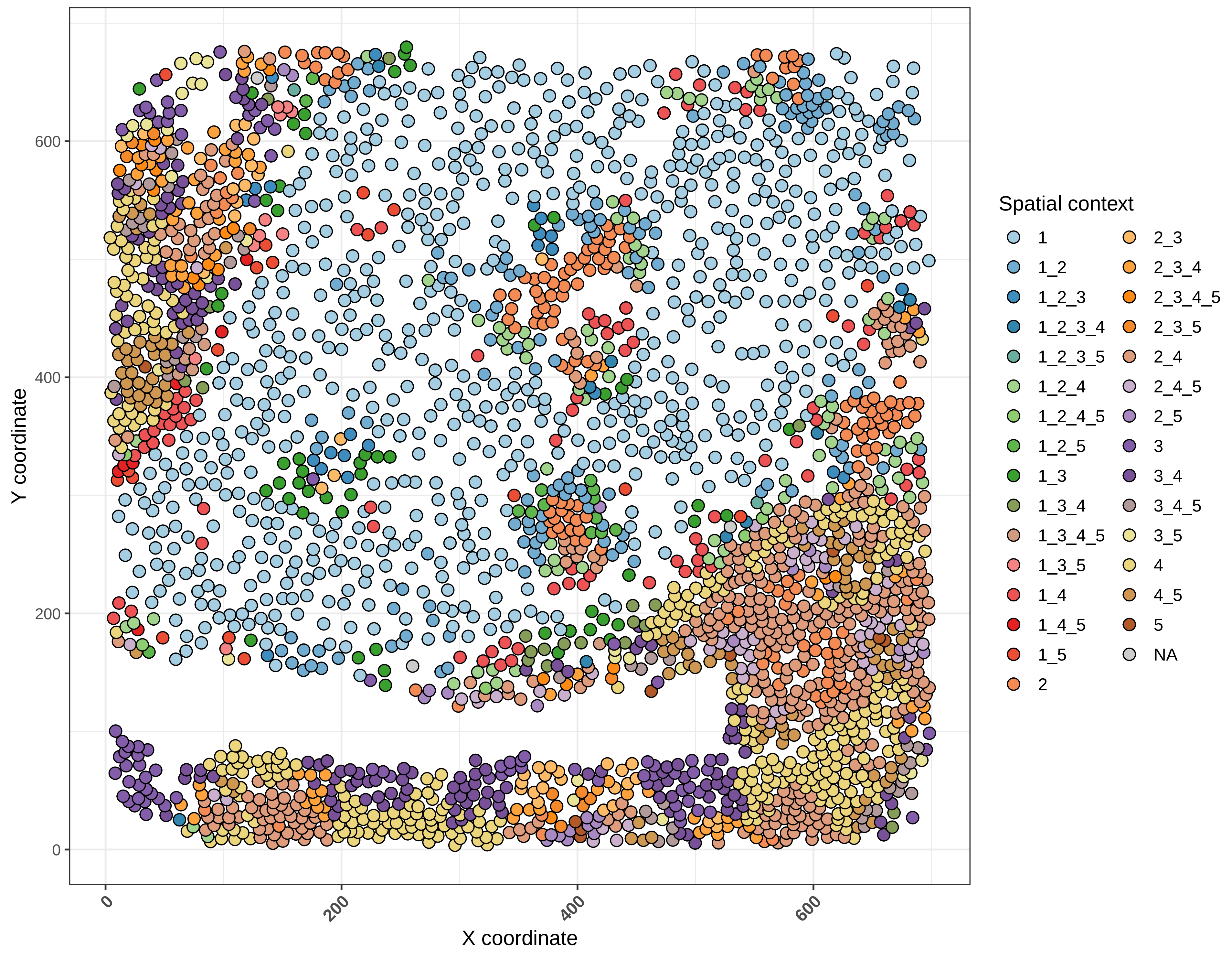

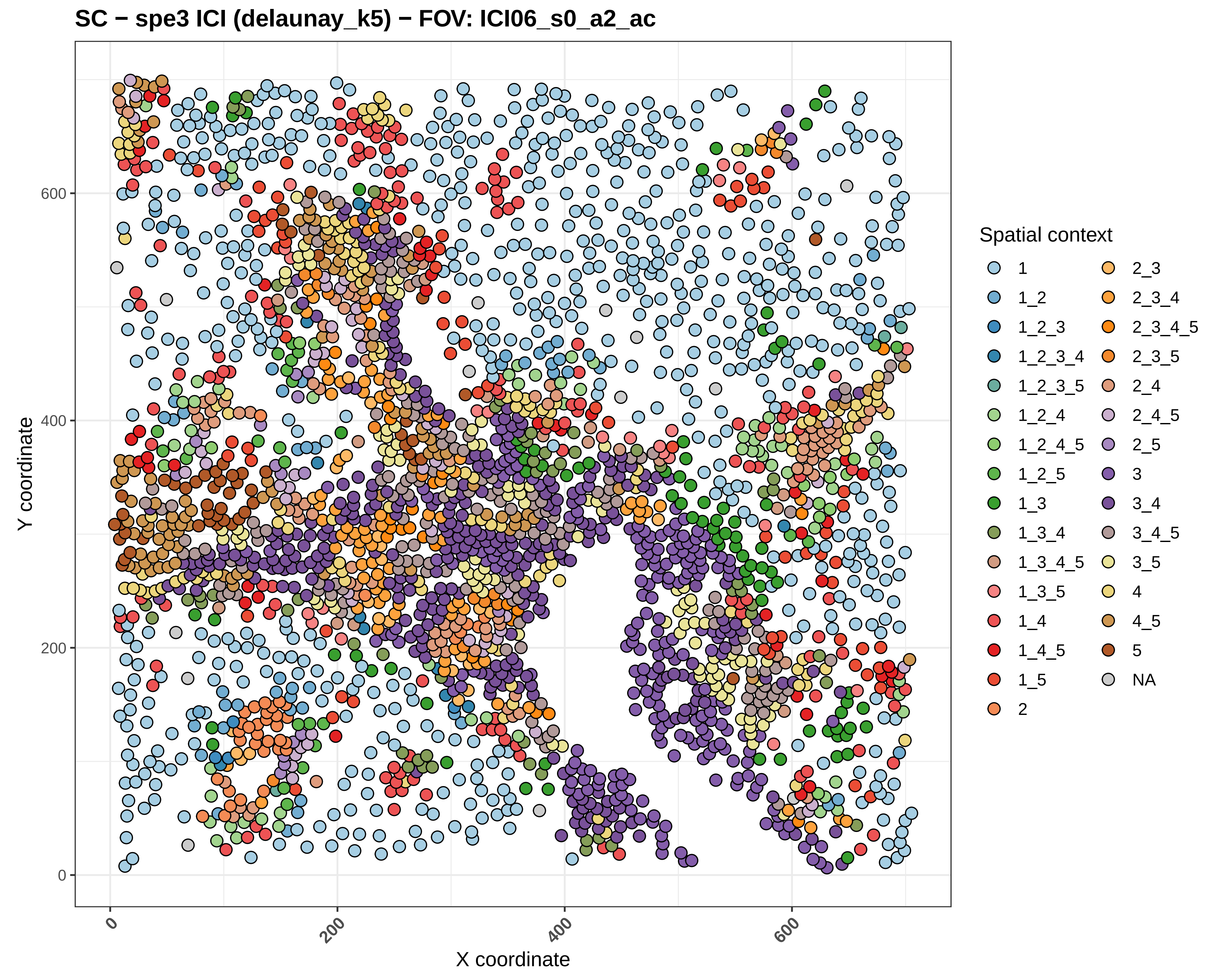
SC − spe3 ICI (delaunay_k5) − FOV: ICI06_s0_a2_ac
Y coordinate
X coordinate
0
200
400
600
Spatial context
1
1_2
1_2_3
1_2_3_4
1_2_3_5
1_2_4
1_2_4_5
1_2_5
1_3
1_3_4
1_3_4_5
1_3_5
1_4
1_4_5
1_5
2
2_3
2_3_4
2_3_4_5
2_3_5
2_4
2_4_5
2_5
3
3_4
3_4_5
3_5
4
4_5
5
NA

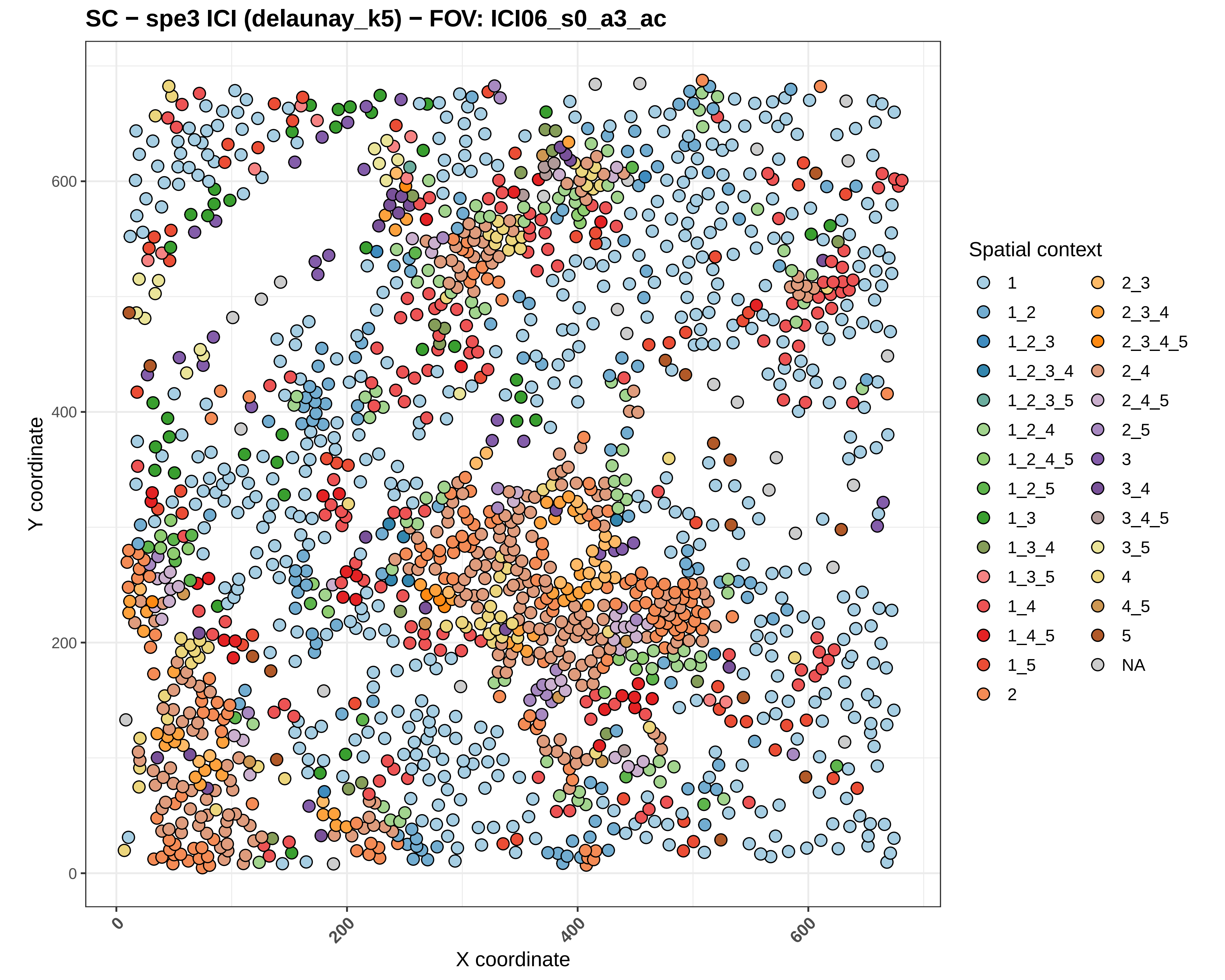
SC − spe3 ICI (delaunay_k5) − FOV: ICI06_s0_a3_ac
Y coordinate
X coordinate
0
200
400
600
Spatial context
1
1_2
1_2_3
1_2_3_4
1_2_3_5
1_2_4
1_2_4_5
1_2_5
1_3
1_3_4
1_3_5
1_4
1_4_5
1_5
2
2_3
2_3_4
2_3_4_5
2_4
2_4_5
2_5
3
3_4
3_4_5
3_5
4
4_5
5
NA

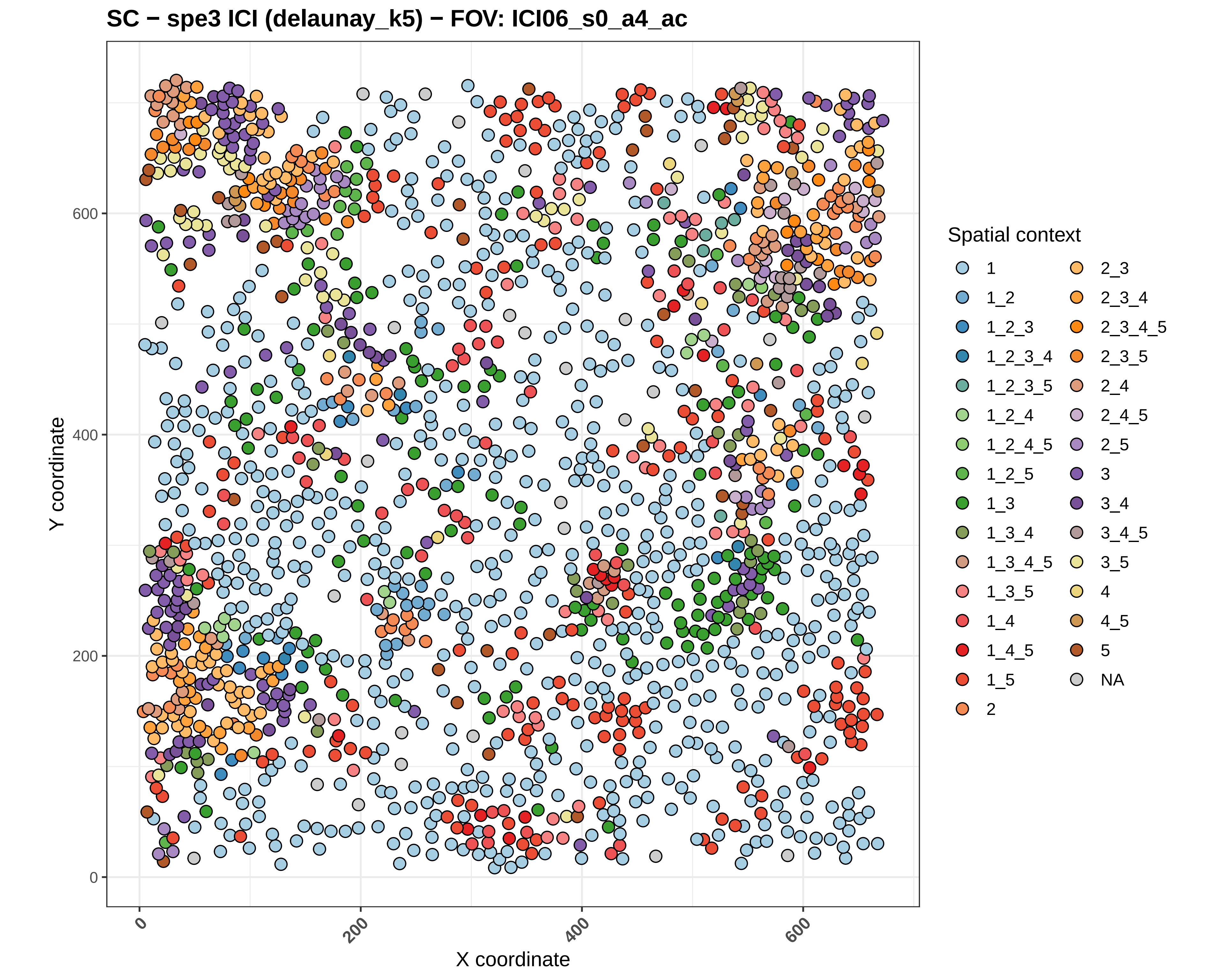

SC − spe3 ICI (delaunay_k5) − FOV: ICI06_s0_a4_ac
Y coordinate
X coordinate
0
200
400
600
Spatial context
1
1_2
1_2_3
1_2_3_4
1_2_3_5
1_2_4
1_2_4_5
1_2_5
1_3
1_3_4
1_3_4_5
1_3_5
1_4
1_4_5
1_5
2
2_3
2_3_4
2_3_4_5
2_3_5
2_4
2_4_5
2_5
3
3_4
3_4_5
3_5
4
4_5
5
NA

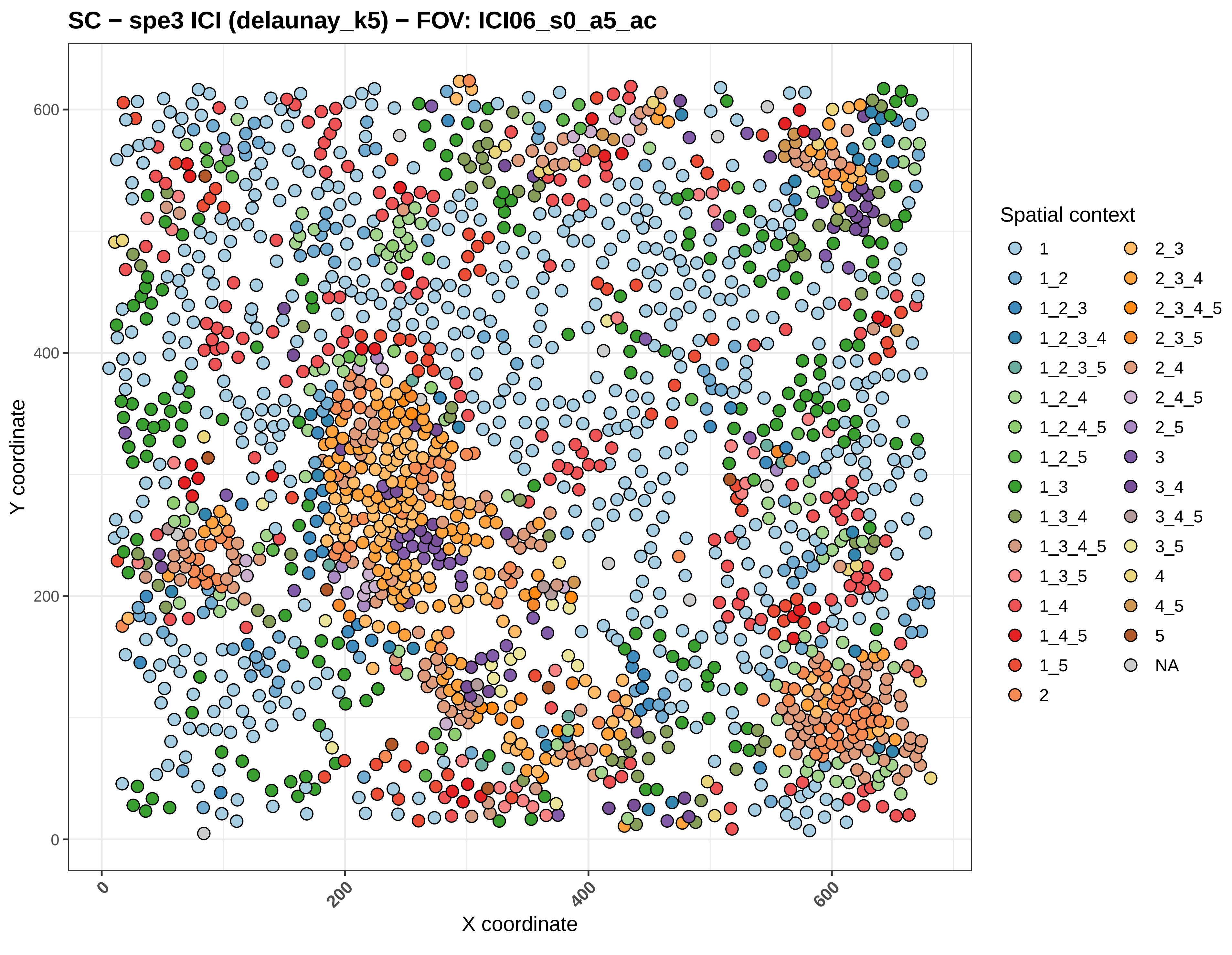

SC − spe3 ICI (delaunay_k5) − FOV: ICI06_s0_a5_ac
Y coordinate
X coordinate
0
200
400
600
Spatial context
1
1_2
1_2_3
1_2_3_4
1_2_3_5
1_2_4
1_2_4_5
1_2_5
1_3
1_3_4
1_3_4_5
1_3_5
1_4
1_4_5
1_5
2
2_3
2_3_4
2_3_4_5
2_3_5
2_4
2_4_5
2_5
3
3_4
3_4_5
3_5
4
4_5
5
NA

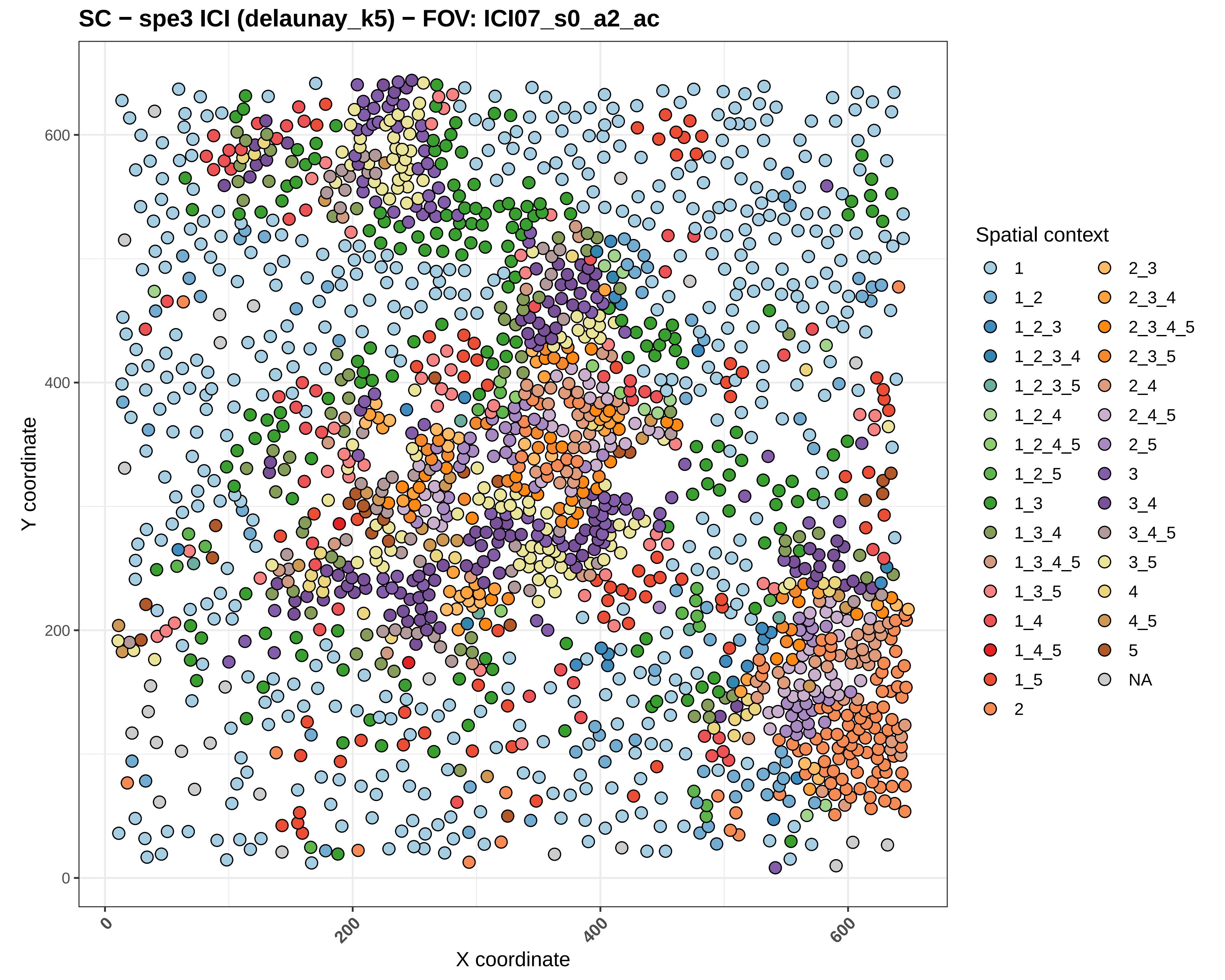

SC − spe3 ICI (delaunay_k5) − FOV: ICI07_s0_a2_ac
Y coordinate
X coordinate
0
200
400
600
Spatial context
1
1_2
1_2_3
1_2_3_4
1_2_3_5
1_2_4
1_2_4_5
1_2_5
1_3
1_3_4
1_3_4_5
1_3_5
1_4
1_4_5
1_5
2
2_3
2_3_4
2_3_4_5
2_3_5
2_4
2_4_5
2_5
3
3_4
3_4_5
3_5
4
4_5
5
NA

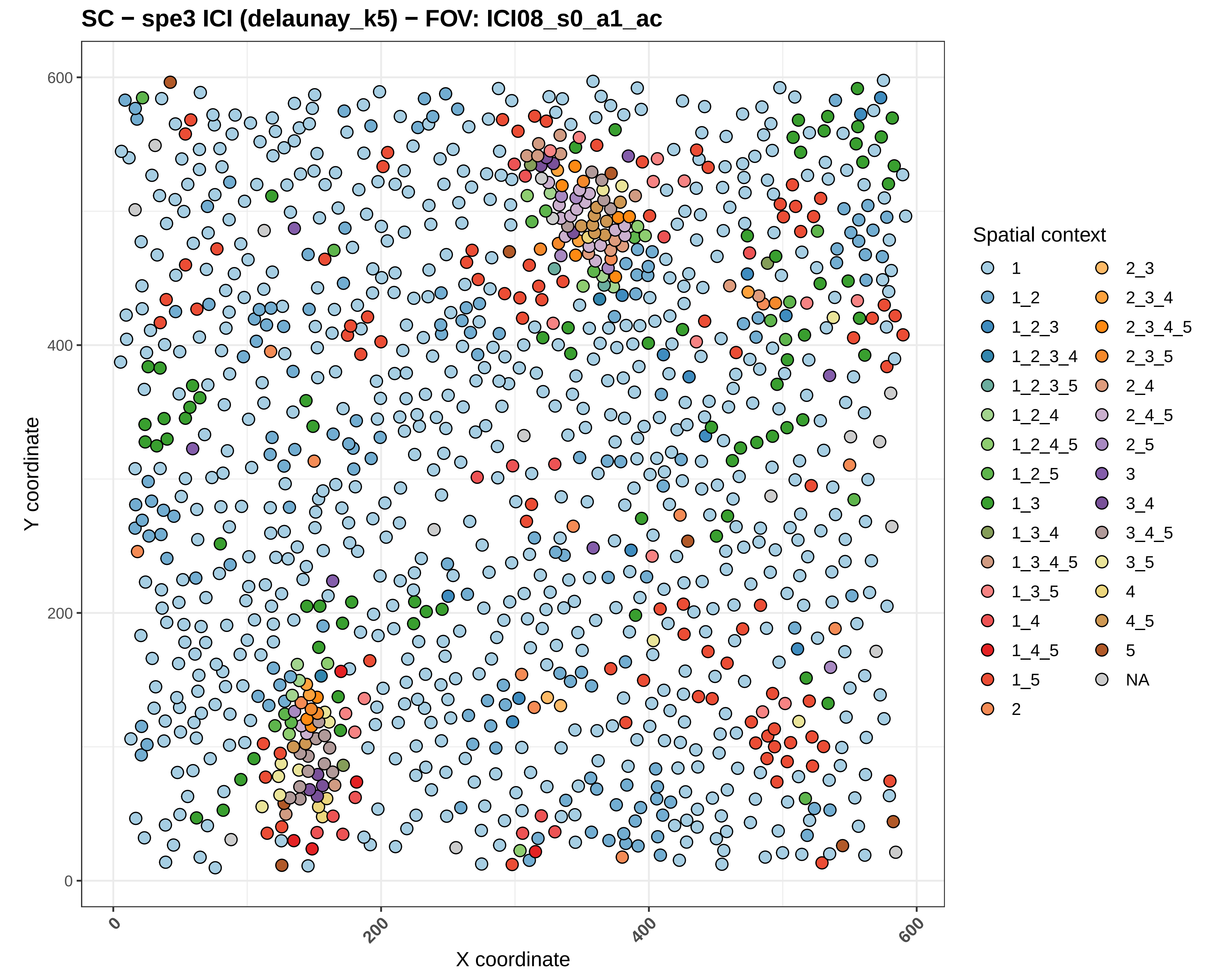
SC − spe3 ICI (delaunay_k5) − FOV: ICI08_s0_a1_ac
Y coordinate
X coordinate
0
200
400
600
0
200
400
600
Spatial context
1
1_2
1_2_3
1_2_3_4
1_2_3_5
1_2_4
1_2_4_5
1_2_5
1_3
1_3_4
1_3_4_5
1_3_5
1_4
1_4_5
1_5
2
2_3
2_3_4
2_3_4_5
2_3_5
2_4
2_4_5
2_5
3
3_4
3_4_5
3_5
4
4_5
5
NA

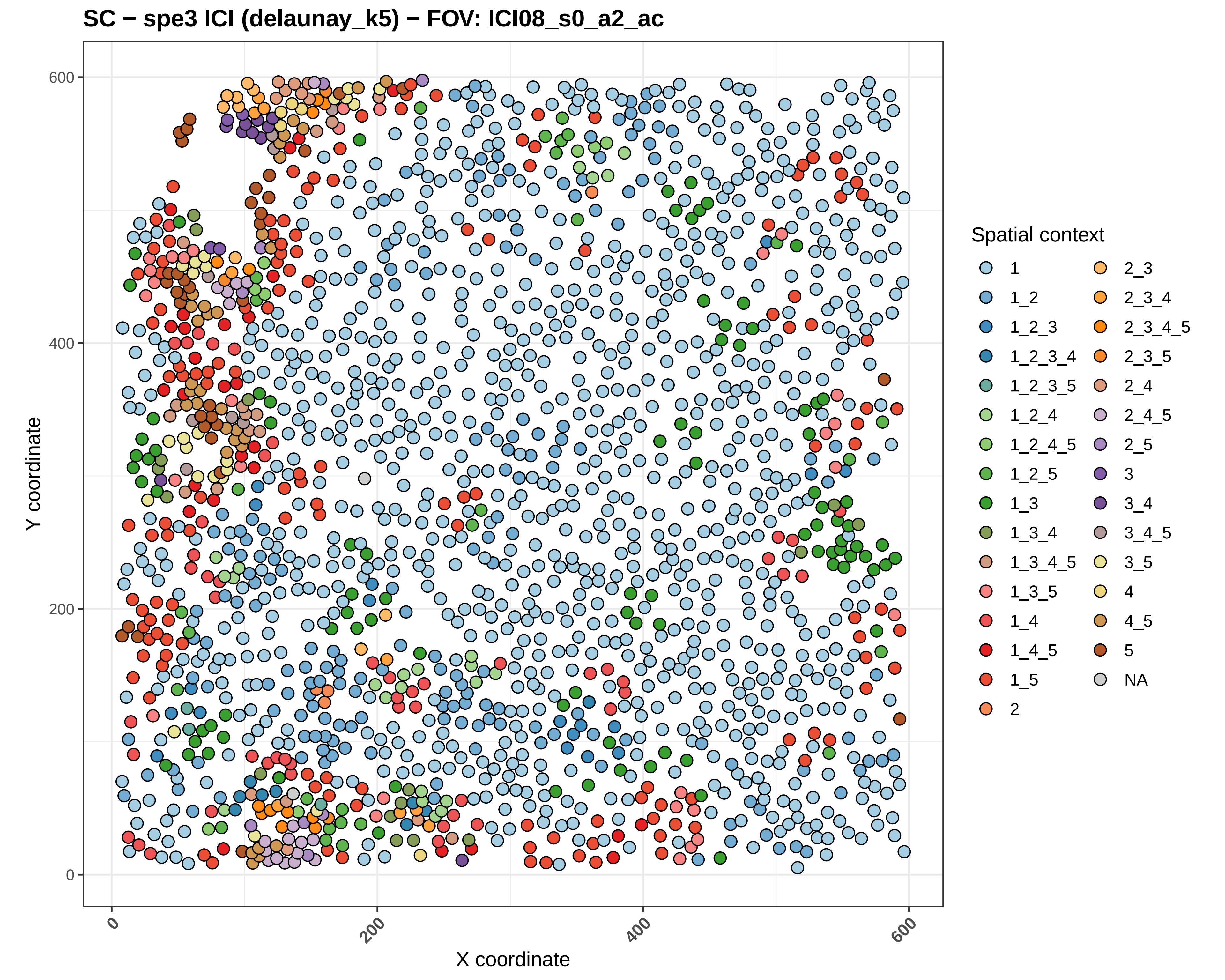
SC − spe3 ICI (delaunay_k5) − FOV: ICI08_s0_a2_ac
Y coordinate
X coordinate
0
200
400
600
Spatial context
1
1_2
1_2_3
1_2_3_4
1_2_3_5
1_2_4
1_2_4_5
1_2_5
1_3
1_3_4
1_3_4_5
1_3_5
1_4
1_4_5
1_5
2
2_3
2_3_4
2_3_4_5
2_3_5
2_4
2_4_5
2_5
3
3_4
3_4_5
3_5
4
4_5
5
NA

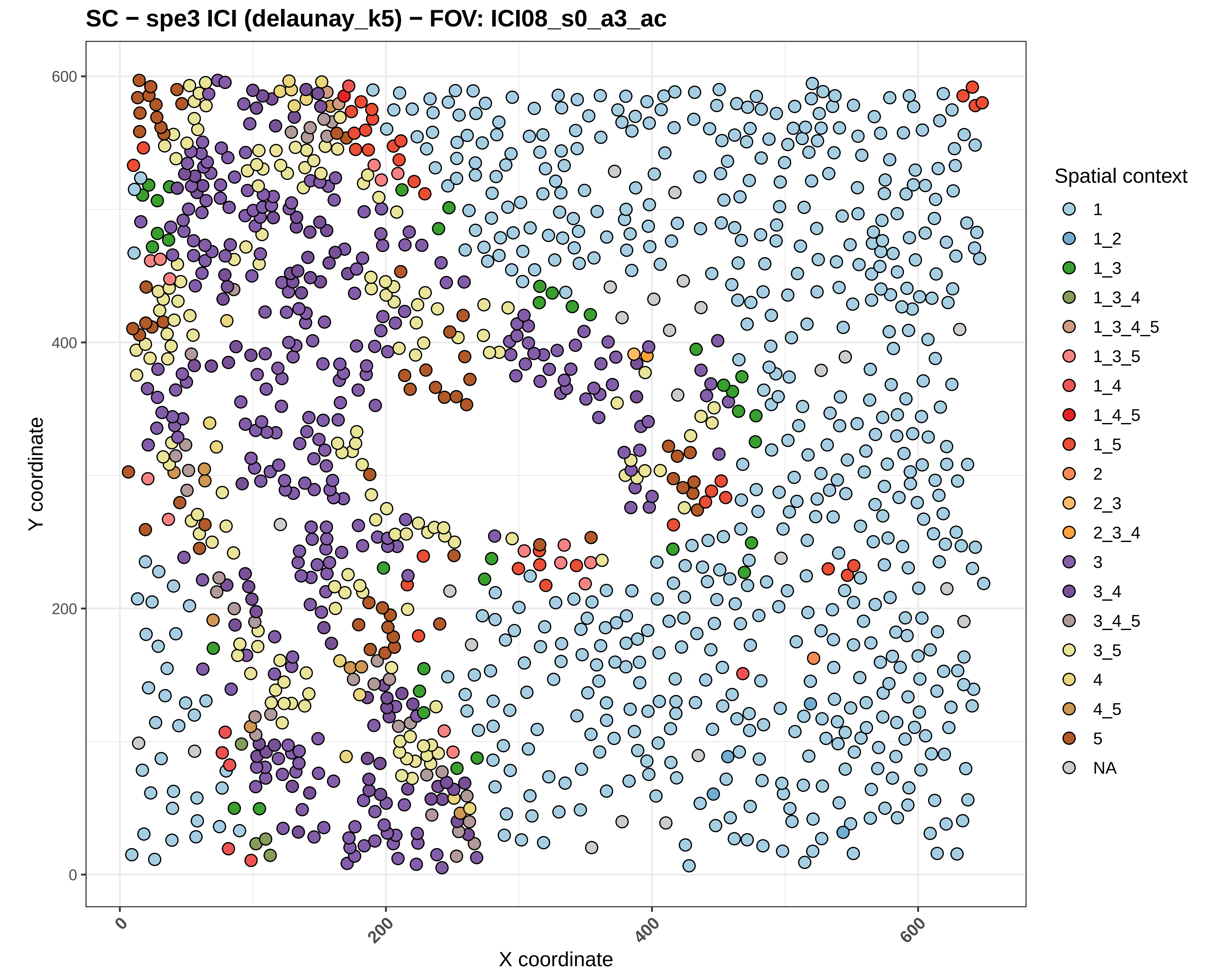

SC − spe3 ICI (delaunay_k5) − FOV: ICI08_s0_a3_ac
Y coordinate
X coordinate
0
200
400
600
Spatial context
1
1_2
1_3
1_3_4
1_3_4_5
1_3_5
1_4
1_4_5
1_5
2
2_3
2_3_4
3
3_4
3_4_5
3_5
4
4_5
5
NA

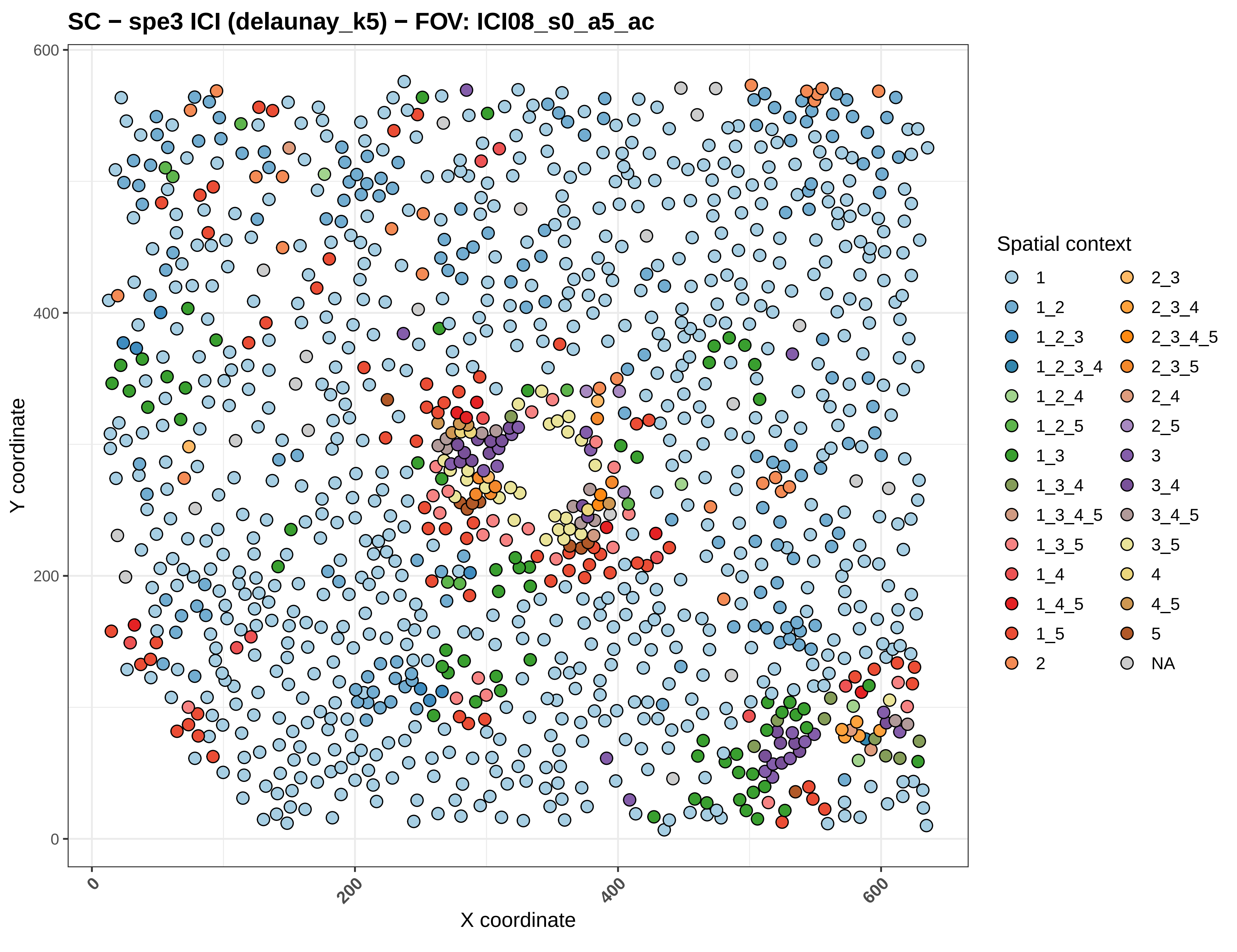

SC − spe3 ICI (delaunay_k5) − FOV: ICI08_s0_a5_ac
Y coordinate
X coordinate
0
200
400
600
Spatial context
1
1_2
1_2_3
1_2_3_4
1_2_4
1_2_5
1_3
1_3_4
1_3_4_5
1_3_5
1_4
1_4_5
1_5
2
2_3
2_3_4
2_3_4_5
2_3_5
2_4
2_5
3
3_4
3_4_5
3_5
4
4_5
5
NA

#### SC − spe3 ICI (delaunay_k5) − FOV: ICI09_s0_a1_ac

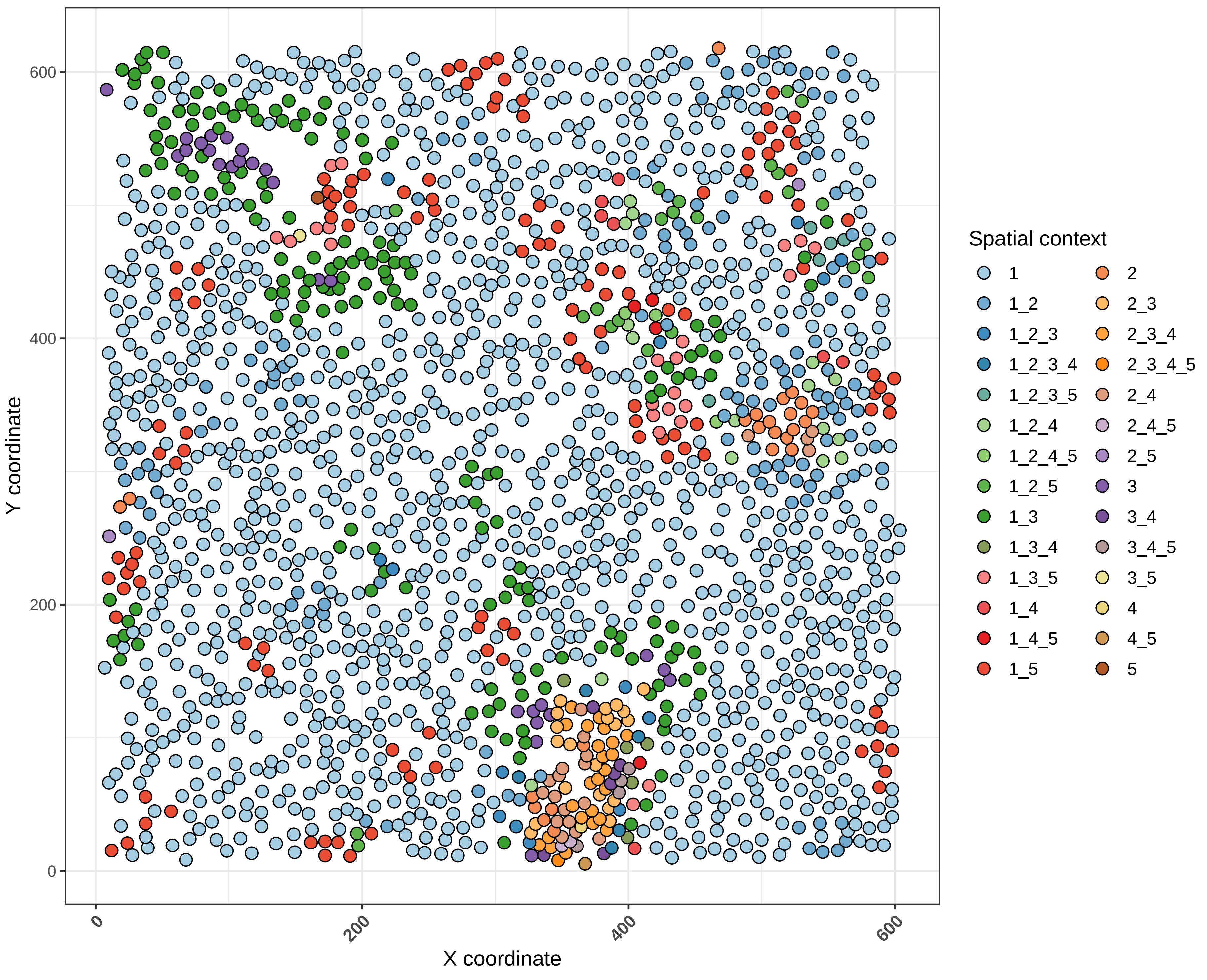

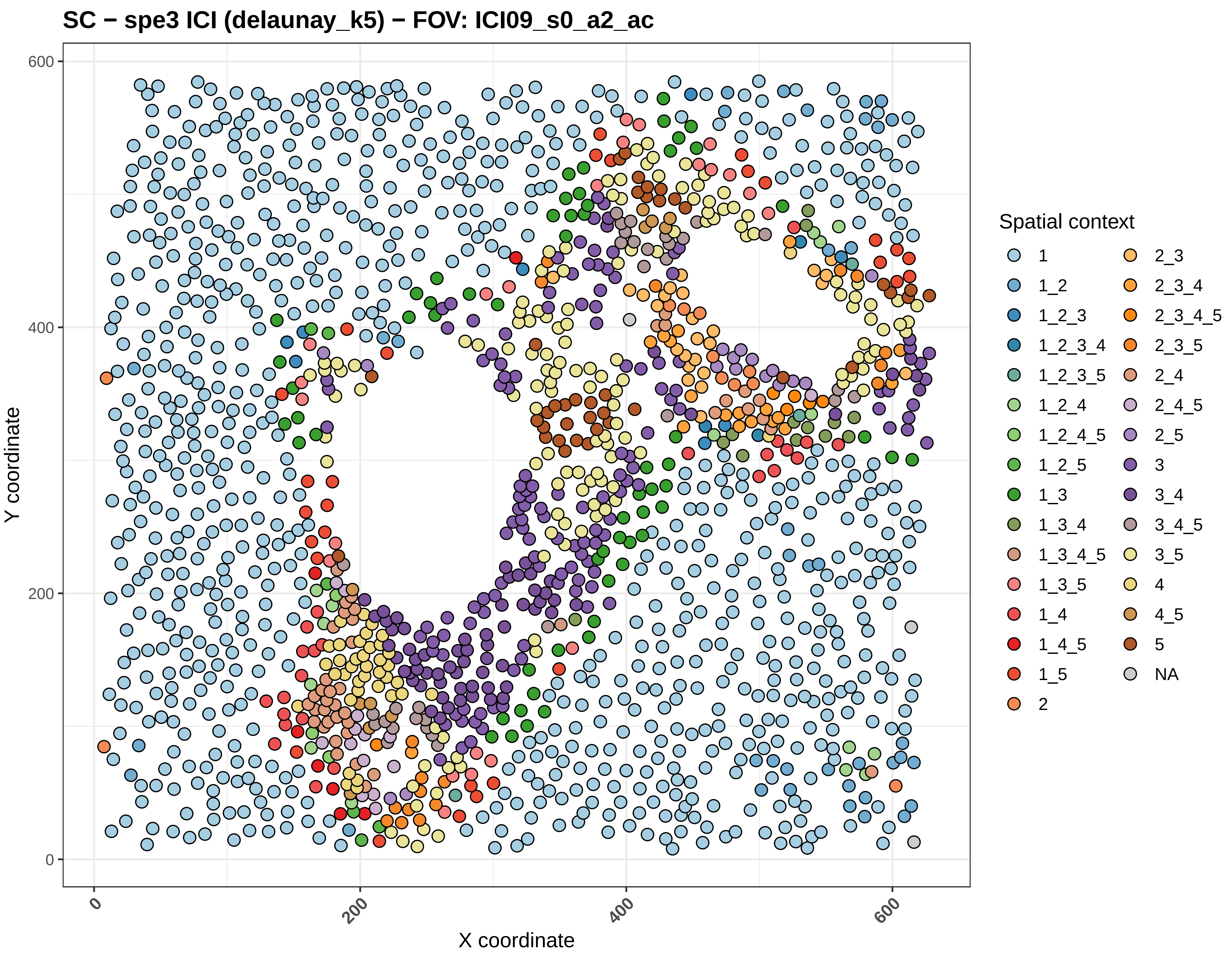

SC − spe3 ICI (delaunay_k5) − FOV: ICI09_s0_a2_ac
Y coordinate
X coordinate
0
200
400
600
Spatial context
1
1_2
1_2_3
1_2_3_4
1_2_3_5
1_2_4
1_2_4_5
1_2_5
1_3
1_3_4
1_3_4_5
1_3_5
1_4
1_4_5
1_5
2
2_3
2_3_4
2_3_4_5
2_3_5
2_4
2_4_5
2_5
3
3_4
3_4_5
3_5
4
4_5
5
NA

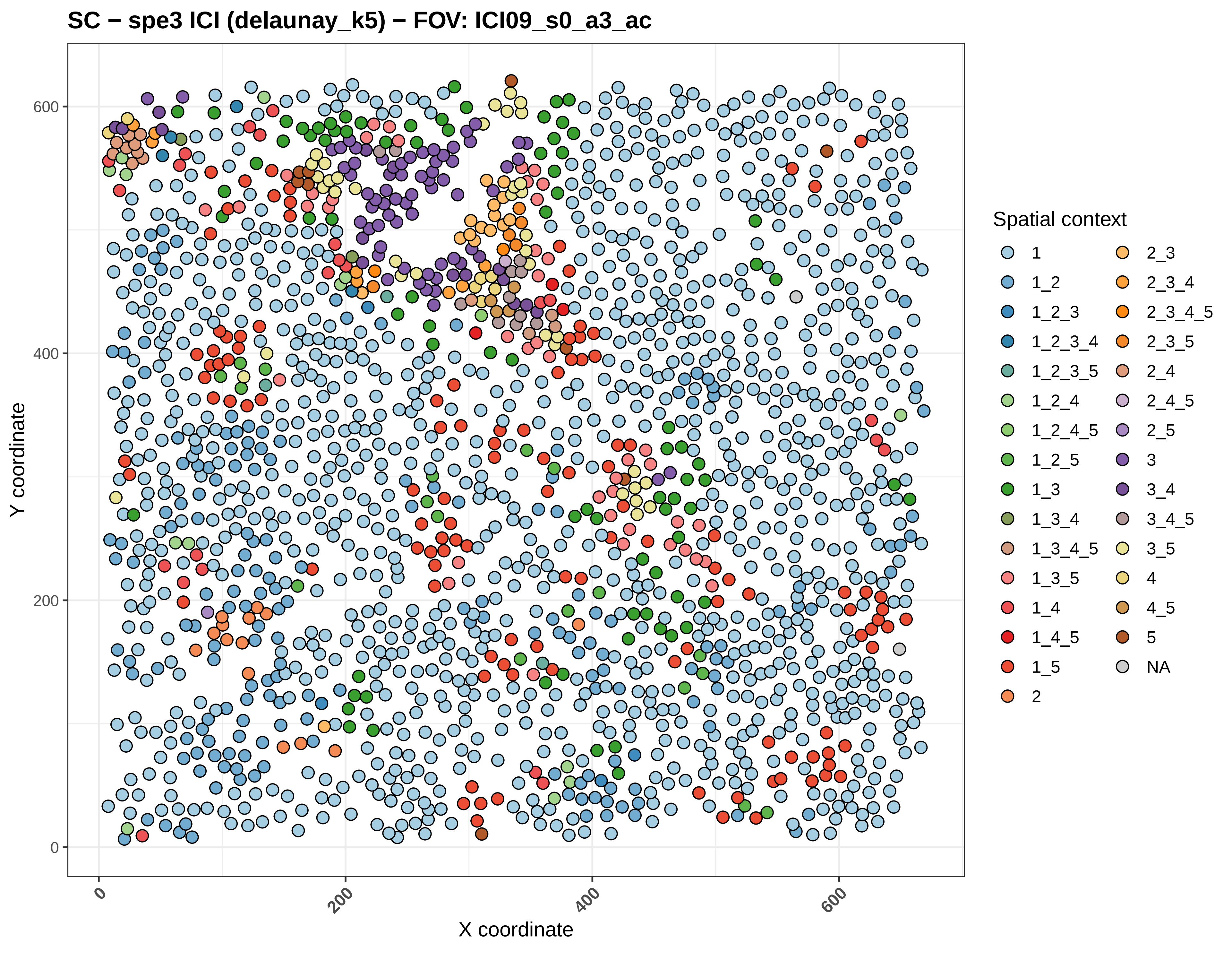

SC − spe3 ICI (delaunay_k5) − FOV: ICI09_s0_a3_ac
Y coordinate
X coordinate
0
200
400
600
Spatial context
1
1_2
1_2_3
1_2_3_4
1_2_3_5
1_2_4
1_2_4_5
1_2_5
1_3
1_3_4
1_3_4_5
1_3_5
1_4
1_4_5
1_5
2
2_3
2_3_4
2_3_4_5
2_3_5
2_4
2_4_5
2_5
3
3_4
3_4_5
3_5
4
4_5
5
NA

#### SC − spe3 ICI (delaunay_k5) − FOV: ICI09_s0_a4_ac

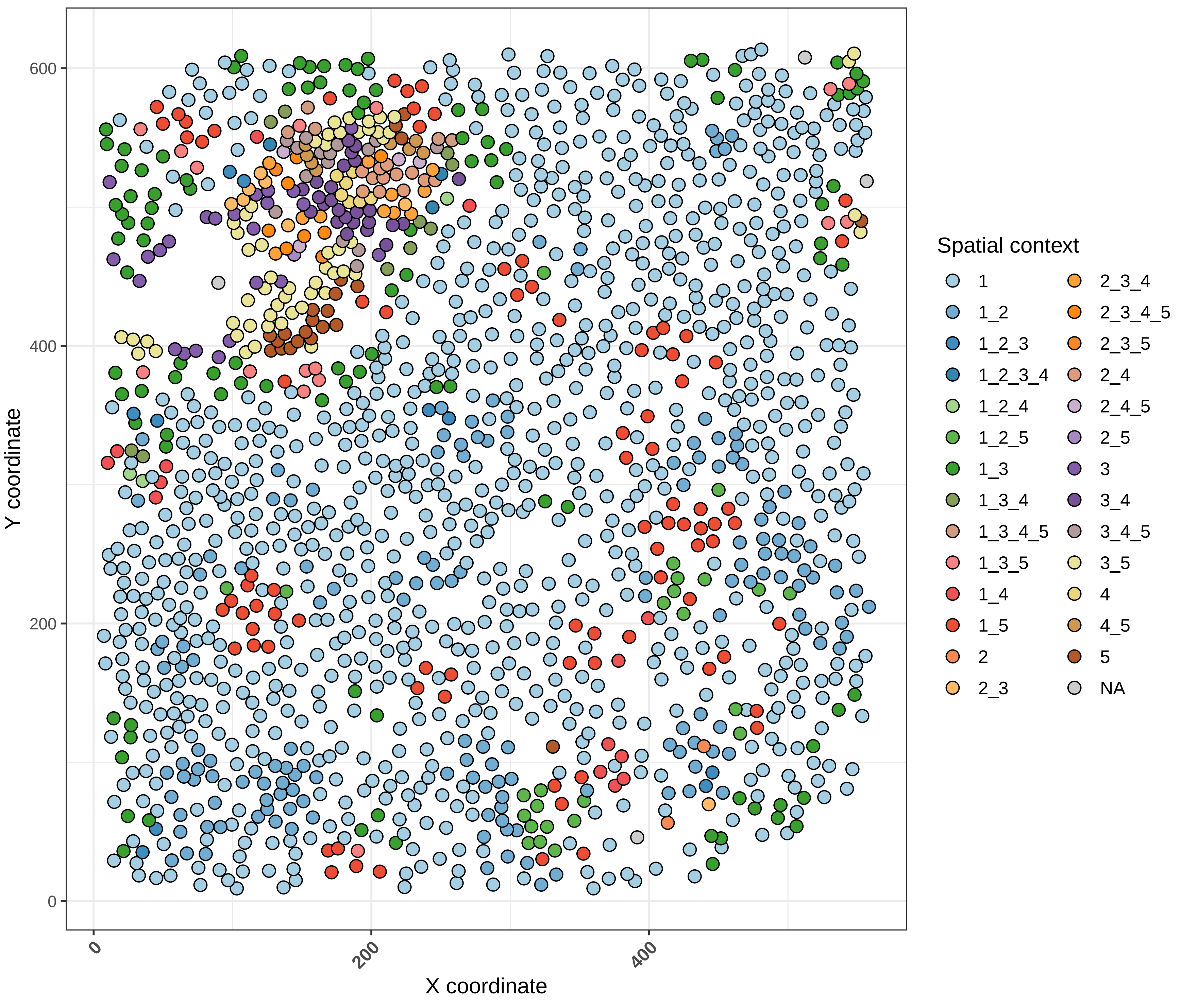

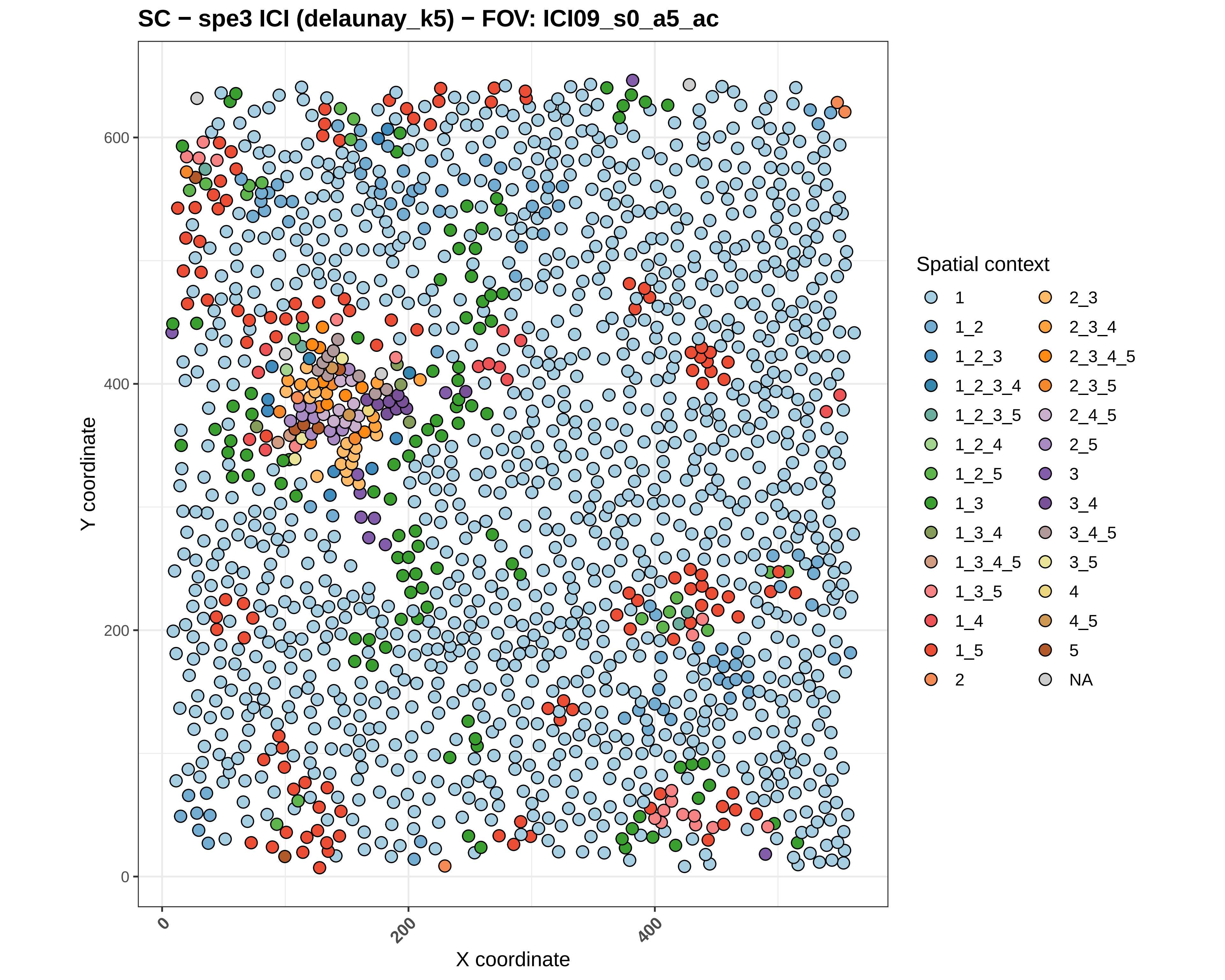

SC − spe3 ICI (delaunay_k5) − FOV: ICI09_s0_a5_ac
Y coordinate
X coordinate
0
200
400
600
Spatial context
1
1_2
1_2_3
1_2_3_4
1_2_3_5
1_2_4
1_2_5
1_3
1_3_4
1_3_4_5
1_3_5
1_4
1_5
2
2_3
2_3_4
2_3_4_5
2_3_5
2_4_5
2_5
3
3_4
3_4_5
3_5
4
4_5
5
NA

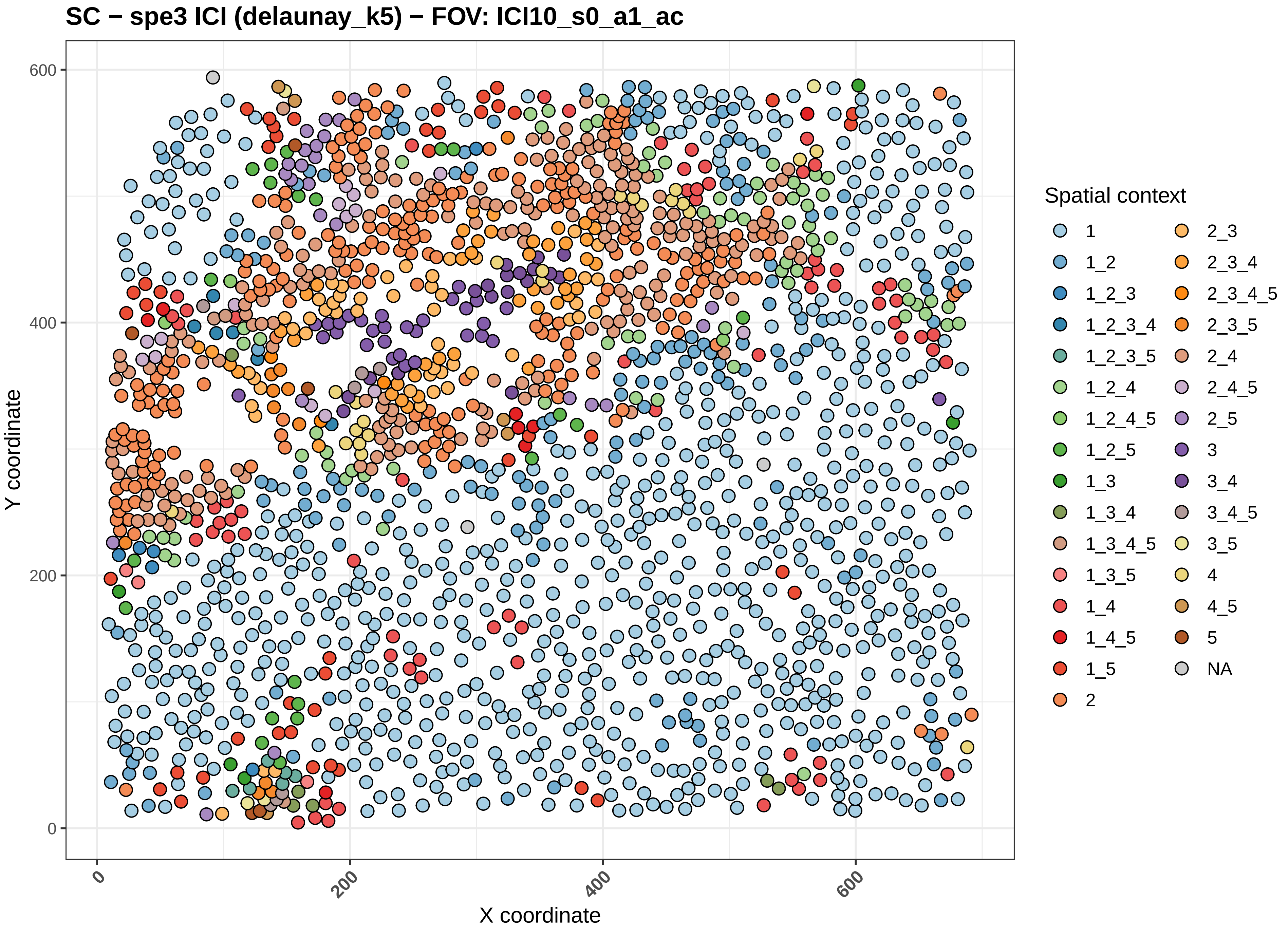
SC − spe3 ICI (delaunay_k5) − FOV: ICI10_s0_a1_ac
Y coordinate
X coordinate
0
200
400
600
Spatial context
1
1_2
1_2_3
1_2_3_4
1_2_3_5
1_2_4
1_2_4_5
1_2_5
1_3
1_3_4
1_3_4_5
1_3_5
1_4
1_4_5
1_5
2
2_3
2_3_4
2_3_4_5
2_3_5
2_4
2_4_5
2_5
3
3_4
3_4_5
3_5
4
4_5
5
NA

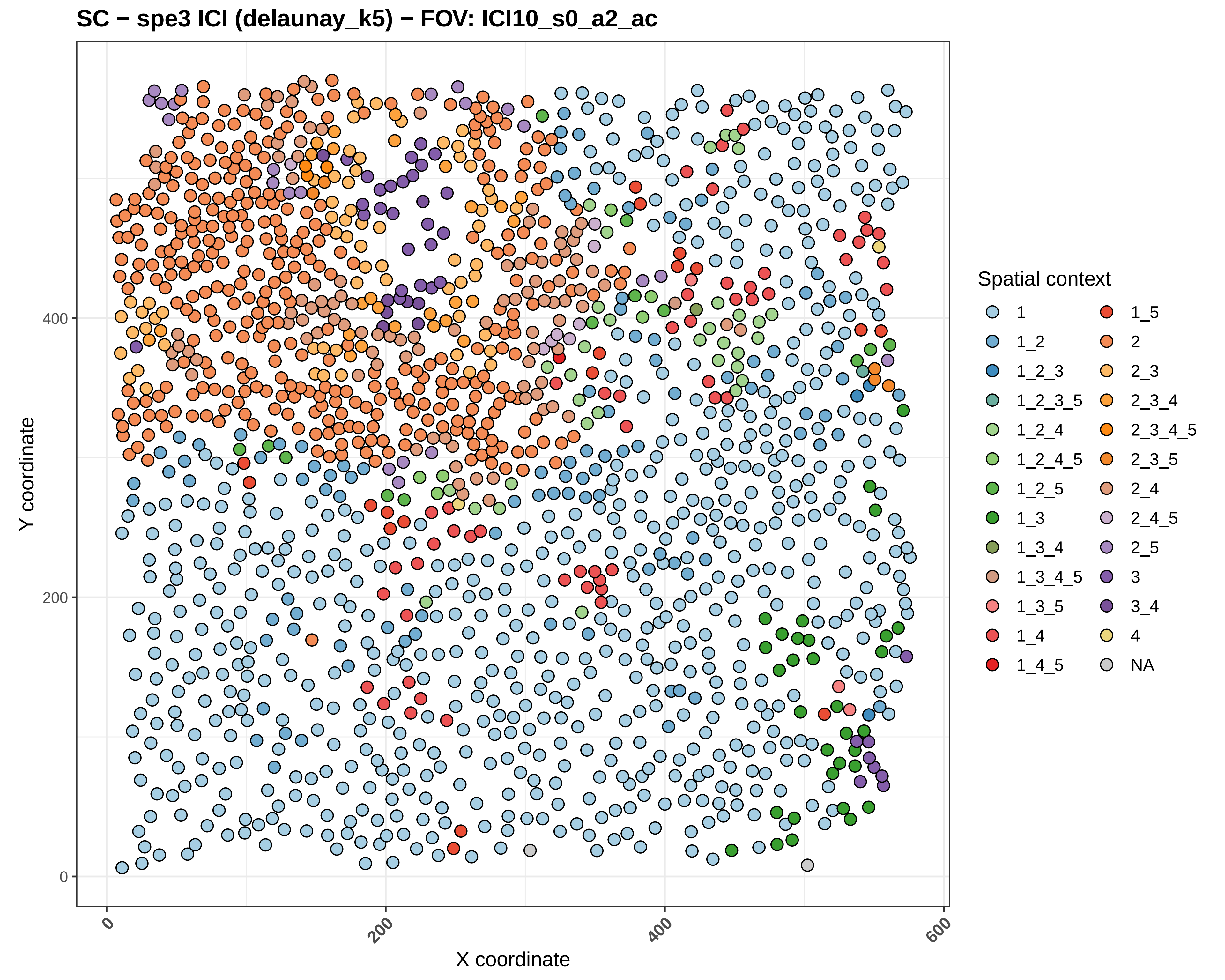
SC − spe3 ICI (delaunay_k5) − FOV: ICI10_s0_a2_ac
Y coordinate
X coordinate
0
200
400
600
Spatial context
1
1_2
1_2_3
1_2_3_5
1_2_4
1_2_4_5
1_2_5
1_3
1_3_4
1_3_4_5
1_3_5
1_4
1_4_5
1_5
2
2_3
2_3_4
2_3_4_5
2_3_5
2_4
2_4_5
2_5
3
3_4
4
NA

SC − spe3 ICI (delaunay_k5) − FOV: ICI10_s0_a3_ac

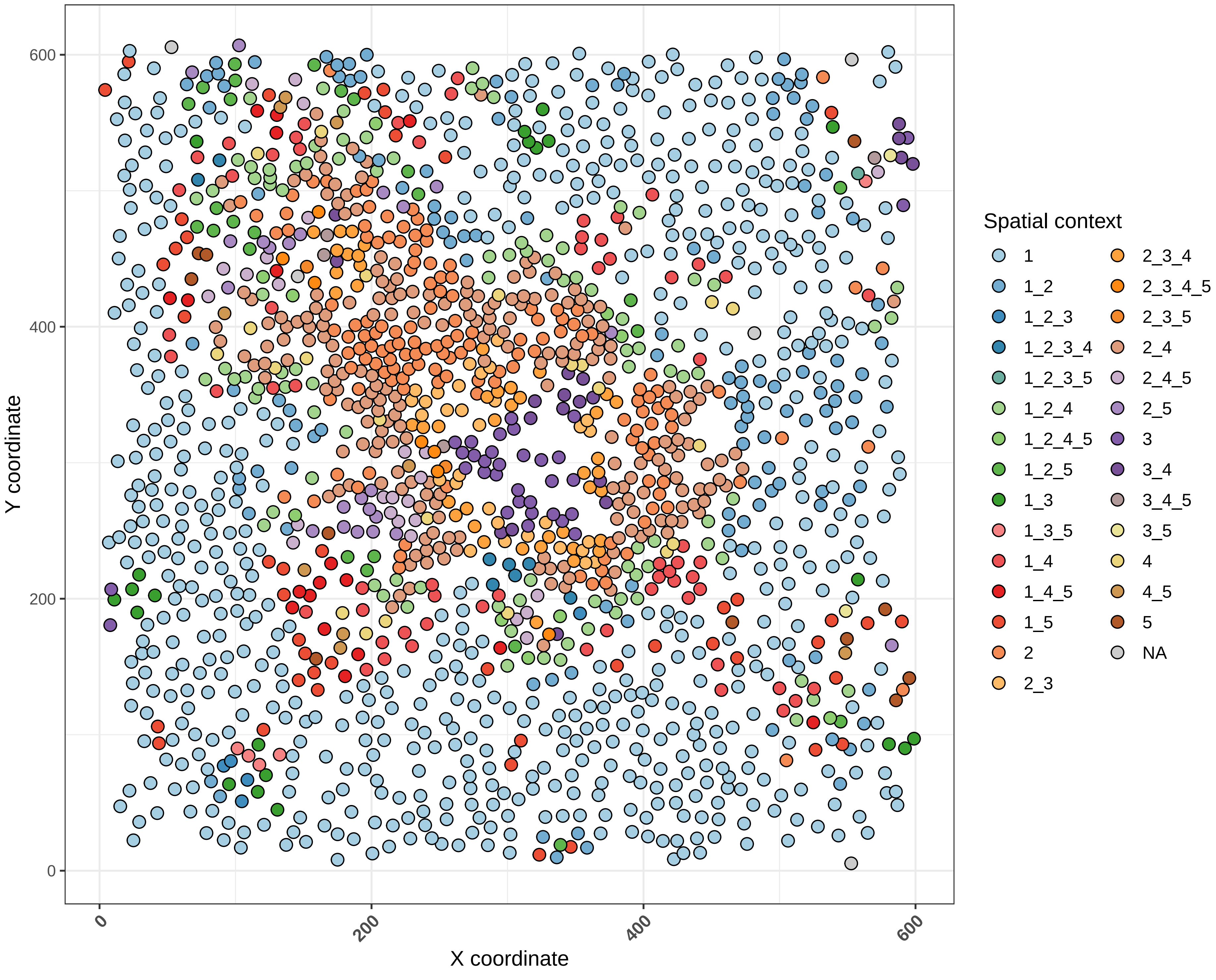

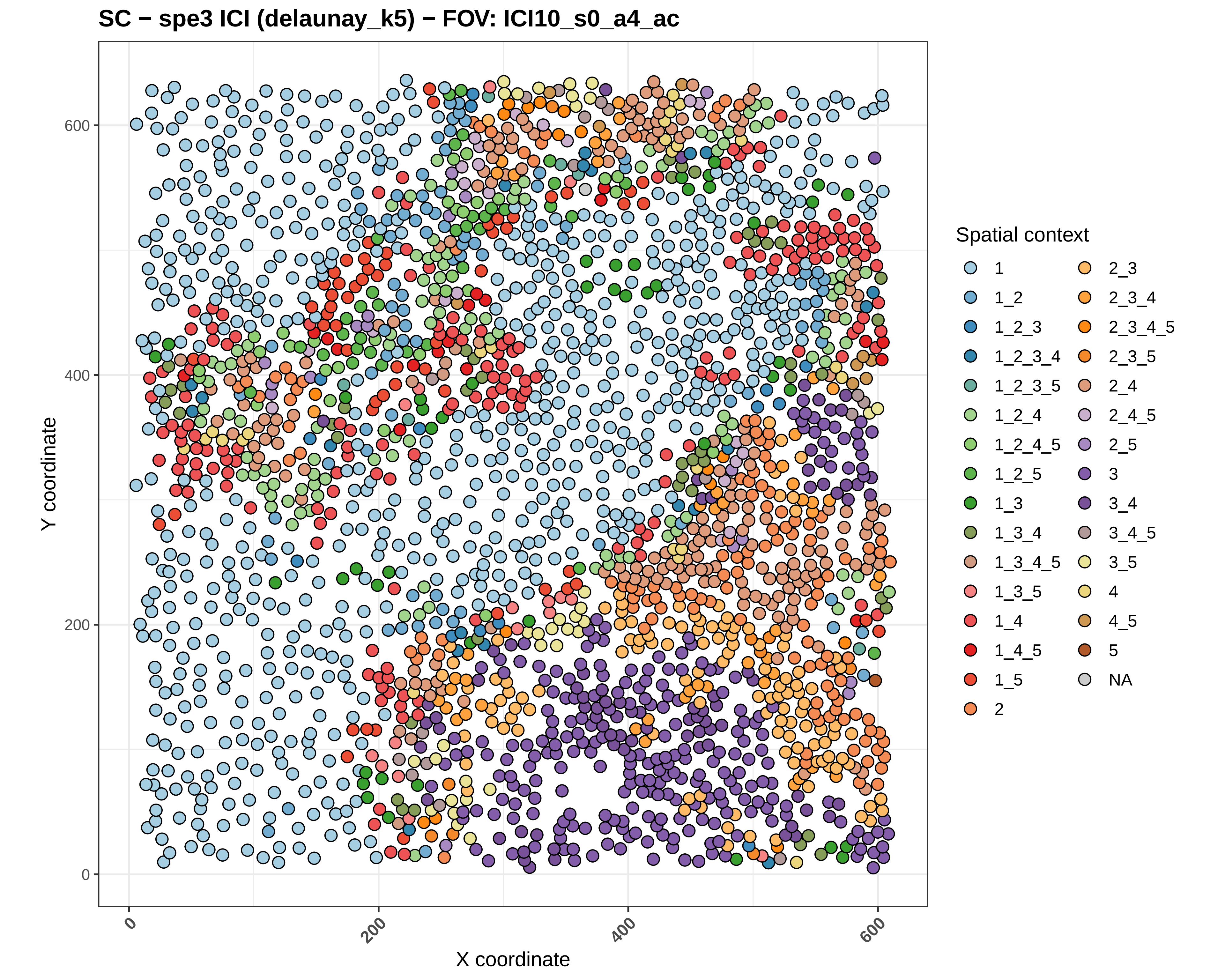

SC − spe3 ICI (delaunay_k5) − FOV: ICI10_s0_a4_ac
Y coordinate
X coordinate
0
200
400
600
Spatial context
1
1_2
1_2_3
1_2_3_4
1_2_3_5
1_2_4
1_2_4_5
1_2_5
1_3
1_3_4
1_3_4_5
1_3_5
1_4
1_4_5
1_5
2
2_3
2_3_4
2_3_4_5
2_3_5
2_4
2_4_5
2_5
3
3_4
3_4_5
3_5
4
4_5
5
NA

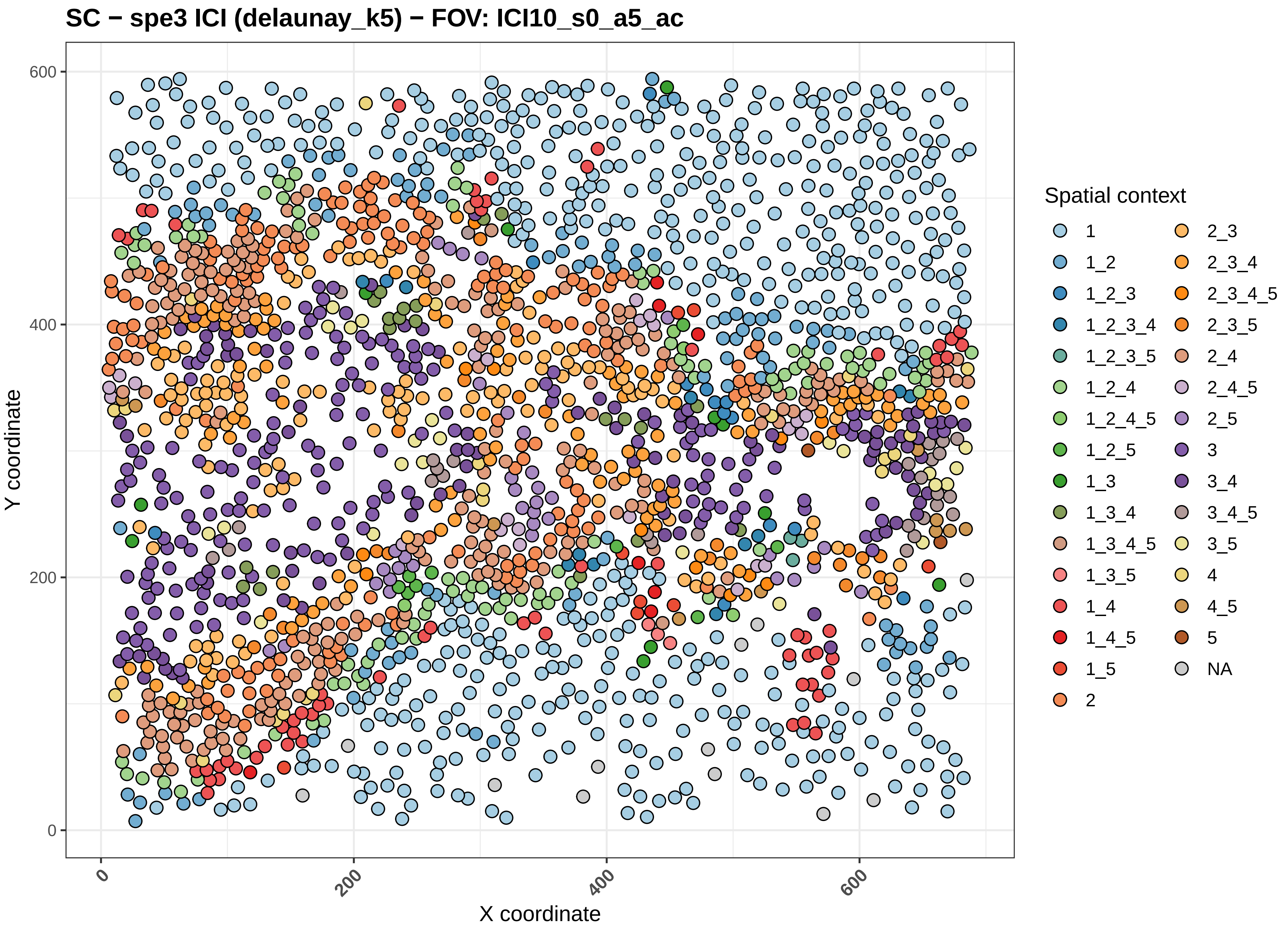
SC − spe3 ICI (delaunay_k5) − FOV: ICI10_s0_a5_ac
Y coordinate
X coordinate
0
200
400
600
0
200
400
600
Spatial context
1
1_2
1_2_3
1_2_3_4
1_2_3_5
1_2_4
1_2_4_5
1_2_5
1_3
1_3_4
1_3_4_5
1_3_5
1_4
1_4_5
1_5
2
2_3
2_3_4
2_3_4_5
2_3_5
2_4
2_4_5
2_5
3
3_4
3_4_5
3_5
4
4_5
5
NA

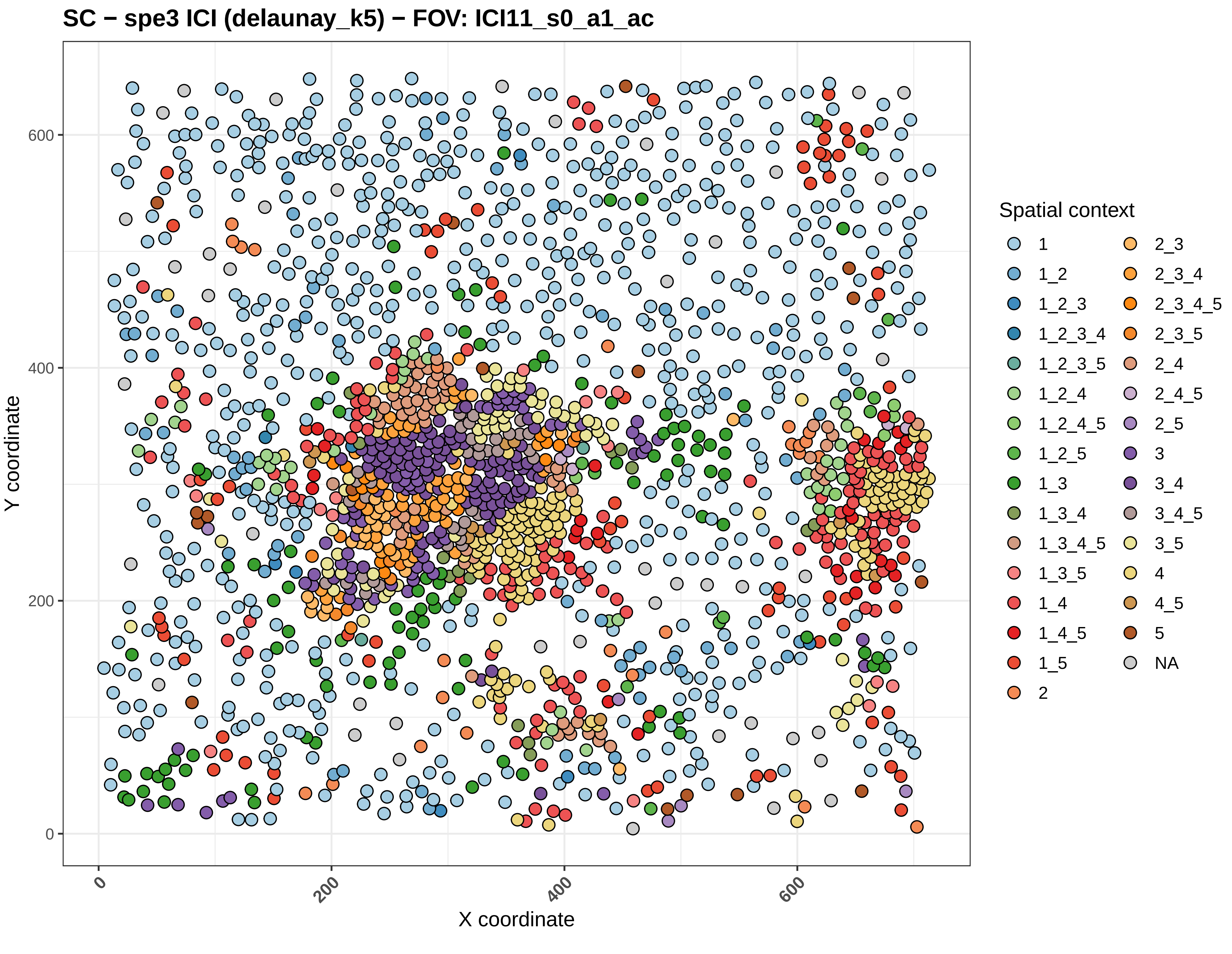
SC − spe3 ICI (delaunay_k5) − FOV: ICI11_s0_a1_ac
Y coordinate
X coordinate
0
200
400
600
Spatial context
1
1_2
1_2_3
1_2_3_4
1_2_3_5
1_2_4
1_2_4_5
1_2_5
1_3
1_3_4
1_3_4_5
1_3_5
1_4
1_4_5
1_5
2
2_3
2_3_4
2_3_4_5
2_3_5
2_4
2_4_5
2_5
3
3_4
3_4_5
3_5
4
4_5
5
NA

SC − spe3 ICI (delaunay_k5) − FOV: ICI11_s0_a4_ac

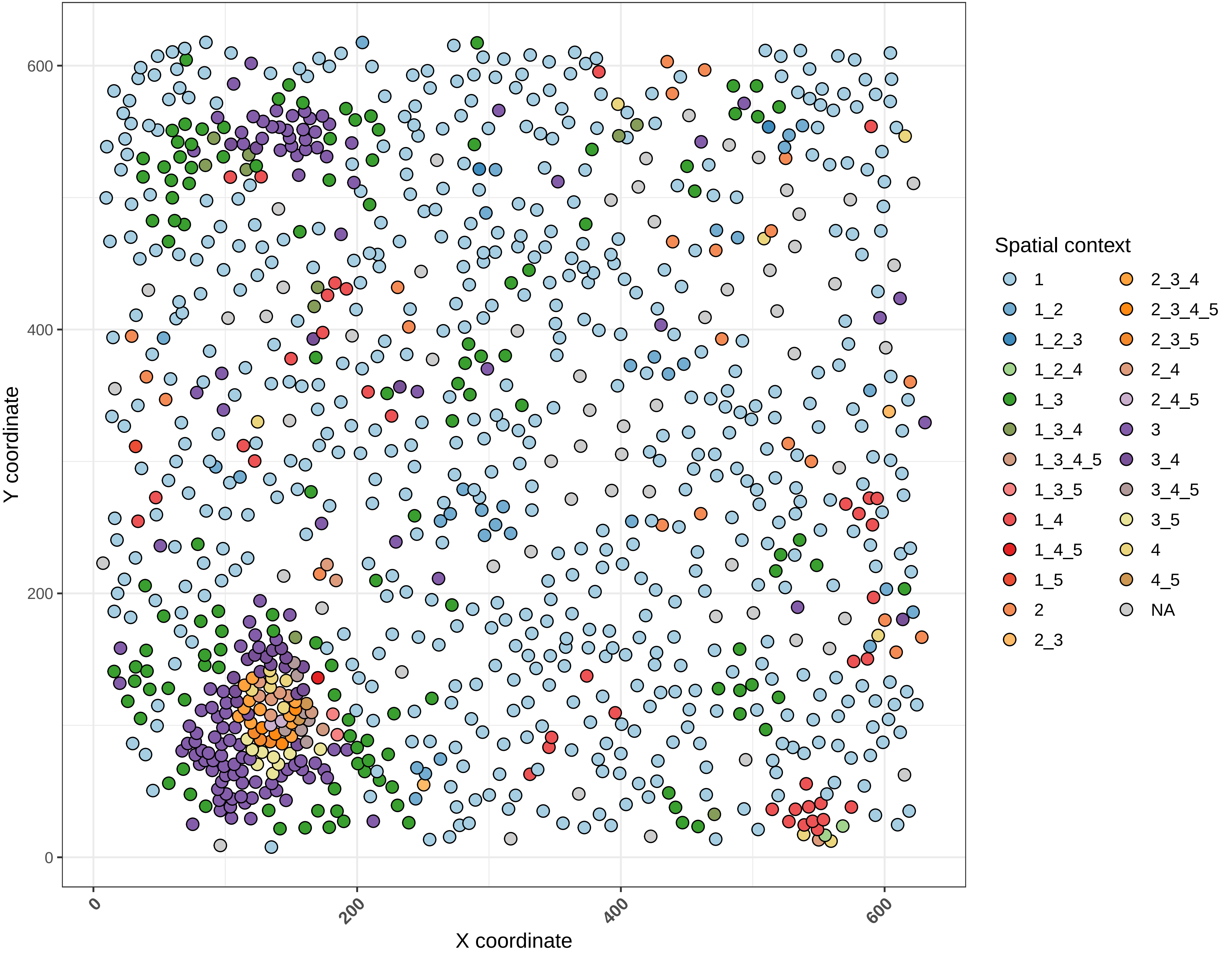

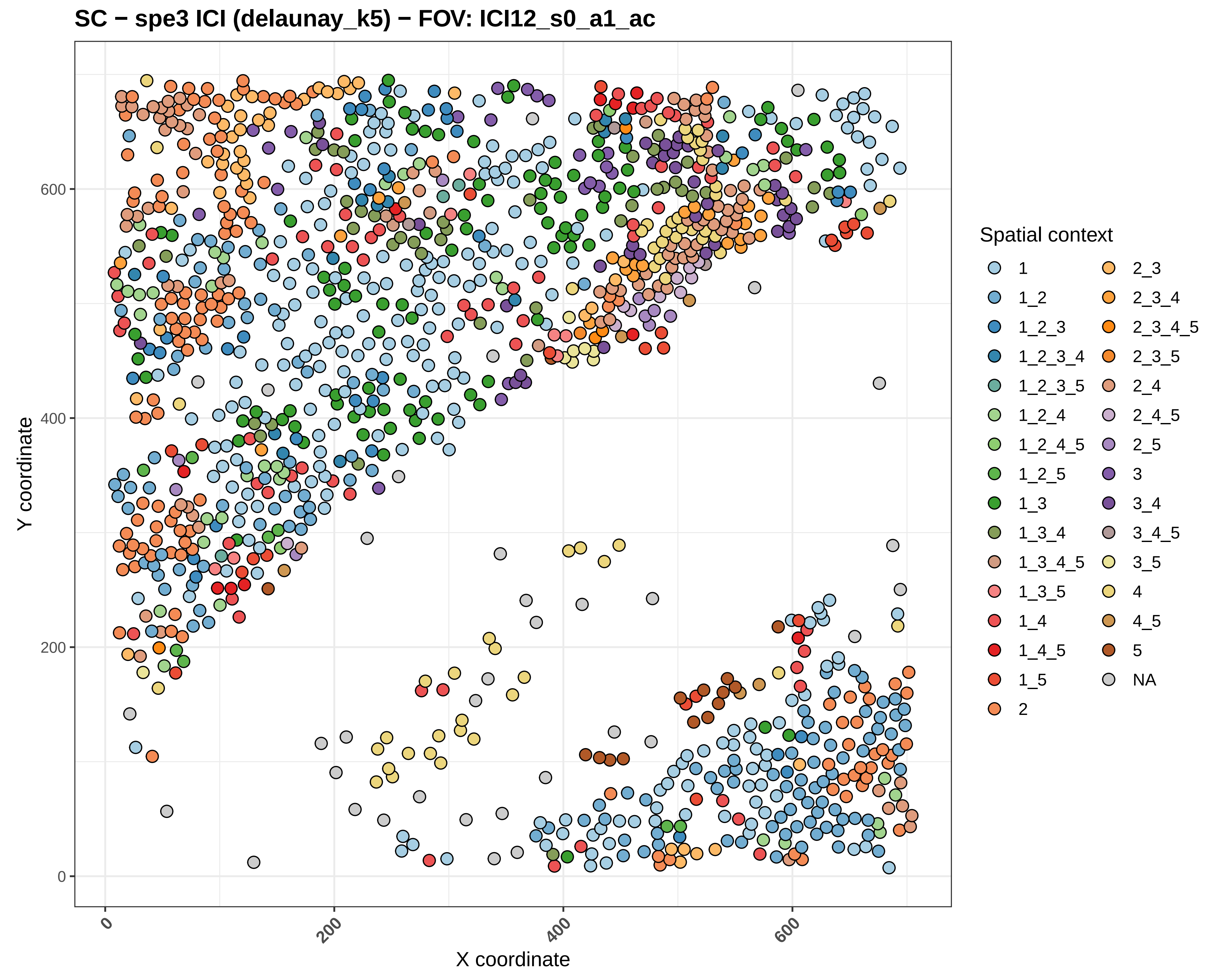
SC − spe3 ICI (delaunay_k5) − FOV: ICI12_s0_a1_ac
Y coordinate
X coordinate
0
200
400
600
Spatial context
1
1_2
1_2_3
1_2_3_4
1_2_3_5
1_2_4
1_2_4_5
1_2_5
1_3
1_3_4
1_3_4_5
1_3_5
1_4
1_4_5
1_5
2
2_3
2_3_4
2_3_4_5
2_3_5
2_4
2_4_5
2_5
3
3_4
3_4_5
3_5
4
4_5
5
NA

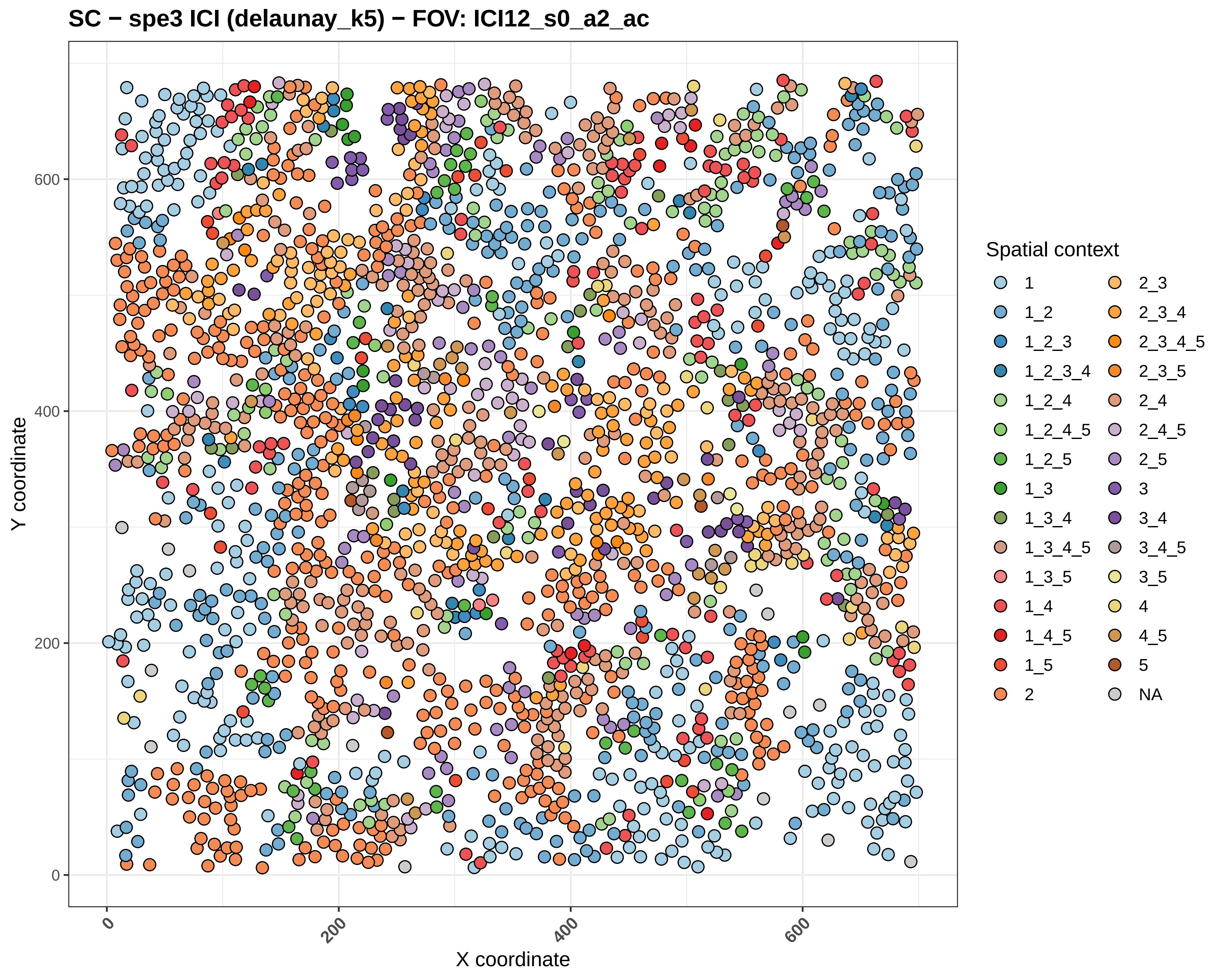

SC − spe3 ICI (delaunay_k5) − FOV: ICI12_s0_a2_ac
Y coordinate
X coordinate
0
200
400
600
Spatial context
1
1_2
1_2_3
1_2_3_4
1_2_4
1_2_4_5
1_2_5
1_3
1_3_4
1_3_4_5
1_3_5
1_4
1_4_5
1_5
2
2_3
2_3_4
2_3_4_5
2_3_5
2_4
2_4_5
2_5
3
3_4
3_4_5
3_5
4
4_5
5
NA

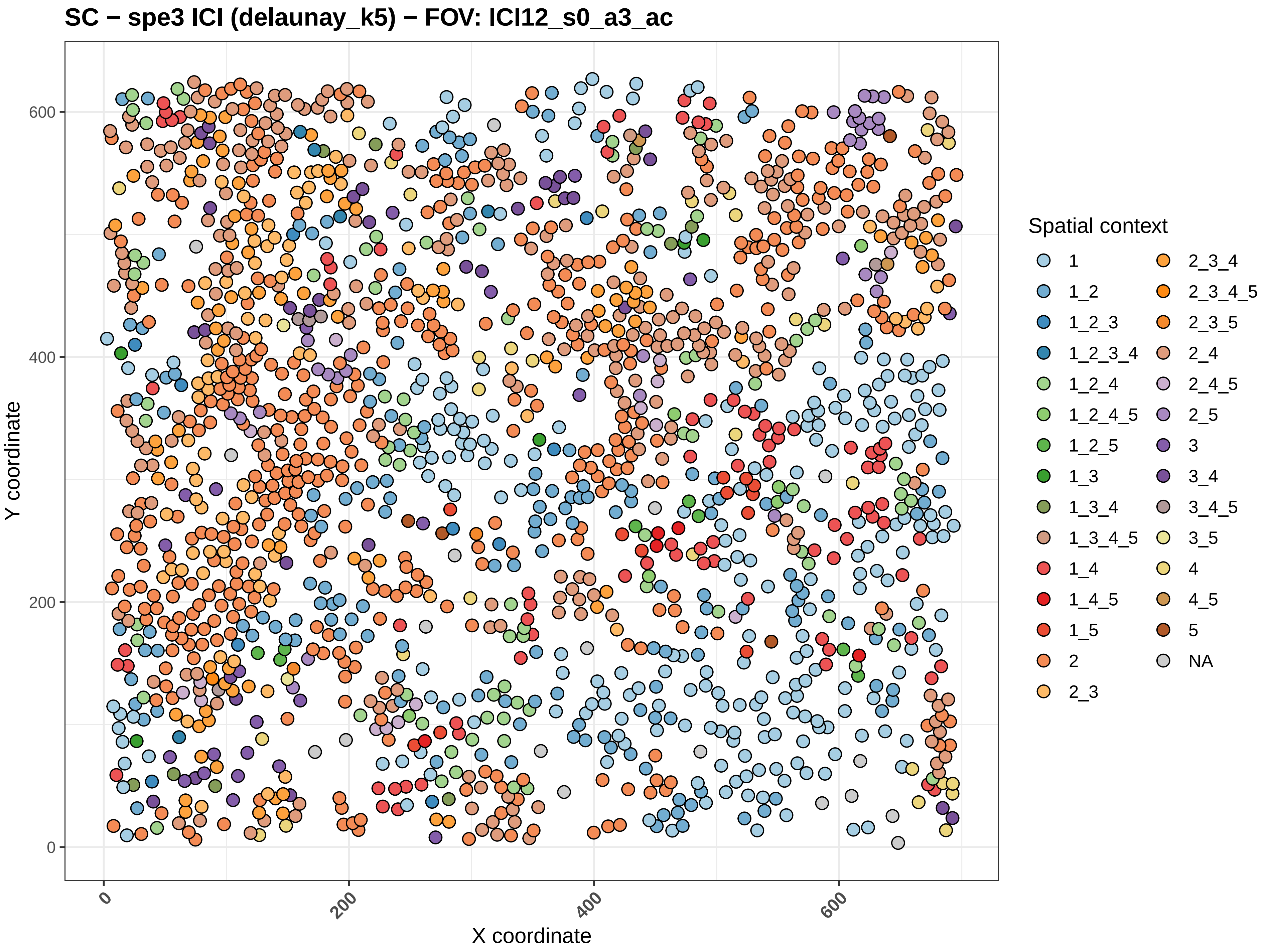
SC − spe3 ICI (delaunay_k5) − FOV: ICI12_s0_a3_ac
Y coordinate
X coordinate
0
200
400
600
Spatial context
1
1_2
1_2_3
1_2_3_4
1_2_4
1_2_4_5
1_2_5
1_3
1_3_4
1_3_4_5
1_4
1_4_5
1_5
2
2_3
2_3_4
2_3_4_5
2_3_5
2_4
2_4_5
2_5
3
3_4
3_4_5
3_5
4
4_5
5
NA

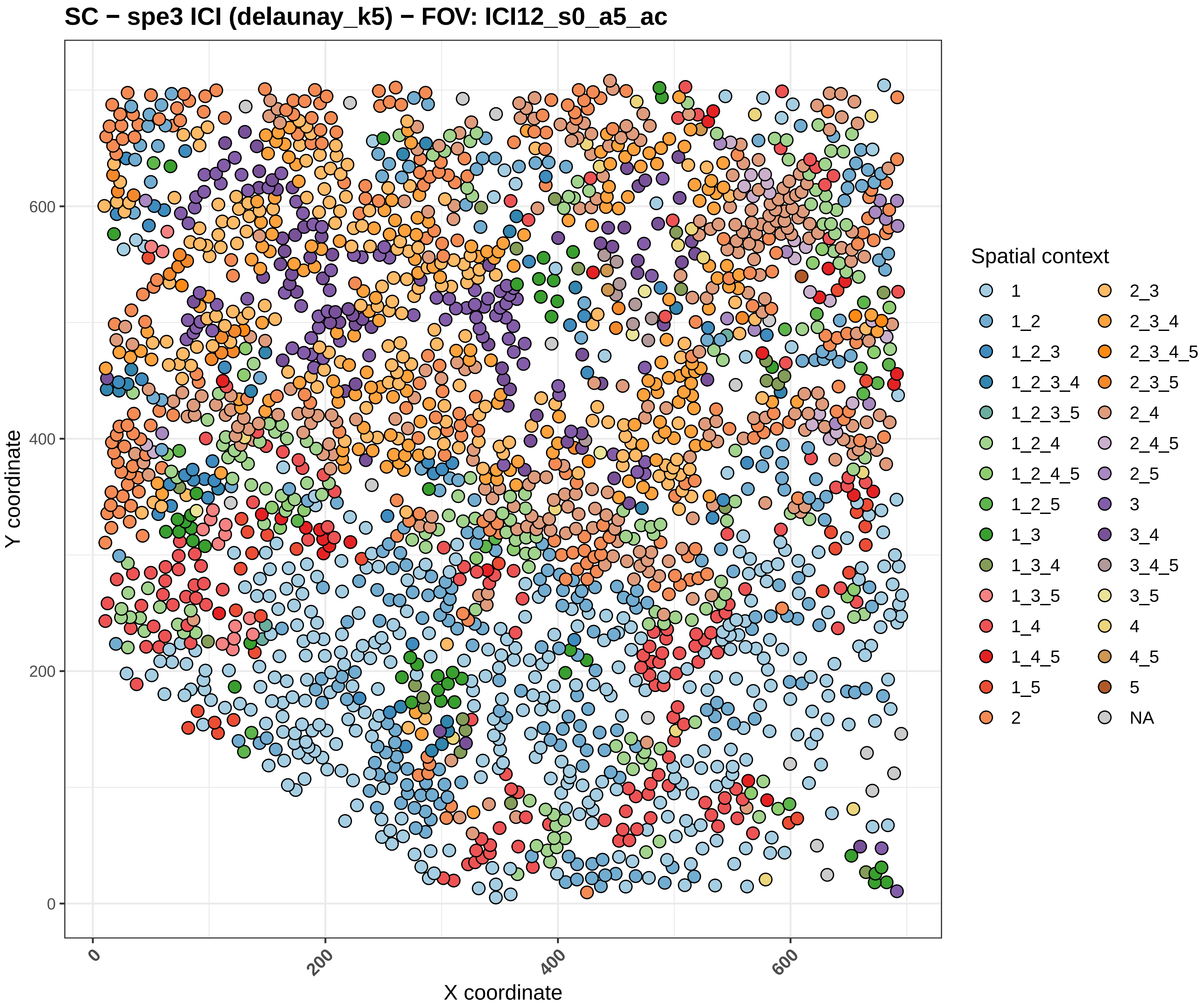

SC − spe3 ICI (delaunay_k5) − FOV: ICI12_s0_a5_ac
Y coordinate
X coordinate
0
200
400
600
Spatial context
1
1_2
1_2_3
1_2_3_4
1_2_3_5
1_2_4
1_2_4_5
1_2_5
1_3
1_3_4
1_3_5
1_4
1_4_5
1_5
2
2_3
2_3_4
2_3_4_5
2_3_5
2_4
2_4_5
2_5
3
3_4
3_4_5
3_5
4
4_5
5
NA

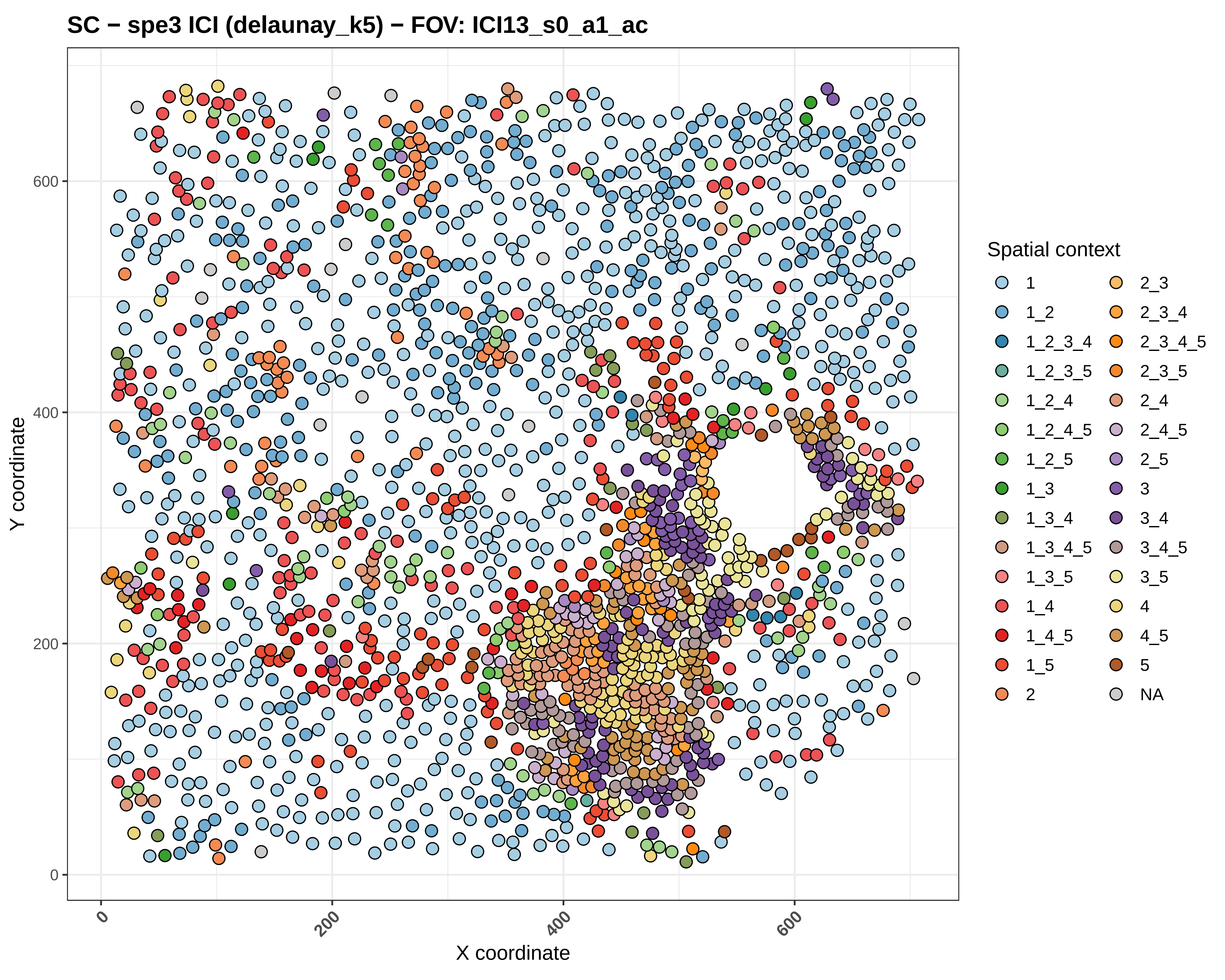

SC − spe3 ICI (delaunay_k5) − FOV: ICI13_s0_a1_ac
Y coordinate
X coordinate
0
200
400
600
Spatial context
1
1_2
1_2_3_4
1_2_3_5
1_2_4
1_2_4_5
1_2_5
1_3
1_3_4
1_3_4_5
1_3_5
1_4
1_4_5
1_5
2
2_3
2_3_4
2_3_4_5
2_3_5
2_4
2_4_5
2_5
3
3_4
3_4_5
3_5
4
4_5
5
NA

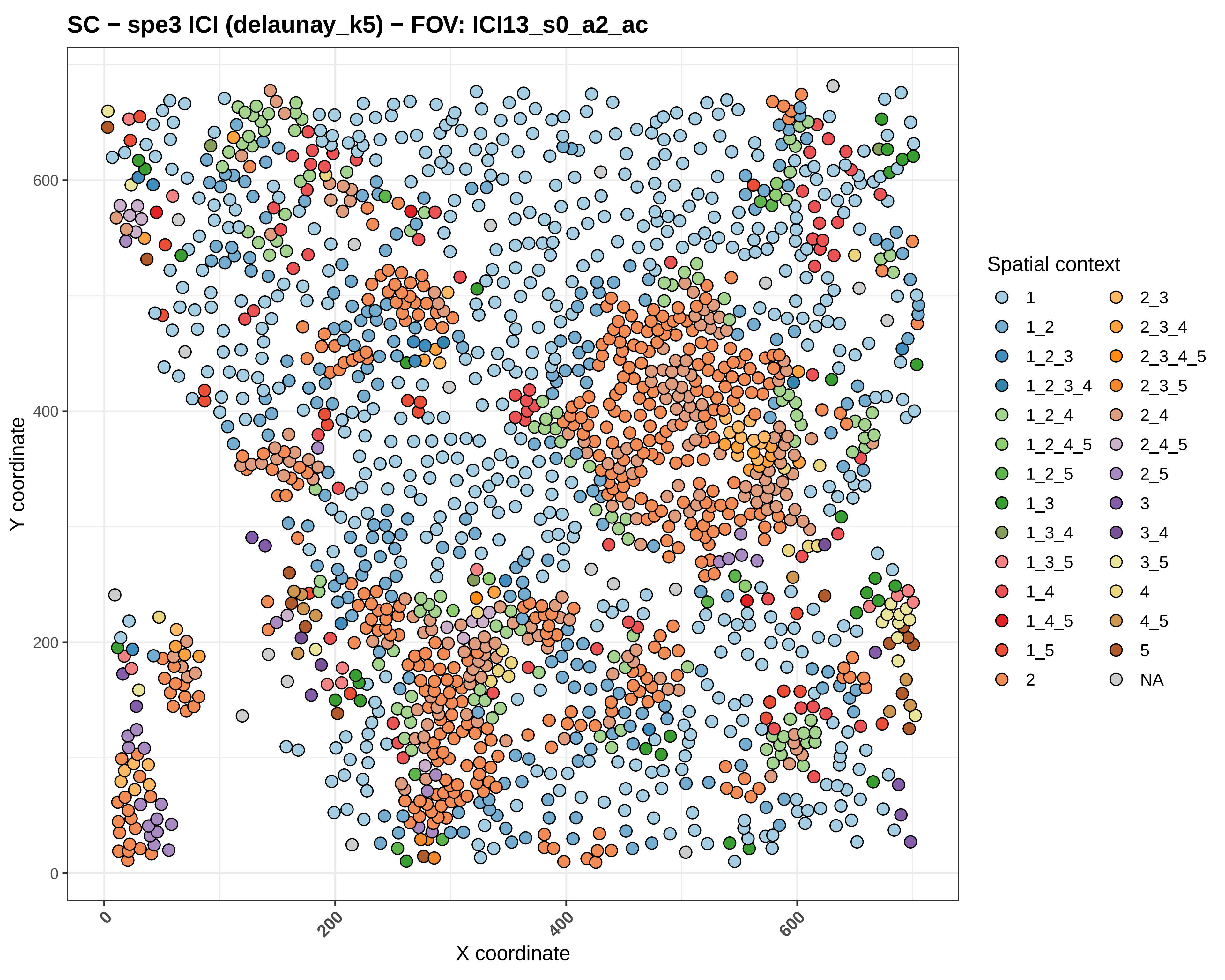

SC − spe3 ICI (delaunay_k5) − FOV: ICI13_s0_a2_ac
Y coordinate
X coordinate
0
200
400
600
Spatial context
1
1_2
1_2_3
1_2_3_4
1_2_4
1_2_4_5
1_2_5
1_3
1_3_4
1_3_5
1_4
1_4_5
1_5
2
2_3
2_3_4
2_3_4_5
2_3_5
2_4
2_4_5
2_5
3
3_4
3_5
4
4_5
5
NA

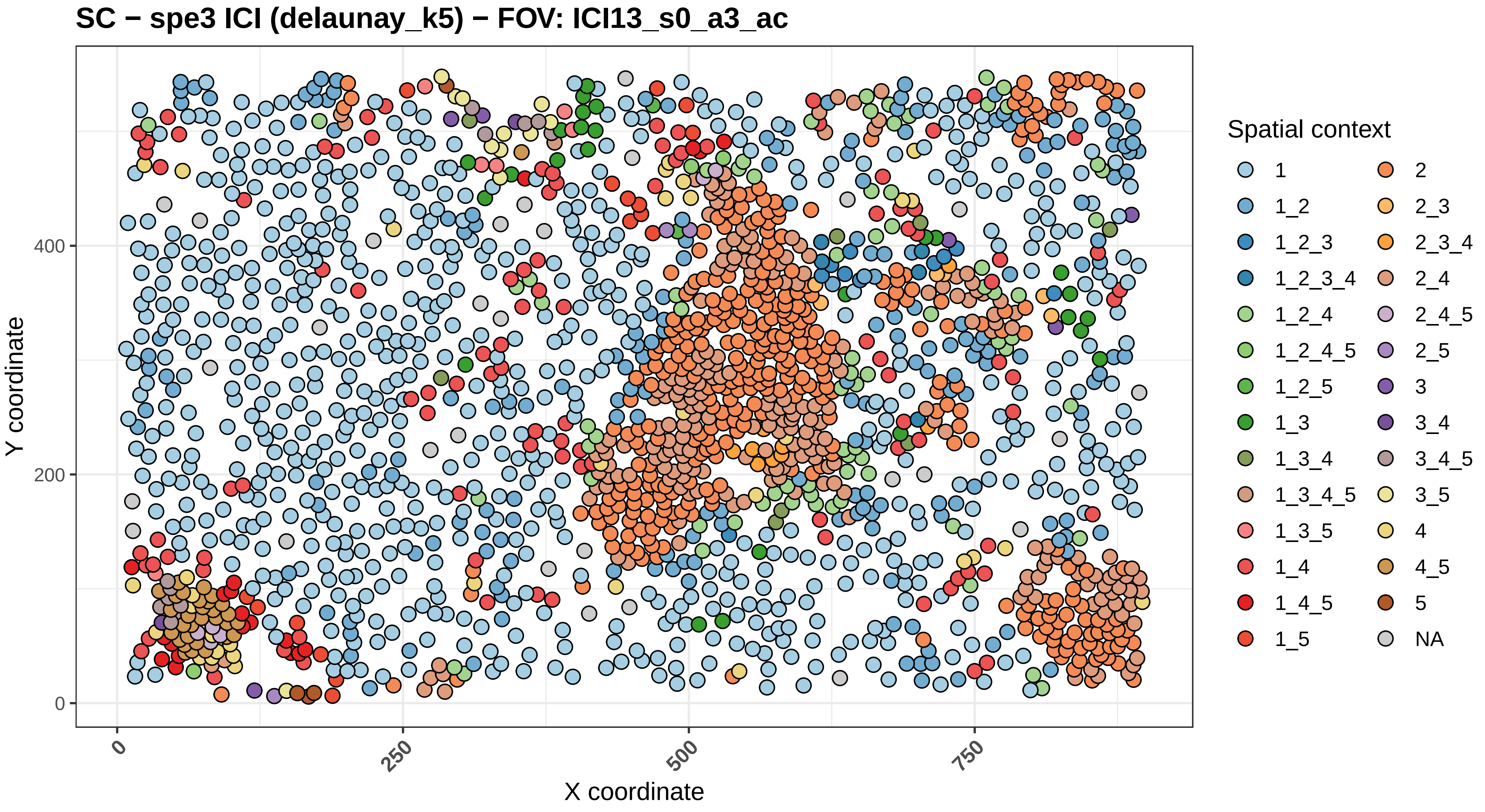
SC − spe3 ICI (delaunay_k5) − FOV: ICI13_s0_a3_ac
Y coordinate
X coordinate
0
200
400
0
250
500
750
Spatial context
1
1_2
1_2_3
1_2_3_4
1_2_4
1_2_4_5
1_2_5
1_3
1_3_4
1_3_4_5
1_3_5
1_4
1_4_5
1_5
2
2_3
2_3_4
2_4
2_4_5
2_5
3
3_4
3_4_5
3_5
4
4_5
5
NA

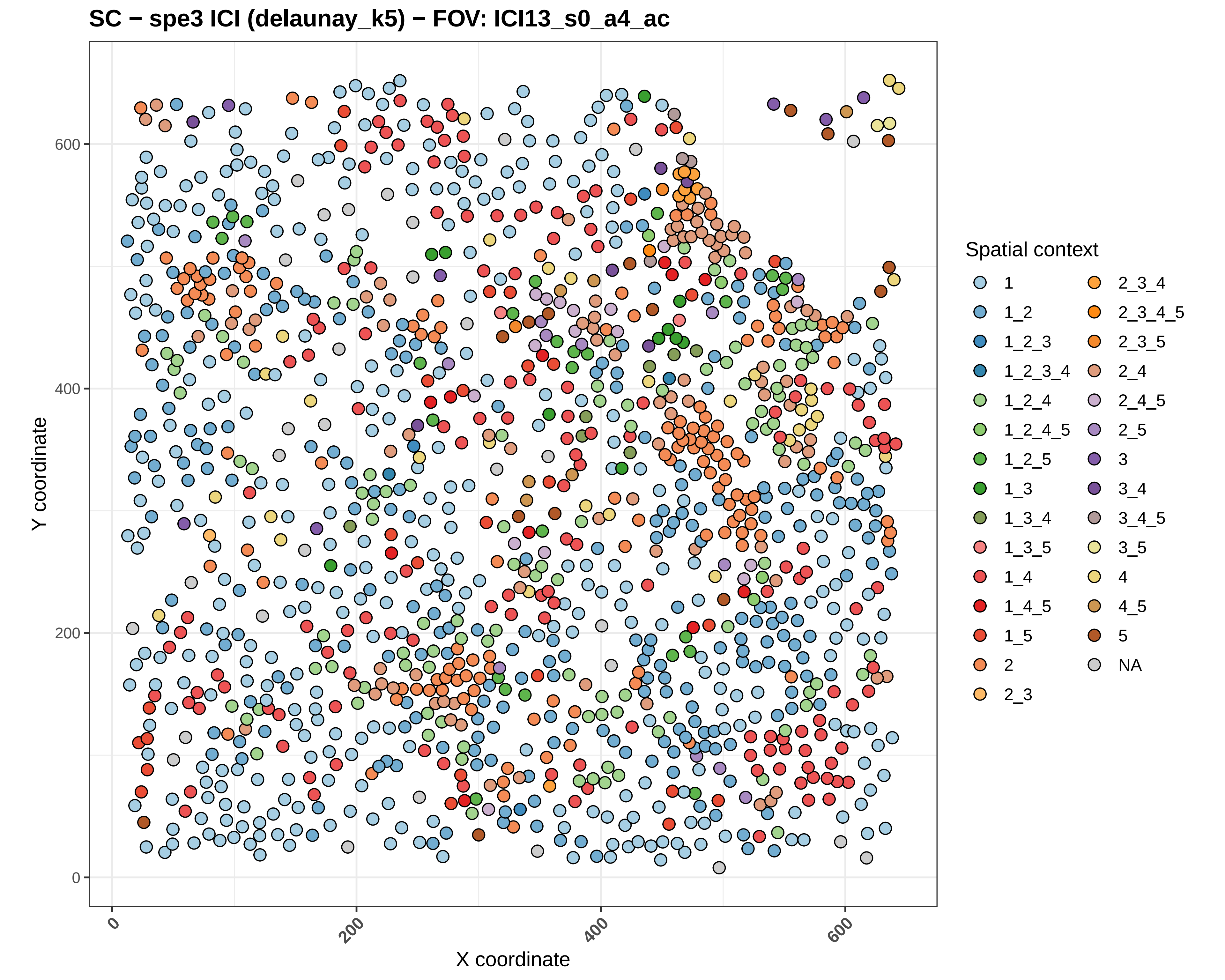
SC − spe3 ICI (delaunay_k5) − FOV: ICI13_s0_a4_ac
Y coordinate
X coordinate
0
200
400
600
Spatial context
1
1_2
1_2_3
1_2_3_4
1_2_4
1_2_4_5
1_2_5
1_3
1_3_4
1_3_5
1_4
1_4_5
1_5
2
2_3
2_3_4
2_3_4_5
2_3_5
2_4
2_4_5
2_5
3
3_4
3_4_5
3_5
4
4_5
5
NA

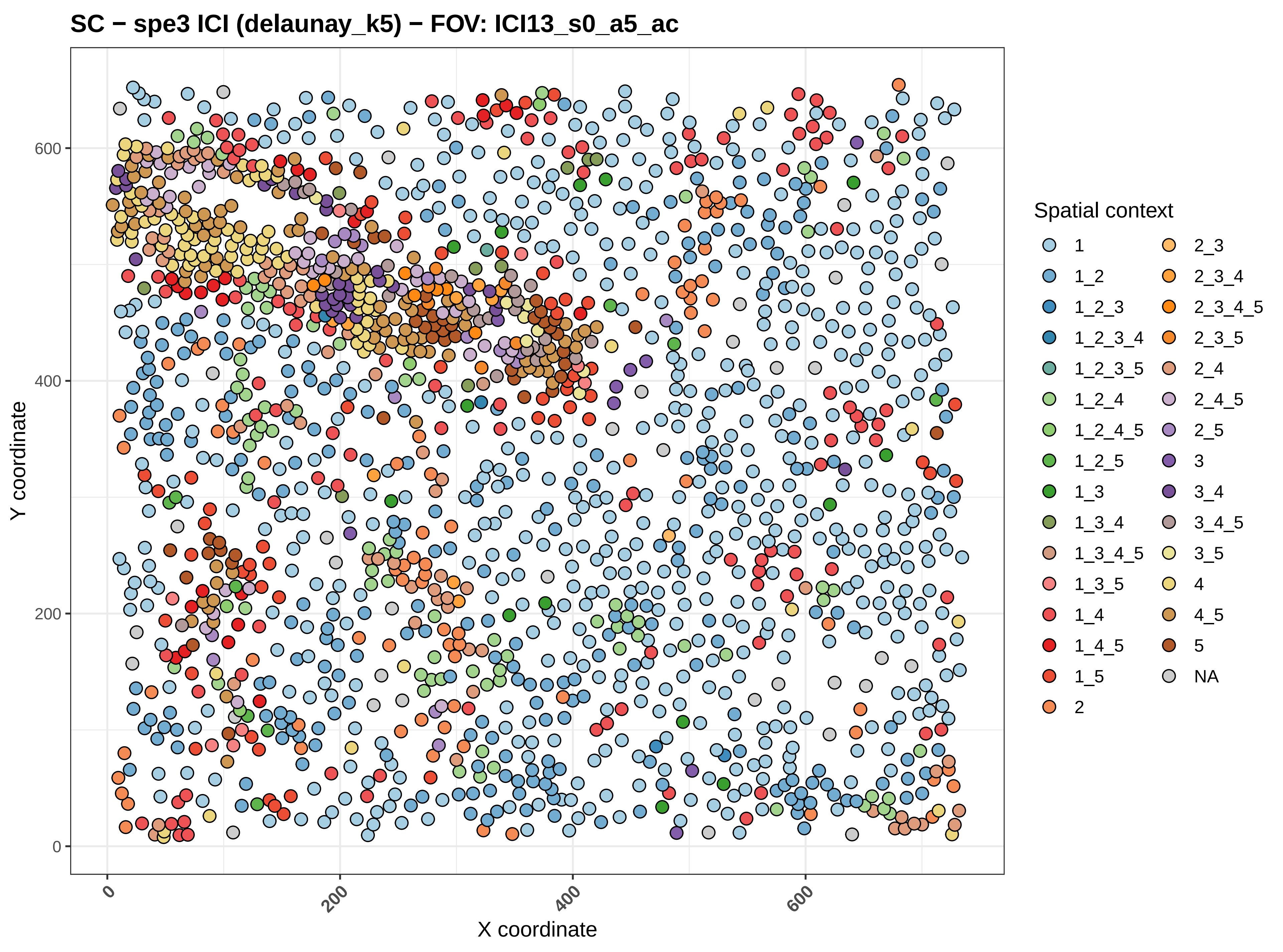
SC − spe3 ICI (delaunay_k5) − FOV: ICI13_s0_a5_ac
Y coordinate
X coordinate
0
200
400
600
Spatial context
1
1_2
1_2_3
1_2_3_4
1_2_3_5
1_2_4
1_2_4_5
1_2_5
1_3
1_3_4
1_3_4_5
1_3_5
1_4
1_4_5
1_5
2
2_3
2_3_4
2_3_4_5
2_3_5
2_4
2_4_5
2_5
3
3_4
3_4_5
3_5
4
4_5
5
NA

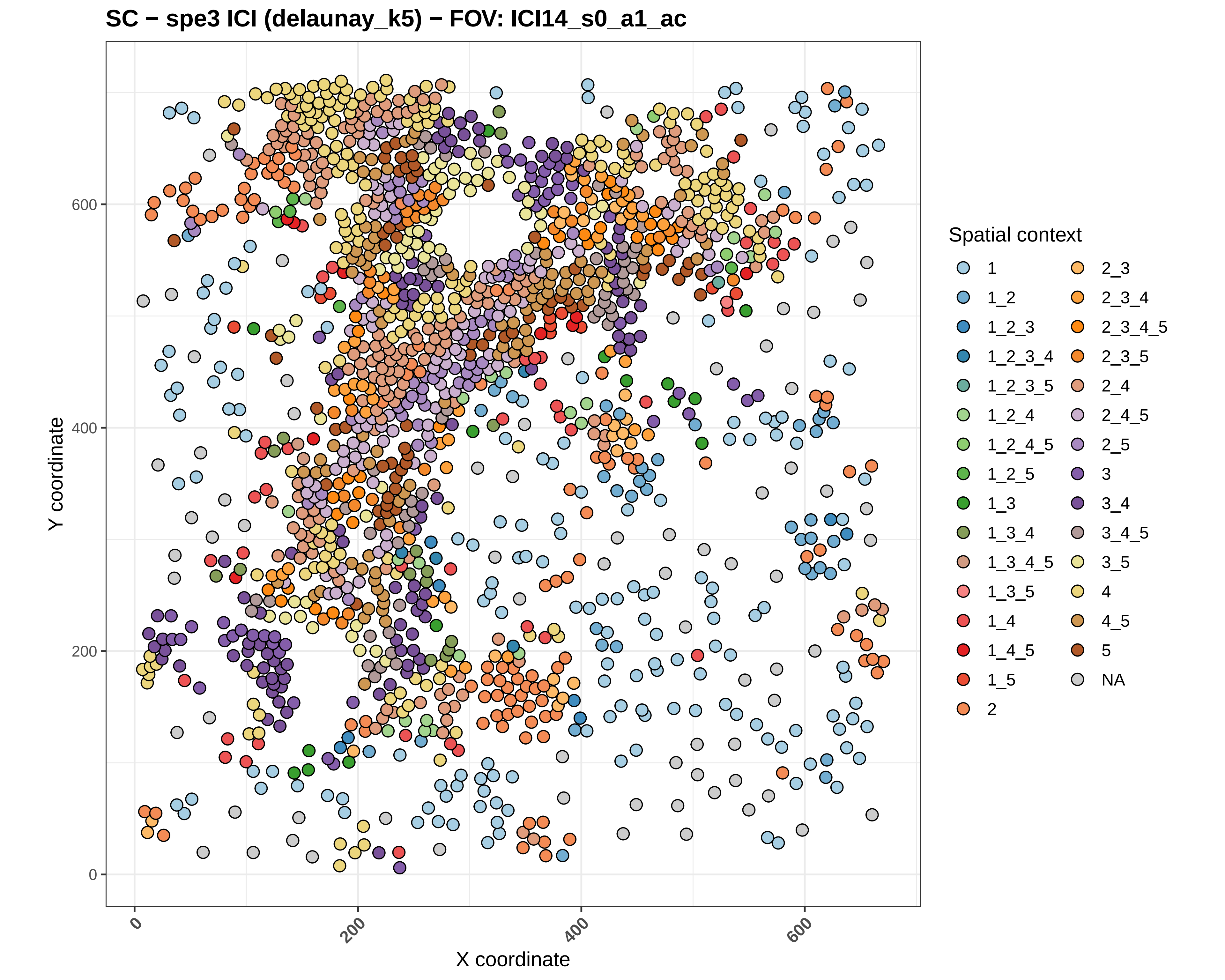

SC − spe3 ICI (delaunay_k5) − FOV: ICI14_s0_a1_ac
Y coordinate
X coordinate
0
200
400
600
Spatial context
1
1_2
1_2_3
1_2_3_4
1_2_3_5
1_2_4
1_2_4_5
1_2_5
1_3
1_3_4
1_3_4_5
1_3_5
1_4
1_4_5
1_5
2
2_3
2_3_4
2_3_4_5
2_3_5
2_4
2_4_5
2_5
3
3_4
3_4_5
3_5
4
4_5
5
NA

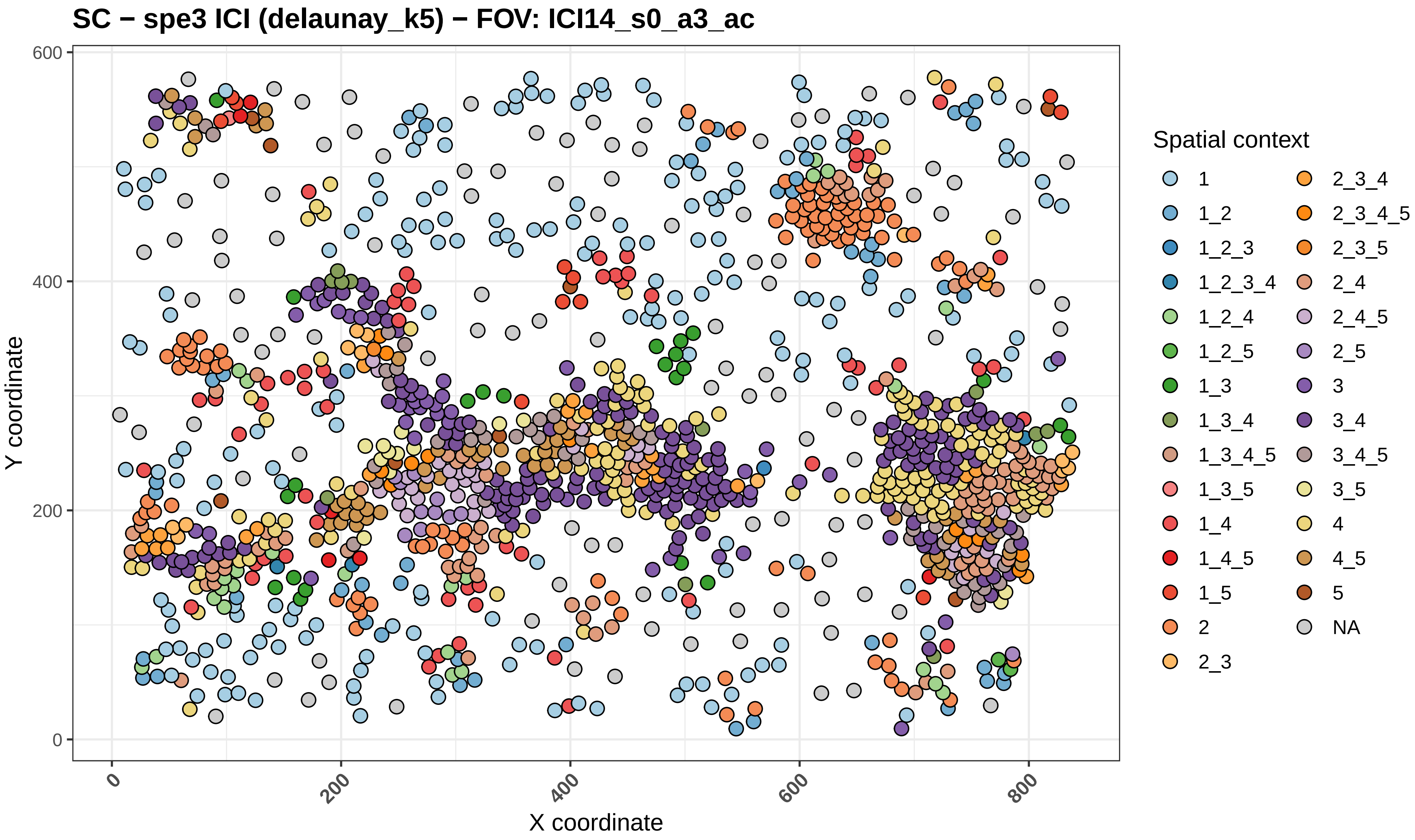

SC − spe3 ICI (delaunay_k5) − FOV: ICI14_s0_a3_ac
Y coordinate
X coordinate
0
200
400
600
800
Spatial context
1
1_2
1_2_3
1_2_3_4
1_2_4
1_2_5
1_3
1_3_4
1_3_4_5
1_3_5
1_4
1_4_5
1_5
2
2_3
2_3_4
2_3_4_5
2_3_5
2_4
2_4_5
2_5
3
3_4
3_4_5
3_5
4
4_5
5
NA

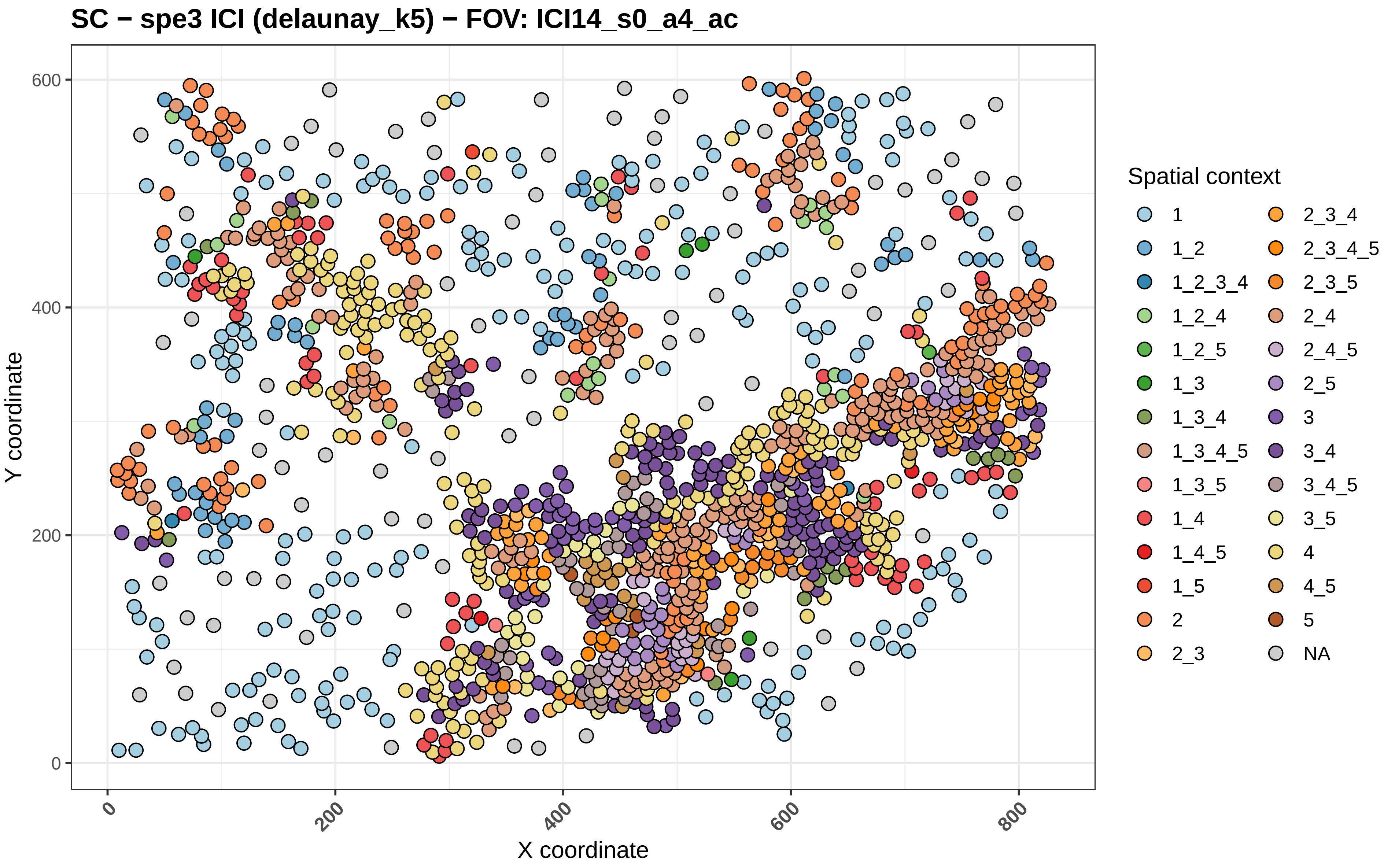
SC − spe3 ICI (delaunay_k5) − FOV: ICI14_s0_a4_ac
Y coordinate
X coordinate
0
200
400
600
800
Spatial context
1
1_2
1_2_3_4
1_2_4
1_2_5
1_3
1_3_4
1_3_4_5
1_3_5
1_4
1_4_5
1_5
2
2_3
2_3_4
2_3_4_5
2_3_5
2_4
2_4_5
2_5
3
3_4
3_4_5
3_5
4
4_5
5
NA

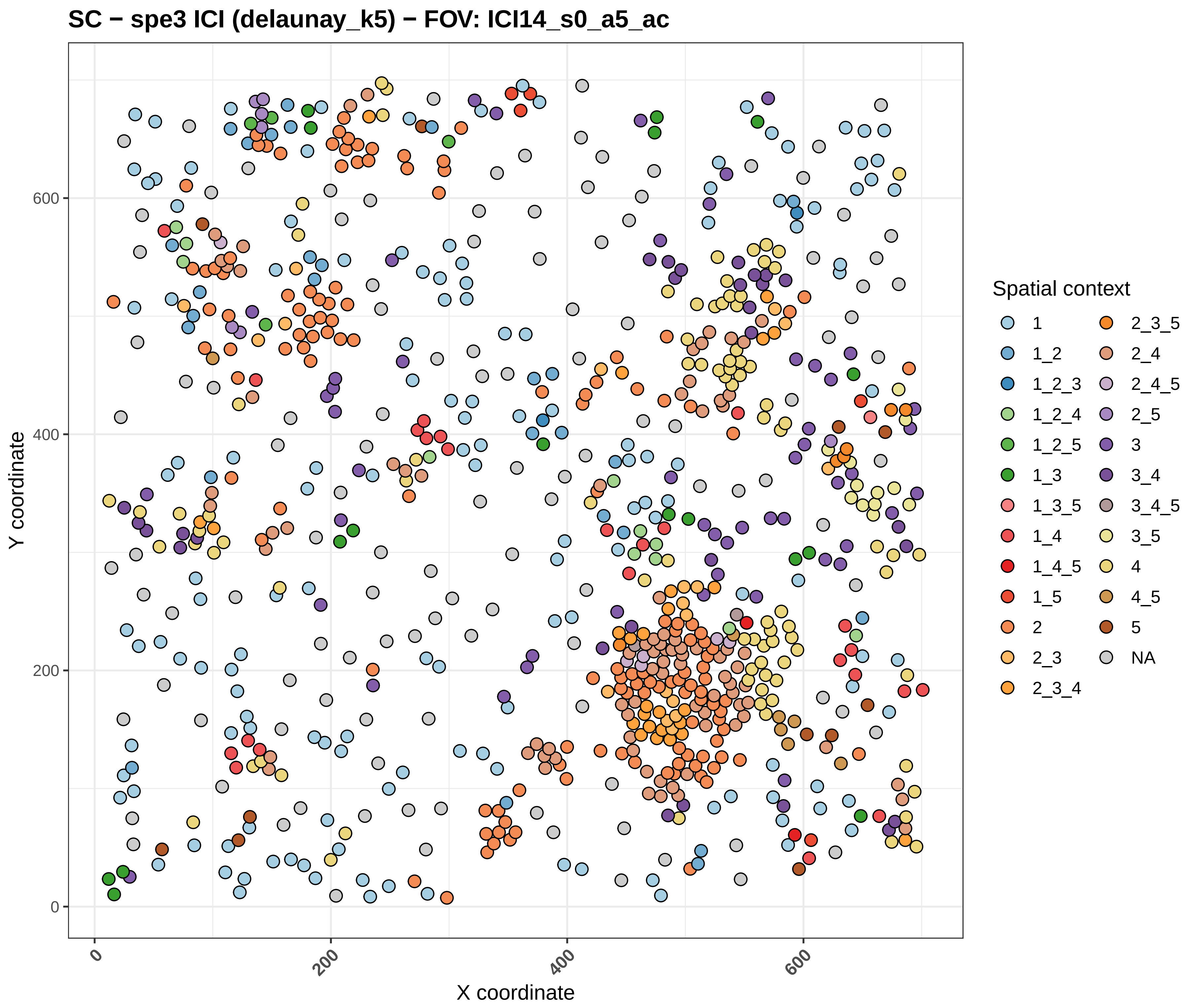
SC − spe3 ICI (delaunay_k5) − FOV: ICI14_s0_a5_ac
Y coordinate
X coordinate
0
200
400
600
Spatial context
1
1_2
1_2_3
1_2_4
1_2_5
1_3
1_3_5
1_4
1_4_5
1_5
2
2_3
2_3_4
2_3_5
2_4
2_4_5
2_5
3
3_4
3_4_5
3_5
4
4_5
5
NA

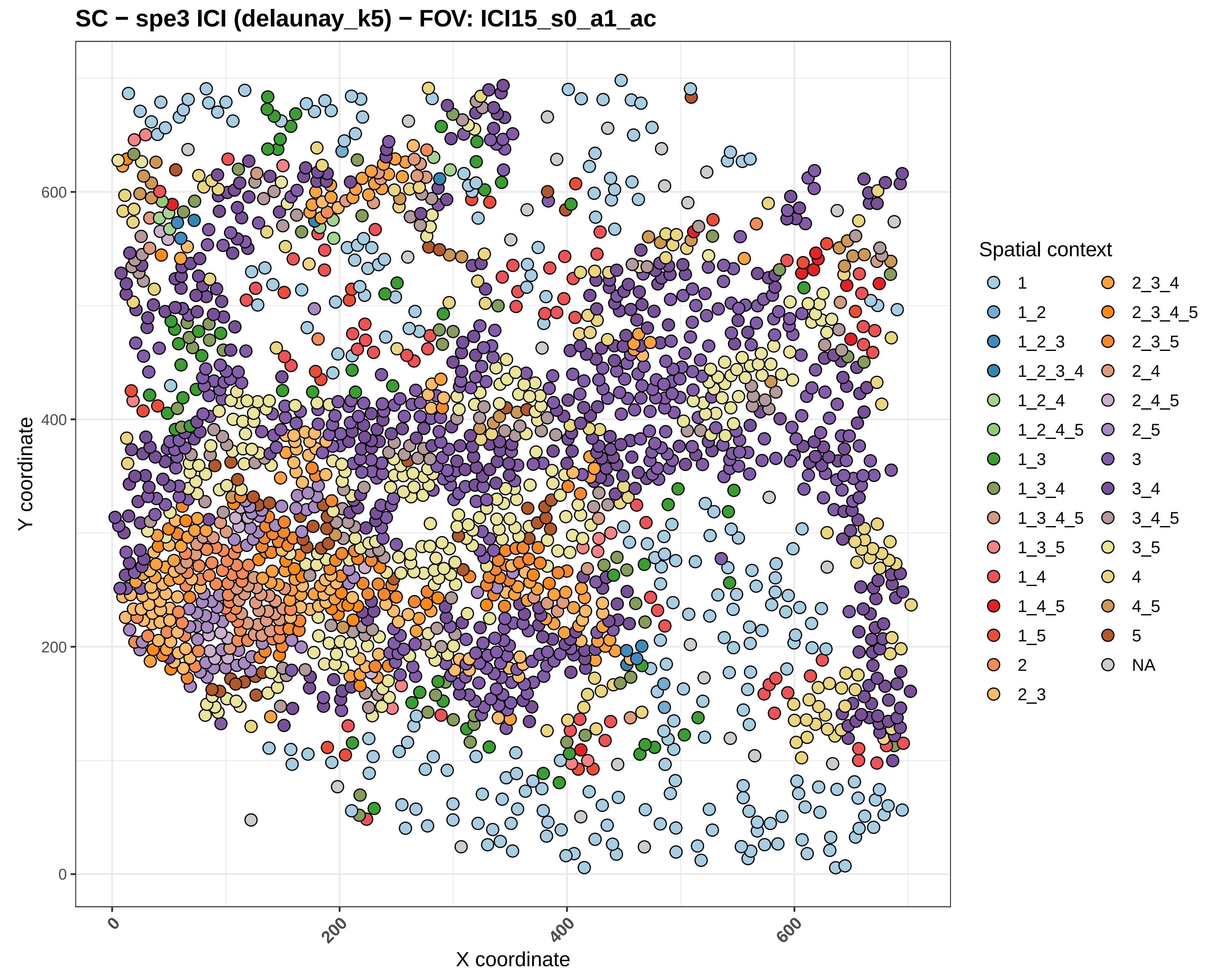
SC − spe3 ICI (delaunay_k5) − FOV: ICI15_s0_a1_ac
Y coordinate
X coordinate
0
200
400
600
Spatial context
1
1_2
1_2_3
1_2_3_4
1_2_4
1_2_4_5
1_3
1_3_4
1_3_4_5
1_3_5
1_4
1_4_5
1_5
2
2_3
2_3_4
2_3_4_5
2_3_5
2_4
2_4_5
2_5
3
3_4
3_4_5
3_5
4
4_5
5
NA

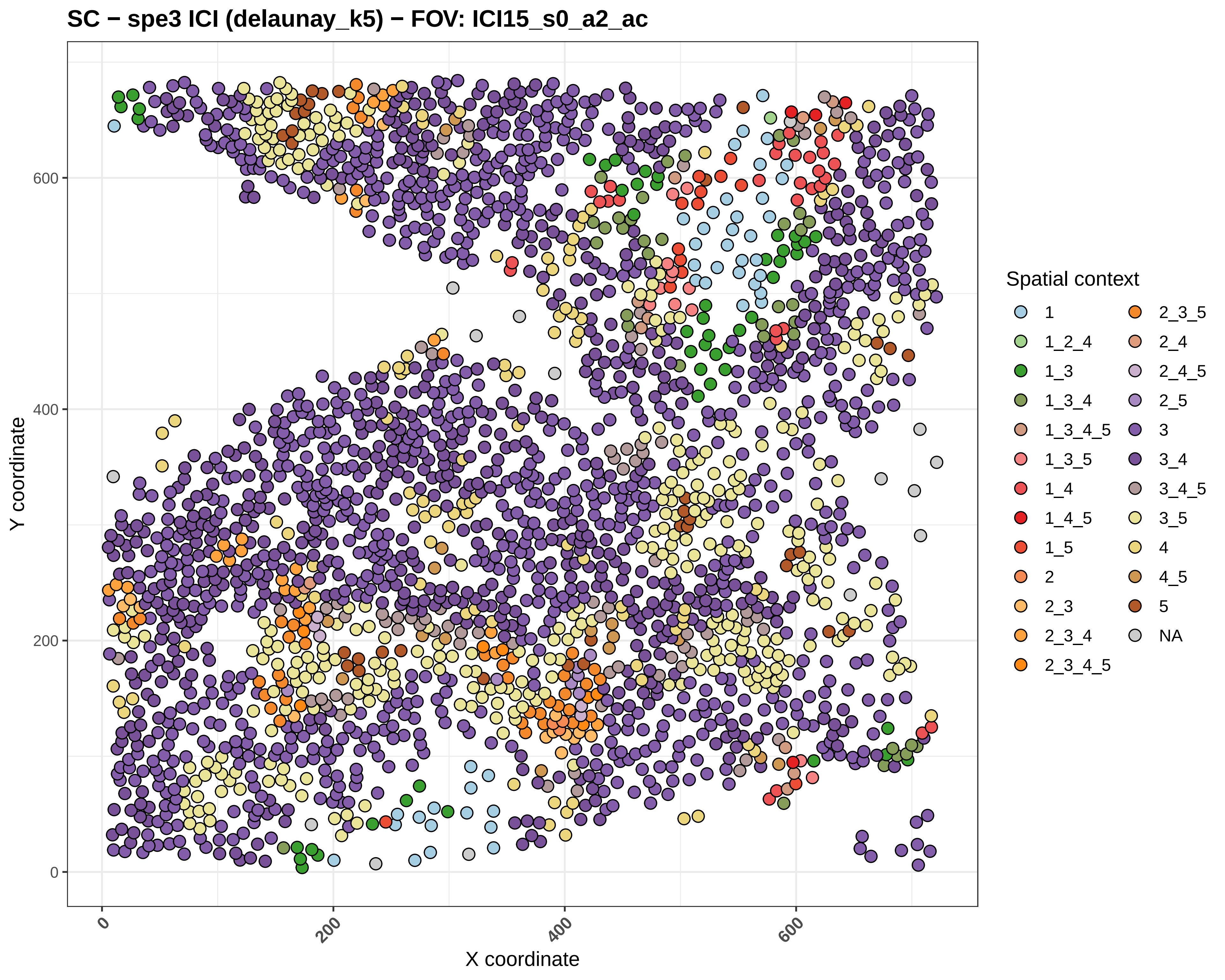
SC − spe3 ICI (delaunay_k5) − FOV: ICI15_s0_a2_ac
Y coordinate
X coordinate
0
200
400
600
Spatial context
1
1_2_4
1_3
1_3_4
1_3_4_5
1_3_5
1_4
1_4_5
1_5
2
2_3
2_3_4
2_3_4_5
2_3_5
2_4
2_4_5
2_5
3
3_4
3_4_5
3_5
4
4_5
5
NA

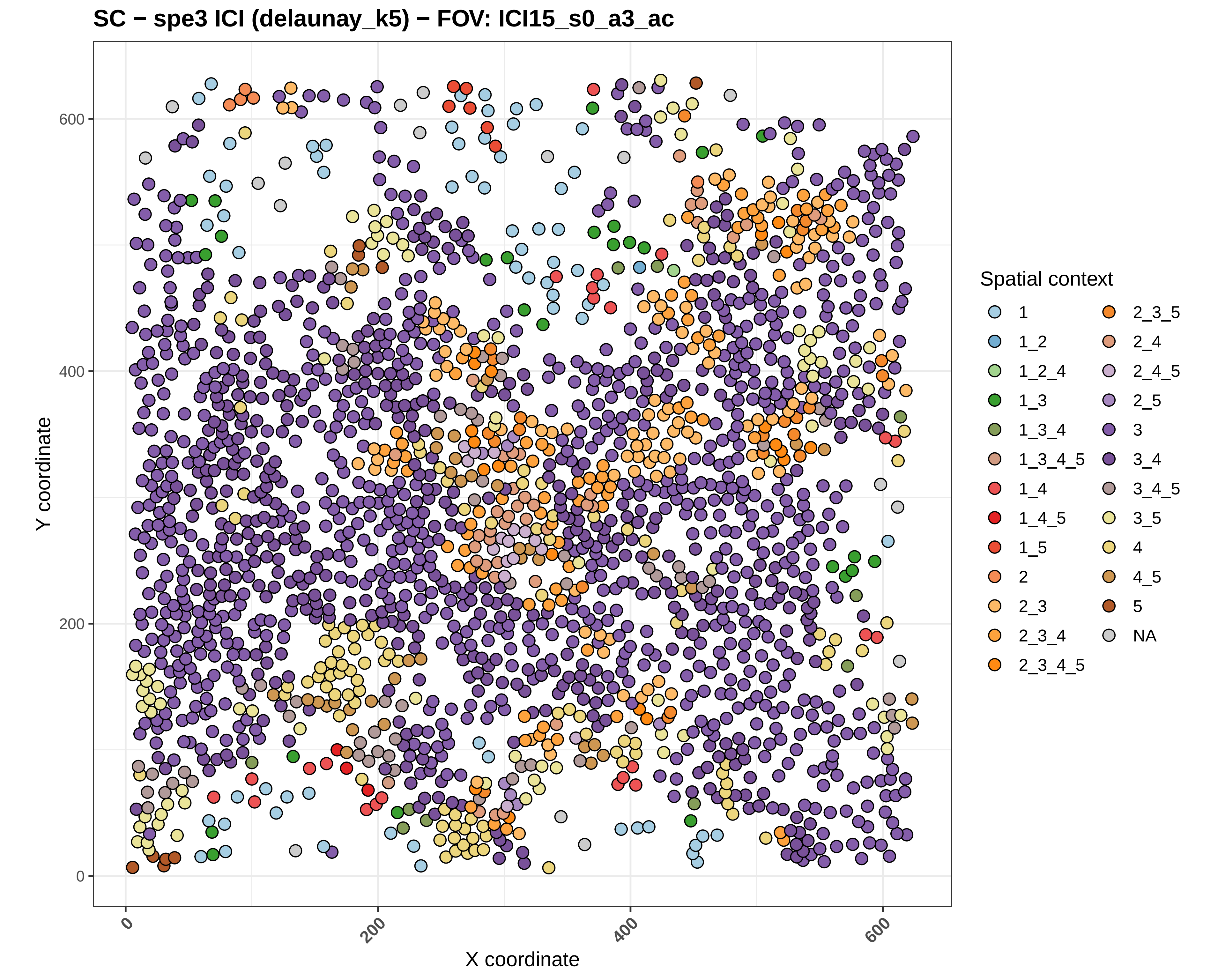
SC − spe3 ICI (delaunay_k5) − FOV: ICI15_s0_a3_ac
Y coordinate
X coordinate
0
200
400
600
Spatial context
1
1_2
1_2_4
1_3
1_3_4
1_3_4_5
1_4
1_4_5
1_5
2
2_3
2_3_4
2_3_4_5
2_3_5
2_4
2_4_5
2_5
3
3_4
3_4_5
3_5
4
4_5
5
NA

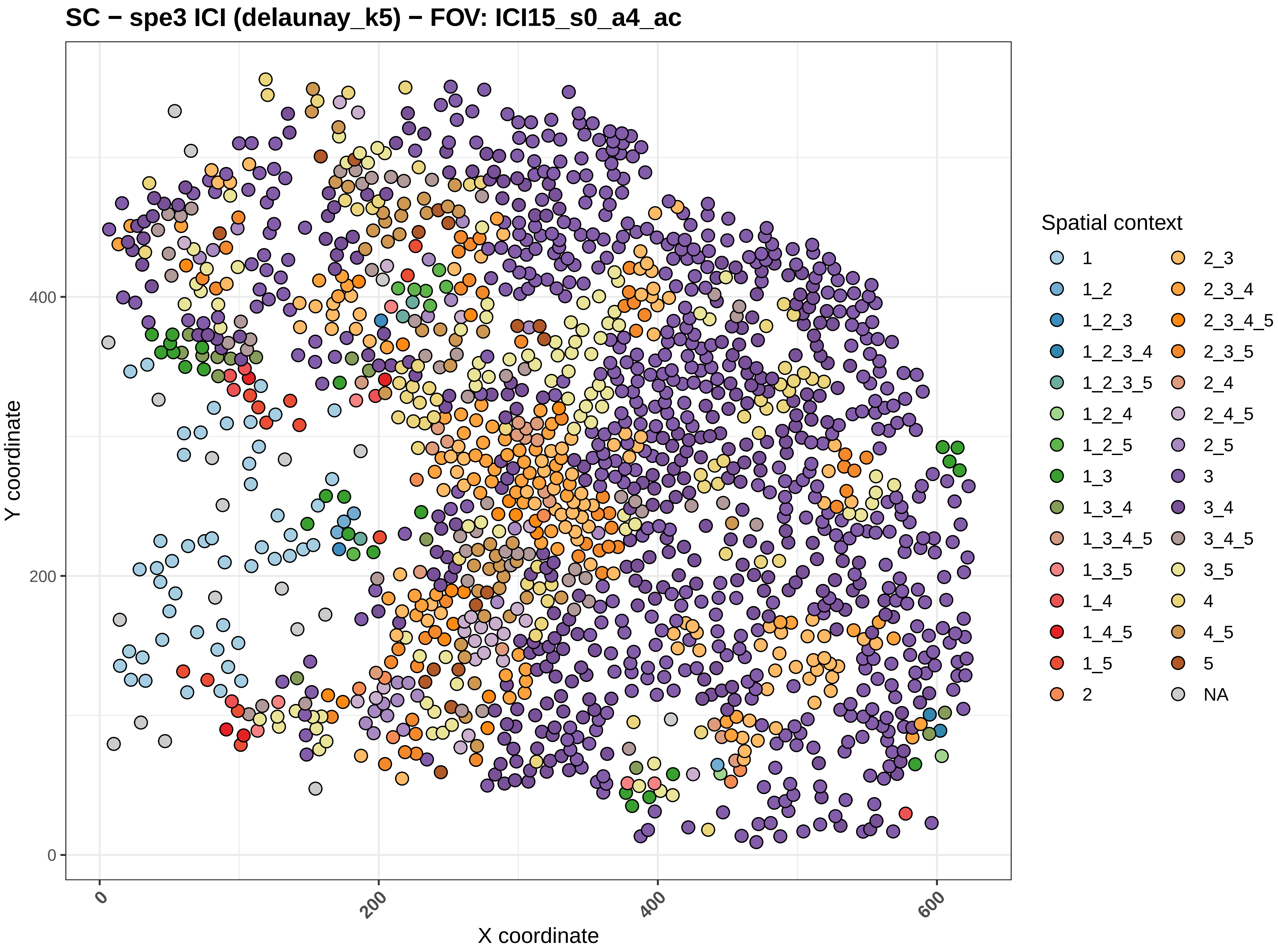
SC − spe3 ICI (delaunay_k5) − FOV: ICI15_s0_a4_ac
Y coordinate
X coordinate
0
200
400
600
Spatial context
1
1_2
1_2_3
1_2_3_4
1_2_3_5
1_2_4
1_2_5
1_3
1_3_4
1_3_4_5
1_3_5
1_4
1_4_5
1_5
2
2_3
2_3_4
2_3_4_5
2_3_5
2_4
2_4_5
2_5
3
3_4
3_4_5
3_5
4
4_5
5
NA

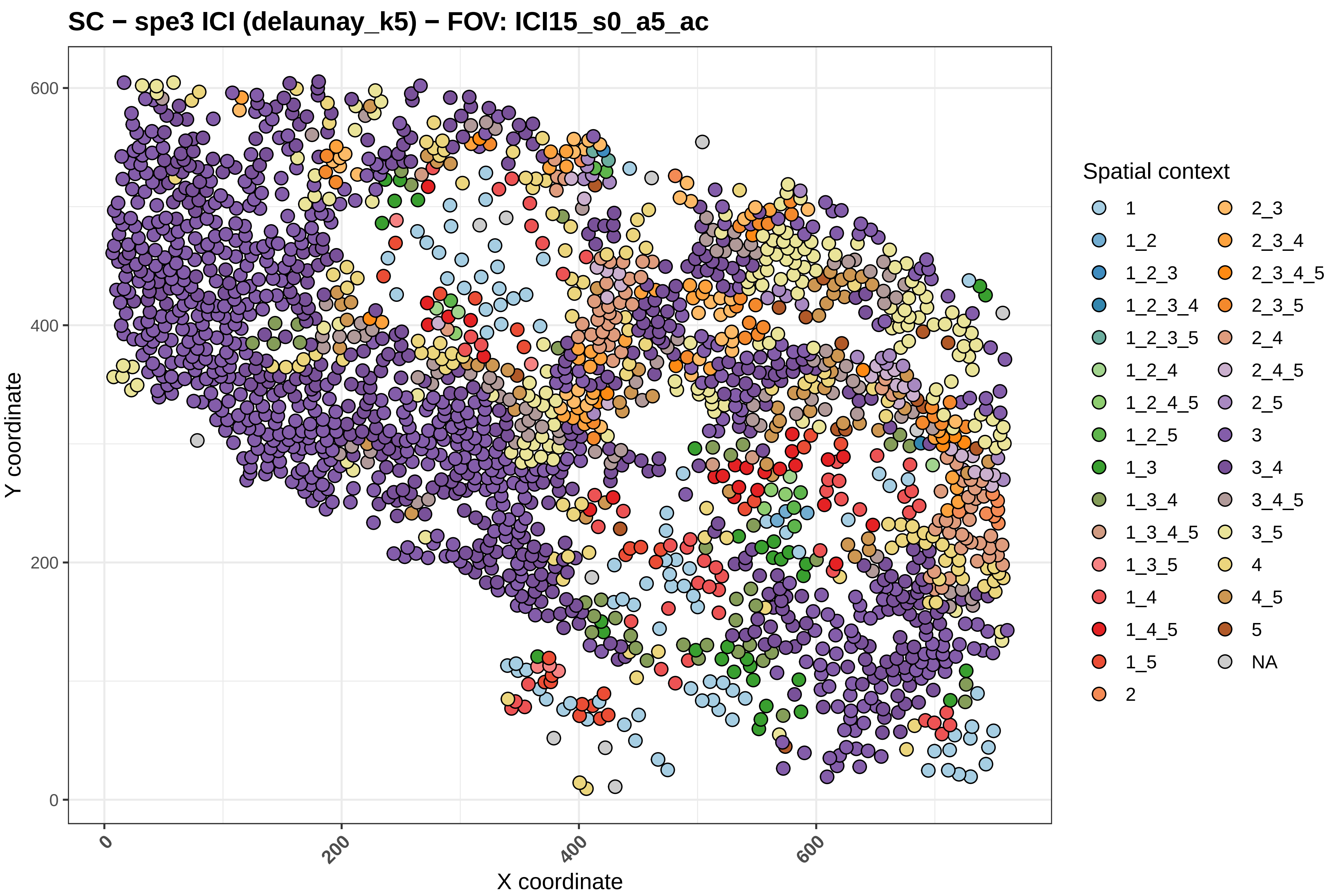
SC − spe3 ICI (delaunay_k5) − FOV: ICI15_s0_a5_ac
Y coordinate
X coordinate
0
200
400
600
Spatial context
1
1_2
1_2_3
1_2_3_4
1_2_3_5
1_2_4
1_2_4_5
1_2_5
1_3
1_3_4
1_3_4_5
1_3_5
1_4
1_4_5
1_5
2
2_3
2_3_4
2_3_4_5
2_3_5
2_4
2_4_5
2_5
3
3_4
3_4_5
3_5
4
4_5
5
NA
