## Supplementary material for "Macrophage-CD8⁺ T Cell Spatial Coupling Defines an Innate-Adaptive Injury Niche in Human Checkpoint Inhibitor Hepatotoxicity": Figure S-8

### Cell Densities + Death Events by DILIN Severity (aggregated, ICI Total)

LMM (1|patient_id) | n=53 FOVs, 12 patients | emmeans BH pairwise

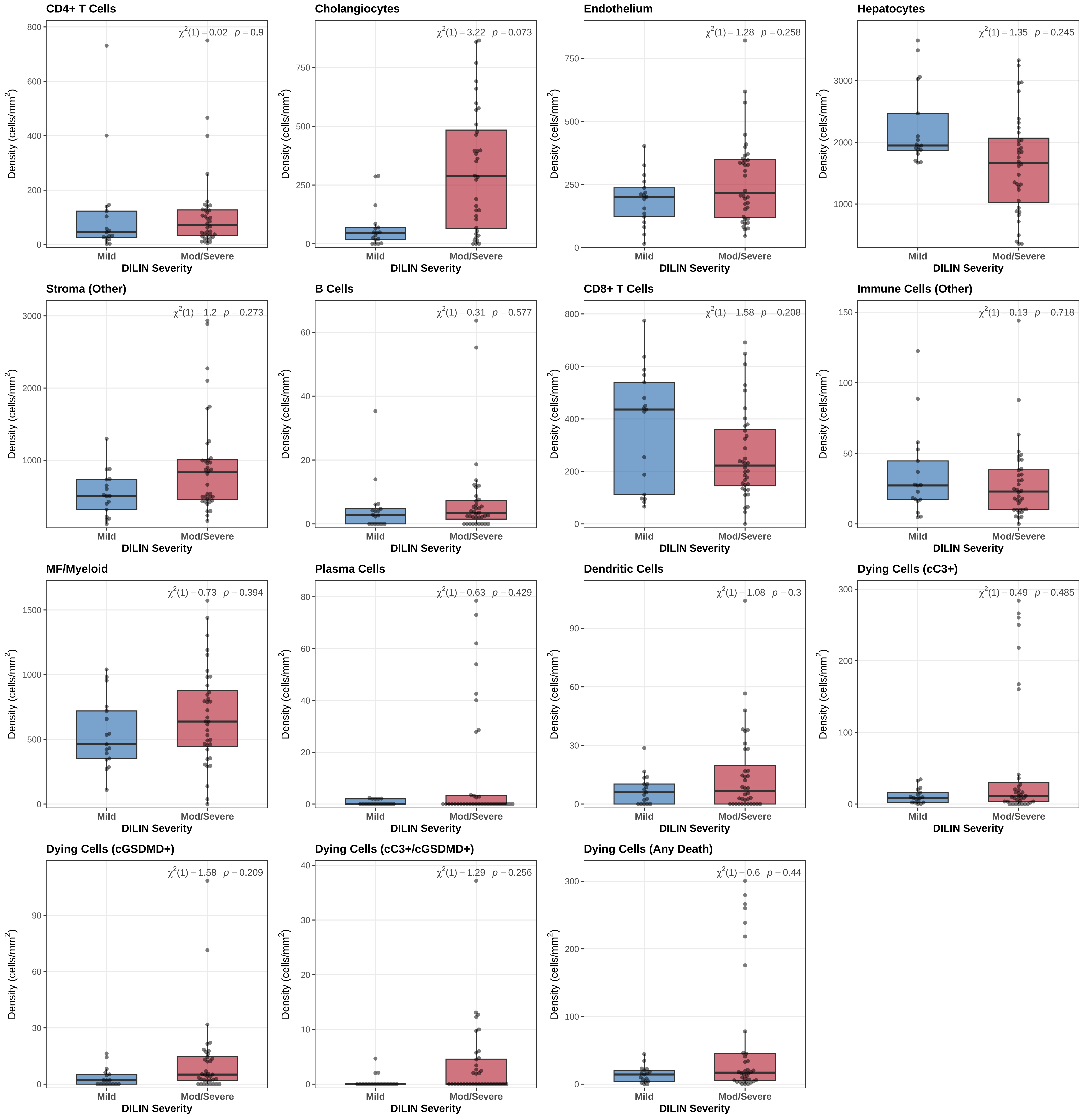

### Cell Densities + Death Events by DILIN Severity (detailed, ICI Total)

LMM (1|patient_id) | n=53 FOVs, 12 patients | emmeans BH pairwise

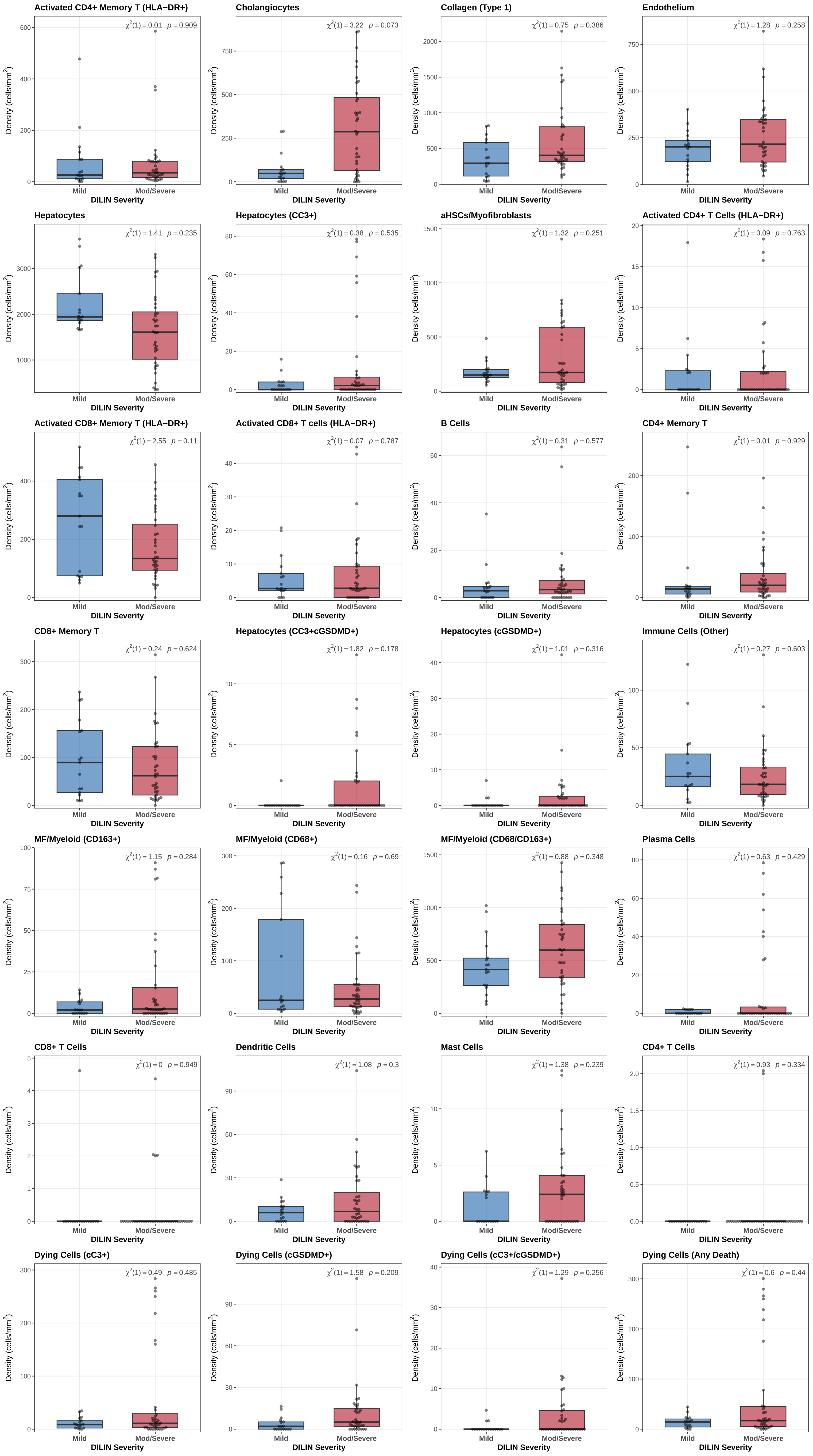
