## Supplementary material for "Macrophage-CD8⁺ T Cell Spatial Coupling Defines an Innate-Adaptive Injury Niche in Human Checkpoint Inhibitor Hepatotoxicity": Figure S-9

#### Ishak vs Cell Density (ICI) [aggregated] — Kendall tau-b (page 1/4)

Kendall tau-b: tie-corrected ordinal correlation | (1|patient_id) not shown

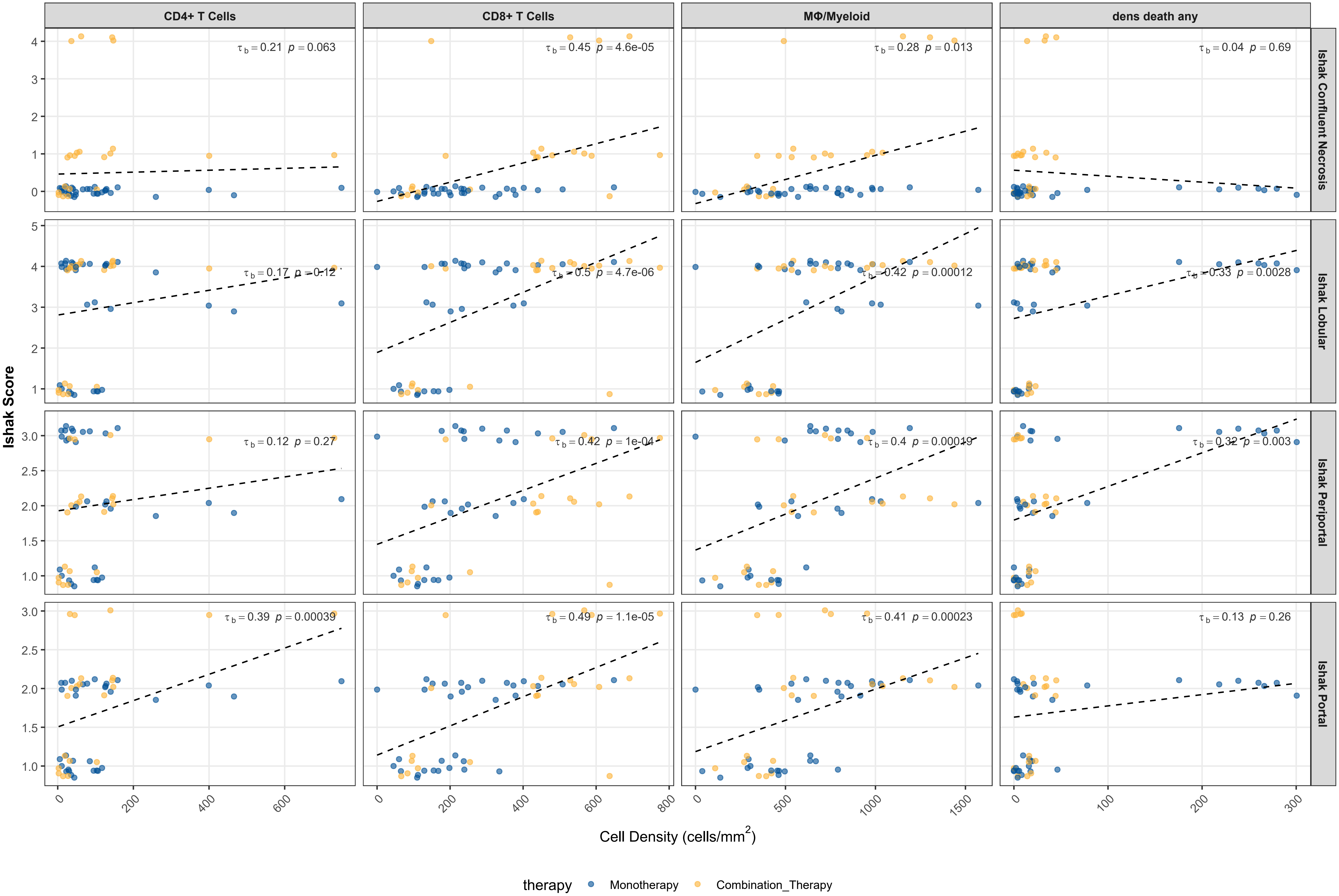

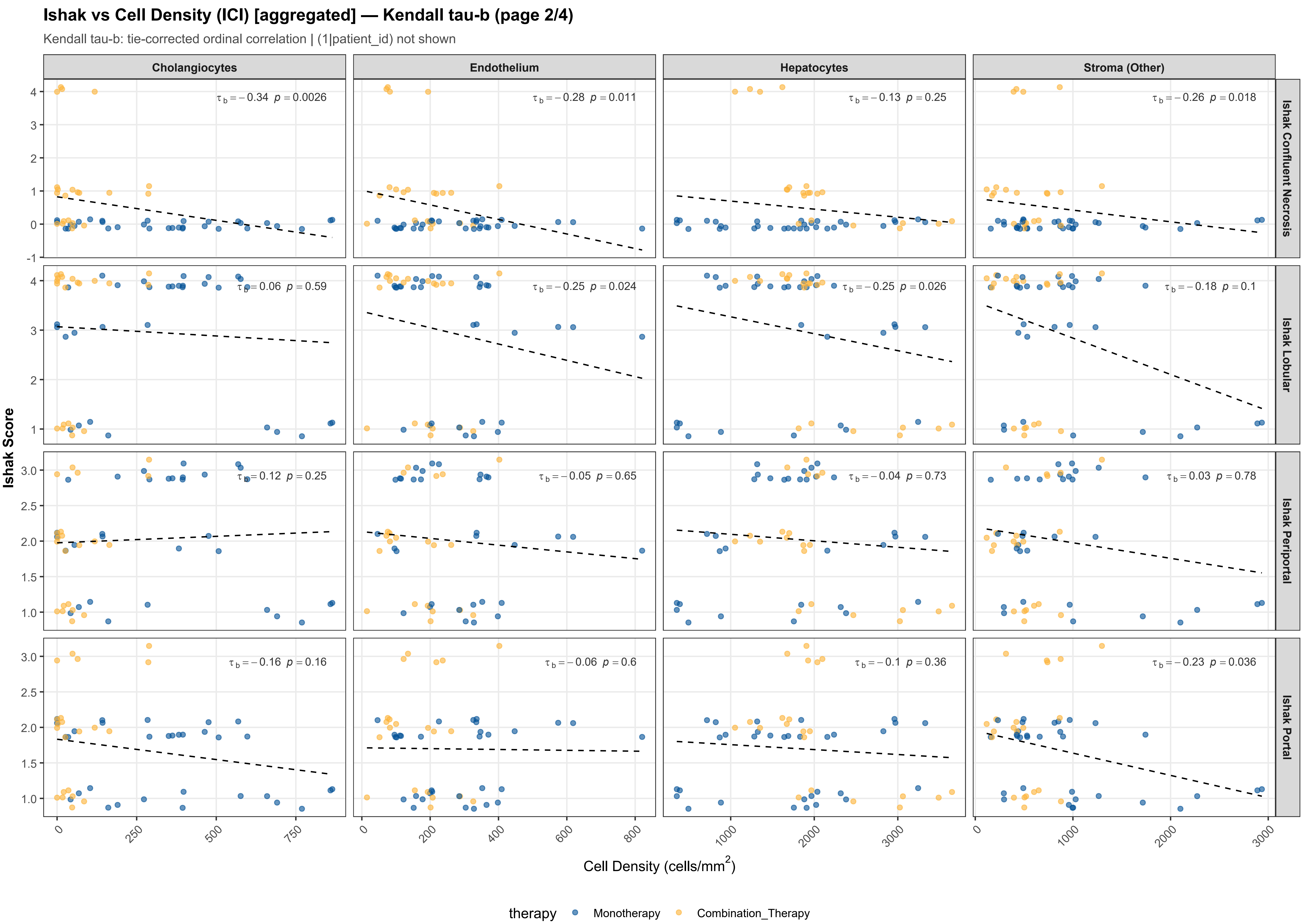

Ishak vs Cell Density (ICI) [aggregated] — Kendall tau-b (page 2/4)
Kendall tau-b: tie-corrected ordinal correlation | (1|patient_id) not shown
Cholangiocytes
Endothelium
Hepatocytes
Stroma (Other)
Ishak Confluent Necrosis
Ishak Lobular
Ishak Periportal
Ishak Portal
$\tau_b = -0.34$ $p = 0.0026$
$\tau_b = -0.28$ $p = 0.011$
$\tau_b = -0.13$ $p = 0.25$
$\tau_b = -0.26$ $p = 0.018$
$\tau_b = 0.06$ $p = 0.59$
$\tau_b = -0.25$ $p = 0.024$
$\tau_b = -0.25$ $p = 0.026$
$\tau_b = -0.18$ $p = 0.1$
$\tau_b = 0.12$ $p = 0.25$
$\tau_b = -0.05$ $p = 0.65$
$\tau_b = -0.04$ $p = 0.73$
$\tau_b = 0.03$ $p = 0.78$
$\tau_b = -0.16$ $p = 0.16$
$\tau_b = -0.06$ $p = 0.6$
$\tau_b = -0.1$ $p = 0.36$
$\tau_b = -0.23$ $p = 0.036$
Ishak Score
Cell Density (cells/mm$^2$)
therapy
Monotherapy
Combination_Therapy

**Ishak vs Cell Density (ICI) [aggregated] — Kendall tau-b (page 3/4)**

Kendall tau-b: tie-corrected ordinal correlation | (1|patient_id) not shown

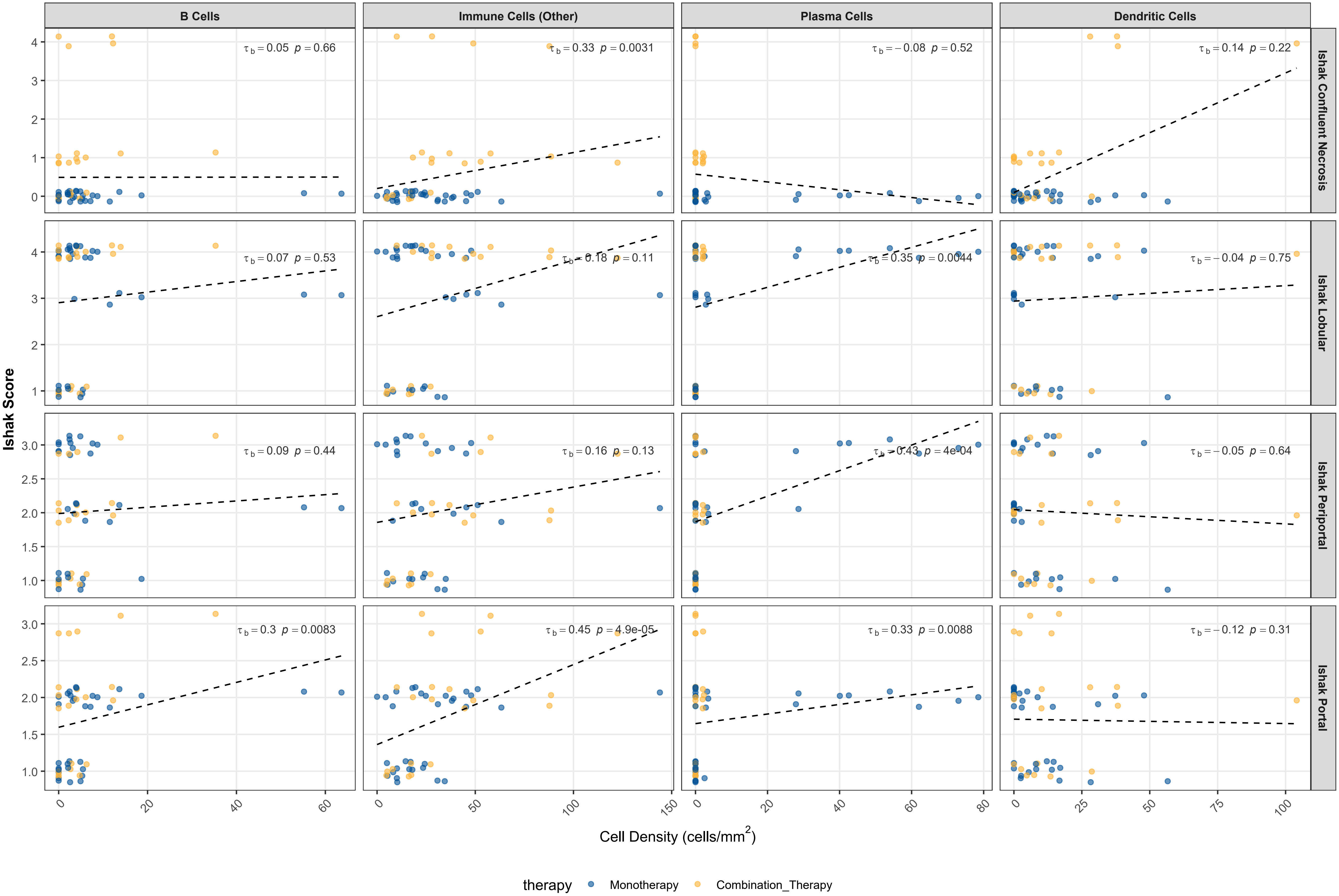

#### Ishak vs Cell Density (ICI) [aggregated] — Kendall tau-b (page 4/4)

Kendall tau-b: tie-corrected ordinal correlation | (1|patient_id) not shown

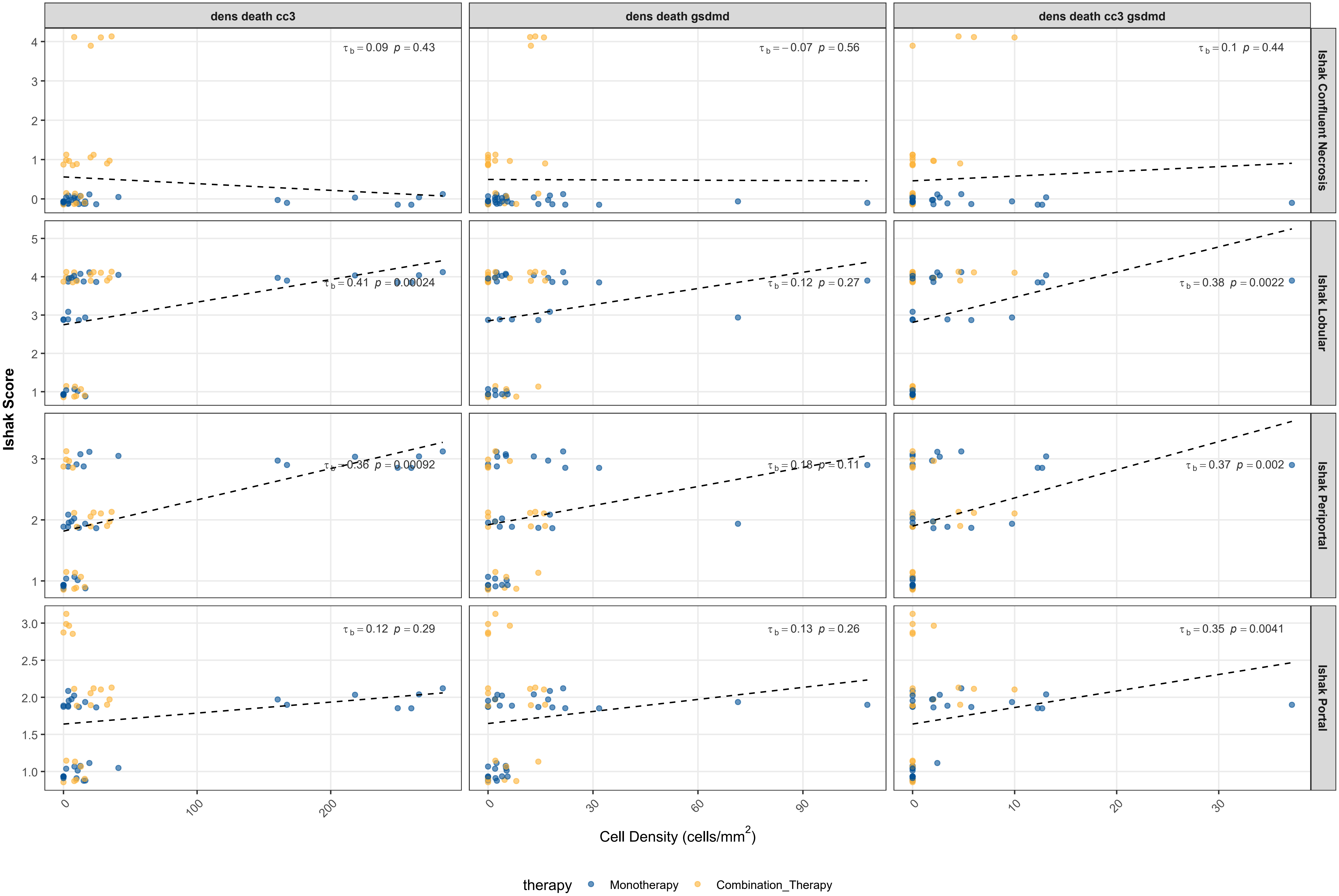

| Ishak | Spatial Feature | Kendall tau-b | p-value | Sig | n |
|---|---|---|---|---|---|
| Ishak Confluent Necrosis | CD8+ T Cells | 0.454 | 4.62e-05 | *** | 53 |
| Ishak Confluent Necrosis | Cholangiocytes | -0.337 | 2.63e-03 | ** | 53 |
| Ishak Confluent Necrosis | Immune Cells (Other) | 0.329 | 3.12e-03 | ** | 53 |
| Ishak Confluent Necrosis | Endothelium | -0.285 | 1.06e-02 | * | 53 |
| Ishak Confluent Necrosis | MΦ/Myeloid | 0.276 | 1.33e-02 | * | 53 |
| Ishak Confluent Necrosis | Stroma (Other) | -0.263 | 1.84e-02 | * | 53 |
| Ishak Confluent Necrosis | CD4+ T Cells | 0.207 | 6.33e-02 | | 53 |
| Ishak Confluent Necrosis | Dendritic Cells | 0.142 | 2.17e-01 | | 53 |
| Ishak Confluent Necrosis | Hepatocytes | -0.129 | 2.47e-01 | | 53 |
| Ishak Confluent Necrosis | dens death cc3 | 0.088 | 4.35e-01 | | 53 |
| Ishak Confluent Necrosis | dens death cc3 gsdmd | 0.097 | 4.35e-01 | | 53 |
| Ishak Confluent Necrosis | Plasma Cells | -0.080 | 5.23e-01 | | 53 |
| Ishak Confluent Necrosis | dens death gsdmd | -0.068 | 5.57e-01 | | 53 |
| Ishak Confluent Necrosis | B Cells | 0.050 | 6.64e-01 | | 53 |
| Ishak Confluent Necrosis | dens death any | 0.045 | 6.89e-01 | | 53 |
| Ishak Lobular | CD8+ T Cells | 0.504 | 4.74e-06 | *** | 53 |
| Ishak Lobular | MΦ/Myeloid | 0.423 | 1.23e-04 | *** | 53 |
| Ishak Lobular | dens death cc3 | 0.409 | 2.39e-04 | *** | 53 |
| Ishak Lobular | dens death cc3 gsdmd | 0.375 | 2.24e-03 | ** | 53 |
| Ishak Lobular | dens death any | 0.331 | 2.78e-03 | ** | 53 |
| Ishak Lobular | Plasma Cells | 0.353 | 4.41e-03 | ** | 53 |
| Ishak Lobular | Endothelium | -0.249 | 2.37e-02 | * | 53 |
| Ishak Lobular | Hepatocytes | -0.245 | 2.60e-02 | * | 53 |
| Ishak Lobular | Stroma (Other) | -0.180 | 1.03e-01 | | 53 |
| Ishak Lobular | Immune Cells (Other) | 0.178 | 1.07e-01 | | 53 |
| Ishak Lobular | CD4+ T Cells | 0.172 | 1.19e-01 | | 53 |
| Ishak Lobular | dens death gsdmd | 0.124 | 2.74e-01 | | 53 |
| Ishak Lobular | B Cells | 0.070 | 5.35e-01 | | 53 |
| Ishak Lobular | Cholangiocytes | 0.059 | 5.93e-01 | | 53 |
| Ishak Lobular | Dendritic Cells | -0.036 | 7.49e-01 | | 53 |
| Ishak Periportal | CD8+ T Cells | 0.419 | 1.02e-04 | *** | 53 |
| Ishak Periportal | MΦ/Myeloid | 0.403 | 1.85e-04 | *** | 53 |
| Ishak Periportal | Plasma Cells | 0.429 | 4.03e-04 | *** | 53 |
| Ishak Periportal | dens death cc3 | 0.361 | 9.20e-04 | *** | 53 |
| Ishak Periportal | dens death cc3 gsdmd | 0.372 | 1.98e-03 | ** | 53 |
| Ishak Periportal | dens death any | 0.322 | 2.96e-03 | ** | 53 |
| Ishak Periportal | dens death gsdmd | 0.176 | 1.14e-01 | | 53 |

### Ishak x Cell Density: Kendall tau-b [aggregated] (2/2)

FOV-level | tau-b corrects for tied ordinal values

| Ishak | Spatial Feature | Kendall tau-b | p-value | Sig | n |
|---|---|---|---|---|---|
| Ishak Periportal | B Cells | 0.085 | 4.43e-01 | | 53 |
| Ishak Periportal | Dendritic Cells | -0.052 | 6.43e-01 | | 53 |
| Ishak Periportal | Endothelium | -0.049 | 6.48e-01 | | 53 |
| Ishak Periportal | Hepatocytes | -0.037 | 7.32e-01 | | 53 |
| Ishak Periportal | Stroma (Other) | 0.030 | 7.81e-01 | | 53 |
| Ishak Portal | CD8+ T Cells | 0.486 | 1.07e-05 | *** | 53 |
| Ishak Portal | Immune Cells (Other) | 0.449 | 4.93e-05 | *** | 53 |
| Ishak Portal | MΦ/Myeloid | 0.407 | 2.32e-04 | *** | 53 |
| Ishak Portal | CD4+ T Cells | 0.392 | 3.94e-04 | *** | 53 |
| Ishak Portal | dens death cc3 gsdmd | 0.353 | 4.09e-03 | ** | 53 |
| Ishak Portal | B Cells | 0.300 | 8.32e-03 | ** | 53 |
| Ishak Portal | Plasma Cells | 0.326 | 8.77e-03 | ** | 53 |
| Ishak Portal | Stroma (Other) | -0.232 | 3.55e-02 | * | 53 |
| Ishak Portal | Cholangiocytes | -0.156 | 1.59e-01 | | 53 |
| Ishak Portal | dens death any | 0.126 | 2.57e-01 | | 53 |
| Ishak Portal | dens death gsdmd | 0.129 | 2.58e-01 | | 53 |
| Ishak Portal | dens death cc3 | 0.118 | 2.90e-01 | | 53 |
| Ishak Portal | Dendritic Cells | -0.115 | 3.13e-01 | | 53 |
| Ishak Portal | Hepatocytes | -0.101 | 3.59e-01 | | 53 |
| Ishak Portal | Endothelium | -0.058 | 6.01e-01 | | 53 |

### Ishak vs Cell Density (ICI) [detailed] — Kendall tau-b (page 1/7)

Kendall tau-b: tie-corrected ordinal correlation | (1|patient_id) not shown

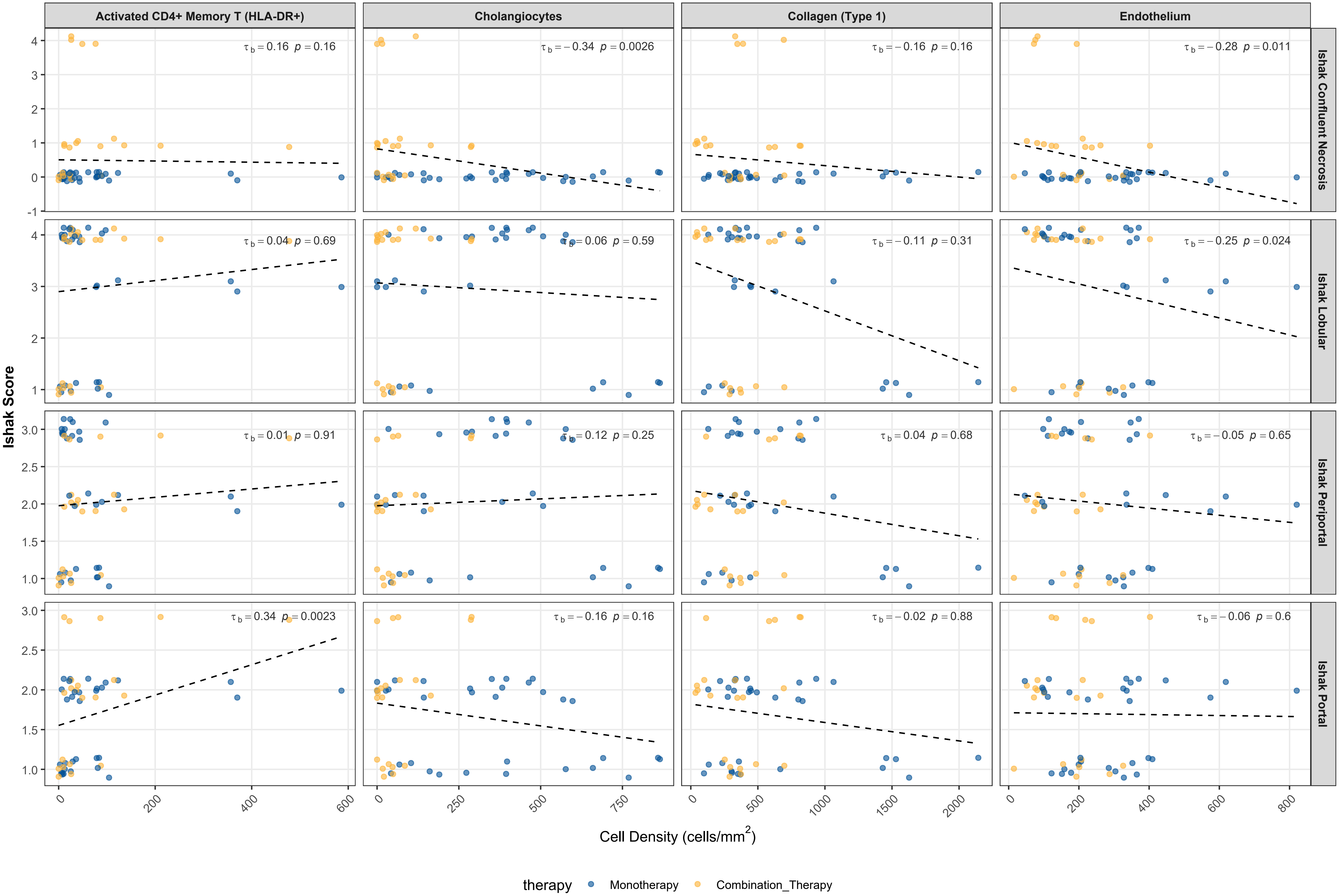

#### Ishak vs Cell Density (ICI) [detailed] — Kendall tau-b (page 2/7)

Kendall tau-b: tie-corrected ordinal correlation | (1|patient_id) not shown

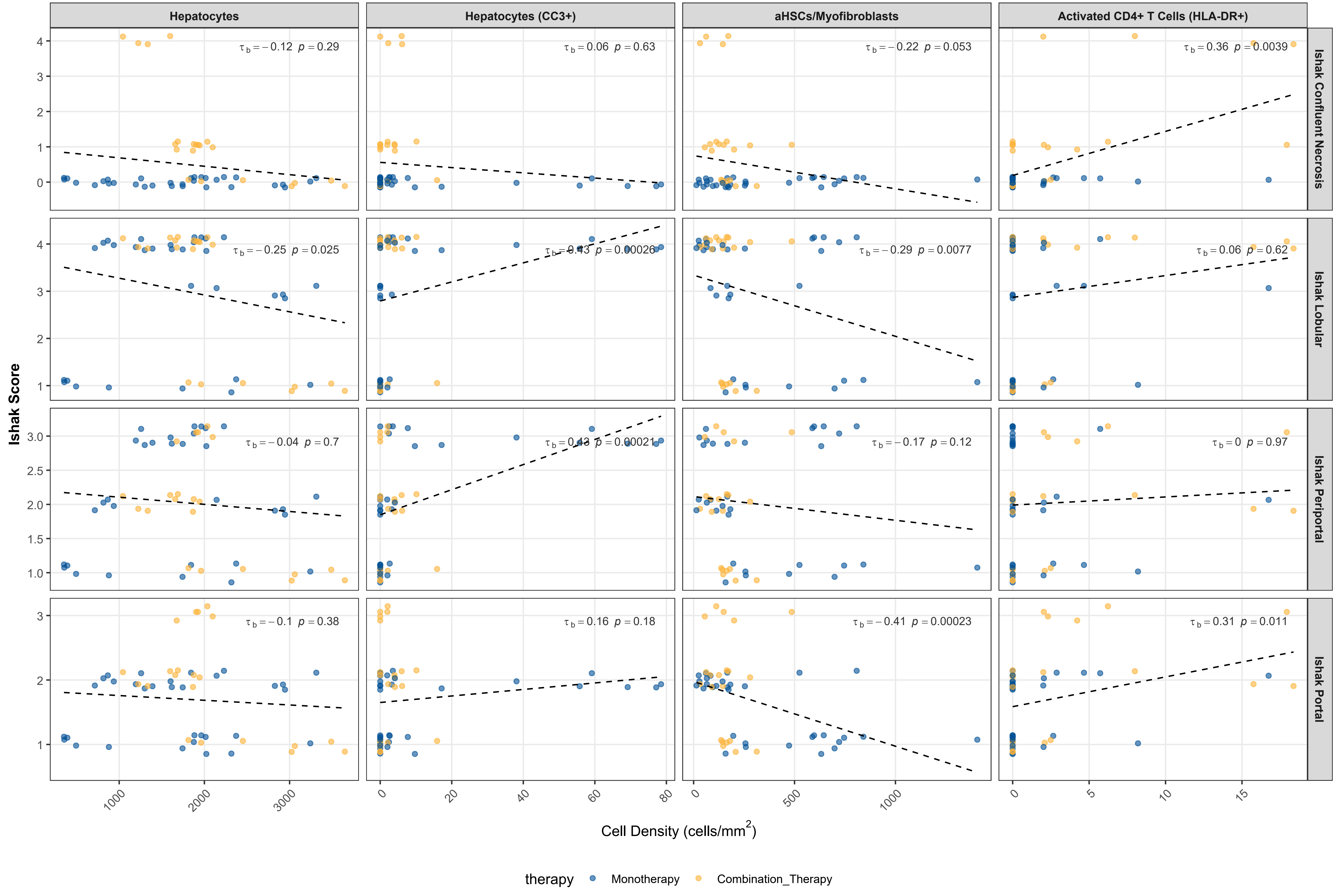

#### Ishak vs Cell Density (ICI) [detailed] — Kendall tau-b (page 3/7)

Kendall tau-b: tie-corrected ordinal correlation | (1|patient_id) not shown

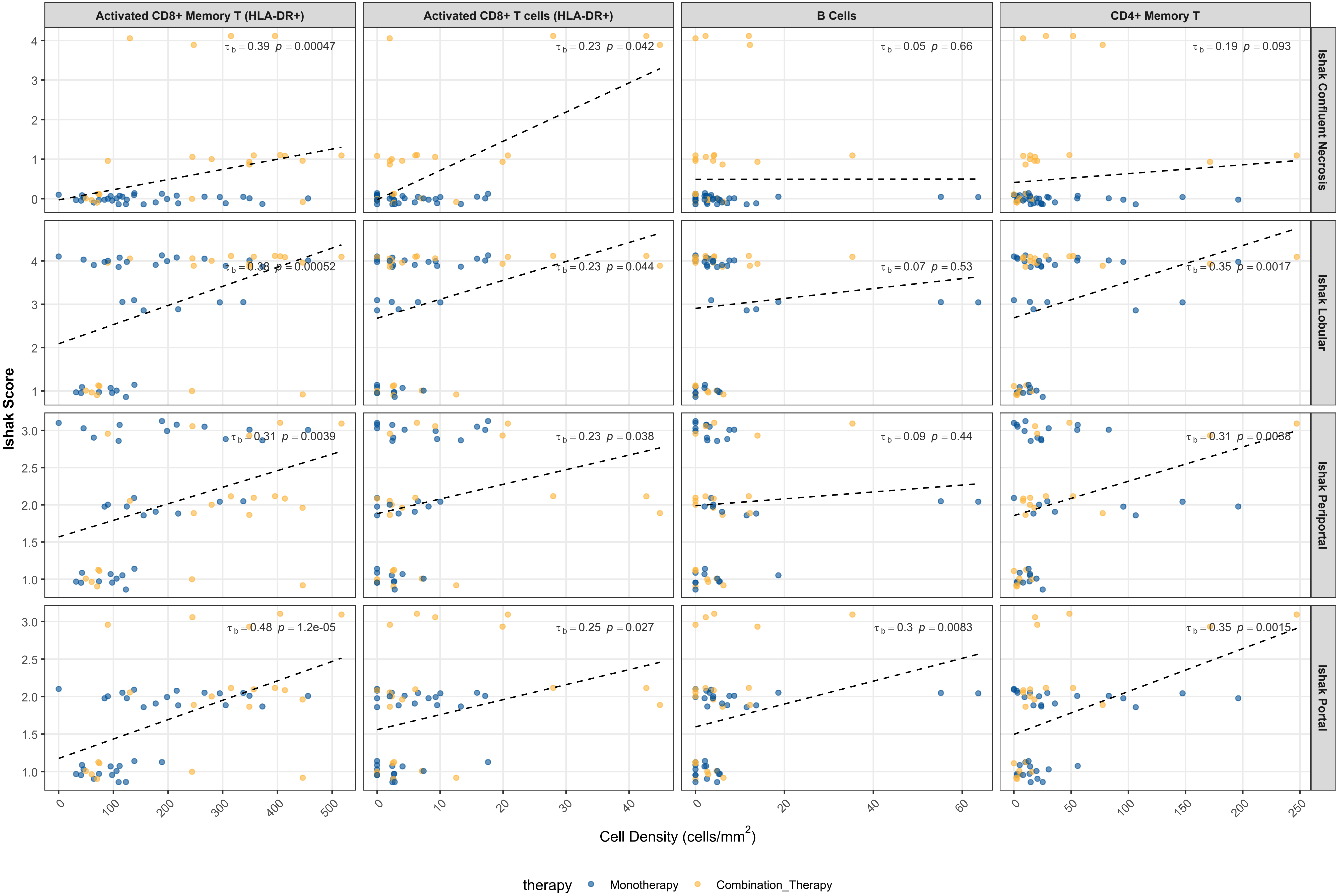

**Ishak vs Cell Density (ICI) [detailed] — Kendall tau-b (page 4/7)**

Kendall tau-b: tie-corrected ordinal correlation | (1|patient_id) not shown

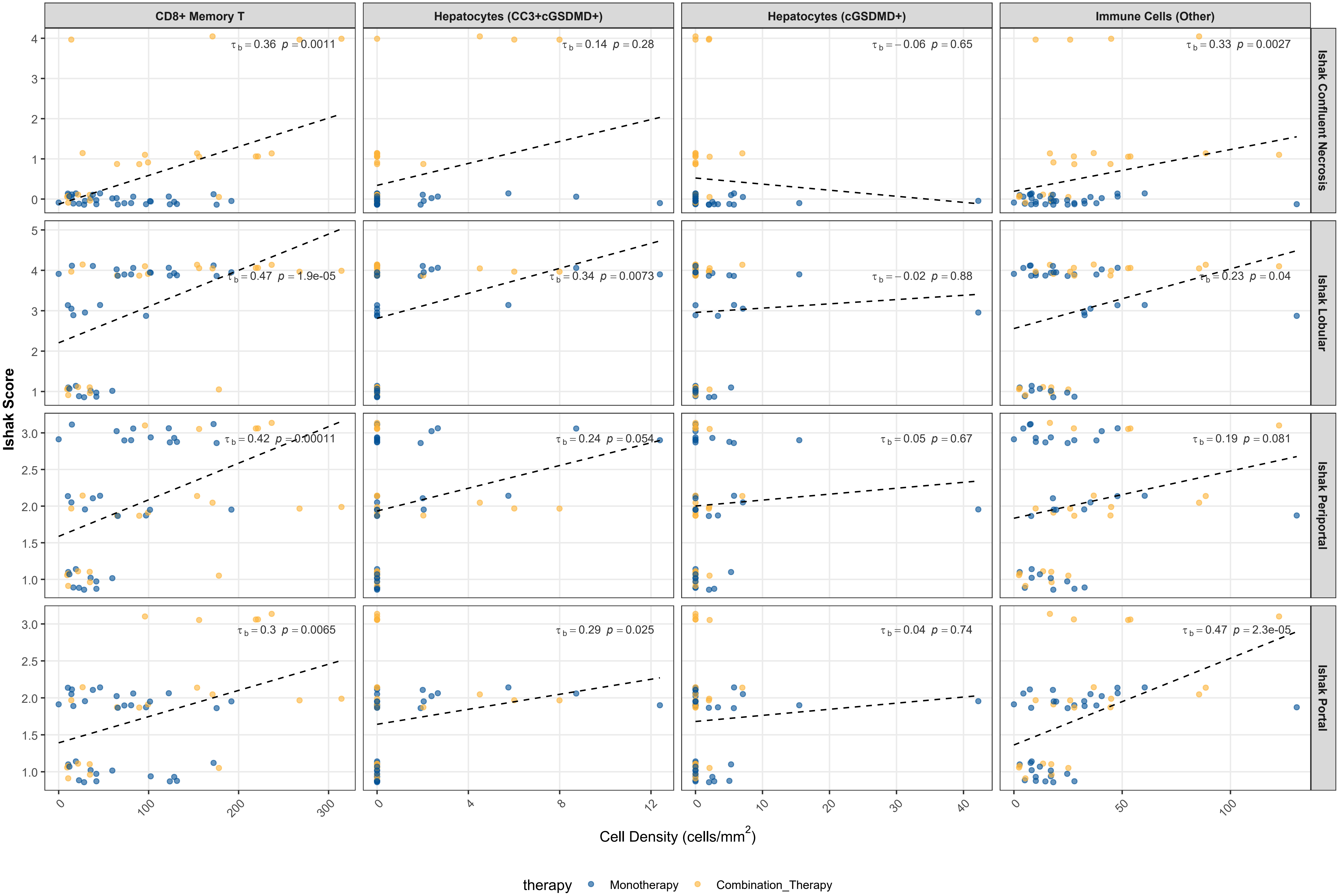

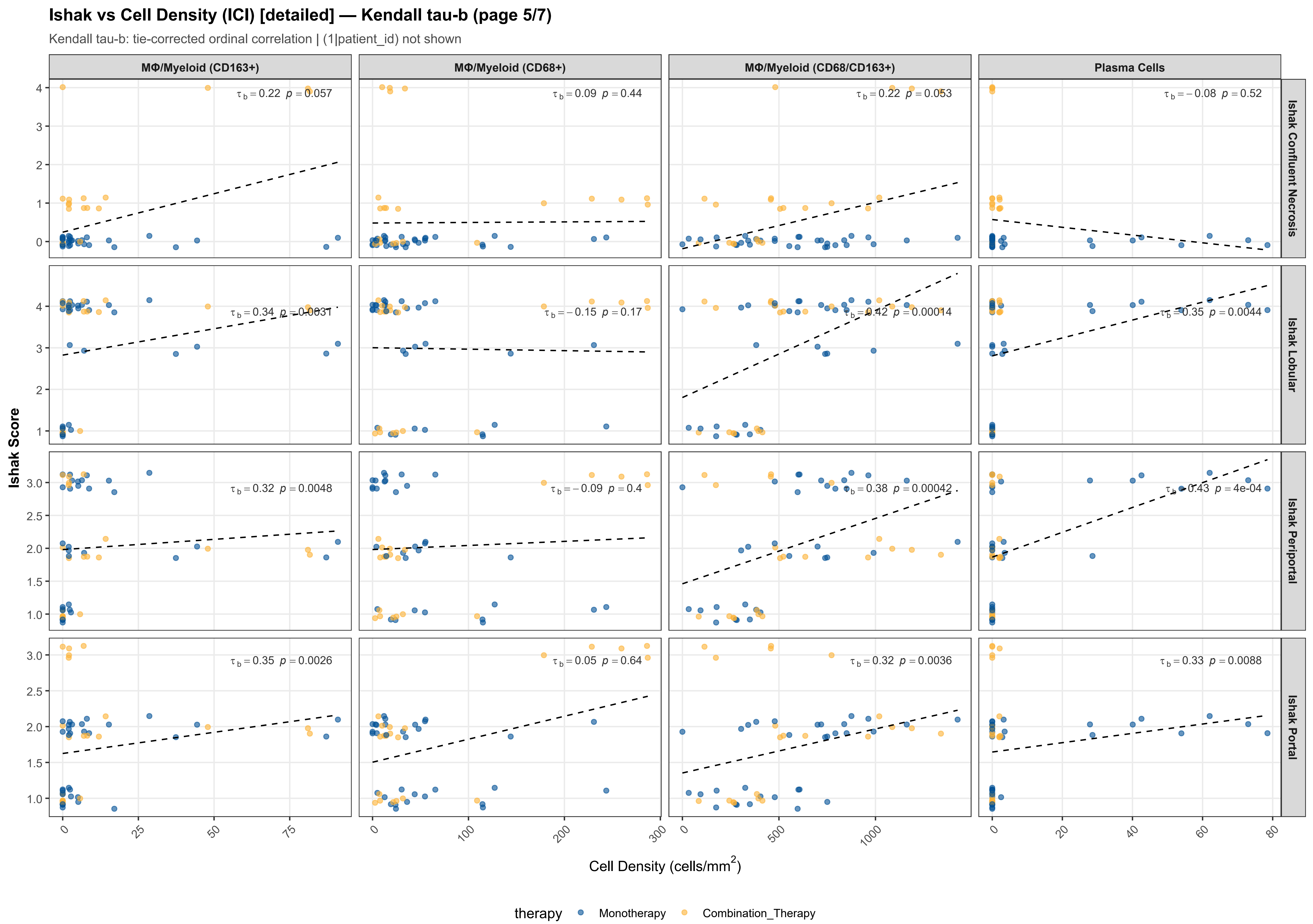

Ishak vs Cell Density (ICI) [detailed] — Kendall tau-b (page 5/7)
Kendall tau-b: tie-corrected ordinal correlation | (1|patient_id) not shown
MΦ/Myeloid (CD163+)
MΦ/Myeloid (CD68+)
MΦ/Myeloid (CD68/CD163+)
Plasma Cells
Ishak Confluent Necrosis
Ishak Lobular
Ishak Periportal
Ishak Portal
$\tau_b = 0.22$ $p = 0.057$
$\tau_b = 0.09$ $p = 0.44$
$\tau_b = 0.22$ $p = 0.053$
$\tau_b = -0.08$ $p = 0.52$
$\tau_b = 0.34$ $p = 0.0031$
$\tau_b = -0.15$ $p = 0.17$
$\tau_b = 0.42$ $p = 0.00014$
$\tau_b = 0.35$ $p = 0.0044$
$\tau_b = 0.32$ $p = 0.0048$
$\tau_b = -0.09$ $p = 0.4$
$\tau_b = 0.38$ $p = 0.00042$
$\tau_b = 0.43$ $p = 4e-04$
$\tau_b = 0.35$ $p = 0.0026$
$\tau_b = 0.05$ $p = 0.64$
$\tau_b = 0.32$ $p = 0.0036$
$\tau_b = 0.33$ $p = 0.0088$
Ishak Score
Cell Density (cells/mm$^2$)
therapy
Monotherapy
Combination_Therapy

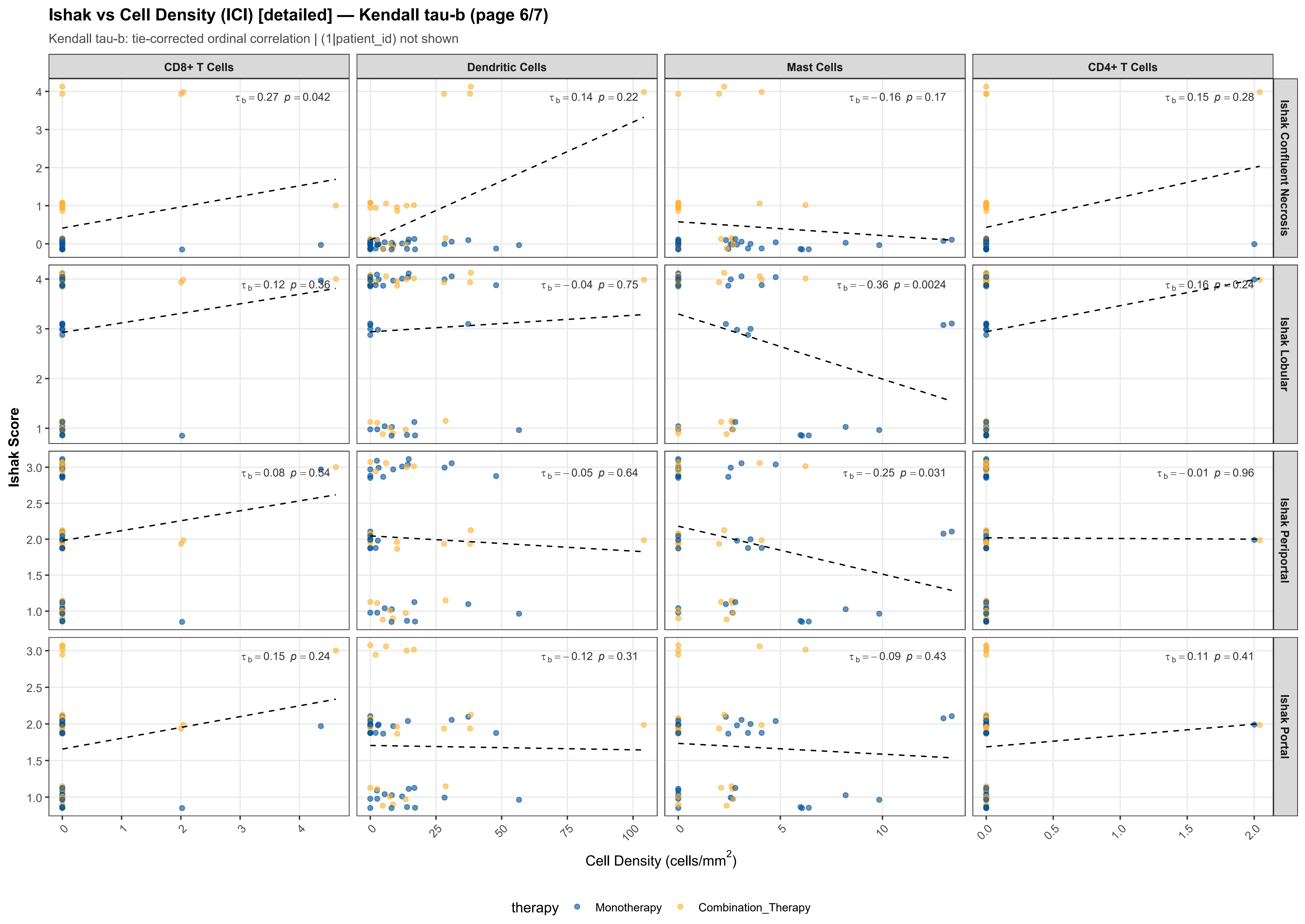

Ishak vs Cell Density (ICI) [detailed] — Kendall tau-b (page 6/7)
Kendall tau-b: tie-corrected ordinal correlation | (1|patient_id) not shown
CD8+ T Cells
Dendritic Cells
Mast Cells
CD4+ T Cells
Ishak Confluent Necrosis
Ishak Lobular
Ishak Periportal
Ishak Portal
τb = 0.27 p = 0.042
τb = 0.14 p = 0.22
τb = −0.16 p = 0.17
τb = 0.15 p = 0.28
τb = 0.12 p = 0.36
τb = −0.04 p = 0.75
τb = −0.36 p = 0.0024
τb = 0.16 p = 0.24
τb = 0.08 p = 0.54
τb = −0.05 p = 0.64
τb = −0.25 p = 0.031
τb = −0.01 p = 0.96
τb = 0.15 p = 0.24
τb = −0.12 p = 0.31
τb = −0.09 p = 0.43
τb = 0.11 p = 0.41
Ishak Score
Cell Density (cells/mm2)
therapy
Monotherapy
Combination_Therapy

#### Ishak vs Cell Density (ICI) [detailed] — Kendall tau-b (page 7/7)

Kendall tau-b: tie-corrected ordinal correlation | (1|patient_id) not shown

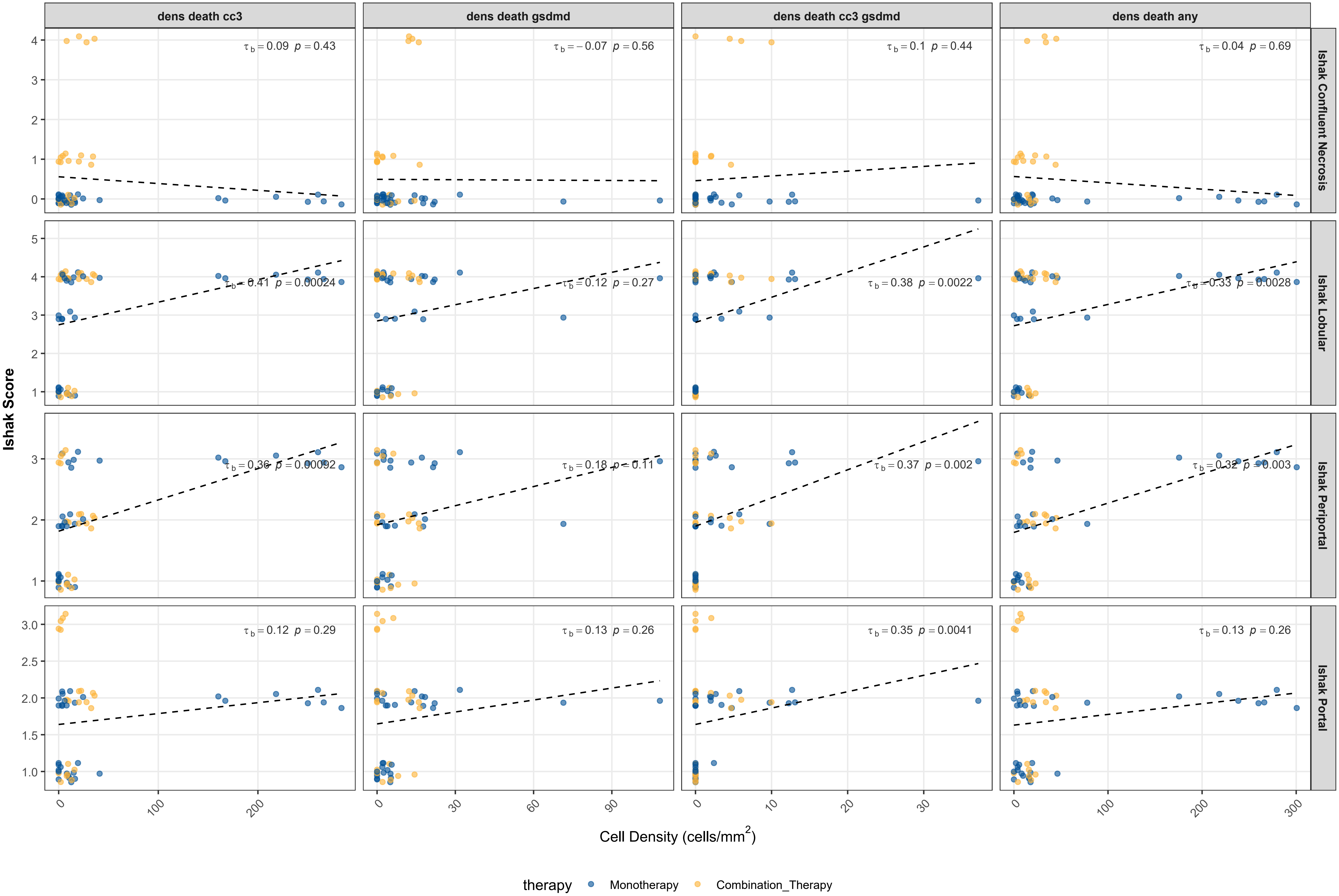

| Ishak | Spatial Feature | Kendall tau-b | p-value | Sig | n |
|---|---|---|---|---|---|
| Ishak Confluent Necrosis | Activated CD8+ Memory T (HLA-DR+) | 0.389 | 4.74e-04 | *** | 53 |
| Ishak Confluent Necrosis | CD8+ Memory T | 0.363 | 1.13e-03 | ** | 53 |
| Ishak Confluent Necrosis | Cholangiocytes | -0.337 | 2.63e-03 | ** | 53 |
| Ishak Confluent Necrosis | Immune Cells (Other) | 0.334 | 2.74e-03 | ** | 53 |
| Ishak Confluent Necrosis | Activated CD4+ T Cells (HLA-DR+) | 0.357 | 3.85e-03 | ** | 53 |
| Ishak Confluent Necrosis | Endothelium | -0.285 | 1.06e-02 | * | 53 |
| Ishak Confluent Necrosis | Activated CD8+ T cells (HLA-DR+) | 0.231 | 4.21e-02 | * | 53 |
| Ishak Confluent Necrosis | CD8+ T Cells | 0.269 | 4.25e-02 | * | 53 |
| Ishak Confluent Necrosis | aHSCs/Myofibroblasts | -0.216 | 5.27e-02 | | 53 |
| Ishak Confluent Necrosis | MΦ/Myeloid (CD68/CD163+) | 0.216 | 5.27e-02 | | 53 |
| Ishak Confluent Necrosis | MΦ/Myeloid (CD163+) | 0.221 | 5.68e-02 | | 53 |
| Ishak Confluent Necrosis | CD4+ Memory T | 0.187 | 9.34e-02 | | 53 |
| Ishak Confluent Necrosis | Activated CD4+ Memory T (HLA-DR+) | 0.158 | 1.56e-01 | | 53 |
| Ishak Confluent Necrosis | Collagen (Type 1) | -0.158 | 1.56e-01 | | 53 |
| Ishak Confluent Necrosis | Mast Cells | -0.164 | 1.70e-01 | | 53 |
| Ishak Confluent Necrosis | Dendritic Cells | 0.142 | 2.17e-01 | | 53 |
| Ishak Confluent Necrosis | CD4+ T Cells | 0.147 | 2.76e-01 | | 53 |
| Ishak Confluent Necrosis | Hepatocytes (CC3+cGSDMD+) | 0.138 | 2.82e-01 | | 53 |
| Ishak Confluent Necrosis | Hepatocytes | -0.118 | 2.90e-01 | | 53 |
| Ishak Confluent Necrosis | dens death cc3 | 0.088 | 4.35e-01 | | 53 |
| Ishak Confluent Necrosis | dens death cc3 gsdmd | 0.097 | 4.35e-01 | | 53 |
| Ishak Confluent Necrosis | MΦ/Myeloid (CD68+) | 0.087 | 4.36e-01 | | 53 |
| Ishak Confluent Necrosis | Plasma Cells | -0.080 | 5.23e-01 | | 53 |
| Ishak Confluent Necrosis | dens death gsdmd | -0.068 | 5.57e-01 | | 53 |
| Ishak Confluent Necrosis | Hepatocytes (CC3+) | 0.058 | 6.27e-01 | | 53 |
| Ishak Confluent Necrosis | Hepatocytes (cGSDMD+) | -0.057 | 6.47e-01 | | 53 |
| Ishak Confluent Necrosis | B Cells | 0.050 | 6.64e-01 | | 53 |
| Ishak Confluent Necrosis | dens death any | 0.045 | 6.89e-01 | | 53 |
| Ishak Lobular | CD8+ Memory T | 0.471 | 1.89e-05 | *** | 53 |
| Ishak Lobular | MΦ/Myeloid (CD68/CD163+) | 0.419 | 1.42e-04 | *** | 53 |
| Ishak Lobular | dens death cc3 | 0.409 | 2.39e-04 | *** | 53 |
| Ishak Lobular | Hepatocytes (CC3+) | 0.430 | 2.61e-04 | *** | 53 |
| Ishak Lobular | Activated CD8+ Memory T (HLA-DR+) | 0.382 | 5.17e-04 | *** | 53 |
| Ishak Lobular | CD4+ Memory T | 0.345 | 1.75e-03 | ** | 53 |
| Ishak Lobular | dens death cc3 gsdmd | 0.375 | 2.24e-03 | ** | 53 |
| Ishak Lobular | Mast Cells | -0.358 | 2.37e-03 | ** | 53 |
| Ishak Lobular | dens death any | 0.331 | 2.78e-03 | ** | 53 |

| Ishak | Spatial Feature | Kendall tau-b | p-value | Sig | n |
|---|---|---|---|---|---|
| Ishak Lobular | aHSCs/Myofibroblasts | -0.294 | 0.007700 | ** | 53 |
| Ishak Lobular | Endothelium | -0.249 | 0.023700 | * | 53 |
| Ishak Lobular | Hepatocytes | -0.247 | 0.024800 | * | 53 |
| Ishak Lobular | Immune Cells (Other) | 0.226 | 0.040200 | * | 53 |
| Ishak Lobular | Activated CD8+ T cells (HLA-DR+) | 0.227 | 0.044000 | * | 53 |
| Ishak Lobular | MΦ/Myeloid (CD68+) | -0.153 | 0.166000 | | 53 |
| Ishak Lobular | CD4+ T Cells | 0.155 | 0.242000 | | 53 |
| Ishak Lobular | dens death gsdmd | 0.124 | 0.274000 | | 53 |
| Ishak Lobular | Collagen (Type 1) | -0.112 | 0.309000 | | 53 |
| Ishak Lobular | CD8+ T Cells | 0.120 | 0.359000 | | 53 |
| Ishak Lobular | B Cells | 0.070 | 0.535000 | | 53 |
| Ishak Lobular | Cholangiocytes | 0.059 | 0.593000 | | 53 |
| Ishak Lobular | Activated CD4+ T Cells (HLA-DR+) | 0.060 | 0.622000 | | 53 |
| Ishak Lobular | Activated CD4+ Memory T (HLA-DR+) | 0.044 | 0.687000 | | 53 |
| Ishak Lobular | Dendritic Cells | -0.036 | 0.749000 | | 53 |
| Ishak Lobular | Hepatocytes (cGSDMD+) | -0.018 | 0.882000 | | 53 |
| Ishak Periportal | CD8+ Memory T | 0.417 | 0.000109 | *** | 53 |
| Ishak Periportal | Hepatocytes (CC3+) | 0.428 | 0.000206 | *** | 53 |
| Ishak Periportal | Plasma Cells | 0.429 | 0.000403 | *** | 53 |
| Ishak Periportal | MΦ/Myeloid (CD68/CD163+) | 0.380 | 0.000422 | *** | 53 |
| Ishak Periportal | dens death cc3 | 0.361 | 0.000920 | *** | 53 |
| Ishak Periportal | dens death cc3 gsdmd | 0.372 | 0.001980 | ** | 53 |
| Ishak Periportal | dens death any | 0.322 | 0.002960 | ** | 53 |
| Ishak Periportal | CD4+ Memory T | 0.313 | 0.003760 | ** | 53 |
| Ishak Periportal | Activated CD8+ Memory T (HLA-DR+) | 0.312 | 0.003860 | ** | 53 |
| Ishak Periportal | MΦ/Myeloid (CD163+) | 0.317 | 0.004750 | ** | 53 |
| Ishak Periportal | Mast Cells | -0.249 | 0.031000 | * | 53 |
| Ishak Periportal | Activated CD8+ T cells (HLA-DR+) | 0.228 | 0.038200 | * | 53 |
| Ishak Periportal | Hepatocytes (CC3+cGSDMD+) | 0.239 | 0.053800 | | 53 |
| Ishak Periportal | Immune Cells (Other) | 0.188 | 0.080700 | | 53 |
| Ishak Periportal | dens death gsdmd | 0.176 | 0.114000 | | 53 |
| Ishak Periportal | aHSCs/Myofibroblasts | -0.166 | 0.125000 | | 53 |
| Ishak Periportal | Cholangiocytes | 0.125 | 0.249000 | | 53 |
| Ishak Periportal | MΦ/Myeloid (CD68+) | -0.092 | 0.396000 | | 53 |
| Ishak Periportal | B Cells | 0.085 | 0.443000 | | 53 |
| Ishak Periportal | CD8+ T Cells | 0.079 | 0.540000 | | 53 |
| Ishak Periportal | Dendritic Cells | -0.052 | 0.643000 | | 53 |

### Ishak x Cell Density: Kendall tau-b [detailed] (3/3)

FOV-level | tau-b corrects for tied ordinal values

| Ishak | Spatial Feature | Kendall tau-b | p-value | Sig | n |
|---|---|---|---|---|---|
| Ishak Periportal | Hepatocytes | -0.042 | 6.95e-01 | | 53 |
| Ishak Periportal | Activated CD4+ Memory T (HLA-DR+) | 0.012 | 9.09e-01 | | 53 |
| Ishak Periportal | CD4+ T Cells | -0.006 | 9.61e-01 | | 53 |
| Ishak Periportal | Activated CD4+ T Cells (HLA-DR+) | 0.004 | 9.70e-01 | | 53 |
| Ishak Portal | Activated CD8+ Memory T (HLA-DR+) | 0.485 | 1.16e-05 | *** | 53 |
| Ishak Portal | Immune Cells (Other) | 0.468 | 2.33e-05 | *** | 53 |
| Ishak Portal | aHSCs/Myofibroblasts | -0.407 | 2.32e-04 | *** | 53 |
| Ishak Portal | CD4+ Memory T | 0.350 | 1.54e-03 | ** | 53 |
| Ishak Portal | Activated CD4+ Memory T (HLA-DR+) | 0.337 | 2.32e-03 | ** | 53 |
| Ishak Portal | MΦ/Myeloid (CD163+) | 0.346 | 2.60e-03 | ** | 53 |
| Ishak Portal | MΦ/Myeloid (CD68/CD163+) | 0.321 | 3.63e-03 | ** | 53 |
| Ishak Portal | dens death cc3 gsdmd | 0.353 | 4.09e-03 | ** | 53 |
| Ishak Portal | CD8+ Memory T | 0.301 | 6.53e-03 | ** | 53 |
| Ishak Portal | B Cells | 0.300 | 8.32e-03 | ** | 53 |
| Ishak Portal | Plasma Cells | 0.326 | 8.77e-03 | ** | 53 |
| Ishak Portal | Activated CD4+ T Cells (HLA-DR+) | 0.313 | 1.07e-02 | * | 53 |
| Ishak Portal | Hepatocytes (CC3+cGSDMD+) | 0.286 | 2.45e-02 | * | 53 |
| Ishak Portal | Activated CD8+ T cells (HLA-DR+) | 0.249 | 2.75e-02 | * | 53 |
| Ishak Portal | Cholangiocytes | -0.156 | 1.59e-01 | | 53 |
| Ishak Portal | Hepatocytes (CC3+) | 0.159 | 1.79e-01 | | 53 |
| Ishak Portal | CD8+ T Cells | 0.154 | 2.43e-01 | | 53 |
| Ishak Portal | dens death any | 0.126 | 2.57e-01 | | 53 |
| Ishak Portal | dens death gsdmd | 0.129 | 2.58e-01 | | 53 |
| Ishak Portal | dens death cc3 | 0.118 | 2.90e-01 | | 53 |
| Ishak Portal | Dendritic Cells | -0.115 | 3.13e-01 | | 53 |
| Ishak Portal | Hepatocytes | -0.098 | 3.77e-01 | | 53 |
| Ishak Portal | CD4+ T Cells | 0.111 | 4.05e-01 | | 53 |
| Ishak Portal | Mast Cells | -0.093 | 4.30e-01 | | 53 |
| Ishak Portal | Endothelium | -0.058 | 6.01e-01 | | 53 |
| Ishak Portal | MΦ/Myeloid (CD68+) | 0.052 | 6.37e-01 | | 53 |
| Ishak Portal | Hepatocytes (cGSDMD+) | 0.041 | 7.40e-01 | | 53 |
| Ishak Portal | Collagen (Type 1) | -0.016 | 8.84e-01 | | 53 |
